# Early Reduction of SARS-CoV-2 Replication in Bronchial Epithelium by Kinin B_2_ Receptor Antagonism

**DOI:** 10.1101/2021.08.13.21262037

**Authors:** Constanze A. Jakwerth, Martin Feuerherd, Ferdinand M. Guerth, Madlen Oelsner, Linda Schellhammer, Johanna Giglberger, Lisa Pechtold, Claudia Jerin, Luisa Kugler, Carolin Mogler, Bernhard Haller, Anna Erb, Barbara Wollenberg, Christoph D. Spinner, Thorsten Buch, Ulrike Protzer, Carsten B. Schmidt-Weber, Ulrich M. Zissler, Adam M. Chaker

## Abstract

**Background:** SARS-CoV2 has evolved to enter the host via the ACE2 receptor which is part of the Kinin-kallirein pathway. This complex pathway is only poorly understood in context of immune regulation but critical to control infection. This study examines SARS-CoV2 infection and epithelial mechanisms of the kinin-kallikrein system at the kinin B_2_ receptor level in SARS-CoV-2 infection that is of direct translational relevance.

**Methods:** From acute SARS-CoV-2-positive patients and -negative controls, transcriptomes of nasal brushings were analyzed. Primary airway epithelial cells (NHBEs) were infected with SARS-CoV-2 and treated with the approved B_2_R antagonist icatibant. SARS-CoV-2 RNA RT-qPCR, cytotoxicity assays, plaque assays and transcriptome analyses were performed. The treatment effect was further studied in a murine airway inflammation model *in vivo*.

**Results:** Here, we report a broad and strong upregulation of kallikreins and the kinin B_2_ receptor (B_2_R) in the nasal mucosa of acutely symptomatic SARS-CoV-2-positive patients. A B_2_R antagonist impeded SARS-CoV-2 replication and spread in NHBEs, as determined in plaque assays on Vero E6 cells. B_2_R antagonism reduced the expression of SARS-CoV-2 entry receptor ACE2 *in vitro* and in a murine airway inflammation model *in vivo*. In addition, it suppressed gene expression broadly, particularly genes involved in G-protein-coupled-receptor signaling and ion transport.

**Conclusions:** In summary, this study provides evidence that treatment with B_2_R antagonists protects airway epithelial cells from SARS-CoV-2 by inhibiting its replication and spread, through the reduction of ACE2 levels and the interference with several cellular signaling processes. Future clinical studies need to shed light on the airway protection potential of approved B_2_R antagonists, like icatibant, in the treatment of early-stage COVID-19.

## 1 INTRODUCTION

SARS-CoV-2 vaccines have been approved worldwide since the end of 2020 and are starting to show their protective effects in public health.^1,2^ Even with vaccines at hand, an important medical need for therapeutic approaches for COVID-19 remains: Parts of the population refrain from vaccination or are not eligible for it, immunocompromised individuals may not mount a sufficient immune response after vaccination and escape variants, such as the currently spreading SARS-CoV-2 variant Delta^3^, may breach the protection afforded by the vaccines.^4-9^ Key factors for SARS-CoV-2 cell entry are two cell surface molecules, the angiotensin-converting enzyme 2 (ACE2) and the transmembrane serine protease (TMPRSS)2.^10^ TMPRSS2 cleaves the coronaviral spike protein and primes it for cell fusion, while ACE2 enables the virus particle to enter the cell by binding of its spike protein.^11,12^ The latter acts as central component in its function as terminal carboxypeptidase in the counter-regulatory axis of the renin-angiotensin system (RAS) and the contact activation system (CAS),^10,13^ which initiates blood coagulation and can additionally activate the kinin-kallikrein system (KKS).^14^ In its role in the RAS, ACE2 has anti-vasoconstrictive and anti-inflammatory effects by hydrolyzing the vasoconstrictive and tissue-damaging angiotensin II, which contributes to airway remodeling and fibrosis,^15,16^ to angiotensin (1-7).^17^ In its role in the KKS, ACE2 further hydrolyzes vasoactive peptides such as des-Arg9-bradykinin (DABK), which activates the inflammatory axis of the KKS.^18,19^ While DABK is the ligand of the inducible kinin B_1_ receptor (*BDKRB1*; B_1_ receptor),^20^ bradykinin, the end product of the KKS cascade, binds to the constitutively expressed kinin B_2_ receptor (*BDKRB2;* B_2_ receptor; B_2_R) and activates it.^21^ Bradykinin mediates its pro-inflammatory effects via the B_2_ receptor, by eliciting a variety of responses, including vasodilation and edema, via the G protein-triggered phosphatidylinositol-calcium second messenger system.^22-26^

The fact that SARS-CoV-2 utilizes ACE2 to enter airway cells along with the fact that ACE2 is a multifunctional enzyme that counter-regulates the ACE-driven mechanisms of the RAS and balances the KKS, may therefore explain the serious course of COVID-19, not only in the lungs but systemically.^27,28^

Recent publications suggest that the KKS could play a role in COVID-19. KKS comes into play particularly in connection with the high prevalence of thromboembolic events in seriously ill COVID-19 patients.^7,18,20,29,30^ A recent study on a cohort of 66 COVID-19 patients admitted to the intensive care unit showed that the KKS was strongly activated, which was reflected in the consumption of factor XII, pre-kallikrein and high-molecular-weight kininogen (HMW-kininogen).^29^ It has further been hypothesized that kinin-dependent “local lung angioedema” involving the kinin B_1_ and B_2_ receptors is an important characteristic of COVID-19.^31-34^

This study examines the potential of interfering with the KKS at the kinin receptor level in SARS-CoV-2 infection with translational relevance. Based on a broad and strong upregulation of kallikreins and of the kinin B_2_ receptor in the nasal mucosa of acutely SARS-CoV-2-positive patients, we hypothesized that the B_2_ receptor antagonism using icatibant, an approved compound for the treatment of hereditary angioedema,^35^ had an early protective effect against SARS-CoV-2-mediated epithelial damage through disruption of ACE2-mediated virus entry.

## 2 MATERIALS AND METHODS

### 2.1 Patients and nasal brushings

The nasal brushings were performed as a part of a larger health professional observational cohort study, which was approved by the ethics commission of the Technical University of Munich (AZ 175/20s). Nasal brushings were subjected to whole-genome microarray transcriptome analysis (see Supp.Info.). All patients gave written, informed consent prior to participation (see Table 1).

**Table 1.**
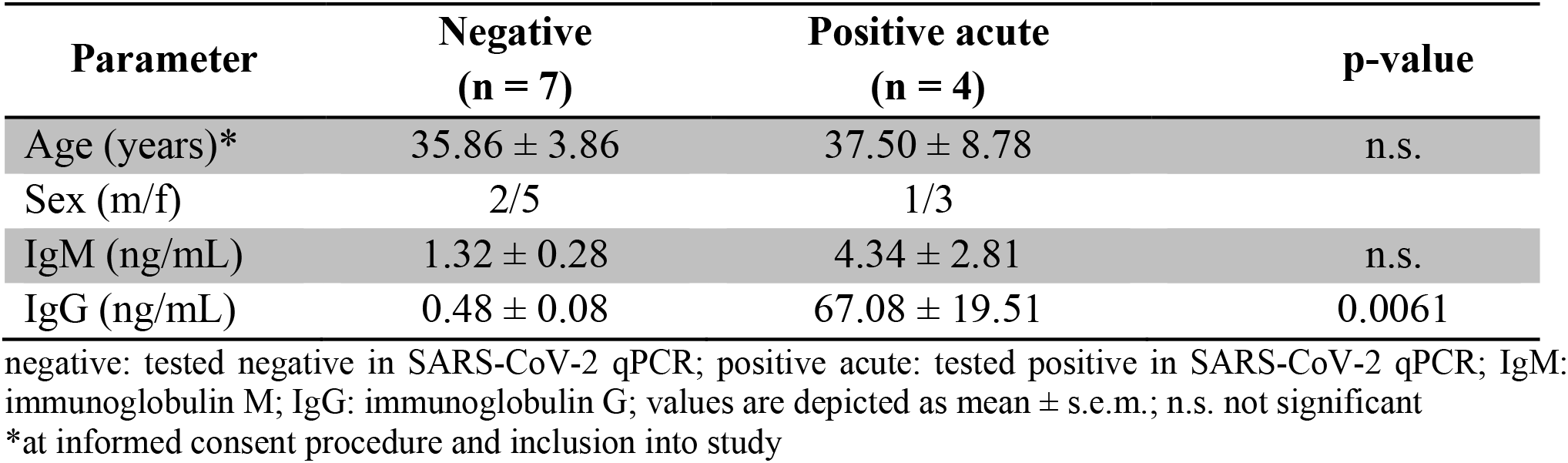
Demographic data of patient cohort

### 2.2 *In vivo* mouse study

Mice received murine IL-12Fc (1 μg of protein in 50 μL PBS) or PBS control intranasally.^36^ Intranasal application was performed under isoflurane anesthesia in two steps of 25 μL per nostril. 48 hours later, the mice received a single subcutaneous injection of icatibant (2 nmol per 10 g body weight; HOE-140 (icatibant), H157, SLBX4410, Sigma) or PBS control. The experiment was terminated by CO_2_ asphyxiation 6 hours or 24 hours after the injection of icatibant. The experiment was carried out twice Organs were snap frozen for protein extraction. Mice enrolled in the experiment were 6-8 weeks old, from either C57BL/6J, BALB/c or C3H HeN strains. Both sexes were included for each strain and means of each mouse type (strain / sex) are depicted as single values in Fig.2A: circle: female; triangle: male. Black: C57BL/6, mid grey: C3H HeN, light grey: BALB/c strain. Experiments were performed and analyzed in a randomized and blinded fashion. Animals were obtained from Janvier Labs (Le Genest-Saint-Isle, France) and housed 5 per cage and sex in individually ventilated cages at Laboratory Animal Service Center of the University of Zurich in Schlieren (Schlieren, Zurich, Switzerland). The animal vivarium was a specific-pathogen-free (SPF) holding room, that was temperature- and humidity-controlled (21 ± 3 °C, 50 ± 10%), with a 12h light/dark cycle. All animals had ad libitum access to the same food and water throughout the entire study. All procedures described in this study had previously been approved by the Cantonal Veterinarian’s Office of Zurich, Switzerland (License ZH096/20), and every effort was made to minimize the number of animals used and their suffering. Experiments were pre-registered at www.animalstudyregistry.org (study title “Effect of drug on ACE2 levels in mice”; doi=10.17590/asr.0000225).

Detailed methods are provided in the online supporting information.

## 3 RESULTS

### 3.1 B_2_ receptor antagonist inhibits replication and spread of SARS-CoV-2

The enzyme ACE2 is the central viral entry receptor for the novel SARS-CoV-2 coronavirus on human epithelial cells of the respiratory tract.^10^ Recent studies showed that this receptor and its co-receptors are not only expressed in the lower respiratory tract, and thus on the alveolar epithelial cells type I and II, but are also present in the upper respiratory tract, but are predominantly expressed in the nasal mucosa.^37^

To investigate the local effect of the acute SARS-CoV-2 infection on the nasal epithelium, we analyzed the transcriptome of nasal brushings from symptomatic patients who tested acutely positive for SARS-CoV-2 and of SARS-CoV-2-negative patients. In a transcriptome analysis, the most strongly induced genes encoding secreted factors included many members of the kallikrein family (Fig.1A, Table S1). In particular, the kallikreins *KLK5, KLK9* and *KLK12* were significantly upregulated in acute SARS-CoV-2-positive patients compared to SARS-CoV-2-negative patients (Fig.1B, Table S2). A subsequent analysis focusing on central factors of the KKS revealed that in addition, the kinin receptor B_2_ (*BDKRB2*; B_2_R) was also significantly increased (Fig.1C, Table S3). This first finding, together with our hypothesis that ACE2, as a central component, is counter-regulated by intervention in the KKS, prompted us to investigate selective kinin B_2_ receptor antagonism in connection with SARS-CoV-2 infection. To circumvent the limitations of cell lines like Vero E6, A549 or Calu-3 cells that are intrinsically impaired to form an interferon response upon viral infection,^38^ we next infected primary bronchial epithelial (NHBE) cells with SARS-CoV-2.

**Figure 1.**
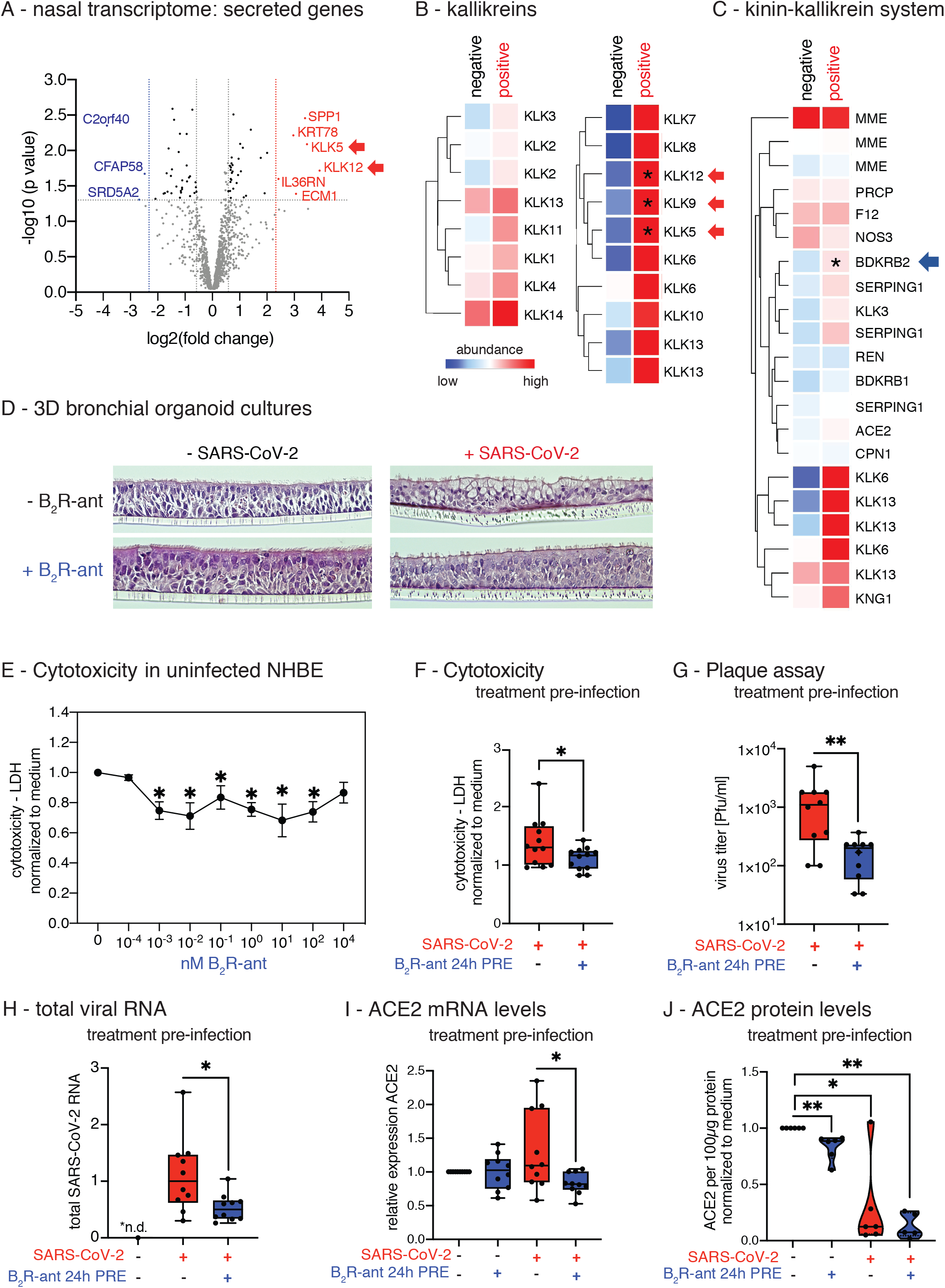
Induction of kallikreins and kinin receptor B_2_ in the nasal mucosa of acutely positive Covid-19 patients. A) Volcano plot of significantly differentially regulated genes (DEGs = differentially expressed genes) in nasal brushings of patients that were acute positive for SARS-CoV-2 compared to healthy individuals (negative) using human miR microarray technology. Highlighted genes have a fold-change (FC)ζ10 with P<0.05, genes in red are up-regulated, genes in blue are downregulated. B) Heat map of gene expression analysis of kallikrein genes and C) of genes of the kinin-kallikrein system (KKS) in nasal brushings comparing SARS-CoV-2 acute-positive patients to healthy controls. All entities with FCζ1.2 are shown. Asterisks indicate significantly regulated genes (P<0.05) in SARS-CoV-2-infected NHBEs compared to medium. Color code indicates Log2-fold change from low (blue) through 0 (white) to high (red). Duplicate gene names indicate the abundance of two or more isoforms of the same gene in the analysis. D) 3D-air-liquid interphase cultures from NHBEs were pre-treated for 24 hours with/without 1 nM B_2_R antagonist from the basal side and subsequently infected with SARS-CoV-2 for 48 hours from the apical side. E) Lactate dehydrogenase (LDH) cytotoxicity assay using the LDH Cytotoxicity Detection Kit PLUS studying the effect of increasing doses of the B_2_R antagonist after 48 hours in primary NHBEs from 4 donors. Results are depicted as mean ± s.e.m. Statistical tests compared each dose of B_2_R antagonist with 0 nM B_2_R antagonist. F) Cytotoxicity assay determining LDH release into the supernatants of cultures of SARS-CoV-2-infected NHBEs from 12 donors that were pre-treated for 24 hours with/without 1 nM B_2_R antagonist. G) Quantification of infectious particles in the supernatants of SARS-CoV-2-infected NHBEs from 10 donors that were pre-treated with/without 1 nM B_2_R antagonist for 24 hours. Supernatants were titrated on Vero E6 cells and plaque assay was quantified 24 hours later. Results are depicted as plaque forming units (PFU) per milliliter. H) qPCR analysis of total SARS-CoV-2 RNA (viral genome and transcripts, which all contain the N1 sequence region) normalized to human *ACTB* of SARS-CoV-2-infected primary NHBE after 24 hours of pre-treatment with/without 1 nM B_2_R antagonist followed by 24 hours of SARS-CoV-2 inoculation. For Figures 1E, F, and H, statistical tests compared SARS-CoV-2-infected versus uninfected samples or B_2_R antagonist-treated versus untreated samples. I) Analysis of human ACE2 gene expression using qPCR (n=10) and J) of human ACE2 protein levels analyzed by ELISA from cell lysates (n=6) after 24 hours of pre-treatment of NHBEs with/without 1 nM B_2_R antagonist, followed by SARS-CoV-2 inoculation for 24 hours.

**Figure 2.**
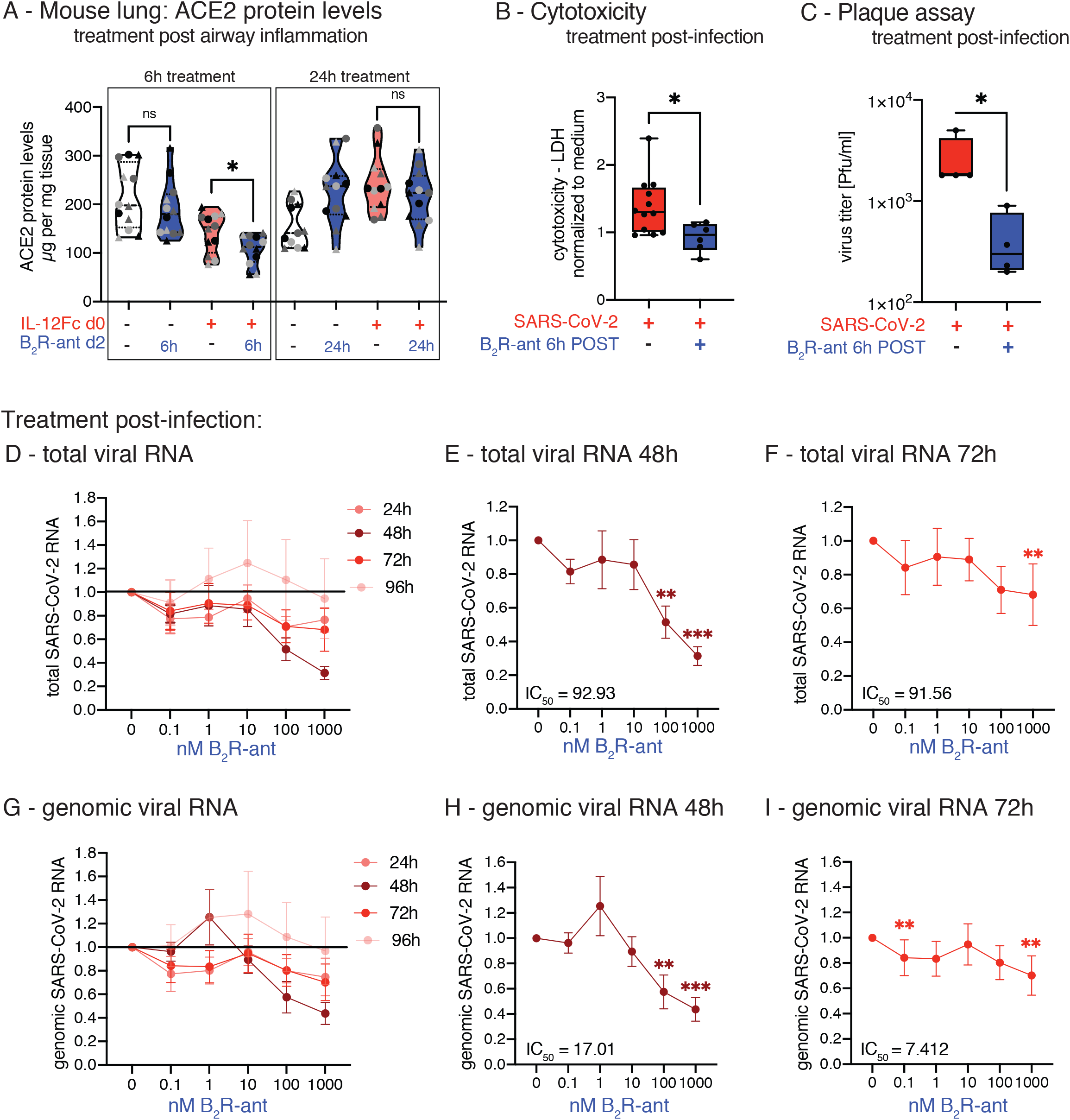
Treatment of NHBE with B_2_R antagonist post-infection in repeated doses inhibits SARS-CoV-2 replication. A) *In vivo* mouse study. 12 sex-matched mice from three different strains per group were treated on day 0 with intranasal application of 1 μg murine IL-12Fc per mouse or PBS as control to mimic virus-induced airway inflammation. After 48h, mice were injected s.c. with 2 nmol of the B_2_R antagonist icatibant per 10 g of body weight or PBS as control. The experiment was terminated either six hours or 24 hours later and murine lung ACE2 protein levels were analyzed by mouse ACE2 ELISA analysis. Circle: female; triangle: male. Black: C57BL/6, mid grey: C3H HeN, light grey: BALB/c strain. The experiment was carried out twice and the data in the figure represent the mean of each mouse type (strain / sex) of both experiments. Statistical tests compared B_2_R antagonist -treated versus untreated groups. B) Cytotoxicity assay determining LDH in supernatants from SARS-CoV-2-infected NHBEs from 12 donors that were treated with/without 1 nM B_2_R antagonist after 6 hours of infection for another 24 hours. C) Quantification of infectious particles in the supernatants from SARS-CoV-2-infected NHBEs from 4 donors that were treated with/without 1 nM B_2_R antagonist after 6 hours of infection for another 24 hours. Supernatants were titrated on Vero E6 cells and plaque assay was quantified 24 hours later. Results are depicted as plaque forming units (PFU) per milliliter. For Figures 2B-C, statistical tests compared B_2_R antagonist-treated versus untreated samples. D) Relative quantification of total SARS-CoV-2 RNA (viral genome and transcripts, which all contain the N1 sequence region) and G) genomic SARS-CoV-2 RNA (containing the *RdRP* gene) normalized to housekeeping gene index of human *ACTB, HPRT, 18S* in NHBEs from 8 independent donors that were infected with SARS-CoV-2 for 6 hours and then treated with increasing doses of the B_2_R antagonist icatibant repeatedly every 24 hours for a total of 96 hours. In cells treated with E) 100 nM and F) 1000 nM icatibant for 48 hours and with H) 100 nM and I) 1000 nM icatibant for 72 hours, total SARS-CoV-2 RNA and genomic SARS-CoV-2 RNA were significantly reduced. Red indicates SARS-CoV-2 infection; blue indicates B_2_R antagonist treatment. PRE indicates pre-treatment; POST indicates post-treatment. In Fig.2D-I, results are depicted as mean ± s.e.m. and statistical tests compared each dose of icatibant with 0 nM icatibant. Statistically significant differences were depicted as p-values *P<0.05, **P<0.01, and ***P<0.001. ns indicates non-significant. + infected / treated; - indicates not infected / not treated.

To examine the effects of SARS-CoV-2 infection and B_2_R antagonist treatment on the microscopic integrity of the airway epithelium, 3D-air-liquid interphase organoid cultures were differentiated from primary NHBEs (see Supp.Info). After complete differentiation, the epithelia were pre-treated from the basal (medium) side with the approved B_2_R antagonist icatibant, and subsequently infected with SARS-CoV-2 from the apical (air) side. The cultures pre-treated with the B_2_R antagonist showed less virus-induced balloon-like structures compared to untreated cultures. The epithelial layers remained qualitatively more intact, which indicated a protective effect of the B_2_R antagonist for the bronchial epithelium (Fig.1D). This finding was further strengthened by cytotoxicity assays: The B_2_R antagonist had no toxic effects on NHBEs even at high doses determined by lactate dehydrogenase (LDH) release, but rather exhibited a cell-protecting effect in uninfected cells (Fig.1E) as well as during SARS-CoV-2 infection (Fig.1F). Next, the supernatants of pre-treated, infected primary NHBEs were collected, titrated onto fresh Vero E6 cell cultures and plaque assays were performed. Strikingly, we found that *in vitro* treatment of NHBEs with the B_2_R antagonist prior to infection reduced the number of plaque forming units (PFU) in a plaque assay by 87% (Fig.1G). In addition, the levels of total SARS-CoV-2 RNA in cells that had been pretreated with the B_2_R antagonist decreased by 52% compared to untreated infected NHBEs (Fig.1H). With regard to the virus entry process, ACE2 was reduced by pretreatment with the B_2_R antagonist on the mRNA level (Fig. 1I) and on the protein level (Fig. 1J). The membrane-standing protease TMPRSS2 cleaves the spike protein for SARS-CoV-2 and primes it therefore for optimized binding to its entry receptor ACE2. In contrast to ACE2, the *TMPRSS2* transcript levels were significantly increased in infected compared to uninfected NHBEs but were not affected by B_2_R antagonist pre-treatment (Fig.S2A). Further experiments on the effect of the B_2_R antagonist on the SARS-CoV-2 infection of NHBE showed that pre-treatment with the B_2_R antagonist significantly reduced infection-mediated cytotoxicity measured by LDH release (Fig.1F).

### 3.2 Repetitive treatment with B_2_R antagonist inhibits SARS-CoV-2 replication and spread post-infection

The finding that B_2_R antagonism leads to a downregulation of ACE2 protein levels in lung epithelial cells was confirmed *in vivo* in a murine airway inflammation model. 24 sex-matched mice from three different strains per group were treated in two blocks intranasally with murine IL-12Fc to mimic virus-induced airway inflammation via activation of the IL-12 / IFN-γ axis.^36^ After 48 hours, mice were injected subcutaneously (s.c.) with the B_2_R antagonist and the experiment was terminated six hours or 24 hours later. IL-12Fc-pretreated mice, which were then further treated with the B_2_R antagonist on day 2, had significantly reduced ACE2 protein levels in the lungs after six hours compared to control mice, which were only treated with PBS on day 2 (Fig.2A). This effect decreased after 24 hours.

Anticipating treatment of SARS-CoV-2 infected patients with the B_2_R antagonist icatibant, NHBEs were first infected with SARS-CoV-2 and then treated with the B_2_R antagonist six hours after infection. Confirming the results of the pre-treatment, post-infection treatment with the B_2_R antagonist also attenuated the cytopathic effect of SARS-CoV-2 (Fig.2B) and reduced the number of PFU in a plaque assay on Vero E6 cells by 84% (Fig.2C).

We also aimed to reflect pharmacokinetics^39^ during treatment of early infection by treating NHBEs post-infection every 24 hours with the B_2_R antagonist repeatedly for a period of 96 hours, reflecting the drug administration of this particular substance in real life. In cells treated post-infection with 100 nM icatibant for 48 hours, the total viral RNA (Fig.2D-F IC_50_(total RNA 48h)=92.93; IC_50_(total RNA 72h)=91.56) and also the genomic viral RNA (Fig.2G-I; IC_50_(total RNA 48h)=17.01; IC_50_(total RNA 72h)=7.412) were significantly reduced by 49% and 42% on average, respectively. Treatment with 1000 nM icatibant for 48 hours led to a reduction of the total SARS-CoV-2 RNA (Fig.2D-F) and also of the genomic SARS-CoV-2 RNA (Fig.2G-I) by 69% and 56% on average, respectively. The genomic viral RNA was detected using RT-qPCR against the sequence of the SARS-CoV-2 RNA-dependent RNA polymerase (*RdRP*), which is only found in virions as well as during the viral replication. On the other hand, total viral RNA was detected with qPCR targeting a sequence of the SARS-CoV-2 *N* gene that is present in the viral genome and also in every SARS-CoV-2 protein-encoding transcript. The reduction in total SARS-CoV-2 RNA and genomic viral RNA during treatment with the B_2_R antagonist was comparable at the indicated concentrations and time points (Fig.2D-I).

### 3.3 B_2_ receptor antagonism broadly silences gene expression in bronchial epithelial cells while maintaining cell-intrinsic antiviral response

Severe cases of COVID-19 develop cytokine storms^40-42^ characterized by excessive systemic release of multiple cytokines including IP-10 (*CXCL10*), IL-6, IL-8 (*CXCL8*), and IL-10.^43-46^ These cases are currently treated with immunomodulating drugs, such as corticosteroids or biologics, like tocilizumab^47^, though these treatments may interfere with or alter the antiviral immune response. We therefore compared the effect of the B_2_R antagonist on gene expression of the bronchial epithelium in the light of SARS-CoV-2 infection with the effect of hydrocortisone. While the B_2_R antagonist mainly suppressed epithelial gene expression during infection, the effects of hydrocortisone on gene induction and gene repression were equal (Fig.3A, Tables S4-5). This finding matches previous reports.^48^ Therefore, conditions were compared in respect to the antiviral epithelial response. In fact, differentially expressed genes (DEGs) induced by SARS-CoV-2 infection in NHBEs encompassed type-I and -III interferons as well as IFN-inducible and antiviral *APOBEC* genes (Fig.S1E, Table S6). In particular, SARS-CoV-2 infection induced the antiviral cytidine deaminases APOBEC3A and B, which we previously described to be induced by type-I interferons in the treatment of hepatitis-B virus infection.^49^ On the other hand, *APOBEC3C* mRNA levels were decreased in SARS-CoV-2-infected NHBE, which could indicate a novel evasion mechanism.^50^ Neither the B_2_R antagonist nor hydrocortisone inhibited the expression of genes with cell-intrinsic antiviral effects and even increased the antiviral factor *APOBEC3A* at the mRNA level (Fig.3B, Table S7).^51^

**Figure 3.**
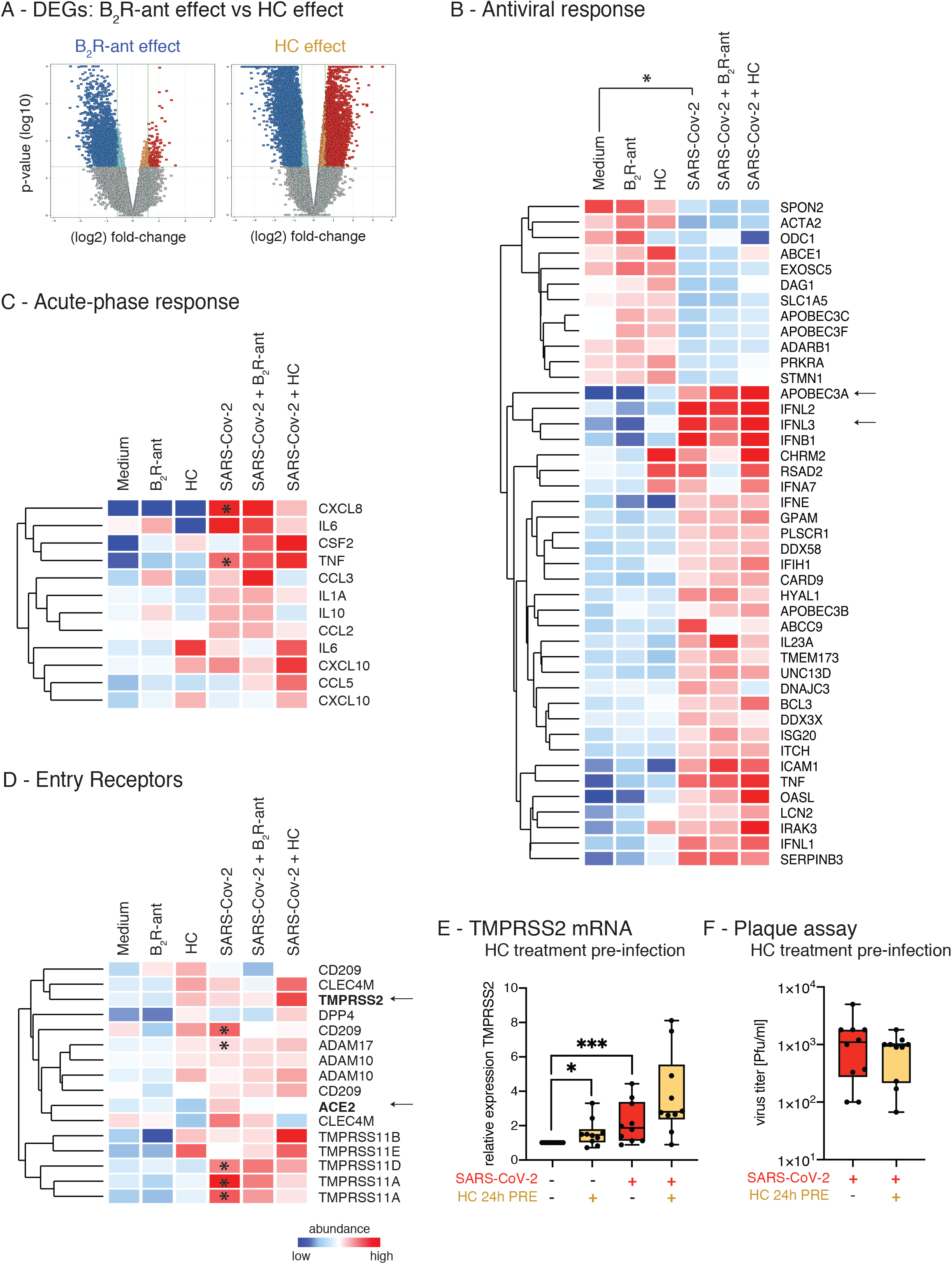
B_2_R antagonism exhibits a protective and suppressive effect on gene expression profile of airway epithelial cells. A) Volcano plots showing global gene expression changes induced by either treatment with B_2_R antagonist or hydrocortisone (HC). Red indicates significantly upregulated entities; blue indicates significantly downregulated entities. Gene expression analysis of pre-treated NHBEs after 24 hours of SARS-CoV-2 infection. B) Heat map of gene expression analysis of genes involved in the epithelial antiviral response, analysis of the effects of SARS-CoV-2 infection. Only entities with significant changes between SARS-CoV-2 infection and medium are shown (gene expression fold change FCζ1.5 with P<0.05). C) Heat map of gene expression analysis of genes involved in the acute phase response are depicted. All entities are shown. Asterisks indicate significantly regulated genes (P<0.05) in SARS-CoV-2 compared to medium. D) Heat map of gene expression analysis of known and potential virus entry receptors are depicted. All entities are shown. Color code indicates Log2-fold change from low (blue) through 0 (white) to high (red). Asterisks indicate significantly regulated genes (P<0.05) in SARS-CoV-2-infected NHBEs compared to medium. Duplicate gene names indicate the presence of two or more isoforms of the same gene in the analysis. E) Analysis of *TMPRSS2* gene expression by qPCR after 24 hours of pre-treatment with/without 10 μM hydrocortisone (HC) followed by 24 hours of SARS-CoV-2 inoculation. Red indicates SARS-CoV-2 infection; yellow indicates pre-treatment with hydrocortisone (HC). Statistical tests compared SARS-CoV-2-infected versus uninfected samples or B_2_R antagonist-treated versus untreated samples. F) Quantification of infectious particles in the supernatants of SARS-CoV-2-infected NHBEs from 10 donors that were pre-treated with/without 10 μM hydrocortisone (HC) for 24 hours. Supernatants were titrated on Vero E6 cells. The plaque assay was quantified 24 hours later. Results are depicted as plaque forming units (PFU) per milliliter.

Our gene expression analysis data show that SARS-CoV-2 infection also induces the expression of acute-phase proteins, such as TNF-α and IL-8 (*CXCL8*),^52,53^ as well as IL-17C, MIP-3α (*CCL20*), IL-36γ^54,55^ and the chemokines CXCL1, -2, -3, -8, -17, CCL2, -3, -5^54^ in primary lung epithelial cells (Fig.3C, Table S8). The induction of these factors most likely contributes to the recruitment and activation of relevant immune cells to the site of infection. Our findings are supported by previous studies of aberrant macrophage and T cell responses underlying immunopathogenesis in bronchoalveolar lavage fluid from COVID-19 patients.^56^ In addition, the gene expression of acute-phase proteins was not significantly affected in airway epithelial cells by the B_2_R antagonist or hydrocortisone treatment (Fig.3C, Table S8). Neither drug interfered with canonical cellular antiviral – and reaction-triggering response mechanisms, showing great potential for treatment options of COVID-19 while maintaining the host’s antiviral immune response.

In addition, we found that SARS-CoV-2 infection increases the expression of three known and postulated entry (co-)receptors: 1) the transmembrane serine protease TMPRSS11A (Fig.S1C, Tables S6,9-13), which was described to prime the MERS coronavirus spike protein^57^, 2) the transmembrane serine protease TMPRSS11D, which was shown to activate SARS-CoV-2 spike protein^58^ and 3) the pathogen-associated molecular pattern-binding C-type lectin receptor DC-SIGN (*CD209*), which was described to serve as entry receptor for SARS-CoV^59^ and has been suggested to act as a receptor also for SARS-CoV-2. The induction of these additional entry receptor candidates triggered by SARS-CoV-2 infection may potentiate the viral spread in the bronchial epithelium and thus represent a pathogenetic approach that needs further research. Overall, treatment with the B_2_R antagonist and hydrocortisone had no significant effects on the expression of these candidate viral entry receptors (Fig.3D, Table S14). On the other hand, hydrocortisone enhanced the expression of TMPRSS proteases (Fig.3D). In particular, when focusing on the known SARS-CoV-2 entry receptors, hydrocortisone treatment of uninfected cells was already sufficient to induce an increase in *TMPRSS2* gene expression (Fig.3E). SARS-CoV-2 infection per se also increased *TMPRSS2* expression, and pre-treatment of SARS-CoV-2-infected NHBEs with hydrocortisone further potentiated this effect. On the other hand, *ACE2* expression showed only a slight upward trend after hydrocortisone pretreatment (Fig.S2B). Finally, the hydrocortisone pre-treatment of SARS-CoV-2-infected NHBEs had no inhibitory effect on the release of infectious particles 24 hours after infection (Fig.3F), which was expected since the treatment of COVID-19 patients with corticosteroids has an immunomodulatory rationale.

### 3.4 B_2_ receptor antagonist counter-balances virus-induced gene expression, particularly genes involved in G protein-coupled receptor (GPCR) signaling and ion transport

With regard to the genes that are attenuated at the mRNA level by B_2_R antagonism, the DEGs were analyzed in a network analysis using the database string to identify enriched cellular processes. B_2_R antagonism reduced the expression levels of 343 membrane-bound receptors significantly in treated versus untreated SARS-CoV-2-infected NHBEs (Table S15). Two particular cellular processes affected by pre-treatment with the B_2_R antagonist were identified, namely G protein-coupled receptor signaling (GO:0007186; Fig.4A, S2C, Tables S4,S17-18) and ion transport (GO:0006811; Fig.4B, Tables S4,17-18). DEGs involved in both processes were significantly downregulated in treated versus untreated SARS-CoV-2-infected NHBEs (Tables 16-18). Notably, all 35 cell surface receptors induced by SARS-CoV-2 infection were downregulated in cells that were treated with the B_2_R antagonist (Fig.4C,D, Tables S19-20).

**Figure 4.**
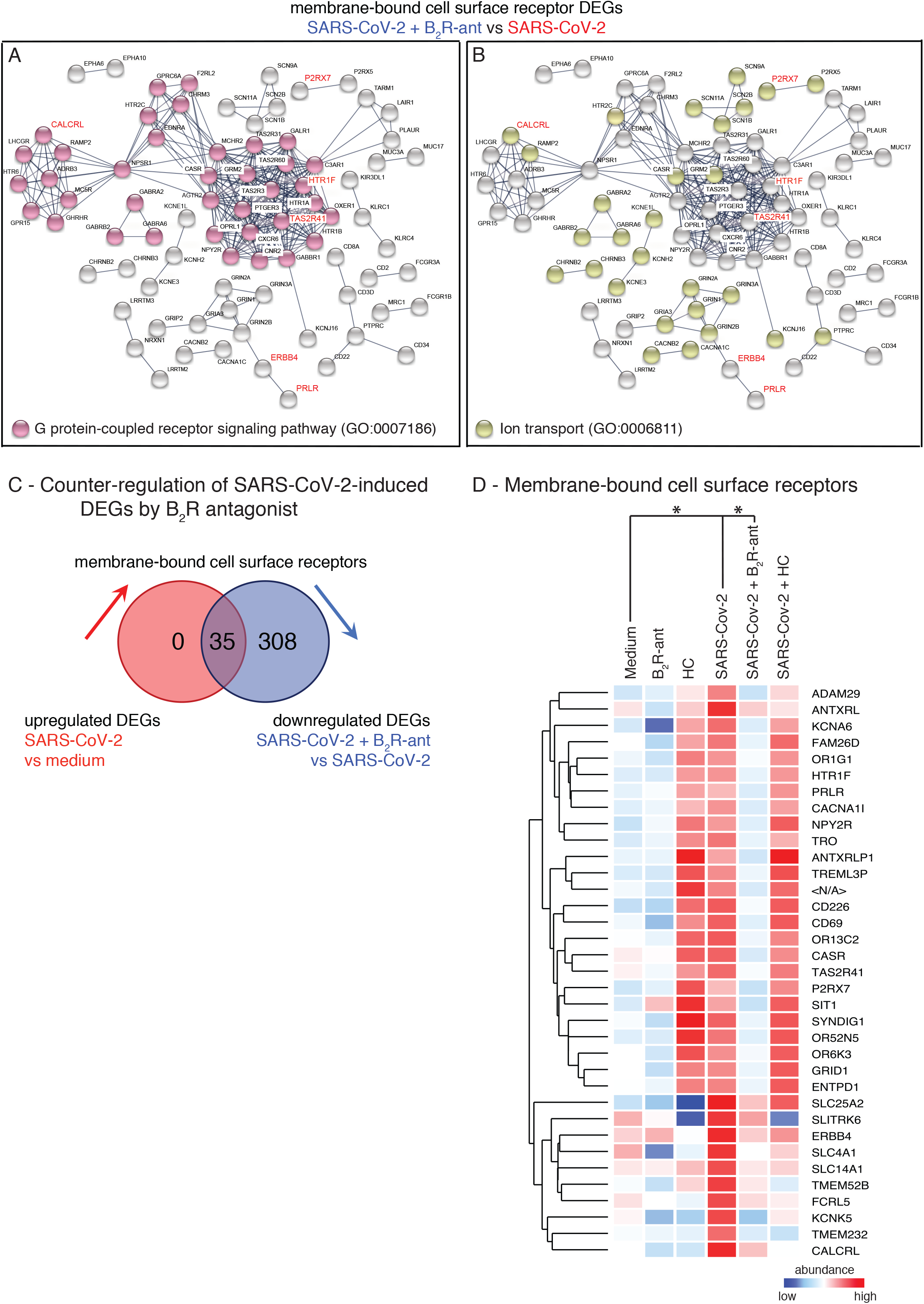
B_2_R antagonism exhibits a protective and suppressive effect on gene expression profile of airway epithelial cells. GO term enrichment analysis, which results from the string network analysis of significant DEGs from the gene expression analysis comparing infected NHBE pretreated with B_2_R antagonist with untreated infected NHBE (SARS-CoV-2 + B_2_R antagonist *versus* SARS-CoV-2). Depicted are enrichment of A) GO-term GO:0007186 “G protein-coupled receptor signaling pathway” and B) GO-term GO:0006811 “Ion transport”. Genes that were significantly upregulated in the comparison SARS-CoV-2 versus medium are highlighted in red. C) Venn diagram showing the cut set of upregulated membrane-bound cell surface receptors in SARS-CoV-2 *versus* medium and of downregulated DEGs in SARS-CoV-2 + icatibant *versus* SARS-CoV-2 (FCζ1.5; P ≤0.05). D) Heat map of gene expression analysis of the 35 membrane-bound cell surface receptors defined in cut set from Fig.4C, all upregulated upon SARS-CoV-2 infection and downregulated upon pre-treatment with B_2_R antagonist are depicted. Only entities with significant changes between SARS-CoV-2 infection and medium (up) and between SARS-CoV-2 + B_2_R antagonist and SARS-CoV-2 (down) are shown (gene expression fold change FCζ1.5 with P<0.05). Color code indicates Log2-fold change from low (blue) through 0 (white) to high (red). Duplicate gene names indicate the abundance of two or more isoforms of the same gene in the analysis.

## 4 DISCUSSION

In this study, we provide evidence for the effect of interference with the KKS at the kinin B_2_ receptor level as a means of protecting the airway epithelium from SARS-CoV-2 infection, while maintaining canonical cell-intrinsic antiviral response.

We initially hypothesized that through KKS interference, either feedback mechanisms or modulated signal transduction target the virus entry receptor ACE2 and thus interfere with the spread of SARS-CoV-2. To this end, the approved B_2_R antagonist icatibant was used in this study. Here, we demonstrate that treatment with a B_2_R antagonist inhibits the replication and spread of SARS-CoV-2 in primary airway epithelial cells, which was determined by a decrease in total and genomic SARS-CoV-2 RNA, resulting in less infectious particles in plaque assays, both when applied pre- and post-infection. While a low concentration of 1 nM B_2_R antagonist was sufficient to reduce viral RNA in primary bronchial epithelial cells when cells were treated pre-infection, 100 nM B_2_R antagonist was required to this effect, when cells were treated post-infection. In addition, the significant reduction in virus load as determined by PCR tapered off after 96 hours. On the one hand, *in vitro* infections are performed with excess amounts of virus particles. On the other hand, the fact that, due to its constitution as a peptide analog, the B_2_R antagonist icatibant used in this study has a short half-life in the human body^39^ but also pharmacological tolerance to interference at receptor level may explain why the effect reached significance after six hours but did not persist. Therefore, a higher dose of the B_2_R antagonist may be required at even shorter repetitive dosing intervals to inhibit virus replication in the long term. Therapeutic application in future dose finding studies should therefore focus on early intervention with at least two doses daily^60^ and on either optimized pharmacokinetics or increased high local tissue concentrations, e.g. through topical application.

Two potential mechanisms of action for suppressing SARS-CoV-2 replication and spread in airway epithelium are revealed by the current study:

1) Treatment with the B_2_R antagonist led to a downregulation of the viral entry receptor ACE2, *in vitro* in primary airway epithelial cells and *in vivo* in a murine airway inflammation model. Since the decrease of genomic SARS-CoV-2 RNA and total SARS-CoV-2 RNA was comparable, we conclude that the B_2_R antagonist icatibant does probably not affect the viral transcription machinery but inhibits the infection rather on the levels of entry, protein synthesis/processing and assembly, maturation or budding.

In comparison, the corticosteroid hydrocortisone even upregulated TMPRSS2 in infected airway epithelial cells. It is noteworthy that hydrocortisone did not change the release of infectious particles from infected airway epithelial cells into the supernatant. Although TMPRSS2 expression was even enhanced by hydrocortisone, our data implicate that this effect on TMPRSS2 alone is insufficient to increase susceptibility for SARS-CoV-2 infection.

2) Treatment with the B_2_R antagonist had a broad suppressive effect on gene expression of multiple cell signaling molecules, in particular on membrane-standing factors involved in GPCR signaling and ion transport.

It has recently been published that SARS-CoV-2 may use cellular GPCR signaling pathways, thereby modulate epithelial transport mechanisms involved in ion transport and thereby cause a local ion imbalance in the airways.^61^ In addition, an extensive recent study described that intracellular SARS-CoV-2 protein interactions include factors involved in intracellular trafficking and transport, particularly ion transport and solute carrier transport.^62^ In fact, the SARS-CoV-2 infection led to a differential regulation of the gene expression of 12 potassium channel (5 upregulated / 7 downregulated), 1 sodium channel (down), but in particular of 55 members of the solute carrier family (24 downregulated / 31 upregulated) in primary airway epithelial cells. On the other hand, B_2_R antagonist treatment of SARS-CoV-2-infected NHBE resulted in a downregulation of 20 potassium channel genes and 6 sodium channel genes, as well as a downregulation of 29 members of the solute carrier family.

We therefore conclude that B_2_R antagonism not only impedes the viral entry process by reducing ACE2, as we had hypothesized, but also counter-regulates cellular processes that include GPCR signaling and transmembrane ion transport, which SARS-CoV-2 may utilize for efficient replication and viral spread.

In conclusion, the results of this study suggest that B_2_ receptor-antagonism protects airway epithelial cells from SARS-CoV-2 spread by reducing ACE2 levels and by interfering with several cellular signaling processes. Further research is needed to elucidate more details about molecular mechanisms involved in the viral life cycle that kinin B_2_ receptor antagonism targets and underlie its effects against SARS-CoV-2 infection. Based on these data, we speculate that the protective effects of B_2_R antagonism could potentially prevent the early stages of COVID-19 from progressing into severe acute respiratory distress syndrome (ARDS) with structural airway damage and fibrotic changes. We therefore propose that the safe approved B_2_R antagonist icatibant be tested in clinical trials for two important aspects: 1) Treatment of early COVID-19 disease targeting the replication and spread of the virus. 2) Optimized dosage regimen to reflect pharmacokinetics and possible pharmacological tolerance at the receptor level. Future controlled clinical trials must provide substantial evidence for optimal dosage regimen, application, efficacy, and safety to investigate, whether KKS interference at the kinin B_2_ receptor level can prevent the escalation of COVID-19 to ARDS and long-term lung damage.

## Data Availability

All data are provided in the supplemental tables.

## Abbreviations

ACE2: angiotensin-converting enzyme 2
ant-B_2_R: kinin B_2_ receptor antagonist
*BDKRB1*: kinin B_1_ receptor (gene name)
*BDKRB2*: kinin B_2_ receptor (gene name)
B_1_R: kinin B_1_ receptor
B_2_R: kinin B_2_ receptor
B_2_R antagonist: kinin B_2_ receptor antagonist
CAS: contact activation system
COVID-19: coronavirus disease of 2019
DABK: des-Arg9-bradykinin
GPCR: G protein-coupled receptor
HC: hydrocortisone
HMW-kininogen: high-molecular-weight-kininogen
IC50: half maximal inhibitory concentration
KKS: kinin-kallikrein system
LDH: lactate dehydrogenase
NHBE: normal human bronchial epithelial cells
RAS: renin-angiotensin system
SARS-CoV-2: Severe Acute Respiratory Syndrome Coronavirus-2
TMPRSS: transmembrane serine protease

