## Supplemental Information for "Early Reduction of SARS-CoV-2 Replication in Bronchial Epithelium by Kinin B_2_ Receptor Antagonism"

### SUPPORTING INFORMATION

MATERIALS AND METHODS

**Cell culture**. Primary normal human bronchial epithelial cells (NHBE, Lonza) from genetically independent donors were grown as monolayers from low passages in serum-free pre-defined bronchial epithelial cell growth medium according to manufacturer's instructions (BEGM BulletKit, Lonza). NHBEs were treated in 12-well plates at 80% confluence. To avoid effects induced by growth factors in the BEGM medium, cells were rested in basal medium without supplements for 12 hours, then stimulated with icatibant at 1 nM or hydrocortisone at 10 μM, followed by further 24 hours of infection with SARS-CoV-2 at the biosafety level 3 laboratory. For RNA analysis, cells were lysed in RLT buffer (Qiagen, Hilden, Germany). For protein analysis, cells were lysed in 1x protein extraction buffer provided with the ACE2 ELISA Kit (Abcam, Cambridge, UK). Cell culture supernatant was collected for further viral and cytolytic analysis.

**Infection with SARS-CoV-2.** The SARS-CoV-2 isolate hCoV-19/Germany/BAV-PL-virotum-nacq/2020 (GISAID accession ID: EPI_ISL_582134) was derived from patient material and amplified on Vero E6 cells (passage 0; ATCC, Manassas, US) and used for infection experiments (passage 4) with an MOI of 0.5 (determined by plaque assay) by adding virus stock to the BEBM culture medium after 24 hours of stimulation. Time of harvest after inoculation was dependent on the experimental setting. For repetitive treatment with icatibant of NHBEs post-infection (Fig.2D-I), NHBEs from 8 genetically independent donors were infected with SARS-CoV-2 with a MOI of 0.5 at T-6 and after 6 hours (T0), treatment with a range of doses of icatibant was started and repeated every 24 hours. Cells were harvested at 24, 48, 72 and 96 hours after treatment start and subjected to RNA extraction and relative viral RNA quantification normalized to a housekeeper index.

**Plaque assay.** Supernatant of SARS-CoV-2-infected NHBEs was titrated (1:5 dilutions of the stock) in Dulbecco's Modified Eagle Medium (DMEM) with 1% penicillin / streptomycin, 1% L-glutamine, 1% non-essential aminoacids, and 1% sodium pyruvate (all Gibco, Thermo Fisher Scientific, Carlsbad, CA, USA) in a total volume/well of 100 µl on 96-well plates containing 70-80 % confluent Vero E6 cells. After 2 hours, the inoculum was discarded, and cells were layered with 100 µl MEM containing 0.5 % carboxymethyl cellulose (1,500 cP, Sigma-Aldrich, St. Louis US). 48 hours post inoculation, 100 µl formaldehyde was added to a final concentration of 5 % for 15 min. Supernatant was taken off, cells were washed with PBS twice and incubated with crystal violet (Sigma-Aldrich) solution for 15 min and washed with PBS again. Plaque-forming units per milliliter pfu/ml were calculated with the formula pfu/mL = number of plaques/dilution x volume of analyzed supernatant.

**RNA isolation and gene expression analysis.** mRNA was extracted from patient nasal brushings and from NHBEs using RNeasy Micro Kit (Qiagen) with on-column DNase digestion (Qiagen). RNA quantification and quality assessment were performed using Nanodrop Technologies (Wilmington) and RNA 6000 Nano Chip Kit with the Agilent 2100 Bioanalyzer (Agilent Technologies, Waldbronn, Germany) according to manufacturer's instructions.

**RT-qPCR-based detection of SARS-CoV-2 RNA.** Isolated RNA was subject to cDNA synthesis by SuperScript III First-Strand Synthesis SuperMix for qRT-PCR (Invitrogen). On a LightCycler 480 II (Roche Diagnostics, Penzberg, Germany) total SARS-CoV-2 RNA was quantified using SARS-CoV-2 N1 primers and probe according to CDC guidelines and a SYBR Green PCR mix (Roche Diagnostics). SARS-CoV-2 genomic RNA was quantified using RdRP primers. The specific primers used in real-time PCR are listed in Table S21. SARS-CoV-2 RNA was normalized with relative quantification to endogenous control ACTB using the 1/ECT formula.

**3D-bronchial air-liquid interphase organoid cultures.** Low passage primary NHBEs were expanded to 95% confluence in PneumaCult-Ex Plus expansion medium (Stemcell) according to manufacturer's instructions. Corning transwell polyester membrane cell culture inserts (type 3460, Merck) were precoated with 1% collagen (Sigma-Aldrich) and the cells were transferred to 12-well plates containing precoated transwell inserts on day 0 with a density of $3x10^5$ cells/$cm^2$. Every other day, PneumaCult-Ex Plus expansion medium (Stemcell) was replenished in apical and basal chambers. On day 3, cells were lifted to air (air-lift) by removing the apical medium. Medium in the basal chamber was exchanged for Pneumacult-ALI maintenance medium (Stemcell). Excess mucus was removed from day 7. Once a week, the transepithelial electrical resistance (TEER) was measured using an EVOM2 instrument (World Precision Instruments). With a TEER of >700, but from day 21 at the earliest, the 3D organoid cultures were regarded as completely differentiated and were then treated and infected with SARS-CoV-2.

**Infection of 3D organoid cultures.**

24 hours prior to treatment of the 3D air-liquid interphase organoid cultures, the supplement hydrocortisone was withdrawn. The cultures were then pretreated from the basal side with/without 1 nM icatibant (HOE 140 (icatibant), Sigma-Aldrich) for 24 hours. Subsequently, SARS-CoV-2 viral suspension (MOI 0.5, strain hCoV-19/Germany/BAV-PL-virotum-nacq/2020) was added from the apical side and incubated for 2 hours, then the virus suspension was removed with PBS washes and the cultures were kept for another 46 hours before harvest.

**FFPE sections of 3D organoid cultures.** Cultures were fixed in 4% paraformaldehyde and embedded in paraffin and sections of 4 μm thickness were produced at the routine diagnostics at the Medical School of the Technical University of Munich, Campus Biederstein. Slides were subjected to standard hematoxylin eosin staining to identify the culture structure by light microscopy.

**Tissue homogenization and murine ACE2 ELISA.** Tissue lysates were prepared by homogenization in ice-cold lysis buffer (Cell Signaling) to a final concentration of 2x lysis buffer in water, containing protease inhibitor (Complete Mini, Roche). 0.1 ml of lysis buffer was added per 100 mg of tissue. Tissue was homogenized using metal beads and Qiagen tissue lyser 20/s for 90 seconds until it was completely homogenized. Samples were centrifuged for 10 minutes at 11,000 x g at 4°C and supernatants were transferred to fresh tubes. Total protein concentration was determined with the Pierce BCA Protein Assay Kit (Thermo Scientific) to determine extraction efficacy across samples, before further analysis by ELISA. Mouse ACE2 ELISA (PicoKine Kit, Boster Bio) was performed according to the manufacturer's instructions using tissue lysate diluted 1:100. Colorimetric readout was acquired using Spark microplate reader (Tecan). Data analysis was performed using Microsoft Excel, R and GraphPad Prism.

**Reverse transcription and quantitative real-time qPCR.** Isolated total RNA was reverse transcribed using a high-capacity cDNA kit (Applied Biosystems) according to manufacturer's instructions. Real-time PCR profiles were visualized using FastStart Universal SYBR Green Mastermix (Roche) and quantified by the ViiA 7 Real-Time PCR System (Applied Biosystems). The specific primers used in real-time PCR are listed in supplementary Table S21. ACE2 and TMPRSS2 mRNA expression was normalized to housekeeping gene index *ACTB* and *HPRT* and the relative quantification was performed using the comparative threshold cycle ($2^{-\Delta\Delta Ct}$) method (relative gene expression). All amplifications were carried out at least in duplicate.

**Gene expression analysis using Agilent microarray technologies.** Microarray experiments were performed using MIAME criteria. Genespring Software GX 14.9.1 (Agilent Technologies) with minimal data reduction constraints (1.5-fold change and $P<0.05$ cutoff) was used to analyze microarray data as previously described.[1,2] Upon data import a standard baseline transformation to the median of all values was performed, including log transformation and computation of fold changes (log2(A/B)=log2(A)-log2(B)). Subsequently, a principle component analysis was conducted, which revealed a homogenous component distribution. Compromised array signals were excluded from further analysis (array spot is non-uniform if pixel noise of feature exceeds threshold or is above saturation threshold). Genes with an absolute log2 fold change larger than 1.5 and a p-value smaller than the testing level of 0.05 by using the Moderated T-Test were defined as significantly differentially expressed hits. The significantly regulated genes were summarized in entity lists (see supplemental tables). These entity lists were analyzed for overlaps using Venn diagrams. Manhattan cityblock on entities (Ward's linkage) was used to cluster changes in gene expression.

Gene Ontology (GO) terms "0007267", "0005125", "0008009", and "0005615" for secreted factors, GO terms "0007267", "0005125", "0008009", and "0005615" for identification of biomarkers, GO terms "0038023", "0004896", "0004888", and "0005887" for surface receptors, GO terms "0009615", "0039528", "0039530", "0039639", "0051607", and "0009597" for anti-viral response. Membrane-bound cell surface receptors were selected from a Venn diagram analysis of the GO-term selection of surface receptors (see above) minus the GO-term selection of secreted genes (see above) in order to isolate surface receptors that are not as well secreted as soluble ligands. The data discussed in this publication will be deposited in NCBI's Gene Expression Omnibus and are accessible through GEO Series accession number pending.

**String network analysis.** Protein-protein interactions were computed using an open-access tool, the string network analysis version 11.0 (string-db.org)[3-13] in order to extract enriched cellular processes and pathways affected by icatibant treatment. The classical classification systems Gene Ontology was relevant for this study. Enriched entities are colored in the interaction plots: violet for GO:0007186 “G protein-coupled receptor signaling pathway” and light green for GO:0006811 “Ion transport” in Figure 2.

**Cytotoxicity assay.** Lactate Dehydrogenase Assay (LDH) assay for assessing the cytotoxicity of virus-infected NHBES in combination with or without icatibant was performed using the LDH-detecting CytoTox 96® Non-Radioactive Cytotoxicity Assay (Promega GmbH) according to manufacturer’s instructions.

**ELISA for human ACE2 levels.** Protein levels of ACE2 in NHBE cell lysates were determined using the human ACE2 ELISA Kit (Abcam). Assays were performed according to manufacturer´s instructions.

**Statistical analysis.** Two-tailed Mann-Whitney U tests were used to determine statistical significance (GraphPad Prism Version 8.4.2, GraphPad Software). For the in vivo mouse study, we performed a prospective power analysis based on human data that predict a 33% reduction in ACE2 following icatibant treatment (mean 1: 1; mean 2: 0.666; SD: 0.26; alpha = 0.05; P = 0.9, power.t.test in R, rounded up). In accordance with the criteria of this *a priori* power analysis, we used a 1-tailed t-test, that compared icatibant-treated groups with untreated groups, resulting in 12 animals per group. Here, we have specifically tested the hypothesis for a downregulation of ACE2 only in mice treated with icatibant compared to untreated mice. The half maximal inhibitory concentration ($IC_{50}$) values were deferred from sigmoidal interpolation of a standard curve using a robust nonlinear regression model in GraphPad Prism. Results are depicted as median with range, if not otherwise indicated in the figure legends. $P<0.05$ was considered statistically significant. Statistically significant differences were depicted as p-values *$P<0.05$, **$P<0.01$, and ***$P<0.001$.

**Data availability statement.** The data discussed in this publication are deposited in NCBI's Gene Expression Omnibus and are accessible under the GEO Series accession number GSE176405.

SUPPLEMENTAL TABLES

Table S1. DEGs from transcriptome analysis filtered on secreted factors from nasal scrapings from SARS-CoV-2 acute positive patients compared to negative patients

Table S2. DEGs from transcriptome analysis filtered on kallikreins from nasal scrapings from SARS-CoV-2 acute positive patients compared to negative patients

Table S3. DEGs from transcriptome analysis filtered on kinin-kallikrein-system members from nasal scrapings from SARS-CoV-2 acute positive patients compared to negative patients

Table S4. DEGs comparing SARS-CoV-2 + B2R antagonist versus SARS-CoV-2

Table S5. DEGs comparing SARS-CoV-2 + hydrocortisone (HC) versus SARS-CoV-2

Table S6. DEGs of antiviral epithelial response comparing SARS-CoV-2 versus medium

Table S7. DEGs of antiviral epithelial response comparing SARS-CoV-2 versus medium

Table S8. Gene expression of members of the acute-phase response comparing SARS-CoV-2 versus medium

Table S9. Gene expression of members of RAS and KKS comparing SARS-CoV-2 versus medium

Table S10. Differentially expressed interleukins comparing SARS-CoV-2 versus medium

Table S11. Potential viral entry receptor DEGs comparing SARS-CoV-2 versus medium

Table S12. Toll-like receptor (TLR) DEGs comparing SARS-CoV-2 versus medium

Table S13. Chemokine DEGs comparing SARS-CoV-2 versus medium

Table S14. Gene expression of potential viral entry receptors comparing SARS-CoV-2 versus medium

Table S15. Membrane-bound receptor DEGs comparing SARS-CoV-2 + B2R antagonist versus SARS-CoV-2

Table S16. Membrane-bound receptor DEGs comparing SARS-CoV-2 + B2R antagonist versus SARS-CoV-2 (P <= 0.05; FC >=2.5)

Table S17. Interactions output of String network analysis of membrane-bound receptor DEGs comparing SARS-CoV-2 + B2R antagonist versus SARS-CoV-2

Table S18. Cellular process enrichment analysis output of String network analysis of membrane-bound receptor DEGs comparing SARS-CoV-2 + B2R antagonist versus SARS-CoV-2

Table S19. Membrane-bound cell surface receptor DEGs, upregulated in SARS-CoV-2 versus medium

Table S20. Cut set: Membrane-bound cell surface receptor DEGs, upregulated in SARS-CoV-2 versus medium and downregulated in SARS-CoV-2 + B2R antagonist versus SARS-CoV-2

Table S21. Reagents

#### SUPPLEMENTAL FIGURE LEGENDS

Figure S1. Epithelial response of primary NHBEs to SARS-CoV-2 infection.

Gene expression analysis of NHBEs after 24 hours of SARS-CoV-2 infection versus medium using microarray technology: A) Heat map of selected genes of the KKS and RAS are depicted independent of significance analysis. All entities are shown. Asterisks indicate significantly regulated genes ($P<0.05$) in SARS-CoV-2 compared to medium. Heat map of gene expression analysis of B) epithelium-derived interleukins, C) a selection of confirmed and potential SARS-CoV-2 entry receptors, D) Toll-like family members, E) factors involved in the immediate antiviral response of infected epithelial cells, and of E) chemokines. In B-F, only significantly regulated entities in SARS-CoV-2-infected versus uninfected NHBEs are shown. Color code indicates Log2-fold change from low (blue) through 0 (white) to high (red). Duplicate gene names indicate the abundance of two or more isoforms of the same gene in the analysis.

Figure S2.

A) Analysis of *TMPRSS2* gene expression by qPCR after 24 hours of pre-treatment of NHBEs of 10 donors with/without 1 nM $B_2R$ antagonist, followed by SARS-CoV-2 inoculation for 24 hours of NHBEs. Results are depicted as median with range. B) *ACE2* gene expression analysis using qPCR after 24 hours of pre-treatment with/without 10 μM hydrocortisone (HC) followed by 24 hours of SARS-CoV-2 infection of NHBEs. Results are depicted as median with range. For Figures S2A and B, statistical tests compared SARS-CoV-2-infected versus uninfected samples or $B_2R$ antagonist-treated versus untreated samples. C) Heat map of gene expression analysis of membrane-bound cell surface receptors included in pathway analysis in Figures 4A,B, the were the highest downregulated upon pre-treatment with $B_2R$ antagonist during SARS-CoV-2 infection of NHBES are depicted. SARS-CoV-2 + $B_2R$ antagonist and SARS-CoV-2 are shown ($FC \geq 2.5$; $P \leq 0.05$). Color code indicates Log2-fold change from low (blue) through 0 (white) to high (red). Duplicate gene names indicate the abundance of two or more isoforms of the same gene in the analysis.
