## Supplementary figures and images for "Early Reduction of SARS-CoV-2 Replication in Bronchial Epithelium by Kinin B_2_ Receptor Antagonism"

### Suplemental Figures

SUPPLEMENTAL FIGURE S1

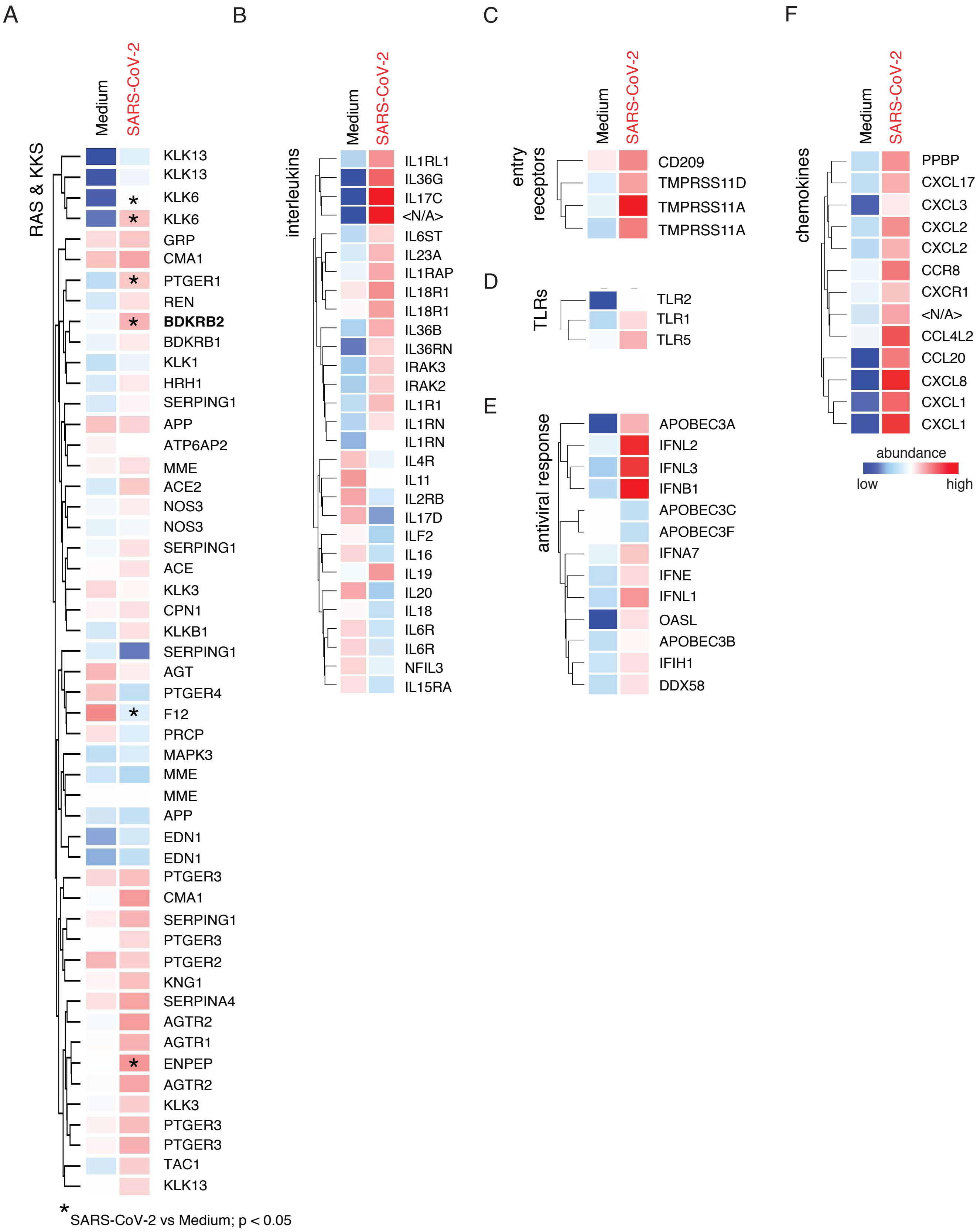

SUPPLEMENTAL FIGURE S2

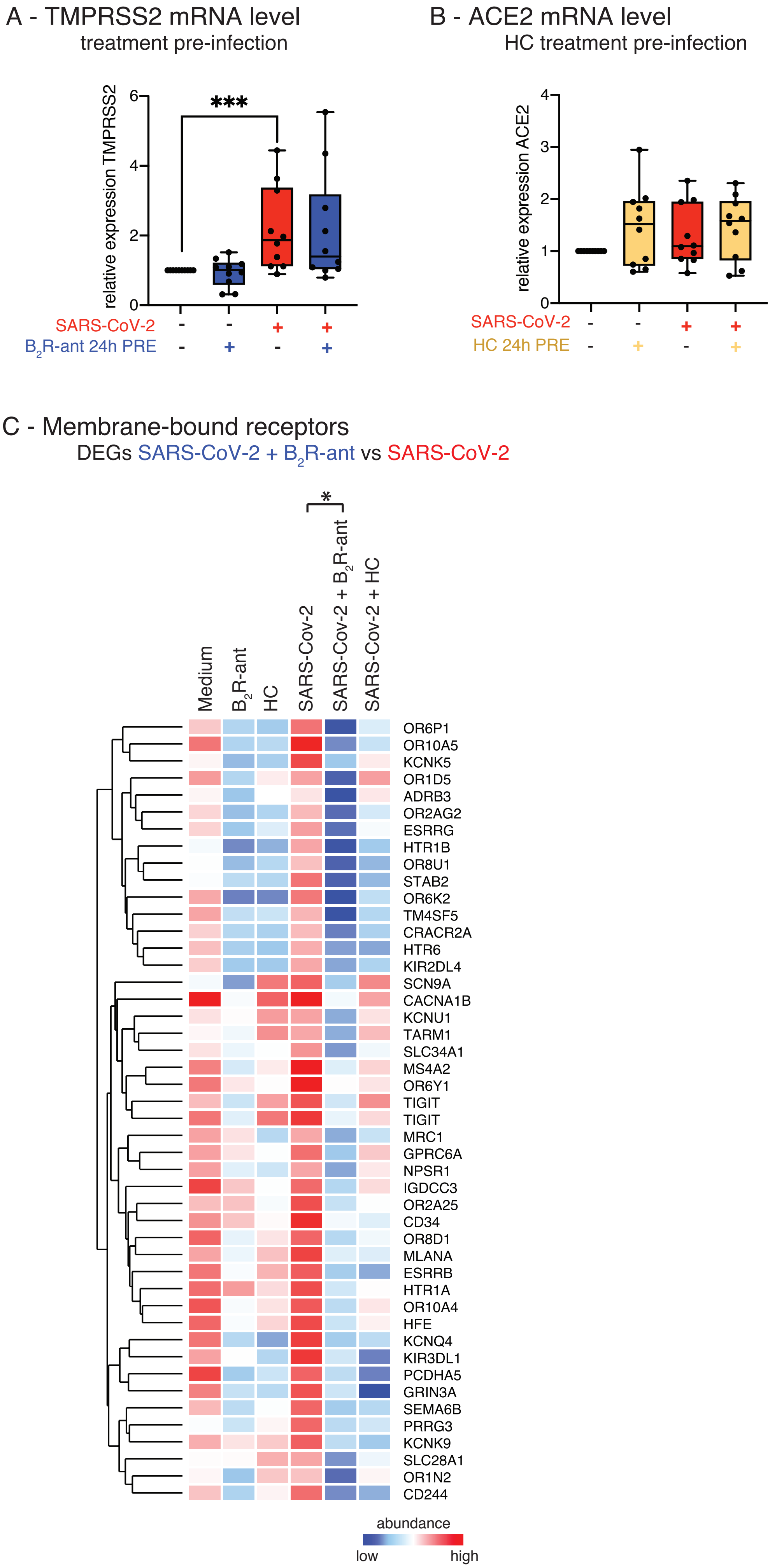
