## Supplementary material for "Early Reduction of SARS-CoV-2 Replication in Bronchial Epithelium by Kinin B_2_ Receptor Antagonism": Supl. Table 1

**Supplemental Table S1.** DEGs from transcriptome analysis filtered on secreted factors from nasal scrapings from SARS-CoV-2 acute positive patients compared to negative patients

| ProbeName | p ([pos acute] | Regulation ([ | FC ([pos acut | Log FC ([pos | GeneSymbol | Description |
|---|---|---|---|---|---|---|
| A_23_P252981 | 0,34020385 | up | 1,1672416 | 0,22310317 | ACE2 | Homo sapiens angiotensin I converting enzyme 2 (ACE2), mRNA [NM_021804] |
| A_33_P3411907 | 0,23858786 | up | 1,3503011 | 0,43328118 | FGF5 | Homo sapiens fibroblast growth factor 5 (FGF5), transcript variant 2, mRNA [NM_033143] |
| A_33_P3525263 | 0,60033906 | up | 1,5155119 | 0,5998051 | A2ML1 | Homo sapiens alpha-2-macroglobulin-like 1 (A2ML1), transcript variant 1, mRNA [NM_144670] |
| A_23_P346093 | 0,08312471 | up | 1,5961888 | 0,6746313 | TMC8 | Homo sapiens transmembrane channel-like 8 (TMC8), mRNA [NM_152468] |
| A_23_P141802 | 0,6342188 | up | 1,1596125 | 0,21364284 | SERPINB7 | Homo sapiens serpin peptidase inhibitor, clade B (ovalbumin), member 7 (SERPINB7), transcript variant 2, mRNA [NM_001040147] |
| A_23_P114232 | 0,8563269 | down | -1,0283316 | -0,0403056 | PRDX4 | Homo sapiens peroxiredoxin 4 (PRDX4), mRNA [NM_006406] |
| A_23_P12128 | 0,21435596 | up | 1,2048411 | 0,26884294 | TSHB | Homo sapiens thyroid stimulating hormone, beta (TSHB), transcript variant 1, mRNA [NM_000549] |
| A_23_P210515 | 0,31126252 | down | -1,1782407 | -0,2366342 | NCOA5 | Homo sapiens nuclear receptor coactivator 5 (NCOA5), mRNA [NM_020967] |
| A_23_P89431 | 0,6829328 | up | 1,2285764 | 0,2969876 | CCL2 | Homo sapiens chemokine (C-C motif) ligand 2 (CCL2), mRNA [NM_002982] |
| A_23_P154643 | 0,18161069 | up | 1,533928 | 0,6172308 | BMP7 | Homo sapiens bone morphogenetic protein 7 (BMP7), mRNA [NM_001719] |
| A_23_P38505 | 0,45073867 | up | 1,1462373 | 0,19690569 | CXCL16 | Homo sapiens chemokine (C-X-C motif) ligand 16 (CXCL16), transcript variant 1, mRNA [NM_022059] |
| A_33_P3350863 | 0,77439594 | down | -1,0995045 | -0,1368535 | RETN | Homo sapiens resistin (RETN), transcript variant 1, mRNA [NM_020415] |
| A_23_P214821 | 0,0720521 | up | 2,3768287 | 1,2490379 | EDN1 | Homo sapiens endothelin 1 (EDN1), transcript variant 1, mRNA [NM_001955] |
| A_23_P98382 | 0,8317378 | down | -1,0278152 | -0,0395809 | TIMM8B | Homo sapiens translocase of inner mitochondrial membrane 8 homolog B (yeast) (TIMM8B), transcript variant 1, mRNA [NM_012459] |
| A_33_P3382924 | 0,26157108 | up | 2,159817 | 1,1109091 | SPARC | Homo sapiens secreted protein, acidic, cysteine-rich (osteonectin) (SPARC), mRNA [NM_003118] |
| A_24_P90900 | 0,62406707 | up | 1,1422311 | 0,19185458 | CTRL | Homo sapiens chymotrypsin-like (CTRL), mRNA [NM_001907] |
| A_23_P94533 | 0,33538678 | up | 1,2000083 | 0,26304436 | CTSL | Homo sapiens cathepsin L (CTSL), transcript variant 1, mRNA [NM_001912] |
| A_24_P55148 | 0,6589927 | down | -1,093799 | -0,1293476 | HIST1H2BJ | Homo sapiens histone cluster 1, H2bj (HIST1H2BJ), mRNA [NM_021058] |
| A_23_P11774 | 0,5802634 | down | -1,0961056 | -0,1323868 | UTP11L | Homo sapiens UTP11-like, U3 small nucleolar ribonucleoprotein (yeast) (UTP11L), mRNA [NM_016037] |
| A_33_P3246833 | 0,2517003 | up | 2,0968657 | 1,0682344 | IL1RN | Homo sapiens interleukin 1 receptor antagonist (IL1RN), transcript variant 4, mRNA [NM_173843] |
| A_23_P114057 | 0,5955839 | down | -1,1266555 | -0,1720464 | SEMA4C | Homo sapiens sema domain, immunoglobulin domain (Ig), transmembrane domain (TM) and short cytoplasmic domain, (semaphorin) 4C (SEMA4C), mRNA [NM_0177 |
| A_23_P301846 | 0,07293202 | up | 1,5677545 | 0,64869964 | CALCA | Homo sapiens calcitonin-related polypeptide alpha (CALCA), transcript variant 2, mRNA [NM_001033952] |
| A_23_P218675 | 0,07808613 | up | 1,6668886 | 0,7371577 | WFDC2 | Homo sapiens WAP four-disulfide core domain 2 (WFDC2), mRNA [NM_006103] |
| A_24_P122137 | 0,28342992 | up | 1,3023006 | 0,38106245 | LIF | Homo sapiens leukemia inhibitory factor (LIF), transcript variant 1, mRNA [NM_002309] |
| A_24_P62783 | 0,37532657 | up | 1,3871528 | 0,4721267 | FABP3 | Homo sapiens fatty acid binding protein 3, muscle and heart (FABP3), mRNA [NM_004102] |
| A_23_P214079 | 0,30000716 | up | 2,1004653 | 1,070709 | SPINK1 | Homo sapiens serine peptidase inhibitor, Kazal type 1 (SPINK1), mRNA [NM_003122] |
| A_23_P88626 | 0,3222333 | down | -2,0550373 | -1,0391645 | ANPEP | Homo sapiens alanyl (membrane) aminopeptidase (ANPEP), mRNA [NM_001150] |
| A_23_P258190 | 0,5863842 | down | -1,3434343 | -0,4259258 | AKR1B1 | Homo sapiens aldo-keto reductase family 1, member B1 (aldose reductase) (AKR1B1), mRNA [NM_001628] |
| A_23_P1594 | 0,2955662 | up | 1,1927941 | 0,254345 | VEGFB | Homo sapiens vascular endothelial growth factor B (VEGFB), transcript variant VEGFB-186, mRNA [NM_003377] |
| A_23_P113393 | 0,0251484 | up | 1,5995178 | 0,67763704 | APLN | Homo sapiens apelin (APLN), mRNA [NM_017413] |
| A_23_P7313 | 0,00349406 | up | 10,502982 | 3,3927271 | SPP1 | Homo sapiens secreted phosphoprotein 1 (SPP1), transcript variant 1, mRNA [NM_001040058] |
| A_32_P170749 | 0,8960107 | up | 1,0255061 | 0,03633613 | STAG3 | Homo sapiens stromal antigen 3 (STAG3), transcript variant 4, mRNA [NM_001282718] |
| A_23_P113111 | 0,10926139 | down | -2,7224672 | -1,4449147 | AR | Homo sapiens androgen receptor (AR), transcript variant 1, mRNA [NM_000044] |
| A_24_P122403 | 0,28822765 | down | -1,1593254 | -0,2132855 | TCEB3 | Homo sapiens transcription elongation factor B (SIII), polypeptide 3 (110kDa, elongin A) (TCEB3), mRNA [NM_003198] |
| A_23_P153964 | 0,20688777 | down | -3,0889125 | -1,6270989 | INHBB | Homo sapiens inhibin, beta B (INHBB), mRNA [NM_002193] |
| A_23_P122216 | 0,15532492 | down | -1,3783735 | -0,4629669 | LOX | Homo sapiens lysyl oxidase (LOX), transcript variant 1, mRNA [NM_002317] |
| A_33_P3210379 | 0,0541494 | down | -1,5626283 | -0,6439746 | SCGB3A1 | Homo sapiens secretoglobin, family 3A, member 1 (SCGB3A1), mRNA [NM_052863] |
| A_33_P3215640 | 0,0322807 | down | -2,4941149 | -1,3185279 | PI16 | Homo sapiens peptidase inhibitor 16 (PI16), transcript variant 1, mRNA [NM_153370] |
| A_23_P106661 | 0,26954055 | up | 1,4148043 | 0,50060254 | CMTM1 | Homo sapiens CKLF-like MARVEL transmembrane domain containing 1 (CMTM1), transcript variant 17, mRNA [NM_052999] |
| A_23_P101505 | 0,16285726 | up | 1,7061496 | 0,77074414 | KLK11 | Homo sapiens kallikrein-related peptidase 11 (KLK11), transcript variant 2, mRNA [NM_144947] |
| A_23_P122924 | 0,1506425 | up | 2,1057057 | 1,0743039 | INHBA | Homo sapiens inhibin, beta A (INHBA), mRNA [NM_002192] |
| A_23_P369237 | 0,87318844 | up | 1,0286412 | 0,04073988 | ADIPOQ | Homo sapiens adiponectin, C1Q and collagen domain containing (ADIPOQ), transcript variant 2, mRNA [NM_004797] |
| A_23_P204847 | 0,39333293 | up | 1,3972312 | 0,48257077 | LCP1 | Homo sapiens lymphocyte cytosolic protein 1 (L-plastin) (LCP1), mRNA [NM_002298] |
| A_23_P24104 | 0,8725616 | up | 1,0661993 | 0,09247714 | PLAU | Homo sapiens plasminogen activator, urokinase (PLAU), transcript variant 1, mRNA [NM_002658] |
| A_23_P88106 | 0,7455171 | down | -1,0779434 | -0,1082815 | MCF2L | Homo sapiens MCF.2 cell line derived transforming sequence-like (MCF2L), transcript variant 2, mRNA [NM_024979] |
| A_33_P3346826 | 0,2416095 | up | 1,4380172 | 0,524081 | IL32 | Homo sapiens interleukin 32 (IL32), transcript variant 4, mRNA [NM_001012633] |
| A_33_P3259092 | 0,7082634 | down | -1,100957 | -0,1387582 | FKTN | Homo sapiens fukutin (FKTN), transcript variant 1, mRNA [NM_001079802] |
| A_23_P118061 | 0,4154726 | down | -1,1452136 | -0,1956167 | CKLF | Homo sapiens chemokine-like factor (CKLF), transcript variant 4, mRNA [NM_181641] |
| A_24_P844984 | 0,8423595 | up | 1,1359301 | 0,18387401 | PIGR | Homo sapiens polymeric immunoglobulin receptor (PIGR), mRNA [NM_002644] |
| A_23_P13907 | 0,3633783 | up | 1,4200653 | 0,50595725 | IGF1 | Homo sapiens insulin-like growth factor 1 (somatomedin C) (IGF1), transcript variant 4, mRNA [NM_000618] |
| A_33_P3413701 | 0,0714303 | down | -1,766756 | -0,8211029 | ERAP1 | Homo sapiens endoplasmic reticulum aminopeptidase 1 (ERAP1), transcript variant 2, mRNA [NM_001040458] |

| | | | | | | |
|---|---|---|---|---|---|---|
| A_24_P382579 | 0,54189306 | up | 1,0794927 | 0,11035347 | OXT | Homo sapiens oxytocin/neurophysin I prepropeptide (OXT), mRNA [NM_000915] |
| A_23_P218111 | 0,5657635 | up | 1,2052906 | 0,26938096 | SERPINA1 | Homo sapiens serpin peptidase inhibitor, clade A (alpha-1 antiproteinase, antitrypsin), member 1 (SERPINA1), transcript variant 2, mRNA [NM_001002236] |
| A_23_P40240 | 0,8870209 | down | -1,032386 | -0,0459824 | CTSZ | Homo sapiens cathepsin Z (CTSZ), mRNA [NM_001336] |
| A_23_P104798 | 0,39669368 | down | -1,2095851 | -0,2745122 | IL18 | Homo sapiens interleukin 18 (IL18), transcript variant 1, mRNA [NM_001562] |
| A_23_P157865 | 0,1928983 | down | -1,8881718 | -0,91699 | TNC | Homo sapiens tenascin C (TNC), mRNA [NM_002160] |
| A_23_P127948 | 0,01269069 | up | 3,4127927 | 1,7709527 | ADM | Homo sapiens adrenomedullin (ADM), mRNA [NM_001124] |
| A_23_P215790 | 0,5866154 | down | -1,1153003 | -0,1574322 | EGFR | Homo sapiens epidermal growth factor receptor (EGFR), transcript variant 1, mRNA [NM_005228] |
| A_23_P73114 | 0,69846743 | down | -1,2543359 | -0,3269237 | PROS1 | Homo sapiens protein S (alpha) (PROS1), mRNA [NM_000313] |
| A_23_P86021 | 0,89991564 | down | -1,0371997 | -0,0526937 | SELENBP1 | Homo sapiens selenium binding protein 1 (SELENBP1), transcript variant 1, mRNA [NM_003944] |
| A_23_P330070 | 0,83532095 | up | 1,06563 | 0,09170654 | TFPI | Homo sapiens tissue factor pathway inhibitor (lipoprotein-associated coagulation inhibitor) (TFPI), transcript variant 2, mRNA [NM_001032281] |
| A_23_P410507 | 0,13451456 | up | 1,2328987 | 0,3020543 | PSPN | Homo sapiens persephin (PSPN), mRNA [NM_004158] |
| A_32_P56392 | 0,68302476 | down | -1,0766046 | -0,1064885 | RBMX | Homo sapiens RNA binding motif protein, X-linked (RBMX), transcript variant 1, mRNA [NM_002139] |
| A_33_P3244843 | 0,59043044 | down | -1,0700521 | -0,0976811 | PRKAG3 | Homo sapiens protein kinase, AMP-activated, gamma 3 non-catalytic subunit (PRKAG3), mRNA [NM_017431] |
| A_32_P75094 | 0,10787725 | down | -1,464529 | -0,5504368 | AIFM2 | Homo sapiens apoptosis-inducing factor, mitochondrion-associated, 2 (AIFM2), transcript variant 2, mRNA [NM_032797] |
| A_23_P94501 | 0,6399172 | up | 1,1158763 | 0,15817712 | ANXA1 | Homo sapiens annexin A1 (ANXA1), mRNA [NM_000700] |
| A_23_P340848 | 0,14594871 | up | 1,7228177 | 0,78477 | PTGIR | Homo sapiens prostaglandin I2 (prostacyclin) receptor (IP) (PTGIR), mRNA [NM_000960] |
| A_23_P41114 | 0,15429671 | up | 2,3939152 | 1,259372 | CSTA | Homo sapiens cystatin A (stefin A) (CSTA), mRNA [NM_005213] |
| A_33_P3290709 | 0,5922341 | down | -1,1187489 | -0,1618863 | EGFL6 | Homo sapiens EGF-like-domain, multiple 6 (EGFL6), transcript variant 2, mRNA [NM_001167890] |
| A_32_P193646 | 0,32866108 | down | -1,1979955 | -0,2606225 | RBMX | Homo sapiens RNA binding motif protein, X-linked (RBMX), transcript variant 1, mRNA [NM_002139] |
| A_23_P430411 | 0,5226412 | up | 1,1144391 | 0,15631782 | ITGB2 | integrin, beta 2 (complement component 3 receptor 3 and 4 subunit) [Source:HGNC Symbol;Acc:HGNC:6155] [ENST00000610622] |
| A_23_P48455 | 0,1589503 | up | 1,2727822 | 0,34798557 | AMN | Homo sapiens amnion associated transmembrane protein (AMN), mRNA [NM_030943] |
| A_23_P405129 | 0,4559726 | up | 1,16988 | 0,22636059 | LTBP2 | Homo sapiens latent transforming growth factor beta binding protein 2 (LTBP2), mRNA [NM_000428] |
| A_23_P121533 | 0,04362308 | up | 2,347726 | 1,2312641 | SPON2 | Homo sapiens spondin 2, extracellular matrix protein (SPON2), transcript variant 1, mRNA [NM_012445] |
| A_24_P416645 | 0,07144744 | up | 6,230724 | 2,6393998 | KLK13 | Homo sapiens kallikrein-related peptidase 13 (KLK13), mRNA [NM_015596] |
| A_23_P113701 | 0,7615045 | down | -1,0813243 | -0,1127993 | PDGFA | Homo sapiens platelet-derived growth factor alpha polypeptide (PDGFA), transcript variant 1, mRNA [NM_002607] |
| A_23_P135769 | 0,17370348 | up | 1,265171 | 0,33933243 | ACTB | Homo sapiens actin, beta (ACTB), mRNA [NM_001101] |
| A_23_P114084 | 0,84269136 | up | 1,034021 | 0,04826551 | PHEX | Homo sapiens phosphate regulating endopeptidase homolog, X-linked (PHEX), transcript variant 1, mRNA [NM_000444] |
| A_23_P167997 | 0,8547732 | down | -1,054214 | -0,0761678 | HIST1H2BG | Homo sapiens histone cluster 1, H2bg (HIST1H2BG), mRNA [NM_003518] |
| A_32_P173662 | 0,3632822 | down | -1,4235327 | -0,5094757 | CRISP2 | Homo sapiens cysteine-rich secretory protein 2 (CRISP2), transcript variant 1, mRNA [NM_003296] |
| A_23_P10121 | 0,08714049 | up | 1,9815224 | 0,9866093 | SFRP1 | Homo sapiens secreted frizzled-related protein 1 (SFRP1), mRNA [NM_003012] |
| A_23_P114626 | 0,5678618 | down | -1,2241974 | -0,2918362 | SERPINC1 | Homo sapiens serpin peptidase inhibitor, clade C (antithrombin), member 1 (SERPINC1), mRNA [NM_000488] |
| A_33_P3270863 | 0,436978 | up | 1,249023 | 0,3208 | XDH | Homo sapiens xanthine dehydrogenase (XDH), mRNA [NM_000379] |
| A_23_P101655 | 0,42922792 | up | 1,143 | 0,1928254 | ACTN4 | Homo sapiens actinin, alpha 4 (ACTN4), mRNA [NM_004924] |
| A_23_P19590 | 0,44284067 | down | -1,1745548 | -0,2321141 | EZR | Homo sapiens ezrin (EZR), transcript variant 1, mRNA [NM_003379] |
| A_23_P115190 | 0,6717296 | up | 1,0715784 | 0,09973738 | NGF | Homo sapiens nerve growth factor (beta polypeptide) (NGF), mRNA [NM_002506] |
| A_23_P90130 | 0,20918906 | up | 1,2444544 | 0,31551334 | NAPSA | Homo sapiens napsin A aspartic peptidase (NAPSA), mRNA [NM_004851] |
| A_33_P3245228 | 0,79901814 | down | -1,3290297 | -0,4103733 | BPIFA1 | Homo sapiens BPI fold containing family A, member 1 (BPIFA1), transcript variant 2, mRNA [NM_130852] |
| A_23_P212617 | 0,6269167 | down | -1,1137285 | -0,1553976 | TFRC | Homo sapiens transferrin receptor (TFRC), transcript variant 1, mRNA [NM_003234] |
| A_23_P82296 | 0,83918583 | down | -1,0265962 | -0,0378688 | GNB2 | Homo sapiens guanine nucleotide binding protein (G protein), beta polypeptide 2 (GNB2), mRNA [NM_005273] |
| A_23_P40956 | 0,6868508 | down | -1,0738301 | -0,1027658 | GHRL | Homo sapiens ghrelin/obestatin prepropeptide (GHRL), transcript variant 1, mRNA [NM_016362] |
| A_23_P94230 | 0,3508384 | down | -1,2771966 | -0,3529807 | LY96 | Homo sapiens lymphocyte antigen 96 (LY96), transcript variant 1, mRNA [NM_015364] |
| A_23_P377299 | 0,16478647 | up | 1,2441227 | 0,31512883 | ECE2 | Homo sapiens endothelin converting enzyme 2 (ECE2), transcript variant 1, mRNA [NM_014693] |
| A_23_P161998 | 0,5103848 | down | -1,2376281 | -0,3075779 | HPX | Homo sapiens hemopexin (HPX), mRNA [NM_000613] |
| A_23_P215549 | 0,48103628 | up | 1,3168023 | 0,39703873 | PON3 | Homo sapiens paraoxonase 3 (PON3), mRNA [NM_000940] |
| A_23_P117298 | 0,11668842 | up | 1,3060594 | 0,38522047 | F7 | Homo sapiens coagulation factor VII (serum prothrombin conversion accelerator) (F7), transcript variant 1, mRNA [NM_000131] |
| A_23_P383118 | 0,22161703 | up | 1,2648332 | 0,33894715 | ZSWIM5 | Homo sapiens zinc finger, SWIM-type containing 5 (ZSWIM5), mRNA [NM_020883] |
| A_23_P216023 | 0,3785029 | up | 1,4029454 | 0,48845884 | ANGPT1 | Homo sapiens angiopoietin 1 (ANGPT1), transcript variant 1, mRNA [NM_001146] |
| A_23_P19663 | 0,15996595 | down | -4,041438 | -2,0148687 | CTGF | Homo sapiens connective tissue growth factor (CTGF), mRNA [NM_001901] |
| A_33_P3292854 | 0,22362651 | up | 1,2267722 | 0,29486737 | CALR | Homo sapiens calreticulin (CALR), mRNA [NM_004343] |
| A_33_P3212555 | 0,58404255 | down | -1,3213733 | -0,4020381 | PROS1 | Homo sapiens protein S (alpha) (PROS1), mRNA [NM_000313] |
| A_32_P466514 | 0,32102555 | up | 1,1794496 | 0,23811372 | IRF2BPL | Homo sapiens interferon regulatory factor 2 binding protein-like (IRF2BPL), mRNA [NM_024496] |
| A_24_P106542 | 0,49028128 | up | 1,1820017 | 0,24123211 | RSPO3 | Homo sapiens R-spondin 3 (RSPO3), mRNA [NM_032784] |
| A_24_P14464 | 0,18195735 | up | 1,5892023 | 0,6683028 | WFDC2 | Homo sapiens WAP four-disulfide core domain 2 (WFDC2), mRNA [NM_006103] |
| A_23_P102117 | 0,6226368 | down | -1,3027637 | -0,3815754 | WNT10A | Homo sapiens wingless-type MMTV integration site family, member 10A (WNT10A), mRNA [NM_025216] |
| A_23_P52761 | 0,17681557 | up | 2,1163561 | 1,0815824 | MMP7 | Homo sapiens matrix metallopeptidase 7 (matrilysin, uterine) (MMP7), mRNA [NM_002423] |

| | | | | | | |
|---|---|---|---|---|---|---|
| A_23_P71530 | 0,42888758 | down | -1,2236215 | -0,2911573 | TNFRSF11B | Homo sapiens tumor necrosis factor receptor superfamily, member 11b (TNFRSF11B), mRNA [NM_002546] |
| A_23_P110234 | 0,04974663 | down | -1,3478093 | -0,4306164 | CSN1S1 | Homo sapiens casein alpha s1 (CSN1S1), transcript variant 1, mRNA [NM_001890] |
| A_24_P141214 | 0,12606637 | down | -1,6287051 | -0,7037255 | STOM | Homo sapiens stomatin (STOM), transcript variant 2, mRNA [NM_198194] |
| A_23_P120316 | 0,33991688 | down | -1,3771346 | -0,4616695 | MTHFD2 | Homo sapiens methylenetetrahydrofolate dehydrogenase (NADP+ dependent) 2, methenyltetrahydrofolate cyclohydrolase (MTHFD2), transcript variant 1, mRNA [NM_0 |
| A_23_P121253 | 0,06613135 | up | 1,4319932 | 0,5180247 | TNFSF10 | Homo sapiens tumor necrosis factor (ligand) superfamily, member 10 (TNFSF10), transcript variant 1, mRNA [NM_003810] |
| A_23_P337800 | 0,7839056 | up | 1,06622 | 0,09250521 | IFNL1 | Homo sapiens interferon, lambda 1 (IFNL1), mRNA [NM_172140] |
| A_23_P370434 | 0,31468028 | down | -1,1833092 | -0,2428271 | C1QBP | Homo sapiens complement component 1, q subcomponent binding protein (C1QBP), mRNA [NM_001212] |
| A_33_P3375934 | 0,57824004 | up | 1,0929403 | 0,12821464 | NAMPT | Homo sapiens nicotinamide phosphoribosyltransferase (NAMPT), mRNA [NM_005746] |
| A_23_P122068 | 0,31232464 | up | 1,1948788 | 0,2568643 | C1QTNF3 | Homo sapiens C1q and tumor necrosis factor related protein 3 (C1QTNF3), transcript variant 2, mRNA [NM_181435] |
| A_33_P3237150 | 0,1257997 | up | 2,224309 | 1,1533571 | BMP2 | Homo sapiens bone morphogenetic protein 2 (BMP2), mRNA [NM_001200] |
| A_23_P78771 | 0,27773327 | up | 1,1640888 | 0,21920116 | NUCB1 | Homo sapiens nucleobindin 1 (NUCB1), mRNA [NM_006184] |
| A_24_P360269 | 0,64975023 | up | 1,1053336 | 0,14448181 | RNASET2 | Homo sapiens ribonuclease T2 (RNASET2), mRNA [NM_003730] |
| A_23_P202658 | 0,5021522 | up | 1,1170897 | 0,1597451 | GSTP1 | Homo sapiens glutathione S-transferase pi 1 (GSTP1), mRNA [NM_000852] |
| A_24_P239177 | 0,05825152 | up | 1,7636298 | 0,8185477 | MUC4 | Homo sapiens mucin 4, cell surface associated (MUC4), transcript variant 1, mRNA [NM_018406] |
| A_24_P870620 | 0,0525393 | down | -3,0150187 | -1,5921669 | PTN | Homo sapiens pleiotrophin (PTN), mRNA [NM_002825] |
| A_23_P101131 | 0,12604871 | up | 1,2785679 | 0,35452878 | GRP | Homo sapiens gastrin-releasing peptide (GRP), transcript variant 1, mRNA [NM_002091] |
| A_23_P203023 | 0,13577837 | down | -1,3265326 | -0,4076601 | RDX | Homo sapiens radixin (RDX), transcript variant 3, mRNA [NM_002906] |
| A_23_P39034 | 0,35876042 | up | 1,1656907 | 0,22118498 | SMARCA4 | Homo sapiens SWI/SNF related, matrix associated, actin dependent regulator of chromatin, subfamily a, member 4 (SMARCA4), transcript variant 3, mRNA [NM_003072 |
| A_33_P3354940 | 0,14443554 | up | 1,2737762 | 0,3491118 | CSF1 | Homo sapiens colony stimulating factor 1 (macrophage) (CSF1), transcript variant 1, mRNA [NM_000757] |
| A_23_P115261 | 0,87652117 | up | 1,0767058 | 0,10662412 | AGT | Homo sapiens angiotensinogen (serpin peptidase inhibitor, clade A, member 8) (AGT), mRNA [NM_000029] |
| A_33_P3397763 | 0,2873617 | up | 1,7145268 | 0,7778104 | TNFSF9 | Homo sapiens tumor necrosis factor (ligand) superfamily, member 9 (TNFSF9), mRNA [NM_003811] |
| A_23_P983 | 0,19014396 | up | 1,1931828 | 0,2548151 | PRDX6 | Homo sapiens peroxiredoxin 6 (PRDX6), mRNA [NM_004905] |
| A_33_P3329088 | 0,01875629 | up | 1,5881642 | 0,66736007 | PRSS8 | Homo sapiens protease, serine, 8 (PRSS8), mRNA [NM_002773] |
| A_33_P3394297 | 0,18468376 | up | 1,3702301 | 0,45441815 | THNSL2 | Homo sapiens threonine synthase-like 2 (S. cerevisiae) (THNSL2), transcript variant 2, mRNA [NM_001244676] |
| A_23_P41145 | 0,21351777 | up | 1,3822936 | 0,46706405 | FAM3D | Homo sapiens family with sequence similarity 3, member D (FAM3D), mRNA [NM_138805] |
| A_23_P31240 | 0,40469095 | up | 1,1493171 | 0,20077695 | GAL3ST4 | Homo sapiens galactose-3-O-sulfotransferase 4 (GAL3ST4), mRNA [NM_024637] |
| A_23_P12989 | 0,32762378 | down | -1,2933294 | -0,3710897 | PRDX5 | Homo sapiens peroxiredoxin 5 (PRDX5), transcript variant 1, mRNA [NM_012094] |
| A_23_P429950 | 0,0592426 | down | -2,8164542 | -1,49388 | KAL1 | Homo sapiens Kallmann syndrome 1 sequence (KAL1), mRNA [NM_000216] |
| A_33_P3332248 | 0,42634466 | up | 1,1473662 | 0,19832587 | ADRA1A | adrenoceptor alpha 1A [Source:HGNC Symbol;Acc:HGNC:277] [ENST00000380573] |
| A_23_P208302 | 0,02219906 | up | 1,9207166 | 0,94164467 | APOC2 | Homo sapiens apolipoprotein C-II (APOC2), mRNA [NM_000483] |
| A_23_P101054 | 0,06361125 | up | 2,5241742 | 1,3358115 | KRT34 | Homo sapiens keratin 34, type I (KRT34), mRNA [NM_021013] |
| A_23_P144071 | 0,66565204 | down | -1,2218152 | -0,2890261 | COL7A1 | Homo sapiens collagen, type VII, alpha 1 (COL7A1), mRNA [NM_000094] |
| A_23_P32500 | 0,07737384 | up | 1,56051 | 0,6420176 | STAB1 | Homo sapiens stabilin 1 (STAB1), mRNA [NM_015136] |
| A_23_P57829 | 0,36515465 | up | 1,1950673 | 0,25709185 | CPNE9 | Homo sapiens copine family member IX (CPNE9), mRNA [NM_153635] |
| A_23_P139912 | 0,28756794 | up | 2,499771 | 1,321796 | IGFBP6 | Homo sapiens insulin-like growth factor binding protein 6 (IGFBP6), mRNA [NM_002178] |
| A_24_P108451 | 0,29746452 | up | 1,1720688 | 0,2290573 | GPI | Homo sapiens glucose-6-phosphate isomerase (GPI), transcript variant 2, mRNA [NM_000175] |
| A_23_P111583 | 0,68210864 | down | -1,1558864 | -0,2089996 | CD36 | Homo sapiens CD36 molecule (thrombospondin receptor) (CD36), transcript variant 2, mRNA [NM_001001547] |
| A_23_P78099 | 0,19795217 | up | 1,3143129 | 0,3943088 | VTN | Homo sapiens vitronectin (VTN), mRNA [NM_000638] |
| A_23_P71037 | 0,0891633 | up | 1,5560391 | 0,6378783 | IL6 | Homo sapiens interleukin 6 (IL6), mRNA [NM_000600] |
| A_33_P3246829 | 0,3360464 | up | 1,829677 | 0,87158895 | IL1RN | Homo sapiens interleukin 1 receptor antagonist (IL1RN), transcript variant 4, mRNA [NM_173843] |
| A_33_P3229032 | 0,13520949 | up | 1,4541255 | 0,5401518 | CLEC11A | Homo sapiens C-type lectin domain family 11, member A (CLEC11A), mRNA [NM_002975] |
| A_23_P259207 | 0,3468186 | up | 1,70534 | 0,7700594 | THNSL2 | Homo sapiens threonine synthase-like 2 (S. cerevisiae) (THNSL2), transcript variant 1, mRNA [NM_018271] |
| A_24_P404840 | 0,33582428 | down | -1,2989664 | -0,3773641 | GJB1 | Homo sapiens gap junction protein, beta 1, 32kDa (GJB1), transcript variant 2, mRNA [NM_000166] |
| A_32_P25273 | 0,06712296 | down | -1,4692304 | -0,5550607 | HSPD1 | Homo sapiens heat shock 60kDa protein 1 (chaperonin) (HSPD1), transcript variant 1, mRNA [NM_002156] |
| A_33_P3340615 | 0,39955354 | up | 1,1239284 | 0,16855016 | PON1 | Homo sapiens paraoxonase 1 (PON1), mRNA [NM_000446] |
| A_32_P197524 | 0,7426789 | down | -1,1096257 | -0,1500731 | EIF2A | Homo sapiens eukaryotic translation initiation factor 2A, 65kDa (EIF2A), mRNA [NM_032025] |
| A_23_P345692 | 0,10754586 | down | -1,4497695 | -0,5358235 | IL17D | Homo sapiens interleukin 17D (IL17D), mRNA [NM_138284] |
| A_33_P3228271 | 0,5531498 | up | 1,1119932 | 0,15314795 | CST3 | Homo sapiens cystatin C (CST3), transcript variant 2, mRNA [NM_001288614] |
| A_33_P3246007 | 0,38756946 | up | 1,1087822 | 0,14897597 | APOA1BP | Homo sapiens apolipoprotein A-I binding protein (APOA1BP), mRNA [NM_144772] |
| A_33_P3233843 | 0,5016104 | down | -1,1420324 | -0,1916036 | IL6ST | Homo sapiens interleukin 6 signal transducer (IL6ST), transcript variant 3, mRNA [NM_001190981] |
| A_23_P113005 | 0,09982387 | up | 1,3912584 | 0,47639036 | EFNA1 | Homo sapiens ephrin-A1 (EFNA1), transcript variant 1, mRNA [NM_004428] |
| A_24_P79054 | 0,7819392 | up | 1,0673789 | 0,09407236 | TGFB1 | Homo sapiens transforming growth factor, beta 1 (TGFB1), mRNA [NM_000660] |
| A_23_P206280 | 0,13620989 | up | 1,2355384 | 0,3051398 | GPR56 | Homo sapiens G protein-coupled receptor 56 (GPR56), transcript variant 3, mRNA [NM_201525] |
| A_23_P258410 | 0,5757498 | up | 1,1149746 | 0,15701087 | WNT7A | Homo sapiens wingless-type MMTV integration site family, member 7A (WNT7A), mRNA [NM_004625] |
| A_23_P109133 | 0,7990355 | up | 1,0559553 | 0,07854875 | AVP | Homo sapiens arginine vasopressin (AVP), mRNA [NM_000490] |

| | | | | | | |
|---|---|---|---|---|---|---|
| A_32_P47701 | 0,2002439 | down | -1,2371444 | -0,3070138 | EEF1A1 | Homo sapiens eukaryotic translation elongation factor 1 alpha 1 (EEF1A1), mRNA [NM_001402] |
| A_23_P213699 | 0,03526172 | down | -2,1589885 | -1,1103555 | NRG2 | Homo sapiens neuregulin 2 (NRG2), transcript variant 3, mRNA [NM_013982] |
| A_23_P214330 | 0,5132956 | up | 1,3454096 | 0,42804548 | SERPINB1 | Homo sapiens serpin peptidase inhibitor, clade B (ovalbumin), member 1 (SERPINB1), transcript variant 1, mRNA [NM_030666] |
| A_23_P62659 | 0,5429938 | down | -1,0942506 | -0,1299431 | PPT1 | Homo sapiens palmitoyl-protein thioesterase 1 (PPT1), transcript variant 1, mRNA [NM_000310] |
| A_23_P122134 | 0,27229956 | down | -1,2396275 | -0,3099066 | NMUR2 | Homo sapiens neuromedin U receptor 2 (NMUR2), mRNA [NM_020167] |
| A_23_P145408 | 0,59058243 | down | -1,0871379 | -0,120535 | FUCA2 | Homo sapiens fucosidase, alpha-L- 2, plasma (FUCA2), mRNA [NM_032020] |
| A_33_P3269203 | 0,46912518 | down | -1,1780806 | -0,2364382 | SERPINH1 | Homo sapiens serpin peptidase inhibitor, clade H (heat shock protein 47), member 1, (collagen binding protein 1) (SERPINH1), transcript variant 1, mRNA [NM_001207[illegible] |
| A_23_P51217 | 0,4975513 | up | 1,2083925 | 0,27308914 | CLCA1 | Homo sapiens chloride channel accessory 1 (CLCA1), mRNA [NM_001285] |
| A_23_P207194 | 0,7919772 | up | 1,0485413 | 0,06838369 | GH1 | Homo sapiens growth hormone 1 (GH1), transcript variant 1, mRNA [NM_000515] |
| A_23_P126735 | 0,64095837 | up | 1,101736 | 0,1397785 | IL10 | Homo sapiens interleukin 10 (IL10), mRNA [NM_000572] |
| A_33_P3364582 | 0,8038665 | up | 1,0317755 | 0,04512906 | TNXB | Homo sapiens tenascin XB (TNXB), transcript variant XB, mRNA [NM_019105] |
| A_33_P3341499 | 0,64057386 | up | 1,1955292 | 0,2576494 | WNT5A | Homo sapiens wingless-type MMTV integration site family, member 5A (WNT5A), transcript variant 1, mRNA [NM_003392] |
| A_19_P00802759 | 0,57393587 | up | 1,0656302 | 0,09170687 | NUDT3 | nudix (nucleoside diphosphate linked moiety X)-type motif 3 [Source:HGNC Symbol;Acc:HGNC:8050] [ENST00000607016] |
| A_24_P206624 | 0,31431708 | down | -1,2659197 | -0,3401859 | FGFR2 | Homo sapiens fibroblast growth factor receptor 2 (FGFR2), transcript variant 2, mRNA [NM_022970] |
| A_23_P121926 | 0,2565482 | up | 1,5297077 | 0,613256 | SEPP1 | Homo sapiens selenoprotein P, plasma, 1 (SEPP1), transcript variant 1, mRNA [NM_005410] |
| A_23_P143526 | 0,20629066 | up | 1,6165016 | 0,6928749 | S100B | Homo sapiens S100 calcium binding protein B (S100B), mRNA [NM_006272] |
| A_23_P31816 | 0,70136887 | up | 1,2370001 | 0,30684564 | DEFA3 | Homo sapiens defensin, alpha 3, neutrophil-specific (DEFA3), mRNA [NM_005217] |
| A_33_P3261818 | 0,32061756 | down | -1,1466265 | -0,1973955 | MDH1 | Homo sapiens malate dehydrogenase 1, NAD (soluble) (MDH1), transcript variant 2, mRNA [NM_005917] |
| A_33_P3324004 | 0,12977064 | down | -2,1588674 | -1,1102746 | CCL15 | Homo sapiens chemokine (C-C motif) ligand 15 (CCL15), mRNA [NM_032965] |
| A_33_P3257728 | 0,2504973 | up | 1,2692962 | 0,34402874 | C1RL | Homo sapiens complement component 1, r subcomponent-like (C1RL), transcript variant 3, mRNA [NM_001297642] |
| A_23_P40174 | 0,44854614 | up | 1,8244399 | 0,8674536 | MMP9 | Homo sapiens matrix metallopeptidase 9 (gelatinase B, 92kDa gelatinase, 92kDa type IV collagenase) (MMP9), mRNA [NM_004994] |
| A_23_P155351 | 0,52252233 | down | -1,1446953 | -0,1949636 | BTD | Homo sapiens biotinidase (BTD), transcript variant 3, mRNA [NM_000060] |
| A_23_P153320 | 0,7261676 | up | 1,1353265 | 0,18310726 | ICAM1 | Homo sapiens intercellular adhesion molecule 1 (ICAM1), mRNA [NM_000201] |
| A_32_P162187 | 0,6074952 | up | 1,1579508 | 0,2115739 | C2 | Homo sapiens complement component 2 (C2), transcript variant 1, mRNA [NM_000063] |
| A_19_P00809660 | 0,22254838 | up | 1,2376976 | 0,30765888 | FGF13 | fibroblast growth factor 13 [Source:HGNC Symbol;Acc:HGNC:3670] [ENST00000421460] |
| A_23_P502312 | 0,27341995 | down | -1,6682338 | -0,7383214 | CD97 | Homo sapiens CD97 molecule (CD97), transcript variant 1, mRNA [NM_078481] |
| A_33_P3392245 | 0,6738635 | up | 1,085441 | 0,1182813 | FAM178A | Homo sapiens family with sequence similarity 178, member A (FAM178A), transcript variant 2, mRNA [NM_001136123] |
| A_33_P3279640 | 0,19153516 | down | -1,2933073 | -0,3710651 | HCN2 | Homo sapiens hyperpolarization activated cyclic nucleotide gated potassium channel 2 (HCN2), mRNA [NM_001194] |
| A_24_P289139 | 0,10480131 | up | 1,4361486 | 0,52220505 | SH3KBP1 | Homo sapiens SH3-domain kinase binding protein 1 (SH3KBP1), transcript variant 2, mRNA [NM_001024666] |
| A_33_P3248539 | 0,23519194 | up | 1,3323644 | 0,41398874 | LCN1 | Homo sapiens lipocalin 1 (LCN1), transcript variant 2, mRNA [NM_001252617] |
| A_23_P120883 | 0,45369956 | up | 1,247473 | 0,3190086 | HMOX1 | Homo sapiens heme oxygenase (decycling) 1 (HMOX1), mRNA [NM_002133] |
| A_23_P383009 | 0,23172355 | down | -2,0226507 | -1,0162472 | IGFBP5 | Homo sapiens insulin-like growth factor binding protein 5 (IGFBP5), mRNA [NM_000599] |
| A_32_P137939 | 0,3571411 | up | 1,1950469 | 0,25706723 | ACTB | Homo sapiens actin, beta (ACTB), mRNA [NM_001101] |
| A_24_P134319 | 0,6061912 | up | 1,0699311 | 0,09751796 | ADNP | Homo sapiens activity-dependent neuroprotector homeobox (ADNP), transcript variant 3, mRNA [NM_001282531] |
| A_23_P208493 | 0,31852493 | up | 1,557862 | 0,6395675 | LILRB2 | Homo sapiens leukocyte immunoglobulin-like receptor, subfamily B (with TM and ITIM domains), member 2 (LILRB2), transcript variant 1, mRNA [NM_005874] |
| A_23_P69497 | 0,45163035 | up | 1,1802026 | 0,23903455 | CLEC3B | Homo sapiens C-type lectin domain family 3, member B (CLEC3B), mRNA [NM_003278] |
| A_24_P245298 | 0,80384195 | up | 1,0492744 | 0,06939207 | TNFSF12 | Homo sapiens tumor necrosis factor (ligand) superfamily, member 12 (TNFSF12), transcript variant 1, mRNA [NM_003809] |
| A_23_P79518 | 0,5824083 | down | -1,5523683 | -0,6344709 | IL1B | Homo sapiens interleukin 1, beta (IL1B), mRNA [NM_000576] |
| A_23_P476 | 0,66028047 | up | 1,0617917 | 0,08650071 | MPZL1 | Homo sapiens myelin protein zero-like 1 (MPZL1), transcript variant 1, mRNA [NM_003953] |
| A_33_P3353242 | 0,17536698 | up | 1,3393894 | 0,4215755 | HSPB1 | Homo sapiens heat shock 27kDa protein 1 (HSPB1), mRNA [NM_001540] |
| A_23_P86599 | 0,8681354 | down | -1,1914651 | -0,2527368 | DMBT1 | Homo sapiens deleted in malignant brain tumors 1 (DMBT1), transcript variant 2, mRNA [NM_007329] |
| A_33_P3277373 | 0,85852623 | down | -1,0380911 | -0,053933 | OSM | Homo sapiens oncostatin M (OSM), mRNA [NM_020530] |
| A_23_P119943 | 0,15755913 | down | -1,4482676 | -0,5343282 | IGFBP2 | Homo sapiens insulin-like growth factor binding protein 2, 36kDa (IGFBP2), mRNA [NM_000597] |
| A_33_P3423570 | 0,537201 | down | -1,2184789 | -0,2850813 | METRN | Homo sapiens meteorin, glial cell differentiation regulator (METRN), mRNA [NM_024042] |
| A_23_P132793 | 0,47497037 | up | 1,1549281 | 0,20780303 | MANF | Homo sapiens mesencephalic astrocyte-derived neurotrophic factor (MANF), mRNA [NM_006010] |
| A_24_P148717 | 0,23587649 | up | 1,4262177 | 0,51219416 | CCR1 | Homo sapiens chemokine (C-C motif) receptor 1 (CCR1), mRNA [NM_001295] |
| A_24_P304423 | 0,5273208 | up | 1,2175506 | 0,28398177 | IGF1 | Homo sapiens insulin-like growth factor 1 (somatomedin C) (IGF1), transcript variant 4, mRNA [NM_000618] |
| A_23_P68910 | 0,33666354 | up | 1,2051014 | 0,26915452 | SSTR3 | Homo sapiens somatostatin receptor 3 (SSTR3), transcript variant 1, mRNA [NM_001051] |
| A_24_P9285 | 0,25274897 | down | -1,1870121 | -0,2473346 | LMAN2 | Homo sapiens lectin, mannose-binding 2 (LMAN2), mRNA [NM_006816] |
| A_23_P119478 | 0,40008697 | up | 1,2596227 | 0,33299166 | EBI3 | Homo sapiens Epstein-Barr virus induced 3 (EBI3), mRNA [NM_005755] |
| A_23_P153480 | 0,00814065 | up | 11,039466 | 3,4645984 | KLK5 | Homo sapiens kallikrein-related peptidase 5 (KLK5), transcript variant 1, mRNA [NM_012427] |
| A_23_P138680 | 0,34676954 | up | 1,1882101 | 0,24879 | IL15RA | Homo sapiens interleukin 15 receptor, alpha (IL15RA), transcript variant 2, mRNA [NM_172200] |
| A_33_P3685216 | 0,06927246 | up | 1,5196508 | 0,60373986 | A1BG | Homo sapiens alpha-1-B glycoprotein (A1BG), mRNA [NM_130786] |
| A_24_P940006 | 0,06353733 | down | -1,8672678 | -0,9009289 | EFNB3 | Homo sapiens ephrin-B3 (EFNB3), mRNA [NM_001406] |
| A_23_P7144 | 0,3847361 | down | -2,1387477 | -1,0967662 | CXCL1 | Homo sapiens chemokine (C-X-C motif) ligand 1 (melanoma growth stimulating activity, alpha) (CXCL1), transcript variant 1, mRNA [NM_001511] |

| | | | | | | |
|---|---|---|---|---|---|---|
| A_19_P00322757 | 0,29809073 | up | 1,211599 | 0,27691227 | MCF2L | MCF.2 cell line derived transforming sequence-like [Source:HGNC Symbol;Acc:HGNC:14576] [ENST00000442625] |
| A_24_P14731 | 0,1654251 | down | -1,3683724 | -0,4524609 | PCSK1N | Homo sapiens proprotein convertase subtilisin/kexin type 1 inhibitor (PCSK1N), mRNA [NM_013271] |
| A_23_P114883 | 0,08924329 | down | -3,5694208 | -1,83569 | FMOD | Homo sapiens fibromodulin (FMOD), transcript variant 1, mRNA [NM_002023] |
| A_23_P395954 | 0,17512895 | up | 1,2851164 | 0,36189908 | SSH2 | slingshot protein phosphatase 2 [Source:HGNC Symbol;Acc:HGNC:30580] [ENST00000394848] |
| A_23_P253791 | 0,00369171 | down | -2,6437979 | -1,4026119 | CAMP | Homo sapiens cathelicidin antimicrobial peptide (CAMP), mRNA [NM_004345] |
| A_23_P203488 | 0,5663369 | up | 1,1242197 | 0,16892394 | SMPD1 | Homo sapiens sphingomyelin phosphodiesterase 1, acid lysosomal (SMPD1), transcript variant 1, mRNA [NM_000543] |
| A_23_P215484 | 0,5059615 | down | -1,460176 | -0,5461423 | CCL26 | Homo sapiens chemokine (C-C motif) ligand 26 (CCL26), mRNA [NM_006072] |
| A_24_P242646 | 0,2975158 | down | -1,2483668 | -0,3200419 | CTSS | Homo sapiens cathepsin S (CTSS), transcript variant 1, mRNA [NM_004079] |
| A_23_P155765 | 0,33783665 | down | -1,3977178 | -0,4830732 | HMGB2 | Homo sapiens high mobility group box 2 (HMGB2), transcript variant 1, mRNA [NM_002129] |
| A_23_P23611 | 0,7065537 | up | 1,1034755 | 0,14205453 | AMY1C | Homo sapiens amylase, alpha 1C (salivary) (AMY1C), mRNA [NM_001008219] |
| A_23_P35092 | 0,33100364 | up | 1,5995077 | 0,6776279 | IL19 | Homo sapiens interleukin 19 (IL19), transcript variant 1, mRNA [NM_153758] |
| A_33_P3225268 | 0,02080355 | up | 1,4273466 | 0,5133357 | QSOX1 | Homo sapiens quiescin Q6 sulfhydryl oxidase 1 (QSOX1), transcript variant 2, mRNA [NM_001004128] |
| A_32_P156963 | 0,88690597 | up | 1,032398 | 0,04599923 | ACTG1 | Homo sapiens actin gamma 1 (ACTG1), transcript variant 2, mRNA [NM_001614] |
| A_33_P3397288 | 0,55481744 | down | -1,1128007 | -0,1541953 | EDN3 | Homo sapiens endothelin 3 (EDN3), transcript variant 2, mRNA [NM_207032] |
| A_33_P3238166 | 0,45189866 | up | 1,5682505 | 0,64915603 | PXDN | Homo sapiens peroxidasin (PXDN), mRNA [NM_012293] |
| A_23_P218646 | 0,25574556 | up | 1,9668758 | 0,97590584 | TNFRSF6B | Homo sapiens tumor necrosis factor receptor superfamily, member 6b, decoy (TNFRSF6B), mRNA [NM_003823] |
| A_23_P101950 | 0,29523283 | down | -1,1611546 | -0,2155601 | MDH1 | Homo sapiens malate dehydrogenase 1, NAD (soluble) (MDH1), transcript variant 2, mRNA [NM_005917] |
| A_23_P385690 | 0,51780844 | down | -1,170818 | -0,2275168 | WNT3A | Homo sapiens wingless-type MMTV integration site family, member 3A (WNT3A), mRNA [NM_033131] |
| A_33_P3359683 | 0,6520237 | up | 1,0820146 | 0,11371991 | IL16 | Homo sapiens interleukin 16 (IL16), transcript variant 3, mRNA [NM_001172128] |
| A_24_P236091 | 0,0402301 | down | -2,8291862 | -1,5003871 | ENO2 | Homo sapiens enolase 2 (gamma, neuronal) (ENO2), mRNA [NM_001975] |
| A_33_P3283480 | 0,2002148 | up | 1,202815 | 0,26641482 | CTSC | Homo sapiens cathepsin C (CTSC), transcript variant 2, mRNA [NM_148170] |
| A_33_P3369371 | 0,11417031 | down | -1,5175962 | -0,601788 | GPX3 | Homo sapiens glutathione peroxidase 3 (plasma) (GPX3), mRNA [NM_002084] |
| A_33_P3256778 | 0,20925272 | down | -1,5347852 | -0,6180367 | CNTF | Homo sapiens ciliary neurotrophic factor (CNTF), mRNA [NM_000614] |
| A_23_P215634 | 0,23609251 | up | 1,7874928 | 0,83793736 | IGFBP3 | Homo sapiens insulin-like growth factor binding protein 3 (IGFBP3), transcript variant 1, mRNA [NM_001013398] |
| A_24_P322771 | 0,37494272 | up | 1,7275747 | 0,7887481 | TFF1 | Homo sapiens trefoil factor 1 (TFF1), mRNA [NM_003225] |
| A_23_P162322 | 0,46904725 | up | 1,2000822 | 0,2631332 | WNT10B | Homo sapiens wingless-type MMTV integration site family, member 10B (WNT10B), mRNA [NM_003394] |
| A_23_P91619 | 0,39663357 | up | 1,1729873 | 0,23018745 | MIF | Homo sapiens macrophage migration inhibitory factor (glycosylation-inhibiting factor) (MIF), mRNA [NM_002415] |
| A_24_P46130 | 0,6979519 | up | 1,1943941 | 0,25627896 | ACPP | Homo sapiens acid phosphatase, prostate (ACPP), transcript variant 1, mRNA [NM_001099] |
| A_24_P364296 | 0,03997383 | down | -3,3344965 | -1,737469 | STX2 | Homo sapiens syntaxin 2 (STX2), transcript variant 1, mRNA [NM_001980] |
| A_33_P3395513 | 0,81235105 | up | 1,0458759 | 0,06471168 | NRG2 | Homo sapiens cDNA FLJ42513 fis, clone BRACE2046295, highly similar to Pro-neuregulin-2, membrane-bound isoform precursor. [AK124504] |
| A_23_P8913 | 0,16434136 | up | 2,4603426 | 1,2988592 | CA2 | Homo sapiens carbonic anhydrase II (CA2), transcript variant 1, mRNA [NM_000067] |
| A_23_P93311 | 0,20194982 | down | -1,2143598 | -0,2801959 | DDR1 | Homo sapiens discoidin domain receptor tyrosine kinase 1 (DDR1), transcript variant 2, mRNA [NM_013993] |
| A_23_P89587 | 0,7006806 | down | -1,0842581 | -0,1167082 | WNT9B | Homo sapiens wingless-type MMTV integration site family, member 9B (WNT9B), mRNA [NM_003396] |
| A_24_P401174 | 0,74820644 | down | -1,0824169 | -0,1142563 | KIAA0556 | Homo sapiens KIAA0556 (KIAA0556), mRNA [NM_015202] |
| A_33_P3351944 | 0,81249005 | down | -1,0456939 | -0,0644606 | EGFR | Homo sapiens epidermal growth factor receptor (EGFR), transcript variant 3, mRNA [NM_201283] |
| A_23_P103104 | 0,57359886 | up | 1,1330259 | 0,18018082 | MFNG | Homo sapiens MFNG O-fucosylpeptide 3-beta-N-acetylglucosaminyltransferase (MFNG), transcript variant 1, mRNA [NM_002405] |
| A_24_P287189 | 0,03805355 | down | -1,4971358 | -0,5822051 | TOLLIP | Homo sapiens toll interacting protein (TOLLIP), mRNA [NM_019009] |
| A_23_P329573 | 0,06282665 | up | 1,8198986 | 0,86385804 | ITGB2 | Homo sapiens integrin, beta 2 (complement component 3 receptor 3 and 4 subunit) (ITGB2), transcript variant 1, mRNA [NM_000211] |
| A_33_P3286278 | 0,01517153 | up | 1,6282812 | 0,7033499 | GRN | Homo sapiens granulin (GRN), mRNA [NM_002087] |
| A_23_P104464 | 0,2516819 | up | 1,4361011 | 0,5221573 | ALOX5 | Homo sapiens arachidonate 5-lipoxygenase (ALOX5), transcript variant 1, mRNA [NM_000698] |
| A_23_P428887 | 0,05947252 | up | 1,5998344 | 0,6779226 | KLHL34 | Homo sapiens kelch-like family member 34 (KLHL34), mRNA [NM_153270] |
| A_33_P3388870 | 0,6317166 | up | 1,0712426 | 0,0992852 | BLOC1S1 | Homo sapiens biogenesis of lysosomal organelles complex-1, subunit 1 (BLOC1S1), transcript variant 1, mRNA [NM_001487] |
| A_24_P129417 | 0,321708 | up | 1,4583763 | 0,544363 | BMP1 | Homo sapiens bone morphogenetic protein 1 (BMP1), transcript variant 1, mRNA [NM_001199] |
| A_24_P390495 | 0,5144565 | down | -1,2989109 | -0,3773024 | CX3CL1 | chemokine (C-X3-C motif) ligand 1 [Source:HGNC Symbol;Acc:HGNC:10647] [ENST00000006053] |
| A_33_P3322363 | 0,26222402 | down | -1,3681809 | -0,4522589 | HMSD | Homo sapiens histocompatibility (minor) serpin domain containing (HMSD), mRNA [NM_001123366] |
| A_23_P51039 | 0,29872593 | up | 1,2239316 | 0,29152286 | INHA | Homo sapiens inhibin, alpha (INHA), mRNA [NM_002191] |
| A_33_P3405213 | 0,8029848 | down | -1,091461 | -0,1262605 | PECAM1 | Homo sapiens platelet/endothelial cell adhesion molecule 1 (PECAM1), mRNA [NM_000442] |
| A_24_P202497 | 0,13624804 | down | -1,2378609 | -0,3078492 | TWSG1 | Homo sapiens twisted gastrulation BMP signaling modulator 1 (TWSG1), mRNA [NM_020648] |
| A_23_P105833 | 0,8761674 | up | 1,0212798 | 0,03037819 | BIVM | Homo sapiens basic, immunoglobulin-like variable motif containing (BIVM), transcript variant 1, mRNA [NM_017693] |
| A_24_P784765 | 0,24952076 | up | 1,3192157 | 0,3996804 | CD59 | Homo sapiens CD59 molecule, complement regulatory protein (CD59), transcript variant 1, mRNA [NM_203330] |
| A_33_P3367692 | 0,66598994 | up | 1,0987316 | 0,13583905 | CFH | Homo sapiens complement factor H (CFH), transcript variant 2, mRNA [NM_001014975] |
| A_23_P44932 | 0,2467943 | down | -1,181934 | -0,2411495 | EIF2A | Homo sapiens eukaryotic translation initiation factor 2A, 65kDa (EIF2A), mRNA [NM_032025] |
| A_23_P69586 | 0,8172932 | up | 1,0860174 | 0,11904718 | FAT1 | Homo sapiens FAT atypical cadherin 1 (FAT1), mRNA [NM_005245] |
| A_23_P159721 | 0,65253556 | up | 1,0961901 | 0,132498 | GPR50 | Homo sapiens G protein-coupled receptor 50 (GPR50), mRNA [NM_004224] |
| A_23_P258246 | 0,18083712 | down | -1,1736488 | -0,2310008 | DDB1 | Homo sapiens damage-specific DNA binding protein 1, 127kDa (DDB1), mRNA [NM_001923] |

| | | | | | | |
|---|---|---|---|---|---|---|
| A_23_P61127 | 0,14308597 | down | -1,3746271 | -0,4590403 | APOO | Homo sapiens apolipoprotein O (APOO), transcript variant 1, mRNA [NM_024122] |
| A_23_P15146 | 0,3371758 | up | 1,4741156 | 0,5598497 | IL32 | Homo sapiens interleukin 32 (IL32), transcript variant 1, mRNA [NM_001012631] |
| A_23_P7212 | 0,0613879 | up | 2,0259044 | 1,0185661 | CFI | Homo sapiens complement factor I (CFI), mRNA [NM_000204] |
| A_33_P3697530 | 0,8358786 | down | -1,0470719 | -0,0663606 | SEMA4D | Homo sapiens sema domain, immunoglobulin domain (Ig), transmembrane domain (TM) and short cytoplasmic domain, (semaphorin) 4D (SEMA4D), transcript variant 2 |
| A_33_P3233040 | 0,65024674 | up | 1,244118 | 0,3151233 | SERPINB11 | Homo sapiens serpin peptidase inhibitor, clade B (ovalbumin), member 11 (gene/pseudogene) (SERPINB11), transcript variant 1, mRNA [NM_080475] |
| A_23_P66608 | 0,6576935 | up | 1,1019773 | 0,14009456 | KAT2A | Homo sapiens K(lysine) acetyltransferase 2A (KAT2A), mRNA [NM_021078] |
| A_23_P202448 | 0,35407692 | up | 1,3065336 | 0,38574418 | CXCL12 | Homo sapiens chemokine (C-X-C motif) ligand 12 (CXCL12), transcript variant 1, mRNA [NM_199168] |
| A_23_P156061 | 0,11683965 | up | 1,3434765 | 0,42597112 | LNPEP | Homo sapiens leucyl/cystinyl aminopeptidase (LNPEP), transcript variant 1, mRNA [NM_005575] |
| A_33_P3368453 | 0,1708443 | down | -1,2940586 | -0,3719029 | MICA | Homo sapiens MHC class I polypeptide-related sequence A (MICA), transcript variant 1*001, mRNA [NM_000247] |
| A_33_P3377194 | 0,22085142 | up | 1,1943074 | 0,25617427 | ADRA1A | Homo sapiens adrenoceptor alpha 1A (ADRA1A), transcript variant 4, mRNA [NM_033304] |
| A_23_P84219 | 0,65930766 | up | 1,1108428 | 0,1516547 | LIPH | Homo sapiens lipase, member H (LIPH), mRNA [NM_139248] |
| A_23_P30884 | 0,44521636 | down | -1,1021602 | -0,140334 | CLIC1 | Homo sapiens chloride intracellular channel 1 (CLIC1), transcript variant 2, mRNA [NM_001288] |
| A_24_P192994 | 0,06294756 | down | -2,3753576 | -1,2481447 | FADS1 | Homo sapiens fatty acid desaturase 1 (FADS1), mRNA [NM_013402] |
| A_32_P223777 | 0,47033617 | down | -1,0932158 | -0,1285783 | IL6ST | Homo sapiens interleukin 6 signal transducer (IL6ST), transcript variant 1, mRNA [NM_002184] |
| A_23_P361544 | 0,033437 | down | -2,158292 | -1,1098901 | CES4A | Homo sapiens carboxylesterase 4A (CES4A), transcript variant 1, mRNA [NM_173815] |
| A_23_P11995 | 0,25228143 | down | -1,2261479 | -0,294133 | PRDX1 | Homo sapiens peroxiredoxin 1 (PRDX1), transcript variant 1, mRNA [NM_002574] |
| A_33_P3233841 | 0,13534483 | down | -1,2694143 | -0,344163 | IL6ST | Homo sapiens interleukin 6 signal transducer (IL6ST), transcript variant 1, mRNA [NM_002184] |
| A_23_P57364 | 0,83465654 | down | -1,0531974 | -0,0747758 | TFF2 | Homo sapiens trefoil factor 2 (TFF2), mRNA [NM_005423] |
| A_33_P3380618 | 0,63125396 | up | 1,0821394 | 0,11388632 | HSPG2 | Homo sapiens heparan sulfate proteoglycan 2 (HSPG2), transcript variant 1, mRNA [NM_001291860] |
| A_23_P32125 | 0,8740183 | down | -1,020747 | -0,0296253 | PMPCA | Homo sapiens peptidase (mitochondrial processing) alpha (PMPCA), transcript variant 1, mRNA [NM_015160] |
| A_33_P3258392 | 0,05227085 | up | 2,0880384 | 1,0621482 | EDN1 | Homo sapiens endothelin 1 (EDN1), transcript variant 1, mRNA [NM_001955] |
| A_24_P261169 | 0,58781123 | down | -1,1451144 | -0,1954918 | SEMA4D | Homo sapiens sema domain, immunoglobulin domain (Ig), transmembrane domain (TM) and short cytoplasmic domain, (semaphorin) 4D (SEMA4D), transcript variant 1 |
| A_23_P39465 | 0,684927 | up | 1,1616111 | 0,21612711 | BST2 | Homo sapiens bone marrow stromal cell antigen 2 (BST2), mRNA [NM_004335] |
| A_23_P8961 | 0,8162647 | down | -1,0827469 | -0,114696 | IL7 | Homo sapiens interleukin 7 (IL7), transcript variant 1, mRNA [NM_000880] |
| A_23_P14774 | 0,8109899 | up | 1,0448828 | 0,0633411 | CTSH | Homo sapiens cathepsin H (CTSH), mRNA [NM_004390] |
| A_23_P435407 | 0,02313798 | down | -1,7620481 | -0,8172533 | GPC4 | Homo sapiens glypican 4 (GPC4), mRNA [NM_001448] |
| A_33_P3254121 | 0,6617376 | up | 1,0702267 | 0,09791639 | RNASET2 | ribonuclease T2 [Source:HGNC Symbol;Acc:HGNC:21686] [ENST00000358165] |
| A_23_P168828 | 0,6258411 | up | 1,1687083 | 0,22491492 | KLF10 | Homo sapiens Kruppel-like factor 10 (KLF10), transcript variant 1, mRNA [NM_005655] |
| A_23_P162918 | 0,85682446 | up | 1,0543584 | 0,0763653 | SERPINA3 | Homo sapiens serpin peptidase inhibitor, clade A (alpha-1 antiproteinase, antitrypsin), member 3 (SERPINA3), mRNA [NM_001085] |
| A_23_P157361 | 0,09985998 | down | -1,7402253 | -0,7992741 | WDR60 | Homo sapiens WD repeat domain 60 (WDR60), mRNA [NM_018051] |
| A_33_P3303542 | 0,33412525 | down | -1,2668612 | -0,3412585 | SSC5D | Homo sapiens scavenger receptor cysteine rich family, 5 domains (SSC5D), transcript variant 1, mRNA [NM_001144950] |
| A_33_P3422822 | 0,03924826 | down | -2,21264 | -1,1457688 | GJC2 | Homo sapiens gap junction protein, gamma 2, 47kDa (GJC2), mRNA [NM_020435] |
| A_23_P162171 | 0,25044632 | down | -1,7549609 | -0,8114389 | MCAM | Homo sapiens melanoma cell adhesion molecule (MCAM), mRNA [NM_006500] |
| A_23_P15734 | 0,1571636 | up | 1,487235 | 0,57263255 | KRT9 | Homo sapiens keratin 9, type I (KRT9), mRNA [NM_000226] |
| A_33_P3286916 | 0,52520806 | up | 1,1290303 | 0,17508426 | PDZD7 | Homo sapiens PDZ domain containing 7 (PDZD7), transcript variant 1, mRNA [NM_001195263] |
| A_33_P3321657 | 0,5998873 | up | 1,0976064 | 0,13436082 | HSPG2 | Homo sapiens heparan sulfate proteoglycan 2 (HSPG2), transcript variant 1, mRNA [NM_001291860] |
| A_23_P68487 | 0,09134058 | up | 2,0375254 | 1,026818 | BMP7 | Homo sapiens bone morphogenetic protein 7 (BMP7), mRNA [NM_001719] |
| A_33_P3349045 | 0,36414886 | up | 1,3198246 | 0,4003462 | IL4R | Homo sapiens interleukin 4 receptor (IL4R), transcript variant 4, mRNA [NM_001257407] |
| A_23_P73589 | 0,74906224 | up | 1,1127942 | 0,15418676 | MSN | Homo sapiens moesin (MSN), mRNA [NM_002444] |
| A_33_P3233834 | 0,56509316 | down | -1,0904009 | -0,1248587 | IL6ST | Homo sapiens interleukin 6 signal transducer (IL6ST), transcript variant 3, mRNA [NM_001190981] |
| A_23_P150053 | 0,5122088 | down | -1,334086 | -0,4158516 | ACTA2 | Homo sapiens actin, alpha 2, smooth muscle, aorta (ACTA2), transcript variant 2, mRNA [NM_001613] |
| A_33_P3405839 | 0,8401941 | down | -1,0309243 | -0,0439384 | TPO | Homo sapiens thyroid peroxidase (TPO), transcript variant 5, mRNA [NM_175722] |
| A_23_P160559 | 0,04083037 | up | 8,362579 | 3,063948 | ECM1 | Homo sapiens extracellular matrix protein 1 (ECM1), transcript variant 1, mRNA [NM_004425] |
| A_24_P38276 | 0,12624162 | up | 1,250821 | 0,32287532 | FZD1 | Homo sapiens frizzled class receptor 1 (FZD1), mRNA [NM_003505] |
| A_33_P3243857 | 0,48866478 | up | 1,1508176 | 0,20265923 | ADAM10 | Homo sapiens ADAM metallopeptidase domain 10 (ADAM10), mRNA [NM_001110] |
| A_24_P88763 | 0,06695586 | up | 1,3922687 | 0,47743762 | LOXL3 | Homo sapiens lysyl oxidase-like 3 (LOXL3), transcript variant 1, mRNA [NM_032603] |
| A_23_P97990 | 0,17067678 | down | -1,2656925 | -0,3399269 | HTRA1 | Homo sapiens HtrA serine peptidase 1 (HTRA1), mRNA [NM_002775] |
| A_24_P911607 | 0,08986841 | up | 1,4645975 | 0,5505042 | WNT7B | Homo sapiens wingless-type MMTV integration site family, member 7B (WNT7B), mRNA [NM_058238] |
| A_33_P3363168 | 0,45477805 | up | 1,109507 | 0,14991872 | SSH2 | Homo sapiens slingshot protein phosphatase 2 (SSH2), transcript variant 1, mRNA [NM_001282129] |
| A_23_P376088 | 0,32648492 | up | 1,2024835 | 0,26601714 | LIME1 | Homo sapiens Lck interacting transmembrane adaptor 1 (LIME1), mRNA [NM_017806] |
| A_23_P130149 | 0,34265518 | up | 1,2961763 | 0,37426198 | ENO3 | Homo sapiens enolase 3 (beta, muscle) (ENO3), transcript variant 1, mRNA [NM_001976] |
| A_33_P3241369 | 0,27644363 | down | -1,3377208 | -0,419777 | UCN3 | Homo sapiens urocortin 3 (UCN3), mRNA [NM_053049] |
| A_33_P3822503 | 0,08374878 | down | -1,5315939 | -0,6150338 | CTF1 | Homo sapiens cardiotrophin 1 (CTF1), transcript variant 1, mRNA [NM_001330] |
| A_33_P3362371 | 0,5196997 | down | -1,0783786 | -0,1088637 | RTN3 | Homo sapiens reticulon 3 (RTN3), transcript variant 7, mRNA [NM_001265591] |
| A_23_P113351 | 0,05453372 | down | -2,0422482 | -1,0301583 | SPARCL1 | Homo sapiens SPARC-like 1 (hevin) (SPARCL1), transcript variant 2, mRNA [NM_004684] |

| | | | | | | |
|---|---|---|---|---|---|---|
| A_33_P3284345 | 0,80258954 | down | -1,1063051 | -0,1457494 | NRG1 | Homo sapiens neuregulin 1 (NRG1), transcript variant HRG-gamma, mRNA [NM_004495] |
| A_23_P133474 | 0,22809277 | down | -1,3125782 | -0,3924034 | GPX3 | Homo sapiens glutathione peroxidase 3 (plasma) (GPX3), mRNA [NM_002084] |
| A_23_P94800 | 0,20810027 | up | 1,3976902 | 0,4830446 | S100A4 | Homo sapiens S100 calcium binding protein A4 (S100A4), transcript variant 1, mRNA [NM_002961] |
| A_33_P3381338 | 0,05035963 | up | 1,4039744 | 0,48951665 | TNXB | Homo sapiens tenascin XB (TNXB), transcript variant XB, mRNA [NM_019105] |
| A_23_P149529 | 0,37922758 | up | 1,1712219 | 0,22801438 | TACSTD2 | Homo sapiens tumor-associated calcium signal transducer 2 (TACSTD2), mRNA [NM_002353] |
| A_23_P136978 | 0,22774617 | up | 1,1971023 | 0,25954646 | SRPX2 | Homo sapiens sushi-repeat containing protein, X-linked 2 (SRPX2), mRNA [NM_014467] |
| A_23_P421423 | 0,12046151 | up | 1,6655777 | 0,7360226 | TNFAIP2 | Homo sapiens tumor necrosis factor, alpha-induced protein 2 (TNFAIP2), mRNA [NM_006291] |
| A_23_P204947 | 0,81850153 | up | 1,1676795 | 0,2236444 | GJB2 | Homo sapiens gap junction protein, beta 2, 26kDa (GJB2), mRNA [NM_004004] |
| A_23_P55828 | 0,1285538 | down | -1,7675353 | -0,821739 | CCL25 | Homo sapiens chemokine (C-C motif) ligand 25 (CCL25), transcript variant 1, mRNA [NM_005624] |
| A_33_P3246885 | 0,88564456 | up | 1,0692121 | 0,09654804 | DMKN | Homo sapiens dermokine (DMKN), transcript variant 9, mRNA [NM_001190348] |
| A_23_P16523 | 0,8482265 | up | 1,0650535 | 0,09092585 | GDF15 | Homo sapiens growth differentiation factor 15 (GDF15), mRNA [NM_004864] |
| A_23_P201636 | 0,12453916 | down | -1,5243695 | -0,6082126 | LAMC2 | Homo sapiens laminin, gamma 2 (LAMC2), transcript variant 1, mRNA [NM_005562] |
| A_23_P254594 | 0,5337992 | up | 1,1434195 | 0,1933548 | GNRH1 | Homo sapiens gonadotropin-releasing hormone 1 (luteinizing-releasing hormone) (GNRH1), transcript variant 1, mRNA [NM_000825] |
| A_23_P57036 | 0,85995346 | down | -1,050379 | -0,07091 | CD40 | Homo sapiens CD40 molecule, TNF receptor superfamily member 5 (CD40), transcript variant 1, mRNA [NM_001250] |
| A_23_P18452 | 0,11056153 | up | 3,9218621 | 1,9715388 | CXCL9 | Homo sapiens chemokine (C-X-C motif) ligand 9 (CXCL9), mRNA [NM_002416] |
| A_23_P155979 | 0,4575603 | up | 1,2863446 | 0,36327723 | EGF | Homo sapiens epidermal growth factor (EGF), transcript variant 1, mRNA [NM_001963] |
| A_23_P256413 | 0,11599319 | down | -1,396649 | -0,4819695 | CMTM7 | Homo sapiens CKLF-like MARVEL transmembrane domain containing 7 (CMTM7), transcript variant 1, mRNA [NM_138410] |
| A_23_P59452 | 0,18332952 | up | 1,5364524 | 0,6196031 | AOC1 | Homo sapiens amine oxidase, copper containing 1 (AOC1), transcript variant 2, mRNA [NM_001091] |
| A_33_P3326634 | 0,50584185 | up | 1,4328234 | 0,5188608 | GPC3 | Homo sapiens glypican 3 (GPC3), transcript variant 1, mRNA [NM_001164617] |
| A_23_P137948 | 0,33624107 | up | 1,1513702 | 0,20335174 | NENF | Homo sapiens neudesin neurotrophic factor (NENF), transcript variant 1, mRNA [NM_013349] |
| A_23_P312300 | 0,18774483 | down | -2,688576 | -1,4268422 | SCGB2A1 | Homo sapiens secretoglobin, family 2A, member 1 (SCGB2A1), mRNA [NM_002407] |
| A_33_P3402489 | 0,36172146 | up | 1,3461794 | 0,42887065 | OAS3 | Homo sapiens 2'-5'-oligoadenylate synthetase 3, 100kDa (OAS3), mRNA [NM_006187] |
| A_32_P95739 | 0,5352109 | up | 1,1238015 | 0,16838719 | TPI1 | Homo sapiens triosephosphate isomerase 1 (TPI1), transcript variant 1, mRNA [NM_000365] |
| A_23_P412321 | 0,527566 | up | 1,2211975 | 0,28829652 | CCR5 | Homo sapiens chemokine (C-C motif) receptor 5 (gene/pseudogene) (CCR5), transcript variant A, mRNA [NM_000579] |
| A_32_P226149 | 0,7400109 | up | 1,056493 | 0,07928327 | YWHAZ | Homo sapiens tyrosine 3-monooxygenase/tryptophan 5-monooxygenase activation protein, zeta (YWHAZ), transcript variant 2, mRNA [NM_145690] |
| A_23_P215060 | 0,65336657 | down | -1,114461 | -0,1563461 | PODXL | Homo sapiens podocalyxin-like (PODXL), transcript variant 1, mRNA [NM_001018111] |
| A_23_P215913 | 0,17409953 | up | 1,5496256 | 0,63191974 | CLU | Homo sapiens clusterin (CLU), transcript variant 1, mRNA [NM_001831] |
| A_33_P3368452 | 0,23594923 | down | -1,2225975 | -0,2899495 | MICA | Homo sapiens MHC class I polypeptide-related sequence A (MICA), transcript variant 1*001, mRNA [NM_000247] |
| A_23_P135248 | 0,5316484 | down | -1,1062382 | -0,1456621 | CCL27 | Homo sapiens chemokine (C-C motif) ligand 27 (CCL27), transcript variant 1, mRNA [NM_006664] |
| A_23_P83328 | 0,4088448 | up | 1,1786308 | 0,23711191 | ENG | Homo sapiens endoglin (ENG), transcript variant 2, mRNA [NM_000118] |
| A_23_P79999 | 0,6024607 | up | 1,0743878 | 0,10351481 | ENTPD6 | Homo sapiens ectonucleoside triphosphate diphosphohydrolase 6 (putative) (ENTPD6), transcript variant 1, mRNA [NM_001247] |
| A_23_P64372 | 0,422513 | up | 1,334031 | 0,41579217 | TCN1 | Homo sapiens transcobalamin I (vitamin B12 binding protein, R binder family) (TCN1), mRNA [NM_001062] |
| A_33_P3296687 | 0,20711632 | up | 1,5016963 | 0,5865931 | KRT33A | Homo sapiens keratin 33A, type I (KRT33A), mRNA [NM_004138] |
| A_33_P3223592 | 0,05829652 | up | 2,1168365 | 1,0819099 | APOE | Homo sapiens apolipoprotein E (APOE), transcript variant 1, mRNA [NM_001302688] |
| A_24_P314451 | 0,246809 | down | -1,3585433 | -0,4420605 | F8 | Homo sapiens coagulation factor VIII, procoagulant component (F8), transcript variant 1, mRNA [NM_000132] |
| A_23_P81825 | 0,37328508 | up | 1,2457545 | 0,31701976 | GUCA1B | Homo sapiens guanylate cyclase activator 1B (retina) (GUCA1B), mRNA [NM_002098] |
| A_23_P146922 | 0,6807926 | down | -1,1255275 | -0,1706013 | GAS6 | Homo sapiens growth arrest-specific 6 (GAS6), mRNA [NM_000820] |
| A_23_P77103 | 0,7574293 | down | -1,1378144 | -0,1862653 | SORD | Homo sapiens sorbitol dehydrogenase (SORD), transcript variant 1, mRNA [NM_003104] |
| A_23_P49759 | 0,83808655 | down | -1,1170428 | -0,1596844 | CCL1 | Homo sapiens chemokine (C-C motif) ligand 1 (CCL1), mRNA [NM_002981] |
| A_23_P154986 | 0,10760969 | down | -1,7029948 | -0,768074 | GGT1 | Homo sapiens gamma-glutamyltransferase 1 (GGT1), transcript variant 6, mRNA [NM_001288833] |
| A_23_P167168 | 0,21242873 | down | -2,0500662 | -1,0356705 | IGJ | Homo sapiens immunoglobulin J polypeptide, linker protein for immunoglobulin alpha and mu polypeptides (IGJ), mRNA [NM_144646] |
| A_33_P3318796 | 0,6682512 | up | 1,0839313 | 0,11627336 | FSTL3 | Homo sapiens follistatin-like 3 (secreted glycoprotein) (FSTL3), mRNA [NM_005860] |
| A_23_P88522 | 0,03004017 | up | 1,4612674 | 0,5472202 | NMB | Homo sapiens neuromedin B (NMB), transcript variant 1, mRNA [NM_021077] |
| A_24_P277367 | 0,10117883 | down | -1,4558461 | -0,5418578 | CXCL5 | Homo sapiens chemokine (C-X-C motif) ligand 5 (CXCL5), mRNA [NM_002994] |
| A_23_P39955 | 0,12021077 | down | -1,5656127 | -0,6467273 | ACTG2 | Homo sapiens actin, gamma 2, smooth muscle, enteric (ACTG2), transcript variant 1, mRNA [NM_001615] |
| A_33_P3215953 | 0,37154123 | up | 1,1244578 | 0,16922957 | MPZL1 | Homo sapiens myelin protein zero-like 1 (MPZL1), transcript variant 2, mRNA [NM_024569] |
| A_23_P1552 | 0,16138077 | up | 1,2163559 | 0,28256544 | CTSC | Homo sapiens cathepsin C (CTSC), transcript variant 1, mRNA [NM_001814] |
| A_23_P49060 | 0,10289617 | up | 1,5221753 | 0,60613453 | SPINT1 | Homo sapiens serine peptidase inhibitor, Kunitz type 1 (SPINT1), transcript variant 1, mRNA [NM_181642] |
| A_23_P70355 | 0,59331805 | down | -1,1144797 | -0,1563703 | SERPINB6 | Homo sapiens serpin peptidase inhibitor, clade B (ovalbumin), member 6 (SERPINB6), transcript variant 1, mRNA [NM_004568] |
| A_23_P218237 | 0,15554999 | down | -1,4648284 | -0,5507317 | LCAT | Homo sapiens lecithin-cholesterol acyltransferase (LCAT), mRNA [NM_000229] |
| A_24_P763243 | 0,77131945 | up | 1,048133 | 0,06782182 | EEF1A1 | Homo sapiens eukaryotic translation elongation factor 1 alpha 1 (EEF1A1), mRNA [NM_001402] |
| A_23_P206396 | 0,32583874 | down | -1,1864451 | -0,2466454 | CKLF | Homo sapiens chemokine-like factor (CKLF), transcript variant 5, mRNA [NM_001040138] |
| A_24_P79755 | 0,32760873 | down | -1,188512 | -0,2491564 | AKR1A1 | Homo sapiens aldo-keto reductase family 1, member A1 (aldehyde reductase) (AKR1A1), transcript variant 1, mRNA [NM_006066] |
| A_33_P3397525 | 0,547459 | up | 1,202754 | 0,26634163 | WNT4 | Homo sapiens wingless-type MMTV integration site family, member 4 (WNT4), mRNA [NM_030761] |
| A_23_P208208 | 0,26661438 | down | -1,2614634 | -0,3350984 | ZNF649 | Homo sapiens zinc finger protein 649 (ZNF649), mRNA [NM_023074] |

| | | | | | | |
|---|---|---|---|---|---|---|
| A_24_P753161 | 0,2945364 | up | 1,1606435 | 0,21492486 | BMPR2 | Homo sapiens bone morphogenetic protein receptor, type II (serine/threonine kinase) (BMPR2), mRNA [NM_001204] |
| A_32_P219279 | 0,6382193 | down | -1,1093186 | -0,1496738 | ELFN2 | Homo sapiens extracellular leucine-rich repeat and fibronectin type III domain containing 2 (ELFN2), transcript variant 1, mRNA [NM_052906] |
| A_33_P3400248 | 0,15910931 | down | -2,445487 | -1,2901218 | FGF20 | Homo sapiens fibroblast growth factor 20 (FGF20), mRNA [NM_019851] |
| A_33_P3287631 | 0,31214112 | up | 1,1921048 | 0,2535111 | CTSB | Homo sapiens cathepsin B (CTSB), transcript variant 2, mRNA [NM_147780] |
| A_23_P88331 | 0,5044598 | down | -1,4588065 | -0,5447885 | DLGAP5 | Homo sapiens discs, large (Drosophila) homolog-associated protein 5 (DLGAP5), transcript variant 1, mRNA [NM_014750] |
| A_23_P5211 | 0,8683344 | up | 1,078711 | 0,10930844 | MUC16 | Homo sapiens mucin 16, cell surface associated (MUC16), mRNA [NM_024690] |
| A_33_P3292478 | 0,817022 | up | 1,0344034 | 0,04879898 | CCL16 | chemokine (C-C motif) ligand 16 [Source:HGNC Symbol;Acc:HGNC:10614] [ENST00000621559] |
| A_23_P169494 | 0,50546277 | up | 1,1228143 | 0,16711934 | ORM1 | Homo sapiens orosomucoid 1 (ORM1), mRNA [NM_000607] |
| A_23_P107465 | 0,18175405 | up | 2,0248537 | 1,0178176 | KRT31 | Homo sapiens keratin 31, type I (KRT31), mRNA [NM_002277] |
| A_32_P44316 | 0,70455444 | down | -1,0715923 | -0,0997562 | EEF1A1 | Homo sapiens eukaryotic translation elongation factor 1 alpha 1 (EEF1A1), mRNA [NM_001402] |
| A_33_P3392250 | 0,42889082 | up | 1,1532393 | 0,20569184 | FAM178A | Homo sapiens family with sequence similarity 178, member A (FAM178A), transcript variant 3, mRNA [NM_001243770] |
| A_33_P3337771 | 0,34030384 | down | -1,145798 | -0,1963527 | TGS1 | Homo sapiens trimethylguanosine synthase 1 (TGS1), mRNA [NM_024831] |
| A_24_P945283 | 0,83303934 | down | -1,0343063 | -0,0486635 | DLG3 | Homo sapiens discs, large homolog 3 (Drosophila) (DLG3), transcript variant 1, mRNA [NM_021120] |
| A_23_P146512 | 0,3159486 | down | -1,278172 | -0,354082 | GOLM1 | Homo sapiens golgi membrane protein 1 (GOLM1), transcript variant 1, mRNA [NM_016548] |
| A_33_P3327277 | 0,33816817 | down | -1,2510933 | -0,3231894 | VWA2 | Homo sapiens von Willebrand factor A domain containing 2 (VWA2), mRNA [NM_001272046] |
| A_33_P3219651 | 0,7743134 | up | 1,1185147 | 0,16158415 | BMPER | Homo sapiens BMP binding endothelial regulator (BMPER), mRNA [NM_133468] |
| A_33_P3243449 | 0,31686118 | down | -1,2199029 | -0,2867663 | CD70 | Homo sapiens CD70 molecule (CD70), mRNA [NM_001252] |
| A_23_P34744 | 0,7772622 | up | 1,0621364 | 0,08696906 | CTSK | Homo sapiens cathepsin K (CTSK), mRNA [NM_000396] |
| A_24_P158903 | 0,482201 | up | 1,1254878 | 0,17055042 | IRAK4 | Homo sapiens interleukin-1 receptor-associated kinase 4 (IRAK4), transcript variant 2, mRNA [NM_016123] |
| A_23_P37441 | 0,514683 | up | 1,1447972 | 0,19509205 | B2M | Homo sapiens beta-2-microglobulin (B2M), mRNA [NM_004048] |
| A_33_P3333282 | 0,12141252 | up | 1,4981747 | 0,5832058 | FGF11 | fibroblast growth factor 11 [Source:HGNC Symbol;Acc:HGNC:3667] [ENST00000293829] |
| A_33_P3376828 | 0,04572126 | down | -1,4671744 | -0,5530404 | CMTM7 | Homo sapiens CKLF-like MARVEL transmembrane domain containing 7 (CMTM7), transcript variant 1, mRNA [NM_138410] |
| A_23_P211631 | 0,8354889 | down | -1,0621327 | -0,086964 | FBLN1 | Homo sapiens fibulin 1 (FBLN1), transcript variant D, mRNA [NM_006486] |
| A_23_P87049 | 0,21226119 | down | -1,5901394 | -0,6691532 | SORL1 | Homo sapiens sortilin-related receptor, L(DLR class) A repeats containing (SORL1), mRNA [NM_003105] |
| A_33_P3245278 | 0,0058253 | down | -1,9188375 | -0,9402326 | PTPRG | Homo sapiens protein tyrosine phosphatase, receptor type, G (PTPRG), mRNA [NM_002841] |
| A_23_P91390 | 0,6414647 | down | -1,3556178 | -0,4389505 | THBD | Homo sapiens thrombomodulin (THBD), mRNA [NM_000361] |
| A_23_P312150 | 0,12912509 | up | 4,6441274 | 2,2154076 | EDN2 | Homo sapiens endothelin 2 (EDN2), transcript variant 1, mRNA [NM_001956] |
| A_23_P252236 | 0,5751759 | down | -1,111766 | -0,1528532 | KLKB1 | Homo sapiens kallikrein B, plasma (Fletcher factor) 1 (KLKB1), mRNA [NM_000892] |
| A_23_P145238 | 0,83970267 | up | 1,0739201 | 0,1028867 | HIST1H2BK | Homo sapiens histone cluster 1, H2bk (HIST1H2BK), mRNA [NM_080593] |
| A_24_P149036 | 0,87256956 | up | 1,122878 | 0,16720113 | DPYSL3 | Homo sapiens dihydropyrimidinase-like 3 (DPYSL3), transcript variant 2, mRNA [NM_001387] |
| A_33_P3409854 | 0,59413385 | up | 1,1135741 | 0,15519762 | LHX1 | Homo sapiens LIM homeobox 1 (LHX1), mRNA [NM_005568] |
| A_33_P3229122 | 0,80134875 | up | 1,1113844 | 0,15235788 | HIST1H2BF | Homo sapiens histone cluster 1, H2bf (HIST1H2BF), mRNA [NM_003522] |
| A_33_P3388501 | 0,35513058 | up | 1,1442572 | 0,19441135 | CHIT1 | Homo sapiens chitinase 1 (chitotriosidase) (CHIT1), transcript variant 1, mRNA [NM_003465] |
| A_32_P331052 | 0,01611313 | up | 1,6718173 | 0,74141717 | RBBP8NL | Homo sapiens RBBP8 N-terminal like (RBBP8NL), mRNA [NM_080833] |
| A_23_P206733 | 0,14200693 | down | -1,80426 | -0,8514072 | CES1 | Homo sapiens carboxylesterase 1 (CES1), transcript variant 3, mRNA [NM_001266] |
| A_23_P160226 | 0,41863644 | up | 1,2233518 | 0,29083937 | MROH7 | Homo sapiens maestro heat-like repeat family member 7 (MROH7), transcript variant 1, mRNA [NM_001039464] |
| A_32_P208823 | 0,075186 | up | 1,7810206 | 0,83270425 | PLXDC1 | Homo sapiens plexin domain containing 1 (PLXDC1), mRNA [NM_020405] |
| A_33_P3354464 | 0,01629794 | down | -1,9229503 | -0,9433215 | LOXL1 | Homo sapiens lysyl oxidase-like 1 (LOXL1), mRNA [NM_005576] |
| A_23_P150693 | 0,04646156 | down | -2,274337 | -1,185446 | FJX1 | Homo sapiens four jointed box 1 (Drosophila) (FJX1), mRNA [NM_014344] |
| A_33_P3303136 | 0,40984744 | down | -1,1871963 | -0,2475584 | SERPINB6 | Homo sapiens serpin peptidase inhibitor, clade B (ovalbumin), member 6 (SERPINB6), transcript variant 2, mRNA [NM_001195291] |
| A_33_P3358731 | 0,7676131 | up | 1,0766618 | 0,10656518 | PCSK5 | Homo sapiens proprotein convertase subtilisin/kexin type 5 (PCSK5), transcript variant 2, mRNA [NM_006200] |
| A_33_P3508822 | 0,1647166 | down | -1,2735553 | -0,3488616 | APP | Homo sapiens amyloid beta (A4) precursor protein (APP), transcript variant 1, mRNA [NM_000484] |
| A_33_P3354955 | 0,84631157 | up | 1,0352143 | 0,04992946 | ESF1 | Homo sapiens ESF1, nucleolar pre-rRNA processing protein, homolog (S. cerevisiae) (ESF1), transcript variant 1, mRNA [NM_016649] |
| A_24_P18270 | 0,5407107 | up | 1,1572067 | 0,21064653 | UCMA | Homo sapiens upper zone of growth plate and cartilage matrix associated (UCMA), transcript variant 1, mRNA [NM_145314] |
| A_23_P207213 | 0,7141255 | down | -1,1535953 | -0,2061372 | ALDH3A1 | Homo sapiens aldehyde dehydrogenase 3 family, member A1 (ALDH3A1), transcript variant 2, mRNA [NM_000691] |
| A_23_P322 | 0,8859272 | up | 1,0212367 | 0,03031723 | EFNA4 | Homo sapiens ephrin-A4 (EFNA4), transcript variant 3, mRNA [NM_182690] |
| A_33_P3276693 | 0,16173162 | up | 2,2456715 | 1,1671469 | PGF | Homo sapiens placental growth factor (PGF), transcript variant 1, mRNA [NM_002632] |
| A_23_P211212 | 0,294792 | up | 1,6099277 | 0,68699586 | COL18A1 | Homo sapiens collagen, type XVIII, alpha 1 (COL18A1), transcript variant 1, mRNA [NM_030582] |
| A_33_P3320943 | 0,54102075 | down | -1,1662813 | -0,2219159 | | Q864S5_BOVIN (Q864S5) Peptidylprolyl isomerase A, partial (97%) [THC2525667] |
| A_23_P314760 | 0,68341684 | down | -1,1136727 | -0,1553253 | PRKAG2 | Homo sapiens protein kinase, AMP-activated, gamma 2 non-catalytic subunit (PRKAG2), transcript variant a, mRNA [NM_016203] |
| A_24_P68783 | 0,02513377 | up | 5,3425293 | 2,417523 | IL36RN | Homo sapiens interleukin 36 receptor antagonist (IL36RN), transcript variant 1, mRNA [NM_012275] |
| A_33_P3297907 | 0,6403286 | down | -1,0957144 | -0,1318719 | FGF22 | Homo sapiens fibroblast growth factor 22 (FGF22), transcript variant 1, mRNA [NM_020637] |
| A_33_P3264163 | 0,36760563 | up | 1,1609969 | 0,21536413 | DDR1 | Homo sapiens discoidin domain receptor tyrosine kinase 1 (DDR1), transcript variant 5, mRNA [NM_001202522] |
| A_33_P3248265 | 0,41943517 | up | 1,8373283 | 0,87760943 | LTB | Homo sapiens lymphotoxin beta (TNF superfamily, member 3) (LTB), transcript variant 1, mRNA [NM_002341] |
| A_33_P3282489 | 0,20226996 | down | -1,2657729 | -0,3400186 | GCNT1 | Homo sapiens glucosaminyl (N-acetyl) transferase 1, core 2 (GCNT1), transcript variant 1, mRNA [NM_001097634] |

| | | | | | | |
|---|---|---|---|---|---|---|
| A_33_P3339231 | 0,4936046 | down | -1,1175994 | -0,1604031 | UBN2 | Homo sapiens ubinuclein 2 (UBN2), mRNA [NM_173569] |
| A_23_P98282 | 0,1904768 | up | 1,4292135 | 0,5152215 | SPTBN2 | Homo sapiens spectrin, beta, non-erythrocytic 2 (SPTBN2), mRNA [NM_006946] |
| A_23_P145289 | 0,67675734 | down | -1,0470519 | -0,066333 | GNL1 | Homo sapiens guanine nucleotide binding protein-like 1 (GNL1), mRNA [NM_005275] |
| A_24_P237486 | 0,8846706 | up | 1,0250686 | 0,03572052 | MECP2 | Homo sapiens methyl CpG binding protein 2 (MECP2), transcript variant 1, mRNA [NM_004992] |
| A_23_P399078 | 0,5040812 | up | 1,3113496 | 0,3910524 | TIMP3 | Homo sapiens TIMP metallopeptidase inhibitor 3 (TIMP3), mRNA [NM_000362] |
| A_23_P23048 | 0,17997593 | up | 3,0334513 | 1,6009601 | S100A9 | Homo sapiens S100 calcium binding protein A9 (S100A9), mRNA [NM_002965] |
| A_23_P139123 | 0,39480665 | up | 1,391765 | 0,47691563 | SERPING1 | Homo sapiens serpin peptidase inhibitor, clade G (C1 inhibitor), member 1 (SERPING1), transcript variant 1, mRNA [NM_000062] |
| A_23_P125977 | 0,23800042 | up | 1,3745737 | 0,45898426 | C1QC | Homo sapiens complement component 1, q subcomponent, C chain (C1QC), transcript variant 2, mRNA [NM_172369] |
| A_23_P41025 | 0,30016905 | down | -1,234757 | -0,3042271 | GNL3 | Homo sapiens guanine nucleotide binding protein-like 3 (nucleolar) (GNL3), transcript variant 1, mRNA [NM_014366] |
| A_33_P3421867 | 0,16666868 | down | -1,3167393 | -0,3969698 | MDGA1 | MAM domain containing glycosylphosphatidylinositol anchor 1 [Source:HGNC Symbol;Acc:HGNC:19267] [ENST00000373401] |
| A_33_P3260733 | 0,71672213 | down | -1,1153253 | -0,1574646 | GHR | Homo sapiens growth hormone receptor (GHR), transcript variant 12, mRNA [NM_001242462] |
| A_24_P245379 | 0,41705942 | up | 1,4610598 | 0,54701525 | SERPINB2 | Homo sapiens serpin peptidase inhibitor, clade B (ovalbumin), member 2 (SERPINB2), transcript variant 2, mRNA [NM_002575] |
| A_23_P201628 | 0,06342369 | down | -1,3214793 | -0,4021539 | LAMC1 | Homo sapiens laminin, gamma 1 (formerly LAMB2) (LAMC1), mRNA [NM_002293] |
| A_33_P3288844 | 0,02729388 | down | -1,9047643 | -0,9296125 | IL6R | Homo sapiens interleukin 6 receptor (IL6R), transcript variant 1, mRNA [NM_000565] |
| A_33_P3226377 | 0,6712728 | up | 1,0866351 | 0,11986757 | PRH2 | Homo sapiens proline-rich protein HaeIII subfamily 2 (PRH2), transcript variant 1, mRNA [NM_005042] |
| A_23_P324327 | 0,6899352 | down | -1,0920358 | -0,1270201 | GPRC5B | Homo sapiens G protein-coupled receptor, class C, group 5, member B (GPRC5B), mRNA [NM_016235] |
| A_23_P103256 | 0,54359174 | up | 1,1467197 | 0,19751278 | CFHR3 | Homo sapiens complement factor H-related 3 (CFHR3), transcript variant 1, mRNA [NM_021023] |
| A_33_P3369336 | 0,02780586 | up | 1,4168198 | 0,5026563 | GCG | Homo sapiens glucagon (GCG), mRNA [NM_002054] |
| A_33_P3212615 | 0,53849864 | up | 1,2354282 | 0,30501118 | TFPI | Homo sapiens tissue factor pathway inhibitor (lipoprotein-associated coagulation inhibitor) (TFPI), transcript variant 1, mRNA [NM_006287] |
| A_33_P3370930 | 0,6187954 | down | -1,0701114 | -0,097761 | LAMB1 | laminin, beta 1 [Source:HGNC Symbol;Acc:HGNC:6486] [ENST00000393559] |
| A_33_P3220911 | 0,62416285 | up | 1,1993065 | 0,2622004 | BST2 | Homo sapiens bone marrow stromal cell antigen 2 (BST2), mRNA [NM_004335] |
| A_24_P307135 | 0,29662985 | down | -1,266321 | -0,3406431 | TNXB | Homo sapiens tenascin XB (TNXB), transcript variant XB, mRNA [NM_019105] |
| A_23_P315815 | 0,5725917 | down | -1,2741623 | -0,3495491 | NRG1 | Homo sapiens neuregulin 1 (NRG1), transcript variant HRG-gamma, mRNA [NM_004495] |
| A_23_P141894 | 0,71018136 | up | 1,069791 | 0,09732892 | PVR | Homo sapiens poliovirus receptor (PVR), transcript variant 1, mRNA [NM_006505] |
| A_23_P43164 | 0,32183 | down | -1,7749964 | -0,8278161 | SULF1 | Homo sapiens sulfatase 1 (SULF1), transcript variant 3, mRNA [NM_015170] |
| A_23_P212508 | 0,10315022 | up | 3,4237823 | 1,775591 | TF | Homo sapiens transferrin (TF), mRNA [NM_001063] |
| A_33_P3287338 | 0,48228914 | down | -1,1217041 | -0,1656922 | IL6ST | Homo sapiens interleukin 6 signal transducer (IL6ST), transcript variant 3, mRNA [NM_001190981] |
| A_19_P00318098 | 0,21640468 | up | 1,5337521 | 0,6170653 | DPYSL3 | dihydropyrimidinase-like 3 [Source:HGNC Symbol;Acc:HGNC:3015] [ENST00000504965] |
| A_23_P4714 | 0,55254036 | up | 1,4125425 | 0,49829423 | MIA | Homo sapiens melanoma inhibitory activity (MIA), transcript variant 1, mRNA [NM_006533] |
| A_33_P3315268 | 0,00610401 | up | 7,7955422 | 2,9626493 | KRT78 | Homo sapiens keratin 78, type II (KRT78), transcript variant 1, mRNA [NM_173352] |
| A_32_P313405 | 0,3667223 | down | -1,5844754 | -0,6640053 | LAMA1 | Homo sapiens laminin, alpha 1 (LAMA1), mRNA [NM_005559] |
| A_23_P77529 | 0,8515763 | down | -1,0993047 | -0,1365913 | MSLN | Homo sapiens mesothelin (MSLN), transcript variant 1, mRNA [NM_005823] |
| A_23_P96827 | 0,28293958 | up | 1,1961287 | 0,25837266 | APCS | Homo sapiens amyloid P component, serum (APCS), mRNA [NM_001639] |
| A_23_P204079 | 0,26090446 | up | 1,2993455 | 0,3777851 | NPFF | Homo sapiens neuropeptide FF-amide peptide precursor (NPFF), mRNA [NM_003717] |
| A_23_P412389 | 0,5977709 | up | 1,1249129 | 0,16981325 | FGF18 | Homo sapiens fibroblast growth factor 18 (FGF18), mRNA [NM_003862] |
| A_33_P3363355 | 0,5517719 | down | -1,1841797 | -0,243888 | ICAM4 | Homo sapiens intercellular adhesion molecule 4 (Landsteiner-Wiener blood group) (ICAM4), transcript variant 2, mRNA [NM_022377] |
| A_23_P27367 | 0,6336555 | down | -1,0713936 | -0,0994886 | KDSR | Homo sapiens 3-ketodihydrosphingosine reductase (KDSR), mRNA [NM_002035] |
| A_23_P53588 | 0,0899206 | up | 1,765761 | 0,8202901 | WNT5B | Homo sapiens wingless-type MMTV integration site family, member 5B (WNT5B), transcript variant 2, mRNA [NM_030775] |
| A_33_P3238433 | 0,77933306 | down | -1,1058673 | -0,1451782 | ALDH3A1 | Homo sapiens aldehyde dehydrogenase 3 family, member A1 (ALDH3A1), transcript variant 1, mRNA [NM_001135168] |
| A_33_P3317589 | 0,728849 | up | 1,0990748 | 0,13628963 | GFRA4 | Homo sapiens GDNF family receptor alpha 4 (GFRA4), transcript variant 2, mRNA [NM_145762] |
| A_23_P86653 | 0,89961046 | up | 1,061644 | 0,08630001 | SRGN | Homo sapiens serglycin (SRGN), transcript variant 1, mRNA [NM_002727] |
| A_24_P140608 | 0,7094733 | up | 1,3242737 | 0,40520132 | HBEGF | Homo sapiens heparin-binding EGF-like growth factor (HBEGF), mRNA [NM_001945] |
| A_23_P139722 | 0,17274459 | up | 1,3230855 | 0,40390635 | TNFRSF1A | Homo sapiens tumor necrosis factor receptor superfamily, member 1A (TNFRSF1A), mRNA [NM_001065] |
| A_33_P3267640 | 0,7926175 | up | 1,0491235 | 0,06918455 | HGFAC | Homo sapiens HGF activator (HGFAC), transcript variant 1, mRNA [NM_001297439] |
| A_23_P166408 | 0,32254505 | down | -1,9829154 | -0,9876231 | OSM | Homo sapiens oncostatin M (OSM), mRNA [NM_020530] |
| A_23_P3911 | 0,15998138 | up | 1,9135087 | 0,9362204 | PLXDC1 | Homo sapiens plexin domain containing 1 (PLXDC1), mRNA [NM_020405] |
| A_23_P93389 | 0,3873352 | up | 1,1398274 | 0,18881534 | NUDT3 | Homo sapiens nudix (nucleoside diphosphate linked moiety X)-type motif 3 (NUDT3), mRNA [NM_006703] |
| A_33_P3250680 | 0,25074315 | down | -1,5987526 | -0,6769467 | CD40LG | Homo sapiens CD40 ligand (CD40LG), mRNA [NM_000074] |
| A_23_P24129 | 0,8464072 | down | -1,1106774 | -0,1514398 | DKK1 | Homo sapiens dickkopf WNT signaling pathway inhibitor 1 (DKK1), mRNA [NM_012242] |
| A_33_P3422330 | 0,23318166 | up | 1,4236554 | 0,5096 | KLHL17 | Homo sapiens kelch-like family member 17 (KLHL17), mRNA [NM_198317] |
| A_23_P101407 | 0,7148887 | down | -1,2037601 | -0,267548 | C3 | Homo sapiens complement component 3 (C3), mRNA [NM_000064] |
| A_33_P3235701 | 0,42559478 | up | 1,1847706 | 0,24460772 | ZCCHC11 | zinc finger, CCHC domain containing 11 [Source:HGNC Symbol;Acc:HGNC:28981] [ENST00000371541] |
| A_23_P13364 | 0,4619889 | down | -1,2383877 | -0,308463 | NUCB2 | Homo sapiens nucleobindin 2 (NUCB2), mRNA [NM_005013] |
| A_23_P78944 | 0,40498602 | up | 1,2561333 | 0,3289896 | AMH | Homo sapiens anti-Mullerian hormone (AMH), mRNA [NM_000479] |
| A_23_P425681 | 0,6281339 | up | 1,2367579 | 0,30656308 | CCK | Homo sapiens cholecystokinin (CCK), transcript variant 1, mRNA [NM_000729] |

| | | | | | | |
|---|---|---|---|---|---|---|
| A_33_P3226810 | 0,11551698 | up | 1,3773315 | 0,46187583 | TNFSF10 | Homo sapiens tumor necrosis factor (ligand) superfamily, member 10 (TNFSF10), transcript variant 1, mRNA [NM_003810] |
| A_23_P81058 | 0,01968712 | up | 2,0667417 | 1,047358 | BMP3 | Homo sapiens bone morphogenetic protein 3 (BMP3), mRNA [NM_001201] |
| A_23_P156327 | 0,3658389 | up | 1,2536155 | 0,32609493 | TGFBI | Homo sapiens transforming growth factor, beta-induced, 68kDa (TGFBI), mRNA [NM_000358] |
| A_32_P198923 | 0,42816192 | up | 1,1855522 | 0,24555923 | YWHAZ | Homo sapiens tyrosine 3-monooxygenase/tryptophan 5-monooxygenase activation protein, zeta (YWHAZ), transcript variant 2, mRNA [NM_145690] |
| A_23_P156880 | 0,30850908 | up | 1,2799122 | 0,3560449 | ENPP1 | Homo sapiens ectonucleotide pyrophosphatase/phosphodiesterase 1 (ENPP1), mRNA [NM_006208] |
| A_23_P14174 | 0,29760534 | up | 1,3391111 | 0,42127565 | TNFSF13B | Homo sapiens tumor necrosis factor (ligand) superfamily, member 13b (TNFSF13B), transcript variant 1, mRNA [NM_006573] |
| A_33_P3210492 | 0,79684067 | up | 1,0683253 | 0,09535098 | C1QTNF4 | Homo sapiens C1q and tumor necrosis factor related protein 4 (C1QTNF4), mRNA [NM_031909] |
| A_23_P258340 | 0,70052075 | down | -1,0911179 | -0,1258069 | PPIA | Homo sapiens peptidylprolyl isomerase A (cyclophilin A) (PPIA), transcript variant 1, mRNA [NM_021130] |
| A_23_P202334 | 0,59628254 | down | -1,1409492 | -0,1902346 | FGFR2 | Homo sapiens fibroblast growth factor receptor 2 (FGFR2), transcript variant 2, mRNA [NM_022970] |
| A_19_P00319513 | 0,19830039 | up | 2,1317146 | 1,0920143 | DPYSL3 | dihydropyrimidinase-like 3 [Source:HGNC Symbol;Acc:HGNC:3015] [ENST00000504965] |
| A_33_P3234043 | 0,6229614 | up | 1,1261799 | 0,17143735 | BMP1 | bone morphogenetic protein 1 [Source:HGNC Symbol;Acc:HGNC:1067] [ENST00000471755] |
| A_23_P63026 | 0,48780465 | down | -1,1079332 | -0,1478709 | LGALS8 | Homo sapiens lectin, galactoside-binding, soluble, 8 (LGALS8), transcript variant 1, mRNA [NM_006499] |
| A_23_P353035 | 0,04054946 | down | -3,4431965 | -1,7837485 | IGFBP7 | Homo sapiens insulin-like growth factor binding protein 7 (IGFBP7), transcript variant 1, mRNA [NM_001553] |
| A_23_P500501 | 0,18789974 | up | 1,4053963 | 0,49097705 | FGFR3 | Homo sapiens fibroblast growth factor receptor 3 (FGFR3), transcript variant 1, mRNA [NM_000142] |
| A_33_P3273552 | 0,3216428 | up | 1,3602248 | 0,44384515 | KRT83 | Homo sapiens keratin 83, type II (KRT83), mRNA [NM_002282] |
| A_23_P332908 | 0,25670207 | up | 1,1746821 | 0,23227042 | ZFC3H1 | Homo sapiens zinc finger, C3H1-type containing (ZFC3H1), mRNA [NM_144982] |
| A_23_P137856 | 0,5007751 | up | 1,1757376 | 0,23356614 | MUC1 | Homo sapiens mucin 1, cell surface associated (MUC1), transcript variant 1, mRNA [NM_002456] |
| A_24_P339944 | 0,7148521 | up | 1,0949467 | 0,1308607 | PDGFB | Homo sapiens platelet-derived growth factor beta polypeptide (PDGFB), transcript variant 1, mRNA [NM_002608] |
| A_23_P254888 | 0,4620465 | up | 1,2417725 | 0,3124009 | ZYX | Homo sapiens zyxin (ZYX), transcript variant 1, mRNA [NM_003461] |
| A_23_P167096 | 0,67867744 | down | -1,1418498 | -0,1913728 | VEGFC | Homo sapiens vascular endothelial growth factor C (VEGFC), mRNA [NM_005429] |
| A_33_P3392784 | 0,06916725 | down | -4,098004 | -2,0349214 | CFAP58 | cilia and flagella associated protein 58 [Source:HGNC Symbol;Acc:HGNC:26676] [ENST00000369703] |
| A_23_P310956 | 0,63448954 | up | 1,1571814 | 0,21061501 | COL6A2 | Homo sapiens collagen, type VI, alpha 2 (COL6A2), transcript variant 2C2a', mRNA [NM_058175] |
| A_33_P3344574 | 0,71975523 | up | 1,0628513 | 0,08793978 | SFTPA2 | Homo sapiens surfactant protein A2 (SFTPA2), mRNA [NM_001098668] |
| A_33_P3361457 | 0,53797287 | down | -1,1191777 | -0,1624391 | IFNAR2 | Homo sapiens interferon (alpha, beta and omega) receptor 2 (IFNAR2), transcript variant 2, mRNA [NM_000874] |
| A_23_P12680 | 0,8358009 | down | -1,0347131 | -0,0492309 | PSAP | Homo sapiens prosaposin (PSAP), transcript variant 2, mRNA [NM_001042465] |
| A_23_P150064 | 0,42311317 | down | -1,2982322 | -0,3765484 | MMRN2 | Homo sapiens multimerin 2 (MMRN2), mRNA [NM_024756] |
| A_33_P3364864 | 0,08727159 | up | 1,3592782 | 0,44284075 | NAMPT | Homo sapiens nicotinamide phosphoribosyltransferase (NAMPT), mRNA [NM_005746] |
| A_23_P252471 | 0,4986255 | up | 1,2662011 | 0,34050658 | PECAM1 | Homo sapiens platelet/endothelial cell adhesion molecule 1 (PECAM1), mRNA [NM_000442] |
| A_24_P65616 | 0,67105025 | down | -1,1403714 | -0,1895038 | PVR | Homo sapiens poliovirus receptor (PVR), transcript variant 1, mRNA [NM_006505] |
| A_23_P166269 | 0,17025042 | up | 1,5054346 | 0,59018004 | FAM3B | Homo sapiens family with sequence similarity 3, member B (FAM3B), transcript variant 1, mRNA [NM_058186] |
| A_23_P76291 | 0,23909448 | down | -3,5364652 | -1,8223081 | PRR4 | Homo sapiens proline rich 4 (lacrimal) (PRR4), transcript variant 2, mRNA [NM_007244] |
| A_24_P169148 | 0,42629266 | down | -1,1645213 | -0,2197371 | HMGB1 | Homo sapiens high mobility group box 1 (HMGB1), mRNA [NM_002128] |
| A_23_P200780 | 0,21727552 | down | -1,4836625 | -0,5691629 | TGFBR3 | Homo sapiens transforming growth factor, beta receptor III (TGFBR3), transcript variant 1, mRNA [NM_003243] |
| A_23_P15357 | 0,8907732 | up | 1,0179379 | 0,02564955 | LGALS3BP | Homo sapiens lectin, galactoside-binding, soluble, 3 binding protein (LGALS3BP), mRNA [NM_005567] |
| A_33_P3279019 | 0,38518643 | up | 1,5907942 | 0,6697472 | SERPINB13 | Homo sapiens serpin peptidase inhibitor, clade B (ovalbumin), member 13 (SERPINB13), mRNA [NM_012397] |
| A_24_P928052 | 0,17805254 | down | -1,4466212 | -0,5326872 | NRP1 | Homo sapiens neuropilin 1 (NRP1), transcript variant 1, mRNA [NM_003873] |
| A_24_P45476 | 0,22676674 | up | 1,9635175 | 0,97344047 | XCL1 | Homo sapiens chemokine (C motif) ligand 1 (XCL1), mRNA [NM_002995] |
| A_23_P157580 | 0,58523315 | up | 1,0961167 | 0,13240135 | SDCBP | Homo sapiens syndecan binding protein (syntenin) (SDCBP), transcript variant 1, mRNA [NM_005625] |
| A_23_P166459 | 0,09492316 | up | 1,4689919 | 0,55482644 | LGALS1 | Homo sapiens lectin, galactoside-binding, soluble, 1 (LGALS1), mRNA [NM_002305] |
| A_23_P88404 | 0,1788001 | down | -1,396456 | -0,4817701 | TGFB3 | Homo sapiens transforming growth factor, beta 3 (TGFB3), mRNA [NM_003239] |
| A_24_P406754 | 0,51376194 | up | 1,2734305 | 0,3487202 | LOXL4 | Homo sapiens lysyl oxidase-like 4 (LOXL4), mRNA [NM_032211] |
| A_23_P64825 | 0,4172175 | up | 1,2832594 | 0,35981283 | LACRT | Homo sapiens lacritin (LACRT), mRNA [NM_033277] |
| A_33_P3278407 | 0,01649434 | up | 1,5588454 | 0,64047784 | MLLT4 | Homo sapiens myeloid/lymphoid or mixed-lineage leukemia (trithorax homolog, Drosophila); translocated to, 4 (MLLT4), transcript variant 1, mRNA [NM_001207008] |
| A_24_P264943 | 0,88884264 | down | -1,0778449 | -0,1081495 | COMP | Homo sapiens cartilage oligomeric matrix protein (COMP), mRNA [NM_000095] |
| A_33_P3209885 | 0,14428182 | up | 1,550408 | 0,63264793 | PLXDC1 | Homo sapiens plexin domain containing 1 (PLXDC1), mRNA [NM_020405] |
| A_23_P40453 | 0,19224717 | up | 1,3783336 | 0,46292508 | CBR3 | Homo sapiens carbonyl reductase 3 (CBR3), mRNA [NM_001236] |
| A_33_P3355732 | 0,5126353 | up | 1,2740101 | 0,34937668 | UMODL1 | uromodulin-like 1 [Source:HGNC Symbol;Acc:HGNC:12560] [ENST00000491559] |
| A_23_P154894 | 0,30575055 | up | 1,4495335 | 0,5355886 | CSTB | Homo sapiens cystatin B (stefin B) (CSTB), mRNA [NM_000100] |
| A_24_P131589 | 0,78418607 | up | 1,1624992 | 0,21722971 | CD86 | Homo sapiens CD86 molecule (CD86), transcript variant 2, mRNA [NM_006889] |
| A_23_P82868 | 0,46300036 | up | 1,7205366 | 0,78285855 | PLAT | Homo sapiens plasminogen activator, tissue (PLAT), transcript variant 1, mRNA [NM_000930] |
| A_33_P3417281 | 0,04972186 | up | 1,826427 | 0,86902404 | MUC4 | Homo sapiens mucin 4, cell surface associated (MUC4), transcript variant 1, mRNA [NM_018406] |
| A_23_P135257 | 0,0812021 | up | 4,878136 | 2,28633 | PRSS3 | Homo sapiens protease, serine, 3 (PRSS3), transcript variant 2, mRNA [NM_002771] |
| A_23_P374844 | 0,5192312 | up | 1,4173948 | 0,5032416 | GAL | Homo sapiens galanin/GMAP prepropeptide (GAL), mRNA [NM_015973] |
| A_23_P100660 | 0,06556968 | down | -1,4921538 | -0,5773962 | SERPINF1 | Homo sapiens serpin peptidase inhibitor, clade F (alpha-2 antiplasmin, pigment epithelium derived factor), member 1 (SERPINF1), mRNA [NM_002615] |
| A_32_P148345 | 0,09021471 | down | -1,4834237 | -0,5689308 | ANXA2 | Homo sapiens annexin A2 (ANXA2), transcript variant 2, mRNA [NM_001002857] |

| | | | | | | |
|---|---|---|---|---|---|---|
| A_33_P3251163 | 0,38399193 | up | 1,2804784 | 0,35668287 | RNASET2 | ribonuclease T2 [Source:HGNC Symbol;Acc:HGNC:21686] [ENST00000510083] |
| A_33_P3415820 | 0,7729727 | down | -1,046976 | -0,0662283 | THBS1 | Homo sapiens thrombospondin 1 (THBS1), mRNA [NM_003246] |
| A_33_P3279470 | 0,02771314 | up | 1,5834537 | 0,6630746 | AGRP | Homo sapiens agouti related neuropeptide (AGRP), mRNA [NM_001138] |
| A_23_P341275 | 0,7813802 | down | -1,06148 | -0,0860773 | POP1 | Homo sapiens processing of precursor 1, ribonuclease P/MRP subunit (S. cerevisiae) (POP1), transcript variant 3, mRNA [NM_015029] |
| A_33_P3404601 | 0,67609966 | up | 1,1612985 | 0,21573886 | C2 | Homo sapiens complement component 2 (C2), transcript variant 5, mRNA [NM_001282458] |
| A_23_P130411 | 0,5747141 | up | 1,2717346 | 0,34679762 | SERPINB11 | Homo sapiens serpin peptidase inhibitor, clade B (ovalbumin), member 11 (gene/pseudogene) (SERPINB11), transcript variant 1, mRNA [NM_080475] |
| A_23_P218442 | 0,07414685 | up | 1,4950187 | 0,58016354 | CEACAM6 | Homo sapiens carcinoembryonic antigen-related cell adhesion molecule 6 (non-specific cross reacting antigen) (CEACAM6), mRNA [NM_002483] |
| A_23_P5983 | 0,48061553 | down | -1,1980724 | -0,2607151 | PLTP | Homo sapiens phospholipid transfer protein (PLTP), transcript variant 1, mRNA [NM_006227] |
| A_32_P75792 | 0,211908 | up | 1,6145357 | 0,6911193 | FAM132A | Homo sapiens family with sequence similarity 132, member A (FAM132A), mRNA [NM_001014980] |
| A_23_P58396 | 0,2918754 | down | -1,4170042 | -0,502844 | PDGFC | Homo sapiens platelet derived growth factor C (PDGFC), transcript variant 1, mRNA [NM_016205] |
| A_24_P246943 | 0,8268626 | down | -1,047952 | -0,0675727 | PPIA | Homo sapiens peptidylprolyl isomerase A (cyclophilin A) (PPIA), transcript variant 1, mRNA [NM_021130] |
| A_24_P135322 | 0,8332009 | down | -1,0810416 | -0,112422 | NRP1 | Homo sapiens neuropilin 1 (NRP1), transcript variant 3, mRNA [NM_001024629] |
| A_33_P3245289 | 0,1772392 | down | -1,2919532 | -0,3695538 | PTPRG | Homo sapiens protein tyrosine phosphatase, receptor type, G (PTPRG), mRNA [NM_002841] |
| A_23_P166848 | 0,399466 | down | -2,8870006 | -1,5295714 | LTF | Homo sapiens lactotransferrin (LTF), transcript variant 1, mRNA [NM_002343] |
| A_23_P207336 | 0,08342782 | up | 1,3269726 | 0,40813857 | PPY | Homo sapiens pancreatic polypeptide (PPY), mRNA [NM_002722] |
| A_23_P97541 | 0,6846668 | up | 1,125375 | 0,17040586 | C4BPA | Homo sapiens complement component 4 binding protein, alpha (C4BPA), mRNA [NM_000715] |
| A_23_P152838 | 0,41842166 | up | 1,7097474 | 0,7737832 | CCL5 | Homo sapiens chemokine (C-C motif) ligand 5 (CCL5), transcript variant 1, mRNA [NM_002985] |
| A_33_P3232692 | 0,32710996 | down | -1,2292585 | -0,2977884 | IL24 | Homo sapiens interleukin 24 (IL24), transcript variant 3, mRNA [NM_001185156] |
| A_23_P208389 | 0,2528569 | up | 1,2489104 | 0,32067 | AXL | Homo sapiens AXL receptor tyrosine kinase (AXL), transcript variant 1, mRNA [NM_021913] |
| A_23_P254741 | 0,80590963 | down | -1,1286623 | -0,174614 | SOD3 | Homo sapiens superoxide dismutase 3, extracellular (SOD3), mRNA [NM_003102] |
| A_33_P3306146 | 0,782985 | up | 1,1264809 | 0,17182289 | PLAU | Homo sapiens plasminogen activator, urokinase (PLAU), transcript variant 2, mRNA [NM_001145031] |
| A_23_P150950 | 0,50691134 | down | -1,1297121 | -0,1759552 | ZFC3H1 | Homo sapiens zinc finger, C3H1-type containing (ZFC3H1), mRNA [NM_144982] |
| A_33_P3365878 | 0,83042586 | up | 1,0417548 | 0,0590158 | BMP8B | Homo sapiens bone morphogenetic protein 8b (BMP8B), mRNA [NM_001720] |
| A_23_P211080 | 0,35480762 | down | -1,1863668 | -0,2465501 | IFNAR2 | Homo sapiens interferon (alpha, beta and omega) receptor 2 (IFNAR2), transcript variant 1, mRNA [NM_207585] |
| A_23_P48886 | 0,82427925 | up | 1,0402596 | 0,0569436 | ADAM10 | Homo sapiens ADAM metallopeptidase domain 10 (ADAM10), mRNA [NM_001110] |
| A_23_P47168 | 0,3218121 | up | 1,2555804 | 0,32835445 | FLRT1 | Homo sapiens fibronectin leucine rich transmembrane protein 1 (FLRT1), mRNA [NM_013280] |
| A_33_P3257708 | 0,601292 | up | 1,062058 | 0,08686252 | APOA1BP | Homo sapiens apolipoprotein A-I binding protein (APOA1BP), mRNA [NM_144772] |
| A_23_P57268 | 0,37170762 | down | -1,130142 | -0,176504 | CXADR | Homo sapiens coxsackie virus and adenovirus receptor (CXADR), transcript variant 1, mRNA [NM_001338] |
| A_33_P3377199 | 0,38406757 | down | -1,1660303 | -0,2216053 | PRDX1 | Homo sapiens peroxiredoxin 1 (PRDX1), transcript variant 1, mRNA [NM_002574] |
| A_32_P84009 | 0,5457338 | down | -1,1413605 | -0,1907546 | CMTM4 | Homo sapiens CKLF-like MARVEL transmembrane domain containing 4 (CMTM4), transcript variant 2, mRNA [NM_181521] |
| A_23_P363769 | 0,21359558 | up | 1,300237 | 0,37877467 | KRT86 | Homo sapiens keratin 86, type II (KRT86), mRNA [NM_002284] |
| A_23_P149545 | 0,08260971 | down | -1,9892255 | -0,9922068 | HIST2H2BE | Homo sapiens histone cluster 2, H2be (HIST2H2BE), mRNA [NM_003528] |
| A_24_P289471 | 0,7866835 | up | 1,070498 | 0,09828208 | RNASET2 | Homo sapiens ribonuclease T2 (RNASET2), mRNA [NM_003730] |
| A_33_P3337540 | 0,78704554 | down | -1,0575761 | -0,0807614 | IFNAR2 | Homo sapiens interferon (alpha, beta and omega) receptor 2 (IFNAR2), transcript variant 2, mRNA [NM_000874] |
| A_33_P3370940 | 0,40496472 | down | -1,2892385 | -0,3665191 | LAMB1 | Homo sapiens laminin, beta 1, mRNA (cDNA clone IMAGE:4889995), containing frame-shift errors. [BC044633] |
| A_33_P3358898 | 0,35263056 | down | -1,4916198 | -0,5768799 | SSC5D | Homo sapiens scavenger receptor cysteine rich family, 5 domains (SSC5D), transcript variant 1, mRNA [NM_001144950] |
| A_23_P167920 | 0,04407441 | down | -1,7650563 | -0,8197141 | DLL1 | Homo sapiens delta-like 1 (Drosophila) (DLL1), mRNA [NM_005618] |
| A_24_P147461 | 0,4505406 | up | 1,229543 | 0,29812214 | SERPINB8 | Homo sapiens serpin peptidase inhibitor, clade B (ovalbumin), member 8 (SERPINB8), transcript variant 3, mRNA [NM_001031848] |
| A_24_P203953 | 0,40182003 | down | -1,1891787 | -0,2499655 | LOC439951 | PREDICTED: Homo sapiens uncharacterized LOC439951 (LOC439951), misc_RNA [XR_171055] |
| A_33_P3211569 | 0,45830736 | up | 1,25475 | 0,32739997 | ERBB3 | Homo sapiens v-erb-b2 avian erythroblastic leukemia viral oncogene homolog 3 (ERBB3), transcript variant s, mRNA [NM_001005915] |
| A_33_P3229288 | 0,560481 | up | 1,1168926 | 0,15949044 | ACE | Homo sapiens angiotensin I converting enzyme (ACE), transcript variant 1, mRNA [NM_000789] |
| A_23_P156683 | 0,5221715 | up | 1,0964736 | 0,13287105 | LTA | Homo sapiens lymphotoxin alpha (LTA), transcript variant 2, mRNA [NM_000595] |
| A_33_P3248903 | 0,8938944 | up | 1,0326307 | 0,04632437 | WNT7B | Homo sapiens wingless-type MMTV integration site family, member 7B (WNT7B), mRNA [NM_058238] |
| A_23_P12463 | 0,01256869 | up | 1,6320908 | 0,7067213 | QSOX1 | Homo sapiens quiescin Q6 sulfhydryl oxidase 1 (QSOX1), transcript variant 1, mRNA [NM_002826] |
| A_32_P195065 | 0,77226883 | up | 1,06297 | 0,08810094 | SEMA4F | Homo sapiens sema domain, immunoglobulin domain (Ig), transmembrane domain (TM) and short cytoplasmic domain, (semaphorin) 4F (SEMA4F), transcript variant 1, |
| A_23_P69329 | 0,09345559 | up | 1,5384706 | 0,6214969 | HYAL1 | Homo sapiens hyaluronoglucosaminidase 1 (HYAL1), transcript variant 8, mRNA [NM_153281] |
| A_33_P3271455 | 0,4886536 | up | 1,530412 | 0,61392003 | PXDN | Homo sapiens peroxidasin (PXDN), mRNA [NM_012293] |
| A_23_P46429 | 0,5342439 | up | 1,2263309 | 0,29434827 | CYR61 | Homo sapiens cysteine-rich, angiogenic inducer, 61 (CYR61), mRNA [NM_001554] |
| A_23_P250122 | 0,4385786 | up | 1,1916096 | 0,2529117 | FAM20C | Homo sapiens family with sequence similarity 20, member C (FAM20C), mRNA [NM_020223] |
| A_23_P2271 | 0,30061272 | up | 2,175163 | 1,1211236 | PTHLH | Homo sapiens parathyroid hormone-like hormone (PTHLH), transcript variant 1, mRNA [NM_198965] |
| A_23_P141035 | 0,3193095 | up | 1,3223356 | 0,40308836 | CHST4 | Homo sapiens carbohydrate (N-acetylglucosamine 6-O) sulfotransferase 4 (CHST4), transcript variant 1, mRNA [NM_005769] |
| A_24_P403459 | 0,3384892 | up | 1,350377 | 0,4333622 | IFNA4 | Homo sapiens interferon, alpha 4 (IFNA4), mRNA [NM_021068] |
| A_33_P3229725 | 0,70838434 | up | 1,0654877 | 0,09151399 | ARSA | Homo sapiens arylsulfatase A (ARSA), transcript variant 1, mRNA [NM_000487] |
| A_33_P3284933 | 0,03730639 | up | 1,5675653 | 0,64852554 | IL27 | Homo sapiens interleukin 27 (IL27), mRNA [NM_145659] |
| A_23_P6066 | 0,3056764 | down | -1,409414 | -0,4950955 | CPXM1 | Homo sapiens carboxypeptidase X (M14 family), member 1 (CPXM1), transcript variant 1, mRNA [NM_019609] |

| | | | | | | |
|---|---|---|---|---|---|---|
| A_23_P49975 | 0,16970846 | up | 1,8850211 | 0,91458064 | KRT10 | Homo sapiens keratin 10, type I (KRT10), mRNA [NM_000421] |
| A_23_P256375 | 0,2590375 | up | 1,1824397 | 0,24176659 | STX4 | Homo sapiens syntaxin 4 (STX4), transcript variant 3, mRNA [NM_004604] |
| A_24_P306726 | 0,5124587 | up | 1,0962893 | 0,13262853 | TPT1 | Homo sapiens tumor protein, translationally-controlled 1 (TPT1), transcript variant 2, mRNA [NM_003295] |
| A_23_P160968 | 0,03965978 | down | -1,5767262 | -0,6569322 | LAMC2 | Homo sapiens laminin, gamma 2 (LAMC2), transcript variant 2, mRNA [NM_018891] |
| A_33_P3246883 | 0,6641942 | down | -1,2283037 | -0,2966673 | DMKN | Homo sapiens dermokine (DMKN), transcript variant 2, mRNA [NM_033317] |
| A_33_P3349637 | 0,23895086 | up | 1,4078681 | 0,4935122 | PCDH1 | Homo sapiens protocadherin 1 (PCDH1), transcript variant 1, mRNA [NM_002587] |
| A_23_P363968 | 0,6078001 | up | 1,075781 | 0,1053844 | C1RL | Homo sapiens complement component 1, r subcomponent-like (C1RL), transcript variant 1, mRNA [NM_016546] |
| A_33_P3376007 | 0,6619983 | up | 1,0640554 | 0,08957332 | TXLNA | Homo sapiens taxilin alpha (TXLNA), mRNA [NM_175852] |
| A_33_P3229246 | 0,5100732 | down | -1,1988593 | -0,2616624 | HIST2H2BE | Homo sapiens histone cluster 2, H2be (HIST2H2BE), mRNA [NM_003528] |
| A_33_P3338698 | 0,01840872 | down | -2,785691 | -1,4780352 | IHH | Homo sapiens indian hedgehog (IHH), mRNA [NM_002181] |
| A_33_P3217559 | 0,38355747 | up | 1,1524204 | 0,2046671 | DLK1 | Homo sapiens delta-like 1 homolog (Drosophila) (DLK1), mRNA [NM_003836] |
| A_23_P41344 | 0,20410588 | up | 3,2787044 | 1,7131258 | EREG | Homo sapiens epiregulin (EREG), mRNA [NM_001432] |
| A_33_P3259878 | 0,21063653 | up | 1,2513833 | 0,32352376 | DDB1 | damage-specific DNA binding protein 1, 127kDa [Source:HGNC Symbol;Acc:HGNC:2717] [ENST00000545894] |
| A_23_P405873 | 0,35305727 | down | -1,3683999 | -0,4524899 | C9orf72 | Homo sapiens chromosome 9 open reading frame 72 (C9orf72), transcript variant 1, mRNA [NM_145005] |
| A_23_P21838 | 0,02033575 | up | 1,3721757 | 0,4564652 | CNP | Homo sapiens 2',3'-cyclic nucleotide 3' phosphodiesterase (CNP), mRNA [NM_033133] |
| A_23_P119916 | 0,632728 | down | -1,1393204 | -0,1881735 | WNT6 | Homo sapiens wingless-type MMTV integration site family, member 6 (WNT6), mRNA [NM_006522] |
| A_24_P333697 | 0,0683104 | up | 5,4190063 | 2,4380283 | KLK13 | Homo sapiens kallikrein-related peptidase 13 (KLK13), mRNA [NM_015596] |
| A_23_P320261 | 0,7880974 | down | -1,1250305 | -0,1699641 | DMKN | Homo sapiens dermokine (DMKN), transcript variant 1, mRNA [NM_001035516] |
| A_23_P128919 | 0,10295703 | up | 1,3114893 | 0,3912061 | LGALS3 | Homo sapiens lectin, galactoside-binding, soluble, 3 (LGALS3), transcript variant 1, mRNA [NM_002306] |
| A_23_P502470 | 0,4368551 | down | -1,1550024 | -0,2078958 | IL6ST | Homo sapiens interleukin 6 signal transducer (IL6ST), transcript variant 1, mRNA [NM_002184] |
| A_23_P129556 | 0,4955782 | up | 1,2267578 | 0,2948504 | IL4R | Homo sapiens interleukin 4 receptor (IL4R), transcript variant 1, mRNA [NM_000418] |
| A_33_P3282018 | 0,4284756 | up | 1,2137663 | 0,2794907 | MCF2L | Homo sapiens MCF.2 cell line derived transforming sequence-like (MCF2L), transcript variant 2, mRNA [NM_024979] |
| A_23_P259071 | 0,29842657 | up | 1,9589301 | 0,97006595 | AREG | Homo sapiens amphiregulin (AREG), mRNA [NM_001657] |
| A_23_P144959 | 0,02631055 | down | -2,1505022 | -1,1046736 | VCAN | Homo sapiens versican (VCAN), transcript variant 1, mRNA [NM_004385] |
| A_23_P102611 | 0,79480326 | up | 1,1460239 | 0,1966371 | WISP2 | Homo sapiens WNT1 inducible signaling pathway protein 2 (WISP2), mRNA [NM_003881] |
| A_24_P354689 | 0,04596365 | up | 2,2133596 | 1,1462379 | SPOCK1 | Homo sapiens sparc/osteonectin, cwcv and kazal-like domains proteoglycan (testican) 1 (SPOCK1), mRNA [NM_004598] |
| A_23_P334021 | 0,5739936 | down | -1,0969574 | -0,1335076 | IGF2R | Homo sapiens insulin-like growth factor 2 receptor (IGF2R), mRNA [NM_000876] |
| A_33_P3389286 | 0,38915217 | up | 1,5072012 | 0,591872 | SFN | Homo sapiens stratifin (SFN), mRNA [NM_006142] |
| A_23_P500010 | 0,01925241 | up | 15,19849 | 3,925856 | KLK12 | Homo sapiens kallikrein-related peptidase 12 (KLK12), transcript variant 2, mRNA [NM_145894] |
| A_23_P153676 | 0,5263597 | up | 1,2131788 | 0,27879214 | TLE2 | Homo sapiens transducin-like enhancer of split 2 (TLE2), transcript variant 1, mRNA [NM_003260] |
| A_33_P3406240 | 0,73250276 | up | 1,0974377 | 0,13413909 | GDF7 | Homo sapiens growth differentiation factor 7 (GDF7), mRNA [NM_182828] |
| A_33_P3358745 | 0,16514464 | up | 1,745358 | 0,80352294 | SEPP1 | Homo sapiens selenoprotein P, plasma, 1 (SEPP1), transcript variant 3, mRNA [NM_001093726] |
| A_32_P77977 | 0,13513838 | down | -1,3085747 | -0,3879963 | UTP11L | Homo sapiens UTP11-like, U3 small nucleolar ribonucleoprotein (yeast) (UTP11L), mRNA [NM_016037] |
| A_23_P138524 | 0,82496345 | up | 1,1494 | 0,20088094 | CPXM2 | Homo sapiens carboxypeptidase X (M14 family), member 2 (CPXM2), mRNA [NM_198148] |
| A_33_P3235776 | 0,04874857 | down | -1,4853683 | -0,5708206 | FAM184A | Homo sapiens mRNA; cDNA DKFZp686I01262 (from clone DKFZp686I01262). [BX640728] |
| A_24_P234838 | 0,24969636 | up | 1,23346 | 0,30271086 | PCDH1 | Homo sapiens protocadherin 1 (PCDH1), transcript variant 2, mRNA [NM_032420] |
| A_23_P158775 | 0,5717862 | up | 1,0727072 | 0,1012563 | UMOD | Homo sapiens uromodulin (UMOD), transcript variant 1, mRNA [NM_003361] |
| A_23_P144348 | 0,2556927 | down | -1,3358827 | -0,4177933 | SLIT2 | Homo sapiens slit homolog 2 (Drosophila) (SLIT2), transcript variant 1, mRNA [NM_004787] |
| A_33_P3382177 | 0,01308363 | up | 1,9477427 | 0,9618031 | TIMP2 | Homo sapiens TIMP metallopeptidase inhibitor 2 (TIMP2), mRNA [NM_003255] |
| A_33_P3250398 | 0,7583395 | up | 1,1204563 | 0,16408643 | CTSZ | cathepsin Z [Source:HGNC Symbol;Acc:HGNC:2547] [ENST00000503833] |
| A_23_P156687 | 0,4163314 | up | 1,3588357 | 0,442371 | CFB | Homo sapiens complement factor B (CFB), mRNA [NM_001710] |
| A_23_P34433 | 0,2762636 | down | -1,2183589 | -0,2849391 | ZCCHC11 | Homo sapiens zinc finger, CCHC domain containing 11 (ZCCHC11), transcript variant 1, mRNA [NM_001009881] |
| A_23_P26325 | 0,75672626 | down | -1,1119218 | -0,1530553 | CCL17 | Homo sapiens chemokine (C-C motif) ligand 17 (CCL17), mRNA [NM_002987] |
| A_33_P3372727 | 0,07576662 | down | -1,691796 | -0,7585555 | SEMA5A | Homo sapiens sema domain, seven thrombospondin repeats (type 1 and type 1-like), transmembrane domain (TM) and short cytoplasmic domain, (semaphorin) 5A (SE |
| A_33_P3419190 | 0,28170082 | up | 1,9492501 | 0,9629192 | AREG | Homo sapiens amphiregulin (AREG), mRNA [NM_001657] |
| A_32_P64200 | 0,8206922 | down | -1,0534322 | -0,0750975 | GUCA1B | Homo sapiens guanylate cyclase activator 1B (retina) (GUCA1B), mRNA [NM_002098] |
| A_23_P303145 | 0,30297124 | down | -1,2815291 | -0,3578662 | FGFR2 | fibroblast growth factor receptor 2 [Source:HGNC Symbol;Acc:HGNC:3689] [ENST00000359354] |
| A_33_P3406962 | 0,57639 | up | 1,1361437 | 0,1841453 | GATA4 | GATA binding protein 4 [Source:HGNC Symbol;Acc:HGNC:4173] [ENST00000532977] |
| A_19_P00800682 | 0,74428135 | up | 1,0662414 | 0,09253408 | RDX | Homo sapiens radixin (RDX), transcript variant 1, mRNA [NM_001260492] |
| A_23_P89270 | 0,11842229 | down | -1,2449404 | -0,3160767 | SERPINF2 | Homo sapiens serpin peptidase inhibitor, clade F (alpha-2 antiplasmin, pigment epithelium derived factor), member 2 (SERPINF2), transcript variant 1, mRNA [NM_000 |
| A_33_P3367917 | 0,2274422 | up | 1,2086918 | 0,27344647 | SSH2 | Homo sapiens slingshot protein phosphatase 2 (SSH2), transcript variant 3, mRNA [NM_001282130] |
| A_33_P3385101 | 0,3453531 | up | 1,2872536 | 0,36429632 | TOLLIP | Homo sapiens toll interacting protein (TOLLIP), mRNA [NM_019009] |
| A_33_P3348719 | 0,06617496 | down | -2,2722015 | -1,1840907 | FGF9 | Homo sapiens fibroblast growth factor 9 (FGF9), mRNA [NM_002010] |
| A_23_P34066 | 0,36099702 | up | 1,126249 | 0,17152576 | IL9R | Homo sapiens interleukin 9 receptor (IL9R), transcript variant 2, mRNA [NM_176786] |
| A_23_P31945 | 0,52742183 | down | -1,2387152 | -0,3088445 | IL33 | Homo sapiens interleukin 33 (IL33), transcript variant 1, mRNA [NM_033439] |

| | | | | | | |
|---|---|---|---|---|---|---|
| A_23_P63681 | 0,4949503 | up | 1,1018362 | 0,13990977 | IDE | Homo sapiens insulin-degrading enzyme (IDE), transcript variant 1, mRNA [NM_004969] |
| A_23_P169437 | 0,2865321 | up | 1,497986 | 0,5830241 | LCN2 | Homo sapiens lipocalin 2 (LCN2), mRNA [NM_005564] |
| A_33_P3287537 | 0,3482802 | up | 1,2006979 | 0,2638732 | PMPCA | peptidase (mitochondrial processing) alpha [Source:HGNC Symbol;Acc:HGNC:18667] [ENST00000371720] |
| A_23_P58588 | 0,6657988 | up | 1,1573452 | 0,2108192 | SLIT3 | Homo sapiens slit homolog 3 (Drosophila) (SLIT3), transcript variant 2, mRNA [NM_003062] |
| A_23_P75283 | 0,05186959 | up | 1,5443143 | 0,62696636 | RBP4 | Homo sapiens retinol binding protein 4, plasma (RBP4), mRNA [NM_006744] |
| A_33_P3345036 | 0,18409589 | down | -1,686143 | -0,7537269 | POMC | Homo sapiens proopiomelanocortin (POMC), transcript variant 1, mRNA [NM_001035256] |
| A_33_P3420816 | 0,5954564 | down | -1,1745563 | -0,2321158 | GDF1 | Homo sapiens growth differentiation factor 1 (GDF1), mRNA [NM_001492] |
| A_23_P35444 | 0,5729875 | up | 1,3178179 | 0,39815104 | INA | Homo sapiens internexin neuronal intermediate filament protein, alpha (INA), mRNA [NM_032727] |
| A_23_P393645 | 0,6095445 | up | 1,0767506 | 0,10668418 | ADAMTS13 | Homo sapiens ADAM metallopeptidase with thrombospondin type 1 motif, 13 (ADAMTS13), transcript variant 1, mRNA [NM_139025] |
| A_33_P3254946 | 0,06388597 | up | 1,4988306 | 0,5838373 | PLEKHH3 | Homo sapiens pleckstrin homology domain containing, family H (with MyTH4 domain) member 3 (PLEKHH3), transcript variant 1, mRNA [NM_024927] |
| A_23_P88099 | 0,670123 | up | 1,1048598 | 0,14386335 | MCF2L | Homo sapiens MCF.2 cell line derived transforming sequence-like (MCF2L), transcript variant 2, mRNA [NM_024979] |
| A_33_P3358228 | 0,76741403 | down | -1,0980643 | -0,1349625 | VWA2 | von Willebrand factor A domain containing 2 [Source:HGNC Symbol;Acc:HGNC:24709] [ENST00000298715] |
| A_33_P3813128 | 0,27299392 | up | 1,226324 | 0,29434013 | SEMA3F | Homo sapiens sema domain, immunoglobulin domain (Ig), short basic domain, secreted, (semaphorin) 3F (SEMA3F), mRNA [NM_004186] |
| A_24_P48495 | 0,3333564 | up | 2,3229408 | 1,2159524 | LYPD3 | Homo sapiens LY6/PLAUR domain containing 3 (LYPD3), mRNA [NM_014400] |
| A_32_P11471 | 0,36461186 | up | 1,1486688 | 0,19996284 | FAU | Homo sapiens Finkel-Biskis-Reilly murine sarcoma virus (FBR-MuSV) ubiquitously expressed (FAU), mRNA [NM_001997] |
| A_33_P3377151 | 0,09478033 | up | 1,5694776 | 0,6502844 | CCL19 | Homo sapiens chemokine (C-C motif) ligand 19 (CCL19), mRNA [NM_006274] |
| A_23_P126706 | 0,00265243 | down | -1,6590347 | -0,7303441 | ANGPTL1 | Homo sapiens angiopoietin-like 1 (ANGPTL1), mRNA [NM_004673] |
| A_23_P114740 | 0,84138006 | up | 1,0375351 | 0,05316011 | CFH | Homo sapiens complement factor H (CFH), transcript variant 1, mRNA [NM_000186] |
| A_33_P3294821 | 0,8033371 | up | 1,044486 | 0,06279322 | OTOP1 | Homo sapiens otopetrin 1 (OTOP1), mRNA [NM_177998] |
| A_33_P3256920 | 0,5127338 | up | 1,1784123 | 0,23684442 | WNT7B | Homo sapiens wingless-type MMTV integration site family, member 7B (WNT7B), mRNA [NM_058238] |
| A_33_P3712341 | 0,24653506 | up | 1,2767123 | 0,35243344 | CXCL12 | Homo sapiens chemokine (C-X-C motif) ligand 12 (CXCL12), transcript variant 3, mRNA [NM_001033886] |
| A_23_P68511 | 0,85836345 | down | -1,0319899 | -0,0454289 | ANGPT4 | Homo sapiens angiopoietin 4 (ANGPT4), mRNA [NM_015985] |
| A_23_P160881 | 0,3915083 | down | -1,188239 | -0,248825 | SMPDL3B | Homo sapiens sphingomyelin phosphodiesterase, acid-like 3B (SMPDL3B), transcript variant 2, mRNA [NM_001009568] |
| A_33_P3306264 | 0,30848613 | up | 2,4107022 | 1,2694534 | LYPD3 | Homo sapiens LY6/PLAUR domain containing 3 (LYPD3), mRNA [NM_014400] |
| A_23_P128503 | 0,82812536 | up | 1,058491 | 0,08200899 | IL26 | Homo sapiens interleukin 26 (IL26), mRNA [NM_018402] |
| A_23_P62752 | 0,03748198 | up | 1,4984361 | 0,5834575 | NPPB | Homo sapiens natriuretic peptide B (NPPB), mRNA [NM_002521] |
| A_33_P3403153 | 0,14104201 | up | 1,295578 | 0,37359586 | C1QTNF2 | Homo sapiens C1q and tumor necrosis factor related protein 2 (C1QTNF2), mRNA [NM_031908] |
| A_23_P62115 | 0,8896978 | up | 1,0375867 | 0,05323188 | TIMP1 | Homo sapiens TIMP metallopeptidase inhibitor 1 (TIMP1), mRNA [NM_003254] |
| A_33_P3334828 | 0,37453774 | up | 1,1793306 | 0,23796819 | INSL3 | Homo sapiens insulin-like 3 (Leydig cell) (INSL3), transcript variant 1, mRNA [NM_001265587] |
| A_24_P225961 | 0,24994639 | down | -1,1747423 | -0,2323444 | DAG1 | Homo sapiens dystroglycan 1 (dystrophin-associated glycoprotein 1) (DAG1), transcript variant 2, mRNA [NM_004393] |
| A_33_P3329149 | 0,45838228 | up | 1,1515462 | 0,20357235 | CDK13 | Homo sapiens cyclin-dependent kinase 13 (CDK13), transcript variant 2, mRNA [NM_031267] |
| A_23_P319598 | 0,7585759 | down | -1,1099176 | -0,1504526 | C4BPB | Homo sapiens complement component 4 binding protein, beta (C4BPB), transcript variant 1, mRNA [NM_000716] |
| A_33_P3329153 | 0,2646967 | up | 1,2128174 | 0,2783624 | CDK13 | Homo sapiens cyclin-dependent kinase 13 (CDK13), transcript variant 2, mRNA [NM_031267] |
| A_24_P233488 | 0,78987676 | up | 1,0746723 | 0,10389686 | LIF | Homo sapiens leukemia inhibitory factor (LIF), transcript variant 1, mRNA [NM_002309] |
| A_33_P3241269 | 0,15326652 | down | -3,8331342 | -1,9385245 | CES1 | Homo sapiens carboxylesterase 1 (CES1), transcript variant 1, mRNA [NM_001025195] |
| A_23_P251562 | 0,37770638 | down | -1,1855508 | -0,2455575 | TUSC2 | Homo sapiens tumor suppressor candidate 2 (TUSC2), mRNA [NM_007275] |
| A_23_P137697 | 0,02278282 | down | -2,8710508 | -1,5215789 | SELP | Homo sapiens selectin P (granule membrane protein 140kDa, antigen CD62) (SELP), mRNA [NM_003005] |
| A_32_P45168 | 0,49527758 | down | -1,2524726 | -0,3247791 | IL6ST | Homo sapiens interleukin 6 signal transducer (IL6ST), transcript variant 1, mRNA [NM_002184] |
| A_23_P201368 | 0,7184853 | down | -1,0629795 | -0,0881137 | CTBS | Homo sapiens chitobiase, di-N-acetyl- (CTBS), mRNA [NM_004388] |
| A_23_P161125 | 0,8795419 | down | -1,0262887 | -0,0374367 | MOV10 | Homo sapiens Mov10 RISC complex RNA helicase (MOV10), transcript variant 1, mRNA [NM_020963] |
| A_33_P3235400 | 0,7850959 | down | -1,0765322 | -0,1063915 | HDGF | Homo sapiens hepatoma-derived growth factor (HDGF), transcript variant 1, mRNA [NM_004494] |
| A_23_P343411 | 0,33334625 | up | 1,1429163 | 0,19271977 | AGRN | Homo sapiens agrin (AGRN), mRNA [NM_198576] |
| A_33_P3318288 | 0,6769505 | up | 1,086215 | 0,11930972 | CFH | Homo sapiens complement factor H (CFH), transcript variant 2, mRNA [NM_001014975] |
| A_23_P85922 | 0,8283161 | down | -1,1240754 | -0,1687388 | BMP8A | Homo sapiens bone morphogenetic protein 8a (BMP8A), mRNA [NM_181809] |
| A_23_P10591 | 0,15794656 | up | 1,7483836 | 0,80602175 | METRNL | Homo sapiens meteorin, glial cell differentiation regulator-like (METRNL), mRNA [NM_001004431] |
| A_23_P4592 | 0,6359686 | up | 1,0977327 | 0,13452674 | SIGLEC6 | Homo sapiens sialic acid binding Ig-like lectin 6 (SIGLEC6), transcript variant 1, mRNA [NM_001245] |
| A_23_P151805 | 0,5805745 | down | -1,2720939 | -0,3472052 | FBLN5 | Homo sapiens fibulin 5 (FBLN5), mRNA [NM_006329] |
| A_24_P374943 | 0,8455259 | up | 1,0424312 | 0,05995222 | CXADR | Homo sapiens coxsackie virus and adenovirus receptor (CXADR), transcript variant 1, mRNA [NM_001338] |
| A_32_P107617 | 0,12109523 | up | 1,3857621 | 0,47067958 | SFTPD | Homo sapiens surfactant protein D (SFTPD), mRNA [NM_003019] |
| A_33_P3225273 | 0,02591554 | up | 1,6181318 | 0,6943291 | QSOX1 | Homo sapiens quiescin Q6 sulfhydryl oxidase 1 (QSOX1), transcript variant 2, mRNA [NM_001004128] |
| A_23_P137391 | 0,5679749 | up | 1,1473656 | 0,19832513 | ENO1 | Homo sapiens enolase 1, (alpha) (ENO1), transcript variant 1, mRNA [NM_001428] |
| A_23_P50946 | 0,3010118 | down | -2,1324046 | -1,0924811 | RAMP1 | Homo sapiens receptor (G protein-coupled) activity modifying protein 1 (RAMP1), mRNA [NM_005855] |
| A_23_P167818 | 0,00579134 | down | -2,2747097 | -1,1856824 | FAM184A | Homo sapiens family with sequence similarity 184, member A (FAM184A), transcript variant 1, mRNA [NM_024581] |
| A_33_P3229083 | 0,7897299 | down | -1,0677998 | -0,0946412 | HIST1H2BK | Homo sapiens histone cluster 1, H2bk (HIST1H2BK), mRNA [NM_080593] |
| A_33_P3369058 | 0,5219757 | up | 1,1812183 | 0,24027556 | LRRK2 | Homo sapiens leucine-rich repeat kinase 2 (LRRK2), mRNA [NM_198578] |

| | | | | | | |
|---|---|---|---|---|---|---|
| A_23_P145978 | 0,1458921 | up | 1,5081737 | 0,5928026 | VIPR2 | Homo sapiens vasoactive intestinal peptide receptor 2 (VIPR2), mRNA [NM_003382] |
| A_33_P3370714 | 0,40390402 | up | 1,1404431 | 0,18959445 | PGC | Homo sapiens progastricsin (pepsinogen C) (PGC), transcript variant 2, mRNA [NM_001166424] |
| A_23_P348028 | 0,01078235 | up | 4,0224257 | 2,0080657 | IL36A | Homo sapiens interleukin 36, alpha (IL36A), mRNA [NM_014440] |
| A_24_P178175 | 0,16577001 | down | -1,4117737 | -0,4975088 | GGT1 | Homo sapiens gamma-glutamyltransferase 1 (GGT1), transcript variant 6, mRNA [NM_001288833] |
| A_24_P12401 | 0,6805001 | up | 1,1282452 | 0,17408068 | VEGFA | Homo sapiens vascular endothelial growth factor A (VEGFA), transcript variant 1, mRNA [NM_001025366] |
| A_23_P4161 | 0,06815421 | down | -1,4992052 | -0,5841979 | ARSG | arylsulfatase G [Source:HGNC Symbol;Acc:HGNC:24102] [ENST00000448504] |
| A_33_P3258274 | 0,52993387 | up | 1,24083 | 0,3113054 | TFPI | tissue factor pathway inhibitor (lipoprotein-associated coagulation inhibitor) [Source:HGNC Symbol;Acc:HGNC:11760] [ENST00000481132] |
| A_32_P101689 | 0,0993974 | up | 1,2799153 | 0,35604838 | FAM3C | Homo sapiens family with sequence similarity 3, member C (FAM3C), transcript variant 1, mRNA [NM_014888] |
| A_19_P00801279 | 0,24581796 | up | 1,2637835 | 0,33774927 | MSN | moesin [Source:HGNC Symbol;Acc:HGNC:7373] [ENST00000429601] |
| A_23_P390518 | 0,40220457 | up | 1,1550874 | 0,20800196 | TNFRSF11A | Homo sapiens tumor necrosis factor receptor superfamily, member 11a, NFKB activator (TNFRSF11A), transcript variant 1, mRNA [NM_003839] |
| A_33_P3219303 | 0,6219149 | down | -1,1327394 | -0,179816 | CEL | Homo sapiens carboxyl ester lipase (CEL), mRNA [NM_001807] |
| A_23_P41314 | 0,6714734 | up | 1,0914263 | 0,12621465 | F11 | Homo sapiens coagulation factor XI (F11), mRNA [NM_000128] |
| A_23_P209904 | 0,3720718 | up | 1,2570806 | 0,3300771 | GPC1 | Homo sapiens glypican 1 (GPC1), mRNA [NM_002081] |
| A_23_P258367 | 0,51736015 | down | -1,1184916 | -0,1615545 | GLE1 | Homo sapiens GLE1 RNA export mediator (GLE1), transcript variant 2, mRNA [NM_001499] |
| A_33_P3389842 | 0,40357858 | down | -2,0033324 | -1,0024018 | PROM1 | Homo sapiens prominin 1 (PROM1), transcript variant 6, mRNA [NM_001145850] |
| A_23_P8702 | 0,07596428 | up | 2,469174 | 1,3040284 | PIP | Homo sapiens prolactin-induced protein (PIP), mRNA [NM_002652] |
| A_23_P30126 | 0,45160148 | up | 1,745951 | 0,80401313 | FGFBP1 | Homo sapiens fibroblast growth factor binding protein 1 (FGFBP1), mRNA [NM_005130] |
| A_33_P3343196 | 0,79308087 | up | 1,1325257 | 0,17954376 | CP | Homo sapiens ceruloplasmin (ferroxidase) (CP), transcript variant 1, mRNA [NM_000096] |
| A_24_P940135 | 0,7608518 | down | -1,0623423 | -0,0872487 | CTBS | Homo sapiens chitobiase, di-N-acetyl- (CTBS), mRNA [NM_004388] |
| A_23_P93180 | 0,6694449 | down | -1,1065236 | -0,1460343 | HIST1H2BC | Homo sapiens histone cluster 1, H2bc (HIST1H2BC), mRNA [NM_003526] |
| A_33_P3349536 | 0,75573003 | down | -1,130644 | -0,1771447 | CHEK1 | Homo sapiens checkpoint kinase 1 (CHEK1), transcript variant 2, mRNA [NM_001114121] |
| A_33_P3354935 | 0,01512028 | up | 1,4845966 | 0,570071 | CSF1 | Homo sapiens colony stimulating factor 1 (macrophage) (CSF1), transcript variant 4, mRNA [NM_172212] |
| A_23_P123853 | 0,5761214 | up | 1,2710243 | 0,34599167 | CCL19 | Homo sapiens chemokine (C-C motif) ligand 19 (CCL19), mRNA [NM_006274] |
| A_24_P142118 | 0,39768353 | up | 1,2352381 | 0,30478913 | THBS1 | Homo sapiens thrombospondin 1 (THBS1), mRNA [NM_003246] |
| A_23_P88963 | 0,23834299 | up | 1,2943769 | 0,3722577 | ALDOA | Homo sapiens aldolase A, fructose-bisphosphate (ALDOA), transcript variant 1, mRNA [NM_000034] |
| A_23_P381203 | 0,2466902 | down | -1,3096784 | -0,3892126 | KIAA0556 | Homo sapiens KIAA0556 (KIAA0556), mRNA [NM_015202] |
| A_23_P139648 | 0,08893253 | up | 1,3965098 | 0,48182565 | IAPP | Homo sapiens islet amyloid polypeptide (IAPP), mRNA [NM_000415] |
| A_33_P3777207 | 0,2873938 | down | -1,184128 | -0,2438251 | MMEL1 | Homo sapiens membrane metallo-endopeptidase-like 1 (MMEL1), mRNA [NM_033467] |
| A_33_P3249872 | 0,7533368 | up | 1,0853441 | 0,11815248 | FBLN1 | Homo sapiens fibulin 1 (FBLN1), transcript variant C, mRNA [NM_001996] |
| A_23_P134419 | 0,18109207 | up | 1,341608 | 0,42396325 | ZP3 | Homo sapiens zona pellucida glycoprotein 3 (sperm receptor) (ZP3), transcript variant 2, mRNA [NM_007155] |
| A_23_P28186 | 0,04939144 | down | -6,4130187 | -2,6810036 | SRD5A2 | Homo sapiens steroid-5-alpha-reductase, alpha polypeptide 2 (3-oxo-5 alpha-steroid delta 4-dehydrogenase alpha 2) (SRD5A2), mRNA [NM_000348] |
| A_33_P3258091 | 0,2159058 | down | -1,2302785 | -0,2989849 | RNPEP | Homo sapiens arginyl aminopeptidase (aminopeptidase B) (RNPEP), mRNA [NM_020216] |
| A_23_P212126 | 0,44878864 | down | -1,2467746 | -0,3182006 | COLQ | Homo sapiens collagen-like tail subunit (single strand of homotrimer) of asymmetric acetylcholinesterase (COLQ), transcript variant II, mRNA [NM_080538] |
| A_24_P310256 | 0,4334904 | down | -1,2413095 | -0,3118629 | LGI4 | Homo sapiens leucine-rich repeat LGI family, member 4 (LGI4), mRNA [NM_139284] |
| A_23_P501007 | 0,06154975 | down | -1,8156925 | -0,86052 | EFEMP1 | Homo sapiens EGF containing fibulin-like extracellular matrix protein 1 (EFEMP1), transcript variant 2, mRNA [NM_001039348] |
| A_24_P224116 | 0,7923767 | down | -1,069996 | -0,0976054 | PLA2G1B | Homo sapiens phospholipase A2, group IB (pancreas) (PLA2G1B), mRNA [NM_000928] |
| A_23_P71270 | 0,7171336 | down | -1,1887666 | -0,2494655 | AZGP1 | Homo sapiens alpha-2-glycoprotein 1, zinc-binding (AZGP1), mRNA [NM_001185] |
| A_33_P3289121 | 0,00446057 | down | -14,534434 | -3,861403 | C2orf40 | Homo sapiens chromosome 2 open reading frame 40 (C2orf40), mRNA [NM_032411] |
| A_23_P200741 | 0,16513768 | up | 1,1974376 | 0,25995052 | DPT | Homo sapiens dermatopontin (DPT), mRNA [NM_001937] |
| A_23_P347169 | 0,6240895 | up | 1,0907024 | 0,12525754 | MTUS1 | Homo sapiens microtubule associated tumor suppressor 1 (MTUS1), transcript variant 1, mRNA [NM_001001924] |
| A_33_P3232523 | 0,10584146 | up | 1,431817 | 0,5178472 | APOL4 | apolipoprotein L, 4 [Source:HGNC Symbol;Acc:HGNC:14867] [ENST00000328429] |
| A_23_P258164 | 0,8018543 | down | -1,0405959 | -0,0574099 | CORT | Homo sapiens cortistatin (CORT), mRNA [NM_001302] |
| A_23_P151895 | 0,5945771 | up | 1,2101682 | 0,27520764 | CILP | Homo sapiens cartilage intermediate layer protein, nucleotide pyrophosphohydrolase (CILP), mRNA [NM_003613] |
| A_23_P40880 | 0,85911644 | up | 1,0350605 | 0,04971513 | CMTM8 | Homo sapiens CKLF-like MARVEL transmembrane domain containing 8 (CMTM8), mRNA [NM_178868] |
| A_23_P117363 | 0,03295006 | down | -1,5169085 | -0,6011341 | SERPINA6 | Homo sapiens serpin peptidase inhibitor, clade A (alpha-1 antiproteinase, antitrypsin), member 6 (SERPINA6), mRNA [NM_001756] |
| A_24_P920125 | 0,21283756 | down | -1,2728367 | -0,3480473 | PPIA | Homo sapiens peptidylprolyl isomerase A (cyclophilin A) (PPIA), transcript variant 1, mRNA [NM_021130] |
| A_24_P91566 | 0,08801466 | up | 1,8728106 | 0,905205 | BMP7 | Homo sapiens bone morphogenetic protein 7 (BMP7), mRNA [NM_001719] |
| A_23_P70398 | 0,10391223 | up | 1,7564192 | 0,8126372 | VEGFA | Homo sapiens vascular endothelial growth factor A (VEGFA), transcript variant 6, mRNA [NM_001025370] |
| A_23_P77000 | 0,06966905 | up | 1,6337533 | 0,70819014 | VASH1 | Homo sapiens vasohibin 1 (VASH1), mRNA [NM_014909] |
| A_33_P3404706 | 0,4042094 | up | 1,2125465 | 0,27804002 | SPN | Homo sapiens sialophorin (SPN), transcript variant 1, mRNA [NM_001030288] |
| A_23_P372874 | 0,67027205 | up | 1,1173208 | 0,16004343 | S100A13 | Homo sapiens S100 calcium binding protein A13 (S100A13), transcript variant 1, mRNA [NM_001024210] |
| A_23_P126836 | 0,83426064 | up | 1,079036 | 0,10974299 | TNFSF4 | Homo sapiens tumor necrosis factor (ligand) superfamily, member 4 (TNFSF4), transcript variant 1, mRNA [NM_003326] |
| A_33_P3401301 | 0,4027336 | up | 1,1685921 | 0,22477144 | RPL39 | Homo sapiens ribosomal protein L39 (RPL39), mRNA [NM_001000] |
| A_23_P130158 | 0,5906038 | down | -1,1650492 | -0,2203909 | WNT3 | Homo sapiens wingless-type MMTV integration site family, member 3 (WNT3), mRNA [NM_030753] |
| A_23_P169503 | 0,11835163 | down | -1,5030733 | -0,5879154 | CEL | Homo sapiens carboxyl ester lipase (CEL), mRNA [NM_001807] |

| | | | | | | |
|---|---|---|---|---|---|---|
| A_24_P141707 | 0,12803572 | up | 1,2605679 | 0,33407384 | INHBE | Homo sapiens inhibin, beta E (INHBE), mRNA [NM_031479] |
| A_23_P251795 | 0,59613025 | up | 1,1137029 | 0,15536441 | GPC2 | glypican 2 [Source:HGNC Symbol;Acc:HGNC:4450] [ENST00000471050] |
| A_23_P257003 | 0,55515826 | down | -1,1446488 | -0,194905 | PCSK5 | Homo sapiens proprotein convertase subtilisin/kexin type 5 (PCSK5), transcript variant 2, mRNA [NM_006200] |
| A_23_P94338 | 0,07157236 | down | -2,1283453 | -1,0897322 | ENPP2 | Homo sapiens ectonucleotide pyrophosphatase/phosphodiesterase 2 (ENPP2), transcript variant 1, mRNA [NM_006209] |
| A_33_P3308105 | 0,8130788 | down | -1,0657431 | -0,0918597 | GGH | Homo sapiens gamma-glutamyl hydrolase (conjugase, folylpolygammaglutamyl hydrolase) (GGH), mRNA [NM_003878] |
| A_33_P3380625 | 0,7221832 | up | 1,1192695 | 0,16255744 | HSPG2 | Homo sapiens heparan sulfate proteoglycan 2 (HSPG2), transcript variant 1, mRNA [NM_001291860] |
| A_23_P142345 | 0,24605535 | up | 1,2227327 | 0,290109 | PRTN3 | Homo sapiens proteinase 3 (PRTN3), mRNA [NM_002777] |
| A_33_P3329522 | 0,22880343 | down | -2,145553 | -1,1013496 | LRRC17 | Homo sapiens leucine rich repeat containing 17 (LRRC17), transcript variant 1, mRNA [NM_001031692] |
| A_23_P254179 | 0,8223881 | down | -1,0380297 | -0,0538477 | ADNP | Homo sapiens activity-dependent neuroprotector homeobox (ADNP), transcript variant 1, mRNA [NM_015339] |
| A_23_P137665 | 0,4617525 | up | 1,951563 | 0,96463 | CHI3L1 | Homo sapiens chitinase 3-like 1 (cartilage glycoprotein-39) (CHI3L1), mRNA [NM_001276] |
| A_33_P3416797 | 0,0889748 | down | -1,6719537 | -0,7415349 | OVOS2 | Homo sapiens mRNA; cDNA DKFZp434C0631 (from clone DKFZp434C0631). [AL831947] |
| A_23_P8339 | 0,5530287 | down | -1,0883772 | -0,1221787 | MRPL18 | Homo sapiens mitochondrial ribosomal protein L18 (MRPL18), mRNA [NM_014161] |
| A_23_P374782 | 0,04932215 | up | 1,3415033 | 0,42385057 | SH3KBP1 | Homo sapiens SH3-domain kinase binding protein 1 (SH3KBP1), transcript variant 2, mRNA [NM_001024666] |
| A_23_P301942 | 0,41970804 | down | -1,6295407 | -0,7044654 | NPPC | Homo sapiens natriuretic peptide C (NPPC), mRNA [NM_024409] |
| A_23_P151907 | 0,8553153 | up | 1,0386598 | 0,05472321 | PCSK6 | Homo sapiens proprotein convertase subtilisin/kexin type 6 (PCSK6), transcript variant 1, mRNA [NM_002570] |
| A_23_P13094 | 0,06081602 | down | -5,4486094 | -2,445888 | MMP10 | Homo sapiens matrix metallopeptidase 10 (stromelysin 2) (MMP10), mRNA [NM_002425] |
| A_23_P218505 | 0,43827215 | up | 1,1427249 | 0,19247809 | LHB | Homo sapiens luteinizing hormone beta polypeptide (LHB), mRNA [NM_000894] |
| A_23_P94030 | 0,00257079 | down | -2,7461228 | -1,4573961 | LAMB1 | Homo sapiens laminin, beta 1 (LAMB1), mRNA [NM_002291] |
| A_23_P32454 | 0,7448028 | up | 1,1340425 | 0,1814747 | TG | Homo sapiens thyroglobulin (TG), mRNA [NM_003235] |
| A_23_P409623 | 0,26326415 | up | 1,2043381 | 0,26824042 | PPFIBP2 | Homo sapiens PTPRF interacting protein, binding protein 2 (liprin beta 2) (PPFIBP2), transcript variant 1, mRNA [NM_003621] |
| A_33_P3273534 | 0,45278168 | down | -1,1279244 | -0,1736704 | KRT81 | Homo sapiens keratin 81, type II (KRT81), mRNA [NM_002281] |
| A_23_P50919 | 0,6705037 | up | 1,2898628 | 0,36721757 | SERPINE2 | Homo sapiens serpin peptidase inhibitor, clade E (nexin, plasminogen activator inhibitor type 1), member 2 (SERPINE2), transcript variant 1, mRNA [NM_006216] |
| A_33_P3321070 | 0,81695294 | down | -1,0591975 | -0,0829717 | WNT4 | wingless-type MMTV integration site family, member 4 [Source:HGNC Symbol;Acc:HGNC:12783] [ENST00000415567] |
| A_24_P215804 | 0,02400749 | down | -1,8088361 | -0,8550617 | CKLF | Homo sapiens chemokine-like factor (CKLF), transcript variant 1, mRNA [NM_016951] |
| A_32_P86150 | 0,1998165 | up | 2,2523708 | 1,1714444 | CTRB2 | Homo sapiens chymotrypsinogen B2 (CTRB2), mRNA [NM_001025200] |
| A_33_P3231953 | 0,72198004 | down | -1,1539358 | -0,206563 | COL12A1 | Homo sapiens collagen, type XII, alpha 1 (COL12A1), transcript variant long, mRNA [NM_004370] |
| A_23_P501722 | 0,38203958 | up | 1,2619864 | 0,33569634 | TSPAN32 | Homo sapiens tetraspanin 32 (TSPAN32), mRNA [NM_139022] |
| A_24_P48204 | 0,17173181 | up | 1,5870416 | 0,66633993 | SECTM1 | Homo sapiens secreted and transmembrane 1 (SECTM1), mRNA [NM_003004] |
| A_33_P3316621 | 0,02881983 | up | 1,8554925 | 0,89180213 | BMP3 | Homo sapiens bone morphogenetic protein 3 (BMP3), mRNA [NM_001201] |
| A_24_P376707 | 0,64295334 | down | -1,0911689 | -0,1258744 | HDGF | Homo sapiens hepatoma-derived growth factor (HDGF), transcript variant 1, mRNA [NM_004494] |
| A_33_P3339036 | 0,38835976 | up | 1,1762421 | 0,23418504 | MECP2 | Homo sapiens methyl CpG binding protein 2 (MECP2), transcript variant 2, mRNA [NM_001110792] |
| A_33_P3245439 | 0,85022277 | up | 1,0680448 | 0,09497213 | CD40 | Homo sapiens CD40 molecule, TNF receptor superfamily member 5 (CD40), transcript variant 1, mRNA [NM_001250] |
| A_23_P105957 | 0,30492133 | down | -1,2792497 | -0,3552979 | ACTN1 | Homo sapiens actinin, alpha 1 (ACTN1), transcript variant 2, mRNA [NM_001102] |
| A_23_P331748 | 0,02073113 | up | 1,6420798 | 0,71552426 | CD33 | Homo sapiens CD33 molecule (CD33), transcript variant 1, mRNA [NM_001772] |
| A_23_P126278 | 0,29043606 | up | 1,2826343 | 0,35910985 | CHIT1 | Homo sapiens chitinase 1 (chitotriosidase) (CHIT1), transcript variant 1, mRNA [NM_003465] |
| A_32_P220307 | 0,82436186 | up | 1,0437423 | 0,06176555 | RPL39 | Homo sapiens ribosomal protein L39 (RPL39), mRNA [NM_001000] |
| A_33_P3390758 | 0,13286002 | down | -1,4013329 | -0,4867997 | HSPA8 | Homo sapiens heat shock 70kDa protein 8 (HSPA8), transcript variant 2, mRNA [NM_153201] |
| A_33_P3395743 | 0,06270421 | up | 1,4942551 | 0,5794264 | VWA1 | Homo sapiens von Willebrand factor A domain containing 1 (VWA1), transcript variant 1, mRNA [NM_022834] |
| A_23_P38427 | 0,25858882 | up | 1,2941059 | 0,37195566 | RAB11FIP4 | RAB11 family interacting protein 4 (class II) [Source:HGNC Symbol;Acc:HGNC:30267] [ENST00000621161] |
| A_23_P160751 | 0,17797971 | down | -1,5748284 | -0,6551946 | FCRL2 | Homo sapiens Fc receptor-like 2 (FCRL2), transcript variant 1, mRNA [NM_030764] |
| A_33_P3420900 | 0,07413913 | down | -1,4631288 | -0,5490568 | PATE2 | Homo sapiens prostate and testis expressed 2 (PATE2), mRNA [NM_212555] |
| A_33_P3219811 | 0,30102584 | up | 1,2174737 | 0,28389066 | PTGDS | prostaglandin D2 synthase 21kDa (brain) [Source:HGNC Symbol;Acc:HGNC:9592] [ENST00000371623] |
| A_33_P3298539 | 0,677058 | down | -1,1035239 | -0,1421178 | APOA1 | Homo sapiens apolipoprotein A-I (APOA1), mRNA [NM_000039] |
| A_23_P360797 | 0,01049556 | down | -2,345924 | -1,2301562 | NTF3 | Homo sapiens neurotrophin 3 (NTF3), transcript variant 2, mRNA [NM_002527] |
| A_24_P410017 | 0,5811428 | down | -1,1351291 | -0,1828564 | POTEI | Homo sapiens POTE ankyrin domain family, member I (POTEI), mRNA [NM_001277406] |
| A_23_P25069 | 0,03732549 | down | -2,2249146 | -1,15375 | OVOS2 | |
| A_33_P3395952 | 0,2504861 | up | 1,222438 | 0,28976128 | COL20A1 | Homo sapiens collagen, type XX, alpha 1 (COL20A1), mRNA [NM_020882] |
| A_33_P3417487 | 0,30207336 | up | 1,5794387 | 0,6594119 | SCUBE1 | signal peptide, CUB domain, EGF-like 1 [Source:HGNC Symbol;Acc:HGNC:13441] [ENST00000290460] |
| A_24_P222655 | 0,1195258 | up | 1,5423589 | 0,62513846 | C1QA | Homo sapiens complement component 1, q subcomponent, A chain (C1QA), mRNA [NM_015991] |
| A_33_P3277178 | 0,88745403 | up | 1,028123 | 0,0400129 | SSPO | SCO-spondin [Source:HGNC Symbol;Acc:HGNC:21998] [ENST00000472850] |
| A_23_P22444 | 0,60848933 | down | -1,1694721 | -0,2258574 | CFP | Homo sapiens complement factor properdin (CFP), transcript variant 1, mRNA [NM_002621] |
| A_24_P13285 | 0,20758834 | up | 1,2795748 | 0,35566443 | PPP1R1A | Homo sapiens protein phosphatase 1, regulatory (inhibitor) subunit 1A (PPP1R1A), mRNA [NM_006741] |
| A_33_P3302255 | 0,28009817 | up | 1,2091353 | 0,27397567 | ITM2B | Homo sapiens integral membrane protein 2B (ITM2B), mRNA [NM_021999] |
| A_33_P3395947 | 0,79866034 | up | 1,1135608 | 0,15518032 | IL4R | Homo sapiens interleukin 4 receptor (IL4R), transcript variant 4, mRNA [NM_001257407] |
| A_24_P133253 | 0,07178813 | down | -1,6279889 | -0,7030909 | KITLG | Homo sapiens KIT ligand (KITLG), transcript variant b, mRNA [NM_000899] |

| | | | | | | |
|---|---|---|---|---|---|---|
| A_33_P3250383 | 0,05453686 | up | 1,250674 | 0,3227058 | CNP | Homo sapiens 2',3'-cyclic nucleotide 3' phosphodiesterase (CNP), mRNA [NM_033133] |
| A_23_P2283 | 0,07695877 | down | -1,7993964 | -0,847513 | TAC3 | Homo sapiens tachykinin 3 (TAC3), transcript variant 1, mRNA [NM_013251] |
| A_24_P164731 | 0,5028796 | down | -1,120706 | -0,1644078 | TMED1 | Homo sapiens transmembrane emp24 protein transport domain containing 1 (TMED1), transcript variant 1, mRNA [NM_006858] |
| A_19_P00806659 | 0,27439347 | up | 1,281636 | 0,35798657 | ITM2B | integral membrane protein 2B [Source:HGNC Symbol;Acc:HGNC:6174] [ENST00000378565] |
| A_23_P17053 | 0,17969406 | up | 4,632178 | 2,2116907 | IL36G | Homo sapiens interleukin 36, gamma (IL36G), transcript variant 1, mRNA [NM_019618] |
| A_23_P60248 | 0,5708135 | up | 1,0909346 | 0,12556466 | TXN | Homo sapiens thioredoxin (TXN), transcript variant 1, mRNA [NM_003329] |
| A_23_P95213 | 0,487733 | up | 1,1480356 | 0,19916743 | SFTPC | Homo sapiens surfactant protein C (SFTPC), transcript variant 1, mRNA [NM_003018] |
| A_33_P3256347 | 0,76318324 | down | -1,044808 | -0,0632379 | TIMM8B | Homo sapiens translocase of inner mitochondrial membrane 8 homolog B (yeast) (TIMM8B), transcript variant 1, mRNA [NM_012459] |
| A_33_P3305571 | 0,14696789 | up | 2,0127962 | 1,009201 | TNFRSF6B | Homo sapiens tumor necrosis factor receptor superfamily, member 6b, decoy (TNFRSF6B), mRNA [NM_003823] |
| A_23_P138760 | 0,8801044 | up | 1,0478337 | 0,06740974 | CLCF1 | Homo sapiens cardiotrophin-like cytokine factor 1 (CLCF1), transcript variant 1, mRNA [NM_013246] |
| A_23_P5654 | 0,5390699 | down | -1,1474082 | -0,1983788 | IL37 | Homo sapiens interleukin 37 (IL37), transcript variant 1, mRNA [NM_014439] |
| A_33_P3360363 | 0,2662149 | down | -1,2793485 | -0,3554093 | GATA4 | Homo sapiens GATA binding protein 4 (GATA4), mRNA [NM_002052] |
| A_23_P140876 | 0,51780075 | up | 1,1936798 | 0,2554159 | ABCA3 | Homo sapiens ATP-binding cassette, sub-family A (ABC1), member 3 (ABCA3), mRNA [NM_001089] |
| A_23_P45099 | 0,2284903 | down | -1,7294619 | -0,7903233 | HLA-DRB5 | Homo sapiens major histocompatibility complex, class II, DR beta 5 (HLA-DRB5), mRNA [NM_002125] |
| A_33_P3310104 | 0,553281 | up | 1,43092 | 0,51694304 | SERPINB5 | Homo sapiens serpin peptidase inhibitor, clade B (ovalbumin), member 5 (SERPINB5), mRNA [NM_002639] |
| A_24_P146683 | 0,03808302 | down | -3,1188772 | -1,6410267 | MSMB | Homo sapiens microseminoprotein, beta- (MSMB), transcript variant PSP94, mRNA [NM_002443] |
| A_33_P3228322 | 0,22573335 | up | 1,3500763 | 0,43304095 | IL18BP | Homo sapiens interleukin 18 binding protein (IL18BP), transcript variant A, mRNA [NM_173042] |
| A_24_P179351 | 0,26968503 | up | 1,1639066 | 0,21897526 | TPT1 | Homo sapiens tumor protein, translationally-controlled 1 (TPT1), transcript variant 2, mRNA [NM_003295] |
| A_23_P9485 | 0,19331434 | up | 1,2422937 | 0,3130063 | ORM2 | Homo sapiens orosomucoid 2 (ORM2), mRNA [NM_000608] |
| A_24_P355944 | 0,16512334 | up | 1,6418251 | 0,71530044 | EFNB2 | Homo sapiens ephrin-B2 (EFNB2), mRNA [NM_004093] |
| A_33_P3399101 | 0,26597327 | up | 1,227568 | 0,29580298 | CTSL | cathepsin L [Source:HGNC Symbol;Acc:HGNC:2537] [ENST00000342020] |
| A_33_P3295358 | 0,10314799 | up | 3,8696668 | 1,9522094 | ANGPTL4 | Homo sapiens angiopoietin-like 4 (ANGPTL4), transcript variant 1, mRNA [NM_139314] |
| A_24_P79403 | 0,8700704 | up | 1,0461112 | 0,06503625 | PF4 | Homo sapiens platelet factor 4 (PF4), mRNA [NM_002619] |
| A_33_P3349883 | 0,47848013 | up | 1,1819345 | 0,24115005 | LGALS8 | lectin, galactoside-binding, soluble, 8 [Source:HGNC Symbol;Acc:HGNC:6569] [ENST00000366583] |
| A_23_P376488 | 0,4001044 | up | 1,4580382 | 0,5440285 | TNF | Homo sapiens tumor necrosis factor (TNF), mRNA [NM_000594] |
| A_24_P257416 | 0,7657004 | down | -1,2361612 | -0,3058669 | CXCL2 | Homo sapiens chemokine (C-X-C motif) ligand 2 (CXCL2), mRNA [NM_002089] |
| A_33_P3215277 | 0,8882146 | down | -1,0470353 | -0,0663101 | TTBK2 | Homo sapiens tau tubulin kinase 2 (TTBK2), mRNA [NM_173500] |
| A_32_P210642 | 0,7509557 | up | 1,1046287 | 0,1435615 | EGFL7 | Homo sapiens EGF-like-domain, multiple 7 (EGFL7), transcript variant 2, mRNA [NM_201446] |
| A_33_P3446495 | 0,3179874 | up | 1,2140023 | 0,2797711 | FRMD4B | Homo sapiens FERM domain containing 4B (FRMD4B), mRNA [NM_015123] |
| A_23_P327361 | 0,46077046 | down | -1,25742 | -0,3304665 | DMXL2 | Homo sapiens Dmx-like 2 (DMXL2), transcript variant 2, mRNA [NM_015263] |
| A_23_P45185 | 0,6647027 | down | -1,0993534 | -0,1366553 | FIGF | Homo sapiens c-fos induced growth factor (vascular endothelial growth factor D) (FIGF), mRNA [NM_004469] |
| A_19_P00317164 | 0,82317036 | down | -1,0588036 | -0,0824349 | RDX | Homo sapiens radixin (RDX), transcript variant 1, mRNA [NM_001260492] |
| A_24_P391368 | 0,3564526 | down | -1,1487865 | -0,2001108 | ATXN10 | Homo sapiens ataxin 10 (ATXN10), transcript variant 1, mRNA [NM_013236] |
| A_23_P154784 | 0,36247012 | down | -2,1018577 | -1,0716649 | BPIFB1 | Homo sapiens BPI fold containing family B, member 1 (BPIFB1), mRNA [NM_033197] |
| A_23_P204144 | 0,04669525 | up | 1,596086 | 0,6745384 | KRT85 | Homo sapiens keratin 85, type II (KRT85), transcript variant 1, mRNA [NM_002283] |
| A_33_P3759611 | 0,17097618 | up | 1,2508175 | 0,32287136 | PDE4C | Homo sapiens phosphodiesterase 4C, cAMP-specific (PDE4C), transcript variant 1, mRNA [NM_000923] |
| A_23_P349416 | 0,55104685 | up | 1,1039332 | 0,1426529 | ERBB3 | Homo sapiens v-erb-b2 avian erythroblastic leukemia viral oncogene homolog 3 (ERBB3), transcript variant 1, mRNA [NM_001982] |
| A_33_P3351955 | 0,3344703 | up | 1,1990498 | 0,2618916 | EGFR | Homo sapiens epidermal growth factor receptor (EGFR), transcript variant 2, mRNA [NM_201282] |
| A_24_P336551 | 0,5078569 | up | 1,1252565 | 0,17025395 | BGLAP | Homo sapiens bone gamma-carboxyglutamate (gla) protein (BGLAP), mRNA [NM_199173] |
| A_33_P3362367 | 0,5941683 | up | 1,1221768 | 0,16629994 | RTN3 | reticulon 3 [Source:HGNC Symbol;Acc:HGNC:10469] [ENST00000338850] |
| A_23_P363778 | 0,11206271 | up | 1,823424 | 0,86665004 | FRZB | Homo sapiens frizzled-related protein (FRZB), mRNA [NM_001463] |
| A_23_P210482 | 0,13323732 | up | 1,6585921 | 0,72995913 | ADA | Homo sapiens adenosine deaminase (ADA), mRNA [NM_000022] |
| A_23_P356139 | 0,24692777 | up | 1,1811075 | 0,2401403 | FAM178A | Homo sapiens family with sequence similarity 178, member A (FAM178A), transcript variant 1, mRNA [NM_018121] |
| A_33_P3278410 | 0,01973846 | up | 1,6688955 | 0,73889357 | MLLT4 | Homo sapiens myeloid/lymphoid or mixed-lineage leukemia (trithorax homolog, Drosophila); translocated to, 4 (MLLT4), transcript variant 1, mRNA [NM_001207008] |
| A_33_P3236177 | 0,65641 | down | -1,2297162 | -0,2983254 | ANG | Homo sapiens angiogenin, ribonuclease, RNase A family, 5 (ANG), transcript variant 1, mRNA [NM_001145] |
| A_23_P6771 | 0,24172048 | down | -1,3757448 | -0,4602129 | LMCD1 | Homo sapiens LIM and cysteine-rich domains 1 (LMCD1), transcript variant 1, mRNA [NM_014583] |
| A_23_P119562 | 0,00915929 | up | 2,1721547 | 1,1191268 | CFD | Homo sapiens complement factor D (adipsin) (CFD), mRNA [NM_001928] |
| A_23_P155755 | 0,64459795 | up | 1,466236 | 0,55211735 | CXCL6 | Homo sapiens chemokine (C-X-C motif) ligand 6 (CXCL6), mRNA [NM_002993] |
| A_33_P3413989 | 0,25225434 | up | 1,7029847 | 0,76806545 | SERPING1 | Homo sapiens serpin peptidase inhibitor, clade G (C1 inhibitor), member 1 (SERPING1), transcript variant 1, mRNA [NM_000062] |
| A_33_P3284508 | 0,05504459 | up | 1,6352898 | 0,7095463 | CD14 | Homo sapiens CD14 molecule (CD14), transcript variant 3, mRNA [NM_001174104] |
| A_24_P211565 | 0,24405786 | up | 1,3514144 | 0,43447018 | C1QTNF6 | Homo sapiens C1q and tumor necrosis factor related protein 6 (C1QTNF6), transcript variant 1, mRNA [NM_031910] |
| A_23_P257834 | 0,41147798 | down | -1,246076 | -0,3173921 | ALB | Homo sapiens albumin (ALB), mRNA [NM_000477] |
| A_23_P129695 | 0,0971243 | up | 1,8978043 | 0,9243312 | VASN | Homo sapiens vasorin (VASN), mRNA [NM_138440] |
| A_33_P3344332 | 0,75814736 | down | -1,0631286 | -0,0883161 | RPL39 | Homo sapiens ribosomal protein L39 (RPL39), mRNA [NM_001000] |
| A_23_P146456 | 0,11079074 | up | 2,6434076 | 1,4023988 | CTSV | Homo sapiens cathepsin V (CTSV), transcript variant 1, mRNA [NM_001333] |

| | | | | | | |
|---|---|---|---|---|---|---|
| A_32_P107029 | 0,30825534 | up | 1,2055022 | 0,26963422 | NAPSA | Homo sapiens napsin A aspartic peptidase (NAPSA), mRNA [NM_004851] |
| A_23_P256473 | 0,34803662 | down | -1,4980923 | -0,5831265 | SEMA3C | Homo sapiens sema domain, immunoglobulin domain (Ig), short basic domain, secreted, (semaphorin) 3C (SEMA3C), mRNA [NM_006379] |
| A_23_P145863 | 0,6150629 | up | 1,0975682 | 0,13431053 | S100A11 | Homo sapiens S100 calcium binding protein A11 (S100A11), mRNA [NM_005620] |
| A_24_P365807 | 0,6087325 | up | 1,20131 | 0,26460853 | EFNB1 | Homo sapiens ephrin-B1 (EFNB1), mRNA [NM_004429] |
| A_23_P61057 | 0,8390848 | down | -1,0529352 | -0,0744167 | IL16 | Homo sapiens interleukin 16 (IL16), transcript variant 1, mRNA [NM_004513] |
| A_33_P3243907 | 0,39003924 | up | 1,1456921 | 0,19621938 | CTSD | Homo sapiens cathepsin D (CTSD), mRNA [NM_001909] |
| A_23_P42282 | 0,8189911 | down | -1,0777144 | -0,107975 | C4B | Homo sapiens complement component 4B (Chido blood group) (C4B), mRNA [NM_001002029] |
| A_23_P145657 | 0,09979409 | down | -1,4685206 | -0,5543636 | STAG3 | Homo sapiens stromal antigen 3 (STAG3), transcript variant 1, mRNA [NM_012447] |
| A_24_P239176 | 0,08429401 | up | 1,689817 | 0,756867 | MUC4 | Homo sapiens mucin 4, cell surface associated (MUC4), transcript variant 1, mRNA [NM_018406] |
| A_23_P28857 | 0,7999505 | up | 1,071452 | 0,09956725 | SIRPG | Homo sapiens signal-regulatory protein gamma (SIRPG), transcript variant 1, mRNA [NM_018556] |
| A_23_P209954 | 0,40411845 | up | 1,68817 | 0,75546014 | GNLY | Homo sapiens granulysin (GNLY), transcript variant 2, mRNA [NM_006433] |
| A_33_P3228266 | 0,2667955 | up | 1,1596209 | 0,2136532 | CST3 | Homo sapiens cystatin C (CST3), transcript variant 1, mRNA [NM_000099] |
| A_23_P164946 | 0,14063843 | down | -1,2393614 | -0,309597 | FKRP | Homo sapiens fukutin related protein (FKRP), transcript variant 2, mRNA [NM_001039885] |
| A_24_P334300 | 0,8958852 | down | -1,0440611 | -0,0622061 | FGF12 | Homo sapiens fibroblast growth factor 12 (FGF12), transcript variant 2, mRNA [NM_004113] |
| A_23_P24616 | 0,31082505 | up | 1,1613384 | 0,21578847 | SIAE | Homo sapiens sialic acid acetylesterase (SIAE), transcript variant 1, mRNA [NM_170601] |
| A_24_P73599 | 0,4642703 | down | -1,2477279 | -0,3193033 | IL16 | Homo sapiens interleukin 16 (IL16), transcript variant 2, mRNA [NM_172217] |
| A_23_P14892 | 0,75147676 | up | 1,0908022 | 0,1253895 | IGFALS | Homo sapiens insulin-like growth factor binding protein, acid labile subunit (IGFALS), transcript variant 2, mRNA [NM_004970] |
| A_24_P60845 | 0,82238597 | up | 1,0299517 | 0,04257667 | ACHE | Homo sapiens acetylcholinesterase (Yt blood group) (ACHE), transcript variant E4-E6, mRNA [NM_000665] |
| A_33_P3340342 | 0,7210401 | up | 1,0677977 | 0,0946383 | CMTM3 | Homo sapiens CKLF-like MARVEL transmembrane domain containing 3 (CMTM3), transcript variant 1, mRNA [NM_144601] |
| A_33_P3235940 | 0,06648849 | up | 11,334522 | 3,5026517 | KLK6 | Homo sapiens kallikrein-related peptidase 6 (KLK6), transcript variant B, mRNA [NM_001012964] |
| A_23_P119222 | 0,7093186 | up | 1,0915871 | 0,1264272 | RETN | Homo sapiens resistin (RETN), transcript variant 1, mRNA [NM_020415] |
| A_23_P98147 | 0,7248737 | down | -1,0510098 | -0,0717761 | CPN1 | Homo sapiens carboxypeptidase N, polypeptide 1 (CPN1), mRNA [NM_001308] |
| A_33_P3423121 | 0,54318035 | down | -1,1855518 | -0,2455587 | RDX | Homo sapiens radixin (RDX), transcript variant 1, mRNA [NM_001260492] |
| A_23_P154840 | 0,04950633 | down | -1,3157058 | -0,3958369 | SOD1 | Homo sapiens superoxide dismutase 1, soluble (SOD1), mRNA [NM_000454] |
| A_33_P3341586 | 0,56605357 | up | 1,1255666 | 0,17065142 | SIL1 | Homo sapiens SIL1 nucleotide exchange factor (SIL1), transcript variant 1, mRNA [NM_001037633] |
| A_23_P99076 | 0,06022827 | up | 1,3900781 | 0,4751659 | PRH2 | Homo sapiens proline-rich protein HaeIII subfamily 2 (PRH2), transcript variant 1, mRNA [NM_005042] |
| A_33_P3387145 | 0,02122792 | up | 1,3932657 | 0,4784704 | SH3KBP1 | Homo sapiens SH3-domain kinase binding protein 1 (SH3KBP1), transcript variant 1, mRNA [NM_031892] |
| A_24_P237175 | 0,8545447 | up | 1,0656685 | 0,09175868 | CST2 | Homo sapiens cystatin SA (CST2), mRNA [NM_001322] |
| A_24_P98161 | 0,17374851 | down | -1,2246224 | -0,2923369 | KRIT1 | Homo sapiens KRIT1, ankyrin repeat containing (KRIT1), transcript variant 4, mRNA [NM_194455] |
| A_23_P70688 | 0,53878653 | up | 1,1443509 | 0,19452949 | LY86 | Homo sapiens lymphocyte antigen 86 (LY86), mRNA [NM_004271] |
| A_24_P71661 | 0,4163749 | up | 1,1685972 | 0,22477776 | CRTAP | Homo sapiens cartilage associated protein (CRTAP), mRNA [NM_006371] |
| A_23_P502047 | 0,77770454 | down | -1,0631459 | -0,0883396 | CHRD | Homo sapiens chordin (CHRD), mRNA [NM_003741] |
| A_23_P133245 | 0,16760693 | down | -1,397838 | -0,4831972 | IK | Homo sapiens IK cytokine, down-regulator of HLA II (IK), mRNA [NM_006083] |
| A_23_P1981 | 0,09353574 | up | 1,2676328 | 0,34213695 | INS | Homo sapiens insulin (INS), transcript variant 1, mRNA [NM_000207] |
| A_19_P00806911 | 0,58475083 | down | -1,067303 | -0,0939697 | NUDT3 | nudix (nucleoside diphosphate linked moiety X)-type motif 3 [Source:HGNC Symbol;Acc:HGNC:8050] [ENST00000607016] |
| A_33_P3380612 | 0,36555845 | up | 1,2363744 | 0,30611566 | HSPG2 | Homo sapiens heparan sulfate proteoglycan 2 (HSPG2), transcript variant 2, mRNA [NM_005529] |
| A_23_P50638 | 0,8456727 | up | 1,0359018 | 0,05088722 | LRG1 | Homo sapiens leucine-rich alpha-2-glycoprotein 1 (LRG1), mRNA [NM_052972] |
| A_33_P3342780 | 0,75334805 | down | -1,0747534 | -0,1040057 | ADAMTS13 | Homo sapiens ADAM metallopeptidase with thrombospondin type 1 motif, 13 (ADAMTS13), transcript variant 3, mRNA [NM_139026] |
| A_33_P3296587 | 0,7458824 | up | 1,1714219 | 0,22826076 | CP | Homo sapiens ceruloplasmin (ferroxidase) (CP), transcript variant 1, mRNA [NM_000096] |
| A_23_P214168 | 0,62458456 | down | -1,3116938 | -0,391431 | COL12A1 | Homo sapiens collagen, type XII, alpha 1 (COL12A1), transcript variant long, mRNA [NM_004370] |
| A_23_P127891 | 0,33433864 | up | 1,3338414 | 0,4155872 | BDNF | Homo sapiens brain-derived neurotrophic factor (BDNF), transcript variant 1, mRNA [NM_170735] |
| A_23_P429363 | 0,11092257 | down | -1,5227097 | -0,6066409 | FGF17 | Homo sapiens fibroblast growth factor 17 (FGF17), mRNA [NM_003867] |
| A_33_P3399267 | 0,646034 | down | -1,1038474 | -0,1425407 | IL15RA | Homo sapiens interleukin 15 receptor, alpha (IL15RA), transcript variant 2, mRNA [NM_172200] |
| A_24_P319647 | 0,38028252 | up | 1,1937774 | 0,2555339 | FCRL2 | Homo sapiens Fc receptor-like 2 (FCRL2), transcript variant 1, mRNA [NM_030764] |
| A_33_P3258801 | 0,04011323 | up | 3,7630959 | 1,9119201 | SLURP1 | Homo sapiens secreted LY6/PLAUR domain containing 1 (SLURP1), mRNA [NM_020427] |
| A_23_P1083 | 0,6175711 | down | -1,1828359 | -0,24225 | GJA4 | Homo sapiens gap junction protein, alpha 4, 37kDa (GJA4), mRNA [NM_002060] |
| A_33_P3348714 | 0,02094986 | down | -3,1954486 | -1,6760185 | FGF9 | Homo sapiens fibroblast growth factor 9 (FGF9), mRNA [NM_002010] |
| A_33_P3210965 | 0,06283026 | down | -2,408892 | -1,2683697 | TCTN1 | Homo sapiens tectonic family member 1 (TCTN1), transcript variant 1, mRNA [NM_001082538] |
| A_33_P3261293 | 0,33264962 | down | -1,4775397 | -0,5631968 | DKK3 | Homo sapiens dickkopf WNT signaling pathway inhibitor 3 (DKK3), transcript variant 1, mRNA [NM_015881] |
| A_33_P3306749 | 0,6172264 | up | 1,123037 | 0,16740543 | OLFM1 | olfactomedin 1 [Source:HGNC Symbol;Acc:HGNC:17187] [ENST00000615948] |
| A_33_P3312730 | 0,08502335 | up | 1,7653186 | 0,8199286 | BHLHA15 | Homo sapiens basic helix-loop-helix family, member a15 (BHLHA15), mRNA [NM_177455] |
| A_23_P135722 | 0,5942069 | down | -1,2097735 | -0,274737 | BTC | Homo sapiens betacellulin (BTC), mRNA [NM_001729] |
| A_23_P34018 | 0,72269154 | up | 1,0646168 | 0,09033424 | RPL39 | Homo sapiens ribosomal protein L39 (RPL39), mRNA [NM_001000] |
| A_33_P3406623 | 0,77747667 | up | 1,046529 | 0,06561237 | TNFSF12 | Homo sapiens tumor necrosis factor (ligand) superfamily, member 12 (TNFSF12), transcript variant 1, mRNA [NM_003809] |
| A_32_P85999 | 0,5064058 | down | -1,6284243 | -0,7034766 | CDH13 | Homo sapiens cadherin 13 (CDH13), transcript variant 1, mRNA [NM_001257] |

| | | | | | | |
|---|---|---|---|---|---|---|
| A_23_P256603 | 0,23182333 | up | 1,3095131 | 0,3890305 | MLLT4 | Homo sapiens myeloid/lymphoid or mixed-lineage leukemia (trithorax homolog, Drosophila); translocated to, 4 (MLLT4), transcript variant 1, mRNA [NM_001207008] |
| A_33_P3249305 | 0,8683143 | down | -1,0252254 | -0,0359411 | CHRD | Homo sapiens chordin (CHRD), mRNA [NM_003741] |
| A_33_P3328609 | 0,0281057 | down | -1,534493 | -0,617762 | PKP4 | Homo sapiens plakophilin 4 (PKP4), transcript variant 1, mRNA [NM_003628] |
| A_23_P156708 | 0,03458871 | up | 1,7020804 | 0,7672992 | TNXB | Homo sapiens tenascin XB (TNXB), transcript variant XB-S, mRNA [NM_032470] |
| A_33_P3330264 | 0,5001052 | down | -1,7203431 | -0,7826963 | CXCL1 | Homo sapiens chemokine (C-X-C motif) ligand 1 (melanoma growth stimulating activity, alpha) (CXCL1), transcript variant 1, mRNA [NM_001511] |
| A_24_P37589 | 0,88032365 | up | 1,0483704 | 0,06814847 | ACPP | Homo sapiens acid phosphatase, prostate (ACPP), transcript variant 1, mRNA [NM_001099] |
| A_24_P382119 | 0,34735253 | up | 1,1258692 | 0,17103916 | MTMR4 | Homo sapiens myotubularin related protein 4 (MTMR4), mRNA [NM_004687] |
| A_33_P3385516 | 0,72256786 | down | -1,0659292 | -0,0921116 | CBLN4 | Homo sapiens cerebellin 4 precursor (CBLN4), mRNA [NM_080617] |
| A_24_P295010 | 0,592242 | up | 1,3589418 | 0,44248366 | SERPINB9 | Homo sapiens serpin peptidase inhibitor, clade B (ovalbumin), member 9 (SERPINB9), mRNA [NM_004155] |
| A_24_P824592 | 0,45077533 | down | -1,158825 | -0,2126628 | RBMX | Homo sapiens RNA binding motif protein, X-linked (RBMX), transcript variant 1, mRNA [NM_002139] |
| A_23_P20022 | 0,68035406 | up | 1,0764817 | 0,10632379 | HILPDA | Homo sapiens hypoxia inducible lipid droplet-associated (HILPDA), transcript variant 1, mRNA [NM_013332] |
| A_33_P3283619 | 0,5633528 | up | 1,2282429 | 0,29659587 | SH2D1A | Homo sapiens SH2 domain containing 1A (SH2D1A), transcript variant 2, mRNA [NM_001114937] |
| A_23_P78742 | 0,03632669 | up | 1,3962021 | 0,48150778 | FLT3LG | Homo sapiens fms-related tyrosine kinase 3 ligand (FLT3LG), transcript variant 3, mRNA [NM_001459] |
| A_23_P16992 | 0,27218512 | down | -1,1462282 | -0,1968943 | PKP4 | Homo sapiens plakophilin 4 (PKP4), transcript variant 1, mRNA [NM_003628] |
| A_24_P382187 | 0,8715293 | down | -1,037345 | -0,0528959 | IGFBP4 | Homo sapiens insulin-like growth factor binding protein 4 (IGFBP4), mRNA [NM_001552] |
| A_33_P3317211 | 0,42748109 | up | 1,1486459 | 0,1999341 | MECP2 | Homo sapiens methyl CpG binding protein 2 (MECP2), transcript variant 2, mRNA [NM_001110792] |
| A_24_P216253 | 0,20817779 | up | 1,1955162 | 0,25763372 | DLGAP4 | Homo sapiens discs, large (Drosophila) homolog-associated protein 4 (DLGAP4), transcript variant 1, mRNA [NM_014902] |
| A_24_P402438 | 0,15223794 | up | 1,8070904 | 0,8536687 | TGFB2 | Homo sapiens transforming growth factor, beta 2 (TGFB2), transcript variant 2, mRNA [NM_003238] |
| A_24_P261417 | 0,10695662 | down | -3,4777765 | -1,7981652 | DKK3 | Homo sapiens dickkopf WNT signaling pathway inhibitor 3 (DKK3), transcript variant 1, mRNA [NM_015881] |
| A_23_P434809 | 0,12560782 | up | 3,9744287 | 1,9907475 | S100A8 | Homo sapiens S100 calcium binding protein A8 (S100A8), mRNA [NM_002964] |
| A_24_P208825 | 0,72921705 | up | 1,0659962 | 0,09220225 | MUC4 | Homo sapiens mucin 4, cell surface associated (MUC4), transcript variant 1, mRNA [NM_018406] |
| A_33_P3234124 | 0,0992045 | up | 1,5054307 | 0,5901763 | FAM132B | Homo sapiens family with sequence similarity 132, member B (FAM132B), mRNA [NM_001291832] |
| A_23_P76102 | 0,4631011 | up | 1,1899055 | 0,25084704 | GDF11 | Homo sapiens growth differentiation factor 11 (GDF11), mRNA [NM_005811] |
| A_24_P353619 | 0,34392866 | down | -1,6542075 | -0,7261402 | ALPL | Homo sapiens alkaline phosphatase, liver/bone/kidney (ALPL), transcript variant 1, mRNA [NM_000478] |
| A_23_P366376 | 6,56E-04 | down | -3,2553055 | -1,7027929 | TDGF1 | Homo sapiens teratocarcinoma-derived growth factor 1 (TDGF1), transcript variant 1, mRNA [NM_003212] |
| A_33_P3347869 | 0,6173055 | down | -1,2893128 | -0,3666024 | C3 | Homo sapiens complement component 3 (C3), mRNA [NM_000064] |
| A_23_P132718 | 0,11257168 | up | 1,4553907 | 0,54140645 | SEMA3B | Homo sapiens sema domain, immunoglobulin domain (Ig), short basic domain, secreted, (semaphorin) 3B (SEMA3B), transcript variant 1, mRNA [NM_004636] |
| A_23_P47885 | 0,6121846 | down | -1,1259966 | -0,1712025 | LRIG3 | Homo sapiens leucine-rich repeats and immunoglobulin-like domains 3 (LRIG3), transcript variant 2, mRNA [NM_153377] |
| A_33_P3214586 | 0,4880669 | up | 1,1726326 | 0,22975104 | ARTN | Homo sapiens artemin (ARTN), transcript variant 4, mRNA [NM_057090] |
| A_23_P154605 | 0,19342607 | down | -1,262889 | -0,3367279 | SULF2 | Homo sapiens sulfatase 2 (SULF2), transcript variant 1, mRNA [NM_018837] |
| A_23_P409438 | 0,0030961 | up | 1,7097596 | 0,77379346 | IFNL2 | Homo sapiens interferon, lambda 2 (IFNL2), mRNA [NM_172138] |
| A_33_P3296181 | 0,6788981 | up | 1,0884646 | 0,12229451 | CCL3L3 | Homo sapiens chemokine (C-C motif) ligand 3-like 3 (CCL3L3), mRNA [NM_001001437] |
| A_23_P202501 | 0,13422044 | down | -1,2272615 | -0,2954427 | RNLS | Homo sapiens renalase, FAD-dependent amine oxidase (RNLS), transcript variant 1, mRNA [NM_001031709] |
| A_33_P3405848 | 0,10257061 | up | 1,6825466 | 0,7506465 | TPO | Homo sapiens thyroid peroxidase (TPO), transcript variant 2, mRNA [NM_175719] |
| A_33_P3364571 | 0,3874441 | up | 1,1433744 | 0,19329795 | TNXB | Homo sapiens tenascin XB (TNXB), transcript variant XB, mRNA [NM_019105] |
| A_23_P126593 | 0,77348995 | up | 1,060691 | 0,08500443 | S100A11 | Homo sapiens S100 calcium binding protein A11 (S100A11), mRNA [NM_005620] |
| A_23_P88865 | 0,5083947 | up | 1,1396811 | 0,1886302 | CMTM3 | Homo sapiens CKLF-like MARVEL transmembrane domain containing 3 (CMTM3), transcript variant 1, mRNA [NM_144601] |
| A_23_P212830 | 0,6916855 | down | -1,1325382 | -0,1795597 | FGFR3 | Homo sapiens fibroblast growth factor receptor 3 (FGFR3), transcript variant 1, mRNA [NM_000142] |
| A_23_P35820 | 0,64608276 | up | 1,0868905 | 0,12020654 | CFL1 | Homo sapiens cofilin 1 (non-muscle) (CFL1), mRNA [NM_005507] |
| A_23_P2920 | 0,523338 | down | -1,3408113 | -0,4231062 | SERPINA3 | Homo sapiens serpin peptidase inhibitor, clade A (alpha-1 antiproteinase, antitrypsin), member 3 (SERPINA3), mRNA [NM_001085] |
| A_24_P87931 | 0,01730811 | up | 1,6006074 | 0,67861944 | APOL1 | Homo sapiens apolipoprotein L, 1 (APOL1), transcript variant 2, mRNA [NM_145343] |
| A_23_P436353 | 0,6597328 | down | -1,0884053 | -0,1222158 | MLLT4 | Homo sapiens myeloid/lymphoid or mixed-lineage leukemia (trithorax homolog, Drosophila); translocated to, 4 (MLLT4), transcript variant 4, mRNA [NM_001291964] |
| A_24_P92472 | 0,09320168 | up | 2,016364 | 1,0117562 | CFI | Homo sapiens complement factor I (CFI), mRNA [NM_000204] |
| A_33_P3316273 | 0,45676517 | up | 1,3526089 | 0,43574476 | CCL3 | Homo sapiens chemokine (C-C motif) ligand 3 (CCL3), mRNA [NM_002983] |
| A_23_P256158 | 0,16100192 | down | -1,6032919 | -0,6810371 | ADRA2C | Homo sapiens adrenoceptor alpha 2C (ADRA2C), mRNA [NM_000683] |
| A_23_P152620 | 0,8481366 | down | -1,049333 | -0,0694726 | TNFSF13 | Homo sapiens tumor necrosis factor (ligand) superfamily, member 13 (TNFSF13), transcript variant gamma, mRNA [NM_172088] |
| A_33_P3418833 | 0,00935525 | down | -3,0313272 | -1,5999496 | FLRT3 | Homo sapiens fibronectin leucine rich transmembrane protein 3 (FLRT3), transcript variant 2, mRNA [NM_198391] |
| A_23_P132826 | 0,33447596 | down | -2,3027737 | -1,2033726 | SERPINI2 | Homo sapiens serpin peptidase inhibitor, clade I (pancpin), member 2 (SERPINI2), transcript variant 2, mRNA [NM_006217] |
| A_23_P89665 | 0,16465548 | up | 1,7423437 | 0,8010292 | KRT33B | Homo sapiens keratin 33B, type I (KRT33B), mRNA [NM_002279] |
| A_19_P00319514 | 0,287372 | up | 1,9946884 | 0,99616337 | DPYSL3 | dihydropyrimidinase-like 3 [Source:HGNC Symbol;Acc:HGNC:3015] [ENST00000504965] |
| A_23_P331928 | 0,28745237 | up | 1,9629182 | 0,973 | CD109 | Homo sapiens CD109 molecule (CD109), transcript variant 1, mRNA [NM_133493] |
| A_24_P122746 | 0,1763144 | up | 1,5682644 | 0,6491688 | VWA1 | Homo sapiens von Willebrand factor A domain containing 1 (VWA1), transcript variant 1, mRNA [NM_022834] |
| A_33_P3407034 | 0,774892 | up | 1,1243284 | 0,16906345 | KIT | Homo sapiens v-kit Hardy-Zuckerman 4 feline sarcoma viral oncogene homolog (KIT), transcript variant 1, mRNA [NM_000222] |
| A_33_P3233906 | 0,30923423 | down | -2,1641803 | -1,1138207 | RAMP1 | Homo sapiens receptor (G protein-coupled) activity modifying protein 1 (RAMP1), mRNA [NM_005855] |
| A_23_P87072 | 0,00803863 | up | 2,0300424 | 1,0215099 | PANX3 | Homo sapiens pannexin 3 (PANX3), mRNA [NM_052959] |

| | | | | | | |
|---|---|---|---|---|---|---|
| A_23_P214544 | 0,09293532 | up | 1,2453116 | 0,3165068 | GPX5 | Homo sapiens glutathione peroxidase 5 (GPX5), transcript variant 1, mRNA [NM_001509] |
| A_33_P3257297 | 0,02126138 | down | -5,5733166 | -2,4785361 | CFAP58 | Homo sapiens cilia and flagella associated protein 58 (CFAP58), mRNA [NM_001008723] |
| A_24_P97526 | 0,49416405 | down | -1,1108797 | -0,1517025 | CMTM6 | Homo sapiens CKLF-like MARVEL transmembrane domain containing 6 (CMTM6), mRNA [NM_017801] |
| A_33_P3313110 | 0,33713353 | up | 1,5872755 | 0,66655254 | MUC16 | Homo sapiens mucin 16, cell surface associated (MUC16), mRNA [NM_024690] |
| A_23_P148297 | 0,7011049 | down | -1,0875611 | -0,1210965 | SH3BGRL | Homo sapiens SH3 domain binding glutamate-rich protein like (SH3BGRL), mRNA [NM_003022] |
| A_33_P3355266 | 0,32370967 | up | 1,2851342 | 0,36191902 | TINAGL1 | Homo sapiens tubulointerstitial nephritis antigen-like 1 (TINAGL1), transcript variant 1, mRNA [NM_022164] |
| A_23_P141362 | 0,19826533 | up | 1,2336552 | 0,30293924 | FZD2 | Homo sapiens frizzled class receptor 2 (FZD2), mRNA [NM_001466] |
| A_23_P379550 | 0,47629362 | down | -1,1601946 | -0,2143669 | YARS | Homo sapiens tyrosyl-tRNA synthetase (YARS), mRNA [NM_003680] |
| A_33_P3399788 | 0,4695204 | down | -1,3531213 | -0,4362912 | SERPINA3 | Homo sapiens serpin peptidase inhibitor, clade A (alpha-1 antiproteinase, antitrypsin), member 3 (SERPINA3), mRNA [NM_001085] |
| A_33_P3232557 | 0,8203894 | up | 1,0568886 | 0,07982329 | DLGAP3 | Homo sapiens discs, large (Drosophila) homolog-associated protein 3 (DLGAP3), mRNA [NM_001080418] |
| A_23_P168288 | 0,14536443 | up | 1,341982 | 0,4243653 | IL22RA2 | Homo sapiens interleukin 22 receptor, alpha 2 (IL22RA2), transcript variant 1, mRNA [NM_052962] |
| A_23_P215296 | 0,43700308 | up | 1,0875741 | 0,12111373 | CDK13 | Homo sapiens cyclin-dependent kinase 13 (CDK13), transcript variant 1, mRNA [NM_003718] |
| A_33_P3288110 | 0,7896866 | down | -1,0378027 | -0,0535322 | PKP4 | Homo sapiens plakophilin 4 (PKP4), transcript variant 2, mRNA [NM_001005476] |
| A_23_P145841 | 0,09685496 | down | -1,387531 | -0,4725201 | SOSTDC1 | Homo sapiens sclerostin domain containing 1 (SOSTDC1), mRNA [NM_015464] |
| A_33_P3235706 | 0,48086086 | down | -1,1763501 | -0,2343175 | ZCCHC11 | Homo sapiens zinc finger, CCHC domain containing 11, mRNA (cDNA clone IMAGE:5505348), with apparent retained intron. [BC048301] |
| A_23_P109269 | 0,4888678 | up | 1,1801448 | 0,23896387 | LAMA5 | Homo sapiens laminin, alpha 5 (LAMA5), mRNA [NM_005560] |
| A_33_P3844650 | 0,8839788 | down | -1,0930297 | -0,1283326 | ANGPTL2 | Homo sapiens angiopoietin-like 2 (ANGPTL2), mRNA [NM_012098] |
| A_33_P3384442 | 0,38080305 | up | 1,1844143 | 0,24417377 | LAMA5 | laminin, alpha 5 [Source:HGNC Symbol;Acc:HGNC:6485] [ENST00000370677] |
| A_33_P3268472 | 0,05866381 | up | 1,4063317 | 0,49193686 | CTSC | Homo sapiens cathepsin C (CTSC), transcript variant 3, mRNA [NM_001114173] |
| A_23_P118203 | 0,8209614 | down | -1,1161977 | -0,1585926 | ZG16B | Homo sapiens zymogen granule protein 16B (ZG16B), mRNA [NM_145252] |
| A_33_P3262635 | 0,37305042 | down | -1,2944251 | -0,3723115 | CECR1 | Homo sapiens cat eye syndrome chromosome region, candidate 1 (CECR1), transcript variant 3, mRNA [NM_001282225] |
| A_23_P79931 | 0,89576656 | up | 1,0262312 | 0,03735575 | ATRN | Homo sapiens attractin (ATRN), transcript variant 2, mRNA [NM_139322] |
| A_33_P3412900 | 0,40820563 | up | 1,1646096 | 0,21984635 | CBLN3 | Homo sapiens cerebellin 3 precursor (CBLN3), mRNA [NM_001039771] |
| A_32_P70315 | 0,07823484 | down | -3,218522 | -1,6863984 | TIMP4 | Homo sapiens TIMP metallopeptidase inhibitor 4 (TIMP4), mRNA [NM_003256] |
| A_33_P3352253 | 0,1772861 | up | 1,2862082 | 0,36312413 | MTUS1 | Homo sapiens microtubule associated tumor suppressor 1 (MTUS1), transcript variant 2, mRNA [NM_001001925] |
| A_23_P256735 | 0,67954034 | down | -1,106944 | -0,1465822 | CPQ | Homo sapiens carboxypeptidase Q (CPQ), mRNA [NM_016134] |
| A_33_P3470781 | 0,24342692 | down | -1,1759146 | -0,2337833 | RALGAPA2 | Homo sapiens Ral GTPase activating protein, alpha subunit 2 (catalytic) (RALGAPA2), mRNA [NM_020343] |
| A_23_P119535 | 0,08461414 | up | 1,3789403 | 0,46356004 | EFNA2 | Homo sapiens ephrin-A2 (EFNA2), mRNA [NM_001405] |
| A_33_P3346635 | 5,89E-05 | up | 2,521466 | 1,3342627 | FAM20C | Homo sapiens cDNA FLJ43291 fis, clone MESAN2015515. [AK125281] |
| A_24_P49199 | 0,08539928 | down | -2,4486847 | -1,292007 | GLDN | Homo sapiens gliomedin (GLDN), mRNA [NM_181789] |
| A_33_P3408203 | 0,48270914 | up | 1,1106793 | 0,15144227 | TGFA | Homo sapiens transforming growth factor, alpha (TGFA), transcript variant 1, mRNA [NM_003236] |
| A_33_P3413483 | 0,7533235 | down | -1,1830748 | -0,2425413 | SORD | sorbitol dehydrogenase [Source:HGNC Symbol;Acc:HGNC:11184] [ENST00000267814] |
| A_23_P371682 | 0,13423292 | up | 1,6137241 | 0,6903939 | GPC6 | Homo sapiens glypican 6 (GPC6), mRNA [NM_005708] |
| A_33_P3413987 | 0,6251195 | up | 1,1221848 | 0,16631022 | SERPING1 | Homo sapiens serpin peptidase inhibitor, clade G (C1 inhibitor), member 1 (SERPING1), transcript variant 1, mRNA [NM_000062] |
| A_23_P107313 | 0,8599073 | up | 1,0307863 | 0,04374524 | SDF2 | Homo sapiens stromal cell-derived factor 2 (SDF2), transcript variant 1, mRNA [NM_006923] |
| A_23_P71480 | 0,5500513 | up | 1,3798883 | 0,46455148 | DEFB1 | Homo sapiens defensin, beta 1 (DEFB1), mRNA [NM_005218] |
| A_23_P334727 | 0,7095305 | up | 1,1585739 | 0,21235003 | NRG4 | Homo sapiens neuregulin 4 (NRG4), mRNA [NM_138573] |
| A_24_P202558 | 0,3543259 | up | 1,1337147 | 0,1810576 | SIPA1L3 | Homo sapiens signal-induced proliferation-associated 1 like 3 (SIPA1L3), mRNA [NM_015073] |
| A_23_P111041 | 0,82251364 | down | -1,0622084 | -0,0870669 | HIST1H2BI | Homo sapiens histone cluster 1, H2bi (HIST1H2BI), mRNA [NM_003525] |
| A_23_P259955 | 0,8985509 | up | 1,0182953 | 0,02615598 | GDF5 | Homo sapiens growth differentiation factor 5 (GDF5), mRNA [NM_000557] |
| A_33_P3231367 | 0,08303475 | down | -1,3863443 | -0,4712856 | ATXN10 | Homo sapiens ataxin 10 (ATXN10), transcript variant 1, mRNA [NM_013236] |
| A_33_P3332215 | 0,7366717 | up | 1,1015995 | 0,13959976 | MUC1 | Homo sapiens mucin 1, cell surface associated (MUC1), transcript variant 7, mRNA [NM_001044392] |
| A_23_P67198 | 0,36909485 | up | 1,339849 | 0,4220704 | CPAMD8 | Homo sapiens C3 and PZP-like, alpha-2-macroglobulin domain containing 8 (CPAMD8), mRNA [NM_015692] |
| A_33_P3260014 | 0,70250493 | up | 1,1374224 | 0,18576817 | TPO | Homo sapiens thyroid peroxidase (TPO), transcript variant 4, mRNA [NM_175721] |
| A_33_P3372666 | 0,79460794 | down | -1,0862074 | -0,1192996 | PDGFA | Homo sapiens platelet-derived growth factor alpha polypeptide (PDGFA), transcript variant 2, mRNA [NM_033023] |
| A_33_P3399263 | 0,61495423 | down | -1,0815102 | -0,1130473 | IL15RA | Homo sapiens interleukin 15 receptor, alpha (IL15RA), transcript variant 3, mRNA [NM_001243539] |
| A_23_P215491 | 0,5552272 | up | 1,101786 | 0,13984406 | CCL24 | Homo sapiens chemokine (C-C motif) ligand 24 (CCL24), mRNA [NM_002991] |
| A_33_P3354646 | 0,05277462 | up | 1,5626273 | 0,6439737 | PNLIPRP1 | Homo sapiens pancreatic lipase-related protein 1 (PNLIPRP1), transcript variant 1, mRNA [NM_006229] |
| A_24_P59220 | 0,31700823 | up | 1,185945 | 0,24603714 | POTEF | Homo sapiens POTE ankyrin domain family, member F (POTEF), mRNA [NM_001099771] |
| A_33_P3416668 | 0,08898424 | up | 1,3770194 | 0,4615489 | VWA1 | Homo sapiens von Willebrand factor A domain containing 1 (VWA1), transcript variant 1, mRNA [NM_022834] |
| A_24_P276628 | 0,38646266 | up | 1,1855582 | 0,24556649 | PPT1 | Homo sapiens palmitoyl-protein thioesterase 1 (PPT1), transcript variant 1, mRNA [NM_000310] |
| A_24_P309317 | 0,54361063 | down | -1,1473416 | -0,198295 | PSAP | Homo sapiens prosaposin (PSAP), transcript variant 2, mRNA [NM_001042465] |
| A_23_P501754 | 0,23369212 | up | 2,7637632 | 1,466634 | CSF3 | Homo sapiens colony stimulating factor 3 (granulocyte) (CSF3), transcript variant 1, mRNA [NM_000759] |
| A_23_P253958 | 0,18079954 | down | -2,0769432 | -1,0544617 | LRRC17 | Homo sapiens leucine rich repeat containing 17 (LRRC17), transcript variant 2, mRNA [NM_005824] |
| A_33_P3230990 | 0,10292421 | up | 1,2566723 | 0,32960844 | SCUBE1 | Homo sapiens signal peptide, CUB domain, EGF-like 1 (SCUBE1), mRNA [NM_173050] |

| | | | | | | |
|---|---|---|---|---|---|---|
| A_33_P3351249 | 0,84909374 | up | 1,0329068 | 0,04671004 | CXCL16 | Homo sapiens chemokine (C-X-C motif) ligand 16 (CXCL16), transcript variant 2, mRNA [NM_001100812] |
| A_33_P3272580 | 0,55729693 | down | -1,0879152 | -0,1215661 | FUCA2 | Homo sapiens fucosidase, alpha-L- 2, plasma (FUCA2), mRNA [NM_032020] |
| A_23_P301476 | 0,87063956 | up | 1,0214499 | 0,03061848 | C3orf33 | Homo sapiens chromosome 3 open reading frame 33 (C3orf33), mRNA [NM_173657] |
| A_23_P380857 | 0,07673352 | up | 1,419726 | 0,50561255 | APOL4 | Homo sapiens apolipoprotein L, 4 (APOL4), transcript variant a, mRNA [NM_030643] |
| A_33_P3339100 | 0,11010718 | down | -1,7666724 | -0,8210345 | SELP | Homo sapiens selectin P (granule membrane protein 140kDa, antigen CD62) (SELP), mRNA [NM_003005] |
| A_23_P392962 | 0,40488476 | up | 1,2885958 | 0,36579978 | GRAP | Homo sapiens GRB2-related adaptor protein (GRAP), mRNA [NM_006613] |
| A_33_P3402091 | 0,89560777 | down | -1,0452079 | -0,0637899 | MERTK | Homo sapiens MER proto-oncogene, tyrosine kinase (MERTK), mRNA [NM_006343] |
| A_24_P6903 | 0,114137 | up | 1,2622811 | 0,33603317 | ACTBL2 | Homo sapiens actin, beta-like 2 (ACTBL2), mRNA [NM_001017992] |
| A_33_P3231653 | 0,5569534 | down | -1,1460892 | -0,1967193 | GOLM1 | Homo sapiens golgi membrane protein 1 (GOLM1), transcript variant 1, mRNA [NM_016548] |
| A_33_P3303245 | 0,7850331 | up | 1,1347094 | 0,18232282 | KIT | Homo sapiens v-kit Hardy-Zuckerman 4 feline sarcoma viral oncogene homolog (KIT), transcript variant 2, mRNA [NM_001093772] |
| A_33_P3372840 | 0,425817 | up | 1,2380005 | 0,30801192 | CXCL12 | Homo sapiens chemokine (C-X-C motif) ligand 12 (CXCL12), transcript variant 3, mRNA [NM_001033886] |
| A_23_P29953 | 0,04873305 | up | 1,4007077 | 0,48615593 | IL15 | Homo sapiens interleukin 15 (IL15), transcript variant 2, mRNA [NM_172175] |
| A_33_P3329622 | 0,3221626 | down | -1,2075226 | -0,2720502 | MROH7 | maestro heat-like repeat family member 7 [Source:HGNC Symbol;Acc:HGNC:24802] [ENST00000395690] |
| A_23_P377291 | 0,3359528 | up | 1,198624 | 0,26137918 | TGFA | Homo sapiens transforming growth factor, alpha (TGFA), transcript variant 1, mRNA [NM_003236] |
| A_33_P3311145 | 0,70167017 | down | -1,1028802 | -0,1412761 | CEP164 | Homo sapiens centrosomal protein 164kDa (CEP164), transcript variant 2, mRNA [NM_001271933] |
| A_24_P371399 | 0,8563764 | down | -1,0319916 | -0,0454312 | C3orf58 | Homo sapiens chromosome 3 open reading frame 58 (C3orf58), transcript variant 1, mRNA [NM_173552] |
| A_23_P3956 | 0,18699248 | up | 1,3430065 | 0,42546627 | C1QTNF1 | Homo sapiens C1q and tumor necrosis factor related protein 1 (C1QTNF1), transcript variant 4, mRNA [NM_198594] |
| A_23_P34888 | 0,17193958 | up | 1,357653 | 0,4411148 | CHIA | Homo sapiens chitinase, acidic (CHIA), transcript variant 2, mRNA [NM_021797] |
| A_23_P28318 | 0,67128927 | down | -1,0470307 | -0,0663037 | NDUFAF7 | Homo sapiens NADH dehydrogenase (ubiquinone) complex I, assembly factor 7 (NDUFAF7), transcript variant 1, mRNA [NM_144736] |
| A_33_P3306163 | 0,17283784 | up | 1,3173583 | 0,39764774 | LGALS3 | Homo sapiens lectin, galactoside-binding, soluble, 3 (LGALS3), transcript variant 3, mRNA [NM_001177388] |
| A_23_P145485 | 0,37670273 | up | 1,5307728 | 0,6142602 | ULBP2 | Homo sapiens UL16 binding protein 2 (ULBP2), mRNA [NM_025217] |
| A_33_P3276615 | 0,634161 | up | 1,1738092 | 0,23119788 | APOL4 | Homo sapiens apolipoprotein L, 4 (APOL4), transcript variant a, mRNA [NM_030643] |
| A_24_P339416 | 0,28651828 | down | -1,1729959 | -0,230198 | ARSG | Homo sapiens arylsulfatase G (ARSG), transcript variant 1, mRNA [NM_014960] |
| A_33_P3233105 | 0,0955682 | up | 1,5760201 | 0,65628594 | MLLT4 | Homo sapiens myeloid/lymphoid or mixed-lineage leukemia (trithorax homolog, Drosophila); translocated to, 4 (MLLT4), transcript variant 1, mRNA [NM_001207008] |
| A_33_P3317058 | 0,4905636 | down | -1,2248179 | -0,2925672 | STAG3 | Homo sapiens stromal antigen 3 (STAG3), transcript variant 3, mRNA [NM_001282717] |
| A_32_P15320 | 0,17994308 | down | -1,2846336 | -0,361357 | EEF1A1 | Homo sapiens eukaryotic translation elongation factor 1 alpha 1 (EEF1A1), mRNA [NM_001402] |
| A_23_P40611 | 0,667017 | down | -1,1626369 | -0,2174006 | TCN2 | Homo sapiens transcobalamin II (TCN2), transcript variant 1, mRNA [NM_000355] |
| A_24_P101114 | 0,48197684 | up | 1,104606 | 0,1435319 | CNOT1 | Homo sapiens CCR4-NOT transcription complex, subunit 1 (CNOT1), transcript variant 2, mRNA [NM_206999] |
| A_24_P335305 | 0,5190497 | up | 1,19342 | 0,25510192 | OAS3 | Homo sapiens 2'-5'-oligoadenylate synthetase 3, 100kDa (OAS3), mRNA [NM_006187] |
| A_33_P3407606 | 0,19080658 | up | 1,3917339 | 0,47688338 | MSN | moesin [Source:HGNC Symbol;Acc:HGNC:7373] [ENST00000447323] |
| A_23_P131588 | 0,36304945 | up | 1,1663558 | 0,222008 | BMP10 | Homo sapiens bone morphogenetic protein 10 (BMP10), mRNA [NM_014482] |
| A_23_P315964 | 0,54541415 | down | -1,2056438 | -0,2698037 | UMODL1 | Homo sapiens uromodulin-like 1 (UMODL1), transcript variant 2, mRNA [NM_173568] |
| A_33_P3392787 | 0,04803197 | down | -4,24121 | -2,084476 | CFAP58 | Homo sapiens cilia and flagella associated protein 58 (CFAP58), mRNA [NM_001008723] |
| A_23_P207125 | 0,43707907 | up | 1,2382698 | 0,30832568 | NLGN2 | Homo sapiens neuroligin 2 (NLGN2), mRNA [NM_020795] |
| A_24_P90881 | 0,45655242 | down | -1,1355046 | -0,1833336 | CES3 | Homo sapiens carboxylesterase 3 (CES3), transcript variant 1, mRNA [NM_024922] |
| A_33_P3280471 | 0,13943313 | up | 1,4299548 | 0,5159695 | ERBB3 | Homo sapiens v-erb-b2 avian erythroblastic leukemia viral oncogene homolog 3 (ERBB3), transcript variant 1, mRNA [NM_001982] |
| A_23_P302060 | 0,7240567 | up | 1,1404339 | 0,18958284 | IFNE | Homo sapiens interferon, epsilon (IFNE), mRNA [NM_176891] |
| A_33_P3259393 | 0,54344565 | up | 1,2966665 | 0,37480748 | HAPLN3 | Homo sapiens hyaluronan and proteoglycan link protein 3 (HAPLN3), mRNA [NM_178232] |
| A_33_P3389872 | 0,05323265 | up | 1,5117273 | 0,59619796 | LRRK2 | Homo sapiens cDNA FLJ45829 fis, clone NT2RP8006452. [AK127729] |
| A_33_P3252286 | 0,22820003 | down | -1,814023 | -0,8591927 | CRLF1 | Homo sapiens cytokine receptor-like factor 1 (CRLF1), mRNA [NM_004750] |
| A_33_P3349927 | 0,8739408 | up | 1,040991 | 0,05795752 | WDR60 | WD repeat domain 60 [Source:HGNC Symbol;Acc:HGNC:21862] [ENST00000467220] |
| A_23_P146644 | 0,80569434 | down | -1,0401087 | -0,0567343 | ANXA2 | Homo sapiens annexin A2 (ANXA2), transcript variant 2, mRNA [NM_001002857] |
| A_33_P3365193 | 0,7900772 | up | 1,0856045 | 0,11849867 | AMY1C | Homo sapiens amylase, alpha 1C (salivary) (AMY1C), mRNA [NM_001008219] |
| A_23_P72117 | 0,08431177 | down | -1,461839 | -0,5477844 | SMPDL3A | Homo sapiens sphingomyelin phosphodiesterase, acid-like 3A (SMPDL3A), transcript variant 1, mRNA [NM_006714] |
| A_33_P3287646 | 0,12882863 | up | 1,4261743 | 0,5121503 | HSPB1 | Homo sapiens heat shock 27kDa protein 1 (HSPB1), mRNA [NM_001540] |
| A_33_P3259708 | 0,8538798 | up | 1,0508524 | 0,07156007 | CMA1 | Homo sapiens chymase 1, mast cell (CMA1), mRNA [NM_001836] |
| A_32_P80850 | 0,00508212 | up | 2,6538956 | 1,4081116 | COL14A1 | Homo sapiens collagen, type XIV, alpha 1 (COL14A1), mRNA [NM_021110] |
| A_33_P3364869 | 0,20327616 | up | 1,5212923 | 0,6052974 | NAMPT | Homo sapiens cDNA FLJ13279 fis, clone OVARC1001055, moderately similar to PRE-B CELL ENHANCING FACTOR PRECURSOR. [AK023341] |
| A_33_P3366064 | 0,8924859 | up | 1,0396273 | 0,05606644 | SOGA1 | Homo sapiens suppressor of glucose, autophagy associated 1 (SOGA1), transcript variant 2, mRNA [NM_199181] |
| A_24_P377124 | 0,48634213 | up | 1,2029711 | 0,26660198 | THPO | Homo sapiens thrombopoietin (THPO), transcript variant 1, mRNA [NM_000460] |
| A_33_P3317442 | 0,6492799 | up | 1,1279675 | 0,17372547 | MCF2L | Homo sapiens MCF.2 cell line derived transforming sequence-like (MCF2L), transcript variant 2, mRNA [NM_024979] |
| A_33_P3334398 | 0,6190877 | up | 1,1044418 | 0,14331734 | CA6 | Homo sapiens carbonic anhydrase VI (CA6), transcript variant 2, mRNA [NM_001270500] |
