## Supplementary material for "Early Reduction of SARS-CoV-2 Replication in Bronchial Epithelium by Kinin B_2_ Receptor Antagonism": Supl. Table 2

**Supplemental Table S2.** DEGs from transcriptome analysis filtered on kallikreins from nasal scrapings from SARS-CoV-2 acute positive patients compared to negative patients

| ProbeName | p ([pos acute] | Regulation ([ | FC (abs) ([po: | FC ([pos acut | Log FC ([pos | GeneSymbol | Description |
|---|---|---|---|---|---|---|---|
| A_33_P3215 | 0,07512057 | up | 1,4754667 | 1,4754667 | 0,56117135 | KLK3 | kallikrein-related peptidase 3 [Source:HGNC Symbol;Acc:HGNC:6364] [ENST00000595151] |
| A_23_P3102 | 1 | up | 1,1165178 | 1,1165178 | 0,15900622 | KLK2 | Homo sapiens kallikrein-related peptidase 2 (KLK2), transcript variant 2, mRNA [NM_001002231] |
| A_24_P5750 | 0,18051589 | up | 1,4157342 | 1,4157342 | 0,5015504 | KLK2 | Homo sapiens kallikrein-related peptidase 2 (KLK2), transcript variant 1, mRNA [NM_005551] |
| A_33_P3251 | 1 | up | 1,2278229 | 1,2278229 | 0,29610246 | KLK13 | kallikrein-related peptidase 13 [Source:HGNC Symbol;Acc:HGNC:6361] [ENST00000602090] |
| A_23_P1015 | 0,16285726 | up | 1,7061496 | 1,7061496 | 0,77074414 | KLK11 | Homo sapiens kallikrein-related peptidase 11 (KLK11), transcript variant 2, mRNA [NM_144947] |
| A_23_P1625 | 1 | up | 1,2936851 | 1,2936851 | 0,37148646 | KLK1 | Homo sapiens kallikrein 1 (KLK1), mRNA [NM_002257] |
| A_24_P3760 | 1 | up | 1,3677056 | 1,3677056 | 0,4517577 | KLK4 | Homo sapiens kallikrein-related peptidase 4 (KLK4), transcript variant 1, mRNA [NM_004917] |
| A_33_P3417 | 1 | up | 5,2178 | 5,2178 | 2,3834417 | KLK14 | Homo sapiens kallikrein-related peptidase 14 (KLK14), mRNA [NM_022046] |
| A_23_P3905 | 1 | up | 14,396585 | 14,396585 | 3,8476548 | KLK7 | Homo sapiens kallikrein-related peptidase 7 (KLK7), transcript variant 1, mRNA [NM_005046] |
| A_23_P3693 | 1 | up | 14,018521 | 14,018521 | 3,8092623 | KLK8 | Homo sapiens kallikrein-related peptidase 8 (KLK8), transcript variant 2, mRNA [NM_144505] |
| A_23_P5000 | 0,01925241 | up | 15,19849 | 15,19849 | 3,925856 | KLK12 | Homo sapiens kallikrein-related peptidase 12 (KLK12), transcript variant 2, mRNA [NM_145894] |
| A_33_P3398 | 0,00995833 | up | 8,533642 | 8,533642 | 3,0931616 | KLK9 | Homo sapiens kallikrein-related peptidase 9 (KLK9), mRNA [NM_012315] |
| A_23_P1534 | 0,00814065 | up | 11,039466 | 11,039466 | 3,4645984 | KLK5 | Homo sapiens kallikrein-related peptidase 5 (KLK5), transcript variant 1, mRNA [NM_012427] |
| A_33_P3235 | 0,06648849 | up | 11,334522 | 11,334522 | 3,5026517 | KLK6 | Homo sapiens kallikrein-related peptidase 6 (KLK6), transcript variant B, mRNA [NM_001012964] |
| A_24_P2369 | 1 | up | 4,5679436 | 4,5679436 | 2,1915448 | KLK6 | Homo sapiens kallikrein-related peptidase 6 (KLK6), transcript variant B, mRNA [NM_001012964] |
| A_33_P3417 | 0,16268907 | up | 3,0919542 | 3,0919542 | 1,6285189 | KLK10 | Homo sapiens kallikrein-related peptidase 10 (KLK10), transcript variant 1, mRNA [NM_002776] |
| A_24_P4166 | 0,07144744 | up | 6,230724 | 6,230724 | 2,6393998 | KLK13 | Homo sapiens kallikrein-related peptidase 13 (KLK13), mRNA [NM_015596] |
| A_24_P3336 | 0,0683104 | up | 5,4190063 | 5,4190063 | 2,4380283 | KLK13 | Homo sapiens kallikrein-related peptidase 13 (KLK13), mRNA [NM_015596] |
