## Supplementary material for "Early Reduction of SARS-CoV-2 Replication in Bronchial Epithelium by Kinin B_2_ Receptor Antagonism": Supl. Table 3

**Supplemental Table S3.** DEGs from transcriptome analysis filtered on kinin-kallikrein-system members from nasal scrapings from SARS-CoV-2 acute positive patients compared to negative patients

| ProbeName | p ([pos post] \ | Regulation ([ | FC (abs) ([pos | FC ([pos post | Log FC ([pos | GeneSymbol | Description |
|---|---|---|---|---|---|---|---|
| A_24_P2601 | 1 | up | 4,4537134 | 4,4537134 | 2,1550088 | MME | Homo sapiens membrane metallo-endopeptidase (MME), transcript variant 2b, mRNA [NM_007289] |
| A_33_P3370 | 0,03633852 | up | 5,9069815 | 5,9069815 | 2,562421 | MME | Homo sapiens membrane metallo-endopeptidase (MME), transcript variant 2b, mRNA [NM_007289] |
| A_33_P3290 | 0,04381223 | up | 3,7117934 | 3,7117934 | 1,8921164 | MME | Homo sapiens membrane metallo-endopeptidase (MME), transcript variant 2b, mRNA [NM_007289] |
| A_23_P1279 | 0,06497923 | down | 1,3449621 | -1,3449621 | -0,4275655 | PRCP | Homo sapiens prolylcarboxypeptidase (angiotensinase C) (PRCP), transcript variant 2, mRNA [NM_199418] |
| A_33_P3233 | 0,04036628 | down | 1,856378 | -1,856378 | -0,8924905 | F12 | Homo sapiens coagulation factor XII (Hageman factor) (F12), mRNA [NM_000505] |
| A_33_P3305 | 0,02187797 | down | 1,8841604 | -1,8841604 | -0,9139218 | NOS3 | Homo sapiens nitric oxide synthase 3 (endothelial cell) (NOS3), transcript variant 1, mRNA [NM_000603] |
| A_23_P3048 | 0,00288877 | up | 1,6989697 | 1,6989697 | 0,7646601 | BDKRB2 | Homo sapiens bradykinin receptor B2 (BDKRB2), mRNA [NM_000623] |
| A_23_P1391 | 0,40203622 | up | 1,4733071 | 1,4733071 | 0,5590582 | SERPING1 | Homo sapiens serpin peptidase inhibitor, clade G (C1 inhibitor), member 1 (SERPING1), transcript variant 1, mRNA [NM_000062] |
| A_33_P3215 | 0,00128211 | up | 1,9917675 | 1,9917675 | 0,99404925 | KLK3 | kallikrein-related peptidase 3 [Source:HGNC Symbol;Acc:HGNC:6364] [ENST00000595151] |
| A_33_P3413 | 0,17071521 | up | 2,0435178 | 2,0435178 | 1,0310549 | SERPING1 | Homo sapiens serpin peptidase inhibitor, clade G (C1 inhibitor), member 1 (SERPING1), transcript variant 1, mRNA [NM_000062] |
| A_23_P3463 | 0,3897618 | up | 1,3292397 | 1,3292397 | 0,41060132 | REN | Homo sapiens renin (REN), mRNA [NM_000537] |
| A_23_P1287 | 0,01012241 | up | 1,7931846 | 1,7931846 | 0,84252405 | BDKRB1 | Homo sapiens bradykinin receptor B1 (BDKRB1), mRNA [NM_000710] |
| A_33_P3413 | 0,08619295 | up | 1,5809374 | 1,5809374 | 0,6607802 | SERPING1 | Homo sapiens serpin peptidase inhibitor, clade G (C1 inhibitor), member 1 (SERPING1), transcript variant 1, mRNA [NM_000062] |
| A_23_P2529 | 0,31028056 | up | 1,2636952 | 1,2636952 | 0,33764857 | ACE2 | Homo sapiens angiotensin I converting enzyme 2 (ACE2), mRNA [NM_021804] |
| A_23_P9814 | 0,1239582 | up | 1,208384 | 1,208384 | 0,27307904 | CPN1 | Homo sapiens carboxypeptidase N, polypeptide 1 (CPN1), mRNA [NM_001308] |
| A_33_P3235 | 0,39434123 | up | 2,7426543 | 2,7426543 | 1,4555727 | KLK6 | Homo sapiens kallikrein-related peptidase 6 (KLK6), transcript variant B, mRNA [NM_001012964] |
| A_24_P4166 | 0,55089855 | up | 1,7370806 | 1,7370806 | 0,79666466 | KLK13 | Homo sapiens kallikrein-related peptidase 13 (KLK13), mRNA [NM_015596] |
| A_24_P3336 | 0,5001939 | up | 1,743478 | 1,743478 | 0,8019681 | KLK13 | Homo sapiens kallikrein-related peptidase 13 (KLK13), mRNA [NM_015596] |
| A_24_P2369 | 1 | up | 1,7377607 | 1,7377607 | 0,7972294 | KLK6 | Homo sapiens kallikrein-related peptidase 6 (KLK6), transcript variant B, mRNA [NM_001012964] |
| A_33_P3251 | 1 | up | 1,9308945 | 1,9308945 | 0,9492693 | KLK13 | kallikrein-related peptidase 13 [Source:HGNC Symbol;Acc:HGNC:6361] [ENST00000602090] |
| A_23_P2122 | 1 | up | 1,7885467 | 1,7885467 | 0,83878773 | KNG1 | Homo sapiens kininogen 1 (KNG1), transcript variant 2, mRNA [NM_000893] |
