## Supplementary material for "Early Reduction of SARS-CoV-2 Replication in Bronchial Epithelium by Kinin B_2_ Receptor Antagonism": Supl. Table 4

**Supplemental Table S4.** DEGs comparing SARS-CoV-2 + B2R antagonist versus SARS-CoV-2

| ProbeName | p ([SARS-CoV-2 | Regulatio | FC ([SARS-Co | GeneSymbol | Description |
|---|---|---|---|---|---|
| A_23_P91970 | 0,031047111 | down | -1,5092839 | AADACL2 | Homo sapiens arylacetamide deacetylase-like 2 (AADACL2), mRNA [NM_207365] |
| A_33_P3366161 | 0,010631708 | down | -2,1879694 | ABAT | Homo sapiens 4-aminobutyrate aminotransferase (ABAT), transcript variant 2, mRNA [NM_000663] |
| A_23_P24774 | 0,019292368 | down | -2,6067395 | ABCC8 | Homo sapiens ATP-binding cassette, sub-family C (CFTR/MRP), member 8 (ABCC8), transcript variant 2, mRNA [NM_000352] |
| A_23_P119763 | 0,033049513 | down | -1,9194201 | ABCG5 | Homo sapiens ATP-binding cassette, sub-family G (WHITE), member 5 (ABCG5), mRNA [NM_022436] |
| A_33_P3333480 | 0,00399051 | down | -1,6237004 | ABHD11-AS1 | Homo sapiens ABHD11 antisense RNA 1 (tail to tail) (ABHD11-AS1), long non-coding RNA [NR_026690] |
| A_23_P366035 | 0,031300526 | down | -1,7492104 | ABHD12B | Homo sapiens abhydrolase domain containing 12B (ABHD12B), transcript variant 1, mRNA [NM_001206673] |
| A_23_P376727 | 0,028580409 | down | -2,0238543 | ACMSD | Homo sapiens aminocarboxymuconate semialdehyde decarboxylase (ACMSD), mRNA [NM_138326] |
| A_33_P3352517 | 3,58E-04 | down | -2,8436701 | ACOT6 | Homo sapiens acyl-CoA thioesterase 6 (ACOT6), mRNA [NM_001037162] |
| A_23_P330419 | 0,03193642 | down | -2,072469 | ACPT | Homo sapiens acid phosphatase, testicular (ACPT), mRNA [NM_033068] |
| A_33_P3382849 | 0,0203917 | down | -1,8869588 | ACR | Homo sapiens acrosin (ACR), mRNA [NM_001097] |
| A_33_P3265129 | 0,008068424 | down | -2,23569 | ACSM2A | Homo sapiens cDNA FLJ34659 fis, clone KIDNE2018863. [AK091978] |
| A_24_P92680 | 0,0381286 | down | -1,7189307 | ACTA2-AS1 | Homo sapiens ACTA2 antisense RNA 1 (ACTA2-AS1), long non-coding RNA [NR_125373] |
| A_33_P3236382 | 0,040037736 | down | -1,5442077 | ACTC1 | Homo sapiens actin, alpha, cardiac muscle 1 (ACTC1), mRNA [NM_005159] |
| A_33_P3345011 | 0,035479486 | down | -1,8101462 | ACTL9 | Homo sapiens actin-like 9 (ACTL9), mRNA [NM_178525] |
| A_33_P3864411 | 0,001478873 | down | -1,9494934 | ACTR3BP5 | Homo sapiens ACTR3B pseudogene 5 (ACTR3BP5), non-coding RNA [NR_045000] |
| A_23_P147311 | 0,002560156 | down | -1,7518705 | ACTRT1 | Homo sapiens actin-related protein T1 (ACTRT1), mRNA [NM_138289] |
| A_23_P148829 | 0,023806369 | down | -1,5880963 | ACTRT2 | Homo sapiens actin-related protein T2 (ACTRT2), mRNA [NM_080431] |
| A_24_P945113 | 0,0275925 | down | -1,5558444 | ACVRL1 | Homo sapiens activin A receptor type II-like 1 (ACVRL1), transcript variant 1, mRNA [NM_000020] |
| A_23_P72527 | 0,04390732 | down | -1,627356 | ADAM18 | Homo sapiens ADAM metallopeptidase domain 18 (ADAM18), transcript variant 1, mRNA [NM_014237] |
| A_23_P367420 | 0,040144276 | up | 1,7933259 | ADAM21 | Homo sapiens ADAM metallopeptidase domain 21 (ADAM21), mRNA [NM_003813] |
| A_23_P95536 | 0,018302761 | down | -2,1320918 | ADAM29 | Homo sapiens ADAM metallopeptidase domain 29 (ADAM29), transcript variant 1, mRNA [NM_014269] |
| A_23_P44648 | 0,034182634 | down | -1,856451 | ADAMTS12 | Homo sapiens ADAM metallopeptidase with thrombospondin type 1 motif, 12 (ADAMTS12), mRNA [NM_030955] |
| A_23_P321307 | 0,006446154 | down | -2,5941637 | ADAMTS2 | Homo sapiens ADAM metallopeptidase with thrombospondin type 1 motif, 2 (ADAMTS2), transcript variant 2, mRNA [NM_021599] |
| A_32_P29954 | 0,012432293 | down | -2,5164652 | ADARB2-AS1 | Homo sapiens ADARB2 antisense RNA 1 (ADARB2-AS1), long non-coding RNA [NR_033387] |
| A_23_P169993 | 0,017135117 | down | -1,6758021 | ADCY8 | Homo sapiens adenylate cyclase 8 (brain) (ADCY8), mRNA [NM_001115] |
| A_23_P81158 | 0,003657068 | down | -3,4838548 | ADH1C | Homo sapiens alcohol dehydrogenase 1C (class I), gamma polypeptide (ADH1C), mRNA [NM_000669] |
| A_32_P423691 | 0,010570923 | down | -1,5737296 | ADIG | Homo sapiens adipogenin (ADIG), mRNA [NM_001018082] |
| A_21_P0000571 | 0,008775 | down | -1,5772151 | ADORA2A-AS1 | Homo sapiens ADORA2A antisense RNA 1 (ADORA2A-AS1), transcript variant 1, long non-coding RNA [NR_028484] |
| A_21_P0000570 | 0,01825944 | down | -1,7880782 | ADORA2A-AS1 | Homo sapiens ADORA2A antisense RNA 1 (ADORA2A-AS1), transcript variant 2, long non-coding RNA [NR_028483] |
| A_23_P168993 | 0,003327001 | down | -3,0895123 | ADRB3 | Homo sapiens adrenoceptor beta 3 (ADRB3), mRNA [NM_000025] |
| A_33_P3293888 | 6,50E-04 | down | -3,0700345 | AFF2 | Homo sapiens AF4/FMR2 family, member 2 (AFF2), transcript variant 1, mRNA [NM_002025] |
| A_33_P3373348 | 0,005118633 | down | -2,6155484 | AFF3 | AF4/FMR2 family, member 3 [Source:HGNC Symbol;Acc:HGNC:6473] [ENST00000483600] |
| A_33_P3378081 | 0,045776542 | down | -1,8695127 | AGBL1 | Homo sapiens ATP/GTP binding protein-like 1 (AGBL1), mRNA [NM_152336] |
| A_33_P3378061 | 0,034953818 | down | -1,910332 | AGBL3 | Homo sapiens ATP/GTP binding protein-like 3 (AGBL3), mRNA [NM_178563] |
| A_23_P62309 | 0,027139816 | down | -1,5687008 | AGTR2 | Homo sapiens angiotensin II receptor, type 2 (AGTR2), mRNA [NM_000686] |
| A_33_P3376493 | 0,033458207 | down | -1,8630021 | AGTR2 | Homo sapiens angiotensin II receptor, type 2 (AGTR2), mRNA [NM_000686] |
| A_24_P933685 | 0,010968552 | down | -1,8081734 | AGVR6190 | PREDICTED: Homo sapiens AGVR6190 (LOC643797), misc_RNA [XR_109223] |
| A_33_P3250857 | 0,015817668 | down | -1,7025341 | AIDA | axin interactor, dorsalization associated [Source:HGNC Symbol;Acc:HGNC:25761] [ENST00000474863] |
| A_33_P3319231 | 0,014342361 | down | -2,0619934 | AIFM2 | apoptosis-inducing factor, mitochondrion-associated, 2 [Source:HGNC Symbol;Acc:HGNC:21411] [ENST00000373248] |
| A_24_P342632 | 0,0286455 | down | -2,01527 | AK5 | Homo sapiens adenylate kinase 5 (AK5), transcript variant 1, mRNA [NM_174858] |
| A_33_P3384078 | 0,009886637 | down | -2,200895 | AK9 | Homo sapiens cDNA FLJ16163 fis, clone BRCAN2014229. [AK131244] |
| A_23_P313652 | 0,046302255 | down | -2,0710914 | AKAP14 | Homo sapiens A kinase (PRKA) anchor protein 14 (AKAP14), transcript variant 1, mRNA [NM_178813] |
| A_33_P3417920 | 1,77E-04 | down | -2,8426435 | AKAP4 | Homo sapiens A kinase (PRKA) anchor protein 4 (AKAP4), transcript variant 1, mRNA [NM_003886] |
| A_33_P3263569 | 0,016882548 | down | -1,7288859 | AKNAD1 | Homo sapiens AKNA domain containing 1 (AKNAD1), transcript variant 1, mRNA [NM_152763] |
| A_33_P3272291 | 0,020097796 | down | -1,9837346 | AKR1C4 | Homo sapiens aldo-keto reductase family 1, member C4 (AKR1C4), mRNA [NM_001818] |

| | | | | | |
|---|---|---|---|---|---|
| A_33_P3392142 | 0,027351584 | down | -1,566315 | AKR1D1 | aldo-keto reductase family 1, member D1 [Source:HGNC Symbol;Acc:HGNC:388] [ENST00000468877] |
| A_24_P218688 | 0,028159836 | down | -1,8972586 | ALDH3B1 | Homo sapiens aldehyde dehydrogenase 3 family, member B1 (ALDH3B1), transcript variant 1, mRNA [NM_000694] |
| A_23_P44867 | 0,03626733 | down | -2,4372706 | AMBN | Homo sapiens ameloblastin (enamel matrix protein) (AMBN), mRNA [NM_016519] |
| A_32_P122226 | 0,046123333 | down | -2,0504458 | AMDHD1 | Homo sapiens amidohydrolase domain containing 1 (AMDHD1), mRNA [NM_152435] |
| A_23_P96478 | 1,25E-04 | down | -2,7986672 | AMELX | Homo sapiens amelogenin, X-linked (AMELX), transcript variant 2, mRNA [NM_182681] |
| A_33_P3295538 | 0,03661888 | down | -1,7001997 | AMIGO3 | Homo sapiens adhesion molecule with Ig-like domain 3 (AMIGO3), mRNA [NM_198722] |
| A_23_P31273 | 0,025942856 | down | -2,722401 | AMPH | Homo sapiens amphiphysin (AMPH), transcript variant 1, mRNA [NM_001635] |
| A_23_P216023 | 5,12E-07 | down | -5,139572 | ANGPT1 | Homo sapiens angiopoietin 1 (ANGPT1), transcript variant 1, mRNA [NM_001146] |
| A_33_P3377703 | 0,039833374 | down | -1,8895715 | ANKRD20A19P | Homo sapiens ankyrin repeat domain 20 family, member A19, pseudogene (ANKRD20A19P), non-coding RNA [NR_073430] |
| A_33_P3343457 | 0,005887221 | down | -2,2402024 | ANKRD30B | Homo sapiens ankyrin repeat domain 30B (ANKRD30B), mRNA [NM_001145029] |
| A_24_P917819 | 0,021567624 | up | 1,7993237 | ANKRD30BP2 | Homo sapiens ankyrin repeat domain 30B pseudogene 2 (ANKRD30BP2), non-coding RNA [NR_026916] |
| A_24_P128713 | 0,024127888 | down | -1,5014349 | ANKRD44 | ankyrin repeat domain 44 [Source:HGNC Symbol;Acc:HGNC:25259] [ENST00000328737] |
| A_33_P3258004 | 0,043464538 | down | -1,6642784 | ANKS1B | Homo sapiens ankyrin repeat and sterile alpha motif domain containing 1B (ANKS1B), transcript variant 1, mRNA [NM_152788] |
| A_33_P3360296 | 0,035806593 | down | -1,9404445 | ANTXRL | Homo sapiens anthrax toxin receptor-like (ANTXRL), mRNA [NM_001278688] |
| A_21_P0010946 | 0,007583805 | down | -1,7793748 | ANTXRLP1 | Homo sapiens anthrax toxin receptor-like pseudogene 1 (ANTXRLP1), transcript variant 2, non-coding RNA [NR_103828] |
| A_21_P0010873 | 0,01194135 | down | -1,8027905 | ANTXRLP1 | Homo sapiens anthrax toxin receptor-like pseudogene 1 (ANTXRLP1), transcript variant 2, non-coding RNA [NR_103828] |
| A_23_P145718 | 0,028939143 | down | -1,5836462 | AOAH | Homo sapiens acyloxyacyl hydrolase (neutrophil) (AOAH), transcript variant 1, mRNA [NM_001637] |
| A_23_P59452 | 0,02375651 | down | -1,6636437 | AOC1 | Homo sapiens amine oxidase, copper containing 1 (AOC1), transcript variant 2, mRNA [NM_001091] |
| A_21_P0000813 | 0,00835472 | down | -1,8337498 | APELA | Homo sapiens apelin receptor early endogenous ligand (APELA), mRNA [NM_001297550] |
| A_23_P208302 | 0,006099036 | down | -2,23923 | APOC2 | Homo sapiens apolipoprotein C-II (APOC2), mRNA [NM_000483] |
| A_23_P26522 | 0,018260319 | down | -2,4486797 | AQP8 | Homo sapiens aquaporin 8 (AQP8), mRNA [NM_001169] |
| A_23_P106362 | 0,024555318 | down | -1,551873 | AQP9 | Homo sapiens aquaporin 9 (AQP9), mRNA [NM_020980] |
| A_33_P3279720 | 0,017673923 | down | -1,5726097 | ARFRP1 | ADP-ribosylation factor related protein 1 [Source:HGNC Symbol;Acc:HGNC:662] [ENST00000612772] |
| A_24_P380061 | 0,018220019 | down | -1,526954 | ARHGAP24 | Homo sapiens Rho GTPase activating protein 24 (ARHGAP24), transcript variant 1, mRNA [NM_001025616] |
| A_23_P72968 | 0,017233046 | down | -1,6389556 | ARHGAP36 | Homo sapiens Rho GTPase activating protein 36 (ARHGAP36), transcript variant 1, mRNA [NM_144967] |
| A_33_P3299110 | 0,003256604 | down | -1,678147 | ARHGAP42 | Homo sapiens Rho GTPase activating protein 42 (ARHGAP42), mRNA [NM_152432] |
| A_33_P3410385 | 0,004025266 | down | -1,7739966 | ARHGAP44 | Homo sapiens Rho GTPase activating protein 44 (ARHGAP44), mRNA [NM_014859] |
| A_33_P3610771 | 0,016006975 | down | -1,5720814 | ARHGEF38 | Homo sapiens Rho guanine nucleotide exchange factor (GEF) 38 (ARHGEF38), transcript variant 1, mRNA [NM_001242729] |
| A_33_P3222009 | 0,04425587 | down | -1,9453231 | ARL14EPL | Homo sapiens ADP-ribosylation factor-like 14 effector protein-like (ARL14EPL), mRNA [NM_001195581] |
| A_23_P86540 | 0,02722762 | down | -1,5565178 | ARMC3 | Homo sapiens armadillo repeat containing 3 (ARMC3), transcript variant 1, mRNA [NM_173081] |
| A_21_P0013851 | 0,038902566 | down | -1,5179518 | ARMCX4 | Homo sapiens armadillo repeat containing, X-linked 4 (ARMCX4), transcript variant 6, non-coding RNA [NR_045863] |
| A_33_P3232334 | 0,03092569 | down | -2,3280091 | ARMCX4 | armadillo repeat containing, X-linked 4 [Source:HGNC Symbol;Acc:HGNC:28615] [ENST00000430461] |
| A_33_P3303785 | 0,011812359 | down | -1,5777488 | ARPP21 | Homo sapiens cAMP-regulated phosphoprotein, 21kDa (ARPP21), transcript variant 4, mRNA [NM_001025069] |
| A_23_P116902 | 0,047357626 | down | -2,4163625 | ART4 | Homo sapiens ADP-ribosyltransferase 4 (Dombrock blood group) (ART4), mRNA [NM_021071] |
| A_33_P3210622 | 0,005283285 | down | -2,1994066 | ASB13 | Homo sapiens ankyrin repeat and SOCS box containing 13 (ASB13), transcript variant 1, mRNA [NM_024701] |
| A_33_P3290162 | 0,043313116 | down | -1,6179641 | ASB18 | Homo sapiens ankyrin repeat and SOCS box containing 18 (ASB18), mRNA [NM_212556] |
| A_33_P3264394 | 0,030635294 | down | -1,6542329 | ASPSCR1 | alveolar soft part sarcoma chromosome region, candidate 1 [Source:HGNC Symbol;Acc:HGNC:13825] [ENST00000583693] |
| A_33_P3220738 | 0,020830175 | down | -1,9815345 | ASTN1 | Homo sapiens astrotactin 1 (ASTN1), transcript variant 2, mRNA [NM_207108] |
| A_33_P3371904 | 8,19E-04 | down | -2,8000426 | ASTN2 | Homo sapiens astrotactin 2 (ASTN2), transcript variant 4, mRNA [NM_198188] |
| A_33_P3217258 | 0,015778339 | down | -1,7088526 | ASZ1 | Homo sapiens ankyrin repeat, SAM and basic leucine zipper domain containing 1 (ASZ1), transcript variant 1, mRNA [NM_130768] |
| A_33_P3708763 | 0,01837297 | down | -2,1448555 | ATP11A-AS1 | Homo sapiens ATP11A antisense RNA 1 (ATP11A-AS1), transcript variant 2, long non-coding RNA [NR_109811] |
| A_33_P3269423 | 0,02057438 | down | -2,9579024 | ATP11AUN | Homo sapiens ATP11A upstream neighbor (ATP11AUN), mRNA [NM_207440] |
| A_33_P3283669 | 0,007015104 | down | -1,7080164 | ATP1A3 | Homo sapiens ATPase, Na+/K+ transporting, alpha 3 polypeptide (ATP1A3), transcript variant 3, mRNA [NM_001256214] |
| A_24_P31275 | 0,009639558 | down | -3,8274825 | ATP1B2 | Homo sapiens ATPase, Na+/K+ transporting, beta 2 polypeptide (ATP1B2), transcript variant 1, mRNA [NM_001678] |
| A_23_P72462 | 0,044738587 | up | 1,6080322 | ATP2A1 | Homo sapiens ATPase, Ca++ transporting, cardiac muscle, fast twitch 1 (ATP2A1), transcript variant b, mRNA [NM_173201] |
| A_23_P23797 | 0,015121167 | down | -1,8671578 | ATP6V1G3 | Homo sapiens ATPase, H+ transporting, lysosomal 13kDa, V1 subunit G3 (ATP6V1G3), transcript variant 2, mRNA [NM_133326] |
| A_23_P163216 | 0,011910637 | down | -1,6608536 | ATP8B4 | Homo sapiens ATPase, class I, type 8B, member 4 (ATP8B4), transcript variant 1, mRNA [NM_024837] |
| A_33_P3343473 | 0,011346163 | down | -1,6308907 | ATRNL1 | Homo sapiens attractin-like 1 (ATRNL1), transcript variant 1, mRNA [NM_207303] |
| A_33_P3330211 | 0,001019423 | down | -2,4211652 | AUTS2 | Homo sapiens autism susceptibility candidate 2 (AUTS2), transcript variant 3, mRNA [NM_001127232] |

| | | | | | |
|---|---|---|---|---|---|
| A_23_P148015 | 0,011985207 | down | -2,3226242 | AXIN2 | Homo sapiens axin 2 (AXIN2), mRNA [NM_004655] |
| A_33_P3279353 | 0,03582275 | down | -1,6726085 | AZU1 | Homo sapiens azurocidin 1 (AZU1), mRNA [NM_001700] |
| A_33_P3286958 | 0,016257666 | down | -2,0075836 | B3GALT2 | Homo sapiens UDP-Gal:betaGlcNAc beta 1,3-galactosyltransferase, polypeptide 2 (B3GALT2), mRNA [NM_003783] |
| A_23_P39755 | 0,001524212 | down | -2,2593756 | B3GNT7 | Homo sapiens UDP-GlcNAc:betaGal beta-1,3-N-acetylglucosaminyltransferase 7 (B3GNT7), mRNA [NM_145236] |
| A_21_P0011660 | 0,020721408 | down | -1,6141876 | B4GALT6 | PREDICTED: Homo sapiens UDP-Gal:betaGlcNAc beta 1,4- galactosyltransferase, polypeptide 6 (B4GALT6), transcript variant X1, m |
| A_21_P0006033 | 0,039523117 | down | -1,5338823 | BARX1-AS1 | BARX1 antisense RNA 1 (head to head) [Source:HGNC Symbol;Acc:HGNC:50673] [ENST00000453045] |
| A_23_P212050 | 0,035425033 | down | -1,6021214 | BCHE | Homo sapiens butyrylcholinesterase (BCHE), mRNA [NM_000055] |
| A_21_P0014929 | 0,011775185 | up | 2,0829556 | BCR | Homo sapiens breakpoint cluster region (BCR), transcript variant 1, mRNA [NM_004327] |
| A_23_P79769 | 0,004277087 | down | -2,0157063 | BIRC7 | Homo sapiens baculoviral IAP repeat containing 7 (BIRC7), transcript variant 2, mRNA [NM_022161] |
| A_23_P154643 | 0,007747203 | down | -2,342536 | BMP7 | Homo sapiens bone morphogenetic protein 7 (BMP7), mRNA [NM_001719] |
| A_33_P3285195 | 0,003501021 | down | -1,6852245 | BMPR1B-AS1 | Homo sapiens BMPR1B antisense RNA 1 (head to head) (BMPR1B-AS1), long non-coding RNA [NR_121610] |
| A_32_P133916 | 0,042214923 | down | -1,8151916 | BNC2 | Homo sapiens basonuclin 2 (BNC2), mRNA [NM_017637] |
| A_23_P43684 | 0,006747896 | down | -1,8317413 | BNC2 | Homo sapiens basonuclin 2 (BNC2), mRNA [NM_017637] |
| A_23_P102083 | 0,027096966 | down | -1,5181345 | BOLL | Homo sapiens boule-like RNA-binding protein (BOLL), transcript variant 2, mRNA [NM_033030] |
| A_23_P254558 | 0,03405087 | down | -1,7469546 | BPESC1 | Homo sapiens blepharophimosis, epicanthus inversus and ptosis, candidate 1 (non-protein coding) (BPESC1), long non-coding RNA |
| A_19_P00321575 | 0,021109326 | down | -2,1044424 | BPESC1 | Homo sapiens blepharophimosis, epicanthus inversus and ptosis, candidate 1 (non-protein coding) (BPESC1), long non-coding RNA |
| A_23_P57222 | 0,018512258 | down | -2,3026552 | BPIFB3 | BPI fold containing family B, member 3 [Source:HGNC Symbol;Acc:HGNC:16178] [ENST00000375494] |
| A_32_P539109 | 0,009570602 | down | -1,9971515 | BPIFB6 | Homo sapiens BPI fold containing family B, member 6 (BPIFB6), mRNA [NM_174897] |
| A_23_P45598 | 0,04977297 | down | -2,075018 | BPY2B | Homo sapiens basic charge, Y-linked, 2B (BPY2B), mRNA [NM_001002760] |
| A_23_P94517 | 0,049273077 | down | -2,1822615 | BRINP1 | Homo sapiens bone morphogenetic protein/retinoic acid inducible neural-specific 1 (BRINP1), mRNA [NM_014618] |
| A_33_P3391915 | 0,041581843 | down | -1,6054517 | BTBD17 | Homo sapiens BTB (POZ) domain containing 17 (BTBD17), mRNA [NM_001080466] |
| A_33_P3301266 | 0,019472696 | down | -1,9919575 | BTNL8 | Homo sapiens butyrophilin-like 8 (BTNL8), transcript variant 4, mRNA [NM_001159708] |
| A_33_P3246198 | 0,04825811 | down | -1,5400901 | C10orf120 | Homo sapiens chromosome 10 open reading frame 120 (C10orf120), mRNA [NM_001010912] |
| A_23_P325700 | 0,023371512 | down | -1,8746072 | C10orf126 | Homo sapiens chromosome 10 open reading frame 126 (C10orf126), mRNA [NM_001278522] |
| A_24_P333635 | 0,028001955 | down | -1,6796358 | C10orf62 | Homo sapiens chromosome 10 open reading frame 62 (C10orf62), mRNA [NM_001009997] |
| A_32_P337442 | 0,04366835 | down | -1,8383653 | C10orf67 | Homo sapiens chromosome 10 open reading frame 67 (C10orf67), mRNA [NM_153714] |
| A_21_P0006760 | 0,024898514 | down | -1,8521272 | C10orf71-AS1 | Homo sapiens C10orf71 antisense RNA 1 (C10orf71-AS1), long non-coding RNA [NR_108038] |
| A_33_P3366659 | 0,01611085 | down | -1,631607 | C12orf54 | chromosome 12 open reading frame 54 [Source:HGNC Symbol;Acc:HGNC:28553] [ENST00000380491] |
| A_32_P741217 | 0,0328319 | down | -1,5199456 | C14orf177 | Homo sapiens chromosome 14 open reading frame 177 (C14orf177), mRNA [NM_182560] |
| A_23_P372356 | 0,00120783 | down | -2,9739976 | C15orf32 | Homo sapiens chromosome 15 open reading frame 32 (C15orf32), transcript variant 1, mRNA [NM_153040] |
| A_23_P325593 | 0,027028698 | down | -1,5072515 | C15orf43 | Homo sapiens chromosome 15 open reading frame 43 (C15orf43), mRNA [NM_152448] |
| A_23_P26024 | 0,048014753 | up | 1,5385834 | C15orf48 | Homo sapiens chromosome 15 open reading frame 48 (C15orf48), transcript variant 2, mRNA [NM_032413] |
| A_33_P3390017 | 0,024789764 | down | -2,0008724 | C15orf54 | Homo sapiens chromosome 15 open reading frame 54 (C15orf54), transcript variant 1, mRNA [NM_207445] |
| A_21_P0008831 | 0,017732698 | down | -1,634158 | C15orf59-AS1 | |
| A_33_P3417840 | 0,028465047 | down | -1,5116956 | C16orf47 | Homo sapiens chromosome 16 open reading frame 47 (C16orf47), mRNA [NM_207385] |
| A_33_P3405193 | 0,04606069 | down | -1,7128793 | C16orf72 | Homo sapiens PRO0149 protein, mRNA (cDNA clone IMAGE:5172419), containing frame-shift errors. [BC029878] |
| A_33_P3417980 | 7,39E-04 | down | -3,3118792 | C16orf86 | chromosome 16 open reading frame 86 [Source:HGNC Symbol;Acc:HGNC:33755] [ENST00000459925] |
| A_21_P0009030 | 0,01749081 | down | -1,5929195 | C16orf97 | Homo sapiens chromosome 16 open reading frame 97 (C16orf97), mRNA [NM_001242473] |
| A_33_P3339187 | 0,025300113 | down | -2,837693 | C17orf112 | Homo sapiens chromosome 17 open reading frame 112 (C17orf112), mRNA [NM_001243552] |
| A_23_P335388 | 0,047291175 | up | 1,72744 | C17orf78 | Homo sapiens chromosome 17 open reading frame 78 (C17orf78), mRNA [NM_173625] |
| A_23_P50052 | 0,011242777 | down | -1,5241826 | C18orf12 | PREDICTED: Homo sapiens chromosome 18 open reading frame 12 (C18orf12), misc_RNA [XR_109479] |
| A_33_P3411945 | 0,014121277 | down | -1,8479769 | C19orf38 | Homo sapiens chromosome 19 open reading frame 38 (C19orf38), mRNA [NM_001136482] |
| A_32_P8925 | 0,019899461 | down | -1,9492047 | C1orf100 | Homo sapiens chromosome 1 open reading frame 100 (C1orf100), transcript variant 1, mRNA [NM_001012970] |
| A_32_P114746 | 0,025109492 | down | -2,0821435 | C1orf111 | Homo sapiens chromosome 1 open reading frame 111 (C1orf111), mRNA [NM_182581] |
| A_33_P3340055 | 0,021914668 | down | -2,376796 | C1orf167 | Homo sapiens chromosome 1 open reading frame 167 (C1orf167), mRNA [NM_001010881] |
| A_24_P94319 | 0,015115822 | down | -1,7896101 | C1orf177 | Homo sapiens chromosome 1 open reading frame 177 (C1orf177), transcript variant 1, mRNA [NM_152607] |
| A_33_P3403560 | 0,029438203 | down | -1,650747 | C1orf185 | Homo sapiens chromosome 1 open reading frame 185 (C1orf185), mRNA [NM_001136508] |
| A_32_P174693 | 0,047867108 | down | -1,8569555 | C1orf234 | Homo sapiens chromosome 1 open reading frame 234 (C1orf234), mRNA [NM_001242521] |
| A_21_P0014079 | 0,005703502 | down | -2,444142 | C1orf61 | chromosome 1 open reading frame 61 [Source:HGNC Symbol;Acc:HGNC:30780] [ENST00000476966] |

| | | | | | |
|---|---|---|---|---|---|
| A_23_P376172 | 0,015287323 | down | -1,6985577 | C1orf64 | Homo sapiens chromosome 1 open reading frame 64 (C1orf64), mRNA [NM_178840] |
| A_23_P137366 | 0,039651893 | down | -2,7504945 | C1QB | Homo sapiens complement component 1, q subcomponent, B chain (C1QB), mRNA [NM_000491] |
| A_33_P3357007 | 0,049568493 | down | -2,1569262 | C1QTNF9B | Homo sapiens C1q and tumor necrosis factor related protein 9B (C1QTNF9B), transcript variant 1, mRNA [NM_001007537] |
| A_32_P162187 | 0,012271603 | down | -1,7030604 | C2 | Homo sapiens complement component 2 (C2), transcript variant 1, mRNA [NM_000063] |
| A_33_P3419831 | 0,04683434 | down | -2,1296825 | C20orf62 | chromosome 20 open reading frame 62 [Source:HGNC Symbol;Acc:HGNC:16195] [ENST00000306731] |
| A_32_P45974 | 0,0219923 | down | -1,7609603 | C2orf27A | Homo sapiens chromosome 2 open reading frame 27, mRNA (cDNA clone IMAGE:6426166). [BC071972] |
| A_33_P3286372 | 0,010050143 | down | -1,9285448 | C2orf48 | Homo sapiens chromosome 2 open reading frame 48 (C2orf48), mRNA [NM_182626] |
| A_33_P3398998 | 0,033948105 | down | -1,7033397 | C2orf50 | Homo sapiens chromosome 2 open reading frame 50 (C2orf50), mRNA [NM_182500] |
| A_33_P3265714 | 0,0170712 | down | -1,9917232 | C2orf61 | Homo sapiens chromosome 2 open reading frame 61 (C2orf61), transcript variant 2, mRNA [NM_173649] |
| A_33_P3237207 | 0,022091318 | down | -2,4677203 | C2orf72 | Homo sapiens chromosome 2 open reading frame 72 (C2orf72), mRNA [NM_001144994] |
| A_33_P3351836 | 0,00362833 | down | -1,8438387 | C2orf73 | chromosome 2 open reading frame 73 [Source:HGNC Symbol;Acc:HGNC:26861] [ENST00000414747] |
| A_23_P17130 | 0,007990896 | down | -1,6558765 | C2orf88 | Homo sapiens chromosome 2 open reading frame 88 (C2orf88), transcript variant 1, mRNA [NM_001042519] |
| A_33_P3416682 | 0,005636004 | down | -1,8367797 | C2orf91 | Homo sapiens chromosome 2 open reading frame 91 (C2orf91), mRNA [NM_001242815] |
| A_23_P2431 | 7,59E-04 | down | -1,9494691 | C3AR1 | Homo sapiens complement component 3a receptor 1 (C3AR1), mRNA [NM_004054] |
| A_33_P3254811 | 0,004196629 | down | -3,2643504 | C3orf70 | Homo sapiens chromosome 3 open reading frame 70 (C3orf70), mRNA [NM_001025266] |
| A_33_P3262351 | 0,048966765 | down | -3,2656286 | C5orf49 | Homo sapiens chromosome 5 open reading frame 49 (C5orf49), mRNA [NM_001089584] |
| A_33_P3660204 | 0,04424352 | down | -1,5158918 | C5orf66-AS2 | Homo sapiens C5orf66 antisense RNA 2 (C5orf66-AS2), long non-coding RNA [NR_037895] |
| A_32_P85106 | 0,039613202 | down | -1,7143147 | C6orf118 | Homo sapiens chromosome 6 open reading frame 118 (C6orf118), mRNA [NM_144980] |
| A_33_P3219755 | 0,04777795 | down | -1,9811188 | C6orf132 | Homo sapiens chromosome 6 open reading frame 132 (C6orf132), mRNA [NM_001164446] |
| A_23_P421054 | 0,016666127 | down | -1,9419198 | C6orf165 | Homo sapiens chromosome 6 open reading frame 165 (C6orf165), mRNA [NM_001031743] |
| A_24_P350303 | 0,029164936 | down | -1,5414382 | C6orf201 | Homo sapiens chromosome 6 open reading frame 201 (C6orf201), transcript variant 1, mRNA [NM_001085401] |
| A_21_P0013066 | 0,02202922 | down | -1,5280143 | C6orf229 | Homo sapiens chromosome 6 open reading frame 229 (C6orf229), mRNA [NM_001282492] |
| A_23_P213857 | 0,002862326 | down | -2,2796526 | C7 | Homo sapiens complement component 7 (C7), mRNA [NM_000587] |
| A_33_P3405349 | 2,05E-04 | down | -2,122237 | C7orf43 | chromosome 7 open reading frame 43 [Source:HGNC Symbol;Acc:HGNC:25604] [ENST00000448720] |
| A_33_P3401397 | 0,010773891 | down | -2,5494263 | C7orf65 | Homo sapiens chromosome 7 open reading frame 65 (C7orf65), mRNA [NM_001123065] |
| A_33_P3396776 | 0,04445598 | down | -1,5308366 | C7orf66 | Homo sapiens chromosome 7 open reading frame 66 (C7orf66), mRNA [NM_001024607] |
| A_33_P3401003 | 0,005982795 | down | -1,7286302 | C7orf71 | Homo sapiens chromosome 7 open reading frame 71 (C7orf71), transcript variant 1, mRNA [NM_001145531] |
| A_33_P3213508 | 0,014024264 | down | -1,8523142 | C7orf72 | Homo sapiens chromosome 7 open reading frame 72 (C7orf72), mRNA [NM_001161834] |
| A_33_P3423854 | 0,02162742 | down | -1,5803097 | C8B | Homo sapiens complement component 8, beta polypeptide (C8B), transcript variant 2, mRNA [NM_001278543] |
| A_33_P3253169 | 0,01887733 | down | -2,5076232 | C8orf86 | chromosome 8 open reading frame 86 [Source:HGNC Symbol;Acc:HGNC:33774] [ENST00000358138] |
| A_33_P3287997 | 0,016095273 | down | -1,9255505 | C9orf106 | Homo sapiens chromosome 9 open reading frame 106 (C9orf106), mRNA [NM_001012715] |
| A_32_P48949 | 0,043982014 | down | -2,0104003 | C9orf129 | Homo sapiens chromosome 9 open reading frame 129 (C9orf129), mRNA [NM_001098808] |
| A_33_P3257703 | 0,017793648 | down | -1,6179146 | C9orf131 | Homo sapiens chromosome 9 open reading frame 131 (C9orf131), transcript variant 1, mRNA [NM_203299] |
| A_33_P3328270 | 0,025886964 | down | -1,8010575 | C9orf172 | Homo sapiens chromosome 9 open reading frame 172 (C9orf172), mRNA [NM_001080482] |
| A_33_P3353662 | 0,016242668 | down | -1,5686653 | C9orf66 | Homo sapiens chromosome 9 open reading frame 66 (C9orf66), mRNA [NM_152569] |
| A_32_P65706 | 0,004236935 | down | -2,0669386 | C9orf92 | Homo sapiens chromosome 9 open reading frame 92 (C9orf92), transcript variant 1, mRNA [NM_001271829] |
| A_23_P4096 | 0,011749357 | down | -2,2866106 | CA4 | Homo sapiens carbonic anhydrase IV (CA4), mRNA [NM_000717] |
| A_24_P241057 | 0,037146978 | down | -1,8314955 | CABP2 | Homo sapiens calcium binding protein 2 (CABP2), mRNA [NM_016366] |
| A_23_P308021 | 0,002962897 | down | -2,41373 | CABP4 | Homo sapiens calcium binding protein 4 (CABP4), transcript variant 1, mRNA [NM_145200] |
| A_33_P3280106 | 0,033068944 | down | -3,169259 | CACNA1B | Homo sapiens calcium channel, voltage-dependent, N type, alpha 1B subunit (CACNA1B), transcript variant 2, mRNA [NM_0012438 |
| A_23_P373031 | 9,21E-04 | down | -2,0299516 | CACNA1C | Homo sapiens calcium channel, voltage-dependent, L type, alpha 1C subunit (CACNA1C), transcript variant 18, mRNA [NM_000719 |
| A_24_P945096 | 0,048186537 | down | -1,8050053 | CACNA1I | Homo sapiens calcium channel, voltage-dependent, T type, alpha 1I subunit (CACNA1I), transcript variant 1, mRNA [NM_021096] |
| A_33_P3285275 | 0,042075284 | down | -1,8045393 | CACNB2 | calcium channel, voltage-dependent, beta 2 subunit [Source:HGNC Symbol;Acc:HGNC:1402] [ENST00000498816] |
| A_33_P3211138 | 0,012632152 | down | -2,2383876 | CADM1 | cell adhesion molecule 1 [Source:HGNC Symbol;Acc:HGNC:5951] [ENST00000452722] |
| A_33_P3261957 | 0,04173563 | down | -1,9484638 | CALCRL | Homo sapiens calcitonin receptor-like (CALCRL), transcript variant 1, mRNA [NM_005795] |
| A_32_P812268 | 0,004539526 | down | -1,9035704 | CALHM1 | Homo sapiens calcium homeostasis modulator 1 (CALHM1), mRNA [NM_001001412] |
| A_24_P380567 | 0,010855469 | down | -1,745703 | CALN1 | Homo sapiens calneuron 1 (CALN1), transcript variant 1, mRNA [NM_031468] |
| A_23_P42882 | 0,011911688 | down | -2,0619926 | CAMK2B | Homo sapiens calcium/calmodulin-dependent protein kinase II beta (CAMK2B), transcript variant 6, mRNA [NM_172082] |
| A_23_P380379 | 0,022989657 | down | -1,935372 | CAPS2 | Homo sapiens calcyphosine 2 (CAPS2), transcript variant 3, mRNA [NM_001286548] |

| | | | | | |
|---|---|---|---|---|---|
| A_33_P3235561 | 0,007799513 | up | 1,6712301 | CAPZB | Homo sapiens capping protein (actin filament) muscle Z-line, beta (CAPZB), transcript variant 3, mRNA [NM_001206541] |
| A_32_P800179 | 0,048436232 | down | -1,76493 | CARD8 | caspase recruitment domain family, member 8 [Source:HGNC Symbol;Acc:HGNC:17057] [ENST00000600800] |
| A_21_P0005629 | 0,012144235 | down | -1,8824894 | CASC11 | cancer susceptibility candidate 11 (non-protein coding) [Source:HGNC Symbol;Acc:HGNC:48939] [ENST00000502463] |
| A_21_P0009156 | 0,007579134 | down | -2,095656 | CASC17 | Homo sapiens cancer susceptibility candidate 17 (non-protein coding) (CASC17), long non-coding RNA [NR_104152] |
| A_32_P61480 | 0,043837786 | down | -1,5960506 | CASC2 | Homo sapiens cancer susceptibility candidate 2 (non-protein coding) (CASC2), transcript variant 3, long non-coding RNA [NR_02694 |
| A_33_P3213362 | 0,00587226 | down | -2,9656525 | CASC2 | Homo sapiens cancer susceptibility candidate 2 (non-protein coding) (CASC2), transcript variant 1, long non-coding RNA [NR_02693 |
| A_21_P0009099 | 0,04300558 | down | -1,5513371 | CASC22 | PREDICTED: Homo sapiens uncharacterized LOC283854 (LOC283854), misc_RNA [XR_243451] |
| A_23_P173 | 0,003097629 | down | -3,1827426 | CASQ1 | Homo sapiens calsequestrin 1 (fast-twitch, skeletal muscle) (CASQ1), mRNA [NM_001231] |
| A_33_P3280646 | 0,006615657 | down | -2,2763705 | CASR | Homo sapiens calcium-sensing receptor (CASR), transcript variant 1, mRNA [NM_001178065] |
| A_23_P29830 | 0,004990423 | up | 1,5528276 | CBLB | Homo sapiens Cbl proto-oncogene B, E3 ubiquitin protein ligase (CBLB), mRNA [NM_170662] |
| A_33_P3343845 | 0,02144335 | down | -1,6561091 | CBX7 | Homo sapiens chromobox homolog 7 (CBX7), mRNA [NM_175709] |
| A_33_P3308686 | 0,036245164 | down | -2,0938563 | CC2D2A | Homo sapiens coiled-coil and C2 domain containing 2A (CC2D2A), transcript variant 2, mRNA [NM_020785] |
| A_33_P3261586 | 0,039363004 | down | -1,5912447 | CCDC108 | Homo sapiens coiled-coil domain containing 108 (CCDC108), transcript variant 1, mRNA [NM_194302] |
| A_33_P3246528 | 0,031469293 | down | -1,9446043 | CCDC114 | Homo sapiens coiled-coil domain containing 114 (CCDC114), mRNA [NM_144577] |
| A_23_P429478 | 0,010966522 | down | -1,6122632 | CCDC140 | Homo sapiens coiled-coil domain containing 140 (CCDC140), mRNA [NM_153038] |
| A_33_P3356230 | 0,03703113 | down | -1,7634814 | CCDC144NL | Homo sapiens coiled-coil domain containing 144 family, N-terminal like (CCDC144NL), mRNA [NM_001004306] |
| A_33_P3238032 | 0,020225203 | down | -2,2519588 | CCDC158 | Homo sapiens coiled-coil domain containing 158 (CCDC158), mRNA [NM_001042784] |
| A_33_P3307008 | 0,021656247 | down | -2,4400527 | CCDC168 | Homo sapiens coiled-coil domain containing 168 (CCDC168), mRNA [NM_001146197] |
| A_21_P0006341 | 0,005785806 | down | -1,8723024 | CCDC171 | coiled-coil domain containing 171 [Source:HGNC Symbol;Acc:HGNC:29828] [ENST00000478913] |
| A_33_P3250865 | 0,040881567 | down | -1,7019154 | CCDC175 | coiled-coil domain containing 175 [Source:HGNC Symbol;Acc:HGNC:19847] [ENST00000281581] |
| A_23_P468 | 0,014091959 | down | -2,2289956 | CCDC181 | Homo sapiens coiled-coil domain containing 181 (CCDC181), transcript variant 2, mRNA [NM_021179] |
| A_23_P344194 | 0,007684334 | down | -2,0479565 | CCDC184 | Homo sapiens coiled-coil domain containing 184 (CCDC184), mRNA [NM_001013635] |
| A_33_P3328726 | 0,015948333 | down | -1,8559755 | CCDC33 | Homo sapiens coiled-coil domain containing 33 (CCDC33), transcript variant 1, mRNA [NM_025055] |
| A_23_P345175 | 0,033578776 | down | -1,7061088 | CCDC63 | Homo sapiens coiled-coil domain containing 63 (CCDC63), transcript variant 1, mRNA [NM_152591] |
| A_23_P356667 | 0,033996154 | down | -1,782858 | CCDC83 | Homo sapiens coiled-coil domain containing 83 (CCDC83), transcript variant 1, mRNA [NM_173556] |
| A_24_P192988 | 0,009657311 | down | -1,5435047 | CCDC89 | coiled-coil domain containing 89 [Source:HGNC Symbol;Acc:HGNC:26762] [ENST00000316398] |
| A_23_P49759 | 0,040338464 | down | -2,415574 | CCL1 | Homo sapiens chemokine (C-C motif) ligand 1 (CCL1), mRNA [NM_002981] |
| A_33_P3324004 | 0,016746009 | down | -1,8994434 | CCL15 | Homo sapiens chemokine (C-C motif) ligand 15 (CCL15), mRNA [NM_032965] |
| A_33_P3285945 | 1,15E-05 | down | -3,4042187 | CCL21 | Homo sapiens chemokine (C-C motif) ligand 21 (CCL21), mRNA [NM_002989] |
| A_24_P97405 | 0,041068777 | down | -1,8425514 | CCRL2 | Homo sapiens chemokine (C-C motif) receptor-like 2 (CCRL2), transcript variant 1, mRNA [NM_003965] |
| A_33_P3343360 | 0,002519094 | down | -2,087378 | CCSER1 | Homo sapiens coiled-coil serine-rich protein 1 (CCSER1), transcript variant 2, mRNA [NM_207491] |
| A_23_P310410 | 0,018581918 | down | -1,6482953 | CD1E | Homo sapiens CD1e molecule (CD1E), transcript variant 2, mRNA [NM_001042583] |
| A_33_P3278013 | 0,033897545 | down | -2,0211592 | CD2 | CD2 molecule [Source:HGNC Symbol;Acc:HGNC:1639] [ENST00000369477] |
| A_23_P121480 | 0,02160561 | up | 1,6621829 | CD200 | Homo sapiens CD200 molecule (CD200), transcript variant 2, mRNA [NM_001004196] |
| A_24_P186539 | 0,035061605 | down | -1,8416742 | CD209 | Homo sapiens CD209 molecule (CD209), transcript variant 1, mRNA [NM_021155] |
| A_23_P209055 | 0,03686409 | down | -1,6131264 | CD22 | Homo sapiens CD22 molecule (CD22), transcript variant 1, mRNA [NM_001771] |
| A_33_P3413558 | 0,034264807 | down | -2,0173213 | CD226 | CD226 molecule [Source:HGNC Symbol;Acc:HGNC:16961] [ENST00000280200] |
| A_23_P85453 | 0,00486417 | down | -3,3950548 | CD244 | Homo sapiens CD244 molecule, natural killer cell receptor 2B4 (CD244), transcript variant 1, mRNA [NM_016382] |
| A_23_P23829 | 0,006846983 | down | -2,085793 | CD34 | Homo sapiens CD34 molecule (CD34), transcript variant 2, mRNA [NM_001773] |
| A_33_P3225046 | 0,034216184 | down | -2,5512295 | CD34 | Homo sapiens CD34 molecule (CD34), transcript variant 1, mRNA [NM_001025109] |
| A_33_P3375541 | 0,001329882 | down | -2,4598093 | CD3D | Homo sapiens CD3d molecule, delta (CD3-TCR complex) (CD3D), transcript variant 1, mRNA [NM_000732] |
| A_33_P3241021 | 0,01925859 | down | -2,1987698 | CD69 | CD69 molecule [Source:HGNC Symbol;Acc:HGNC:1694] [ENST00000416624] |
| A_24_P320033 | 0,04820995 | down | -2,115845 | CD80 | Homo sapiens CD80 molecule (CD80), mRNA [NM_005191] |
| A_33_P3228837 | 0,007806487 | down | -1,7269272 | CD8A | Homo sapiens CD8a molecule (CD8A), transcript variant 3, mRNA [NM_001145873] |
| A_32_P56001 | 0,025102844 | down | -1,6389058 | CD93 | Homo sapiens CD93 molecule (CD93), mRNA [NM_012072] |
| A_23_P374281 | 0,02590909 | down | -1,5849372 | CDC20B | Homo sapiens cell division cycle 20B (CDC20B), transcript variant 1, mRNA [NM_152623] |
| A_33_P3397383 | 0,002665454 | down | -2,147482 | CDC20B | cell division cycle 20B [Source:HGNC Symbol;Acc:HGNC:24222] [ENST00000507931] |
| A_21_P0004295 | 0,045147456 | down | -1,9102632 | CDC42SE2 | CDC42 small effector 2 [Source:HGNC Symbol;Acc:HGNC:18547] [ENST00000515533] |
| A_23_P55586 | 0,023311725 | down | -1,5579506 | CDH20 | Homo sapiens cadherin 20, type 2 (CDH20), mRNA [NM_031891] |

| | | | | | |
|---|---|---|---|---|---|
| A_23_P92999 | 0,014992202 | down | -3,3550658 | CDH9 | Homo sapiens cadherin 9, type 2 (T1-cadherin) (CDH9), mRNA [NM_016279] |
| A_24_P51683 | 0,014340207 | down | -2,7784083 | CDK5R2 | Homo sapiens cyclin-dependent kinase 5, regulatory subunit 2 (p39) (CDK5R2), mRNA [NM_003936] |
| A_23_P30294 | 0,028361538 | down | -1,5275115 | CDO1 | Homo sapiens cysteine dioxygenase type 1 (CDO1), mRNA [NM_001801] |
| A_33_P3796201 | 0,018720688 | down | -2,0009687 | CDRT7 | Homo sapiens CMT1A duplicated region transcript 7 (non-protein coding) (CDRT7), long non-coding RNA [NR_033371] |
| A_33_P3249818 | 0,001202056 | down | -2,4600298 | CEACAM18 | Homo sapiens carcinoembryonic antigen-related cell adhesion molecule 18 (CEACAM18), mRNA [NM_001278392] |
| A_23_P78526 | 0,009063659 | down | -1,841083 | CEACAM19 | Homo sapiens carcinoembryonic antigen-related cell adhesion molecule 19 (CEACAM19), transcript variant 2, mRNA [NM_020219] |
| A_21_P0011782 | 0,03170947 | down | -1,6683136 | CEACAM22P | carcinoembryonic antigen-related cell adhesion molecule 2, pseudogene [Source:HGNC Symbol;Acc:HGNC:38029] [ENST0000045 |
| A_33_P3265309 | 0,02638436 | down | -1,658292 | CEACAM4 | Homo sapiens carcinoembryonic antigen-related cell adhesion molecule 4 (CEACAM4), mRNA [NM_001817] |
| A_23_P211326 | 0,010494229 | down | -2,0310352 | CECR2 | Homo sapiens cat eye syndrome chromosome region, candidate 2 (CECR2), transcript variant 1, mRNA [NM_001290047] |
| A_21_P0012243 | 0,006165844 | down | -1,599026 | CECR7 | cat eye syndrome chromosome region, candidate 7 (non-protein coding) [Source:HGNC Symbol;Acc:HGNC:1845] [ENST000006099 |
| A_33_P3313810 | 0,042647574 | down | -1,7413269 | CELA1 | Homo sapiens chymotrypsin-like elastase family, member 1 (CELA1), mRNA [NM_001971] |
| A_33_P3376872 | 0,008131525 | down | -1,5721785 | CELA3B | Homo sapiens chymotrypsin-like elastase family, member 3B (CELA3B), mRNA [NM_007352] |
| A_21_P0012348 | 0,030732593 | down | -1,6901684 | CES5A | Homo sapiens carboxylesterase 5A (CES5A), transcript variant 2, mRNA [NM_145024] |
| A_33_P3276303 | 0,039331276 | down | -1,872592 | CES5AP1 | Homo sapiens carboxylesterase 5A pseudogene 1 (CES5AP1), non-coding RNA [NR_037839] |
| A_23_P4074 | 0,01197975 | down | -1,9024229 | CFAP52 | Homo sapiens cilia and flagella associated protein 52 (CFAP52), transcript variant 2, mRNA [NM_145054] |
| A_33_P3336262 | 0,025655484 | down | -2,3982623 | CFAP74 | cilia and flagella associated protein 74 [Source:HGNC Symbol;Acc:HGNC:29368] [ENST00000412120] |
| A_24_P111406 | 0,04216407 | down | -1,5375454 | CFHR5 | Homo sapiens complement factor H-related 5 (CFHR5), mRNA [NM_030787] |
| A_23_P46894 | 0,007760174 | down | -2,4157164 | CHAT | Homo sapiens choline O-acetyltransferase (CHAT), transcript variant M, mRNA [NM_020549] |
| A_21_P0013981 | 0,02963747 | down | -1,7642552 | CHRM3 | cholinergic receptor, muscarinic 3 [Source:HGNC Symbol;Acc:HGNC:1952] [ENST00000481779] |
| A_33_P3215635 | 0,014044057 | down | -1,6950067 | CHRNB2 | Homo sapiens cholinergic receptor, nicotinic, beta 2 (neuronal) (CHRNB2), mRNA [NM_000748] |
| A_23_P123424 | 0,03585705 | down | -1,8078041 | CHRNB3 | Homo sapiens cholinergic receptor, nicotinic, beta 3 (neuronal) (CHRNB3), mRNA [NM_000749] |
| A_21_P0007879 | 0,040652055 | down | -1,5503924 | CISTR | Homo sapiens chondrogenesis-associated transcript (CISTR), transcript variant 1, long non-coding RNA [NR_104332] |
| A_23_P101683 | 0,044061646 | down | -1,7502303 | CLC | Homo sapiens Charcot-Leyden crystal galectin (CLC), mRNA [NM_001828] |
| A_23_P59772 | 0,002632682 | down | -2,0110087 | CLCN1 | Homo sapiens chloride channel, voltage-sensitive 1 (CLCN1), transcript variant 1, mRNA [NM_000083] |
| A_33_P3218089 | 0,012613019 | down | -2,398188 | CLDN24 | Homo sapiens claudin 24 (CLDN24), mRNA [NM_001185149] |
| A_23_P37942 | 0,006116098 | down | -2,4962442 | CLDN6 | Homo sapiens claudin 6 (CLDN6), mRNA [NM_021195] |
| A_33_P3303519 | 0,011558063 | down | -2,6391098 | CLEC12B | Homo sapiens C-type lectin domain family 12, member B (CLEC12B), transcript variant 2, mRNA [NM_205852] |
| A_33_P3332955 | 0,021471735 | down | -2,0002813 | CLEC1B | Homo sapiens C-type lectin domain family 1, member B (CLEC1B), transcript variant 1, mRNA [NM_016509] |
| A_33_P3295348 | 0,022303764 | down | -1,8181782 | CLEC2A | Homo sapiens C-type lectin domain family 2, member A (CLEC2A), transcript variant 1, mRNA [NM_001130711] |
| A_23_P42931 | 0,04913469 | down | -2,0257864 | CLEC2L | Homo sapiens C-type lectin domain family 2, member L (CLEC2L), mRNA [NM_001080511] |
| A_24_P227415 | 0,020447036 | down | -1,5718691 | CLEC7A | Homo sapiens C-type lectin domain family 7, member A (CLEC7A), transcript variant 6, mRNA [NM_197954] |
| A_23_P321984 | 0,026650889 | down | -1,8684766 | CLECL1 | Homo sapiens C-type lectin-like 1 (CLECL1), transcript variant 1, mRNA [NM_172004] |
| A_23_P311895 | 0,015129729 | down | -1,6527369 | CLIC5 | Homo sapiens chloride intracellular channel 5 (CLIC5), transcript variant 2, mRNA [NM_016929] |
| A_23_P364567 | 0,036356688 | down | -2,1880867 | CLRN1 | Homo sapiens clarin 1 (CLRN1), transcript variant 4, mRNA [NM_052995] |
| A_33_P3236077 | 0,018208487 | down | -2,8857257 | CLRN2 | Homo sapiens clarin 2 (CLRN2), mRNA [NM_001079827] |
| A_23_P106042 | 0,029507997 | down | -2,0352478 | CMTM5 | Homo sapiens CKLF-like MARVEL transmembrane domain containing 5 (CMTM5), transcript variant 3, mRNA [NM_001037288] |
| A_33_P3334615 | 0,022438606 | down | -1,8690815 | CNGA2 | Homo sapiens cyclic nucleotide gated channel alpha 2 (CNGA2), mRNA [NM_005140] |
| A_33_P3220347 | 0,014361652 | down | -1,5842836 | CNPY1 | Homo sapiens canopy FGF signaling regulator 1 (CNPY1), mRNA [NM_001103176] |
| A_23_P310931 | 0,043720756 | down | -2,2412853 | CNR2 | Homo sapiens cannabinoid receptor 2 (macrophage) (CNR2), mRNA [NM_001841] |
| A_24_P257478 | 0,025207188 | down | -2,3886561 | COL25A1 | Homo sapiens collagen, type XXV, alpha 1 (COL25A1), transcript variant 1, mRNA [NM_198721] |
| A_23_P134627 | 0,009714678 | down | -2,6834006 | COL26A1 | Homo sapiens collagen, type XXVI, alpha 1 (COL26A1), transcript variant 2, mRNA [NM_133457] |
| A_23_P500464 | 0,02084132 | down | -1,5156122 | COL2A1 | Homo sapiens collagen, type II, alpha 1 (COL2A1), transcript variant 1, mRNA [NM_001844] |
| A_33_P3298830 | 0,04802027 | down | -1,5998613 | COL4A2-AS2 | PREDICTED: Homo sapiens COL4A2 antisense RNA 2 (COL4A2-AS2), misc_RNA [XR_158875] |
| A_33_P3268734 | 0,013409594 | down | -1,889081 | COL5A1-AS1 | CR748640 Soares_NFL_T_GBC_S1 Homo sapiens cDNA clone IMAGp971L0898 ; IMAGE:2348699 5', mRNA sequence [CR74864 |
| A_33_P3363470 | 0,044834632 | down | -1,859182 | COL6A4P1 | collagen, type VI, alpha 4 pseudogene 1 [Source:HGNC Symbol;Acc:HGNC:33484] [ENST00000508169] |
| A_23_P404698 | 0,008921796 | down | -1,8799448 | COL6A5 | Homo sapiens collagen, type VI, alpha 5 (COL6A5), transcript variant 2, mRNA [NM_153264] |
| A_33_P3367748 | 0,047925796 | down | -1,6672007 | COL9A2 | collagen, type IX, alpha 2 [Source:HGNC Symbol;Acc:HGNC:2218] [ENST00000461118] |
| A_23_P258164 | 0,002234575 | down | -1,7419977 | CORT | Homo sapiens cortistatin (CORT), mRNA [NM_001302] |
| A_33_P3279610 | 0,006845136 | down | -2,68844 | COX6A2 | Homo sapiens cytochrome c oxidase subunit VIa polypeptide 2 (COX6A2), mRNA [NM_005205] |

| | | | | | |
|---|---|---|---|---|---|
| A_23_P31161 | 0,021397408 | down | -2,2954264 | CPA2 | Homo sapiens carboxypeptidase A2 (pancreatic) (CPA2), mRNA [NM_001869] |
| A_33_P3572454 | 0,0469599 | down | -1,8450403 | CPEB1-AS1 | Homo sapiens CPEB1 antisense RNA 1 (CPEB1-AS1), long non-coding RNA [NR_046096] |
| A_24_P187799 | 0,011223841 | down | -1,6574252 | CPED1 | Homo sapiens cadherin-like and PC-esterase domain containing 1 (CPED1), transcript variant 1, mRNA [NM_024913] |
| A_33_P3288135 | 0,03557338 | down | -1,6077269 | CPLX2 | complexin 2 [Source:HGNC Symbol;Acc:HGNC:2310] [ENST00000506642] |
| A_33_P3415300 | 3,22E-04 | down | -1,8253644 | CPLX2 | Homo sapiens complexin 2 (CPLX2), transcript variant 1, mRNA [NM_006650] |
| A_23_P16694 | 0,001492748 | down | -2,3275268 | CPS1-IT1 | Homo sapiens CPS1 intronic transcript 1 (non-protein coding) (CPS1-IT1), long non-coding RNA [NR_002763] |
| A_23_P6066 | 0,005217733 | down | -2,4568622 | CPXM1 | Homo sapiens carboxypeptidase X (M14 family), member 1 (CPXM1), transcript variant 1, mRNA [NM_019609] |
| A_33_P3314151 | 5,65E-04 | down | -2,5952857 | CRACR2A | Homo sapiens calcium release activated channel regulator 2A (CRACR2A), transcript variant 1, mRNA [NM_001144958] |
| A_32_P384562 | 0,040731587 | up | 1,8812089 | CROCCP3 | Homo sapiens ciliary rootlet coiled-coil, rootletin pseudogene 3 (CROCCP3), non-coding RNA [NR_023386] |
| A_33_P3317670 | 0,03860851 | down | -1,6800821 | CRP | Homo sapiens C-reactive protein, pentraxin-related (CRP), mRNA [NM_000567] |
| A_23_P132147 | 0,02914792 | down | -1,5891044 | CRYAA | Homo sapiens crystallin, alpha A (CRYAA), mRNA [NM_000394] |
| A_24_P11061 | 0,015850281 | down | -2,6343973 | CSAG1 | Homo sapiens chondrosarcoma associated gene 1 (CSAG1), transcript variant a, mRNA [NM_153478] |
| A_33_P3341442 | 0,023133043 | down | -2,068512 | CSF2RB | Homo sapiens colony stimulating factor 2 receptor, beta, low-affinity (granulocyte-macrophage) (CSF2RB), mRNA [NM_000395] |
| A_24_P263284 | 0,023453342 | down | -1,5171199 | CSMD2 | Homo sapiens CUB and Sushi multiple domains 2 (CSMD2), transcript variant 2, mRNA [NM_052896] |
| A_21_P0011331 | 0,028115246 | down | -2,1181018 | CSPG4 | Homo sapiens chondroitin sulfate proteoglycan 4 (CSPG4), mRNA [NM_001897] |
| A_33_P3397670 | 0,016647432 | down | -1,6904874 | CSPG4P1Y | Homo sapiens chondroitin sulfate proteoglycan 4 pseudogene 1, Y-linked (CSPG4P1Y), non-coding RNA [NR_001554] |
| A_33_P3344046 | 0,04674656 | down | -1,6149155 | CST13P | Homo sapiens cystatin 13, pseudogene (CST13P), non-coding RNA [NR_001279] |
| A_23_P68601 | 0,007800755 | down | -1,7359333 | CST7 | Homo sapiens cystatin F (leukocystatin) (CST7), mRNA [NM_003650] |
| A_23_P96633 | 0,007769989 | down | -3,34114 | CT55 | Homo sapiens cancer/testis antigen 55 (CT55), transcript variant 1, mRNA [NM_001031705] |
| A_33_P3318343 | 0,014935905 | down | -3,2943294 | CTAG2 | Homo sapiens cancer/testis antigen 2 (CTAG2), transcript variant 2, mRNA [NM_020994] |
| A_19_P00317835 | 0,030327663 | down | -1,6346669 | CTD-3080P12.3 | Homo sapiens uncharacterized LOC101928857 (CTD-3080P12.3), long non-coding RNA [NR_109911] |
| A_33_P3302191 | 0,028284203 | down | -1,5007458 | CTNNA3 | Homo sapiens catenin (cadherin-associated protein), alpha 3 (CTNNA3), transcript variant 1, mRNA [NM_013266] |
| A_23_P431139 | 0,012986339 | down | -1,637884 | CTRB1 | Homo sapiens chymotrypsinogen B1 (CTRB1), mRNA [NM_001906] |
| A_33_P3712341 | 0,029308213 | down | -1,9806979 | CXCL12 | Homo sapiens chemokine (C-X-C motif) ligand 12 (CXCL12), transcript variant 3, mRNA [NM_001033886] |
| A_23_P109913 | 0,026102223 | down | -1,5543842 | CXCR6 | Homo sapiens chemokine (C-X-C motif) receptor 6 (CXCR6), mRNA [NM_006564] |
| A_23_P62227 | 0,00877738 | down | -1,9773695 | CXorf21 | Homo sapiens chromosome X open reading frame 21 (CXorf21), mRNA [NM_025159] |
| A_33_P3224100 | 0,026299298 | down | -1,5186739 | CXorf22 | Homo sapiens chromosome X open reading frame 22 (CXorf22), mRNA [NM_152632] |
| A_33_P3256063 | 0,009198226 | down | -1,9772589 | CXorf30 | chromosome X open reading frame 30 [Source:HGNC Symbol;Acc:HGNC:27298] [ENST00000378653] |
| A_33_P3250308 | 0,007533446 | down | -1,6791451 | CXorf38 | chromosome X open reading frame 38 [Source:HGNC Symbol;Acc:HGNC:28589] [ENST00000378421] |
| A_33_P3302275 | 0,024101624 | down | -1,7963018 | CXorf66 | Homo sapiens chromosome X open reading frame 66 (CXorf66), mRNA [NM_001013403] |
| A_24_P400457 | 0,0206838 | down | -1,834865 | CYB5RL | Homo sapiens cytochrome b5 reductase-like (CYB5RL), mRNA [NM_001031672] |
| A_23_P129169 | 0,041386057 | down | -1,5027083 | CYP11A1 | Homo sapiens cytochrome P450, family 11, subfamily A, polypeptide 1 (CYP11A1), transcript variant 1, mRNA [NM_000781] |
| A_23_P215997 | 0,002778934 | down | -1,977991 | CYP11B2 | Homo sapiens cytochrome P450, family 11, subfamily B, polypeptide 2 (CYP11B2), mRNA [NM_000498] |
| A_32_P86289 | 0,024997668 | down | -2,0580502 | CYP19A1 | Homo sapiens cytochrome P450, family 19, subfamily A, polypeptide 1 (CYP19A1), transcript variant 2, mRNA [NM_031226] |
| A_33_P3294764 | 0,032925136 | down | -1,7042578 | CYP1B1-AS1 | Homo sapiens CYP1B1 antisense RNA 1 (CYP1B1-AS1), long non-coding RNA [NR_027252] |
| A_23_P210109 | 0,035814576 | down | -1,7915407 | CYP26B1 | Homo sapiens cytochrome P450, family 26, subfamily B, polypeptide 1 (CYP26B1), transcript variant 1, mRNA [NM_019885] |
| A_33_P3264667 | 0,033385508 | down | -2,4411025 | CYP26C1 | Homo sapiens cytochrome P450, family 26, subfamily C, polypeptide 1 (CYP26C1), mRNA [NM_183374] |
| A_23_P143734 | 0,008938405 | up | 1,5135046 | CYP2D6 | Homo sapiens cytochrome P450, family 2, subfamily D, polypeptide 6 (CYP2D6), transcript variant 1, mRNA [NM_000106] |
| A_23_P89981 | 0,002488869 | down | -2,5774465 | CYP2F1 | Homo sapiens cytochrome P450, family 2, subfamily F, polypeptide 1 (CYP2F1), mRNA [NM_000774] |
| A_33_P3225562 | 0,036734223 | up | 1,9440111 | CYP2G1P | Homo sapiens cytochrome P450, family 2, subfamily G, polypeptide 1 pseudogene (CYP2G1P), non-coding RNA [NR_040249] |
| A_33_P3251342 | 0,016320314 | down | -2,0000348 | CYP3A4 | Homo sapiens cytochrome P450, family 3, subfamily A, polypeptide 4 (CYP3A4), transcript variant 1, mRNA [NM_017460] |
| A_33_P3318117 | 0,039689284 | down | -1,8858395 | CYP3A7 | Homo sapiens cytochrome P450, family 3, subfamily A, polypeptide 7 (CYP3A7), mRNA [NM_000765] |
| A_33_P3382471 | 0,003357379 | down | -2,6018145 | CYP4A22 | Homo sapiens cytochrome P450, family 4, subfamily A, polypeptide 22 (CYP4A22), mRNA [NM_001010969] |
| A_21_P0010756 | 0,022373417 | down | -1,8354546 | CYP4Z1 | Homo sapiens cytochrome P450, family 4, subfamily Z, polypeptide 1 (CYP4Z1), mRNA [NM_178134] |
| A_33_P3762733 | 0,006513609 | down | -1,5170356 | D21S2088E | Homo sapiens D21S2088E (D21S2088E), long non-coding RNA [NR_040254] |
| A_33_P3288104 | 0,020269053 | down | -2,5339923 | DAAM2 | dishevelled associated activator of morphogenesis 2 [Source:HGNC Symbol;Acc:HGNC:18143] [ENST00000405961] |
| A_24_P203134 | 0,025951901 | down | -1,6571937 | DCAF12L1 | Homo sapiens DDB1 and CUL4 associated factor 12-like 1 (DCAF12L1), mRNA [NM_178470] |
| A_33_P3309999 | 0,04150654 | down | -1,8166949 | DCAF12L2 | Homo sapiens DDB1 and CUL4 associated factor 12-like 2 (DCAF12L2), mRNA [NM_001013628] |
| A_33_P3250740 | 0,036202684 | down | -1,8845214 | DCAF8L1 | Homo sapiens DDB1 and CUL4 associated factor 8-like 1 (DCAF8L1), mRNA [NM_001017930] |

| | | | | | |
|---|---|---|---|---|---|
| A_33_P3291748 | 0,02801496 | down | -1,7237383 | DCDC5 | Homo sapiens doublecortin domain containing 5 (DCDC5), mRNA [NM_020869] |
| A_33_P3298236 | 0,04807997 | down | -1,5764133 | DCHS2 | Homo sapiens dachsous cadherin-related 2 (DCHS2), transcript variant 2, mRNA [NM_001142552] |
| A_33_P3302777 | 0,049691927 | down | -2,275277 | DCLK1 | Homo sapiens doublecortin-like kinase 1 (DCLK1), transcript variant 1, mRNA [NM_004734] |
| A_23_P314798 | 0,005699699 | down | -2,054663 | DCST2 | Homo sapiens DC-STAMP domain containing 2 (DCST2), mRNA [NM_144622] |
| A_33_P3406686 | 2,43E-04 | down | -2,0665302 | DCTN1 | Homo sapiens dynactin 1 (DCTN1), transcript variant 7, non-coding RNA [NR_033935] |
| A_23_P2317 | 0,014728752 | down | -1,9737356 | DDN | Homo sapiens dendrin (DDN), mRNA [NM_015086] |
| A_33_P3284621 | 0,032165885 | down | -1,7418137 | DDR2 | discoidin domain receptor tyrosine kinase 2 [Source:HGNC Symbol;Acc:HGNC:2731] [ENST00000367922] |
| A_33_P3322307 | 0,03985162 | up | 1,6604378 | DDX11 | Homo sapiens DEAD/H (Asp-Glu-Ala-Asp/His) box helicase 11 (DDX11), transcript variant 5, mRNA [NM_001257145] |
| A_33_P3766913 | 0,013309958 | down | -1,6925575 | DEFA10P | Homo sapiens defensin, alpha 10 pseudogene (DEFA10P), non-coding RNA [NR_029386] |
| A_24_P363711 | 0,008533821 | down | -1,5578359 | DEFA6 | Homo sapiens defensin, alpha 6, Paneth cell-specific (DEFA6), mRNA [NM_001926] |
| A_33_P3358815 | 0,02766882 | down | -1,5146904 | DEFB103A | Homo sapiens defensin, beta 103A (DEFB103A), mRNA [NM_001081551] |
| A_23_P9018 | 0,03737975 | down | -1,6921226 | DEFB104B | Homo sapiens defensin, beta 104B (DEFB104B), mRNA [NM_001040702] |
| A_23_P425814 | 0,001431951 | down | -2,1869211 | DEFB106B | Homo sapiens defensin, beta 106B (DEFB106B), mRNA [NM_001040704] |
| A_33_P3370454 | 0,029209057 | down | -1,6868416 | DEFB108B | Homo sapiens defensin, beta 108B (DEFB108B), mRNA [NM_001002035] |
| A_33_P3400552 | 0,014533272 | down | -2,0616431 | DEFB108B | Homo sapiens defensin, beta 108B (DEFB108B), mRNA [NM_001002035] |
| A_33_P3272274 | 0,03854113 | down | -1,5066607 | DEFB110 | Homo sapiens defensin, beta 110 locus (DEFB110), transcript variant 2, mRNA [NM_001037728] |
| A_33_P3354201 | 0,008366687 | down | -1,9586437 | DEFB114 | Homo sapiens defensin, beta 114 (DEFB114), mRNA [NM_001037499] |
| A_33_P3362331 | 0,033105385 | up | 1,5374974 | DEFB130 | Homo sapiens defensin, beta 130 (DEFB130), mRNA [NM_001037804] |
| A_33_P3397545 | 0,012391587 | down | -1,6570518 | DEFB135 | Homo sapiens defensin, beta 135 (DEFB135), mRNA [NM_001033017] |
| A_23_P320124 | 0,027572913 | down | -1,6056577 | DEFT1P | Homo sapiens defensin, theta 1 pseudogene (DEFT1P), non-coding RNA [NR_036686] |
| A_33_P3312686 | 0,037015967 | down | -1,5057763 | DGAT2L6 | Homo sapiens diacylglycerol O-acyltransferase 2-like 6 (DGAT2L6), mRNA [NM_198512] |
| A_33_P3310164 | 0,04859753 | down | -1,7201254 | DGKB | Homo sapiens diacylglycerol kinase, beta 90kDa (DGKB), transcript variant 1, mRNA [NM_004080] |
| A_33_P3310172 | 0,007656447 | down | -3,0667126 | DGKB | Homo sapiens diacylglycerol kinase, beta 90kDa (DGKB), transcript variant 2, mRNA [NM_145695] |
| A_21_P0000776 | 0,03796289 | down | -1,7554238 | DIO2-AS1 | Homo sapiens DIO2 antisense RNA 1 (DIO2-AS1), long non-coding RNA [NR_038355] |
| A_33_P3246513 | 0,023300625 | down | -1,5247034 | DIO3 | Homo sapiens deiodinase, iodothyronine, type III (DIO3), mRNA [NM_001362] |
| A_33_P3400903 | 0,04463355 | down | -1,8349433 | DISC2 | Homo sapiens disrupted in schizophrenia 2 (non-protein coding) (DISC2), long non-coding RNA [NR_002227] |
| A_32_P327679 | 0,012974867 | down | -2,1272712 | DKFZP434K028 | Homo sapiens uncharacterized LOC26070 (DKFZP434K028), long non-coding RNA [NR_026882] |
| A_33_P3328782 | 0,031162087 | down | -1,568641 | DKFZP434L187 | Homo sapiens uncharacterized LOC26082 (DKFZP434L187), long non-coding RNA [NR_026771] |
| A_33_P3852239 | 0,003500327 | down | -2,5923839 | DKFZp686K1684 | Homo sapiens uncharacterized LOC440034 (DKFZp686K1684), long non-coding RNA [NR_033971] |
| A_33_P3419032 | 0,01332148 | down | -1,7779032 | DLG2 | Homo sapiens cDNA FLJ45207 fis, clone BRCAN2010665, highly similar to Channel associated protein of synapse-110. [AK127150] |
| A_23_P392317 | 0,007740615 | down | -1,8077958 | DLGAP2 | Homo sapiens discs, large (Drosophila) homolog-associated protein 2 (DLGAP2), transcript variant 1, mRNA [NM_004745] |
| A_33_P3297813 | 0,029551955 | down | -1,6679462 | DMD | Homo sapiens dystrophin, mRNA (cDNA clone IMAGE:5274415), complete cds. [BC036103] |
| A_23_P54612 | 0,041041426 | down | -1,6262472 | DNAAF1 | Homo sapiens dynein, axonemal, assembly factor 1 (DNAAF1), mRNA [NM_178452] |
| A_33_P3250338 | 0,04739361 | down | -1,6457335 | DNAH10 | dynein, axonemal, heavy chain 10 [Source:HGNC Symbol;Acc:HGNC:2941] [ENST00000614082] |
| A_33_P3415124 | 0,025411185 | down | -1,6654321 | DNAH10OS | Homo sapiens cDNA FLJ45278 fis, clone BRHIP3001141. [AK127211] |
| A_21_P0011612 | 0,038796496 | down | -1,6803192 | DNAH17 | Homo sapiens dynein, axonemal, heavy chain 17 (DNAH17), mRNA [NM_173628] |
| A_21_P0011613 | 0,01007361 | down | -1,7394402 | DNAH17 | Homo sapiens dynein, axonemal, heavy chain 17 (DNAH17), mRNA [NM_173628] |
| A_21_P0011629 | 0,005205224 | down | -3,5004113 | DNAH17-AS1 | Homo sapiens DNAH17 antisense RNA 1 (DNAH17-AS1), long non-coding RNA [NR_102401] |
| A_23_P351168 | 0,023482013 | down | -1,5141796 | DNAJB7 | Homo sapiens DnaJ (Hsp40) homolog, subfamily B, member 7 (DNAJB7), mRNA [NM_145174] |
| A_33_P3225066 | 0,018797975 | down | -1,7043399 | DNAJC12 | Homo sapiens DnaJ (Hsp40) homolog, subfamily C, member 12 (DNAJC12), transcript variant 2, mRNA [NM_201262] |
| A_24_P324368 | 0,02482619 | down | -1,5848008 | DNAJC14 | Homo sapiens DnaJ (Hsp40) homolog, subfamily C, member 14 (DNAJC14), mRNA [NM_032364] |
| A_33_P3389216 | 0,030128539 | down | -2,2425702 | DNM3OS | Homo sapiens DNM3 opposite strand/antisense RNA (DNM3OS), transcript variant 1, long non-coding RNA [NR_038397] |
| A_23_P17673 | 0,007485096 | down | -1,655853 | DNMT3L | Homo sapiens DNA (cytosine-5-)-methyltransferase 3-like (DNMT3L), transcript variant 1, mRNA [NM_013369] |
| A_21_P0000604 | 0,0333518 | down | -1,6873538 | DOC2GP | Homo sapiens double C2-like domains, gamma, pseudogene (DOC2GP), non-coding RNA [NR_033791] |
| A_24_P263653 | 0,0242788 | up | 1,5383873 | DOCK2 | Homo sapiens dedicator of cytokinesis 2 (DOCK2), mRNA [NM_004946] |
| A_21_P0014196 | 0,011244023 | down | -1,5759659 | DOCK4-AS1 | Homo sapiens DOCK4 antisense RNA 1 (DOCK4-AS1), long non-coding RNA [NR_103806] |
| A_23_P152262 | 0,026854055 | down | -2,005611 | DPEP1 | Homo sapiens dipeptidase 1 (renal) (DPEP1), transcript variant 1, mRNA [NM_004413] |
| A_23_P129413 | 0,04406817 | down | -1,8766406 | DPEP3 | Homo sapiens dipeptidase 3 (DPEP3), transcript variant 1, mRNA [NM_022357] |
| A_33_P3211356 | 0,024918947 | down | -1,8567276 | DPP10 | Homo sapiens dipeptidyl-peptidase 10 (non-functional) (DPP10), transcript variant 4, mRNA [NM_001178037] |

| | | | | | |
|---|---|---|---|---|---|
| A_33_P3423556 | 0,0450772 | down | -1,6422315 | DPP6 | dipeptidyl-peptidase 6 [Source:HGNC Symbol;Acc:HGNC:3010] [ENST00000462622] |
| A_33_P3402963 | 0,011867359 | down | -2,3022385 | DPRX | Homo sapiens divergent-paired related homeobox (DPRX), mRNA [NM_001012728] |
| A_23_P210224 | 0,03756614 | down | -1,6590161 | DPYSL5 | Homo sapiens dihydropyrimidinase-like 5 (DPYSL5), transcript variant 1, mRNA [NM_020134] |
| A_23_P401691 | 0,016863316 | down | -1,7007902 | DPYSL5 | Homo sapiens dihydropyrimidinase-like 5 (DPYSL5), transcript variant 1, mRNA [NM_020134] |
| A_33_P3230841 | 0,005046258 | down | -2,2503648 | DUPD1 | Homo sapiens dual specificity phosphatase and pro isomerase domain containing 1 (DUPD1), mRNA [NM_001003892] |
| A_21_P0014945 | 0,0229177 | down | -1,6244951 | DUX4 | PREDICTED: Homo sapiens double homeobox 4-like (DUX4L), partial ncRNA [XR_431831] |
| A_33_P3308586 | 0,012136263 | down | -2,7378514 | DUXA | Homo sapiens double homeobox A (DUXA), mRNA [NM_001012729] |
| A_23_P360990 | 0,013973192 | down | -1,7379928 | DYDC1 | Homo sapiens DPY30 domain containing 1 (DYDC1), transcript variant 1, mRNA [NM_138812] |
| A_23_P391396 | 0,014558342 | down | -1,7974476 | EBF3 | Homo sapiens early B-cell factor 3 (EBF3), mRNA [NM_001005463] |
| A_33_P3401701 | 0,04848501 | down | -1,7141898 | EDNRA | Homo sapiens endothelin receptor type A (EDNRA), transcript variant 1, mRNA [NM_001957] |
| A_33_P3785051 | 0,014296101 | down | -1,5280104 | EFCAB10 | Homo sapiens cDNA clone IMAGE:6616931, partial cds. [BC062748] |
| A_33_P3341399 | 0,028450297 | down | -2,275723 | EFCAB5 | Homo sapiens EF-hand calcium binding domain 5 (EFCAB5), transcript variant 1, mRNA [NM_198529] |
| A_33_P3242878 | 0,003934616 | down | -2,8881428 | EFCAB6 | Homo sapiens EF-hand calcium binding domain 6 (EFCAB6), transcript variant 1, mRNA [NM_022785] |
| A_33_P3289596 | 0,034285527 | down | -1,7455719 | EFR3B | Homo sapiens EFR3 homolog B (S. cerevisiae) (EFR3B), mRNA [NM_014971] |
| A_21_P0012412 | 0,04386375 | down | -1,8561323 | EGFEM1P | Homo sapiens EGF-like and EMI domain containing 1, pseudogene (EGFEM1P), non-coding RNA [NR_021485] |
| A_33_P3367577 | 0,025148116 | down | -1,6013718 | EIF4ENIF1 | eukaryotic translation initiation factor 4E nuclear import factor 1 [Source:HGNC Symbol;Acc:HGNC:16687] [ENST00000397520] |
| A_33_P3265504 | 0,008006132 | down | -1,5530111 | EIF5AL1 | Homo sapiens eukaryotic translation initiation factor 5A-like 1 (EIF5AL1), mRNA [NM_001099692] |
| A_33_P3235204 | 0,013914954 | down | -1,718016 | ELMOD3 | Homo sapiens ELMO/CED-12 domain containing 3 (ELMOD3), transcript variant 1, mRNA [NM_032213] |
| A_23_P215454 | 0,04263823 | down | -2,2999966 | ELN | Homo sapiens elastin (ELN), transcript variant 13, mRNA [NM_001278939] |
| A_23_P382065 | 0,036078747 | down | -1,7763911 | EMCN | Homo sapiens endomucin (EMCN), transcript variant 1, mRNA [NM_016242] |
| A_24_P52887 | 0,017854396 | down | -1,5313406 | ENDOU | Homo sapiens endonuclease, polyU-specific (ENDOU), transcript variant 2, mRNA [NM_006025] |
| A_33_P3417626 | 0,047544826 | down | -2,0249577 | ENHO | Homo sapiens energy homeostasis associated (ENHO), mRNA [NM_198573] |
| A_33_P3411338 | 0,015863022 | down | -1,6056409 | ENPEP | Homo sapiens glutamyl aminopeptidase (aminopeptidase A) (ENPEP), mRNA [NM_001977] |
| A_23_P410115 | 0,015176682 | down | -2,5132942 | ENTHD1 | Homo sapiens ENTH domain containing 1 (ENTHD1), mRNA [NM_152512] |
| A_33_P3218975 | 0,042596944 | down | -1,5564153 | ENTPD1 | Homo sapiens ectonucleoside triphosphate diphosphohydrolase 1 (ENTPD1), transcript variant 1, mRNA [NM_001776] |
| A_33_P3218980 | 0,008420228 | down | -1,7830722 | ENTPD1 | Homo sapiens ectonucleoside triphosphate diphosphohydrolase 1 (ENTPD1), transcript variant 1, mRNA [NM_001776] |
| A_24_P97374 | 0,03289334 | down | -1,5794563 | EOMES | Homo sapiens eomesodermin (EOMES), transcript variant 2, mRNA [NM_005442] |
| A_23_P385199 | 0,002411813 | down | -1,9690264 | EPHA10 | Homo sapiens EPH receptor A10 (EPHA10), transcript variant 2, mRNA [NM_173641] |
| A_23_P18342 | 0,03890482 | down | -1,6254228 | EPHA6 | Homo sapiens EPH receptor A6 (EPHA6), transcript variant 3, mRNA [NM_001278300] |
| A_33_P3408782 | 0,045393586 | down | -1,774031 | EPHA6 | Homo sapiens EPH receptor A6 (EPHA6), transcript variant 1, mRNA [NM_001080448] |
| A_33_P3317168 | 9,59E-05 | down | -2,4836824 | EPS8L3 | Homo sapiens EPS8-like 3 (EPS8L3), transcript variant 1, mRNA [NM_139053] |
| A_32_P183765 | 0,04826975 | down | -2,016431 | ERBB4 | Homo sapiens erb-b2 receptor tyrosine kinase 4 (ERBB4), transcript variant JM-a/CVT-1, mRNA [NM_005235] |
| A_33_P3527931 | 0,006620627 | down | -1,7783923 | ERICH4 | Homo sapiens glutamate-rich 4 (ERICH4), mRNA [NM_001130514] |
| A_33_P3250165 | 0,02719581 | down | -1,6521742 | ERMN | ermin, ERM-like protein [Source:HGNC Symbol;Acc:HGNC:29208] [ENST00000411762] |
| A_23_P144843 | 0,048472755 | down | -1,6798735 | ESM1 | Homo sapiens endothelial cell-specific molecule 1 (ESM1), transcript variant 1, mRNA [NM_007036] |
| A_24_P383478 | 0,02022964 | down | -1,5490509 | ESR1 | Homo sapiens estrogen receptor 1 (ESR1), transcript variant 1, mRNA [NM_000125] |
| A_33_P3347911 | 3,01E-04 | down | -3,872853 | ESR2 | Homo sapiens estrogen receptor 2 (ER beta) (ESR2), transcript variant g, mRNA [NM_001271877] |
| A_23_P391857 | 1,56E-04 | down | -3,0335839 | ESRRB | Homo sapiens estrogen-related receptor beta (ESRRB), mRNA [NM_004452] |
| A_24_P7965 | 0,01767049 | down | -3,0306885 | ESRRG | Homo sapiens estrogen-related receptor gamma (ESRRG), transcript variant 2, mRNA [NM_206594] |
| A_33_P3295098 | 0,04833084 | up | 1,7559806 | ESYT3 | extended synaptotagmin-like protein 3 [Source:HGNC Symbol;Acc:HGNC:24295] [ENST00000289135] |
| A_33_P3356325 | 2,89E-04 | down | -1,8960326 | ETV6 | Homo sapiens ets variant 6 (ETV6), mRNA [NM_001987] |
| A_19_P00322662 | 0,032146204 | up | 1,9007689 | EWSAT1 | Homo sapiens Ewing sarcoma associated transcript 1 (EWSAT1), long non-coding RNA [NR_026949] |
| A_23_P415820 | 0,014562849 | down | -1,5625918 | EXOC6B | Homo sapiens mRNA for KIAA0919 protein, partial cds. [AB023136] |
| A_24_P944973 | 0,035306223 | down | -1,667021 | EYS | Homo sapiens eyes shut homolog (Drosophila) (EYS), transcript variant 3, mRNA [NM_198283] |
| A_23_P41314 | 0,001322001 | down | -2,6187067 | F11 | Homo sapiens coagulation factor XI (F11), mRNA [NM_000128] |
| A_32_P60065 | 0,044208974 | down | -1,7265391 | F2RL2 | Homo sapiens coagulation factor II (thrombin) receptor-like 2 (F2RL2), transcript variant 1, mRNA [NM_004101] |
| A_32_P41604 | 0,031596623 | down | -1,6880432 | F5 | Homo sapiens coagulation factor V (proaccelerin, labile factor) (F5), mRNA [NM_000130] |
| A_23_P79562 | 0,029691687 | down | -1,843774 | FABP1 | Homo sapiens fatty acid binding protein 1, liver (FABP1), mRNA [NM_001443] |
| A_24_P237927 | 0,04237508 | down | -1,956004 | FAM102B | Homo sapiens family with sequence similarity 102, member B (FAM102B), mRNA [NM_001010883] |

| | | | | | |
|---|---|---|---|---|---|
| A_21_P0000066 | 0,023593895 | down | -1,8376143 | FAM104B | Homo sapiens family with sequence similarity 104, member B (FAM104B), transcript variant 6, mRNA [NM_001166703] |
| A_19_P00319160 | 0,009505627 | down | -1,7981164 | FAM107A | Homo sapiens family with sequence similarity 107, member A (FAM107A), transcript variant 4, mRNA [NM_001282714] |
| A_23_P345799 | 0,042048596 | down | -2,1316698 | FAM129C | Homo sapiens family with sequence similarity 129, member C (FAM129C), transcript variant 1, mRNA [NM_173544] |
| A_33_P3350259 | 0,003618136 | down | -2,302685 | FAM129C | Homo sapiens family with sequence similarity 129, member C (FAM129C), transcript variant 2, mRNA [NM_001098524] |
| A_33_P3270668 | 0,016839528 | down | -1,5188427 | FAM131B | Homo sapiens family with sequence similarity 131, member B (FAM131B), transcript variant a, mRNA [NM_001031690] |
| A_32_P923011 | 3,34E-04 | down | -2,1873875 | FAM138A | Homo sapiens family with sequence similarity 138, member A (FAM138A), long non-coding RNA [NR_026818] |
| A_33_P3222501 | 0,01594983 | up | 1,5802441 | FAM157A | Homo sapiens family with sequence similarity 157, member A (FAM157A), mRNA [NM_001145248] |
| A_33_P3295786 | 6,02E-04 | down | -2,5948565 | FAM159A | Homo sapiens family with sequence similarity 159, member A (FAM159A), mRNA [NM_001042693] |
| A_33_P3212849 | 0,028809644 | down | -1,5776689 | FAM170B-AS1 | Homo sapiens FAM170B antisense RNA 1 (FAM170B-AS1), long non-coding RNA [NR_038973] |
| A_33_P3354076 | 0,036654413 | down | -1,827489 | FAM172BP | Homo sapiens family with sequence similarity 172, member B, pseudogene (FAM172BP), non-coding RNA [NR_036433] |
| A_21_P0008784 | 0,004297308 | down | -1,8934579 | FAM174B | family with sequence similarity 174, member B [Source:HGNC Symbol;Acc:HGNC:34339] [ENST00000557545] |
| A_24_P231546 | 0,017320273 | down | -1,5601337 | FAM178B | Homo sapiens family with sequence similarity 178, member B (FAM178B), transcript variant B, mRNA [NM_016490] |
| A_33_P3267435 | 0,020403026 | down | -1,7600558 | FAM183CP | Homo sapiens family with sequence similarity 183, member C, pseudogene (FAM183CP), long non-coding RNA [NR_024473] |
| A_24_P298716 | 0,02700291 | down | -2,2583587 | FAM197Y2 | Homo sapiens family with sequence similarity 197, Y-linked, member 2, pseudogene (FAM197Y2), non-coding RNA [NR_001553] |
| A_23_P304489 | 0,01741108 | down | -1,7784556 | FAM19A5 | Homo sapiens family with sequence similarity 19 (chemokine (C-C motif)-like), member A5 (FAM19A5), transcript variant 2, mRNA [N |
| A_33_P3374180 | 0,013498561 | down | -2,2330728 | FAM209B | PREDICTED: Homo sapiens family with sequence similarity 209, member B (FAM209B), transcript variant X1, mRNA [XM_00526041 |
| A_33_P3346635 | 0,02594708 | down | -1,7511193 | FAM20C | Homo sapiens cDNA FLJ43291 fis, clone MESAN2015515. [AK125281] |
| A_24_P751492 | 0,043571632 | down | -1,8563595 | FAM221B | Homo sapiens family with sequence similarity 221, member B (FAM221B), transcript variant 1, mRNA [NM_001012446] |
| A_33_P3354111 | 0,012318901 | down | -1,5293885 | FAM227B | Homo sapiens family with sequence similarity 227, member B (FAM227B), mRNA [NM_152647] |
| A_33_P3361758 | 0,029240668 | down | -2,503681 | FAM227B | Homo sapiens family with sequence similarity 227, member B (FAM227B), mRNA [NM_152647] |
| A_33_P3340762 | 0,027402744 | down | -2,1253355 | FAM230A | PREDICTED: Homo sapiens family with sequence similarity 230, member A (FAM230A), mRNA [XM_006726832] |
| A_33_P3348794 | 0,01017505 | down | -1,7918137 | FAM231A | Homo sapiens family with sequence similarity 231, member A (FAM231A), mRNA [NM_001282321] |
| A_33_P3270104 | 0,021062791 | down | -1,9156722 | FAM26D | Homo sapiens family with sequence similarity 26, member D (FAM26D), transcript variant 1, mRNA [NM_001256887] |
| A_33_P3764636 | 0,03276288 | down | -1,7257786 | FAM41AY1 | Homo sapiens family with sequence similarity 41, member A, Y-linked 1 (FAM41AY1), long non-coding RNA [NR_028083] |
| A_32_P426044 | 0,014757811 | down | -2,9857864 | FAM47A | Homo sapiens family with sequence similarity 47, member A (FAM47A), mRNA [NM_203408] |
| A_33_P3249160 | 0,001634686 | down | -2,3019555 | FAM47C | Homo sapiens family with sequence similarity 47, member C (FAM47C), mRNA [NM_001013736] |
| A_23_P163711 | 0,033805527 | down | -1,5389714 | FAM57B | Homo sapiens family with sequence similarity 57, member B (FAM57B), mRNA [NM_031478] |
| A_23_P360354 | 0,027717715 | down | -1,5071845 | FAM81B | Homo sapiens family with sequence similarity 81, member B (FAM81B), mRNA [NM_152548] |
| A_33_P3212630 | 0,003910977 | down | -1,7625624 | FAM90A1 | Homo sapiens family with sequence similarity 90, member A1 (FAM90A1), mRNA [NM_018088] |
| A_33_P3230822 | 0,006938295 | down | -1,6147572 | FAM90A27P | family with sequence similarity 90, member A27, pseudogene [Source:HGNC Symbol;Acc:HGNC:43617] [ENST00000593323] |
| A_33_P3378777 | 0,019280918 | down | -1,8016951 | FAM99B | Homo sapiens family with sequence similarity 99, member B (non-protein coding) (FAM99B), long non-coding RNA [NR_026642] |
| A_32_P203029 | 0,011705165 | up | 2,05842 | FAM9B | Homo sapiens family with sequence similarity 9, member B (FAM9B), mRNA [NM_205849] |
| A_21_P0012091 | 0,033953514 | down | -1,6005809 | FAR2P2 | Homo sapiens fatty acyl CoA reductase 2 pseudogene 2 (FAR2P2), transcript variant 1, long non-coding RNA [NR_046258] |
| A_21_P0011966 | 0,03946446 | down | -2,303114 | FAR2P2 | Homo sapiens fatty acyl CoA reductase 2 pseudogene 2 (FAR2P2), transcript variant 1, long non-coding RNA [NR_046258] |
| A_21_P0014038 | 0,037732787 | down | -1,8651702 | FBRSL1 | Homo sapiens fibrosin-like 1 (FBRSL1), mRNA [NM_001142641] |
| A_24_P7600 | 0,016325645 | down | -1,7963687 | FBXL7 | Homo sapiens F-box and leucine-rich repeat protein 7 (FBXL7), transcript variant 1, mRNA [NM_012304] |
| A_23_P395911 | 7,70E-04 | down | -1,9733268 | FBXO17 | F-box protein 17 [Source:HGNC Symbol;Acc:HGNC:18754] [ENST00000601394] |
| A_24_P280684 | 0,04529091 | down | -1,6275228 | FBXO40 | Homo sapiens F-box protein 40 (FBXO40), mRNA [NM_016298] |
| A_33_P3415475 | 0,007016006 | down | -1,7912946 | FBXO47 | Homo sapiens F-box protein 47 (FBXO47), mRNA [NM_001008777] |
| A_33_P3382606 | 0,031009002 | down | -3,421359 | FBXW12 | Homo sapiens F-box and WD repeat domain containing 12 (FBXW12), transcript variant 1, mRNA [NM_207102] |
| A_21_P0010561 | 0,031492133 | down | -1,549648 | FCGR1B | Homo sapiens Fc fragment of IgG, high affinity Ib, receptor (CD64) (FCGR1B), transcript variant 3, mRNA [NM_001244910] |
| A_23_P200728 | 0,009134237 | down | -2,0077603 | FCGR3A | Homo sapiens Fc fragment of IgG, low affinity IIIa, receptor (CD16a) (FCGR3A), transcript variant 1, mRNA [NM_000569] |
| A_23_P157875 | 0,02370182 | down | -2,793071 | FCN1 | Homo sapiens ficolin (collagen/fibrinogen domain containing) 1 (FCN1), mRNA [NM_002003] |
| A_24_P319647 | 0,006348877 | down | -2,4273787 | FCRL2 | Homo sapiens Fc receptor-like 2 (FCRL2), transcript variant 1, mRNA [NM_030764] |
| A_23_P115200 | 0,013404521 | down | -2,1123626 | FCRL4 | Homo sapiens Fc receptor-like 4 (FCRL4), mRNA [NM_031282] |
| A_33_P3335506 | 0,02240652 | down | -1,5982019 | FCRL5 | Fc receptor-like 5 [Source:HGNC Symbol;Acc:HGNC:18508] [ENST00000368190] |
| A_33_P3335511 | 0,021325678 | down | -1,8054705 | FCRL5 | Homo sapiens Fc receptor-like 5 (FCRL5), transcript variant 2, mRNA [NM_001195388] |
| A_24_P276576 | 0,041088227 | down | -1,506987 | FCRLA | Homo sapiens Fc receptor-like A (FCRLA), transcript variant 2, mRNA [NM_032738] |
| A_19_P00318142 | 0,03127272 | down | -1,6595783 | FENDRR | Homo sapiens FOXF1 adjacent non-coding developmental regulatory RNA (FENDRR), transcript variant 2, long non-coding RNA [NF |

| | | | | | |
|---|---|---|---|---|---|
| A_23_P422849 | 0,004525175 | down | -3,2448816 | FERD3L | Homo sapiens Fer3-like bHLH transcription factor (FERD3L), mRNA [NM_152898] |
| A_33_P3337574 | 0,00257823 | down | -3,0048683 | FGF14-IT1 | Homo sapiens FGF14 intronic transcript 1 (non-protein coding) (FGF14-IT1), long non-coding RNA [NR_036486] |
| A_23_P427587 | 0,04863393 | down | -1,5415174 | FGF19 | Homo sapiens fibroblast growth factor 19 (FGF19), mRNA [NM_005117] |
| A_33_P3400248 | 0,031653225 | down | -1,5195261 | FGF20 | Homo sapiens fibroblast growth factor 20 (FGF20), mRNA [NM_019851] |
| A_23_P153878 | 0,03816743 | down | -1,7560352 | FGF22 | Homo sapiens fibroblast growth factor 22 (FGF22), transcript variant 1, mRNA [NM_020637] |
| A_33_P3348719 | 0,017074728 | down | -1,8904246 | FGF9 | Homo sapiens fibroblast growth factor 9 (FGF9), mRNA [NM_002010] |
| A_24_P226069 | 0,012855742 | down | -2,200535 | FGFBP2 | Homo sapiens fibroblast growth factor binding protein 2 (FGFBP2), mRNA [NM_031950] |
| A_23_P92754 | 0,001855567 | down | -1,6196762 | FGFR4 | Homo sapiens fibroblast growth factor receptor 4 (FGFR4), transcript variant 3, mRNA [NM_213647] |
| A_33_P3244585 | 0,010611074 | down | -1,7724673 | FHAD1 | Homo sapiens forkhead-associated (FHA) phosphopeptide binding domain 1 (FHAD1), mRNA [NM_052929] |
| A_33_P3849275 | 0,008364818 | down | -1,5197634 | FHL1 | Homo sapiens four and a half LIM domains 1 (FHL1), transcript variant 4, mRNA [NM_001159704] |
| A_33_P3347099 | 0,04606654 | down | -2,0981262 | FLJ36000 | Homo sapiens uncharacterized FLJ36000 (FLJ36000), long non-coding RNA [NR_027084] |
| A_33_P3249893 | 0,017608857 | down | -1,6668963 | FLJ37786 | Homo sapiens cDNA FLJ37786 fis, clone BRHIP2028480. [AK095105] |
| A_33_P3347437 | 0,00698914 | down | -2,2816217 | FLJ39080 | Homo sapiens uncharacterized LOC441355 (FLJ39080), long non-coding RNA [NR_033830] |
| A_33_P3282291 | 0,004138898 | down | -2,082982 | FLJ40039 | Homo sapiens cDNA FLJ40039 fis, clone SYNOV2000397. [AK097358] |
| A_32_P764462 | 0,009408258 | down | -1,7255485 | FLJ40536 | Homo sapiens cDNA FLJ40536 fis, clone TESTI2047930. [AK097855] |
| A_33_P3294861 | 0,010472008 | down | -1,5649143 | FLJ40712 | Homo sapiens cDNA FLJ40712 fis, clone THYMU2027249. [AK098031] |
| A_33_P3259112 | 0,039675087 | down | -1,8512163 | FLJ41278 | Homo sapiens uncharacterized LOC400046 (FLJ41278), long non-coding RNA [NR_033988] |
| A_24_P390668 | 0,045774035 | up | 1,7456115 | FMNL1 | Homo sapiens formin-like 1 (FMNL1), mRNA [NM_005892] |
| A_33_P3382303 | 0,007252289 | up | 1,5629487 | FMNL1 | Homo sapiens formin-like 1 (FMNL1), mRNA [NM_005892] |
| A_33_P3247559 | 0,041324694 | down | -1,6715589 | FNDC7 | Homo sapiens fibronectin type III domain containing 7 (FNDC7), mRNA [NM_001144937] |
| A_33_P3264331 | 0,03634445 | down | -1,5333498 | FOXD3 | Homo sapiens forkhead box D3 (FOXD3), mRNA [NM_012183] |
| A_23_P118254 | 0,038415223 | down | -1,5392998 | FOXF1 | Homo sapiens forkhead box F1 (FOXF1), mRNA [NM_001451] |
| A_23_P151150 | 0,023757206 | down | -1,749514 | FOXM1 | Homo sapiens forkhead box M1 (FOXM1), transcript variant 1, mRNA [NM_202002] |
| A_23_P159709 | 0,047998704 | down | -1,7458783 | FOXP3 | Homo sapiens forkhead box P3 (FOXP3), transcript variant 1, mRNA [NM_014009] |
| A_33_P3412798 | 0,013932908 | down | -1,6052852 | FRG2B | Homo sapiens FSHD region gene 2 family, member B (FRG2B), mRNA [NM_001080998] |
| A_33_P3361831 | 0,015000658 | down | -1,6056765 | FRG2C | Homo sapiens FSHD region gene 2 family, member C (FRG2C), mRNA [NM_001124759] |
| A_23_P135132 | 0,032041963 | down | -1,6696563 | FRMD3 | Homo sapiens FERM domain containing 3 (FRMD3), transcript variant 1, mRNA [NM_174938] |
| A_33_P3296772 | 0,022907246 | up | 1,8994278 | FRMD6-AS1 | Homo sapiens FRMD6 antisense RNA 1 (FRMD6-AS1), long non-coding RNA [NR_037676] |
| A_33_P3603812 | 0,026724193 | down | -1,7006004 | FRMD8 | Homo sapiens FERM domain containing 8 (FRMD8), transcript variant 3, mRNA [NM_001300833] |
| A_24_P940275 | 0,01671353 | down | -2,124028 | FRMPD4 | Homo sapiens FERM and PDZ domain containing 4 (FRMPD4), mRNA [NM_014728] |
| A_33_P3223497 | 0,017560655 | down | -1,8986789 | FRY | Homo sapiens furry homolog (Drosophila) (FRY), mRNA [NM_023037] |
| A_21_P0014679 | 0,019822974 | down | -1,8882841 | FRY-AS1 | Homo sapiens FRY antisense RNA 1 (FRY-AS1), long non-coding RNA [NR_103839] |
| A_33_P3390918 | 0,019934693 | down | -2,867776 | FSD2 | Homo sapiens fibronectin type III and SPRY domain containing 2 (FSD2), transcript variant 1, mRNA [NM_001007122] |
| A_23_P385017 | 0,020011328 | down | -1,5888841 | G6PC | Homo sapiens glucose-6-phosphatase, catalytic subunit (G6PC), transcript variant 1, mRNA [NM_000151] |
| A_23_P93302 | 0,023259068 | up | 1,5119251 | GABBR1 | Homo sapiens gamma-aminobutyric acid (GABA) B receptor, 1 (GABBR1), transcript variant 1, mRNA [NM_001470] |
| A_23_P167121 | 0,021553127 | down | -1,5914401 | GABRA2 | Homo sapiens gamma-aminobutyric acid (GABA) A receptor, alpha 2 (GABRA2), transcript variant 1, mRNA [NM_000807] |
| A_23_P41847 | 0,030457659 | down | -1,6563475 | GABRA6 | Homo sapiens gamma-aminobutyric acid (GABA) A receptor, alpha 6 (GABRA6), mRNA [NM_000811] |
| A_33_P3336715 | 0,03190766 | down | -1,5230186 | GABRB2 | Homo sapiens gamma-aminobutyric acid (GABA) A receptor, beta 2 (GABRB2), transcript variant 1, mRNA [NM_021911] |
| A_33_P3409266 | 0,004422575 | down | -2,6119633 | GAFA2 | Homo sapiens FGF-2 activity-associated protein 2 (GAFA2) mRNA, complete cds. [AF220234] |
| A_24_P943370 | 0,042670358 | down | -1,6299446 | GAGE1 | Homo sapiens G antigen 1 (GAGE1), transcript variant 2, mRNA [NM_001040663] |
| A_24_P212539 | 0,043055326 | down | -1,903442 | GALM | Homo sapiens galactose mutarotase (aldose 1-epimerase) (GALM), mRNA [NM_138801] |
| A_33_P3239387 | 0,043270163 | down | -1,8653852 | GALNT13 | Homo sapiens polypeptide N-acetylgalactosaminyltransferase 13 (GALNT13), transcript variant 2, mRNA [NM_001301627] |
| A_23_P76749 | 0,012300794 | down | -2,224294 | GALNT16 | Homo sapiens polypeptide N-acetylgalactosaminyltransferase 16 (GALNT16), transcript variant 2, mRNA [NM_020692] |
| A_33_P3320017 | 0,03129499 | down | -1,5777336 | GALNTL6 | Homo sapiens polypeptide N-acetylgalactosaminyltransferase-like 6 (GALNTL6), mRNA [NM_001034845] |
| A_33_P3320022 | 0,042286485 | down | -1,6095455 | GALNTL6 | Homo sapiens polypeptide N-acetylgalactosaminyltransferase-like 6 (GALNTL6), mRNA [NM_001034845] |
| A_23_P153155 | 0,021664225 | down | -1,6480209 | GALR1 | Homo sapiens galanin receptor 1 (GALR1), mRNA [NM_001480] |
| A_23_P253446 | 0,025692504 | down | -1,5552802 | GAP43 | Homo sapiens growth associated protein 43 (GAP43), transcript variant 2, mRNA [NM_002045] |
| A_23_P67646 | 0,03465304 | down | -1,5277807 | GAPDHS | Homo sapiens glyceraldehyde-3-phosphate dehydrogenase, spermatogenic (GAPDHS), mRNA [NM_014364] |
| A_24_P136522 | 0,001867754 | down | -2,1369407 | GARNL3 | Homo sapiens cDNA FLJ38360 fis, clone FEBRA2000462, weakly similar to RAP1 GTPASE ACTIVATING PROTEIN 1. [AK095679] |

| | | | | | |
|---|---|---|---|---|---|
| A_32_P24140 | 0,024756469 | down | -1,8699211 | GAS2 | Homo sapiens growth arrest-specific 2 (GAS2), transcript variant 1, mRNA [NM_005256] |
| A_33_P3266192 | 0,035987016 | down | -1,6881009 | GAS6-AS1 | Homo sapiens hypothetical LOC650669, mRNA (cDNA clone MGC:169000 IMAGE:9021377), complete cds. [BC137379] |
| A_24_P374244 | 0,022576097 | down | -1,8060682 | GATA1 | Homo sapiens GATA binding protein 1 (globin transcription factor 1) (GATA1), mRNA [NM_002049] |
| A_33_P3276329 | 0,001178022 | down | -2,329487 | GATSL2 | Homo sapiens GATS protein-like 2 (GATSL2), mRNA [NM_001145064] |
| A_24_P280664 | 0,018959697 | down | -1,543666 | GBP7 | Homo sapiens guanylate binding protein 7 (GBP7), mRNA [NM_207398] |
| A_33_P3423969 | 0,029979857 | down | -2,2224796 | GBX2 | Homo sapiens gastrulation brain homeobox 2 (GBX2), transcript variant 2, mRNA [NM_001301687] |
| A_23_P119886 | 0,011360985 | up | 2,6298027 | GCKR | Homo sapiens glucokinase (hexokinase 4) regulator (GCKR), mRNA [NM_001486] |
| A_24_P49896 | 0,00807113 | down | -3,2250502 | GDPD4 | Homo sapiens glycerophosphodiester phosphodiesterase domain containing 4 (GDPD4), mRNA [NM_182833] |
| A_33_P3335895 | 0,02800477 | down | -1,8848543 | GFAP | Homo sapiens glial fibrillary acidic protein (GFAP), transcript variant 3, mRNA [NM_001242376] |
| A_23_P8497 | 0,033993203 | up | 1,8711636 | GHRHR | Homo sapiens growth hormone releasing hormone receptor (GHRHR), mRNA [NM_000823] |
| A_23_P427023 | 0,01127144 | down | -1,7846636 | GIMAP1 | Homo sapiens GTPase, IMAP family member 1 (GIMAP1), mRNA [NM_130759] |
| A_24_P22943 | 0,027324371 | down | -1,803808 | GIPC3 | Homo sapiens GIPC PDZ domain containing family, member 3 (GIPC3), mRNA [NM_133261] |
| A_23_P416666 | 0,03871483 | down | -1,7628706 | GJA10 | Homo sapiens gap junction protein, alpha 10, 62kDa (GJA10), mRNA [NM_032602] |
| A_32_P205241 | 1,61E-04 | down | -3,7297597 | GJA3 | Homo sapiens gap junction protein, alpha 3, 46kDa (GJA3), mRNA [NM_021954] |
| A_23_P44436 | 0,04893549 | down | -2,1812942 | GKN1 | Homo sapiens gastrokine 1 (GKN1), mRNA [NM_019617] |
| A_23_P61317 | 0,023280963 | down | -1,5414157 | GKN2 | Homo sapiens gastrokine 2 (GKN2), mRNA [NM_182536] |
| A_23_P209246 | 0,032072127 | up | 2,425932 | GLI2 | Homo sapiens GLI family zinc finger 2 (GLI2), mRNA [NM_005270] |
| A_23_P317839 | 0,04206719 | down | -1,5144175 | GLIPR1L1 | Homo sapiens GLI pathogenesis-related 1 like 1 (GLIPR1L1), mRNA [NM_152779] |
| A_23_P425324 | 0,040394273 | down | -2,0531516 | GMCL1P1 | Homo sapiens germ cell-less, spermatogenesis associated 1 pseudogene 1 (GMCL1P1), non-coding RNA [NR_003281] |
| A_33_P3409939 | 0,013645762 | down | -1,76237 | GNAT3 | Homo sapiens guanine nucleotide binding protein, alpha transducing 3 (GNAT3), mRNA [NM_001102386] |
| A_33_P3284004 | 4,19E-04 | down | -2,2367973 | GNN | Homo sapiens Grp94 neighboring nucleotidase pseudogene (GNN), non-coding RNA [NR_027249] |
| A_33_P3271460 | 0,001068325 | down | -2,3771665 | GOLGA1 | golgin A1 [Source:HGNC Symbol;Acc:HGNC:4424] [ENST00000373551] |
| A_21_P0011355 | 0,006758985 | down | -1,6644135 | GOLGA8K | Homo sapiens golgin A8 family, member K (GOLGA8K), mRNA [NM_001282493] |
| A_23_P9075 | 0,0267155 | down | -1,5356686 | GOT1L1 | Homo sapiens glutamic-oxaloacetic transaminase 1-like 1 (GOT1L1), mRNA [NM_152413] |
| A_24_P944964 | 0,01974662 | down | -2,1319976 | GP5 | Homo sapiens glycoprotein V (platelet) (GP5), mRNA [NM_004488] |
| A_24_P319374 | 0,012251008 | down | -1,620831 | GPA33 | Homo sapiens glycoprotein A33 (transmembrane) (GPA33), mRNA [NM_005814] |
| A_23_P68240 | 4,87E-04 | down | -2,0198312 | GPAT2 | Homo sapiens glycerol-3-phosphate acyltransferase 2, mitochondrial (GPAT2), mRNA [NM_207328] |
| A_23_P336678 | 0,001395157 | down | -1,8374106 | GPHB5 | Homo sapiens glycoprotein hormone beta 5 (GPHB5), mRNA [NM_145171] |
| A_33_P3343250 | 0,001750255 | down | -2,2864676 | GPR1 | Homo sapiens G protein-coupled receptor 1 (GPR1), transcript variant 4, mRNA [NM_001261453] |
| A_23_P415706 | 0,03804463 | down | -1,501283 | GPR133 | Homo sapiens G protein-coupled receptor 133 (GPR133), mRNA [NM_198827] |
| A_23_P6943 | 0,046938103 | down | -2,3869176 | GPR15 | Homo sapiens G protein-coupled receptor 15 (GPR15), mRNA [NM_005290] |
| A_33_P3332576 | 0,006585927 | down | -2,063662 | GPR151 | Homo sapiens G protein-coupled receptor 151 (GPR151), mRNA [NM_194251] |
| A_33_P3236813 | 0,036663827 | down | -1,6711134 | GPR19 | Homo sapiens G protein-coupled receptor 19 (GPR19), mRNA [NM_006143] |
| A_23_P434289 | 0,010999681 | down | -1,8822633 | GPR62 | Homo sapiens G protein-coupled receptor 62 (GPR62), mRNA [NM_080865] |
| A_23_P214727 | 0,03161659 | down | -1,8933461 | GPR63 | Homo sapiens G protein-coupled receptor 63 (GPR63), transcript variant 2, mRNA [NM_030784] |
| A_23_P81683 | 0,005961971 | down | -2,900122 | GPRC6A | Homo sapiens G protein-coupled receptor, class C, group 6, member A (GPRC6A), transcript variant 1, mRNA [NM_148963] |
| A_33_P3288832 | 0,036066066 | down | -1,7577938 | GPRIN1 | Homo sapiens G protein regulated inducer of neurite outgrowth 1 (GPRIN1), mRNA [NM_052899] |
| A_23_P49643 | 0,021874988 | down | -1,7441924 | GRAP | Homo sapiens GRB2-related adaptor protein (GRAP), mRNA [NM_006613] |
| A_23_P329768 | 0,027018417 | down | -1,6921754 | GREB1 | Homo sapiens growth regulation by estrogen in breast cancer 1 (GREB1), transcript variant a, mRNA [NM_014668] |
| A_24_P229025 | 0,031088475 | down | -2,0338626 | GRIA3 | Homo sapiens glutamate receptor, ionotropic, AMPA 3 (GRIA3), transcript variant 3, mRNA [NM_001256743] |
| A_24_P941896 | 0,011640321 | down | -1,7608724 | GRID1 | Homo sapiens glutamate receptor, ionotropic, delta 1 (GRID1), mRNA [NM_017551] |
| A_23_P257962 | 0,018429821 | down | -1,8486652 | GRIN1 | Homo sapiens glutamate receptor, ionotropic, N-methyl D-aspartate 1 (GRIN1), transcript variant GluN1-1a, mRNA [NM_007327] |
| A_32_P169114 | 0,03321869 | down | -1,513263 | GRIN2A | Homo sapiens glutamate receptor, ionotropic, N-methyl D-aspartate 2A (GRIN2A), transcript variant 1, mRNA [NM_001134407] |
| A_24_P381844 | 0,009016948 | down | -1,9644018 | GRIN2A | Homo sapiens glutamate receptor, ionotropic, N-methyl D-aspartate 2A (GRIN2A), transcript variant 2, mRNA [NM_000833] |
| A_23_P151264 | 0,02073521 | down | -1,7591082 | GRIN2B | Homo sapiens glutamate receptor, ionotropic, N-methyl D-aspartate 2B (GRIN2B), mRNA [NM_000834] |
| A_23_P347541 | 0,00443829 | down | -2,5857582 | GRIN3A | Homo sapiens glutamate receptor, ionotropic, N-methyl-D-aspartate 3A (GRIN3A), mRNA [NM_133445] |
| A_33_P3279276 | 0,042579386 | down | -2,21045 | GRIP2 | Homo sapiens glutamate receptor interacting protein 2 (GRIP2), mRNA [NM_001080423] |
| A_23_P313542 | 0,03216801 | down | -2,166786 | GRK1 | Homo sapiens G protein-coupled receptor kinase 1 (GRK1), mRNA [NM_002929] |
| A_33_P3240867 | 0,01873725 | down | -2,384537 | GRK1 | Homo sapiens G protein-coupled receptor kinase 1 (GRK1), mRNA [NM_002929] |

| | | | | | |
|---|---|---|---|---|---|
| A_33_P3358943 | 0,015442268 | up | 1,8035592 | GRM2 | Homo sapiens glutamate receptor, metabotropic 2 (GRM2), transcript variant 1, mRNA [NM_000839] |
| A_21_P0012432 | 0,043566234 | down | -1,5768262 | GRM7-AS3 | GRM7 antisense RNA 3 [Source:HGNC Symbol;Acc:HGNC:42444] [ENST00000455623] |
| A_33_P3224867 | 0,002138784 | down | -1,9573493 | GSG1 | Homo sapiens germ cell associated 1 (GSG1), transcript variant 5, mRNA [NM_001206842] |
| A_32_P58872 | 0,039238624 | down | -2,7026865 | GSG1L | Homo sapiens GSG1-like (GSG1L), transcript variant 1, mRNA [NM_001109763] |
| A_23_P97606 | 0,010904098 | down | -1,9429742 | GSTM5 | Homo sapiens glutathione S-transferase mu 5 (GSTM5), mRNA [NM_000851] |
| A_21_P0012351 | 0,023995766 | down | -1,6061655 | GSTTP1 | Homo sapiens glutathione S-transferase theta pseudogene 1 (GSTTP1), non-coding RNA [NR_003081] |
| A_23_P29163 | 0,012940131 | down | -1,6909525 | GSTTP1 | Homo sapiens glutathione S-transferase theta pseudogene 1 (GSTTP1), non-coding RNA [NR_003081] |
| A_23_P92453 | 0,023681872 | down | -1,5070332 | GSX2 | Homo sapiens GS homeobox 2 (GSX2), mRNA [NM_133267] |
| A_23_P57020 | 0,004884807 | down | -1,8387595 | GTSF1L | Homo sapiens gametocyte specific factor 1-like (GTSF1L), transcript variant 2, mRNA [NM_001008901] |
| A_23_P350001 | 0,033693388 | down | -1,6945769 | GUCY1A2 | Homo sapiens guanylate cyclase 1, soluble, alpha 2 (GUCY1A2), transcript variant 2, mRNA [NM_000855] |
| A_33_P3253687 | 0,03129755 | down | -1,6448917 | GVINP1 | Homo sapiens GTPase, very large interferon inducible pseudogene 1 (GVINP1), non-coding RNA [NR_003945] |
| A_21_P0014007 | 0,045544066 | down | -2,184219 | GVQW1 | Homo sapiens cDNA FLJ25547 fis, clone JTH01487. [AK098413] |
| A_23_P130836 | 0,014197438 | down | -1,7214528 | GZMM | Homo sapiens granzyme M (lymphocyte met-ase 1) (GZMM), transcript variant 1, mRNA [NM_005317] |
| A_23_P26457 | 0,043562904 | down | -1,7985305 | HBA2 | Homo sapiens hemoglobin, alpha 2 (HBA2), mRNA [NM_000517] |
| A_23_P47665 | 0,022798194 | down | -1,9191141 | HBE1 | Homo sapiens hemoglobin, epsilon 1 (HBE1), mRNA [NM_005330] |
| A_21_P0004638 | 0,016782567 | down | -1,5101095 | HCG14 | Homo sapiens HLA complex group 14 (non-protein coding) (HCG14), long non-coding RNA [NR_104117] |
| A_33_P3225418 | 0,003910848 | down | -2,7474208 | HCG9 | Homo sapiens HLA complex group 9 (non-protein coding) (HCG9), long non-coding RNA [NR_028032] |
| A_24_P365526 | 0,034141503 | down | -2,1688495 | HCK | Homo sapiens HCK proto-oncogene, Src family tyrosine kinase (HCK), transcript variant 1, mRNA [NM_002110] |
| A_33_P3290677 | 0,004864968 | down | -2,2523031 | HELT | Homo sapiens helt bHLH transcription factor (HELT), transcript variant 1, mRNA [NM_001300781] |
| A_24_P363408 | 0,01041187 | down | -1,946091 | HEY2 | Homo sapiens hes-related family bHLH transcription factor with YRPW motif 2 (HEY2), mRNA [NM_012259] |
| A_24_P289648 | 0,013605773 | down | -2,7294364 | HFE | Homo sapiens hemochromatosis (HFE), transcript variant 11, mRNA [NM_139011] |
| A_33_P3328883 | 0,022730727 | down | -1,5646136 | HGC6.3 | Homo sapiens uncharacterized LOC100128124 (HGC6.3), mRNA [NM_001129895] |
| A_33_P3267640 | 3,50E-04 | down | -1,8998362 | HGFAC | Homo sapiens HGF activator (HGFAC), transcript variant 1, mRNA [NM_001297439] |
| A_19_P00316839 | 0,021494688 | down | -1,566805 | HHLA1 | Homo sapiens HERV-H LTR-associating 1 (HHLA1), mRNA [NM_001145095] |
| A_19_P00321196 | 0,012742592 | down | -1,6750767 | HHLA1 | Homo sapiens HERV-H LTR-associating 1 (HHLA1), mRNA [NM_001145095] |
| A_24_P414658 | 0,026600687 | down | -1,784871 | HIST1H2AG | histone cluster 1, H2ag [Source:HGNC Symbol;Acc:HGNC:4737] [ENST00000359193] |
| A_33_P3400578 | 0,049615394 | down | -2,2382545 | HLF | Homo sapiens hepatic leukemia factor (HLF), mRNA [NM_002126] |
| A_23_P30204 | 0,031031884 | down | -1,7356155 | HMHB1 | Homo sapiens histocompatibility (minor) HB-1 (HMHB1), mRNA [NM_021182] |
| A_24_P314515 | 0,001820662 | down | -3,7560997 | HNF1A-AS1 | Homo sapiens HNF1A antisense RNA 1 (HNF1A-AS1), long non-coding RNA [NR_024345] |
| A_24_P330822 | 0,013213486 | down | -1,9828106 | HNF1B | HNF1 homeobox B [Source:HGNC Symbol;Acc:HGNC:11630] [ENST00000621123] |
| A_21_P0010501 | 0,032022167 | down | -1,7861295 | HOTAIR | BROAD Institute lincRNA (HOTAIR), lincRNA [TCONS_00079053_HOTAIR] |
| A_21_P0010502 | 0,029631104 | down | -1,9983487 | HOTAIR | Homo sapiens HOX transcript antisense RNA (HOTAIR), transcript variant 3, long non-coding RNA [NR_047518] |
| A_21_P0010497 | 0,024294227 | down | -2,1730926 | HOTTIP | Homo sapiens HOXA distal transcript antisense RNA (HOTTIP), long non-coding RNA [NR_037843] |
| A_33_P3645079 | 0,02658369 | down | -1,6168351 | HOXA-AS3 | Homo sapiens HOXA cluster antisense RNA 3 (HOXA-AS3), transcript variant 1, long non-coding RNA [NR_038831] |
| A_33_P3272957 | 0,007225943 | down | -1,6565276 | HOXA-AS3 | Homo sapiens HOXA cluster antisense RNA 3 (HOXA-AS3), transcript variant 1, long non-coding RNA [NR_038831] |
| A_33_P3303956 | 0,006335457 | down | -1,9405333 | HOXA-AS3 | Homo sapiens HOXA cluster antisense RNA 3 (HOXA-AS3), transcript variant 1, long non-coding RNA [NR_038831] |
| A_23_P93772 | 0,04898771 | down | -1,9598454 | HOXA5 | Homo sapiens homeobox A5 (HOXA5), mRNA [NM_019102] |
| A_23_P118675 | 0,031270694 | down | -1,7261444 | HOXB1 | Homo sapiens homeobox B1 (HOXB1), mRNA [NM_002144] |
| A_23_P370588 | 0,04663072 | down | -1,6035045 | HOXB8 | Homo sapiens homeobox B8 (HOXB8), mRNA [NM_024016] |
| A_24_P124558 | 0,02194761 | down | -1,5636178 | HOXC8 | Homo sapiens homeobox C8 (HOXC8), mRNA [NM_022658] |
| A_23_P143029 | 0,044551004 | down | -1,9668746 | HOXD11 | Homo sapiens homeobox D11 (HOXD11), mRNA [NM_021192] |
| A_23_P210164 | 0,020883225 | down | -1,9059907 | HOXD8 | Homo sapiens homeobox D8 (HOXD8), transcript variant 1, mRNA [NM_019558] |
| A_23_P406782 | 0,019799199 | down | -1,7807523 | HPN | Homo sapiens hepsin (HPN), transcript variant 1, mRNA [NM_182983] |
| A_23_P61707 | 0,01613162 | down | -2,0155325 | HPSE2 | Homo sapiens heparanase 2 (inactive) (HPSE2), transcript variant 1, mRNA [NM_021828] |
| A_23_P57658 | 0,03175189 | down | -1,854061 | HRASLS | Homo sapiens HRAS-like suppressor (HRASLS), mRNA [NM_020386] |
| A_23_P53018 | 5,47E-04 | down | -1,8466473 | HRASLS5 | Homo sapiens HRAS-like suppressor family, member 5 (HRASLS5), transcript variant 1, mRNA [NM_054108] |
| A_24_P226508 | 0,026348215 | down | -1,6322298 | HS3ST5 | Homo sapiens heparan sulfate (glucosamine) 3-O-sulfotransferase 5 (HS3ST5), mRNA [NM_153612] |
| A_23_P351295 | 0,038088456 | down | -1,7974523 | HS3ST5 | Homo sapiens heparan sulfate (glucosamine) 3-O-sulfotransferase 5 (HS3ST5), mRNA [NM_153612] |
| A_23_P25030 | 0,036976084 | down | -1,6988587 | HSD17B6 | Homo sapiens hydroxysteroid (17-beta) dehydrogenase 6 (HSD17B6), mRNA [NM_003725] |

| | | | | | |
|---|---|---|---|---|---|
| A_23_P51580 | 0,035305064 | down | -2,6408658 | HSD3B2 | Homo sapiens hydroxy-delta-5-steroid dehydrogenase, 3 beta- and steroid delta-isomerase 2 (HSD3B2), transcript variant 1, mRNA [ |
| A_33_P3380762 | 0,01645385 | down | -2,218521 | HSD52 | Homo sapiens uncharacterized LOC729467 (HSD52), long non-coding RNA [NR_027120] |
| A_23_P96497 | 0,010038016 | down | -2,2703576 | HSFX1 | Homo sapiens heat shock transcription factor family, X linked 1 (HSFX1), mRNA [NM_016153] |
| A_33_P3262660 | 0,024654528 | down | -1,5785424 | HSFY1P1 | Homo sapiens heat shock transcription factor, Y-linked 1 pseudogene 1 (HSFY1P1), non-coding RNA [NR_003607] |
| A_23_P62446 | 0,017856548 | down | -1,6148437 | HSFY2 | Homo sapiens heat shock transcription factor, Y linked 2 (HSFY2), transcript variant 2, mRNA [NM_001001877] |
| A_24_P308029 | 0,008991133 | down | -2,0571911 | HSPB6 | Homo sapiens heat shock protein, alpha-crystallin-related, B6 (HSPB6), mRNA [NM_144617] |
| A_24_P97687 | 0,019812126 | down | -2,5974538 | HTR1A | Homo sapiens 5-hydroxytryptamine (serotonin) receptor 1A, G protein-coupled (HTR1A), mRNA [NM_000524] |
| A_23_P156824 | 8,35E-06 | down | -3,658279 | HTR1B | Homo sapiens 5-hydroxytryptamine (serotonin) receptor 1B, G protein-coupled (HTR1B), mRNA [NM_000863] |
| A_33_P3406836 | 0,009645983 | down | -1,7121874 | HTR1F | Homo sapiens 5-hydroxytryptamine (serotonin) receptor 1F, G protein-coupled (HTR1F), mRNA [NM_000866] |
| A_23_P433586 | 0,033621114 | down | -1,6894854 | HTR2C | 5-hydroxytryptamine (serotonin) receptor 2C, G protein-coupled [Source:HGNC Symbol;Acc:HGNC:5295] [ENST00000276198] |
| A_33_P3345016 | 0,004005227 | down | -2,5235734 | HTR6 | Homo sapiens 5-hydroxytryptamine (serotonin) receptor 6, G protein-coupled (HTR6), mRNA [NM_000871] |
| A_33_P3402983 | 0,009742701 | down | -1,5278882 | HYDIN | Homo sapiens HYDIN, axonemal central pair apparatus protein (HYDIN), transcript variant 1, mRNA [NM_001270974] |
| A_33_P3276856 | 1,56E-05 | down | -2,86866 | HYDIN | Homo sapiens HYDIN, axonemal central pair apparatus protein (HYDIN), transcript variant 3, mRNA [NM_001198542] |
| A_33_P3343010 | 0,001765987 | down | -2,9950593 | HYPK | huntingtin interacting protein K [Source:HGNC Symbol;Acc:HGNC:18418] [ENST00000620261] |
| A_24_P281872 | 0,04829999 | down | -1,9377159 | HYPM | Homo sapiens huntingtin interacting protein M (HYPM), mRNA [NM_012274] |
| A_23_P144549 | 0,015709674 | down | -1,7027826 | IBSP | Homo sapiens integrin-binding sialoprotein (IBSP), mRNA [NM_004967] |
| A_24_P12690 | 0,04858266 | down | -1,765571 | IDO2 | Homo sapiens indoleamine 2,3-dioxygenase 2 (IDO2), mRNA [NM_194294] |
| A_23_P151294 | 0,021711312 | down | -1,5524135 | IFNG | Homo sapiens interferon, gamma (IFNG), mRNA [NM_000619] |
| A_33_P3234020 | 0,002116963 | down | -2,629176 | IGDCC3 | Homo sapiens immunoglobulin superfamily, DCC subclass, member 3 (IGDCC3), mRNA [NM_004884] |
| A_23_P116435 | 0,014167596 | down | -2,0407908 | IGF2-AS | Homo sapiens IGF2 antisense RNA (IGF2-AS), transcript variant 1, long non-coding RNA [NR_028044] |
| A_33_P3312819 | 0,007605148 | down | -2,091783 | IGF2-AS | Homo sapiens IGF2 antisense RNA (IGF2-AS), transcript variant 1, long non-coding RNA [NR_028044] |
| A_33_P3243486 | 0,021501858 | down | -1,9799732 | IGFL4 | Homo sapiens IGF-like family member 4 (IGFL4), mRNA [NM_001002923] |
| A_33_P3761915 | 0,035674144 | down | -1,8249285 | IGKV1D-13 | 602502772F1 NIH_MGC_77 Homo sapiens cDNA clone IMAGE:4616320 5', mRNA sequence [BG482625] |
| A_23_P333683 | 0,009766041 | down | -2,1946976 | IGSF10 | Homo sapiens immunoglobulin superfamily, member 10 (IGSF10), transcript variant 1, mRNA [NM_178822] |
| A_23_P376060 | 0,020424858 | down | -1,7723099 | IKZF3 | Homo sapiens IKAROS family zinc finger 3 (Aiolos) (IKZF3), transcript variant 1, mRNA [NM_012481] |
| A_23_P167479 | 0,037269745 | down | -1,8862203 | IL17B | Homo sapiens interleukin 17B (IL17B), mRNA [NM_014443] |
| A_23_P35092 | 0,002405184 | down | -1,8314921 | IL19 | Homo sapiens interleukin 19 (IL19), transcript variant 1, mRNA [NM_153758] |
| A_23_P56604 | 0,022398826 | down | -1,5287845 | IL1RL2 | Homo sapiens interleukin 1 receptor-like 2 (IL1RL2), mRNA [NM_003854] |
| A_33_P3422124 | 0,01851935 | down | -2,0862327 | IL22RA2 | Homo sapiens interleukin 22 receptor, alpha 2 (IL22RA2), transcript variant 3, mRNA [NM_181310] |
| A_33_P3295917 | 0,02525328 | down | -2,2710023 | IL7 | interleukin 7 [Source:HGNC Symbol;Acc:HGNC:6023] [ENST00000518982] |
| A_33_P3734378 | 0,037677806 | up | 1,6150934 | INHBA-AS1 | Homo sapiens INHBA antisense RNA 1 (INHBA-AS1), transcript variant 1, long non-coding RNA [NR_027118] |
| A_24_P155502 | 0,013609498 | down | -2,3382971 | INHBC | Homo sapiens inhibin, beta C (INHBC), mRNA [NM_005538] |
| A_23_P1981 | 0,004859632 | down | -2,212384 | INS | Homo sapiens insulin (INS), transcript variant 1, mRNA [NM_000207] |
| A_23_P112220 | 0,008226439 | down | -1,5063162 | INSL4 | Homo sapiens insulin-like 4 (placenta) (INSL4), mRNA [NM_002195] |
| A_21_P0014041 | 0,014197269 | up | 1,7122842 | IPO13 | PREDICTED: Homo sapiens importin 13 (IPO13), mRNA [XM_003846624] |
| A_32_P13756 | 0,010204908 | down | -1,6868024 | IQCF6 | Homo sapiens IQ motif containing F6 (IQCF6), mRNA [NM_001143833] |
| A_24_P275585 | 0,021433104 | down | -1,5442313 | IQCH | Homo sapiens IQ motif containing H (IQCH), transcript variant 1, mRNA [NM_001031715] |
| A_33_P3343120 | 0,047475763 | down | -1,6006099 | IRF8 | Homo sapiens interferon regulatory factor 8 (IRF8), mRNA [NM_002163] |
| A_23_P85039 | 0,006505657 | down | -1,6968335 | IRS4 | Homo sapiens insulin receptor substrate 4 (IRS4), mRNA [NM_003604] |
| A_23_P3312 | 0,005662673 | down | -2,3090434 | ISLR | Homo sapiens immunoglobulin superfamily containing leucine-rich repeat (ISLR), transcript variant 1, mRNA [NM_005545] |
| A_21_P0000848 | 0,041238997 | down | -1,615331 | ISPD-AS1 | Homo sapiens ISPD antisense RNA 1 (ISPD-AS1), transcript variant 1, long non-coding RNA [NR_038946] |
| A_33_P3316115 | 0,011325472 | down | -1,8391839 | ITFG1-AS1 | Homo sapiens ITFG1 antisense RNA 1 (ITFG1-AS1), transcript variant 1, long non-coding RNA [NR_110903] |
| A_23_P354151 | 0,032291017 | down | -1,636231 | ITK | Homo sapiens IL2-inducible T-cell kinase (ITK), mRNA [NM_005546] |
| A_33_P3239614 | 0,003455637 | down | -2,602169 | IYD | Homo sapiens iodotyrosine deiodinase (IYD), transcript variant 2, mRNA [NM_203395] |
| A_33_P3352088 | 0,020647027 | down | -2,0071373 | JAKMIP3 | Homo sapiens Janus kinase and microtubule interacting protein 3 (JAKMIP3), mRNA [NM_001105521] |
| A_33_P3249976 | 0,003087562 | down | -2,360399 | JAM2 | Homo sapiens junctional adhesion molecule 2 (JAM2), transcript variant 1, mRNA [NM_021219] |
| A_23_P68423 | 0,04000083 | down | -2,1109946 | JPH2 | Homo sapiens junctophilin 2 (JPH2), transcript variant 2, mRNA [NM_175913] |
| A_33_P3394252 | 0,002462881 | up | 2,1443913 | KANSL1L | Homo sapiens KAT8 regulatory NSL complex subunit 1-like (KANSL1L), mRNA [NM_152519] |
| A_21_P0014696 | 0,026084138 | down | -2,1703508 | KATNAL2 | PREDICTED: Homo sapiens katanin p60 subunit A-like 2 (KATNAL2), transcript variant X6, mRNA [XM_006722554] |

| | | | | | |
|---|---|---|---|---|---|
| A_33_P3396129 | 0,001627864 | down | -2,174469 | KATNB1 | katanin p80 (WD repeat containing) subunit B 1 [Source:HGNC Symbol;Acc:HGNC:6217] [ENST00000566611] |
| A_33_P3214061 | 0,007263868 | down | -1,6704192 | KATNBL1P6 | Homo sapiens katanin p80 subunit B-like 1 pseudogene 6 (KATNBL1P6), non-coding RNA [NR_003954] |
| A_23_P126528 | 0,02976564 | down | -2,3561606 | KCNA10 | Homo sapiens potassium channel, voltage gated shaker related subfamily A, member 10 (KCNA10), mRNA [NM_005549] |
| A_23_P417173 | 0,038633503 | down | -1,7254565 | KCNA5 | Homo sapiens potassium channel, voltage gated shaker related subfamily A, member 5 (KCNA5), mRNA [NM_002234] |
| A_33_P3415012 | 0,018916216 | down | -2,1078854 | KCNA6 | Homo sapiens potassium channel, voltage gated shaker related subfamily A, member 6 (KCNA6), mRNA [NM_002235] |
| A_24_P31627 | 0,018577732 | down | -1,5557013 | KCNB1 | Homo sapiens potassium channel, voltage gated Shab related subfamily B, member 1 (KCNB1), mRNA [NM_004975] |
| A_33_P3375766 | 0,019758724 | down | -1,9204632 | KCNC2 | Homo sapiens potassium channel, voltage gated Shaw related subfamily C, member 2 (KCNC2), transcript variant 3, mRNA [NM_15[illegible] |
| A_23_P24948 | 0,012424674 | down | -1,6533204 | KCNE3 | Homo sapiens potassium channel, voltage gated subfamily E regulatory beta subunit 3 (KCNE3), mRNA [NM_005472] |
| A_23_P256641 | 0,027132004 | down | -1,7334518 | KCNE5 | Homo sapiens potassium channel, voltage gated subfamily E regulatory beta subunit 5 (KCNE5), mRNA [NM_012282] |
| A_23_P168403 | 0,014747651 | down | -1,5590942 | KCNH2 | Homo sapiens potassium voltage-gated channel, subfamily H (eag-related), member 2 (KCNH2), transcript variant 1, mRNA [NM_00[illegible] |
| A_23_P377882 | 0,029400332 | down | -2,4876564 | KCNH2 | Homo sapiens potassium voltage-gated channel, subfamily H (eag-related), member 2 (KCNH2), transcript variant 2, mRNA [NM_17[illegible] |
| A_33_P3352906 | 0,011806937 | down | -2,4861965 | KCNIP4-IT1 | Homo sapiens KCNIP4 intronic transcript 1 (non-protein coding) (KCNIP4-IT1), long non-coding RNA [NR_002813] |
| A_23_P501193 | 0,036005966 | down | -1,5049622 | KCNJ16 | Homo sapiens potassium channel, inwardly rectifying subfamily J, member 16 (KCNJ16), transcript variant 2, mRNA [NM_170741] |
| A_23_P64879 | 0,04161025 | down | -1,5252799 | KCNJ8 | Homo sapiens potassium channel, inwardly rectifying subfamily J, member 8 (KCNJ8), mRNA [NM_004982] |
| A_23_P65629 | 0,030017061 | down | -1,5310405 | KCNK10 | Homo sapiens potassium channel, two pore domain subfamily K, member 10 (KCNK10), transcript variant 1, mRNA [NM_021161] |
| A_33_P3278789 | 0,042921234 | down | -2,1384544 | KCNK16 | Homo sapiens potassium channel, two pore domain subfamily K, member 16 (KCNK16), transcript variant 3, mRNA [NM_001135106 |
| A_23_P319423 | 0,001031155 | down | -3,493477 | KCNK5 | Homo sapiens potassium channel, two pore domain subfamily K, member 5 (KCNK5), mRNA [NM_003740] |
| A_23_P111978 | 0,026515633 | down | -2,6335762 | KCNK9 | Homo sapiens potassium channel, two pore domain subfamily K, member 9 (KCNK9), transcript variant 1, mRNA [NM_001282534] |
| A_21_P0012333 | 0,002524239 | down | -1,7794586 | KCNMB3 | Homo sapiens potassium channel subfamily M regulatory beta subunit 3 (KCNMB3), transcript variant 2, mRNA [NM_171829] |
| A_23_P34424 | 0,012569945 | down | -3,5274222 | KCNQ4 | Homo sapiens potassium channel, voltage gated KQT-like subfamily Q, member 4 (KCNQ4), transcript variant 1, mRNA [NM_004700 |
| A_33_P3243008 | 0,003794023 | down | -2,5622954 | KCNU1 | Homo sapiens potassium channel, subfamily U, member 1 (KCNU1), mRNA [NM_001031836] |
| A_23_P94902 | 0,020633692 | down | -1,8646433 | KCTD8 | Homo sapiens potassium channel tetramerization domain containing 8 (KCTD8), mRNA [NM_198353] |
| A_23_P386442 | 0,031378444 | down | -1,5431198 | KIAA0087 | Homo sapiens KIAA0087 (KIAA0087), long non-coding RNA [NR_022006] |
| A_21_P0012955 | 0,014261086 | down | -1,5745742 | KIAA1024L | Homo sapiens KIAA1024-like (KIAA1024L), mRNA [NM_001257308] |
| A_24_P600377 | 0,016548693 | down | -2,2791271 | KIAA1024L | Homo sapiens KIAA1024-like (KIAA1024L), mRNA [NM_001257308] |
| A_33_P3396746 | 0,014812454 | down | -1,6421765 | KIF26B | kinesin family member 26B [Source:HGNC Symbol;Acc:HGNC:25484] [ENST00000479506] |
| A_23_P218025 | 0,025891636 | down | -1,8120186 | KIF5A | Homo sapiens kinesin family member 5A (KIF5A), mRNA [NM_004984] |
| A_32_P154473 | 0,043823462 | down | -1,6224418 | KIF5C | Homo sapiens kinesin family member 5C (KIF5C), transcript variant 1, mRNA [NM_004522] |
| A_24_P297816 | 0,021040173 | down | -1,5233433 | KIF6 | Homo sapiens kinesin family member 6 (KIF6), transcript variant 1, mRNA [NM_145027] |
| A_24_P350622 | 0,021689557 | down | -2,5401216 | KIR2DL4 | Homo sapiens killer cell immunoglobulin-like receptor, two domains, long cytoplasmic tail, 4 (KIR2DL4), transcript variant 1, mRNA [[illegible] |
| A_24_P117147 | 0,005054997 | down | -2,8706326 | KIR3DL1 | Homo sapiens killer cell immunoglobulin-like receptor, three domains, long cytoplasmic tail, 1 (KIR3DL1), mRNA [NM_013289] |
| A_33_P3407034 | 0,025277419 | down | -1,8000169 | KIT | Homo sapiens v-kit Hardy-Zuckerman 4 feline sarcoma viral oncogene homolog (KIT), transcript variant 1, mRNA [NM_000222] |
| A_23_P139654 | 0,008360908 | down | -2,1316981 | KLRC1 | Homo sapiens killer cell lectin-like receptor subfamily C, member 1 (KLRC1), transcript variant 2, mRNA [NM_007328] |
| A_23_P218058 | 0,023808677 | down | -2,2714703 | KLRC4 | Homo sapiens killer cell lectin-like receptor subfamily C, member 4 (KLRC4), mRNA [NM_013431] |
| A_23_P66854 | 0,04591667 | down | -1,841054 | KRT20 | Homo sapiens keratin 20, type I (KRT20), mRNA [NM_019010] |
| A_21_P0000016 | 0,025162458 | down | -1,5859189 | KRT72 | Homo sapiens keratin 72, type II (KRT72), transcript variant 2, mRNA [NM_001146225] |
| A_33_P3249274 | 0,004156508 | down | -1,6956166 | KRT73-AS1 | Homo sapiens KRT73 antisense RNA 1 (KRT73-AS1), long non-coding RNA [NR_126005] |
| A_23_P130241 | 0,027745107 | down | -1,7036868 | KRTAP1-3 | Homo sapiens keratin associated protein 1-3 (KRTAP1-3), mRNA [NM_030966] |
| A_33_P3334877 | 0,014509118 | down | -1,8587502 | KRTAP10-4 | Homo sapiens keratin associated protein 10-4 (KRTAP10-4), mRNA [NM_198687] |
| A_33_P3293760 | 0,043526623 | down | -1,6800654 | KRTAP10-8 | Homo sapiens keratin associated protein 10-8 (KRTAP10-8), mRNA [NM_198695] |
| A_33_P3354399 | 0,007530211 | down | -2,2296572 | KRTAP12-3 | Homo sapiens keratin associated protein 12-3 (KRTAP12-3), mRNA [NM_198697] |
| A_24_P314534 | 0,006929359 | down | -2,5464656 | KRTAP13-2 | Homo sapiens keratin associated protein 13-2 (KRTAP13-2), mRNA [NM_181621] |
| A_24_P357386 | 0,02522473 | down | -1,9479643 | KRTAP13-3 | Homo sapiens keratin associated protein 13-3 (KRTAP13-3), mRNA [NM_181622] |
| A_24_P348845 | 0,034238644 | down | -1,5535786 | KRTAP15-1 | Homo sapiens keratin associated protein 15-1 (KRTAP15-1), mRNA [NM_181623] |
| A_33_P3311258 | 0,039156202 | down | -1,668654 | KRTAP19-4 | Homo sapiens keratin associated protein 19-4 (KRTAP19-4), mRNA [NM_181610] |
| A_33_P3263935 | 0,04657903 | down | -1,5338427 | KRTAP22-2 | Homo sapiens keratin associated protein 22-2 (KRTAP22-2), mRNA [NM_001164434] |
| A_33_P3635527 | 0,007063348 | down | -1,5939873 | KRTAP24-1 | Homo sapiens keratin associated protein 24-1 (KRTAP24-1), mRNA [NM_001085455] |
| A_33_P3306619 | 0,00388131 | down | -2,5663044 | KRTAP25-1 | Homo sapiens keratin associated protein 25-1 (KRTAP25-1), mRNA [NM_001128598] |
| A_33_P3382399 | 0,019608565 | down | -1,8537794 | KRTAP3-1 | Homo sapiens keratin associated protein 3-1 (KRTAP3-1), mRNA [NM_031958] |

| | | | | | |
|---|---|---|---|---|---|
| A_33_P3245348 | 0,042631235 | down | -1,6363318 | KRTAP4-1 | Homo sapiens keratin associated protein 4-1 (KRTAP4-1), mRNA [NM_033060] |
| A_23_P66844 | 0,030058116 | down | -1,5933099 | KRTAP4-7 | Homo sapiens keratin associated protein 4-7 (KRTAP4-7), mRNA [NM_033061] |
| A_33_P3304691 | 0,019832622 | down | -2,4583423 | KRTAP5-1 | Homo sapiens keratin associated protein 5-1 (KRTAP5-1), mRNA [NM_001005922] |
| A_33_P3229472 | 0,02375219 | down | -2,0219884 | KRTAP5-10 | Homo sapiens keratin associated protein 5-10 (KRTAP5-10), mRNA [NM_001012710] |
| A_24_P400130 | 0,032737307 | down | -1,6066586 | KRTAP7-1 | Homo sapiens keratin associated protein 7-1 (gene/pseudogene) (KRTAP7-1), mRNA [NM_181606] |
| A_33_P3404123 | 0,047075003 | down | -2,0333507 | KRTAP9-7 | Homo sapiens keratin associated protein 9-7 (KRTAP9-7), mRNA [NM_001277332] |
| A_33_P3292164 | 0,001767325 | down | -3,2989824 | KRTAP9-7 | Homo sapiens keratin associated protein 9-7 (KRTAP9-7), mRNA [NM_001277332] |
| A_32_P216635 | 0,025187433 | down | -1,8526232 | KSR2 | Homo sapiens kinase suppressor of ras 2 (KSR2), mRNA [NM_173598] |
| A_33_P3412493 | 0,01686806 | down | -1,6489053 | KY | kyphoscoliosis peptidase [Source:HGNC Symbol;Acc:HGNC:26576] [ENST00000506319] |
| A_33_P3355230 | 0,008185254 | down | -2,241749 | LAIR1 | Homo sapiens leukocyte-associated immunoglobulin-like receptor 1 (LAIR1), transcript variant a, mRNA [NM_002287] |
| A_23_P120227 | 0,03560222 | up | 1,7144676 | LBH | Homo sapiens limb bud and heart development (LBH), mRNA [NM_030915] |
| A_33_P3302125 | 0,046494123 | down | -2,247747 | LCE2A | Homo sapiens late cornified envelope 2A (LCE2A), mRNA [NM_178428] |
| A_24_P207828 | 0,008816589 | down | -2,2170775 | LCE2B | Homo sapiens late cornified envelope 2B (LCE2B), mRNA [NM_014357] |
| A_23_P32115 | 0,004413821 | down | -1,745348 | LCN12 | Homo sapiens lipocalin 12 (LCN12), mRNA [NM_178536] |
| A_33_P3357678 | 0,02369719 | down | -1,5485321 | LCTL | Homo sapiens lactase-like (LCTL), transcript variant 1, mRNA [NM_207338] |
| A_23_P125253 | 0,031528335 | down | -1,9757904 | LDB2 | LIM domain binding 2 [Source:HGNC Symbol;Acc:HGNC:6533] [ENST00000509803] |
| A_23_P54357 | 0,012811312 | down | -1,764326 | LDHAL6B | Homo sapiens lactate dehydrogenase A-like 6B (LDHAL6B), mRNA [NM_033195] |
| A_21_P0000886 | 0,029880458 | down | -2,7892067 | LDLRAD4-AS1 | Homo sapiens LDLRAD4 antisense RNA 1 (LDLRAD4-AS1), long non-coding RNA [NR_040031] |
| A_24_P213788 | 0,029186301 | down | -1,6411464 | LEF1-AS1 | Homo sapiens LEF1 antisense RNA 1 (LEF1-AS1), transcript variant 1, long non-coding RNA [NR_029373] |
| A_23_P5031 | 0,004856758 | down | -2,1486547 | LGALS13 | Homo sapiens lectin, galactoside-binding, soluble, 13 (LGALS13), mRNA [NM_013268] |
| A_23_P153662 | 0,023752682 | down | -2,0830572 | LGALS14 | Homo sapiens lectin, galactoside-binding, soluble, 14 (LGALS14), transcript variant 2, mRNA [NM_203471] |
| A_33_P3393655 | 0,01835152 | down | -2,0721805 | LGI2 | Homo sapiens leucine-rich repeat LGI family, member 2 (LGI2), mRNA [NM_018176] |
| A_23_P93169 | 0,022633133 | down | -1,5956634 | LGSN | Homo sapiens lengsin, lens protein with glutamine synthetase domain (LGSN), transcript variant 1, mRNA [NM_016571] |
| A_33_P3315959 | 0,015295078 | down | -1,6939301 | LGSN | Homo sapiens lengsin, lens protein with glutamine synthetase domain (LGSN), transcript variant 1, mRNA [NM_016571] |
| A_33_P3266769 | 0,029083842 | down | -1,6246543 | LHCGR | luteinizing hormone/choriogonadotropin receptor [Source:HGNC Symbol;Acc:HGNC:6585] [ENST00000477576] |
| A_23_P32165 | 0,003005201 | down | -2,0424867 | LHX2 | Homo sapiens LIM homeobox 2 (LHX2), mRNA [NM_004789] |
| A_33_P3339246 | 0,013194697 | down | -1,8058815 | LHX8 | Homo sapiens LIM homeobox 8 (LHX8), transcript variant 1, mRNA [NM_001001933] |
| A_23_P327562 | 0,018744908 | down | -1,7599863 | LHX9 | Homo sapiens LIM homeobox 9 (LHX9), transcript variant 1, mRNA [NM_020204] |
| A_23_P90497 | 0,030422596 | down | -1,6590503 | LILRA4 | Homo sapiens leukocyte immunoglobulin-like receptor, subfamily A (with TM domain), member 4 (LILRA4), mRNA [NM_012276] |
| A_33_P3397599 | 0,02240254 | down | -1,5443267 | LILRA6 | Homo sapiens leukocyte immunoglobulin-like receptor, subfamily A (with TM domain), member 6 (LILRA6), transcript variant 2, non-c |
| A_19_P00808088 | 0,01331415 | down | -2,1073465 | LMCD1-AS1 | Homo sapiens LMCD1 antisense RNA 1 (head to head) (LMCD1-AS1), long non-coding RNA [NR_033378] |
| A_21_P0000009 | 0,012701068 | down | -1,7340429 | LMNTD1 | Homo sapiens lamin tail domain containing 1 (LMNTD1), transcript variant 4, mRNA [NM_001145729] |
| A_23_P87310 | 0,02378552 | down | -1,9068073 | LMO1 | Homo sapiens LIM domain only 1 (rhombotin 1) (LMO1), transcript variant 1, mRNA [NM_002315] |
| A_33_P3295261 | 0,02405326 | down | -2,0753076 | LMOD1 | Homo sapiens leiomodin 1 (smooth muscle) (LMOD1), mRNA [NM_012134] |
| A_21_P0013116 | 0,03655672 | down | -2,2979639 | LPAL2 | Homo sapiens lipoprotein, Lp(a)-like 2, pseudogene (LPAL2), transcript variant 2, non-coding RNA [NR_028093] |
| A_23_P10980 | 0,021006359 | down | -1,6632141 | LPHN3 | Homo sapiens latrophilin 3 (LPHN3), mRNA [NM_015236] |
| A_33_P3384958 | 0,046123818 | down | -1,7408122 | LPPR4 | Homo sapiens lipid phosphate phosphatase-related protein type 4 (LPPR4), transcript variant 1, mRNA [NM_014839] |
| A_33_P3313215 | 0,032946598 | down | -1,5020438 | LRIT2 | Homo sapiens leucine-rich repeat, immunoglobulin-like and transmembrane domains 2 (LRIT2), transcript variant 1, mRNA [NM_001 |
| A_33_P3253538 | 0,033262815 | down | -1,6693532 | LRRC14B | Homo sapiens leucine rich repeat containing 14B (LRRC14B), mRNA [NM_001080478] |
| A_24_P827037 | 0,04940006 | down | -1,5716008 | LRRC15 | Homo sapiens leucine rich repeat containing 15 (LRRC15), transcript variant 2, mRNA [NM_130830] |
| A_33_P3329522 | 0,035971925 | down | -1,8807086 | LRRC17 | Homo sapiens leucine rich repeat containing 17 (LRRC17), transcript variant 1, mRNA [NM_001031692] |
| A_33_P3325763 | 0,03961499 | down | -1,8114413 | LRRC3 | leucine rich repeat containing 3 [Source:HGNC Symbol;Acc:HGNC:14965] [ENST00000291592] |
| A_33_P3412013 | 0,02789616 | down | -1,8558396 | LRRC37A5P | Homo sapiens leucine rich repeat containing 37, member A5, pseudogene (LRRC37A5P), long non-coding RNA [NR_034087] |
| A_23_P103877 | 0,025501208 | down | -1,5451796 | LRRC38 | Homo sapiens leucine rich repeat containing 38 (LRRC38), mRNA [NM_001010847] |
| A_33_P3228739 | 0,030499687 | down | -1,6381553 | LRRC3C | Homo sapiens leucine rich repeat containing 3C (LRRC3C), mRNA [NM_001195545] |
| A_33_P3263212 | 0,03668526 | down | -1,5966007 | LRRC7 | Homo sapiens leucine rich repeat containing 7 (LRRC7), mRNA [NM_020794] |
| A_32_P222149 | 0,01813859 | down | -2,1223886 | LRRC72 | Homo sapiens leucine rich repeat containing 72 (LRRC72), mRNA [NM_001195280] |
| A_33_P3471466 | 0,024049615 | down | -1,8462199 | LRRC74A | Homo sapiens leucine rich repeat containing 74A (LRRC74A), mRNA [NM_194287] |
| A_21_P0011248 | 0,013530348 | down | -1,540009 | LRRC9 | Homo sapiens leucine rich repeat containing 9 (LRRC9), non-coding RNA [NR_075071] |

| | | | | | |
|---|---|---|---|---|---|
| A_33_P3228082 | 0,028227016 | down | -1,755979 | LRRC9 | Homo sapiens leucine rich repeat containing 9 (LRRC9), non-coding RNA [NR_075071] |
| A_33_P3333762 | 0,010576146 | down | -2,3159719 | LRRIQ4 | Homo sapiens leucine-rich repeats and IQ motif containing 4 (LRRIQ4), mRNA [NM_001080460] |
| A_33_P3369058 | 0,025662275 | down | -2,1408231 | LRRK2 | Homo sapiens leucine-rich repeat kinase 2 (LRRK2), mRNA [NM_198578] |
| A_33_P3251024 | 0,003865806 | down | -2,1450882 | LRRTM2 | Homo sapiens leucine rich repeat transmembrane neuronal 2 (LRRTM2), mRNA [NM_015564] |
| A_24_P295465 | 0,04664632 | down | -1,7114917 | LRRTM3 | Homo sapiens leucine rich repeat transmembrane neuronal 3 (LRRTM3), transcript variant 1, mRNA [NM_178011] |
| A_21_P0004421 | 0,001957095 | down | -3,0867379 | LUCAT1 | Homo sapiens lung cancer associated transcript 1 (non-protein coding) (LUCAT1), transcript variant 1, long non-coding RNA [NR_10 |
| A_33_P3410232 | 0,00843408 | down | -2,8161294 | LY6G6E | Homo sapiens lymphocyte antigen 6 complex, locus G6E (pseudogene) (LY6G6E), transcript variant 1, non-coding RNA [NR_02454 |
| A_32_P178758 | 0,01623019 | down | -1,6708554 | LY86-AS1 | Homo sapiens LY86 antisense RNA 1 (LY86-AS1), long non-coding RNA [NR_026970] |
| A_24_P164998 | 0,04618982 | down | -1,8268793 | LYPD8 | Homo sapiens LY6/PLAUR domain containing 8 (LYPD8), transcript variant 1, mRNA [NM_001085474] |
| A_21_P0010798 | 0,03150783 | down | -2,3285651 | LYPD8 | LY6/PLAUR domain containing 8 [Source:HGNC Symbol;Acc:HGNC:44208] [ENST00000566597] |
| A_33_P3312611 | 0,02163946 | down | -1,5357232 | LYPLAL1-AS1 | Homo sapiens LYPLAL1 antisense RNA 1 (head to head) (LYPLAL1-AS1), long non-coding RNA [NR_038845] |
| A_33_P3288484 | 0,019724475 | down | -1,6927693 | LYRM2 | Homo sapiens LYR motif containing 2 (LYRM2), transcript variant 1, mRNA [NM_020466] |
| A_23_P130056 | 0,044759832 | down | -1,668994 | LYZL6 | Homo sapiens lysozyme-like 6 (LYZL6), transcript variant 2, mRNA [NM_020426] |
| A_32_P55871 | 4,34E-04 | down | -2,0672731 | MAATS1 | Homo sapiens MYCBP-associated, testis expressed 1 (MAATS1), mRNA [NM_033364] |
| A_23_P53884 | 0,011463768 | down | -1,8319464 | MAB21L1 | Homo sapiens mab-21-like 1 (C. elegans) (MAB21L1), mRNA [NM_005584] |
| A_33_P3359250 | 0,009580674 | down | -1,8293854 | MAD2L2 | MAD2 mitotic arrest deficient-like 2 (yeast) [Source:HGNC Symbol;Acc:HGNC:6764] [ENST00000376664] |
| A_33_P3406828 | 0,043331243 | down | -1,6847068 | MAFIP | Homo sapiens MAFF interacting protein (pseudogene) (MAFIP), transcript variant 1, non-coding RNA [NR_046439] |
| A_23_P136870 | 0,038517393 | down | -1,838736 | MAGEA6 | Homo sapiens melanoma antigen family A, 6 (MAGEA6), transcript variant 2, mRNA [NM_175868] |
| A_23_P96432 | 0,024737077 | down | -1,5041717 | MAGEB3 | Homo sapiens melanoma antigen family B, 3 (MAGEB3), mRNA [NM_002365] |
| A_33_P3365860 | 0,009986353 | down | -2,5970807 | MAGEB5 | Homo sapiens melanoma antigen family B, 5 (MAGEB5), mRNA [NM_001271752] |
| A_23_P502081 | 0,025154505 | down | -2,347159 | MAGEC3 | Homo sapiens melanoma antigen family C, 3 (MAGEC3), transcript variant 1, mRNA [NM_138702] |
| A_24_P330112 | 0,040388357 | up | 1,5208633 | MAGOHB | Homo sapiens mago-nashi homolog B (Drosophila) (MAGOHB), transcript variant 1, mRNA [NM_018048] |
| A_21_P0010808 | 0,01438448 | down | -1,7226753 | MALRD1 | Homo sapiens MAM and LDL receptor class A domain containing 1 (MALRD1), mRNA [NM_001142308] |
| A_23_P308483 | 0,00961127 | down | -1,8202497 | MAP3K15 | Homo sapiens mitogen-activated protein kinase kinase kinase 15 (MAP3K15), mRNA [NM_001001671] |
| A_33_P3226835 | 0,015095647 | down | -1,5610665 | MAP3K19 | Homo sapiens mitogen-activated protein kinase kinase kinase 19 (MAP3K19), transcript variant 1, mRNA [NM_025052] |
| A_23_P5002 | 0,016190061 | down | -1,9600558 | MAP4K1 | Homo sapiens mitogen-activated protein kinase kinase kinase kinase 1 (MAP4K1), transcript variant 1, mRNA [NM_001042600] |
| A_24_P33508 | 0,03066948 | down | -1,90427 | MARCH11 | Homo sapiens membrane-associated ring finger (C3HC4) 11 (MARCH11), mRNA [NM_001102562] |
| A_24_P555473 | 0,003589794 | down | -1,7051879 | MARCH3 | Homo sapiens membrane-associated ring finger (C3HC4) 3, E3 ubiquitin protein ligase (MARCH3), mRNA [NM_178450] |
| A_23_P310 | 0,036913984 | up | 1,7013352 | MARCKSL1 | Homo sapiens MARCKS-like 1 (MARCKSL1), transcript variant 1, mRNA [NM_023009] |
| A_33_P3327010 | 0,018821258 | down | -1,906458 | MAST2 | Homo sapiens microtubule associated serine/threonine kinase 2 (MAST2), mRNA [NM_015112] |
| A_23_P35529 | 0,016076481 | down | -1,7424669 | MBL2 | Homo sapiens mannose-binding lectin (protein C) 2, soluble (MBL2), mRNA [NM_000242] |
| A_23_P50039 | 0,024530353 | down | -2,340947 | MC5R | Homo sapiens melanocortin 5 receptor (MC5R), mRNA [NM_005913] |
| A_24_P98006 | 0,012918294 | down | -1,5489992 | MCHR2 | Homo sapiens melanin-concentrating hormone receptor 2 (MCHR2), transcript variant 1, mRNA [NM_001040179] |
| A_33_P3421867 | 0,025253518 | down | -1,9638449 | MDGA1 | MAM domain containing glycosylphosphatidylinositol anchor 1 [Source:HGNC Symbol;Acc:HGNC:19267] [ENST00000373401] |
| A_24_P106624 | 0,03683387 | down | -1,5203565 | MEOX2 | Homo sapiens mesenchyme homeobox 2 (MEOX2), mRNA [NM_005924] |
| A_33_P3418877 | 0,03303317 | down | -1,8649106 | MESTIT1 | Homo sapiens MEST intronic transcript 1, antisense RNA (MESTIT1), long non-coding RNA [NR_004382] |
| A_24_P64653 | 0,039237116 | down | -2,3458757 | METTL7B | Homo sapiens methyltransferase like 7B (METTL7B), mRNA [NM_152637] |
| A_23_P10062 | 0,018048776 | down | -1,7168759 | MGAT4C | Homo sapiens MGAT4 family, member C (MGAT4C), mRNA [NM_013244] |
| A_21_P0000533 | 0,036055263 | down | -1,6930189 | MGC27382 | Homo sapiens uncharacterized MGC27382 (MGC27382), long non-coding RNA [NR_027310] |
| A_19_P00322705 | 0,014285381 | down | -1,8138497 | MIAT | Homo sapiens myocardial infarction associated transcript (non-protein coding) (MIAT), transcript variant 1, long non-coding RNA [NR |
| A_19_P00315524 | 0,014834112 | down | -1,8691691 | MIAT | Homo sapiens myocardial infarction associated transcript (non-protein coding) (MIAT), transcript variant 1, long non-coding RNA [NR |
| A_19_P00321068 | 0,016492907 | down | -1,8953818 | MIAT | Homo sapiens myocardial infarction associated transcript (non-protein coding) (MIAT), transcript variant 1, long non-coding RNA [NR |
| A_19_P00319573 | 0,002692486 | down | -2,857709 | MIAT | myocardial infarction associated transcript (non-protein coding) [Source:HGNC Symbol;Acc:HGNC:33425] [ENST00000418918] |
| A_21_P0006525 | 0,038374793 | down | -1,5076687 | MID1 | PREDICTED: Homo sapiens midline 1 (Opitz/BBB syndrome) (MID1), transcript variant X2, misc_RNA [XR_247299] |
| A_21_P0006407 | 0,030546632 | down | -1,9590634 | MID1 | PREDICTED: Homo sapiens midline 1 (Opitz/BBB syndrome) (MID1), transcript variant X3, misc_RNA [XR_247300] |
| A_33_P3341479 | 0,016142607 | down | -2,253803 | MID1IP1-AS1 | Homo sapiens MID1IP1 antisense RNA 1 (MID1IP1-AS1), long non-coding RNA [NR_046706] |
| A_33_P3230951 | 0,02890488 | down | -2,2230616 | MIMT1 | Homo sapiens MER1 repeat containing imprinted transcript 1 (non-protein coding) (MIMT1), long non-coding RNA [NR_024059] |
| A_33_P3267612 | 0,01706529 | down | -1,6215658 | MIR143HG | Homo sapiens MIR143 host gene (non-protein coding) (MIR143HG), transcript variant 1, long non-coding RNA [NR_105059] |
| A_19_P00322941 | 0,020048054 | down | -2,4096763 | MIR143HG | MIR143 host gene (non-protein coding) [Source:HGNC Symbol;Acc:HGNC:42872] [ENST00000518014] |

| | | | | | |
|---|---|---|---|---|---|
| A_21_P0010972 | 0,018481193 | down | -1,5706773 | MIR670HG | PREDICTED: Homo sapiens MIR670 host gene (non-protein coding) (MIR670HG), transcript variant X1, ncRNA [XR_109044] |
| A_24_P346855 | 0,02566544 | down | -1,5026174 | MKI67 | Homo sapiens marker of proliferation Ki-67 (MKI67), transcript variant 1, mRNA [NM_002417] |
| A_23_P43350 | 0,018209439 | down | -2,549153 | MLANA | Homo sapiens melan-A (MLANA), mRNA [NM_005511] |
| A_33_P3240578 | 0,04776173 | up | 1,6836269 | MLLT4-AS1 | Homo sapiens MLLT4 antisense RNA 1 (head to head) (MLLT4-AS1), long non-coding RNA [NR_027906] |
| A_23_P19529 | 0,025830306 | down | -2,3999329 | MLN | Homo sapiens motilin (MLN), transcript variant 2, mRNA [NM_001040109] |
| A_33_P3343260 | 0,019171745 | down | -1,5242124 | MMP25-AS1 | Homo sapiens MMP25 antisense RNA 1 (MMP25-AS1), long non-coding RNA [NR_123723] |
| A_33_P3398316 | 0,032978766 | down | -1,6535074 | MMP27 | Homo sapiens matrix metallopeptidase 27 (MMP27), mRNA [NM_022122] |
| A_33_P3212257 | 0,04559144 | down | -1,6133904 | MMRN1 | Homo sapiens multimerin 1 (MMRN1), mRNA [NM_007351] |
| A_33_P3391439 | 0,03167116 | down | -1,5740826 | MOBP | Homo sapiens myelin-associated oligodendrocyte basic protein (MOBP), transcript variant 6, non-coding RNA [NR_103505] |
| A_33_P3350202 | 0,00280205 | down | -1,7730066 | MOCS3 | Homo sapiens molybdenum cofactor synthesis 3 (MOCS3), mRNA [NM_014484] |
| A_33_P3353866 | 0,03079916 | down | -2,3416293 | MOGAT2 | Homo sapiens monoacylglycerol O-acyltransferase 2 (MOGAT2), mRNA [NM_025098] |
| A_33_P3326984 | 0,026959056 | up | 1,7924637 | MOK | MOK protein kinase [Source:HGNC Symbol;Acc:HGNC:9833] [ENST00000520252] |
| A_33_P3308626 | 0,01815931 | down | -1,6070156 | MON1B | Homo sapiens MON1 secretory trafficking family member B (MON1B), transcript variant 1, mRNA [NM_014940] |
| A_33_P3301221 | 0,0424404 | down | -1,5947183 | MORN1 | Homo sapiens MORN repeat containing 1 (MORN1), transcript variant 3, non-coding RNA [NR_125361] |
| A_23_P143774 | 0,009422177 | down | -1,9976432 | MOV10L1 | Homo sapiens Mov10 RISC complex RNA helicase like 1 (MOV10L1), transcript variant 1, mRNA [NM_018995] |
| A_33_P3315390 | 0,005439034 | down | -1,6178117 | MPPED2 | Homo sapiens metallophosphoesterase domain containing 2 (MPPED2), transcript variant 2, mRNA [NM_001145399] |
| A_33_P3368991 | 0,021981671 | down | -1,5019486 | MPZ | Homo sapiens myelin protein zero (MPZ), mRNA [NM_000530] |
| A_23_P12746 | 0,017510623 | down | -2,5118883 | MRC1 | Homo sapiens mannose receptor, C type 1 (MRC1), mRNA [NM_002438] |
| A_24_P315638 | 0,02326975 | down | -2,3199162 | MRGPRE | Homo sapiens MAS-related GPR, member E (MRGPRE), mRNA [NM_001039165] |
| A_33_P3414187 | 0,005624386 | down | -1,8189058 | MRGPRX2 | MAS-related GPR, member X2 [Source:HGNC Symbol;Acc:HGNC:17983] [ENST00000329773] |
| A_32_P49309 | 0,012127081 | down | -1,5549365 | MRO | Homo sapiens maestro (MRO), transcript variant 1, mRNA [NM_031939] |
| A_23_P4545 | 0,04316003 | down | -2,013822 | MRO | Homo sapiens maestro (MRO), transcript variant 1, mRNA [NM_031939] |
| A_21_P0007310 | 0,016397106 | down | -1,9900146 | MRPL23-AS1 | Homo sapiens MRPL23 antisense RNA 1 (MRPL23-AS1), long non-coding RNA [NR_024471] |
| A_33_P3265444 | 0,007131707 | down | -1,9921964 | MRS2P2 | Homo sapiens MRS2 pseudogene 2 (MRS2P2), non-coding RNA [NR_024072] |
| A_23_P127760 | 0,02509917 | down | -1,8254985 | MS4A12 | Homo sapiens membrane-spanning 4-domains, subfamily A, member 12 (MS4A12), transcript variant 1, mRNA [NM_017716] |
| A_33_P3386681 | 0,023245169 | down | -1,5094343 | MS4A13 | Homo sapiens membrane-spanning 4-domains, subfamily A, member 13 (MS4A13), transcript variant 1, mRNA [NM_001012417] |
| A_23_P98565 | 0,009936024 | down | -1,5753117 | MS4A14 | Homo sapiens membrane-spanning 4-domains, subfamily A, member 14 (MS4A14), transcript variant 1, mRNA [NM_032597] |
| A_33_P3295313 | 2,88E-04 | down | -3,3100839 | MS4A2 | Homo sapiens membrane-spanning 4-domains, subfamily A, member 2 (MS4A2), transcript variant 1, mRNA [NM_000139] |
| A_23_P75769 | 0,009824608 | down | -2,2440243 | MS4A4A | Homo sapiens membrane-spanning 4-domains, subfamily A, member 4A (MS4A4A), transcript variant 2, mRNA [NM_024021] |
| A_33_P3330872 | 0,001458078 | down | -1,969483 | MSL3 | Homo sapiens male-specific lethal 3 homolog (Drosophila) (MSL3), transcript variant 1, mRNA [NM_078629] |
| A_32_P361185 | 0,011174598 | up | 2,106383 | MSS51 | Homo sapiens MSS51 mitochondrial translational activator (MSS51), mRNA [NM_001024593] |
| A_21_P0010518 | 0,025333704 | down | -1,676155 | MST1P2 | Homo sapiens macrophage stimulating 1 (hepatocyte growth factor-like) pseudogene 2 (MST1P2), non-coding RNA [NR_027504] |
| A_21_P0000110 | 0,003094848 | down | -2,1579723 | MTRNR2L10 | Homo sapiens MT-RNR2-like 10 (MTRNR2L10), mRNA [NM_001190708] |
| A_23_P306610 | 0,026377171 | down | -1,7250291 | MUC17 | Homo sapiens mucin 17, cell surface associated (MUC17), mRNA [NM_001040105] |
| A_21_P0007832 | 0,02856536 | down | -1,8988714 | MUC19 | Homo sapiens mucin 19, oligomeric (MUC19), mRNA [NM_173600] |
| A_23_P256784 | 0,007262185 | down | -1,6538298 | MUC2 | Homo sapiens mucin 2, oligomeric mucus/gel-forming (MUC2), mRNA [NM_002457] |
| A_21_P0004641 | 0,001457015 | down | -2,7669964 | MUC22 | Homo sapiens mucin 22 (MUC22), mRNA [NM_001198815] |
| A_23_P313278 | 0,03299773 | down | -2,2489772 | MUC3A | Homo sapiens MUC3A mRNA for intestinal mucin, partial cds. [AB038784] |
| A_33_P3216570 | 6,84E-04 | down | -2,7739484 | MUC5AC | PREDICTED: Homo sapiens mucin 5AC, oligomeric mucus/gel-forming (MUC5AC), partial mRNA [XM_006709945] |
| A_23_P121614 | 0,04446174 | down | -1,8414402 | MUC7 | Homo sapiens mucin 7, secreted (MUC7), transcript variant 3, mRNA [NM_152291] |
| A_23_P71649 | 0,04060037 | down | -1,913628 | MUSK | Homo sapiens muscle, skeletal, receptor tyrosine kinase (MUSK), transcript variant 1, mRNA [NM_005592] |
| A_33_P3354703 | 0,041456167 | down | -1,5524842 | MYADML | Homo sapiens myeloid-associated differentiation marker-like (pseudogene) (MYADML), non-coding RNA [NR_003143] |
| A_23_P143190 | 0,022219589 | down | -1,6488377 | MYBL2 | Homo sapiens v-myb avian myeloblastosis viral oncogene homolog-like 2 (MYBL2), transcript variant 1, mRNA [NM_002466] |
| A_33_P3361388 | 0,007121021 | down | -1,9170288 | MYCBPAP | Homo sapiens MYCBP associated protein (MYCBPAP), mRNA [NM_032133] |
| A_21_P0002574 | 0,033023104 | down | -1,6460961 | MYCNUT | Homo sapiens MYCN upstream transcript (non-protein coding) (MYCNUT), long non-coding RNA [NR_125783] |
| A_24_P131646 | 0,045093197 | down | -1,93593 | MYL3 | Homo sapiens myosin, light chain 3, alkali; ventricular, skeletal, slow (MYL3), mRNA [NM_000258] |
| A_24_P195669 | 0,047109917 | up | 1,7565132 | MYO15B | myosin XVB pseudogene [Source:HGNC Symbol;Acc:HGNC:14083] [ENST00000581866] |
| A_33_P3298216 | 0,017748736 | down | -1,7841884 | MYO16 | Homo sapiens myosin XVI (MYO16), transcript variant 1, mRNA [NM_001198950] |
| A_23_P86411 | 0,013175471 | down | -2,981841 | MYO3A | Homo sapiens myosin IIIA (MYO3A), mRNA [NM_017433] |

| | | | | | |
|---|---|---|---|---|---|
| A_23_P417853 | 0,003943107 | down | -3,089567 | MYO3B | Homo sapiens myosin IIIB (MYO3B), transcript variant 2, mRNA [NM_138995] |
| A_23_P422350 | 0,008919479 | down | -2,6658723 | MYO7A | Homo sapiens myosin VIIA (MYO7A), transcript variant 1, mRNA [NM_000260] |
| A_23_P160438 | 4,85E-04 | down | -2,1827946 | MYOG | Homo sapiens myogenin (myogenic factor 4) (MYOG), mRNA [NM_002479] |
| A_33_P3265526 | 0,0274323 | down | -1,6611817 | NAA16 | Homo sapiens N(alpha)-acetyltransferase 16, NatA auxiliary subunit (NAA16), transcript variant 3, mRNA [NM_001110798] |
| A_23_P161583 | 0,034985024 | down | -1,5058839 | NAALAD2 | Homo sapiens N-acetylated alpha-linked acidic dipeptidase 2 (NAALAD2), transcript variant 1, mRNA [NM_005467] |
| A_33_P3374987 | 0,038903512 | down | -1,7453496 | NAT16 | Homo sapiens N-acetyltransferase 16 (GCN5-related, putative) (NAT16), mRNA [NM_198571] |
| A_23_P31798 | 0,007207064 | down | -1,7664478 | NAT2 | Homo sapiens N-acetyltransferase 2 (arylamine N-acetyltransferase) (NAT2), mRNA [NM_000015] |
| A_24_P308506 | 0,013390812 | down | -2,895042 | NAT8B | Homo sapiens N-acetyltransferase 8B (GCN5-related, putative, gene/pseudogene) (NAT8B), mRNA [NM_016347] |
| A_33_P3336273 | 0,011199677 | down | -2,6443272 | NAV3 | Homo sapiens neuron navigator 3 (NAV3), transcript variant 1, mRNA [NM_001024383] |
| A_33_P3229328 | 0,009837533 | down | -1,8517557 | NBPF6 | Homo sapiens neuroblastoma breakpoint family, member 6 (NBPF6), transcript variant 2, mRNA [NM_001143988] |
| A_33_P3363799 | 0,024858827 | down | -1,6049216 | NCAM1 | Homo sapiens neural cell adhesion molecule 1 (NCAM1), transcript variant 5, mRNA [NM_001242607] |
| A_33_P3363804 | 0,018789183 | down | -2,021061 | NCAM1 | Homo sapiens neural cell adhesion molecule 1 (NCAM1), transcript variant 4, mRNA [NM_001242608] |
| A_33_P3239884 | 0,002841969 | down | -2,5537057 | NCAM2 | Homo sapiens neural cell adhesion molecule 2 (NCAM2), mRNA [NM_004540] |
| A_23_P153797 | 0,014627793 | down | -1,7978258 | NCAN | Homo sapiens neurocan (NCAN), mRNA [NM_004386] |
| A_33_P3344423 | 0,03575051 | down | -2,0579205 | NCR3 | Homo sapiens natural cytotoxicity triggering receptor 3 (NCR3), transcript variant 3, mRNA [NM_001145467] |
| A_24_P944756 | 0,030244118 | down | -1,5702604 | NECAB1 | Homo sapiens N-terminal EF-hand calcium binding protein 1 (NECAB1), mRNA [NM_022351] |
| A_21_P0014167 | 0,005656122 | down | -2,0893974 | NEDD9 | neural precursor cell expressed, developmentally down-regulated 9 [Source:HGNC Symbol;Acc:HGNC:7733] [ENST00000379433] |
| A_33_P3341970 | 0,01064762 | down | -1,7429734 | NEGR1 | Homo sapiens neuronal growth regulator 1 (NEGR1), mRNA [NM_173808] |
| A_23_P155711 | 0,02755507 | down | -1,7996886 | NEIL3 | Homo sapiens nei endonuclease VIII-like 3 (E. coli) (NEIL3), mRNA [NM_018248] |
| A_33_P3250521 | 0,040740874 | down | -1,8263547 | NEK5 | Homo sapiens NIMA-related kinase 5 (NEK5), mRNA [NM_199289] |
| A_23_P251151 | 0,03332935 | down | -1,9204824 | NELL1 | Homo sapiens NEL-like 1 (chicken) (NELL1), transcript variant 1, mRNA [NM_006157] |
| A_33_P3651948 | 0,002632919 | down | -2,4755828 | NEO1 | Homo sapiens neogenin 1 (NEO1), transcript variant 1, mRNA [NM_002499] |
| A_23_P218626 | 0,015912347 | down | -1,6067183 | NEU4 | Homo sapiens sialidase 4 (NEU4), transcript variant 1, mRNA [NM_080741] |
| A_23_P24485 | 0,02755005 | up | 1,6015421 | NFRKB | Homo sapiens nuclear factor related to kappaB binding protein (NFRKB), transcript variant 2, mRNA [NM_006165] |
| A_33_P3374258 | 0,011579833 | down | -1,5482773 | NHLH2 | nescient helix loop helix 2 [Source:HGNC Symbol;Acc:HGNC:7818] [ENST00000369506] |
| A_23_P119042 | 0,031219978 | down | -1,8689405 | NKG7 | Homo sapiens natural killer cell granule protein 7 (NKG7), mRNA [NM_005601] |
| A_33_P3359801 | 0,028437251 | down | -1,5990512 | NKX2-4 | Homo sapiens NK2 homeobox 4 (NKX2-4), mRNA [NM_033176] |
| A_23_P18123 | 0,04825984 | down | -1,5699302 | NLGN1 | Homo sapiens neuroligin 1 (NLGN1), mRNA [NM_014932] |
| A_19_P00318172 | 0,00433782 | down | -1,9935366 | NOL4L | Homo sapiens nucleolar protein 4-like (NOL4L), transcript variant 1, mRNA [NM_001256798] |
| A_33_P3347697 | 0,046253055 | down | -2,3265584 | NOVA1 | Homo sapiens neuro-oncological ventral antigen 1 (NOVA1), transcript variant 1, mRNA [NM_002515] |
| A_33_P3224324 | 0,01927524 | down | -1,5609238 | NOX4 | Homo sapiens NADPH oxidase 4 (NOX4), transcript variant NOX4B, mRNA [NM_001143836] |
| A_21_P0010973 | 7,82E-04 | down | -2,288842 | NOX4 | Homo sapiens NADPH oxidase 4 (NOX4), transcript variant 1, mRNA [NM_016931] |
| A_33_P3372389 | 0,003357381 | down | -2,1958215 | NPHP4 | Homo sapiens nephronophthisis 4 (NPHP4), transcript variant 1, mRNA [NM_015102] |
| A_33_P3356990 | 0,03562931 | down | -1,7808349 | NPIPB3 | Homo sapiens nuclear pore complex interacting protein family, member B3 (NPIPB3), mRNA [NM_130464] |
| A_23_P62752 | 0,049293585 | down | -1,5550296 | NPPB | Homo sapiens natriuretic peptide B (NPPB), mRNA [NM_002521] |
| A_33_P3417113 | 0,014879715 | down | -2,569186 | NPSR1 | Homo sapiens neuropeptide S receptor 1 (NPSR1), transcript variant 4, mRNA [NM_001300934] |
| A_33_P3232403 | 0,020753045 | down | -1,8655186 | NPSR1-AS1 | Homo sapiens NPSR1 antisense RNA 1 (NPSR1-AS1), transcript variant 3, long non-coding RNA [NR_015356] |
| A_23_P124905 | 0,025623603 | down | -1,8063793 | NPTX1 | Homo sapiens neuronal pentraxin I (NPTX1), mRNA [NM_002522] |
| A_33_P3396244 | 0,03147338 | down | -1,7422631 | NPY2R | Homo sapiens neuropeptide Y receptor Y2 (NPY2R), mRNA [NM_000910] |
| A_33_P3229412 | 0,01686717 | down | -2,3086514 | NRG3 | Homo sapiens neuregulin 3 (NRG3), transcript variant 1, mRNA [NM_001010848] |
| A_32_P486693 | 0,04066332 | down | -1,8055965 | NRIP3 | Homo sapiens nuclear receptor interacting protein 3 (NRIP3), mRNA [NM_020645] |
| A_23_P255618 | 0,027217021 | down | -1,6472136 | NRSN1 | Homo sapiens neurensin 1 (NRSN1), mRNA [NM_080723] |
| A_33_P3273679 | 0,019523714 | down | -1,5049766 | NRXN1 | Homo sapiens neurexin 1, mRNA (cDNA clone IMAGE:4815048), complete cds. [BC046631] |
| A_23_P205900 | 0,004602406 | down | -2,3870409 | NTRK3 | Homo sapiens neurotrophic tyrosine kinase, receptor, type 3 (NTRK3), transcript variant 1, mRNA [NM_001012338] |
| A_23_P36882 | 0,004819464 | down | -1,8925977 | NTS | Homo sapiens neurotensin (NTS), mRNA [NM_006183] |
| A_24_P626981 | 0,01359252 | down | -1,5124943 | NUDT16P1 | Homo sapiens nudix (nucleoside diphosphate linked moiety X)-type motif 16 pseudogene 1 (NUDT16P1), transcript variant 2, non-cc |
| A_23_P393955 | 0,025015803 | down | -1,6212423 | NUTM1 | Homo sapiens NUT midline carcinoma, family member 1 (NUTM1), transcript variant 3, mRNA [NM_175741] |
| A_23_P148568 | 0,03134269 | down | -2,4392092 | NXF2 | Homo sapiens nuclear RNA export factor 2 (NXF2), mRNA [NM_022053] |
| A_23_P171336 | 0,00904082 | down | -1,8883972 | NXF3 | Homo sapiens nuclear RNA export factor 3 (NXF3), mRNA [NM_022052] |

| | | | | | |
|---|---|---|---|---|---|
| A_33_P3423506 | 0,038413778 | down | -1,6355782 | NXPE2 | Homo sapiens neurexophilin and PC-esterase domain family, member 2 (NXPE2), mRNA [NM_182495] |
| A_33_P3243917 | 0,005221182 | down | -1,650293 | OBP2B | Homo sapiens odorant binding protein 2B (OBP2B), transcript variant alpha, mRNA [NM_014581] |
| A_33_P3405399 | 0,017602803 | down | -1,6110272 | ODF2L | Homo sapiens outer dense fiber of sperm tails 2-like (ODF2L), transcript variant 3, mRNA [NM_001184765] |
| A_24_P4736 | 0,014130244 | down | -2,758079 | ODF3 | Homo sapiens outer dense fiber of sperm tails 3 (ODF3), transcript variant 1, mRNA [NM_053280] |
| A_33_P3377256 | 0,001153963 | up | 2,0249357 | OGFRP1 | Homo sapiens opioid growth factor receptor pseudogene 1 (OGFRP1), non-coding RNA [NR_036498] |
| A_33_P3791263 | 0,030990012 | down | -1,627912 | OLFM3 | Homo sapiens olfactomedin 3 (OLFM3), transcript variant 3, mRNA [NM_001288823] |
| A_23_P55286 | 0,025392123 | down | -1,5167581 | OMG | Homo sapiens oligodendrocyte myelin glycoprotein (OMG), mRNA [NM_002544] |
| A_33_P3355886 | 0,009158146 | down | -2,1629128 | OOSP1 | oocyte secreted protein 1 [Source:HGNC Symbol;Acc:HGNC:49233] [ENST00000398992] |
| A_23_P317820 | 0,024143169 | down | -1,5902946 | OOSP2 | Homo sapiens oocyte secreted protein 2 (OOSP2), mRNA [NM_173801] |
| A_33_P3404974 | 0,015888244 | down | -2,425905 | OPN5 | Homo sapiens opsin 5 (OPN5), transcript variant 1, mRNA [NM_181744] |
| A_33_P3359647 | 0,007100266 | down | -1,9313471 | OPRL1 | Homo sapiens opiate receptor-like 1 (OPRL1), transcript variant 1, mRNA [NM_182647] |
| A_23_P75867 | 0,020425553 | down | -2,7280943 | OR10A4 | Homo sapiens olfactory receptor, family 10, subfamily A, member 4 (OR10A4), mRNA [NM_207186] |
| A_23_P315991 | 2,55E-04 | down | -4,965023 | OR10A5 | Homo sapiens olfactory receptor, family 10, subfamily A, member 5 (OR10A5), mRNA [NM_178168] |
| A_33_P3273906 | 0,04627171 | down | -1,9127558 | OR10G4 | Homo sapiens olfactory receptor, family 10, subfamily G, member 4 (OR10G4), mRNA [NM_001004462] |
| A_33_P3290792 | 0,024469726 | down | -2,045246 | OR10G9 | Homo sapiens olfactory receptor, family 10, subfamily G, member 9 (OR10G9), mRNA [NM_001001953] |
| A_33_P3359383 | 0,0329296 | down | -2,2303607 | OR10H5 | Homo sapiens olfactory receptor, family 10, subfamily H, member 5 (OR10H5), mRNA [NM_001004466] |
| A_23_P705 | 0,020731745 | down | -1,5605893 | OR10R2 | Homo sapiens olfactory receptor, family 10, subfamily R, member 2 (OR10R2), mRNA [NM_001004472] |
| A_33_P3381097 | 0,03317082 | down | -1,9448466 | OR10V1 | Homo sapiens olfactory receptor, family 10, subfamily V, member 1 (OR10V1), mRNA [NM_001005324] |
| A_33_P3326271 | 0,015069996 | down | -2,1182187 | OR13C2 | Homo sapiens olfactory receptor, family 13, subfamily C, member 2 (OR13C2), mRNA [NM_001004481] |
| A_33_P3244215 | 0,015015913 | down | -2,1482882 | OR13C5 | Homo sapiens olfactory receptor, family 13, subfamily C, member 5 (OR13C5), mRNA [NM_001004482] |
| A_33_P3266489 | 0,004965589 | down | -2,2513986 | OR13H1 | Homo sapiens olfactory receptor, family 13, subfamily H, member 1 (OR13H1), mRNA [NM_001004486] |
| A_33_P3270252 | 0,026854103 | down | -1,6455377 | OR1B1 | Homo sapiens olfactory receptor, family 1, subfamily B, member 1 (gene/pseudogene) (OR1B1), mRNA [NM_001004450] |
| A_24_P299769 | 0,03918802 | down | -1,680728 | OR1C1 | Homo sapiens olfactory receptor, family 1, subfamily C, member 1 (OR1C1), mRNA [NM_012353] |
| A_23_P15832 | 2,04E-04 | down | -3,2454913 | OR1D5 | Homo sapiens olfactory receptor, family 1, subfamily D, member 5 (OR1D5), mRNA [NM_014566] |
| A_23_P38634 | 0,043097183 | down | -1,5717353 | OR1G1 | Homo sapiens olfactory receptor, family 1, subfamily G, member 1 (OR1G1), mRNA [NM_003555] |
| A_23_P135226 | 6,52E-04 | down | -2,69251 | OR1N2 | Homo sapiens olfactory receptor, family 1, subfamily N, member 2 (OR1N2), mRNA [NM_001004457] |
| A_23_P59783 | 0,019589493 | down | -1,591302 | OR2A14 | Homo sapiens olfactory receptor, family 2, subfamily A, member 14 (OR2A14), mRNA [NM_001001659] |
| A_24_P52293 | 0,010447596 | down | -2,7451172 | OR2A25 | Homo sapiens olfactory receptor, family 2, subfamily A, member 25 (OR2A25), mRNA [NM_001004488] |
| A_33_P3262069 | 0,005720638 | down | -2,7597675 | OR2AG2 | Homo sapiens olfactory receptor, family 2, subfamily AG, member 2 (OR2AG2), mRNA [NM_001004490] |
| A_23_P19428 | 0,021217631 | up | 1,666443 | OR2J2 | Homo sapiens olfactory receptor, family 2, subfamily J, member 2 (OR2J2), mRNA [NM_030905] |
| A_32_P310503 | 0,044940356 | down | -2,1407912 | OR2M2 | Homo sapiens olfactory receptor, family 2, subfamily M, member 2 (OR2M2), mRNA [NM_001004688] |
| A_33_P3260659 | 0,004821391 | down | -2,3211706 | OR2T1 | Homo sapiens olfactory receptor, family 2, subfamily T, member 1 (OR2T1), mRNA [NM_030904] |
| A_33_P3386203 | 0,013911681 | down | -2,0066903 | OR2T10 | Homo sapiens olfactory receptor, family 2, subfamily T, member 10 (OR2T10), mRNA [NM_001004693] |
| A_24_P281439 | 0,029881543 | down | -1,895704 | OR2T5 | Homo sapiens olfactory receptor, family 2, subfamily T, member 5 (OR2T5), mRNA [NM_001004697] |
| A_33_P3260684 | 0,00407762 | down | -2,4415846 | OR2T6 | Homo sapiens olfactory receptor, family 2, subfamily T, member 6 (OR2T6), mRNA [NM_001005471] |
| A_33_P3346529 | 0,035443623 | down | -2,147902 | OR2W5 | Homo sapiens olfactory receptor, family 2, subfamily W, member 5 (gene/pseudogene) (OR2W5), mRNA [NM_001004698] |
| A_33_P3265709 | 0,037284248 | down | -2,1869175 | OR2Y1 | Homo sapiens olfactory receptor, family 2, subfamily Y, member 1 (OR2Y1), mRNA [NM_001001657] |
| A_33_P3249229 | 0,028405152 | down | -2,1856308 | OR3A4P | Homo sapiens olfactory receptor, family 3, subfamily A, member 4 pseudogene (OR3A4P), non-coding RNA [NR_024128] |
| A_33_P3420605 | 0,03833315 | down | -2,3175163 | OR4D1 | Homo sapiens olfactory receptor, family 4, subfamily D, member 1 (OR4D1), mRNA [NM_012374] |
| A_33_P3222469 | 0,046416048 | down | -1,5104042 | OR4K13 | Homo sapiens olfactory receptor, family 4, subfamily K, member 13 (OR4K13), mRNA [NM_001004714] |
| A_33_P3349746 | 0,043825023 | down | -1,5753626 | OR4K2 | Homo sapiens olfactory receptor, family 4, subfamily K, member 2 (OR4K2), mRNA [NM_001005501] |
| A_33_P3349751 | 0,010313837 | down | -1,5279223 | OR4K5 | Homo sapiens olfactory receptor, family 4, subfamily K, member 5 (OR4K5), mRNA [NM_001005483] |
| A_23_P13479 | 0,023037136 | down | -1,939346 | OR4S1 | Homo sapiens olfactory receptor, family 4, subfamily S, member 1 (OR4S1), mRNA [NM_001004725] |
| A_23_P128023 | 0,025714723 | down | -1,6901866 | OR51B4 | Homo sapiens olfactory receptor, family 51, subfamily B, member 4 (OR51B4), mRNA [NM_033179] |
| A_21_P0000762 | 0,025036069 | down | -1,5392176 | OR51B5 | Homo sapiens olfactory receptor, family 51, subfamily B, member 5 (OR51B5), transcript variant 2, non-coding RNA [NR_038321] |
| A_33_P3385696 | 0,01805648 | down | -1,9870719 | OR52D1 | Homo sapiens olfactory receptor, family 52, subfamily D, member 1 (OR52D1), mRNA [NM_001005163] |
| A_33_P3385716 | 0,027173834 | down | -1,5179464 | OR52K1 | Homo sapiens olfactory receptor, family 52, subfamily K, member 1 (OR52K1), mRNA [NM_001005171] |
| A_33_P3363725 | 0,006460224 | down | -1,9825865 | OR52N5 | Homo sapiens olfactory receptor, family 52, subfamily N, member 5 (OR52N5), mRNA [NM_001001922] |
| A_33_P3383246 | 3,16E-04 | down | -2,1851623 | OR52R1 | Homo sapiens olfactory receptor, family 52, subfamily R, member 1 (gene/pseudogene) (OR52R1), mRNA [NM_001005177] |

| | | | | | |
|---|---|---|---|---|---|
| A_33_P3359483 | 0,030861493 | down | -1,6683838 | OR5AC2 | Homo sapiens olfactory receptor, family 5, subfamily AC, member 2 (OR5AC2), mRNA [NM_054106] |
| A_23_P1863 | 0,004353148 | down | -1,633698 | OR5AK2 | Homo sapiens olfactory receptor, family 5, subfamily AK, member 2 (OR5AK2), mRNA [NM_001005323] |
| A_33_P3321462 | 0,034324516 | down | -2,1516585 | OR5B21 | Homo sapiens olfactory receptor, family 5, subfamily B, member 21 (OR5B21), mRNA [NM_001005218] |
| A_33_P3224535 | 0,024968455 | down | -1,6096613 | OR5D18 | Homo sapiens olfactory receptor, family 5, subfamily D, member 18 (OR5D18), mRNA [NM_001001952] |
| A_33_P3362652 | 0,030694908 | down | -1,8591263 | OR5H14 | Homo sapiens olfactory receptor, family 5, subfamily H, member 14 (OR5H14), mRNA [NM_001005514] |
| A_33_P3318631 | 0,04450053 | down | -1,5870581 | OR5H2 | Homo sapiens olfactory receptor, family 5, subfamily H, member 2 (OR5H2), mRNA [NM_001005482] |
| A_33_P3419551 | 0,0297491 | down | -1,683478 | OR5K1 | Homo sapiens olfactory receptor, family 5, subfamily K, member 1 (OR5K1), mRNA [NM_001004736] |
| A_33_P3419562 | 0,018924652 | down | -1,7824667 | OR5K2 | Homo sapiens olfactory receptor, family 5, subfamily K, member 2 (OR5K2), mRNA [NM_001004737] |
| A_23_P42241 | 0,011878522 | down | -1,8668864 | OR5V1 | Homo sapiens olfactory receptor, family 5, subfamily V, member 1 (OR5V1), mRNA [NM_030876] |
| A_33_P3222124 | 0,040385414 | down | -1,528293 | OR6B3 | Homo sapiens olfactory receptor, family 6, subfamily B, member 3 (OR6B3), mRNA [NM_173351] |
| A_33_P3379190 | 0,03627465 | down | -1,5157654 | OR6C4 | Homo sapiens olfactory receptor, family 6, subfamily C, member 4 (OR6C4), mRNA [NM_001005494] |
| A_33_P3330886 | 0,019117502 | down | -1,5555087 | OR6C65 | Homo sapiens olfactory receptor, family 6, subfamily C, member 65 (OR6C65), mRNA [NM_001005518] |
| A_33_P3330881 | 0,043449238 | down | -1,5133283 | OR6C75 | Homo sapiens olfactory receptor, family 6, subfamily C, member 75 (OR6C75), mRNA [NM_001005497] |
| A_23_P51761 | 0,008216991 | down | -5,356401 | OR6K2 | Homo sapiens olfactory receptor, family 6, subfamily K, member 2 (OR6K2), mRNA [NM_001005279] |
| A_33_P3211488 | 0,029434578 | down | -1,640576 | OR6K3 | Homo sapiens olfactory receptor, family 6, subfamily K, member 3 (OR6K3), mRNA [NM_001005327] |
| A_33_P3258206 | 0,044189624 | down | -1,780782 | OR6N2 | Homo sapiens olfactory receptor, family 6, subfamily N, member 2 (OR6N2), mRNA [NM_001005278] |
| A_33_P3259693 | 0,007073275 | down | -4,365351 | OR6P1 | Homo sapiens olfactory receptor, family 6, subfamily P, member 1 (OR6P1), mRNA [NM_001160325] |
| A_23_P85963 | 0,007849532 | down | -2,8825476 | OR6Y1 | Homo sapiens olfactory receptor, family 6, subfamily Y, member 1 (OR6Y1), mRNA [NM_001005189] |
| A_23_P16630 | 0,015471651 | down | -1,6853212 | OR7A5 | Homo sapiens olfactory receptor, family 7, subfamily A, member 5 (OR7A5), mRNA [NM_017506] |
| A_33_P3330418 | 0,028292958 | down | -2,0804675 | OR7A5 | Homo sapiens olfactory receptor, family 7, subfamily A, member 5 (OR7A5), mRNA [NM_017506] |
| A_33_P3254246 | 0,024128508 | down | -1,9104643 | OR7C1 | Homo sapiens olfactory receptor, family 7, subfamily C, member 1 (OR7C1), mRNA [NM_198944] |
| A_33_P3300610 | 0,022892961 | down | -1,8640034 | OR7G2 | Homo sapiens olfactory receptor, family 7, subfamily G, member 2 (OR7G2), mRNA [NM_001005193] |
| A_23_P127662 | 0,007351937 | down | -2,6807091 | OR8D1 | Homo sapiens olfactory receptor, family 8, subfamily D, member 1 (OR8D1), mRNA [NM_001002917] |
| A_23_P13195 | 0,030000508 | down | -1,9745032 | OR8G1 | Homo sapiens olfactory receptor, family 8, subfamily G, member 1 (gene/pseudogene) (OR8G1), transcript variant 1, mRNA [NM_001 |
| A_23_P75707 | 0,01577365 | down | -2,4640841 | OR8H1 | Homo sapiens olfactory receptor, family 8, subfamily H, member 1 (OR8H1), mRNA [NM_001005199] |
| A_33_P3305388 | 0,008085789 | down | -1,750339 | OR8K5 | Homo sapiens olfactory receptor, family 8, subfamily K, member 5 (OR8K5), mRNA [NM_001004058] |
| A_23_P24676 | 0,010718057 | down | -2,850212 | OR8U1 | Homo sapiens olfactory receptor, family 8, subfamily U, member 1 (OR8U1), mRNA [NM_001005204] |
| A_33_P3252583 | 0,035934936 | down | -1,714341 | OR9I1 | Homo sapiens olfactory receptor, family 9, subfamily I, member 1 (OR9I1), mRNA [NM_001005211] |
| A_32_P52519 | 0,039870366 | down | -2,125239 | OTOA | Homo sapiens otoancorin (OTOA), transcript variant 2, mRNA [NM_170664] |
| A_33_P3340260 | 0,04455294 | down | -2,3873303 | OTOG | Homo sapiens otogelin (OTOG), transcript variant 1, mRNA [NM_001277269] |
| A_23_P90997 | 0,01919352 | down | -1,5597974 | OTOS | Homo sapiens otospiralin (OTOS), mRNA [NM_148961] |
| A_21_P0013524 | 0,015546041 | down | -1,7492126 | OTUD6B-AS1 | PREDICTED: Homo sapiens uncharacterized LOC100506365 (GS1-251I9.4), transcript variant X3, ncRNA [XR_432334] |
| A_33_P3222203 | 0,046216812 | down | -1,5790596 | OXER1 | Homo sapiens oxoeicosanoid (OXE) receptor 1 (OXER1), mRNA [NM_148962] |
| A_23_P413760 | 0,00141387 | down | -2,0794134 | P2RX5 | Homo sapiens purinergic receptor P2X, ligand gated ion channel, 5 (P2RX5), transcript variant 2, mRNA [NM_175080] |
| A_24_P319113 | 0,016156284 | down | -1,7449347 | P2RX7 | Homo sapiens purinergic receptor P2X, ligand-gated ion channel, 7 (P2RX7), transcript variant 1, mRNA [NM_002562] |
| A_33_P3838128 | 0,004812387 | down | -2,7561374 | PABPC5-AS1 | 5000ECD06 Fetal Brain 18 Homo sapiens cDNA 5', mRNA sequence [DR033788] |
| A_33_P3358213 | 0,003705944 | down | -2,299844 | PADI6 | Homo sapiens peptidyl arginine deiminase, type VI (PADI6), mRNA [NM_207421] |
| A_24_P254506 | 0,016567204 | down | -1,8143921 | PAGE4 | Homo sapiens P antigen family, member 4 (prostate associated) (PAGE4), mRNA [NM_007003] |
| A_23_P161481 | 0,045414384 | down | -1,5948199 | PALD1 | Homo sapiens phosphatase domain containing, paladin 1 (PALD1), mRNA [NM_014431] |
| A_33_P3258061 | 0,008578483 | down | -1,6063448 | PALM3 | Homo sapiens paralemmin 3 (PALM3), mRNA [NM_001145028] |
| A_33_P3258056 | 0,002306606 | down | -2,869416 | PALM3 | Homo sapiens paralemmin 3 (PALM3), mRNA [NM_001145028] |
| A_23_P87072 | 0,001147769 | down | -2,2309823 | PANX3 | Homo sapiens pannexin 3 (PANX3), mRNA [NM_052959] |
| A_21_P0004005 | 0,013925897 | down | -1,6637568 | PART1 | Homo sapiens prostate androgen-regulated transcript 1 (non-protein coding) (PART1), transcript variant 3, long non-coding RNA [NR |
| A_23_P404059 | 0,04943351 | down | -1,8155209 | PASD1 | Homo sapiens PAS domain containing 1 (PASD1), mRNA [NM_173493] |
| A_33_P3381378 | 0,046911106 | down | -1,8302609 | PAX1 | Homo sapiens paired box 1 (PAX1), transcript variant 2, mRNA [NM_001257096] |
| A_23_P209499 | 0,04743609 | down | -2,408312 | PAX3 | Homo sapiens paired box 3 (PAX3), transcript variant PAX3D, mRNA [NM_181458] |
| A_23_P500985 | 0,022460757 | down | -1,786175 | PAX7 | Homo sapiens paired box 7 (PAX7), transcript variant 2, mRNA [NM_013945] |
| A_21_P0000824 | 0,02732842 | down | -1,6523719 | PAXBP1-AS1 | Homo sapiens PAXBP1 antisense RNA 1 (PAXBP1-AS1), transcript variant 2, long non-coding RNA [NR_038880] |
| A_33_P3297040 | 0,004619776 | down | -2,057278 | PBOV1 | Homo sapiens prostate and breast cancer overexpressed 1 (PBOV1), mRNA [NM_021635] |

| | | | | | |
|---|---|---|---|---|---|
| A_24_P247849 | 0,031391047 | down | -1,7146211 | PBX1 | pre-B-cell leukemia homeobox 1 [Source:HGNC Symbol;Acc:HGNC:8632] [ENST00000474046] |
| A_23_P90419 | 0,03150541 | up | 1,7349693 | PBX4 | Homo sapiens pre-B-cell leukemia homeobox 4 (PBX4), transcript variant 1, mRNA [NM_025245] |
| A_24_P111147 | 0,008438322 | down | -1,6621444 | PCDH15 | protocadherin-related 15 [Source:HGNC Symbol;Acc:HGNC:14674] [ENST00000373955] |
| A_23_P161331 | 0,0409735 | down | -1,6947165 | PCDH15 | Homo sapiens protocadherin-related 15 (PCDH15), transcript variant C, mRNA [NM_033056] |
| A_33_P3250348 | 0,032262944 | down | -1,5183926 | PCDH18 | Homo sapiens protocadherin 18 (PCDH18), transcript variant 1, mRNA [NM_019035] |
| A_33_P3423230 | 0,019828225 | down | -1,601106 | PCDH19 | Homo sapiens protocadherin 19 (PCDH19), transcript variant 3, mRNA [NM_001184880] |
| A_24_P419039 | 5,07E-04 | down | -2,4353812 | PCDH19 | Homo sapiens protocadherin 19 (PCDH19), transcript variant 2, mRNA [NM_020766] |
| A_33_P3258546 | 0,004212022 | down | -2,5884864 | PCDHA5 | Homo sapiens protocadherin alpha 5 (PCDHA5), transcript variant 2, mRNA [NM_031501] |
| A_23_P144627 | 0,006557134 | down | -1,5764344 | PCDHB13 | Homo sapiens protocadherin beta 13 (PCDHB13), mRNA [NM_018933] |
| A_23_P58464 | 0,034339726 | down | -1,567382 | PCDHB6 | Homo sapiens protocadherin beta 6 (PCDHB6), transcript variant 1, mRNA [NM_018939] |
| A_23_P303101 | 0,002023251 | down | -2,8505929 | PCDHGC4 | Homo sapiens protocadherin gamma subfamily C, 4 (PCDHGC4), transcript variant 2, mRNA [NM_032406] |
| A_33_P3380361 | 0,03516438 | down | -1,714423 | PCGEM1 | Homo sapiens PCGEM1, prostate-specific transcript (non-protein coding) (PCGEM1), long non-coding RNA [NR_002769] |
| A_32_P109876 | 0,001757039 | down | -2,1307557 | PCLO | Homo sapiens piccolo presynaptic cytomatrix protein (PCLO), transcript variant 1, mRNA [NM_033026] |
| A_23_P136405 | 0,008593064 | down | -1,5554755 | PDCD1 | Homo sapiens programmed cell death 1 (PDCD1), mRNA [NM_005018] |
| A_23_P363301 | 0,035008356 | down | -2,0084164 | PDCL2 | Homo sapiens phosducin-like 2 (PDCL2), mRNA [NM_152401] |
| A_32_P116857 | 0,024232931 | down | -1,5943699 | PDE11A | Homo sapiens phosphodiesterase 11A (PDE11A), transcript variant 4, mRNA [NM_016953] |
| A_24_P43144 | 0,028661901 | down | -1,9567113 | PDE11A | Homo sapiens phosphodiesterase 11A (PDE11A), transcript variant 2, mRNA [NM_001077358] |
| A_24_P208436 | 0,001804135 | down | -3,6961048 | PDE1A | Homo sapiens phosphodiesterase 1A, calmodulin-dependent (PDE1A), transcript variant 2, mRNA [NM_001003683] |
| A_23_P74278 | 0,045435812 | down | -2,1193247 | PDE4B | Homo sapiens phosphodiesterase 4B, cAMP-specific (PDE4B), transcript variant d, mRNA [NM_001037341] |
| A_33_P3759611 | 0,04839397 | down | -1,883238 | PDE4C | Homo sapiens phosphodiesterase 4C, cAMP-specific (PDE4C), transcript variant 1, mRNA [NM_000923] |
| A_23_P10743 | 0,016055655 | down | -2,5153842 | PDE6B | Homo sapiens phosphodiesterase 6B, cGMP-specific, rod, beta (PDE6B), transcript variant 1, mRNA [NM_000283] |
| A_23_P98070 | 0,021761954 | down | -2,0409334 | PDE6C | Homo sapiens phosphodiesterase 6C, cGMP-specific, cone, alpha prime (PDE6C), mRNA [NM_006204] |
| A_24_P70906 | 0,02700309 | down | -1,7115198 | PDILT | Homo sapiens protein disulfide isomerase-like, testis expressed (PDILT), mRNA [NM_174924] |
| A_23_P65189 | 0,021605043 | down | -1,7435932 | PDX1 | Homo sapiens pancreatic and duodenal homeobox 1 (PDX1), mRNA [NM_000209] |
| A_33_P3229402 | 0,046958726 | down | -1,5581459 | PECAM1 | Homo sapiens platelet/endothelial cell adhesion molecule 1 (PECAM1), mRNA [NM_000442] |
| A_23_P252471 | 0,029860396 | down | -1,58969 | PECAM1 | Homo sapiens platelet/endothelial cell adhesion molecule 1 (PECAM1), mRNA [NM_000442] |
| A_33_P3271325 | 0,007715667 | down | -2,5816746 | PER4 | Homo sapiens Per4 pseudogene, mRNA sequence. [AF348410] |
| A_23_P259003 | 0,040193457 | down | -1,7555931 | PEX5L | Homo sapiens peroxisomal biogenesis factor 5-like (PEX5L), transcript variant 1, mRNA [NM_016559] |
| A_24_P120907 | 0,033085685 | down | -1,6302824 | PGM5 | Homo sapiens phosphoglucomutase 5 (PGM5), mRNA [NM_021965] |
| A_21_P0011953 | 0,003449355 | down | -2,1517708 | PGM5P3-AS1 | Homo sapiens PGM5P3 antisense RNA 1 (PGM5P3-AS1), transcript variant 3, long non-coding RNA [NR_121190] |
| A_21_P0012084 | 0,047295164 | down | -1,6837814 | PGM5P4-AS1 | Homo sapiens PGM5P4 antisense RNA 1 (PGM5P4-AS1), transcript variant 1, long non-coding RNA [NR_121185] |
| A_23_P138938 | 0,03896207 | down | -1,7553858 | PGR | Homo sapiens progesterone receptor (PGR), transcript variant 2, mRNA [NM_000926] |
| A_32_P49199 | 0,03604469 | down | -2,151365 | PGR | Homo sapiens progesterone receptor (PGR), transcript variant 2, mRNA [NM_000926] |
| A_33_P3333627 | 0,011927024 | up | 1,6296293 | PHACTR1 | Homo sapiens phosphatase and actin regulator 1 (PHACTR1), transcript variant 1, mRNA [NM_030948] |
| A_33_P3294302 | 0,001482816 | down | -2,2683 | PHLDA1 | pleckstrin homology-like domain, family A, member 1 [Source:HGNC Symbol;Acc:HGNC:8933] [ENST00000266671] |
| A_33_P3884610 | 0,035532076 | down | -1,6387417 | PIP5K1P1 | Homo sapiens phosphatidylinositol-4-phosphate 5-kinase, type I, pseudogene 1 (PIP5K1P1), non-coding RNA [NR_027712] |
| A_24_P221858 | 0,009242502 | down | -1,9021239 | PIWIL3 | Homo sapiens piwi-like RNA-mediated gene silencing 3 (PIWIL3), transcript variant 1, mRNA [NM_001008496] |
| A_32_P92489 | 0,014702252 | down | -1,6848819 | PKD1L2 | Homo sapiens polycystic kidney disease 1-like 2 (gene/pseudogene) (PKD1L2), transcript variant 1, mRNA [NM_052892] |
| A_23_P129332 | 0,042786203 | down | -1,9189802 | PKD1L2 | Homo sapiens polycystic kidney disease 1-like 2 (gene/pseudogene) (PKD1L2), transcript variant 3, mRNA [NM_001076780] |
| A_33_P3254136 | 0,029659249 | down | -1,50774 | PKHD1L1 | Homo sapiens polycystic kidney and hepatic disease 1 (autosomal recessive)-like 1 (PKHD1L1), mRNA [NM_177531] |
| A_23_P145529 | 0,0375816 | up | 1,6652653 | PKIB | Homo sapiens protein kinase (cAMP-dependent, catalytic) inhibitor beta (PKIB), transcript variant 1, mRNA [NM_181795] |
| A_33_P3242829 | 0,006623486 | up | 1,9147525 | PKN2 | protein kinase N2 [Source:HGNC Symbol;Acc:HGNC:9406] [ENST00000370505] |
| A_23_P300100 | 0,04553141 | down | -1,9364897 | PLA2G2D | phospholipase A2, group IID [Source:HGNC Symbol;Acc:HGNC:9033] [ENST00000375105] |
| A_21_P0008628 | 0,01404575 | down | -1,5317932 | PLA2G4E-AS1 | Homo sapiens PLA2G4E antisense RNA 1 (PLA2G4E-AS1), long non-coding RNA [NR_120334] |
| A_23_P16469 | 0,03658775 | up | 2,0255094 | PLAUR | Homo sapiens plasminogen activator, urokinase receptor (PLAUR), transcript variant 3, mRNA [NM_001005377] |
| A_33_P3381623 | 0,031122813 | down | -2,1630507 | PLB1 | Homo sapiens phospholipase B1 (PLB1), transcript variant 1, mRNA [NM_153021] |
| A_21_P0014399 | 0,014517419 | down | -2,2547927 | PLCG1-AS1 | Homo sapiens PLCG1 antisense RNA 1 (PLCG1-AS1), long non-coding RNA [NR_109889] |
| A_33_P3334180 | 0,008099051 | down | -2,2826352 | PLCH2 | phospholipase C, eta 2 [Source:HGNC Symbol;Acc:HGNC:29037] [ENST00000473964] |
| A_21_P0014779 | 0,005460852 | down | -2,3653927 | PLIN2 | perilipin 2 [Source:HGNC Symbol;Acc:HGNC:248] [ENST00000494753] |

| | | | | | |
|---|---|---|---|---|---|
| A_33_P3209885 | 0,033807635 | down | -1,9375224 | PLXDC1 | Homo sapiens plexin domain containing 1 (PLXDC1), mRNA [NM_020405] |
| A_23_P321223 | 0,049739998 | down | -1,9099728 | PMCH | Homo sapiens pro-melanin-concentrating hormone (PMCH), mRNA [NM_002674] |
| A_23_P24083 | 0,022197308 | down | -1,8066955 | PNLIPRP2 | Homo sapiens pancreatic lipase-related protein 2 (PNLIPRP2), transcript variant 1, mRNA [NM_005396] |
| A_33_P3328863 | 9,70E-04 | down | -2,1913567 | PNMAL2 | Homo sapiens paraneoplastic Ma antigen family-like 2 (PNMAL2), mRNA [NM_020709] |
| A_23_P218827 | 0,024349459 | down | -1,7427821 | POLQ | Homo sapiens polymerase (DNA directed), theta (POLQ), mRNA [NM_199420] |
| A_33_P3273819 | 0,032691617 | down | -1,6763145 | POT1-AS1 | Homo sapiens POT1 antisense RNA 1 (POT1-AS1), transcript variant 2, long non-coding RNA [NR_125719] |
| A_23_P420348 | 0,030440873 | down | -1,6018723 | POTED | Homo sapiens POTE ankyrin domain family, member D (POTED), mRNA [NM_174981] |
| A_24_P825874 | 0,042660728 | down | -1,818898 | POTEI | Homo sapiens POTE ankyrin domain family, member I (POTEI), mRNA [NM_001277406] |
| A_23_P125505 | 0,02732394 | down | -2,2345095 | PPEF1 | Homo sapiens protein phosphatase, EF-hand calcium binding domain 1 (PPEF1), transcript variant 1, mRNA [NM_006240] |
| A_23_P402936 | 0,03402005 | down | -2,4539344 | PPFIA2 | Homo sapiens protein tyrosine phosphatase, receptor type, f polypeptide (PTPRF), interacting protein (liprin), alpha 2 (PPFIA2), trans |
| A_33_P3239122 | 0,003219153 | down | -1,9094028 | PPIH | peptidylprolyl isomerase H (cyclophilin H) [Source:HGNC Symbol;Acc:HGNC:14651] [ENST00000372550] |
| A_23_P352535 | 0,012496718 | down | -1,8338985 | PPP1R16B | Homo sapiens protein phosphatase 1, regulatory subunit 16B (PPP1R16B), transcript variant 1, mRNA [NM_015568] |
| A_24_P13285 | 0,03417862 | down | -2,4174519 | PPP1R1A | Homo sapiens protein phosphatase 1, regulatory (inhibitor) subunit 1A (PPP1R1A), mRNA [NM_006741] |
| A_33_P3383471 | 2,56E-04 | down | -2,637356 | PPP1R1A | Homo sapiens cDNA FLJ41975 fis, clone SKNMC2006998, moderately similar to PROTEIN PHOSPHATASE INHIBITOR 1. [AK12396 |
| A_23_P98571 | 0,020380264 | up | 1,557742 | PPP1R32 | Homo sapiens protein phosphatase 1, regulatory subunit 32 (PPP1R32), transcript variant 1, mRNA [NM_145017] |
| A_33_P3213179 | 0,016068015 | down | -1,7629584 | PPP1R3A | Homo sapiens protein phosphatase 1, regulatory subunit 3A (PPP1R3A), mRNA [NM_002711] |
| A_24_P177604 | 0,01069072 | down | -1,5203142 | PPP1R3F | Homo sapiens protein phosphatase 1, regulatory subunit 3F (PPP1R3F), transcript variant 1, mRNA [NM_033215] |
| A_23_P207336 | 0,020148592 | down | -2,1200163 | PPY | Homo sapiens pancreatic polypeptide (PPY), mRNA [NM_002722] |
| A_33_P3308456 | 0,003020654 | down | -2,3672547 | PRAC2 | Homo sapiens prostate cancer susceptibility candidate 2 (PRAC2), transcript variant 2, mRNA [NM_001282275] |
| A_33_P3303594 | 0,033531234 | down | -1,5788069 | PRAMEF11 | Homo sapiens PRAME family member 11 (PRAMEF11), mRNA [NM_001146344] |
| A_23_P126658 | 0,00163893 | down | -1,9134471 | PRAMEF8 | Homo sapiens PRAME family member 8 (PRAMEF8), mRNA [NM_001012276] |
| A_23_P256581 | 0,020443026 | down | -1,5253493 | PRDM13 | Homo sapiens PR domain containing 13 (PRDM13), mRNA [NM_021620] |
| A_23_P123488 | 0,033076752 | down | -1,6322678 | PRDM14 | Homo sapiens PR domain containing 14 (PRDM14), mRNA [NM_024504] |
| A_33_P3394993 | 0,011087554 | down | -3,464921 | PREX2 | Homo sapiens phosphatidylinositol-3,4,5-trisphosphate-dependent Rac exchange factor 2 (PREX2), transcript variant 2, mRNA [NM_ |
| A_23_P1473 | 0,00211822 | down | -2,901628 | PRF1 | Homo sapiens perforin 1 (pore forming protein) (PRF1), transcript variant 1, mRNA [NM_005041] |
| A_23_P47466 | 0,021974158 | down | -1,7820842 | PRG3 | Homo sapiens proteoglycan 3 (PRG3), mRNA [NM_006093] |
| A_33_P3369178 | 0,0368909 | down | -1,5547136 | PRG4 | Homo sapiens proteoglycan 4 (PRG4), transcript variant A, mRNA [NM_005807] |
| A_23_P16189 | 0,012960743 | down | -2,2491133 | PRKCG | Homo sapiens protein kinase C, gamma (PRKCG), mRNA [NM_002739] |
| A_21_P0006825 | 0,030877028 | down | -1,7936928 | PRKCQ-AS1 | PRKCQ antisense RNA 1 [Source:HGNC Symbol;Acc:HGNC:44689] [ENST00000608526] |
| A_24_P250765 | 0,012637203 | down | -2,5618713 | PRKG1 | Homo sapiens protein kinase, cGMP-dependent, type I (PRKG1), transcript variant 2, mRNA [NM_006258] |
| A_33_P3416757 | 0,032219727 | down | -1,5744605 | PRLR | Homo sapiens prolactin receptor (PRLR), transcript variant 2, mRNA [NM_001204315] |
| A_23_P167468 | 0,026686197 | down | -1,6078331 | PRLR | Homo sapiens prolactin receptor (PRLR), transcript variant 1, mRNA [NM_000949] |
| A_23_P100189 | 0,036417995 | down | -1,5783534 | PRM1 | Homo sapiens protamine 1 (PRM1), mRNA [NM_002761] |
| A_33_P3389842 | 0,024132708 | down | -1,590863 | PROM1 | Homo sapiens prominin 1 (PROM1), transcript variant 6, mRNA [NM_001145850] |
| A_23_P140074 | 0,028099801 | down | -1,5098361 | PROZ | Homo sapiens protein Z, vitamin K-dependent plasma glycoprotein (PROZ), transcript variant 2, mRNA [NM_003891] |
| A_23_P214459 | 0,029717173 | down | -2,1787717 | PRPH2 | Homo sapiens peripherin 2 (retinal degeneration, slow) (PRPH2), mRNA [NM_000322] |
| A_33_P3334708 | 0,029183386 | down | -1,6497078 | PRR23A | Homo sapiens proline rich 23A (PRR23A), mRNA [NM_001134659] |
| A_23_P351328 | 0,001161686 | down | -3,0344086 | PRR30 | Homo sapiens proline rich 30 (PRR30), mRNA [NM_178553] |
| A_23_P387537 | 0,003706914 | down | -2,592161 | PRRG3 | Homo sapiens proline rich Gla (G-carboxyglutamic acid) 3 (transmembrane) (PRRG3), transcript variant 1, mRNA [NM_024082] |
| A_23_P400298 | 0,034459036 | up | 1,6042997 | PRSS22 | Homo sapiens protease, serine, 22 (PRSS22), mRNA [NM_022119] |
| A_23_P350719 | 4,59E-04 | up | 2,4456582 | PRSS30P | Homo sapiens protease, serine, 30, pseudogene (PRSS30P), non-coding RNA [NR_026864] |
| A_24_P327084 | 0,034861688 | down | -2,00523 | PRSS33 | Homo sapiens protease, serine, 33 (PRSS33), mRNA [NM_152891] |
| A_33_P3332166 | 0,042544346 | down | -2,235401 | PRSS38 | Homo sapiens protease, serine, 38 (PRSS38), mRNA [NM_183062] |
| A_23_P406025 | 0,021081101 | down | -2,046563 | PRUNE2 | Homo sapiens prune homolog 2 (Drosophila) (PRUNE2), mRNA [NM_015225] |
| A_23_P11408 | 6,76E-04 | down | -2,7966206 | PRY2 | Homo sapiens PTPN13-like, Y-linked 2 (PRY2), mRNA [NM_001002758] |
| A_33_P3383004 | 0,028017398 | down | -1,5436888 | PSAPL1 | Homo sapiens prosaposin-like 1 (gene/pseudogene) (PSAPL1), mRNA [NM_001085382] |
| A_23_P355311 | 0,0201157 | down | -2,0062895 | PTCH2 | Homo sapiens patched 2 (PTCH2), transcript variant 1, mRNA [NM_003738] |
| A_23_P103328 | 0,014799698 | down | -1,5368288 | PTGER3 | Homo sapiens prostaglandin E receptor 3 (subtype EP3) (PTGER3), transcript variant 4, mRNA [NM_198714] |
| A_24_P135875 | 0,03493849 | down | -2,0114586 | PTGER4P2-CDK2AP2P2 | Homo sapiens PTGER4P2-CDK2AP2P2 readthrough transcribed pseudogene (PTGER4P2-CDK2AP2P2), non-coding RNA [NR_02 |

| | | | | | |
|---|---|---|---|---|---|
| A_23_P12392 | 0,021553507 | down | -1,5038058 | PTPRC | Homo sapiens protein tyrosine phosphatase, receptor type, C (PTPRC), transcript variant 4, non-coding RNA [NR_052021] |
| A_33_P3364811 | 0,009837327 | down | -2,0276864 | PTPRC | Homo sapiens protein tyrosine phosphatase, receptor type, C (PTPRC), transcript variant 1, mRNA [NM_002838] |
| A_24_P639665 | 0,03575465 | down | -1,5028753 | PTPRD-AS1 | Homo sapiens PTPRD antisense RNA 1 (PTPRD-AS1), transcript variant 1, long non-coding RNA [NR_121599] |
| A_33_P3344911 | 0,046657693 | down | -1,65717 | PTPRQ | Homo sapiens protein tyrosine phosphatase, receptor type, Q (PTPRQ), mRNA [NM_001145026] |
| A_23_P111919 | 0,044083185 | down | -1,5362079 | PURG | Homo sapiens purine-rich element binding protein G (PURG), transcript variant A, mRNA [NM_013357] |
| A_21_P0014107 | 0,03620525 | down | -1,7610126 | PVRL3-AS1 | Homo sapiens PVRL3 antisense RNA 1 (PVRL3-AS1), long non-coding RNA [NR_045114] |
| A_33_P3352293 | 0,0342977 | down | -1,5516156 | PWRN2 | Homo sapiens Prader-Willi region non-protein coding RNA 2 (PWRN2), long non-coding RNA [NR_026647] |
| A_23_P333022 | 0,01464301 | down | -1,5140634 | PXT1 | Homo sapiens peroxisomal, testis specific 1 (PXT1), mRNA [NM_152990] |
| A_33_P3307568 | 0,011544356 | down | -1,9397079 | PYDC2 | Homo sapiens pyrin domain containing 2 (PYDC2), mRNA [NM_001083308] |
| A_24_P233078 | 0,003553038 | down | -1,5936139 | PYY2 | Homo sapiens peptide YY, 2 (pseudogene) (PYY2), non-coding RNA [NR_003064] |
| A_33_P3257866 | 0,020632543 | down | -1,7823973 | PYY2 | Homo sapiens peptide YY, 2 (pseudogene) (PYY2), non-coding RNA [NR_003064] |
| A_23_P9565 | 0,034188855 | down | -1,9924444 | RAB33B | Homo sapiens RAB33B, member RAS oncogene family (RAB33B), mRNA [NM_031296] |
| A_33_P3665739 | 0,006331355 | down | -2,84996 | RAB43 | AGENCOURT_6606420 NIH_MGC_106 Homo sapiens cDNA clone IMAGE:5483928 5', mRNA sequence [BM917410] |
| A_21_P0014768 | 0,009542707 | up | 1,5444516 | RABGEF1 | Homo sapiens RAB guanine nucleotide exchange factor (GEF) 1 (RABGEF1), transcript variant 1, mRNA [NM_001287060] |
| A_23_P94141 | 0,033640523 | down | -1,5639945 | RAD54B | Homo sapiens RAD54 homolog B (S. cerevisiae) (RAD54B), transcript variant 2, mRNA [NM_001205262] |
| A_33_P3361227 | 0,031787764 | down | -1,523814 | RAD9B | RAD9 homolog B (S. pombe) [Source:HGNC Symbol;Acc:HGNC:21700] [ENST00000409461] |
| A_23_P111860 | 0,027753724 | down | -1,5966492 | RADIL | Homo sapiens Ras association and DIL domains (RADIL), mRNA [NM_018059] |
| A_23_P64525 | 0,013250612 | down | -1,7757229 | RAG2 | Homo sapiens recombination activating gene 2 (RAG2), transcript variant 1, mRNA [NM_000536] |
| A_24_P116710 | 0,024685333 | down | -1,5919541 | RAMP2 | Homo sapiens receptor (G protein-coupled) activity modifying protein 2 (RAMP2), mRNA [NM_005854] |
| A_33_P3390367 | 0,046583578 | down | -1,5535983 | RAPGEF4-AS1 | Homo sapiens RAPGEF4 antisense RNA 1 (RAPGEF4-AS1), long non-coding RNA [NR_026995] |
| A_33_P3209950 | 6,31E-04 | down | -1,975907 | RASGRP2 | RAS guanyl releasing protein 2 (calcium and DAG-regulated) [Source:HGNC Symbol;Acc:HGNC:9879] [ENST00000377494] |
| A_24_P288448 | 0,00798671 | down | -2,8284748 | RASSF2 | Ras association (RalGDS/AF-6) domain family member 2 [Source:HGNC Symbol;Acc:HGNC:9883] [ENST00000478553] |
| A_21_P0013891 | 0,007988425 | down | -1,6082373 | RBMY1B | Homo sapiens RNA binding motif protein, Y-linked, family 1, member B (RBMY1B), mRNA [NM_001006121] |
| A_24_P400604 | 0,03346542 | down | -1,6095589 | RBMY1B | Homo sapiens RNA binding motif protein, Y-linked, family 1, member B (RBMY1B), mRNA [NM_001006121] |
| A_33_P3382648 | 0,03444707 | down | -2,2658005 | RBMY1B | Homo sapiens RNA binding motif protein, Y-linked, family 1, member B (RBMY1B), mRNA [NM_001006121] |
| A_24_P357406 | 0,036492184 | down | -3,09754 | RBMY1B | Homo sapiens RNA binding motif protein, Y-linked, family 1, member B (RBMY1B), mRNA [NM_001006121] |
| A_24_P794648 | 0,04699977 | down | -1,9063456 | RBMY2EP | Homo sapiens RNA binding motif protein, Y-linked, family 2, member E pseudogene (RBMY2EP), non-coding RNA [NR_001574] |
| A_33_P3332180 | 0,015776986 | down | -1,6796435 | RBP3 | Homo sapiens retinol binding protein 3, interstitial (RBP3), mRNA [NM_002900] |
| A_23_P389500 | 0,016380938 | down | -2,0475123 | REG1B | Homo sapiens regenerating islet-derived 1 beta (REG1B), mRNA [NM_006507] |
| A_23_P119936 | 0,00250462 | down | -2,6922388 | REG3A | Homo sapiens regenerating islet-derived 3 alpha (REG3A), transcript variant 2, mRNA [NM_138938] |
| A_32_P65628 | 0,040335067 | down | -1,5178937 | REG3G | Homo sapiens regenerating islet-derived 3 gamma (REG3G), transcript variant 1, mRNA [NM_001008387] |
| A_33_P3259861 | 4,70E-04 | down | -1,6689343 | REREP3 | Homo sapiens arginine-glutamic acid dipeptide (RE) repeats pseudogene 3 (REREP3), non-coding RNA [NR_033735] |
| A_24_P321525 | 0,019384399 | down | -2,7674422 | RERG | Homo sapiens RAS-like, estrogen-regulated, growth inhibitor (RERG), transcript variant 1, mRNA [NM_032918] |
| A_33_P3562537 | 0,009784841 | down | -1,727199 | RET | Homo sapiens ret proto-oncogene (RET), transcript variant 4, mRNA [NM_020630] |
| A_23_P92196 | 0,008156353 | down | -2,1651845 | RETNLB | Homo sapiens resistin like beta (RETNLB), mRNA [NM_032579] |
| A_32_P118568 | 0,04419269 | down | -1,7633426 | RFPL1S | Homo sapiens RFPL1 antisense RNA 1 (RFPL1S), antisense RNA [NR_002727] |
| A_33_P3397835 | 0,028696736 | down | -1,5436453 | RFPL4B | Homo sapiens ret finger protein-like 4B (RFPL4B), mRNA [NM_001013734] |
| A_21_P0014387 | 0,0481745 | up | 1,9874269 | RFX2 | regulatory factor X, 2 (influences HLA class II expression) [Source:HGNC Symbol;Acc:HGNC:9983] [ENST00000587700] |
| A_33_P3281741 | 4,93E-05 | down | -3,6344013 | RFX8 | Homo sapiens RFX family member 8, lacking RFX DNA binding domain (RFX8), mRNA [NM_001145664] |
| A_33_P3379606 | 0,004223689 | down | -2,1260705 | RGS7BP | Homo sapiens regulator of G-protein signaling 7 binding protein (RGS7BP), transcript variant 1, mRNA [NM_001029875] |
| A_21_P0000057 | 0,04901155 | down | -1,7914121 | RGS9 | Homo sapiens regulator of G-protein signaling 9 (RGS9), transcript variant 3, mRNA [NM_001165933] |
| A_33_P3364661 | 0,004959233 | down | -1,7044164 | RHOA | ras homolog family member A [Source:HGNC Symbol;Acc:HGNC:667] [ENST00000265538] |
| A_24_P162226 | 0,004750913 | down | -1,9699838 | RIMBP2 | Homo sapiens RIMS binding protein 2 (RIMBP2), mRNA [NM_015347] |
| A_32_P38645 | 0,028343802 | down | -2,2389953 | RIMS4 | Homo sapiens regulating synaptic membrane exocytosis 4 (RIMS4), transcript variant 2, mRNA [NM_182970] |
| A_33_P3303291 | 0,003236403 | up | 1,6796671 | RIPK3 | Homo sapiens receptor-interacting serine-threonine kinase 3 (RIPK3), mRNA [NM_006871] |
| A_24_P99066 | 0,01730723 | down | -1,611368 | RNF17 | Homo sapiens ring finger protein 17 (RNF17), transcript variant 1, mRNA [NM_031277] |
| A_23_P126248 | 3,00E-04 | down | -2,6069722 | RNF186 | Homo sapiens ring finger protein 186 (RNF186), mRNA [NM_019062] |
| A_33_P3324775 | 0,020811899 | down | -1,7813789 | RORB-AS1 | Homo sapiens RORB antisense RNA 1 (RORB-AS1), long non-coding RNA [NR_125791] |
| A_33_P3421028 | 0,017858334 | down | -1,6955128 | ROS1 | ROS proto-oncogene 1 , receptor tyrosine kinase [Source:HGNC Symbol;Acc:HGNC:10261] [ENST00000403284] |

| | | | | | |
|---|---|---|---|---|---|
| A_33_P3249329 | 0,045990188 | down | -2,0362694 | RPEL1 | Homo sapiens ribulose-5-phosphate-3-epimerase-like 1 (RPEL1), mRNA [NM_001143909] |
| A_33_P3226420 | 0,01062754 | down | -2,2720504 | RSAD2 | radical S-adenosyl methionine domain containing 2 [Source:HGNC Symbol;Acc:HGNC:30908] [ENST00000474872] |
| A_23_P130653 | 0,04245527 | down | -1,5393481 | RTBDN | Homo sapiens retbindin (RTBDN), transcript variant 2, mRNA [NM_031429] |
| A_23_P432056 | 0,00145678 | down | -1,9153086 | RTN4RL1 | Homo sapiens reticulon 4 receptor-like 1 (RTN4RL1), mRNA [NM_178568] |
| A_33_P3296997 | 0,032993436 | up | 1,5298659 | RUFY4 | Homo sapiens RUN and FYVE domain containing 4 (RUFY4), transcript variant 1, mRNA [NM_198483] |
| A_23_P23292 | 0,011359981 | down | -1,8885309 | RXRG | Homo sapiens retinoid X receptor, gamma (RXRG), transcript variant 1, mRNA [NM_006917] |
| A_33_P3409086 | 0,016069941 | down | -1,9586259 | S100A1 | Homo sapiens S100 calcium binding protein A1 (S100A1), mRNA [NM_006271] |
| A_23_P143526 | 0,01568885 | down | -1,7065295 | S100B | Homo sapiens S100 calcium binding protein B (S100B), mRNA [NM_006272] |
| A_21_P0014499 | 0,04908468 | down | -1,8375996 | SACS-AS1 | Homo sapiens SACS antisense RNA 1 (SACS-AS1), long non-coding RNA [NR_103450] |
| A_23_P5853 | 0,017460572 | down | -1,6240827 | SAG | Homo sapiens S-antigen; retina and pineal gland (arrestin) (SAG), mRNA [NM_000541] |
| A_23_P21943 | 5,22E-04 | down | -2,4187655 | SAGE1 | Homo sapiens sarcoma antigen 1 (SAGE1), mRNA [NM_018666] |
| A_23_P349025 | 0,012938356 | down | -1,5576007 | SAMD15 | Homo sapiens sterile alpha motif domain containing 15 (SAMD15), mRNA [NM_001010860] |
| A_24_P383523 | 0,032800946 | up | 1,6207551 | SAMD4A | Homo sapiens sterile alpha motif domain containing 4A (SAMD4A), transcript variant 1, mRNA [NM_015589] |
| A_33_P3344292 | 0,026692951 | down | -1,917629 | SAMD4A | sterile alpha motif domain containing 4A [Source:HGNC Symbol;Acc:HGNC:23023] [ENST00000554335] |
| A_24_P6449 | 0,02592097 | down | -1,9460803 | SAMD7 | sterile alpha motif domain containing 7 [Source:HGNC Symbol;Acc:HGNC:25394] [ENST00000487910] |
| A_33_P3370763 | 0,022473752 | down | -1,9249504 | SARM1 | Homo sapiens sterile alpha and TIR motif containing 1 (SARM1), mRNA [NM_015077] |
| A_23_P94103 | 0,03416655 | up | 1,9073746 | SCARA5 | Homo sapiens scavenger receptor class A, member 5 (SCARA5), mRNA [NM_173833] |
| A_24_P210569 | 0,01826427 | down | -2,189779 | SCG3 | Homo sapiens secretogranin III (SCG3), transcript variant 1, mRNA [NM_013243] |
| A_33_P3340869 | 6,46E-04 | down | -2,163698 | SCGB1B2P | Homo sapiens secretoglobin, family 1B, member 2, pseudogene (SCGB1B2P), non-coding RNA [NR_027620] |
| A_23_P150555 | 0,024509488 | down | -2,2670386 | SCGB1D2 | Homo sapiens secretoglobin, family 1D, member 2 (SCGB1D2), mRNA [NM_006551] |
| A_24_P223018 | 0,04327273 | down | -1,6534114 | SCN11A | Homo sapiens sodium channel, voltage gated, type XI alpha subunit (SCN11A), transcript variant 1, mRNA [NM_014139] |
| A_23_P79015 | 0,022667388 | down | -2,3576698 | SCN1B | Homo sapiens sodium channel, voltage gated, type I beta subunit (SCN1B), transcript variant b, mRNA [NM_199037] |
| A_33_P3394699 | 0,011821868 | down | -1,8345557 | SCN2B | sodium channel, voltage-gated, type II, beta subunit [Source:HGNC Symbol;Acc:HGNC:10589] [ENST00000278947] |
| A_32_P187571 | 0,036437433 | down | -1,8836282 | SCN2B | Homo sapiens sodium channel, voltage gated, type II beta subunit (SCN2B), mRNA [NM_004588] |
| A_24_P792124 | 0,006706269 | down | -2,949434 | SCN9A | Homo sapiens sodium channel, voltage gated, type IX alpha subunit (SCN9A), mRNA [NM_002977] |
| A_23_P46412 | 0,004065272 | down | -1,7652537 | SCNN1D | Homo sapiens sodium channel, non voltage gated 1 delta subunit (SCNN1D), transcript variant 1, mRNA [NM_001130413] |
| A_33_P3230990 | 0,002914288 | down | -1,8658291 | SCUBE1 | Homo sapiens signal peptide, CUB domain, EGF-like 1 (SCUBE1), mRNA [NM_173050] |
| A_33_P3371219 | 0,02899139 | down | -1,5448829 | SDC2 | Homo sapiens syndecan 2 (SDC2), mRNA [NM_002998] |
| A_21_P0000891 | 0,026608245 | down | -1,5076152 | SDCBP2-AS1 | Homo sapiens SDCBP2 antisense RNA 1 (SDCBP2-AS1), transcript variant 1, long non-coding RNA [NR_040047] |
| A_23_P2645 | 0,013407113 | down | -2,7873142 | SDS | Homo sapiens serine dehydratase (SDS), mRNA [NM_006843] |
| A_33_P3421053 | 0,015919976 | down | -1,6199515 | SELE | Homo sapiens selectin E (SELE), mRNA [NM_000450] |
| A_33_P3339100 | 0,04074181 | down | -1,7279708 | SELP | Homo sapiens selectin P (granule membrane protein 140kDa, antigen CD62) (SELP), mRNA [NM_003005] |
| A_33_P3243138 | 0,04191208 | down | -2,2884147 | SELV | Homo sapiens selenoprotein V (SELV), mRNA [NM_182704] |
| A_33_P3263651 | 0,007370222 | down | -2,925967 | SEMA6B | Homo sapiens sema domain, transmembrane domain (TM), and cytoplasmic domain, (semaphorin) 6B (SEMA6B), mRNA [NM_0321 |
| A_21_P0013419 | 0,020110438 | down | -1,9440056 | SEPT7-AS1 | SEPT7 antisense RNA 1 (head to head) [Source:HGNC Symbol;Acc:HGNC:51153] [ENST00000424194] |
| A_23_P117363 | 0,02775725 | down | -1,6676384 | SERPINA6 | Homo sapiens serpin peptidase inhibitor, clade A (alpha-1 antiproteinase, antitrypsin), member 6 (SERPINA6), mRNA [NM_001756] |
| A_33_P3413993 | 0,016059466 | down | -1,6616057 | SERPING1 | serpin peptidase inhibitor, clade G (C1 inhibitor), member 1 [Source:HGNC Symbol;Acc:HGNC:1228] [ENST00000405496] |
| A_33_P3374076 | 0,003019857 | down | -2,1672196 | SEZ6 | Homo sapiens seizure related 6 homolog (mouse) (SEZ6), transcript variant 2, mRNA [NM_001098635] |
| A_33_P3387455 | 0,022483964 | down | -1,5039475 | SFMBT2 | Homo sapiens Scm-like with four mbt domains 2 (SFMBT2), transcript variant 1, mRNA [NM_001029880] |
| A_23_P62881 | 0,036320515 | down | -2,7144587 | SGIP1 | Homo sapiens SH3-domain GRB2-like (endophilin) interacting protein 1 (SGIP1), mRNA [NM_032291] |
| A_32_P527371 | 0,015741695 | down | -2,8667238 | SGSM1 | small G protein signaling modulator 1 [Source:HGNC Symbol;Acc:HGNC:29410] [ENST00000480523] |
| A_24_P203103 | 0,030777708 | down | -1,8644733 | SH2D1A | Homo sapiens SH2 domain containing 1A (SH2D1A), transcript variant 1, mRNA [NM_002351] |
| A_21_P0000575 | 0,010813881 | down | -1,5378398 | SH3RF3-AS1 | Homo sapiens SH3RF3 antisense RNA 1 (SH3RF3-AS1), long non-coding RNA [NR_029193] |
| A_23_P208636 | 0,016278734 | down | -2,7216 | SHANK1 | Homo sapiens SH3 and multiple ankyrin repeat domains 1 (SHANK1), mRNA [NM_016148] |
| A_21_P0000018 | 0,03708357 | down | -2,3851333 | SHBG | Homo sapiens sex hormone-binding globulin (SHBG), transcript variant 4, mRNA [NM_001146281] |
| A_23_P115573 | 5,64E-04 | down | -1,7605057 | SHISA4 | Homo sapiens shisa family member 4 (SHISA4), transcript variant 1, mRNA [NM_198149] |
| A_23_P17481 | 0,004950251 | down | -1,779463 | SIGLEC1 | Homo sapiens sialic acid binding Ig-like lectin 1, sialoadhesin (SIGLEC1), mRNA [NM_023068] |
| A_23_P164596 | 0,019069891 | down | -1,9004545 | SIGLEC12 | Homo sapiens sialic acid binding Ig-like lectin 12 (gene/pseudogene) (SIGLEC12), transcript variant 1, mRNA [NM_053003] |
| A_33_P3382498 | 0,047635347 | down | -1,8773823 | SIGLEC14 | Homo sapiens sialic acid binding Ig-like lectin 14 (SIGLEC14), mRNA [NM_001098612] |

| | | | | | |
|---|---|---|---|---|---|
| A_32_P80816 | 0,022218926 | down | -1,5690434 | SIGLECL1 | Homo sapiens SIGLEC family like 1 (SIGLECL1), transcript variant 1, mRNA [NM_173635] |
| A_23_P301886 | 0,019425863 | down | -1,5216206 | SIM2 | Homo sapiens single-minded family bHLH transcription factor 2 (SIM2), transcript variant SIM2s, mRNA [NM_009586] |
| A_23_P17456 | 0,013554475 | down | -1,6302507 | SIRPB1 | Homo sapiens signal-regulatory protein beta 1 (SIRPB1), transcript variant 1, mRNA [NM_006065] |
| A_33_P3356577 | 0,023766683 | down | -1,943252 | SIRPB1 | signal-regulatory protein beta 1 [Source:HGNC Symbol;Acc:HGNC:15928] [ENST00000381596] |
| A_21_P0012133 | 0,001266331 | down | -2,9247816 | SIRPB2 | signal-regulatory protein beta 2 [Source:HGNC Symbol;Acc:HGNC:16247] [ENST00000486775] |
| A_23_P43369 | 0,009741781 | down | -1,8951069 | SIT1 | Homo sapiens signaling threshold regulating transmembrane adaptor 1 (SIT1), mRNA [NM_014450] |
| A_24_P34611 | 0,014306084 | down | -1,7785677 | SIX3 | Homo sapiens SIX homeobox 3 (SIX3), mRNA [NM_005413] |
| A_21_P0002046 | 0,022115024 | down | -2,3738728 | SIX3-AS1 | Homo sapiens SIX3 antisense RNA 1 (SIX3-AS1), transcript variant 1, long non-coding RNA [NR_103785] |
| A_33_P3352827 | 0,04088375 | down | -1,5143107 | SLAMF1 | Homo sapiens signaling lymphocytic activation molecule family member 1 (SLAMF1), transcript variant 1, mRNA [NM_003037] |
| A_23_P355377 | 0,026464036 | down | -1,9830261 | SLC12A5 | Homo sapiens solute carrier family 12 (potassium/chloride transporter), member 5 (SLC12A5), transcript variant 2, mRNA [NM_0207( |
| A_33_P3210399 | 0,038037594 | down | -1,8543372 | SLC14A1 | Homo sapiens solute carrier family 14 (urea transporter), member 1 (Kidd blood group) (SLC14A1), transcript variant 4, mRNA [NM_( |
| A_23_P363313 | 0,033226755 | down | -1,6353767 | SLC16A11 | Homo sapiens solute carrier family 16, member 11 (SLC16A11), mRNA [NM_153357] |
| A_23_P42189 | 0,03699989 | down | -1,5042356 | SLC17A1 | Homo sapiens solute carrier family 17 (organic anion transporter), member 1 (SLC17A1), mRNA [NM_005074] |
| A_23_P255695 | 0,029835057 | down | -2,376025 | SLC17A3 | Homo sapiens solute carrier family 17 (organic anion transporter), member 3 (SLC17A3), transcript variant 2, mRNA [NM_006632] |
| A_23_P24294 | 0,04997865 | down | -1,6079974 | SLC17A6 | Homo sapiens solute carrier family 17 (vesicular glutamate transporter), member 6 (SLC17A6), mRNA [NM_020346] |
| A_24_P350759 | 0,045736626 | down | -2,0317166 | SLC1A2 | Homo sapiens solute carrier family 1 (glial high affinity glutamate transporter), member 2 (SLC1A2), transcript variant 1, mRNA [NM_( |
| A_23_P161968 | 0,012990549 | down | -2,0607798 | SLC22A10 | Homo sapiens solute carrier family 22, member 10 (SLC22A10), mRNA [NM_001039752] |
| A_23_P111395 | 0,028384248 | down | -1,834713 | SLC22A2 | Homo sapiens solute carrier family 22 (organic cation transporter), member 2 (SLC22A2), mRNA [NM_003058] |
| A_23_P98616 | 0,007975384 | up | 1,9063076 | SLC22A6 | Homo sapiens solute carrier family 22 (organic anion transporter), member 6 (SLC22A6), transcript variant 3, mRNA [NM_153277] |
| A_24_P304311 | 0,024703661 | down | -1,5111704 | SLC22A8 | Homo sapiens solute carrier family 22 (organic anion transporter), member 8 (SLC22A8), transcript variant 1, mRNA [NM_004254] |
| A_23_P21990 | 0,021435626 | down | -2,3075705 | SLC23A1 | Homo sapiens solute carrier family 23 (ascorbic acid transporter), member 1 (SLC23A1), transcript variant 2, mRNA [NM_152685] |
| A_33_P3390637 | 0,001712626 | down | -2,0518823 | SLC23A3 | Homo sapiens solute carrier family 23, member 3 (SLC23A3), transcript variant 1, mRNA [NM_144712] |
| A_23_P92650 | 0,008077113 | down | -2,16979 | SLC25A2 | Homo sapiens solute carrier family 25 (mitochondrial carrier; ornithine transporter) member 2 (SLC25A2), mRNA [NM_031947] |
| A_33_P3378634 | 0,010233933 | down | -1,6054789 | SLC25A30-AS1 | Homo sapiens SLC25A30 antisense RNA 1 (SLC25A30-AS1), long non-coding RNA [NR_047031] |
| A_33_P3357853 | 0,036125734 | down | -1,5698785 | SLC25A48 | solute carrier family 25, member 48 [Source:HGNC Symbol;Acc:HGNC:30451] [ENST00000274513] |
| A_33_P3400700 | 0,034226507 | down | -1,6804116 | SLC26A5 | Homo sapiens solute carrier family 26 (anion exchanger), member 5 (SLC26A5), transcript variant d, mRNA [NM_206885] |
| A_23_P14667 | 0,021093618 | down | -2,6855488 | SLC28A1 | Homo sapiens solute carrier family 28 (concentrative nucleoside transporter), member 1 (SLC28A1), transcript variant 1, mRNA [NM_ |
| A_24_P405705 | 0,029763723 | down | -1,5351261 | SLC2A2 | Homo sapiens solute carrier family 2 (facilitated glucose transporter), member 2 (SLC2A2), transcript variant 1, mRNA [NM_000340] |
| A_23_P58729 | 0,009434443 | down | -2,8295608 | SLC34A1 | Homo sapiens solute carrier family 34 (type II sodium/phosphate cotransporter), member 1 (SLC34A1), transcript variant 1, mRNA [NI |
| A_33_P3780983 | 0,029410128 | down | -1,6129428 | SLC34A3 | Homo sapiens solute carrier family 34 (type II sodium/phosphate cotransporter), member 3 (SLC34A3), transcript variant 2, mRNA [NI |
| A_23_P31996 | 0,029256124 | down | -1,7108716 | SLC46A2 | Homo sapiens solute carrier family 46, member 2 (SLC46A2), mRNA [NM_033051] |
| A_33_P3403399 | 0,037178632 | down | -1,6168706 | SLC47A1 | Homo sapiens solute carrier family 47 (multidrug and toxin extrusion), member 1 (SLC47A1), mRNA [NM_018242] |
| A_24_P142503 | 0,006304974 | down | -2,2221107 | SLC47A1 | Homo sapiens solute carrier family 47 (multidrug and toxin extrusion), member 1 (SLC47A1), mRNA [NM_018242] |
| A_33_P3400217 | 0,022825863 | down | -2,310932 | SLC4A1 | Homo sapiens solute carrier family 4 (anion exchanger), member 1 (Diego blood group) (SLC4A1), mRNA [NM_000342] |
| A_24_P930111 | 0,01632282 | down | -1,7870566 | SLC4A10 | Homo sapiens solute carrier family 4, sodium bicarbonate transporter, member 10 (SLC4A10), transcript variant 2, mRNA [NM_02205 |
| A_24_P242581 | 0,0472428 | down | -1,5385278 | SLC5A9 | Homo sapiens solute carrier family 5 (sodium/sugar cotransporter), member 9 (SLC5A9), transcript variant 2, mRNA [NM_001011547 |
| A_24_P212234 | 0,012272978 | down | -2,2397532 | SLC6A18 | Homo sapiens solute carrier family 6 (neutral amino acid transporter), member 18 (SLC6A18), mRNA [NM_182632] |
| A_24_P24770 | 0,010167176 | down | -2,4188416 | SLC9C1 | Homo sapiens solute carrier family 9, subfamily C (Na+-transporting carboxylic acid decarboxylase), member 1 (SLC9C1), mRNA [NI |
| A_32_P485915 | 0,023882182 | down | -1,8074466 | SLC9C2 | Homo sapiens solute carrier family 9, member C2 (putative) (SLC9C2), mRNA [NM_178527] |
| A_33_P3261468 | 0,024255997 | down | -1,5493028 | SLFN12L | Homo sapiens schlafen family member 12-like (SLFN12L), mRNA [NM_001195790] |
| A_33_P3358049 | 0,007637858 | down | -2,1126182 | SLFN14 | Homo sapiens schlafen family member 14 (SLFN14), mRNA [NM_001129820] |
| A_33_P3411021 | 0,023191443 | down | -2,4643996 | SLIT1 | Homo sapiens slit homolog 1 (Drosophila), mRNA (cDNA clone IMAGE:5247200), with apparent retained intron. [BC028105] |
| A_21_P0000767 | 0,032174986 | down | -1,5992131 | SLIT1-AS1 | Homo sapiens SLIT1 antisense RNA 1 (SLIT1-AS1), long non-coding RNA [NR_038330] |
| A_24_P66605 | 0,03905139 | down | -1,6498523 | SLITRK2 | Homo sapiens SLIT and NTRK-like family, member 2 (SLITRK2), transcript variant 1, mRNA [NM_032539] |
| A_33_P3230189 | 0,0163531 | down | -1,5824893 | SLITRK6 | Homo sapiens SLIT and NTRK-like family, member 6 (SLITRK6), mRNA [NM_032229] |
| A_33_P3877739 | 0,034608416 | down | -1,8168641 | SMCR2 | ag52h12.x5 Gessler Wilms tumor Homo sapiens cDNA clone IMAGE:1126631 3', mRNA sequence [AI821758] |
| A_33_P3442301 | 0,005258334 | down | -2,9398596 | SMG1P3 | 602363146F1 NIH_MGC_90 Homo sapiens cDNA clone IMAGE:4471450 5', mRNA sequence [BG250912] |
| A_21_P0011718 | 0,02753268 | down | -1,5028747 | SMIM17 | Homo sapiens small integral membrane protein 17 (SMIM17), mRNA [NM_001193628] |
| A_21_P0000170 | 0,023633376 | down | -1,593824 | SMIM18 | Homo sapiens small integral membrane protein 18 (SMIM18), mRNA [NM_001206847] |

| | | | | | |
|---|---|---|---|---|---|
| A_23_P151361 | 0,028034547 | down | -2,04211 | SMIM2 | Homo sapiens small integral membrane protein 2 (SMIM2), mRNA [NM_024058] |
| A_19_P00810806 | 0,009850499 | down | -1,8672627 | SMIM2-AS1 | Homo sapiens SMIM2 antisense RNA 1 (SMIM2-AS1), transcript variant 1, long non-coding RNA [NR_104064] |
| A_24_P555066 | 0,010583305 | down | -1,7624078 | SMTNL2 | Homo sapiens smoothelin-like 2 (SMTNL2), transcript variant 2, mRNA [NM_198501] |
| A_24_P389285 | 0,032019325 | down | -1,6541474 | SND1-IT1 | Homo sapiens SND1 intronic transcript 1 (non-protein coding) (SND1-IT1), long non-coding RNA [NR_027330] |
| A_33_P3711426 | 0,028850287 | down | -1,5505359 | SNHG8 | AGENCOURT_6644733 NIH_MGC_122 Homo sapiens cDNA clone IMAGE:5766924 5', mRNA sequence [BM926530] |
| A_21_P0000327 | 0,004324663 | down | -1,7134421 | SNORA71E | Homo sapiens small nucleolar RNA, H/ACA box 71E (SNORA71E), small nucleolar RNA [NR_002972] |
| A_33_P3579391 | 0,0441276 | down | -1,516591 | SNORA77 | DB361496 MAMMA1 Homo sapiens cDNA clone MAMMA1001810 3', mRNA sequence [DB361496] |
| A_33_P3604591 | 0,020365998 | down | -1,5485392 | SNORA78 | qj88d05.x1 NCI_CGAP_Kid3 Homo sapiens cDNA clone IMAGE:1866537 3', mRNA sequence [AI271839] |
| A_21_P0000457 | 0,013652135 | down | -1,9589225 | SNORD115-27 | Homo sapiens small nucleolar RNA, C/D box 115-27 (SNORD115-27), small nucleolar RNA [NR_003496] |
| A_21_P0000441 | 0,045578953 | up | 1,74759 | SNORD115-5 | Homo sapiens small nucleolar RNA, C/D box 115-5 (SNORD115-5), small nucleolar RNA [NR_003297] |
| A_33_P3414967 | 0,042055704 | down | -1,5740342 | SNTG1 | Homo sapiens syntrophin, gamma 1 (SNTG1), transcript variant 1, mRNA [NM_018967] |
| A_23_P251132 | 0,01099353 | down | -2,4376757 | SNTG2 | Homo sapiens syntrophin, gamma 2 (SNTG2), mRNA [NM_018968] |
| A_24_P13831 | 0,008098613 | down | -1,9385316 | SNX20 | Homo sapiens sorting nexin 20 (SNX20), transcript variant 1, mRNA [NM_182854] |
| A_33_P3284453 | 0,021567157 | down | -1,985824 | SOGA3 | Homo sapiens SOGA family member 3 (SOGA3), mRNA [NM_001012279] |
| A_24_P307964 | 0,045259863 | down | -2,1442137 | SOHLH1 | Homo sapiens spermatogenesis and oogenesis specific basic helix-loop-helix 1 (SOHLH1), transcript variant 2, mRNA [NM_0010124 |
| A_21_P0000167 | 0,04515371 | down | -1,5759147 | SORCS1 | Homo sapiens sortilin-related VPS10 domain containing receptor 1 (SORCS1), transcript variant 5, mRNA [NM_001206571] |
| A_33_P3414192 | 0,020702502 | down | -2,2150536 | SORCS1 | Homo sapiens sortilin-related VPS10 domain containing receptor 1 (SORCS1), transcript variant 6, mRNA [NM_001206572] |
| A_23_P434416 | 0,040382847 | down | -1,5515503 | SOX1 | Homo sapiens SRY (sex determining region Y)-box 1 (SOX1), mRNA [NM_005986] |
| A_23_P80662 | 0,036862474 | down | -1,5133303 | SOX14 | Homo sapiens SRY (sex determining region Y)-box 14 (SOX14), mRNA [NM_004189] |
| A_33_P3226761 | 0,032501493 | down | -1,56753 | SOX18 | Homo sapiens SRY (sex determining region Y)-box 18 (SOX18), mRNA [NM_018419] |
| A_24_P756289 | 0,0130065 | down | -1,5947951 | SOX2-OT | Homo sapiens SOX2 overlapping transcript (SOX2-OT), transcript variant 1, long non-coding RNA [NR_075091] |
| A_23_P85218 | 0,017003726 | down | -4,103596 | SOX3 | Homo sapiens SRY (sex determining region Y)-box 3 (SOX3), mRNA [NM_005634] |
| A_23_P69970 | 0,022561891 | down | -1,6421124 | SOX30 | Homo sapiens SRY (sex determining region Y)-box 30 (SOX30), transcript variant 1, mRNA [NM_178424] |
| A_23_P317729 | 0,033817295 | down | -1,61861 | SP7 | Homo sapiens Sp7 transcription factor (SP7), transcript variant 2, mRNA [NM_152860] |
| A_33_P3383184 | 0,024169628 | down | -1,562946 | SP9 | Sp9 transcription factor [Source:HGNC Symbol;Acc:HGNC:30690] [ENST00000394967] |
| A_24_P272254 | 0,014332896 | down | -1,525989 | SPACA5 | Homo sapiens sperm acrosome associated 5 (SPACA5), mRNA [NM_205856] |
| A_23_P65302 | 0,027455894 | down | -1,5659392 | SPACA7 | Homo sapiens sperm acrosome associated 7 (SPACA7), mRNA [NM_145248] |
| A_23_P324319 | 0,01768081 | down | -2,1850088 | SPAG11B | Homo sapiens sperm associated antigen 11B (SPAG11B), transcript variant C, mRNA [NM_058203] |
| A_33_P3346966 | 0,049103294 | down | -1,8825275 | SPAG16 | Homo sapiens sperm associated antigen 16 (SPAG16), transcript variant 1, mRNA [NM_024532] |
| A_21_P0006840 | 0,021653082 | down | -1,5073831 | SPAG6 | sperm associated antigen 6 [Source:HGNC Symbol;Acc:HGNC:11215] [ENST00000487973] |
| A_23_P329956 | 0,045111276 | down | -1,5097711 | SPATA3 | Homo sapiens spermatogenesis associated 3 (SPATA3), mRNA [NM_139073] |
| A_21_P0001802 | 0,019296184 | down | -1,6256552 | SPATA3-AS1 | Homo sapiens SPATA3 antisense RNA 1 (head to head) (SPATA3-AS1), long non-coding RNA [NR_033879] |
| A_21_P0001801 | 0,026842715 | down | -1,6465759 | SPATA3-AS1 | Homo sapiens SPATA3 antisense RNA 1 (head to head) (SPATA3-AS1), long non-coding RNA [NR_033879] |
| A_33_P3217364 | 0,02341446 | down | -2,2228775 | SPATA31A3 | Homo sapiens SPATA31 subfamily A, member 3 (SPATA31A3), mRNA [NM_001083124] |
| A_33_P3242609 | 0,014653626 | down | -1,634781 | SPATA31D1 | Homo sapiens SPATA31 subfamily D, member 1 (SPATA31D1), mRNA [NM_001001670] |
| A_33_P3239242 | 0,018988892 | down | -1,5935154 | SPATA6 | Homo sapiens spermatogenesis associated 6 (SPATA6), transcript variant 1, mRNA [NM_019073] |
| A_32_P217390 | 0,02975649 | up | 1,7394167 | SPATA8 | Homo sapiens spermatogenesis associated 8 (SPATA8), mRNA [NM_173499] |
| A_33_P3276450 | 0,033931054 | down | -1,5588677 | SPATC1 | Homo sapiens spermatogenesis and centriole associated 1 (SPATC1), transcript variant 1, mRNA [NM_198572] |
| A_19_P00806499 | 0,012199066 | down | -2,5847063 | SPG20-AS1 | Homo sapiens SPG20 antisense RNA 1 (SPG20-AS1), transcript variant 1, long non-coding RNA [NR_045180] |
| A_32_P148275 | 0,017867608 | down | -1,9993777 | SPIC | Homo sapiens Spi-C transcription factor (Spi-1/PU.1 related) (SPIC), mRNA [NM_152323] |
| A_33_P3388006 | 0,022059584 | up | 1,6597683 | SPICE1 | Homo sapiens spindle and centriole associated protein 1 (SPICE1), mRNA [NM_144718] |
| A_33_P3239620 | 0,04304656 | down | -2,234494 | SPINT3 | Homo sapiens serine peptidase inhibitor, Kunitz type, 3 (SPINT3), mRNA [NM_006652] |
| A_32_P142779 | 0,012638428 | down | -2,3252904 | SPPL2C | Homo sapiens signal peptide peptidase like 2C (SPPL2C), mRNA [NM_175882] |
| A_33_P3401981 | 0,013331353 | up | 2,2982929 | SPRED3 | Homo sapiens sprouty-related, EVH1 domain containing 3 (SPRED3), transcript variant 1, mRNA [NM_001042522] |
| A_33_P3338423 | 0,04898628 | down | -1,7929358 | SPRY4 | Homo sapiens sprouty homolog 4 (Drosophila) (SPRY4), transcript variant 3, mRNA [NM_001293289] |
| A_21_P0014846 | 0,012128036 | down | -1,7351543 | SRGAP3-AS3 | Homo sapiens SRGAP3 antisense RNA 3 (SRGAP3-AS3), long non-coding RNA [NR_103443] |
| A_33_P3393200 | 9,40E-04 | down | -2,0401747 | SRRM4 | Homo sapiens serine/arginine repetitive matrix 4 (SRRM4), mRNA [NM_194286] |
| A_33_P3393931 | 4,34E-04 | down | -3,198034 | SSBP3-AS1 | Homo sapiens SSBP3 antisense RNA 1 (SSBP3-AS1), long non-coding RNA [NR_103541] |
| A_23_P33875 | 0,03749426 | down | -1,6057633 | SSX4B | Homo sapiens synovial sarcoma, X breakpoint 4B (SSX4B), transcript variant 1, mRNA [NM_001034832] |

| | | | | | |
|---|---|---|---|---|---|
| A_21_P0000162 | 0,043983694 | down | -1,5549469 | SSX6 | synovial sarcoma, X breakpoint 6 (pseudogene) [Source:HGNC Symbol;Acc:HGNC:19652] [ENST00000412590] |
| A_23_P431923 | 0,017879179 | down | -2,0143034 | SSX8 | Homo sapiens synovial sarcoma, X breakpoint 8 (SSX8), non-coding RNA [NR_027250] |
| A_23_P429425 | 0,025924178 | down | -1,9006276 | ST6GAL2 | Homo sapiens ST6 beta-galactosamide alpha-2,6-sialyltranferase 2 (ST6GAL2), transcript variant 1, mRNA [NM_032528] |
| A_24_P664850 | 0,02258923 | down | -2,2057276 | ST8SIA2 | Homo sapiens ST8 alpha-N-acetyl-neuraminide alpha-2,8-sialyltransferase 2 (ST8SIA2), mRNA [NM_006011] |
| A_23_P162607 | 6,94E-04 | down | -3,8409832 | STAB2 | Homo sapiens stabilin 2 (STAB2), mRNA [NM_017564] |
| A_33_P3365760 | 0,00873082 | down | -2,2722902 | STAP1 | Homo sapiens signal transducing adaptor family member 1 (STAP1), mRNA [NM_012108] |
| A_24_P214231 | 0,040103983 | down | -1,7594398 | STIL | Homo sapiens SCL/TAL1 interrupting locus (STIL), transcript variant 1, mRNA [NM_001048166] |
| A_32_P85676 | 0,028793484 | down | -1,5987798 | STK32B | Homo sapiens serine/threonine kinase 32B (STK32B), mRNA [NM_018401] |
| A_32_P126229 | 0,014228804 | down | -2,0438733 | STMND1 | Homo sapiens stathmin domain containing 1 (STMND1), mRNA [NM_001190766] |
| A_33_P3357935 | 0,001949755 | down | -1,6140078 | STRC | Homo sapiens stereocilin (STRC), mRNA [NM_153700] |
| A_33_P3212456 | 0,035511613 | up | 1,9275006 | STX1A | Homo sapiens syntaxin 1A (brain) (STX1A), transcript variant 2, mRNA [NM_001165903] |
| A_23_P418785 | 0,03741636 | down | -1,7757784 | STXBP5L | Homo sapiens syntaxin binding protein 5-like (STXBP5L), mRNA [NM_014980] |
| A_21_P0002970 | 0,03441257 | down | -1,66391 | SUCLG2-AS1 | Homo sapiens SUCLG2 antisense RNA 1 (head to head) (SUCLG2-AS1), transcript variant 1, long non-coding RNA [NR_109992] |
| A_23_P145711 | 0,03152687 | down | -1,7065339 | SUGCT | Homo sapiens succinyl-CoA:glutarate-CoA transferase (SUGCT), transcript variant 4, mRNA [NM_024728] |
| A_23_P147070 | 0,043297544 | up | 1,500111 | SUN1 | Sad1 and UNC84 domain containing 1 [Source:HGNC Symbol;Acc:HGNC:18587] [ENST00000340926] |
| A_23_P140050 | 0,02568759 | down | -1,5557477 | SUPT20H | Homo sapiens suppressor of Ty 20 homolog (S. cerevisiae) (SUPT20H), transcript variant 2, mRNA [NM_017569] |
| A_33_P3347152 | 0,04633341 | down | -1,5735486 | SUPT3H | suppressor of Ty 3 homolog (S. cerevisiae) [Source:HGNC Symbol;Acc:HGNC:11466] [ENST00000371458] |
| A_33_P3314301 | 0,003081162 | down | -2,9637141 | SV2C | Homo sapiens synaptic vesicle glycoprotein 2C (SV2C), transcript variant 1, mRNA [NM_014979] |
| A_23_P112982 | 0,03930959 | down | -1,670812 | SVOP | Homo sapiens SV2 related protein homolog (rat) (SVOP), mRNA [NM_018711] |
| A_23_P722 | 0,011971025 | down | -2,126776 | SYCP1 | Homo sapiens synaptonemal complex protein 1 (SYCP1), transcript variant 1, mRNA [NM_003176] |
| A_23_P76374 | 0,025963977 | down | -2,016444 | SYCP3 | Homo sapiens synaptonemal complex protein 3 (SYCP3), transcript variant 2, mRNA [NM_153694] |
| A_33_P3400239 | 0,016009161 | down | -2,0678427 | SYNDIG1 | PREDICTED: Homo sapiens synapse differentiation inducing 1 (SYNDIG1), transcript variant X1, mRNA [XM_006723626] |
| A_23_P55917 | 0,006564947 | down | -2,0010567 | SYT3 | Homo sapiens synaptotagmin III (SYT3), transcript variant 1, mRNA [NM_032298] |
| A_23_P78849 | 0,030389708 | down | -1,6106217 | SYT5 | Homo sapiens synaptotagmin V (SYT5), transcript variant 1, mRNA [NM_003180] |
| A_33_P3277298 | 0,010476894 | down | -2,276835 | SYT6 | Homo sapiens synaptotagmin VI (SYT6), transcript variant 3, mRNA [NM_001270805] |
| A_33_P3373134 | 0,023886966 | down | -1,7469172 | SZT2 | Homo sapiens seizure threshold 2 homolog (mouse) (SZT2), mRNA [NM_015284] |
| A_33_P3253877 | 0,032428876 | down | -1,6475282 | TACC1 | transforming, acidic coiled-coil containing protein 1 [Source:HGNC Symbol;Acc:HGNC:11522] [ENST00000518415] |
| A_24_P376441 | 0,017705858 | up | 1,5187011 | TAF5L | Homo sapiens TAF5-like RNA polymerase II, p300/CBP-associated factor (PCAF)-associated factor, 65kDa (TAF5L), transcript varian |
| A_33_P3405769 | 6,09E-04 | down | -2,5182621 | TARM1 | Homo sapiens T cell-interacting, activating receptor on myeloid cells 1 (TARM1), mRNA [NM_001135686] |
| A_23_P416191 | 5,99E-04 | down | -2,2355773 | TAS2R31 | Homo sapiens taste receptor, type 2, member 31 (TAS2R31), mRNA [NM_176885] |
| A_23_P359746 | 0,029495725 | down | -1,6928108 | TAS2R38 | Homo sapiens taste receptor, type 2, member 38 (TAS2R38), mRNA [NM_176817] |
| A_23_P337867 | 0,019129733 | down | -1,8654202 | TAS2R41 | Homo sapiens taste receptor, type 2, member 41 (TAS2R41), mRNA [NM_176883] |
| A_23_P367013 | 0,034210492 | down | -1,915556 | TAS2R60 | Homo sapiens taste receptor, type 2, member 60 (TAS2R60), mRNA [NM_177437] |
| A_21_P0014430 | 0,04677206 | down | -2,0643628 | TBC1D22A-AS1 | Homo sapiens TBC1D22A antisense RNA 1 (TBC1D22A-AS1), long non-coding RNA [NR_122047] |
| A_24_P896205 | 0,03571733 | down | -1,846406 | TBX2-AS1 | Homo sapiens TBX2 antisense RNA 1 (TBX2-AS1), transcript variant 1, long non-coding RNA [NR_125749] |
| A_23_P25176 | 0,03920474 | down | -2,0042367 | TBX5 | Homo sapiens T-box 5 (TBX5), transcript variant 1, mRNA [NM_000192] |
| A_33_P3424122 | 0,017793663 | down | -1,5505353 | TCHHL1 | Homo sapiens trichohyalin-like 1 (TCHHL1), mRNA [NM_001008536] |
| A_23_P205348 | 0,033034436 | down | -1,6842132 | TCL6 | Homo sapiens T-cell leukemia/lymphoma 6 (non-protein coding) (TCL6), long non-coding RNA [NR_028288] |
| A_33_P3387164 | 0,04923833 | down | -2,036251 | TCP11 | Homo sapiens t-complex 11, testis-specific (TCP11), transcript variant 6, mRNA [NM_001261820] |
| A_33_P3267248 | 0,024514845 | down | -1,5303752 | TDRD10 | Homo sapiens tudor domain containing 10 (TDRD10), transcript variant 2, mRNA [NM_182499] |
| A_33_P3421664 | 0,029505037 | down | -1,9344068 | TDRD5 | Homo sapiens tudor domain containing 5 (TDRD5), transcript variant 1, mRNA [NM_001199085] |
| A_24_P37712 | 0,013865687 | down | -1,9171915 | TEC | Homo sapiens tec protein tyrosine kinase (TEC), mRNA [NM_003215] |
| A_23_P375354 | 0,01789843 | down | -1,9865586 | TECTA | Homo sapiens tectorin alpha (TECTA), mRNA [NM_005422] |
| A_23_P97826 | 0,0270659 | down | -1,583307 | TECTB | Homo sapiens tectorin beta (TECTB), mRNA [NM_058222] |
| A_23_P74723 | 0,014890608 | down | -1,685172 | TEX35 | Homo sapiens testis expressed 35 (TEX35), transcript variant 1, mRNA [NM_032126] |
| A_24_P315056 | 0,014173012 | down | -1,791677 | TEX36 | Homo sapiens testis expressed 36 (TEX36), mRNA [NM_001128202] |
| A_32_P120127 | 0,028518107 | down | -1,5341239 | TEX36-AS1 | Homo sapiens TEX36 antisense RNA 1 (TEX36-AS1), long non-coding RNA [NR_023362] |
| A_23_P432005 | 0,006189491 | down | -2,2041304 | TEX37 | Homo sapiens testis expressed 37 (TEX37), mRNA [NM_152670] |
| A_24_P484965 | 0,018883005 | down | -1,6665107 | TEX41 | testis expressed 41 (non-protein coding) [Source:HGNC Symbol;Acc:HGNC:48667] [ENST00000423031] |

| | | | | | |
|---|---|---|---|---|---|
| A_21_P0011860 | 0,034430943 | down | -1,7206877 | TEX41 | Homo sapiens testis expressed 41 (non-protein coding) (TEX41), long non-coding RNA [NR_033870] |
| A_24_P377124 | 0,015288389 | down | -1,9978459 | THPO | Homo sapiens thrombopoietin (THPO), transcript variant 1, mRNA [NM_000460] |
| A_21_P0003309 | 0,012410848 | down | -2,8761206 | THPO | Homo sapiens thrombopoietin (THPO), transcript variant 8, mRNA [NM_001290027] |
| A_33_P3342056 | 0,002911164 | down | -2,5372047 | TIGIT | Homo sapiens T cell immunoreceptor with Ig and ITIM domains (TIGIT), mRNA [NM_173799] |
| A_33_P3235213 | 0,015364205 | down | -2,5404625 | TIGIT | Homo sapiens T cell immunoreceptor with Ig and ITIM domains (TIGIT), mRNA [NM_173799] |
| A_24_P201153 | 0,042924162 | up | 1,5147619 | TJP2 | Homo sapiens tight junction protein 2 (TJP2), transcript variant 2, mRNA [NM_201629] |
| A_33_P3274955 | 0,022220744 | down | -1,9257457 | TKTL2 | Homo sapiens transketolase-like 2 (TKTL2), mRNA [NM_032136] |
| A_33_P3780311 | 0,025073512 | down | -1,7692645 | TLR8-AS1 | Homo sapiens TLR8 antisense RNA 1 (TLR8-AS1), long non-coding RNA [NR_030727] |
| A_23_P114008 | 0,02951521 | down | -1,514669 | TM4SF20 | Homo sapiens transmembrane 4 L six family member 20 (TM4SF20), mRNA [NM_024795] |
| A_23_P27107 | 0,003577291 | down | -3,6172636 | TM4SF5 | Homo sapiens transmembrane 4 L six family member 5 (TM4SF5), mRNA [NM_003963] |
| A_32_P87649 | 0,0370746 | down | -2,2158432 | TMCO2 | Homo sapiens transmembrane and coiled-coil domains 2 (TMCO2), mRNA [NM_001008740] |
| A_23_P254688 | 0,02878435 | down | -2,3979728 | TMEM108 | Homo sapiens transmembrane protein 108 (TMEM108), transcript variant 1, mRNA [NM_023943] |
| A_23_P308839 | 0,03741295 | down | -1,7640392 | TMEM132D | Homo sapiens transmembrane protein 132D (TMEM132D), mRNA [NM_133448] |
| A_23_P252082 | 0,006004837 | down | -1,7340733 | TMEM176A | Homo sapiens transmembrane protein 176A (TMEM176A), mRNA [NM_018487] |
| A_33_P3226060 | 0,005579385 | down | -2,483791 | TMEM178B | Homo sapiens transmembrane protein 178B (TMEM178B), mRNA [NM_001195278] |
| A_33_P3422248 | 0,002868369 | down | -1,7062064 | TMEM200C | transmembrane protein 200C [Source:HGNC Symbol;Acc:HGNC:37208] [ENST00000581347] |
| A_23_P26062 | 0,04506961 | down | -1,85466 | TMEM202 | Homo sapiens transmembrane protein 202 (TMEM202), mRNA [NM_001080462] |
| A_33_P3363341 | 0,022621458 | down | -1,5413668 | TMEM212 | Homo sapiens transmembrane protein 212 (TMEM212), mRNA [NM_001164436] |
| A_32_P489986 | 0,023459831 | down | -2,1372492 | TMEM232 | Homo sapiens transmembrane protein 232 (TMEM232), mRNA [NM_001039763] |
| A_23_P430747 | 0,015087839 | down | -1,5232787 | TMEM257 | Homo sapiens transmembrane protein 257 (TMEM257), mRNA [NM_004709] |
| A_23_P382240 | 0,038378276 | down | -1,5911989 | TMEM26 | Homo sapiens transmembrane protein 26 (TMEM26), mRNA [NM_178505] |
| A_23_P318938 | 0,028179064 | down | -2,0037775 | TMEM52B | Homo sapiens transmembrane protein 52B (TMEM52B), transcript variant 1, mRNA [NM_153022] |
| A_33_P3277611 | 0,001163601 | down | -1,8976617 | TMEM8C | Homo sapiens transmembrane protein 8C (TMEM8C), mRNA [NM_001080483] |
| A_33_P3368776 | 0,029106585 | down | -1,5082635 | TMIGD1 | Homo sapiens transmembrane and immunoglobulin domain containing 1 (TMIGD1), mRNA [NM_206832] |
| A_33_P3286923 | 0,040494617 | up | 1,6092532 | TMPRSS5 | Homo sapiens transmembrane protease, serine 5 (TMPRSS5), transcript variant 4, mRNA [NM_001288751] |
| A_21_P0010506 | 0,023786785 | down | -1,8435849 | TNFRSF14 | Homo sapiens tumor necrosis factor receptor superfamily, member 14 (TNFRSF14), transcript variant 1, mRNA [NM_003820] |
| A_33_P3286157 | 0,02185078 | down | -1,5149355 | TNFRSF4 | Homo sapiens tumor necrosis factor receptor superfamily, member 4 (TNFRSF4), mRNA [NM_003327] |
| A_33_P3305571 | 0,029245658 | up | 1,6943535 | TNFRSF6B | Homo sapiens tumor necrosis factor receptor superfamily, member 6b, decoy (TNFRSF6B), mRNA [NM_003823] |
| A_24_P415680 | 0,020926481 | down | -1,7976235 | TNNI3K | Homo sapiens TNNI3 interacting kinase (TNNI3K), mRNA [NM_015978] |
| A_23_P127824 | 0,006474461 | down | -2,5665646 | TNNT3 | Homo sapiens troponin T type 3 (skeletal, fast) (TNNT3), transcript variant 3, mRNA [NM_001042780] |
| A_32_P19000 | 0,035113588 | up | 1,6654769 | TNRC6C-AS1 | Homo sapiens TNRC6C antisense RNA 1 (TNRC6C-AS1), long non-coding RNA [NR_040071] |
| A_24_P911362 | 8,10E-04 | up | 2,7653973 | TNXB | tenascin XB [Source:HGNC Symbol;Acc:HGNC:11976] [ENST00000427181] |
| A_24_P307135 | 0,006712705 | down | -1,8947989 | TNXB | Homo sapiens tenascin XB (TNXB), transcript variant XB, mRNA [NM_019105] |
| A_24_P117942 | 0,01944567 | down | -1,5644218 | TOMM20L | Homo sapiens translocase of outer mitochondrial membrane 20 homolog (yeast)-like (TOMM20L), mRNA [NM_207377] |
| A_33_P3247403 | 4,89E-04 | down | -2,3949142 | TOR3A | Homo sapiens torsin family 3, member A (TOR3A), mRNA [NM_022371] |
| A_24_P413470 | 0,034744814 | down | -1,6178033 | TP73 | Homo sapiens tumor protein p73 (TP73), transcript variant 1, mRNA [NM_005427] |
| A_23_P37702 | 0,030871052 | down | -1,8448529 | TPSAB1 | Homo sapiens tryptase alpha/beta 1 (TPSAB1), mRNA [NM_003294] |
| A_24_P118052 | 0,037084557 | down | -2,0904982 | TPTE2 | Homo sapiens transmembrane phosphoinositide 3-phosphatase and tensin homolog 2 (TPTE2), transcript variant 3, mRNA [NM_199 |
| A_23_P18518 | 0,035332363 | up | 1,7811774 | TRAM1L1 | Homo sapiens translocation associated membrane protein 1-like 1 (TRAM1L1), mRNA [NM_152402] |
| A_24_P942132 | 0,034116253 | down | -1,5879163 | TRAPPC6B | Homo sapiens trafficking protein particle complex 6B (TRAPPC6B), transcript variant 1, mRNA [NM_001079537] |
| A_21_P0000178 | 0,028545864 | down | -1,6577682 | TREM1 | Homo sapiens triggering receptor expressed on myeloid cells 1 (TREM1), transcript variant 2, mRNA [NM_001242589] |
| A_24_P170439 | 0,024402061 | down | -1,7213168 | TREML3P | Homo sapiens triggering receptor expressed on myeloid cells-like 3, pseudogene (TREML3P), non-coding RNA [NR_027256] |
| A_24_P655458 | 0,043449752 | down | -1,666827 | TREML4 | Homo sapiens triggering receptor expressed on myeloid cells-like 4 (TREML4), mRNA [NM_198153] |
| A_33_P3311493 | 0,017985418 | down | -2,043727 | TRHDE-AS1 | Homo sapiens TRHDE antisense RNA 1 (TRHDE-AS1), transcript variant 2, long non-coding RNA [NR_026836] |
| A_21_P0007724 | 0,018857677 | down | -2,6892905 | TRHDE-AS1 | Homo sapiens TRHDE antisense RNA 1 (TRHDE-AS1), transcript variant 1, long non-coding RNA [NR_026837] |
| A_33_P3271594 | 0,03771064 | down | -1,8189481 | TRIM54 | Homo sapiens tripartite motif containing 54 (TRIM54), transcript variant 1, mRNA [NM_032546] |
| A_33_P3220247 | 0,015958285 | down | -2,6647294 | TRIM6 | Homo sapiens tripartite motif containing 6 (TRIM6), transcript variant 1, mRNA [NM_001003818] |
| A_32_P429687 | 0,02452206 | down | -1,5121688 | TRIM72 | Homo sapiens tripartite motif containing 72, E3 ubiquitin protein ligase (TRIM72), mRNA [NM_001008274] |
| A_33_P3325748 | 0,033234823 | down | -1,7895725 | TRIOBP | Homo sapiens TRIO and F-actin binding protein (TRIOBP), transcript variant 2, mRNA [NM_138632] |

| | | | | | |
|---|---|---|---|---|---|
| A_33_P3340014 | 0,009521623 | down | -1,9314481 | TRO | Homo sapiens trophinin (TRO), transcript variant 3, mRNA [NM_016157] |
| A_33_P3330074 | 0,014877556 | down | -1,6581982 | TRPC2 | transient receptor potential cation channel, subfamily C, member 2, pseudogene [Source:HGNC Symbol;Acc:HGNC:12334] [ENST00 |
| A_23_P129225 | 0,048240725 | down | -1,5774928 | TRPM1 | Homo sapiens transient receptor potential cation channel, subfamily M, member 1 (TRPM1), transcript variant 2, mRNA [NM_002420 |
| A_23_P250694 | 0,023217982 | down | -1,756289 | TSGA13 | Homo sapiens testis specific, 13 (TSGA13), mRNA [NM_052933] |
| A_24_P213643 | 0,020759711 | down | -1,855485 | TSPAN10 | Homo sapiens tetraspanin 10 (TSPAN10), transcript variant 2, mRNA [NM_031945] |
| A_24_P13083 | 0,015068628 | up | 1,9005895 | TSPAN18 | Homo sapiens tetraspanin 18 (TSPAN18), mRNA [NM_130783] |
| A_23_P40515 | 0,036352996 | down | -1,5586011 | TSSK2 | Homo sapiens testis-specific serine kinase 2 (TSSK2), mRNA [NM_053006] |
| A_23_P399681 | 0,03151081 | down | -1,5436629 | TSSK3 | Homo sapiens testis-specific serine kinase 3 (TSSK3), mRNA [NM_052841] |
| A_33_P3280811 | 0,003975987 | down | -1,6676342 | TTC16 | Homo sapiens tetratricopeptide repeat domain 16 (TTC16), mRNA [NM_144965] |
| A_23_P73150 | 0,046419665 | down | -1,800721 | TTC25 | Homo sapiens tetratricopeptide repeat domain 25 (TTC25), transcript variant 1, mRNA [NM_031421] |
| A_33_P3385993 | 0,012815143 | down | -1,895446 | TTLL9 | Homo sapiens tubulin tyrosine ligase-like family member 9 (TTLL9), mRNA [NM_001008409] |
| A_21_P0013907 | 0,017969057 | down | -1,8344084 | TTTY10 | Homo sapiens testis-specific transcript, Y-linked 10 (non-protein coding) (TTTY10), long non-coding RNA [NR_001542] |
| A_24_P331693 | 0,02313415 | down | -1,82485 | TTTY22 | Homo sapiens testis-specific transcript, Y-linked 22 (non-protein coding) (TTTY22), long non-coding RNA [NR_001539] |
| A_24_P340227 | 0,015995963 | down | -1,5865815 | TTTY23 | Homo sapiens testis-specific transcript, Y-linked 23 (non-protein coding) (TTTY23), long non-coding RNA [NR_001540] |
| A_23_P171409 | 0,028324999 | down | -1,9723618 | TTTY6 | Homo sapiens testis-specific transcript, Y-linked 6 (non-protein coding) (TTTY6), long non-coding RNA [NR_001527] |
| A_23_P434398 | 0,026975011 | down | -1,5758218 | TXLNB | Homo sapiens taxilin beta (TXLNB), mRNA [NM_153235] |
| A_33_P3344956 | 0,011818254 | down | -3,1964319 | TXNL1 | thioredoxin-like 1 [Source:HGNC Symbol;Acc:HGNC:12436] [ENST00000587807] |
| A_23_P312505 | 0,030452294 | down | -1,5068733 | TYR | Homo sapiens tyrosinase (TYR), mRNA [NM_000372] |
| A_23_P6293 | 0,041261557 | down | -1,6167684 | UBASH3A | Homo sapiens ubiquitin associated and SH3 domain containing A (UBASH3A), transcript variant 1, mRNA [NM_018961] |
| A_23_P365327 | 0,012106837 | down | -1,5911689 | UBE3D | ubiquitin protein ligase E3D [Source:HGNC Symbol;Acc:HGNC:21381] [ENST00000369746] |
| A_21_P0003740 | 0,036029145 | down | -1,7385359 | UCHL1-AS1 | Homo sapiens UCHL1 antisense RNA 1 (head to head) (UCHL1-AS1), long non-coding RNA [NR_102709] |
| A_21_P0012591 | 0,04274333 | down | -1,5638993 | UGT2B11 | Homo sapiens UDP glucuronosyltransferase 2 family, polypeptide B11 (UGT2B11), mRNA [NM_001073] |
| A_24_P251950 | 0,032581035 | down | -1,6749815 | UGT3A2 | Homo sapiens UDP glycosyltransferase 3 family, polypeptide A2 (UGT3A2), transcript variant 1, mRNA [NM_174914] |
| A_33_P3379454 | 0,023420928 | down | -1,5715766 | UHRF1 | Homo sapiens ubiquitin-like with PHD and ring finger domains 1 (UHRF1), transcript variant 3, mRNA [NM_001290050] |
| A_33_P3355724 | 0,028034156 | down | -1,6178106 | UMODL1 | uromodulin-like 1 [Source:HGNC Symbol;Acc:HGNC:12560] [ENST00000475047] |
| A_33_P3355739 | 0,00532971 | down | -2,5786617 | UMODL1 | Homo sapiens uromodulin-like 1 (UMODL1), transcript variant 4, mRNA [NM_001199528] |
| A_23_P131036 | 0,03945001 | down | -2,0490565 | UPK1A-AS1 | Homo sapiens UPK1A antisense RNA 1 (UPK1A-AS1), antisense RNA [NR_046420] |
| A_23_P422240 | 0,00716381 | down | -2,1393716 | UROC1 | Homo sapiens urocanate hydratase 1 (UROC1), transcript variant 1, mRNA [NM_144639] |
| A_23_P116430 | 0,037442725 | down | -2,0437229 | USH1C | Homo sapiens Usher syndrome 1C (autosomal recessive, severe) (USH1C), transcript variant 1, mRNA [NM_005709] |
| A_32_P189592 | 0,017352445 | down | -1,9643154 | USHBP1 | Homo sapiens Usher syndrome 1C binding protein 1 (USHBP1), transcript variant 1, mRNA [NM_031941] |
| A_24_P366859 | 0,017748082 | down | -2,0791984 | USHBP1 | Homo sapiens Usher syndrome 1C binding protein 1 (USHBP1), transcript variant 1, mRNA [NM_031941] |
| A_23_P400449 | 0,012869964 | down | -1,5607202 | VAT1L | Homo sapiens vesicle amine transport 1-like (VAT1L), mRNA [NM_020927] |
| A_23_P303238 | 0,008245124 | down | -2,2485015 | VN1R5 | Homo sapiens vomeronasal 1 receptor 5 (gene/pseudogene) (VN1R5), mRNA [NM_173858] |
| A_33_P3297030 | 0,040567037 | down | -1,6205988 | VPS53 | vacuolar protein sorting 53 homolog (S. cerevisiae) [Source:HGNC Symbol;Acc:HGNC:25608] [ENST00000570359] |
| A_33_P3514487 | 0,03688778 | down | -2,765136 | VSTM1 | Homo sapiens V-set and transmembrane domain containing 1 (VSTM1), transcript variant 1, mRNA [NM_198481] |
| A_23_P157027 | 0,025446316 | down | -1,5663916 | VSTM2A | Homo sapiens V-set and transmembrane domain containing 2A (VSTM2A), transcript variant 1, mRNA [NM_182546] |
| A_23_P304509 | 0,031054812 | down | -1,7640876 | VSTM4 | Homo sapiens V-set and transmembrane domain containing 4 (VSTM4), transcript variant 2, mRNA [NM_144984] |
| A_33_P3381608 | 0,02186642 | down | -1,5735759 | VWA5B1 | Homo sapiens von Willebrand factor A domain containing 5B1 (VWA5B1), mRNA [NM_001039500] |
| A_21_P0000868 | 0,005715421 | down | -1,7696235 | VWA8-AS1 | Homo sapiens VWA8 antisense RNA 1 (head to head) (VWA8-AS1), long non-coding RNA [NR_039974] |
| A_33_P3304878 | 5,81E-04 | down | -2,8109832 | WDFY4 | Homo sapiens WDFY family member 4 (WDFY4), mRNA [NM_020945] |
| A_33_P3843415 | 0,011776608 | down | -1,781742 | WDR11-AS1 | Homo sapiens WDR11 antisense RNA 1 (WDR11-AS1), long non-coding RNA [NR_033850] |
| A_21_P0010586 | 0,008913534 | down | -1,6191868 | WDR64 | Homo sapiens WD repeat domain 64 (WDR64), mRNA [NM_144625] |
| A_33_P3415491 | 0,045268893 | down | -2,1741624 | WDR90 | Homo sapiens WD repeat domain 90 (WDR90), mRNA [NM_145294] |
| A_33_P3285815 | 0,011177463 | down | -2,7388449 | WFDC6 | WAP four-disulfide core domain 6 [Source:HGNC Symbol;Acc:HGNC:16164] [ENST00000372665] |
| A_24_P942600 | 0,03254825 | down | -1,8915997 | WNK2 | Homo sapiens WNK lysine deficient protein kinase 2 (WNK2), transcript variant 1, mRNA [NM_001282394] |
| A_33_P3377994 | 0,001511849 | down | -1,5869852 | WNK4 | Homo sapiens WNK lysine deficient protein kinase 4 (WNK4), mRNA [NM_032387] |
| A_23_P162322 | 1,84E-04 | down | -2,9866734 | WNT10B | Homo sapiens wingless-type MMTV integration site family, member 10B (WNT10B), mRNA [NM_003394] |
| A_33_P3397525 | 0,037170377 | down | -1,685212 | WNT4 | Homo sapiens wingless-type MMTV integration site family, member 4 (WNT4), mRNA [NM_030761] |
| A_21_P0010968 | 0,019532476 | down | -1,905548 | WT1-AS | Homo sapiens WT1 antisense RNA (WT1-AS), transcript variant 4, long non-coding RNA [NR_120548] |

| | | | | | |
|---|---|---|---|---|---|
| A_32_P345659 | 0,003531585 | down | -2,0300136 | XIRP1 | Homo sapiens xin actin-binding repeat containing 1 (XIRP1), transcript variant 1, mRNA [NM_194293] |
| A_23_P393163 | 0,034352224 | down | -1,6251616 | XIRP2 | Homo sapiens xin actin-binding repeat containing 2 (XIRP2), transcript variant 1, mRNA [NM_152381] |
| A_23_P34183 | 0,02866348 | down | -1,5105858 | XKRY2 | Homo sapiens XK, Kell blood group complex subunit-related, Y-linked 2 (XKRY2), mRNA [NM_001002906] |
| A_33_P3420392 | 0,035549026 | down | -1,504413 | XPNPEP2 | X-prolyl aminopeptidase (aminopeptidase P) 2, membrane-bound [Source:HGNC Symbol;Acc:HGNC:12823] [ENST00000371105] |
| A_24_P21770 | 0,017571697 | down | -2,0008323 | YPEL4 | Homo sapiens yippee-like 4 (Drosophila) (YPEL4), mRNA [NM_145008] |
| A_33_P3283237 | 0,026418805 | down | -1,6907194 | YY2 | Homo sapiens YY2 transcription factor (YY2), mRNA [NM_206923] |
| A_33_P3289167 | 3,82E-04 | down | -2,0359247 | ZBTB32 | Homo sapiens zinc finger and BTB domain containing 32 (ZBTB32), mRNA [NM_014383] |
| A_33_P3339961 | 0,009845624 | down | -1,6638288 | ZBTB8OS | Homo sapiens zinc finger and BTB domain containing 8 opposite strand (ZBTB8OS), mRNA [NM_178547] |
| A_33_P3307820 | 0,040118076 | down | -2,7631676 | ZC2HC1B | Homo sapiens zinc finger, C2HC-type containing 1B (ZC2HC1B), mRNA [NM_001013623] |
| A_33_P3419399 | 0,03198294 | down | -1,7944478 | ZC3H12D | Homo sapiens zinc finger CCCH-type containing 12D (ZC3H12D), mRNA [NM_207360] |
| A_33_P3420500 | 0,047823306 | down | -2,085041 | ZDHHC20 | Homo sapiens zinc finger, DHHC-type containing 20, mRNA (cDNA clone IMAGE:4824131), complete cds. [BC034944] |
| A_21_P0000590 | 0,035697006 | down | -1,5158571 | ZEB2 | Homo sapiens zinc finger E-box binding homeobox 2 (ZEB2), transcript variant 3, non-coding RNA [NR_033258] |
| A_21_P0009057 | 0,015923569 | down | -1,6044217 | ZFHX3 | zinc finger homeobox 3 [Source:HGNC Symbol;Acc:HGNC:777] [ENST00000558842] |
| A_33_P3341851 | 0,027893983 | down | -1,8647861 | ZGRF1 | zinc finger, GRF-type containing 1 [Source:HGNC Symbol;Acc:HGNC:25654] [ENST00000309071] |
| A_23_P327910 | 0,039211668 | down | -1,5859483 | ZIC3 | Homo sapiens Zic family member 3 (ZIC3), mRNA [NM_003413] |
| A_23_P78479 | 2,25E-04 | down | -3,4076486 | ZIM2 | Homo sapiens zinc finger, imprinted 2 (ZIM2), transcript variant 1, mRNA [NM_015363] |
| A_19_P00320274 | 0,035131957 | down | -1,9689277 | ZMIZ1-AS1 | Homo sapiens ZMIZ1 antisense RNA 1 (ZMIZ1-AS1), transcript variant 1, long non-coding RNA [NR_024431] |
| A_21_P0002294 | 0,019555937 | down | -1,6401422 | ZNF112 | zinc finger protein 112 [Source:HGNC Symbol;Acc:HGNC:12892] [ENST00000590687] |
| A_33_P3360912 | 0,024400972 | down | -1,7903016 | ZNF236 | zinc finger protein 236 [Source:HGNC Symbol;Acc:HGNC:13028] [ENST00000543926] |
| A_33_P3399638 | 0,026274096 | down | -2,028254 | ZNF257 | Homo sapiens zinc finger protein 257 (ZNF257), mRNA [NM_033468] |
| A_33_P3629930 | 0,021978818 | down | -2,635195 | ZNF32-AS3 | Homo sapiens ZNF32 antisense RNA 3 (ZNF32-AS3), long non-coding RNA [NR_038867] |
| A_23_P407096 | 0,037482087 | down | -1,633855 | ZNF366 | Homo sapiens zinc finger protein 366 (ZNF366), mRNA [NM_152625] |
| A_21_P0013971 | 0,013959267 | down | -1,9742739 | ZNF385D | zinc finger protein 385D [Source:HGNC Symbol;Acc:HGNC:26191] [ENST00000497570] |
| A_24_P917306 | 0,005543311 | down | -2,5536773 | ZNF385D | zinc finger protein 385D [Source:HGNC Symbol;Acc:HGNC:26191] [ENST00000494108] |
| A_24_P393565 | 0,008360088 | down | -1,9465516 | ZNF396 | zinc finger protein 396 [Source:HGNC Symbol;Acc:HGNC:18824] [ENST00000589332] |
| A_21_P0006771 | 0,011780021 | down | -1,5669092 | ZNF503 | Homo sapiens zinc finger protein 503 (ZNF503), transcript variant 2, non-coding RNA [NR_120651] |
| A_21_P0014669 | 0,03184181 | up | 1,7214284 | ZNF518A | zinc finger protein 518A [Source:HGNC Symbol;Acc:HGNC:29009] [ENST00000563195] |
| A_33_P3345126 | 0,04562894 | down | -1,8368274 | ZNF568 | Homo sapiens zinc finger protein 568 (ZNF568), transcript variant 1, mRNA [NM_198539] |
| A_33_P3245784 | 0,010288716 | down | -1,7738867 | ZNF620 | Homo sapiens zinc finger protein 620 (ZNF620), transcript variant 1, mRNA [NM_175888] |
| A_33_P3243248 | 2,33E-04 | down | -3,6258764 | ZNFX1 | zinc finger, NFX1-type containing 1 [Source:HGNC Symbol;Acc:HGNC:29271] [ENST00000469991] |
| A_23_P89132 | 0,02538322 | down | -1,9572493 | ZP2 | Homo sapiens zona pellucida glycoprotein 2 (sperm receptor) (ZP2), transcript variant 1, mRNA [NM_003460] |
| A_23_P168726 | 0,010678448 | down | -2,6407816 | ZPBP | Homo sapiens zona pellucida binding protein (ZPBP), transcript variant 1, mRNA [NM_007009] |
| A_33_P3406651 | 0,017996123 | down | -2,621658 | ZSCAN5B | Homo sapiens zinc finger and SCAN domain containing 5B (ZSCAN5B), mRNA [NM_001080456] |
