## Supplementary material for "Early Reduction of SARS-CoV-2 Replication in Bronchial Epithelium by Kinin B_2_ Receptor Antagonism": Supl. Table 5

**Supplemental Table S5.** DEGs comparing SARS-CoV-2 + hydrocortisone (HC) versus SARS-CoV-2

| ProbeName | p ([Cov2+HC] \ | Regulatic | FC ([Cov2+H | GeneSymbol | Description |
|---|---|---|---|---|---|
| A_33_P3525263 | 0,029426465 | up | 3,530448 | A2ML1 | Homo sapiens alpha-2-macroglobulin-like 1 (A2ML1), transcript variant 1, mRNA [NM_144670] |
| A_21_P0000904 | 0,010344355 | up | 2,4416256 | A2MP1 | Homo sapiens alpha-2-macroglobulin pseudogene 1 (A2MP1), non-coding RNA [NR_040112] |
| A_33_P3307960 | 0,001662964 | down | -2,1020954 | AADACL3 | Homo sapiens arylacetamide deacetylase-like 3 (AADACL3), transcript variant 1, mRNA [NM_001103170] |
| A_33_P3289204 | 0,005918358 | down | -1,5758368 | AAK1 | Homo sapiens AP2 associated kinase 1 (AAK1), mRNA [NM_014911] |
| A_32_P181527 | 3,85E-04 | down | -2,363122 | AARD | Homo sapiens alanine and arginine rich domain containing protein (AARD), mRNA [NM_001025357] |
| A_33_P3343981 | 6,39E-05 | down | -2,5489085 | AATK | apoptosis-associated tyrosine kinase [Source:HGNC Symbol;Acc:HGNC:21] [ENST00000572798] |
| A_33_P3504522 | 0,031712905 | up | 1,8714286 | AATK-AS1 | Homo sapiens AATK antisense RNA 1 (AATK-AS1), long non-coding RNA [NR_027255] |
| A_33_P3366161 | 0,00659302 | down | -2,394082 | ABAT | Homo sapiens 4-aminobutyrate aminotransferase (ABAT), transcript variant 2, mRNA [NM_000663] |
| A_33_P3239052 | 0,004630672 | up | 1,9099368 | ABCA17P | Homo sapiens ATP-binding cassette, sub-family A (ABC1), member 17, pseudogene (ABCA17P), non-coding RNA [NR_003574] |
| A_33_P3417081 | 0,002699567 | up | 1,7335106 | ABCA17P | Homo sapiens ATP-binding cassette, sub-family A (ABC1), member 17, pseudogene (ABCA17P), non-coding RNA [NR_003574] |
| A_33_P3251672 | 1,92E-04 | down | -2,1948128 | ABCA2 | Homo sapiens ATP-binding cassette, sub-family A (ABC1), member 2 (ABCA2), transcript variant 1, mRNA [NM_001606] |
| A_33_P3342305 | 0,002783157 | up | 1,5834169 | ABCA8 | Homo sapiens ATP-binding cassette, sub-family A (ABC1), member 8 (ABCA8), transcript variant 2, mRNA [NM_007168] |
| A_33_P3332414 | 0,024288127 | up | 2,5256684 | ABCB1 | Homo sapiens ATP-binding cassette, sub-family B (MDR/TAP), member 1 (ABCB1), mRNA [NM_000927] |
| A_33_P3337554 | 0,005962792 | up | 2,5511692 | ABCB5 | Homo sapiens ATP-binding cassette, sub-family B (MDR/TAP), member 5 (ABCB5), transcript variant 4, mRNA [NM_001163993] |
| A_23_P111662 | 0,0417194 | up | 2,0366604 | ABCB5 | Homo sapiens ATP-binding cassette, sub-family B (MDR/TAP), member 5 (ABCB5), transcript variant 2, mRNA [NM_178559] |
| A_33_P3372563 | 0,007973543 | down | -1,7968081 | ABCC10 | Homo sapiens ATP-binding cassette, sub-family C (CFTR/MRP), member 10 (ABCC10), transcript variant MRP7A, mRNA [NM_033450] |
| A_33_P3389518 | 0,02783587 | up | 2,5090327 | ABCC2 | ATP-binding cassette, sub-family C (CFTR/MRP), member 2 [Source:HGNC Symbol;Acc:HGNC:53] [ENST00000370434] |
| A_23_P207507 | 2,26E-04 | up | 3,4624865 | ABCC3 | Homo sapiens ATP-binding cassette, sub-family C (CFTR/MRP), member 3 (ABCC3), transcript variant 1, mRNA [NM_003786] |
| A_33_P3298024 | 4,10E-04 | up | 1,7537702 | ABCC3 | Homo sapiens ATP-binding cassette, sub-family C (CFTR/MRP), member 3 (ABCC3), transcript variant 2, mRNA [NM_001144070] |
| A_23_P15272 | 0,045632645 | down | -1,5595915 | ABCC6 | Homo sapiens ATP-binding cassette, sub-family C (CFTR/MRP), member 6 (ABCC6), transcript variant 2, mRNA [NM_001079528] |
| A_23_P100539 | 0,005381806 | down | -1,7245983 | ABCC6 | Homo sapiens ATP-binding cassette, sub-family C (CFTR/MRP), member 6 (ABCC6), transcript variant 1, mRNA [NM_001171] |
| A_23_P24774 | 0,036295537 | down | -2,1536572 | ABCC8 | Homo sapiens ATP-binding cassette, sub-family C (CFTR/MRP), member 8 (ABCC8), transcript variant 2, mRNA [NM_000352] |
| A_23_P368691 | 0,004399675 | up | 1,5568287 | ABCC9 | Homo sapiens ATP-binding cassette, sub-family C (CFTR/MRP), member 9 (ABCC9), transcript variant SUR2A, mRNA [NM_005691] |
| A_32_P108474 | 0,00827288 | up | 1,5606675 | ABCE1 | Homo sapiens ATP-binding cassette, sub-family E (OABP), member 1 (ABCE1), transcript variant 1, mRNA [NM_002940] |
| A_23_P119763 | 0,014607829 | down | -1,8482424 | ABCG5 | Homo sapiens ATP-binding cassette, sub-family G (WHITE), member 5 (ABCG5), mRNA [NM_022436] |
| A_33_P3333480 | 0,001818699 | down | -1,9954699 | ABHD11-AS1 | Homo sapiens ABHD11 antisense RNA 1 (tail to tail) (ABHD11-AS1), long non-coding RNA [NR_026690] |
| A_33_P3289461 | 6,40E-05 | down | -2,6793847 | ABHD16B | Homo sapiens abhydrolase domain containing 16B (ABHD16B), mRNA [NM_080622] |
| A_23_P369701 | 0,010576213 | up | 2,0081685 | ABHD17C | Homo sapiens abhydrolase domain containing 17C (ABHD17C), mRNA [NM_021214] |
| A_23_P395172 | 0,049621515 | up | 1,5313747 | ABHD2 | Homo sapiens abhydrolase domain containing 2 (ABHD2), transcript variant 1, mRNA [NM_007011] |
| A_23_P211850 | 0,004822654 | down | -1,5097221 | ABHD6 | Homo sapiens abhydrolase domain containing 6 (ABHD6), mRNA [NM_020676] |
| A_24_P933418 | 0,016648674 | up | 1,5108805 | ABI2 | Homo sapiens abl-interactor 2 (ABI2), transcript variant 3, mRNA [NM_005759] |
| A_23_P141429 | 2,31E-04 | down | -1,7596201 | ABI3 | Homo sapiens ABI family, member 3 (ABI3), transcript variant 1, mRNA [NM_016428] |
| A_33_P3411975 | 0,022768965 | down | -2,5573123 | ABI3BP | Homo sapiens ABI family, member 3 (NESH) binding protein (ABI3BP), mRNA [NM_015429] |
| A_33_P3388651 | 0,044935524 | down | -1,9597416 | ABLIM1 | Homo sapiens actin binding LIM protein 1 (ABLIM1), transcript variant 3, mRNA [NM_001003408] |
| A_21_P0006397 | 0,023092113 | down | -1,5499774 | ABO | ABO blood group (transferase A, alpha 1-3-N-acetylgalactosaminyltransferase; transferase B, alpha 1-3-galactosyltransferase) [Source:HGNC Symbol;Acc:HGNC:79] [ENST00000453660] |
| A_23_P356616 | 1,41E-04 | up | 2,1220593 | ABTB2 | Homo sapiens ankyrin repeat and BTB (POZ) domain containing 2 (ABTB2), mRNA [NM_145804] |
| A_23_P87791 | 0,013618453 | down | -1,8317808 | ACACB | Homo sapiens acetyl-CoA carboxylase beta (ACACB), mRNA [NM_001093] |
| A_24_P227450 | 4,12E-04 | down | -1,6584389 | ACAD10 | Homo sapiens acyl-CoA dehydrogenase family, member 10 (ACAD10), transcript variant 2, mRNA [NM_025247] |
| A_23_P47426 | 0,019761082 | up | 1,6119752 | ACAD8 | Homo sapiens acyl-CoA dehydrogenase family, member 8 (ACAD8), mRNA [NM_014384] |
| A_23_P307310 | 0,005543658 | down | -1,845507 | ACAN | Homo sapiens aggrecan (ACAN), transcript variant 2, mRNA [NM_013227] |
| A_33_P3500167 | 0,02227594 | down | -2,0672276 | ACAN | Homo sapiens aggrecan (ACAN), transcript variant 1, mRNA [NM_001135] |
| A_24_P366122 | 9,64E-04 | down | -2,126014 | ACBD4 | Homo sapiens acyl-CoA binding domain containing 4 (ACBD4), transcript variant 2, mRNA [NM_024722] |
| A_23_P72584 | 0,018283261 | down | -1,7503034 | ACBD7 | Homo sapiens acyl-CoA binding domain containing 7 (ACBD7), mRNA [NM_001039844] |
| A_33_P3229288 | 0,023987522 | down | -1,5539296 | ACE | Homo sapiens angiotensin I converting enzyme (ACE), transcript variant 1, mRNA [NM_000789] |
| A_23_P38952 | 0,00350111 | down | -2,871732 | ACER1 | Homo sapiens alkaline ceramidase 1 (ACER1), mRNA [NM_133492] |
| A_33_P3788618 | 0,04782368 | down | -1,7643745 | ACER2 | Homo sapiens alkaline ceramidase 2 (ACER2), mRNA [NM_001010887] |
| A_33_P3413759 | 0,003940203 | down | -1,8196955 | ACER2 | Homo sapiens alkaline ceramidase 2 (ACER2), mRNA [NM_001010887] |
| A_23_P115161 | 0,020368934 | down | -1,6849034 | ACKR1 | Homo sapiens atypical chemokine receptor 1 (Duffy blood group) (ACKR1), transcript variant 2, mRNA [NM_002036] |
| A_23_P9415 | 0,043564785 | up | 1,6299031 | ACO1 | Homo sapiens aconitase 1, soluble (ACO1), transcript variant 2, mRNA [NM_002197] |
| A_23_P325726 | 0,009921301 | down | -1,9248482 | ACOT11 | Homo sapiens acyl-CoA thioesterase 11 (ACOT11), transcript variant 1, mRNA [NM_015547] |
| A_23_P44207 | 4,67E-04 | down | -2,633721 | ACOT12 | Homo sapiens acyl-CoA thioesterase 12 (ACOT12), mRNA [NM_130767] |
| A_33_P3382849 | 0,001268477 | down | -2,1774144 | ACR | Homo sapiens acrosin (ACR), mRNA [NM_001097] |
| A_21_P0014045 | 0,019600458 | up | 1,7970951 | ACR | Homo sapiens acrosin (ACR), mRNA [NM_001097] |
| A_24_P941736 | 0,001584907 | up | 1,7576125 | ACSBG1 | acyl-CoA synthetase bubblegum family member 1 [Source:HGNC Symbol;Acc:HGNC:29567] [ENST00000258873] |
| A_24_P248606 | 0,002776695 | up | 1,5534928 | ACSL3 | Homo sapiens acyl-CoA synthetase long-chain family member 3 (ACSL3), transcript variant 1, mRNA [NM_004457] |
| A_33_P3217983 | 4,68E-04 | down | -3,8882313 | ACSL5 | Homo sapiens acyl-CoA synthetase long-chain family member 5 (ACSL5), transcript variant 3, mRNA [NM_203380] |
| A_33_P3395083 | 0,030824587 | up | 1,6948549 | ACSM4 | Homo sapiens acyl-CoA synthetase medium-chain family member 4 (ACSM4), mRNA [NM_001080454] |

| | | | | | |
|---|---|---|---|---|---|
| A_33_P3339276 | 6,28E-05 | down | -3,4135482 | ACSM5 | Homo sapiens acyl-CoA synthetase medium-chain family member 5 (ACSM5), mRNA [NM_017888] |
| A_33_P3395081 | 0,010583664 | up | 2,189848 | ACSM5 | Homo sapiens acyl-CoA synthetase medium-chain family member 5 (ACSM5), mRNA [NM_017888] |
| A_33_P3400253 | 0,011227565 | up | 1,5034304 | ACSM6 | acyl-CoA synthetase medium-chain family member 6 [Source:HGNC Symbol;Acc:HGNC:31665] [ENST00000327739] |
| A_33_P3255165 | 0,001244836 | down | -2,0750618 | ACTA2-AS1 | Homo sapiens ACTA2 antisense RNA 1 (ACTA2-AS1), long non-coding RNA [NR_125373] |
| A_24_P6903 | 0,008853097 | down | -2,0334089 | ACTBL2 | Homo sapiens actin, beta-like 2 (ACTBL2), mRNA [NM_001017992] |
| A_23_P145681 | 1,97E-04 | down | -2,4584684 | ACTL6B | Homo sapiens actin-like 6B (ACTL6B), mRNA [NM_016188] |
| A_23_P138881 | 0,031939603 | down | -1,5926933 | ACTN3 | Homo sapiens actinin, alpha 3 (gene/pseudogene) (ACTN3), transcript variant 1, coding, mRNA [NM_001104] |
| A_33_P3864411 | 1,10E-04 | down | -3,3315203 | ACTR3BP5 | Homo sapiens ACTR3B pseudogene 5 (ACTR3BP5), non-coding RNA [NR_045000] |
| A_23_P148829 | 3,73E-05 | down | -2,8337343 | ACTRT2 | Homo sapiens actin-related protein T2 (ACTRT2), mRNA [NM_080431] |
| A_33_P3360665 | 0,004474744 | up | 1,5591735 | ACVR1 | Homo sapiens activin A receptor, type I (ACVR1), transcript variant 1, mRNA [NM_001105] |
| A_23_P153930 | 0,017644241 | up | 1,5079288 | ACVR2A | Homo sapiens activin A receptor, type IIA (ACVR2A), transcript variant 2, mRNA [NM_001616] |
| A_33_P3244478 | 0,004662937 | down | -2,028014 | ACVR2B-AS1 | Homo sapiens ACVR2B antisense RNA 1 (ACVR2B-AS1), long non-coding RNA [NR_028389] |
| A_33_P3272402 | 0,02633608 | down | -1,8789531 | ADAD2 | Homo sapiens adenosine deaminase domain containing 2 (ADAD2), transcript variant 1, mRNA [NM_139174] |
| A_21_P0000708 | 4,31E-05 | up | 2,3538947 | ADAM20P1 | Homo sapiens ADAM metallopeptidase domain 20 pseudogene 1 (ADAM20P1), non-coding RNA [NR_037933] |
| A_33_P3216458 | 0,008956674 | down | -2,0490108 | ADAM23 | Homo sapiens ADAM metallopeptidase domain 23 (ADAM23), mRNA [NM_003812] |
| A_23_P126363 | 0,00159325 | up | 3,4996753 | ADAM30 | Homo sapiens ADAM metallopeptidase domain 30 (ADAM30), mRNA [NM_021794] |
| A_23_P44665 | 0,01791134 | up | 1,5582592 | ADAM32 | Homo sapiens ADAM metallopeptidase domain 32 (ADAM32), mRNA [NM_145004] |
| A_33_P3342653 | 3,49E-04 | down | -2,3681595 | ADAM33 | Homo sapiens ADAM metallopeptidase domain 33 (ADAM33), transcript variant 2, mRNA [NM_153202] |
| A_21_P0013537 | 0,045523345 | up | 2,835412 | ADAM5 | ADAM metallopeptidase domain 5, pseudogene [Source:HGNC Symbol;Acc:HGNC:212] [ENST00000497773] |
| A_24_P230026 | 0,004090494 | up | 1,7438173 | ADAM5 | Homo sapiens ADAM metallopeptidase domain 5 (pseudogene) (ADAM5), non-coding RNA [NR_001448] |
| A_23_P94296 | 0,005643754 | down | -2,2715983 | ADAM7 | Homo sapiens ADAM metallopeptidase domain 7 (ADAM7), mRNA [NM_003817] |
| A_33_P3390172 | 0,014125242 | up | 1,5661411 | ADAMDEC1 | Homo sapiens ADAM-like, decysin 1 (ADAMDEC1), transcript variant 2, mRNA [NM_001145271] |
| A_33_P3342780 | 0,005330453 | down | -2,3838043 | ADAMTS13 | Homo sapiens ADAM metallopeptidase with thrombospondin type 1 motif, 13 (ADAMTS13), transcript variant 3, mRNA [NM_139026] |
| A_23_P321307 | 0,001061425 | down | -2,2114859 | ADAMTS2 | Homo sapiens ADAM metallopeptidase with thrombospondin type 1 motif, 2 (ADAMTS2), transcript variant 2, mRNA [NM_021599] |
| A_23_P420359 | 0,002968004 | up | 1,5588328 | ADAMTS20 | Homo sapiens ADAM metallopeptidase with thrombospondin type 1 motif, 20 (ADAMTS20), mRNA [NM_025003] |
| A_33_P3210762 | 0,044484977 | down | -2,289858 | ADAMTS7 | Homo sapiens ADAM metallopeptidase with thrombospondin type 1 motif, 7 (ADAMTS7), mRNA [NM_014272] |
| A_33_P3215557 | 0,001233639 | down | -3,2593195 | ADAMTS7 | Homo sapiens ADAM metallopeptidase with thrombospondin type 1 motif, 7 (ADAMTS7), mRNA [NM_014272] |
| A_23_P11005 | 8,97E-04 | down | -2,1541433 | ADAMTS7 | Homo sapiens ADAM metallopeptidase with thrombospondin type 1 motif, 7 (ADAMTS7), mRNA [NM_014272] |
| A_33_P3286722 | 0,029972412 | up | 1,6552258 | ADAMTS7P1 | Homo sapiens FLJ00317 protein (LOC390660), non-coding RNA [NR_045529] |
| A_32_P196263 | 0,03066857 | up | 3,4846997 | ADAMTS9 | Homo sapiens ADAM metallopeptidase with thrombospondin type 1 motif, 9 (ADAMTS9), mRNA [NM_182920] |
| A_21_P0000741 | 0,00957908 | down | -2,1032197 | ADAMTS9-AS2 | Homo sapiens ADAMTS9 antisense RNA 2 (ADAMTS9-AS2), long non-coding RNA [NR_038264] |
| A_23_P342138 | 0,026067015 | down | -1,7030599 | ADAMTSL1 | Homo sapiens ADAMTS-like 1 (ADAMTSL1), transcript variant 4, mRNA [NM_001040272] |
| A_33_P3312509 | 0,004562189 | down | -1,8096466 | ADAMTSL2 | Homo sapiens ADAMTS-like 2 (ADAMTSL2), transcript variant 1, mRNA [NM_014694] |
| A_23_P20697 | 9,63E-04 | down | -1,6589369 | ADAMTSL2 | Homo sapiens ADAMTS-like 2 (ADAMTSL2), transcript variant 1, mRNA [NM_014694] |
| A_23_P115011 | 0,04471734 | up | 1,6426224 | ADAMTSL4 | Homo sapiens ADAMTS-like 4 (ADAMTSL4), transcript variant 1, mRNA [NM_019032] |
| A_33_P3245489 | 0,010311194 | down | -2,0192525 | ADAMTSL5 | Homo sapiens ADAMTS-like 5 (ADAMTSL5), mRNA [NM_213604] |
| A_33_P3399061 | 0,001725975 | down | -1,5418559 | ADARB1 | Homo sapiens adenosine deaminase, RNA-specific, B1 (ADARB1), transcript variant 4, non-coding RNA [NR_027673] |
| A_32_P29954 | 0,043392982 | down | -1,8932555 | ADARB2-AS1 | Homo sapiens ADARB2 antisense RNA 1 (ADARB2-AS1), long non-coding RNA [NR_033387] |
| A_23_P67864 | 0,013476179 | up | 1,5341223 | ADCY3 | Homo sapiens adenylate cyclase 3 (ADCY3), mRNA [NM_004036] |
| A_23_P501547 | 0,003848887 | up | 1,7392433 | ADCY6 | Homo sapiens adenylate cyclase 6 (ADCY6), transcript variant 1, mRNA [NM_015270] |
| A_33_P3241782 | 0,009068268 | up | 2,8748028 | ADD2 | Homo sapiens adducin 2 (beta) (ADD2), transcript variant 1, mRNA [NM_001617] |
| A_23_P421436 | 0,030845629 | up | 1,5464028 | ADD2 | Homo sapiens adducin 2 (beta) (ADD2), transcript variant 4, mRNA [NM_017488] |
| A_23_P70643 | 0,002273397 | up | 1,8131804 | ADGB | Homo sapiens androglobin (ADGB), mRNA [NM_024694] |
| A_23_P81158 | 2,92E-07 | down | -7,077289 | ADH1C | Homo sapiens alcohol dehydrogenase 1C (class I), gamma polypeptide (ADH1C), mRNA [NM_000669] |
| A_33_P3300747 | 0,03590694 | down | -1,8467563 | ADHFE1 | Homo sapiens alcohol dehydrogenase, iron containing, 1 (ADHFE1), mRNA [NM_144650] |
| A_23_P369237 | 0,008858528 | down | -1,7243252 | ADIPOQ | Homo sapiens adiponectin, C1Q and collagen domain containing (ADIPOQ), transcript variant 2, mRNA [NM_004797] |
| A_33_P3248765 | 0,04501125 | down | -1,5417329 | ADIPOR1 | Homo sapiens adiponectin receptor 1 (ADIPOR1), transcript variant 3, mRNA [NM_001290557] |
| A_23_P127948 | 0,01370018 | up | 1,6644799 | ADM | Homo sapiens adrenomedullin (ADM), mRNA [NM_001124] |
| A_33_P3413910 | 0,012669004 | down | -1,5888822 | ADM2 | Homo sapiens adrenomedullin 2 (ADM2), transcript variant 1, mRNA [NM_024866] |
| A_23_P254179 | 1,19E-04 | up | 1,7140367 | ADNP | Homo sapiens activity-dependent neuroprotector homeobox (ADNP), transcript variant 1, mRNA [NM_015339] |
| A_24_P237270 | 0,013966318 | up | 2,6195738 | ADORA2A | Homo sapiens adenosine A2a receptor (ADORA2A), transcript variant 3, mRNA [NM_000675] |
| A_21_P0000570 | 0,0036484 | down | -2,696237 | ADORA2A-AS1 | Homo sapiens ADORA2A antisense RNA 1 (ADORA2A-AS1), transcript variant 2, long non-coding RNA [NR_028483] |
| A_23_P14716 | 3,24E-04 | up | 1,907414 | ADPGK | Homo sapiens ADP-dependent glucokinase (ADPGK), transcript variant 1, mRNA [NM_031284] |
| A_23_P138706 | 0,012365628 | up | 2,0762808 | ADRA2A | Homo sapiens adrenoceptor alpha 2A (ADRA2A), mRNA [NM_000681] |
| A_23_P145024 | 0,002872571 | up | 1,7600032 | ADRB2 | Homo sapiens adrenoceptor beta 2, surface (ADRB2), mRNA [NM_000024] |
| A_23_P201432 | 4,62E-04 | up | 1,6679738 | ADSS | Homo sapiens adenylosuccinate synthase (ADSS), mRNA [NM_001126] |
| A_23_P206077 | 0,015299762 | up | 1,7301587 | AEN | Homo sapiens apoptosis enhancing nuclease (AEN), mRNA [NM_022767] |
| A_24_P414332 | 1,98E-04 | up | 2,9045258 | AFF1 | Homo sapiens AF4/FMR2 family, member 1 (AFF1), transcript variant 2, mRNA [NM_005935] |
| A_33_P3293888 | 0,004467628 | down | -2,0620875 | AFF2 | Homo sapiens AF4/FMR2 family, member 2 (AFF2), transcript variant 1, mRNA [NM_002025] |
| A_33_P3373348 | 0,015525769 | down | -2,2335885 | AFF3 | AF4/FMR2 family, member 3 [Source:HGNC Symbol;Acc:HGNC:6473] [ENST00000483600] |
| A_21_P0014083 | 0,029103717 | down | -2,4790573 | AGAP1-IT1 | PREDICTED: Homo sapiens AGAP1 intronic transcript 1 (non-protein coding) (AGAP1-IT1), ncRNA [XR_108380] |

| | | | | | |
|---|---|---|---|---|---|
| A_33_P3375435 | 0,003405481 | down | -1,5671016 | AGAP11 | Homo sapiens ankyrin repeat and GTPase domain Arf GTPase activating protein 11 (AGAP11), mRNA [NM_133447] |
| A_32_P41526 | 0,006955841 | up | 1,6513537 | AGAP5 | Homo sapiens ArfGAP with GTPase domain, ankyrin repeat and PH domain 5 (AGAP5), mRNA [NM_001144000] |
| A_33_P3355418 | 0,03584851 | down | -1,5230759 | AGBL1 | Homo sapiens ATP/GTP binding protein-like 1 (AGBL1), mRNA [NM_152336] |
| A_32_P82623 | 0,006135705 | down | -1,6526597 | AGBL3 | Homo sapiens ATP/GTP binding protein-like 3 (AGBL3), mRNA [NM_178563] |
| A_33_P3378101 | 0,008360048 | down | -1,606262 | AGBL4 | Homo sapiens ATP/GTP binding protein-like 4 (AGBL4), mRNA [NM_032785] |
| A_33_P3386572 | 0,008979002 | down | -2,5810676 | AGBL4 | ATP/GTP binding protein-like 4 [Source:HGNC Symbol;Acc:HGNC:25892] [ENST00000497451] |
| A_33_P3238993 | 0,001415354 | up | 1,7832907 | AGFG1 | Homo sapiens ArfGAP with FG repeats 1 (AGFG1), transcript variant 1, mRNA [NM_001135187] |
| A_23_P200298 | 0,011998847 | up | 1,5337645 | AGL | Homo sapiens amylo-alpha-1, 6-glucosidase, 4-alpha-glucanotransferase (AGL), transcript variant 4, mRNA [NM_000028] |
| A_33_P3402116 | 0,002777017 | down | -2,3888633 | AGO1 | Homo sapiens argonaute RISC catalytic component 1 (AGO1), mRNA [NM_012199] |
| A_23_P31407 | 0,003832783 | down | -2,7187066 | AGR2 | Homo sapiens anterior gradient 2 (AGR2), mRNA [NM_006408] |
| A_33_P3215797 | 0,037382234 | down | -1,7725457 | AHDC1 | Homo sapiens AT hook, DNA binding motif, containing 1 (AHDC1), mRNA [NM_001029882] |
| A_24_P943393 | 0,001716839 | up | 1,5964024 | AHNAK | Homo sapiens AHNAK nucleoprotein (AHNAK), transcript variant 1, mRNA [NM_001620] |
| A_33_P3222380 | 0,007465863 | down | -1,8567657 | AHNAK2 | Homo sapiens AHNAK nucleoprotein 2 (AHNAK2), mRNA [NM_138420] |
| A_33_P3368560 | 8,44E-06 | down | -4,076132 | AHSA2 | AHA1, activator of heat shock 90kDa protein ATPase homolog 2 (yeast) [Source:HGNC Symbol;Acc:HGNC:20437] [ENST00000394457] |
| A_23_P155514 | 0,01874555 | down | -2,168846 | AHSG | Homo sapiens alpha-2-HS-glycoprotein (AHSG), mRNA [NM_001622] |
| A_33_P3250857 | 2,24E-04 | down | -2,1244879 | AIDA | axin interactor, dorsalization associated [Source:HGNC Symbol;Acc:HGNC:25761] [ENST00000474863] |
| A_23_P214627 | 0,022656925 | up | 2,3285694 | AIF1 | Homo sapiens allograft inflammatory factor 1 (AIF1), transcript variant 2, mRNA [NM_004847] |
| A_24_P308229 | 0,011156531 | up | 1,6835155 | AIM1L | Homo sapiens absent in melanoma 1-like (AIM1L), mRNA [NM_001039775] |
| A_23_P68740 | 0,006984205 | down | -2,614945 | AIRE | Homo sapiens autoimmune regulator (AIRE), mRNA [NM_000383] |
| A_33_P3307253 | 2,50E-04 | down | -2,5982091 | AK5 | Homo sapiens adenylate kinase 5 (AK5), transcript variant 2, mRNA [NM_012093] |
| A_23_P83200 | 7,96E-04 | down | -2,3167734 | AK8 | Homo sapiens adenylate kinase 8 (AK8), mRNA [NM_152572] |
| A_23_P348749 | 0,03900661 | up | 1,5676278 | AK9 | Homo sapiens adenylate kinase 9 (AK9), transcript variant 2, mRNA [NM_145025] |
| A_24_P142095 | 5,81E-06 | up | 2,2052712 | AKAP13 | Homo sapiens A kinase (PRKA) anchor protein 13 (AKAP13), transcript variant 1, mRNA [NM_006738] |
| A_33_P3417920 | 0,001822032 | down | -2,6352181 | AKAP4 | Homo sapiens A kinase (PRKA) anchor protein 4 (AKAP4), transcript variant 1, mRNA [NM_003886] |
| A_33_P3220047 | 0,024796136 | down | -1,7948828 | AKNA | Homo sapiens AT-hook transcription factor (AKNA), mRNA [NM_030767] |
| A_33_P3297853 | 0,012681669 | up | 2,072086 | AKNA | Homo sapiens AT-hook transcription factor (AKNA), mRNA [NM_030767] |
| A_33_P3365117 | 0,02401498 | down | -2,3901718 | AKR1C1 | aldo-keto reductase family 1, member C1 [Source:HGNC Symbol;Acc:HGNC:384] [ENST00000380859] |
| A_33_P3329028 | 0,048200592 | down | -1,573146 | AKR1C8P | Homo sapiens aldo-keto reductase family 1, member C8, pseudogene (AKR1C8P), non-coding RNA [NR_027916] |
| A_23_P382000 | 0,007212819 | up | 1,5880212 | AKR1D1 | Homo sapiens aldo-keto reductase family 1, member D1 (AKR1D1), transcript variant 1, mRNA [NM_005989] |
| A_33_P3218004 | 0,006336816 | up | 1,8036532 | AKR7L | Homo sapiens aldo-keto reductase family 7-like (gene/pseudogene) (AKR7L), transcript variant 1, non-coding RNA [NR_040288] |
| A_21_P0008700 | 0,002198689 | down | -3,0510697 | ALDH1A3 | PREDICTED: Homo sapiens uncharacterized LOC145757 (RP11-66B24.1), transcript variant X1, misc_RNA [XR_109231] |
| A_23_P205959 | 0,016692081 | up | 2,1798975 | ALDH1A3 | Homo sapiens aldehyde dehydrogenase 1 family, member A3 (ALDH1A3), transcript variant 1, mRNA [NM_000693] |
| A_33_P3282634 | 0,005763746 | down | -2,8167398 | ALDH1L2 | Homo sapiens aldehyde dehydrogenase 1 family, member L2 (ALDH1L2), transcript variant 1, mRNA [NM_001034173] |
| A_23_P32143 | 1,40E-04 | down | -2,5945826 | ALDOB | Homo sapiens aldolase B, fructose-bisphosphate (ALDOB), mRNA [NM_000035] |
| A_24_P795594 | 8,92E-04 | up | 1,8017913 | ALG9 | Homo sapiens ALG9, alpha-1,2-mannosyltransferase (ALG9), transcript variant 3, mRNA [NM_001077691] |
| A_21_P0000832 | 0,046364613 | up | 2,166625 | ALKBH3-AS1 | Homo sapiens ALKBH3 antisense RNA 1 (ALKBH3-AS1), long non-coding RNA [NR_038907] |
| A_23_P371876 | 0,001106634 | down | -1,6448562 | ALKBH8 | Homo sapiens alkB, alkylation repair homolog 8 (E. coli) (ALKBH8), transcript variant 2, mRNA [NM_138775] |
| A_33_P3328445 | 0,003449616 | up | 1,5680072 | ALOX12-AS1 | Homo sapiens cDNA FLJ90099 fis, clone HEMBA1006016. [AK074580] |
| A_21_P0011615 | 0,019961353 | down | -1,5215989 | ALOX12P2 | Homo sapiens arachidonate 12-lipoxygenase pseudogene 2 (ALOX12P2), transcript variant 1, non-coding RNA [NR_002710] |
| A_32_P440768 | 0,016271768 | down | -2,0284593 | ALOX12P2 | Homo sapiens arachidonate 12-lipoxygenase pseudogene 2 (ALOX12P2), transcript variant 1, non-coding RNA [NR_002710] |
| A_21_P0011510 | 0,018983087 | down | -1,910281 | ALOX12P2 | Homo sapiens arachidonate 12-lipoxygenase pseudogene 2 (ALOX12P2), transcript variant 1, non-coding RNA [NR_002710] |
| A_23_P60627 | 7,64E-05 | up | 2,623692 | ALOX15B | Homo sapiens arachidonate 15-lipoxygenase, type B (ALOX15B), transcript variant d, mRNA [NM_001141] |
| A_24_P347378 | 0,001915561 | down | -2,8134346 | ALOX5AP | Homo sapiens arachidonate 5-lipoxygenase-activating protein (ALOX5AP), transcript variant 1, mRNA [NM_001629] |
| A_23_P144531 | 0,01764902 | down | -1,5003316 | ALPK1 | Homo sapiens cDNA: FLJ22670 fis, clone HSI08684. [AK026323] |
| A_33_P3210099 | 7,71E-05 | up | 2,0061088 | ALPK3 | Homo sapiens alpha-kinase 3 (ALPK3), mRNA [NM_020778] |
| A_23_P79587 | 7,87E-04 | down | -1,917896 | ALPP | alkaline phosphatase, placental [Source:HGNC Symbol;Acc:HGNC:439] [ENST00000485563] |
| A_24_P131580 | 0,001577825 | down | -2,1451337 | ALPPL2 | Homo sapiens alkaline phosphatase, placental-like 2 (ALPPL2), mRNA [NM_031313] |
| A_32_P420009 | 0,001209005 | up | 1,7932116 | ALS2CL | Homo sapiens ALS2 C-terminal like (ALS2CL), transcript variant 1, mRNA [NM_147129] |
| A_21_P0000072 | 0,001730534 | up | 1,8883623 | ALS2CR11 | Homo sapiens amyotrophic lateral sclerosis 2 (juvenile) chromosome region, candidate 11 (ALS2CR11), transcript variant 3, mRNA [NM_001168217] |
| A_23_P256504 | 0,038529847 | up | 1,5191715 | AMBP | Homo sapiens alpha-1-microglobulin/bikunin precursor (AMBP), mRNA [NM_001633] |
| A_23_P353742 | 0,001261096 | up | 1,5682698 | AMBRA1 | Homo sapiens autophagy/beclin-1 regulator 1 (AMBRA1), transcript variant 2, mRNA [NM_017749] |
| A_33_P3395369 | 0,031860378 | down | -1,5717801 | AMD1 | Homo sapiens adenosylmethionine decarboxylase 1 (AMD1), transcript variant 1, mRNA [NM_001634] |
| A_32_P122226 | 0,005964098 | down | -1,9636006 | AMDHD1 | Homo sapiens amidohydrolase domain containing 1 (AMDHD1), mRNA [NM_152435] |
| A_23_P96478 | 0,008781098 | down | -1,5819442 | AMELX | Homo sapiens amelogenin, X-linked (AMELX), transcript variant 2, mRNA [NM_182681] |
| A_23_P48455 | 0,002895262 | down | -2,2500978 | AMN | Homo sapiens amnion associated transmembrane protein (AMN), mRNA [NM_030943] |
| A_33_P3248152 | 0,035998587 | up | 1,6223426 | AMPD1 | Homo sapiens adenosine monophosphate deaminase 1 (AMPD1), transcript variant 1, mRNA [NM_000036] |
| A_23_P31273 | 1,93E-04 | down | -3,3505127 | AMPH | Homo sapiens amphiphysin (AMPH), transcript variant 1, mRNA [NM_001635] |
| A_23_P23611 | 0,010142086 | up | 1,6799506 | AMY1C | Homo sapiens amylase, alpha 1C (salivary) (AMY1C), mRNA [NM_001008219] |
| A_33_P3380837 | 0,003610633 | down | -1,5970545 | AMZ1 | Homo sapiens archaelysin family metallopeptidase 1 (AMZ1), transcript variant 1, mRNA [NM_133463] |
| A_23_P44643 | 0,001730785 | up | 1,7816619 | ANAPC7 | Homo sapiens anaphase promoting complex subunit 7 (ANAPC7), transcript variant 1, mRNA [NM_016238] |
| A_23_P216023 | 2,87E-05 | down | -2,6406276 | ANGPT1 | Homo sapiens angiopoietin 1 (ANGPT1), transcript variant 1, mRNA [NM_001146] |

| | | | | | |
|---|---|---|---|---|---|
| A_23_P60079 | 1,69E-04 | up | 3,1915596 | ANGPT2 | Homo sapiens angiopoietin 2 (ANGPT2), transcript variant 1, mRNA [NM_001147] |
| A_23_P68511 | 9,79E-04 | down | -2,526863 | ANGPT4 | Homo sapiens angiopoietin 4 (ANGPT4), mRNA [NM_015985] |
| A_24_P199484 | 0,011069021 | up | 1,6644821 | ANGPTL1 | Homo sapiens angiopoietin-like 1 (ANGPTL1), mRNA [NM_004673] |
| A_33_P3295358 | 0,003626651 | up | 4,529974 | ANGPTL4 | Homo sapiens angiopoietin-like 4 (ANGPTL4), transcript variant 1, mRNA [NM_139314] |
| A_23_P216108 | 0,033264194 | down | -1,6853294 | ANK1 | Homo sapiens ankyrin 1, erythrocytic (ANK1), transcript variant 3, mRNA [NM_000037] |
| A_33_P3376321 | 0,03833149 | up | 1,763552 | ANK1 | Homo sapiens ankyrin 1, erythrocytic (ANK1), transcript variant 2, mRNA [NM_020477] |
| A_24_P162173 | 0,03494861 | down | -1,5669458 | ANK3 | Homo sapiens ankyrin 3, node of Ranvier (ankyrin G) (ANK3), transcript variant 2, mRNA [NM_001149] |
| A_23_P202269 | 0,012309408 | up | 1,595854 | ANK3 | Homo sapiens ankyrin 3, node of Ranvier (ankyrin G) (ANK3), transcript variant 1, mRNA [NM_020987] |
| A_23_P365086 | 5,27E-04 | down | -2,2261803 | ANKEF1 | ankyrin repeat and EF-hand domain containing 1 [Source:HGNC Symbol;Acc:HGNC:15803] [ENST00000378392] |
| A_23_P350698 | 0,04327582 | up | 1,7760044 | ANKFN1 | Homo sapiens ankyrin-repeat and fibronectin type III domain containing 1 (ANKFN1), mRNA [NM_153228] |
| A_33_P3300142 | 0,003123338 | down | -1,9835418 | ANKFY1 | Homo sapiens ankyrin repeat and FYVE domain containing 1 (ANKFY1), transcript variant 3, mRNA [NM_001257999] |
| A_24_P151834 | 1,59E-04 | up | 1,9421685 | ANKFY1 | ankyrin repeat and FYVE domain containing 1 [Source:HGNC Symbol;Acc:HGNC:20763] [ENST00000570934] |
| A_23_P355623 | 2,98E-04 | up | 1,8783357 | ANKIB1 | Homo sapiens ankyrin repeat and IBR domain containing 1 (ANKIB1), mRNA [NM_019004] |
| A_33_P3629247 | 2,15E-05 | down | -2,8871994 | ANKMY1 | Homo sapiens ankyrin repeat and MYND domain containing 1 (ANKMY1), transcript variant 2, mRNA [NM_017844] |
| A_23_P20002 | 7,59E-04 | down | -2,0567007 | ANKMY2 | Homo sapiens ankyrin repeat and MYND domain containing 2 (ANKMY2), mRNA [NM_020319] |
| A_23_P205046 | 0,002181487 | up | 1,8088802 | ANKRD10 | Homo sapiens ankyrin repeat domain 10 (ANKRD10), transcript variant 1, mRNA [NM_017664] |
| A_24_P324405 | 0,038336452 | down | -1,5627742 | ANKRD11 | Homo sapiens ankyrin repeat domain 11 (ANKRD11), transcript variant 2, mRNA [NM_013275] |
| A_24_P98975 | 0,003115999 | up | 2,1638896 | ANKRD13A | Homo sapiens ankyrin repeat domain 13A (ANKRD13A), mRNA [NM_033121] |
| A_23_P380881 | 0,018299524 | up | 1,5613228 | ANKRD13B | Homo sapiens ankyrin repeat domain 13B (ANKRD13B), mRNA [NM_152345] |
| A_33_P3238941 | 0,023405317 | down | -1,6474557 | ANKRD18A | Homo sapiens ankyrin repeat domain 18A (ANKRD18A), mRNA [NM_147195] |
| A_33_P3236698 | 0,02733214 | down | -1,5753028 | ANKRD19P | Homo sapiens cDNA FLJ77364 complete cds, highly similar to Homo sapiens ankyrin repeat domain 19 (ANKRD19), mRNA. [AK292218] |
| A_32_P79190 | 0,02829047 | down | -1,6032921 | ANKRD20A12P | Homo sapiens ankyrin repeat domain 20 family, member A12, pseudogene (ANKRD20A12P), non-coding RNA [NR_046228] |
| A_21_P0010552 | 0,010922704 | down | -2,678001 | ANKRD20A2 | Homo sapiens ankyrin repeat domain 20 family, member A2 (ANKRD20A2), mRNA [NM_001012421] |
| A_21_P0013636 | 0,011390762 | down | -2,130555 | ANKRD20A8P | Homo sapiens ankyrin repeat domain 20 family, member A8, pseudogene (ANKRD20A8P), non-coding RNA [NR_003366] |
| A_33_P3283599 | 0,021296803 | down | -1,5555745 | ANKRD20A8P | Homo sapiens ankyrin repeat domain 20 family, member A8, pseudogene (ANKRD20A8P), non-coding RNA [NR_003366] |
| A_23_P153640 | 7,21E-04 | up | 1,589114 | ANKRD27 | Homo sapiens ankyrin repeat domain 27 (VPS9 domain) (ANKRD27), mRNA [NM_032139] |
| A_33_P3368985 | 0,004434611 | up | 1,6193372 | ANKRD30A | Homo sapiens ankyrin repeat domain 30A (ANKRD30A), mRNA [NM_052997] |
| A_33_P3343452 | 0,010613969 | up | 1,6300339 | ANKRD30B | Homo sapiens ankyrin repeat domain 30B (ANKRD30B), mRNA [NM_001145029] |
| A_24_P917819 | 3,37E-04 | up | 3,1309278 | ANKRD30BP2 | Homo sapiens ankyrin repeat domain 30B pseudogene 2 (ANKRD30BP2), non-coding RNA [NR_026916] |
| A_21_P0000612 | 0,026807783 | up | 1,5672113 | ANKRD30BP3 | Homo sapiens ankyrin repeat domain 30B pseudogene 3 (ANKRD30BP3), non-coding RNA [NR_033891] |
| A_32_P516342 | 0,005262137 | down | -1,5526334 | ANKRD33 | Homo sapiens ankyrin repeat domain 33 (ANKRD33), transcript variant 2, mRNA [NM_182608] |
| A_33_P3213468 | 0,047288768 | up | 1,7614316 | ANKRD33B | Homo sapiens ankyrin repeat domain 33B (ANKRD33B), mRNA [NM_001164440] |
| A_33_P3386581 | 0,002379512 | up | 1,5572565 | ANKRD40 | Homo sapiens ankyrin repeat domain 40 (ANKRD40), mRNA [NM_052855] |
| A_33_P3315325 | 0,04124088 | up | 2,2684429 | ANKRD46 | Homo sapiens ankyrin repeat domain 46 (ANKRD46), transcript variant 4, mRNA [NM_001270379] |
| A_33_P3267305 | 0,017259978 | up | 1,5573055 | ANKRD52 | Homo sapiens ankyrin repeat domain 52 (ANKRD52), mRNA [NM_173595] |
| A_33_P3278455 | 0,01209993 | up | 1,5690844 | ANKRD6 | Homo sapiens ankyrin repeat domain 6 (ANKRD6), transcript variant 2, mRNA [NM_001242811] |
| A_23_P131875 | 0,02429793 | down | -1,7589577 | ANKRD60 | ankyrin repeat domain 60 [Source:HGNC Symbol;Acc:HGNC:16217] [ENST00000457363] |
| A_33_P3321522 | 0,019463852 | down | -1,9857504 | ANKRD61 | Homo sapiens ankyrin repeat domain 61 (ANKRD61), mRNA [NM_001271700] |
| A_21_P0014906 | 0,008241392 | up | 2,303644 | ANKRD62 | Homo sapiens ankyrin repeat domain 62 (ANKRD62), mRNA [NM_001277333] |
| A_33_P3215298 | 0,00331886 | up | 1,6017898 | ANKRD62 | Homo sapiens ankyrin repeat domain 62 (ANKRD62), mRNA [NM_001277333] |
| A_21_P0000033 | 0,001027453 | down | -1,7841551 | ANKRD66 | Homo sapiens ankyrin repeat domain 66 (ANKRD66), mRNA [NM_001162435] |
| A_33_P3315510 | 0,00700174 | down | -2,2131946 | ANKS1A | Homo sapiens ankyrin repeat and sterile alpha motif domain containing 1A, mRNA (cDNA clone MGC:42354 IMAGE:4819948), complete cds. [BC031934] |
| A_33_P3258003 | 0,012545034 | up | 2,5905426 | ANKS1B | Homo sapiens ankyrin repeat and sterile alpha motif domain containing 1B (ANKS1B), transcript variant 1, mRNA [NM_152788] |
| A_23_P119907 | 0,005921348 | up | 1,5092171 | ANKZF1 | Homo sapiens ankyrin repeat and zinc finger domain containing 1 (ANKZF1), transcript variant 1, mRNA [NM_018089] |
| A_24_P87036 | 0,047388364 | up | 1,8801037 | ANO1 | Homo sapiens anoctamin 1, calcium activated chloride channel (ANO1), transcript variant 1, mRNA [NM_018043] |
| A_23_P409093 | 0,01690907 | up | 2,5766165 | ANO4 | Homo sapiens anoctamin 4 (ANO4), transcript variant 3, mRNA [NM_178826] |
| A_33_P3247342 | 0,026236312 | down | -1,945768 | ANO7 | Homo sapiens anoctamin 7 (ANO7), transcript variant NGEP-S, mRNA [NM_001001666] |
| A_23_P14649 | 3,28E-04 | down | -1,9868567 | ANP32A-IT1 | Homo sapiens ANP32A intronic transcript 1 (non-protein coding) (ANP32A-IT1), long non-coding RNA [NR_026808] |
| A_23_P92520 | 0,005396168 | down | -2,222245 | ANP32C | Homo sapiens acidic (leucine-rich) nuclear phosphoprotein 32 family, member C (ANP32C), mRNA [NM_012403] |
| A_33_P3334803 | 0,016679745 | down | -1,8048469 | ANTXRL | PREDICTED: Homo sapiens anthrax toxin receptor-like (ANTXRL), misc_RNA [XR_159158] |
| A_33_P3360296 | 0,007221441 | down | -2,1171846 | ANTXRL | Homo sapiens anthrax toxin receptor-like (ANTXRL), mRNA [NM_001278688] |
| A_21_P0010873 | 0,007487498 | up | 2,0783598 | ANTXRLP1 | Homo sapiens anthrax toxin receptor-like pseudogene 1 (ANTXRLP1), transcript variant 2, non-coding RNA [NR_103828] |
| A_32_P150632 | 3,38E-05 | up | 2,3079174 | ANXA11 | annexin A11 [Source:HGNC Symbol;Acc:HGNC:535] [ENST00000438331] |
| A_32_P148345 | 4,11E-04 | down | -3,3306816 | ANXA2 | Homo sapiens annexin A2 (ANXA2), transcript variant 2, mRNA [NM_001002857] |
| A_24_P323114 | 0,026825327 | down | -1,5443736 | ANXA2P3 | Homo sapiens annexin A2 pseudogene 3 (ANXA2P3), non-coding RNA [NR_001446] |
| A_23_P426305 | 0,004401853 | up | 1,8137885 | AOC3 | Homo sapiens amine oxidase, copper containing 3 (AOC3), transcript variant 1, mRNA [NM_003734] |
| A_33_P3354414 | 0,03652855 | up | 2,1845255 | AOX1 | Homo sapiens aldehyde oxidase 1 (AOX1), mRNA [NM_001159] |
| A_21_P0012040 | 0,040383544 | down | -1,5373157 | AOX2P | Homo sapiens aldehyde oxidase 2 pseudogene (AOX2P), non-coding RNA [NR_001557] |
| A_19_P00322315 | 0,005855204 | up | 1,8821987 | AOX2P | Homo sapiens aldehyde oxidase 2 pseudogene (AOX2P), non-coding RNA [NR_001557] |
| A_21_P0012322 | 0,04839082 | down | -1,5199943 | AP1B1P1 | Homo sapiens adaptor-related protein complex 1, beta 1 subunit pseudogene 1 (AP1B1P1), non-coding RNA [NR_040114] |
| A_24_P937306 | 0,01365073 | up | 1,8422337 | AP1S2 | adaptor-related protein complex 1, sigma 2 subunit [Source:HGNC Symbol;Acc:HGNC:560] [ENST00000380291] |

| | | | | | |
|---|---|---|---|---|---|
| A_24_P105747 | 0,015969923 | up | 1,5530336 | AP1S3 | Homo sapiens adaptor-related protein complex 1, sigma 3 subunit (AP1S3), transcript variant 1, mRNA [NM_001039569] |
| A_24_P9883 | 0,003459801 | up | 1,8960917 | AP5B1 | Homo sapiens adaptor-related protein complex 5, beta 1 subunit (AP5B1), mRNA [NM_138368] |
| A_33_P3295655 | 0,005078261 | up | 2,317328 | APBA1 | Homo sapiens amyloid beta (A4) precursor protein-binding, family A, member 1 (APBA1), mRNA [NM_001163] |
| A_33_P3251901 | 0,001223244 | down | -1,6067699 | APBB2 | Homo sapiens amyloid beta (A4) precursor protein-binding, family B, member 2 (APBB2), transcript variant 1, mRNA [NM_004307] |
| A_23_P96827 | 0,003259553 | down | -1,6744944 | APCS | Homo sapiens amyloid P component, serum (APCS), mRNA [NM_001639] |
| A_24_P312692 | 0,001868962 | up | 1,6060324 | API5 | Homo sapiens apoptosis inhibitor 5 (API5), transcript variant 2, mRNA [NM_006595] |
| A_23_P334246 | 0,004398041 | up | 1,5317397 | APLF | Homo sapiens aprataxin and PNKP like factor (APLF), mRNA [NM_173545] |
| A_23_P113393 | 0,023416903 | down | -1,6063217 | APLN | Homo sapiens apelin (APLN), mRNA [NM_017413] |
| A_32_P9543 | 0,002336655 | up | 4,6148553 | APOBEC3A | Homo sapiens apolipoprotein B mRNA editing enzyme, catalytic polypeptide-like 3A (APOBEC3A), transcript variant 1, mRNA [NM_145699] |
| A_32_P347617 | 0,013836287 | up | 1,50879 | APOBEC3H | Homo sapiens apolipoprotein B mRNA editing enzyme, catalytic polypeptide-like 3H (APOBEC3H), transcript variant SV-183, mRNA [NM_181773] |
| A_24_P109214 | 0,002347909 | down | -1,508793 | APOC1 | Homo sapiens apolipoprotein C-I (APOC1), mRNA [NM_001645] |
| A_23_P87491 | 0,037531596 | up | 2,5087702 | APOF | Homo sapiens apolipoprotein F (APOF), mRNA [NM_001638] |
| A_33_P3232523 | 0,011679252 | down | -2,3184888 | APOL4 | apolipoprotein L, 4 [Source:HGNC Symbol;Acc:HGNC:14867] [ENST00000328429] |
| A_23_P380857 | 0,02434637 | down | -2,7622771 | APOL4 | Homo sapiens apolipoprotein L, 4 (APOL4), transcript variant a, mRNA [NM_030643] |
| A_24_P941167 | 0,014520964 | down | -2,7115307 | APOL6 | Homo sapiens apolipoprotein L, 6 (APOL6), mRNA [NM_030641] |
| A_23_P155052 | 0,025400255 | up | 1,746567 | APOL6 | Homo sapiens apolipoprotein L, 6 (APOL6), mRNA [NM_030641] |
| A_33_P3246108 | 9,03E-04 | down | -2,2307947 | APOLD1 | Homo sapiens apolipoprotein L domain containing 1 (APOLD1), transcript variant 1, mRNA [NM_001130415] |
| A_23_P42265 | 0,023211027 | down | -1,5572853 | APOM | Homo sapiens apolipoprotein M (APOM), transcript variant 1, mRNA [NM_019101] |
| A_33_P3369565 | 0,003553233 | down | -1,5081879 | APTX | Homo sapiens aprataxin (APTX), transcript variant 11, mRNA [NM_001195252] |
| A_23_P126613 | 0,03618346 | down | -2,076147 | AQP10 | Homo sapiens aquaporin 10 (AQP10), mRNA [NM_080429] |
| A_33_P3217786 | 0,008911069 | down | -1,9877217 | AQP7P1 | Homo sapiens aquaporin 7 pseudogene 1 (AQP7P1), non-coding RNA [NR_002817] |
| A_21_P0013643 | 0,016923526 | up | 2,3540564 | AQP7P3 | Homo sapiens aquaporin 7 pseudogene 3 (AQP7P3), non-coding RNA [NR_026558] |
| A_24_P316305 | 0,00225965 | up | 1,5464362 | AQR | aquarius intron-binding spliceosomal factor [Source:HGNC Symbol;Acc:HGNC:29513] [ENST00000156471] |
| A_24_P280378 | 2,23E-05 | up | 1,9253452 | AQR | Homo sapiens aquarius intron-binding spliceosomal factor (AQR), mRNA [NM_014691] |
| A_33_P3224020 | 0,03939753 | down | -1,6893276 | ARAFP2 | Homo sapiens v-raf murine sarcoma 3611 viral oncogene homolog pseudogene, mRNA (cDNA clone IMAGE:5295529). [BC033982] |
| A_33_P3416583 | 0,035444465 | down | -1,6926707 | ARAP1 | Homo sapiens ArfGAP with RhoGAP domain, ankyrin repeat and PH domain 1 (ARAP1), transcript variant 4, mRNA [NM_001135190] |
| A_32_P83784 | 0,014772935 | up | 1,7045891 | ARAP2 | Homo sapiens ArfGAP with RhoGAP domain, ankyrin repeat and PH domain 2 (ARAP2), mRNA [NM_015230] |
| A_23_P167389 | 0,005662123 | up | 1,5801344 | ARAP3 | Homo sapiens ArfGAP with RhoGAP domain, ankyrin repeat and PH domain 3 (ARAP3), mRNA [NM_022481] |
| A_33_P3356216 | 0,003940393 | down | -1,7887914 | ARCN1 | Homo sapiens archain 1 (ARCN1), transcript variant 1, mRNA [NM_001655] |
| A_33_P3419190 | 0,013995755 | down | -1,6401076 | AREG | Homo sapiens amphiregulin (AREG), mRNA [NM_001657] |
| A_23_P259071 | 0,001904874 | down | -1,9908347 | AREG | Homo sapiens amphiregulin (AREG), mRNA [NM_001657] |
| A_33_P3271925 | 6,00E-05 | up | 1,7544965 | ARFGEF2 | Homo sapiens ADP-ribosylation factor guanine nucleotide-exchange factor 2 (brefeldin A-inhibited) (ARFGEF2), mRNA [NM_006420] |
| A_33_P3209706 | 0,005001717 | down | -1,6233346 | ARGFX | Homo sapiens arginine-fifty homeobox (ARGFX), mRNA [NM_001012659] |
| A_33_P3349289 | 0,005745707 | up | 2,2340586 | ARGFXP2 | Homo sapiens arginine-fifty homeobox pseudogene 2 (ARGFXP2), non-coding RNA [NR_002222] |
| A_24_P401739 | 4,67E-05 | up | 2,1889362 | ARHGAP17 | Homo sapiens Rho GTPase activating protein 17 (ARHGAP17), transcript variant 1, mRNA [NM_001006634] |
| A_33_P3411025 | 0,0017887 | down | -1,8095853 | ARHGAP19 | Homo sapiens Rho GTPase activating protein 19 (ARHGAP19), transcript variant 2, mRNA [NM_001204300] |
| A_33_P3319401 | 0,001254876 | up | 1,9754615 | ARHGAP21 | Rho GTPase activating protein 21 [Source:HGNC Symbol;Acc:HGNC:23725] [ENST00000476499] |
| A_23_P115608 | 0,012737622 | up | 1,522334 | ARHGAP21 | Homo sapiens Rho GTPase activating protein 21 (ARHGAP21), mRNA [NM_020824] |
| A_33_P3236030 | 0,006813917 | up | 1,9398439 | ARHGAP24 | Rho GTPase activating protein 24 [Source:HGNC Symbol;Acc:HGNC:25361] [ENST00000509709] |
| A_33_P3228305 | 0,03848363 | down | -1,9320827 | ARHGAP26 | Homo sapiens Rho GTPase activating protein 26 (ARHGAP26), transcript variant 1, mRNA [NM_015071] |
| A_33_P3228558 | 0,008444564 | up | 1,7969031 | ARHGAP27 | Homo sapiens Rho GTPase activating protein 27 (ARHGAP27), transcript variant 4, mRNA [NM_001282290] |
| A_23_P35045 | 0,043800388 | up | 2,0699978 | ARHGAP30 | Homo sapiens Rho GTPase activating protein 30 (ARHGAP30), transcript variant 1, mRNA [NM_001025598] |
| A_24_P349039 | 0,031615544 | up | 1,5194767 | ARHGAP31 | Homo sapiens Rho GTPase activating protein 31 (ARHGAP31), mRNA [NM_020754] |
| A_23_P161686 | 0,023598269 | up | 1,6576315 | ARHGAP32 | Homo sapiens Rho GTPase activating protein 32 (ARHGAP32), transcript variant 2, mRNA [NM_014715] |
| A_33_P3313411 | 0,001277117 | down | -2,8624554 | ARHGAP33 | Homo sapiens Rho GTPase activating protein 33 (ARHGAP33), transcript variant 2, mRNA [NM_001172630] |
| A_23_P72968 | 0,040057488 | up | 1,5615122 | ARHGAP36 | Homo sapiens Rho GTPase activating protein 36 (ARHGAP36), transcript variant 1, mRNA [NM_144967] |
| A_33_P3299110 | 1,39E-05 | down | -4,104027 | ARHGAP42 | Homo sapiens Rho GTPase activating protein 42 (ARHGAP42), mRNA [NM_152432] |
| A_33_P3243332 | 0,046721507 | down | -1,5194172 | ARHGAP42 | Homo sapiens Rho GTPase activating protein 42 (ARHGAP42), mRNA [NM_152432] |
| A_33_P3410385 | 1,96E-05 | down | -5,0895834 | ARHGAP44 | Homo sapiens Rho GTPase activating protein 44 (ARHGAP44), mRNA [NM_014859] |
| A_24_P41864 | 0,002207561 | up | 3,7018497 | ARHGAP6 | Homo sapiens Rho GTPase activating protein 6 (ARHGAP6), transcript variant 3, mRNA [NM_006125] |
| A_24_P64661 | 0,004935765 | down | -2,982005 | ARHGAP9 | Homo sapiens Rho GTPase activating protein 9 (ARHGAP9), transcript variant 1, mRNA [NM_032496] |
| A_24_P202581 | 0,008445428 | up | 1,6829821 | ARHGEF1 | Homo sapiens Rho guanine nucleotide exchange factor (GEF) 1 (ARHGEF1), transcript variant 1, mRNA [NM_199002] |
| A_33_P3351356 | 0,023686832 | up | 1,5129212 | ARHGEF10 | Rho guanine nucleotide exchange factor (GEF) 10 [Source:HGNC Symbol;Acc:HGNC:14103] [ENST00000398560] |
| A_33_P3799936 | 0,033584837 | up | 1,5934123 | ARHGEF10L | Homo sapiens Rho guanine nucleotide exchange factor (GEF) 10-like (ARHGEF10L), transcript variant 1, mRNA [NM_018125] |
| A_33_P3215575 | 0,03050873 | up | 1,6481 | ARHGEF10L | Homo sapiens Rho guanine nucleotide exchange factor (GEF) 10-like (ARHGEF10L), transcript variant 1, mRNA [NM_018125] |
| A_23_P49674 | 0,010564775 | down | -2,28264 | ARHGEF15 | Homo sapiens Rho guanine nucleotide exchange factor (GEF) 15 (ARHGEF15), transcript variant 1, mRNA [NM_173728] |
| A_23_P50357 | 0,012724575 | up | 1,5723478 | ARHGEF18 | Homo sapiens Rho/Rac guanine nucleotide exchange factor (GEF) 18 (ARHGEF18), transcript variant 1, mRNA [NM_015318] |
| A_23_P69100 | 2,16E-04 | up | 2,165012 | ARHGEF26 | Homo sapiens Rho guanine nucleotide exchange factor (GEF) 26 (ARHGEF26), transcript variant 2, mRNA [NM_015595] |
| A_33_P3405680 | 0,01847384 | down | -2,929358 | ARHGEF3-AS1 | Homo sapiens ARHGEF3 antisense RNA 1 (ARHGEF3-AS1), long non-coding RNA [NR_046572] |
| A_33_P3233099 | 0,006118679 | down | -1,6284099 | ARHGEF4 | Homo sapiens Rho guanine nucleotide exchange factor (GEF) 4 (ARHGEF4), transcript variant 1, mRNA [NM_015320] |
| A_23_P417891 | 0,033574887 | up | 1,7245998 | ARHGEF7 | Homo sapiens Rho guanine nucleotide exchange factor (GEF) 7 (ARHGEF7), transcript variant 2, mRNA [NM_145735] |

| | | | | | |
|---|---|---|---|---|---|
| A_23_P321855 | 2,72E-04 | up | 1,687822 | ARHGEF7 | Homo sapiens Rho guanine nucleotide exchange factor (GEF) 7 (ARHGEF7), transcript variant 1, mRNA [NM_003899] |
| A_33_P3288904 | 0,006244665 | up | 1,5550225 | ARHGEF7 | Homo sapiens Rho guanine nucleotide exchange factor (GEF) 7 (ARHGEF7), transcript variant 1, mRNA [NM_003899] |
| A_24_P92952 | 4,22E-04 | up | 1,7907203 | ARID1A | Homo sapiens AT rich interactive domain 1A (SWI-like) (ARID1A), transcript variant 1, mRNA [NM_006015] |
| A_33_P3291877 | 0,003798425 | down | -1,530412 | ARID1B | Homo sapiens AT rich interactive domain 1B (SWI1-like) (ARID1B), transcript variant 2, mRNA [NM_020732] |
| A_33_P3272921 | 0,003682695 | up | 1,9864254 | ARID3A | Homo sapiens AT rich interactive domain 3A (BRIGHT-like) (ARID3A), mRNA [NM_005224] |
| A_33_P3389342 | 5,62E-04 | down | -1,7345865 | ARID5A | Homo sapiens AT rich interactive domain 5A (MRF1-like) (ARID5A), mRNA [NM_212481] |
| A_24_P85478 | 0,001448648 | up | 1,5707865 | ARIH1 | Homo sapiens ariadne RBR E3 ubiquitin protein ligase 1 (ARIH1), mRNA [NM_005744] |
| A_33_P3375553 | 0,013572644 | up | 1,6597234 | ARL13A | Homo sapiens ADP-ribosylation factor-like 13A (ARL13A), transcript variant 3, mRNA [NM_001162491] |
| A_24_P303199 | 0,003442107 | down | -2,497777 | ARL15 | Homo sapiens ADP-ribosylation factor-like 15 (ARL15), mRNA [NM_019087] |
| A_23_P118150 | 4,26E-04 | up | 1,7309519 | ARL6IP1 | Homo sapiens ADP-ribosylation factor-like 6 interacting protein 1 (ARL6IP1), mRNA [NM_015161] |
| A_32_P146113 | 0,015494243 | up | 2,5578406 | ARMC12 | Homo sapiens armadillo repeat containing 12 (ARMC12), transcript variant 1, mRNA [NM_145028] |
| A_23_P86540 | 0,007605813 | up | 1,9427668 | ARMC3 | Homo sapiens armadillo repeat containing 3 (ARMC3), transcript variant 1, mRNA [NM_173081] |
| A_33_P3364826 | 0,04771689 | up | 2,087339 | ARMC4 | armadillo repeat containing 4 [Source:HGNC Symbol;Acc:HGNC:25583] [ENST00000434029] |
| A_23_P15375 | 0,002954034 | down | -1,5836288 | ARMC7 | armadillo repeat containing 7 [Source:HGNC Symbol;Acc:HGNC:26168] [ENST00000245543] |
| A_33_P3425356 | 4,75E-04 | up | 1,5505869 | ARMC8 | Homo sapiens armadillo repeat containing 8 (ARMC8), transcript variant 2, mRNA [NM_015396] |
| A_33_P3232334 | 0,00419129 | down | -3,2951999 | ARMCX4 | armadillo repeat containing, X-linked 4 [Source:HGNC Symbol;Acc:HGNC:28615] [ENST00000430461] |
| A_23_P162037 | 0,012158245 | up | 1,5645332 | ARNTL | Homo sapiens aryl hydrocarbon receptor nuclear translocator-like (ARNTL), transcript variant 3, mRNA [NM_001030273] |
| A_24_P37540 | 0,028799871 | up | 1,5324749 | ARPC4-TTLL3 | Homo sapiens ARPC4-TTLL3 readthrough (ARPC4-TTLL3), mRNA [NM_001198793] |
| A_24_P313262 | 8,15E-04 | up | 1,8738422 | ARPP19 | Homo sapiens cAMP-regulated phosphoprotein, 19kDa (ARPP19), mRNA [NM_006628] |
| A_24_P344976 | 0,00827331 | down | -2,1638367 | ARR3 | Homo sapiens arrestin 3, retinal (X-arrestin) (ARR3), mRNA [NM_004312] |
| A_23_P391607 | 1,54E-04 | up | 1,6721184 | ARRDC1 | Homo sapiens arrestin domain containing 1 (ARRDC1), mRNA [NM_152285] |
| A_24_P22976 | 1,10E-05 | up | 2,403279 | ARRDC2 | Homo sapiens arrestin domain containing 2 (ARRDC2), transcript variant 2, mRNA [NM_001025604] |
| A_24_P339416 | 0,001750056 | down | -2,5286026 | ARSG | Homo sapiens arylsulfatase G (ARSG), transcript variant 1, mRNA [NM_014960] |
| A_33_P3298349 | 0,011733079 | down | -2,3836076 | ART5 | Homo sapiens ADP-ribosyltransferase 5 (ART5), transcript variant 3, mRNA [NM_001297668] |
| A_24_P416961 | 0,025857667 | down | -2,5112412 | ARVCF | Homo sapiens armadillo repeat gene deleted in velocardiofacial syndrome (ARVCF), mRNA [NM_001670] |
| A_23_P306223 | 0,007069324 | down | -2,377259 | ARX | Homo sapiens aristaless related homeobox (ARX), mRNA [NM_139058] |
| A_23_P146325 | 0,002491754 | up | 1,8358874 | ASAP1-IT1 | Homo sapiens ASAP1 intronic transcript 1 (non-protein coding) (ASAP1-IT1), long non-coding RNA [NR_002765] |
| A_24_P362540 | 0,003638036 | up | 2,1051774 | ASAP2 | Homo sapiens ArfGAP with SH3 domain, ankyrin repeat and PH domain 2 (ASAP2), transcript variant 1, mRNA [NM_003887] |
| A_33_P3263614 | 2,93E-04 | down | -2,066154 | ASAP3 | Homo sapiens ArfGAP with SH3 domain, ankyrin repeat and PH domain 3 (ASAP3), transcript variant 2, mRNA [NM_001143778] |
| A_23_P114689 | 0,028896725 | up | 1,522755 | ASAP3 | Homo sapiens ArfGAP with SH3 domain, ankyrin repeat and PH domain 3 (ASAP3), transcript variant 1, mRNA [NM_017707] |
| A_33_P3290124 | 0,001264147 | down | -1,989607 | ASB10 | Homo sapiens ankyrin repeat and SOCS box containing 10 (ASB10), transcript variant 1, mRNA [NM_001142459] |
| A_33_P3210622 | 3,12E-06 | down | -3,8629391 | ASB13 | Homo sapiens ankyrin repeat and SOCS box containing 13 (ASB13), transcript variant 1, mRNA [NM_024701] |
| A_33_P3290139 | 0,009687145 | up | 2,646106 | ASB15 | Homo sapiens ankyrin repeat and SOCS box containing 15 (ASB15), transcript variant 2, mRNA [NM_001290258] |
| A_33_P3290162 | 1,71E-06 | down | -4,200646 | ASB18 | Homo sapiens ankyrin repeat and SOCS box containing 18 (ASB18), mRNA [NM_212556] |
| A_23_P501319 | 0,001133729 | up | 3,2339606 | ASB4 | Homo sapiens ankyrin repeat and SOCS box containing 4 (ASB4), transcript variant 1, mRNA [NM_016116] |
| A_24_P920979 | 1,43E-04 | up | 1,8665576 | ASB7 | Homo sapiens ankyrin repeat and SOCS box containing 7 (ASB7), transcript variant 2, mRNA [NM_198243] |
| A_32_P122240 | 0,005807095 | down | -2,551313 | ASCL5 | Homo sapiens achaete-scute family bHLH transcription factor 5 (ASCL5), mRNA [NM_001270601] |
| A_33_P3250730 | 5,04E-05 | down | -2,2522976 | ASIC4 | Homo sapiens acid sensing (proton gated) ion channel family member 4 (ASIC4), mRNA [NM_182847] |
| A_24_P690811 | 0,044123385 | down | -2,2649004 | ASPDH | Homo sapiens aspartate dehydrogenase domain containing (ASPDH), transcript variant 2, mRNA [NM_001024656] |
| A_23_P91640 | 0,004178115 | up | 1,6600933 | ASPHD2 | Homo sapiens aspartate beta-hydroxylase domain containing 2 (ASPHD2), mRNA [NM_020437] |
| A_33_P3264394 | 0,04293084 | down | -1,8767622 | ASPSCR1 | alveolar soft part sarcoma chromosome region, candidate 1 [Source:HGNC Symbol;Acc:HGNC:13825] [ENST00000583693] |
| A_33_P3231820 | 0,018145066 | down | -1,7157686 | ASTL | Homo sapiens astacin-like metallo-endopeptidase (M12 family) (ASTL), mRNA [NM_001002036] |
| A_23_P301021 | 8,89E-04 | up | 2,6595945 | ASTN1 | Homo sapiens astrotactin 1 (ASTN1), transcript variant 1, mRNA [NM_004319] |
| A_24_P83738 | 0,019653236 | down | -1,7760829 | ASTN2 | Homo sapiens astrotactin 2 (ASTN2), transcript variant 4, mRNA [NM_198188] |
| A_23_P36464 | 0,001936744 | up | 1,6653413 | ASUN | Homo sapiens asunder spermatogenesis regulator (ASUN), mRNA [NM_018164] |
| A_33_P3217238 | 0,027898796 | up | 1,5011556 | ATAD2 | Homo sapiens ATPase family, AAA domain containing 2 (ATAD2), mRNA [NM_014109] |
| A_32_P191004 | 2,70E-04 | up | 1,6246657 | ATAD2B | Homo sapiens ATPase family, AAA domain containing 2B (ATAD2B), transcript variant 1, mRNA [NM_017552] |
| A_24_P113144 | 0,022431966 | up | 2,133683 | ATAD5 | Homo sapiens ATPase family, AAA domain containing 5 (ATAD5), mRNA [NM_024857] |
| A_33_P3305458 | 0,015403907 | up | 1,5969318 | ATF6B | Homo sapiens activating transcription factor 6 beta (ATF6B), transcript variant 1, mRNA [NM_004381] |
| A_19_P00809417 | 0,045554437 | down | -1,6165559 | ATF7IP2 | activating transcription factor 7 interacting protein 2 [Source:HGNC Symbol;Acc:HGNC:20397] [ENST00000570163] |
| A_23_P36305 | 0,002848527 | up | 1,581645 | ATG16L2 | Homo sapiens autophagy related 16-like 2 (S. cerevisiae) (ATG16L2), mRNA [NM_033388] |
| A_33_P3342160 | 2,55E-04 | up | 1,8004694 | ATG2B | Homo sapiens autophagy related 2B (ATG2B), mRNA [NM_018036] |
| A_33_P3396807 | 0,004599845 | down | -1,6846886 | ATG4C | Homo sapiens autophagy related 4C, cysteine peptidase (ATG4C), transcript variant 1, mRNA [NM_032852] |
| A_32_P154053 | 0,003425434 | up | 3,0233526 | ATG9B | Homo sapiens autophagy related 9B (ATG9B), transcript variant 1, mRNA [NM_173681] |
| A_33_P3357591 | 0,014687812 | up | 1,7591987 | ATHL1 | Homo sapiens ATH1, acid trehalase-like 1 (yeast) (ATHL1), mRNA [NM_025092] |
| A_23_P13885 | 0,015656002 | up | 1,5485855 | ATN1 | Homo sapiens atrophin 1 (ATN1), transcript variant 1, mRNA [NM_001007026] |
| A_33_P3392740 | 0,04319735 | down | -1,9271152 | ATOH8 | Homo sapiens mRNA; cDNA DKFZp761E2117 (from clone DKFZp761E2117). [AL831857] |
| A_24_P161973 | 0,001358852 | up | 1,6857262 | ATP11A | Homo sapiens ATPase, class VI, type 11A (ATP11A), transcript variant 1, mRNA [NM_015205] |
| A_33_P3269423 | 0,014485754 | down | -2,0106888 | ATP11AUN | Homo sapiens ATP11A upstream neighbor (ATP11AUN), mRNA [NM_207440] |
| A_24_P399942 | 0,001956185 | up | 1,8498672 | ATP11C | Homo sapiens ATPase, class VI, type 11C (ATP11C), transcript variant 1, mRNA [NM_173694] |
| A_23_P1072 | 0,016523637 | up | 1,6457088 | ATP1A1 | Homo sapiens ATPase, Na+/K+ transporting, alpha 1 polypeptide (ATP1A1), transcript variant 1, mRNA [NM_000701] |

| | | | | | |
|---|---|---|---|---|---|
| A_33_P3283669 | 9,24E-05 | down | -3,0959156 | ATP1A3 | Homo sapiens ATPase, Na+/K+ transporting, alpha 3 polypeptide (ATP1A3), transcript variant 3, mRNA [NM_001256214] |
| A_24_P91832 | 0,007252164 | up | 1,5988394 | ATP1B4 | Homo sapiens ATPase, Na+/K+ transporting, beta 4 polypeptide (ATP1B4), transcript variant 2, mRNA [NM_012069] |
| A_33_P3342830 | 5,22E-04 | down | -2,1896355 | ATP2A1 | Homo sapiens ATPase, Ca++ transporting, cardiac muscle, fast twitch 1 (ATP2A1), transcript variant b, mRNA [NM_173201] |
| A_24_P141786 | 0,029243087 | down | -1,5651772 | ATP2A2 | Homo sapiens ATPase, Ca++ transporting, cardiac muscle, slow twitch 2 (ATP2A2), transcript variant a, mRNA [NM_001681] |
| A_24_P202319 | 0,010893703 | down | -1,6152818 | ATP2A3 | Homo sapiens ATPase, Ca++ transporting, ubiquitous (ATP2A3), transcript variant 7, mRNA [NM_174958] |
| A_23_P207632 | 0,004446442 | up | 1,5122726 | ATP2A3 | Homo sapiens ATPase, Ca++ transporting, ubiquitous (ATP2A3), transcript variant 5, mRNA [NM_174953] |
| A_33_P3267185 | 0,008981112 | down | -1,8199997 | ATP2B1 | Homo sapiens ATPase, Ca++ transporting, plasma membrane 1 (ATP2B1), transcript variant 1, mRNA [NM_001001323] |
| A_33_P3393851 | 4,88E-05 | down | -5,0949497 | ATP2B2 | ATPase, Ca++ transporting, plasma membrane 2 [Source:HGNC Symbol;Acc:HGNC:815] [ENST00000467702] |
| A_33_P3400973 | 1,19E-05 | down | -4,057551 | ATP2B2 | Homo sapiens ATPase, Ca++ transporting, plasma membrane 2 (ATP2B2), transcript variant 1, mRNA [NM_001001331] |
| A_24_P405205 | 6,25E-04 | up | 2,2330663 | ATP2B4 | Homo sapiens ATPase, Ca++ transporting, plasma membrane 4 (ATP2B4), transcript variant 1, mRNA [NM_001001396] |
| A_19_P00317360 | 0,03898048 | down | -3,1237462 | ATP2C1 | Homo sapiens ATPase, Ca++ transporting, type 2C, member 1 (ATP2C1), transcript variant 8, mRNA [NM_001199182] |
| A_23_P146146 | 0,006398266 | down | -1,8038149 | ATP6V0D2 | Homo sapiens ATPase, H+ transporting, lysosomal 38kDa, V0 subunit d2 (ATP6V0D2), mRNA [NM_152565] |
| A_33_P3210885 | 0,029625334 | down | -1,5526893 | ATP7A | Homo sapiens ATPase, Cu++ transporting, alpha polypeptide (ATP7A), transcript variant 2, mRNA [NM_001282224] |
| A_33_P3327772 | 0,001335359 | down | -2,3949463 | ATP7A | Homo sapiens ATPase, Cu++ transporting, alpha polypeptide (ATP7A), transcript variant 1, mRNA [NM_000052] |
| A_23_P79108 | 0,006407671 | up | 2,5874522 | ATP8B3 | Homo sapiens ATPase, aminophospholipid transporter, class I, type 8B, member 3 (ATP8B3), transcript variant 1, mRNA [NM_138813] |
| A_33_P3362498 | 0,006385105 | down | -1,6338238 | ATR | Homo sapiens ATR serine/threonine kinase (ATR), mRNA [NM_001184] |
| A_24_P371425 | 1,60E-04 | down | -1,9645659 | ATRIP | Homo sapiens ATR interacting protein (ATRIP), transcript variant 2, mRNA [NM_032166] |
| A_33_P3343473 | 4,20E-05 | down | -3,3012915 | ATRNL1 | Homo sapiens attractin-like 1 (ATRNL1), transcript variant 1, mRNA [NM_207303] |
| A_33_P3612589 | 0,022663072 | up | 1,5475882 | ATRX | Homo sapiens alpha thalassemia/mental retardation syndrome X-linked (ATRX), transcript variant 1, mRNA [NM_000489] |
| A_33_P3409099 | 1,20E-04 | up | 2,1457882 | ATXN1 | Homo sapiens ataxin 1 (ATXN1), transcript variant 1, mRNA [NM_000332] |
| A_33_P3256500 | 0,002576867 | up | 1,8549869 | ATXN2 | ataxin 2 [Source:HGNC Symbol;Acc:HGNC:10555] [ENST00000392645] |
| A_23_P389907 | 1,56E-04 | up | 1,8955917 | ATXN2 | Homo sapiens ataxin 2 (ATXN2), mRNA [NM_002973] |
| A_23_P317207 | 0,040375687 | down | -1,7810861 | ATXN7L2 | Homo sapiens ataxin 7-like 2 (ATXN7L2), mRNA [NM_153340] |
| A_24_P263956 | 6,70E-05 | up | 2,0723948 | ATXN7L3B | Homo sapiens ataxin 7-like 3B (ATXN7L3B), mRNA [NM_001136262] |
| A_33_P3869455 | 0,003416931 | down | -2,258131 | ATXN8 | Homo sapiens ataxin 8 (ATXN8) mRNA, partial cds. [DQ641254] |
| A_33_P3330211 | 0,033744603 | down | -1,6613653 | AUTS2 | Homo sapiens autism susceptibility candidate 2 (AUTS2), transcript variant 3, mRNA [NM_001127232] |
| A_23_P72340 | 0,035358205 | down | -1,5781926 | AVPR1B | Homo sapiens arginine vasopressin receptor 1B (AVPR1B), mRNA [NM_000707] |
| A_21_P0013915 | 0,003318351 | down | -2,197826 | AZFP | Homo sapiens AML-associated zinc finger protein (AZFP) mRNA, partial cds. [AY371499] |
| A_21_P0014818 | 0,004663016 | up | 1,8332663 | AZFP | Homo sapiens AML-associated zinc finger protein (AZFP) mRNA, partial cds. [AY371499] |
| A_24_P267664 | 0,005560591 | down | -1,7212967 | B3GALT5-AS1 | Homo sapiens B3GALT5 antisense RNA 1 (B3GALT5-AS1), transcript variant 2, long non-coding RNA [NR_026543] |
| A_33_P3266536 | 9,14E-04 | up | 2,9250572 | B3GALT5-AS1 | Homo sapiens B3GALT5 antisense RNA 1 (B3GALT5-AS1), transcript variant 2, long non-coding RNA [NR_026543] |
| A_23_P500956 | 1,71E-04 | up | 1,937736 | B3GNT2 | Homo sapiens UDP-GlcNAc:betaGal beta-1,3-N-acetylglucosaminyltransferase 2 (B3GNT2), mRNA [NM_006577] |
| A_23_P18372 | 3,91E-04 | up | 1,830105 | B3GNT5 | Homo sapiens UDP-GlcNAc:betaGal beta-1,3-N-acetylglucosaminyltransferase 5 (B3GNT5), mRNA [NM_032047] |
| A_23_P39755 | 8,00E-06 | down | -2,4359066 | B3GNT7 | Homo sapiens UDP-GlcNAc:betaGal beta-1,3-N-acetylglucosaminyltransferase 7 (B3GNT7), mRNA [NM_145236] |
| A_23_P331211 | 0,009388549 | down | -3,4637957 | B4GALNT2 | Homo sapiens beta-1,4-N-acetyl-galactosaminyl transferase 2 (B4GALNT2), transcript variant 1, mRNA [NM_153446] |
| A_23_P152678 | 0,012109761 | down | -1,7475814 | B9D1 | Homo sapiens B9 protein domain 1 (B9D1), transcript variant 2, mRNA [NM_015681] |
| A_24_P176714 | 0,001396947 | down | -1,8995298 | B9D1 | Homo sapiens B9 protein domain 1 (B9D1), transcript variant 2, mRNA [NM_015681] |
| A_33_P3282973 | 0,022270218 | down | -1,616956 | BAALC | brain and acute leukemia, cytoplasmic [Source:HGNC Symbol;Acc:HGNC:14333] [ENST00000297574] |
| A_33_P3460043 | 0,008309009 | down | -1,8965774 | BAALC-AS2 | Homo sapiens BAALC antisense RNA 2 (BAALC-AS2), long non-coding RNA [NR_027071] |
| A_23_P211047 | 0,006471936 | up | 1,5013947 | BACH1 | Homo sapiens BTB and CNC homology 1, basic leucine zipper transcription factor 1 (BACH1), transcript variant 1, mRNA [NM_206866] |
| A_33_P3277714 | 1,30E-04 | up | 2,7034454 | BACH2 | PREDICTED: Homo sapiens BTB and CNC homology 1, basic leucine zipper transcription factor 2 (BACH2), transcript variant X3, mRNA [XM_006715545] |
| A_23_P30634 | 0,012915991 | up | 1,8272383 | BACH2 | Homo sapiens BTB and CNC homology 1, basic leucine zipper transcription factor 2 (BACH2), transcript variant 1, mRNA [NM_021813] |
| A_24_P126262 | 0,002597489 | down | -1,9638956 | BAGE4 | Homo sapiens B melanoma antigen family, member 4 (BAGE4), mRNA [NM_181704] |
| A_23_P49539 | 3,87E-04 | up | 1,9214773 | BAHCC1 | Homo sapiens BAH domain and coiled-coil containing 1 (BAHCC1), mRNA [NM_001291324] |
| A_23_P61810 | 0,006985195 | up | 1,5212247 | BAIAP2 | Homo sapiens BAI1-associated protein 2 (BAIAP2), transcript variant 1, mRNA [NM_017450] |
| A_23_P315836 | 0,020893224 | up | 1,8045979 | BAIAP2 | Homo sapiens BAI1-associated protein 2 (BAIAP2), transcript variant 2, mRNA [NM_017451] |
| A_33_P3262575 | 0,018705232 | up | 1,5705003 | BAIAP2L1 | Homo sapiens BAI1-associated protein 2-like 1 (BAIAP2L1), mRNA [NM_018842] |
| A_23_P10232 | 0,045140605 | down | -1,588413 | BANK1 | Homo sapiens B-cell scaffold protein with ankyrin repeats 1 (BANK1), transcript variant 1, mRNA [NM_017935] |
| A_33_P3341144 | 0,001898305 | up | 1,6344742 | BAX | Homo sapiens BCL2-associated X protein (BAX), transcript variant beta, mRNA [NM_004324] |
| A_23_P215449 | 2,30E-04 | up | 1,7666466 | BAZ1B | Homo sapiens bromodomain adjacent to zinc finger domain, 1B (BAZ1B), mRNA [NM_032408] |
| A_33_P3321577 | 3,60E-04 | up | 1,5970042 | BAZ1B | Homo sapiens bromodomain adjacent to zinc finger domain, 1B (BAZ1B), mRNA [NM_032408] |
| A_23_P203841 | 4,26E-05 | up | 2,1134968 | BAZ2A | Homo sapiens bromodomain adjacent to zinc finger domain, 2A (BAZ2A), transcript variant 1, mRNA [NM_013449] |
| A_23_P120048 | 3,92E-05 | up | 1,9111196 | BAZ2B | Homo sapiens bromodomain adjacent to zinc finger domain, 2B (BAZ2B), transcript variant 1, mRNA [NM_013450] |
| A_23_P5785 | 0,034858283 | down | -1,5196817 | BBS5 | Homo sapiens Bardet-Biedl syndrome 5 (BBS5), mRNA [NM_152384] |
| A_33_P3360087 | 0,01705025 | down | -2,2011354 | BBS9 | Homo sapiens Bardet-Biedl syndrome 9 (BBS9), transcript variant 1, mRNA [NM_014451] |
| A_33_P3369034 | 0,005917535 | up | 1,5353501 | BCAR1 | Homo sapiens breast cancer anti-estrogen resistance 1 (BCAR1), transcript variant 1, mRNA [NM_001170714] |
| A_33_P3395738 | 0,017151585 | up | 1,8121217 | BCAR1 | Homo sapiens cDNA FLJ45059 fis, clone BRAWH3023274. [AK127004] |
| A_23_P97394 | 0,00139124 | up | 2,4571218 | BCAR3 | Homo sapiens breast cancer anti-estrogen resistance 3 (BCAR3), transcript variant 2, mRNA [NM_003567] |
| A_33_P3389852 | 7,88E-04 | down | -2,2945647 | BCAS3 | Homo sapiens breast carcinoma amplified sequence 3 (BCAS3), transcript variant 2, mRNA [NM_017679] |
| A_24_P1255 | 0,012659968 | up | 1,6957777 | BCCIP | Homo sapiens BRCA2 and CDKN1A interacting protein (BCCIP), transcript variant C, mRNA [NM_078469] |
| A_21_P0000540 | 0,007890617 | down | -1,6423104 | BCDIN3D-AS1 | Homo sapiens BCDIN3D antisense RNA 1 (BCDIN3D-AS1), transcript variant 3, long non-coding RNA [NR_027500] |

| | | | | | |
|---|---|---|---|---|---|
| A_23_P212050 | 7,41E-04 | down | -3,1648295 | BCHE | Homo sapiens butyrylcholinesterase (BCHE), mRNA [NM_000055] |
| A_33_P3236628 | 0,030094031 | up | 2,240844 | BCKDHA | branched chain keto acid dehydrogenase E1, alpha polypeptide [Source:HGNC Symbol;Acc:HGNC:986] [ENST00000595085] |
| A_33_P3355185 | 0,03931582 | down | -1,6604134 | BCL2 | Homo sapiens B-cell CLL/lymphoma 2 (BCL2), transcript variant beta, mRNA [NM_000657] |
| A_23_P152002 | 0,003998983 | up | 3,5884342 | BCL2A1 | Homo sapiens BCL2-related protein A1 (BCL2A1), transcript variant 1, mRNA [NM_004049] |
| A_23_P321703 | 0,029859778 | up | 1,5435159 | BCL2A1 | Homo sapiens BCL2-related protein A1 (BCL2A1), transcript variant 1, mRNA [NM_004049] |
| A_23_P210886 | 0,008138884 | up | 1,8574662 | BCL2L1 | Homo sapiens BCL2-like 1 (BCL2L1), transcript variant 1, mRNA [NM_138578] |
| A_24_P122921 | 0,04997349 | up | 1,5144498 | BCL2L11 | Homo sapiens BCL2-like 11 (apoptosis facilitator) (BCL2L11), transcript variant 1, mRNA [NM_138621] |
| A_33_P3373775 | 7,53E-04 | down | -2,0600271 | BCL2L15 | Homo sapiens BCL2-like 15 (BCL2L15), mRNA [NM_001010922] |
| A_33_P3408177 | 0,009076105 | down | -2,2645876 | BCL2L15 | Homo sapiens BCL2-like 15 (BCL2L15), mRNA [NM_001010922] |
| A_23_P57856 | 0,001324288 | up | 1,8052293 | BCL6 | Homo sapiens B-cell CLL/lymphoma 6 (BCL6), transcript variant 2, mRNA [NM_001130845] |
| A_23_P324813 | 0,031495452 | up | 2,3570411 | BCL6B | Homo sapiens B-cell CLL/lymphoma 6, member B (BCL6B), mRNA [NM_181844] |
| A_23_P382602 | 0,004127504 | down | -1,8549685 | BCL9 | Homo sapiens B-cell CLL/lymphoma 9 (BCL9), mRNA [NM_004326] |
| A_24_P134816 | 0,002411928 | up | 1,8462732 | BCL9L | Homo sapiens B-cell CLL/lymphoma 9-like (BCL9L), mRNA [NM_182557] |
| A_23_P124300 | 0,019574344 | down | -2,192181 | BCO1 | Homo sapiens beta-carotene oxygenase 1 (BCO1), mRNA [NM_017429] |
| A_33_P3261353 | 0,007539886 | down | -1,7592189 | BCORP1 | Homo sapiens BCL6 corepressor pseudogene 1 (BCORP1), transcript variant 1, non-coding RNA [NR_033732] |
| A_21_P0014929 | 0,001336881 | up | 2,708813 | BCR | Homo sapiens breakpoint cluster region (BCR), transcript variant 1, mRNA [NM_004327] |
| A_32_P56713 | 0,03094396 | up | 1,5761756 | BCR | Homo sapiens breakpoint cluster region (BCR), transcript variant 1, mRNA [NM_004327] |
| A_24_P23951 | 5,67E-04 | up | 1,8380742 | BCRP2 | Homo sapiens breakpoint cluster region pseudogene 2 (BCRP2), non-coding RNA [NR_037566] |
| A_24_P940776 | 6,57E-04 | up | 1,6054363 | BDP1 | Homo sapiens B double prime 1, subunit of RNA polymerase III transcription initiation factor IIIB (BDP1), mRNA [NM_018429] |
| A_21_P0000137 | 0,035239317 | down | -1,6254718 | BEAN1 | Homo sapiens brain expressed, associated with NEDD4, 1 (BEAN1), transcript variant 4, mRNA [NM_001197224] |
| A_33_P3418234 | 0,003316664 | down | -2,173397 | BECN1P1 | Homo sapiens beclin 1, autophagy related, pseudogene 1 (BECN1P1), mRNA [NM_001290693] |
| A_33_P3322539 | 0,013588826 | down | -2,0744846 | BEND2 | Homo sapiens BEN domain containing 2 (BEND2), transcript variant 2, mRNA [NM_001184767] |
| A_23_P904 | 0,014026511 | up | 1,8615069 | BEND5 | Homo sapiens BEN domain containing 5 (BEND5), transcript variant 1, mRNA [NM_024603] |
| A_23_P376188 | 0,047684956 | up | 1,5018283 | BEND7 | Homo sapiens BEN domain containing 7 (BEND7), transcript variant 2, mRNA [NM_001100912] |
| A_23_P16225 | 0,002999725 | down | -5,7396145 | BEST2 | Homo sapiens bestrophin 2 (BEST2), mRNA [NM_017682] |
| A_33_P3330384 | 0,002154321 | down | -2,3778203 | BET1L | Homo sapiens Bet1 golgi vesicular membrane trafficking protein-like (BET1L), transcript variant 2, mRNA [NM_016526] |
| A_33_P3245178 | 0,04616858 | down | -2,3291366 | BEX2 | Homo sapiens brain expressed X-linked 2 (BEX2), transcript variant 1, mRNA [NM_001168399] |
| A_24_P57898 | 0,009359345 | down | -2,2560744 | BHLHE23 | Homo sapiens basic helix-loop-helix family, member e23 (BHLHE23), mRNA [NM_080606] |
| A_24_P268676 | 4,18E-04 | up | 2,0268736 | BHLHE40 | Homo sapiens basic helix-loop-helix family, member e40 (BHLHE40), mRNA [NM_003670] |
| A_24_P231829 | 0,042339105 | up | 1,9852791 | BHMT2 | Homo sapiens betaine--homocysteine S-methyltransferase 2 (BHMT2), transcript variant 1, mRNA [NM_017614] |
| A_23_P98350 | 5,47E-05 | up | 4,6481695 | BIRC3 | Homo sapiens baculoviral IAP repeat containing 3 (BIRC3), transcript variant 1, mRNA [NM_001165] |
| A_23_P419624 | 0,005530147 | up | 1,5084311 | BLCAP | Homo sapiens bladder cancer associated protein (BLCAP), transcript variant 1, mRNA [NM_006698] |
| A_33_P3398548 | 0,001195801 | up | 1,7616756 | BLZF1 | Homo sapiens basic leucine zipper nuclear factor 1 (BLZF1), mRNA [NM_003666] |
| A_23_P314115 | 1,51E-04 | up | 1,7129883 | BMI1 | Homo sapiens BMI1 proto-oncogene, polycomb ring finger (BMI1), mRNA [NM_005180] |
| A_33_P3234043 | 0,04506046 | down | -1,6399586 | BMP1 | bone morphogenetic protein 1 [Source:HGNC Symbol;Acc:HGNC:1067] [ENST00000471755] |
| A_23_P131588 | 0,044946168 | down | -1,9618815 | BMP10 | Homo sapiens bone morphogenetic protein 10 (BMP10), mRNA [NM_014482] |
| A_33_P3237150 | 0,005409889 | up | 1,909463 | BMP2 | Homo sapiens bone morphogenetic protein 2 (BMP2), mRNA [NM_001200] |
| A_23_P81058 | 0,035789225 | up | 1,8841637 | BMP3 | Homo sapiens bone morphogenetic protein 3 (BMP3), mRNA [NM_001201] |
| A_23_P154643 | 0,016173737 | down | -2,2606778 | BMP7 | Homo sapiens bone morphogenetic protein 7 (BMP7), mRNA [NM_001719] |
| A_33_P3365878 | 0,001274611 | down | -1,6426723 | BMP8B | Homo sapiens bone morphogenetic protein 8b (BMP8B), mRNA [NM_001720] |
| A_21_P0011284 | 0,009933102 | down | -1,762271 | BMS1P20 | Homo sapiens BMS1 pseudogene 20 (BMS1P20), non-coding RNA [NR_027293] |
| A_23_P429689 | 2,67E-04 | up | 1,8814974 | BOD1L1 | Homo sapiens biorientation of chromosomes in cell division 1-like 1 (BOD1L1), mRNA [NM_148894] |
| A_33_P3407195 | 2,04E-04 | up | 1,7545211 | BOD1L1 | Homo sapiens biorientation of chromosomes in cell division 1-like 1 (BOD1L1), mRNA [NM_148894] |
| A_33_P3368301 | 0,012896141 | down | -1,6117138 | BOLA3 | Homo sapiens bolA family member 3 (BOLA3), transcript variant 1, mRNA [NM_212552] |
| A_21_P0002242 | 0,018850703 | down | -1,995305 | BOLA3-AS1 | Homo sapiens BOLA3 antisense RNA 1 (head to head) (BOLA3-AS1), transcript variant 3, long non-coding RNA [NR_045637] |
| A_23_P254797 | 2,15E-04 | down | -3,7931802 | BPIFB2 | Homo sapiens BPI fold containing family B, member 2 (BPIFB2), mRNA [NM_025227] |
| A_24_P122636 | 0,019227099 | down | -1,839708 | BPNT1 | Homo sapiens 3'(2'), 5'-bisphosphate nucleotidase 1 (BPNT1), transcript variant 1, mRNA [NM_006085] |
| A_33_P3269453 | 8,88E-05 | up | 1,922633 | BPTF | PREDICTED: Homo sapiens bromodomain PHD finger transcription factor (BPTF), transcript variant X14, mRNA [XM_006721770] |
| A_33_P3240674 | 9,58E-04 | down | -1,5187112 | BRD3 | Homo sapiens bromodomain containing 3 (BRD3), mRNA [NM_007371] |
| A_23_P216689 | 7,17E-04 | up | 1,7414857 | BRD3 | Homo sapiens bromodomain containing 3 (BRD3), mRNA [NM_007371] |
| A_23_P425104 | 0,003342068 | up | 1,5089451 | BRD4 | Homo sapiens bromodomain containing 4 (BRD4), transcript variant long, mRNA [NM_058243] |
| A_23_P136589 | 0,006057689 | up | 1,6135938 | BRDT | Homo sapiens bromodomain, testis-specific (BRDT), transcript variant 1, mRNA [NM_207189] |
| A_33_P3363016 | 0,008024241 | down | -1,8357172 | BRF1 | Homo sapiens BRF1, RNA polymerase III transcription initiation factor 90 kDa subunit (BRF1), transcript variant 8, mRNA [NM_001242790] |
| A_23_P250948 | 9,26E-04 | up | 1,7824018 | BRPF1 | Homo sapiens bromodomain and PHD finger containing, 1 (BRPF1), transcript variant 1, mRNA [NM_001003694] |
| A_24_P414712 | 0,009892587 | up | 1,8548483 | BRPF3 | Homo sapiens bromodomain and PHD finger containing, 3 (BRPF3), mRNA [NM_015695] |
| A_33_P3375077 | 0,030378807 | up | 2,156738 | BRPF3 | Homo sapiens bromodomain and PHD finger containing, 3 (BRPF3), mRNA [NM_015695] |
| A_32_P489130 | 0,015476787 | up | 1,6531242 | BRWD3 | Homo sapiens bromodomain and WD repeat domain containing 3 (BRWD3), mRNA [NM_153252] |
| A_23_P408675 | 5,03E-04 | up | 1,6197914 | BTAF1 | Homo sapiens BTAF1 RNA polymerase II, B-TFIID transcription factor-associated, 170kDa (BTAF1), mRNA [NM_003972] |
| A_23_P13438 | 0,001888376 | up | 1,6038172 | BTBD10 | Homo sapiens BTB (POZ) domain containing 10 (BTBD10), transcript variant 2, mRNA [NM_032320] |
| A_33_P3354823 | 0,00290407 | up | 2,5527203 | BTBD19 | Homo sapiens BTB (POZ) domain containing 19 (BTBD19), mRNA [NM_001136537] |
| A_24_P134356 | 0,001516066 | up | 1,6596701 | BTBD3 | Homo sapiens BTB (POZ) domain containing 3 (BTBD3), transcript variant 1, mRNA [NM_014962] |

| | | | | | |
|---|---|---|---|---|---|
| A_32_P113566 | 0,02323099 | down | -1,5212191 | BTBD9 | Homo sapiens BTB (POZ) domain containing 9 (BTBD9), transcript variant 3, mRNA [NM_152733] |
| A_33_P3277514 | 0,029109191 | up | 1,7217766 | BTK | Homo sapiens Bruton agammaglobulinemia tyrosine kinase (BTK), transcript variant 3, mRNA [NM_001287344] |
| A_33_P3358923 | 0,002692393 | up | 1,5316614 | BTLA | Homo sapiens B and T lymphocyte associated (BTLA), transcript variant 1, mRNA [NM_181780] |
| A_33_P3370959 | 0,002619321 | down | -2,8369935 | BTN1A1 | Homo sapiens butyrophilin, subfamily 1, member A1 (BTN1A1), mRNA [NM_001732] |
| A_33_P3281577 | 0,007148892 | up | 1,6669568 | BTN1A1 | Homo sapiens butyrophilin, subfamily 1, member A1 (BTN1A1), mRNA [NM_001732] |
| A_21_P0000138 | 0,021222197 | up | 1,8736105 | BTN2A2 | Homo sapiens butyrophilin, subfamily 2, member A2 (BTN2A2), transcript variant 4, mRNA [NM_001197238] |
| A_23_P145218 | 0,004865241 | down | -1,5944017 | BTN2A3P | butyrophilin, subfamily 2, member A3, pseudogene [Source:HGNC Symbol;Acc:HGNC:13229] [ENST00000377662] |
| A_24_P136470 | 0,001189107 | down | -1,71171 | BTN2A3P | Homo sapiens butyrophilin, subfamily 2, member A3, pseudogene (BTN2A3P), non-coding RNA [NR_027795] |
| A_33_P3264731 | 5,50E-04 | down | -1,7097157 | BTNL10 | Homo sapiens butyrophilin-like 10 (BTNL10), mRNA [NM_001287262] |
| A_24_P713668 | 0,001830209 | up | 1,8137326 | BZW1 | Homo sapiens basic leucine zipper and W2 domains 1 (BZW1), transcript variant 4, mRNA [NM_014670] |
| A_33_P3376806 | 0,015165294 | down | -2,23423 | C10orf67 | Homo sapiens chromosome 10 open reading frame 67 (C10orf67), mRNA [NM_153714] |
| A_21_P0006995 | 0,04882063 | up | 1,8024592 | C10orf71-AS1 | Homo sapiens C10orf71 antisense RNA 1 (C10orf71-AS1), long non-coding RNA [NR_108038] |
| A_33_P3284132 | 0,016933754 | up | 2,883197 | C10orf90 | Homo sapiens chromosome 10 open reading frame 90 (C10orf90), mRNA [NM_001004298] |
| A_32_P191441 | 0,004085322 | up | 1,5091099 | C10orf90 | Homo sapiens chromosome 10 open reading frame 90 (C10orf90), mRNA [NM_001004298] |
| A_33_P3268863 | 0,001032587 | down | -3,2098944 | C11orf44 | Homo sapiens chromosome 11 open reading frame 44 (C11orf44), mRNA [NM_001271983] |
| A_23_P24529 | 0,001431932 | up | 1,5920888 | C11orf57 | Homo sapiens chromosome 11 open reading frame 57 (C11orf57), transcript variant 1, mRNA [NM_018195] |
| A_24_P364057 | 0,004171371 | down | -2,026147 | C11orf70 | Homo sapiens chromosome 11 open reading frame 70 (C11orf70), transcript variant 1, mRNA [NM_032930] |
| A_33_P3247489 | 1,24E-04 | down | -2,005608 | C11orf85 | Homo sapiens chromosome 11 open reading frame 85 (C11orf85), transcript variant 1, mRNA [NM_001037225] |
| A_24_P392151 | 0,034566443 | up | 2,1127703 | C11orf86 | Homo sapiens chromosome 11 open reading frame 86 (C11orf86), mRNA [NM_001136485] |
| A_33_P3285354 | 0,021742122 | down | -1,5483687 | C11orf95 | Homo sapiens chromosome 11 open reading frame 95 (C11orf95), mRNA [NM_001144936] |
| A_24_P876408 | 7,03E-04 | up | 1,8578663 | C11orf95 | Homo sapiens chromosome 11 open reading frame 95 (C11orf95), mRNA [NM_001144936] |
| A_32_P74409 | 0,049087975 | up | 2,3700185 | C11orf96 | Homo sapiens chromosome 11 open reading frame 96 (C11orf96), mRNA [NM_001145033] |
| A_33_P3384148 | 0,014294409 | down | -2,119955 | C12orf40 | Homo sapiens chromosome 12 open reading frame 40 (C12orf40), mRNA [NM_001031748] |
| A_32_P228167 | 0,008204586 | up | 1,5817872 | C12orf50 | Homo sapiens chromosome 12 open reading frame 50 (C12orf50), mRNA [NM_152589] |
| A_24_P769672 | 0,001613886 | up | 1,6958622 | C12orf73 | Homo sapiens chromosome 12 open reading frame 73 (C12orf73), mRNA [NM_001135570] |
| A_32_P163089 | 0,011926737 | down | -1,7622283 | C12orf75 | Homo sapiens chromosome 12 open reading frame 75 (C12orf75), mRNA [NM_001145199] |
| A_33_P3379811 | 0,008081278 | up | 1,7449297 | C12orf77 | Homo sapiens chromosome 12 open reading frame 77 (C12orf77), mRNA [NM_001101339] |
| A_33_P3606685 | 0,038166415 | up | 2,2252226 | C12orf80 | Homo sapiens chromosome 12 open reading frame 80 (C12orf80), mRNA [NM_001242696] |
| A_33_P3449815 | 0,005411675 | up | 3,3264139 | C14orf144 | Homo sapiens, clone IMAGE:4098392, mRNA, partial cds. [BC007749] |
| A_23_P379692 | 0,004892612 | up | 1,5022277 | C14orf178 | Homo sapiens chromosome 14 open reading frame 178 (C14orf178), transcript variant 1, mRNA [NM_174943] |
| A_23_P372356 | 9,64E-04 | down | -2,7665613 | C15orf32 | Homo sapiens chromosome 15 open reading frame 32 (C15orf32), transcript variant 1, mRNA [NM_153040] |
| A_23_P26024 | 0,006526126 | up | 1,8586736 | C15orf48 | Homo sapiens chromosome 15 open reading frame 48 (C15orf48), transcript variant 2, mRNA [NM_032413] |
| A_23_P163467 | 0,015977236 | up | 1,6525888 | C15orf52 | Homo sapiens chromosome 15 open reading frame 52 (C15orf52), mRNA [NM_207380] |
| A_32_P189211 | 0,00385368 | up | 1,6179454 | C15orf56 | Homo sapiens chromosome 15 open reading frame 56 (C15orf56), mRNA [NM_001039905] |
| A_33_P3407994 | 0,033229604 | up | 1,6081665 | C15orf59 | Homo sapiens chromosome 15 open reading frame 59 (C15orf59), transcript variant 1, mRNA [NM_001039614] |
| A_21_P0008654 | 0,030906381 | up | 2,2401686 | C15orf59-AS1 | |
| A_21_P0014325 | 0,00792768 | down | -2,7033167 | C16orf47 | Homo sapiens chromosome 16 open reading frame 47 (C16orf47), mRNA [NM_207385] |
| A_21_P0000053 | 0,009089874 | up | 1,6763681 | C16orf52 | Homo sapiens chromosome 16 open reading frame 52 (C16orf52), mRNA [NM_001164579] |
| A_23_P414281 | 0,003419568 | down | -2,4759495 | C16orf71 | Homo sapiens chromosome 16 open reading frame 71 (C16orf71), mRNA [NM_139170] |
| A_33_P3561747 | 0,005827852 | up | 3,1427622 | C16orf82 | Homo sapiens chromosome 16 open reading frame 82 (C16orf82), mRNA [NM_001145545] |
| A_33_P3417980 | 0,019703312 | down | -1,8801054 | C16orf86 | chromosome 16 open reading frame 86 [Source:HGNC Symbol;Acc:HGNC:33755] [ENST00000459925] |
| A_33_P3295690 | 0,00491537 | down | -1,570722 | C16orf90 | Homo sapiens chromosome 16 open reading frame 90 (C16orf90), mRNA [NM_001080524] |
| A_33_P3309201 | 0,029863568 | up | 1,892012 | C16orf95 | chromosome 16 open reading frame 95 [Source:HGNC Symbol;Acc:HGNC:40033] [ENST00000562840] |
| A_32_P212095 | 7,57E-04 | down | -2,2403986 | C17orf105 | Homo sapiens chromosome 17 open reading frame 105 (C17orf105), mRNA [NM_001136483] |
| A_33_P3339187 | 2,17E-04 | down | -3,5336208 | C17orf112 | Homo sapiens chromosome 17 open reading frame 112 (C17orf112), mRNA [NM_001243552] |
| A_19_P00320517 | 0,01833869 | down | -2,2703955 | C17orf51 | Homo sapiens chromosome 17 open reading frame 51 (C17orf51), mRNA [NM_001113434] |
| A_19_P00320842 | 0,008737954 | down | -2,0490916 | C17orf51 | Homo sapiens chromosome 17 open reading frame 51 (C17orf51), mRNA [NM_001113434] |
| A_24_P59607 | 0,03321925 | down | -1,7336564 | C17orf51 | Homo sapiens chromosome 17 open reading frame 51 (C17orf51), mRNA [NM_001113434] |
| A_19_P00316405 | 0,017694099 | down | -1,9292026 | C17orf51 | Homo sapiens chromosome 17 open reading frame 51 (C17orf51), mRNA [NM_001113434] |
| A_33_P3295838 | 0,003848652 | down | -2,3658662 | C17orf53 | Homo sapiens chromosome 17 open reading frame 53 (C17orf53), transcript variant 2, mRNA [NM_001171251] |
| A_23_P164100 | 0,036524523 | up | 2,5161004 | C17orf64 | Homo sapiens chromosome 17 open reading frame 64 (C17orf64), mRNA [NM_181707] |
| A_24_P301547 | 0,013102852 | down | -1,5883107 | C17orf80 | Homo sapiens chromosome 17 open reading frame 80 (C17orf80), transcript variant 1, mRNA [NM_017941] |
| A_33_P3410935 | 0,001054743 | down | -1,7327513 | C17orf89 | Homo sapiens chromosome 17 open reading frame 89 (C17orf89), mRNA [NM_001086521] |
| A_23_P50052 | 0,003982777 | down | -1,8659004 | C18orf12 | PREDICTED: Homo sapiens chromosome 18 open reading frame 12 (C18orf12), misc_RNA [XR_109479] |
| A_33_P3417667 | 0,031383574 | down | -1,6995401 | C18orf15 | Homo sapiens cDNA FLJ31338 fis, clone MAMGL1000184. [AK055900] |
| A_23_P345361 | 0,010653761 | up | 2,0523012 | C18orf25 | Homo sapiens chromosome 18 open reading frame 25 (C18orf25), transcript variant 1, mRNA [NM_145055] |
| A_33_P3266978 | 0,011262254 | up | 1,5762713 | C18orf42 | Homo sapiens chromosome 18 open reading frame 42 (C18orf42), mRNA [NM_001145194] |
| A_21_P0009620 | 0,003890228 | down | -2,7433755 | C18orf61 | Homo sapiens uncharacterized LOC497259 (C18orf61), long non-coding RNA [NR_049896] |
| A_33_P3300680 | 0,00242119 | down | -1,5668111 | C19orf12 | Homo sapiens chromosome 19 open reading frame 12 (C19orf12), transcript variant 3, mRNA [NM_001256046] |
| A_24_P373885 | 0,002294859 | down | -1,6359339 | C19orf25 | Homo sapiens chromosome 19 open reading frame 25 (C19orf25), mRNA [NM_152482] |
| A_23_P308632 | 0,025408313 | down | -1,6422756 | C19orf26 | Homo sapiens chromosome 19 open reading frame 26 (C19orf26), mRNA [NM_152769] |

| | | | | | |
|---|---|---|---|---|---|
| A_33_P3411945 | 0,033334192 | down | -1,7053002 | C19orf38 | Homo sapiens chromosome 19 open reading frame 38 (C19orf38), mRNA [NM_001136482] |
| A_23_P107801 | 0,001735886 | down | -1,9899907 | C19orf44 | Homo sapiens chromosome 19 open reading frame 44 (C19orf44), transcript variant 1, mRNA [NM_032207] |
| A_33_P3389363 | 0,006787806 | down | -1,877193 | C19orf54 | chromosome 19 open reading frame 54 [Source:HGNC Symbol;Acc:HGNC:24758] [ENST00000597507] |
| A_24_P247286 | 0,027553683 | up | 2,1465638 | C19orf71 | Homo sapiens chromosome 19 open reading frame 71 (C19orf71), mRNA [NM_001135580] |
| A_21_P0000177 | 0,015290594 | down | -1,6555092 | C19orf83 | chromosome 19 open reading frame 83 [Source:HGNC Symbol;Acc:HGNC:48331] [ENST00000591087] |
| A_33_P3395859 | 1,02E-05 | down | -2,3398092 | C19orf84 | Homo sapiens chromosome 19 open reading frame 84 (C19orf84), transcript variant 1, mRNA [NM_001193623] |
| A_23_P67992 | 0,002809603 | up | 1,5477301 | C1D | Homo sapiens C1D nuclear receptor corepressor (C1D), transcript variant 1, mRNA [NM_006333] |
| A_33_P3250830 | 0,013743729 | up | 2,24282 | C1orf110 | PREDICTED: Homo sapiens chromosome 1 open reading frame 110 (C1orf110), transcript variant X3, mRNA [XM_005245126] |
| A_33_P3280317 | 0,041153792 | up | 1,7212596 | C1orf137 | Homo sapiens chromosome 1 open reading frame 137 (C1orf137), mRNA [NM_001013643] |
| A_33_P3295091 | 0,003839824 | down | -5,1490974 | C1orf140 | Homo sapiens uncharacterized LOC400804 (C1orf140), long non-coding RNA [NR_024236] |
| A_33_P3395562 | 0,009299512 | up | 2,427006 | C1orf141 | Homo sapiens chromosome 1 open reading frame 141 (C1orf141), transcript variant 1, mRNA [NM_001276351] |
| A_33_P3260193 | 0,007036821 | up | 1,5297849 | C1orf141 | Homo sapiens chromosome 1 open reading frame 141 (C1orf141), transcript variant 2, mRNA [NM_001276352] |
| A_23_P74668 | 0,002065316 | down | -2,011021 | C1orf158 | Homo sapiens chromosome 1 open reading frame 158 (C1orf158), mRNA [NM_152290] |
| A_33_P3299081 | 4,04E-04 | up | 2,6546507 | C1orf159 | chromosome 1 open reading frame 159 [Source:HGNC Symbol;Acc:HGNC:26062] [ENST00000379319] |
| A_23_P412562 | 0,002009431 | up | 1,7276855 | C1orf162 | Homo sapiens chromosome 1 open reading frame 162 (C1orf162), transcript variant 1, mRNA [NM_174896] |
| A_33_P3295278 | 0,005062784 | up | 1,9322988 | C1orf185 | Homo sapiens chromosome 1 open reading frame 185 (C1orf185), mRNA [NM_001136508] |
| A_33_P3359724 | 0,015535068 | down | -1,8766551 | C1orf194 | chromosome 1 open reading frame 194 [Source:HGNC Symbol;Acc:HGNC:32331] [ENST00000369942] |
| A_23_P321377 | 0,017205432 | up | 1,9437603 | C1orf216 | Homo sapiens chromosome 1 open reading frame 216 (C1orf216), mRNA [NM_152374] |
| A_32_P453321 | 0,030099818 | down | -1,5776931 | C1orf228 | Homo sapiens chromosome 1 open reading frame 228 (C1orf228), mRNA [NM_001145636] |
| A_24_P234116 | 1,09E-05 | up | 2,1553545 | C1orf56 | Homo sapiens chromosome 1 open reading frame 56 (C1orf56), mRNA [NM_017860] |
| A_21_P0014079 | 4,27E-04 | down | -3,8189242 | C1orf61 | chromosome 1 open reading frame 61 [Source:HGNC Symbol;Acc:HGNC:30780] [ENST00000476966] |
| A_23_P376172 | 3,23E-05 | down | -2,8939626 | C1orf64 | Homo sapiens chromosome 1 open reading frame 64 (C1orf64), mRNA [NM_178840] |
| A_33_P3359354 | 0,00227733 | down | -1,687056 | C1orf86 | Homo sapiens chromosome 1 open reading frame 86 (C1orf86), transcript variant 7, mRNA [NM_001282671] |
| A_24_P49214 | 0,014438279 | up | 1,504375 | C1orf86 | Homo sapiens chromosome 1 open reading frame 86 (C1orf86), transcript variant 7, mRNA [NM_001282671] |
| A_21_P0001087 | 0,015655853 | up | 1,6109104 | C1orf86 | chromosome 1 open reading frame 86 [Source:HGNC Symbol;Acc:HGNC:26428] [ENST00000420964] |
| A_23_P50928 | 0,0209881 | up | 2,9088085 | C1QL2 | Homo sapiens complement component 1, q subcomponent-like 2 (C1QL2), mRNA [NM_182528] |
| A_23_P259292 | 0,011675671 | down | -1,6602038 | C1QTNF5 | Homo sapiens C1q and tumor necrosis factor related protein 5 (C1QTNF5), transcript variant 1, mRNA [NM_015645] |
| A_23_P400505 | 0,037478536 | up | 1,8946884 | C1QTNF9 | Homo sapiens C1q and tumor necrosis factor related protein 9 (C1QTNF9), transcript variant 2, mRNA [NM_178540] |
| A_33_P3230478 | 0,002951325 | down | -2,517501 | C1S | Homo sapiens complement component 1, s subcomponent (C1S), transcript variant 1, mRNA [NM_201442] |
| A_33_P3399875 | 0,002426516 | down | -2,303656 | C20orf144 | Homo sapiens chromosome 20 open reading frame 144 (C20orf144), mRNA [NM_080825] |
| A_23_P314835 | 9,88E-05 | down | -3,350432 | C20orf85 | Homo sapiens chromosome 20 open reading frame 85 (C20orf85), mRNA [NM_178456] |
| A_23_P132139 | 5,35E-05 | down | -3,4410741 | C21orf58 | Homo sapiens chromosome 21 open reading frame 58 (C21orf58), transcript variant 2, mRNA [NM_001286462] |
| A_23_P166431 | 0,027236141 | up | 2,2669718 | C22orf24 | Homo sapiens chromosome 22 open reading frame 24 (C22orf24), transcript variant 1, mRNA [NM_001302819] |
| A_24_P940149 | 6,92E-05 | up | 1,9645551 | C2CD2 | Homo sapiens C2 calcium-dependent domain containing 2 (C2CD2), transcript variant 1, mRNA [NM_015500] |
| A_33_P3215412 | 0,007303437 | up | 1,6405735 | C2CD4C | Homo sapiens C2 calcium-dependent domain containing 4C (C2CD4C), mRNA [NM_001136263] |
| A_21_P0000205 | 3,79E-04 | down | -3,7655904 | C2orf48 | Homo sapiens chromosome 2 open reading frame 48 (C2orf48), mRNA [NM_182626] |
| A_24_P883629 | 4,42E-04 | up | 2,2073674 | C2orf49 | Homo sapiens chromosome 2 open reading frame 49 (C2orf49), transcript variant 1, mRNA [NM_024093] |
| A_33_P3398998 | 5,66E-04 | down | -2,6267378 | C2orf50 | Homo sapiens chromosome 2 open reading frame 50 (C2orf50), mRNA [NM_182500] |
| A_24_P297526 | 0,026571114 | up | 1,5326529 | C2orf50 | chromosome 2 open reading frame 50 [Source:HGNC Symbol;Acc:HGNC:26324] [ENST00000381585] |
| A_23_P60990 | 0,027017508 | up | 2,402772 | C2orf54 | Homo sapiens chromosome 2 open reading frame 54 (C2orf54), transcript variant 2, mRNA [NM_024861] |
| A_33_P3265714 | 3,98E-05 | down | -7,859415 | C2orf61 | Homo sapiens chromosome 2 open reading frame 61 (C2orf61), transcript variant 2, mRNA [NM_173649] |
| A_24_P413941 | 0,003602079 | up | 1,546933 | C2orf69 | Homo sapiens chromosome 2 open reading frame 69 (C2orf69), mRNA [NM_153689] |
| A_33_P3416682 | 0,011818631 | down | -1,5430188 | C2orf91 | Homo sapiens chromosome 2 open reading frame 91 (C2orf91), mRNA [NM_001242815] |
| A_23_P2431 | 0,001250125 | down | -2,1110663 | C3AR1 | Homo sapiens complement component 3a receptor 1 (C3AR1), mRNA [NM_004054] |
| A_23_P395585 | 7,72E-04 | down | -2,2261655 | C3orf22 | chromosome 3 open reading frame 22 [Source:HGNC Symbol;Acc:HGNC:28534] [ENST00000318225] |
| A_32_P72492 | 0,003761428 | up | 1,5031688 | C3orf30 | Homo sapiens chromosome 3 open reading frame 30 (C3orf30), mRNA [NM_152539] |
| A_23_P301476 | 0,0331316 | up | 1,585251 | C3orf33 | Homo sapiens chromosome 3 open reading frame 33 (C3orf33), mRNA [NM_173657] |
| A_32_P430695 | 0,013790251 | up | 1,5842432 | C3orf35 | Homo sapiens chromosome 3 open reading frame 35 (C3orf35), transcript variant B, mRNA [NM_178339] |
| A_33_P3238548 | 0,003659779 | up | 2,674651 | C3orf36 | Homo sapiens chromosome 3 open reading frame 36 (C3orf36), mRNA [NM_025041] |
| A_33_P3237507 | 0,030876918 | up | 2,255131 | C4orf50 | chromosome 4 open reading frame 50 [Source:HGNC Symbol;Acc:HGNC:33766] [ENST00000531445] |
| A_33_P3223002 | 0,013509703 | up | 1,5923036 | C5AR2 | Homo sapiens complement component 5a receptor 2 (C5AR2), transcript variant 1, mRNA [NM_001271749] |
| A_23_P403081 | 0,001000011 | up | 3,1656044 | C5orf34 | Homo sapiens chromosome 5 open reading frame 34 (C5orf34), mRNA [NM_198566] |
| A_33_P3262351 | 0,009470932 | down | -3,2358284 | C5orf49 | Homo sapiens chromosome 5 open reading frame 49 (C5orf49), mRNA [NM_001089584] |
| A_21_P0000174 | 1,50E-04 | down | -5,5968323 | C5orf56 | Homo sapiens cDNA, FLJ99232. [AK309191] |
| A_33_P3296198 | 0,002799848 | down | -1,5142679 | C5orf63 | Homo sapiens chromosome 5 open reading frame 63 (C5orf63), transcript variant 1, mRNA [NM_001164479] |
| A_33_P3660204 | 0,017682819 | down | -1,9066949 | C5orf66-AS2 | Homo sapiens C5orf66 antisense RNA 2 (C5orf66-AS2), long non-coding RNA [NR_037895] |
| A_33_P3404739 | 0,01775147 | up | 1,8065815 | C6orf141 | PREDICTED: Homo sapiens chromosome 6 open reading frame 141 (C6orf141), transcript variant X2, mRNA [XM_005248851] |
| A_33_P3422374 | 0,007171255 | up | 1,631184 | C6orf163 | Homo sapiens chromosome 6 open reading frame 163 (C6orf163), mRNA [NM_001010868] |
| A_23_P308042 | 0,006316056 | up | 1,618673 | C6orf195 | Homo sapiens chromosome 6 open reading frame 195 (C6orf195), mRNA [NM_152554] |
| A_23_P81934 | 0,037040733 | up | 1,6466604 | C6orf25 | Homo sapiens chromosome 6 open reading frame 25 (C6orf25), transcript variant 1, mRNA [NM_025260] |
| A_23_P93602 | 0,04388638 | down | -1,7513932 | C6orf58 | Homo sapiens chromosome 6 open reading frame 58 (C6orf58), mRNA [NM_001010905] |

| | | | | | |
|---|---|---|---|---|---|
| A_21_P0014171 | 0,009925813 | down | -1,7401906 | C6orf62 | PREDICTED: Homo sapiens chromosome 6 open reading frame 62 (C6orf62), transcript variant X1, mRNA [XM_005249433] |
| A_23_P213857 | 1,46E-04 | down | -3,2527456 | C7 | Homo sapiens complement component 7 (C7), mRNA [NM_000587] |
| A_33_P3226710 | 0,00100096 | down | -1,7926316 | C7orf13 | Homo sapiens chromosome 7 open reading frame 13 (C7orf13), long non-coding RNA [NR_026865] |
| A_23_P157316 | 0,008317599 | down | -1,8962457 | C7orf34 | Homo sapiens chromosome 7 open reading frame 34 (C7orf34), mRNA [NM_178829] |
| A_33_P3405349 | 0,002907356 | down | -2,0664725 | C7orf43 | chromosome 7 open reading frame 43 [Source:HGNC Symbol;Acc:HGNC:25604] [ENST00000448720] |
| A_23_P93704 | 0,009477842 | down | -1,9422268 | C7orf61 | Homo sapiens chromosome 7 open reading frame 61 (C7orf61), mRNA [NM_001004323] |
| A_33_P3401397 | 1,19E-04 | down | -2,5869439 | C7orf65 | Homo sapiens chromosome 7 open reading frame 65 (C7orf65), mRNA [NM_001123065] |
| A_33_P3401003 | 0,005568145 | up | 2,0440712 | C7orf71 | Homo sapiens chromosome 7 open reading frame 71 (C7orf71), transcript variant 1, mRNA [NM_001145531] |
| A_33_P3222822 | 0,006868362 | up | 1,5260484 | C7orf76 | Homo sapiens chromosome 7 open reading frame 76 (C7orf76), transcript variant 1, mRNA [NM_001201450] |
| A_23_P46639 | 0,006902554 | down | -2,498289 | C8A | Homo sapiens complement component 8, alpha polypeptide (C8A), mRNA [NM_000562] |
| A_33_P3319006 | 4,48E-04 | down | -1,7982696 | C8G | Homo sapiens complement component 8, gamma polypeptide (C8G), mRNA [NM_000606] |
| A_21_P0000720 | 0,049018677 | up | 2,0051517 | C8orf37-AS1 | Homo sapiens C8orf37 antisense RNA 1 (C8orf37-AS1), transcript variant 5, long non-coding RNA [NR_038205] |
| A_33_P3410849 | 2,17E-05 | down | -2,0052426 | C8orf58 | Homo sapiens chromosome 8 open reading frame 58 (C8orf58), transcript variant 1, mRNA [NM_001013842] |
| A_33_P3416513 | 2,53E-04 | down | -3,1108584 | C8orf74 | Homo sapiens chromosome 8 open reading frame 74 (C8orf74), mRNA [NM_001040032] |
| A_33_P3287997 | 3,08E-05 | down | -3,2246013 | C9orf106 | Homo sapiens chromosome 9 open reading frame 106 (C9orf106), mRNA [NM_001012715] |
| A_23_P312646 | 0,001104734 | up | 1,5398448 | C9orf142 | Homo sapiens chromosome 9 open reading frame 142 (C9orf142), mRNA [NM_183241] |
| A_24_P212860 | 0,00370285 | down | -1,775879 | C9orf156 | Homo sapiens chromosome 9 open reading frame 156 (C9orf156), mRNA [NM_016481] |
| A_23_P331092 | 0,003785853 | down | -1,6645281 | C9orf163 | Homo sapiens chromosome 9 open reading frame 163 (C9orf163), mRNA [NM_152571] |
| A_33_P3354847 | 0,005497535 | up | 1,5073663 | C9orf171 | Homo sapiens chromosome 9 open reading frame 171 (C9orf171), transcript variant 1, mRNA [NM_207417] |
| A_33_P3359223 | 0,011334027 | down | -1,7157632 | C9orf173 | Homo sapiens chromosome 9 open reading frame 173 (C9orf173), transcript variant 1, mRNA [NM_001256699] |
| A_24_P303874 | 0,001281957 | down | -2,0948942 | C9orf62 | Homo sapiens chromosome 9 open reading frame 62 (C9orf62), mRNA [NM_173520] |
| A_33_P3353662 | 0,005055436 | up | 1,8618675 | C9orf66 | Homo sapiens chromosome 9 open reading frame 66 (C9orf66), mRNA [NM_152569] |
| A_33_P3267745 | 0,001190263 | down | -2,0081553 | C9orf69 | Homo sapiens chromosome 9 open reading frame 69 (C9orf69), transcript variant 2, mRNA [NM_001256526] |
| A_32_P341615 | 0,004157216 | up | 3,114894 | C9orf84 | Homo sapiens chromosome 9 open reading frame 84 (C9orf84), transcript variant 1, mRNA [NM_173521] |
| A_23_P4096 | 0,013583898 | down | -1,6254097 | CA4 | Homo sapiens carbonic anhydrase IV (CA4), mRNA [NM_000717] |
| A_19_P00320384 | 0,001966541 | down | -1,973875 | CA5BP1 | carbonic anhydrase VB pseudogene 1 [Source:HGNC Symbol;Acc:HGNC:29544] [ENST00000380336] |
| A_33_P3334398 | 0,011373877 | down | -1,6157086 | CA6 | Homo sapiens carbonic anhydrase VI (CA6), transcript variant 2, mRNA [NM_001270500] |
| A_23_P83838 | 0,00373772 | down | -2,4077115 | CA8 | Homo sapiens carbonic anhydrase VIII (CA8), mRNA [NM_004056] |
| A_33_P3213685 | 0,022332134 | down | -1,758655 | CAB39L | Homo sapiens calcium binding protein 39-like (CAB39L), transcript variant 4, mRNA [NM_001287339] |
| A_33_P3302354 | 4,99E-04 | up | 2,092675 | CABIN1 | Homo sapiens calcineurin binding protein 1 (CABIN1), transcript variant 1, mRNA [NM_001199281] |
| A_24_P241057 | 0,045949005 | up | 1,9105861 | CABP2 | Homo sapiens calcium binding protein 2 (CABP2), mRNA [NM_016366] |
| A_23_P308021 | 0,00464069 | down | -1,8930149 | CABP4 | Homo sapiens calcium binding protein 4 (CABP4), transcript variant 1, mRNA [NM_145200] |
| A_23_P21747 | 0,010721117 | down | -1,5862623 | CABP5 | Homo sapiens calcium binding protein 5 (CABP5), mRNA [NM_019855] |
| A_33_P3348061 | 0,006846719 | up | 2,160392 | CABP7 | Homo sapiens calcium binding protein 7 (CABP7), mRNA [NM_182527] |
| A_33_P3280106 | 0,029164301 | down | -2,0901303 | CACNA1B | Homo sapiens calcium channel, voltage-dependent, N type, alpha 1B subunit (CACNA1B), transcript variant 2, mRNA [NM_001243812] |
| A_32_P153773 | 1,44E-05 | down | -3,7969358 | CACNA1B | Homo sapiens calcium channel, voltage-dependent, N type, alpha 1B subunit (CACNA1B), transcript variant 1, mRNA [NM_000718] |
| A_23_P373031 | 8,25E-06 | down | -2,7767072 | CACNA1C | Homo sapiens calcium channel, voltage-dependent, L type, alpha 1C subunit (CACNA1C), transcript variant 18, mRNA [NM_000719] |
| A_33_P3350566 | 3,73E-04 | down | -2,3052325 | CACNA1I | Homo sapiens calcium channel, voltage-dependent, T type, alpha 1I subunit (CACNA1I), transcript variant 1, mRNA [NM_021096] |
| A_33_P3373259 | 0,006270819 | up | 3,3288553 | CACNA2D3 | Homo sapiens calcium channel, voltage-dependent, alpha 2/delta subunit 3 (CACNA2D3), mRNA [NM_018398] |
| A_24_P402825 | 0,001253673 | up | 1,8601954 | CACNA2D3 | Homo sapiens calcium channel, voltage-dependent, alpha 2/delta subunit 3 (CACNA2D3), mRNA [NM_018398] |
| A_24_P91165 | 0,009269845 | down | -1,9518722 | CACNB1 | Homo sapiens calcium channel, voltage-dependent, beta 1 subunit (CACNB1), transcript variant 3, mRNA [NM_199248] |
| A_33_P3221989 | 0,013163974 | up | 2,7490482 | CACNB4 | Homo sapiens calcium channel, voltage-dependent, beta 4 subunit (CACNB4), transcript variant 1, mRNA [NM_001005747] |
| A_23_P66180 | 5,42E-04 | down | -4,5994687 | CACNG3 | Homo sapiens calcium channel, voltage-dependent, gamma subunit 3 (CACNG3), mRNA [NM_006539] |
| A_24_P70303 | 7,39E-04 | down | -2,302264 | CACNG4 | Homo sapiens calcium channel, voltage-dependent, gamma subunit 4 (CACNG4), mRNA [NM_014405] |
| A_23_P4782 | 0,00147378 | down | -2,0308728 | CACNG7 | Homo sapiens calcium channel, voltage-dependent, gamma subunit 7 (CACNG7), mRNA [NM_031896] |
| A_33_P3356290 | 0,042340543 | up | 2,519178 | CADPS | Homo sapiens Ca++-dependent secretion activator (CADPS), transcript variant 3, mRNA [NM_183393] |
| A_33_P3354001 | 0,048061475 | down | -2,105878 | CADPS2 | Homo sapiens Ca++-dependent secretion activator 2 (CADPS2), transcript variant 3, mRNA [NM_001167940] |
| A_23_P98995 | 3,51E-04 | up | 2,2245557 | CALCOCO1 | Homo sapiens calcium binding and coiled-coil domain 1 (CALCOCO1), transcript variant 1, mRNA [NM_020898] |
| A_33_P3306103 | 5,78E-04 | down | -3,8219736 | CALCRL | Homo sapiens calcitonin receptor-like (CALCRL), transcript variant 1, mRNA [NM_005795] |
| A_33_P3261957 | 0,005408529 | down | -2,4857736 | CALCRL | Homo sapiens calcitonin receptor-like (CALCRL), transcript variant 1, mRNA [NM_005795] |
| A_33_P3306110 | 0,03281209 | down | -2,0890841 | CALCRL | Homo sapiens calcitonin receptor-like (CALCRL), transcript variant 1, mRNA [NM_005795] |
| A_32_P812268 | 1,33E-07 | down | -4,322479 | CALHM1 | Homo sapiens calcium homeostasis modulator 1 (CALHM1), mRNA [NM_001001412] |
| A_24_P313186 | 0,001296904 | up | 2,2438834 | CALM1 | Homo sapiens calmodulin 1 (phosphorylase kinase, delta) (CALM1), mRNA [NM_006888] |
| A_23_P393080 | 0,010239303 | down | -1,542385 | CALML3 | Homo sapiens calmodulin-like 3 (CALML3), mRNA [NM_005185] |
| A_33_P3397935 | 0,031458963 | down | -1,8161145 | CALML3-AS1 | Homo sapiens CALML3 antisense RNA 1 (CALML3-AS1), transcript variant 1, long non-coding RNA [NR_120496] |
| A_23_P129188 | 0,004761655 | up | 1,5682592 | CALML4 | Homo sapiens calmodulin-like 4 (CALML4), transcript variant 1, mRNA [NM_033429] |
| A_33_P3292854 | 0,038584128 | down | -1,6172713 | CALR | Homo sapiens calreticulin (CALR), mRNA [NM_004343] |
| A_33_P3294149 | 0,008315786 | down | -2,1864622 | CALY | calcyon neuron-specific vesicular protein [Source:HGNC Symbol;Acc:HGNC:17938] [ENST00000467611] |
| A_33_P3294153 | 0,003345588 | down | -1,8922855 | CALY | calcyon neuron-specific vesicular protein [Source:HGNC Symbol;Acc:HGNC:17938] [ENST00000368555] |
| A_33_P3318646 | 1,64E-05 | down | -4,245599 | CALY | Homo sapiens calcyon neuron-specific vesicular protein (CALY), mRNA [NM_015722] |
| A_24_P141332 | 0,006458008 | up | 1,6508731 | CAMK2G | Homo sapiens calcium/calmodulin-dependent protein kinase II gamma (CAMK2G), transcript variant 1, mRNA [NM_172171] |

| | | | | | |
|---|---|---|---|---|---|
| A_23_P11800 | 0,003929476 | up | 1,9782262 | CAMK2N1 | Homo sapiens calcium/calmodulin-dependent protein kinase II inhibitor 1 (CAMK2N1), mRNA [NM_018584] |
| A_33_P3244283 | 0,010771492 | down | -1,8027769 | CAMK2N2 | Homo sapiens calcium/calmodulin-dependent protein kinase II inhibitor 2 (CAMK2N2), mRNA [NM_033259] |
| A_23_P29680 | 0,002454664 | up | 1,7136111 | CAMKV | Homo sapiens CaM kinase-like vesicle-associated (CAMKV), mRNA [NM_024046] |
| A_23_P253791 | 0,019852001 | down | -3,2464259 | CAMP | Homo sapiens cathelicidin antimicrobial peptide (CAMP), mRNA [NM_004345] |
| A_24_P175460 | 0,006971636 | up | 1,6087166 | CAMSAP1 | Homo sapiens calmodulin regulated spectrin-associated protein 1 (CAMSAP1), mRNA [NM_015447] |
| A_24_P47681 | 2,43E-04 | down | -3,8831863 | CAND1 | cullin-associated and neddylation-dissociated 1 [Source:HGNC Symbol;Acc:HGNC:30688] [ENST00000545606] |
| A_33_P3216938 | 0,032763 | down | -1,9497224 | CAND1.11 | Homo sapiens uncharacterized LOC100130460 (CAND1.11), long non-coding RNA [NR_103765] |
| A_24_P116669 | 0,025507515 | down | -1,50759 | CANT1 | Homo sapiens calcium activated nucleotidase 1 (CANT1), transcript variant 1, mRNA [NM_138793] |
| A_33_P3325131 | 0,027410096 | down | -1,720297 | CANX | Homo sapiens calnexin (CANX), transcript variant 1, mRNA [NM_001746] |
| A_33_P3739260 | 0,012642053 | down | -1,7218921 | CAP2 | Homo sapiens CAP, adenylate cyclase-associated protein, 2 (yeast) (CAP2), mRNA [NM_006366] |
| A_32_P167076 | 0,001388048 | up | 2,767674 | CAPN14 | Homo sapiens calpain 14 (CAPN14), mRNA [NM_001145122] |
| A_24_P282210 | 0,003317926 | up | 1,6330211 | CAPN15 | Homo sapiens calpain 15 (CAPN15), mRNA [NM_005632] |
| A_23_P23924 | 0,014474309 | up | 1,5532141 | CAPN2 | Homo sapiens calpain 2, (m/II) large subunit (CAPN2), transcript variant 1, mRNA [NM_001748] |
| A_33_P3342752 | 6,61E-04 | up | 3,0172226 | CAPN8 | calpain 8 [Source:HGNC Symbol;Acc:HGNC:1485] [ENST00000366873] |
| A_24_P56281 | 0,030871319 | up | 2,0110452 | CAPRIN2 | Homo sapiens caprin family member 2 (CAPRIN2), transcript variant 1, mRNA [NM_001002259] |
| A_32_P209208 | 0,01743113 | down | -2,6199665 | CAPS2 | Homo sapiens calcyphosine 2 (CAPS2), transcript variant 1, mRNA [NM_032606] |
| A_23_P133536 | 0,010121712 | down | -2,2008352 | CAPSL | Homo sapiens calcyphosine-like (CAPSL), transcript variant 1, mRNA [NM_144647] |
| A_23_P87727 | 9,79E-04 | up | 2,1819143 | CAPZA3 | Homo sapiens capping protein (actin filament) muscle Z-line, alpha 3 (CAPZA3), mRNA [NM_033328] |
| A_33_P3235561 | 0,045450002 | up | 1,7201736 | CAPZB | Homo sapiens capping protein (actin filament) muscle Z-line, beta (CAPZB), transcript variant 3, mRNA [NM_001206541] |
| A_33_P3302687 | 0,004589391 | down | -1,9119489 | CARKD | Homo sapiens carbohydrate kinase domain containing (CARKD), transcript variant 2, mRNA [NM_001242881] |
| A_23_P95231 | 6,82E-05 | up | 3,9417884 | CASC1 | Homo sapiens cancer susceptibility candidate 1 (CASC1), transcript variant 1, mRNA [NM_018272] |
| A_33_P3538104 | 0,009089293 | down | -3,6091278 | CASC22 | cancer susceptibility candidate 22 (non-protein coding) [Source:HGNC Symbol;Acc:HGNC:50627] [ENST00000569713] |
| A_21_P0009099 | 4,61E-04 | up | 2,8797824 | CASC22 | PREDICTED: Homo sapiens uncharacterized LOC283854 (LOC283854), misc_RNA [XR_243451] |
| A_21_P0013146 | 0,00601698 | up | 1,6315575 | CASC6 | Homo sapiens cancer susceptibility candidate 6 (non-protein coding) (CASC6), long non-coding RNA [NR_104154] |
| A_19_P00316700 | 0,004191788 | up | 1,529514 | CASC8 | Homo sapiens cancer susceptibility candidate 8 (non-protein coding) (CASC8), transcript variant 2, long non-coding RNA [NR_024393] |
| A_19_P00316699 | 0,008812192 | up | 1,6398703 | CASC8 | Homo sapiens cancer susceptibility candidate 8 (non-protein coding) (CASC8), transcript variant 2, long non-coding RNA [NR_024393] |
| A_24_P69379 | 0,007251092 | up | 1,8421323 | CASD1 | Homo sapiens CAS1 domain containing 1 (CASD1), mRNA [NM_022900] |
| A_33_P3349552 | 0,012569573 | down | -1,7314166 | CASKIN1 | Homo sapiens CASK interacting protein 1 (CASKIN1), mRNA [NM_020764] |
| A_24_P219378 | 1,33E-04 | up | 1,7895368 | CASKIN1 | Homo sapiens CASK interacting protein 1 (CASKIN1), mRNA [NM_020764] |
| A_24_P3045 | 0,03393119 | down | -1,5223879 | CASP10 | Homo sapiens caspase 10, apoptosis-related cysteine peptidase (CASP10), transcript variant 2, mRNA [NM_032974] |
| A_23_P13102 | 2,76E-04 | down | -3,3307505 | CASP12 | Homo sapiens caspase 12 (gene/pseudogene) (CASP12), transcript variant 1, mRNA [NM_001191016] |
| A_33_P3308387 | 0,014873353 | down | -1,6307902 | CASP2 | Homo sapiens caspase 2, apoptosis-related cysteine peptidase (CASP2), transcript variant 1, mRNA [NM_032982] |
| A_33_P3278118 | 0,008159657 | down | -1,5568461 | CASP3 | Homo sapiens caspase 3, apoptosis-related cysteine peptidase (CASP3), transcript variant alpha, mRNA [NM_004346] |
| A_33_P3382959 | 4,07E-04 | up | 1,7423341 | CASP8AP2 | Homo sapiens caspase 8 associated protein 2 (CASP8AP2), transcript variant 1, mRNA [NM_012115] |
| A_33_P3338300 | 0,01847792 | up | 1,7063329 | CASZ1 | Homo sapiens castor zinc finger 1 (CASZ1), transcript variant 1, mRNA [NM_001079843] |
| A_33_P3335257 | 0,015389154 | down | -1,5475271 | CATSPERD | Homo sapiens catsper channel auxiliary subunit delta (CATSPERD), mRNA [NM_152784] |
| A_33_P3335262 | 7,41E-05 | down | -2,245983 | CATSPERD | Homo sapiens cDNA FLJ46209 fis, clone TESTI4011616. [AK128088] |
| A_24_P925040 | 0,00365774 | down | -2,072815 | CAV2 | Homo sapiens caveolin 2 (CAV2), transcript variant 1, mRNA [NM_001233] |
| A_23_P212715 | 0,013763597 | up | 1,5976015 | CBLB | Homo sapiens Cbl proto-oncogene B, E3 ubiquitin protein ligase (CBLB), mRNA [NM_170662] |
| A_33_P3385516 | 0,035538558 | down | -2,1539981 | CBLN4 | Homo sapiens cerebellin 4 precursor (CBLN4), mRNA [NM_080617] |
| A_24_P298174 | 0,00341654 | down | -1,5970433 | CBX1 | Homo sapiens chromobox homolog 1 (CBX1), transcript variant 1, mRNA [NM_006807] |
| A_33_P3423954 | 3,93E-05 | down | -2,3307319 | CBX2 | Homo sapiens chromobox homolog 2 (CBX2), transcript variant 2, mRNA [NM_032647] |
| A_33_P3270485 | 0,00533068 | up | 1,8114058 | CBX6 | chromobox homolog 6 [Source:HGNC Symbol;Acc:HGNC:1556] [ENST00000407418] |
| A_33_P3343845 | 0,019814452 | down | -1,8198328 | CBX7 | Homo sapiens chromobox homolog 7 (CBX7), mRNA [NM_175709] |
| A_23_P348383 | 0,04713254 | down | -1,9929665 | CC2D2A | Homo sapiens coiled-coil and C2 domain containing 2A (CC2D2A), transcript variant 1, mRNA [NM_001080522] |
| A_33_P3308686 | 0,03714166 | down | -1,8379667 | CC2D2A | Homo sapiens coiled-coil and C2 domain containing 2A (CC2D2A), transcript variant 2, mRNA [NM_020785] |
| A_33_P3347343 | 4,37E-04 | down | -2,2603095 | CCDC102B | Homo sapiens coiled-coil domain containing 102B (CCDC102B), transcript variant 1, mRNA [NM_001093729] |
| A_23_P26928 | 0,015572618 | down | -1,9606558 | CCDC103 | family with sequence similarity 187, member A [Source:HGNC Symbol;Acc:HGNC:35153] [ENST00000331733] |
| A_33_P3309741 | 0,010708613 | down | -2,4800415 | CCDC108 | Homo sapiens coiled-coil domain containing 108 (CCDC108), transcript variant 3, mRNA [NM_001278295] |
| A_23_P383325 | 0,002781586 | up | 1,5832578 | CCDC110 | Homo sapiens coiled-coil domain containing 110 (CCDC110), transcript variant 1, mRNA [NM_152775] |
| A_23_P342108 | 0,023967449 | up | 1,7094778 | CCDC116 | Homo sapiens coiled-coil domain containing 116 (CCDC116), mRNA [NM_152612] |
| A_23_P432034 | 0,002155626 | up | 1,6987933 | CCDC117 | Homo sapiens coiled-coil domain containing 117 (CCDC117), transcript variant 1, mRNA [NM_173510] |
| A_23_P73540 | 0,004863574 | up | 1,5290366 | CCDC120 | Homo sapiens coiled-coil domain containing 120 (CCDC120), transcript variant 3, mRNA [NM_033626] |
| A_33_P3399593 | 0,026075276 | down | -2,056184 | CCDC134 | Homo sapiens coiled-coil domain containing 134 (CCDC134), mRNA [NM_024821] |
| A_23_P311144 | 0,043209735 | up | 1,637733 | CCDC138 | Homo sapiens coiled-coil domain containing 138 (CCDC138), transcript variant 1, mRNA [NM_144978] |
| A_33_P3348884 | 0,004441567 | down | -2,8710835 | CCDC141 | Homo sapiens coiled-coil domain containing 141 (CCDC141), mRNA [NM_173648] |
| A_33_P3350753 | 0,00642123 | up | 1,7318728 | CCDC144A | Homo sapiens coiled-coil domain containing 144A (CCDC144A), transcript variant 1, mRNA [NM_014695] |
| A_33_P3472460 | 0,021069707 | up | 1,7286515 | CCDC144NL-AS1 | Homo sapiens CCDC144NL antisense RNA 1 (CCDC144NL-AS1), long non-coding RNA [NR_104185] |
| A_21_P0006788 | 0,03376422 | up | 2,3587623 | CCDC147-AS1 | Homo sapiens CCDC147 antisense RNA 1 (head to head) (CCDC147-AS1), long non-coding RNA [NR_108036] |
| A_33_P3608195 | 0,026017535 | up | 2,391328 | CCDC148-AS1 | Homo sapiens CCDC148 antisense RNA 1 (CCDC148-AS1), long non-coding RNA [NR_038850] |
| A_33_P3345294 | 0,00559675 | down | -2,1172876 | CCDC162P | coiled-coil domain containing 162, pseudogene [Source:HGNC Symbol;Acc:HGNC:21565] [ENST00000368966] |

| | | | | | |
|---|---|---|---|---|---|
| A_24_P937790 | 0,001516081 | down | -3,460304 | CCDC166 | Homo sapiens coiled-coil domain containing 166 (CCDC166), mRNA [NM_001162914] |
| A_33_P3260100 | 0,002700094 | down | -1,5527667 | CCDC167 | Homo sapiens coiled-coil domain containing 167 (CCDC167), mRNA [NM_138493] |
| A_33_P3307008 | 0,001274365 | down | -2,9033213 | CCDC168 | Homo sapiens coiled-coil domain containing 168 (CCDC168), mRNA [NM_001146197] |
| A_33_P3351686 | 0,02654928 | up | 1,656789 | CCDC17 | Homo sapiens coiled-coil domain containing 17 (CCDC17), transcript variant 1, mRNA [NM_001114938] |
| A_19_P00321342 | 0,001306799 | down | -1,7376709 | CCDC171 | coiled-coil domain containing 171 [Source:HGNC Symbol;Acc:HGNC:29828] [ENST00000478913] |
| A_24_P195081 | 0,036035486 | down | -2,2094705 | CCDC171 | Homo sapiens coiled-coil domain containing 171 (CCDC171), mRNA [NM_173550] |
| A_32_P395879 | 0,041576877 | down | -2,6437542 | CCDC171 | Homo sapiens coiled-coil domain containing 171 (CCDC171), mRNA [NM_173550] |
| A_19_P00322813 | 3,38E-04 | down | -2,3783627 | CCDC171 | coiled-coil domain containing 171 [Source:HGNC Symbol;Acc:HGNC:29828] [ENST00000478913] |
| A_33_P3409337 | 0,026217945 | down | -2,1595662 | CCDC176 | Homo sapiens coiled-coil domain containing 176 (CCDC176), mRNA [NM_025057] |
| A_24_P232790 | 0,001236759 | down | -2,334553 | CCDC177 | Homo sapiens coiled-coil domain containing 177 (CCDC177), mRNA [NM_001271507] |
| A_33_P3402838 | 0,029053606 | up | 1,6959014 | CCDC177 | Homo sapiens coiled-coil domain containing 177 (CCDC177), mRNA [NM_001271507] |
| A_33_P3246623 | 0,037874065 | down | -1,7895004 | CCDC18 | Homo sapiens coiled-coil domain containing 18 (CCDC18), mRNA [NM_206886] |
| A_23_P468 | 0,006259782 | down | -1,7990108 | CCDC181 | Homo sapiens coiled-coil domain containing 181 (CCDC181), transcript variant 2, mRNA [NM_021179] |
| A_24_P68031 | 0,004809663 | up | 1,86876 | CCDC186 | coiled-coil domain containing 186 [Source:HGNC Symbol;Acc:HGNC:24349] [ENST00000369285] |
| A_23_P381478 | 0,037384126 | up | 1,9621594 | CCDC27 | Homo sapiens coiled-coil domain containing 27 (CCDC27), mRNA [NM_152492] |
| A_33_P3271725 | 0,001256424 | down | -2,6084304 | CCDC33 | Homo sapiens coiled-coil domain containing 33 (CCDC33), transcript variant 1, mRNA [NM_025055] |
| A_33_P3328726 | 0,03785341 | down | -1,668882 | CCDC33 | Homo sapiens coiled-coil domain containing 33 (CCDC33), transcript variant 1, mRNA [NM_025055] |
| A_23_P402176 | 0,025902273 | up | 2,9043152 | CCDC42 | Homo sapiens coiled-coil domain containing 42 (CCDC42), transcript variant 1, mRNA [NM_144681] |
| A_24_P940620 | 0,026630698 | down | -1,5328398 | CCDC57 | coiled-coil domain containing 57 [Source:HGNC Symbol;Acc:HGNC:27564] [ENST00000327026] |
| A_33_P3355247 | 0,001883982 | down | -1,7335064 | CCDC69 | Homo sapiens coiled-coil domain containing 69 (CCDC69), mRNA [NM_015621] |
| A_33_P3366833 | 0,033117328 | up | 2,343939 | CCDC73 | Homo sapiens coiled-coil domain containing 73 (CCDC73), mRNA [NM_001008391] |
| A_24_P680657 | 0,029661966 | up | 2,3946407 | CCDC73 | Homo sapiens coiled-coil domain containing 73 (CCDC73), mRNA [NM_001008391] |
| A_23_P128008 | 0,04504393 | up | 1,9336817 | CCDC81 | Homo sapiens coiled-coil domain containing 81 (CCDC81), transcript variant 2, mRNA [NM_021827] |
| A_24_P636332 | 0,019390367 | up | 1,501096 | CCDC84 | Homo sapiens coiled-coil domain containing 84 (CCDC84), transcript variant 1, mRNA [NM_198489] |
| A_23_P37391 | 0,014914638 | up | 1,9355748 | CCDC85C | Homo sapiens coiled-coil domain containing 85C (CCDC85C), mRNA [NM_001144995] |
| A_33_P3351735 | 9,73E-07 | down | -5,8407197 | CCDC87 | Homo sapiens coiled-coil domain containing 87 (CCDC87), mRNA [NM_018219] |
| A_24_P192988 | 4,67E-06 | up | 4,36685 | CCDC89 | coiled-coil domain containing 89 [Source:HGNC Symbol;Acc:HGNC:26762] [ENST00000316398] |
| A_23_P79661 | 0,003301081 | up | 1,5144651 | CCDC93 | Homo sapiens coiled-coil domain containing 93 (CCDC93), mRNA [NM_019044] |
| A_23_P425681 | 7,88E-04 | down | -2,7723253 | CCK | Homo sapiens cholecystokinin (CCK), transcript variant 1, mRNA [NM_000729] |
| A_23_P162010 | 0,027224194 | up | 1,5794512 | CCKBR | Homo sapiens cholecystokinin B receptor (CCKBR), mRNA [NM_176875] |
| A_23_P218369 | 0,003124131 | up | 2,3154924 | CCL14 | Homo sapiens chemokine (C-C motif) ligand 14 (CCL14), transcript variant 3, mRNA [NM_032963] |
| A_33_P3324004 | 6,44E-04 | down | -2,0327985 | CCL15 | Homo sapiens chemokine (C-C motif) ligand 15 (CCL15), mRNA [NM_032965] |
| A_23_P55270 | 0,011740174 | up | 1,6595211 | CCL18 | Homo sapiens chemokine (C-C motif) ligand 18 (pulmonary and activation-regulated) (CCL18), mRNA [NM_002988] |
| A_23_P17065 | 0,003304017 | up | 3,2915943 | CCL20 | Homo sapiens chemokine (C-C motif) ligand 20 (CCL20), transcript variant 1, mRNA [NM_004591] |
| A_33_P3285945 | 0,022399796 | down | -1,5837394 | CCL21 | Homo sapiens chemokine (C-C motif) ligand 21 (CCL21), mRNA [NM_002989] |
| A_33_P3273623 | 0,001517262 | up | 2,7256968 | CCL21 | Homo sapiens chemokine (C-C motif) ligand 21 (CCL21), mRNA [NM_002989] |
| A_24_P313418 | 0,002711889 | up | 2,312314 | CCL22 | Homo sapiens chemokine (C-C motif) ligand 22 (CCL22), mRNA [NM_002990] |
| A_23_P215491 | 0,0339459 | down | -1,6506284 | CCL24 | Homo sapiens chemokine (C-C motif) ligand 24 (CCL24), mRNA [NM_002991] |
| A_23_P78037 | 0,006466149 | up | 1,7884573 | CCL7 | Homo sapiens chemokine (C-C motif) ligand 7 (CCL7), mRNA [NM_006273] |
| A_33_P3380772 | 0,006543832 | down | -3,1228104 | CCM2L | Homo sapiens cerebral cavernous malformation 2-like (CCM2L), mRNA [NM_080625] |
| A_23_P202837 | 0,043515656 | up | 1,549355 | CCND1 | Homo sapiens cyclin D1 (CCND1), mRNA [NM_053056] |
| A_23_P24176 | 0,007845075 | up | 1,6680956 | CCNJ | Homo sapiens cyclin J (CCNJ), transcript variant 2, mRNA [NM_019084] |
| A_24_P51588 | 0,007735003 | up | 1,5011964 | CCNL2 | Homo sapiens cyclin L2 (CCNL2), transcript variant 1, mRNA [NM_030937] |
| A_23_P98963 | 2,62E-05 | up | 1,7875533 | CCNT1 | Homo sapiens cyclin T1 (CCNT1), transcript variant b, mRNA [NM_001277842] |
| A_33_P3329922 | 0,014569812 | up | 1,9338719 | CCNYL2 | Homo sapiens cyclin Y-like 2, pseudogene (CCNYL2), non-coding RNA [NR_103829] |
| A_23_P250302 | 0,011863328 | up | 2,0989566 | CCR3 | Homo sapiens chemokine (C-C motif) receptor 3 (CCR3), transcript variant 1, mRNA [NM_001837] |
| A_23_P331928 | 0,042595293 | up | 1,9395188 | CD109 | Homo sapiens CD109 molecule (CD109), transcript variant 1, mRNA [NM_133493] |
| A_23_P61466 | 0,001437014 | down | -2,6993966 | CD163L1 | Homo sapiens CD163 molecule-like 1 (CD163L1), transcript variant 2, mRNA [NM_174941] |
| A_33_P3232080 | 0,037687495 | up | 2,0245526 | CD177 | Homo sapiens CD177 molecule (CD177), mRNA [NM_020406] |
| A_33_P3232086 | 6,48E-04 | up | 3,7797058 | CD177 | Homo sapiens mRNA for NB1 glycoprotein (NB1 gene), negative phenotype #1. [AJ310433] |
| A_23_P113572 | 5,92E-05 | down | -2,1480355 | CD19 | Homo sapiens CD19 molecule (CD19), transcript variant 2, mRNA [NM_001770] |
| A_23_P402670 | 5,37E-06 | down | -3,7468238 | CD1A | Homo sapiens CD1a molecule (CD1A), mRNA [NM_001763] |
| A_33_P3408655 | 0,004719882 | down | -2,4622576 | CD2 | Homo sapiens cDNA, FLJ17718. [AK310676] |
| A_23_P161076 | 2,48E-04 | down | -3,5607443 | CD2 | Homo sapiens CD2 molecule (CD2), mRNA [NM_001767] |
| A_24_P186539 | 0,007357374 | down | -1,7667044 | CD209 | Homo sapiens CD209 molecule (CD209), transcript variant 1, mRNA [NM_021155] |
| A_23_P85453 | 2,12E-04 | down | -3,1463666 | CD244 | Homo sapiens CD244 molecule, natural killer cell receptor 2B4 (CD244), transcript variant 1, mRNA [NM_016382] |
| A_33_P3337485 | 4,24E-06 | down | -6,5178323 | CD248 | Homo sapiens CD248 molecule, endosialin (CD248), mRNA [NM_020404] |
| A_33_P3381513 | 0,006298159 | down | -2,4580994 | CD274 | Homo sapiens CD274 molecule (CD274), transcript variant 1, mRNA [NM_014143] |
| A_23_P338479 | 0,009201013 | down | -2,061184 | CD274 | Homo sapiens CD274 molecule (CD274), transcript variant 1, mRNA [NM_014143] |
| A_24_P930975 | 0,007111243 | up | 1,8312323 | CD300LD | Homo sapiens CD300 molecule-like family member d (CD300LD), mRNA [NM_001115152] |
| A_24_P305050 | 0,006615171 | down | -2,1730132 | CD300LG | Homo sapiens CD300 molecule-like family member g (CD300LG), transcript variant 1, mRNA [NM_145273] |

| | | | | | |
|---|---|---|---|---|---|
| A_21_P0000094 | 0,046811096 | down | -2,1490846 | CD33 | Homo sapiens CD33 molecule (CD33), transcript variant 3, mRNA [NM_001177608] |
| A_23_P331748 | 0,013609384 | up | 2,5934112 | CD33 | Homo sapiens CD33 molecule (CD33), transcript variant 1, mRNA [NM_001772] |
| A_33_P3225046 | 5,56E-04 | down | -2,8348346 | CD34 | Homo sapiens CD34 molecule (CD34), transcript variant 1, mRNA [NM_001025109] |
| A_33_P3375541 | 3,04E-04 | down | -4,321667 | CD3D | Homo sapiens CD3d molecule, delta (CD3-TCR complex) (CD3D), transcript variant 1, mRNA [NM_000732] |
| A_23_P208310 | 0,008759373 | up | 1,5025588 | CD3EAP | Homo sapiens CD3e molecule, epsilon associated protein (CD3EAP), transcript variant 2, mRNA [NM_012099] |
| A_24_P295999 | 0,00230018 | down | -2,0094502 | CD4 | Homo sapiens CD4 molecule (CD4), transcript variant 1, mRNA [NM_000616] |
| A_24_P784765 | 0,00454094 | up | 1,9941286 | CD59 | Homo sapiens CD59 molecule, complement regulatory protein (CD59), transcript variant 1, mRNA [NM_203330] |
| A_23_P87879 | 0,024972405 | up | 1,7708118 | CD69 | Homo sapiens CD69 molecule (CD69), mRNA [NM_001781] |
| A_33_P3417215 | 0,037861183 | up | 2,156851 | CD72 | CD72 molecule [Source:HGNC Symbol;Acc:HGNC:1696] [ENST00000378431] |
| A_23_P70095 | 0,026682056 | up | 1,5889329 | CD74 | Homo sapiens CD74 molecule, major histocompatibility complex, class II invariant chain (CD74), transcript variant 3, mRNA [NM_001025158] |
| A_21_P0007312 | 0,010089157 | up | 2,360503 | CD81-AS1 | Homo sapiens CD81 antisense RNA 1 (CD81-AS1), long non-coding RNA [NR_108080] |
| A_33_P3228837 | 0,027940307 | down | -1,7684418 | CD8A | Homo sapiens CD8a molecule (CD8A), transcript variant 3, mRNA [NM_001145873] |
| A_32_P163247 | 8,32E-06 | down | -4,4355097 | CD8A | Homo sapiens CD8a molecule (CD8A), transcript variant 1, mRNA [NM_001768] |
| A_24_P68222 | 0,005252489 | up | 1,6286755 | CD99P1 | Homo sapiens CD99 molecule pseudogene 1 (CD99P1), transcript variant 2, non-coding RNA [NR_033381] |
| A_33_P3231557 | 0,005462163 | down | -1,5598055 | CDC14B | Homo sapiens cell division cycle 14B (CDC14B), transcript variant 1, mRNA [NM_003671] |
| A_23_P149200 | 0,001298583 | down | -2,1254947 | CDC20 | Homo sapiens cell division cycle 20 (CDC20), mRNA [NM_001255] |
| A_23_P210726 | 0,032197572 | up | 1,6517742 | CDC25B | Homo sapiens cell division cycle 25B (CDC25B), transcript variant 1, mRNA [NM_021873] |
| A_23_P1602 | 0,009650124 | up | 1,7577429 | CDC42EP2 | Homo sapiens CDC42 effector protein (Rho GTPase binding) 2 (CDC42EP2), mRNA [NM_006779] |
| A_33_P3331307 | 0,033897676 | down | -1,5629555 | CDC42EP3 | Homo sapiens CDC42 effector protein (Rho GTPase binding) 3 (CDC42EP3), transcript variant 1, mRNA [NM_006449] |
| A_23_P66891 | 0,01923892 | up | 1,5023733 | CDC42EP4 | Homo sapiens CDC42 effector protein (Rho GTPase binding) 4 (CDC42EP4), mRNA [NM_012121] |
| A_33_P3342235 | 7,86E-04 | down | -2,2596657 | CDC42EP5 | Homo sapiens CDC42 effector protein (Rho GTPase binding) 5 (CDC42EP5), mRNA [NM_145057] |
| A_23_P320887 | 0,005005766 | up | 1,7695841 | CDC42SE1 | Homo sapiens CDC42 small effector 1 (CDC42SE1), transcript variant 1, mRNA [NM_001038707] |
| A_33_P3218450 | 0,030113665 | down | -2,2372382 | CDC6 | Homo sapiens cell division cycle 6 (CDC6), mRNA [NM_001254] |
| A_33_P3296169 | 0,038098224 | down | -1,56226 | CDCA7 | Homo sapiens cell division cycle associated 7 (CDCA7), transcript variant 1, mRNA [NM_031942] |
| A_23_P251421 | 0,013571911 | up | 1,9711822 | CDCA7 | Homo sapiens cell division cycle associated 7 (CDCA7), transcript variant 1, mRNA [NM_031942] |
| A_19_P00808208 | 1,00E-04 | up | 2,5704663 | CDCA7L | Homo sapiens cell division cycle associated 7-like (CDCA7L), transcript variant 2, mRNA [NM_001127370] |
| A_32_P25357 | 5,02E-04 | down | -7,389543 | CDH15 | Homo sapiens cadherin 15, type 1, M-cadherin (myotubule) (CDH15), mRNA [NM_004933] |
| A_23_P55586 | 0,02983519 | up | 1,8347712 | CDH20 | Homo sapiens cadherin 20, type 2 (CDH20), mRNA [NM_031891] |
| A_23_P40192 | 0,005879337 | down | -1,7318448 | CDH22 | Homo sapiens cadherin 22, type 2 (CDH22), mRNA [NM_021248] |
| A_33_P3728167 | 0,019367564 | down | -1,7221159 | CDH5 | Homo sapiens cadherin 5, type 2 (vascular endothelium) (CDH5), mRNA [NM_001795] |
| A_24_P382287 | 1,17E-05 | down | -4,0800266 | CDH7 | Homo sapiens cadherin 7, type 2 (CDH7), transcript variant b, mRNA [NM_004361] |
| A_23_P92999 | 0,023771705 | down | -2,7576294 | CDH9 | Homo sapiens cadherin 9, type 2 (T1-cadherin) (CDH9), mRNA [NM_016279] |
| A_24_P358805 | 0,006928236 | up | 1,6619374 | CDHR1 | Homo sapiens cadherin-related family member 1 (CDHR1), transcript variant 2, mRNA [NM_001171971] |
| A_23_P133338 | 0,004773735 | down | -2,2586293 | CDHR2 | Homo sapiens cadherin-related family member 2 (CDHR2), transcript variant 2, mRNA [NM_017675] |
| A_24_P352388 | 0,007394767 | up | 1,5371187 | CDHR5 | Homo sapiens cadherin-related family member 5 (CDHR5), transcript variant 3, mRNA [NM_031264] |
| A_33_P3333360 | 0,004100029 | down | -1,6626391 | CDIP1 | Homo sapiens cell death-inducing p53 target 1 (CDIP1), transcript variant 1, mRNA [NM_001199054] |
| A_24_P865226 | 0,009444265 | up | 2,686029 | CDIPT-AS1 | Homo sapiens CDIPT antisense RNA 1 (head to head) (CDIPT-AS1), transcript variant 1, long non-coding RNA [NR_015396] |
| A_33_P3288180 | 0,004274086 | up | 1,5713006 | CDK11A | Homo sapiens cyclin-dependent kinase 11A (CDK11A), transcript variant 1, mRNA [NM_024011] |
| A_24_P198820 | 9,45E-04 | up | 1,5067244 | CDK12 | Homo sapiens cyclin-dependent kinase 12 (CDK12), transcript variant 1, mRNA [NM_016507] |
| A_23_P89155 | 1,47E-04 | down | -2,0411613 | CDK3 | Homo sapiens cyclin-dependent kinase 3 (CDK3), mRNA [NM_001258] |
| A_33_P3226985 | 0,023623267 | up | 1,7322665 | CDK5R1 | Homo sapiens cyclin-dependent kinase 5, regulatory subunit 1 (p35) (CDK5R1), mRNA [NM_003885] |
| A_24_P51683 | 4,38E-04 | down | -4,809555 | CDK5R2 | Homo sapiens cyclin-dependent kinase 5, regulatory subunit 2 (p39) (CDK5R2), mRNA [NM_003936] |
| A_23_P44781 | 9,20E-04 | up | 2,2092495 | CDKAL1 | Homo sapiens CDK5 regulatory subunit associated protein 1-like 1 (CDKAL1), mRNA [NM_017774] |
| A_24_P282343 | 0,015573337 | up | 1,5056881 | CDKL5 | Homo sapiens cyclin-dependent kinase-like 5 (CDKL5), transcript variant I, mRNA [NM_003159] |
| A_23_P204696 | 4,62E-04 | down | -1,7872244 | CDKN1B | Homo sapiens cyclin-dependent kinase inhibitor 1B (p27, Kip1) (CDKN1B), mRNA [NM_004064] |
| A_23_P307502 | 0,00501539 | down | -1,904884 | CDKN2A-AS1 | Homo sapiens CDKN2A antisense RNA 1 (head to head) (CDKN2A-AS1), long non-coding RNA [NR_024274] |
| A_33_P3307903 | 0,009691667 | up | 2,482482 | CDKN3 | Homo sapiens cyclin-dependent kinase inhibitor 3 (CDKN3), transcript variant 1, mRNA [NM_005192] |
| A_23_P30294 | 0,04234366 | up | 1,5243511 | CDO1 | Homo sapiens cysteine dioxygenase type 1 (CDO1), mRNA [NM_001801] |
| A_21_P0009302 | 0,007560723 | up | 2,4652426 | CDRT1 | Homo sapiens CMT1A duplicated region transcript 1 (CDRT1), transcript variant 2, mRNA [NM_001282540] |
| A_21_P0011514 | 6,57E-04 | down | -2,6141567 | CDRT15L2 | Homo sapiens CMT1A duplicated region transcript 15-like 2 (CDRT15L2), mRNA [NM_001190790] |
| A_33_P3413895 | 1,86E-06 | down | -3,3361976 | CDRT15P1 | Homo sapiens CMT1A duplicated region transcript 15 pseudogene 1 (CDRT15P1), non-coding RNA [NR_003261] |
| A_21_P0000608 | 1,16E-06 | down | -3,8243082 | CDRT15P2 | Homo sapiens CMT1A duplicated region transcript 15 pseudogene 2 (CDRT15P2), non-coding RNA [NR_033865] |
| A_33_P3629131 | 0,001670588 | down | -1,6157066 | CDRT3 | DKFZp686P15240_r1 686 (synonym: hlcc3) Homo sapiens cDNA clone DKFZp686P15240 5', mRNA sequence [BX484257] |
| A_33_P3629151 | 0,039265983 | down | -2,067086 | CDRT8 | qf35h11.x1 Soares_testis_NHT Homo sapiens cDNA clone IMAGE:1752069 3', mRNA sequence [AI150443] |
| A_23_P7250 | 0,012445125 | up | 1,7029341 | CDS1 | Homo sapiens CDP-diacylglycerol synthase (phosphatidate cytidylyltransferase) 1 (CDS1), mRNA [NM_001263] |
| A_33_P3386262 | 0,042680535 | up | 1,7525376 | CDT1 | Homo sapiens chromatin licensing and DNA replication factor 1 (CDT1), mRNA [NM_030928] |
| A_23_P58788 | 1,52E-05 | down | -4,2847047 | CDX1 | Homo sapiens caudal type homeobox 1 (CDX1), mRNA [NM_001804] |
| A_32_P401091 | 0,040518202 | up | 1,5491887 | CDY2A | Homo sapiens chromodomain protein, Y-linked, 2A (CDY2A), mRNA [NM_004825] |
| A_23_P78526 | 0,001392291 | down | -2,4651396 | CEACAM19 | Homo sapiens carcinoembryonic antigen-related cell adhesion molecule 19 (CEACAM19), transcript variant 2, mRNA [NM_020219] |
| A_24_P118376 | 0,023620382 | down | -1,735538 | CEACAM20 | Homo sapiens carcinoembryonic antigen-related cell adhesion molecule 20 (CEACAM20), transcript variant 4S, mRNA [NM_001102598] |
| A_33_P3265309 | 0,015854158 | down | -2,123054 | CEACAM4 | Homo sapiens carcinoembryonic antigen-related cell adhesion molecule 4 (CEACAM4), mRNA [NM_001817] |

| | | | | | |
|---|---|---|---|---|---|
| A_33_P3241489 | 0,005456397 | down | -2,92738 | CEBPZOS | Homo sapiens CEBPZ opposite strand (CEBPZOS), transcript variant 1, long non-coding RNA [NR_037879] |
| A_23_P211326 | 4,57E-04 | down | -3,2235022 | CECR2 | Homo sapiens cat eye syndrome chromosome region, candidate 2 (CECR2), transcript variant 1, mRNA [NM_001290047] |
| A_23_P46238 | 0,003807392 | down | -1,6638838 | CELA2A | Homo sapiens chymotrypsin-like elastase family, member 2A (CELA2A), mRNA [NM_033440] |
| A_23_P115645 | 0,013375069 | down | -1,6101028 | CELF2 | Homo sapiens CUGBP, Elav-like family member 2 (CELF2), transcript variant 1, mRNA [NM_001025076] |
| A_23_P337849 | 0,004098706 | down | -1,6718253 | CELF3 | Homo sapiens CUGBP, Elav-like family member 3 (CELF3), transcript variant 1, mRNA [NM_007185] |
| A_24_P84396 | 0,003567911 | down | -1,8202022 | CEMIP | Homo sapiens cell migration inducing protein, hyaluronan binding (CEMIP), transcript variant 3, mRNA [NM_018689] |
| A_23_P155989 | 0,028260088 | up | 1,8596663 | CENPK | Homo sapiens centromere protein K (CENPK), transcript variant 1, mRNA [NM_022145] |
| A_33_P3387856 | 0,001934981 | up | 1,7731589 | CENPN | Homo sapiens centromere protein N (CENPN), transcript variant 1, mRNA [NM_001100625] |
| A_33_P3387931 | 0,008390521 | up | 1,7065606 | CENPP | Homo sapiens centromere protein P (CENPP), transcript variant 1, mRNA [NM_001012267] |
| A_33_P3316318 | 2,33E-04 | down | -3,0072806 | CEP120 | centrosomal protein 120kDa [Source:HGNC Symbol;Acc:HGNC:26690] [ENST00000513565] |
| A_23_P89710 | 0,00422314 | up | 1,5953163 | CEP192 | Homo sapiens centrosomal protein 192kDa (CEP192), mRNA [NM_032142] |
| A_33_P3308905 | 0,04473067 | up | 1,5482205 | CEP57L1 | Homo sapiens centrosomal protein 57kDa-like 1 (CEP57L1), transcript variant 4, mRNA [NM_001271853] |
| A_33_P3265224 | 0,017436305 | up | 1,7378798 | CEP68 | centrosomal protein 68kDa [Source:HGNC Symbol;Acc:HGNC:29076] [ENST00000537589] |
| A_33_P3349496 | 0,028558487 | up | 1,737262 | CEP83 | PREDICTED: Homo sapiens coiled-coil domain containing 41 (CCDC41), transcript variant X4, misc_RNA [XR_429106] |
| A_23_P329798 | 0,005093244 | up | 1,6173794 | CER1 | Homo sapiens cerberus 1, DAN family BMP antagonist (CER1), mRNA [NM_005454] |
| A_24_P62237 | 0,011137427 | up | 1,6700625 | CERK | Homo sapiens ceramide kinase (CERK), mRNA [NM_022766] |
| A_33_P3355538 | 0,008629097 | up | 1,5983876 | CERKL | Homo sapiens ceramide kinase-like (CERKL), transcript variant 2, mRNA [NM_001030311] |
| A_23_P206733 | 0,030430181 | down | -1,8463308 | CES1 | Homo sapiens carboxylesterase 1 (CES1), transcript variant 3, mRNA [NM_001266] |
| A_21_P0011496 | 0,003903005 | up | 2,5043967 | CES1 | Homo sapiens carboxylesterase 1 (CES1), transcript variant 1, mRNA [NM_001025195] |
| A_23_P374892 | 0,030947937 | up | 1,6062461 | CES1P1 | Homo sapiens carboxylesterase 1 pseudogene 1 (CES1P1), non-coding RNA [NR_003276] |
| A_21_P0012348 | 0,002149856 | down | -1,7301201 | CES5A | Homo sapiens carboxylesterase 5A (CES5A), transcript variant 2, mRNA [NM_145024] |
| A_23_P66967 | 0,018129745 | down | -2,2123945 | CETN1 | centrin, EF-hand protein, 1 [Source:HGNC Symbol;Acc:HGNC:1866] [ENST00000327228] |
| A_33_P3233160 | 0,006843818 | down | -1,6671313 | CFAP36 | Homo sapiens cilia and flagella associated protein 36 (CFAP36), transcript variant 1, mRNA [NM_001282761] |
| A_33_P3246193 | 0,02343372 | down | -1,889717 | CFAP46 | Homo sapiens cilia and flagella associated protein 46 (CFAP46), mRNA [NM_001200049] |
| A_23_P75179 | 0,046678837 | up | 2,0267203 | CFAP46 | Homo sapiens cilia and flagella associated protein 46 (CFAP46), mRNA [NM_001200049] |
| A_33_P3341648 | 0,004549889 | up | 3,442468 | CFAP46 | Homo sapiens cilia and flagella associated protein 46 (CFAP46), mRNA [NM_001200049] |
| A_33_P3395804 | 0,002122528 | down | -1,7715048 | CFAP61 | Homo sapiens cilia and flagella associated protein 61 (CFAP61), transcript variant 2, mRNA [NM_001167816] |
| A_33_P3420083 | 0,019846821 | down | -1,9947184 | CFAP74 | Homo sapiens cDNA FLJ37323 fis, clone BRAMY2018273. [AK094642] |
| A_33_P3336262 | 0,001368449 | down | -4,9972706 | CFAP74 | cilia and flagella associated protein 74 [Source:HGNC Symbol;Acc:HGNC:29368] [ENST00000412120] |
| A_33_P3410123 | 0,001914744 | up | 2,3813667 | CFAP74 | cilia and flagella associated protein 74 [Source:HGNC Symbol;Acc:HGNC:29368] [ENST00000378592] |
| A_33_P3265965 | 0,014522439 | down | -1,5846591 | CFAP97 | Homo sapiens cilia and flagella associated protein 97 (CFAP97), transcript variant 2, mRNA [NM_001292033] |
| A_33_P3292230 | 0,013785527 | up | 2,631982 | CFAP99 | Homo sapiens cilia and flagella associated protein 99 (CFAP99), mRNA [NM_001193282] |
| A_24_P92472 | 0,018447837 | up | 2,5921743 | CFI | Homo sapiens complement factor I (CFI), mRNA [NM_000204] |
| A_21_P0001952 | 0,001209235 | up | 2,143366 | CFLAR | Homo sapiens CASP8 and FADD-like apoptosis regulator (CFLAR), transcript variant 8, mRNA [NM_001202519] |
| A_24_P120115 | 9,73E-04 | up | 1,6651971 | CFLAR | Homo sapiens CASP8 and FADD-like apoptosis regulator (CFLAR), transcript variant 1, mRNA [NM_003879] |
| A_33_P3626816 | 0,006243705 | up | 2,457341 | CFLAR-AS1 | Homo sapiens CFLAR antisense RNA 1 (CFLAR-AS1), long non-coding RNA [NR_040030] |
| A_23_P27709 | 0,002698615 | down | -3,251489 | CGB2 | Homo sapiens chorionic gonadotropin, beta polypeptide 2 (CGB2), mRNA [NM_033378] |
| A_21_P0013947 | 0,04918295 | down | -1,5645117 | CGB7 | chorionic gonadotropin, beta polypeptide 7 [Source:HGNC Symbol;Acc:HGNC:16451] [ENST00000598442] |
| A_23_P363163 | 9,00E-04 | up | 1,6188563 | CGGBP1 | Homo sapiens CGG triplet repeat binding protein 1 (CGGBP1), transcript variant 1, mRNA [NM_001008390] |
| A_33_P3329974 | 0,030297454 | up | 1,6807488 | CGN | Homo sapiens cingulin (CGN), mRNA [NM_020770] |
| A_21_P0000061 | 3,41E-04 | down | -1,7977105 | CGREF1 | Homo sapiens cell growth regulator with EF-hand domain 1 (CGREF1), transcript variant 5, mRNA [NM_001301324] |
| A_33_P3281850 | 0,014829683 | down | -2,0037453 | CGREF1 | cell growth regulator with EF-hand domain 1 [Source:HGNC Symbol;Acc:HGNC:16962] [ENST00000440612] |
| A_32_P194264 | 0,007264215 | down | -2,3087862 | CHAC2 | Homo sapiens ChaC, cation transport regulator homolog 2 (E. coli) (CHAC2), mRNA [NM_001008708] |
| A_33_P3416366 | 0,018308803 | up | 1,5319493 | CHAF1A | Homo sapiens chromatin assembly factor 1, subunit A (p150) (CHAF1A), mRNA [NM_005483] |
| A_23_P46894 | 1,96E-06 | down | -4,754198 | CHAT | Homo sapiens choline O-acetyltransferase (CHAT), transcript variant M, mRNA [NM_020549] |
| A_24_P400376 | 0,001360349 | down | -1,7831373 | CHCHD2 | Homo sapiens coiled-coil-helix-coiled-coil-helix domain containing 2 (CHCHD2), mRNA [NM_016139] |
| A_24_P379727 | 0,001662958 | down | -1,5655147 | CHCHD5 | coiled-coil-helix-coiled-coil-helix domain containing 5 [Source:HGNC Symbol;Acc:HGNC:17840] [ENST00000489052] |
| A_33_P3315258 | 0,003169751 | down | -1,6488701 | CHD1L | Homo sapiens chromodomain helicase DNA binding protein 1-like (CHD1L), transcript variant 1, mRNA [NM_004284] |
| A_24_P85317 | 0,02884298 | down | -1,6420432 | CHD2 | Homo sapiens chromodomain helicase DNA binding protein 2 (CHD2), transcript variant 2, mRNA [NM_001042572] |
| A_24_P264664 | 0,002320646 | up | 1,760656 | CHD2 | Homo sapiens chromodomain helicase DNA binding protein 2 (CHD2), transcript variant 1, mRNA [NM_001271] |
| A_24_P289383 | 0,005215926 | up | 1,612137 | CHD7 | Homo sapiens chromodomain helicase DNA binding protein 7 (CHD7), mRNA [NM_017780] |
| A_23_P34888 | 1,90E-06 | down | -9,308448 | CHIA | Homo sapiens chitinase, acidic (CHIA), transcript variant 2, mRNA [NM_021797] |
| A_33_P3347528 | 0,006059417 | down | -2,744952 | CHIAP2 | chitinase, acidic pseudogene 2 [Source:HGNC Symbol;Acc:HGNC:44463] [ENST00000369743] |
| A_23_P110345 | 0,00926203 | up | 1,7784241 | CHIC2 | Homo sapiens cysteine-rich hydrophobic domain 2 (CHIC2), mRNA [NM_012110] |
| A_23_P314120 | 0,037599258 | up | 1,5856392 | CHKB | Homo sapiens choline kinase beta (CHKB), mRNA [NM_005198] |
| A_24_P287780 | 0,004640854 | down | -1,6041877 | CHMP1A | Homo sapiens charged multivesicular body protein 1A (CHMP1A), transcript variant 2, mRNA [NM_002768] |
| A_21_P0013823 | 0,016939819 | up | 1,7130407 | CHMP1B2P | Homo sapiens charged multivesicular body protein 1B2, pseudogene (CHMP1B2P), non-coding RNA [NR_110646] |
| A_24_P945293 | 0,049607385 | down | -1,6153692 | CHMP3 | Homo sapiens charged multivesicular body protein 3 (CHMP3), transcript variant 1, mRNA [NM_016079] |
| A_33_P3287348 | 4,00E-04 | down | -3,3547635 | CHN2 | Homo sapiens chimerin 2 (CHN2), transcript variant 4, mRNA [NM_001293070] |
| A_33_P3217198 | 0,036514025 | up | 1,7209828 | CHODL-AS1 | Homo sapiens CHODL antisense RNA 1 (CHODL-AS1), long non-coding RNA [NR_024354] |
| A_23_P138805 | 0,001178233 | up | 1,6891029 | CHORDC1 | Homo sapiens cysteine and histidine-rich domain (CHORD) containing 1 (CHORDC1), transcript variant 1, mRNA [NM_012124] |

| | | | | | |
|---|---|---|---|---|---|
| A_23_P13548 | 0,019218529 | up | 1,9116877 | CHRDL2 | chordin-like 2 [Source:HGNC Symbol;Acc:HGNC:24168] [ENST00000376332] |
| A_33_P3351298 | 0,023953466 | up | 1,9089944 | CHRM2 | cholinergic receptor, muscarinic 2 [Source:HGNC Symbol;Acc:HGNC:1951] [ENST00000480591] |
| A_21_P0013981 | 9,74E-04 | down | -2,0198975 | CHRM3 | cholinergic receptor, muscarinic 3 [Source:HGNC Symbol;Acc:HGNC:1952] [ENST00000481779] |
| A_23_P401472 | 0,047449134 | up | 2,0814574 | CHRM3 | Homo sapiens cholinergic receptor, muscarinic 3 (CHRM3), mRNA [NM_000740] |
| A_33_P3367850 | 0,006948927 | down | -2,2006845 | CHRM4 | Homo sapiens cholinergic receptor, muscarinic 4 (CHRM4), mRNA [NM_000741] |
| A_33_P3388147 | 0,014153615 | up | 2,4225132 | CHRNA3 | Homo sapiens cholinergic receptor, nicotinic, alpha 3 (neuronal) (CHRNA3), transcript variant 2, mRNA [NM_001166694] |
| A_23_P88691 | 0,011515127 | down | -1,9405705 | CHRNA5 | Homo sapiens cholinergic receptor, nicotinic, alpha 5 (neuronal) (CHRNA5), mRNA [NM_000745] |
| A_23_P157593 | 0,027669668 | down | -1,8169965 | CHRNA6 | Homo sapiens cholinergic receptor, nicotinic, alpha 6 (neuronal) (CHRNA6), transcript variant 1, mRNA [NM_004198] |
| A_23_P123424 | 0,00230288 | down | -1,9914637 | CHRNB3 | Homo sapiens cholinergic receptor, nicotinic, beta 3 (neuronal) (CHRNB3), mRNA [NM_000749] |
| A_23_P304279 | 0,010832148 | down | -2,641827 | CHRNE | Homo sapiens cholinergic receptor, nicotinic, epsilon (muscle) (CHRNE), mRNA [NM_000080] |
| A_23_P98205 | 0,0047318 | down | -2,126139 | CHST1 | Homo sapiens carbohydrate (keratan sulfate Gal-6) sulfotransferase 1 (CHST1), mRNA [NM_003654] |
| A_23_P80473 | 3,76E-04 | down | -2,3302994 | CHST13 | Homo sapiens carbohydrate (chondroitin 4) sulfotransferase 13 (CHST13), mRNA [NM_152889] |
| A_23_P430670 | 0,002399346 | down | -1,694095 | CHST5 | Homo sapiens carbohydrate (N-acetylglucosamine 6-O) sulfotransferase 5 (CHST5), mRNA [NM_024533] |
| A_23_P90273 | 1,65E-04 | down | -1,9655869 | CHST8 | Homo sapiens carbohydrate (N-acetylgalactosamine 4-0) sulfotransferase 8 (CHST8), transcript variant 3, mRNA [NM_022467] |
| A_23_P368886 | 0,011733693 | up | 2,614214 | CHSY3 | Homo sapiens chondroitin sulfate synthase 3 (CHSY3), mRNA [NM_175856] |
| A_23_P410717 | 0,001488702 | up | 1,750577 | CIART | Homo sapiens circadian associated repressor of transcription (CIART), transcript variant 1, mRNA [NM_144697] |
| A_23_P37359 | 0,02879568 | down | -1,6747525 | CIDEB | Homo sapiens cell death-inducing DFFA-like effector b (CIDEB), mRNA [NM_014430] |
| A_33_P3324394 | 0,004653296 | down | -2,6218495 | CIITA | Homo sapiens class II, major histocompatibility complex, transactivator (CIITA), transcript variant 3, mRNA [NM_001286403] |
| A_23_P372925 | 0,009690589 | up | 1,6191005 | CIPC | Homo sapiens CLOCK-interacting pacemaker (CIPC), mRNA [NM_033426] |
| A_33_P3362636 | 0,002627636 | down | -2,1129327 | CISD3 | Homo sapiens CDGSH iron sulfur domain 3 (CISD3), mRNA [NM_001136498] |
| A_33_P3312301 | 0,01122574 | up | 1,6481724 | CIT | Homo sapiens citron rho-interacting serine/threonine kinase (CIT), transcript variant 1, mRNA [NM_001206999] |
| A_23_P73526 | 0,036486465 | down | -1,8135424 | CITED1 | Homo sapiens Cbp/p300-interacting transactivator, with Glu/Asp-rich carboxy-terminal domain, 1 (CITED1), transcript variant 1, mRNA [NM_004143] |
| A_24_P215804 | 0,003729039 | down | -2,6928515 | CKLF | Homo sapiens chemokine-like factor (CKLF), transcript variant 1, mRNA [NM_016951] |
| A_23_P50250 | 0,002254428 | up | 1,7011378 | CKM | Homo sapiens creatine kinase, muscle (CKM), mRNA [NM_001824] |
| A_24_P618401 | 0,012608502 | down | -1,5598154 | CKMT2-AS1 | Homo sapiens CKMT2 antisense RNA 1 (CKMT2-AS1), transcript variant 1, long non-coding RNA [NR_034121] |
| A_23_P311232 | 6,11E-05 | up | 2,224919 | CLASP1 | Homo sapiens cytoplasmic linker associated protein 1 (CLASP1), transcript variant 1, mRNA [NM_015282] |
| A_33_P3224250 | 0,016650908 | down | -1,662424 | CLCC1 | Homo sapiens chloride channel CLIC-like 1 (CLCC1), transcript variant 1, mRNA [NM_001048210] |
| A_23_P59772 | 0,001642692 | down | -2,0665634 | CLCN1 | Homo sapiens chloride channel, voltage-sensitive 1 (CLCN1), transcript variant 1, mRNA [NM_000083] |
| A_21_P0013622 | 0,002283342 | down | -2,4269674 | CLCN3 | Homo sapiens chloride channel, voltage-sensitive 3 (CLCN3), transcript variant a, mRNA [NM_001243372] |
| A_33_P3415092 | 0,02116882 | up | 1,7488639 | CLCN5 | Homo sapiens chloride channel, voltage-sensitive 5 (CLCN5), transcript variant 1, mRNA [NM_001127899] |
| A_24_P2361 | 0,013707844 | down | -2,1040394 | CLCNKA | Homo sapiens chloride channel, voltage-sensitive Ka (CLCNKA), transcript variant 1, mRNA [NM_004070] |
| A_24_P224966 | 0,0354239 | down | -1,8670157 | CLCNKB | Homo sapiens chloride channel, voltage-sensitive Kb (CLCNKB), transcript variant 1, mRNA [NM_000085] |
| A_33_P3284036 | 0,002234023 | up | 1,670852 | CLDN14 | claudin 14 [Source:HGNC Symbol;Acc:HGNC:2035] [ENST00000478313] |
| A_33_P3249046 | 0,004168338 | up | 2,31434 | CLDN2 | Homo sapiens claudin 2 (CLDN2), transcript variant 2, mRNA [NM_001171092] |
| A_33_P3410816 | 0,04481422 | up | 1,5299324 | CLDN20 | Homo sapiens claudin 20 (CLDN20), mRNA [NM_001001346] |
| A_33_P3280750 | 0,005409367 | up | 2,0741956 | CLDN25 | Homo sapiens claudin 25 (CLDN25), mRNA [NM_001101389] |
| A_33_P3378272 | 1,31E-04 | down | -3,5065687 | CLDN4 | claudin 4 [Source:HGNC Symbol;Acc:HGNC:2046] [ENST00000431918] |
| A_23_P128470 | 0,018434938 | up | 1,6665734 | CLEC12A | Homo sapiens C-type lectin domain family 12, member A (CLEC12A), transcript variant 1, mRNA [NM_138337] |
| A_32_P194246 | 5,58E-04 | up | 1,5906976 | CLEC16A | Homo sapiens C-type lectin domain family 16, member A (CLEC16A), transcript variant 1, mRNA [NM_015226] |
| A_33_P3394362 | 0,022667361 | down | -1,5451049 | CLEC19A | Homo sapiens C-type lectin domain family 19, member A (CLEC19A), mRNA [NM_001256720] |
| A_33_P3236065 | 0,015412676 | up | 2,551869 | CLEC1B | Homo sapiens C-type lectin domain family 1, member B (CLEC1B), transcript variant 1, mRNA [NM_016509] |
| A_33_P3381255 | 0,004459971 | down | -2,4468186 | CLEC2A | Homo sapiens C-type lectin domain family 2, member A (CLEC2A), transcript variant 2, mRNA [NM_207375] |
| A_23_P48029 | 0,006869038 | up | 1,5041587 | CLEC4A | Homo sapiens C-type lectin domain family 4, member A (CLEC4A), transcript variant 1, mRNA [NM_016184] |
| A_33_P3213235 | 0,00830558 | down | -2,4781516 | CLEC4G | Homo sapiens C-type lectin domain family 4, member G (CLEC4G), transcript variant 1, mRNA [NM_198492] |
| A_23_P208482 | 0,002154949 | down | -2,4334755 | CLEC4M | Homo sapiens C-type lectin domain family 4, member M (CLEC4M), transcript variant 2, mRNA [NM_001144904] |
| A_23_P304356 | 0,005967342 | up | 1,5990499 | CLEC5A | Homo sapiens C-type lectin domain family 5, member A (CLEC5A), transcript variant 1, mRNA [NM_013252] |
| A_23_P257417 | 0,004748156 | up | 1,7403588 | CLHC1 | Homo sapiens clathrin heavy chain linker domain containing 1 (CLHC1), transcript variant 1, mRNA [NM_152385] |
| A_23_P385067 | 0,026932927 | up | 2,883143 | CLIC6 | Homo sapiens chloride intracellular channel 6 (CLIC6), mRNA [NM_053277] |
| A_21_P0014278 | 6,49E-04 | up | 1,9078076 | CLIP1-AS1 | Homo sapiens CLIP1 antisense RNA 1 (CLIP1-AS1), long non-coding RNA [NR_045382] |
| A_24_P67027 | 0,004377851 | up | 1,6833444 | CLK2 | Homo sapiens CDC-like kinase 2 (CLK2), transcript variant 1, mRNA [NM_001294338] |
| A_32_P20691 | 0,007664878 | down | -1,7279457 | CLK3 | Homo sapiens CDC-like kinase 3 (CLK3), transcript variant 2, mRNA [NM_003992] |
| A_33_P3267989 | 0,002575991 | up | 1,5702981 | CLNK | Homo sapiens cytokine-dependent hematopoietic cell linker (CLNK), mRNA [NM_052964] |
| A_23_P419038 | 0,001218614 | up | 1,5543065 | CLOCK | Homo sapiens clock circadian regulator (CLOCK), transcript variant 2, mRNA [NM_004898] |
| A_23_P8142 | 0,048650343 | down | -1,733737 | CLPS | Homo sapiens colipase, pancreatic (CLPS), transcript variant 1, mRNA [NM_001832] |
| A_33_P3394347 | 0,005423827 | down | -1,9430237 | CLRN1-AS1 | Homo sapiens CLRN1 antisense RNA 1 (CLRN1-AS1), non-coding RNA [NR_024066] |
| A_24_P61753 | 0,006105791 | up | 1,5314384 | CLUH | Homo sapiens clustered mitochondria (cluA/CLU1) homolog (CLUH), mRNA [NM_015229] |
| A_33_P3323238 | 0,003737203 | up | 1,5907513 | CLUL1 | Homo sapiens clusterin-like 1 (retinal) (CLUL1), transcript variant 3, mRNA [NM_001289036] |
| A_24_P269551 | 0,008712972 | up | 1,5566236 | CLVS1 | Homo sapiens clavesin 1 (CLVS1), mRNA [NM_173519] |
| A_21_P0008110 | 0,001238613 | down | -1,7236023 | CLYBL | Homo sapiens citrate lyase beta like (CLYBL), transcript variant 2, non-coding RNA [NR_104592] |
| A_33_P3259708 | 0,002686116 | down | -3,5881226 | CMA1 | Homo sapiens chymase 1, mast cell (CMA1), mRNA [NM_001836] |
| A_21_P0000542 | 0,003812905 | up | 1,5547174 | CMAHP | Homo sapiens cytidine monophospho-N-acetylneuraminic acid hydroxylase, pseudogene (CMAHP), transcript variant 2, non-coding RNA [NR_027626] |

| | | | | | |
|---|---|---|---|---|---|
| A_23_P371239 | 0,00102019 | up | 1,8268139 | CMIP | Homo sapiens c-Maf inducing protein (CMIP), transcript variant 1, mRNA [NM_198390] |
| A_23_P115366 | 0,006778779 | up | 1,8284441 | CMPK1 | Homo sapiens cytidine monophosphate (UMP-CMP) kinase 1, cytosolic (CMPK1), transcript variant 1, mRNA [NM_016308] |
| A_23_P106042 | 0,001992758 | down | -2,0395648 | CMTM5 | Homo sapiens CKLF-like MARVEL transmembrane domain containing 5 (CMTM5), transcript variant 3, mRNA [NM_001037288] |
| A_23_P374149 | 0,016214821 | up | 1,584496 | CMTR1 | Homo sapiens cap methyltransferase 1 (CMTR1), mRNA [NM_015050] |
| A_23_P124946 | 0,008985943 | up | 2,017345 | CMYA5 | Homo sapiens cardiomyopathy associated 5 (CMYA5), mRNA [NM_153610] |
| A_23_P9869 | 0,04576728 | down | -1,8986223 | CNDP1 | Homo sapiens carnosine dipeptidase 1 (metallopeptidase M20 family) (CNDP1), mRNA [NM_032649] |
| A_33_P3403549 | 4,01E-04 | up | 1,9591597 | CNEP1R1 | Homo sapiens CTD nuclear envelope phosphatase 1 regulatory subunit 1 (CNEP1R1), transcript variant 1, mRNA [NM_153261] |
| A_33_P3295738 | 0,012137467 | down | -1,5438634 | CNGB1 | Homo sapiens cyclic nucleotide gated channel beta 1 (CNGB1), transcript variant 2, mRNA [NM_001135639] |
| A_23_P216376 | 0,029464684 | up | 2,607827 | CNGB3 | Homo sapiens cyclic nucleotide gated channel beta 3 (CNGB3), mRNA [NM_019098] |
| A_33_P3370634 | 0,010660959 | up | 1,7067461 | CNIH1 | cornichon family AMPA receptor auxiliary protein 1 [Source:HGNC Symbol;Acc:HGNC:19431] [ENST00000557659] |
| A_24_P203315 | 0,002542454 | down | -2,6691172 | CNIH2 | Homo sapiens cornichon family AMPA receptor auxiliary protein 2 (CNIH2), transcript variant 1, mRNA [NM_182553] |
| A_33_P3240722 | 0,011586685 | up | 1,548701 | CNKSR2 | Homo sapiens connector enhancer of kinase suppressor of Ras 2 (CNKSR2), transcript variant 1, mRNA [NM_014927] |
| A_23_P138168 | 0,023698628 | up | 1,7258986 | CNN3 | Homo sapiens calponin 3, acidic (CNN3), transcript variant 1, mRNA [NM_001839] |
| A_24_P93896 | 0,00249374 | up | 2,7414887 | CNNM2 | Homo sapiens cyclin and CBS domain divalent metal cation transport mediator 2 (CNNM2), transcript variant 3, mRNA [NM_199077] |
| A_33_P3271171 | 0,009612307 | down | -1,5576261 | CNOT3 | CCR4-NOT transcription complex, subunit 3 [Source:HGNC Symbol;Acc:HGNC:7879] [ENST00000447684] |
| A_24_P941922 | 7,48E-05 | up | 2,131063 | CNOT4 | CCR4-NOT transcription complex, subunit 4 [Source:HGNC Symbol;Acc:HGNC:7880] [ENST00000428680] |
| A_23_P110846 | 7,83E-04 | up | 1,5898323 | CNOT8 | Homo sapiens CCR4-NOT transcription complex, subunit 8 (CNOT8), transcript variant 1, mRNA [NM_004779] |
| A_23_P310931 | 3,80E-04 | down | -2,771332 | CNR2 | Homo sapiens cannabinoid receptor 2 (macrophage) (CNR2), mRNA [NM_001841] |
| A_33_P3315320 | 0,004322523 | up | 1,7382574 | CNTD1 | Homo sapiens cyclin N-terminal domain containing 1 (CNTD1), mRNA [NM_173478] |
| A_33_P3409090 | 7,06E-04 | down | -2,982998 | CNTN1 | Homo sapiens contactin 1 (CNTN1), transcript variant 4, mRNA [NM_001256064] |
| A_23_P390700 | 0,007623092 | down | -2,287456 | CNTN1 | Homo sapiens contactin 1 (CNTN1), transcript variant 1, mRNA [NM_001843] |
| A_24_P114142 | 0,006959795 | down | -2,3191767 | CNTN2 | Homo sapiens contactin 2 (axonal) (CNTN2), mRNA [NM_005076] |
| A_23_P503059 | 0,007743829 | up | 1,9012928 | CNTN5 | Homo sapiens contactin 5 (CNTN5), transcript variant 1, mRNA [NM_014361] |
| A_33_P3329378 | 0,004564306 | down | -2,433023 | CNTN6 | Homo sapiens contactin 6 (CNTN6), transcript variant 1, mRNA [NM_014461] |
| A_21_P0013630 | 2,96E-05 | down | -2,12658 | CNTNAP3 | Homo sapiens contactin associated protein-like 3 (CNTNAP3), mRNA [NM_033655] |
| A_23_P56787 | 0,022395354 | down | -2,1013687 | CNTNAP5 | Homo sapiens contactin associated protein-like 5 (CNTNAP5), mRNA [NM_130773] |
| A_33_P3287379 | 0,003205908 | down | -1,6007304 | COG8 | Homo sapiens component of oligomeric golgi complex 8 (COG8), mRNA [NM_032382] |
| A_23_P214144 | 0,002074588 | down | -1,833848 | COL10A1 | Homo sapiens collagen, type X, alpha 1 (COL10A1), mRNA [NM_000493] |
| A_33_P3216448 | 0,004516994 | down | -2,815546 | COL11A2 | Homo sapiens collagen, type XI, alpha 2 (COL11A2), transcript variant 4, mRNA [NM_001163771] |
| A_32_P80850 | 0,003720608 | down | -3,8221629 | COL14A1 | Homo sapiens collagen, type XIV, alpha 1 (COL14A1), mRNA [NM_021110] |
| A_33_P3395952 | 0,027361266 | down | -1,675836 | COL20A1 | Homo sapiens collagen, type XX, alpha 1 (COL20A1), mRNA [NM_020882] |
| A_23_P31124 | 0,013298092 | down | -5,7974954 | COL21A1 | Homo sapiens collagen, type XXI, alpha 1 (COL21A1), mRNA [NM_030820] |
| A_23_P134627 | 5,91E-05 | down | -3,6059496 | COL26A1 | Homo sapiens collagen, type XXVI, alpha 1 (COL26A1), transcript variant 2, mRNA [NM_133457] |
| A_33_P3398236 | 0,022317115 | down | -2,4709964 | COL28A1 | Homo sapiens partial mRNA for collagen, type XXVIII (COL28 gene), isoform 2. [AJ890452] |
| A_33_P3262969 | 0,00637459 | down | -2,0853295 | COL4A6 | collagen, type IV, alpha 6 [Source:HGNC Symbol;Acc:HGNC:2208] [ENST00000461897] |
| A_23_P217379 | 0,016557684 | up | 1,8328223 | COL4A6 | Homo sapiens collagen, type IV, alpha 6 (COL4A6), transcript variant B, mRNA [NM_033641] |
| A_21_P0012396 | 3,78E-04 | down | -3,3985467 | COL6A4P2 | Homo sapiens collagen, type VI, alpha 4 pseudogene 2 (COL6A4P2), non-coding RNA [NR_027898] |
| A_33_P3311611 | 0,001709913 | up | 1,9748062 | COL6A4P2 | Homo sapiens collagen, type VI, alpha 4 pseudogene 2 (COL6A4P2), non-coding RNA [NR_027898] |
| A_24_P311856 | 0,009996332 | down | -4,2215304 | COL9A1 | Homo sapiens collagen, type IX, alpha 1 (COL9A1), transcript variant 1, mRNA [NM_001851] |
| A_23_P45786 | 0,010780422 | down | -1,8327225 | COL9A2 | Homo sapiens collagen, type IX, alpha 2 (COL9A2), mRNA [NM_001852] |
| A_33_P3294459 | 0,00420817 | up | 2,0978785 | COPG2IT1 | Homo sapiens cDNA FLJ41646 fis, clone FEBRA2024019. [AK123640] |
| A_23_P79725 | 0,02389704 | up | 1,5501678 | COPS7B | Homo sapiens COP9 signalosome subunit 7B (COPS7B), transcript variant 2, mRNA [NM_022730] |
| A_23_P101093 | 6,34E-04 | down | -2,065991 | COPZ2 | Homo sapiens coatomer protein complex, subunit zeta 2 (COPZ2), mRNA [NM_016429] |
| A_23_P117694 | 0,001165197 | down | -1,8303889 | CORO2B | Homo sapiens coronin, actin binding protein, 2B (CORO2B), transcript variant 1, mRNA [NM_006091] |
| A_23_P258164 | 3,44E-04 | down | -2,002615 | CORT | Homo sapiens cortistatin (CORT), mRNA [NM_001302] |
| A_23_P55123 | 0,00783863 | up | 1,6106086 | COX10 | Homo sapiens COX10 heme A:farnesyltransferase cytochrome c oxidase assembly factor (COX10), mRNA [NM_001303] |
| A_33_P3289113 | 0,03914508 | down | -1,5612668 | COX11 | cytochrome c oxidase assembly homolog 11 (yeast) [Source:HGNC Symbol;Acc:HGNC:2261] [ENST00000572088] |
| A_33_P3279610 | 1,13E-05 | down | -7,1665487 | COX6A2 | Homo sapiens cytochrome c oxidase subunit VIa polypeptide 2 (COX6A2), mRNA [NM_005205] |
| A_23_P78571 | 0,001849674 | down | -2,2612944 | COX6B2 | Homo sapiens cytochrome c oxidase subunit VIb polypeptide 2 (testis) (COX6B2), mRNA [NM_144613] |
| A_23_P136246 | 0,010888245 | up | 2,2285357 | COX7B2 | Homo sapiens cytochrome c oxidase subunit VIIb2 (COX7B2), mRNA [NM_130902] |
| A_23_P18017 | 0,03582283 | down | -1,8536375 | CPA3 | Homo sapiens carboxypeptidase A3 (mast cell) (CPA3), mRNA [NM_001870] |
| A_23_P19754 | 9,44E-04 | up | 3,1427877 | CPA4 | Homo sapiens carboxypeptidase A4 (CPA4), transcript variant 1, mRNA [NM_016352] |
| A_24_P53282 | 0,003089754 | up | 1,6997094 | CPD | Homo sapiens carboxypeptidase D (CPD), transcript variant 1, mRNA [NM_001304] |
| A_33_P3284448 | 0,003018205 | up | 2,572973 | CPEB2-AS1 | Homo sapiens CPEB2 antisense RNA 1 (head to head) (CPEB2-AS1), long non-coding RNA [NR_038857] |
| A_24_P51909 | 0,013993435 | down | -1,9890718 | CPLX1 | Homo sapiens complexin 1 (CPLX1), mRNA [NM_006651] |
| A_33_P3288135 | 0,037153814 | down | -1,6413679 | CPLX2 | complexin 2 [Source:HGNC Symbol;Acc:HGNC:2310] [ENST00000506642] |
| A_33_P3415300 | 2,02E-06 | down | -4,6049824 | CPLX2 | Homo sapiens complexin 2 (CPLX2), transcript variant 1, mRNA [NM_006650] |
| A_24_P391868 | 0,021307105 | up | 2,8690038 | CPLX2 | Homo sapiens complexin 2 (CPLX2), transcript variant 1, mRNA [NM_006650] |
| A_23_P98147 | 0,011320182 | down | -1,5981624 | CPN1 | Homo sapiens carboxypeptidase N, polypeptide 1 (CPN1), mRNA [NM_001308] |
| A_24_P137461 | 0,00256872 | down | -2,7807517 | CPN2 | Homo sapiens carboxypeptidase N, polypeptide 2 (CPN2), transcript variant 1, mRNA [NM_001080513] |
| A_33_P3334688 | 0,020761188 | up | 1,9068142 | CPNE2 | copine II [Source:HGNC Symbol;Acc:HGNC:2315] [ENST00000565951] |

| | | | | | |
|---|---|---|---|---|---|
| A_23_P360804 | 0,003789434 | down | -2,440039 | CPNE5 | Homo sapiens copine V (CPNE5), mRNA [NM_020939] |
| A_33_P3262191 | 0,001115283 | up | 2,401006 | CPNE7 | Homo sapiens copine VII (CPNE7), transcript variant 2, mRNA [NM_014427] |
| A_24_P56240 | 0,018007193 | up | 1,6318351 | CPNE8 | Homo sapiens copine VIII (CPNE8), mRNA [NM_153634] |
| A_23_P16694 | 0,018302524 | down | -1,5629363 | CPS1-IT1 | Homo sapiens CPS1 intronic transcript 1 (non-protein coding) (CPS1-IT1), long non-coding RNA [NR_002763] |
| A_24_P225518 | 0,03812364 | down | -1,5799567 | CPTP | Homo sapiens ceramide-1-phosphate transfer protein (CPTP), mRNA [NM_001029885] |
| A_23_P6066 | 3,63E-05 | down | -3,599711 | CPXM1 | Homo sapiens carboxypeptidase X (M14 family), member 1 (CPXM1), transcript variant 1, mRNA [NM_019609] |
| A_23_P256821 | 0,019834466 | up | 1,7066884 | CR1 | Homo sapiens complement component (3b/4b) receptor 1 (Knops blood group) (CR1), transcript variant S, mRNA [NM_000651] |
| A_33_P3303697 | 0,004147958 | up | 1,5217868 | CR2 | Homo sapiens complement component (3d/Epstein Barr virus) receptor 2 (CR2), transcript variant 1, mRNA [NM_001006658] |
| A_33_P3314151 | 0,001233501 | down | -2,0131567 | CRACR2A | Homo sapiens calcium release activated channel regulator 2A (CRACR2A), transcript variant 1, mRNA [NM_001144958] |
| A_23_P432545 | 0,002512852 | down | -2,0230112 | CRACR2B | Homo sapiens calcium release activated channel regulator 2B (CRACR2B), transcript variant 2, mRNA [NM_173584] |
| A_23_P12357 | 0,002369137 | up | 1,6480929 | CRB1 | Homo sapiens crumbs family member 1, photoreceptor morphogenesis associated (CRB1), transcript variant 1, mRNA [NM_201253] |
| A_24_P916656 | 1,84E-06 | up | 3,7066298 | CRCP | Homo sapiens CGRP receptor component (CRCP), transcript variant 1, mRNA [NM_014478] |
| A_21_P0013318 | 0,01183599 | up | 1,5585951 | CRCP | Homo sapiens CGRP receptor component (CRCP), transcript variant 4, mRNA [NM_001142414] |
| A_23_P79231 | 8,34E-05 | up | 2,0454664 | CREB1 | Homo sapiens cAMP responsive element binding protein 1 (CREB1), transcript variant B, mRNA [NM_134442] |
| A_23_P150407 | 0,004115873 | down | -1,6781266 | CREB3L1 | Homo sapiens cAMP responsive element binding protein 3-like 1 (CREB3L1), mRNA [NM_052854] |
| A_23_P108082 | 0,035483632 | up | 2,2954984 | CREB3L3 | Homo sapiens cAMP responsive element binding protein 3-like 3 (CREB3L3), transcript variant 1, mRNA [NM_032607] |
| A_33_P3240229 | 0,002460218 | up | 1,505764 | CREBBP | Homo sapiens CREB binding protein (CREBBP), transcript variant 1, mRNA [NM_004380] |
| A_23_P404606 | 0,0161229 | up | 1,5361679 | CREBRF | Homo sapiens CREB3 regulatory factor (CREBRF), transcript variant 1, mRNA [NM_153607] |
| A_23_P203645 | 6,03E-04 | up | 1,7799864 | CREBZF | Homo sapiens CREB/ATF bZIP transcription factor (CREBZF), transcript variant 1, mRNA [NM_001039618] |
| A_23_P33465 | 0,02989304 | down | -1,9775258 | CRELD2 | Homo sapiens cysteine-rich with EGF-like domains 2 (CRELD2), transcript variant 2, mRNA [NM_024324] |
| A_21_P0004513 | 0,010518785 | up | 2,5552926 | CRHBP | corticotropin releasing hormone binding protein [Source:HGNC Symbol;Acc:HGNC:2356] [ENST00000514258] |
| A_33_P3354965 | 1,41E-05 | down | -4,3568997 | CRIP1 | cysteine-rich protein 1 (intestinal) [Source:HGNC Symbol;Acc:HGNC:2360] [ENST00000330233] |
| A_23_P44674 | 0,00833867 | down | -1,5923656 | CRIP1 | Homo sapiens cysteine-rich protein 1 (intestinal) (CRIP1), mRNA [NM_001311] |
| A_33_P3325404 | 1,25E-04 | down | -3,2360277 | CRIP3 | Homo sapiens cysteine-rich protein 3 (CRIP3), mRNA [NM_206922] |
| A_23_P133722 | 0,012978573 | up | 1,578372 | CRISP1 | Homo sapiens cysteine-rich secretory protein 1 (CRISP1), transcript variant 1, mRNA [NM_001131] |
| A_33_P3359943 | 0,002260854 | up | 1,7673469 | CRISP2 | cysteine-rich secretory protein 2 [Source:HGNC Symbol;Acc:HGNC:12024] [ENST00000618917] |
| A_24_P270814 | 0,001086293 | up | 1,7294685 | CRK | Homo sapiens v-crk avian sarcoma virus CT10 oncogene homolog (CRK), transcript variant II, mRNA [NM_016823] |
| A_33_P3298139 | 2,22E-04 | down | -3,0805817 | CRLF2 | Homo sapiens cytokine receptor-like factor 2 (CRLF2), transcript variant 1, mRNA [NM_022148] |
| A_23_P113237 | 2,37E-06 | down | -2,9314392 | CRLF2 | Homo sapiens cytokine receptor-like factor 2 (CRLF2), transcript variant 2, mRNA [NM_001012288] |
| A_32_P149546 | 0,03569096 | up | 1,6532187 | CROCC | Homo sapiens ciliary rootlet coiled-coil, rootletin (CROCC), mRNA [NM_014675] |
| A_32_P384562 | 0,029093489 | up | 2,2857208 | CROCCP3 | Homo sapiens ciliary rootlet coiled-coil, rootletin pseudogene 3 (CROCCP3), non-coding RNA [NR_023386] |
| A_33_P3331882 | 0,005035898 | down | -1,8579845 | CRTC1 | Homo sapiens CREB regulated transcription coactivator 1 (CRTC1), transcript variant 3, mRNA [NM_001098482] |
| A_33_P3420204 | 0,024275467 | up | 1,5119295 | CRTC1 | Homo sapiens CREB regulated transcription coactivator 1 (CRTC1), transcript variant 3, mRNA [NM_001098482] |
| A_33_P3370707 | 0,022243343 | down | -2,0506284 | CRX | Homo sapiens cone-rod homeobox (CRX), mRNA [NM_000554] |
| A_24_P407235 | 0,00270968 | up | 1,7429675 | CRY1 | Homo sapiens cryptochrome circadian clock 1 (CRY1), mRNA [NM_004075] |
| A_23_P132147 | 0,021310885 | up | 1,6263691 | CRYAA | Homo sapiens crystallin, alpha A (CRYAA), mRNA [NM_000394] |
| A_23_P4254 | 0,0174314 | up | 2,266853 | CRYBA1 | Homo sapiens crystallin, beta A1 (CRYBA1), mRNA [NM_005208] |
| A_23_P103034 | 0,043820303 | up | 1,510905 | CRYBA4 | Homo sapiens crystallin, beta A4 (CRYBA4), mRNA [NM_001886] |
| A_19_P00321202 | 0,006751498 | down | -1,5842283 | CRYBG3 | Homo sapiens beta-gamma crystallin domain containing 3 (CRYBG3), mRNA [NM_153605] |
| A_23_P165380 | 0,002211608 | down | -2,2930126 | CRYGA | Homo sapiens crystallin, gamma A (CRYGA), mRNA [NM_014617] |
| A_23_P414842 | 0,004491135 | up | 1,5110959 | CRYGB | Homo sapiens crystallin, gamma B (CRYGB), mRNA [NM_005210] |
| A_33_P3461948 | 0,003799858 | down | -3,7659984 | CRYGN | Homo sapiens crystallin, gamma N (CRYGN), mRNA [NM_144727] |
| A_23_P132738 | 0,041580934 | up | 1,5530294 | CRYGS | Homo sapiens crystallin, gamma S (CRYGS), mRNA [NM_017541] |
| A_33_P3354940 | 0,018383762 | down | -1,6621056 | CSF1 | Homo sapiens colony stimulating factor 1 (macrophage) (CSF1), transcript variant 1, mRNA [NM_000757] |
| A_33_P3222424 | 0,016774347 | up | 1,8983201 | CSF3 | Homo sapiens colony stimulating factor 3 (granulocyte) (CSF3), transcript variant 1, mRNA [NM_000759] |
| A_23_P381351 | 0,03866832 | down | -1,9257097 | CSH1 | Homo sapiens chorionic somatomammotropin hormone 1 (placental lactogen) (CSH1), mRNA [NM_001317] |
| A_23_P207174 | 9,76E-05 | down | -1,8567106 | CSH2 | Homo sapiens chorionic somatomammotropin hormone 2 (CSH2), transcript variant 2, mRNA [NM_022644] |
| A_33_P3230239 | 0,002314063 | up | 1,7356757 | CSMD1 | Homo sapiens CUB and Sushi multiple domains 1 (CSMD1), mRNA [NM_033225] |
| A_23_P110234 | 0,00987252 | down | -1,6113873 | CSN1S1 | Homo sapiens casein alpha s1 (CSN1S1), transcript variant 1, mRNA [NM_001890] |
| A_23_P252882 | 0,004350505 | up | 2,8656228 | CSN3 | Homo sapiens casein kappa (CSN3), mRNA [NM_005212] |
| A_24_P338788 | 9,53E-04 | down | -3,3007724 | CSNK1A1L | Homo sapiens casein kinase 1, alpha 1-like (CSNK1A1L), mRNA [NM_145203] |
| A_33_P3324765 | 0,003004634 | up | 1,669857 | CSNK1D | Homo sapiens casein kinase 1, delta (CSNK1D), transcript variant 1, mRNA [NM_001893] |
| A_24_P234792 | 5,73E-04 | up | 1,6014132 | CSNK1G3 | Homo sapiens casein kinase 1, gamma 3 (CSNK1G3), transcript variant 4, mRNA [NM_001044723] |
| A_23_P502575 | 9,75E-04 | down | -1,7419225 | CSNK2A1 | Homo sapiens casein kinase 2, alpha 1 polypeptide (CSNK2A1), transcript variant 1, mRNA [NM_177559] |
| A_21_P0011330 | 2,67E-04 | down | -2,354427 | CSPG4 | Homo sapiens chondroitin sulfate proteoglycan 4 (CSPG4), mRNA [NM_001897] |
| A_23_P71537 | 3,15E-04 | up | 2,0833828 | CSPP1 | Homo sapiens centrosome and spindle pole associated protein 1 (CSPP1), transcript variant 2, mRNA [NM_001291339] |
| A_23_P121011 | 0,010259474 | up | 1,9384537 | CSRNP1 | Homo sapiens cysteine-serine-rich nuclear protein 1 (CSRNP1), mRNA [NM_033027] |
| A_33_P3224070 | 0,042454362 | up | 1,6007551 | CSRNP1 | Homo sapiens cysteine-serine-rich nuclear protein 1 (CSRNP1), mRNA [NM_033027] |
| A_23_P151209 | 0,008302202 | up | 1,5274146 | CSRNP2 | Homo sapiens cysteine-serine-rich nuclear protein 2 (CSRNP2), transcript variant 1, mRNA [NM_030809] |
| A_23_P143365 | 0,004069459 | up | 1,5013725 | CST9L | Homo sapiens cystatin 9-like (CST9L), mRNA [NM_080610] |
| A_33_P3311621 | 0,011371057 | down | -2,1211188 | CSTF3-AS1 | Homo sapiens CSTF3 antisense RNA 1 (head to head) (CSTF3-AS1), long non-coding RNA [NR_034027] |

| | | | | | |
|---|---|---|---|---|---|
| A_23_P210807 | 9,77E-04 | down | -2,4248476 | CSTL1 | Homo sapiens cystatin-like 1 (CSTL1), mRNA [NM_138283] |
| A_32_P207124 | 1,67E-04 | down | -2,0936832 | CT47A11 | Homo sapiens cancer/testis antigen family 47, member A11 (CT47A11), mRNA [NM_173571] |
| A_33_P3247165 | 0,012858363 | up | 2,6368213 | CT62 | Homo sapiens cancer/testis antigen 62 (CT62), mRNA [NM_001102658] |
| A_33_P3318343 | 2,80E-05 | down | -3,1020103 | CTAG2 | Homo sapiens cancer/testis antigen 2 (CTAG2), transcript variant 2, mRNA [NM_020994] |
| A_23_P402952 | 0,038284265 | up | 1,5308574 | CTAGE15 | Homo sapiens CTAGE family, member 15 (CTAGE15), mRNA [NM_001008747] |
| A_23_P41292 | 4,19E-04 | up | 1,6454359 | CTBP1 | Homo sapiens C-terminal binding protein 1 (CTBP1), transcript variant 2, mRNA [NM_001012614] |
| A_19_P00319655 | 0,009483689 | down | -2,103435 | CTD-3080P12.3 | Homo sapiens uncharacterized LOC101928857 (CTD-3080P12.3), long non-coding RNA [NR_109911] |
| A_19_P00318243 | 0,001164722 | down | -1,6152066 | CTD-3080P12.3 | Homo sapiens uncharacterized LOC101928857 (CTD-3080P12.3), long non-coding RNA [NR_109911] |
| A_19_P00317835 | 9,74E-04 | down | -2,428418 | CTD-3080P12.3 | Homo sapiens uncharacterized LOC101928857 (CTD-3080P12.3), long non-coding RNA [NR_109911] |
| A_24_P142473 | 0,006302226 | up | 1,5723447 | CTDNEP1 | Homo sapiens CTD nuclear envelope phosphatase 1 (CTDNEP1), transcript variant 1, mRNA [NM_015343] |
| A_23_P110624 | 1,40E-05 | down | -2,704117 | CTNND2 | Homo sapiens catenin (cadherin-associated protein), delta 2 (CTNND2), transcript variant 1, mRNA [NM_001332] |
| A_33_P3254331 | 0,029978069 | up | 1,7602673 | CTRC | Homo sapiens chymotrypsin C (caldecrin) (CTRC), mRNA [NM_007272] |
| A_23_P10291 | 4,25E-05 | down | -2,1754255 | CTSE | Homo sapiens cathepsin E (CTSE), transcript variant 1, mRNA [NM_001910] |
| A_23_P140384 | 0,006731194 | down | -2,4162056 | CTSG | Homo sapiens cathepsin G (CTSG), mRNA [NM_001911] |
| A_23_P40240 | 0,007260038 | up | 2,5232852 | CTSZ | Homo sapiens cathepsin Z (CTSZ), mRNA [NM_001336] |
| A_33_P3219939 | 0,029734585 | up | 1,6534452 | CUBN | Homo sapiens cubilin (intrinsic factor-cobalamin receptor) (CUBN), mRNA [NM_001081] |
| A_24_P252794 | 0,006025991 | up | 1,5586597 | CUL2 | Homo sapiens cullin 2 (CUL2), transcript variant 3, mRNA [NM_003591] |
| A_23_P148463 | 0,001255646 | up | 1,758439 | CUL4B | Homo sapiens cullin 4B (CUL4B), transcript variant 1, mRNA [NM_003588] |
| A_24_P220454 | 0,002197961 | up | 1,6262196 | CUX1 | Homo sapiens cut-like homeobox 1 (CUX1), transcript variant 1, mRNA [NM_181552] |
| A_23_P2543 | 0,013578005 | up | 4,612537 | CUX2 | Homo sapiens cut-like homeobox 2 (CUX2), mRNA [NM_015267] |
| A_24_P374943 | 0,004246139 | down | -1,5352498 | CXADR | Homo sapiens coxsackie virus and adenovirus receptor (CXADR), transcript variant 1, mRNA [NM_001338] |
| A_33_P3330264 | 0,017882597 | down | -1,9021653 | CXCL1 | Homo sapiens chemokine (C-X-C motif) ligand 1 (melanoma growth stimulating activity, alpha) (CXCL1), transcript variant 1, mRNA [NM_001511] |
| A_23_P202448 | 0,012672961 | down | -1,5311905 | CXCL12 | Homo sapiens chemokine (C-X-C motif) ligand 12 (CXCL12), transcript variant 1, mRNA [NM_199168] |
| A_33_P3712341 | 0,004769301 | down | -2,1816678 | CXCL12 | Homo sapiens chemokine (C-X-C motif) ligand 12 (CXCL12), transcript variant 3, mRNA [NM_001033886] |
| A_33_P3375859 | 0,007276913 | up | 1,5187298 | CXCR2P1 | Homo sapiens chemokine (C-X-C motif) receptor 2 pseudogene 1 (CXCR2P1), non-coding RNA [NR_002712] |
| A_23_P114299 | 0,024957158 | down | -1,5800143 | CXCR3 | Homo sapiens chemokine (C-X-C motif) receptor 3 (CXCR3), transcript variant 1, mRNA [NM_001504] |
| A_23_P102000 | 0,006605382 | up | 2,2860897 | CXCR4 | Homo sapiens chemokine (C-X-C motif) receptor 4 (CXCR4), transcript variant 1, mRNA [NM_001008540] |
| A_23_P109913 | 0,017001104 | up | 1,6446344 | CXCR6 | Homo sapiens chemokine (C-X-C motif) receptor 6 (CXCR6), mRNA [NM_006564] |
| A_33_P3293925 | 4,97E-04 | down | -2,2571478 | CXorf36 | Homo sapiens chromosome X open reading frame 36 (CXorf36), transcript variant 1, mRNA [NM_176819] |
| A_24_P36745 | 0,004067759 | up | 1,6072811 | CXorf38 | Homo sapiens chromosome X open reading frame 38 (CXorf38), mRNA [NM_144970] |
| A_33_P3650224 | 0,008596472 | down | -1,6693398 | CXorf49B | Homo sapiens chromosome X open reading frame 49B (CXorf49B), mRNA [NM_001145139] |
| A_33_P3209170 | 0,005274218 | up | 1,5505161 | CXorf51A | Homo sapiens chromosome X open reading frame 51A (CXorf51A), mRNA [NM_001144064] |
| A_23_P413157 | 0,030987509 | up | 1,6192452 | CXorf58 | Homo sapiens chromosome X open reading frame 58 (CXorf58), transcript variant 1, mRNA [NM_152761] |
| A_33_P3255324 | 0,017143449 | up | 1,5634949 | CXXC1P1 | Homo sapiens CXXC finger protein 1 pseudogene 1 (CXXC1P1), non-coding RNA [NR_033924] |
| A_33_P3388564 | 0,002705569 | down | -1,6247121 | CYB5D1 | Homo sapiens cytochrome b5 domain containing 1 (CYB5D1), mRNA [NM_144607] |
| A_33_P3316686 | 1,49E-04 | down | -1,8561141 | CYB5R4 | Homo sapiens cytochrome b5 reductase 4 (CYB5R4), mRNA [NM_016230] |
| A_24_P365767 | 0,03592154 | down | -1,7528096 | CYBB | Homo sapiens cytochrome b-245, beta polypeptide (CYBB), mRNA [NM_000397] |
| A_23_P167509 | 0,021687586 | down | -2,0427537 | CYFIP2 | Homo sapiens cytoplasmic FMR1 interacting protein 2 (CYFIP2), transcript variant 3, mRNA [NM_014376] |
| A_33_P3285868 | 1,40E-05 | down | -3,9301143 | CYGB | Homo sapiens cytoglobin (CYGB), mRNA [NM_134268] |
| A_24_P158285 | 0,046856362 | up | 2,0379727 | CYLC2 | cylicin, basic protein of sperm head cytoskeleton 2 [Source:HGNC Symbol;Acc:HGNC:2583] [ENST00000612124] |
| A_23_P129169 | 0,0026079 | down | -2,9085817 | CYP11A1 | Homo sapiens cytochrome P450, family 11, subfamily A, polypeptide 1 (CYP11A1), transcript variant 1, mRNA [NM_000781] |
| A_23_P215997 | 0,048540805 | up | 1,7009579 | CYP11B2 | Homo sapiens cytochrome P450, family 11, subfamily B, polypeptide 2 (CYP11B2), mRNA [NM_000498] |
| A_33_P3376478 | 0,007228423 | down | -2,1758037 | CYP17A1 | Homo sapiens cytochrome P450, family 17, subfamily A, polypeptide 1 (CYP17A1), mRNA [NM_000102] |
| A_19_P00316135 | 0,007894971 | up | 1,643985 | CYP1B1-AS1 | Homo sapiens CYP1B1 antisense RNA 1 (CYP1B1-AS1), long non-coding RNA [NR_027252] |
| A_33_P3411279 | 0,014393191 | down | -1,7415445 | CYP21A2 | Homo sapiens cytochrome P450, family 21, subfamily A, polypeptide 2 (CYP21A2), transcript variant 2, mRNA [NM_001128590] |
| A_23_P210109 | 0,014521415 | up | 1,878929 | CYP26B1 | Homo sapiens cytochrome P450, family 26, subfamily B, polypeptide 1 (CYP26B1), transcript variant 1, mRNA [NM_019885] |
| A_33_P3264667 | 1,72E-04 | down | -3,784117 | CYP26C1 | Homo sapiens cytochrome P450, family 26, subfamily C, polypeptide 1 (CYP26C1), mRNA [NM_183374] |
| A_23_P36397 | 0,012570703 | up | 2,182729 | CYP27B1 | Homo sapiens cytochrome P450, family 27, subfamily B, polypeptide 1 (CYP27B1), mRNA [NM_000785] |
| A_23_P55779 | 0,009765516 | down | -1,7268119 | CYP2A13 | Homo sapiens cytochrome P450, family 2, subfamily A, polypeptide 13 (CYP2A13), mRNA [NM_000766] |
| A_23_P208373 | 0,003408433 | down | -1,9334961 | CYP2B6 | Homo sapiens cytochrome P450, family 2, subfamily B, polypeptide 6 (CYP2B6), mRNA [NM_000767] |
| A_24_P394940 | 0,030274356 | up | 2,3021047 | CYP2E1 | Homo sapiens cytochrome P450, family 2, subfamily E, polypeptide 1 (CYP2E1), mRNA [NM_000773] |
| A_21_P0014273 | 0,039196245 | down | -1,6374826 | CYP2R1 | cytochrome P450, family 2, subfamily R, polypeptide 1 [Source:HGNC Symbol;Acc:HGNC:20580] [ENST00000532805] |
| A_24_P191013 | 5,27E-04 | down | -2,3335712 | CYP4A11 | Homo sapiens cytochrome P450, family 4, subfamily A, polypeptide 11 (CYP4A11), mRNA [NM_000778] |
| A_33_P3303474 | 6,19E-04 | down | -2,9305618 | CYP4A11 | Homo sapiens cytochrome P450, family 4, subfamily A, polypeptide 11 (CYP4A11), mRNA [NM_000778] |
| A_33_P3382471 | 0,04377312 | down | -1,8437686 | CYP4A22 | Homo sapiens cytochrome P450, family 4, subfamily A, polypeptide 22 (CYP4A22), mRNA [NM_001010969] |
| A_21_P0000967 | 0,01111107 | up | 1,7298989 | CYP4B1 | cytochrome P450, family 4, subfamily B, polypeptide 1 [Source:HGNC Symbol;Acc:HGNC:2644] [ENST00000546128] |
| A_21_P0011168 | 0,013058359 | down | -2,217668 | CYP4F35P | Homo sapiens cytochrome P450, family 4, subfamily F, polypeptide 35, pseudogene (CYP4F35P), non-coding RNA [NR_026756] |
| A_24_P28811 | 0,004201054 | down | -2,5955667 | CYP4F62P | Homo sapiens cytochrome P450, family 4, subfamily F, polypeptide 62, pseudogene (CYP4F62P), non-coding RNA [NR_103761] |
| A_23_P131060 | 3,88E-04 | down | -2,656675 | CYP4F8 | Homo sapiens cytochrome P450, family 4, subfamily F, polypeptide 8 (CYP4F8), mRNA [NM_007253] |
| A_24_P293530 | 0,026695231 | up | 2,3638124 | CYP4X1 | Homo sapiens cytochrome P450, family 4, subfamily X, polypeptide 1 (CYP4X1), mRNA [NM_178033] |
| A_21_P0010756 | 0,0263843 | up | 2,1172006 | CYP4Z1 | Homo sapiens cytochrome P450, family 4, subfamily Z, polypeptide 1 (CYP4Z1), mRNA [NM_178134] |

| | | | | | |
|---|---|---|---|---|---|
| A_33_P3873533 | 0,005243063 | up | 1,5229244 | CYP51A1-AS1 | Homo sapiens CYP51A1 antisense RNA 1 (CYP51A1-AS1), transcript variant 1, long non-coding RNA [NR_122109] |
| A_23_P146198 | 0,009408837 | down | -1,964436 | CYP7A1 | Homo sapiens cytochrome P450, family 7, subfamily A, polypeptide 1 (CYP7A1), mRNA [NM_000780] |
| A_23_P169092 | 0,03852742 | down | -1,9439952 | CYP7B1 | Homo sapiens cytochrome P450, family 7, subfamily B, polypeptide 1 (CYP7B1), mRNA [NM_004820] |
| A_24_P208704 | 0,004966459 | up | 1,5003439 | CYP8B1 | Homo sapiens cytochrome P450, family 8, subfamily B, polypeptide 1 (CYP8B1), mRNA [NM_004391] |
| A_23_P83781 | 6,53E-04 | up | 1,6800588 | CYTH1 | Homo sapiens cytohesin 1 (CYTH1), transcript variant 1, mRNA [NM_004762] |
| A_33_P3762733 | 9,65E-04 | down | -1,9852363 | D21S2088E | Homo sapiens D21S2088E (D21S2088E), long non-coding RNA [NR_040254] |
| A_21_P0010273 | 0,006046833 | down | -2,4261594 | D21S2088E | Homo sapiens D21S2088E (D21S2088E), long non-coding RNA [NR_040254] |
| A_33_P3355296 | 0,003476603 | down | -1,6510508 | D2HGDH | D-2-hydroxyglutarate dehydrogenase [Source:HGNC Symbol;Acc:HGNC:28358] [ENST00000467427] |
| A_33_P3288104 | 6,03E-06 | down | -3,9056752 | DAAM2 | dishevelled associated activator of morphogenesis 2 [Source:HGNC Symbol;Acc:HGNC:18143] [ENST00000405961] |
| A_32_P52785 | 7,74E-04 | down | -3,3082118 | DAAM2 | Homo sapiens dishevelled associated activator of morphogenesis 2 (DAAM2), transcript variant 2, mRNA [NM_015345] |
| A_23_P23850 | 0,001730926 | down | -2,1302145 | DAB1 | Homo sapiens Dab, reelin signal transducer, homolog 1 (Drosophila) (DAB1), mRNA [NM_021080] |
| A_33_P3375334 | 0,004435388 | up | 1,6764264 | DAB1 | Dab, reelin signal transducer, homolog 1 (Drosophila) [Source:HGNC Symbol;Acc:HGNC:2661] [ENST00000371230] |
| A_23_P123848 | 0,007713168 | up | 1,7693135 | DAB2IP | Homo sapiens DAB2 interacting protein (DAB2IP), transcript variant 1, mRNA [NM_032552] |
| A_24_P289260 | 0,005719409 | up | 2,305007 | DACT2 | Homo sapiens dishevelled-binding antagonist of beta-catenin 2 (DACT2), transcript variant 1, mRNA [NM_214462] |
| A_24_P225961 | 0,007524025 | up | 1,5787312 | DAG1 | Homo sapiens dystroglycan 1 (dystrophin-associated glycoprotein 1) (DAG1), transcript variant 2, mRNA [NM_004393] |
| A_24_P115967 | 0,002508643 | up | 1,8557249 | DAGLA | Homo sapiens diacylglycerol lipase, alpha (DAGLA), mRNA [NM_006133] |
| A_24_P902052 | 0,00250415 | up | 1,7481979 | DANCR | Homo sapiens differentiation antagonizing non-protein coding RNA (DANCR), long non-coding RNA [NR_024031] |
| A_23_P435636 | 0,006239211 | up | 1,8311049 | DAND5 | Homo sapiens DAN domain family member 5, BMP antagonist (DAND5), mRNA [NM_152654] |
| A_24_P10233 | 6,09E-04 | up | 3,2715988 | DAPK2 | Homo sapiens death-associated protein kinase 2 (DAPK2), mRNA [NM_014326] |
| A_23_P148984 | 5,67E-05 | up | 1,8133062 | DARS2 | Homo sapiens aspartyl-tRNA synthetase 2, mitochondrial (DARS2), mRNA [NM_018122] |
| A_23_P28466 | 0,033617917 | up | 2,3074393 | DAW1 | Homo sapiens dynein assembly factor with WDR repeat domains 1 (DAW1), mRNA [NM_178821] |
| A_33_P3682006 | 0,023167709 | down | -2,5146136 | DBH-AS1 | Homo sapiens DBH antisense RNA 1 (DBH-AS1), long non-coding RNA [NR_102735] |
| A_23_P351535 | 0,023917466 | up | 2,0434322 | DBIL5P | Homo sapiens diazepam binding inhibitor-like 5, pseudogene (DBIL5P), non-coding RNA [NR_024120] |
| A_33_P3291594 | 0,03220782 | up | 2,0969286 | DBT | dihydrolipoamide branched chain transacylase E2 [Source:HGNC Symbol;Acc:HGNC:2698] [ENST00000370131] |
| A_33_P3407350 | 0,001911914 | up | 4,610509 | DBX1 | Homo sapiens developing brain homeobox 1 (DBX1), mRNA [NM_001029865] |
| A_33_P3309999 | 0,002543547 | down | -4,4681644 | DCAF12L2 | Homo sapiens DDB1 and CUL4 associated factor 12-like 2 (DCAF12L2), mRNA [NM_001013628] |
| A_33_P3327290 | 0,016412854 | up | 1,576158 | DCAF4L1 | Homo sapiens DDB1 and CUL4 associated factor 4-like 1 (DCAF4L1), mRNA [NM_001029955] |
| A_33_P3768992 | 0,008012371 | down | -2,7903297 | DCAF4L2 | Homo sapiens DDB1 and CUL4 associated factor 4-like 2 (DCAF4L2), mRNA [NM_152418] |
| A_21_P0014056 | 0,008895797 | down | -2,3837156 | DCAF8 | DDB1 and CUL4 associated factor 8 [Source:HGNC Symbol;Acc:HGNC:24891] [ENST00000481831] |
| A_33_P3250740 | 0,023464588 | up | 2,0537393 | DCAF8L1 | Homo sapiens DDB1 and CUL4 associated factor 8-like 1 (DCAF8L1), mRNA [NM_001017930] |
| A_24_P58331 | 0,001359299 | down | -1,7749751 | DCAKD | Homo sapiens dephospho-CoA kinase domain containing (DCAKD), transcript variant 1, mRNA [NM_024819] |
| A_33_P3246028 | 0,04465046 | up | 2,2831223 | DCDC2B | Homo sapiens doublecortin domain containing 2B (DCDC2B), mRNA [NM_001099434] |
| A_33_P3298226 | 0,021359531 | up | 2,4176476 | DCHS2 | dachsous cadherin-related 2 [Source:HGNC Symbol;Acc:HGNC:23111] [ENST00000456341] |
| A_24_P54131 | 0,039073016 | up | 1,5488048 | DCLRE1B | Homo sapiens DNA cross-link repair 1B (DCLRE1B), mRNA [NM_022836] |
| A_24_P886040 | 2,93E-04 | up | 1,8939086 | DCP2 | Homo sapiens decapping mRNA 2 (DCP2), transcript variant 1, mRNA [NM_152624] |
| A_23_P314798 | 0,032032058 | down | -1,6891665 | DCST2 | Homo sapiens DC-STAMP domain containing 2 (DCST2), mRNA [NM_144622] |
| A_23_P76622 | 9,10E-04 | down | -5,0089407 | DCT | Homo sapiens dopachrome tautomerase (DCT), transcript variant 1, mRNA [NM_001922] |
| A_33_P3406686 | 0,014732851 | down | -1,7680238 | DCTN1 | Homo sapiens dynactin 1 (DCTN1), transcript variant 7, non-coding RNA [NR_033935] |
| A_32_P43812 | 0,0301917 | up | 1,5114195 | DCUN1D4 | Homo sapiens DCN1, defective in cullin neddylation 1, domain containing 4 (DCUN1D4), transcript variant 1, mRNA [NM_001040402] |
| A_33_P3259878 | 0,001668798 | down | -2,148791 | DDB1 | damage-specific DNA binding protein 1, 127kDa [Source:HGNC Symbol;Acc:HGNC:2717] [ENST00000545894] |
| A_33_P3283038 | 0,002054396 | up | 1,640494 | DDC-AS1 | Homo sapiens DDC antisense RNA 1 (DDC-AS1), long non-coding RNA [NR_033845] |
| A_21_P0000039 | 0,027385073 | up | 1,6010927 | DDHD2 | Homo sapiens DDHD domain containing 2 (DDHD2), transcript variant 3, mRNA [NM_001164234] |
| A_32_P93149 | 0,009067026 | down | -1,9642712 | DDX10 | Homo sapiens DEAD (Asp-Glu-Ala-Asp) box polypeptide 10 (DDX10), mRNA [NM_004398] |
| A_33_P3322307 | 0,04876158 | up | 1,6677231 | DDX11 | Homo sapiens DEAD/H (Asp-Glu-Ala-Asp/His) box helicase 11 (DDX11), transcript variant 5, mRNA [NM_001257145] |
| A_32_P227317 | 0,007571008 | down | -1,5109097 | DDX11L2 | Homo sapiens DEAD/H (Asp-Glu-Ala-Asp/His) box helicase 11 like 2 (DDX11L2), transcript variant 1, non-coding RNA [NR_024004] |
| A_23_P57534 | 0,02238735 | up | 1,6512882 | DDX17 | Homo sapiens DEAD (Asp-Glu-Ala-Asp) box helicase 17 (DDX17), transcript variant 1, mRNA [NM_006386] |
| A_24_P512054 | 0,025132263 | up | 2,2537367 | DDX50 | Homo sapiens DEAD (Asp-Glu-Ala-Asp) box polypeptide 50 (DDX50), mRNA [NM_024045] |
| A_33_P3372874 | 0,003122786 | up | 1,511794 | DDX53 | Homo sapiens DEAD (Asp-Glu-Ala-Asp) box polypeptide 53 (DDX53), mRNA [NM_182699] |
| A_33_P3461258 | 0,01619388 | up | 2,8916602 | DEFA11P | Homo sapiens defensin, alpha 11 pseudogene (DEFA11P), non-coding RNA [NR_073421] |
| A_23_P9018 | 9,17E-05 | down | -2,9044347 | DEFB104B | Homo sapiens defensin, beta 104B (DEFB104B), mRNA [NM_001040702] |
| A_33_P3265803 | 0,008213288 | down | -1,50702 | DEFB108B | Homo sapiens defensin, beta 108B (DEFB108B), mRNA [NM_001002035] |
| A_33_P3412678 | 0,001509245 | up | 2,1219666 | DEFB112 | Homo sapiens defensin, beta 112 (DEFB112), mRNA [NM_001037498] |
| A_33_P3328352 | 0,042822607 | up | 2,0151548 | DEFB115 | Homo sapiens defensin, beta 115 (DEFB115), mRNA [NM_001037730] |
| A_23_P312285 | 4,23E-04 | up | 2,5822425 | DEFB119 | Homo sapiens defensin, beta 119 (DEFB119), transcript variant 1, mRNA [NM_153289] |
| A_33_P3305446 | 0,028459717 | down | -1,8651847 | DEFB121 | Homo sapiens defensin, beta 121 (DEFB121), transcript variant 1, mRNA [NM_001011878] |
| A_33_P3316522 | 3,07E-04 | down | -2,4777787 | DEFB124 | Homo sapiens defensin, beta 124 (DEFB124), mRNA [NM_001037500] |
| A_33_P3272962 | 0,01496626 | down | -2,4502509 | DEFB128 | Homo sapiens defensin, beta 128 (DEFB128), mRNA [NM_001037732] |
| A_33_P3316475 | 3,44E-04 | down | -2,4996994 | DEFB132 | Homo sapiens defensin, beta 132 (DEFB132), mRNA [NM_207469] |
| A_23_P254702 | 0,016124837 | up | 1,5328605 | DEK | Homo sapiens DEK proto-oncogene (DEK), transcript variant 1, mRNA [NM_003472] |
| A_23_P320897 | 0,03877517 | up | 1,5515177 | DENND1B | Homo sapiens DENN/MADD domain containing 1B (DENND1B), transcript variant 3, mRNA [NM_001195215] |
| A_33_P3248629 | 0,017658008 | down | -1,7057351 | DENND2A | DENN/MADD domain containing 2A [Source:HGNC Symbol;Acc:HGNC:22212] [ENST00000492720] |

| | | | | | |
|---|---|---|---|---|---|
| A_33_P3321136 | 0,034672167 | down | -1,6051948 | DENND3 | Homo sapiens DENN/MADD domain containing 3 (DENND3), mRNA [NM_014957] |
| A_33_P3321130 | 0,011235802 | up | 2,3258991 | DENND3 | DENN/MADD domain containing 3 [Source:HGNC Symbol;Acc:HGNC:29134] [ENST00000520482] |
| A_32_P175301 | 0,04668635 | up | 1,9659216 | DENND3 | Homo sapiens DENN/MADD domain containing 3 (DENND3), mRNA [NM_014957] |
| A_23_P157726 | 0,00855777 | up | 1,5042038 | DENND4C | DENN/MADD domain containing 4C [Source:HGNC Symbol;Acc:HGNC:26079] [ENST00000380437] |
| A_24_P659036 | 1,89E-04 | down | -3,4460607 | DENND6B | Homo sapiens DENN/MADD domain containing 6B (DENND6B), mRNA [NM_001001794] |
| A_33_P3409672 | 0,005486531 | up | 1,7143492 | DEPDC5 | Homo sapiens DEP domain containing 5 (DEPDC5), transcript variant 4, mRNA [NM_001242896] |
| A_33_P3724155 | 0,001755328 | down | -1,7662646 | DERL3 | Homo sapiens derlin 3 (DERL3), transcript variant 3, mRNA [NM_198440] |
| A_33_P3275801 | 0,004862028 | down | -1,7054603 | DES | Homo sapiens desmin (DES), mRNA [NM_001927] |
| A_23_P44546 | 0,005835782 | up | 2,0363755 | DFFB | Homo sapiens DNA fragmentation factor, 40kDa, beta polypeptide (caspase-activated DNase) (DFFB), transcript variant 1, mRNA [NM_001282669] |
| A_24_P125881 | 0,002112263 | up | 1,5512819 | DGCR2 | Homo sapiens DiGeorge syndrome critical region gene 2 (DGCR2), transcript variant 1, mRNA [NM_005137] |
| A_21_P0012310 | 0,023877386 | down | -1,6884927 | DGCR5 | Homo sapiens DiGeorge syndrome critical region gene 5 (non-protein coding) (DGCR5), transcript variant 3, non-coding RNA [NR_110533] |
| A_33_P3263359 | 0,03161778 | down | -2,4003706 | DGKA | diacylglycerol kinase, alpha 80kDa [Source:HGNC Symbol;Acc:HGNC:2849] [ENST00000546995] |
| A_33_P3421728 | 0,001897034 | down | -3,2951567 | DGKG | Homo sapiens diacylglycerol kinase, gamma 90kDa (DGKG), transcript variant 1, mRNA [NM_001346] |
| A_24_P343095 | 0,021350173 | down | -1,8172582 | DHFR | Homo sapiens dihydrofolate reductase (DHFR), transcript variant 1, mRNA [NM_000791] |
| A_33_P3338740 | 0,013261414 | up | 2,067898 | DHH | Homo sapiens desert hedgehog (DHH), mRNA [NM_021044] |
| A_23_P33759 | 0,016533468 | up | 1,9122171 | DHRS3 | Homo sapiens dehydrogenase/reductase (SDR family) member 3 (DHRS3), mRNA [NM_004753] |
| A_33_P3403459 | 0,008537592 | down | -1,5253488 | DHRS4-AS1 | Homo sapiens DHRS4 antisense RNA 1 (DHRS4-AS1), transcript variant 1, long non-coding RNA [NR_023921] |
| A_23_P81463 | 0,006843912 | up | 1,5645705 | DHX29 | Homo sapiens DEAH (Asp-Glu-Ala-His) box polypeptide 29 (DHX29), mRNA [NM_019030] |
| A_23_P47004 | 0,002685192 | up | 1,641429 | DHX32 | Homo sapiens DEAH (Asp-Glu-Ala-His) box polypeptide 32 (DHX32), mRNA [NM_018180] |
| A_24_P364970 | 7,66E-04 | up | 1,8028526 | DHX33 | Homo sapiens DEAH (Asp-Glu-Ala-His) box polypeptide 33 (DHX33), transcript variant 1, mRNA [NM_020162] |
| A_23_P38011 | 0,007193404 | up | 1,6014931 | DHX38 | Homo sapiens DEAH (Asp-Glu-Ala-His) box polypeptide 38 (DHX38), mRNA [NM_014003] |
| A_21_P0011150 | 0,001096528 | up | 1,6416379 | DHX9 | Homo sapiens DEAH (Asp-Glu-Ala-His) box helicase 9 (DHX9), transcript variant 1, mRNA [NM_001357] |
| A_33_P3330125 | 0,001311651 | down | -2,7022662 | DIABLO | Homo sapiens diablo, IAP-binding mitochondrial protein (DIABLO), transcript variant 5, mRNA [NM_001278302] |
| A_33_P3243683 | 0,013784593 | up | 1,5663291 | DIAPH1 | Homo sapiens diaphanous-related formin 1 (DIAPH1), transcript variant 1, mRNA [NM_005219] |
| A_23_P395426 | 0,001265822 | up | 1,6745187 | DIDO1 | Homo sapiens death inducer-obliterator 1 (DIDO1), transcript variant 1, mRNA [NM_022105] |
| A_24_P944144 | 0,004435291 | up | 2,2270083 | DIDO1 | Homo sapiens death inducer-obliterator 1 (DIDO1), transcript variant 3, mRNA [NM_080797] |
| A_23_P34946 | 0,003886883 | up | 1,5197195 | DIEXF | Homo sapiens digestive organ expansion factor homolog (zebrafish) (DIEXF), mRNA [NM_014388] |
| A_33_P3325700 | 0,005312665 | up | 1,6405076 | DIO2 | Homo sapiens deiodinase, iodothyronine, type II (DIO2), transcript variant 4, mRNA [NM_001242502] |
| A_23_P401380 | 0,001001802 | up | 1,8645418 | DIP2B | Homo sapiens DIP2 disco-interacting protein 2 homolog B (Drosophila) (DIP2B), mRNA [NM_173602] |
| A_33_P3238484 | 0,049390856 | down | -2,3921447 | DIP2C | Homo sapiens DIP2 disco-interacting protein 2 homolog C (Drosophila) (DIP2C), mRNA [NM_014974] |
| A_23_P131139 | 0,003401689 | down | -2,455574 | DIRC1 | Homo sapiens disrupted in renal carcinoma 1 (DIRC1), mRNA [NM_052952] |
| A_33_P3234521 | 0,02290145 | down | -2,3625288 | DIS3L | DIS3 like exosome 3'-5' exoribonuclease [Source:HGNC Symbol;Acc:HGNC:28698] [ENST00000564909] |
| A_33_P3270203 | 5,77E-05 | down | -2,018802 | DIS3L2 | Homo sapiens DIS3 like 3'-5' exoribonuclease 2 (DIS3L2), transcript variant 2, mRNA [NM_001257281] |
| A_33_P3270192 | 0,0024941 | up | 1,5860902 | DIS3L2 | DIS3 like 3'-5' exoribonuclease 2 [Source:HGNC Symbol;Acc:HGNC:28648] [ENST00000409401] |
| A_33_P3400918 | 0,035515938 | down | -2,173556 | DISC1 | Homo sapiens disrupted in schizophrenia 1 (DISC1), transcript variant m, mRNA [NM_001164550] |
| A_24_P83787 | 0,001158951 | down | -1,9296458 | DISC1 | Homo sapiens disrupted in schizophrenia 1 (DISC1), transcript variant S, mRNA [NM_001012959] |
| A_21_P0000050 | 0,008982472 | up | 2,19026 | DISC1 | Homo sapiens disrupted in schizophrenia 1 (DISC1), transcript variant l, mRNA [NM_001164549] |
| A_21_P0000049 | 0,036681984 | up | 1,6988332 | DISC1 | Homo sapiens disrupted in schizophrenia 1 (DISC1), transcript variant b, mRNA [NM_001164538] |
| A_21_P0009654 | 0,008276031 | up | 1,5006328 | DKFZp434J0226 | uncharacterized LOC93429 [Source:EntrezGene;Acc:93429] [ENST00000593980] |
| A_21_P0005080 | 0,006843387 | up | 1,6353315 | DKFZp451B082 | Homo sapiens uncharacterized LOC401282 (DKFZp451B082), long non-coding RNA [NR_033862] |
| A_33_P3567892 | 0,012691454 | up | 2,2013688 | DKFZP547J0410 | Homo sapiens mRNA; cDNA DKFZp564J0482 (from clone DKFZp564J0482) [AL050263] |
| A_23_P403398 | 0,012210114 | up | 1,6544875 | DKFZP586I1420 | Homo sapiens uncharacterized protein DKFZp586I1420 (DKFZP586I1420), non-coding RNA [NR_002186] |
| A_21_P0014245 | 0,001610669 | up | 1,7190765 | DKFZp667F0711 | PREDICTED: Homo sapiens uncharacterized protein DKFZp667F0711 (DKFZp667F0711), misc_RNA [XR_247499] |
| A_33_P3736195 | 0,006251018 | up | 1,8365387 | DKFZp686M1136 | Homo sapiens mRNA; cDNA DKFZp686M1136 (from clone DKFZp686M1136). [BX647230] |
| A_32_P189790 | 0,004931542 | down | -2,5081434 | DKFZp779M0652 | Homo sapiens uncharacterized DKFZp779M0652 (DKFZp779M0652), long non-coding RNA [NR_027134] |
| A_21_P0000139 | 8,21E-04 | down | -2,3586934 | DKKL1 | Homo sapiens dickkopf-like 1 (DKKL1), transcript variant 1, mRNA [NM_014419] |
| A_23_P130743 | 0,019536445 | up | 2,9366312 | DKKL1 | Homo sapiens dickkopf-like 1 (DKKL1), transcript variant 1, mRNA [NM_014419] |
| A_19_P00321577 | 0,019539066 | down | -3,16847 | DLEU1 | Homo sapiens deleted in lymphocytic leukemia 1 (non-protein coding) (DLEU1), transcript variant 3, long non-coding RNA [NR_109974] |
| A_33_P3329462 | 0,005266239 | down | -2,635833 | DLEU1-AS1 | Homo sapiens DLEU1 antisense RNA 1 (DLEU1-AS1), long non-coding RNA [NR_125753] |
| A_21_P0000753 | 0,005680444 | down | -1,6917849 | DLG1-AS1 | Homo sapiens DLG1 antisense RNA 1 (DLG1-AS1), long non-coding RNA [NR_038289] |
| A_32_P229618 | 0,008384389 | down | -2,4926064 | DLG2 | Homo sapiens discs, large homolog 2 (Drosophila) (DLG2), transcript variant 2, mRNA [NM_001364] |
| A_24_P129632 | 0,002488149 | up | 1,7774208 | DLG5 | Homo sapiens discs, large homolog 5 (Drosophila) (DLG5), mRNA [NM_004747] |
| A_24_P62367 | 0,007281127 | up | 1,5816133 | DLGAP1 | Homo sapiens discs, large (Drosophila) homolog-associated protein 1 (DLGAP1), transcript variant 1, mRNA [NM_004746] |
| A_33_P3368785 | 4,30E-04 | down | -1,6626756 | DLGAP1-AS1 | Homo sapiens DLGAP1 antisense RNA 1 (DLGAP1-AS1), long non-coding RNA [NR_024101] |
| A_33_P3511777 | 0,020363208 | down | -2,1924493 | DLGAP1-AS3 | Homo sapiens DLGAP1 antisense RNA 3 (DLGAP1-AS3), long non-coding RNA [NR_038895] |
| A_33_P3232557 | 0,003702205 | down | -1,5786244 | DLGAP3 | Homo sapiens discs, large (Drosophila) homolog-associated protein 3 (DLGAP3), mRNA [NM_001080418] |
| A_23_P205697 | 3,54E-05 | up | 2,1944447 | DLST | Homo sapiens dihydrolipoamide S-succinyltransferase (E2 component of 2-oxo-glutarate complex) (DLST), transcript variant 1, mRNA [NM_001933] |
| A_23_P86599 | 0,012810119 | down | -1,5343366 | DMBT1 | Homo sapiens deleted in malignant brain tumors 1 (DMBT1), transcript variant 2, mRNA [NM_007329] |
| A_23_P50535 | 0,002093551 | up | 1,6997447 | DMPK | Homo sapiens dystrophia myotonica-protein kinase (DMPK), transcript variant 2, mRNA [NM_004409] |
| A_23_P169293 | 8,46E-05 | down | -3,0850255 | DMRT1 | Homo sapiens doublesex and mab-3 related transcription factor 1 (DMRT1), mRNA [NM_021951] |
| A_32_P213459 | 0,019670954 | down | -2,3889842 | DMRT2 | Homo sapiens doublesex and mab-3 related transcription factor 2 (DMRT2), transcript variant 1, mRNA [NM_006557] |

| | | | | | |
|---|---|---|---|---|---|
| A_23_P8664 | 1,48E-04 | up | 2,0007913 | DMTF1 | Homo sapiens cyclin D binding myb-like transcription factor 1 (DMTF1), transcript variant 1, mRNA [NM_021145] |
| A_23_P250571 | 0,001191842 | up | 1,6223614 | DMXL1 | Homo sapiens Dmx-like 1 (DMXL1), transcript variant 2, mRNA [NM_005509] |
| A_23_P54612 | 0,04848233 | up | 1,7026728 | DNAAF1 | Homo sapiens dynein, axonemal, assembly factor 1 (DNAAF1), mRNA [NM_178452] |
| A_33_P3379801 | 3,70E-06 | down | -2,5171723 | DNAAF2 | Homo sapiens dynein, axonemal, assembly factor 2 (DNAAF2), transcript variant 2, mRNA [NM_001083908] |
| A_23_P18213 | 0,030150713 | up | 1,5424603 | DNAH1 | Homo sapiens dynein, axonemal, heavy chain 1 (DNAH1), mRNA [NM_015512] |
| A_32_P19806 | 0,003302187 | up | 2,409575 | DNAH11 | Homo sapiens dynein, axonemal, heavy chain 11 (DNAH11), mRNA [NM_001277115] |
| A_33_P3422010 | 0,004826859 | down | -1,7272685 | DNAH12 | Homo sapiens dynein, axonemal, heavy chain 12 (DNAH12), transcript variant 3, mRNA [NM_001291661] |
| A_24_P127691 | 0,023188455 | down | -1,6056577 | DNAH14 | dynein, axonemal, heavy chain 14 [Source:HGNC Symbol;Acc:HGNC:2945] [ENST00000495456] |
| A_32_P439662 | 0,006191589 | down | -2,1076164 | DNAH17 | Homo sapiens dynein, axonemal, heavy chain 17 (DNAH17), mRNA [NM_173628] |
| A_21_P0011612 | 0,012521409 | down | -1,7481616 | DNAH17 | Homo sapiens dynein, axonemal, heavy chain 17 (DNAH17), mRNA [NM_173628] |
| A_21_P0011611 | 0,015868079 | down | -2,6220064 | DNAH17 | Homo sapiens dynein, axonemal, heavy chain 17 (DNAH17), mRNA [NM_173628] |
| A_21_P0011629 | 0,03889971 | down | -2,205363 | DNAH17-AS1 | Homo sapiens DNAH17 antisense RNA 1 (DNAH17-AS1), long non-coding RNA [NR_102401] |
| A_33_P3285629 | 0,040795118 | down | -1,9019712 | DNAI2 | Homo sapiens dynein, axonemal, intermediate chain 2 (DNAI2), transcript variant 1, mRNA [NM_023036] |
| A_33_P3419239 | 0,007283832 | up | 2,2648532 | DNAI2 | Homo sapiens dynein, axonemal, intermediate chain 2 (DNAI2), transcript variant 1, mRNA [NM_023036] |
| A_23_P90062 | 0,020638434 | up | 1,5062263 | DNAJB1 | Homo sapiens DnaJ (Hsp40) homolog, subfamily B, member 1 (DNAJB1), transcript variant 1, mRNA [NM_006145] |
| A_23_P215227 | 5,62E-05 | up | 2,568141 | DNAJB6 | Homo sapiens DnaJ (Hsp40) homolog, subfamily B, member 6 (DNAJB6), transcript variant 2, mRNA [NM_005494] |
| A_23_P385063 | 7,80E-04 | up | 1,9519597 | DNAJB6 | Homo sapiens DnaJ (Hsp40) homolog, subfamily B, member 6 (DNAJB6), transcript variant 1, mRNA [NM_058246] |
| A_23_P351168 | 0,011160938 | up | 1,9290311 | DNAJB7 | Homo sapiens DnaJ (Hsp40) homolog, subfamily B, member 7 (DNAJB7), mRNA [NM_145174] |
| A_24_P225604 | 0,009710436 | down | -2,0190687 | DNAJC10 | Homo sapiens DnaJ (Hsp40) homolog, subfamily C, member 10 (DNAJC10), transcript variant 1, mRNA [NM_018981] |
| A_23_P140698 | 0,00551278 | up | 1,557554 | DNAJC17 | Homo sapiens DnaJ (Hsp40) homolog, subfamily C, member 17 (DNAJC17), mRNA [NM_018163] |
| A_33_P3310533 | 0,013973764 | down | -1,6050729 | DNAJC21 | Homo sapiens DnaJ (Hsp40) homolog, subfamily C, member 21 (DNAJC21), transcript variant 2, mRNA [NM_001012339] |
| A_32_P407245 | 0,001390776 | down | -2,958204 | DNAJC22 | Homo sapiens DnaJ (Hsp40) homolog, subfamily C, member 22 (DNAJC22), mRNA [NM_024902] |
| A_33_P3238623 | 0,001455533 | down | -2,9765627 | DNAJC27-AS1 | Homo sapiens DNAJC27 antisense RNA 1 (DNAJC27-AS1), long non-coding RNA [NR_034113] |
| A_33_P3462960 | 0,034826424 | down | -1,7510014 | DNAJC3 | Homo sapiens DnaJ (Hsp40) homolog, subfamily C, member 3 (DNAJC3), mRNA [NM_006260] |
| A_23_P417921 | 0,002681698 | down | -3,4832234 | DNAJC5G | Homo sapiens DnaJ (Hsp40) homolog, subfamily C, member 5 gamma (DNAJC5G), transcript variant 1, mRNA [NM_173650] |
| A_33_P3234202 | 0,003853421 | down | -3,5173056 | DNASE1L3 | Homo sapiens deoxyribonuclease I-like 3 (DNASE1L3), transcript variant 1, mRNA [NM_004944] |
| A_33_P3329419 | 0,011025995 | down | -1,7211856 | DNM1 | Homo sapiens dynamin 1 (DNM1), transcript variant 5, mRNA [NM_001288739] |
| A_21_P0014010 | 0,024666678 | up | 1,9884621 | DNM1P35 | Homo sapiens FKSG88 mRNA, complete cds. [AF357221] |
| A_33_P3378384 | 0,04026399 | down | -1,8548464 | DNM1P46 | Homo sapiens DNM1 pseudogene 46 (DNM1P46), non-coding RNA [NR_003260] |
| A_33_P3264364 | 0,04994858 | down | -1,5548985 | DNM3 | Homo sapiens dynamin 3 (DNM3), transcript variant 3, mRNA [NM_001278252] |
| A_21_P0000785 | 0,004500289 | down | -1,5831301 | DNM3OS | Homo sapiens DNM3 opposite strand/antisense RNA (DNM3OS), transcript variant 1, long non-coding RNA [NR_038397] |
| A_23_P328206 | 0,022542162 | up | 1,5521035 | DNMBP | Homo sapiens dynamin binding protein (DNMBP), mRNA [NM_015221] |
| A_33_P3329187 | 0,022357736 | up | 1,556207 | DNMT1 | Homo sapiens DNA (cytosine-5-)-methyltransferase 1 (DNMT1), transcript variant 1, mRNA [NM_001130823] |
| A_33_P3359508 | 0,013587849 | up | 2,4637272 | DNMT3B | Homo sapiens mRNA for DNA cytosine-5 methyltransferase 3 beta isoform 6 variant protein. [AB208880] |
| A_23_P45059 | 0,001120868 | up | 1,6661294 | DOCK1 | Homo sapiens dedicator of cytokinesis 1 (DOCK1), transcript variant 2, mRNA [NM_001380] |
| A_23_P16722 | 0,009644154 | down | -2,4588943 | DOCK10 | Homo sapiens dedicator of cytokinesis 10 (DOCK10), transcript variant DOCK10.1, mRNA [NM_014689] |
| A_23_P59637 | 0,007658771 | up | 2,1747725 | DOCK4 | Homo sapiens dedicator of cytokinesis 4 (DOCK4), mRNA [NM_014705] |
| A_21_P0014196 | 0,003022428 | up | 1,9664681 | DOCK4-AS1 | Homo sapiens DOCK4 antisense RNA 1 (DOCK4-AS1), long non-coding RNA [NR_103806] |
| A_24_P203622 | 0,047245428 | down | -1,7191191 | DOCK7 | Homo sapiens dedicator of cytokinesis 7 (DOCK7), transcript variant 2, mRNA [NM_033407] |
| A_23_P353888 | 0,006473901 | up | 1,666996 | DOCK8 | Homo sapiens dedicator of cytokinesis 8 (DOCK8), transcript variant 1, mRNA [NM_203447] |
| A_33_P3410279 | 0,03036796 | up | 1,6931001 | DOCK9 | dedicator of cytokinesis 9 [Source:HGNC Symbol;Acc:HGNC:14132] [ENST00000340449] |
| A_33_P3228564 | 6,51E-04 | down | -2,3601186 | DOK3 | Homo sapiens docking protein 3 (DOK3), transcript variant 2, mRNA [NM_001144875] |
| A_23_P40217 | 0,04773601 | down | -2,0114987 | DOK5 | Homo sapiens docking protein 5 (DOK5), transcript variant 1, mRNA [NM_018431] |
| A_32_P160561 | 0,022296073 | up | 1,6394849 | DOK6 | Homo sapiens docking protein 6 (DOK6), mRNA [NM_152721] |
| A_33_P3416009 | 0,013051764 | up | 2,2344027 | DONSON | Homo sapiens cDNA FLJ10412 fis, clone NT2RP1000040. [AK001274] |
| A_33_P3274164 | 0,016668975 | down | -2,2119913 | DOT1L | PREDICTED: Homo sapiens DOT1-like histone H3K79 methyltransferase (DOT1L), transcript variant X2, mRNA [XM_005259660] |
| A_23_P122508 | 0,032821715 | up | 2,2440035 | DPCR1 | Homo sapiens diffuse panbronchiolitis critical region 1 (DPCR1), mRNA [NM_080870] |
| A_23_P129413 | 0,006410284 | down | -2,1258245 | DPEP3 | Homo sapiens dipeptidase 3 (DPEP3), transcript variant 1, mRNA [NM_022357] |
| A_33_P3317277 | 0,001422871 | down | -2,062667 | DPF1 | Homo sapiens D4, zinc and double PHD fingers family 1 (DPF1), transcript variant 4, mRNA [NM_001289978] |
| A_24_P140057 | 0,004362118 | up | 1,5014815 | DPP10 | Homo sapiens dipeptidyl-peptidase 10 (non-functional) (DPP10), transcript variant 1, mRNA [NM_020868] |
| A_23_P170888 | 0,019059066 | up | 2,4859843 | DPP6 | Homo sapiens dipeptidyl-peptidase 6 (DPP6), transcript variant 3, mRNA [NM_001039350] |
| A_24_P288323 | 0,028084772 | up | 1,580212 | DPP9 | Homo sapiens dipeptidyl-peptidase 9 (DPP9), mRNA [NM_139159] |
| A_23_P200741 | 7,57E-04 | down | -2,8650231 | DPT | Homo sapiens dermatopontin (DPT), mRNA [NM_001937] |
| A_21_P0000652 | 0,006175277 | up | 1,7490034 | DPY19L2P3 | Homo sapiens DPY19L2 pseudogene 3 (DPY19L2P3), non-coding RNA [NR_036482] |
| A_33_P3705275 | 0,026975082 | down | -1,7875768 | DPY19L3 | Homo sapiens dpy-19-like 3 (C. elegans) (DPY19L3), transcript variant 1, mRNA [NM_207325] |
| A_21_P0000189 | 7,98E-04 | up | 1,607012 | DR1 | Homo sapiens down-regulator of transcription 1, TBP-binding (negative cofactor 2) (DR1), mRNA [NM_001938] |
| A_24_P109417 | 0,001425563 | down | -2,1778865 | DRAXIN | Homo sapiens dorsal inhibitory axon guidance protein (DRAXIN), mRNA [NM_198545] |
| A_23_P308722 | 0,02663354 | up | 1,5768726 | DRC1 | Homo sapiens dynein regulatory complex subunit 1 (DRC1), mRNA [NM_145038] |
| A_33_P3373765 | 0,021281824 | up | 1,6649033 | DRD4 | Homo sapiens dopamine receptor D4 (DRD4), mRNA [NM_000797] |
| A_32_P54274 | 0,007347291 | down | -2,555898 | DRD5 | Homo sapiens dopamine receptor D5 (DRD5), mRNA [NM_000798] |
| A_33_P3345344 | 0,040082086 | up | 1,6131399 | DRGX | dorsal root ganglia homeobox [Source:HGNC Symbol;Acc:HGNC:21536] [ENST00000374139] |

| | | | | | |
|---|---|---|---|---|---|
| A_33_P3244753 | 0,048566543 | down | -1,5562943 | DRP2 | Homo sapiens dystrophin related protein 2 (DRP2), transcript variant 1, mRNA [NM_001939] |
| A_23_P4494 | 9,89E-04 | down | -1,9485718 | DSC2 | Homo sapiens desmocollin 2 (DSC2), transcript variant Dsc2a, mRNA [NM_024422] |
| A_33_P3266898 | 0,008562752 | down | -1,6537274 | DSC3 | Homo sapiens desmocollin 3 (DSC3), transcript variant Dsc3a, mRNA [NM_001941] |
| A_23_P47340 | 0,00395136 | up | 1,5103059 | DSCAML1 | Homo sapiens Down syndrome cell adhesion molecule like 1 (DSCAML1), mRNA [NM_020693] |
| A_33_P3436732 | 0,005906404 | down | -1,6795889 | DSCR10 | Homo sapiens Down syndrome critical region 10 (non-protein coding) (DSCR10), long non-coding RNA [NR_027695] |
| A_23_P6252 | 0,010442387 | up | 2,0846872 | DSCR8 | Homo sapiens Down syndrome critical region 8 (DSCR8), transcript variant 4, long non-coding RNA [NR_026838] |
| A_33_P3254996 | 0,04244527 | down | -1,5011464 | DSCR9 | Homo sapiens Down syndrome critical region 9 (non-protein coding) (DSCR9), long non-coding RNA [NR_026719] |
| A_23_P165937 | 0,001956194 | up | 1,7212129 | DSN1 | Homo sapiens DSN1, MIS12 kinetochore complex component (DSN1), transcript variant 3, mRNA [NM_024918] |
| A_24_P145316 | 7,57E-04 | up | 1,8062468 | DTNBP1 | Homo sapiens dystrobrevin binding protein 1 (DTNBP1), transcript variant 2, mRNA [NM_183040] |
| A_24_P70117 | 0,002419105 | down | -1,5647241 | DTWD1 | Homo sapiens DTW domain containing 1 (DTWD1), transcript variant 1, mRNA [NM_020234] |
| A_23_P54291 | 0,006314754 | up | 1,8059084 | DUOX1 | Homo sapiens dual oxidase 1 (DUOX1), transcript variant 1, mRNA [NM_017434] |
| A_23_P151851 | 0,008428302 | up | 1,5826753 | DUOX2 | Homo sapiens dual oxidase 2 (DUOX2), mRNA [NM_014080] |
| A_23_P110712 | 0,003292607 | up | 2,6109247 | DUSP1 | Homo sapiens dual specificity phosphatase 1 (DUSP1), mRNA [NM_004417] |
| A_24_P182494 | 0,004330461 | up | 1,9164082 | DUSP10 | Homo sapiens dual specificity phosphatase 10 (DUSP10), transcript variant 1, mRNA [NM_007207] |
| A_23_P207537 | 0,015296159 | up | 1,5188663 | DUSP14 | Homo sapiens dual specificity phosphatase 14 (DUSP14), mRNA [NM_007026] |
| A_23_P369090 | 0,00373022 | up | 1,7598325 | DUSP15 | dual specificity phosphatase 15 [Source:HGNC Symbol;Acc:HGNC:16236] [ENST00000278979] |
| A_23_P120254 | 0,00730953 | up | 1,5154352 | DUSP22 | Homo sapiens dual specificity phosphatase 22 (DUSP22), transcript variant 2, mRNA [NM_020185] |
| A_23_P146134 | 4,07E-05 | down | -3,01515 | DUSP26 | Homo sapiens dual specificity phosphatase 26 (putative) (DUSP26), mRNA [NM_024025] |
| A_33_P3294645 | 0,019835008 | up | 2,487461 | DUSP27 | Homo sapiens dual specificity phosphatase 27 (putative) (DUSP27), mRNA [NM_001080426] |
| A_32_P98298 | 0,01600302 | down | -1,573665 | DUSP8 | Homo sapiens dual specificity phosphatase 8 (DUSP8), mRNA [NM_004420] |
| A_24_P417189 | 0,006913159 | down | -1,58841 | DUSP9 | Homo sapiens dual specificity phosphatase 9 (DUSP9), mRNA [NM_001395] |
| A_33_P3256105 | 0,001834772 | down | -2,7120774 | DUX4 | Homo sapiens double homeobox 4 (DUX4), mRNA [NM_001293798] |
| A_21_P0014945 | 0,00490812 | down | -1,5617869 | DUX4 | PREDICTED: Homo sapiens double homeobox 4-like (DUX4L), partial ncRNA [XR_431831] |
| A_33_P3308586 | 0,011966401 | down | -2,86793 | DUXA | Homo sapiens double homeobox A (DUXA), mRNA [NM_001012729] |
| A_23_P55342 | 0,005366131 | up | 1,6559526 | DVL2 | Homo sapiens dishevelled segment polarity protein 2 (DVL2), mRNA [NM_004422] |
| A_23_P380309 | 0,006360615 | up | 1,5730883 | DYNAP | Homo sapiens dynactin associated protein (DYNAP), mRNA [NM_173629] |
| A_33_P3228340 | 0,002299959 | down | -1,8857358 | DYNC2LI1 | Homo sapiens dynein, cytoplasmic 2, light intermediate chain 1, mRNA (cDNA clone IMAGE:5268633), complete cds. [BC040558] |
| A_33_P3260445 | 0,013770338 | up | 1,63385 | DYNLT1 | dynein, light chain, Tctex-type 1 [Source:HGNC Symbol;Acc:HGNC:11697] [ENST00000367088] |
| A_23_P211126 | 4,48E-04 | up | 1,7837408 | DYRK1A | Homo sapiens dual-specificity tyrosine-(Y)-phosphorylation regulated kinase 1A (DYRK1A), transcript variant 2, mRNA [NM_130436] |
| A_24_P345209 | 0,01952586 | down | -1,6890571 | DYRK3 | Homo sapiens dual-specificity tyrosine-(Y)-phosphorylation regulated kinase 3 (DYRK3), transcript variant 2, mRNA [NM_001004023] |
| A_33_P3318861 | 6,12E-07 | down | -3,8533163 | DYTN | Homo sapiens dystrotelin (DYTN), mRNA [NM_001093730] |
| A_33_P3344169 | 0,008045947 | down | -1,7103448 | DYX1C1 | Homo sapiens dyslexia susceptibility 1 candidate 1 (DYX1C1), transcript variant 3, mRNA [NM_001033560] |
| A_21_P0000077 | 0,003504455 | up | 2,1751118 | DZIP1L | Homo sapiens DAZ interacting zinc finger protein 1-like (DZIP1L), transcript variant 2, mRNA [NM_001170538] |
| A_33_P3267651 | 0,049999043 | down | -1,6429102 | DZIP3 | Homo sapiens DAZ interacting zinc finger protein 3 (DZIP3), mRNA [NM_014648] |
| A_23_P152218 | 0,002492347 | up | 1,7194058 | E2F4 | Homo sapiens E2F transcription factor 4, p107/p130-binding (E2F4), mRNA [NM_001950] |
| A_33_P3318661 | 0,014770975 | down | -2,820066 | E2F7 | Homo sapiens E2F transcription factor 7 (E2F7), mRNA [NM_203394] |
| A_23_P119478 | 0,03369076 | up | 2,0352004 | EBI3 | Homo sapiens Epstein-Barr virus induced 3 (EBI3), mRNA [NM_005755] |
| A_23_P6561 | 0,006324521 | up | 1,527278 | EBLN2 | Homo sapiens endogenous Bornavirus-like nucleoprotein 2 (EBLN2), mRNA [NM_018029] |
| A_23_P128554 | 0,02712596 | up | 1,5203484 | EBPL | Homo sapiens emopamil binding protein-like (EBPL), transcript variant 1, mRNA [NM_032565] |
| A_32_P155666 | 2,44E-06 | down | -3,5224547 | ECEL1 | Homo sapiens endothelin converting enzyme-like 1 (ECEL1), transcript variant 1, mRNA [NM_004826] |
| A_33_P3610406 | 0,035881113 | down | -1,9595158 | ECEL1P2 | Homo sapiens endothelin converting enzyme-like 1, pseudogene 2 (ECEL1P2), non-coding RNA [NR_028501] |
| A_23_P303671 | 0,045427993 | down | -2,4793918 | ECM2 | Homo sapiens extracellular matrix protein 2, female organ and adipocyte specific (ECM2), transcript variant 1, mRNA [NM_001393] |
| A_33_P3398386 | 0,008423259 | up | 1,677102 | ECM2 | Homo sapiens extracellular matrix protein 2, female organ and adipocyte specific (ECM2), transcript variant 3, mRNA [NM_001197296] |
| A_23_P214821 | 0,015242857 | up | 2,0308316 | EDN1 | Homo sapiens endothelin 1 (EDN1), transcript variant 1, mRNA [NM_001955] |
| A_33_P3258392 | 0,004991615 | up | 2,2229662 | EDN1 | Homo sapiens endothelin 1 (EDN1), transcript variant 1, mRNA [NM_001955] |
| A_23_P312150 | 0,004822036 | up | 4,1389832 | EDN2 | Homo sapiens endothelin 2 (EDN2), transcript variant 1, mRNA [NM_001956] |
| A_23_P17438 | 0,002242122 | up | 1,643979 | EDN3 | Homo sapiens endothelin 3 (EDN3), transcript variant 2, mRNA [NM_207032] |
| A_33_P3401701 | 0,001845509 | down | -2,4981432 | EDNRA | Homo sapiens endothelin receptor type A (EDNRA), transcript variant 1, mRNA [NM_001957] |
| A_24_P217572 | 0,007951487 | down | -2,4608543 | EDNRA | Homo sapiens endothelin receptor type A (EDNRA), transcript variant 1, mRNA [NM_001957] |
| A_24_P330263 | 0,011145772 | up | 3,0162613 | EDNRB | Homo sapiens endothelin receptor type B (EDNRB), transcript variant 2, mRNA [NM_003991] |
| A_23_P144202 | 0,01286239 | down | -1,999893 | EEFSEC | Homo sapiens eukaryotic elongation factor, selenocysteine-tRNA-specific (EEFSEC), mRNA [NM_021937] |
| A_33_P3785051 | 4,40E-05 | down | -2,860614 | EFCAB10 | Homo sapiens cDNA clone IMAGE:6616931, partial cds. [BC062748] |
| A_32_P99753 | 0,004179734 | down | -2,0296485 | EFCAB12 | Homo sapiens EF-hand calcium binding domain 12 (EFCAB12), mRNA [NM_207307] |
| A_24_P374445 | 0,001638302 | up | 1,728249 | EFCAB13 | Homo sapiens EF-hand calcium binding domain 13 (EFCAB13), transcript variant A, mRNA [NM_152347] |
| A_23_P149476 | 0,018866753 | down | -1,699196 | EFCAB2 | Homo sapiens EF-hand calcium binding domain 2 (EFCAB2), transcript variant 1, mRNA [NM_032328] |
| A_33_P3341399 | 0,03724218 | up | 2,0360973 | EFCAB5 | Homo sapiens EF-hand calcium binding domain 5 (EFCAB5), transcript variant 1, mRNA [NM_198529] |
| A_24_P144439 | 0,040022194 | up | 1,67512 | EFHB | Homo sapiens EF-hand domain family, member B (EFHB), mRNA [NM_144715] |
| A_33_P3418716 | 0,02257096 | up | 2,0486677 | EFHD1 | Homo sapiens EF-hand domain family, member D1 (EFHD1), transcript variant 1, mRNA [NM_025202] |
| A_23_P154338 | 0,028402377 | up | 1,7657197 | EFHD1 | Homo sapiens EF-hand domain family, member D1 (EFHD1), transcript variant 1, mRNA [NM_025202] |
| A_24_P365807 | 0,0458631 | up | 1,5034336 | EFNB1 | Homo sapiens ephrin-B1 (EFNB1), mRNA [NM_004429] |
| A_33_P3373614 | 0,026753431 | up | 2,1357713 | EFR3B | Homo sapiens EFR3 homolog B (S. cerevisiae) (EFR3B), mRNA [NM_014971] |

| | | | | | |
|---|---|---|---|---|---|
| A_21_P0000670 | 0,002861952 | down | -1,809165 | EFTUD1 | Homo sapiens elongation factor Tu GTP binding domain containing 1 (EFTUD1), transcript variant 1, mRNA [NM_024580] |
| A_21_P0014141 | 0,015739515 | down | -2,5724912 | EGFLAM-AS2 | Homo sapiens EGFLAM antisense RNA 2 (EGFLAM-AS2), transcript variant 1, long non-coding RNA [NR_102749] |
| A_23_P46936 | 0,011283875 | down | -2,1333718 | EGR2 | Homo sapiens early growth response 2 (EGR2), transcript variant 1, mRNA [NM_000399] |
| A_23_P79251 | 0,042895146 | up | 1,5655627 | EHD3 | Homo sapiens EH-domain containing 3 (EHD3), mRNA [NM_014600] |
| A_23_P163458 | 0,030832924 | up | 1,793863 | EHD4 | Homo sapiens EH-domain containing 4 (EHD4), mRNA [NM_139265] |
| A_24_P123119 | 0,023222499 | down | -2,0867867 | EHHADH | Homo sapiens enoyl-CoA, hydratase/3-hydroxyacyl CoA dehydrogenase (EHHADH), transcript variant 1, mRNA [NM_001966] |
| A_33_P3570218 | 0,002554313 | up | 2,1769478 | EHHADH-AS1 | Homo sapiens EHHADH antisense RNA 1 (EHHADH-AS1), long non-coding RNA [NR_038990] |
| A_33_P3420841 | 0,002072258 | up | 1,7297856 | EIF3D | eukaryotic translation initiation factor 3, subunit D [Source:HGNC Symbol;Acc:HGNC:3278] [ENST00000402116] |
| A_23_P121250 | 2,15E-04 | up | 1,7888389 | EIF4A2 | Homo sapiens eukaryotic translation initiation factor 4A2 (EIF4A2), mRNA [NM_001967] |
| A_24_P218970 | 0,009203413 | up | 1,6621192 | EIF4B | Homo sapiens eukaryotic translation initiation factor 4B (EIF4B), transcript variant 2, mRNA [NM_001417] |
| A_24_P115621 | 0,003624963 | up | 1,7561288 | EIF4EBP2 | Homo sapiens eukaryotic translation initiation factor 4E binding protein 2 (EIF4EBP2), mRNA [NM_004096] |
| A_33_P3367577 | 0,014299153 | down | -1,918253 | EIF4ENIF1 | eukaryotic translation initiation factor 4E nuclear import factor 1 [Source:HGNC Symbol;Acc:HGNC:16687] [ENST00000397520] |
| A_33_P3306545 | 0,007566222 | up | 1,6055776 | EIF4G2 | Homo sapiens eukaryotic translation initiation factor 4 gamma, 2 (EIF4G2), transcript variant 3, mRNA [NM_001172705] |
| A_33_P3265504 | 4,05E-04 | down | -2,0771916 | EIF5AL1 | Homo sapiens eukaryotic translation initiation factor 5A-like 1 (EIF5AL1), mRNA [NM_001099692] |
| A_23_P104188 | 0,015842427 | up | 1,9937354 | ELF3 | Homo sapiens E74-like factor 3 (ets domain transcription factor, epithelial-specific ) (ELF3), transcript variant 1, mRNA [NM_004433] |
| A_33_P3210848 | 6,92E-04 | down | -2,8148923 | ELFN1 | Homo sapiens extracellular leucine-rich repeat and fibronectin type III domain containing 1 (ELFN1), mRNA [NM_001128636] |
| A_23_P171054 | 0,002846356 | up | 2,5292606 | ELK1 | Homo sapiens ELK1, member of ETS oncogene family (ELK1), transcript variant 2, mRNA [NM_005229] |
| A_23_P338325 | 0,04199136 | up | 1,8026216 | ELK3 | ELK3, ETS-domain protein (SRF accessory protein 2) [Source:HGNC Symbol;Acc:HGNC:3325] [ENST00000549985] |
| A_33_P3365010 | 3,51E-04 | down | -1,811571 | ELK4 | Homo sapiens ELK4, ETS-domain protein (SRF accessory protein 1) (ELK4), transcript variant b, mRNA [NM_021795] |
| A_23_P161098 | 3,15E-04 | down | -1,7513969 | ELK4 | Homo sapiens ELK4, ETS-domain protein (SRF accessory protein 1) (ELK4), transcript variant a, mRNA [NM_001973] |
| A_23_P16538 | 0,002874676 | up | 1,5056599 | ELL | Homo sapiens elongation factor RNA polymerase II (ELL), mRNA [NM_006532] |
| A_23_P58506 | 0,009220957 | down | -1,510791 | ELL2 | Homo sapiens elongation factor, RNA polymerase II, 2 (ELL2), mRNA [NM_012081] |
| A_33_P3235204 | 5,92E-04 | down | -2,2281508 | ELMOD3 | Homo sapiens ELMO/CED-12 domain containing 3 (ELMOD3), transcript variant 1, mRNA [NM_032213] |
| A_33_P3297305 | 0,038555056 | up | 1,6188455 | ELMOD3 | Homo sapiens ELMO/CED-12 domain containing 3 (ELMOD3), transcript variant 1, mRNA [NM_032213] |
| A_23_P215454 | 0,017970193 | down | -2,1434155 | ELN | Homo sapiens elastin (ELN), transcript variant 13, mRNA [NM_001278939] |
| A_23_P62741 | 0,01272562 | down | -1,6796091 | ELTD1 | Homo sapiens EGF, latrophilin and seven transmembrane domain containing 1 (ELTD1), mRNA [NM_022159] |
| A_23_P384698 | 0,001464986 | up | 1,8395041 | EMC3-AS1 | Homo sapiens EMC3 antisense RNA 1 (EMC3-AS1), long non-coding RNA [NR_103821] |
| A_33_P3333455 | 0,006747321 | down | -1,6864041 | EMILIN1 | Homo sapiens elastin microfibril interfacer 1 (EMILIN1), mRNA [NM_007046] |
| A_33_P3230558 | 0,0178784 | down | -2,9097154 | EML5 | Homo sapiens echinoderm microtubule associated protein like 5 (EML5), mRNA [NM_183387] |
| A_23_P76488 | 0,001533294 | up | 2,7437348 | EMP1 | Homo sapiens epithelial membrane protein 1 (EMP1), mRNA [NM_001423] |
| A_23_P502336 | 0,001613648 | up | 3,0525634 | EMR2 | Homo sapiens egf-like module containing, mucin-like, hormone receptor-like 2 (EMR2), transcript variant 1, mRNA [NM_013447] |
| A_33_P3343715 | 0,007534971 | down | -1,7811725 | EMX1 | Homo sapiens empty spiracles homeobox 1, mRNA (cDNA clone IMAGE:5198260), complete cds. [BC037242] |
| A_23_P315122 | 0,008555609 | down | -1,6279536 | EMX1 | Homo sapiens empty spiracles homeobox 1 (EMX1), mRNA [NM_004097] |
| A_24_P892472 | 0,00470722 | up | 1,533044 | EMX2OS | Homo sapiens EMX2 opposite strand/antisense RNA (EMX2OS), long non-coding RNA [NR_002791] |
| A_23_P134433 | 0,004071393 | down | -2,1234019 | EN2 | Homo sapiens engrailed homeobox 2 (EN2), mRNA [NM_001427] |
| A_33_P3283525 | 1,72E-04 | down | -2,8275383 | ENAH | enabled homolog (Drosophila) [Source:HGNC Symbol;Acc:HGNC:18271] [ENST00000498108] |
| A_33_P3274245 | 2,47E-05 | down | -3,2017236 | ENDOV | Homo sapiens endonuclease V (ENDOV), transcript variant 1, mRNA [NM_173627] |
| A_33_P3405114 | 0,011816337 | down | -1,7067217 | ENHO | Homo sapiens energy homeostasis associated (ENHO), mRNA [NM_198573] |
| A_33_P3268478 | 0,03362398 | up | 2,4324758 | ENO4 | PREDICTED: Homo sapiens enolase family member 4 (ENO4), transcript variant X3, mRNA [XM_006717835] |
| A_33_P3314407 | 0,004169174 | up | 2,05936 | ENO4 | Homo sapiens enolase family member 4 (ENO4), mRNA [NM_001242699] |
| A_33_P3383491 | 0,015615943 | up | 2,1667993 | ENPP7 | Homo sapiens ectonucleotide pyrophosphatase/phosphodiesterase 7 (ENPP7), mRNA [NM_178543] |
| A_23_P410115 | 0,02173277 | up | 2,7167153 | ENTHD1 | Homo sapiens ENTH domain containing 1 (ENTHD1), mRNA [NM_152512] |
| A_33_P3267395 | 0,00668312 | up | 1,5455282 | ENTHD1 | Homo sapiens ENTH domain containing 1 (ENTHD1), mRNA [NM_152512] |
| A_33_P3262580 | 0,001272671 | down | -1,9607317 | ENTPD1 | ectonucleoside triphosphate diphosphohydrolase 1 [Source:HGNC Symbol;Acc:HGNC:3363] [ENST00000371206] |
| A_23_P40693 | 7,63E-08 | up | 3,0056467 | EP300 | Homo sapiens E1A binding protein p300 (EP300), mRNA [NM_001429] |
| A_23_P253158 | 4,81E-04 | up | 1,9169128 | EP400 | Homo sapiens E1A binding protein p400 (EP400), mRNA [NM_015409] |
| A_24_P185709 | 0,035125125 | up | 1,8212844 | EPB41L1 | Homo sapiens erythrocyte membrane protein band 4.1-like 1 (EPB41L1), transcript variant 1, mRNA [NM_012156] |
| A_24_P944640 | 3,59E-05 | up | 2,6409175 | EPB41L5 | Homo sapiens erythrocyte membrane protein band 4.1 like 5 (EPB41L5), transcript variant 1, mRNA [NM_020909] |
| A_23_P149281 | 0,001811384 | up | 2,2461278 | EPHA2 | Homo sapiens EPH receptor A2 (EPHA2), mRNA [NM_004431] |
| A_23_P147839 | 0,003417709 | up | 1,6498951 | EPHA5 | Homo sapiens EPH receptor A5 (EPHA5), transcript variant 1, mRNA [NM_004439] |
| A_24_P34031 | 0,002845326 | up | 1,5979704 | EPHA7 | Homo sapiens EPH receptor A7 (EPHA7), transcript variant 3, mRNA [NM_001288630] |
| A_23_P166993 | 0,013252611 | up | 1,6178619 | EPHB1 | Homo sapiens EPH receptor B1 (EPHB1), mRNA [NM_004441] |
| A_33_P3880302 | 0,0379964 | up | 1,7301103 | EPHB2 | Homo sapiens EPH receptor B2 (EPHB2), transcript variant 2, mRNA [NM_004442] |
| A_23_P8834 | 0,04492877 | down | -1,6941019 | EPHX2 | Homo sapiens epoxide hydrolase 2, cytoplasmic (EPHX2), transcript variant 1, mRNA [NM_001979] |
| A_33_P3317168 | 0,004511012 | down | -1,8272046 | EPS8L3 | Homo sapiens EPS8-like 3 (EPS8L3), transcript variant 1, mRNA [NM_139053] |
| A_33_P3418371 | 0,0117395 | down | -2,476254 | EPWW6493 | Homo sapiens clone DNA142975 EPWW6493 (UNQ6493) mRNA, complete cds. [AY358234] |
| A_24_P314585 | 0,02443491 | up | 1,5268553 | ERAS | ES cell expressed Ras [Source:HGNC Symbol;Acc:HGNC:5174] [ENST00000338270] |
| A_23_P30175 | 0,003749256 | up | 1,5263602 | ERBB2IP | Homo sapiens erbb2 interacting protein (ERBB2IP), transcript variant 2, mRNA [NM_018695] |
| A_23_P349416 | 0,007924241 | up | 2,006668 | ERBB3 | Homo sapiens v-erb-b2 avian erythroblastic leukemia viral oncogene homolog 3 (ERBB3), transcript variant 1, mRNA [NM_001982] |
| A_33_P3211558 | 0,002223508 | up | 1,7141474 | ERBB4 | Homo sapiens erb-b2 receptor tyrosine kinase 4 (ERBB4), transcript variant JM-a/CVT-1, mRNA [NM_005235] |
| A_24_P374759 | 0,03658763 | up | 2,3656187 | ERC2 | Homo sapiens ELKS/RAB6-interacting/CAST family member 2 (ERC2), mRNA [NM_015576] |

| | | | | | |
|---|---|---|---|---|---|
| A_23_P18055 | 0,001127566 | down | -1,8585036 | ERC2-IT1 | Homo sapiens ERC2 intronic transcript 1 (non-protein coding) (ERC2-IT1), long non-coding RNA [NR_024615] |
| A_24_P401990 | 0,006906077 | down | -1,5723 | ERCC2 | Homo sapiens excision repair cross-complementation group 2 (ERCC2), transcript variant 1, mRNA [NM_000400] |
| A_23_P5325 | 0,003661848 | up | 1,531169 | ERCC3 | Homo sapiens excision repair cross-complementation group 3 (ERCC3), transcript variant 1, mRNA [NM_000122] |
| A_33_P3243364 | 8,19E-04 | down | -1,7636925 | ERCC4 | Homo sapiens excision repair cross-complementation group 4 (ERCC4), mRNA [NM_005236] |
| A_32_P96325 | 0,001393546 | down | -3,3412685 | ERCC6L | Homo sapiens excision repair cross-complementation group 6-like (ERCC6L), mRNA [NM_017669] |
| A_23_P301414 | 0,009147773 | up | 1,5297977 | ERG | Homo sapiens v-ets avian erythroblastosis virus E26 oncogene homolog (ERG), transcript variant 2, mRNA [NM_004449] |
| A_32_P26969 | 0,00157811 | up | 1,6210351 | ERICH1 | Homo sapiens glutamate-rich 1 (ERICH1), transcript variant 1, mRNA [NM_207332] |
| A_21_P0001935 | 0,02320879 | down | -1,706053 | ERICH2 | Homo sapiens glutamate-rich 2 (ERICH2), transcript variant 3, mRNA [NM_001290031] |
| A_32_P88231 | 0,021358388 | down | -2,302917 | ERICH2 | Homo sapiens glutamate-rich 2 (ERICH2), transcript variant 3, mRNA [NM_001290031] |
| A_24_P522998 | 0,011366876 | up | 1,6873815 | ERICH3 | Homo sapiens glutamate-rich 3 (ERICH3), mRNA [NM_001002912] |
| A_33_P3527931 | 0,044285484 | up | 1,9202657 | ERICH4 | Homo sapiens glutamate-rich 4 (ERICH4), mRNA [NM_001130514] |
| A_23_P346431 | 0,039495114 | up | 2,0218446 | ERICH6 | Homo sapiens glutamate-rich 6 (ERICH6), mRNA [NM_152394] |
| A_23_P52219 | 3,41E-04 | down | -1,7161516 | ERLIN1 | Homo sapiens ER lipid raft associated 1 (ERLIN1), mRNA [NM_006459] |
| A_33_P3363061 | 7,53E-04 | down | -1,5709026 | ERLIN2 | Homo sapiens ER lipid raft associated 2 (ERLIN2), transcript variant 2, mRNA [NM_001003790] |
| A_24_P276873 | 0,007341505 | up | 2,7847862 | ERMN | ermin, ERM-like protein [Source:HGNC Symbol;Acc:HGNC:29208] [ENST00000409395] |
| A_23_P164042 | 0,027926857 | down | -1,5825058 | ERN1 | Homo sapiens endoplasmic reticulum to nucleus signaling 1 (ERN1), mRNA [NM_001433] |
| A_33_P3391521 | 0,04915751 | up | 1,9391391 | ERO1LB | ERO1-like beta (S. cerevisiae) [Source:HGNC Symbol;Acc:HGNC:14355] [ENST00000366589] |
| A_23_P46470 | 6,29E-04 | up | 2,5742476 | ERRFI1 | Homo sapiens ERBB receptor feedback inhibitor 1 (ERRFI1), mRNA [NM_018948] |
| A_33_P3274179 | 4,51E-05 | up | 2,8315766 | ERV18-1 | Homo sapiens cDNA FLJ44837 fis, clone BRACE3048677. [AK126787] |
| A_24_P924261 | 0,013387012 | up | 1,7997565 | ERVFRD-1 | Homo sapiens endogenous retrovirus group FRD, member 1 (ERVFRD-1), mRNA [NM_207582] |
| A_21_P0011452 | 0,005507411 | up | 1,8732511 | ERVK13-1 | Homo sapiens endogenous retrovirus group K13, member 1 (ERVK13-1), long non-coding RNA [NR_040023] |
| A_21_P0011451 | 0,00183766 | up | 1,8008957 | ERVK13-1 | Homo sapiens endogenous retrovirus group K13, member 1 (ERVK13-1), long non-coding RNA [NR_040023] |
| A_23_P47410 | 0,001001694 | down | -2,3179276 | ESAM | Homo sapiens endothelial cell adhesion molecule (ESAM), mRNA [NM_138961] |
| A_23_P317347 | 0,040918473 | up | 1,5483316 | ESCO1 | Homo sapiens establishment of sister chromatid cohesion N-acetyltransferase 1 (ESCO1), mRNA [NM_052911] |
| A_24_P323598 | 0,009181517 | up | 1,5034969 | ESCO2 | Homo sapiens establishment of sister chromatid cohesion N-acetyltransferase 2 (ESCO2), mRNA [NM_001017420] |
| A_32_P901770 | 0,006881032 | up | 1,6445563 | ESPN | Homo sapiens espin (ESPN), mRNA [NM_031475] |
| A_24_P383478 | 0,001982417 | down | -2,0436792 | ESR1 | Homo sapiens estrogen receptor 1 (ESR1), transcript variant 1, mRNA [NM_000125] |
| A_33_P3347911 | 8,79E-07 | down | -3,446456 | ESR2 | Homo sapiens estrogen receptor 2 (ER beta) (ESR2), transcript variant g, mRNA [NM_001271877] |
| A_23_P1585 | 0,008307762 | up | 1,5605787 | ESRRA | Homo sapiens estrogen-related receptor alpha (ESRRA), transcript variant 1, mRNA [NM_004451] |
| A_23_P391857 | 1,70E-05 | down | -3,4335802 | ESRRB | Homo sapiens estrogen-related receptor beta (ESRRB), mRNA [NM_004452] |
| A_24_P132470 | 0,005093907 | up | 1,6161141 | ESYT2 | Homo sapiens extended synaptotagmin-like protein 2 (ESYT2), mRNA [NM_020728] |
| A_33_P3295098 | 0,01704706 | up | 1,7905717 | ESYT3 | extended synaptotagmin-like protein 3 [Source:HGNC Symbol;Acc:HGNC:24295] [ENST00000289135] |
| A_33_P3357954 | 0,001653464 | down | -4,349572 | ETV1 | Homo sapiens ets variant 1 (ETV1), transcript variant 1, mRNA [NM_004956] |
| A_33_P3343106 | 0,025167542 | down | -1,5154656 | ETV2 | Homo sapiens ets variant 2 (ETV2), transcript variant 1, mRNA [NM_014209] |
| A_33_P3370848 | 0,04854229 | down | -1,5260133 | ETV2 | Homo sapiens ets variant 2 (ETV2), transcript variant 1, mRNA [NM_014209] |
| A_33_P3321303 | 0,003607692 | down | -2,0873415 | ETV3 | Homo sapiens ets variant 3 (ETV3), transcript variant 1, mRNA [NM_001145312] |
| A_33_P3324860 | 0,006528006 | down | -1,5857948 | ETV3 | Homo sapiens ets variant 3 (ETV3), transcript variant 1, mRNA [NM_001145312] |
| A_32_P96000 | 0,012005357 | up | 1,5192149 | ETV3L | Homo sapiens ets variant 3-like (ETV3L), mRNA [NM_001004341] |
| A_33_P3356325 | 2,13E-05 | down | -2,4202154 | ETV6 | Homo sapiens ets variant 6 (ETV6), mRNA [NM_001987] |
| A_24_P649388 | 0,034770023 | up | 1,8993859 | EVA1C | Homo sapiens eva-1 homolog C (C. elegans) (EVA1C), transcript variant 1, mRNA [NM_058187] |
| A_23_P78092 | 0,01885017 | down | -2,3012228 | EVI2A | Homo sapiens ecotropic viral integration site 2A (EVI2A), transcript variant 1, mRNA [NM_001003927] |
| A_24_P368943 | 1,03E-05 | down | -2,8284686 | EVX1 | Homo sapiens even-skipped homeobox 1 (EVX1), mRNA [NM_001989] |
| A_19_P00322896 | 0,004588202 | down | -1,6236471 | EWSAT1 | Homo sapiens Ewing sarcoma associated transcript 1 (EWSAT1), long non-coding RNA [NR_026949] |
| A_19_P00321618 | 0,010617553 | down | -1,6290148 | EWSAT1 | Homo sapiens Ewing sarcoma associated transcript 1 (EWSAT1), long non-coding RNA [NR_026949] |
| A_21_P0011328 | 0,004875637 | down | -2,4133215 | EWSAT1 | Homo sapiens Ewing sarcoma associated transcript 1 (EWSAT1), long non-coding RNA [NR_026949] |
| A_19_P00319776 | 0,004002255 | up | 1,5214367 | EWSAT1 | Homo sapiens Ewing sarcoma associated transcript 1 (EWSAT1), long non-coding RNA [NR_026949] |
| A_23_P23303 | 0,022069998 | up | 2,4264603 | EXO1 | Homo sapiens exonuclease 1 (EXO1), transcript variant 3, mRNA [NM_003686] |
| A_23_P214354 | 0,020419754 | up | 1,629443 | EXOC2 | Homo sapiens exocyst complex component 2 (EXOC2), transcript variant 1, mRNA [NM_018303] |
| A_23_P424734 | 1,83E-04 | down | -2,222643 | EXOC3L1 | Homo sapiens exocyst complex component 3-like 1 (EXOC3L1), mRNA [NM_178516] |
| A_24_P7946 | 0,003724593 | down | -1,5062692 | EXOC4 | Homo sapiens exocyst complex component 4 (EXOC4), transcript variant 2, mRNA [NM_001037126] |
| A_23_P14464 | 0,001477657 | down | -1,6260372 | EXOC5 | Homo sapiens exocyst complex component 5 (EXOC5), mRNA [NM_006544] |
| A_23_P415820 | 0,019158224 | up | 1,637326 | EXOC6B | Homo sapiens mRNA for KIAA0919 protein, partial cds. [AB023136] |
| A_33_P3390024 | 0,011119891 | up | 1,6800578 | EXOC7 | Homo sapiens exocyst complex component 7 (EXOC7), transcript variant 7, mRNA [NM_001282314] |
| A_23_P430161 | 0,003501352 | up | 1,5293694 | EXOC8 | Homo sapiens exocyst complex component 8 (EXOC8), mRNA [NM_175876] |
| A_33_P3276813 | 8,35E-04 | down | -2,452158 | EYS | Homo sapiens eyes shut homolog (Drosophila) (EYS), transcript variant 1, mRNA [NM_001142800] |
| A_19_P00321802 | 0,014316908 | up | 2,1438918 | EYS | Homo sapiens eyes shut homolog (Drosophila) (EYS), transcript variant 1, mRNA [NM_001142800] |
| A_19_P00321803 | 0,008098099 | up | 1,5118567 | EYS | Homo sapiens eyes shut homolog (Drosophila) (EYS), transcript variant 1, mRNA [NM_001142800] |
| A_33_P3319302 | 0,002557066 | up | 1,6588356 | F10 | coagulation factor X [Source:HGNC Symbol;Acc:HGNC:3528] [ENST00000410083] |
| A_23_P41314 | 0,004248398 | down | -2,9510171 | F11 | Homo sapiens coagulation factor XI (F11), mRNA [NM_000128] |
| A_32_P140139 | 1,22E-07 | down | -5,374175 | F13A1 | Homo sapiens coagulation factor XIII, A1 polypeptide (F13A1), mRNA [NM_000129] |
| A_33_P3416097 | 0,041908305 | up | 1,5978563 | F13A1 | Homo sapiens coagulation factor XIII, A1 polypeptide (F13A1), mRNA [NM_000129] |

| | | | | | |
|---|---|---|---|---|---|
| A_33_P3232955 | 7,80E-04 | down | -2,064049 | F2RL3 | Homo sapiens coagulation factor II (thrombin) receptor-like 3 (F2RL3), mRNA [NM_003950] |
| A_24_P314451 | 0,004525866 | down | -1,507068 | F8 | Homo sapiens coagulation factor VIII, procoagulant component (F8), transcript variant 1, mRNA [NM_000132] |
| A_33_P3422499 | 0,014774672 | down | -1,6071633 | FAAHP1 | fatty acid amide hydrolase pseudogene 1 [Source:HGNC Symbol;Acc:HGNC:50679] [ENST00000606395] |
| A_24_P558365 | 0,001601827 | up | 2,3001192 | FABP12 | Homo sapiens fatty acid binding protein 12 (FABP12), mRNA [NM_001105281] |
| A_23_P391711 | 0,004679105 | up | 1,5643895 | FABP2 | Homo sapiens fatty acid binding protein 2, intestinal (FABP2), mRNA [NM_000134] |
| A_24_P62783 | 0,010720147 | up | 2,422172 | FABP3 | Homo sapiens fatty acid binding protein 3, muscle and heart (FABP3), mRNA [NM_004102] |
| A_33_P3348348 | 0,035426676 | up | 1,9435431 | FABP9 | Homo sapiens fatty acid binding protein 9, testis (FABP9), mRNA [NM_001080526] |
| A_24_P294719 | 4,91E-04 | up | 1,7289597 | FAF2 | Homo sapiens Fas associated factor family member 2 (FAF2), mRNA [NM_014613] |
| A_33_P3248982 | 0,005491264 | down | -2,2872682 | FAIM2 | Homo sapiens Fas apoptotic inhibitory molecule 2 (FAIM2), mRNA [NM_012306] |
| A_23_P415061 | 0,04715793 | down | -1,7052219 | FAM104B | Homo sapiens family with sequence similarity 104, member B (FAM104B), transcript variant 1, mRNA [NM_138362] |
| A_23_P66827 | 0,03630291 | up | 1,716627 | FAM106A | Homo sapiens family with sequence similarity 106, member A (FAM106A), long non-coding RNA [NR_026809] |
| A_33_P3273125 | 0,03173592 | up | 1,860961 | FAM106CP | Homo sapiens family with sequence similarity 106, member C, pseudogene (FAM106CP), long non-coding RNA [NR_026810] |
| A_24_P164505 | 0,020702105 | up | 1,9274954 | FAM106CP | Homo sapiens family with sequence similarity 106, member C, pseudogene (FAM106CP), long non-coding RNA [NR_026810] |
| A_33_P3367361 | 0,003260006 | up | 1,7052593 | FAM107B | Homo sapiens family with sequence similarity 107, member B (FAM107B), transcript variant 1, mRNA [NM_001282695] |
| A_33_P3330443 | 0,020055912 | down | -1,5823969 | FAM110B | Homo sapiens family with sequence similarity 110, member B (FAM110B), mRNA [NM_147189] |
| A_24_P141629 | 0,004969107 | down | -1,7736809 | FAM111A | Homo sapiens family with sequence similarity 111, member A (FAM111A), transcript variant 1, mRNA [NM_022074] |
| A_23_P41908 | 7,05E-05 | up | 1,8649619 | FAM114A2 | Homo sapiens family with sequence similarity 114, member A2 (FAM114A2), mRNA [NM_018691] |
| A_33_P3424861 | 0,03609376 | down | -1,6719481 | FAM118A | Homo sapiens family with sequence similarity 118, member A (FAM118A), transcript variant 1, mRNA [NM_001104595] |
| A_24_P941505 | 7,10E-05 | up | 1,811265 | FAM120A | Homo sapiens family with sequence similarity 120A (FAM120A), transcript variant 1, mRNA [NM_014612] |
| A_23_P340188 | 0,019435441 | up | 1,7801194 | FAM122C | Homo sapiens family with sequence similarity 122C (FAM122C), transcript variant 2, mRNA [NM_138819] |
| A_33_P3350583 | 0,006401117 | up | 1,6635574 | FAM122C | Homo sapiens family with sequence similarity 122C (FAM122C), transcript variant 2, mRNA [NM_138819] |
| A_23_P344568 | 0,001125732 | up | 1,9776001 | FAM124A | Homo sapiens family with sequence similarity 124A (FAM124A), transcript variant 1, mRNA [NM_145019] |
| A_33_P3307795 | 0,011735971 | down | -2,2420087 | FAM124B | family with sequence similarity 124B [Source:HGNC Symbol;Acc:HGNC:26224] [ENST00000243806] |
| A_24_P233786 | 0,008132864 | down | -2,2358842 | FAM129A | Homo sapiens family with sequence similarity 129, member A (FAM129A), mRNA [NM_052966] |
| A_33_P3226167 | 0,008619238 | up | 1,8443828 | FAM129B | Homo sapiens family with sequence similarity 129, member B (FAM129B), transcript variant 1, mRNA [NM_022833] |
| A_23_P345799 | 3,56E-05 | down | -3,2906842 | FAM129C | Homo sapiens family with sequence similarity 129, member C (FAM129C), transcript variant 1, mRNA [NM_173544] |
| A_24_P169073 | 0,006848342 | down | -1,7508674 | FAM131C | Homo sapiens family with sequence similarity 131, member C (FAM131C), mRNA [NM_182623] |
| A_32_P75792 | 0,004318236 | up | 1,6423882 | FAM132A | Homo sapiens family with sequence similarity 132, member A (FAM132A), mRNA [NM_001014980] |
| A_24_P247536 | 1,20E-04 | down | -2,7078838 | FAM133B | Homo sapiens family with sequence similarity 133, member B (FAM133B), transcript variant 2, mRNA [NM_001040057] |
| A_24_P158065 | 0,001728036 | up | 1,527433 | FAM133B | Homo sapiens family with sequence similarity 133, member B (FAM133B), transcript variant 2, mRNA [NM_001040057] |
| A_32_P233938 | 0,003294548 | up | 1,5074688 | FAM133B | Homo sapiens family with sequence similarity 133, member B (FAM133B), transcript variant 2, mRNA [NM_001040057] |
| A_32_P923011 | 6,32E-04 | down | -2,1686156 | FAM138A | Homo sapiens family with sequence similarity 138, member A (FAM138A), long non-coding RNA [NR_026818] |
| A_21_P0001906 | 5,43E-04 | down | -2,9384544 | FAM138D | family with sequence similarity 138, member D [Source:EntrezGene;Acc:677784] [ENST00000546223] |
| A_33_P3691916 | 0,005662591 | up | 1,7100301 | FAM13A | Homo sapiens family with sequence similarity 13, member A (FAM13A), transcript variant 1, mRNA [NM_014883] |
| A_33_P3218905 | 0,02768031 | up | 1,6890384 | FAM13A-AS1 | Homo sapiens FAM13A antisense RNA 1 (FAM13A-AS1), long non-coding RNA [NR_002806] |
| A_24_P935794 | 0,030304937 | up | 2,239764 | FAM150A | Homo sapiens family with sequence similarity 150, member A (FAM150A), mRNA [NM_207413] |
| A_33_P3320197 | 0,021728544 | down | -1,7152566 | FAM150B | Homo sapiens family with sequence similarity 150, member B (FAM150B), mRNA [NM_001002919] |
| A_33_P3728979 | 0,02997819 | down | -1,7160997 | FAM151B | Homo sapiens family with sequence similarity 151, member B (FAM151B), mRNA [NM_205548] |
| A_33_P3411414 | 0,029891273 | up | 1,5925311 | FAM156B | Homo sapiens family with sequence similarity 156, member B (FAM156B), mRNA [NM_001099684] |
| A_24_P190873 | 0,013957304 | up | 3,0950873 | FAM163A | Homo sapiens family with sequence similarity 163, member A (FAM163A), mRNA [NM_173509] |
| A_33_P3268378 | 0,005588337 | down | -2,0505102 | FAM167A | Homo sapiens family with sequence similarity 167, member A (FAM167A), mRNA [NM_053279] |
| A_33_P3354076 | 0,013553762 | down | -1,8828101 | FAM172BP | Homo sapiens family with sequence similarity 172, member B, pseudogene (FAM172BP), non-coding RNA [NR_036433] |
| A_33_P3234138 | 2,92E-07 | down | -3,9809952 | FAM172BP | Homo sapiens family with sequence similarity 172, member B, pseudogene (FAM172BP), non-coding RNA [NR_036433] |
| A_21_P0008784 | 0,018996093 | down | -1,9660755 | FAM174B | family with sequence similarity 174, member B [Source:HGNC Symbol;Acc:HGNC:34339] [ENST00000557545] |
| A_33_P3367396 | 8,08E-04 | up | 4,04873 | FAM177B | Homo sapiens family with sequence similarity 177, member B (FAM177B), mRNA [NM_207468] |
| A_24_P231546 | 0,003861708 | down | -1,9854071 | FAM178B | Homo sapiens family with sequence similarity 178, member B (FAM178B), transcript variant B, mRNA [NM_016490] |
| A_24_P595717 | 0,002702448 | up | 2,1180608 | FAM179A | Homo sapiens family with sequence similarity 179, member A (FAM179A), mRNA [NM_199280] |
| A_21_P0012164 | 0,019883346 | down | -1,8290044 | FAM182B | Homo sapiens family with sequence similarity 182, member B (FAM182B), transcript variant 1, long non-coding RNA [NR_026714] |
| A_23_P301572 | 0,001022237 | up | 2,5910285 | FAM184B | Homo sapiens family with sequence similarity 184, member B (FAM184B), mRNA [NM_015688] |
| A_32_P225415 | 0,002356326 | down | -2,5659325 | FAM187B | Homo sapiens family with sequence similarity 187, member B (FAM187B), mRNA [NM_152481] |
| A_23_P213863 | 0,004919835 | up | 1,6486899 | FAM193B | Homo sapiens family with sequence similarity 193, member B (FAM193B), transcript variant 3, mRNA [NM_001190946] |
| A_32_P202013 | 0,001994301 | up | 1,9106536 | FAM196A | Homo sapiens family with sequence similarity 196, member A (FAM196A), mRNA [NM_001039762] |
| A_33_P3375636 | 0,046729024 | up | 2,5227191 | FAM196B | Homo sapiens family with sequence similarity 196, member B (FAM196B), mRNA [NM_001129891] |
| A_33_P3407494 | 0,007401437 | up | 2,4989254 | FAM205A | Homo sapiens family with sequence similarity 205, member A (FAM205A), mRNA [NM_001141917] |
| A_32_P108254 | 0,015012047 | down | -2,2508464 | FAM20A | Homo sapiens family with sequence similarity 20, member A (FAM20A), transcript variant 1, mRNA [NM_017565] |
| A_24_P352952 | 0,005664714 | up | 1,6598387 | FAM20A | Homo sapiens family with sequence similarity 20, member A (FAM20A), transcript variant 1, mRNA [NM_017565] |
| A_33_P3346635 | 0,0034861 | down | -3,0008318 | FAM20C | Homo sapiens cDNA FLJ43291 fis, clone MESAN2015515. [AK125281] |
| A_24_P336577 | 0,002253953 | down | -2,1234443 | FAM212B | Homo sapiens family with sequence similarity 212, member B (FAM212B), transcript variant 1, mRNA [NM_019099] |
| A_23_P99853 | 0,001009749 | up | 1,9668612 | FAM214A | Homo sapiens family with sequence similarity 214, member A (FAM214A), transcript variant 1, mRNA [NM_019600] |
| A_33_P3348204 | 0,016489772 | down | -1,76026 | FAM218A | Homo sapiens family with sequence similarity 218, member A (FAM218A), mRNA [NM_153027] |
| A_23_P372771 | 4,13E-04 | up | 1,8678917 | FAM219B | Homo sapiens family with sequence similarity 219, member B (FAM219B), mRNA [NM_020447] |

| | | | | | |
|---|---|---|---|---|---|
| A_33_P3279124 | 6,94E-04 | down | -2,241831 | FAM21C | Homo sapiens family with sequence similarity 21, member C (FAM21C), transcript variant 3, mRNA [NM_001169107] |
| A_23_P376799 | 0,001749602 | up | 1,5378025 | FAM21C | Homo sapiens family with sequence similarity 21, member C (FAM21C), transcript variant 1, mRNA [NM_015262] |
| A_21_P0014781 | 0,023249382 | down | -1,5580695 | FAM220A | Homo sapiens family with sequence similarity 220, member A (FAM220A), mRNA [NM_001037163] |
| A_33_P3371402 | 0,031571973 | up | 1,5144013 | FAM223A | Homo sapiens family with sequence similarity 223, member A (non-protein coding) (FAM223A), long non-coding RNA [NR_027401] |
| A_33_P3347037 | 0,01996657 | down | -1,7251548 | FAM226A | Homo sapiens family with sequence similarity 226, member A (non-protein coding) (FAM226A), long non-coding RNA [NR_026595] |
| A_33_P3825317 | 0,01775975 | up | 1,6009078 | FAM227A | Homo sapiens family with sequence similarity 227, member A (FAM227A), transcript variant 2, mRNA [NM_001291030] |
| A_33_P3217490 | 3,54E-06 | down | -4,2254405 | FAM230B | Homo sapiens family with sequence similarity 230, member B (non-protein coding) (FAM230B), long non-coding RNA [NR_108107] |
| A_33_P3307535 | 0,002818223 | up | 3,7065685 | FAM230C | Homo sapiens family with sequence similarity 230, member C (FAM230C), long non-coding RNA [NR_027278] |
| A_33_P3348794 | 0,00178906 | down | -2,6549115 | FAM231A | Homo sapiens family with sequence similarity 231, member A (FAM231A), mRNA [NM_001282321] |
| A_21_P0000707 | 0,003016254 | up | 1,569634 | FAM24B | Homo sapiens family with sequence similarity 24, member B (FAM24B), transcript variant 1, mRNA [NM_152644] |
| A_33_P3301219 | 0,021696292 | down | -2,624242 | FAM27C | Homo sapiens family with sequence similarity 27, member C (FAM27C), long non-coding RNA [NR_027421] |
| A_24_P342096 | 0,005607381 | down | -1,8374298 | FAM27C | Homo sapiens family with sequence similarity 27, member C (FAM27C), long non-coding RNA [NR_027421] |
| A_32_P942508 | 0,010624141 | up | 2,2929559 | FAM27L | Homo sapiens family with sequence similarity 27-like (FAM27L), long non-coding RNA [NR_028336] |
| A_21_P0013755 | 0,030722627 | up | 1,6322308 | FAM3C | Homo sapiens family with sequence similarity 3, member C (FAM3C), transcript variant 1, mRNA [NM_014888] |
| A_33_P3410589 | 0,006085541 | up | 1,7659036 | FAM43A | Homo sapiens family with sequence similarity 43, member A (FAM43A), mRNA [NM_153690] |
| A_33_P3290573 | 0,044492114 | down | -1,590135 | FAM46A | Homo sapiens family with sequence similarity 46, member A (FAM46A), mRNA [NM_017633] |
| A_23_P12199 | 0,006724544 | up | 1,7883741 | FAM46B | Homo sapiens family with sequence similarity 46, member B (FAM46B), mRNA [NM_052943] |
| A_32_P105381 | 0,004591356 | up | 1,5013219 | FAM47B | Homo sapiens family with sequence similarity 47, member B (FAM47B), mRNA [NM_152631] |
| A_23_P163711 | 0,006363601 | down | -2,2620392 | FAM57B | Homo sapiens family with sequence similarity 57, member B (FAM57B), mRNA [NM_031478] |
| A_23_P379327 | 0,028445618 | up | 1,5911783 | FAM63B | Homo sapiens family with sequence similarity 63, member B (FAM63B), transcript variant 1, mRNA [NM_001040450] |
| A_33_P3276918 | 0,011296997 | up | 1,725439 | FAM64A | Homo sapiens family with sequence similarity 64, member A (FAM64A), transcript variant 2, mRNA [NM_001195228] |
| A_33_P3316410 | 0,027176771 | down | -1,5786875 | FAM66A | Homo sapiens family with sequence similarity 66, member A (FAM66A), long non-coding RNA [NR_026789] |
| A_21_P0013467 | 0,008598109 | down | -2,208449 | FAM66C | Homo sapiens family with sequence similarity 66, member C (FAM66C), long non-coding RNA [NR_026788] |
| A_32_P203404 | 0,005996903 | up | 1,5676231 | FAM69C | Homo sapiens family with sequence similarity 69, member C (FAM69C), mRNA [NM_001044369] |
| A_23_P328145 | 0,04128869 | up | 2,4037752 | FAM71D | Homo sapiens family with sequence similarity 71, member D (FAM71D), mRNA [NM_173526] |
| A_24_P349807 | 0,017690929 | down | -1,7958219 | FAM71E2 | Homo sapiens family with sequence similarity 71, member E2 (FAM71E2), mRNA [NM_001145402] |
| A_23_P134204 | 0,031037144 | up | 2,2574735 | FAM71F1 | Homo sapiens family with sequence similarity 71, member F1 (FAM71F1), transcript variant 1, mRNA [NM_032599] |
| A_32_P112894 | 6,67E-04 | down | -1,8343762 | FAM76A | Homo sapiens family with sequence similarity 76, member A (FAM76A), transcript variant 3, mRNA [NM_152660] |
| A_23_P321473 | 0,00437168 | up | 1,5002732 | FAM76A | Homo sapiens family with sequence similarity 76, member A (FAM76A), transcript variant 3, mRNA [NM_152660] |
| A_23_P360354 | 0,030963572 | up | 1,5352794 | FAM81B | Homo sapiens family with sequence similarity 81, member B (FAM81B), mRNA [NM_152548] |
| A_23_P216052 | 0,016607704 | up | 1,6889387 | FAM83A | Homo sapiens family with sequence similarity 83, member A (FAM83A), transcript variant 1, mRNA [NM_032899] |
| A_24_P43810 | 5,89E-04 | up | 2,3330913 | FAM83A | Homo sapiens family with sequence similarity 83, member A (FAM83A), transcript variant 2, mRNA [NM_207006] |
| A_23_P421175 | 0,002968998 | up | 1,5633337 | FAM83H | Homo sapiens family with sequence similarity 83, member H (FAM83H), mRNA [NM_198488] |
| A_23_P306215 | 0,037779916 | up | 1,5431579 | FAM84A | Homo sapiens family with sequence similarity 84, member A (FAM84A), mRNA [NM_145175] |
| A_33_P3421203 | 0,04362825 | down | -1,5116765 | FAM86B3P | Homo sapiens family with sequence similarity 86, member B3, pseudogene (FAM86B3P), transcript variant 1, non-coding RNA [NR_024362] |
| A_21_P0011081 | 0,01365763 | down | -1,5429541 | FAM86C1 | Homo sapiens family with sequence similarity 86, member C1 (FAM86C1), transcript variant 3, mRNA [NM_001099653] |
| A_33_P3239087 | 0,041032266 | down | -1,8037536 | FAM87A | Homo sapiens family with sequence similarity 87, member A (FAM87A), long non-coding RNA [NR_103537] |
| A_33_P3330906 | 0,014818569 | up | 1,9203304 | FAM87B | Homo sapiens family with sequence similarity 87, member B (FAM87B), long non-coding RNA [NR_103536] |
| A_23_P133648 | 0,034307715 | up | 1,6330312 | FAM8A1 | Homo sapiens family with sequence similarity 8, member A1 (FAM8A1), mRNA [NM_016255] |
| A_33_P3212630 | 0,001747256 | down | -3,3527334 | FAM90A1 | Homo sapiens family with sequence similarity 90, member A1 (FAM90A1), mRNA [NM_018088] |
| A_21_P0003568 | 0,001939059 | up | 1,8156425 | FAM90A1 | Homo sapiens family with sequence similarity 90, member A1 (FAM90A1), mRNA [NM_018088] |
| A_23_P394323 | 0,043266747 | up | 1,9558101 | FAM98B | Homo sapiens cDNA FLJ38426 fis, clone FEBRA2012507. [AK095745] |
| A_33_P3378785 | 0,024480637 | down | -1,5395018 | FAM99A | family with sequence similarity 99, member A (non-protein coding) [Source:HGNC Symbol;Acc:HGNC:32368] [ENST00000538190] |
| A_32_P12202 | 0,01610589 | up | 1,6216207 | FAM9C | family with sequence similarity 9, member C [Source:HGNC Symbol;Acc:HGNC:18405] [ENST00000333995] |
| A_23_P206441 | 0,004933883 | up | 1,9991549 | FANCA | Homo sapiens Fanconi anemia, complementation group A (FANCA), transcript variant 1, mRNA [NM_000135] |
| A_23_P143994 | 0,04938628 | up | 1,629476 | FANCD2 | Homo sapiens Fanconi anemia, complementation group D2 (FANCD2), transcript variant 2, mRNA [NM_001018115] |
| A_23_P318989 | 0,004417549 | down | -1,5394521 | FANCD2OS | Homo sapiens FANCD2 opposite strand (FANCD2OS), transcript variant 1, mRNA [NM_173472] |
| A_33_P3388331 | 0,016305903 | up | 1,5709159 | FARP1 | Homo sapiens FERM, RhoGEF (ARHGEF) and pleckstrin domain protein 1 (chondrocyte-derived) (FARP1), transcript variant 1, mRNA [NM_005766] |
| A_24_P16833 | 0,020856353 | up | 1,6721623 | FAT3 | Homo sapiens FAT atypical cadherin 3 (FAT3), mRNA [NM_001008781] |
| A_23_P22565 | 0,041596003 | up | 1,845195 | FATE1 | Homo sapiens fetal and adult testis expressed 1 (FATE1), mRNA [NM_033085] |
| A_23_P501831 | 0,00733359 | down | -1,7351876 | FAXDC2 | Homo sapiens fatty acid hydroxylase domain containing 2 (FAXDC2), mRNA [NM_032385] |
| A_24_P87458 | 0,035986487 | up | 2,1667743 | FBF1 | Homo sapiens Fas (TNFRSF6) binding factor 1 (FBF1), mRNA [NM_001080542] |
| A_23_P337934 | 0,04515116 | up | 1,5523353 | FBLIM1 | Homo sapiens filamin binding LIM protein 1 (FBLIM1), transcript variant 1, mRNA [NM_017556] |
| A_33_P3268954 | 0,03212055 | down | -1,6624596 | FBLL1 | Homo sapiens fibrillarin-like 1 (FBLL1), non-coding RNA [NR_024356] |
| A_33_P3334248 | 0,002271559 | down | -1,6398602 | FBRS | fibrosin [Source:HGNC Symbol;Acc:HGNC:20442] [ENST00000484152] |
| A_21_P0014038 | 7,27E-04 | down | -1,9793236 | FBRSL1 | Homo sapiens fibrosin-like 1 (FBRSL1), mRNA [NM_001142641] |
| A_24_P280497 | 7,37E-07 | down | -4,9480095 | FBRSL1 | Homo sapiens fibrosin-like 1 (FBRSL1), mRNA [NM_001142641] |
| A_23_P379071 | 9,06E-04 | down | -2,8962922 | FBXL13 | Homo sapiens F-box and leucine-rich repeat protein 13 (FBXL13), transcript variant 1, mRNA [NM_145032] |
| A_23_P406385 | 0,005929473 | up | 1,8685771 | FBXL16 | Homo sapiens F-box and leucine-rich repeat protein 16 (FBXL16), mRNA [NM_153350] |
| A_24_P7600 | 5,01E-05 | down | -3,129352 | FBXL7 | Homo sapiens F-box and leucine-rich repeat protein 7 (FBXL7), transcript variant 1, mRNA [NM_012304] |
| A_33_P3273822 | 0,013145187 | up | 1,5259279 | FBXO11 | F-box protein 11 [Source:HGNC Symbol;Acc:HGNC:13590] [ENST00000434234] |

| | | | | | |
|---|---|---|---|---|---|
| A_23_P395911 | 0,009411264 | down | -1,6269808 | FBXO17 | F-box protein 17 [Source:HGNC Symbol;Acc:HGNC:18754] [ENST00000601394] |
| A_24_P13715 | 0,008266107 | down | -2,0570483 | FBXO22 | Homo sapiens F-box protein 22 (FBXO22), transcript variant 1, mRNA [NM_147188] |
| A_33_P3352941 | 0,004730377 | down | -2,0010238 | FBXO22-AS1 | Homo sapiens FBXO22 antisense RNA 1 (FBXO22-AS1), long non-coding RNA [NR_003136] |
| A_33_P3319593 | 0,022781504 | down | -1,8319618 | FBXO31 | Homo sapiens F-box protein 31 (FBXO31), transcript variant 1, mRNA [NM_024735] |
| A_24_P41021 | 0,001233473 | up | 1,5855432 | FBXO33 | Homo sapiens F-box protein 33 (FBXO33), mRNA [NM_203301] |
| A_33_P3237225 | 4,28E-04 | up | 1,8605698 | FBXO34 | Homo sapiens F-box protein 34 (FBXO34), transcript variant 2, mRNA [NM_152231] |
| A_23_P146811 | 8,67E-04 | up | 1,5966741 | FBXO38 | Homo sapiens F-box protein 38 (FBXO38), transcript variant 1, mRNA [NM_030793] |
| A_23_P27744 | 0,003682372 | up | 1,758386 | FBXO46 | Homo sapiens F-box protein 46 (FBXO46), mRNA [NM_001080469] |
| A_33_P3214745 | 7,46E-04 | down | -2,5513668 | FBXO48 | Homo sapiens F-box protein 48 (FBXO48), mRNA [NM_001024680] |
| A_33_P3268686 | 0,033927105 | down | -1,9083179 | FBXW12 | Homo sapiens F-box and WD repeat domain containing 12 (FBXW12), transcript variant 1, mRNA [NM_207102] |
| A_33_P3382606 | 9,33E-05 | down | -6,1770062 | FBXW12 | Homo sapiens F-box and WD repeat domain containing 12 (FBXW12), transcript variant 1, mRNA [NM_207102] |
| A_33_P3364488 | 0,044976752 | up | 2,3534455 | FCAMR | Homo sapiens Fc receptor, IgA, IgM, high affinity (FCAMR), transcript variant 3, mRNA [NM_001170631] |
| A_24_P348265 | 0,011067308 | up | 1,5136819 | FCAR | Homo sapiens Fc fragment of IgA, receptor for (FCAR), transcript variant 3, mRNA [NM_133271] |
| A_33_P3272628 | 0,032828458 | down | -1,8110564 | FCER1G | Fc fragment of IgE, high affinity I, receptor for; gamma polypeptide [Source:HGNC Symbol;Acc:HGNC:3611] [ENST00000367992] |
| A_23_P349083 | 1,31E-04 | up | 2,490516 | FCHO2 | Homo sapiens FCH domain only 2 (FCHO2), transcript variant 1, mRNA [NM_138782] |
| A_23_P157875 | 8,58E-04 | down | -3,77583 | FCN1 | Homo sapiens ficolin (collagen/fibrinogen domain containing) 1 (FCN1), mRNA [NM_002003] |
| A_23_P216756 | 0,01066284 | up | 2,2674217 | FCN2 | Homo sapiens ficolin (collagen/fibrinogen domain containing lectin) 2 (FCN2), transcript variant SV0, mRNA [NM_004108] |
| A_33_P3294504 | 0,001223975 | down | -2,2018769 | FCRL1 | Homo sapiens Fc receptor-like 1 (FCRL1), transcript variant 2, mRNA [NM_001159397] |
| A_23_P103803 | 0,023062773 | up | 1,6132985 | FCRL3 | Homo sapiens Fc receptor-like 3 (FCRL3), mRNA [NM_052939] |
| A_33_P3335511 | 0,00894197 | down | -1,9285803 | FCRL5 | Homo sapiens Fc receptor-like 5 (FCRL5), transcript variant 2, mRNA [NM_001195388] |
| A_33_P3363933 | 1,52E-05 | down | -5,8658667 | FCRL6 | Homo sapiens Fc receptor-like 6 (FCRL6), transcript variant 2, mRNA [NM_001284217] |
| A_23_P46039 | 0,010873776 | down | -3,3095942 | FCRLA | Homo sapiens Fc receptor-like A (FCRLA), transcript variant 2, mRNA [NM_032738] |
| A_32_P151933 | 3,10E-04 | down | -3,2773395 | FECH | Homo sapiens ferrochelatase (FECH), transcript variant 1, mRNA [NM_001012515] |
| A_23_P151970 | 0,019278765 | up | 1,9405661 | FEM1B | Homo sapiens fem-1 homolog b (C. elegans) (FEM1B), mRNA [NM_015322] |
| A_19_P00322654 | 0,010244543 | up | 1,7101922 | FENDRR | Homo sapiens FOXF1 adjacent non-coding developmental regulatory RNA (FENDRR), transcript variant 2, long non-coding RNA [NR_036444] |
| A_19_P00322440 | 0,011039838 | up | 1,5175792 | FENDRR | Homo sapiens FOXF1 adjacent non-coding developmental regulatory RNA (FENDRR), transcript variant 2, long non-coding RNA [NR_036444] |
| A_33_P3613688 | 0,027775241 | down | -1,6935943 | FER1L6-AS2 | 601564882F1 NIH_MGC_20 Homo sapiens cDNA clone IMAGE:3925989 5', mRNA sequence [BE729513] |
| A_33_P3262949 | 0,014979539 | up | 1,9489259 | FERMT1 | fermitin family member 1 [Source:HGNC Symbol;Acc:HGNC:15889] [ENST00000378844] |
| A_33_P3217009 | 0,014866857 | down | -1,6326511 | FEZF1-AS1 | Homo sapiens FEZF1 antisense RNA 1 (FEZF1-AS1), long non-coding RNA [NR_036484] |
| A_24_P405631 | 0,008987518 | up | 1,5396821 | FEZF2 | Homo sapiens FEZ family zinc finger 2 (FEZF2), mRNA [NM_018008] |
| A_23_P397391 | 3,95E-04 | down | -3,838275 | FFAR2 | Homo sapiens free fatty acid receptor 2 (FFAR2), mRNA [NM_005306] |
| A_33_P3298810 | 0,002698344 | up | 1,6344917 | FFAR3 | Homo sapiens free fatty acid receptor 3 (FFAR3), mRNA [NM_005304] |
| A_24_P129588 | 0,006758017 | down | -2,2586274 | FFAR4 | Homo sapiens free fatty acid receptor 4 (FFAR4), transcript variant 1, mRNA [NM_181745] |
| A_24_P282251 | 0,00283166 | up | 1,5249639 | FGA | Homo sapiens fibrinogen alpha chain (FGA), transcript variant alpha, mRNA [NM_021871] |
| A_33_P3345886 | 0,007376475 | down | -2,216262 | FGD2 | FYVE, RhoGEF and PH domain containing 2 [Source:HGNC Symbol;Acc:HGNC:3664] [ENST00000493635] |
| A_24_P937325 | 0,002067509 | up | 1,9224793 | FGD4 | FYVE, RhoGEF and PH domain containing 4 [Source:HGNC Symbol;Acc:HGNC:19125] [ENST00000472289] |
| A_21_P0014935 | 9,60E-04 | down | -1,6134119 | FGD5-AS1 | Homo sapiens FGD5 antisense RNA 1 (FGD5-AS1), transcript variant 1, long non-coding RNA [NR_046251] |
| A_33_P3333282 | 2,94E-04 | up | 1,8044817 | FGF11 | fibroblast growth factor 11 [Source:HGNC Symbol;Acc:HGNC:3667] [ENST00000293829] |
| A_23_P429363 | 0,026263436 | up | 2,443448 | FGF17 | Homo sapiens fibroblast growth factor 17 (FGF17), mRNA [NM_003867] |
| A_23_P412389 | 0,031494185 | up | 1,5014063 | FGF18 | Homo sapiens fibroblast growth factor 18 (FGF18), mRNA [NM_003862] |
| A_33_P3379886 | 0,016049786 | down | -2,9597058 | FGF2 | Homo sapiens fibroblast growth factor 2 (basic) (FGF2), mRNA [NM_002006] |
| A_33_P3419696 | 0,014554832 | down | -4,3174977 | FGF2 | Homo sapiens fibroblast growth factor 2 (basic) (FGF2), mRNA [NM_002006] |
| A_23_P395404 | 0,002446821 | up | 1,5789539 | FGF20 | Homo sapiens fibroblast growth factor 20 (FGF20), mRNA [NM_019851] |
| A_24_P355720 | 2,02E-04 | down | -3,3695152 | FGF4 | Homo sapiens fibroblast growth factor 4 (FGF4), mRNA [NM_002007] |
| A_23_P390734 | 0,022992566 | up | 1,640866 | FGFR1OP2 | Homo sapiens FGFR1 oncogene partner 2 (FGFR1OP2), transcript variant 1, mRNA [NM_015633] |
| A_23_P212830 | 0,0019482 | up | 2,7260854 | FGFR3 | Homo sapiens fibroblast growth factor receptor 3 (FGFR3), transcript variant 1, mRNA [NM_000142] |
| A_23_P500501 | 0,001470698 | up | 2,3992887 | FGFR3 | Homo sapiens fibroblast growth factor receptor 3 (FGFR3), transcript variant 1, mRNA [NM_000142] |
| A_23_P92754 | 1,05E-06 | down | -3,103521 | FGFR4 | Homo sapiens fibroblast growth factor receptor 4 (FGFR4), transcript variant 3, mRNA [NM_213647] |
| A_23_P20484 | 1,52E-06 | down | -3,35292 | FGL1 | Homo sapiens fibrinogen-like 1 (FGL1), transcript variant 4, mRNA [NM_201553] |
| A_23_P328259 | 0,004330946 | up | 1,6456351 | FHDC1 | Homo sapiens FH2 domain containing 1 (FHDC1), mRNA [NM_033393] |
| A_33_P3849275 | 5,33E-06 | down | -2,8817616 | FHL1 | Homo sapiens four and a half LIM domains 1 (FHL1), transcript variant 4, mRNA [NM_001159704] |
| A_33_P3398392 | 0,00401109 | up | 1,7874069 | FHL1 | four and a half LIM domains 1 [Source:HGNC Symbol;Acc:HGNC:3702] [ENST00000370674] |
| A_33_P3237552 | 0,03620623 | down | -1,7886882 | FIBCD1 | Homo sapiens fibrinogen C domain containing 1 (FIBCD1), transcript variant 1, mRNA [NM_032843] |
| A_23_P45185 | 0,04419588 | down | -2,0750341 | FIGF | Homo sapiens c-fos induced growth factor (vascular endothelial growth factor D) (FIGF), mRNA [NM_004469] |
| A_33_P3315944 | 0,015261022 | up | 1,5539688 | FIGLA | Homo sapiens folliculogenesis specific basic helix-loop-helix (FIGLA), mRNA [NM_001004311] |
| A_24_P203407 | 8,90E-04 | down | -1,8672665 | FITM2 | Homo sapiens fat storage-inducing transmembrane protein 2 (FITM2), mRNA [NM_001080472] |
| A_21_P0000010 | 2,16E-04 | up | 4,4326086 | FKBP5 | Homo sapiens FK506 binding protein 5 (FKBP5), transcript variant 4, mRNA [NM_001145777] |
| A_24_P38081 | 1,37E-05 | up | 7,811647 | FKBP5 | Homo sapiens FK506 binding protein 5 (FKBP5), transcript variant 1, mRNA [NM_004117] |
| A_33_P3259092 | 0,010087859 | down | -1,8276203 | FKTN | Homo sapiens fukutin (FKTN), transcript variant 1, mRNA [NM_001079802] |
| A_33_P3359763 | 0,020268353 | up | 2,289631 | FLCN | folliculin [Source:HGNC Symbol;Acc:HGNC:27310] [ENST00000466317] |
| A_24_P173124 | 0,001927992 | up | 2,1148808 | FLCN | Homo sapiens folliculin (FLCN), transcript variant 1, mRNA [NM_144997] |

| | | | | | |
|---|---|---|---|---|---|
| A_21_P0014075 | 0,003158539 | down | -2,2762873 | FLG-AS1 | PREDICTED: Homo sapiens FLG antisense RNA 1 (FLG-AS1), misc_RNA [XR_108352] |
| A_33_P3335740 | 0,020990472 | up | 1,5296347 | FLJ10038 | Homo sapiens uncharacterized protein FLJ10038 (FLJ10038), long non-coding RNA [NR_026891] |
| A_24_P516246 | 0,015995814 | down | -1,7149222 | FLJ12825 | Homo sapiens uncharacterized LOC440101 (FLJ12825), long non-coding RNA [NR_026655] |
| A_33_P3367606 | 0,01777231 | down | -2,1124868 | FLJ13224 | Homo sapiens uncharacterized LOC79857 (FLJ13224), long non-coding RNA [NR_026806] |
| A_19_P00322224 | 0,003457243 | down | -1,6482079 | FLJ16779 | Homo sapiens uncharacterized LOC100192386 (FLJ16779), long non-coding RNA [NR_024389] |
| A_33_P3394584 | 0,004035913 | down | -2,9776568 | FLJ16779 | Homo sapiens uncharacterized LOC100192386 (FLJ16779), long non-coding RNA [NR_024389] |
| A_23_P42684 | 0,019135697 | down | -2,2966716 | FLJ20712 | PREDICTED: Homo sapiens uncharacterized FLJ20712 (FLJ20712), misc_RNA [XR_108766] |
| A_33_P3301752 | 9,41E-04 | down | -2,1593962 | FLJ21369 | Homo sapiens cDNA: FLJ21369 fis, clone COL03091. [AK025022] |
| A_33_P3257518 | 0,03100291 | up | 1,5447198 | FLJ22447 | Homo sapiens uncharacterized LOC400221 (FLJ22447), long non-coding RNA [NR_039985] |
| A_33_P3555368 | 0,023832219 | up | 1,6851894 | FLJ26086 | Homo sapiens cDNA FLJ26086 fis, clone RCT05358. [AK129597] |
| A_33_P3255314 | 0,040211808 | up | 1,5473405 | FLJ26245 | Homo sapiens uncharacterized LOC400533 (FLJ26245), long non-coding RNA [NR_033985] |
| A_21_P0000908 | 0,00369099 | down | -3,0145388 | FLJ27354 | Homo sapiens uncharacterized LOC400761 (FLJ27354), long non-coding RNA [NR_033981] |
| A_19_P00320314 | 0,017562265 | down | -1,5386207 | FLJ32255 | Homo sapiens uncharacterized LOC643977 (FLJ32255), long non-coding RNA [NR_104643] |
| A_19_P00320757 | 1,30E-04 | down | -2,5267632 | FLJ32255 | Homo sapiens uncharacterized LOC643977 (FLJ32255), long non-coding RNA [NR_104643] |
| A_24_P307724 | 7,91E-06 | down | -3,1189468 | FLJ33360 | Homo sapiens FLJ33360 protein (FLJ33360), long non-coding RNA [NR_028351] |
| A_33_P3338215 | 0,006682403 | up | 1,6131512 | FLJ33544 | Homo sapiens cDNA FLJ33544 fis, clone BRAMY2008333. [AK090863] |
| A_33_P3377763 | 3,27E-04 | down | -1,9197592 | FLJ36000 | Homo sapiens uncharacterized FLJ36000 (FLJ36000), long non-coding RNA [NR_027084] |
| A_33_P3347099 | 0,04967377 | down | -2,3025231 | FLJ36000 | Homo sapiens uncharacterized FLJ36000 (FLJ36000), long non-coding RNA [NR_027084] |
| A_21_P0003889 | 3,89E-06 | down | -5,948334 | FLJ36777 | uncharacterized LOC730971 [Source:EntrezGene;Acc:730971] [ENST00000504402] |
| A_21_P0007120 | 0,013756256 | up | 1,6984112 | FLJ37035 | uncharacterized LOC399821 [Source:EntrezGene;Acc:399821] [ENST00000531977] |
| A_33_P3249893 | 2,27E-04 | down | -3,184977 | FLJ37786 | Homo sapiens cDNA FLJ37786 fis, clone BRHIP2028480. [AK095105] |
| A_24_P194661 | 0,003836976 | up | 1,7033844 | FLJ38379 | PREDICTED: Homo sapiens uncharacterized FLJ38379 (FLJ38379), misc_RNA [XR_108456] |
| A_33_P3325871 | 0,018945675 | down | -1,5846035 | FLJ38668 | PREDICTED: Homo sapiens uncharacterized LOC644903 (FLJ38668), misc_RNA [XR_244911] |
| A_33_P3347437 | 0,039484907 | down | -1,6276728 | FLJ39080 | Homo sapiens uncharacterized LOC441355 (FLJ39080), long non-coding RNA [NR_033830] |
| A_33_P3325843 | 0,004427913 | down | -1,8404075 | FLJ40039 | Homo sapiens cDNA FLJ40039 fis, clone SYNOV2000397. [AK097358] |
| A_33_P3282291 | 1,44E-04 | down | -2,8809133 | FLJ40039 | Homo sapiens cDNA FLJ40039 fis, clone SYNOV2000397. [AK097358] |
| A_19_P00810894 | 9,68E-04 | down | -1,9498805 | FLJ40288 | uncharacterized FLJ40288 [Source:EntrezGene;Acc:286023] [ENST00000573229] |
| A_24_P929322 | 0,007296097 | up | 1,7061266 | FLJ42393 | Homo sapiens uncharacterized LOC401105 (FLJ42393), long non-coding RNA [NR_024413] |
| A_21_P0008871 | 0,011668701 | up | 1,8319113 | FLJ42627 | Homo sapiens uncharacterized LOC645644 (FLJ42627), long non-coding RNA [NR_024492] |
| A_33_P3229452 | 0,00183783 | down | -2,3216498 | FLJ44477 | Homo sapiens cDNA FLJ44477 fis, clone UTERU2031703. [AK126441] |
| A_33_P3229171 | 0,0102221 | up | 1,6474235 | FLJ44635 | Homo sapiens TPT1-like protein (FLJ44635), mRNA [NM_207422] |
| A_33_P3325029 | 0,043246366 | up | 1,770173 | FLJ44674 | Homo sapiens cDNA FLJ44674 fis, clone BRACE3007649. [AK128747] |
| A_33_P3356525 | 0,017334156 | up | 1,9351741 | FLJ45482 | Homo sapiens cDNA FLJ45482 fis, clone BRTHA2001953. [AK127393] |
| A_33_P3340808 | 0,003869844 | down | -2,694643 | FLJ46026 | PREDICTED: Homo sapiens FLJ46026 protein (FLJ46026), misc_RNA [XR_159298] |
| A_23_P211878 | 0,044934873 | up | 1,7573129 | FLNB | Homo sapiens filamin B, beta (FLNB), transcript variant 2, mRNA [NM_001457] |
| A_24_P253818 | 0,01833832 | down | -1,6042352 | FLOT2 | Homo sapiens flotillin 2 (FLOT2), mRNA [NM_004475] |
| A_21_P0000024 | 0,002637014 | up | 2,4566004 | FLT1 | Homo sapiens fms-related tyrosine kinase 1 (FLT1), transcript variant 4, mRNA [NM_001160031] |
| A_23_P356070 | 1,39E-04 | down | -3,4461544 | FLT4 | Homo sapiens fms-related tyrosine kinase 4 (FLT4), transcript variant 2, mRNA [NM_002020] |
| A_24_P417596 | 0,04290036 | down | -2,0989215 | FLYWCH2 | Homo sapiens FLYWCH family member 2 (FLYWCH2), transcript variant 1, mRNA [NM_138439] |
| A_33_P3404749 | 9,43E-04 | down | -1,7353859 | FMN1 | Homo sapiens formin 1 (FMN1), transcript variant 1, mRNA [NM_001277313] |
| A_33_P3219821 | 0,00569851 | up | 1,5503658 | FMN2 | Homo sapiens formin 2 (FMN2), mRNA [NM_020066] |
| A_24_P390668 | 0,031658538 | up | 1,8615139 | FMNL1 | Homo sapiens formin-like 1 (FMNL1), mRNA [NM_005892] |
| A_32_P193080 | 0,019397354 | up | 1,7342943 | FMNL2 | Homo sapiens formin-like 2 (FMNL2), mRNA [NM_052905] |
| A_23_P149613 | 0,002766131 | down | -3,7661257 | FMO1 | Homo sapiens flavin containing monooxygenase 1 (FMO1), transcript variant 2, mRNA [NM_002021] |
| A_33_P3285271 | 0,022303024 | down | -2,2103994 | FMO6P | Homo sapiens flavin containing monooxygenase 6 pseudogene (FMO6P), non-coding RNA [NR_002601] |
| A_24_P40417 | 0,005229462 | up | 1,806462 | FMR1 | Homo sapiens fragile X mental retardation 1 (FMR1), transcript variant ISO1, mRNA [NM_002024] |
| A_33_P3238785 | 7,42E-04 | down | -2,579514 | FMR1-AS1 | Homo sapiens FMR1 antisense RNA 1 (FMR1-AS1), transcript variant 1, long non-coding RNA [NR_024499] |
| A_23_P89215 | 1,19E-05 | down | -2,4766564 | FN3K | Homo sapiens fructosamine 3 kinase (FN3K), mRNA [NM_022158] |
| A_23_P417942 | 0,001492011 | up | 1,7552344 | FNBP1L | Homo sapiens formin binding protein 1-like (FNBP1L), transcript variant 1, mRNA [NM_001024948] |
| A_23_P395595 | 0,004693421 | up | 1,5741659 | FNBP4 | Homo sapiens formin binding protein 4 (FNBP4), mRNA [NM_015308] |
| A_33_P3385656 | 0,004327302 | down | -2,3949888 | FNDC9 | Homo sapiens fibronectin type III domain containing 9 (FNDC9), mRNA [NM_001001343] |
| A_23_P47709 | 0,003013448 | down | -4,063029 | FOLR2 | Homo sapiens folate receptor 2 (fetal) (FOLR2), transcript variant 1, mRNA [NM_000803] |
| A_24_P32935 | 0,015293999 | down | -2,112469 | FOLR2 | Homo sapiens folate receptor 2 (fetal) (FOLR2), transcript variant 1, mRNA [NM_000803] |
| A_23_P37127 | 0,001005047 | up | 1,8111105 | FOXA1 | Homo sapiens forkhead box A1 (FOXA1), mRNA [NM_004496] |
| A_23_P413585 | 0,037297953 | down | -1,8295522 | FOXD2 | Homo sapiens forkhead box D2 (FOXD2), mRNA [NM_004474] |
| A_33_P3293403 | 0,009727813 | down | -2,0014288 | FOXD4L5 | Homo sapiens forkhead box D4-like 5 (FOXD4L5), mRNA [NM_001126334] |
| A_23_P118254 | 6,41E-05 | down | -3,0930185 | FOXF1 | Homo sapiens forkhead box F1 (FOXF1), mRNA [NM_001451] |
| A_23_P82959 | 0,004538706 | down | -1,9072067 | FOXH1 | Homo sapiens forkhead box H1 (FOXH1), mRNA [NM_003923] |
| A_24_P15391 | 0,002316359 | down | -1,8997567 | FOXI3 | Homo sapiens forkhead box I3 (FOXI3), mRNA [NM_001135649] |
| A_24_P21985 | 0,003618163 | up | 1,7873597 | FOXJ2 | Homo sapiens forkhead box J2 (FOXJ2), mRNA [NM_018416] |
| A_24_P115199 | 3,85E-04 | up | 2,0135167 | FOXK1 | Homo sapiens forkhead box K1 (FOXK1), mRNA [NM_001037165] |

| | | | | | |
|---|---|---|---|---|---|
| A_33_P3240392 | 0,023361523 | up | 1,7184272 | FOXO3 | Homo sapiens forkhead box O3 (FOXO3), transcript variant 1, mRNA [NM_001455] |
| A_33_P3375665 | 0,023842458 | up | 1,7107227 | FOXO3 | Homo sapiens forkhead box O3 (FOXO3), transcript variant 1, mRNA [NM_001455] |
| A_33_P3214303 | 0,002168996 | down | -2,2630866 | FOXP1 | Homo sapiens forkhead box P1 (FOXP1), transcript variant 3, mRNA [NM_001244808] |
| A_33_P3284019 | 1,58E-05 | down | -6,1766624 | FOXP4 | Homo sapiens forkhead box P4 (FOXP4), transcript variant 1, mRNA [NM_001012426] |
| A_32_P164246 | 0,048351653 | up | 1,6154212 | FOXQ1 | Homo sapiens forkhead box Q1 (FOXQ1), mRNA [NM_033260] |
| A_24_P204011 | 0,018628534 | up | 1,7592133 | FOXR1 | Homo sapiens forkhead box R1 (FOXR1), mRNA [NM_181721] |
| A_23_P55649 | 0,007350548 | down | -3,8598385 | FPR2 | Homo sapiens formyl peptide receptor 2 (FPR2), transcript variant 1, mRNA [NM_001462] |
| A_33_P3247042 | 0,001502167 | down | -2,5937939 | FPR3 | Homo sapiens formyl peptide receptor 3 (FPR3), mRNA [NM_002030] |
| A_33_P3261173 | 0,047491986 | up | 1,9127512 | FRAT1 | Homo sapiens frequently rearranged in advanced T-cell lymphomas 1 (FRAT1), mRNA [NM_005479] |
| A_33_P3212394 | 0,007186733 | down | -2,02655 | FRG2C | Homo sapiens FSHD region gene 2 family, member C (FRG2C), mRNA [NM_001124759] |
| A_23_P133665 | 7,54E-04 | up | 2,4344623 | FRK | Homo sapiens fyn-related Src family tyrosine kinase (FRK), mRNA [NM_002031] |
| A_23_P81717 | 0,00256881 | down | -3,4555852 | FRMD1 | Homo sapiens FERM domain containing 1 (FRMD1), transcript variant 1, mRNA [NM_024919] |
| A_33_P3406072 | 0,02642051 | up | 1,8330715 | FRMD3 | Homo sapiens FERM domain containing 3 (FRMD3), transcript variant 2, mRNA [NM_001244959] |
| A_24_P67988 | 0,001290035 | up | 1,7496171 | FRMD8 | Homo sapiens FERM domain containing 8 (FRMD8), transcript variant 1, mRNA [NM_031904] |
| A_32_P129669 | 1,06E-05 | down | -2,286693 | FRMPD3 | Homo sapiens FERM and PDZ domain containing 3 (FRMPD3), mRNA [NM_032428] |
| A_23_P140342 | 0,001789877 | up | 2,88348 | FSCB | Homo sapiens fibrous sheath CABYR binding protein (FSCB), mRNA [NM_032135] |
| A_33_P3241541 | 0,047440782 | up | 1,5365906 | FSD1L | Homo sapiens fibronectin type III and SPRY domain containing 1-like (FSD1L), transcript variant 3, mRNA [NM_001145313] |
| A_33_P3298326 | 0,008770271 | up | 1,5313987 | FSHB | Homo sapiens follicle stimulating hormone, beta polypeptide (FSHB), transcript variant 1, mRNA [NM_000510] |
| A_33_P3293773 | 0,010431117 | down | -1,7891737 | FSIP2 | Homo sapiens fibrous sheath interacting protein 2 (FSIP2), mRNA [NM_173651] |
| A_24_P251734 | 0,03935817 | up | 1,845808 | FSTL5 | Homo sapiens follistatin-like 5 (FSTL5), transcript variant 1, mRNA [NM_020116] |
| A_19_P00812340 | 0,028096678 | down | -1,7524016 | FTL | Homo sapiens ferritin, light polypeptide (FTL), mRNA [NM_000146] |
| A_23_P435833 | 8,07E-04 | down | -1,85859 | FUBP3 | Homo sapiens far upstream element (FUSE) binding protein 3 (FUBP3), mRNA [NM_003934] |
| A_23_P106887 | 0,005512263 | up | 1,9124972 | FUS | Homo sapiens FUS RNA binding protein (FUS), transcript variant 1, mRNA [NM_004960] |
| A_33_P3748714 | 0,02899889 | down | -1,7335144 | FUT10 | Homo sapiens fucosyltransferase 10 (alpha (1,3) fucosyltransferase) (FUT10), mRNA [NM_032664] |
| A_33_P3354713 | 0,02359254 | down | -1,5208355 | FUT11 | Homo sapiens fucosyltransferase 11 (alpha (1,3) fucosyltransferase) (FUT11), transcript variant 2, mRNA [NM_001284194] |
| A_23_P170534 | 2,54E-04 | down | -2,4924881 | FUT7 | Homo sapiens fucosyltransferase 7 (alpha (1,3) fucosyltransferase) (FUT7), mRNA [NM_004479] |
| A_23_P313632 | 0,007588525 | down | -1,978376 | FUT8 | Homo sapiens fucosyltransferase 8 (alpha (1,6) fucosyltransferase) (FUT8), transcript variant 1, mRNA [NM_178155] |
| A_24_P463891 | 0,00441719 | up | 2,0976708 | FUT9 | Homo sapiens fucosyltransferase 9 (alpha (1,3) fucosyltransferase) (FUT9), mRNA [NM_006581] |
| A_33_P3228435 | 0,047189698 | down | -2,3280106 | FXYD1 | Homo sapiens FXYD domain containing ion transport regulator 1 (FXYD1), transcript variant c, mRNA [NM_001278717] |
| A_23_P161769 | 1,09E-05 | down | -4,1996922 | FXYD2 | Homo sapiens FXYD domain containing ion transport regulator 2 (FXYD2), transcript variant b, mRNA [NM_021603] |
| A_33_P3228445 | 0,045499567 | up | 1,7076968 | FXYD2 | Homo sapiens FXYD domain containing ion transport regulator 2 (FXYD2), transcript variant a, mRNA [NM_001680] |
| A_23_P119611 | 3,11E-04 | down | -2,6833901 | FXYD7 | Homo sapiens FXYD domain containing ion transport regulator 7 (FXYD7), mRNA [NM_022006] |
| A_21_P0007806 | 1,24E-04 | up | 3,559904 | FZD10-AS1 | Homo sapiens FZD10 antisense RNA 1 (head to head) (FZD10-AS1), long non-coding RNA [NR_033834] |
| A_21_P0007804 | 0,016386146 | up | 1,5877942 | FZD10-AS1 | FZD10 antisense RNA 1 (head to head) [Source:HGNC Symbol;Acc:HGNC:48632] [ENST00000505807] |
| A_24_P941759 | 0,015737474 | up | 1,8846071 | G2E3 | Homo sapiens G2/M-phase specific E3 ubiquitin protein ligase (G2E3), mRNA [NM_017769] |
| A_23_P203743 | 0,040838942 | up | 1,5975051 | GAB2 | Homo sapiens GRB2-associated binding protein 2 (GAB2), transcript variant 2, mRNA [NM_012296] |
| A_33_P3251369 | 0,010169972 | up | 1,6214709 | GAB3 | Homo sapiens GRB2-associated binding protein 3 (GAB3), transcript variant 1, mRNA [NM_001081573] |
| A_23_P93302 | 0,001983317 | up | 2,007925 | GABBR1 | Homo sapiens gamma-aminobutyric acid (GABA) B receptor, 1 (GABBR1), transcript variant 1, mRNA [NM_001470] |
| A_24_P176255 | 6,24E-05 | up | 2,0033505 | GABPB1 | Homo sapiens GA binding protein transcription factor, beta subunit 1 (GABPB1), transcript variant beta-1, mRNA [NM_005254] |
| A_23_P125618 | 0,002921711 | down | -2,8683338 | GABRA3 | Homo sapiens gamma-aminobutyric acid (GABA) A receptor, alpha 3 (GABRA3), mRNA [NM_000808] |
| A_32_P25514 | 0,048979037 | down | -1,523449 | GABRG2 | Homo sapiens gamma-aminobutyric acid (GABA) A receptor, gamma 2 (GABRG2), transcript variant 1, mRNA [NM_198904] |
| A_33_P3210805 | 0,025627023 | up | 2,3591926 | GABRG3 | Homo sapiens gamma-aminobutyric acid (GABA) A receptor, gamma 3 (GABRG3), transcript variant 1, mRNA [NM_033223] |
| A_23_P93105 | 0,005064974 | up | 2,7371933 | GABRR2 | Homo sapiens gamma-aminobutyric acid (GABA) A receptor, rho 2 (GABRR2), mRNA [NM_002043] |
| A_21_P0001900 | 5,92E-04 | down | -1,922291 | GACAT1 | Homo sapiens gastric cancer associated transcript 1 (non-protein coding) (GACAT1), transcript variant 1, long non-coding RNA [NR_126369] |
| A_24_P239606 | 1,85E-05 | up | 2,6496363 | GADD45B | Homo sapiens growth arrest and DNA-damage-inducible, beta (GADD45B), mRNA [NM_015675] |
| A_33_P3409266 | 0,001433723 | down | -3,112106 | GAFA2 | Homo sapiens FGF-2 activity-associated protein 2 (GAFA2) mRNA, complete cds. [AF220234] |
| A_24_P228796 | 0,02810782 | up | 2,7754824 | GAGE7 | Homo sapiens G antigen 7 (GAGE7), mRNA [NM_021123] |
| A_33_P3275390 | 0,006148054 | up | 1,989173 | GAL3ST3 | Homo sapiens galactose-3-O-sulfotransferase 3 (GAL3ST3), mRNA [NM_033036] |
| A_23_P212119 | 0,002652666 | up | 2,5316937 | GALNT15 | Homo sapiens polypeptide N-acetylgalactosaminyltransferase 15 (GALNT15), mRNA [NM_054110] |
| A_23_P76749 | 0,00141386 | down | -3,1896255 | GALNT16 | Homo sapiens polypeptide N-acetylgalactosaminyltransferase 16 (GALNT16), transcript variant 2, mRNA [NM_020692] |
| A_24_P353794 | 0,016806483 | up | 2,1543794 | GALNT2 | Homo sapiens polypeptide N-acetylgalactosaminyltransferase 2 (GALNT2), transcript variant 1, mRNA [NM_004481] |
| A_24_P942370 | 0,022877907 | up | 1,8416846 | GALNT4 | Homo sapiens polypeptide N-acetylgalactosaminyltransferase 4 (GALNT4), mRNA [NM_003774] |
| A_23_P65100 | 0,043815847 | up | 1,6851047 | GALNT8 | Homo sapiens polypeptide N-acetylgalactosaminyltransferase 8 (GALNT8), mRNA [NM_017417] |
| A_33_P3355933 | 0,009397322 | down | -1,7762246 | GALNT9 | polypeptide N-acetylgalactosaminyltransferase 9 [Source:HGNC Symbol;Acc:HGNC:4131] [ENST00000614360] |
| A_19_P00322242 | 0,00233029 | down | -1,6707494 | GALNT9 | Homo sapiens polypeptide N-acetylgalactosaminyltransferase 9 (GALNT9), transcript variant A, mRNA [NM_001122636] |
| A_19_P00327126 | 0,01112599 | down | -2,2553372 | GALNT9 | Homo sapiens polypeptide N-acetylgalactosaminyltransferase 9 (GALNT9), transcript variant A, mRNA [NM_001122636] |
| A_23_P429601 | 0,03693305 | up | 1,5900459 | GALNTL5 | Homo sapiens polypeptide N-acetylgalactosaminyltransferase-like 5 (GALNTL5), transcript variant 1, mRNA [NM_145292] |
| A_23_P77756 | 0,015098353 | down | -1,8882959 | GALR2 | Homo sapiens galanin receptor 2 (GALR2), mRNA [NM_003857] |
| A_23_P256663 | 0,003986001 | down | -2,35315 | GALR3 | Homo sapiens galanin receptor 3 (GALR3), mRNA [NM_003614] |
| A_33_P3265159 | 0,047689028 | up | 1,5113626 | GAPVD1 | Homo sapiens cDNA clone IMAGE:5203707, containing frame-shift errors. [BC110990] |
| A_23_P372660 | 9,18E-05 | up | 1,8971967 | GAPVD1 | Homo sapiens GTPase activating protein and VPS9 domains 1 (GAPVD1), transcript variant 4, mRNA [NM_015635] |

| | | | | | |
|---|---|---|---|---|---|
| A_21_P0000680 | 0,021143526 | up | 1,6370286 | GAS5-AS1 | Homo sapiens GAS5 antisense RNA 1 (GAS5-AS1), long non-coding RNA [NR_037605] |
| A_33_P3266192 | 0,008735375 | up | 2,2319102 | GAS6-AS1 | Homo sapiens hypothetical LOC650669, mRNA (cDNA clone MGC:169000 IMAGE:9021377), complete cds. [BC137379] |
| A_33_P3352522 | 8,76E-04 | up | 3,0857325 | GAS6-AS1 | Homo sapiens cDNA FLJ35543 fis, clone SPLEN2002957. [AK092862] |
| A_23_P159191 | 0,014197458 | down | -1,9814205 | GAST | Homo sapiens gastrin (GAST), mRNA [NM_000805] |
| A_33_P3550894 | 0,007920567 | up | 3,3181272 | GATA2 | Homo sapiens GATA binding protein 2 (GATA2), transcript variant 1, mRNA [NM_001145661] |
| A_21_P0014114 | 0,031014336 | up | 2,3775504 | GATA2-AS1 | Homo sapiens GATA2 antisense RNA 1 (GATA2-AS1), long non-coding RNA [NR_125398] |
| A_33_P3613516 | 0,005997577 | up | 1,9766632 | GATA2-AS1 | Homo sapiens cDNA: FLJ21000 fis, clone CAE03359. [AK024653] |
| A_21_P0006622 | 0,007651915 | up | 1,5847104 | GATA3-AS1 | Homo sapiens GATA3 antisense RNA 1 (GATA3-AS1), transcript variant 1, long non-coding RNA [NR_104327] |
| A_23_P131020 | 0,007460038 | up | 1,6193429 | GATAD2A | Homo sapiens GATA zinc finger domain containing 2A (GATAD2A), transcript variant 2, mRNA [NM_017660] |
| A_23_P327777 | 0,022931254 | up | 1,907041 | GATS | Homo sapiens GATS, stromal antigen 3 opposite strand (GATS), transcript variant 1, mRNA [NM_178831] |
| A_24_P341187 | 8,76E-04 | up | 2,0459318 | GBA2 | Homo sapiens glucosidase, beta (bile acid) 2 (GBA2), mRNA [NM_020944] |
| A_23_P18672 | 0,002874911 | down | -2,0285633 | GBA3 | Homo sapiens glucosidase, beta, acid 3 (gene/pseudogene) (GBA3), transcript variant 1, mRNA [NM_020973] |
| A_19_P00808588 | 0,001536843 | down | -2,772137 | GBP1P1 | Homo sapiens guanylate binding protein 1, interferon-inducible pseudogene 1 (GBP1P1), non-coding RNA [NR_003133] |
| A_24_P309069 | 0,026806865 | up | 1,9763068 | GBX1 | Homo sapiens gastrulation brain homeobox 1 (GBX1), mRNA [NM_001098834] |
| A_24_P85942 | 0,031765655 | up | 1,7461706 | GCC2 | Homo sapiens GRIP and coiled-coil domain containing 2 (GCC2), transcript variant 1, mRNA [NM_181453] |
| A_21_P0000149 | 0,023415644 | down | -1,515101 | GCFC2 | Homo sapiens GC-rich sequence DNA-binding factor 2 (GCFC2), transcript variant 3, mRNA [NM_001201335] |
| A_33_P3298460 | 0,001627946 | up | 1,7978699 | GCG | Homo sapiens glucagon (GCG), mRNA [NM_002054] |
| A_33_P3342260 | 0,025767755 | down | -1,618017 | GCK | Homo sapiens glucokinase (hexokinase 4) (GCK), transcript variant 1, mRNA [NM_000162] |
| A_23_P119886 | 0,009353855 | up | 2,7956314 | GCKR | Homo sapiens glucokinase (hexokinase 4) regulator (GCKR), mRNA [NM_001486] |
| A_32_P98732 | 0,001170277 | down | -1,7013136 | GCM1 | Homo sapiens glial cells missing homolog 1 (Drosophila) (GCM1), mRNA [NM_003643] |
| A_33_P3411204 | 3,24E-04 | down | -1,8942883 | GCNT2 | Homo sapiens glucosaminyl (N-acetyl) transferase 2, I-branching enzyme (I blood group) (GCNT2), transcript variant 2, mRNA [NM_001491] |
| A_33_P3282522 | 0,011933794 | up | 1,5805343 | GCNT7 | Homo sapiens glucosaminyl (N-acetyl) transferase family member 7 (GCNT7), mRNA [NM_080615] |
| A_24_P393571 | 0,033864994 | up | 2,6382973 | GDA | Homo sapiens guanine deaminase (GDA), transcript variant 2, mRNA [NM_004293] |
| A_33_P3331767 | 0,01592533 | up | 1,5423998 | GDAP1L1 | Homo sapiens ganglioside induced differentiation associated protein 1-like 1 (GDAP1L1), transcript variant 5, mRNA [NM_001256740] |
| A_23_P46755 | 0,00619766 | down | -2,0526762 | GDF2 | Homo sapiens growth differentiation factor 2 (GDF2), mRNA [NM_016204] |
| A_23_P72817 | 0,004251664 | up | 2,0742042 | GDF3 | Homo sapiens growth differentiation factor 3 (GDF3), mRNA [NM_020634] |
| A_23_P259955 | 0,00495404 | down | -1,8451531 | GDF5 | Homo sapiens growth differentiation factor 5 (GDF5), mRNA [NM_000557] |
| A_33_P3406240 | 8,33E-04 | down | -2,432216 | GDF7 | Homo sapiens growth differentiation factor 7 (GDF7), mRNA [NM_182828] |
| A_23_P161446 | 0,007686908 | up | 1,6648022 | GDI2 | Homo sapiens GDP dissociation inhibitor 2 (GDI2), transcript variant 1, mRNA [NM_001494] |
| A_19_P00322166 | 0,044912465 | up | 3,2015655 | GDNF-AS1 | Homo sapiens GDNF antisense RNA 1 (head to head) (GDNF-AS1), transcript variant 1, mRNA [NM_001278094] |
| A_21_P0004501 | 3,79E-04 | up | 1,8827144 | GDNF-AS1 | Homo sapiens GDNF antisense RNA 1 (head to head) (GDNF-AS1), transcript variant GDNFOS, non-coding RNA [NR_103442] |
| A_33_P3258330 | 0,002125959 | down | -1,8680649 | GDPD5 | Homo sapiens glycerophosphodiester phosphodiesterase domain containing 5 (GDPD5), mRNA [NM_030792] |
| A_23_P89910 | 0,001146262 | up | 1,5790973 | GEMIN7 | Homo sapiens gem (nuclear organelle) associated protein 7 (GEMIN7), transcript variant 1, mRNA [NM_024707] |
| A_23_P257365 | 0,00192961 | up | 1,8662931 | GFI1 | Homo sapiens growth factor independent 1 transcription repressor (GFI1), transcript variant 1, mRNA [NM_005263] |
| A_33_P3639068 | 0,012118743 | down | -2,452989 | GFRA1 | Homo sapiens GDNF family receptor alpha 1 (GFRA1), transcript variant 1, mRNA [NM_005264] |
| A_33_P3293336 | 0,013639861 | down | -1,9114004 | GFRA1 | Homo sapiens GDNF family receptor alpha 1 (GFRA1), transcript variant 1, mRNA [NM_005264] |
| A_33_P3317580 | 0,00260289 | down | -1,5988168 | GFRA2 | Homo sapiens GDNF family receptor alpha 2 (GFRA2), transcript variant 1, mRNA [NM_001495] |
| A_33_P3345051 | 0,03539785 | down | -1,8015491 | GFY | Homo sapiens golgi-associated, olfactory signaling regulator (GFY), mRNA [NM_001195256] |
| A_24_P926400 | 9,36E-05 | up | 2,0868938 | GGA1 | golgi-associated, gamma adaptin ear containing, ARF binding protein 1 [Source:HGNC Symbol;Acc:HGNC:17842] [ENST00000489772] |
| A_33_P3301485 | 0,027455045 | up | 1,570191 | GGA1 | golgi-associated, gamma adaptin ear containing, ARF binding protein 1 [Source:HGNC Symbol;Acc:HGNC:17842] [ENST00000491295] |
| A_23_P38181 | 6,09E-05 | up | 1,8353864 | GGA3 | Homo sapiens golgi-associated, gamma adaptin ear containing, ARF binding protein 3 (GGA3), transcript variant long, mRNA [NM_138619] |
| A_19_P00809029 | 0,004376513 | down | -1,9679946 | GGACT | Homo sapiens gamma-glutamylamine cyclotransferase (GGACT), transcript variant 2, mRNA [NM_001195087] |
| A_33_P3369790 | 0,001721148 | down | -2,172858 | GGN | Homo sapiens gametogenetin (GGN), mRNA [NM_152657] |
| A_24_P606538 | 3,50E-04 | down | -7,4479575 | GGNBP1 | Homo sapiens gametogenetin binding protein 1 (pseudogene) (GGNBP1), non-coding RNA [NR_028361] |
| A_21_P0012283 | 0,005808626 | down | -1,8766586 | GGT1 | Homo sapiens gamma-glutamyltransferase 1 (GGT1), transcript variant 6, mRNA [NM_001288833] |
| A_33_P3255304 | 0,01671425 | up | 3,1028264 | GGT5 | Homo sapiens gamma-glutamyltransferase 5 (GGT5), transcript variant 1, mRNA [NM_001099781] |
| A_23_P112452 | 0,00118785 | down | -2,6855805 | GGTA1P | Homo sapiens glycoprotein, alpha-galactosyltransferase 1 pseudogene (GGTA1P), transcript variant 1, non-coding RNA [NR_003191] |
| A_33_P3389008 | 2,26E-04 | down | -2,9363604 | GHITM | Homo sapiens growth hormone inducible transmembrane protein (GHITM), mRNA [NM_014394] |
| A_33_P3260733 | 0,017230097 | down | -1,8564405 | GHR | Homo sapiens growth hormone receptor (GHR), transcript variant 12, mRNA [NM_001242462] |
| A_33_P3391826 | 9,20E-04 | down | -2,3660154 | GHRH | Homo sapiens growth hormone releasing hormone (GHRH), transcript variant 1, mRNA [NM_021081] |
| A_33_P3232655 | 0,04055595 | down | -1,7404616 | GHRHR | Homo sapiens growth hormone releasing hormone receptor (GHRHR), mRNA [NM_000823] |
| A_23_P40956 | 0,016927265 | down | -1,7253838 | GHRL | Homo sapiens ghrelin/obestatin prepropeptide (GHRL), transcript variant 1, mRNA [NM_016362] |
| A_33_P3261620 | 0,007403389 | down | -1,9328339 | GHSR | Homo sapiens growth hormone secretagogue receptor (GHSR), transcript variant 1b, mRNA [NM_004122] |
| A_24_P274831 | 0,016501527 | up | 1,5041035 | GIMAP7 | Homo sapiens GTPase, IMAP family member 7 (GIMAP7), mRNA [NM_153236] |
| A_21_P0012784 | 0,013513634 | up | 1,5857635 | GIMD1 | Homo sapiens GIMAP family P-loop NTPase domain containing 1 (GIMD1), mRNA [NM_001195138] |
| A_21_P0000123 | 0,004458814 | up | 1,5180712 | GIMD1 | Homo sapiens GIMAP family P-loop NTPase domain containing 1 (GIMD1), mRNA [NM_001195138] |
| A_33_P3340025 | 0,01172148 | up | 1,5093861 | GINS1 | Homo sapiens GINS complex subunit 1 (Psf1 homolog) (GINS1), mRNA [NM_021067] |
| A_33_P3340040 | 0,001791175 | up | 2,1294875 | GINS4 | Homo sapiens GINS complex subunit 4 (Sld5 homolog) (GINS4), mRNA [NM_032336] |
| A_24_P118527 | 0,003569122 | down | -2,7951174 | GIPC3 | Homo sapiens GIPC PDZ domain containing family, member 3 (GIPC3), mRNA [NM_133261] |
| A_24_P373768 | 0,003562748 | down | -1,9710605 | GIPR | Homo sapiens gastric inhibitory polypeptide receptor (GIPR), mRNA [NM_000164] |
| A_33_P3401673 | 0,011017551 | down | -1,5706959 | GIT1 | Homo sapiens G protein-coupled receptor kinase interacting ArfGAP 1 (GIT1), transcript variant 2, mRNA [NM_014030] |

| | | | | | |
|---|---|---|---|---|---|
| A_23_P204436 | 0,001229069 | up | 1,6251062 | GIT2 | Homo sapiens G protein-coupled receptor kinase interacting ArfGAP 2 (GIT2), transcript variant 4, mRNA [NM_139201] |
| A_23_P371729 | 7,55E-04 | up | 3,0301702 | GJA5 | Homo sapiens gap junction protein, alpha 5, 40kDa (GJA5), transcript variant A, mRNA [NM_005266] |
| A_23_P23037 | 0,001049121 | down | -4,8213058 | GJA8 | Homo sapiens gap junction protein, alpha 8, 50kDa (GJA8), mRNA [NM_005267] |
| A_23_P74828 | 0,009174958 | down | -1,5511122 | GJA9 | Homo sapiens gap junction protein, alpha 9, 59kDa (GJA9), mRNA [NM_030772] |
| A_24_P404840 | 0,036449272 | up | 2,46556 | GJB1 | Homo sapiens gap junction protein, beta 1, 32kDa (GJB1), transcript variant 2, mRNA [NM_000166] |
| A_24_P379233 | 0,008738674 | up | 1,7165059 | GJB3 | Homo sapiens gap junction protein, beta 3, 31kDa (GJB3), transcript variant 1, mRNA [NM_024009] |
| A_33_P3273777 | 0,003252058 | up | 1,7110896 | GJB7 | Homo sapiens gap junction protein, beta 7, 25kDa (GJB7), mRNA [NM_198568] |
| A_23_P30983 | 0,008841389 | up | 2,3704643 | GJB7 | Homo sapiens gap junction protein, beta 7, 25kDa (GJB7), mRNA [NM_198568] |
| A_24_P43723 | 0,00458878 | down | -2,595548 | GJC3 | Homo sapiens gap junction protein, gamma 3, 30.2kDa (GJC3), mRNA [NM_181538] |
| A_33_P3264790 | 0,003311693 | down | -2,27699 | GJD4 | Homo sapiens gap junction protein, delta 4, 40.1kDa, mRNA (cDNA clone IMAGE:5240397), partial cds. [BC035898] |
| A_32_P172848 | 0,03154166 | down | -1,6203784 | GK | Homo sapiens glycerol kinase (GK), transcript variant 2, mRNA [NM_000167] |
| A_33_P3779229 | 7,77E-05 | up | 1,819968 | GK5 | Homo sapiens glycerol kinase 5 (putative) (GK5), transcript variant 1, mRNA [NM_001039547] |
| A_23_P218579 | 0,013944304 | down | -1,7987121 | GLB1L | Homo sapiens galactosidase, beta 1-like (GLB1L), transcript variant 2, mRNA [NM_024506] |
| A_23_P151870 | 0,013529988 | up | 1,708478 | GLCE | Homo sapiens glucuronic acid epimerase (GLCE), mRNA [NM_015554] |
| A_23_P105251 | 0,044350248 | down | -2,2113159 | GLI1 | Homo sapiens GLI family zinc finger 1 (GLI1), transcript variant 1, mRNA [NM_005269] |
| A_33_P3285715 | 3,72E-04 | down | -2,8884506 | GLI4 | GLI family zinc finger 4 [Source:HGNC Symbol;Acc:HGNC:4320] [ENST00000523812] |
| A_23_P317839 | 0,014000998 | up | 1,7146568 | GLIPR1L1 | Homo sapiens GLI pathogenesis-related 1 like 1 (GLIPR1L1), mRNA [NM_152779] |
| A_33_P3295749 | 0,045004267 | down | -1,7896079 | GLIPR1L2 | Homo sapiens GLI pathogenesis-related 1 like 2 (GLIPR1L2), transcript variant 3, non-coding RNA [NR_072995] |
| A_23_P316612 | 0,006853018 | down | -1,9681526 | GLIS1 | Homo sapiens GLIS family zinc finger 1 (GLIS1), mRNA [NM_147193] |
| A_33_P3326568 | 0,03171113 | down | -1,9440285 | GLIS3 | GLIS family zinc finger 3 [Source:HGNC Symbol;Acc:HGNC:28510] [ENST00000467497] |
| A_23_P312837 | 0,023104616 | up | 2,65419 | GLIS3-AS1 | Homo sapiens GLIS3 antisense RNA 1 (GLIS3-AS1), long non-coding RNA [NR_026663] |
| A_23_P69908 | 0,027346017 | up | 1,9260375 | GLRX | Homo sapiens glutaredoxin (thioltransferase) (GLRX), transcript variant 1, mRNA [NM_002064] |
| A_33_P3379916 | 0,008340247 | up | 1,581214 | GLS | Homo sapiens glutaminase (GLS), transcript variant 1, mRNA [NM_014905] |
| A_33_P3390391 | 0,013024476 | up | 1,5270689 | GLTSCR1 | Homo sapiens glioma tumor suppressor candidate region gene 1 (GLTSCR1), mRNA [NM_015711] |
| A_33_P3295148 | 0,001163628 | down | -2,2113628 | GLUD1P3 | Homo sapiens glutamate dehydrogenase 1 pseudogene 3 (GLUD1P3), non-coding RNA [NR_048575] |
| A_21_P0010921 | 0,002934405 | down | -1,5741236 | GLUD1P7 | Homo sapiens glutamate dehydrogenase 1 pseudogene 7 (GLUD1P7), non-coding RNA [NR_111968] |
| A_24_P53976 | 0,001629973 | up | 3,2393265 | GLUL | Homo sapiens glutamate-ammonia ligase (GLUL), transcript variant 1, mRNA [NM_002065] |
| A_24_P385092 | 0,020599626 | up | 1,9363577 | GLYCAM1 | Homo sapiens glycosylation dependent cell adhesion molecule 1 (pseudogene) (GLYCAM1), non-coding RNA [NR_003039] |
| A_33_P3227440 | 0,001511871 | up | 2,3171763 | GM140 | Homo sapiens uncharacterized LOC100287948 (GM140), long non-coding RNA [NR_046249] |
| A_21_P0004821 | 0,011309111 | down | -2,141949 | GMDS-AS1 | Homo sapiens GMDS antisense RNA 1 (head to head) (GMDS-AS1), long non-coding RNA [NR_046229] |
| A_21_P0004594 | 0,018287726 | down | -1,7500677 | GMDS-AS1 | Homo sapiens GMDS antisense RNA 1 (head to head) (GMDS-AS1), long non-coding RNA [NR_046229] |
| A_21_P0004819 | 0,00762791 | down | -2,883862 | GMDS-AS1 | Homo sapiens GMDS antisense RNA 1 (head to head) (GMDS-AS1), long non-coding RNA [NR_046229] |
| A_23_P208866 | 0,049594626 | down | -1,9756324 | GMFG | Homo sapiens glia maturation factor, gamma (GMFG), transcript variant 1, mRNA [NM_004877] |
| A_23_P92202 | 0,012280559 | down | -1,5003216 | GMPPB | Homo sapiens GDP-mannose pyrophosphorylase B (GMPPB), transcript variant 2, mRNA [NM_021971] |
| A_24_P277657 | 0,00635613 | up | 3,2374477 | GMPR | Homo sapiens guanosine monophosphate reductase (GMPR), mRNA [NM_006877] |
| A_24_P150874 | 1,52E-04 | up | 1,83957 | GNA13 | Homo sapiens guanine nucleotide binding protein (G protein), alpha 13 (GNA13), transcript variant 1, mRNA [NM_006572] |
| A_24_P415150 | 0,008021111 | up | 1,6190535 | GNA14 | Homo sapiens guanine nucleotide binding protein (G protein), alpha 14 (GNA14), mRNA [NM_004297] |
| A_33_P3419339 | 0,005463831 | up | 1,5613573 | GNAI3 | Homo sapiens guanine nucleotide binding protein (G protein), alpha inhibiting activity polypeptide 3 (GNAI3), mRNA [NM_006496] |
| A_24_P393611 | 5,31E-04 | down | -1,9059224 | GNAO1 | Homo sapiens guanine nucleotide binding protein (G protein), alpha activating activity polypeptide O (GNAO1), transcript variant 2, mRNA [NM_138736] |
| A_33_P3404814 | 0,014360598 | up | 1,9089407 | GNAO1 | Homo sapiens guanine nucleotide binding protein (G protein), alpha activating activity polypeptide O (GNAO1), transcript variant 1, mRNA [NM_020988] |
| A_24_P418809 | 0,02605408 | up | 1,5298349 | GNAS | Homo sapiens GNAS complex locus (GNAS), transcript variant 7, mRNA [NM_001077489] |
| A_23_P166884 | 0,002147384 | up | 2,1037886 | GNAT1 | Homo sapiens guanine nucleotide binding protein (G protein), alpha transducing activity polypeptide 1 (GNAT1), transcript variant 2, mRNA [NM_000172] |
| A_33_P3232937 | 4,10E-05 | down | -2,4412806 | GNG12-AS1 | Homo sapiens GNG12 antisense RNA 1 (GNG12-AS1), long non-coding RNA [NR_040077] |
| A_23_P26994 | 0,013854917 | up | 1,5970473 | GNGT2 | Homo sapiens guanine nucleotide binding protein (G protein), gamma transducing activity polypeptide 2 (GNGT2), transcript variant 1, mRNA [NM_031498] |
| A_23_P41025 | 0,008901925 | up | 1,6051912 | GNL3 | Homo sapiens guanine nucleotide binding protein-like 3 (nucleolar) (GNL3), transcript variant 1, mRNA [NM_014366] |
| A_23_P254594 | 0,016588513 | up | 1,8471657 | GNRH1 | Homo sapiens gonadotropin-releasing hormone 1 (luteinizing-releasing hormone) (GNRH1), transcript variant 1, mRNA [NM_000825] |
| A_33_P3418790 | 9,95E-04 | down | -2,535362 | GNRH2 | gonadotropin-releasing hormone 2 [Source:HGNC Symbol;Acc:HGNC:4420] [ENST00000380347] |
| A_24_P341535 | 0,006689819 | down | -1,5202242 | GOLGA2P7 | Homo sapiens golgin A2 pseudogene 7 (GOLGA2P7), non-coding RNA [NR_027001] |
| A_33_P3220020 | 0,00261392 | down | -2,3369923 | GOLGA2P9 | Homo sapiens golgin A2 pseudogene 9 (GOLGA2P9), non-coding RNA [NR_033899] |
| A_33_P3259440 | 0,008460073 | up | 2,6569686 | GOLGA6A | Homo sapiens golgin A6 family, member A (GOLGA6A), mRNA [NM_001038640] |
| A_33_P3250268 | 2,97E-04 | down | -2,456583 | GOLGA6L7P | golgin A6 family-like 7, pseudogene [Source:HGNC Symbol;Acc:HGNC:37442] [ENST00000567390] |
| A_33_P3383796 | 0,011694315 | down | -2,3258135 | GOLGA7B | Homo sapiens golgin A7 family, member B (GOLGA7B), mRNA [NM_001010917] |
| A_32_P119569 | 0,017129513 | down | -2,1709645 | GOLGA8EP | Homo sapiens golgin A8 family, member E, pseudogene (GOLGA8EP), non-coding RNA [NR_033350] |
| A_21_P0011355 | 0,007065025 | down | -1,7510203 | GOLGA8K | Homo sapiens golgin A8 family, member K (GOLGA8K), mRNA [NM_001282493] |
| A_23_P140614 | 0,007897202 | down | -1,8720249 | GOLGA8O | Homo sapiens golgin A8 family, member O (GOLGA8O), mRNA [NM_001277308] |
| A_23_P146512 | 5,16E-05 | up | 1,9917469 | GOLM1 | Homo sapiens golgi membrane protein 1 (GOLM1), transcript variant 1, mRNA [NM_016548] |
| A_23_P63825 | 0,037946623 | down | -1,5756027 | GOT1 | Homo sapiens glutamic-oxaloacetic transaminase 1, soluble (GOT1), mRNA [NM_002079] |
| A_33_P3461416 | 0,043583944 | up | 1,8938205 | GP6 | Homo sapiens glycoprotein VI (platelet) (GP6), transcript variant 1, mRNA [NM_001083899] |
| A_33_P3383226 | 5,82E-06 | down | -3,3479235 | GP9 | Homo sapiens glycoprotein IX (platelet) (GP9), mRNA [NM_000174] |
| A_23_P68240 | 5,58E-04 | down | -2,119454 | GPAT2 | Homo sapiens glycerol-3-phosphate acyltransferase 2, mitochondrial (GPAT2), mRNA [NM_207328] |
| A_33_P3210671 | 0,001814864 | down | -2,1337194 | GPAT2 | Homo sapiens glycerol-3-phosphate acyltransferase 2, mitochondrial (GPAT2), mRNA [NM_207328] |

| | | | | | |
|---|---|---|---|---|---|
| A_33_P3218301 | 0,001165541 | down | -2,2128723 | GPAT2 | PREDICTED: Homo sapiens glycerol-3-phosphate acyltransferase 2, mitochondrial (GPAT2), transcript variant X14, mRNA [XM_006712308] |
| A_33_P3292812 | 0,004987279 | up | 1,7472413 | GPATCH2L | Homo sapiens G patch domain containing 2-like (GPATCH2L), transcript variant 2, mRNA [NM_017972] |
| A_24_P359117 | 0,001313573 | up | 1,5446315 | GPATCH8 | Homo sapiens G patch domain containing 8 (GPATCH8), transcript variant 1, mRNA [NM_001002909] |
| A_23_P593 | 0,004384307 | up | 1,5407459 | GPBP1L1 | Homo sapiens GC-rich promoter binding protein 1-like 1 (GPBP1L1), mRNA [NM_021639] |
| A_33_P3230264 | 1,50E-04 | down | -2,9927914 | GPC3 | Homo sapiens glypican 3 (GPC3), transcript variant 1, mRNA [NM_001164617] |
| A_23_P371682 | 0,042187512 | down | -2,015897 | GPC6 | Homo sapiens glypican 6 (GPC6), mRNA [NM_005708] |
| A_23_P8640 | 0,024457503 | down | -2,1534204 | GPER1 | Homo sapiens G protein-coupled estrogen receptor 1 (GPER1), transcript variant 3, mRNA [NM_001039966] |
| A_33_P3345816 | 0,00127806 | down | -2,682225 | GPER1 | Homo sapiens G protein-coupled estrogen receptor 1 (GPER1), transcript variant 3, mRNA [NM_001039966] |
| A_23_P47181 | 0,002771839 | down | -2,5075548 | GPHA2 | Homo sapiens glycoprotein hormone alpha 2 (GPHA2), mRNA [NM_130769] |
| A_23_P214267 | 0,010044016 | up | 1,5787207 | GPR110 | Homo sapiens G protein-coupled receptor 110 (GPR110), transcript variant 1, mRNA [NM_153840] |
| A_24_P85881 | 2,66E-05 | down | -3,1183107 | GPR113 | Homo sapiens G protein-coupled receptor 113 (GPR113), transcript variant 3, mRNA [NM_153835] |
| A_23_P21425 | 1,42E-04 | down | -2,163482 | GPR119 | Homo sapiens G protein-coupled receptor 119 (GPR119), mRNA [NM_178471] |
| A_24_P411749 | 0,003951041 | up | 1,5663688 | GPR126 | Homo sapiens G protein-coupled receptor 126 (GPR126), transcript variant a1, mRNA [NM_020455] |
| A_23_P128808 | 0,019218048 | down | -2,0662374 | GPR132 | Homo sapiens G protein-coupled receptor 132 (GPR132), transcript variant 2, mRNA [NM_013345] |
| A_33_P3391135 | 0,027703986 | down | -1,663206 | GPR133 | Homo sapiens G protein-coupled receptor 133 (GPR133), mRNA [NM_198827] |
| A_33_P3237110 | 0,009549045 | up | 1,80922 | GPR137C | Homo sapiens G protein-coupled receptor 137C (GPR137C), mRNA [NM_001099652] |
| A_33_P3317508 | 0,001893246 | up | 1,8895388 | GPR148 | Homo sapiens G protein-coupled receptor 148 (GPR148), mRNA [NM_207364] |
| A_23_P6943 | 0,01268652 | down | -2,360718 | GPR15 | Homo sapiens G protein-coupled receptor 15 (GPR15), mRNA [NM_005290] |
| A_33_P3243429 | 0,001381921 | down | -1,7274574 | GPR152 | Homo sapiens G protein-coupled receptor 152 (GPR152), mRNA [NM_206997] |
| A_33_P3236846 | 0,015782857 | down | -1,7127675 | GPR17 | Homo sapiens G protein-coupled receptor 17 (GPR17), transcript variant 1, mRNA [NM_001161415] |
| A_23_P37676 | 4,31E-05 | down | -3,264633 | GPR176 | Homo sapiens G protein-coupled receptor 176 (GPR176), transcript variant 1, mRNA [NM_007223] |
| A_23_P135219 | 0,001668293 | down | -2,1915994 | GPR21 | Homo sapiens G protein-coupled receptor 21 (GPR21), mRNA [NM_005294] |
| A_23_P316531 | 0,005174169 | down | -1,5560902 | GPR25 | Homo sapiens G protein-coupled receptor 25 (GPR25), mRNA [NM_005298] |
| A_23_P385835 | 0,016364418 | up | 1,7857974 | GPR27 | Homo sapiens G protein-coupled receptor 27 (GPR27), mRNA [NM_018971] |
| A_23_P35000 | 0,029483926 | down | -1,6234807 | GPR3 | Homo sapiens G protein-coupled receptor 3 (GPR3), mRNA [NM_005281] |
| A_21_P0000136 | 0,002551375 | up | 1,5681198 | GPR33 | Homo sapiens G protein-coupled receptor 33 (gene/pseudogene) (GPR33), transcript variant 1, mRNA [NM_001197184] |
| A_23_P131534 | 0,043139283 | down | -2,7964563 | GPR45 | Homo sapiens G protein-coupled receptor 45 (GPR45), mRNA [NM_007227] |
| A_23_P159721 | 3,25E-04 | down | -3,7544026 | GPR50 | Homo sapiens G protein-coupled receptor 50 (GPR50), mRNA [NM_004224] |
| A_33_P3345469 | 0,006971665 | up | 1,563693 | GPR6 | Homo sapiens G protein-coupled receptor 6 (GPR6), transcript variant 1, mRNA [NM_001286099] |
| A_23_P214727 | 0,009047546 | down | -1,8481054 | GPR63 | Homo sapiens G protein-coupled receptor 63 (GPR63), transcript variant 2, mRNA [NM_030784] |
| A_23_P253692 | 0,027462259 | up | 2,6838884 | GPR64 | Homo sapiens G protein-coupled receptor 64 (GPR64), transcript variant 1, mRNA [NM_001079858] |
| A_33_P3222942 | 0,002005759 | down | -3,2473733 | GPR78 | Homo sapiens G protein-coupled receptor 78 (GPR78), transcript variant 1, mRNA [NM_080819] |
| A_23_P36825 | 4,03E-04 | up | 1,996425 | GPRC5A | Homo sapiens G protein-coupled receptor, class C, group 5, member A (GPRC5A), mRNA [NM_003979] |
| A_24_P160466 | 0,003367896 | up | 1,7671115 | GPRIN1 | Homo sapiens G protein regulated inducer of neurite outgrowth 1 (GPRIN1), mRNA [NM_052899] |
| A_32_P215938 | 0,011651007 | up | 1,8216562 | GPSM1 | Homo sapiens G-protein signaling modulator 1 (GPSM1), transcript variant 1, mRNA [NM_001145638] |
| A_23_P3038 | 0,025707763 | down | -1,6158806 | GPX2 | Homo sapiens glutathione peroxidase 2 (gastrointestinal) (GPX2), transcript variant 1, mRNA [NM_002083] |
| A_23_P214544 | 0,007145948 | down | -2,376403 | GPX5 | Homo sapiens glutathione peroxidase 5 (GPX5), transcript variant 1, mRNA [NM_001509] |
| A_24_P876522 | 0,03760453 | down | -1,6779771 | GPX8 | Homo sapiens glutathione peroxidase 8 (putative) (GPX8), mRNA [NM_001008397] |
| A_21_P0011002 | 0,03726861 | up | 2,078209 | GRAMD1B | PREDICTED: Homo sapiens GRAM domain containing 1B (GRAMD1B), transcript variant X3, mRNA [XM_005271621] |
| A_23_P22350 | 0,002180151 | up | 2,2492068 | GRAMD3 | Homo sapiens GRAM domain containing 3 (GRAMD3), transcript variant 2, mRNA [NM_023927] |
| A_24_P23258 | 0,002392013 | up | 1,5778039 | GRAMD4 | Homo sapiens GRAM domain containing 4 (GRAMD4), mRNA [NM_015124] |
| A_24_P299355 | 0,047998834 | down | -1,6007717 | GRAPL | Homo sapiens GRB2-related adaptor protein-like (GRAPL), mRNA [NM_001129778] |
| A_33_P3299416 | 0,004154743 | up | 1,6788518 | GRASP | Homo sapiens GRP1 (general receptor for phosphoinositides 1)-associated scaffold protein (GRASP), transcript variant 1, mRNA [NM_181711] |
| A_24_P407717 | 0,008658212 | up | 1,5316979 | GRB2 | Homo sapiens growth factor receptor-bound protein 2 (GRB2), transcript variant 1, mRNA [NM_002086] |
| A_23_P163992 | 0,014755414 | up | 1,5689802 | GRB7 | Homo sapiens growth factor receptor-bound protein 7 (GRB7), transcript variant 1, mRNA [NM_005310] |
| A_33_P3321642 | 0,001378008 | down | -2,161356 | GRHL2 | Homo sapiens grainyhead-like 2 (Drosophila) (GRHL2), mRNA [NM_024915] |
| A_33_P3292660 | 2,40E-05 | down | -2,932343 | GRIA3 | Homo sapiens glutamate receptor, ionotropic, AMPA 3 (GRIA3), transcript variant 3, mRNA [NM_001256743] |
| A_33_P3307147 | 0,013407583 | down | -1,8931065 | GRIK2 | Homo sapiens glutamate receptor, ionotropic, kainate 2 (GRIK2), transcript variant 3, mRNA [NM_001166247] |
| A_33_P3280074 | 0,0133658 | up | 2,6377692 | GRIK4 | Homo sapiens glutamate receptor, ionotropic, kainate 4 (GRIK4), transcript variant 1, mRNA [NM_001282470] |
| A_23_P16063 | 0,028954387 | down | -1,8834364 | GRIK5 | Homo sapiens glutamate receptor, ionotropic, kainate 5 (GRIK5), transcript variant 2, mRNA [NM_002088] |
| A_33_P3348127 | 0,023589134 | up | 1,5021366 | GRIN1 | Homo sapiens glutamate receptor, ionotropic, N-methyl D-aspartate 1 (GRIN1), transcript variant GluN1-5b, mRNA [NM_001185091] |
| A_33_P3334895 | 0,001110548 | down | -2,4044151 | GRIN2A | Homo sapiens glutamate receptor, ionotropic, N-methyl D-aspartate 2A (GRIN2A), transcript variant 1, mRNA [NM_001134407] |
| A_23_P49546 | 0,002041802 | down | -2,0675054 | GRIN2C | Homo sapiens glutamate receptor, ionotropic, N-methyl D-aspartate 2C (GRIN2C), transcript variant 1, mRNA [NM_000835] |
| A_23_P347541 | 4,49E-05 | down | -9,993847 | GRIN3A | Homo sapiens glutamate receptor, ionotropic, N-methyl-D-aspartate 3A (GRIN3A), mRNA [NM_133445] |
| A_33_P3279276 | 0,008239197 | down | -2,549413 | GRIP2 | Homo sapiens glutamate receptor interacting protein 2 (GRIP2), mRNA [NM_001080423] |
| A_33_P3240560 | 0,023142906 | up | 2,0222685 | GRIP2 | Homo sapiens glutamate receptor interacting protein 2 (GRIP2), mRNA [NM_001080423] |
| A_33_P3240867 | 0,049338836 | down | -1,7402186 | GRK1 | Homo sapiens G protein-coupled receptor kinase 1 (GRK1), mRNA [NM_002929] |
| A_23_P313542 | 0,00270107 | down | -2,5159054 | GRK1 | Homo sapiens G protein-coupled receptor kinase 1 (GRK1), mRNA [NM_002929] |
| A_33_P3396578 | 0,032518364 | up | 1,9688488 | GRK5 | Homo sapiens G protein-coupled receptor kinase 5, mRNA (cDNA clone IMAGE:3912936), with apparent retained intron. [BC018116] |
| A_24_P83899 | 2,92E-05 | down | -3,4445233 | GRM5 | Homo sapiens glutamate receptor, metabotropic 5 (GRM5), transcript variant b, mRNA [NM_000842] |
| A_21_P0012431 | 0,005928879 | up | 1,8135818 | GRM7-AS3 | GRM7 antisense RNA 3 [Source:HGNC Symbol;Acc:HGNC:42444] [ENST00000445675] |

| | | | | | |
|---|---|---|---|---|---|
| A_33_P3241135 | 0,009890405 | up | 1,5681392 | GRM8 | Homo sapiens glutamate receptor, metabotropic 8 (GRM8), transcript variant 1, mRNA [NM_000845] |
| A_23_P101131 | 7,65E-05 | down | -2,3068554 | GRP | Homo sapiens gastrin-releasing peptide (GRP), transcript variant 1, mRNA [NM_002091] |
| A_21_P0014853 | 7,56E-06 | down | -2,3666143 | GS1-24F4.2 | Homo sapiens uncharacterized LOC100652791 (GS1-24F4.2), long non-coding RNA [NR_045217] |
| A_21_P0013399 | 0,007130324 | down | -1,7207018 | GS1-259H13.2 | Homo sapiens transmembrane protein 225-like (GS1-259H13.2), transcript variant 1, mRNA [NM_001195541] |
| A_23_P109382 | 0,021052513 | down | -2,0178795 | GSC2 | Homo sapiens goosecoid homeobox 2 (GSC2), mRNA [NM_005315] |
| A_33_P3221999 | 0,034279637 | up | 1,6704557 | GSDMB | Homo sapiens gasdermin B (GSDMB), transcript variant 3, mRNA [NM_001165958] |
| A_33_P3224867 | 0,009566514 | down | -2,0657132 | GSG1 | Homo sapiens germ cell associated 1 (GSG1), transcript variant 5, mRNA [NM_001206842] |
| A_33_P3226650 | 0,006539121 | up | 1,5069734 | GSTA7P | Homo sapiens glutathione S-transferase alpha 7, pseudogene (GSTA7P), non-coding RNA [NR_033760] |
| A_23_P58869 | 0,013358886 | down | -1,9213274 | GSTM2P1 | Homo sapiens glutathione S-transferase mu 2 (muscle) pseudogene 1 (GSTM2P1), non-coding RNA [NR_002932] |
| A_23_P97606 | 4,85E-04 | down | -2,9533257 | GSTM5 | Homo sapiens glutathione S-transferase mu 5 (GSTM5), mRNA [NM_000851] |
| A_23_P106204 | 0,015576639 | down | -1,5231328 | GSTZ1 | Homo sapiens glutathione S-transferase zeta 1 (GSTZ1), transcript variant 1, mRNA [NM_145870] |
| A_32_P470728 | 0,002160123 | down | -1,7798007 | GSX1 | Homo sapiens GS homeobox 1 (GSX1), mRNA [NM_145657] |
| A_21_P0013240 | 0,018550523 | up | 1,5613441 | GTF2IRD1 | Homo sapiens GTF2I repeat domain containing 1 (GTF2IRD1), transcript variant 3, mRNA [NM_001199207] |
| A_23_P111621 | 0,00506415 | up | 1,8155928 | GTF2IRD1 | Homo sapiens GTF2I repeat domain containing 1 (GTF2IRD1), transcript variant 2, mRNA [NM_005685] |
| A_33_P3410454 | 0,012598364 | up | 1,5307513 | GTF2IRD2B | GTF2I repeat domain containing 2B [Source:HGNC Symbol;Acc:HGNC:33125] [ENST00000394939] |
| A_33_P3299140 | 2,27E-05 | down | -5,3311434 | GTF3C5 | general transcription factor IIIC, polypeptide 5, 63kDa [Source:HGNC Symbol;Acc:HGNC:4668] [ENST00000435745] |
| A_33_P3375646 | 0,00275325 | up | 2,2431738 | GTPBP6 | Homo sapiens cDNA FLJ56662 complete cds, highly similar to Homo sapiens GTP binding protein 6 (GTPBP6), mRNA. [AK296003] |
| A_21_P0011663 | 0,001609878 | down | -7,8402534 | GTSCR1 | Homo sapiens Gilles de la Tourette syndrome chromosome region, candidate 1 (non-protein coding) (GTSCR1), mRNA [NM_001278515] |
| A_33_P3372682 | 0,007449273 | down | -1,5325682 | GTSE1-AS1 | Homo sapiens GTSE1 antisense RNA 1 (head to head) (GTSE1-AS1), long non-coding RNA [NR_024009] |
| A_23_P81825 | 0,039147038 | up | 1,6118363 | GUCA1B | Homo sapiens guanylate cyclase activator 1B (retina) (GUCA1B), mRNA [NM_002098] |
| A_33_P3269109 | 0,001351021 | up | 2,6242332 | GUCY1A3 | guanylate cyclase 1, soluble, alpha 3 [Source:HGNC Symbol;Acc:HGNC:4685] [ENST00000512983] |
| A_33_P3378354 | 0,005223789 | down | -2,9228108 | GUCY1B2 | guanylate cyclase 1, soluble, beta 2 (pseudogene) [Source:HGNC Symbol;Acc:HGNC:4686] [ENST00000389600] |
| A_23_P360302 | 0,001830586 | up | 1,6599213 | GUCY2F | Homo sapiens guanylate cyclase 2F, retinal (GUCY2F), mRNA [NM_001522] |
| A_23_P202034 | 0,004412488 | down | -2,9978898 | GUCY2GP | Homo sapiens guanylate cyclase 2G, pseudogene (GUCY2GP), non-coding RNA [NR_028134] |
| A_21_P0010955 | 0,004966741 | down | -2,1678271 | GUCY2GP | Homo sapiens guanylate cyclase 2G, pseudogene (GUCY2GP), non-coding RNA [NR_028134] |
| A_33_P3244369 | 0,002367541 | down | -1,9065975 | GXYLT1P3 | Homo sapiens glucoside xylosyltransferase 1 pseudogene 3 (GXYLT1P3), non-coding RNA [NR_121572] |
| A_23_P29422 | 0,002791555 | down | -1,6761544 | GYG1 | Homo sapiens glycogenin 1 (GYG1), transcript variant 1, mRNA [NM_004130] |
| A_21_P0013518 | 0,001322587 | down | -1,6444604 | GYG1 | Homo sapiens glycogenin 1 (GYG1), transcript variant 3, mRNA [NM_001184721] |
| A_33_P3362933 | 0,004218669 | down | -2,2235413 | GYPA | Homo sapiens glycophorin A (MNS blood group) (GYPA), mRNA [NM_002099] |
| A_24_P139901 | 0,030936023 | down | -1,9003744 | GYPC | Homo sapiens glycophorin C (Gerbich blood group) (GYPC), transcript variant 1, mRNA [NM_002101] |
| A_23_P41304 | 0,008740234 | down | -2,0800996 | GYPE | Homo sapiens glycophorin E (MNS blood group) (GYPE), transcript variant 1, mRNA [NM_002102] |
| A_24_P156288 | 0,004882542 | up | 1,6046172 | GZF1 | Homo sapiens GDNF-inducible zinc finger protein 1 (GZF1), mRNA [NM_022482] |
| A_23_P130836 | 4,96E-04 | down | -2,4099512 | GZMM | Homo sapiens granzyme M (lymphocyte met-ase 1) (GZMM), transcript variant 1, mRNA [NM_005317] |
| A_32_P202859 | 0,014048261 | down | -1,740487 | H1FNT | Homo sapiens H1 histone family, member N, testis-specific (H1FNT), mRNA [NM_181788] |
| A_33_P3222689 | 0,023441153 | up | 2,264142 | H2BFM | Homo sapiens H2B histone family, member M (H2BFM), mRNA [NM_001164416] |
| A_24_P626850 | 0,03205002 | up | 1,5978588 | H6PD | Homo sapiens hexose-6-phosphate dehydrogenase (glucose 1-dehydrogenase) (H6PD), transcript variant 2, mRNA [NM_004285] |
| A_23_P28697 | 0,007936003 | down | -2,297879 | HAAO | Homo sapiens 3-hydroxyanthranilate 3,4-dioxygenase (HAAO), mRNA [NM_012205] |
| A_33_P3209511 | 0,039172057 | up | 1,7965009 | HAAO | 3-hydroxyanthranilate 3,4-dioxygenase [Source:HGNC Symbol;Acc:HGNC:4796] [ENST00000402268] |
| A_21_P0001940 | 0,014738527 | up | 2,3864875 | HAGLROS | Homo sapiens HAGLR opposite strand (non-protein coding) (HAGLROS), long non-coding RNA [NR_110457] |
| A_23_P61637 | 0,020553652 | up | 1,8487067 | HAL | Homo sapiens histidine ammonia-lyase (HAL), transcript variant 1, mRNA [NM_002108] |
| A_33_P3336720 | 0,010704094 | down | -2,052485 | HAMP | Homo sapiens hepcidin antimicrobial peptide (HAMP), mRNA [NM_021175] |
| A_23_P58770 | 0,009310278 | down | -1,9913946 | HAND1 | Homo sapiens heart and neural crest derivatives expressed 1 (HAND1), mRNA [NM_004821] |
| A_23_P22129 | 0,034578875 | down | -1,7861642 | HAP1 | Homo sapiens huntingtin-associated protein 1 (HAP1), transcript variant 2, mRNA [NM_177977] |
| A_33_P3318946 | 1,19E-07 | down | -5,0862484 | HAPLN2 | Homo sapiens hyaluronan and proteoglycan link protein 2 (HAPLN2), mRNA [NM_021817] |
| A_33_P3259393 | 0,032889005 | up | 1,668276 | HAPLN3 | Homo sapiens hyaluronan and proteoglycan link protein 3 (HAPLN3), mRNA [NM_178232] |
| A_32_P226498 | 0,00157735 | down | -1,7585773 | HAR1A | Homo sapiens highly accelerated region 1A (non-protein coding) (HAR1A), long non-coding RNA [NR_003244] |
| A_19_P00325498 | 0,003086957 | up | 1,5631231 | HAR1A | Homo sapiens highly accelerated region 1A (non-protein coding) (HAR1A), long non-coding RNA [NR_003244] |
| A_23_P349771 | 2,05E-05 | up | 2,9714763 | HAUS5 | Homo sapiens HAUS augmin-like complex, subunit 5 (HAUS5), mRNA [NM_015302] |
| A_33_P3271945 | 2,08E-04 | up | 2,1224012 | HAUS6 | HAUS augmin-like complex, subunit 6 [Source:HGNC Symbol;Acc:HGNC:25948] [ENST00000380496] |
| A_24_P411561 | 0,015606083 | up | 1,9283028 | HAVCR2 | Homo sapiens hepatitis A virus cellular receptor 2 (HAVCR2), mRNA [NM_032782] |
| A_23_P26457 | 8,85E-07 | down | -4,2518497 | HBA2 | Homo sapiens hemoglobin, alpha 2 (HBA2), mRNA [NM_000517] |
| A_33_P3258362 | 0,00554746 | down | -2,4367406 | HBA2 | Homo sapiens hemoglobin, alpha 2 (HBA2), mRNA [NM_000517] |
| A_24_P732099 | 0,001044986 | down | -2,124324 | HBBP1 | Homo sapiens hemoglobin, beta pseudogene 1 (HBBP1), non-coding RNA [NR_001589] |
| A_24_P75190 | 0,004833977 | down | -1,9641873 | HBD | Homo sapiens hemoglobin, delta (HBD), mRNA [NM_000519] |
| A_24_P140608 | 0,03690173 | up | 1,7344575 | HBEGF | Homo sapiens heparin-binding EGF-like growth factor (HBEGF), mRNA [NM_001945] |
| A_33_P3329078 | 5,61E-04 | down | -1,8830012 | HBG1 | Homo sapiens hemoglobin, gamma A (HBG1), mRNA [NM_000559] |
| A_23_P49254 | 0,001044818 | down | -1,956365 | HBQ1 | Homo sapiens hemoglobin, theta 1 (HBQ1), mRNA [NM_005331] |
| A_33_P3246293 | 0,020663312 | down | -1,5968391 | HBS1L | Homo sapiens HBS1-like translational GTPase (HBS1L), transcript variant 3, mRNA [NM_001145207] |
| A_23_P429092 | 0,01645119 | up | 1,5286416 | HBS1L | Homo sapiens HBS1-like translational GTPase (HBS1L), transcript variant 3, mRNA [NM_001145207] |
| A_33_P3421827 | 0,005143235 | down | -1,764401 | HBZ | Homo sapiens hemoglobin, zeta (HBZ), mRNA [NM_005332] |
| A_21_P0011413 | 0,018512594 | down | -2,0111082 | HBZ | Homo sapiens hemoglobin, zeta (HBZ), mRNA [NM_005332] |

| | | | | |
|---|---|---|---|---|
| A_33_P3380472 | 0,03402353 | down | -1,9062929 | HCAR1 | Homo sapiens hydroxycarboxylic acid receptor 1 (HCAR1), mRNA [NM_032554] |
| A_33_P3418511 | 6,39E-04 | down | -2,3196747 | HCCAT5 | Homo sapiens hepatocellular carcinoma associated transcript 5 (non-protein coding) (HCCAT5), long non-coding RNA [NR_027756] |
| A_33_P3350207 | 0,005258909 | down | -1,6405324 | HCFC1R1 | Homo sapiens host cell factor C1 regulator 1 (XPO1 dependent) (HCFC1R1), transcript variant 1, mRNA [NM_017885] |
| A_33_P3281036 | 0,03585449 | down | -1,5077711 | HCG11 | Homo sapiens HLA complex group 11 (non-protein coding) (HCG11), long non-coding RNA [NR_026790] |
| A_21_P0004638 | 0,010193622 | down | -1,8063086 | HCG14 | Homo sapiens HLA complex group 14 (non-protein coding) (HCG14), long non-coding RNA [NR_104117] |
| A_33_P3242453 | 0,019196037 | up | 4,08366 | HCG22 | Homo sapiens HLA complex group 22 (HCG22), long non-coding RNA [NR_003948] |
| A_23_P359376 | 1,18E-04 | down | -3,0499578 | HCG4B | Homo sapiens HLA complex group 4B (non-protein coding) (HCG4B), long non-coding RNA [NR_001317] |
| A_24_P255874 | 7,03E-04 | down | -2,3337145 | HCG4B | Homo sapiens HLA complex group 4B (non-protein coding) (HCG4B), long non-coding RNA [NR_001317] |
| A_33_P3279640 | 1,81E-04 | down | -5,0119853 | HCN2 | Homo sapiens hyperpolarization activated cyclic nucleotide gated potassium channel 2 (HCN2), mRNA [NM_001194] |
| A_24_P17870 | 0,001412229 | down | -2,2590015 | HCP5 | Homo sapiens HLA complex P5 (non-protein coding) (HCP5), long non-coding RNA [NR_040662] |
| A_33_P3306624 | 0,028612057 | down | -1,9453716 | HCRT | Homo sapiens hypocretin (orexin) neuropeptide precursor (HCRT), mRNA [NM_001524] |
| A_33_P3360249 | 0,03511759 | up | 2,3344207 | HCRTR1 | Homo sapiens hypocretin (orexin) receptor 1 (HCRTR1), mRNA [NM_001525] |
| A_23_P404162 | 0,021066705 | up | 1,8764596 | HDAC9 | Homo sapiens histone deacetylase 9 (HDAC9), transcript variant 3, mRNA [NM_014707] |
| A_23_P117662 | 0,030056873 | down | -1,5614154 | HDC | Homo sapiens histidine decarboxylase (HDC), mRNA [NM_002112] |
| A_33_P3319640 | 0,04668592 | down | -1,5353616 | HDGFRP2 | Hepatoma-derived growth factor-related protein 2 [Source:UniProtKB/Swiss-Prot;Acc:Q7Z4V5] [ENST00000615225] |
| A_33_P3337831 | 0,013048367 | up | 1,8463705 | HEATR1 | HEAT repeat containing 1 [Source:HGNC Symbol;Acc:HGNC:25517] [ENST00000366579] |
| A_33_P3251332 | 0,01359861 | up | 1,546485 | HEATR5A | Homo sapiens HEAT repeat containing 5A (HEATR5A), mRNA [NM_015473] |
| A_32_P389118 | 0,003363523 | up | 1,6498227 | HEATR5B | Homo sapiens HEAT repeat containing 5B (HEATR5B), mRNA [NM_019024] |
| A_23_P77965 | 0,004311132 | up | 1,5872011 | HEATR6 | Homo sapiens HEAT repeat containing 6 (HEATR6), mRNA [NM_022070] |
| A_23_P430792 | 0,006442114 | down | -2,2311523 | HEATR9 | Homo sapiens HEAT repeat containing 9 (HEATR9), mRNA [NM_152781] |
| A_23_P54006 | 0,028086223 | up | 1,541669 | HECTD1 | Homo sapiens HECT domain containing E3 ubiquitin protein ligase 1 (HECTD1), mRNA [NM_015382] |
| A_33_P3221343 | 0,003550776 | down | -1,7615312 | HECTD2 | Homo sapiens HECT domain containing E3 ubiquitin protein ligase 2 (HECTD2), transcript variant 2, mRNA [NM_173497] |
| A_23_P149221 | 0,012305411 | up | 1,5016406 | HECTD3 | Homo sapiens HECT domain containing E3 ubiquitin protein ligase 3 (HECTD3), mRNA [NM_024602] |
| A_23_P162702 | 7,18E-06 | up | 2,101758 | HECTD4 | Homo sapiens HECT domain containing E3 ubiquitin protein ligase 4 (HECTD4), mRNA [NM_001109662] |
| A_33_P3258117 | 2,80E-04 | down | -2,1395042 | HELLS | Homo sapiens helicase, lymphoid-specific (HELLS), transcript variant 7, mRNA [NM_001289072] |
| A_33_P3290677 | 0,04067902 | up | 1,9901832 | HELT | Homo sapiens helt bHLH transcription factor (HELT), transcript variant 1, mRNA [NM_001300781] |
| A_33_P3400374 | 0,001200643 | up | 1,8917596 | HELZ2 | Homo sapiens helicase with zinc finger 2, transcriptional coactivator (HELZ2), transcript variant 1, mRNA [NM_001037335] |
| A_24_P213950 | 0,00455544 | down | -2,582786 | HEPACAM | Homo sapiens hepatic and glial cell adhesion molecule (HEPACAM), mRNA [NM_152722] |
| A_24_P399980 | 0,045907665 | down | -1,5664026 | HEPH | Homo sapiens hephaestin (HEPH), transcript variant 2, mRNA [NM_014799] |
| A_33_P3298120 | 0,008743118 | down | -2,1430945 | HEPN1 | Homo sapiens hepatocellular carcinoma, down-regulated 1 (HEPN1), mRNA [NM_001037558] |
| A_23_P257503 | 5,55E-04 | up | 1,8905501 | HERC1 | Homo sapiens HECT and RLD domain containing E3 ubiquitin protein ligase family member 1 (HERC1), mRNA [NM_003922] |
| A_33_P3334343 | 0,016264822 | down | -1,6356195 | HERC2 | Homo sapiens HECT and RLD domain containing E3 ubiquitin protein ligase 2 (HERC2), mRNA [NM_004667] |
| A_23_P250358 | 0,032947358 | up | 2,206426 | HERC6 | Homo sapiens HECT and RLD domain containing E3 ubiquitin protein ligase family member 6 (HERC6), transcript variant 1, mRNA [NM_017912] |
| A_23_P6596 | 0,014158235 | up | 1,5965549 | HES1 | Homo sapiens hes family bHLH transcription factor 1 (HES1), mRNA [NM_005524] |
| A_33_P3322864 | 3,43E-04 | down | -2,4609735 | HES6 | Homo sapiens hes family bHLH transcription factor 6 (HES6), transcript variant 1, mRNA [NM_018645] |
| A_33_P3322859 | 0,004990507 | down | -1,7943754 | HES6 | Homo sapiens hes family bHLH transcription factor 6 (HES6), transcript variant 1, mRNA [NM_018645] |
| A_24_P356601 | 0,004516445 | up | 1,5153377 | HEXIM1 | Homo sapiens hexamethylene bis-acetamide inducible 1 (HEXIM1), mRNA [NM_006460] |
| A_32_P83845 | 0,019446112 | down | -2,099766 | HEY1 | Homo sapiens hes-related family bHLH transcription factor with YRPW motif 1 (HEY1), transcript variant 2, mRNA [NM_001040708] |
| A_24_P289648 | 0,02001032 | down | -1,9472069 | HFE | Homo sapiens hemochromatosis (HFE), transcript variant 11, mRNA [NM_139011] |
| A_32_P34046 | 0,004946376 | up | 1,8117332 | HFM1 | Homo sapiens HFM1, ATP-dependent DNA helicase homolog (S. cerevisiae) (HFM1), mRNA [NM_001017975] |
| A_33_P3328883 | 0,005486569 | up | 1,8799044 | HGC6.3 | Homo sapiens uncharacterized LOC100128124 (HGC6.3), mRNA [NM_001129895] |
| A_32_P115050 | 0,003764059 | down | -2,1888003 | HHIP-AS1 | Homo sapiens HHIP antisense RNA 1 (HHIP-AS1), long non-coding RNA [NR_037595] |
| A_23_P86064 | 0,002286235 | up | 2,309257 | HHIPL2 | Homo sapiens HHIP-like 2 (HHIPL2), mRNA [NM_024746] |
| A_19_P00321196 | 0,009329415 | down | -1,991794 | HHLA1 | Homo sapiens HERV-H LTR-associating 1 (HHLA1), mRNA [NM_001145095] |
| A_33_P3301040 | 0,003741551 | down | -3,1783168 | HHLA2 | Homo sapiens HERV-H LTR-associating 2 (HHLA2), transcript variant 4, mRNA [NM_001282558] |
| A_23_P83073 | 0,001278458 | up | 1,72005 | HIATL1 | Homo sapiens hippocampus abundant transcript-like 1 (HIATL1), mRNA [NM_032558] |
| A_33_P3379017 | 0,003426349 | up | 1,5664546 | HIC2 | Homo sapiens hypermethylated in cancer 2 (HIC2), mRNA [NM_015094] |
| A_23_P142187 | 0,019951386 | down | -1,6122245 | HIF3A | Homo sapiens hypoxia inducible factor 3, alpha subunit (HIF3A), transcript variant 2, mRNA [NM_022462] |
| A_33_P3338152 | 2,40E-05 | down | -3,5523608 | HIF3A | hypoxia inducible factor 3, alpha subunit [Source:HGNC Symbol;Acc:HGNC:15825] [ENST00000475432] |
| A_23_P338534 | 0,023420548 | up | 1,9893645 | HIF3A | Homo sapiens hypoxia inducible factor 3, alpha subunit (HIF3A), transcript variant 2, mRNA [NM_022462] |
| A_24_P548866 | 0,034563087 | down | -1,7748989 | HIGD1A | Homo sapiens HIG1 hypoxia inducible domain family, member 1A (HIGD1A), transcript variant 1, mRNA [NM_001099668] |
| A_23_P398294 | 6,48E-04 | up | 1,635008 | HIP1R | Homo sapiens huntingtin interacting protein 1 related (HIP1R), transcript variant 1, mRNA [NM_003959] |
| A_24_P229164 | 0,010091365 | up | 1,5303259 | HIP1R | Homo sapiens huntingtin interacting protein 1 related (HIP1R), transcript variant 1, mRNA [NM_003959] |
| A_24_P681011 | 1,09E-06 | up | 2,618245 | HIPK2 | Homo sapiens homeodomain interacting protein kinase 2 (HIPK2), transcript variant 1, mRNA [NM_022740] |
| A_23_P7976 | 0,005376293 | down | -1,8306426 | HIST1H1E | Homo sapiens histone cluster 1, H1e (HIST1H1E), mRNA [NM_005321] |
| A_23_P355439 | 0,009021338 | down | -1,8383144 | HIST1H2AA | Homo sapiens histone cluster 1, H2aa (HIST1H2AA), mRNA [NM_170745] |
| A_23_P372860 | 5,88E-05 | down | -2,105557 | HIST1H2AC | Homo sapiens histone cluster 1, H2ac (HIST1H2AC), mRNA [NM_003512] |
| A_21_P0000197 | 1,06E-06 | down | -3,0707626 | HIST1H2AG | Homo sapiens histone cluster 1, H2ag (HIST1H2AG), mRNA [NM_021064] |
| A_23_P81859 | 0,017578103 | down | -1,949021 | HIST1H2AH | Homo sapiens histone cluster 1, H2ah (HIST1H2AH), mRNA [NM_080596] |
| A_33_P3360216 | 0,001496088 | down | -3,2797215 | HIST1H2AI | Homo sapiens histone cluster 1, H2ai (HIST1H2AI), mRNA [NM_003509] |
| A_23_P323823 | 0,021067059 | up | 2,4526615 | HIST1H2BA | Homo sapiens histone cluster 1, H2ba (HIST1H2BA), mRNA [NM_170610] |

| | | | | | |
|---|---|---|---|---|---|
| A_33_P3229122 | 1,82E-04 | down | -2,492722 | HIST1H2BF | Homo sapiens histone cluster 1, H2bf (HIST1H2BF), mRNA [NM_003522] |
| A_33_P3229067 | 0,001685746 | down | -1,8567146 | HIST1H2BN | Homo sapiens histone cluster 1, H2bn (HIST1H2BN), mRNA [NM_003520] |
| A_23_P59069 | 0,038922857 | down | -1,5408846 | HIST1H2BO | Homo sapiens histone cluster 1, H2bo (HIST1H2BO), mRNA [NM_003527] |
| A_23_P93258 | 2,22E-04 | down | -2,0724356 | HIST1H3B | Homo sapiens histone cluster 1, H3b (HIST1H3B), mRNA [NM_003537] |
| A_24_P217834 | 3,23E-04 | down | -1,8191965 | HIST1H3D | Homo sapiens histone cluster 1, H3d (HIST1H3D), mRNA [NM_003530] |
| A_23_P30799 | 0,00404298 | down | -1,5358788 | HIST1H3F | Homo sapiens histone cluster 1, H3f (HIST1H3F), mRNA [NM_021018] |
| A_24_P9321 | 1,22E-04 | down | -1,989284 | HIST1H3I | Homo sapiens histone cluster 1, H3i (HIST1H3I), mRNA [NM_003533] |
| A_23_P93282 | 0,020880792 | down | -1,829476 | HIST1H3J | Homo sapiens histone cluster 1, H3j (HIST1H3J), mRNA [NM_003535] |
| A_33_P3344229 | 6,77E-07 | down | -2,9686506 | HIST1H4A | Homo sapiens histone cluster 1, H4a (HIST1H4A), mRNA [NM_003538] |
| A_24_P166407 | 0,015382585 | down | -1,5926392 | HIST1H4B | Homo sapiens histone cluster 1, H4b (HIST1H4B), mRNA [NM_003544] |
| A_33_P3378880 | 0,00558101 | down | -1,8124152 | HIST1H4C | histone cluster 1, H4c [Source:HGNC Symbol;Acc:HGNC:4787] [ENST00000377803] |
| A_33_P3410836 | 4,16E-05 | down | -2,1101665 | HIST1H4D | Homo sapiens histone cluster 1, H4d (HIST1H4D), mRNA [NM_003539] |
| A_23_P395374 | 0,002317277 | down | -1,6843828 | HIST1H4D | Homo sapiens histone cluster 1, H4d (HIST1H4D), mRNA [NM_003539] |
| A_23_P415411 | 4,82E-05 | down | -2,0333576 | HIST1H4E | Homo sapiens histone cluster 1, H4e (HIST1H4E), mRNA [NM_003545] |
| A_23_P323685 | 0,003074857 | down | -1,9407462 | HIST1H4H | Homo sapiens histone cluster 1, H4h (HIST1H4H), mRNA [NM_003543] |
| A_24_P20873 | 5,58E-04 | down | -1,92553 | HIST1H4I | Homo sapiens histone cluster 1, H4i (HIST1H4I), mRNA [NM_003495] |
| A_33_P3299865 | 1,55E-04 | down | -1,9213991 | HIST1H4K | Homo sapiens histone cluster 1, H4k (HIST1H4K), mRNA [NM_003541] |
| A_33_P3351851 | 0,001617059 | down | -1,798097 | HIST1H4L | Homo sapiens histone cluster 1, H4l (HIST1H4L), mRNA [NM_003546] |
| A_24_P68631 | 6,35E-05 | down | -3,0025165 | HIST2H2AB | Homo sapiens histone cluster 2, H2ab (HIST2H2AB), mRNA [NM_175065] |
| A_23_P301247 | 1,54E-05 | down | -2,2656443 | HIST2H2AC | Homo sapiens histone cluster 2, H2ac (HIST2H2AC), mRNA [NM_003517] |
| A_33_P3584591 | 0,003896507 | up | 1,6012734 | HIST2H2BA | AGENCOURT_8230913 Lupski_dorsal_root_ganglion Homo sapiens cDNA clone IMAGE:6182568 5', mRNA sequence [BQ880825] |
| A_23_P436281 | 0,003828814 | down | -1,5297269 | HIST2H4B | Homo sapiens histone cluster 2, H4b (HIST2H4B), mRNA [NM_001034077] |
| A_33_P3229335 | 0,029833786 | down | -2,263397 | HIST3H2BB | Homo sapiens histone cluster 3, H2bb (HIST3H2BB), mRNA [NM_175055] |
| A_23_P388871 | 0,010389054 | down | -2,3241265 | HIST4H4 | Homo sapiens histone cluster 4, H4 (HIST4H4), mRNA [NM_175054] |
| A_23_P214766 | 0,001141353 | up | 1,8541164 | HIVEP2 | Homo sapiens human immunodeficiency virus type I enhancer binding protein 2 (HIVEP2), mRNA [NM_006734] |
| A_33_P3335233 | 4,16E-04 | down | -1,8245721 | HKDC1 | Homo sapiens hexokinase domain containing 1 (HKDC1), mRNA [NM_025130] |
| A_23_P429020 | 0,001320261 | up | 1,8938237 | HKR1 | Homo sapiens HKR1, GLI-Kruppel zinc finger family member (HKR1), mRNA [NM_181786] |
| A_33_P3237574 | 7,14E-05 | down | -4,014172 | HLA-A | Homo sapiens major histocompatibility complex, class I, A (HLA-A), transcript variant 2 (A*01:01:01:01 allele), mRNA [NM_001242758] |
| A_23_P42306 | 0,004525662 | down | -1,61178 | HLA-DMA | Homo sapiens major histocompatibility complex, class II, DM alpha (HLA-DMA), mRNA [NM_006120] |
| A_32_P356316 | 0,00209728 | down | -2,0558805 | HLA-DOA | Homo sapiens major histocompatibility complex, class II, DO alpha (HLA-DOA), mRNA [NM_002119] |
| A_33_P3234277 | 0,007440937 | down | -1,6656693 | HLA-DPA1 | Homo sapiens major histocompatibility complex, class II, DP alpha 1 (HLA-DPA1), transcript variant 2, mRNA [NM_001242524] |
| A_33_P3271634 | 0,004205641 | down | -1,8915035 | HLA-DPB1 | major histocompatibility complex, class II, DP beta 1 [Source:HGNC Symbol;Acc:HGNC:4940] [ENST00000466949] |
| A_24_P166443 | 0,031168299 | down | -1,6067256 | HLA-DPB1 | Homo sapiens major histocompatibility complex, class II, DP beta 1 (HLA-DPB1), mRNA [NM_002121] |
| A_33_P3271651 | 0,004004741 | down | -2,2556922 | HLA-DPB1 | Homo sapiens major histocompatibility complex, class II, DP beta 1 (HLA-DPB1), mRNA [NM_002121] |
| A_32_P505730 | 0,019204423 | down | -1,7966197 | HLA-DPB2 | major histocompatibility complex, class II, DP beta 2 (pseudogene) [Source:HGNC Symbol;Acc:HGNC:4941] [ENST00000304890] |
| A_33_P3273884 | 0,041243464 | down | -1,814365 | HLA-DQA1 | Homo sapiens major histocompatibility complex, class II, DQ alpha 1 (HLA-DQA1), mRNA [NM_002122] |
| A_33_P3293049 | 0,048472874 | up | 1,8870652 | HLA-DQA1 | Homo sapiens major histocompatibility complex, class II, DQ alpha 1 (HLA-DQA1), mRNA [NM_002122] |
| A_32_P87697 | 0,01924299 | up | 2,5127823 | HLA-DRA | Homo sapiens major histocompatibility complex, class II, DR alpha (HLA-DRA), mRNA [NM_019111] |
| A_19_P00317913 | 0,001436618 | down | -1,9975991 | HLA-F-AS1 | Homo sapiens HLA-F antisense RNA 1 (HLA-F-AS1), transcript variant 2, long non-coding RNA [NR_026973] |
| A_32_P76811 | 0,025307573 | down | -1,6350058 | HLA-F-AS1 | Homo sapiens HLA-F antisense RNA 1 (HLA-F-AS1), transcript variant 1, long non-coding RNA [NR_026972] |
| A_23_P134684 | 0,027169727 | up | 1,5220101 | HMBOX1 | Homo sapiens homeobox containing 1 (HMBOX1), transcript variant 1, mRNA [NM_024567] |
| A_21_P0013605 | 0,035852224 | down | -1,5103077 | HMCN2 | Homo sapiens hemicentin 2 (HMCN2), mRNA [NM_001291815] |
| A_33_P3224730 | 0,009908535 | down | -1,5565445 | HMCN2 | Homo sapiens hemicentin 2 (HMCN2), mRNA [NM_001291815] |
| A_33_P3421530 | 0,001042806 | up | 2,0377245 | HMCN2 | Homo sapiens cDNA FLJ46139 fis, clone TESTI2052670, weakly similar to Basement membrane-specific heparan sulfate proteoglycan core protein precursor. [AK128020] |
| A_24_P75127 | 0,004806795 | up | 1,6700169 | HMCN2 | Homo sapiens hemicentin 2 (HMCN2), mRNA [NM_001291815] |
| A_21_P0000710 | 0,002478603 | down | -1,8883015 | HMGA1P7 | Homo sapiens high mobility group AT-hook 1 pseudogene 7 (HMGA1P7), non-coding RNA [NR_037938] |
| A_33_P3211078 | 0,028041389 | down | -1,6358004 | HMGA2 | Homo sapiens high mobility group AT-hook 2 (HMGA2), transcript variant 3, mRNA [NM_001300918] |
| A_33_P3237359 | 0,01537244 | up | 1,559645 | HMGB3 | Homo sapiens high mobility group box 3 (HMGB3), transcript variant 1, mRNA [NM_001301228] |
| A_33_P3319041 | 0,010771482 | up | 1,7462195 | HMGB3 | Homo sapiens high mobility group box 3 (HMGB3), transcript variant 1, mRNA [NM_001301228] |
| A_33_P3388283 | 0,007038314 | down | -1,663555 | HMGCL | 3-hydroxymethyl-3-methylglutaryl-CoA lyase [Source:HGNC Symbol;Acc:HGNC:5005] [ENST00000496907] |
| A_32_P49508 | 0,010349119 | down | -2,4455123 | HMGCLL1 | Homo sapiens 3-hydroxymethyl-3-methylglutaryl-CoA lyase-like 1 (HMGCLL1), transcript variant 1, mRNA [NM_019036] |
| A_33_P3214012 | 0,018191556 | down | -2,0473406 | HMGCLL1 | Homo sapiens 3-hydroxymethyl-3-methylglutaryl-CoA lyase-like 1 (HMGCLL1), transcript variant 4, mRNA [NM_001287746] |
| A_21_P0014727 | 0,015775474 | up | 1,8452727 | HMGN3-AS1 | Homo sapiens HMGN3 antisense RNA 1 (HMGN3-AS1), long non-coding RNA [NR_040671] |
| A_23_P431981 | 7,33E-04 | up | 1,5532157 | HMGXB4 | Homo sapiens HMG box domain containing 4 (HMGXB4), transcript variant 1, mRNA [NM_001003681] |
| A_33_P3318414 | 0,021162491 | up | 1,528641 | HMHA1 | Homo sapiens histocompatibility (minor) HA-1 (HMHA1), transcript variant 1, mRNA [NM_012292] |
| A_23_P30204 | 9,84E-04 | down | -2,650677 | HMHB1 | Homo sapiens histocompatibility (minor) HB-1 (HMHB1), mRNA [NM_021182] |
| A_23_P319005 | 0,001155234 | down | -2,1018016 | HMP19 | Homo sapiens HMP19 protein (HMP19), mRNA [NM_015980] |
| A_33_P3279009 | 0,03273818 | down | -1,5753238 | HMX1 | Homo sapiens H6 family homeobox 1 (HMX1), mRNA [NM_018942] |
| A_33_P3335940 | 4,05E-05 | down | -2,3070016 | HN1L | Homo sapiens hematological and neurological expressed 1-like (HN1L), mRNA [NM_144570] |
| A_24_P314515 | 9,94E-05 | down | -3,1421816 | HNF1A-AS1 | Homo sapiens HNF1A antisense RNA 1 (HNF1A-AS1), long non-coding RNA [NR_024345] |
| A_24_P330822 | 3,02E-04 | down | -3,4937778 | HNF1B | HNF1 homeobox B [Source:HGNC Symbol;Acc:HGNC:11630] [ENST00000621123] |

| | | | | | |
|---|---|---|---|---|---|
| A_23_P28761 | 0,03739146 | up | 2,0764575 | HNF4A | Homo sapiens hepatocyte nuclear factor 4, alpha (HNF4A), transcript variant 6, mRNA [NM_001030004] |
| A_23_P251453 | 0,006440555 | up | 1,6304468 | HNF4G | Homo sapiens hepatocyte nuclear factor 4, gamma (HNF4G), mRNA [NM_004133] |
| A_24_P517901 | 0,009569193 | down | -2,0638452 | HNRNPA1 | Homo sapiens heterogeneous nuclear ribonucleoprotein A1 (HNRNPA1), transcript variant 1, mRNA [NM_002136] |
| A_32_P8015 | 0,001842846 | up | 1,5374743 | HNRNPA1L2 | Homo sapiens heterogeneous nuclear ribonucleoprotein A1-like 2 (HNRNPA1L2), transcript variant 1, mRNA [NM_001011724] |
| A_24_P269895 | 5,79E-04 | down | -1,7847922 | HNRNPA3 | Homo sapiens heterogeneous nuclear ribonucleoprotein A3 (HNRNPA3), mRNA [NM_194247] |
| A_33_P3422170 | 9,78E-06 | down | -3,2656121 | HNRNPA3 | Homo sapiens heterogeneous nuclear ribonucleoprotein A3 (HNRNPA3), mRNA [NM_194247] |
| A_24_P706314 | 0,008936658 | up | 1,5978988 | HNRNPA3 | Homo sapiens heterogeneous nuclear ribonucleoprotein A3 (HNRNPA3), mRNA [NM_194247] |
| A_24_P136161 | 2,97E-04 | down | -3,1033063 | HNRNPCL1 | Homo sapiens heterogeneous nuclear ribonucleoprotein C-like 1 (HNRNPCL1), mRNA [NM_001013631] |
| A_23_P11286 | 0,009180428 | up | 1,5284913 | HNRNPH2 | Homo sapiens heterogeneous nuclear ribonucleoprotein H2 (H') (HNRNPH2), transcript variant 1, mRNA [NM_019597] |
| A_33_P3380161 | 0,008996571 | up | 1,7229505 | HNRNPU-AS1 | Homo sapiens HNRNPU antisense RNA 1 (HNRNPU-AS1), long non-coding RNA [NR_026778] |
| A_23_P99906 | 5,02E-04 | down | -1,6013273 | HOMER2 | Homo sapiens homer homolog 2 (Drosophila) (HOMER2), transcript variant 2, mRNA [NM_199330] |
| A_33_P3350673 | 0,023553602 | down | -3,9377358 | HOPX | Homo sapiens HOP homeobox (HOPX), transcript variant 5, mRNA [NM_001145460] |
| A_19_P00330464 | 0,009405563 | down | -2,0319715 | HOTAIR | Homo sapiens HOX transcript antisense RNA (HOTAIR), transcript variant 3, long non-coding RNA [NR_047518] |
| A_19_P00329806 | 0,007263965 | down | -3,069209 | HOTAIR | Homo sapiens HOX transcript antisense RNA (HOTAIR), transcript variant 3, long non-coding RNA [NR_047518] |
| A_21_P0010502 | 0,013289575 | up | 2,5695338 | HOTAIR | Homo sapiens HOX transcript antisense RNA (HOTAIR), transcript variant 3, long non-coding RNA [NR_047518] |
| A_19_P00330814 | 0,019982494 | up | 1,8002514 | HOTAIR | Homo sapiens HOX transcript antisense RNA (HOTAIR), transcript variant 3, long non-coding RNA [NR_047518] |
| A_21_P0000690 | 0,005479139 | down | -1,7699054 | HOTTIP | Homo sapiens HOXA distal transcript antisense RNA (HOTTIP), long non-coding RNA [NR_037843] |
| A_23_P303978 | 0,007206545 | up | 3,384505 | HOXA-AS2 | PREDICTED: Homo sapiens HOXA cluster antisense RNA 2 (HOXA-AS2), transcript variant X3, misc_RNA [XR_426154] |
| A_33_P3288649 | 2,56E-06 | down | -3,6138618 | HOXA10 | Homo sapiens homeobox A10 (HOXA10), transcript variant 1, mRNA [NM_018951] |
| A_23_P93772 | 0,008921609 | down | -1,6315465 | HOXA5 | Homo sapiens homeobox A5 (HOXA5), mRNA [NM_019102] |
| A_33_P3416231 | 0,0287716 | down | -1,5529215 | HOXA9 | Homo sapiens homeobox A9 (HOXA9), mRNA [NM_152739] |
| A_21_P0000588 | 1,47E-04 | down | -2,2999432 | HOXB-AS3 | Homo sapiens HOXB cluster antisense RNA 3 (HOXB-AS3), transcript variant 8, long non-coding RNA [NR_110331] |
| A_33_P3256760 | 0,010889646 | up | 2,1349678 | HOXB-AS3 | Homo sapiens HOXB cluster antisense RNA 3 (HOXB-AS3), transcript variant 1, long non-coding RNA [NR_033201] |
| A_23_P55281 | 0,010001525 | up | 2,893467 | HOXB7 | Homo sapiens homeobox B7 (HOXB7), mRNA [NM_004502] |
| A_23_P335981 | 0,013365089 | up | 2,8749177 | HOXC12 | Homo sapiens homeobox C12 (HOXC12), mRNA [NM_173860] |
| A_33_P3300965 | 0,03152497 | down | -2,2361546 | HOXC6 | Homo sapiens homeobox C6 (HOXC6), transcript variant 1, mRNA [NM_004503] |
| A_21_P0000804 | 0,012406716 | up | 2,0707414 | HOXD-AS2 | Homo sapiens HOXD cluster antisense RNA 2 (HOXD-AS2), long non-coding RNA [NR_038435] |
| A_23_P429383 | 1,74E-05 | down | -3,625333 | HOXD9 | Homo sapiens homeobox D9 (HOXD9), mRNA [NM_014213] |
| A_33_P3333238 | 0,002704259 | up | 3,0535908 | HOXD9 | Homo sapiens homeobox D9 (HOXD9), mRNA [NM_014213] |
| A_23_P206760 | 0,009594685 | down | -1,5299377 | HP | Homo sapiens haptoglobin (HP), transcript variant 1, mRNA [NM_005143] |
| A_33_P3246133 | 0,006856144 | up | 1,5597274 | HPCA | Homo sapiens hippocalcin (HPCA), mRNA [NM_002143] |
| A_33_P3244568 | 8,42E-04 | down | -2,7627234 | HPN-AS1 | Homo sapiens HPN antisense RNA 1 (HPN-AS1), transcript variant 1, long non-coding RNA [NR_024561] |
| A_23_P109442 | 0,002167652 | up | 1,6348284 | HPS4 | Homo sapiens Hermansky-Pudlak syndrome 4 (HPS4), transcript variant 1, mRNA [NM_022081] |
| A_24_P103893 | 0,019114943 | up | 1,57316 | HPSE2 | Homo sapiens heparanase 2 (inactive) (HPSE2), transcript variant 1, mRNA [NM_021828] |
| A_33_P3390955 | 0,002757762 | down | -3,8761911 | HPVC1 | Homo sapiens human papillomavirus (type 18) E5 central sequence-like 1 (HPVC1), long non-coding RNA [NR_004422] |
| A_33_P3324206 | 0,025605764 | up | 2,5948868 | HR | Homo sapiens hair growth associated (HR), transcript variant 1, mRNA [NM_005144] |
| A_23_P105012 | 0,037146356 | up | 2,8592906 | HRASLS2 | Homo sapiens HRAS-like suppressor 2 (HRASLS2), mRNA [NM_017878] |
| A_23_P53018 | 1,78E-04 | down | -2,2246108 | HRASLS5 | Homo sapiens HRAS-like suppressor family, member 5 (HRASLS5), transcript variant 1, mRNA [NM_054108] |
| A_23_P416314 | 0,009686125 | down | -2,8701372 | HRASLS5 | Homo sapiens HRAS-like suppressor family, member 5 (HRASLS5), transcript variant 1, mRNA [NM_054108] |
| A_23_P386310 | 0,005372807 | up | 1,617661 | HRH4 | Homo sapiens histamine receptor H4 (HRH4), transcript variant 1, mRNA [NM_021624] |
| A_24_P8220 | 2,57E-04 | up | 1,7303046 | HS6ST1 | Homo sapiens heparan sulfate 6-O-sulfotransferase 1 (HS6ST1), mRNA [NM_004807] |
| A_33_P3269328 | 0,03421331 | up | 1,783919 | HS6ST2-AS1 | Homo sapiens HS6ST2 antisense RNA 1 (HS6ST2-AS1), long non-coding RNA [NR_046691] |
| A_23_P32629 | 0,049749732 | up | 2,1596513 | HS6ST3 | Homo sapiens heparan sulfate 6-O-sulfotransferase 3 (HS6ST3), mRNA [NM_153456] |
| A_33_P3309231 | 2,52E-04 | down | -1,789975 | HSCB | Homo sapiens HscB mitochondrial iron-sulfur cluster co-chaperone (HSCB), mRNA [NM_172002] |
| A_23_P14986 | 0,003507377 | up | 2,675781 | HSD11B2 | Homo sapiens hydroxysteroid (11-beta) dehydrogenase 2 (HSD11B2), mRNA [NM_000196] |
| A_33_P3335042 | 0,002377223 | down | -1,5743148 | HSD17B12 | Homo sapiens hydroxysteroid (17-beta) dehydrogenase 12 (HSD17B12), mRNA [NM_016142] |
| A_23_P78762 | 0,035159968 | down | -1,6458961 | HSD17B14 | Homo sapiens hydroxysteroid (17-beta) dehydrogenase 14 (HSD17B14), mRNA [NM_016246] |
| A_23_P81973 | 0,006237744 | down | -1,649735 | HSD17B8 | Homo sapiens hydroxysteroid (17-beta) dehydrogenase 8 (HSD17B8), mRNA [NM_014234] |
| A_24_P350397 | 0,005073893 | up | 2,3265784 | HSD3B1 | Homo sapiens hydroxy-delta-5-steroid dehydrogenase, 3 beta- and steroid delta-isomerase 1 (HSD3B1), mRNA [NM_000862] |
| A_23_P51580 | 0,03669871 | down | -1,9014846 | HSD3B2 | Homo sapiens hydroxy-delta-5-steroid dehydrogenase, 3 beta- and steroid delta-isomerase 2 (HSD3B2), transcript variant 1, mRNA [NM_000198] |
| A_23_P206324 | 0,004155602 | up | 1,5423712 | HSDL1 | Homo sapiens hydroxysteroid dehydrogenase like 1 (HSDL1), transcript variant 1, mRNA [NM_031463] |
| A_23_P143512 | 0,04611166 | down | -1,7398057 | HSF2BP | Homo sapiens heat shock transcription factor 2 binding protein (HSF2BP), mRNA [NM_007031] |
| A_23_P96497 | 0,04778582 | down | -1,838555 | HSFX1 | Homo sapiens heat shock transcription factor family, X linked 1 (HSFX1), mRNA [NM_016153] |
| A_33_P3262660 | 0,012649986 | up | 1,8607665 | HSFY1P1 | Homo sapiens heat shock transcription factor, Y-linked 1 pseudogene 1 (HSFY1P1), non-coding RNA [NR_003607] |
| A_21_P0012242 | 0,014924848 | up | 1,6260266 | HSFY1P1 | Homo sapiens heat shock transcription factor, Y-linked 1 pseudogene 1 (HSFY1P1), non-coding RNA [NR_003607] |
| A_23_P45579 | 0,009164658 | up | 1,6165117 | HSFY2 | Homo sapiens heat shock transcription factor, Y linked 2 (HSFY2), transcript variant 1, mRNA [NM_153716] |
| A_23_P153372 | 0,031574614 | up | 1,5518726 | HSH2D | Homo sapiens hematopoietic SH2 domain containing (HSH2D), transcript variant 1, mRNA [NM_032855] |
| A_33_P3408212 | 0,04187005 | down | -1,8331393 | HSP90B1 | Homo sapiens heat shock protein 90kDa beta (Grp94), member 1 (HSP90B1), mRNA [NM_003299] |
| A_23_P70547 | 0,027085125 | up | 1,5092076 | HSPA1L | Homo sapiens heat shock 70kDa protein 1-like (HSPA1L), mRNA [NM_005527] |
| A_24_P308029 | 0,005017571 | down | -3,0115173 | HSPB6 | Homo sapiens heat shock protein, alpha-crystallin-related, B6 (HSPB6), mRNA [NM_144617] |
| A_24_P307580 | 0,035332657 | up | 1,7057303 | HTATIP2 | Homo sapiens HIV-1 Tat interactive protein 2, 30kDa (HTATIP2), transcript variant 5, mRNA [NM_001098523] |

| | | | | | |
|---|---|---|---|---|---|
| A_33_P3212665 | 7,37E-04 | down | -2,0434484 | HTATSF1P2 | Homo sapiens HIV-1 Tat specific factor 1 pseudogene 2 (HTATSF1P2), non-coding RNA [NR_033884] |
| A_24_P97687 | 0,01675707 | down | -2,0159528 | HTR1A | Homo sapiens 5-hydroxytryptamine (serotonin) receptor 1A, G protein-coupled (HTR1A), mRNA [NM_000524] |
| A_23_P156824 | 0,028811073 | down | -2,3053637 | HTR1B | Homo sapiens 5-hydroxytryptamine (serotonin) receptor 1B, G protein-coupled (HTR1B), mRNA [NM_000863] |
| A_33_P3319176 | 0,002895417 | up | 1,5695919 | HTR2C | 5-hydroxytryptamine (serotonin) receptor 2C, G protein-coupled [Source:HGNC Symbol;Acc:HGNC:5295] [ENST00000371951] |
| A_24_P150068 | 2,32E-04 | down | -2,7729204 | HTR3A | Homo sapiens 5-hydroxytryptamine (serotonin) receptor 3A, ionotropic (HTR3A), transcript variant 1, mRNA [NM_213621] |
| A_23_P87007 | 0,02688598 | up | 1,809479 | HTR3B | Homo sapiens 5-hydroxytryptamine (serotonin) receptor 3B, ionotropic (HTR3B), mRNA [NM_006028] |
| A_24_P32139 | 0,004944449 | down | -2,0978289 | HTR3E | Homo sapiens 5-hydroxytryptamine (serotonin) receptor 3E, ionotropic (HTR3E), transcript variant 1, mRNA [NM_182589] |
| A_23_P133637 | 0,002001995 | down | -3,1842644 | HTR4 | Homo sapiens 5-hydroxytryptamine (serotonin) receptor 4, G protein-coupled (HTR4), transcript variant d, mRNA [NM_001040172] |
| A_33_P3345016 | 0,01483987 | down | -2,499635 | HTR6 | Homo sapiens 5-hydroxytryptamine (serotonin) receptor 6, G protein-coupled (HTR6), mRNA [NM_000871] |
| A_24_P160070 | 0,006929396 | up | 1,7487901 | HUNK | Homo sapiens hormonally up-regulated Neu-associated kinase (HUNK), mRNA [NM_014586] |
| A_33_P3811287 | 2,54E-04 | up | 1,665934 | HUWE1 | Homo sapiens HECT, UBA and WWE domain containing 1, E3 ubiquitin protein ligase (HUWE1), mRNA [NM_031407] |
| A_33_P3276856 | 0,002949173 | down | -2,9663448 | HYDIN | Homo sapiens HYDIN, axonemal central pair apparatus protein (HYDIN), transcript variant 3, mRNA [NM_001198542] |
| A_33_P3402983 | 0,009250621 | up | 1,6178532 | HYDIN | Homo sapiens HYDIN, axonemal central pair apparatus protein (HYDIN), transcript variant 1, mRNA [NM_001270974] |
| A_33_P3257538 | 0,002038517 | down | -1,8686112 | HYKK | Homo sapiens hydroxylysine kinase (HYKK), transcript variant 2, mRNA [NM_001083612] |
| A_33_P3308481 | 0,004046568 | up | 2,1251822 | HYMAI | Homo sapiens hydatidiform mole associated and imprinted (non-protein coding) (HYMAI), long non-coding RNA [NR_002768] |
| A_23_P139648 | 4,48E-05 | down | -2,112569 | IAPP | Homo sapiens islet amyloid polypeptide (IAPP), mRNA [NM_000415] |
| A_33_P3209346 | 0,021434665 | down | -1,7575938 | IARS | Homo sapiens isoleucyl-tRNA synthetase (IARS), transcript variant 2, mRNA [NM_013417] |
| A_24_P211797 | 0,041219816 | down | -1,7998426 | ICA1L | Homo sapiens islet cell autoantigen 1,69kDa-like (ICA1L), transcript variant 5, mRNA [NM_001288624] |
| A_23_P430168 | 0,003701409 | up | 1,5722562 | ICA1L | islet cell autoantigen 1,69kDa-like [Source:HGNC Symbol;Acc:HGNC:14442] [ENST00000484561] |
| A_21_P0014085 | 0,018689714 | down | -1,9524608 | ID2-AS1 | Homo sapiens ID2 antisense RNA 1 (head to head) (ID2-AS1), transcript variant 2, long non-coding RNA [NR_110154] |
| A_21_P0014098 | 0,03247594 | down | -1,7489321 | IDH1-AS1 | Homo sapiens IDH1 antisense RNA 1 (IDH1-AS1), long non-coding RNA [NR_046452] |
| A_24_P261929 | 0,001310512 | down | -1,602756 | IFI27L1 | Homo sapiens interferon, alpha-inducible protein 27-like 1 (IFI27L1), transcript variant 2, mRNA [NM_206949] |
| A_23_P152782 | 0,01758987 | down | -1,5925759 | IFI35 | Homo sapiens interferon-induced protein 35 (IFI35), mRNA [NM_005533] |
| A_23_P157920 | 0,006975321 | up | 1,6171018 | IFNA16 | Homo sapiens interferon, alpha 16 (IFNA16), mRNA [NM_002173] |
| A_24_P403459 | 0,015372827 | down | -1,6661605 | IFNA4 | Homo sapiens interferon, alpha 4 (IFNA4), mRNA [NM_021068] |
| A_23_P29036 | 0,002323007 | up | 1,5480002 | IFNGR2 | Homo sapiens interferon gamma receptor 2 (interferon gamma transducer 1) (IFNGR2), mRNA [NM_005534] |
| A_23_P74112 | 0,037152972 | up | 1,5013652 | IFNLR1 | Homo sapiens interferon, lambda receptor 1 (IFNLR1), transcript variant 1, mRNA [NM_170743] |
| A_24_P362881 | 0,03465929 | up | 2,0243392 | IFT57 | Homo sapiens intraflagellar transport 57 (IFT57), mRNA [NM_018010] |
| A_23_P48339 | 0,019701743 | down | -1,6018773 | IFT88 | Homo sapiens intraflagellar transport 88 (IFT88), transcript variant 1, mRNA [NM_175605] |
| A_33_P3312819 | 0,019067045 | down | -1,7233582 | IGF2-AS | Homo sapiens IGF2 antisense RNA (IGF2-AS), transcript variant 1, long non-coding RNA [NR_028044] |
| A_23_P116435 | 1,47E-06 | down | -4,0888147 | IGF2-AS | Homo sapiens IGF2 antisense RNA (IGF2-AS), transcript variant 1, long non-coding RNA [NR_028044] |
| A_23_P250156 | 3,46E-04 | up | 1,9915376 | IGF2BP2 | Homo sapiens insulin-like growth factor 2 mRNA binding protein 2 (IGF2BP2), transcript variant 1, mRNA [NM_006548] |
| A_33_P3324949 | 0,010903144 | down | -2,0044863 | IGF2BP2-AS1 | Homo sapiens cDNA FLJ32900 fis, clone TESTI2005492. [AK057462] |
| A_23_P42868 | 0,033802975 | up | 1,7320195 | IGFBP1 | Homo sapiens insulin-like growth factor binding protein 1 (IGFBP1), mRNA [NM_000596] |
| A_23_P383009 | 0,031098576 | down | -2,0642736 | IGFBP5 | Homo sapiens insulin-like growth factor binding protein 5 (IGFBP5), mRNA [NM_000599] |
| A_33_P3240602 | 9,14E-04 | down | -2,6778626 | IGFL2 | Homo sapiens cDNA FLJ43766 fis, clone TESTI2049246. [AK125754] |
| A_24_P110558 | 0,002433789 | up | 1,778843 | IGIP | Homo sapiens IgA-inducing protein (IGIP), mRNA [NM_001007189] |
| A_33_P3761915 | 0,001529081 | down | -1,9244936 | IGKV1D-13 | 602502772F1 NIH_MGC_77 Homo sapiens cDNA clone IMAGE:4616320 5', mRNA sequence [BG482625] |
| A_24_P239076 | 0,01606543 | up | 1,5508348 | IGLL1 | Homo sapiens immunoglobulin lambda-like polypeptide 1 (IGLL1), transcript variant 1, mRNA [NM_020070] |
| A_23_P125303 | 0,010426924 | up | 2,3475902 | IGSF1 | Homo sapiens immunoglobulin superfamily, member 1 (IGSF1), transcript variant 1, mRNA [NM_001555] |
| A_21_P0011816 | 0,006135663 | down | -1,6586212 | IGSF3 | Homo sapiens immunoglobulin superfamily, member 3 (IGSF3), transcript variant 2, mRNA [NM_001007237] |
| A_21_P0012337 | 0,012646686 | down | -1,9985874 | IGSF3 | Homo sapiens immunoglobulin superfamily, member 3 (IGSF3), transcript variant 2, mRNA [NM_001007237] |
| A_33_P3372004 | 0,001524354 | up | 3,0648525 | IGSF6 | Homo sapiens immunoglobulin superfamily, member 6 (IGSF6), mRNA [NM_005849] |
| A_24_P132518 | 0,001148596 | up | 1,6198292 | IKBKB | Homo sapiens inhibitor of kappa light polypeptide gene enhancer in B-cells, kinase beta (IKBKB), transcript variant 1, mRNA [NM_001556] |
| A_33_P3241756 | 0,010314094 | up | 1,745149 | IKBKB | inhibitor of kappa light polypeptide gene enhancer in B-cells, kinase beta [Source:HGNC Symbol;Acc:HGNC:5960] [ENST00000517917] |
| A_33_P3224795 | 0,007069314 | up | 1,6368266 | IKZF5 | Homo sapiens IKAROS family zinc finger 5 (Pegasus) (IKZF5), transcript variant 1, mRNA [NM_001271840] |
| A_33_P3421520 | 0,00315331 | up | 1,6816151 | IKZF5 | Homo sapiens IKAROS family zinc finger 5 (Pegasus) (IKZF5), transcript variant 1, mRNA [NM_001271840] |
| A_23_P126735 | 0,045100823 | down | -1,6721869 | IL10 | Homo sapiens interleukin 10 (IL10), mRNA [NM_000572] |
| A_23_P91943 | 0,008145737 | up | 1,6686361 | IL12A | Homo sapiens interleukin 12A (IL12A), mRNA [NM_000882] |
| A_23_P72077 | 0,030955851 | up | 2,240614 | IL12RB2 | Homo sapiens interleukin 12 receptor, beta 2 (IL12RB2), transcript variant 1, mRNA [NM_001559] |
| A_33_P3399267 | 0,03253483 | up | 1,9193676 | IL15RA | Homo sapiens interleukin 15 receptor, alpha (IL15RA), transcript variant 2, mRNA [NM_172200] |
| A_24_P299423 | 0,010321686 | up | 1,5832777 | IL16 | interleukin 16 [Source:HGNC Symbol;Acc:HGNC:5980] [ENST00000560241] |
| A_23_P332820 | 1,38E-05 | down | -5,28419 | IL17A | Homo sapiens interleukin 17A (IL17A), mRNA [NM_002190] |
| A_23_P167882 | 0,015199741 | down | -2,2103593 | IL17F | Homo sapiens interleukin 17F (IL17F), mRNA [NM_052872] |
| A_33_P3224809 | 0,03583215 | up | 1,5187645 | IL17RA | Homo sapiens interleukin 17 receptor A (IL17RA), transcript variant 1, mRNA [NM_014339] |
| A_32_P188860 | 0,009478212 | up | 1,9564458 | IL17RD | Homo sapiens interleukin 17 receptor D (IL17RD), mRNA [NM_017563] |
| A_23_P104798 | 0,002976272 | up | 1,8122756 | IL18 | Homo sapiens interleukin 18 (IL18), transcript variant 1, mRNA [NM_001562] |
| A_33_P3251876 | 0,016259922 | down | -2,0582955 | IL18R1 | Homo sapiens interleukin 18 receptor 1 (IL18R1), transcript variant 1, mRNA [NM_003855] |
| A_33_P3211666 | 0,00116263 | down | -2,300176 | IL18R1 | Homo sapiens interleukin 18 receptor 1 (IL18R1), transcript variant 1, mRNA [NM_003855] |
| A_24_P208567 | 0,01877997 | down | -1,7998474 | IL18R1 | Homo sapiens interleukin 18 receptor 1 (IL18R1), transcript variant 1, mRNA [NM_003855] |
| A_33_P3221960 | 0,011177207 | down | -2,0176184 | IL18RAP | Homo sapiens interleukin 18 receptor accessory protein (IL18RAP), mRNA [NM_003853] |

| | | | | | |
|---|---|---|---|---|---|
| A_21_P0000070 | 0,0157305 | down | -1,8858325 | IL1RAP | Homo sapiens interleukin 1 receptor accessory protein (IL1RAP), transcript variant 6, mRNA [NM_001167931] |
| A_33_P3246829 | 0,044434246 | up | 1,716623 | IL1RN | Homo sapiens interleukin 1 receptor antagonist (IL1RN), transcript variant 4, mRNA [NM_173843] |
| A_23_P46482 | 0,034308128 | up | 2,0915449 | IL20 | Homo sapiens interleukin 20 (IL20), mRNA [NM_018724] |
| A_33_P3677020 | 2,17E-04 | down | -2,045414 | IL21R-AS1 | Homo sapiens IL21R antisense RNA 1 (IL21R-AS1), long non-coding RNA [NR_037158] |
| A_33_P3290780 | 0,002509799 | up | 3,5172255 | IL24 | Homo sapiens interleukin 24 (IL24), transcript variant 3, mRNA [NM_001185156] |
| A_33_P3284933 | 6,81E-04 | down | -1,7218941 | IL27 | Homo sapiens interleukin 27 (IL27), mRNA [NM_145659] |
| A_23_P148473 | 0,004959405 | down | -2,2535973 | IL2RG | Homo sapiens interleukin 2 receptor, gamma (IL2RG), mRNA [NM_000206] |
| A_33_P3691168 | 0,00113698 | down | -2,4043922 | IL31RA | Homo sapiens interleukin 31 receptor A (IL31RA), transcript variant 7, mRNA [NM_001297572] |
| A_32_P217750 | 0,001589689 | up | 2,2130914 | IL3RA | Homo sapiens interleukin 3 receptor, alpha (low affinity) (IL3RA), transcript variant 1, mRNA [NM_002183] |
| A_23_P213706 | 0,003143349 | up | 1,8149196 | IL4 | Homo sapiens interleukin 4 (IL4), transcript variant 1, mRNA [NM_000589] |
| A_33_P3328254 | 0,002448818 | up | 1,607555 | IL5RA | Homo sapiens interleukin 5 receptor, alpha (IL5RA), transcript variant 3, mRNA [NM_175725] |
| A_21_P0000171 | 0,025667274 | up | 1,6281902 | IL6R | Homo sapiens interleukin 6 receptor (IL6R), transcript variant 3, mRNA [NM_001206866] |
| A_23_P404494 | 0,004672748 | down | -3,3858967 | IL7R | Homo sapiens interleukin 7 receptor (IL7R), transcript variant 1, mRNA [NM_002185] |
| A_23_P109907 | 0,014406681 | down | -1,6118205 | ILDR1 | Homo sapiens immunoglobulin-like domain containing receptor 1 (ILDR1), transcript variant 2, mRNA [NM_175924] |
| A_33_P3219745 | 0,00181664 | up | 3,1036162 | ILF2 | interleukin enhancer binding factor 2 [Source:HGNC Symbol;Acc:HGNC:6037] [ENST00000368681] |
| A_23_P434944 | 0,001673754 | up | 1,6360632 | ILF3 | Homo sapiens interleukin enhancer binding factor 3, 90kDa (ILF3), transcript variant 2, mRNA [NM_004516] |
| A_33_P3314468 | 0,003313095 | down | -1,5327259 | IMMT | Homo sapiens inner membrane protein, mitochondrial (IMMT), transcript variant 3, mRNA [NM_001100170] |
| A_23_P321034 | 7,31E-04 | up | 1,5692724 | INADL | Homo sapiens InaD-like (Drosophila) (INADL), mRNA [NM_176877] |
| A_23_P116387 | 0,00631718 | up | 1,6778177 | INCENP | Homo sapiens inner centromere protein antigens 135/155kDa (INCENP), transcript variant 1, mRNA [NM_001040694] |
| A_23_P153964 | 0,034590907 | up | 1,6755359 | INHBB | Homo sapiens inhibin, beta B (INHBB), mRNA [NM_002193] |
| A_24_P155502 | 2,80E-04 | down | -4,241902 | INHBC | Homo sapiens inhibin, beta C (INHBC), mRNA [NM_005538] |
| A_24_P141707 | 0,003215164 | down | -3,1024585 | INHBE | Homo sapiens inhibin, beta E (INHBE), mRNA [NM_031479] |
| A_33_P3326822 | 0,03087595 | down | -1,5025284 | INMT | indolethylamine N-methyltransferase [Source:HGNC Symbol;Acc:HGNC:6069] [ENST00000461246] |
| A_33_P3330716 | 0,001911539 | up | 1,5748763 | INO80D | Homo sapiens INO80 complex subunit D (INO80D), mRNA [NM_017759] |
| A_24_P317835 | 0,001065744 | up | 1,6896206 | INPP5E | Homo sapiens inositol polyphosphate-5-phosphatase, 72 kDa (INPP5E), mRNA [NM_019892] |
| A_23_P1981 | 0,005422625 | down | -2,7124164 | INS | Homo sapiens insulin (INS), transcript variant 1, mRNA [NM_000207] |
| A_33_P3814721 | 0,006613183 | up | 3,1322021 | INSC | Homo sapiens inscuteable homolog (Drosophila) (INSC), transcript variant 1, mRNA [NM_001031853] |
| A_24_P944458 | 8,63E-04 | up | 1,5913138 | INSIG2 | Homo sapiens insulin induced gene 2 (INSIG2), mRNA [NM_016133] |
| A_23_P83012 | 0,003850275 | up | 1,6707414 | INSL6 | Homo sapiens insulin-like 6 (INSL6), mRNA [NM_007179] |
| A_33_P3335725 | 0,002509327 | up | 2,00769 | INSR | Homo sapiens insulin receptor (INSR), transcript variant 1, mRNA [NM_000208] |
| A_23_P420269 | 0,001734262 | up | 1,769534 | INTS2 | Homo sapiens integrator complex subunit 2 (INTS2), transcript variant 1, mRNA [NM_020748] |
| A_23_P126757 | 0,003251835 | up | 1,6706553 | INTS3 | Homo sapiens integrator complex subunit 3 (INTS3), mRNA [NM_023015] |
| A_33_P3320804 | 5,93E-04 | down | -1,8044443 | IP6K1 | Homo sapiens inositol hexakisphosphate kinase 1 (IP6K1), transcript variant 3, mRNA [NM_001242829] |
| A_24_P940059 | 0,008110121 | up | 1,7877446 | IPCEF1 | Homo sapiens interaction protein for cytohesin exchange factors 1 (IPCEF1), transcript variant 3, mRNA [NM_015553] |
| A_23_P115036 | 0,003704742 | up | 1,5571901 | IPO13 | Homo sapiens importin 13 (IPO13), mRNA [NM_014652] |
| A_19_P00802141 | 0,007965564 | up | 1,5040046 | IPO7 | Homo sapiens importin 7 (IPO7), mRNA [NM_006391] |
| A_33_P3360917 | 0,03173101 | up | 1,606928 | IPO9 | Homo sapiens importin 9 (IPO9), mRNA [NM_018085] |
| A_23_P19852 | 0,030193271 | up | 1,6077543 | IQCE | Homo sapiens IQ motif containing E (IQCE), transcript variant 1, mRNA [NM_152558] |
| A_32_P165933 | 8,46E-04 | down | -2,9810503 | IQCF2 | Homo sapiens IQ motif containing F2 (IQCF2), mRNA [NM_203424] |
| A_21_P0000726 | 0,020541223 | up | 2,6988564 | IQCF4 | Homo sapiens IQ motif containing F5 pseudogene (IQCF4), transcript variant 3, non-coding RNA [NR_038215] |
| A_33_P3209581 | 7,34E-04 | down | -3,0256817 | IQSEC3 | Homo sapiens IQ motif and Sec7 domain 3 (IQSEC3), transcript variant 1, mRNA [NM_001170738] |
| A_24_P203298 | 0,004053007 | up | 1,5862942 | IQUB | Homo sapiens IQ motif and ubiquitin domain containing (IQUB), transcript variant 1, mRNA [NM_178827] |
| A_32_P21474 | 0,006831886 | down | -1,7194406 | IRAK1BP1 | Homo sapiens interleukin-1 receptor-associated kinase 1 binding protein 1 (IRAK1BP1), mRNA [NM_001010844] |
| A_33_P3352970 | 0,001574826 | up | 1,9624866 | IRAK2 | Homo sapiens interleukin-1 receptor-associated kinase 2 (IRAK2), mRNA [NM_001570] |
| A_23_P162300 | 0,034158435 | up | 2,292764 | IRAK3 | Homo sapiens interleukin-1 receptor-associated kinase 3 (IRAK3), transcript variant 1, mRNA [NM_007199] |
| A_24_P497186 | 2,89E-04 | up | 1,6401263 | IRF2BP2 | Homo sapiens interferon regulatory factor 2 binding protein 2 (IRF2BP2), transcript variant 1, mRNA [NM_182972] |
| A_23_P354547 | 8,60E-04 | up | 2,4298775 | IRF9 | interferon regulatory factor 9 [Source:HGNC Symbol;Acc:HGNC:6131] [ENST00000558468] |
| A_23_P65442 | 5,45E-04 | up | 1,7469696 | IRF9 | Homo sapiens interferon regulatory factor 9 (IRF9), mRNA [NM_006084] |
| A_33_P3359022 | 0,006748874 | up | 3,2121844 | IRGC | Homo sapiens immunity-related GTPase family, cinema (IRGC), mRNA [NM_019612] |
| A_33_P3364038 | 3,44E-04 | up | 3,423585 | IRGM | Homo sapiens immunity-related GTPase family, M (IRGM), mRNA [NM_001145805] |
| A_24_P225679 | 0,04138997 | up | 1,5800693 | IRS1 | Homo sapiens insulin receptor substrate 1 (IRS1), mRNA [NM_005544] |
| A_23_P156025 | 0,00359778 | up | 1,6172497 | IRX2 | Homo sapiens iroquois homeobox 2 (IRX2), transcript variant 1, mRNA [NM_033267] |
| A_33_P3297562 | 0,001465897 | up | 1,8113352 | IRX2 | Homo sapiens iroquois homeobox 2 (IRX2), transcript variant 1, mRNA [NM_033267] |
| A_24_P387609 | 0,020372847 | up | 1,9411589 | ISCA1 | Homo sapiens iron-sulfur cluster assembly 1 (ISCA1), mRNA [NM_030940] |
| A_23_P81529 | 0,019934908 | down | -2,0678053 | ISL1 | Homo sapiens ISL LIM homeobox 1 (ISL1), mRNA [NM_002202] |
| A_23_P3312 | 0,020137306 | down | -2,125707 | ISLR | Homo sapiens immunoglobulin superfamily containing leucine-rich repeat (ISLR), transcript variant 1, mRNA [NM_005545] |
| A_23_P206759 | 4,51E-04 | up | 2,1346786 | IST1 | Homo sapiens increased sodium tolerance 1 homolog (yeast) (IST1), transcript variant 2, mRNA [NM_014761] |
| A_23_P206022 | 8,76E-04 | down | -1,8895589 | ITGA11 | Homo sapiens integrin, alpha 11 (ITGA11), mRNA [NM_001004439] |
| A_23_P252193 | 0,001025169 | down | -1,9867948 | ITGA9 | Homo sapiens integrin, alpha 9 (ITGA9), mRNA [NM_002207] |
| A_24_P366652 | 1,16E-04 | down | -2,513293 | ITGAD | Homo sapiens integrin, alpha D (ITGAD), mRNA [NM_005353] |
| A_23_P124108 | 0,001232402 | down | -3,5196462 | ITGAM | Homo sapiens integrin, alpha M (complement component 3 receptor 3 subunit) (ITGAM), transcript variant 2, mRNA [NM_000632] |

| | | | | | |
|---|---|---|---|---|---|
| A_23_P312132 | 0,013147151 | down | -2,0367305 | ITGAX | Homo sapiens integrin, alpha X (complement component 3 receptor 4 subunit) (ITGAX), transcript variant 2, mRNA [NM_000887] |
| A_33_P3216227 | 0,025302926 | up | 2,444929 | ITGB1BP1 | integrin beta 1 binding protein 1 [Source:HGNC Symbol;Acc:HGNC:23927] [ENST00000360635] |
| A_23_P45424 | 0,013169362 | up | 1,5521921 | ITGB1BP2 | Homo sapiens integrin beta 1 binding protein (melusin) 2 (ITGB1BP2), transcript variant 1, mRNA [NM_012278] |
| A_23_P430411 | 0,022813078 | down | -1,6540679 | ITGB2 | integrin, beta 2 (complement component 3 receptor 3 and 4 subunit) [Source:HGNC Symbol;Acc:HGNC:6155] [ENST00000610622] |
| A_21_P0000759 | 1,75E-06 | down | -5,2900457 | ITGB2-AS1 | Homo sapiens ITGB2 antisense RNA 1 (ITGB2-AS1), transcript variant 1, long non-coding RNA [NR_038311] |
| A_24_P220855 | 0,008061946 | up | 1,6843562 | ITIH5 | Homo sapiens inter-alpha-trypsin inhibitor heavy chain family, member 5 (ITIH5), transcript variant 3, mRNA [NM_001001851] |
| A_24_P292253 | 0,005352265 | up | 1,5454142 | ITIH5 | Homo sapiens inter-alpha-trypsin inhibitor heavy chain family, member 5 (ITIH5), transcript variant 1, mRNA [NM_030569] |
| A_33_P3363071 | 7,18E-05 | down | -2,4234066 | ITIH6 | Homo sapiens inter-alpha-trypsin inhibitor heavy chain family, member 6 (ITIH6), mRNA [NM_198510] |
| A_24_P202567 | 3,43E-04 | up | 1,7551434 | ITPKC | Homo sapiens inositol-trisphosphate 3-kinase C (ITPKC), mRNA [NM_025194] |
| A_23_P19517 | 7,02E-05 | up | 1,9720839 | ITPR3 | Homo sapiens inositol 1,4,5-trisphosphate receptor, type 3 (ITPR3), mRNA [NM_002224] |
| A_33_P3269718 | 0,04000611 | down | -1,5930502 | IVD | Homo sapiens isovaleryl-CoA dehydrogenase (IVD), transcript variant 2, mRNA [NM_001159508] |
| A_24_P206702 | 0,015672725 | down | -1,8684233 | IZUMO1R | Homo sapiens IZUMO1 receptor, JUNO (IZUMO1R), mRNA [NM_001199206] |
| A_23_P101521 | 0,005823297 | down | -2,1478875 | IZUMO2 | Homo sapiens IZUMO family member 2 (IZUMO2), mRNA [NM_152358] |
| A_33_P3282230 | 0,030880097 | up | 2,845994 | IZUMO3 | Homo sapiens IZUMO family member 3 (IZUMO3), mRNA [NM_001271706] |
| A_33_P3221114 | 0,0016005 | up | 2,7967262 | JAKMIP1 | Homo sapiens janus kinase and microtubule interacting protein 1 (JAKMIP1), transcript variant 2, mRNA [NM_144720] |
| A_33_P3352088 | 0,001488103 | up | 3,230823 | JAKMIP3 | Homo sapiens Janus kinase and microtubule interacting protein 3 (JAKMIP3), mRNA [NM_001105521] |
| A_33_P3340224 | 0,018708527 | down | -1,8299342 | JAZF1-AS1 | Homo sapiens JAZF1 antisense RNA 1 (JAZF1-AS1), long non-coding RNA [NR_034097] |
| A_23_P311616 | 4,48E-04 | up | 1,807664 | JMJD6 | Homo sapiens jumonji domain containing 6 (JMJD6), transcript variant 2, mRNA [NM_015167] |
| A_33_P3220929 | 0,022468219 | down | -1,7113514 | JOSD2 | Homo sapiens Josephin domain containing 2 (JOSD2), transcript variant 3, mRNA [NM_001270641] |
| A_23_P394395 | 2,51E-04 | up | 2,8895512 | JPH2 | Homo sapiens junctophilin 2 (JPH2), transcript variant 1, mRNA [NM_020433] |
| A_23_P397320 | 0,013712353 | up | 2,936744 | JSRP1 | Homo sapiens junctional sarcoplasmic reticulum protein 1 (JSRP1), mRNA [NM_144616] |
| A_24_P241815 | 0,022797745 | up | 1,844299 | JUNB | Homo sapiens jun B proto-oncogene (JUNB), mRNA [NM_002229] |
| A_23_P500130 | 0,00268663 | up | 1,6832846 | KANK1 | Homo sapiens KN motif and ankyrin repeat domains 1 (KANK1), transcript variant 2, mRNA [NM_153186] |
| A_23_P50426 | 8,93E-05 | up | 1,9528491 | KANK2 | Homo sapiens KN motif and ankyrin repeat domains 2 (KANK2), transcript variant 1, mRNA [NM_015493] |
| A_33_P3326020 | 0,012822941 | down | -1,8918476 | KANK3 | KN motif and ankyrin repeat domains 3 [Source:HGNC Symbol;Acc:HGNC:24796] [ENST00000593649] |
| A_19_P00809076 | 0,023317711 | down | -1,5426443 | KANSL1-AS1 | Homo sapiens KANSL1 antisense RNA 1 (KANSL1-AS1), long non-coding RNA [NR_034172] |
| A_32_P159651 | 0,038649257 | up | 2,015811 | KAT2B | Homo sapiens K(lysine) acetyltransferase 2B (KAT2B), mRNA [NM_003884] |
| A_21_P0014695 | 7,40E-04 | down | -2,180699 | KATNAL2 | PREDICTED: Homo sapiens katanin p60 subunit A-like 2 (KATNAL2), transcript variant X2, mRNA [XM_005258358] |
| A_33_P3396129 | 0,01530918 | down | -1,8324571 | KATNB1 | katanin p80 (WD repeat containing) subunit B 1 [Source:HGNC Symbol;Acc:HGNC:6217] [ENST00000566611] |
| A_33_P3295029 | 0,027394993 | up | 2,070759 | KBTBD12 | Homo sapiens kelch repeat and BTB (POZ) domain containing 12 (KBTBD12), mRNA [NM_207335] |
| A_33_P3306192 | 5,56E-04 | down | -1,7332128 | KBTBD13 | Homo sapiens kelch repeat and BTB (POZ) domain containing 13 (KBTBD13), mRNA [NM_001101362] |
| A_24_P12539 | 2,84E-05 | up | 2,2051759 | KBTBD2 | Homo sapiens kelch repeat and BTB (POZ) domain containing 2 (KBTBD2), mRNA [NM_015483] |
| A_23_P25605 | 0,00165452 | up | 1,7885392 | KBTBD7 | Homo sapiens kelch repeat and BTB (POZ) domain containing 7 (KBTBD7), mRNA [NM_032138] |
| A_23_P48229 | 0,030473076 | up | 2,183206 | KCNA1 | Homo sapiens potassium channel, voltage gated shaker related subfamily A, member 1 (KCNA1), mRNA [NM_000217] |
| A_24_P498652 | 0,03311174 | up | 1,5751976 | KCNA1 | Homo sapiens potassium channel, voltage gated shaker related subfamily A, member 1 (KCNA1), mRNA [NM_000217] |
| A_23_P126528 | 0,025002083 | down | -1,8944923 | KCNA10 | Homo sapiens potassium channel, voltage gated shaker related subfamily A, member 10 (KCNA10), mRNA [NM_005549] |
| A_19_P00803558 | 0,017993955 | down | -1,8183241 | KCNA6 | Homo sapiens potassium channel, voltage gated shaker related subfamily A, member 6 (KCNA6), mRNA [NM_002235] |
| A_24_P182929 | 0,003607938 | down | -2,2805445 | KCNAB1 | Homo sapiens potassium channel, voltage gated subfamily A regulatory beta subunit 1 (KCNAB1), transcript variant 2, mRNA [NM_003471] |
| A_33_P3237379 | 0,005980671 | up | 1,528615 | KCNC1 | Homo sapiens potassium channel, voltage gated Shaw related subfamily C, member 1 (KCNC1), transcript variant 2, mRNA [NM_004976] |
| A_33_P3338793 | 0,002044239 | down | -2,766663 | KCNC3 | Homo sapiens potassium channel, voltage gated Shaw related subfamily C, member 3 (KCNC3), mRNA [NM_004977] |
| A_23_P101516 | 0,004299892 | down | -1,802982 | KCNC3 | Homo sapiens potassium channel, voltage gated Shaw related subfamily C, member 3 (KCNC3), mRNA [NM_004977] |
| A_23_P12079 | 0,002350793 | down | -1,6121246 | KCNC4 | Homo sapiens potassium channel, voltage gated Shaw related subfamily C, member 4 (KCNC4), transcript variant 3, mRNA [NM_001039574] |
| A_21_P0001165 | 0,007664676 | up | 2,2003453 | KCNC4-AS1 | Homo sapiens KCNC4 antisense RNA 1 (head to head) (KCNC4-AS1), long non-coding RNA [NR_046546] |
| A_23_P154855 | 0,024369469 | down | -1,735821 | KCNE1 | Homo sapiens potassium voltage-gated channel, Isk-related family, member 1 (KCNE1), transcript variant 2, mRNA [NM_000219] |
| A_33_P3246448 | 1,69E-05 | down | -2,2173588 | KCNE4 | Homo sapiens potassium channel, voltage gated subfamily E regulatory beta subunit 4 (KCNE4), mRNA [NM_080671] |
| A_23_P256641 | 0,002490502 | down | -2,5361445 | KCNE5 | Homo sapiens potassium channel, voltage gated subfamily E regulatory beta subunit 5 (KCNE5), mRNA [NM_012282] |
| A_23_P377882 | 9,42E-04 | down | -3,8824236 | KCNH2 | Homo sapiens potassium voltage-gated channel, subfamily H (eag-related), member 2 (KCNH2), transcript variant 2, mRNA [NM_172056] |
| A_33_P3253672 | 0,010540944 | down | -1,7743999 | KCNH3 | Homo sapiens potassium channel, voltage gated eag related subfamily H, member 3 (KCNH3), mRNA [NM_012284] |
| A_33_P3314857 | 0,002971171 | up | 1,7871902 | KCNJ13 | Homo sapiens potassium channel, inwardly rectifying subfamily J, member 13 (KCNJ13), transcript variant 1, mRNA [NM_002242] |
| A_23_P130764 | 0,004105751 | up | 1,5269767 | KCNJ14 | Homo sapiens potassium channel, inwardly rectifying subfamily J, member 14 (KCNJ14), mRNA [NM_013348] |
| A_23_P501193 | 0,00730458 | down | -1,7539414 | KCNJ16 | Homo sapiens potassium channel, inwardly rectifying subfamily J, member 16 (KCNJ16), transcript variant 2, mRNA [NM_170741] |
| A_33_P3374778 | 0,001621819 | down | -2,4872687 | KCNJ2-AS1 | Homo sapiens KCNJ2 antisense RNA 1 (head to head) (KCNJ2-AS1), long non-coding RNA [NR_036534] |
| A_23_P29057 | 3,65E-04 | up | 2,7525916 | KCNJ6 | Homo sapiens potassium channel, inwardly rectifying subfamily J, member 6 (KCNJ6), mRNA [NM_002240] |
| A_24_P112032 | 0,001694284 | up | 1,9100335 | KCNK17 | Homo sapiens potassium channel, two pore domain subfamily K, member 17 (KCNK17), transcript variant 1, mRNA [NM_031460] |
| A_23_P319423 | 0,022692606 | down | -1,9197071 | KCNK5 | Homo sapiens potassium channel, two pore domain subfamily K, member 5 (KCNK5), mRNA [NM_003740] |
| A_23_P111978 | 0,003435899 | down | -3,1017575 | KCNK9 | Homo sapiens potassium channel, two pore domain subfamily K, member 9 (KCNK9), transcript variant 1, mRNA [NM_001282534] |
| A_24_P206121 | 0,001872998 | down | -1,6520807 | KCNMB1 | Homo sapiens potassium channel subfamily M regulatory beta subunit 1 (KCNMB1), mRNA [NM_004137] |
| A_23_P500353 | 0,025271948 | down | -2,0468364 | KCNN2 | Homo sapiens potassium channel, calcium activated intermediate/small conductance subfamily N alpha, member 2 (KCNN2), transcript variant 1, mRNA [NM_021614] |
| A_23_P67529 | 0,040988214 | up | 2,169673 | KCNN4 | Homo sapiens potassium channel, calcium activated intermediate/small conductance subfamily N alpha, member 4 (KCNN4), mRNA [NM_002250] |
| A_33_P3247331 | 0,02588216 | up | 2,013642 | KCNQ1 | potassium voltage-gated channel, KQT-like subfamily, member 1 [Source:HGNC Symbol;Acc:HGNC:6294] [ENST00000345015] |
| A_33_P3360753 | 0,003169136 | down | -1,5962949 | KCNQ2 | potassium voltage-gated channel, KQT-like subfamily, member 2 [Source:HGNC Symbol;Acc:HGNC:6296] [ENST00000370221] |

| | | | | | |
|---|---|---|---|---|---|
| A_33_P3395823 | 7,59E-05 | down | -5,3014545 | KCNQ2 | Homo sapiens potassium channel, voltage gated KQT-like subfamily Q, member 2 (KCNQ2), transcript variant 5, mRNA [NM_172109] |
| A_23_P34424 | 0,004937258 | down | -3,1257198 | KCNQ4 | Homo sapiens potassium channel, voltage gated KQT-like subfamily Q, member 4 (KCNQ4), transcript variant 1, mRNA [NM_004700] |
| A_33_P3339481 | 0,00879766 | up | 2,02252 | KCNQ5 | potassium voltage-gated channel, KQT-like subfamily, member 5 [Source:HGNC Symbol;Acc:HGNC:6299] [ENST00000370392] |
| A_23_P394377 | 0,001783268 | up | 2,0775702 | KCNRG | Homo sapiens potassium channel regulator (KCNRG), transcript variant 2, mRNA [NM_199464] |
| A_33_P3281613 | 0,005022806 | down | -1,7147918 | KCNT1 | Homo sapiens cDNA FLJ41282 fis, clone BRAMY2037823. [AK123276] |
| A_23_P162165 | 0,048158426 | down | -1,9417099 | KCTD14 | Homo sapiens potassium channel tetramerization domain containing 14 (KCTD14), transcript variant 1, mRNA [NM_023930] |
| A_24_P373844 | 0,005817683 | up | 1,7453177 | KCTD15 | Homo sapiens potassium channel tetramerization domain containing 15 (KCTD15), transcript variant 1, mRNA [NM_024076] |
| A_24_P307808 | 0,00391396 | down | -2,4891002 | KCTD16 | Homo sapiens potassium channel tetramerization domain containing 16 (KCTD16), mRNA [NM_020768] |
| A_33_P3255131 | 0,001361013 | down | -2,552722 | KCTD19 | Homo sapiens potassium channel tetramerization domain containing 19 (KCTD19), mRNA [NM_001100915] |
| A_24_P324011 | 0,008922031 | down | -1,5242391 | KCTD2 | Homo sapiens potassium channel tetramerization domain containing 2 (KCTD2), transcript variant 1, mRNA [NM_015353] |
| A_23_P160406 | 0,003268658 | up | 1,5889231 | KCTD3 | Homo sapiens potassium channel tetramerization domain containing 3 (KCTD3), mRNA [NM_016121] |
| A_23_P385246 | 0,001701486 | up | 1,6992344 | KCTD6 | Homo sapiens potassium channel tetramerization domain containing 6 (KCTD6), transcript variant 1, mRNA [NM_153331] |
| A_33_P3298960 | 0,048527516 | down | -1,7296618 | KDELC2 | Homo sapiens KDEL (Lys-Asp-Glu-Leu) containing 2 (KDELC2), mRNA [NM_153705] |
| A_24_P93754 | 0,003555759 | up | 2,234351 | KDM4C | lysine (K)-specific demethylase 4C [Source:HGNC Symbol;Acc:HGNC:17071] [ENST00000381306] |
| A_33_P3340718 | 0,031532235 | up | 1,5025992 | KDM6A | Homo sapiens lysine (K)-specific demethylase 6A (KDM6A), transcript variant 1, mRNA [NM_001291415] |
| A_24_P942630 | 0,006956785 | up | 1,68332 | KDM6B | Homo sapiens lysine (K)-specific demethylase 6B (KDM6B), mRNA [NM_001080424] |
| A_23_P359897 | 0,003200207 | up | 1,9619479 | KDM7A | Homo sapiens lysine (K)-specific demethylase 7A (KDM7A), mRNA [NM_030647] |
| A_33_P3806721 | 0,031026712 | down | -1,5671064 | KDM8 | Homo sapiens lysine (K)-specific demethylase 8 (KDM8), transcript variant 1, mRNA [NM_001145348] |
| A_32_P86905 | 3,27E-05 | down | -3,2722602 | KGFLP2 | Homo sapiens keratinocyte growth factor-like protein 2 (KGFLP2), non-coding RNA [NR_003670] |
| A_19_P00325810 | 0,023336338 | down | -1,9546651 | KHDC1 | Homo sapiens KH homology domain containing 1 (KHDC1), transcript variant 3, non-coding RNA [NR_027005] |
| A_33_P3394727 | 0,011118636 | up | 2,2089422 | KHDRBS3 | Homo sapiens KH domain containing, RNA binding, signal transduction associated 3 (KHDRBS3), mRNA [NM_006558] |
| A_24_P134235 | 0,017630188 | down | -1,5254782 | KHSRP | Homo sapiens KH-type splicing regulatory protein (KHSRP), mRNA [NM_003685] |
| A_24_P79617 | 0,016722454 | up | 1,7737057 | KIAA0040 | Homo sapiens KIAA0040 (KIAA0040), transcript variant 2, mRNA [NM_014656] |
| A_23_P213369 | 0,00168678 | down | -1,6433421 | KIAA0141 | Homo sapiens KIAA0141 (KIAA0141), transcript variant 1, mRNA [NM_014773] |
| A_23_P304171 | 0,001105014 | up | 1,9433224 | KIAA0226 | Homo sapiens KIAA0226 (KIAA0226), transcript variant 1, mRNA [NM_001145642] |
| A_23_P324490 | 0,001412007 | up | 1,5876878 | KIAA0355 | Homo sapiens KIAA0355 (KIAA0355), mRNA [NM_014686] |
| A_24_P912372 | 5,64E-05 | up | 2,714933 | KIAA0754 | Homo sapiens KIAA0754 (KIAA0754), mRNA [NM_015038] |
| A_23_P74467 | 4,24E-04 | up | 2,1971421 | KIAA0907 | Homo sapiens KIAA0907 (KIAA0907), mRNA [NM_014949] |
| A_24_P325176 | 2,45E-04 | up | 1,8764067 | KIAA1109 | Homo sapiens KIAA1109 (KIAA1109), mRNA [NM_015312] |
| A_23_P393880 | 0,009121237 | up | 1,8931862 | KIAA1244 | Homo sapiens KIAA1244 (KIAA1244), mRNA [NM_020340] |
| A_32_P163739 | 0,021933738 | down | -1,9898101 | KIAA1257 | Homo sapiens KIAA1257 (KIAA1257), mRNA [NM_020741] |
| A_33_P3256793 | 0,011772674 | down | -2,008441 | KIAA1324 | Homo sapiens KIAA1324 (KIAA1324), transcript variant 1, mRNA [NM_020775] |
| A_23_P64121 | 0,048027918 | up | 1,8075274 | KIAA1549L | Homo sapiens KIAA1549-like (KIAA1549L), mRNA [NM_012194] |
| A_23_P98930 | 0,003942034 | up | 1,7568319 | KIAA1551 | Homo sapiens KIAA1551 (KIAA1551), mRNA [NM_018169] |
| A_24_P101047 | 0,034226704 | down | -2,5048814 | KIAA1586 | Homo sapiens KIAA1586 (KIAA1586), transcript variant 1, mRNA [NM_020931] |
| A_33_P3382236 | 0,012024356 | down | -1,6863831 | KIAA1598 | Homo sapiens KIAA1598 (KIAA1598), transcript variant 4, mRNA [NM_001258299] |
| A_32_P176594 | 0,002390355 | down | -2,447257 | KIAA1614 | Homo sapiens KIAA1614 (KIAA1614), mRNA [NM_020950] |
| A_23_P309619 | 1,93E-04 | up | 2,2849314 | KIAA1671 | Homo sapiens KIAA1671 (KIAA1671), mRNA [NM_001145206] |
| A_19_P00805272 | 0,009414494 | up | 1,5493217 | KIAA1671 | Homo sapiens KIAA1671 (KIAA1671), mRNA [NM_001145206] |
| A_33_P3267482 | 0,018449912 | down | -2,229552 | KIAA1804 | Homo sapiens mixed lineage kinase 4 (KIAA1804), mRNA [NM_032435] |
| A_24_P265265 | 0,008679732 | up | 2,0363975 | KIAA1804 | Mitogen-activated protein kinase kinase kinase MLK4 [Source:UniProtKB/Swiss-Prot;Acc:Q5TCX8] [ENST00000366623] |
| A_33_P3290955 | 0,043903414 | down | -1,5877137 | KIAA1875 | Homo sapiens KIAA1875 (KIAA1875), long non-coding RNA [NR_024207] |
| A_33_P3307945 | 0,009050739 | up | 1,922386 | KIAA2012 | Homo sapiens KIAA2012 (KIAA2012), mRNA [NM_001277372] |
| A_24_P97145 | 0,014629508 | up | 1,6042181 | KIDINS220 | Homo sapiens kinase D-interacting substrate, 220kDa (KIDINS220), mRNA [NM_020738] |
| A_23_P147388 | 0,007999312 | up | 2,1238241 | KIF13B | Homo sapiens kinesin family member 13B (KIF13B), mRNA [NM_015254] |
| A_23_P400515 | 0,04488299 | up | 1,629718 | KIF17 | Homo sapiens kinesin family member 17 (KIF17), transcript variant 1, mRNA [NM_020816] |
| A_24_P233014 | 0,013659262 | up | 2,5639615 | KIF19 | Homo sapiens kinesin family member 19 (KIF19), mRNA [NM_153209] |
| A_24_P145066 | 3,91E-06 | up | 2,443308 | KIF1B | Homo sapiens kinesin family member 1B (KIF1B), transcript variant 2, mRNA [NM_183416] |
| A_23_P201287 | 0,001660282 | up | 1,5956471 | KIF1B | Homo sapiens kinesin family member 1B (KIF1B), transcript variant 1, mRNA [NM_015074] |
| A_33_P3221665 | 0,001838957 | down | -2,2452626 | KIF21B | Homo sapiens kinesin family member 21B (KIF21B), transcript variant 3, mRNA [NM_001252102] |
| A_33_P3350638 | 0,003604109 | down | -1,8013489 | KIF22 | kinesin family member 22 [Source:HGNC Symbol;Acc:HGNC:6391] [ENST00000563666] |
| A_33_P3396746 | 8,85E-05 | down | -2,7450712 | KIF26B | kinesin family member 26B [Source:HGNC Symbol;Acc:HGNC:25484] [ENST00000479506] |
| A_33_P3382565 | 0,001647349 | down | -2,0453825 | KIF26B | Homo sapiens kinesin family member 26B (KIF26B), mRNA [NM_018012] |
| A_23_P218025 | 0,029503416 | down | -1,6846617 | KIF5A | Homo sapiens kinesin family member 5A (KIF5A), mRNA [NM_004984] |
| A_24_P93948 | 0,004013184 | up | 1,9720426 | KIF5A | kinesin family member 5A [Source:HGNC Symbol;Acc:HGNC:6323] [ENST00000455537] |
| A_24_P225878 | 0,048428725 | up | 2,017116 | KIF9 | Homo sapiens kinesin family member 9 (KIF9), transcript variant 1, mRNA [NM_022342] |
| A_24_P350622 | 0,007968188 | down | -2,1430786 | KIR2DL4 | Homo sapiens killer cell immunoglobulin-like receptor, two domains, long cytoplasmic tail, 4 (KIR2DL4), transcript variant 1, mRNA [NM_002255] |
| A_23_P502590 | 0,011922343 | down | -2,290757 | KIR2DS4 | Homo sapiens killer cell immunoglobulin-like receptor, two domains, short cytoplasmic tail, 4 (KIR2DS4), transcript variant 1, mRNA [NM_012314] |
| A_24_P117147 | 1,25E-06 | down | -4,5118985 | KIR3DL1 | Homo sapiens killer cell immunoglobulin-like receptor, three domains, long cytoplasmic tail, 1 (KIR3DL1), mRNA [NM_013289] |
| A_33_P3233010 | 8,38E-06 | down | -2,2276783 | KIR3DL2 | Homo sapiens killer cell immunoglobulin-like receptor, three domains, long cytoplasmic tail, 2 (KIR3DL2), transcript variant 2, mRNA [NM_001242867] |
| A_23_P370574 | 0,02456653 | down | -2,3750045 | KIR3DL3 | Homo sapiens killer cell immunoglobulin-like receptor, three domains, long cytoplasmic tail, 3 (KIR3DL3), mRNA [NM_153443] |

| | | | | | |
|---|---|---|---|---|---|
| A_33_P3340926 | 0,022898423 | down | -1,6445712 | KIR3DX1 | Homo sapiens killer cell immunoglobulin-like receptor, three domains, X1 (KIR3DX1), transcript variant 1, non-coding RNA [NR_026716] |
| A_23_P124892 | 0,01763964 | down | -1,9058623 | KISS1 | Homo sapiens KiSS-1 metastasis-suppressor (KISS1), mRNA [NM_002256] |
| A_24_P133253 | 0,017830791 | down | -2,431596 | KITLG | Homo sapiens KIT ligand (KITLG), transcript variant b, mRNA [NM_000899] |
| A_23_P503064 | 0,038456183 | up | 1,6591477 | KL | Homo sapiens klotho (KL), mRNA [NM_004795] |
| A_24_P227211 | 0,001971516 | down | -1,7002665 | KLC2 | Homo sapiens kinesin light chain 2 (KLC2), transcript variant 1, mRNA [NM_022822] |
| A_33_P3288805 | 0,008660758 | down | -1,6146739 | KLC2 | Homo sapiens kinesin light chain 2 (KLC2), transcript variant 2, mRNA [NM_001134774] |
| A_33_P3410925 | 4,26E-04 | down | -2,221677 | KLF1 | Homo sapiens Kruppel-like factor 1 (erythroid) (KLF1), mRNA [NM_006563] |
| A_23_P168828 | 0,011095034 | up | 1,8240104 | KLF10 | Homo sapiens Kruppel-like factor 10 (KLF10), transcript variant 1, mRNA [NM_005655] |
| A_23_P44505 | 0,0092587 | up | 1,9485897 | KLF11 | Homo sapiens Kruppel-like factor 11 (KLF11), transcript variant 1, mRNA [NM_003597] |
| A_19_P00321061 | 0,007539997 | up | 2,6221404 | KLF13 | Kruppel-like factor 13 [Source:HGNC Symbol;Acc:HGNC:13672] [ENST00000500533] |
| A_23_P30163 | 0,00698861 | up | 2,1173794 | KLF3-AS1 | Homo sapiens KLF3 antisense RNA 1 (KLF3-AS1), long non-coding RNA [NR_026804] |
| A_23_P53891 | 0,030135188 | up | 1,6753246 | KLF5 | Homo sapiens Kruppel-like factor 5 (intestinal) (KLF5), transcript variant 1, mRNA [NM_001730] |
| A_23_P63798 | 0,002779183 | up | 2,418726 | KLF6 | Homo sapiens Kruppel-like factor 6 (KLF6), transcript variant A, mRNA [NM_001300] |
| A_23_P415401 | 4,71E-05 | up | 2,4729486 | KLF9 | Homo sapiens Kruppel-like factor 9 (KLF9), mRNA [NM_001206] |
| A_24_P343255 | 0,010856547 | up | 1,535066 | KLHDC3 | Homo sapiens kelch domain containing 3 (KLHDC3), transcript variant 1, mRNA [NM_057161] |
| A_33_P3417765 | 0,006523673 | down | -2,2455633 | KLHDC7A | Homo sapiens kelch domain containing 7A (KLHDC7A), mRNA [NM_152375] |
| A_24_P242609 | 0,004774055 | up | 1,5175129 | KLHL12 | Homo sapiens kelch-like family member 12 (KLHL12), transcript variant 2, mRNA [NM_021633] |
| A_23_P159974 | 2,76E-04 | down | -2,1351867 | KLHL13 | Homo sapiens kelch-like family member 13 (KLHL13), transcript variant 1, mRNA [NM_033495] |
| A_23_P72503 | 0,042915124 | up | 1,8162782 | KLHL2 | Homo sapiens kelch-like family member 2 (KLHL2), transcript variant 1, mRNA [NM_007246] |
| A_23_P341223 | 4,62E-04 | up | 1,7054403 | KLHL21 | Homo sapiens kelch-like family member 21 (KLHL21), mRNA [NM_014851] |
| A_24_P521994 | 0,00211257 | up | 1,8394332 | KLHL24 | Homo sapiens kelch-like family member 24 (KLHL24), mRNA [NM_017644] |
| A_33_P3399618 | 0,04681998 | down | -1,5641695 | KLHL33 | Homo sapiens kelch-like family member 33 (KLHL33), mRNA [NM_001109997] |
| A_24_P645765 | 9,31E-05 | up | 1,737309 | KLHL42 | Homo sapiens kelch-like family member 42 (KLHL42), mRNA [NM_020782] |
| A_24_P944253 | 0,018423297 | down | -2,3165352 | KLHL6 | Homo sapiens kelch-like family member 6 (KLHL6), mRNA [NM_130446] |
| A_23_P324994 | 0,001881584 | up | 1,5058286 | KLHL7 | Homo sapiens kelch-like family member 7 (KLHL7), transcript variant 2, mRNA [NM_018846] |
| A_33_P3251065 | 0,027889302 | up | 1,6546538 | KLK13 | kallikrein-related peptidase 13 [Source:HGNC Symbol;Acc:HGNC:6361] [ENST00000602090] |
| A_33_P3417640 | 0,018454894 | up | 3,137429 | KLK14 | Homo sapiens kallikrein-related peptidase 14 (KLK14), mRNA [NM_022046] |
| A_23_P310257 | 2,88E-04 | down | -2,764699 | KLK2 | Homo sapiens kallikrein-related peptidase 2 (KLK2), transcript variant 2, mRNA [NM_001002231] |
| A_33_P3215843 | 0,001132771 | up | 1,8901061 | KLK3 | Homo sapiens kallikrein-related peptidase 3 (KLK3), transcript variant 3, mRNA [NM_001030047] |
| A_24_P376047 | 0,021676516 | down | -1,970522 | KLK4 | Homo sapiens kallikrein-related peptidase 4 (KLK4), transcript variant 1, mRNA [NM_004917] |
| A_24_P236935 | 0,023031501 | up | 2,1099305 | KLK6 | Homo sapiens kallikrein-related peptidase 6 (KLK6), transcript variant B, mRNA [NM_001012964] |
| A_21_P0007870 | 3,60E-04 | down | -1,8859109 | KLRAP1 | Homo sapiens killer cell lectin-like receptor subfamily A pseudogene 1 (KLRAP1), non-coding RNA [NR_028045] |
| A_23_P99275 | 0,032236006 | up | 1,6653702 | KLRB1 | Homo sapiens killer cell lectin-like receptor subfamily B, member 1 (KLRB1), mRNA [NM_002258] |
| A_23_P139654 | 0,003491028 | down | -2,4679215 | KLRC1 | Homo sapiens killer cell lectin-like receptor subfamily C, member 1 (KLRC1), transcript variant 2, mRNA [NM_007328] |
| A_23_P128281 | 1,42E-04 | up | 3,0703955 | KLRC3 | Homo sapiens killer cell lectin-like receptor subfamily C, member 3 (KLRC3), transcript variant 2, mRNA [NM_007333] |
| A_23_P218058 | 0,001312448 | down | -2,3314788 | KLRC4 | Homo sapiens killer cell lectin-like receptor subfamily C, member 4 (KLRC4), mRNA [NM_013431] |
| A_23_P64898 | 0,004674304 | down | -2,2530773 | KLRG1 | Homo sapiens killer cell lectin-like receptor subfamily G, member 1 (KLRG1), mRNA [NM_005810] |
| A_33_P3306207 | 0,001092403 | down | -1,993609 | KLRG2 | Homo sapiens killer cell lectin-like receptor subfamily G, member 2 (KLRG2), mRNA [NM_198508] |
| A_23_P200838 | 2,40E-05 | down | -2,4878786 | KMO | Homo sapiens kynurenine 3-monooxygenase (kynurenine 3-hydroxylase) (KMO), mRNA [NM_003679] |
| A_24_P77082 | 1,04E-04 | down | -4,0521417 | KMO | Homo sapiens kynurenine 3-monooxygenase (kynurenine 3-hydroxylase) (KMO), mRNA [NM_003679] |
| A_33_P3307307 | 0,005004669 | up | 1,6311291 | KMT2A | Homo sapiens lysine (K)-specific methyltransferase 2A (KMT2A), transcript variant 1, mRNA [NM_001197104] |
| A_23_P142421 | 0,00408004 | up | 1,5771136 | KMT2B | Homo sapiens lysine (K)-specific methyltransferase 2B (KMT2B), mRNA [NM_014727] |
| A_33_P3340254 | 0,005059445 | up | 1,608228 | KMT2D | Homo sapiens lysine (K)-specific methyltransferase 2D (KMT2D), mRNA [NM_003482] |
| A_33_P3277407 | 0,008625941 | down | -1,6158193 | KMT2E | Homo sapiens lysine (K)-specific methyltransferase 2E (KMT2E), transcript variant 1, mRNA [NM_182931] |
| A_33_P3275435 | 0,007296328 | down | -1,5644714 | KMT2E-AS1 | Homo sapiens KMT2E antisense RNA 1 (head to head) (KMT2E-AS1), long non-coding RNA [NR_024586] |
| A_33_P3401262 | 0,043342073 | down | -1,6312026 | KNCN | Homo sapiens kinocilin (KNCN), mRNA [NM_001097611] |
| A_23_P136817 | 0,028807037 | up | 1,5946918 | KNTC1 | Homo sapiens kinetochore associated 1 (KNTC1), mRNA [NM_014708] |
| A_23_P156443 | 0,03007271 | down | -2,0611014 | KPNA5 | Homo sapiens karyopherin alpha 5 (importin alpha 6) (KPNA5), mRNA [NM_002269] |
| A_21_P0011637 | 0,023166556 | up | 1,8288679 | KRT16P3 | Homo sapiens keratin 16 pseudogene 3 (KRT16P3), non-coding RNA [NR_029393] |
| A_33_P3398618 | 0,008033396 | down | -2,4820197 | KRT17P5 | Homo sapiens keratin 17 pseudogene 5 (KRT17P5), non-coding RNA [NR_001443] |
| A_33_P3221898 | 0,00906708 | down | -1,9525985 | KRT17P5 | Homo sapiens keratin 17 pseudogene 5 (KRT17P5), non-coding RNA [NR_001443] |
| A_32_P489662 | 0,031928908 | down | -1,55269 | KRT28 | Homo sapiens keratin 28, type I (KRT28), mRNA [NM_181535] |
| A_33_P3341490 | 0,045643225 | down | -1,6641976 | KRT42P | Homo sapiens keratin 42 pseudogene (KRT42P), non-coding RNA [NR_033415] |
| A_33_P3249274 | 0,007293653 | up | 1,8417765 | KRT73-AS1 | Homo sapiens KRT73 antisense RNA 1 (KRT73-AS1), long non-coding RNA [NR_126005] |
| A_33_P3315284 | 0,001877142 | down | -1,7517412 | KRT74 | Homo sapiens keratin 74, type II (KRT74), mRNA [NM_175053] |
| A_24_P142495 | 0,027835682 | down | -1,7227523 | KRTAP1-3 | Homo sapiens keratin associated protein 1-3 (KRTAP1-3), mRNA [NM_030966] |
| A_33_P3254579 | 0,03133491 | down | -2,3173804 | KRTAP1-5 | Homo sapiens keratin associated protein 1-5 (KRTAP1-5), mRNA [NM_031957] |
| A_33_P3293785 | 0,003217299 | down | -1,6902558 | KRTAP10-3 | Homo sapiens keratin associated protein 10-3 (KRTAP10-3), mRNA [NM_198696] |
| A_33_P3334877 | 0,002084116 | down | -3,317111 | KRTAP10-4 | Homo sapiens keratin associated protein 10-4 (KRTAP10-4), mRNA [NM_198687] |
| A_33_P3256272 | 2,14E-04 | down | -3,9907413 | KRTAP10-5 | Homo sapiens keratin associated protein 10-5 (KRTAP10-5), mRNA [NM_198694] |
| A_24_P66312 | 4,27E-04 | down | -3,0264258 | KRTAP12-1 | Homo sapiens keratin associated protein 12-1 (KRTAP12-1), mRNA [NM_181686] |

| | | | | | |
|---|---|---|---|---|---|
| A_33_P3354404 | 0,026921423 | down | -1,5019562 | KRTAP12-2 | Homo sapiens keratin associated protein 12-2 (KRTAP12-2), mRNA [NM_181684] |
| A_24_P314534 | 0,006242979 | down | -2,003012 | KRTAP13-2 | Homo sapiens keratin associated protein 13-2 (KRTAP13-2), mRNA [NM_181621] |
| A_24_P246107 | 0,011898583 | down | -2,3112679 | KRTAP13-4 | Homo sapiens keratin associated protein 13-4 (KRTAP13-4), mRNA [NM_181600] |
| A_33_P3340666 | 0,002101731 | down | -2,1085093 | KRTAP19-5 | Homo sapiens keratin associated protein 19-5 (KRTAP19-5), mRNA [NM_181611] |
| A_24_P76854 | 9,40E-04 | down | -2,5346448 | KRTAP2-1 | Homo sapiens keratin associated protein 2-1 (KRTAP2-1), mRNA [NM_001123387] |
| A_23_P153022 | 0,018012684 | down | -1,896541 | KRTAP2-4 | Homo sapiens keratin associated protein 2-4 (KRTAP2-4), mRNA [NM_033184] |
| A_33_P3247473 | 0,041269552 | down | -2,176554 | KRTAP23-1 | Homo sapiens keratin associated protein 23-1 (KRTAP23-1), mRNA [NM_181624] |
| A_33_P3306619 | 0,020986319 | down | -1,6541213 | KRTAP25-1 | Homo sapiens keratin associated protein 25-1 (KRTAP25-1), mRNA [NM_001128598] |
| A_33_P3382399 | 0,03041392 | down | -1,6884477 | KRTAP3-1 | Homo sapiens keratin associated protein 3-1 (KRTAP3-1), mRNA [NM_031958] |
| A_23_P107454 | 0,003035267 | down | -1,8454458 | KRTAP3-1 | Homo sapiens keratin associated protein 3-1 (KRTAP3-1), mRNA [NM_031958] |
| A_33_P3411315 | 0,002207418 | down | -2,6068997 | KRTAP3-3 | Homo sapiens keratin associated protein 3-3 (KRTAP3-3), mRNA [NM_033185] |
| A_33_P3245348 | 0,006456472 | down | -1,9707732 | KRTAP4-1 | Homo sapiens keratin associated protein 4-1 (KRTAP4-1), mRNA [NM_033060] |
| A_23_P4400 | 0,013716091 | down | -2,6203098 | KRTAP4-11 | Homo sapiens keratin associated protein 4-11 (KRTAP4-11), mRNA [NM_033059] |
| A_23_P27197 | 0,007224562 | down | -2,0422816 | KRTAP4-5 | Homo sapiens keratin associated protein 4-5 (KRTAP4-5), mRNA [NM_033188] |
| A_33_P3256257 | 0,025008855 | down | -2,2223895 | KRTAP4-7 | Homo sapiens keratin associated protein 4-7 (KRTAP4-7), mRNA [NM_033061] |
| A_33_P3317752 | 5,47E-04 | down | -2,6964154 | KRTAP4-7 | Homo sapiens keratin associated protein 4-7 (KRTAP4-7), mRNA [NM_033061] |
| A_33_P3209615 | 0,020038685 | down | -1,8541687 | KRTAP4-8 | Homo sapiens keratin associated protein 4-8 (KRTAP4-8), mRNA [NM_031960] |
| A_33_P3304691 | 0,01030086 | down | -2,1921768 | KRTAP5-1 | Homo sapiens keratin associated protein 5-1 (KRTAP5-1), mRNA [NM_001005922] |
| A_33_P3363310 | 0,004814841 | down | -1,712829 | KRTAP5-10 | keratin associated protein 5-10 [Source:HGNC Symbol;Acc:HGNC:23605] [ENST00000376536] |
| A_33_P3229472 | 0,029148912 | down | -2,0889318 | KRTAP5-10 | Homo sapiens keratin associated protein 5-10 (KRTAP5-10), mRNA [NM_001012710] |
| A_33_P3372742 | 0,013315971 | down | -1,5124658 | KRTAP5-11 | keratin associated protein 5-11 [Source:HGNC Symbol;Acc:HGNC:23606] [ENST00000617152] |
| A_33_P3317317 | 0,001959605 | down | -2,0561798 | KRTAP5-4 | Homo sapiens keratin associated protein 5-4 (KRTAP5-4), mRNA [NM_001012709] |
| A_33_P3304576 | 0,029869568 | down | -1,8333087 | KRTAP5-5 | Homo sapiens keratin associated protein 5-5 (KRTAP5-5), mRNA [NM_001001480] |
| A_33_P3228709 | 2,96E-04 | down | -2,5869048 | KRTAP5-7 | Homo sapiens keratin associated protein 5-7 (KRTAP5-7), mRNA [NM_001012503] |
| A_33_P3263664 | 0,010709509 | down | -2,2726455 | KRTAP6-2 | Homo sapiens keratin associated protein 6-2 (KRTAP6-2), mRNA [NM_181604] |
| A_33_P3352340 | 0,03306264 | down | -1,9157394 | KRTAP9-1 | Homo sapiens keratin associated protein 9-1 (KRTAP9-1), mRNA [NM_001190460] |
| A_33_P3406806 | 1,76E-04 | down | -3,4069166 | KRTAP9-1 | Homo sapiens keratin associated protein 9-1 (KRTAP9-1), mRNA [NM_001190460] |
| A_33_P3358908 | 6,45E-04 | down | -1,9955626 | KRTAP9-6 | Homo sapiens keratin associated protein 9-6 (KRTAP9-6), mRNA [NM_001277331] |
| A_33_P3404123 | 0,01403093 | down | -2,0465748 | KRTAP9-7 | Homo sapiens keratin associated protein 9-7 (KRTAP9-7), mRNA [NM_001277332] |
| A_33_P3307457 | 0,01141333 | up | 1,6556492 | KRTAP9-8 | Homo sapiens keratin associated protein 9-8 (KRTAP9-8), mRNA [NM_031963] |
| A_23_P433132 | 0,005324503 | up | 1,5113947 | KRTCAP3 | Homo sapiens keratinocyte associated protein 3 (KRTCAP3), transcript variant 2, mRNA [NM_173853] |
| A_23_P207774 | 0,040963635 | up | 2,0151267 | KSR1 | Homo sapiens kinase suppressor of ras 1 (KSR1), mRNA [NM_014238] |
| A_33_P3267195 | 0,00120319 | down | -1,6540695 | KU-MEL-3 | Homo sapiens uncharacterized LOC497048 (KU-MEL-3), long non-coding RNA [NR_122107] |
| A_19_P00316107 | 0,027886016 | up | 1,6285424 | L3MBTL1 | Homo sapiens mRNA for KIAA0681 protein, partial cds. [AB014581] |
| A_23_P311885 | 0,005044981 | up | 1,6372061 | L3MBTL3 | Homo sapiens l(3)mbt-like 3 (Drosophila) (L3MBTL3), transcript variant 1, mRNA [NM_032438] |
| A_23_P374322 | 5,62E-05 | down | -2,3650563 | LACC1 | Homo sapiens laccase (multicopper oxidoreductase) domain containing 1 (LACC1), transcript variant 2, mRNA [NM_153218] |
| A_33_P3347482 | 0,014284395 | down | -1,5816962 | LACTBL1 | Homo sapiens lactamase, beta-like 1 (LACTBL1), mRNA [NM_001289974] |
| A_33_P3355230 | 5,85E-04 | down | -4,6951303 | LAIR1 | Homo sapiens leukocyte-associated immunoglobulin-like receptor 1 (LAIR1), transcript variant a, mRNA [NM_002287] |
| A_33_P3391596 | 7,24E-04 | down | -2,8605895 | LAMA4 | Homo sapiens laminin, alpha 4 (LAMA4), transcript variant 1, mRNA [NM_001105206] |
| A_23_P334328 | 0,002092414 | up | 2,2642705 | LAMB4 | Homo sapiens laminin, beta 4 (LAMB4), mRNA [NM_007356] |
| A_33_P3338413 | 0,03535476 | up | 2,676047 | LAMB4 | Homo sapiens laminin, beta 4 (LAMB4), mRNA [NM_007356] |
| A_23_P82979 | 0,010396963 | down | -2,8568778 | LAMC3 | Homo sapiens laminin, gamma 3 (LAMC3), mRNA [NM_006059] |
| A_33_P3238007 | 0,013150224 | down | -1,5923891 | LARP1B | Homo sapiens La ribonucleoprotein domain family, member 1B (LARP1B), transcript variant 2, mRNA [NM_178043] |
| A_23_P117782 | 0,04579296 | down | -1,5046333 | LARP6 | Homo sapiens La ribonucleoprotein domain family, member 6 (LARP6), transcript variant 1, mRNA [NM_018357] |
| A_33_P3237977 | 0,00461089 | down | -1,7431139 | LARP6 | Homo sapiens La ribonucleoprotein domain family, member 6 (LARP6), transcript variant 2, mRNA [NM_197958] |
| A_24_P70002 | 4,06E-04 | up | 1,9345651 | LATS2 | Homo sapiens large tumor suppressor kinase 2 (LATS2), mRNA [NM_014572] |
| A_23_P127565 | 0,013901345 | up | 1,6796892 | LAYN | Homo sapiens layilin (LAYN), transcript variant 2, mRNA [NM_178834] |
| A_23_P120227 | 0,005677398 | up | 2,5130944 | LBH | Homo sapiens limb bud and heart development (LBH), mRNA [NM_030915] |
| A_23_P86493 | 0,002892892 | down | -2,0821714 | LBX1 | Homo sapiens ladybird homeobox 1 (LBX1), mRNA [NM_006562] |
| A_33_P3392517 | 0,015042769 | down | -1,8043176 | LBX2 | Homo sapiens ladybird homeobox 2 (LBX2), transcript variant 2, mRNA [NM_001009812] |
| A_23_P319640 | 0,009354573 | down | -1,594739 | LBX2-AS1 | Homo sapiens LBX2 antisense RNA 1 (LBX2-AS1), long non-coding RNA [NR_024606] |
| A_33_P3356821 | 0,018369041 | down | -1,971476 | LCE1C | Homo sapiens late cornified envelope 1C (LCE1C), transcript variant 1, mRNA [NM_178351] |
| A_23_P375524 | 3,18E-04 | down | -2,7382565 | LCE1D | Homo sapiens late cornified envelope 1D (LCE1D), mRNA [NM_178352] |
| A_33_P3302125 | 0,04598352 | down | -1,936221 | LCE2A | Homo sapiens late cornified envelope 2A (LCE2A), mRNA [NM_178428] |
| A_33_P3365072 | 4,77E-04 | down | -4,2183623 | LCE6A | Homo sapiens late cornified envelope 6A (LCE6A), mRNA [NM_001128600] |
| A_23_P103361 | 6,61E-05 | down | -2,857248 | LCK | Homo sapiens LCK proto-oncogene, Src family tyrosine kinase (LCK), transcript variant 2, mRNA [NM_005356] |
| A_33_P3248539 | 0,015191632 | down | -1,7442003 | LCN1 | Homo sapiens lipocalin 1 (LCN1), transcript variant 2, mRNA [NM_001252617] |
| A_33_P3263955 | 0,01092718 | up | 1,7500861 | LCN10 | Homo sapiens lipocalin 10 (LCN10), mRNA [NM_001001712] |
| A_23_P32115 | 5,30E-04 | down | -2,6447988 | LCN12 | Homo sapiens lipocalin 12 (LCN12), mRNA [NM_178536] |
| A_33_P3263938 | 0,005069436 | down | -2,3223047 | LCN15 | Homo sapiens lipocalin 15 (LCN15), mRNA [NM_203347] |
| A_33_P3390557 | 0,005539991 | down | -2,3481658 | LCNL1 | Homo sapiens lipocalin-like 1 (LCNL1), mRNA [NM_207510] |

| | | | | | |
|---|---|---|---|---|---|
| A_23_P328836 | 0,003371461 | up | 1,5761234 | LCOR | Homo sapiens ligand dependent nuclear receptor corepressor (LCOR), transcript variant 1, mRNA [NM_032440] |
| A_23_P30547 | 0,008231903 | up | 1,5660498 | LCP2 | Homo sapiens lymphocyte cytosolic protein 2 (SH2 domain containing leukocyte protein of 76kDa) (LCP2), mRNA [NM_005565] |
| A_32_P98227 | 0,002758769 | down | -2,713767 | LDB3 | Homo sapiens LIM domain binding 3 (LDB3), transcript variant 5, mRNA [NM_001171610] |
| A_32_P44568 | 0,002949719 | up | 1,9103214 | LDHA | Homo sapiens lactate dehydrogenase A (LDHA), transcript variant 1, mRNA [NM_005566] |
| A_33_P3236441 | 8,47E-04 | down | -2,6096644 | LDHAL6A | Homo sapiens lactate dehydrogenase A-like 6A (LDHAL6A), transcript variant 1, mRNA [NM_144972] |
| A_23_P53039 | 7,35E-06 | down | -3,9985337 | LDHC | Homo sapiens lactate dehydrogenase C (LDHC), transcript variant 1, mRNA [NM_002301] |
| A_21_P0000886 | 0,008492159 | down | -2,2101512 | LDLRAD4-AS1 | Homo sapiens LDLRAD4 antisense RNA 1 (LDLRAD4-AS1), long non-coding RNA [NR_040031] |
| A_23_P25587 | 0,013101074 | down | -1,585457 | LECT1 | Homo sapiens leukocyte cell derived chemotaxin 1 (LECT1), transcript variant 1, mRNA [NM_007015] |
| A_23_P137573 | 0,031823214 | up | 1,5584595 | LEFTY2 | Homo sapiens left-right determination factor 2 (LEFTY2), transcript variant 1, mRNA [NM_003240] |
| A_24_P397817 | 0,032652948 | up | 2,0830197 | LEP | Homo sapiens leptin (LEP), mRNA [NM_000230] |
| A_24_P231104 | 0,01611238 | up | 1,543655 | LEPR | Homo sapiens leptin receptor (LEPR), transcript variant 3, mRNA [NM_001003679] |
| A_33_P3339820 | 0,001513058 | up | 1,524049 | LEPR | Homo sapiens leptin receptor (LEPR), transcript variant 2, mRNA [NM_001003680] |
| A_24_P148094 | 8,73E-04 | up | 1,8079225 | LEPROT | Homo sapiens leptin receptor overlapping transcript (LEPROT), transcript variant 1, mRNA [NM_017526] |
| A_23_P434518 | 5,19E-04 | down | -2,062575 | LFNG | Homo sapiens LFNG O-fucosylpeptide 3-beta-N-acetylglucosaminyltransferase (LFNG), transcript variant 2, mRNA [NM_001040168] |
| A_23_P120902 | 0,049549673 | up | 2,075476 | LGALS2 | Homo sapiens lectin, galactoside-binding, soluble, 2 (LGALS2), mRNA [NM_006498] |
| A_23_P15357 | 9,32E-04 | down | -1,6210023 | LGALS3BP | Homo sapiens lectin, galactoside-binding, soluble, 3 binding protein (LGALS3BP), mRNA [NM_005567] |
| A_23_P127267 | 0,009676578 | up | 1,5536923 | LGI1 | Homo sapiens leucine-rich, glioma inactivated 1 (LGI1), mRNA [NM_005097] |
| A_24_P310256 | 0,00538868 | up | 1,6410364 | LGI4 | Homo sapiens leucine-rich repeat LGI family, member 4 (LGI4), mRNA [NM_139284] |
| A_33_P3266769 | 0,03548057 | up | 1,6033709 | LHCGR | luteinizing hormone/choriogonadotropin receptor [Source:HGNC Symbol;Acc:HGNC:6585] [ENST00000477576] |
| A_23_P255104 | 0,010824876 | up | 1,6277791 | LHFPL2 | Homo sapiens lipoma HMGIC fusion partner-like 2 (LHFPL2), mRNA [NM_005779] |
| A_32_P158083 | 4,69E-06 | down | -10,581505 | LHFPL3 | Homo sapiens lipoma HMGIC fusion partner-like 3 (LHFPL3), mRNA [NM_199000] |
| A_32_P394491 | 0,021738604 | up | 2,4792063 | LHFPL3-AS1 | Homo sapiens LHFPL3 antisense RNA 1 (LHFPL3-AS1), transcript variant 2, long non-coding RNA [NR_034141] |
| A_33_P3692756 | 0,03698587 | down | -1,5285515 | LHFPL3-AS2 | Homo sapiens LHFPL3 antisense RNA 2 (LHFPL3-AS2), long non-coding RNA [NR_027374] |
| A_33_P3409854 | 6,32E-05 | down | -3,0190783 | LHX1 | Homo sapiens LIM homeobox 1 (LHX1), mRNA [NM_005568] |
| A_33_P3223678 | 0,007198805 | down | -3,3527548 | LHX3 | Homo sapiens LIM homeobox 3 (LHX3), transcript variant 2, mRNA [NM_014564] |
| A_23_P23869 | 0,023427997 | up | 1,5366815 | LHX4 | Homo sapiens LIM homeobox 4 (LHX4), mRNA [NM_033343] |
| A_23_P47967 | 0,00821214 | up | 1,9770395 | LHX5 | Homo sapiens LIM homeobox 5 (LHX5), mRNA [NM_022363] |
| A_24_P415845 | 0,024534483 | down | -1,9664092 | LIG4 | Homo sapiens ligase IV, DNA, ATP-dependent (LIG4), transcript variant 1, mRNA [NM_002312] |
| A_24_P348989 | 0,02411375 | down | -1,7900414 | LILRA1 | Homo sapiens leukocyte immunoglobulin-like receptor, subfamily A (with TM domain), member 1 (LILRA1), transcript variant 1, mRNA [NM_006863] |
| A_23_P90497 | 0,013204076 | down | -2,2014155 | LILRA4 | Homo sapiens leukocyte immunoglobulin-like receptor, subfamily A (with TM domain), member 4 (LILRA4), mRNA [NM_012276] |
| A_23_P215461 | 0,004292857 | up | 2,013782 | LIMK1 | Homo sapiens LIM domain kinase 1 (LIMK1), transcript variant 1, mRNA [NM_002314] |
| A_33_P3220615 | 0,02750677 | down | -1,6926328 | LIN28B | Homo sapiens lin-28 homolog B (C. elegans) (LIN28B), mRNA [NM_001004317] |
| A_23_P99253 | 0,013196187 | up | 1,721298 | LIN7A | Homo sapiens lin-7 homolog A (C. elegans) (LIN7A), mRNA [NM_004664] |
| A_23_P301995 | 0,009299332 | down | -1,7733628 | LIN9 | Homo sapiens lin-9 DREAM MuvB core complex component (LIN9), transcript variant 1, mRNA [NM_173083] |
| A_33_P3399228 | 0,009695206 | up | 1,9365015 | LINGO1 | Homo sapiens leucine rich repeat and Ig domain containing 1 (LINGO1), transcript variant 5, mRNA [NM_001301191] |
| A_23_P47019 | 0,00143648 | up | 1,6181887 | LIPF | Homo sapiens lipase, gastric (LIPF), transcript variant 2, mRNA [NM_004190] |
| A_33_P3401422 | 0,008592036 | down | -2,7452915 | LIPM | Homo sapiens lipase, family member M (LIPM), mRNA [NM_001128215] |
| A_33_P3323939 | 2,72E-05 | down | -9,259584 | LIPN | Homo sapiens lipase, family member N (LIPN), mRNA [NM_001102469] |
| A_23_P3532 | 0,003011246 | up | 1,8339392 | LITAF | Homo sapiens lipopolysaccharide-induced TNF factor (LITAF), transcript variant 1, mRNA [NM_004862] |
| A_32_P116606 | 0,003818828 | up | 1,5201497 | LIX1 | Homo sapiens Lix1 homolog (chicken) (LIX1), mRNA [NM_153234] |
| A_33_P3387756 | 2,78E-04 | down | -2,6778126 | LKAAEAR1 | Homo sapiens LKAAEAR motif containing 1 (LKAAEAR1), mRNA [NM_001007125] |
| A_33_P3337131 | 0,003904029 | down | -1,6552787 | LL22NC03-63E9.3 | Homo sapiens uncharacterized LOC648691 (LL22NC03-63E9.3), long non-coding RNA [NR_027426] |
| A_33_P3337124 | 0,001019795 | down | -4,4124637 | LL22NC03-63E9.3 | Homo sapiens uncharacterized LOC648691 (LL22NC03-63E9.3), long non-coding RNA [NR_027426] |
| A_23_P140630 | 0,01173853 | down | -1,6752245 | LMAN1L | Homo sapiens lectin, mannose-binding, 1 like (LMAN1L), mRNA [NM_021819] |
| A_23_P150931 | 0,012808262 | up | 1,5421113 | LMBR1L | Homo sapiens limb development membrane protein 1-like (LMBR1L), transcript variant 1, mRNA [NM_018113] |
| A_23_P6771 | 0,003662108 | down | -2,2806568 | LMCD1 | Homo sapiens LIM and cysteine-rich domains 1 (LMCD1), transcript variant 1, mRNA [NM_014583] |
| A_19_P00808088 | 0,005403922 | down | -2,3141427 | LMCD1-AS1 | Homo sapiens LMCD1 antisense RNA 1 (head to head) (LMCD1-AS1), long non-coding RNA [NR_033378] |
| A_33_P3883985 | 0,02948677 | up | 2,2318318 | LMF1 | Homo sapiens lipase maturation factor 1 (LMF1), transcript variant 4, non-coding RNA [NR_036442] |
| A_23_P170959 | 0,001367361 | down | -2,6275523 | LMLN | Homo sapiens leishmanolysin-like (metallopeptidase M8 family) (LMLN), transcript variant 2, mRNA [NM_033029] |
| A_23_P87310 | 2,76E-06 | down | -4,842292 | LMO1 | Homo sapiens LIM domain only 1 (rhombotin 1) (LMO1), transcript variant 1, mRNA [NM_002315] |
| A_33_P3280805 | 0,01143042 | up | 2,3695478 | LMO7 | Homo sapiens LIM domain 7 (LMO7), transcript variant 2, mRNA [NM_015842] |
| A_32_P795513 | 0,04081982 | up | 1,6023687 | LMOD3 | leiomodin 3 (fetal) [Source:HGNC Symbol;Acc:HGNC:6649] [ENST00000420581] |
| A_33_P3266928 | 0,046466764 | up | 1,5932807 | LMTK3 | Homo sapiens lemur tyrosine kinase 3 (LMTK3), mRNA [NM_001080434] |
| A_33_P3215739 | 0,001677505 | down | -2,3665833 | LNP1 | Homo sapiens leukemia NUP98 fusion partner 1 (LNP1), mRNA [NM_001085451] |
| A_23_P213137 | 0,033163324 | up | 2,526658 | LNX1 | Homo sapiens ligand of numb-protein X 1, E3 ubiquitin protein ligase (LNX1), transcript variant 2, mRNA [NM_032622] |
| A_32_P198810 | 9,32E-04 | down | -1,9636338 | LOH12CR2 | Homo sapiens loss of heterozygosity, 12, chromosomal region 2 (non-protein coding) (LOH12CR2), long non-coding RNA [NR_024061] |
| A_23_P94216 | 0,003931487 | up | 1,6968296 | LONRF1 | Homo sapiens LON peptidase N-terminal domain and ring finger 1 (LONRF1), mRNA [NM_152271] |
| A_33_P3315519 | 0,032252204 | down | -1,7960142 | LONRF2 | Homo sapiens LON peptidase N-terminal domain and ring finger 2 (LONRF2), mRNA [NM_198461] |
| A_23_P114414 | 0,00101968 | down | -2,0328484 | LONRF3 | Homo sapiens LON peptidase N-terminal domain and ring finger 3 (LONRF3), transcript variant 1, mRNA [NM_001031855] |
| A_23_P386888 | 7,77E-04 | down | -2,3508594 | LPA | Homo sapiens lipoprotein, Lp(a) (LPA), mRNA [NM_005577] |
| A_21_P0013164 | 0,021936392 | up | 2,7585485 | LPAL2 | Homo sapiens lipoprotein, Lp(a)-like 2, pseudogene (LPAL2), transcript variant 2, non-coding RNA [NR_028093] |

| | | | | | |
|---|---|---|---|---|---|
| A_24_P406006 | 4,13E-05 | up | 2,4378085 | LPCAT1 | Homo sapiens lysophosphatidylcholine acyltransferase 1 (LPCAT1), mRNA [NM_024830] |
| A_23_P49009 | 7,72E-04 | up | 2,0237591 | LPCAT4 | Homo sapiens lysophosphatidylcholine acyltransferase 4 (LPCAT4), mRNA [NM_153613] |
| A_33_P3356070 | 0,034210503 | up | 1,6612626 | LPIN1 | Homo sapiens lipin 1 (LPIN1), transcript variant 3, mRNA [NM_001261428] |
| A_32_P52609 | 0,002807883 | up | 1,731044 | LPIN1 | Homo sapiens lipin 1 (LPIN1), transcript variant 1, mRNA [NM_145693] |
| A_23_P146233 | 0,047277145 | up | 2,1292322 | LPL | Homo sapiens lipoprotein lipase (LPL), mRNA [NM_000237] |
| A_33_P3384958 | 0,015253551 | down | -1,8476499 | LPPR4 | Homo sapiens lipid phosphate phosphatase-related protein type 4 (LPPR4), transcript variant 1, mRNA [NM_014839] |
| A_24_P360078 | 0,00276564 | up | 1,633135 | LRBA | Homo sapiens LPS-responsive vesicle trafficking, beach and anchor containing (LRBA), transcript variant 2, mRNA [NM_006726] |
| A_24_P170774 | 0,03429293 | up | 1,5892844 | LRCH1 | Homo sapiens leucine-rich repeats and calponin homology (CH) domain containing 1 (LRCH1), transcript variant 2, mRNA [NM_015116] |
| A_21_P0011105 | 0,01440034 | up | 2,291546 | LRCOL1 | leucine rich colipase-like 1 [Source:HGNC Symbol;Acc:HGNC:44160] [ENST00000616042] |
| A_33_P3368925 | 0,04829474 | down | -1,5186015 | LRFN3 | leucine rich repeat and fibronectin type III domain containing 3 [Source:HGNC Symbol;Acc:HGNC:28370] [ENST00000588831] |
| A_33_P3280094 | 0,036396194 | up | 1,5753541 | LRFN4 | Homo sapiens leucine rich repeat and fibronectin type III domain containing 4 (LRFN4), mRNA [NM_024036] |
| A_23_P163195 | 0,007201625 | up | 1,5282245 | LRFN5 | Homo sapiens leucine rich repeat and fibronectin type III domain containing 5 (LRFN5), mRNA [NM_152447] |
| A_23_P16415 | 0,003698964 | down | -1,7087084 | LRP3 | Homo sapiens low density lipoprotein receptor-related protein 3 (LRP3), mRNA [NM_002333] |
| A_24_P403561 | 0,00458768 | up | 1,7837663 | LRP4 | Homo sapiens low density lipoprotein receptor-related protein 4 (LRP4), mRNA [NM_002334] |
| A_21_P0003885 | 0,047736116 | up | 1,5249289 | LRPAP1 | Homo sapiens low density lipoprotein receptor-related protein associated protein 1 (LRPAP1), transcript variant 1, mRNA [NM_002337] |
| A_33_P3286208 | 0,025328742 | down | -1,525203 | LRR1 | Homo sapiens leucine rich repeat protein 1 (LRR1), transcript variant 3, mRNA [NM_203467] |
| A_23_P19369 | 0,002109183 | up | 1,7184225 | LRRC16A | Homo sapiens leucine rich repeat containing 16A (LRRC16A), transcript variant 1, mRNA [NM_017640] |
| A_23_P165136 | 1,51E-04 | down | -4,3166943 | LRRC25 | Homo sapiens leucine rich repeat containing 25 (LRRC25), mRNA [NM_145256] |
| A_23_P49181 | 0,04385628 | down | -1,5634527 | LRRC29 | Homo sapiens leucine rich repeat containing 29 (LRRC29), transcript variant 1, mRNA [NM_012163] |
| A_33_P3325763 | 3,85E-04 | down | -2,3088548 | LRRC3 | leucine rich repeat containing 3 [Source:HGNC Symbol;Acc:HGNC:14965] [ENST00000291592] |
| A_21_P0010217 | 0,030037235 | down | -2,1358535 | LRRC3-AS1 | Homo sapiens LRRC3 antisense RNA 1 (head to head) (LRRC3-AS1), long non-coding RNA [NR_047476] |
| A_24_P281036 | 0,002265659 | up | 1,7054558 | LRRC31 | Homo sapiens leucine rich repeat containing 31 (LRRC31), transcript variant 1, mRNA [NM_024727] |
| A_23_P41194 | 0,020677513 | up | 1,7861514 | LRRC34 | Homo sapiens leucine rich repeat containing 34 (LRRC34), transcript variant 3, mRNA [NM_153353] |
| A_33_P3335183 | 0,01658908 | down | -1,6600022 | LRRC37A2 | Homo sapiens leucine rich repeat containing 37, member A2 (LRRC37A2), mRNA [NM_001006607] |
| A_32_P28402 | 0,005212484 | up | 1,5398017 | LRRC37BP1 | Homo sapiens leucine rich repeat containing 37B pseudogene 1 (LRRC37BP1), non-coding RNA [NR_015341] |
| A_33_P3228739 | 3,88E-04 | down | -2,6524434 | LRRC3C | Homo sapiens leucine rich repeat containing 3C (LRRC3C), mRNA [NM_001195545] |
| A_33_P3263217 | 9,58E-04 | up | 1,8331597 | LRRC4 | Homo sapiens leucine rich repeat containing 4 (LRRC4), mRNA [NM_022143] |
| A_23_P381461 | 8,78E-05 | down | -2,0442166 | LRRC45 | leucine rich repeat containing 45 [Source:HGNC Symbol;Acc:HGNC:28302] [ENST00000306688] |
| A_23_P129174 | 0,021765538 | down | -1,7467071 | LRRC49 | Homo sapiens leucine rich repeat containing 49 (LRRC49), transcript variant 2, mRNA [NM_017691] |
| A_33_P3344399 | 0,038031247 | down | -1,5649277 | LRRC49 | Homo sapiens leucine rich repeat containing 49 (LRRC49), transcript variant 3, mRNA [NM_001199018] |
| A_33_P3228862 | 0,034672726 | down | -1,5541698 | LRRC69 | Homo sapiens leucine rich repeat containing 69 (LRRC69), mRNA [NM_001129890] |
| A_32_P222149 | 2,45E-04 | down | -2,3933353 | LRRC72 | Homo sapiens leucine rich repeat containing 72 (LRRC72), mRNA [NM_001195280] |
| A_33_P3258339 | 0,00401824 | down | -1,5922018 | LRRC75A | Homo sapiens leucine rich repeat containing 75A (LRRC75A), transcript variant 1, mRNA [NM_001113567] |
| A_23_P368484 | 6,69E-05 | up | 3,0368626 | LRRC75A | Homo sapiens leucine rich repeat containing 75A (LRRC75A), transcript variant 2, mRNA [NM_207387] |
| A_21_P0000529 | 0,008317401 | up | 2,49926 | LRRC75A-AS1 | Homo sapiens LRRC75A antisense RNA 1 (LRRC75A-AS1), transcript variant 20, long non-coding RNA [NR_027176] |
| A_33_P3420446 | 1,53E-04 | down | -7,0295076 | LRRD1 | Homo sapiens leucine-rich repeats and death domain containing 1 (LRRD1), mRNA [NM_001161528] |
| A_33_P3358648 | 0,003372378 | up | 1,5558778 | LRRFIP1 | Homo sapiens leucine rich repeat (in FLII) interacting protein 1 (LRRFIP1), transcript variant 3, mRNA [NM_001137552] |
| A_33_P3292387 | 2,14E-05 | down | -6,217423 | LRRIQ3 | Homo sapiens leucine-rich repeats and IQ motif containing 3 (LRRIQ3), mRNA [NM_001105659] |
| A_33_P3381478 | 0,009728421 | up | 1,5799105 | LRRIQ3 | Homo sapiens leucine-rich repeats and IQ motif containing 3 (LRRIQ3), mRNA [NM_001105659] |
| A_23_P351913 | 0,01982202 | down | -1,8701682 | LRRN4 | Homo sapiens leucine rich repeat neuronal 4 (LRRN4), mRNA [NM_152611] |
| A_23_P131330 | 0,040538747 | down | -1,8405358 | LRRTM1 | Homo sapiens leucine rich repeat transmembrane neuronal 1 (LRRTM1), mRNA [NM_178839] |
| A_33_P3251024 | 6,27E-05 | down | -2,4655702 | LRRTM2 | Homo sapiens leucine rich repeat transmembrane neuronal 2 (LRRTM2), mRNA [NM_015564] |
| A_24_P140569 | 0,030658914 | up | 1,8946028 | LRRTM2 | Homo sapiens leucine rich repeat transmembrane neuronal 2 (LRRTM2), mRNA [NM_015564] |
| A_24_P666105 | 0,013896322 | up | 2,4680288 | LRRTM4 | Homo sapiens leucine rich repeat transmembrane neuronal 4 (LRRTM4), transcript variant 1, mRNA [NM_001134745] |
| A_33_P3216651 | 0,037950173 | down | -2,4116886 | LRTOMT | Homo sapiens leucine rich transmembrane and O-methyltransferase domain containing (LRTOMT), transcript variant 6, mRNA [NM_001145310] |
| A_23_P368195 | 0,036415886 | up | 1,5082973 | LSM11 | Homo sapiens LSM11, U7 small nuclear RNA associated (LSM11), mRNA [NM_173491] |
| A_21_P0013035 | 0,007954476 | down | -1,8766198 | LSM14A | Homo sapiens LSM14A, SCD6 homolog A (S. cerevisiae) (LSM14A), transcript variant 2, mRNA [NM_015578] |
| A_23_P323783 | 7,80E-04 | down | -1,5597228 | LSM14B | Homo sapiens LSM14B, SCD6 homolog B (S. cerevisiae) (LSM14B), mRNA [NM_144703] |
| A_33_P3349776 | 3,50E-04 | down | -1,7401884 | LSM14B | Homo sapiens LSM14B, SCD6 homolog B (S. cerevisiae) (LSM14B), mRNA [NM_144703] |
| A_32_P466877 | 0,036480986 | up | 2,4479024 | LSMEM2 | leucine-rich single-pass membrane protein 2 [Source:HGNC Symbol;Acc:HGNC:26781] [ENST00000316436] |
| A_23_P142389 | 0,001212374 | up | 1,6296514 | LSR | Homo sapiens lipolysis stimulated lipoprotein receptor (LSR), transcript variant 2, mRNA [NM_205834] |
| A_23_P156683 | 0,023978427 | down | -2,0891063 | LTA | Homo sapiens lymphotoxin alpha (LTA), transcript variant 2, mRNA [NM_000595] |
| A_23_P163173 | 0,002780756 | down | -1,9424365 | LTB4R2 | Homo sapiens leukotriene B4 receptor 2 (LTB4R2), transcript variant 1, mRNA [NM_019839] |
| A_33_P3278826 | 0,004520775 | down | -2,2895336 | LTK | leukocyte receptor tyrosine kinase [Source:HGNC Symbol;Acc:HGNC:6721] [ENST00000563518] |
| A_23_P122624 | 0,011912887 | up | 1,5103949 | LTV1 | Homo sapiens LTV1 ribosome biogenesis factor (LTV1), mRNA [NM_032860] |
| A_33_P3291569 | 0,016530352 | up | 1,5058268 | LUC7L3 | Homo sapiens LUC7-like 3 (S. cerevisiae) (LUC7L3), transcript variant 1, mRNA [NM_016424] |
| A_33_P3246997 | 7,02E-04 | up | 1,7699775 | LUC7L3 | LUC7-like 3 (S. cerevisiae) [Source:HGNC Symbol;Acc:HGNC:24309] [ENST00000506686] |
| A_21_P0004568 | 0,03822696 | down | -1,5382239 | LUCAT1 | lung cancer associated transcript 1 (non-protein coding) [Source:HGNC Symbol;Acc:HGNC:48498] [ENST00000513626] |
| A_24_P649282 | 0,009919231 | up | 1,5013951 | LUZP2 | Homo sapiens leucine zipper protein 2 (LUZP2), transcript variant 1, mRNA [NM_001009909] |
| A_33_P3230090 | 6,64E-04 | down | -1,8521699 | LUZP6 | Homo sapiens leucine zipper protein 6 (LUZP6), mRNA [NM_001128619] |
| A_24_P7584 | 0,007470159 | down | -2,3366644 | LY6G5C | Homo sapiens lymphocyte antigen 6 complex, locus G5C (LY6G5C), mRNA [NM_025262] |

| | | | | | |
|---|---|---|---|---|---|
| A_33_P3275722 | 2,97E-04 | down | -2,375565 | LY6G6D | Homo sapiens lymphocyte antigen 6 complex, locus G6D (LY6G6D), mRNA [NM_021246] |
| A_33_P3410232 | 0,005701698 | down | -2,9748836 | LY6G6E | Homo sapiens lymphocyte antigen 6 complex, locus G6E (pseudogene) (LY6G6E), transcript variant 1, non-coding RNA [NR_024541] |
| A_33_P3277097 | 0,002263848 | down | -2,4196107 | LY6G6E | Homo sapiens lymphocyte antigen 6 complex, locus G6E (pseudogene) (LY6G6E), transcript variant 2, non-coding RNA [NR_003673] |
| A_23_P70688 | 0,009006206 | down | -1,6368446 | LY86 | Homo sapiens lymphocyte antigen 86 (LY86), mRNA [NM_004271] |
| A_24_P324674 | 1,00E-03 | down | -3,2344613 | LY9 | Homo sapiens lymphocyte antigen 9 (LY9), transcript variant 1, mRNA [NM_002348] |
| A_33_P3215422 | 0,0011589 | down | -2,4098272 | LYL1 | Homo sapiens lymphoblastic leukemia associated hematopoiesis regulator 1 (LYL1), mRNA [NM_005583] |
| A_23_P311780 | 0,020435786 | down | -2,1101465 | LYPD4 | Homo sapiens LY6/PLAUR domain containing 4 (LYPD4), transcript variant 1, mRNA [NM_173506] |
| A_21_P0009657 | 0,013676779 | up | 1,6043092 | LYPD5 | LY6/PLAUR domain containing 5 [Source:HGNC Symbol;Acc:HGNC:26397] [ENST00000599397] |
| A_33_P3258510 | 0,008422169 | up | 1,6364477 | LYPLA2 | Homo sapiens lysophospholipase II (LYPLA2), mRNA [NM_007260] |
| A_24_P276490 | 0,044998635 | up | 1,5612335 | LYPLA2 | Homo sapiens lysophospholipase II (LYPLA2), mRNA [NM_007260] |
| A_33_P3312611 | 0,018514456 | up | 2,0208094 | LYPLAL1-AS1 | Homo sapiens LYPLAL1 antisense RNA 1 (head to head) (LYPLAL1-AS1), long non-coding RNA [NR_038845] |
| A_33_P3288484 | 0,008144885 | down | -1,6318748 | LYRM2 | Homo sapiens LYR motif containing 2 (LYRM2), transcript variant 1, mRNA [NM_020466] |
| A_24_P26554 | 0,013847774 | up | 1,5175674 | LYRM4 | Homo sapiens LYR motif containing 4 (LYRM4), transcript variant 1, mRNA [NM_020408] |
| A_24_P648176 | 0,015015048 | down | -2,484134 | LYRM5 | Homo sapiens LYR motif containing 5 (LYRM5), mRNA [NM_001001660] |
| A_23_P354074 | 0,006300472 | up | 1,8371681 | LYST | Homo sapiens lysosomal trafficking regulator (LYST), transcript variant 1, mRNA [NM_000081] |
| A_23_P130056 | 0,019320456 | up | 1,9696681 | LYZL6 | Homo sapiens lysozyme-like 6 (LYZL6), transcript variant 2, mRNA [NM_020426] |
| A_33_P3212302 | 0,036582347 | up | 1,5158855 | LZTS1 | Homo sapiens leucine zipper, putative tumor suppressor 1 (LZTS1), mRNA [NM_021020] |
| A_33_P3254666 | 0,016913267 | down | -1,587856 | LZTS2 | Homo sapiens cDNA FLJ40678 fis, clone THYMU2022660. [AK097997] |
| A_24_P160696 | 0,029818017 | up | 1,5104735 | MAATS1 | Homo sapiens MYCBP-associated, testis expressed 1 (MAATS1), mRNA [NM_033364] |
| A_23_P343366 | 0,008271859 | down | -1,6528074 | MAB21L3 | Homo sapiens mab-21-like 3 (C. elegans) (MAB21L3), mRNA [NM_152367] |
| A_23_P391778 | 0,001112525 | up | 1,9133213 | MACF1 | Homo sapiens microtubule-actin crosslinking factor 1 (MACF1), mRNA [NM_012090] |
| A_24_P418637 | 0,029811347 | up | 1,6760046 | MACF1 | Homo sapiens microtubule-actin crosslinking factor 1 (MACF1), mRNA [NM_012090] |
| A_33_P3359253 | 0,037550688 | down | -1,7886592 | MAD2L2 | MAD2 mitotic arrest deficient-like 2 (yeast) [Source:HGNC Symbol;Acc:HGNC:6764] [ENST00000376655] |
| A_33_P3359250 | 5,01E-04 | down | -2,6350017 | MAD2L2 | MAD2 mitotic arrest deficient-like 2 (yeast) [Source:HGNC Symbol;Acc:HGNC:6764] [ENST00000376664] |
| A_33_P3416767 | 0,001696284 | up | 1,7473966 | MAFG | Homo sapiens v-maf avian musculoaponeurotic fibrosarcoma oncogene homolog G (MAFG), transcript variant 2, mRNA [NM_032711] |
| A_33_P3241325 | 0,003155357 | down | -1,5587494 | MAFG-AS1 | Homo sapiens MAFG antisense RNA 1 (head to head) (MAFG-AS1), long non-coding RNA [NR_015454] |
| A_33_P3406828 | 1,80E-04 | down | -2,787117 | MAFIP | Homo sapiens MAFF interacting protein (pseudogene) (MAFIP), transcript variant 1, non-coding RNA [NR_046439] |
| A_33_P3272823 | 0,003409972 | down | -2,8012407 | MAG | Homo sapiens myelin associated glycoprotein (MAG), transcript variant 1, mRNA [NM_002361] |
| A_23_P96291 | 0,007732341 | up | 1,5023415 | MAGEA1 | Homo sapiens melanoma antigen family A, 1 (MAGEA1), mRNA [NM_004988] |
| A_24_P185945 | 0,01996031 | up | 1,5547098 | MAGEA4 | Homo sapiens melanoma antigen family A, 4 (MAGEA4), transcript variant 2, mRNA [NM_002362] |
| A_23_P136870 | 5,82E-05 | down | -3,1001205 | MAGEA6 | Homo sapiens melanoma antigen family A, 6 (MAGEA6), transcript variant 2, mRNA [NM_175868] |
| A_23_P254831 | 9,46E-04 | down | -1,8332686 | MAGEB2 | Homo sapiens melanoma antigen family B, 2 (MAGEB2), mRNA [NM_002364] |
| A_33_P3405509 | 0,006540382 | up | 1,5670403 | MAGEB5 | Homo sapiens melanoma antigen family B, 5 (MAGEB5), mRNA [NM_001271752] |
| A_33_P3213204 | 0,02218047 | up | 1,6637679 | MAGEF1 | Homo sapiens melanoma antigen family F, 1 (MAGEF1), mRNA [NM_022149] |
| A_23_P99996 | 6,10E-04 | down | -3,0282664 | MAGEL2 | Homo sapiens MAGE-like 2 (MAGEL2), mRNA [NM_019066] |
| A_33_P3369535 | 0,027161095 | down | -2,0121102 | MAGI1 | Homo sapiens membrane associated guanylate kinase, WW and PDZ domain containing 1 (MAGI1), transcript variant 2, mRNA [NM_004742] |
| A_33_P3369530 | 0,006354068 | up | 1,8876338 | MAGI1 | Homo sapiens membrane associated guanylate kinase, WW and PDZ domain containing 1 (MAGI1), transcript variant 1, mRNA [NM_015520] |
| A_24_P75220 | 0,011313551 | up | 1,5294299 | MAGI1 | Homo sapiens membrane associated guanylate kinase, WW and PDZ domain containing 1 (MAGI1), transcript variant 2, mRNA [NM_004742] |
| A_23_P200216 | 0,012985399 | up | 1,5253428 | MAGOH | Homo sapiens mago-nashi homolog, proliferation-associated (Drosophila) (MAGOH), mRNA [NM_002370] |
| A_23_P148273 | 0,00637643 | down | -1,8399441 | MAGT1 | Homo sapiens magnesium transporter 1 (MAGT1), mRNA [NM_032121] |
| A_33_P3244279 | 6,30E-04 | up | 2,759087 | MAGT1 | Homo sapiens clone FLB3024 PRO0756 mRNA, complete cds. [AF130104] |
| A_21_P0010808 | 0,042503312 | up | 1,6721302 | MALRD1 | Homo sapiens MAM and LDL receptor class A domain containing 1 (MALRD1), mRNA [NM_001142308] |
| A_21_P0010807 | 0,008914619 | up | 1,5071123 | MALRD1 | Homo sapiens MAM and LDL receptor class A domain containing 1 (MALRD1), mRNA [NM_001142308] |
| A_24_P116909 | 0,001067091 | up | 1,842436 | MALT1 | Homo sapiens MALT1 paracaspase (MALT1), transcript variant 1, mRNA [NM_006785] |
| A_33_P3220149 | 0,014194852 | down | -1,6238451 | MAML1 | mastermind-like 1 (Drosophila) [Source:HGNC Symbol;Acc:HGNC:13632] [ENST00000503050] |
| A_23_P110606 | 0,002284292 | up | 1,6225219 | MAML1 | Homo sapiens mastermind-like 1 (Drosophila) (MAML1), mRNA [NM_014757] |
| A_33_P3335371 | 0,03594006 | down | -1,5356039 | MAML3 | Homo sapiens mastermind-like 3 (Drosophila) (MAML3), mRNA [NM_018717] |
| A_33_P3357573 | 0,031085104 | down | -1,5982143 | MAML3 | Homo sapiens mastermind-like 3 (Drosophila) (MAML3), mRNA [NM_018717] |
| A_24_P297537 | 0,003216366 | down | -1,7668031 | MAMSTR | Homo sapiens MEF2 activating motif and SAP domain containing transcriptional regulator (MAMSTR), transcript variant 2, mRNA [NM_182574] |
| A_24_P187921 | 0,011465751 | down | -2,072286 | MANEA | Homo sapiens mannosidase, endo-alpha (MANEA), mRNA [NM_024641] |
| A_23_P132793 | 0,044265483 | down | -1,6440865 | MANF | Homo sapiens mesencephalic astrocyte-derived neurotrophic factor (MANF), mRNA [NM_006010] |
| A_33_P3242543 | 0,001709668 | up | 1,7940465 | MAOA | Homo sapiens monoamine oxidase A (MAOA), transcript variant 2, mRNA [NM_001270458] |
| A_23_P163455 | 0,015003612 | down | -2,039289 | MAP1A | Homo sapiens microtubule-associated protein 1A (MAP1A), mRNA [NM_002373] |
| A_23_P20248 | 8,31E-04 | up | 1,8232006 | MAP2K1 | Homo sapiens mitogen-activated protein kinase kinase 1 (MAP2K1), mRNA [NM_002755] |
| A_23_P118427 | 0,005329644 | up | 1,8167671 | MAP2K3 | Homo sapiens mitogen-activated protein kinase kinase 3 (MAP2K3), transcript variant B, mRNA [NM_145109] |
| A_24_P296698 | 0,028025737 | up | 1,5437715 | MAP2K3 | Homo sapiens mitogen-activated protein kinase kinase 3 (MAP2K3), transcript variant B, mRNA [NM_145109] |
| A_23_P58031 | 0,017667292 | up | 1,6788749 | MAP3K13 | Homo sapiens mitogen-activated protein kinase kinase kinase 13 (MAP3K13), transcript variant 1, mRNA [NM_004721] |
| A_19_P00324286 | 0,007768023 | up | 1,5396519 | MAP3K13 | Homo sapiens mitogen-activated protein kinase kinase kinase 13 (MAP3K13), transcript variant 4, non-coding RNA [NR_038322] |
| A_24_P63078 | 0,005491082 | up | 1,8178045 | MAP3K19 | Homo sapiens mitogen-activated protein kinase kinase kinase 19 (MAP3K19), transcript variant 1, mRNA [NM_025052] |
| A_23_P134125 | 0,024195522 | up | 1,5069194 | MAP3K5 | Homo sapiens mitogen-activated protein kinase kinase kinase 5 (MAP3K5), mRNA [NM_005923] |
| A_23_P500773 | 0,00186588 | up | 1,5652468 | MAP3K7 | Homo sapiens mitogen-activated protein kinase kinase kinase 7 (MAP3K7), transcript variant A, mRNA [NM_003188] |

| | | | | | |
|---|---|---|---|---|---|
| A_33_P3238310 | 0,001371857 | down | -1,7792462 | MAP3K7CL | Homo sapiens MAP3K7 C-terminal like (MAP3K7CL), transcript variant 8, mRNA [NM_001286623] |
| A_23_P57277 | 0,005593776 | up | 2,377055 | MAP3K7CL | Homo sapiens MAP3K7 C-terminal like (MAP3K7CL), transcript variant 1, mRNA [NM_020152] |
| A_33_P3246505 | 0,017297868 | up | 2,5002482 | MAP3K8 | mitogen-activated protein kinase kinase kinase 8 [Source:HGNC Symbol;Acc:HGNC:6860] [ENST00000375322] |
| A_23_P23947 | 0,009185061 | up | 2,4190743 | MAP3K8 | Homo sapiens mitogen-activated protein kinase kinase kinase 8 (MAP3K8), transcript variant 1, mRNA [NM_005204] |
| A_33_P3323435 | 4,99E-04 | up | 2,2391062 | MAP3K9 | Homo sapiens mitogen-activated protein kinase kinase kinase 9 (MAP3K9), transcript variant 1, mRNA [NM_033141] |
| A_33_P3379571 | 0,033511776 | up | 1,607734 | MAP3K9 | Homo sapiens mitogen-activated protein kinase kinase kinase 9 (MAP3K9), transcript variant 1, mRNA [NM_033141] |
| A_33_P3346573 | 6,07E-05 | down | -2,064614 | MAP4 | Homo sapiens microtubule-associated protein 4 (MAP4), transcript variant 4, mRNA [NM_001134364] |
| A_33_P3354137 | 5,89E-04 | down | -2,6352262 | MAP4 | Homo sapiens microtubule-associated protein 4 (MAP4), transcript variant 3, mRNA [NM_030885] |
| A_23_P5002 | 0,001021598 | down | -2,840189 | MAP4K1 | Homo sapiens mitogen-activated protein kinase kinase kinase kinase 1 (MAP4K1), transcript variant 1, mRNA [NM_001042600] |
| A_23_P90804 | 0,004319808 | up | 1,8926725 | MAP4K4 | Homo sapiens mitogen-activated protein kinase kinase kinase kinase 4 (MAP4K4), transcript variant 2, mRNA [NM_145686] |
| A_33_P3279841 | 0,035786662 | up | 1,5834215 | MAPK8IP3 | Homo sapiens mitogen-activated protein kinase 8 interacting protein 3 (MAPK8IP3), transcript variant 1, mRNA [NM_015133] |
| A_23_P216894 | 0,037850536 | up | 1,5968746 | MAPKAP1 | Homo sapiens mitogen-activated protein kinase associated protein 1 (MAPKAP1), transcript variant 1, mRNA [NM_001006617] |
| A_24_P124662 | 0,003858582 | up | 1,5630234 | MAPKAPK5 | Homo sapiens mitogen-activated protein kinase-activated protein kinase 5 (MAPKAPK5), transcript variant 2, mRNA [NM_139078] |
| A_23_P99204 | 0,005419465 | up | 1,6991384 | MAPKAPK5-AS1 | Homo sapiens MAPKAPK5 antisense RNA 1 (MAPKAPK5-AS1), long non-coding RNA [NR_015404] |
| A_21_P0007761 | 0,007063204 | up | 1,5074999 | MAPKAPK5-AS1 | Homo sapiens MAPKAPK5 antisense RNA 1 (MAPKAPK5-AS1), long non-coding RNA [NR_015404] |
| A_21_P0014032 | 0,002965167 | down | -1,8313522 | MAPT-IT1 | Homo sapiens MAPT intronic transcript 1 (non-protein coding) (MAPT-IT1), long non-coding RNA [NR_024560] |
| A_23_P110492 | 0,012284086 | up | 1,5136778 | MARCH6 | Homo sapiens membrane-associated ring finger (C3HC4) 6, E3 ubiquitin protein ligase (MARCH6), transcript variant 1, mRNA [NM_005885] |
| A_23_P310 | 0,013619931 | up | 1,9864509 | MARCKSL1 | Homo sapiens MARCKS-like 1 (MARCKSL1), transcript variant 1, mRNA [NM_023009] |
| A_24_P914495 | 0,011688267 | down | -1,5616211 | MARK2 | Homo sapiens MAP/microtubule affinity-regulating kinase 2 (MARK2), transcript variant 3, mRNA [NM_004954] |
| A_23_P101480 | 0,009107015 | down | -1,7050976 | MAST1 | Homo sapiens microtubule associated serine/threonine kinase 1 (MAST1), mRNA [NM_014975] |
| A_24_P84340 | 0,0280801 | down | -2,154636 | MAST4 | Homo sapiens microtubule associated serine/threonine kinase family member 4 (MAST4), transcript variant 2, mRNA [NM_198828] |
| A_33_P3403044 | 0,002129745 | up | 1,6060967 | MAT2A | Homo sapiens methionine adenosyltransferase II, alpha (MAT2A), mRNA [NM_005911] |
| A_23_P401568 | 3,29E-04 | up | 1,9942687 | MAT2A | Homo sapiens methionine adenosyltransferase II, alpha (MAT2A), mRNA [NM_005911] |
| A_23_P50678 | 0,032507036 | up | 1,7295928 | MATK | Homo sapiens megakaryocyte-associated tyrosine kinase (MATK), transcript variant 1, mRNA [NM_139355] |
| A_32_P902988 | 0,012739374 | down | -1,823178 | MATN1 | Homo sapiens matrilin 1, cartilage matrix protein (MATN1), mRNA [NM_002379] |
| A_23_P71328 | 0,00441701 | down | -2,5255136 | MATN2 | Homo sapiens matrilin 2 (MATN2), transcript variant 2, mRNA [NM_030583] |
| A_19_P00319465 | 0,00855925 | up | 1,520238 | MAVS | Homo sapiens mitochondrial antiviral signaling protein (MAVS), transcript variant 1, mRNA [NM_020746] |
| A_23_P415395 | 0,02552089 | down | -1,8821733 | MBD3L1 | Homo sapiens methyl-CpG binding domain protein 3-like 1 (MBD3L1), mRNA [NM_145208] |
| A_23_P378450 | 6,15E-04 | up | 1,8693514 | MBD3L2 | Homo sapiens methyl-CpG binding domain protein 3-like 2 (MBD3L2), mRNA [NM_144614] |
| A_33_P3269388 | 0,004076998 | up | 1,6246194 | MBOAT7 | Homo sapiens membrane bound O-acyltransferase domain containing 7 (MBOAT7), transcript variant 4, mRNA [NM_001146082] |
| A_23_P208085 | 0,033940904 | up | 2,306321 | MBP | Homo sapiens myelin basic protein (MBP), transcript variant 7, mRNA [NM_001025101] |
| A_33_P3227990 | 0,00146357 | up | 1,8949925 | MBP | Homo sapiens myelin basic protein (MBP), transcript variant 7, mRNA [NM_001025101] |
| A_24_P402080 | 0,004132152 | up | 1,5414695 | MBP | Homo sapiens myelin basic protein (MBP), transcript variant 8, mRNA [NM_001025100] |
| A_32_P58937 | 0,001633452 | up | 1,5885934 | MBTD1 | Homo sapiens mbt domain containing 1 (MBTD1), mRNA [NM_017643] |
| A_23_P14948 | 0,003072893 | up | 1,5503612 | MBTPS1 | Homo sapiens membrane-bound transcription factor peptidase, site 1 (MBTPS1), mRNA [NM_003791] |
| A_23_P114095 | 0,002888899 | down | -1,5812219 | MBTPS2 | Homo sapiens membrane-bound transcription factor peptidase, site 2 (MBTPS2), mRNA [NM_015884] |
| A_33_P3318642 | 0,012690086 | down | -2,237437 | MC1R | melanocortin 1 receptor (alpha melanocyte stimulating hormone receptor) [Source:HGNC Symbol;Acc:HGNC:6929] [ENST00000539976] |
| A_23_P38649 | 0,005708573 | down | -2,6989307 | MC2R | Homo sapiens melanocortin 2 receptor (adrenocorticotropic hormone) (MC2R), mRNA [NM_000529] |
| A_23_P50039 | 0,001365967 | down | -2,9424393 | MC5R | Homo sapiens melanocortin 5 receptor (MC5R), mRNA [NM_005913] |
| A_23_P330561 | 5,33E-05 | down | -3,7693567 | MCEMP1 | Homo sapiens mast cell-expressed membrane protein 1 (MCEMP1), mRNA [NM_174918] |
| A_23_P88099 | 0,001494988 | down | -2,0908046 | MCF2L | Homo sapiens MCF.2 cell line derived transforming sequence-like (MCF2L), transcript variant 2, mRNA [NM_024979] |
| A_19_P00322757 | 1,56E-05 | down | -7,4011693 | MCF2L | MCF.2 cell line derived transforming sequence-like [Source:HGNC Symbol;Acc:HGNC:14576] [ENST00000442625] |
| A_24_P103060 | 0,004736973 | up | 1,7019986 | MCFD2 | Homo sapiens multiple coagulation factor deficiency 2 (MCFD2), transcript variant 1, mRNA [NM_139279] |
| A_33_P3393311 | 0,007953781 | down | -2,2043447 | MCHR1 | Homo sapiens melanin-concentrating hormone receptor 1 (MCHR1), mRNA [NM_005297] |
| A_24_P336759 | 0,01271178 | up | 1,6743331 | MCL1 | Homo sapiens myeloid cell leukemia 1 (MCL1), transcript variant 1, mRNA [NM_021960] |
| A_23_P397347 | 0,011300852 | up | 1,583166 | MCM9 | Homo sapiens minichromosome maintenance complex component 9 (MCM9), transcript variant 2, mRNA [NM_153255] |
| A_24_P244944 | 0,024573645 | down | -1,6570293 | MCTP2 | Homo sapiens multiple C2 domains, transmembrane 2 (MCTP2), transcript variant 1, mRNA [NM_018349] |
| A_33_P3296193 | 0,025928678 | down | -1,7219698 | MCTS2P | Homo sapiens malignant T cell amplified sequence 2, pseudogene (MCTS2P), non-coding RNA [NR_003677] |
| A_23_P346405 | 0,002731906 | up | 1,9808964 | MCU | Homo sapiens mitochondrial calcium uniporter (MCU), transcript variant 1, mRNA [NM_138357] |
| A_33_P3421867 | 5,77E-05 | down | -3,20775 | MDGA1 | MAM domain containing glycosylphosphatidylinositol anchor 1 [Source:HGNC Symbol;Acc:HGNC:19267] [ENST00000373401] |
| A_23_P105730 | 0,017192103 | down | -1,7158177 | MDM1 | Homo sapiens Mdm1 nuclear protein homolog (mouse) (MDM1), transcript variant 2, mRNA [NM_020128] |
| A_33_P3361707 | 0,007522723 | down | -1,9325333 | MDM2 | Homo sapiens MDM2 proto-oncogene, E3 ubiquitin protein ligase (MDM2), transcript variant 1, mRNA [NM_002392] |
| A_23_P502750 | 0,001828127 | up | 1,691623 | MDM2 | Homo sapiens MDM2 proto-oncogene, E3 ubiquitin protein ligase (MDM2), transcript variant 1, mRNA [NM_002392] |
| A_19_P00320719 | 0,001351993 | up | 1,769058 | MDM4 | Homo sapiens MDM4, p53 regulator (MDM4), transcript variant 1, mRNA [NM_002393] |
| A_23_P365060 | 0,003667409 | up | 1,9061389 | MDN1 | Homo sapiens MDN1, midasin homolog (yeast) (MDN1), mRNA [NM_014611] |
| A_23_P323196 | 0,002701155 | up | 1,5520177 | MDS2 | Homo sapiens myelodysplastic syndrome 2 translocation associated (MDS2), long non-coding RNA [NR_027042] |
| A_32_P32905 | 0,006834899 | up | 1,8450972 | MEAF6 | Homo sapiens MYST/Esa1-associated factor 6 (MEAF6), transcript variant 1, mRNA [NM_022756] |
| A_24_P320103 | 0,001471606 | down | -1,6215931 | MECOM | Homo sapiens MDS1 and EVI1 complex locus (MECOM), transcript variant 4, mRNA [NM_004991] |
| A_33_P3888629 | 0,009934093 | up | 1,7395353 | MECOM | Homo sapiens MDS1 and EVI1 complex locus (MECOM), transcript variant 6, mRNA [NM_001164000] |
| A_33_P3317211 | 0,001435448 | down | -1,6290061 | MECP2 | Homo sapiens methyl CpG binding protein 2 (MECP2), transcript variant 2, mRNA [NM_001110792] |
| A_24_P237486 | 0,005128513 | up | 1,5629245 | MECP2 | Homo sapiens methyl CpG binding protein 2 (MECP2), transcript variant 1, mRNA [NM_004992] |

| | | | | | |
|---|---|---|---|---|---|
| A_23_P73702 | 0,009597085 | up | 1,5282973 | MED12 | Homo sapiens mediator complex subunit 12 (MED12), mRNA [NM_005120] |
| A_32_P701268 | 0,005207061 | up | 1,6475548 | MED12L | Homo sapiens mediator complex subunit 12-like (MED12L), mRNA [NM_053002] |
| A_33_P3223759 | 0,005624732 | up | 1,5288526 | MED13 | Homo sapiens mediator complex subunit 13 (MED13), mRNA [NM_005121] |
| A_23_P21120 | 0,018708179 | up | 1,5747808 | MED14 | mediator complex subunit 14 [Source:HGNC Symbol;Acc:HGNC:2370] [ENST00000324817] |
| A_24_P207727 | 0,014027731 | up | 1,7538003 | MED15 | Homo sapiens mediator complex subunit 15 (MED15), transcript variant 1, mRNA [NM_001003891] |
| A_23_P138105 | 1,61E-05 | down | -2,8731956 | MED18 | Homo sapiens mediator complex subunit 18 (MED18), transcript variant 1, mRNA [NM_017638] |
| A_23_P164737 | 0,005338272 | up | 1,5029979 | MED26 | Homo sapiens mediator complex subunit 26 (MED26), mRNA [NM_004831] |
| A_33_P3354771 | 1,82E-04 | down | -2,06769 | MEF2BNB | MEF2B neighbor [Source:HGNC Symbol;Acc:HGNC:37247] [ENST00000591566] |
| A_24_P914940 | 0,009790556 | down | -1,536852 | MEF2BNB | Homo sapiens MEF2B neighbor (MEF2BNB), transcript variant 1, mRNA [NM_001145784] |
| A_33_P3341656 | 0,030461814 | up | 1,5093846 | MEF2D | Homo sapiens myocyte enhancer factor 2D (MEF2D), transcript variant 1, mRNA [NM_005920] |
| A_23_P140967 | 0,012199974 | down | -3,3862178 | MEFV | Homo sapiens Mediterranean fever (MEFV), transcript variant 1, mRNA [NM_000243] |
| A_32_P129894 | 0,009926206 | up | 1,6731288 | MEGF9 | Homo sapiens multiple EGF-like-domains 9 (MEGF9), mRNA [NM_001080497] |
| A_23_P94422 | 0,001898999 | up | 1,6393847 | MELK | Homo sapiens maternal embryonic leucine zipper kinase (MELK), transcript variant 1, mRNA [NM_014791] |
| A_23_P155931 | 0,037688866 | up | 2,6468644 | MEPE | Homo sapiens matrix extracellular phosphoglycoprotein (MEPE), transcript variant 2, mRNA [NM_020203] |
| A_23_P99891 | 0,001541975 | up | 1,6707851 | MESDC1 | Homo sapiens mesoderm development candidate 1 (MESDC1), mRNA [NM_022566] |
| A_24_P141736 | 0,019215584 | up | 1,9375159 | METAP2 | Homo sapiens methionyl aminopeptidase 2 (METAP2), mRNA [NM_006838] |
| A_19_P00804755 | 0,002161007 | down | -1,604823 | METTL15 | Homo sapiens methyltransferase like 15 (METTL15), transcript variant 2, mRNA [NM_152636] |
| A_24_P212074 | 0,006087913 | up | 1,6536155 | METTL21C | Homo sapiens methyltransferase like 21C (METTL21C), mRNA [NM_001010977] |
| A_33_P3384322 | 0,010277949 | up | 1,6992383 | METTL22 | methyltransferase like 22 [Source:HGNC Symbol;Acc:HGNC:28368] [ENST00000381920] |
| A_24_P64653 | 0,003744374 | down | -2,4848907 | METTL7B | Homo sapiens methyltransferase like 7B (METTL7B), mRNA [NM_152637] |
| A_23_P39453 | 0,025701828 | down | -1,8752239 | MEX3D | Homo sapiens mex-3 RNA binding family member D (MEX3D), transcript variant 1, mRNA [NM_203304] |
| A_33_P3256680 | 0,013176124 | up | 1,8891271 | MFHAS1 | Homo sapiens malignant fibrous histiocytoma amplified sequence 1 (MFHAS1), mRNA [NM_004225] |
| A_33_P3320277 | 0,038532715 | down | -1,5010661 | MFN2 | Homo sapiens mitofusin 2 (MFN2), transcript variant 1, mRNA [NM_014874] |
| A_23_P41246 | 0,001961342 | up | 1,5904489 | MFSD10 | Homo sapiens major facilitator superfamily domain containing 10 (MFSD10), transcript variant 1, mRNA [NM_001120] |
| A_23_P10062 | 0,010827144 | up | 1,7645781 | MGAT4C | Homo sapiens MGAT4 family, member C (MGAT4C), mRNA [NM_013244] |
| A_21_P0010572 | 0,028876297 | up | 2,6149817 | MGAT4EP | Homo sapiens MGAT4 family, member E, pseudogene (MGAT4EP), non-coding RNA [NR_038135] |
| A_21_P0000716 | 0,00376305 | up | 1,5229214 | MGAT4EP | Homo sapiens MGAT4 family, member E, pseudogene (MGAT4EP), non-coding RNA [NR_038135] |
| A_33_P3526315 | 5,23E-04 | up | 1,8847977 | MGC12488 | Homo sapiens, clone IMAGE:3932794, mRNA. [BC005372] |
| A_23_P410469 | 0,004003068 | up | 1,5547153 | MGC15885 | Homo sapiens uncharacterized protein MGC15885 (MGC15885), long non-coding RNA [NR_026897] |
| A_21_P0001689 | 9,15E-04 | down | -1,9717506 | MGC27382 | Homo sapiens uncharacterized MGC27382 (MGC27382), long non-coding RNA [NR_027310] |
| A_21_P0009655 | 2,26E-04 | down | -2,2472506 | MGC45922 | Homo sapiens uncharacterized LOC284365 (MGC45922), long non-coding RNA [NR_038359] |
| A_33_P3283213 | 0,016931621 | up | 1,6418042 | MGC57346 | Homo sapiens ADP-ribosylation factor pseudogene (MGC57346), transcript variant 2, non-coding RNA [NR_027295] |
| A_33_P3402817 | 0,002759489 | down | -2,1947258 | MGC57346-CRHR1 | Homo sapiens MGC57346-CRHR1 readthrough (MGC57346-CRHR1), transcript variant 6, mRNA [NM_001303016] |
| A_24_P67898 | 0,008551984 | up | 1,7535005 | MGEA5 | Homo sapiens meningioma-expressed antigen 5s splice variant mRNA, complete cds. [AF307332] |
| A_21_P0014058 | 7,40E-04 | up | 2,9577873 | MGST3 | microsomal glutathione S-transferase 3 [Source:HGNC Symbol;Acc:HGNC:7064] [ENST00000461759] |
| A_19_P00316194 | 0,04972662 | down | -1,9060684 | MIAT | Homo sapiens myocardial infarction associated transcript (non-protein coding) (MIAT), transcript variant 1, long non-coding RNA [NR_003491] |
| A_19_P00322828 | 0,004351653 | down | -1,736303 | MIAT | Homo sapiens myocardial infarction associated transcript (non-protein coding) (MIAT), transcript variant 1, long non-coding RNA [NR_003491] |
| A_19_P00322705 | 0,010855231 | down | -2,0042849 | MIAT | Homo sapiens myocardial infarction associated transcript (non-protein coding) (MIAT), transcript variant 1, long non-coding RNA [NR_003491] |
| A_19_P00319573 | 0,003389827 | down | -2,2257543 | MIAT | myocardial infarction associated transcript (non-protein coding) [Source:HGNC Symbol;Acc:HGNC:33425] [ENST00000418918] |
| A_19_P00322706 | 3,32E-05 | down | -2,6597984 | MIAT | Homo sapiens myocardial infarction associated transcript (non-protein coding) (MIAT), transcript variant 1, long non-coding RNA [NR_003491] |
| A_19_P00318175 | 0,010241238 | up | 1,7715418 | MIAT | Homo sapiens myocardial infarction associated transcript (non-protein coding) (MIAT), transcript variant 2, long non-coding RNA [NR_033319] |
| A_33_P3589819 | 5,94E-04 | down | -2,072167 | MIATNB | Homo sapiens cDNA: FLJ22849 fis, clone KAIA987. [AK026502] |
| A_33_P3315190 | 8,18E-04 | down | -2,0224578 | MICAL2 | Homo sapiens microtubule associated monooxygenase, calponin and LIM domain containing 2 (MICAL2), transcript variant 5, mRNA [NM_001282666] |
| A_33_P3277238 | 0,013301073 | up | 2,444692 | MICU1 | mitochondrial calcium uptake 1 [Source:HGNC Symbol;Acc:HGNC:1530] [ENST00000603173] |
| A_21_P0006407 | 0,040905643 | down | -1,7332528 | MID1 | PREDICTED: Homo sapiens midline 1 (Opitz/BBB syndrome) (MID1), transcript variant X3, misc_RNA [XR_247300] |
| A_21_P0006408 | 0,004775461 | up | 3,311436 | MID1 | PREDICTED: Homo sapiens midline 1 (Opitz/BBB syndrome) (MID1), transcript variant X1, misc_RNA [XR_247298] |
| A_32_P128656 | 0,021887286 | up | 1,8113108 | MID1 | Homo sapiens midline 1 (MID1), transcript variant 1, mRNA [NM_000381] |
| A_33_P3375528 | 0,002755834 | down | -1,5285896 | MIER3 | Homo sapiens mesoderm induction early response 1, family member 3 (MIER3), transcript variant 3, mRNA [NM_152622] |
| A_23_P18993 | 0,001299637 | down | -2,1060343 | MIER3 | Homo sapiens mesoderm induction early response 1, family member 3 (MIER3), transcript variant 3, mRNA [NM_152622] |
| A_24_P33982 | 1,82E-04 | down | -3,1755064 | MILR1 | Homo sapiens mast cell immunoglobulin-like receptor 1 (MILR1), transcript variant S2, mRNA [NM_001291317] |
| A_33_P3389133 | 0,030689828 | down | -1,7485473 | MINPP1 | Homo sapiens multiple inositol-polyphosphate phosphatase 1 (MINPP1), transcript variant 2, mRNA [NM_001178117] |
| A_23_P68998 | 0,003807367 | down | -1,6993451 | MIOX | Homo sapiens myo-inositol oxygenase (MIOX), mRNA [NM_017584] |
| A_19_P00322941 | 4,41E-07 | down | -4,0554934 | MIR143HG | MIR143 host gene (non-protein coding) [Source:HGNC Symbol;Acc:HGNC:42872] [ENST00000518014] |
| A_19_P00316284 | 0,04593981 | down | -1,7991551 | MIR143HG | MIR143 host gene (non-protein coding) [Source:HGNC Symbol;Acc:HGNC:42872] [ENST00000614517] |
| A_33_P3267612 | 4,36E-05 | down | -3,003199 | MIR143HG | Homo sapiens MIR143 host gene (non-protein coding) (MIR143HG), transcript variant 1, long non-coding RNA [NR_105059] |
| A_19_P00321399 | 0,003079232 | down | -2,0186248 | MIR143HG | MIR143 host gene (non-protein coding) [Source:HGNC Symbol;Acc:HGNC:42872] [ENST00000505254] |
| A_21_P0004073 | 0,04853896 | up | 2,2245014 | MIR143HG | Homo sapiens MIR143 host gene (non-protein coding) (MIR143HG), transcript variant 1, long non-coding RNA [NR_105059] |
| A_24_P253723 | 0,005076202 | down | -1,5041326 | MIR22HG | Homo sapiens MIR22 host gene (non-protein coding) (MIR22HG), transcript variant 1, long non-coding RNA [NR_028502] |
| A_21_P0009446 | 0,010370562 | up | 1,6666968 | MIR3976HG | MIR3976 host gene (non-protein coding) [Source:HGNC Symbol;Acc:HGNC:51104] [ENST00000580378] |
| A_23_P216935 | 0,021649934 | up | 1,977444 | MIR600HG | Homo sapiens MIR600 host gene (non-protein coding) (MIR600HG), long non-coding RNA [NR_026677] |
| A_21_P0012131 | 0,002684358 | down | -2,72532 | MIR646HG | Homo sapiens MIR646 host gene (non-protein coding) (MIR646HG), long non-coding RNA [NR_046099] |

| | | | | | |
|---|---|---|---|---|---|
| A_33_P3215288 | 0,016712015 | down | -1,8486344 | MIR646HG | Homo sapiens cDNA FLJ46426 fis, clone THYMU3013897. [AK128288] |
| A_32_P16053 | 0,022840051 | up | 1,576171 | MIR663AHG | Homo sapiens MIR663A host gene (non-protein coding) (MIR663AHG), long non-coding RNA [NR_040095] |
| A_21_P0010972 | 0,022541068 | up | 1,6325399 | MIR670HG | PREDICTED: Homo sapiens MIR670 host gene (non-protein coding) (MIR670HG), transcript variant X1, ncRNA [XR_109044] |
| A_23_P353667 | 0,007241065 | down | -2,7483463 | MIR7-3HG | Homo sapiens MIR7-3 host gene (non-protein coding) (MIR7-3HG), long non-coding RNA [NR_027148] |
| A_33_P3265111 | 0,018409802 | up | 1,9715332 | MIR7-3HG | Homo sapiens MIR7-3 host gene (non-protein coding) (MIR7-3HG), long non-coding RNA [NR_027148] |
| A_33_P3312384 | 7,73E-04 | down | -3,0419917 | MIR99AHG | Homo sapiens mir-99a-let-7c cluster host gene (non-protein coding) (MIR99AHG), transcript variant 1, long non-coding RNA [NR_027790] |
| A_21_P0010141 | 0,006614401 | down | -1,9844428 | MIR99AHG | mir-99a-let-7c cluster host gene (non-protein coding) [Source:HGNC Symbol;Acc:HGNC:1274] [ENST00000435697] |
| A_33_P3345479 | 0,01243664 | up | 1,7927265 | MIRLET7BHG | Homo sapiens MIRLET7B host gene (non-protein coding) (MIRLET7BHG), transcript variant 1, long non-coding RNA [NR_027033] |
| A_23_P364107 | 4,38E-05 | up | 1,9610615 | MIS18BP1 | Homo sapiens MIS18 binding protein 1 (MIS18BP1), mRNA [NM_018353] |
| A_24_P346855 | 0,007944907 | up | 1,645899 | MKI67 | Homo sapiens marker of proliferation Ki-67 (MKI67), transcript variant 1, mRNA [NM_002417] |
| A_21_P0000789 | 0,002583901 | down | -2,3862584 | MKNK1-AS1 | Homo sapiens MKNK1 antisense RNA 1 (MKNK1-AS1), long non-coding RNA [NR_038403] |
| A_23_P43350 | 0,0119388 | down | -2,5683417 | MLANA | Homo sapiens melan-A (MLANA), mRNA [NM_005511] |
| A_21_P0002521 | 0,009276412 | down | -2,2020385 | MLK7-AS1 | Homo sapiens MLK7 antisense RNA 1 (MLK7-AS1), long non-coding RNA [NR_033882] |
| A_23_P61050 | 0,006309668 | up | 1,5196759 | MLKL | Homo sapiens mixed lineage kinase domain-like (MLKL), transcript variant 1, mRNA [NM_152649] |
| A_23_P436353 | 0,003042342 | up | 1,8923386 | MLLT4 | Homo sapiens myeloid/lymphoid or mixed-lineage leukemia (trithorax homolog, Drosophila); translocated to, 4 (MLLT4), transcript variant 4, mRNA [NM_001291964] |
| A_21_P0004803 | 0,007520729 | up | 2,4520588 | MLLT4-AS1 | Homo sapiens MLLT4 antisense RNA 1 (head to head) (MLLT4-AS1), long non-coding RNA [NR_027906] |
| A_21_P0004591 | 0,01339912 | up | 1,6700851 | MLLT4-AS1 | Homo sapiens MLLT4 antisense RNA 1 (head to head) (MLLT4-AS1), long non-coding RNA [NR_027906] |
| A_32_P98683 | 0,03706403 | up | 1,5619074 | MLLT6 | Homo sapiens myeloid/lymphoid or mixed-lineage leukemia (trithorax homolog, Drosophila); translocated to, 6 (MLLT6), mRNA [NM_005937] |
| A_23_P19529 | 5,31E-04 | down | -2,238329 | MLN | Homo sapiens motilin (MLN), transcript variant 2, mRNA [NM_001040109] |
| A_23_P165783 | 0,008579084 | up | 1,9938464 | MLPH | Homo sapiens melanophilin (MLPH), transcript variant 1, mRNA [NM_024101] |
| A_23_P164148 | 0,011515264 | up | 1,5282754 | MLX | Homo sapiens MLX, MAX dimerization protein (MLX), transcript variant 3, mRNA [NM_170607] |
| A_23_P9823 | 0,001913563 | up | 1,8534033 | MLXIP | Homo sapiens MLX interacting protein (MLXIP), mRNA [NM_014938] |
| A_33_P3390643 | 0,009352758 | down | -2,150833 | MMACHC | Homo sapiens methylmalonic aciduria (cobalamin deficiency) cblC type, with homocystinuria (MMACHC), mRNA [NM_015506] |
| A_33_P3221203 | 0,004004173 | up | 2,487406 | MMP13 | Homo sapiens matrix metallopeptidase 13 (collagenase 3) (MMP13), mRNA [NM_002427] |
| A_23_P136413 | 0,013363938 | down | -1,582584 | MMP17 | Homo sapiens matrix metallopeptidase 17 (membrane-inserted) (MMP17), mRNA [NM_016155] |
| A_33_P3262124 | 0,010596427 | down | -1,7448127 | MMP25 | Homo sapiens matrix metallopeptidase 25 (MMP25), mRNA [NM_022468] |
| A_23_P150064 | 0,030921496 | down | -2,1384914 | MMRN2 | Homo sapiens multimerin 2 (MMRN2), mRNA [NM_024756] |
| A_23_P46982 | 0,00389012 | up | 1,6282198 | MMS19 | Homo sapiens MMS19 nucleotide excision repair homolog (S. cerevisiae) (MMS19), transcript variant 1, mRNA [NM_022362] |
| A_24_P83678 | 0,013596161 | up | 1,5543054 | MMS22L | Homo sapiens MMS22-like, DNA repair protein (MMS22L), mRNA [NM_198468] |
| A_32_P95914 | 0,018637527 | up | 1,6134751 | MMS22L | Homo sapiens MMS22-like, DNA repair protein (MMS22L), mRNA [NM_198468] |
| A_23_P133123 | 0,002373149 | up | 1,9124838 | MND1 | Homo sapiens meiotic nuclear divisions 1 homolog (S. cerevisiae) (MND1), transcript variant 1, mRNA [NM_032117] |
| A_23_P341443 | 0,006977398 | up | 1,6077263 | MNT | Homo sapiens MAX network transcriptional repressor (MNT), mRNA [NM_020310] |
| A_33_P3347622 | 0,015678398 | down | -1,5132842 | MOB1B | Homo sapiens MOB kinase activator 1B (MOB1B), transcript variant 3, mRNA [NM_001244767] |
| A_23_P130919 | 7,18E-04 | up | 1,6796283 | MOB3A | Homo sapiens MOB kinase activator 3A (MOB3A), mRNA [NM_130807] |
| A_33_P3718014 | 0,014855491 | up | 2,4699988 | MOBP | Homo sapiens myelin-associated oligodendrocyte basic protein (MOBP), transcript variant 6, non-coding RNA [NR_103505] |
| A_33_P3391439 | 0,03170417 | up | 1,6865513 | MOBP | Homo sapiens myelin-associated oligodendrocyte basic protein (MOBP), transcript variant 6, non-coding RNA [NR_103505] |
| A_23_P109614 | 0,006464911 | up | 1,7945963 | MOBP | Homo sapiens myelin-associated oligodendrocyte basic protein (MOBP), transcript variant 3, mRNA [NM_182935] |
| A_33_P3350202 | 1,21E-04 | down | -2,2234743 | MOCS3 | Homo sapiens molybdenum cofactor synthesis 3 (MOCS3), mRNA [NM_014484] |
| A_33_P3414242 | 0,0011432 | down | -2,440265 | MOG | Homo sapiens myelin oligodendrocyte glycoprotein (MOG), transcript variant beta2, mRNA [NM_001008229] |
| A_33_P3353866 | 0,002421997 | down | -2,320081 | MOGAT2 | Homo sapiens monoacylglycerol O-acyltransferase 2 (MOGAT2), mRNA [NM_025098] |
| A_23_P317796 | 4,69E-04 | down | -2,7246964 | MOGAT3 | Homo sapiens monoacylglycerol O-acyltransferase 3 (MOGAT3), transcript variant 1, mRNA [NM_178176] |
| A_33_P3326984 | 0,023765568 | up | 1,7533274 | MOK | MOK protein kinase [Source:HGNC Symbol;Acc:HGNC:9833] [ENST00000520252] |
| A_33_P3308626 | 0,00661672 | down | -2,3428545 | MON1B | Homo sapiens MON1 secretory trafficking family member B (MON1B), transcript variant 1, mRNA [NM_014940] |
| A_24_P419028 | 0,006457241 | down | -2,4797826 | MOP-1 | Homo sapiens mRNA for MOP-1, complete cds. [AB014771] |
| A_24_P100234 | 2,60E-04 | up | 1,796292 | MORC2 | Homo sapiens MORC family CW-type zinc finger 2 (MORC2), transcript variant 3, mRNA [NM_014941] |
| A_32_P122793 | 5,45E-04 | up | 2,050473 | MORC2-AS1 | Homo sapiens MORC2 antisense RNA 1 (MORC2-AS1), long non-coding RNA [NR_026920] |
| A_23_P325501 | 0,004822488 | up | 1,5546386 | MORC3 | Homo sapiens MORC family CW-type zinc finger 3 (MORC3), mRNA [NM_015358] |
| A_24_P165205 | 9,06E-04 | down | -4,1114454 | MORN1 | Homo sapiens MORN repeat containing 1 (MORN1), transcript variant 1, mRNA [NM_024848] |
| A_33_P3308434 | 0,003482816 | down | -1,6105392 | MORN1 | Homo sapiens MORN repeat containing 1 (MORN1), transcript variant 2, mRNA [NM_001301060] |
| A_23_P73835 | 0,001923207 | up | 1,5501475 | MOSPD1 | Homo sapiens motile sperm domain containing 1 (MOSPD1), mRNA [NM_019556] |
| A_24_P231010 | 0,02556332 | up | 2,4785457 | MOV10L1 | Homo sapiens Mov10 RISC complex RNA helicase like 1 (MOV10L1), transcript variant 1, mRNA [NM_018995] |
| A_23_P31064 | 0,007477664 | down | -2,7419415 | MOXD1 | Homo sapiens monooxygenase, DBH-like 1 (MOXD1), mRNA [NM_015529] |
| A_33_P3301778 | 0,013726661 | down | -2,774984 | MOXD2P | Homo sapiens monooxygenase, DBH-like 2, pseudogene (MOXD2P), non-coding RNA [NR_024346] |
| A_23_P71598 | 0,002726142 | up | 2,0989473 | MPDZ | Homo sapiens multiple PDZ domain protein (MPDZ), transcript variant 1, mRNA [NM_003829] |
| A_33_P3212232 | 0,012612782 | up | 2,5327194 | MPEG1 | Homo sapiens macrophage expressed 1 (MPEG1), mRNA [NM_001039396] |
| A_24_P398500 | 0,033549447 | down | -1,8777893 | MPHOSPH9 | Homo sapiens M-phase phosphoprotein 9 (MPHOSPH9), transcript variant 1, mRNA [NM_022782] |
| A_24_P390833 | 0,011031075 | up | 1,7277579 | MPPE1 | Homo sapiens metallophosphoesterase 1 (MPPE1), transcript variant 1, mRNA [NM_023075] |
| A_19_P00317887 | 0,024228279 | up | 1,9214084 | MPRIP | Homo sapiens myosin phosphatase Rho interacting protein (MPRIP), transcript variant 2, mRNA [NM_201274] |
| A_19_P00803575 | 5,16E-04 | up | 3,039543 | MPRIP | Homo sapiens myosin phosphatase Rho interacting protein (MPRIP), transcript variant 2, mRNA [NM_201274] |
| A_19_P00811812 | 0,019071246 | up | 1,9129765 | MPRIP | Homo sapiens myosin phosphatase Rho interacting protein (MPRIP), transcript variant 2, mRNA [NM_201274] |
| A_33_P3230528 | 0,017230224 | up | 1,9477117 | MPRIP | myosin phosphatase Rho interacting protein [Source:HGNC Symbol;Acc:HGNC:30321] [ENST00000395807] |

| | | | | | |
|---|---|---|---|---|---|
| A_23_P15348 | 0,00136183 | up | 2,2845612 | MPRIP | Homo sapiens myosin phosphatase Rho interacting protein (MPRIP), transcript variant 1, mRNA [NM_015134] |
| A_23_P357351 | 0,003569885 | down | -1,8905705 | MPV17L | Homo sapiens MPV17 mitochondrial membrane protein-like (MPV17L), transcript variant 2, mRNA [NM_173803] |
| A_23_P165130 | 0,001454862 | down | -1,5287973 | MPV17L2 | Homo sapiens MPV17 mitochondrial membrane protein-like 2 (MPV17L2), mRNA [NM_032683] |
| A_23_P138089 | 2,99E-04 | down | -2,2869017 | MPZ | Homo sapiens myelin protein zero (MPZ), mRNA [NM_000530] |
| A_24_P126210 | 0,011105979 | up | 2,0579045 | MRAP | Homo sapiens melanocortin 2 receptor accessory protein (MRAP), transcript variant 1, mRNA [NM_178817] |
| A_33_P3303176 | 0,004048911 | down | -1,5862625 | MRGPRG | Homo sapiens MAS-related GPR, member G (MRGPRG), mRNA [NM_001164377] |
| A_23_P365725 | 0,001653301 | down | -1,9150255 | MRGPRX1 | Homo sapiens MAS-related GPR, member X1 (MRGPRX1), mRNA [NM_147199] |
| A_21_P0014055 | 0,006977658 | up | 1,7973994 | MROH1 | Homo sapiens maestro heat-like repeat family member 1 (MROH1), transcript variant 4, mRNA [NM_001288814] |
| A_23_P407080 | 0,002028554 | up | 2,174302 | MROH2B | Homo sapiens maestro heat-like repeat family member 2B (MROH2B), mRNA [NM_173489] |
| A_33_P3227686 | 1,76E-06 | down | -4,0350814 | MROH5 | Homo sapiens maestro heat-like repeat family member 5 (MROH5), transcript variant 1, mRNA [NM_207414] |
| A_21_P0007310 | 0,004111955 | down | -2,6106217 | MRPL23-AS1 | Homo sapiens MRPL23 antisense RNA 1 (MRPL23-AS1), long non-coding RNA [NR_024471] |
| A_33_P3410259 | 0,021648256 | up | 1,6411436 | MRPL23-AS1 | Homo sapiens MRPL23 antisense RNA 1 (MRPL23-AS1), long non-coding RNA [NR_024471] |
| A_33_P3304252 | 2,37E-04 | up | 1,7480198 | MRPL32 | Homo sapiens mitochondrial ribosomal protein L32 (MRPL32), mRNA [NM_031903] |
| A_32_P16489 | 0,001954969 | up | 1,5613405 | MRPL35 | Homo sapiens mitochondrial ribosomal protein L35 (MRPL35), transcript variant 2, mRNA [NM_145644] |
| A_33_P3316233 | 0,012672559 | down | -2,088455 | MRPS18C | Homo sapiens mitochondrial ribosomal protein S18C (MRPS18C), transcript variant 1, mRNA [NM_016067] |
| A_21_P0011187 | 0,001542367 | down | -1,7500587 | MRPS31P5 | Homo sapiens mitochondrial ribosomal protein S31 pseudogene 5 (MRPS31P5), transcript variant 1, non-coding RNA [NR_002816] |
| A_21_P0011186 | 0,004924431 | down | -2,0679207 | MRPS31P5 | mitochondrial ribosomal protein S31 pseudogene 5 [Source:HGNC Symbol;Acc:HGNC:39253] [ENST00000606619] |
| A_33_P3255814 | 0,00120186 | up | 1,5465524 | MRS2 | Homo sapiens MRS2 magnesium transporter (MRS2), transcript variant 1, mRNA [NM_001286264] |
| A_33_P3290577 | 0,02732534 | down | -2,1473665 | MS4A14 | Homo sapiens membrane-spanning 4-domains, subfamily A, member 14 (MS4A14), transcript variant 4, mRNA [NM_001261828] |
| A_33_P3402257 | 0,004964944 | down | -5,3955584 | MS4A18 | PREDICTED: Homo sapiens membrane-spanning 4-domains, subfamily A, member 18 (MS4A18), mRNA [XM_006709948] |
| A_33_P3295313 | 1,24E-04 | down | -2,3610694 | MS4A2 | Homo sapiens membrane-spanning 4-domains, subfamily A, member 2 (MS4A2), transcript variant 1, mRNA [NM_000139] |
| A_23_P104972 | 1,70E-04 | down | -2,050914 | MS4A5 | Homo sapiens membrane-spanning 4-domains, subfamily A, member 5 (MS4A5), mRNA [NM_023945] |
| A_23_P139146 | 0,018656688 | up | 1,6567833 | MS4A8 | Homo sapiens membrane-spanning 4-domains, subfamily A, member 8 (MS4A8), mRNA [NM_031457] |
| A_33_P3418798 | 0,005807373 | down | -1,5711458 | MSANTD1 | Homo sapiens Myb/SANT-like DNA-binding domain containing 1 (MSANTD1), mRNA [NM_001042690] |
| A_23_P52885 | 0,001690496 | up | 1,846882 | MSANTD2 | Homo sapiens Myb/SANT-like DNA-binding domain containing 2 (MSANTD2), transcript variant 1, mRNA [NM_024631] |
| A_21_P0011802 | 5,11E-04 | down | -1,9990914 | MSH6 | mutS homolog 6 [Source:HGNC Symbol;Acc:HGNC:7329] [ENST00000454137] |
| A_23_P132438 | 2,86E-04 | up | 1,7242018 | MSL2 | Homo sapiens male-specific lethal 2 homolog (Drosophila) (MSL2), transcript variant 1, mRNA [NM_018133] |
| A_19_P00801279 | 2,42E-04 | down | -3,2195396 | MSN | moesin [Source:HGNC Symbol;Acc:HGNC:7373] [ENST00000429601] |
| A_21_P0010521 | 0,048516165 | down | -1,9153457 | MST1 | macrophage stimulating 1 (hepatocyte growth factor-like) [Source:HGNC Symbol;Acc:HGNC:7380] [ENST00000481055] |
| A_21_P0010517 | 0,023180293 | down | -1,8026766 | MST1 | macrophage stimulating 1 (hepatocyte growth factor-like) [Source:HGNC Symbol;Acc:HGNC:7380] [ENST00000490966] |
| A_33_P3256550 | 0,001982672 | down | -2,2743616 | MST152 | Homo sapiens MSTP152 (MST152) mRNA, complete cds. [AF190155] |
| A_21_P0010692 | 0,018854633 | down | -1,5109818 | MST1P2 | Homo sapiens macrophage stimulating 1 (hepatocyte growth factor-like) pseudogene 2 (MST1P2), non-coding RNA [NR_027504] |
| A_33_P3351934 | 0,002086713 | down | -1,709015 | MSTO1 | Homo sapiens misato homolog 1 (Drosophila), mRNA (cDNA clone IMAGE:4828998). [BC070067] |
| A_24_P132008 | 5,24E-04 | down | -2,3607066 | MSX2 | Homo sapiens msh homeobox 2 (MSX2), mRNA [NM_002449] |
| A_23_P54840 | 0,045061547 | up | 1,9189141 | MT1A | Homo sapiens metallothionein 1A (MT1A), mRNA [NM_005946] |
| A_33_P3780505 | 0,004210844 | up | 1,5617253 | MT1DP | Homo sapiens metallothionein 1D, pseudogene (functional) (MT1DP), transcript variant 1, non-coding RNA [NR_003658] |
| A_23_P420942 | 0,015363483 | up | 2,442063 | MT1E | PREDICTED: Homo sapiens metallothionein 1E (MT1E), transcript variant X1, mRNA [XM_005255956] |
| A_21_P0000671 | 0,016916355 | up | 2,0678828 | MT1E | Homo sapiens metallothionein 1E (MT1E), mRNA [NM_175617] |
| A_33_P3315314 | 0,01369161 | up | 2,364064 | MT1HL1 | Homo sapiens metallothionein 1H-like 1 (MT1HL1), mRNA [NM_001276687] |
| A_33_P3236868 | 0,03354789 | up | 1,7321408 | MT1X | Homo sapiens metallothionein 1X (MT1X), mRNA [NM_005952] |
| A_23_P97195 | 6,49E-04 | up | 1,6560745 | MTFR1L | Homo sapiens mitochondrial fission regulator 1-like (MTFR1L), transcript variant 1, mRNA [NM_019557] |
| A_24_P382630 | 0,026771845 | down | -1,5056959 | MTG2 | Homo sapiens mitochondrial ribosome-associated GTPase 2 (MTG2), mRNA [NM_015666] |
| A_23_P161507 | 0,025734615 | up | 1,8815855 | MTL5 | Homo sapiens metallothionein-like 5, testis-specific (tesmin) (MTL5), transcript variant 1, mRNA [NM_004923] |
| A_23_P73530 | 1,85E-05 | up | 2,0140069 | MTMR1 | Homo sapiens myotubularin related protein 1 (MTMR1), mRNA [NM_003828] |
| A_23_P127418 | 0,01296101 | up | 2,4161127 | MTNR1B | Homo sapiens melatonin receptor 1B (MTNR1B), mRNA [NM_005959] |
| A_21_P0000102 | 0,015824515 | down | -1,5537988 | MTRNR2L2 | Homo sapiens MT-RNR2-like 2 (MTRNR2L2), mRNA [NM_001190470] |
| A_21_P0003137 | 6,50E-04 | down | -1,6873052 | MTRNR2L2 | Homo sapiens MT-RNR2-like 2 (MTRNR2L2), mRNA [NM_001190470] |
| A_21_P0000104 | 0,002941521 | down | -2,8835225 | MTRNR2L4 | Homo sapiens MT-RNR2-like 4 (MTRNR2L4), mRNA [NM_001190476] |
| A_21_P0000109 | 0,001559489 | down | -2,458362 | MTRNR2L9 | Homo sapiens MT-RNR2-like 9 (pseudogene) (MTRNR2L9), mRNA [NM_001190706] |
| A_23_P347632 | 0,007445417 | up | 1,8250667 | MTSS1 | Homo sapiens metastasis suppressor 1 (MTSS1), transcript variant 2, mRNA [NM_014751] |
| A_33_P3354256 | 0,042932354 | up | 1,6628829 | MTSS1 | Homo sapiens metastasis suppressor 1 (MTSS1), transcript variant 1, mRNA [NM_001282971] |
| A_23_P403745 | 0,015737431 | up | 1,8079444 | MTSS1L | Homo sapiens metastasis suppressor 1-like (MTSS1L), mRNA [NM_138383] |
| A_33_P3356846 | 0,01568203 | down | -1,5823991 | MTX3 | Homo sapiens metaxin 3 (MTX3), transcript variant 2, mRNA [NM_001010891] |
| A_23_P137856 | 0,033491045 | up | 2,250113 | MUC1 | Homo sapiens mucin 1, cell surface associated (MUC1), transcript variant 1, mRNA [NM_002456] |
| A_33_P3352038 | 0,018443268 | up | 1,7946509 | MUC12 | Homo sapiens mucin 12, cell surface associated (MUC12), mRNA [NM_001164462] |
| A_21_P0014035 | 0,001519333 | up | 1,8026568 | MUC19 | Homo sapiens mucin 19, oligomeric (MUC19), mRNA [NM_173600] |
| A_23_P256784 | 4,67E-06 | down | -2,5710428 | MUC2 | Homo sapiens mucin 2, oligomeric mucus/gel-forming (MUC2), mRNA [NM_002457] |
| A_33_P3226775 | 0,003156152 | up | 2,1462955 | MUC20 | Homo sapiens mucin 20, cell surface associated (MUC20), transcript variant S, mRNA [NM_001098516] |
| A_33_P3275035 | 0,001146135 | up | 6,4944553 | MUC21 | mucin-21 precursor [Source:RefSeq peptide;Acc:NP_001010909] [ENST00000551904] |
| A_21_P0004641 | 2,94E-07 | down | -4,0721626 | MUC22 | Homo sapiens mucin 22 (MUC22), mRNA [NM_001198815] |
| A_33_P3236902 | 1,56E-04 | down | -2,4708188 | MUC3A | Homo sapiens mucin 3A, cell surface associated (MUC3A), mRNA [NM_005960] |

| | | | | | |
|---|---|---|---|---|---|
| A_23_P313278 | 2,47E-06 | down | -3,7260416 | MUC3A | Homo sapiens MUC3A mRNA for intestinal mucin, partial cds. [AB038784] |
| A_33_P3331752 | 0,009544668 | down | -2,3752792 | MUC3A | Homo sapiens mucin 3A, cell surface associated (MUC3A), mRNA [NM_005960] |
| A_24_P208825 | 0,001948918 | up | 2,7457297 | MUC4 | Homo sapiens mucin 4, cell surface associated (MUC4), transcript variant 1, mRNA [NM_018406] |
| A_24_P917886 | 0,03567067 | down | -1,8528497 | MUC5AC | PREDICTED: Homo sapiens mucin 5AC, oligomeric mucus/gel-forming (MUC5AC), partial mRNA [XM_006709947] |
| A_33_P3314500 | 0,00168808 | down | -2,3630898 | MUC6 | Homo sapiens mucin 6, oligomeric mucus/gel-forming (MUC6), mRNA [NM_005961] |
| A_24_P88079 | 0,028105382 | down | -2,2698305 | MUC6 | Homo sapiens mucin 6, oligomeric mucus/gel-forming (MUC6), mRNA [NM_005961] |
| A_23_P161634 | 0,001427964 | up | 1,5634254 | MUS81 | Homo sapiens MUS81 structure-specific endonuclease subunit (MUS81), mRNA [NM_025128] |
| A_33_P3230788 | 6,73E-04 | down | -2,6201136 | MUSK | muscle, skeletal, receptor tyrosine kinase [Source:HGNC Symbol;Acc:HGNC:7525] [ENST00000374439] |
| A_23_P71649 | 0,012849936 | down | -1,946675 | MUSK | Homo sapiens muscle, skeletal, receptor tyrosine kinase (MUSK), transcript variant 1, mRNA [NM_005592] |
| A_23_P152125 | 0,03000515 | down | -1,5058655 | MVD | Homo sapiens mevalonate (diphospho) decarboxylase (MVD), mRNA [NM_002461] |
| A_23_P161399 | 0,016381186 | up | 1,675401 | MXI1 | Homo sapiens MAX interactor 1, dimerization protein (MXI1), transcript variant 2, mRNA [NM_130439] |
| A_24_P134195 | 0,035933394 | up | 1,58576 | MYADM | Homo sapiens myeloid-associated differentiation marker (MYADM), transcript variant 1, mRNA [NM_001020818] |
| A_33_P3354703 | 0,026728408 | up | 1,8965278 | MYADML | Homo sapiens myeloid-associated differentiation marker-like (pseudogene) (MYADML), non-coding RNA [NR_003143] |
| A_24_P110331 | 0,021348467 | up | 2,2610419 | MYADML2 | Homo sapiens myeloid-associated differentiation marker-like 2 (MYADML2), mRNA [NM_001145113] |
| A_23_P143190 | 0,009680659 | down | -2,3747523 | MYBL2 | Homo sapiens v-myb avian myeloblastosis viral oncogene homolog-like 2 (MYBL2), transcript variant 1, mRNA [NM_002466] |
| A_23_P128362 | 0,019101294 | up | 2,9619708 | MYBPC1 | Homo sapiens myosin binding protein C, slow type (MYBPC1), transcript variant 2, mRNA [NM_206819] |
| A_23_P215956 | 0,006527643 | up | 1,7077779 | MYC | Homo sapiens v-myc avian myelocytomatosis viral oncogene homolog (MYC), mRNA [NM_002467] |
| A_23_P67952 | 4,03E-04 | down | -2,131788 | MYCNOS | Homo sapiens MYCN opposite strand (MYCNOS), transcript variant 2, long non-coding RNA [NR_026766] |
| A_21_P0002574 | 9,75E-05 | down | -2,601155 | MYCNUT | Homo sapiens MYCN upstream transcript (non-protein coding) (MYCNUT), long non-coding RNA [NR_125783] |
| A_21_P0001830 | 0,03766443 | up | 2,6764069 | MYCNUT | Homo sapiens MYCN upstream transcript (non-protein coding) (MYCNUT), long non-coding RNA [NR_125783] |
| A_23_P76460 | 0,025056493 | down | -2,0821393 | MYF6 | Homo sapiens myogenic factor 6 (herculin) (MYF6), mRNA [NM_002469] |
| A_23_P206920 | 0,001735333 | up | 2,8108711 | MYH11 | Homo sapiens myosin, heavy chain 11, smooth muscle (MYH11), transcript variant SM1B, mRNA [NM_001040114] |
| A_23_P89334 | 0,03966881 | up | 2,2639663 | MYH13 | Homo sapiens myosin, heavy chain 13, skeletal muscle (MYH13), mRNA [NM_003802] |
| A_24_P323545 | 7,87E-05 | down | -2,9340646 | MYH14 | Homo sapiens myosin, heavy chain 14, non-muscle (MYH14), transcript variant 1, mRNA [NM_001077186] |
| A_23_P66530 | 0,009032233 | up | 1,7220699 | MYH4 | Homo sapiens myosin, heavy chain 4, skeletal muscle (MYH4), mRNA [NM_017533] |
| A_21_P0013396 | 0,008437656 | up | 1,8822068 | MYH6 | Homo sapiens myosin, heavy chain 6, cardiac muscle, alpha (MYH6), mRNA [NM_002471] |
| A_24_P408424 | 0,047443025 | up | 1,8105723 | MYH9 | Homo sapiens myosin, heavy chain 9, non-muscle (MYH9), mRNA [NM_002473] |
| A_23_P393015 | 0,008702905 | up | 2,471978 | MYL10 | Homo sapiens myosin, light chain 10, regulatory (MYL10), mRNA [NM_138403] |
| A_23_P59738 | 0,002927366 | down | -1,8630004 | MYL7 | Homo sapiens myosin, light chain 7, regulatory (MYL7), mRNA [NM_021223] |
| A_23_P143817 | 0,025058301 | down | -2,069211 | MYLK | Homo sapiens myosin light chain kinase (MYLK), transcript variant 1, mRNA [NM_053025] |
| A_24_P319923 | 0,03428257 | down | -2,2214272 | MYLK | Homo sapiens myosin light chain kinase (MYLK), transcript variant 1, mRNA [NM_053025] |
| A_23_P80008 | 0,038759314 | down | -1,5519135 | MYLK2 | Homo sapiens myosin light chain kinase 2 (MYLK2), mRNA [NM_033118] |
| A_33_P3300217 | 0,003170568 | down | -2,0050418 | MYLK3 | Homo sapiens myosin light chain kinase 3 (MYLK3), mRNA [NM_182493] |
| A_24_P195669 | 0,035682242 | up | 1,714218 | MYO15B | myosin XVB pseudogene [Source:HGNC Symbol;Acc:HGNC:14083] [ENST00000581866] |
| A_23_P162288 | 2,63E-04 | down | -2,5615146 | MYO1A | Homo sapiens myosin IA (MYO1A), transcript variant 2, mRNA [NM_005379] |
| A_24_P7143 | 0,001319849 | down | -1,8134135 | MYO1B | Homo sapiens myosin IB (MYO1B), transcript variant 2, mRNA [NM_012223] |
| A_33_P3297978 | 0,002334527 | up | 1,9452218 | MYO1E | Homo sapiens myosin IE (MYO1E), mRNA [NM_004998] |
| A_24_P206343 | 0,013487754 | down | -1,8169161 | MYO1G | Homo sapiens myosin IG (MYO1G), mRNA [NM_033054] |
| A_23_P86411 | 5,30E-04 | down | -12,323614 | MYO3A | Homo sapiens myosin IIIA (MYO3A), mRNA [NM_017433] |
| A_33_P3351617 | 0,003282587 | up | 1,5235124 | MYO3A | myosin IIIA [Source:HGNC Symbol;Acc:HGNC:7601] [ENST00000376302] |
| A_24_P182696 | 0,003192708 | up | 1,800706 | MYO3B | Homo sapiens myosin IIIB (MYO3B), transcript variant 2, mRNA [NM_138995] |
| A_24_P255218 | 0,009917685 | down | -1,5152389 | MYO5A | Homo sapiens myosin VA (heavy chain 12, myoxin) (MYO5A), transcript variant 1, mRNA [NM_000259] |
| A_23_P140434 | 0,042948928 | up | 1,7287023 | MYO5C | Homo sapiens myosin VC (MYO5C), mRNA [NM_018728] |
| A_23_P209799 | 0,03066883 | down | -1,7837999 | MYO7B | Homo sapiens myosin VIIB (MYO7B), mRNA [NM_001080527] |
| A_23_P205841 | 1,20E-04 | up | 1,7710788 | MYO9A | Homo sapiens myosin IXA (MYO9A), mRNA [NM_006901] |
| A_24_P246173 | 0,023108976 | up | 1,5593519 | MYO9B | Homo sapiens myosin IXB (MYO9B), transcript variant 1, mRNA [NM_004145] |
| A_33_P3334773 | 0,036099665 | down | -2,1809733 | MYOD1 | Homo sapiens myogenic differentiation 1 (MYOD1), mRNA [NM_002478] |
| A_23_P1320 | 0,037886802 | up | 2,3622386 | MYOZ1 | Homo sapiens myozenin 1 (MYOZ1), mRNA [NM_021245] |
| A_23_P75790 | 1,13E-04 | down | -2,1697874 | MYRF | Homo sapiens myelin regulatory factor (MYRF), transcript variant 1, mRNA [NM_013279] |
| A_23_P210835 | 0,016813977 | up | 2,4232779 | MYT1 | Homo sapiens myelin transcription factor 1 (MYT1), mRNA [NM_004535] |
| A_33_P3351290 | 0,002003248 | down | -4,2394276 | MZB1 | marginal zone B and B1 cell-specific protein [Source:HGNC Symbol;Acc:HGNC:30125] [ENST00000509591] |
| A_24_P140666 | 0,012045578 | up | 2,3837895 | N4BP3 | Homo sapiens NEDD4 binding protein 3 (N4BP3), mRNA [NM_015111] |
| A_23_P251785 | 0,017950911 | up | 1,5032552 | NAA16 | Homo sapiens N(alpha)-acetyltransferase 16, NatA auxiliary subunit (NAA16), transcript variant 1, mRNA [NM_024561] |
| A_24_P66679 | 0,030297631 | up | 1,5428978 | NAA30 | Homo sapiens N(alpha)-acetyltransferase 30, NatC catalytic subunit (NAA30), mRNA [NM_001011713] |
| A_23_P38206 | 0,00798504 | up | 2,6496568 | NAA38 | Homo sapiens N(alpha)-acetyltransferase 38, NatC auxiliary subunit (NAA38), mRNA [NM_032356] |
| A_33_P3273854 | 0,005689734 | down | -1,8182738 | NAALADL2 | Homo sapiens N-acetylated alpha-linked acidic dipeptidase-like 2 (NAALADL2), mRNA [NM_207015] |
| A_21_P0003061 | 3,08E-05 | down | -3,2124703 | NAALADL2 | N-acetylated alpha-linked acidic dipeptidase-like 2 [Source:HGNC Symbol;Acc:HGNC:23219] [ENST00000481679] |
| A_33_P3269631 | 4,24E-06 | down | -3,1281722 | NAALADL2 | N-acetylated alpha-linked acidic dipeptidase-like 2 [Source:HGNC Symbol;Acc:HGNC:23219] [ENST00000495900] |
| A_21_P0002709 | 0,013174832 | up | 1,571184 | NAALADL2 | N-acetylated alpha-linked acidic dipeptidase-like 2 [Source:HGNC Symbol;Acc:HGNC:23219] [ENST00000491329] |
| A_23_P59888 | 0,015714463 | down | -2,9707885 | NACAP1 | Homo sapiens nascent-polypeptide-associated complex alpha polypeptide pseudogene 1 (NACAP1), non-coding RNA [NR_002182] |
| A_23_P200056 | 0,013418793 | up | 1,5280752 | NADK | Homo sapiens NAD kinase (NADK), transcript variant 1, mRNA [NM_023018] |

| | | | | | |
|---|---|---|---|---|---|
| A_33_P3233649 | 2,58E-04 | down | -1,820731 | NADK2 | Homo sapiens NAD kinase 2, mitochondrial (NADK2), transcript variant 1, mRNA [NM_001085411] |
| A_24_P408457 | 0,001723348 | up | 1,7763001 | NAGA | Homo sapiens N-acetylgalactosaminidase, alpha- (NAGA), mRNA [NM_000262] |
| A_33_P3269159 | 8,09E-04 | down | -2,6755955 | NALCN | sodium leak channel, non-selective [Source:HGNC Symbol;Acc:HGNC:19082] [ENST00000470333] |
| A_33_P3364869 | 0,03938168 | up | 1,5717431 | NAMPT | Homo sapiens cDNA FLJ13279 fis, clone OVARC1001055, moderately similar to PRE-B CELL ENHANCING FACTOR PRECURSOR. [AK023341] |
| A_33_P3388453 | 0,0346105 | down | -1,5534446 | NANOS2 | Homo sapiens nanos homolog 2 (Drosophila) (NANOS2), mRNA [NM_001029861] |
| A_24_P132950 | 1,81E-04 | up | 2,2192233 | NAP1L4 | Homo sapiens nucleosome assembly protein 1-like 4 (NAP1L4), mRNA [NM_005969] |
| A_23_P144369 | 4,74E-04 | down | -2,9375165 | NAP1L5 | Homo sapiens nucleosome assembly protein 1-like 5 (NAP1L5), mRNA [NM_153757] |
| A_23_P90130 | 0,022546481 | down | -1,6918042 | NAPSA | Homo sapiens napsin A aspartic peptidase (NAPSA), mRNA [NM_004851] |
| A_19_P00811284 | 0,002538711 | down | -2,5038776 | NASP | Homo sapiens nuclear autoantigenic sperm protein (histone-binding) (NASP), transcript variant 2, mRNA [NM_002482] |
| A_23_P31798 | 4,56E-04 | down | -2,052926 | NAT2 | Homo sapiens N-acetyltransferase 2 (arylamine N-acetyltransferase) (NAT2), mRNA [NM_000015] |
| A_24_P308506 | 2,97E-04 | down | -4,1338882 | NAT8B | Homo sapiens N-acetyltransferase 8B (GCN5-related, putative, gene/pseudogene) (NAT8B), mRNA [NM_016347] |
| A_33_P3885807 | 0,029832877 | up | 2,16845 | NAV2-AS5 | Homo sapiens NAV2 antisense RNA 5 (NAV2-AS5), long non-coding RNA [NR_049725] |
| A_33_P3336273 | 0,001017289 | down | -3,477483 | NAV3 | Homo sapiens neuron navigator 3 (NAV3), transcript variant 1, mRNA [NM_001024383] |
| A_19_P00322653 | 0,03376436 | down | -1,5001124 | NBAT1 | Homo sapiens neuroblastoma associated transcript 1 (NBAT1), long non-coding RNA [NR_034143] |
| A_24_P272160 | 0,005362199 | down | -1,9399726 | NBEAL2 | PREDICTED: Homo sapiens neurobeachin-like 2 (NBEAL2), transcript variant X5, mRNA [XM_006713072] |
| A_24_P278126 | 0,035867598 | up | 1,5225801 | NBN | Homo sapiens nibrin (NBN), mRNA [NM_002485] |
| A_33_P3368800 | 0,003929 | up | 1,5129112 | NBPF12 | Homo sapiens neuroblastoma breakpoint family, member 12 (NBPF12), mRNA [NM_001278141] |
| A_23_P359430 | 4,45E-05 | up | 1,9607549 | NBPF14 | Homo sapiens neuroblastoma breakpoint family, member 14 (NBPF14), mRNA [NM_015383] |
| A_33_P3229328 | 0,037570205 | down | -1,5957004 | NBPF6 | Homo sapiens neuroblastoma breakpoint family, member 6 (NBPF6), transcript variant 2, mRNA [NM_001143988] |
| A_33_P3279162 | 1,05E-04 | down | -2,4012783 | NBPF6 | Homo sapiens neuroblastoma breakpoint family, member 6 (NBPF6), transcript variant 1, mRNA [NM_001143987] |
| A_33_P3279167 | 0,034026597 | up | 2,3573923 | NBPF7 | Homo sapiens neuroblastoma breakpoint family, member 7 (NBPF7), mRNA [NM_001047980] |
| A_23_P153797 | 6,87E-05 | down | -2,359089 | NCAN | Homo sapiens neurocan (NCAN), mRNA [NM_004386] |
| A_32_P71447 | 0,002607989 | up | 1,6109287 | NCAPD3 | Homo sapiens non-SMC condensin II complex, subunit D3 (NCAPD3), mRNA [NM_015261] |
| A_33_P3659876 | 0,018900212 | up | 1,9627743 | NCAPG2 | Homo sapiens non-SMC condensin II complex, subunit G2 (NCAPG2), transcript variant 2, mRNA [NM_001281932] |
| A_23_P138194 | 0,004033855 | down | -1,7657692 | NCF2 | Homo sapiens neutrophil cytosolic factor 2 (NCF2), transcript variant 1, mRNA [NM_000433] |
| A_23_P73239 | 0,009923887 | up | 1,5089098 | NCKAP1 | Homo sapiens NCK-associated protein 1 (NCKAP1), transcript variant 2, mRNA [NM_205842] |
| A_23_P360079 | 0,026163474 | down | -2,1442404 | NCKAP5 | Homo sapiens NCK-associated protein 5 (NCKAP5), transcript variant 1, mRNA [NM_207363] |
| A_33_P3211198 | 0,020378586 | up | 2,657094 | NCMAP | Homo sapiens noncompact myelin associated protein (NCMAP), mRNA [NM_001010980] |
| A_33_P3270009 | 0,013990724 | up | 1,7548628 | NCOA1 | Homo sapiens nuclear receptor coactivator 1 (NCOA1), transcript variant 2, mRNA [NM_147223] |
| A_23_P120442 | 3,45E-06 | up | 2,3146632 | NCOA3 | Homo sapiens nuclear receptor coactivator 3 (NCOA3), transcript variant 1, mRNA [NM_181659] |
| A_23_P210515 | 4,56E-04 | up | 2,0261898 | NCOA5 | Homo sapiens nuclear receptor coactivator 5 (NCOA5), mRNA [NM_020967] |
| A_24_P237661 | 0,005510134 | down | -2,3107212 | NCOR1 | Homo sapiens nuclear receptor corepressor 1 (NCOR1), transcript variant 2, mRNA [NM_001190438] |
| A_33_P3571824 | 0,017173065 | down | -1,6457217 | NCR1 | Homo sapiens natural cytotoxicity triggering receptor 1 (NCR1), transcript variant 1, mRNA [NM_004829] |
| A_33_P3329063 | 0,00876898 | up | 2,9441502 | NCR1 | Homo sapiens natural cytotoxicity triggering receptor 1 (NCR1), transcript variant 1, mRNA [NM_004829] |
| A_33_P3344423 | 0,015026493 | down | -2,2144315 | NCR3 | Homo sapiens natural cytotoxicity triggering receptor 3 (NCR3), transcript variant 3, mRNA [NM_001145467] |
| A_23_P251881 | 0,0266625 | up | 1,5593257 | NCR3 | Homo sapiens natural cytotoxicity triggering receptor 3 (NCR3), transcript variant 1, mRNA [NM_147130] |
| A_21_P0005720 | 0,003437661 | down | -2,6269855 | NCRNA00249 | PREDICTED: Homo sapiens uncharacterized LOC101926947 (NCRNA00249), transcript variant X4, ncRNA [XR_426332] |
| A_21_P0005941 | 7,97E-04 | down | -4,310279 | NCRNA00249 | PREDICTED: Homo sapiens uncharacterized LOC101926947 (NCRNA00249), transcript variant X6, ncRNA [XR_426334] |
| A_21_P0005954 | 0,012923341 | up | 1,5654821 | NCRNA00250 | Homo sapiens non-protein coding RNA 250 (NCRNA00250), long non-coding RNA [NR_126028] |
| A_24_P182122 | 0,006851737 | up | 1,6987934 | ND1 | mitochondrially encoded NADH dehydrogenase 1 [Source:HGNC Symbol;Acc:HGNC:7455] [ENST00000361390] |
| A_23_P431853 | 0,002769837 | up | 1,6081934 | ND2 | mitochondrially encoded NADH dehydrogenase 2 [Source:HGNC Symbol;Acc:HGNC:7456] [ENST00000361453] |
| A_23_P406928 | 0,003309238 | up | 1,7042087 | ND5 | mitochondrially encoded NADH dehydrogenase 5 [Source:HGNC Symbol;Acc:HGNC:7461] [ENST00000361567] |
| A_23_P317056 | 7,15E-04 | down | -2,0310848 | ND6 | mitochondrially encoded NADH dehydrogenase 6 [Source:HGNC Symbol;Acc:HGNC:7462] [ENST00000361681] |
| A_24_P185604 | 0,01608457 | up | 1,8107717 | NDRG3 | Homo sapiens NDRG family member 3 (NDRG3), transcript variant 1, mRNA [NM_032013] |
| A_21_P0014051 | 0,021705482 | down | -1,8673599 | NDUFA6-AS1 | Homo sapiens NDUFA6 antisense RNA 1 (head to head) (NDUFA6-AS1), long non-coding RNA [NR_034118] |
| A_33_P3440264 | 0,030728243 | down | -1,5836536 | NDUFA6-AS1 | Homo sapiens NDUFA6 antisense RNA 1 (head to head) (NDUFA6-AS1), long non-coding RNA [NR_034118] |
| A_32_P93328 | 0,011637275 | down | -2,349616 | NDUFB2-AS1 | Homo sapiens NDUFB2 antisense RNA 1 (NDUFB2-AS1), long non-coding RNA [NR_024454] |
| A_32_P103614 | 0,007229638 | down | -2,2269628 | NEBL-AS1 | Homo sapiens NEBL antisense RNA 1 (NEBL-AS1), transcript variant 1, long non-coding RNA [NR_046283] |
| A_33_P3247629 | 0,002574228 | down | -1,6961678 | NECAP1 | Homo sapiens NECAP endocytosis associated 1 (NECAP1), transcript variant 1, mRNA [NM_015509] |
| A_32_P144920 | 0,016973576 | up | 1,8109 | NECAP1 | Homo sapiens NECAP endocytosis associated 1 (NECAP1), transcript variant 1, mRNA [NM_015509] |
| A_21_P0014167 | 0,024223035 | down | -2,4402568 | NEDD9 | neural precursor cell expressed, developmentally down-regulated 9 [Source:HGNC Symbol;Acc:HGNC:7733] [ENST00000379433] |
| A_33_P3408152 | 0,03880187 | up | 1,9315583 | NEFH | Homo sapiens neurofilament, heavy polypeptide (NEFH), mRNA [NM_021076] |
| A_33_P3341970 | 0,001092668 | down | -2,1198213 | NEGR1 | Homo sapiens neuronal growth regulator 1 (NEGR1), mRNA [NM_173808] |
| A_23_P129157 | 0,015869478 | up | 1,7004247 | NEIL1 | Homo sapiens nei endonuclease VIII-like 1 (E. coli) (NEIL1), transcript variant 2, mRNA [NM_024608] |
| A_23_P155301 | 0,00247403 | down | -1,7221266 | NEK11 | Homo sapiens NIMA-related kinase 11 (NEK11), transcript variant 2, mRNA [NM_145910] |
| A_33_P3354203 | 0,010914825 | down | -2,0274506 | NELL1 | NEL-like 1 (chicken) [Source:HGNC Symbol;Acc:HGNC:7750] [ENST00000534263] |
| A_33_P3651948 | 2,78E-06 | down | -5,6157737 | NEO1 | Homo sapiens neogenin 1 (NEO1), transcript variant 1, mRNA [NM_002499] |
| A_33_P3358233 | 0,015215811 | down | -1,7082175 | NES | Homo sapiens nestin (NES), mRNA [NM_006617] |
| A_23_P83931 | 0,007473439 | up | 1,7759846 | NET1 | Homo sapiens neuroepithelial cell transforming 1 (NET1), transcript variant 1, mRNA [NM_001047160] |
| A_33_P3210085 | 0,003138731 | up | 2,3310716 | NET1 | Homo sapiens neuroepithelial cell transforming 1 (NET1), transcript variant 1, mRNA [NM_001047160] |
| A_23_P4489 | 0,014171049 | down | -2,248982 | NETO1 | Homo sapiens neuropilin (NRP) and tolloid (TLL)-like 1 (NETO1), transcript variant 3, mRNA [NM_138966] |

| | | | | | |
|---|---|---|---|---|---|
| A_23_P203716 | 0,018916348 | down | -1,661997 | NEU3 | Homo sapiens sialidase 3 (membrane sialidase) (NEU3), mRNA [NM_006656] |
| A_33_P3293524 | 0,001673997 | down | -4,031506 | NEURL1 | Homo sapiens neuralized E3 ubiquitin protein ligase 1 (NEURL1), mRNA [NM_004210] |
| A_33_P3222218 | 1,06E-04 | down | -6,7120357 | NEURL1B | Homo sapiens neuralized E3 ubiquitin protein ligase 1B (NEURL1B), mRNA [NM_001142651] |
| A_23_P360179 | 9,02E-04 | up | 1,6177408 | NEURL4 | Homo sapiens neuralized E3 ubiquitin protein ligase 4 (NEURL4), transcript variant 1, mRNA [NM_032442] |
| A_33_P3856171 | 0,004834694 | up | 1,5211654 | NF1P2 | Homo sapiens neurofibromin 1 pseudogene 2 (NF1P2), non-coding RNA [NR_028506] |
| A_33_P3406866 | 0,019105682 | down | -1,5455644 | NFASC | Homo sapiens neurofascin (NFASC), transcript variant 1, mRNA [NM_001005388] |
| A_33_P3406873 | 0,00964368 | down | -1,697259 | NFASC | Homo sapiens neurofascin (NFASC), transcript variant 5, mRNA [NM_001005389] |
| A_23_P359647 | 0,001177051 | up | 1,9955714 | NFAT5 | Homo sapiens nuclear factor of activated T-cells 5, tonicity-responsive (NFAT5), transcript variant 1, mRNA [NM_138714] |
| A_33_P3225600 | 0,003511366 | up | 1,5280656 | NFE2L2 | Homo sapiens nuclear factor, erythroid 2-like 2 (NFE2L2), transcript variant 1, mRNA [NM_006164] |
| A_23_P30024 | 0,004475336 | up | 1,5669713 | NFKB1 | Homo sapiens nuclear factor of kappa light polypeptide gene enhancer in B-cells 1 (NFKB1), transcript variant 1, mRNA [NM_003998] |
| A_23_P106002 | 6,57E-04 | up | 1,9049616 | NFKBIA | Homo sapiens nuclear factor of kappa light polypeptide gene enhancer in B-cells inhibitor, alpha (NFKBIA), mRNA [NM_020529] |
| A_33_P3386506 | 0,020065878 | down | -1,8353041 | NFKBIL1 | Homo sapiens nuclear factor of kappa light polypeptide gene enhancer in B-cells inhibitor-like 1 (NFKBIL1), transcript variant 2, mRNA [NM_001144961] |
| A_23_P212089 | 0,010140986 | up | 1,8924223 | NFKBIZ | Homo sapiens nuclear factor of kappa light polypeptide gene enhancer in B-cells inhibitor, zeta (NFKBIZ), transcript variant 1, mRNA [NM_031419] |
| A_23_P304543 | 0,002166071 | down | -1,507857 | NFX1 | Homo sapiens nuclear transcription factor, X-box binding 1 (NFX1), transcript variant 2, mRNA [NM_147133] |
| A_33_P3355877 | 3,60E-04 | down | -2,5798106 | NFXL1 | Homo sapiens nuclear transcription factor, X-box binding-like 1 (NFXL1), transcript variant 4, non-coding RNA [NR_103795] |
| A_33_P3368706 | 0,026360476 | up | 1,6471084 | NFYC | Homo sapiens nuclear transcription factor Y, gamma (NFYC), transcript variant 5, mRNA [NM_001142590] |
| A_23_P115190 | 0,001995173 | down | -1,6254513 | NGF | Homo sapiens nerve growth factor (beta polypeptide) (NGF), mRNA [NM_002506] |
| A_21_P0000551 | 0,034179255 | down | -1,9869114 | NHEG1 | Homo sapiens neuroblastoma highly expressed 1 (NHEG1), long non-coding RNA [NR_027994] |
| A_24_P251381 | 6,11E-04 | up | 1,713977 | NHEJ1 | Homo sapiens nonhomologous end-joining factor 1 (NHEJ1), mRNA [NM_024782] |
| A_33_P3374258 | 1,28E-04 | down | -2,2045033 | NHLH2 | nescient helix loop helix 2 [Source:HGNC Symbol;Acc:HGNC:7818] [ENST00000369506] |
| A_33_P3264116 | 0,00448698 | down | -1,7100171 | NHLRC1 | Homo sapiens NHL repeat containing E3 ubiquitin protein ligase 1 (NHLRC1), mRNA [NM_198586] |
| A_33_P3260747 | 0,003276244 | down | -2,59381 | NHSL2 | NHS-like 2 [Source:HGNC Symbol;Acc:HGNC:33737] [ENST00000373677] |
| A_33_P3283906 | 0,005401195 | down | -2,1007211 | NIP7 | Homo sapiens NIP7, nucleolar pre-rRNA processing protein (NIP7), transcript variant 1, mRNA [NM_016101] |
| A_23_P170213 | 4,12E-04 | down | -4,72848 | NKAIN2 | Homo sapiens Na+/K+ transporting ATPase interacting 2 (NKAIN2), transcript variant 1, mRNA [NM_001040214] |
| A_33_P3417880 | 0,025176277 | down | -1,5129054 | NKAPL | Homo sapiens NFKB activating protein-like (NKAPL), mRNA [NM_001007531] |
| A_21_P0006584 | 0,005895584 | down | -2,2634559 | NKAPP1 | Homo sapiens NFKB activating protein pseudogene 1 (NKAPP1), non-coding RNA [NR_027131] |
| A_23_P41804 | 0,004860871 | up | 2,1013956 | NKD2 | Homo sapiens naked cuticle homolog 2 (Drosophila) (NKD2), transcript variant 1, mRNA [NM_033120] |
| A_23_P119042 | 0,010292025 | down | -2,5601726 | NKG7 | Homo sapiens natural killer cell granule protein 7 (NKG7), mRNA [NM_005601] |
| A_24_P256155 | 7,83E-04 | down | -1,8861685 | NKX1-2 | Homo sapiens NK1 homeobox 2 (NKX1-2), mRNA [NM_001146340] |
| A_24_P38702 | 0,001249734 | down | -2,038627 | NKX2-3 | Homo sapiens NK2 homeobox 3 (NKX2-3), mRNA [NM_145285] |
| A_33_P3299599 | 9,04E-04 | down | -3,27701 | NKX2-5 | Homo sapiens NK2 homeobox 5 (NKX2-5), transcript variant 2, mRNA [NM_001166175] |
| A_23_P340019 | 0,001037749 | up | 2,2344067 | NLRC3 | Homo sapiens NLR family, CARD domain containing 3 (NLRC3), transcript variant 1, mRNA [NM_178844] |
| A_23_P89550 | 0,04720287 | up | 1,5038708 | NLRP1 | Homo sapiens NLR family, pyrin domain containing 1 (NLRP1), transcript variant 1, mRNA [NM_033004] |
| A_23_P47579 | 0,025943493 | down | -1,7777244 | NLRP14 | Homo sapiens NLR family, pyrin domain containing 14 (NLRP14), mRNA [NM_176822] |
| A_33_P3406047 | 0,030552896 | down | -1,6208805 | NLRP4 | Homo sapiens NLR family, pyrin domain containing 4 (NLRP4), mRNA [NM_134444] |
| A_23_P311632 | 0,029547643 | up | 2,5289338 | NLRP6 | Homo sapiens NLR family, pyrin domain containing 6 (NLRP6), transcript variant 1, mRNA [NM_138329] |
| A_23_P156402 | 0,027857557 | down | -1,9117494 | NME5 | Homo sapiens NME/NM23 family member 5 (NME5), mRNA [NM_003551] |
| A_23_P304652 | 0,006490581 | down | -2,663762 | NME9 | Homo sapiens NME/NM23 family member 9 (NME9), mRNA [NM_178130] |
| A_23_P122134 | 0,019939475 | down | -1,6099445 | NMUR2 | Homo sapiens neuromedin U receptor 2 (NMUR2), mRNA [NM_020167] |
| A_23_P336015 | 0,003438368 | up | 1,7506347 | NOC2L | Homo sapiens nucleolar complex associated 2 homolog (S. cerevisiae) (NOC2L), mRNA [NM_015658] |
| A_23_P138058 | 0,001841893 | up | 1,626602 | NOC2L | Homo sapiens nucleolar complex associated 2 homolog (S. cerevisiae) (NOC2L), mRNA [NM_015658] |
| A_19_P00318172 | 0,003904414 | down | -1,7540485 | NOL4L | Homo sapiens nucleolar protein 4-like (NOL4L), transcript variant 1, mRNA [NM_001256798] |
| A_24_P101800 | 0,044917855 | down | -1,9421266 | NOL4L | nucleolar protein 4-like [Source:HGNC Symbol;Acc:HGNC:16106] [ENST00000485364] |
| A_24_P118938 | 2,03E-04 | up | 2,3503451 | NOL4L | Homo sapiens nucleolar protein 4-like (NOL4L), transcript variant 1, mRNA [NM_001256798] |
| A_23_P372096 | 0,007688603 | up | 1,7032671 | NOL4L | nucleolar protein 4-like [Source:HGNC Symbol;Acc:HGNC:16106] [ENST00000326071] |
| A_24_P21410 | 0,009327647 | up | 1,5646846 | NOL6 | Homo sapiens nucleolar protein 6 (RNA-associated) (NOL6), transcript variant alpha, mRNA [NM_022917] |
| A_33_P3278298 | 0,02167171 | up | 2,3758404 | NOM1 | Homo sapiens cDNA FLJ16401 fis, clone UTERU2004299. [AK131362] |
| A_33_P3378430 | 1,34E-04 | up | 1,8212479 | NOM1 | Homo sapiens nucleolar protein with MIF4G domain 1 (NOM1), mRNA [NM_138400] |
| A_24_P151727 | 0,001767795 | up | 1,5035468 | NONO | Homo sapiens non-POU domain containing, octamer-binding (NONO), transcript variant 2, mRNA [NM_007363] |
| A_23_P204364 | 0,009575786 | up | 1,6311073 | NOP2 | Homo sapiens NOP2 nucleolar protein (NOP2), transcript variant 2, mRNA [NM_001033714] |
| A_23_P79360 | 0,03721539 | down | -1,7113781 | NOSTRIN | Homo sapiens nitric oxide synthase trafficking (NOSTRIN), transcript variant 1, mRNA [NM_052946] |
| A_23_P200792 | 0,004471285 | up | 1,5070963 | NOTCH2 | Homo sapiens notch 2 (NOTCH2), transcript variant 1, mRNA [NM_024408] |
| A_33_P3264771 | 0,020159049 | up | 1,9521946 | NOTO | Homo sapiens notochord homeobox (NOTO), mRNA [NM_001134462] |
| A_24_P270496 | 0,003984513 | up | 1,5070605 | NOVA1 | Homo sapiens neuro-oncological ventral antigen 1 (NOVA1), transcript variant 3, mRNA [NM_006491] |
| A_33_P3224324 | 0,03931472 | up | 1,6053014 | NOX4 | Homo sapiens NADPH oxidase 4 (NOX4), transcript variant NOX4B, mRNA [NM_001143836] |
| A_23_P140475 | 0,037340295 | up | 1,9854882 | NOX5 | Homo sapiens NADPH oxidase, EF-hand calcium binding domain 5 (NOX5), transcript variant 1, mRNA [NM_024505] |
| A_23_P300740 | 0,024487728 | down | -2,076812 | NPB | Homo sapiens neuropeptide B (NPB), mRNA [NM_148896] |
| A_33_P3419970 | 0,001119719 | down | -2,2751696 | NPB | Homo sapiens neuropeptide B (NPB), mRNA [NM_148896] |
| A_23_P107587 | 0,002938846 | up | 1,6709594 | NPC1 | Homo sapiens Niemann-Pick disease, type C1 (NPC1), mRNA [NM_000271] |
| A_23_P146572 | 0,003155774 | up | 1,5817384 | NPDC1 | Homo sapiens neural proliferation, differentiation and control, 1 (NPDC1), mRNA [NM_015392] |
| A_23_P84836 | 6,00E-05 | up | 2,0663073 | NPEPPS | Homo sapiens aminopeptidase puromycin sensitive (NPEPPS), mRNA [NM_006310] |

| | | | | | |
|---|---|---|---|---|---|
| A_21_P0014949 | 0,002389173 | up | 1,8118553 | NPEPPS | Homo sapiens aminopeptidase puromycin sensitive (NPEPPS), mRNA [NM_006310] |
| A_23_P1452 | 0,024722338 | down | -1,7921013 | NPFFR1 | Homo sapiens neuropeptide FF receptor 1 (NPFFR1), mRNA [NM_022146] |
| A_33_P3372389 | 0,04861259 | down | -2,197123 | NPHP4 | Homo sapiens nephronophthisis 4 (NPHP4), transcript variant 1, mRNA [NM_015102] |
| A_33_P3212823 | 0,003022953 | down | -1,6415645 | NPIPB5 | Homo sapiens nuclear pore complex interacting protein family, member B5 (NPIPB5), mRNA [NM_001135865] |
| A_24_P204675 | 0,004452141 | up | 1,5356759 | NPIPB5 | Homo sapiens nuclear pore complex interacting protein family, member B5 (NPIPB5), mRNA [NM_001135865] |
| A_23_P339954 | 0,004380181 | up | 1,5442699 | NPIPB9 | Homo sapiens nuclear pore complex interacting protein family, member B9 (NPIPB9), transcript variant 2, mRNA [NM_001287251] |
| A_33_P3316456 | 0,01212238 | down | -2,190545 | NPL | Homo sapiens N-acetylneuraminate pyruvate lyase (dihydrodipicolinate synthase) (NPL), transcript variant 2, mRNA [NM_001200050] |
| A_21_P0000687 | 0,040265396 | up | 2,233231 | NPPA-AS1 | Homo sapiens NPPA antisense RNA 1 (NPPA-AS1), antisense RNA [NR_037806] |
| A_33_P3362008 | 0,01879841 | down | -1,5872176 | NPPB | Homo sapiens natriuretic peptide B (NPPB), mRNA [NM_002521] |
| A_23_P62752 | 0,015774854 | down | -2,422663 | NPPB | Homo sapiens natriuretic peptide B (NPPB), mRNA [NM_002521] |
| A_24_P95822 | 0,011762762 | down | -1,5874593 | NPTN | Homo sapiens neuroplastin (NPTN), transcript variant b, mRNA [NM_012428] |
| A_23_P82651 | 0,034062736 | down | -1,9872 | NPTX2 | Homo sapiens neuronal pentraxin II (NPTX2), mRNA [NM_002523] |
| A_23_P29282 | 1,67E-04 | down | -3,4055805 | NPTXR | Homo sapiens neuronal pentraxin receptor (NPTXR), mRNA [NM_014293] |
| A_23_P69699 | 0,001302117 | up | 1,8257021 | NPY1R | Homo sapiens neuropeptide Y receptor Y1 (NPY1R), mRNA [NM_000909] |
| A_24_P364042 | 0,00744355 | down | -1,8327472 | NPY4R | Homo sapiens neuropeptide Y receptor Y4 (NPY4R), transcript variant 1, mRNA [NM_005972] |
| A_24_P411573 | 0,001925317 | down | -3,0978665 | NPY6R | Homo sapiens neuropeptide Y receptor Y6 (pseudogene) (NPY6R), non-coding RNA [NR_002713] |
| A_23_P160800 | 0,003145441 | down | -3,083117 | NR0B2 | Homo sapiens nuclear receptor subfamily 0, group B, member 2 (NR0B2), mRNA [NM_021969] |
| A_24_P943472 | 0,02581247 | up | 1,6233679 | NR1D2 | Homo sapiens nuclear receptor subfamily 1, group D, member 2 (NR1D2), transcript variant 1, mRNA [NM_005126] |
| A_23_P25396 | 0,013305876 | up | 2,1403549 | NR1H4 | Homo sapiens nuclear receptor subfamily 1, group H, member 4 (NR1H4), transcript variant 2, mRNA [NM_005123] |
| A_23_P205867 | 0,03358383 | up | 1,9315988 | NR2E3 | Homo sapiens nuclear receptor subfamily 2, group E, member 3 (NR2E3), transcript variant 2, mRNA [NM_014249] |
| A_33_P3421318 | 0,007383967 | down | -2,06435 | NR2F2-AS1 | Homo sapiens NR2F2 antisense RNA 1 (NR2F2-AS1), transcript variant 3, long non-coding RNA [NR_125738] |
| A_33_P3386150 | 0,010957693 | up | 2,0393243 | NRADDP | Homo sapiens neurotrophin receptor associated death domain, pseudogene (NRADDP), non-coding RNA [NR_024046] |
| A_24_P242132 | 0,030017272 | up | 1,6964096 | NRBP2 | Homo sapiens nuclear receptor binding protein 2 (NRBP2), mRNA [NM_178564] |
| A_33_P3301514 | 0,03276224 | up | 2,2194393 | NRCAM | Homo sapiens neuronal cell adhesion molecule (NRCAM), transcript variant 4, mRNA [NM_001193582] |
| A_24_P252364 | 0,001700091 | up | 7,682432 | NRCAM | Homo sapiens neuronal cell adhesion molecule (NRCAM), transcript variant 1, mRNA [NM_001037132] |
| A_33_P3234048 | 0,004813931 | up | 2,5418632 | NRDE2 | NRDE-2, necessary for RNA interference, domain containing [Source:HGNC Symbol;Acc:HGNC:20186] [ENST00000354366] |
| A_23_P213699 | 0,048292883 | up | 1,7348461 | NRG2 | Homo sapiens neuregulin 2 (NRG2), transcript variant 3, mRNA [NM_013982] |
| A_23_P334727 | 0,001957601 | down | -1,9204177 | NRG4 | Homo sapiens neuregulin 4 (NRG4), mRNA [NM_138573] |
| A_23_P211007 | 0,00452449 | up | 1,6498637 | NRIP1 | Homo sapiens nuclear receptor interacting protein 1 (NRIP1), mRNA [NM_003489] |
| A_21_P0002006 | 1,22E-04 | up | 2,4948225 | NRIR | Homo sapiens negative regulator of interferon response (non-protein coding) (NRIR), long non-coding RNA [NR_126359] |
| A_33_P3338631 | 9,95E-04 | down | -1,9091784 | NRL | Homo sapiens neural retina leucine zipper (NRL), mRNA [NM_006177] |
| A_23_P132856 | 1,19E-04 | down | -2,7387319 | NRROS | Homo sapiens negative regulator of reactive oxygen species (NRROS), mRNA [NM_198565] |
| A_33_P3252221 | 0,011974739 | up | 1,5278227 | NRSN1 | Homo sapiens neurensin 1 (NRSN1), mRNA [NM_080723] |
| A_33_P3325275 | 0,001726396 | down | -2,093054 | NRSN2 | Homo sapiens neurensin 2 (NRSN2), mRNA [NM_024958] |
| A_33_P3273669 | 7,58E-04 | down | -2,451953 | NRXN1 | Homo sapiens neurexin 1 (NRXN1), transcript variant alpha1, mRNA [NM_004801] |
| A_33_P3273684 | 0,037389204 | up | 2,021645 | NRXN1 | Homo sapiens neurexin 1 (NRXN1), transcript variant alpha2, mRNA [NM_001135659] |
| A_24_P918384 | 0,002243104 | up | 2,7493532 | NSD1 | Homo sapiens nuclear receptor binding SET domain protein 1 (NSD1), transcript variant 2, mRNA [NM_022455] |
| A_33_P3325087 | 3,66E-04 | up | 1,7453665 | NSRP1 | Homo sapiens nuclear speckle splicing regulatory protein 1 (NSRP1), transcript variant 1, mRNA [NM_032141] |
| A_23_P251505 | 9,84E-04 | up | 1,5778569 | NSUN5 | Homo sapiens NOP2/Sun domain family, member 5 (NSUN5), transcript variant 1, mRNA [NM_148956] |
| A_33_P3271261 | 1,62E-04 | down | -3,1890185 | NTN3 | Homo sapiens netrin 3 (NTN3), mRNA [NM_006181] |
| A_33_P3221414 | 0,029822819 | down | -2,901232 | NTNG1 | netrin G1 [Source:HGNC Symbol;Acc:HGNC:23319] [ENST00000370068] |
| A_33_P3277784 | 0,002766848 | up | 2,088362 | NTNG2 | netrin G2 [Source:HGNC Symbol;Acc:HGNC:14288] [ENST00000372179] |
| A_24_P265506 | 0,012382316 | down | -2,2533813 | NTRK1 | Homo sapiens neurotrophic tyrosine kinase, receptor, type 1 (NTRK1), transcript variant 2, mRNA [NM_002529] |
| A_33_P3322784 | 0,003157703 | up | 1,5411966 | NTRK2 | Homo sapiens neurotrophic tyrosine kinase, receptor, type 2 (NTRK2), transcript variant d, mRNA [NM_001018065] |
| A_33_P3260134 | 0,001824246 | down | -2,3722246 | NTRK3 | Homo sapiens neurotrophic tyrosine kinase, receptor, type 3 (NTRK3), transcript variant 2, mRNA [NM_002530] |
| A_23_P371039 | 0,02495735 | down | -2,355455 | NTSR1 | Homo sapiens neurotensin receptor 1 (high affinity) (NTSR1), mRNA [NM_002531] |
| A_24_P145122 | 0,00261807 | down | -1,5965099 | NUCKS1 | Homo sapiens nuclear casein kinase and cyclin-dependent kinase substrate 1 (NUCKS1), mRNA [NM_022731] |
| A_24_P626981 | 0,031781968 | up | 1,5822601 | NUDT16P1 | Homo sapiens nudix (nucleoside diphosphate linked moiety X)-type motif 16 pseudogene 1 (NUDT16P1), transcript variant 2, non-coding RNA [NR_002949] |
| A_33_P3336810 | 0,005153067 | up | 1,6095084 | NUDT17 | Homo sapiens nudix (nucleoside diphosphate linked moiety X)-type motif 17 (NUDT17), mRNA [NM_001012758] |
| A_24_P50753 | 0,024144895 | down | -1,6164465 | NUDT4 | Homo sapiens nudix (nucleoside diphosphate linked moiety X)-type motif 4 (NUDT4), transcript variant 2, mRNA [NM_199040] |
| A_33_P3416503 | 0,011599925 | down | -1,7402624 | NUDT8 | Homo sapiens nudix (nucleoside diphosphate linked moiety X)-type motif 8 (NUDT8), transcript variant 2, mRNA [NM_181843] |
| A_24_P791829 | 6,45E-04 | up | 1,6766021 | NUFIP2 | Homo sapiens nuclear fragile X mental retardation protein interacting protein 2 (NUFIP2), mRNA [NM_020772] |
| A_33_P3246598 | 0,019162731 | down | -2,739721 | NUGGC | Homo sapiens nuclear GTPase, germinal center associated (NUGGC), mRNA [NM_001010906] |
| A_33_P3246593 | 0,005528355 | up | 3,158021 | NUGGC | Homo sapiens nuclear GTPase, germinal center associated (NUGGC), mRNA [NM_001010906] |
| A_23_P43726 | 0,00478638 | up | 1,5267423 | NUP160 | Homo sapiens nucleoporin 160kDa (NUP160), mRNA [NM_015231] |
| A_23_P211627 | 0,024939364 | up | 1,5134053 | NUP50 | Homo sapiens nucleoporin 50kDa (NUP50), transcript variant 2, mRNA [NM_007172] |
| A_23_P26777 | 0,001040732 | up | 1,6789508 | NUP85 | Homo sapiens nucleoporin 85kDa (NUP85), transcript variant 1, mRNA [NM_024844] |
| A_21_P0011128 | 0,005162046 | down | -1,6331309 | NUS1 | Homo sapiens nuclear undecaprenyl pyrophosphate synthase 1 homolog (S. cerevisiae) (NUS1), mRNA [NM_138459] |
| A_23_P93082 | 0,010448776 | down | -1,5846817 | NUS1 | Homo sapiens nuclear undecaprenyl pyrophosphate synthase 1 homolog (S. cerevisiae) (NUS1), mRNA [NM_138459] |
| A_33_P3311353 | 0,03827941 | up | 2,0389698 | NUTM1 | Homo sapiens NUT midline carcinoma, family member 1 (NUTM1), transcript variant 3, mRNA [NM_175741] |
| A_21_P0010949 | 0,02935524 | up | 1,683138 | NUTM2B-AS1 | Homo sapiens NUTM2B antisense RNA 1 (NUTM2B-AS1), transcript variant 1, long non-coding RNA [NR_120611] |

| | | | | | |
|---|---|---|---|---|---|
| A_21_P0010892 | 0,015135157 | up | 1,7938089 | NUTM2B-AS1 | Homo sapiens NUTM2B antisense RNA 1 (NUTM2B-AS1), transcript variant 1, long non-coding RNA [NR_120611] |
| A_33_P3235098 | 0,014196672 | down | -2,679955 | NUTM2D | Homo sapiens NUT family member 2D (NUTM2D), non-coding RNA [NR_075100] |
| A_33_P3311994 | 0,001173437 | down | -1,8333664 | NWD2 | Homo sapiens NACHT and WD repeat domain containing 2 (NWD2), mRNA [NM_001144990] |
| A_23_P148568 | 0,037205 | down | -1,9863268 | NXF2 | Homo sapiens nuclear RNA export factor 2 (NXF2), mRNA [NM_022053] |
| A_23_P114353 | 6,13E-04 | down | -2,3162537 | NXF5 | Homo sapiens nuclear RNA export factor 5 (NXF5), transcript variant 1, mRNA [NM_032946] |
| A_33_P3335661 | 0,031974666 | up | 1,8642734 | NXPE2 | Homo sapiens neurexophilin and PC-esterase domain family, member 2 (NXPE2), mRNA [NM_182495] |
| A_24_P91916 | 0,026278896 | up | 1,6167156 | NXT2 | Homo sapiens nuclear transport factor 2-like export factor 2 (NXT2), transcript variant 1, mRNA [NM_018698] |
| A_23_P407654 | 0,003780992 | up | 1,7647666 | NYAP1 | Homo sapiens neuronal tyrosine-phosphorylated phosphoinositide-3-kinase adaptor 1 (NYAP1), mRNA [NM_173564] |
| A_33_P3382835 | 0,010550587 | down | -2,5840642 | NYNRIN | Homo sapiens NYN domain and retroviral integrase containing (NYNRIN), mRNA [NM_025081] |
| A_23_P139786 | 0,026100144 | up | 2,2244084 | OASL | Homo sapiens 2'-5'-oligoadenylate synthetase-like (OASL), transcript variant 1, mRNA [NM_003733] |
| A_24_P759674 | 0,016734585 | up | 1,606538 | OBFC1 | Homo sapiens oligonucleotide/oligosaccharide-binding fold containing 1 (OBFC1), mRNA [NM_024928] |
| A_33_P3350575 | 0,005921053 | down | -1,9476157 | OC90 | Homo sapiens otoconin 90 (OC90), mRNA [NM_001080399] |
| A_23_P129133 | 0,005745504 | up | 3,0998487 | OCA2 | Homo sapiens oculocutaneous albinism II (OCA2), transcript variant 1, mRNA [NM_000275] |
| A_23_P92672 | 0,027097465 | up | 2,2307148 | OCLN | Homo sapiens occludin (OCLN), transcript variant 1, mRNA [NM_002538] |
| A_24_P4736 | 0,011389444 | down | -2,1563425 | ODF3 | Homo sapiens outer dense fiber of sperm tails 3 (ODF3), transcript variant 1, mRNA [NM_053280] |
| A_24_P134653 | 0,010780783 | up | 1,6965927 | OFD1 | Homo sapiens oral-facial-digital syndrome 1 (OFD1), mRNA [NM_003611] |
| A_33_P3227284 | 0,001088555 | up | 1,7004446 | OGT | Homo sapiens O-linked N-acetylglucosamine (GlcNAc) transferase (OGT), transcript variant 1, mRNA [NM_181672] |
| A_21_P0008626 | 6,92E-04 | down | -1,6473643 | OIP5-AS1 | Homo sapiens OIP5 antisense RNA 1 (OIP5-AS1), long non-coding RNA [NR_026757] |
| A_23_P63777 | 0,006828985 | up | 1,613796 | OIT3 | Homo sapiens oncoprotein induced transcript 3 (OIT3), transcript variant 1, mRNA [NM_152635] |
| A_33_P3232269 | 0,02799962 | up | 1,6209966 | OK/SW-CL.36 | Homo sapiens mRNA for OK/SW-CL.36, complete cds. [AB064670] |
| A_33_P3383205 | 0,003990809 | down | -2,3293228 | OK/SW-CL.58 | Homo sapiens mRNA for OK/SW-CL.58, complete cds. [AB064667] |
| A_33_P3248354 | 0,00591839 | up | 2,42535 | OLFM1 | Homo sapiens olfactomedin 1 (OLFM1), transcript variant 2, mRNA [NM_006334] |
| A_33_P3248137 | 0,002558445 | down | -1,9415053 | OLFML2A | Homo sapiens olfactomedin-like 2A (OLFML2A), transcript variant 1, mRNA [NM_182487] |
| A_23_P137689 | 0,004640715 | down | -2,430831 | OLFML2B | Homo sapiens olfactomedin-like 2B (OLFML2B), transcript variant 2, mRNA [NM_015441] |
| A_32_P124245 | 0,030166123 | down | -1,9891382 | OLIG3 | Homo sapiens oligodendrocyte transcription factor 3 (OLIG3), mRNA [NM_175747] |
| A_32_P124708 | 0,005780972 | down | -2,8720434 | ONECUT2 | Homo sapiens one cut homeobox 2 (ONECUT2), mRNA [NM_004852] |
| A_33_P3238969 | 0,004369856 | down | -2,2201133 | ONECUT3 | Homo sapiens one cut homeobox 3 (ONECUT3), mRNA [NM_001080488] |
| A_23_P96623 | 0,012263366 | down | -2,3588285 | OPN1MW | Homo sapiens opsin 1 (cone pigments), medium-wave-sensitive (OPN1MW), mRNA [NM_000513] |
| A_23_P24234 | 0,002182704 | down | -3,3130405 | OPN4 | Homo sapiens opsin 4 (OPN4), transcript variant 2, mRNA [NM_001030015] |
| A_33_P3272075 | 0,005009085 | down | -2,2159388 | OPRD1 | Homo sapiens opioid receptor, delta 1 (OPRD1), mRNA [NM_000911] |
| A_23_P345564 | 0,010220286 | down | -2,2855544 | OPRL1 | Homo sapiens opiate receptor-like 1 (OPRL1), transcript variant 1, mRNA [NM_182647] |
| A_33_P3359647 | 9,37E-04 | down | -2,4119186 | OPRL1 | Homo sapiens opiate receptor-like 1 (OPRL1), transcript variant 1, mRNA [NM_182647] |
| A_23_P97096 | 0,01646894 | down | -2,0072184 | OPTC | Homo sapiens opticin (OPTC), mRNA [NM_014359] |
| A_33_P3285032 | 0,022763161 | up | 2,753948 | OR10A3 | Homo sapiens olfactory receptor, family 10, subfamily A, member 3 (OR10A3), mRNA [NM_001003745] |
| A_23_P315991 | 5,25E-04 | down | -3,4114103 | OR10A5 | Homo sapiens olfactory receptor, family 10, subfamily A, member 5 (OR10A5), mRNA [NM_178168] |
| A_33_P3391370 | 0,012181696 | up | 2,1010926 | OR10AG1 | Homo sapiens olfactory receptor, family 10, subfamily AG, member 1 (OR10AG1), mRNA [NM_001005491] |
| A_24_P60680 | 0,007067597 | down | -2,3801374 | OR10C1 | Homo sapiens olfactory receptor, family 10, subfamily C, member 1 (gene/pseudogene) (OR10C1), mRNA [NM_013941] |
| A_33_P3225507 | 0,032634925 | down | -1,7341892 | OR10G2 | Homo sapiens olfactory receptor, family 10, subfamily G, member 2 (OR10G2), mRNA [NM_001005466] |
| A_33_P3225378 | 0,041694187 | down | -1,6911439 | OR10G3 | Homo sapiens olfactory receptor, family 10, subfamily G, member 3 (OR10G3), mRNA [NM_001005465] |
| A_33_P3273906 | 0,001179225 | down | -4,756754 | OR10G4 | Homo sapiens olfactory receptor, family 10, subfamily G, member 4 (OR10G4), mRNA [NM_001004462] |
| A_23_P125204 | 0,001710379 | down | -1,7427746 | OR10G8 | Homo sapiens olfactory receptor, family 10, subfamily G, member 8 (OR10G8), mRNA [NM_001004464] |
| A_33_P3290792 | 7,95E-04 | down | -3,8022256 | OR10G9 | Homo sapiens olfactory receptor, family 10, subfamily G, member 9 (OR10G9), mRNA [NM_001001953] |
| A_23_P50799 | 0,001227054 | down | -2,0943136 | OR10H2 | Homo sapiens olfactory receptor, family 10, subfamily H, member 2 (OR10H2), mRNA [NM_013939] |
| A_33_P3381067 | 0,004953384 | up | 1,9446958 | OR10Q1 | Homo sapiens olfactory receptor, family 10, subfamily Q, member 1 (OR10Q1), mRNA [NM_001004471] |
| A_33_P3766667 | 0,03623942 | down | -2,179765 | OR10R3P | Homo sapiens cDNA FLJ16108 fis, clone THYMU2009425, moderately similar to OLFACTORY RECEPTOR-LIKE PROTEIN HGMP07J. [AK122667] |
| A_33_P3313250 | 0,04290828 | up | 1,866483 | OR10X1 | Homo sapiens olfactory receptor, family 10, subfamily X, member 1 (gene/pseudogene) (OR10X1), mRNA [NM_001004477] |
| A_23_P59099 | 7,95E-04 | down | -2,4706976 | OR11A1 | Homo sapiens olfactory receptor, family 11, subfamily A, member 1 (OR11A1), mRNA [NM_013937] |
| A_23_P169207 | 0,031254042 | down | -2,2509897 | OR13C4 | Homo sapiens olfactory receptor, family 13, subfamily C, member 4 (OR13C4), mRNA [NM_001001919] |
| A_33_P3266484 | 0,012560548 | up | 1,7373185 | OR13F1 | Homo sapiens olfactory receptor, family 13, subfamily F, member 1 (OR13F1), mRNA [NM_001004485] |
| A_33_P3266489 | 0,004778009 | down | -3,0014067 | OR13H1 | Homo sapiens olfactory receptor, family 13, subfamily H, member 1 (OR13H1), mRNA [NM_001004486] |
| A_33_P3345464 | 0,005645712 | up | 1,5151366 | OR14C36 | Homo sapiens olfactory receptor, family 14, subfamily C, member 36 (OR14C36), mRNA [NM_001001918] |
| A_23_P130281 | 1,06E-04 | down | -3,1400967 | OR1A2 | Homo sapiens olfactory receptor, family 1, subfamily A, member 2 (OR1A2), mRNA [NM_012352] |
| A_23_P382506 | 3,37E-04 | down | -2,6885407 | OR1F2P | Homo sapiens olfactory receptor, family 1, subfamily F, member 2 (OR1F2P), non-coding RNA [NR_002169] |
| A_33_P3338382 | 0,005240858 | down | -2,4790673 | OR1K1 | Homo sapiens olfactory receptor, family 1, subfamily K, member 1 (OR1K1), mRNA [NM_080859] |
| A_33_P3299619 | 0,013759104 | up | 3,1697783 | OR1L1 | Homo sapiens olfactory receptor, family 1, subfamily L, member 1 (OR1L1), mRNA [NM_001005236] |
| A_33_P3223739 | 0,001311063 | down | -2,2281668 | OR1N1 | Homo sapiens olfactory receptor, family 1, subfamily N, member 1 (OR1N1), mRNA [NM_012363] |
| A_24_P506816 | 0,037139498 | down | -2,056911 | OR2A20P | Homo sapiens olfactory receptor, family 2, subfamily A, member 20 pseudogene (OR2A20P), non-coding RNA [NR_002158] |
| A_24_P52293 | 0,005205235 | down | -2,048576 | OR2A25 | Homo sapiens olfactory receptor, family 2, subfamily A, member 25 (OR2A25), mRNA [NM_001004488] |
| A_33_P3423335 | 0,039884757 | up | 1,9075545 | OR2AE1 | Homo sapiens olfactory receptor, family 2, subfamily AE, member 1 (OR2AE1), mRNA [NM_001005276] |
| A_23_P36439 | 0,001674779 | down | -1,9889965 | OR2AP1 | Homo sapiens olfactory receptor, family 2, subfamily AP, member 1 (OR2AP1), mRNA [NM_001258285] |
| A_33_P3365621 | 0,016764833 | up | 2,057348 | OR2AT4 | Homo sapiens olfactory receptor, family 2, subfamily AT, member 4 (OR2AT4), mRNA [NM_001005285] |

| | | | | | |
|---|---|---|---|---|---|
| A_33_P3310377 | 0,04275748 | up | 1,6165267 | OR2B11 | Homo sapiens olfactory receptor, family 2, subfamily B, member 11 (OR2B11), mRNA [NM_001004492] |
| A_23_P111088 | 7,69E-05 | down | -2,227896 | OR2B2 | Homo sapiens olfactory receptor, family 2, subfamily B, member 2 (OR2B2), mRNA [NM_033057] |
| A_33_P3264193 | 0,010380799 | up | 2,694678 | OR2D2 | Homo sapiens olfactory receptor, family 2, subfamily D, member 2 (OR2D2), mRNA [NM_003700] |
| A_23_P8716 | 5,57E-05 | down | -3,4915853 | OR2F1 | Homo sapiens olfactory receptor, family 2, subfamily F, member 1 (gene/pseudogene) (OR2F1), mRNA [NM_012369] |
| A_33_P3267769 | 0,045258008 | up | 2,3939729 | OR2G3 | Homo sapiens olfactory receptor, family 2, subfamily G, member 3 (OR2G3), mRNA [NM_001001914] |
| A_33_P3365082 | 0,006239188 | up | 1,614535 | OR2G6 | Homo sapiens olfactory receptor, family 2, subfamily G, member 6 (OR2G6), mRNA [NM_001013355] |
| A_23_P111092 | 0,007200058 | down | -1,9487162 | OR2H1 | Homo sapiens olfactory receptor, family 2, subfamily H, member 1 (OR2H1), mRNA [NM_030883] |
| A_33_P3387781 | 0,009608569 | down | -1,6519259 | OR2L1P | Homo sapiens olfactory receptor, family 2, subfamily L, member 1 pseudogene (OR2L1P), non-coding RNA [NR_002145] |
| A_33_P3369979 | 1,72E-04 | down | -2,8982615 | OR2L2 | Homo sapiens olfactory receptor, family 2, subfamily L, member 2 (OR2L2), mRNA [NM_001004686] |
| A_33_P3331952 | 0,02389502 | down | -2,0886557 | OR2M3 | Homo sapiens olfactory receptor, family 2, subfamily M, member 3 (OR2M3), mRNA [NM_001004689] |
| A_33_P3331966 | 0,005469095 | up | 2,8720655 | OR2M4 | Homo sapiens olfactory receptor, family 2, subfamily M, member 4 (OR2M4), mRNA [NM_017504] |
| A_33_P3311046 | 9,18E-06 | down | -3,580687 | OR2M7 | Homo sapiens olfactory receptor, family 2, subfamily M, member 7 (OR2M7), mRNA [NM_001004691] |
| A_33_P3230886 | 0,002598262 | up | 1,7961943 | OR2T11 | Homo sapiens olfactory receptor, family 2, subfamily T, member 11 (gene/pseudogene) (OR2T11), mRNA [NM_001001964] |
| A_33_P3350086 | 0,020657936 | up | 2,1281207 | OR2T33 | Homo sapiens olfactory receptor, family 2, subfamily T, member 33 (OR2T33), mRNA [NM_001004695] |
| A_33_P3260667 | 0,013380126 | up | 2,1196475 | OR2T34 | Homo sapiens olfactory receptor, family 2, subfamily T, member 34 (OR2T34), mRNA [NM_001001821] |
| A_23_P107483 | 2,83E-04 | down | -5,1384115 | OR3A3 | Homo sapiens olfactory receptor, family 3, subfamily A, member 3 (OR3A3), mRNA [NM_012373] |
| A_33_P3282693 | 6,99E-04 | down | -1,8982828 | OR3A4P | Homo sapiens olfactory receptor, family 3, subfamily A, member 4 pseudogene (OR3A4P), non-coding RNA [NR_024128] |
| A_23_P139099 | 2,99E-04 | down | -5,8948903 | OR4A15 | Homo sapiens olfactory receptor, family 4, subfamily A, member 15 (OR4A15), mRNA [NM_001005275] |
| A_33_P3231715 | 0,017567094 | up | 1,5125734 | OR4A47 | Homo sapiens olfactory receptor, family 4, subfamily A, member 47 (OR4A47), mRNA [NM_001005512] |
| A_33_P3213144 | 0,048408054 | up | 1,8001727 | OR4C11 | Homo sapiens olfactory receptor, family 4, subfamily C, member 11 (OR4C11), mRNA [NM_001004700] |
| A_33_P3210637 | 0,04597922 | up | 2,1822824 | OR4C16 | Homo sapiens olfactory receptor, family 4, subfamily C, member 16 (gene/pseudogene) (OR4C16), mRNA [NM_001004701] |
| A_23_P2077 | 0,007677946 | down | -2,4164681 | OR4C3 | Homo sapiens olfactory receptor, family 4, subfamily C, member 3 (OR4C3), mRNA [NM_001004702] |
| A_33_P3239278 | 0,002466427 | down | -3,6244109 | OR4D10 | Homo sapiens olfactory receptor, family 4, subfamily D, member 10 (OR4D10), mRNA [NM_001004705] |
| A_33_P3420635 | 0,002055555 | down | -2,0595808 | OR4D6 | Homo sapiens olfactory receptor, family 4, subfamily D, member 6 (OR4D6), mRNA [NM_001004708] |
| A_33_P3420640 | 0,011772635 | up | 3,005202 | OR4D9 | Homo sapiens olfactory receptor, family 4, subfamily D, member 9 (OR4D9), mRNA [NM_001004711] |
| A_33_P3308142 | 8,31E-04 | down | -2,1169314 | OR4F13P | Homo sapiens olfactory receptor, family 4, subfamily F, member 13 pseudogene (OR4F13P), non-coding RNA [NR_046417] |
| A_33_P3290874 | 0,009642881 | up | 1,8540063 | OR4F15 | Homo sapiens olfactory receptor, family 4, subfamily F, member 15 (OR4F15), mRNA [NM_001001674] |
| A_33_P3218659 | 0,002312795 | up | 1,8027846 | OR4N2 | Homo sapiens olfactory receptor, family 4, subfamily N, member 2 (OR4N2), mRNA [NM_001004723] |
| A_33_P3312529 | 0,003014722 | up | 1,6246563 | OR4N3P | Homo sapiens olfactory receptor, family 4, subfamily N, member 3 pseudogene (OR4N3P), non-coding RNA [NR_028067] |
| A_33_P3315979 | 4,50E-04 | down | -2,681299 | OR4Q3 | Homo sapiens olfactory receptor, family 4, subfamily Q, member 3 (OR4Q3), mRNA [NM_172194] |
| A_23_P13479 | 1,88E-04 | down | -2,5835233 | OR4S1 | Homo sapiens olfactory receptor, family 4, subfamily S, member 1 (OR4S1), mRNA [NM_001004725] |
| A_33_P3210653 | 0,008885331 | up | 1,5214895 | OR51A4 | Homo sapiens olfactory receptor, family 51, subfamily A, member 4 (OR51A4), mRNA [NM_001005329] |
| A_23_P13455 | 0,003595567 | down | -3,2587981 | OR51A7 | Homo sapiens olfactory receptor, family 51, subfamily A, member 7 (OR51A7), mRNA [NM_001004749] |
| A_24_P58204 | 0,00119164 | down | -2,6878226 | OR51B5 | Homo sapiens olfactory receptor, family 51, subfamily B, member 5 (OR51B5), transcript variant 1, mRNA [NM_001005567] |
| A_23_P24898 | 6,97E-06 | down | -2,724301 | OR51D1 | Homo sapiens olfactory receptor, family 51, subfamily D, member 1 (OR51D1), mRNA [NM_001004751] |
| A_33_P3424507 | 1,02E-04 | down | -3,147332 | OR51F1 | Homo sapiens olfactory receptor, family 51, subfamily F, member 1 (gene/pseudogene) (OR51F1), mRNA [NM_001004752] |
| A_23_P105118 | 2,42E-04 | down | -2,3055463 | OR51G1 | Homo sapiens olfactory receptor, family 51, subfamily G, member 1 (gene/pseudogene) (OR51G1), mRNA [NM_001005237] |
| A_33_P3322769 | 0,025163736 | down | -2,3581405 | OR51G2 | Homo sapiens olfactory receptor, family 51, subfamily G, member 2 (OR51G2), mRNA [NM_001005238] |
| A_33_P3333648 | 7,76E-04 | down | -4,8825912 | OR51I2 | Homo sapiens olfactory receptor, family 51, subfamily I, member 2 (OR51I2), mRNA [NM_001004754] |
| A_33_P3208951 | 0,017625254 | up | 1,6265873 | OR51L1 | Homo sapiens olfactory receptor, family 51, subfamily L, member 1 (OR51L1), mRNA [NM_001004755] |
| A_23_P36203 | 0,003401211 | down | -2,4920852 | OR52B2 | Homo sapiens olfactory receptor, family 52, subfamily B, member 2 (OR52B2), mRNA [NM_001004052] |
| A_33_P3423375 | 0,031025419 | up | 2,1908667 | OR52B6 | Homo sapiens olfactory receptor, family 52, subfamily B, member 6 (OR52B6), mRNA [NM_001005162] |
| A_33_P3303905 | 0,029343242 | up | 2,6550999 | OR52E4 | Homo sapiens olfactory receptor, family 52, subfamily E, member 4 (OR52E4), mRNA [NM_001005165] |
| A_33_P3281990 | 0,018217295 | up | 2,9310203 | OR52E8 | Homo sapiens olfactory receptor, family 52, subfamily E, member 8 (OR52E8), mRNA [NM_001005168] |
| A_33_P3289401 | 0,025732711 | up | 2,242614 | OR52J3 | Homo sapiens olfactory receptor, family 52, subfamily J, member 3 (OR52J3), mRNA [NM_001001916] |
| A_23_P98744 | 0,007172618 | down | -1,6615777 | OR52K2 | Homo sapiens olfactory receptor, family 52, subfamily K, member 2 (OR52K2), mRNA [NM_001005172] |
| A_33_P3229042 | 0,001611285 | up | 1,8852211 | OR52N1 | Homo sapiens olfactory receptor, family 52, subfamily N, member 1 (OR52N1), mRNA [NM_001001913] |
| A_33_P3303945 | 0,012066483 | up | 1,5312277 | OR52N4 | Homo sapiens olfactory receptor, family 52, subfamily N, member 4 (gene/pseudogene) (OR52N4), mRNA [NM_001005175] |
| A_33_P3306342 | 0,005794936 | down | -2,6158733 | OR52N5 | Homo sapiens olfactory receptor, family 52, subfamily N, member 5 (OR52N5), mRNA [NM_001001922] |
| A_33_P3383246 | 0,001139946 | down | -1,9188237 | OR52R1 | Homo sapiens olfactory receptor, family 52, subfamily R, member 1 (gene/pseudogene) (OR52R1), mRNA [NM_001005177] |
| A_33_P3366584 | 6,72E-04 | down | -2,6991994 | OR56A4 | Homo sapiens olfactory receptor, family 56, subfamily A, member 4 (OR56A4), mRNA [NM_001005179] |
| A_23_P75764 | 1,33E-04 | down | -5,2734756 | OR5A1 | Homo sapiens olfactory receptor, family 5, subfamily A, member 1 (OR5A1), mRNA [NM_001004728] |
| A_33_P3389673 | 0,03434065 | up | 2,3966675 | OR5A2 | Homo sapiens olfactory receptor, family 5, subfamily A, member 2 (OR5A2), mRNA [NM_001001954] |
| A_33_P3389713 | 0,009704749 | up | 1,5092523 | OR5AR1 | Homo sapiens olfactory receptor, family 5, subfamily AR, member 1 (gene/pseudogene) (OR5AR1), mRNA [NM_001004730] |
| A_33_P3380452 | 0,018418264 | up | 2,2278075 | OR5B12 | Homo sapiens olfactory receptor, family 5, subfamily B, member 12 (OR5B12), mRNA [NM_001004733] |
| A_33_P3417985 | 0,007080638 | up | 2,1670299 | OR5B17 | Homo sapiens olfactory receptor, family 5, subfamily B, member 17 (OR5B17), mRNA [NM_001005489] |
| A_33_P3258602 | 6,69E-04 | up | 2,7529066 | OR5B2 | Homo sapiens olfactory receptor, family 5, subfamily B, member 2 (OR5B2), mRNA [NM_001005566] |
| A_33_P3313065 | 0,005894904 | down | -2,896391 | OR5C1 | Homo sapiens olfactory receptor, family 5, subfamily C, member 1 (OR5C1), mRNA [NM_001001923] |
| A_33_P3363949 | 0,003275975 | up | 1,5561773 | OR5D13 | Homo sapiens olfactory receptor, family 5, subfamily D, member 13 (gene/pseudogene) (OR5D13), mRNA [NM_001001967] |
| A_33_P3224535 | 0,001004434 | up | 2,5785334 | OR5D18 | Homo sapiens olfactory receptor, family 5, subfamily D, member 18 (OR5D18), mRNA [NM_001001952] |
| A_33_P3356557 | 0,019931592 | up | 1,792738 | OR5D18 | Homo sapiens olfactory receptor, family 5, subfamily D, member 18 (OR5D18), mRNA [NM_001001952] |

| | | | | | |
|---|---|---|---|---|---|
| A_23_P127684 | 9,19E-04 | down | -3,293139 | OR5F1 | Homo sapiens olfactory receptor, family 5, subfamily F, member 1 (OR5F1), mRNA [NM_003697] |
| A_33_P3242064 | 0,017754449 | up | 3,432592 | OR5J2 | Homo sapiens olfactory receptor, family 5, subfamily J, member 2 (OR5J2), mRNA [NM_001005492] |
| A_33_P3419551 | 0,009239809 | up | 2,369195 | OR5K1 | Homo sapiens olfactory receptor, family 5, subfamily K, member 1 (OR5K1), mRNA [NM_001004736] |
| A_33_P3343526 | 0,014641221 | up | 2,2335374 | OR5M9 | Homo sapiens olfactory receptor, family 5, subfamily M, member 9 (OR5M9), mRNA [NM_001004743] |
| A_33_P3271221 | 0,03979505 | up | 1,8698763 | OR5T1 | Homo sapiens olfactory receptor, family 5, subfamily T, member 1 (OR5T1), mRNA [NM_001004745] |
| A_33_P3271226 | 0,004299074 | up | 1,5457038 | OR5T3 | Homo sapiens olfactory receptor, family 5, subfamily T, member 3 (OR5T3), mRNA [NM_001004747] |
| A_23_P42241 | 0,005591505 | down | -2,303802 | OR5V1 | Homo sapiens olfactory receptor, family 5, subfamily V, member 1 (OR5V1), mRNA [NM_030876] |
| A_33_P3316470 | 0,02196035 | up | 1,5681694 | OR6B1 | Homo sapiens olfactory receptor, family 6, subfamily B, member 1 (OR6B1), mRNA [NM_001005281] |
| A_33_P3383271 | 0,031749576 | up | 1,7139796 | OR6C3 | Homo sapiens olfactory receptor, family 6, subfamily C, member 3 (OR6C3), mRNA [NM_054104] |
| A_33_P3400678 | 0,015540849 | up | 1,5931393 | OR6C4 | Homo sapiens olfactory receptor, family 6, subfamily C, member 4 (OR6C4), mRNA [NM_001005494] |
| A_33_P3271166 | 2,77E-04 | down | -3,511871 | OR6C74 | Homo sapiens olfactory receptor, family 6, subfamily C, member 74 (OR6C74), mRNA [NM_001005490] |
| A_33_P3330881 | 0,03026075 | up | 1,6353365 | OR6C75 | Homo sapiens olfactory receptor, family 6, subfamily C, member 75 (OR6C75), mRNA [NM_001005497] |
| A_33_P3283320 | 0,011310394 | up | 1,8658123 | OR6F1 | Homo sapiens olfactory receptor, family 6, subfamily F, member 1 (OR6F1), mRNA [NM_001005286] |
| A_23_P51761 | 0,013125476 | down | -2,3204024 | OR6K2 | Homo sapiens olfactory receptor, family 6, subfamily K, member 2 (OR6K2), mRNA [NM_001005279] |
| A_33_P3255706 | 0,038161643 | up | 2,404831 | OR6K6 | Homo sapiens olfactory receptor, family 6, subfamily K, member 6 (OR6K6), mRNA [NM_001005184] |
| A_23_P23502 | 0,008681828 | down | -2,1089737 | OR6N1 | Homo sapiens olfactory receptor, family 6, subfamily N, member 1 (OR6N1), mRNA [NM_001005185] |
| A_33_P3258206 | 0,008781507 | up | 2,601408 | OR6N2 | Homo sapiens olfactory receptor, family 6, subfamily N, member 2 (OR6N2), mRNA [NM_001005278] |
| A_33_P3342469 | 0,001189114 | down | -1,7908281 | OR6T1 | Homo sapiens olfactory receptor, family 6, subfamily T, member 1 (OR6T1), mRNA [NM_001005187] |
| A_33_P3265244 | 0,020334423 | down | -2,293603 | OR6V1 | Homo sapiens olfactory receptor, family 6, subfamily V, member 1 (OR6V1), mRNA [NM_001001667] |
| A_23_P123164 | 0,002433654 | down | -2,3287656 | OR6W1P | Homo sapiens olfactory receptor, family 6, subfamily W, member 1 pseudogene (OR6W1P), non-coding RNA [NR_002140] |
| A_23_P85963 | 1,32E-04 | down | -2,6328394 | OR6Y1 | Homo sapiens olfactory receptor, family 6, subfamily Y, member 1 (OR6Y1), mRNA [NM_001005189] |
| A_23_P28023 | 1,92E-04 | down | -2,8394368 | OR7A17 | Homo sapiens olfactory receptor, family 7, subfamily A, member 17 (OR7A17), mRNA [NM_030901] |
| A_33_P3867391 | 0,011385617 | down | -2,438094 | OR7E19P | AGENCOURT_6564958 NIH_MGC_119 Homo sapiens cDNA clone IMAGE:5743918 5', mRNA sequence [BM563292] |
| A_21_P0014676 | 1,85E-04 | down | -3,8823173 | OR7E47P | Homo sapiens olfactory receptor, family 7, subfamily E, member 47 pseudogene (OR7E47P), transcript variant 4, non-coding RNA [NR_120438] |
| A_23_P408930 | 0,020716727 | down | -2,4416728 | OR7E5P | Homo sapiens olfactory receptor, family 7, subfamily E, member 5 pseudogene (OR7E5P), non-coding RNA [NR_027688] |
| A_33_P3300610 | 0,020876633 | down | -2,1259708 | OR7G2 | Homo sapiens olfactory receptor, family 7, subfamily G, member 2 (OR7G2), mRNA [NM_001005193] |
| A_23_P141939 | 0,04465835 | down | -1,8934103 | OR7G3 | Homo sapiens olfactory receptor, family 7, subfamily G, member 3 (OR7G3), mRNA [NM_001001958] |
| A_33_P3345683 | 0,030609543 | up | 2,415741 | OR8B12 | Homo sapiens olfactory receptor, family 8, subfamily B, member 12 (OR8B12), mRNA [NM_001005195] |
| A_33_P3253574 | 0,04884437 | down | -2,0726106 | OR8B3 | Homo sapiens olfactory receptor, family 8, subfamily B, member 3 (OR8B3), mRNA [NM_001005467] |
| A_23_P127662 | 0,0286937 | down | -1,9781832 | OR8D1 | Homo sapiens olfactory receptor, family 8, subfamily D, member 1 (OR8D1), mRNA [NM_001002917] |
| A_23_P139050 | 8,67E-04 | down | -2,507722 | OR8D2 | Homo sapiens olfactory receptor, family 8, subfamily D, member 2 (gene/pseudogene) (OR8D2), mRNA [NM_001002918] |
| A_23_P75707 | 2,73E-04 | down | -2,9737904 | OR8H1 | Homo sapiens olfactory receptor, family 8, subfamily H, member 1 (OR8H1), mRNA [NM_001005199] |
| A_33_P3300864 | 0,038115393 | down | -1,9922559 | OR8H2 | Homo sapiens olfactory receptor, family 8, subfamily H, member 2 (OR8H2), mRNA [NM_001005200] |
| A_33_P3300858 | 0,008135446 | down | -2,3250535 | OR8H3 | Homo sapiens olfactory receptor, family 8, subfamily H, member 3 (OR8H3), mRNA [NM_001005201] |
| A_33_P3305388 | 0,032834895 | up | 1,7348851 | OR8K5 | Homo sapiens olfactory receptor, family 8, subfamily K, member 5 (OR8K5), mRNA [NM_001004058] |
| A_23_P24676 | 0,034402426 | down | -2,2296932 | OR8U1 | Homo sapiens olfactory receptor, family 8, subfamily U, member 1 (OR8U1), mRNA [NM_001005204] |
| A_33_P3361991 | 3,73E-04 | down | -3,465477 | OR9A2 | Homo sapiens olfactory receptor, family 9, subfamily A, member 2 (OR9A2), mRNA [NM_001001658] |
| A_33_P3348569 | 0,004469313 | up | 2,8144004 | OR9G4 | Homo sapiens olfactory receptor, family 9, subfamily G, member 4 (OR9G4), mRNA [NM_001005284] |
| A_24_P104091 | 0,004035441 | up | 1,5304418 | ORAOV1 | Homo sapiens oral cancer overexpressed 1 (ORAOV1), mRNA [NM_153451] |
| A_33_P3381410 | 0,001305408 | down | -1,6660612 | ORC2 | Homo sapiens origin recognition complex, subunit 2 (ORC2), transcript variant 1, mRNA [NM_006190] |
| A_23_P39616 | 0,002860244 | up | 1,5174649 | ORC2 | Homo sapiens origin recognition complex, subunit 2 (ORC2), transcript variant 1, mRNA [NM_006190] |
| A_23_P169494 | 0,012584661 | up | 1,8092697 | ORM1 | Homo sapiens orosomucoid 1 (ORM1), mRNA [NM_000607] |
| A_23_P129829 | 5,09E-04 | down | -1,7993774 | ORMDL3 | Homo sapiens ORMDL sphingolipid biosynthesis regulator 3 (ORMDL3), mRNA [NM_139280] |
| A_33_P3394972 | 0,005441499 | up | 1,8170481 | OSBPL5 | Homo sapiens oxysterol binding protein-like 5 (OSBPL5), transcript variant 1, mRNA [NM_020896] |
| A_23_P53081 | 0,002773081 | up | 1,6820652 | OSBPL5 | Homo sapiens oxysterol binding protein-like 5 (OSBPL5), transcript variant 1, mRNA [NM_020896] |
| A_33_P3255194 | 4,57E-04 | down | -1,957056 | OSBPL6 | Homo sapiens oxysterol binding protein-like 6 (OSBPL6), transcript variant 5, mRNA [NM_001201482] |
| A_33_P3277373 | 0,001996342 | down | -2,3197296 | OSM | Homo sapiens oncostatin M (OSM), mRNA [NM_020530] |
| A_24_P55496 | 0,004719114 | down | -1,7628931 | OSR2 | Homo sapiens odd-skipped related transciption factor 2 (OSR2), transcript variant 2, mRNA [NM_053001] |
| A_24_P317450 | 3,44E-04 | down | -2,3754592 | OSTCP1 | Homo sapiens oligosaccharyltransferase complex subunit pseudogene 1 (OSTCP1), non-coding RNA [NR_028496] |
| A_21_P0004602 | 0,023023602 | up | 1,7287875 | OSTM1-AS1 | OSTM1 antisense RNA 1 [Source:HGNC Symbol;Acc:HGNC:43666] [ENST00000441184] |
| A_23_P257355 | 0,014776409 | up | 2,547601 | OTC | Homo sapiens ornithine carbamoyltransferase (OTC), mRNA [NM_000531] |
| A_33_P3364308 | 3,83E-04 | down | -3,4251778 | OTOA | otoancorin [Source:HGNC Symbol;Acc:HGNC:16378] [ENST00000563871] |
| A_23_P254852 | 4,14E-04 | down | -1,9700263 | OTOF | Homo sapiens otoferlin (OTOF), transcript variant 1, mRNA [NM_194248] |
| A_33_P3326643 | 2,59E-04 | down | -2,3997157 | OTOG | Homo sapiens otogelin (OTOG), transcript variant 1, mRNA [NM_001277269] |
| A_21_P0011057 | 0,02961104 | down | -2,4803238 | OTOGL | Homo sapiens otogelin-like (OTOGL), mRNA [NM_173591] |
| A_23_P90997 | 0,001843449 | down | -2,459764 | OTOS | Homo sapiens otospiralin (OTOS), mRNA [NM_148961] |
| A_24_P402898 | 3,39E-05 | up | 2,095009 | OTUD4 | Homo sapiens OTU deubiquitinase 4 (OTUD4), transcript variant 3, mRNA [NM_001102653] |
| A_32_P169574 | 9,17E-04 | up | 1,7905633 | OTUD5 | Homo sapiens OTU deubiquitinase 5 (OTUD5), transcript variant 1, mRNA [NM_017602] |
| A_19_P00329048 | 0,041767642 | down | -1,9538356 | OTUD6B-AS1 | Homo sapiens OTUD6B antisense RNA 1 (head to head) (OTUD6B-AS1), transcript variant 1, long non-coding RNA [NR_110438] |
| A_21_P0011090 | 0,023074916 | up | 2,0532122 | OVOS2 | |
| A_33_P3447304 | 0,009017119 | up | 1,9566851 | OXR1 | Homo sapiens oxidation resistance 1 (OXR1), transcript variant 4, mRNA [NM_001198533] |

| | | | | | |
|---|---|---|---|---|---|
| A_23_P418413 | 0,010517987 | up | 1,7307795 | OXSR1 | Homo sapiens oxidative stress responsive 1 (OXSR1), mRNA [NM_005109] |
| A_24_P382579 | 0,004912084 | down | -1,5769534 | OXT | Homo sapiens oxytocin/neurophysin I prepropeptide (OXT), mRNA [NM_000915] |
| A_23_P372848 | 0,001458099 | down | -2,3753898 | P2RX1 | Homo sapiens purinergic receptor P2X, ligand gated ion channel, 1 (P2RX1), mRNA [NM_002558] |
| A_23_P124003 | 4,84E-06 | down | -2,092846 | P2RX2 | Homo sapiens purinergic receptor P2X, ligand gated ion channel, 2 (P2RX2), transcript variant 4, mRNA [NM_170683] |
| A_23_P127721 | 9,14E-04 | down | -1,910056 | P2RX3 | Homo sapiens purinergic receptor P2X, ligand gated ion channel, 3 (P2RX3), mRNA [NM_002559] |
| A_23_P346376 | 6,89E-04 | down | -2,7239776 | P2RY10 | Homo sapiens purinergic receptor P2Y, G-protein coupled, 10 (P2RY10), transcript variant 1, mRNA [NM_014499] |
| A_24_P165864 | 0,002766851 | up | 1,8962711 | P2RY14 | Homo sapiens purinergic receptor P2Y, G-protein coupled, 14 (P2RY14), transcript variant 2, mRNA [NM_014879] |
| A_23_P114307 | 0,003070912 | down | -1,7766659 | P2RY4 | Homo sapiens pyrimidinergic receptor P2Y, G-protein coupled, 4 (P2RY4), mRNA [NM_002565] |
| A_24_P340128 | 0,012922802 | up | 1,6336266 | P2RY8 | Homo sapiens purinergic receptor P2Y, G-protein coupled, 8 (P2RY8), mRNA [NM_178129] |
| A_23_P69179 | 0,033441156 | up | 1,9956447 | P3H2 | Homo sapiens prolyl 3-hydroxylase 2 (P3H2), transcript variant 1, mRNA [NM_018192] |
| A_21_P0003077 | 0,003273956 | up | 2,0127995 | P3H2-AS1 | PREDICTED: Homo sapiens uncharacterized LOC101929152 (LOC101929152), ncRNA [XR_241635] |
| A_24_P290286 | 0,035311103 | up | 2,0112612 | P4HA3 | Homo sapiens prolyl 4-hydroxylase, alpha polypeptide III (P4HA3), transcript variant 1, mRNA [NM_182904] |
| A_24_P827491 | 0,005337297 | down | -2,3202643 | PA2G4 | Homo sapiens proliferation-associated 2G4, 38kDa (PA2G4), mRNA [NM_006191] |
| A_33_P3413808 | 0,031272847 | down | -1,9377786 | PABPC1L | Homo sapiens poly(A) binding protein, cytoplasmic 1-like (PABPC1L), mRNA [NM_001124756] |
| A_24_P162454 | 0,004072938 | down | -2,3940876 | PABPC1L2B | Homo sapiens poly(A) binding protein, cytoplasmic 1-like 2B (PABPC1L2B), mRNA [NM_001042506] |
| A_23_P48307 | 0,014632347 | down | -2,675061 | PABPC3 | Homo sapiens poly(A) binding protein, cytoplasmic 3 (PABPC3), mRNA [NM_030979] |
| A_33_P3838128 | 0,013666823 | down | -2,1025033 | PABPC5-AS1 | 5000ECD06 Fetal Brain 18 Homo sapiens cDNA 5', mRNA sequence [DR033788] |
| A_23_P170901 | 0,00222703 | down | -2,7807186 | PACRG | Homo sapiens PARK2 co-regulated (PACRG), transcript variant 1, mRNA [NM_152410] |
| A_33_P3778618 | 0,004979041 | up | 1,7290627 | PACRG-AS1 | Homo sapiens PACRG antisense RNA 1 (PACRG-AS1), long non-coding RNA [NR_028390] |
| A_23_P126869 | 3,80E-04 | down | -2,579594 | PADI3 | Homo sapiens peptidyl arginine deiminase, type III (PADI3), mRNA [NM_016233] |
| A_21_P0011841 | 0,01970487 | up | 1,5961697 | PAFAH1B1 | Homo sapiens platelet-activating factor acetylhydrolase 1b, regulatory subunit 1 (45kDa) (PAFAH1B1), mRNA [NM_000430] |
| A_33_P3216890 | 0,026805284 | up | 1,8420718 | PAG1 | Homo sapiens phosphoprotein membrane anchor with glycosphingolipid microdomains 1 (PAG1), mRNA [NM_018440] |
| A_32_P109683 | 0,018066635 | up | 2,259293 | PAGE2B | Homo sapiens P antigen family, member 2B (PAGE2B), mRNA [NM_001015038] |
| A_24_P254506 | 0,03886721 | up | 1,9550252 | PAGE4 | Homo sapiens P antigen family, member 4 (prostate associated) (PAGE4), mRNA [NM_007003] |
| A_24_P344666 | 0,002971523 | up | 1,5864477 | PAK7 | Homo sapiens p21 protein (Cdc42/Rac)-activated kinase 7 (PAK7), transcript variant 1, mRNA [NM_020341] |
| A_23_P161481 | 6,34E-06 | down | -6,7687163 | PALD1 | Homo sapiens phosphatase domain containing, paladin 1 (PALD1), mRNA [NM_014431] |
| A_33_P3423984 | 0,002264323 | down | -2,8922403 | PALLD | palladin, cytoskeletal associated protein [Source:HGNC Symbol;Acc:HGNC:17068] [ENST00000508898] |
| A_33_P3258061 | 0,028902292 | down | -1,5411907 | PALM3 | Homo sapiens paralemmin 3 (PALM3), mRNA [NM_001145028] |
| A_23_P64689 | 0,015131655 | up | 1,6021582 | PAN2 | Homo sapiens PAN2 poly(A) specific ribonuclease subunit (PAN2), transcript variant 3, mRNA [NM_014871] |
| A_23_P385938 | 1,24E-05 | up | 1,9873462 | PAN3 | Homo sapiens PAN3 poly(A) specific ribonuclease subunit (PAN3), mRNA [NM_175854] |
| A_33_P3303380 | 8,17E-04 | down | -1,7011992 | PARD3 | par-3 family cell polarity regulator [Source:HGNC Symbol;Acc:HGNC:16051] [ENST00000374768] |
| A_21_P0014253 | 0,01991493 | down | -2,4691706 | PARD3-AS1 | Homo sapiens PARD3 antisense RNA 1 (PARD3-AS1), long non-coding RNA [NR_108043] |
| A_32_P205637 | 0,003827347 | up | 3,0286245 | PARD6B | Homo sapiens par-6 family cell polarity regulator beta (PARD6B), mRNA [NM_032521] |
| A_33_P3314974 | 6,12E-05 | down | -2,912085 | PARD6G-AS1 | Homo sapiens PARD6G antisense RNA 1 (PARD6G-AS1), transcript variant 1, long non-coding RNA [NR_028339] |
| A_23_P147465 | 0,005256948 | down | -1,9135871 | PARK2 | Homo sapiens parkin RBR E3 ubiquitin protein ligase (PARK2), transcript variant 1, mRNA [NM_004562] |
| A_24_P191781 | 0,036214277 | up | 2,062362 | PARM1 | Homo sapiens prostate androgen-regulated mucin-like protein 1 (PARM1), mRNA [NM_015393] |
| A_32_P144342 | 0,010089459 | down | -2,299694 | PARP4 | Homo sapiens poly (ADP-ribose) polymerase family, member 4 (PARP4), mRNA [NM_006437] |
| A_21_P0004005 | 0,04683815 | up | 1,6630085 | PART1 | Homo sapiens prostate androgen-regulated transcript 1 (non-protein coding) (PART1), transcript variant 3, long non-coding RNA [NR_028509] |
| A_33_P3413404 | 4,61E-05 | down | -4,5607347 | PARVB | parvin, beta [Source:HGNC Symbol;Acc:HGNC:14653] [ENST00000402876] |
| A_23_P250413 | 0,022017727 | down | -2,0139523 | PARVG | Homo sapiens parvin, gamma (PARVG), transcript variant 1, mRNA [NM_022141] |
| A_33_P3223338 | 0,0041816 | up | 1,592895 | PARVG | parvin, gamma [Source:HGNC Symbol;Acc:HGNC:14654] [ENST00000453888] |
| A_33_P3420904 | 0,041639313 | down | -1,7522591 | PATE3 | Homo sapiens prostate and testis expressed 3 (PATE3), mRNA [NM_001129883] |
| A_24_P211558 | 0,025770394 | down | -1,8822757 | PATZ1 | Homo sapiens POZ (BTB) and AT hook containing zinc finger 1 (PATZ1), transcript variant 4, mRNA [NM_032051] |
| A_23_P414211 | 0,036456045 | down | -1,7500744 | PAX4 | Homo sapiens paired box 4 (PAX4), mRNA [NM_006193] |
| A_23_P500985 | 2,95E-05 | down | -4,420688 | PAX7 | Homo sapiens paired box 7 (PAX7), transcript variant 2, mRNA [NM_013945] |
| A_33_P3291579 | 0,014668189 | up | 1,7677433 | PAX7 | Homo sapiens paired box 7 (PAX7), transcript variant 3, mRNA [NM_001135254] |
| A_33_P3231076 | 0,034398902 | down | -1,6347736 | PAXIP1-AS2 | Homo sapiens PAXIP1 antisense RNA 2 (PAXIP1-AS2), transcript variant 2, long non-coding RNA [NR_024477] |
| A_24_P312519 | 1,89E-04 | down | -3,0984168 | PBLD | Homo sapiens phenazine biosynthesis-like protein domain containing (PBLD), transcript variant 2, mRNA [NM_001033083] |
| A_23_P332260 | 0,005954117 | down | -3,610085 | PBOV1 | Homo sapiens prostate and breast cancer overexpressed 1 (PBOV1), mRNA [NM_021635] |
| A_24_P173823 | 0,030713713 | down | -1,8760438 | PBX1 | Homo sapiens pre-B-cell leukemia homeobox 1 (PBX1), transcript variant 1, mRNA [NM_002585] |
| A_23_P90419 | 0,029757544 | up | 1,8889253 | PBX4 | Homo sapiens pre-B-cell leukemia homeobox 4 (PBX4), transcript variant 1, mRNA [NM_025245] |
| A_24_P79808 | 0,045399994 | up | 1,6084005 | PBXIP1 | Homo sapiens pre-B-cell leukemia homeobox interacting protein 1 (PBXIP1), mRNA [NM_020524] |
| A_33_P3252381 | 0,002013713 | up | 1,669291 | PCA3 | Homo sapiens prostate cancer associated 3 (non-protein coding) (PCA3), long non-coding RNA [NR_015342] |
| A_33_P3219095 | 0,003217471 | up | 1,571677 | PCAT18 | Homo sapiens prostate cancer associated transcript 18 (non-protein coding) (PCAT18), long non-coding RNA [NR_024259] |
| A_19_P00320718 | 0,021550383 | down | -1,6819419 | PCAT19 | Homo sapiens prostate cancer associated transcript 19 (non-protein coding) (PCAT19), long non-coding RNA [NR_040109] |
| A_19_P00315824 | 0,003078699 | down | -2,0715327 | PCAT19 | Homo sapiens prostate cancer associated transcript 19 (non-protein coding) (PCAT19), long non-coding RNA [NR_040109] |
| A_19_P00317660 | 0,006214581 | down | -3,5947058 | PCAT19 | Homo sapiens prostate cancer associated transcript 19 (non-protein coding) (PCAT19), long non-coding RNA [NR_040109] |
| A_19_P00808044 | 0,005521487 | up | 1,7006618 | PCAT19 | Homo sapiens prostate cancer associated transcript 19 (non-protein coding) (PCAT19), long non-coding RNA [NR_040109] |
| A_21_P0008650 | 0,012084872 | up | 1,9711802 | PCAT29 | Homo sapiens prostate cancer associated transcript 29 (non-protein coding) (PCAT29), transcript variant 2, long non-coding RNA [NR_126438] |
| A_33_P3224362 | 0,008755856 | up | 2,0739584 | PCBP3 | Homo sapiens poly(rC) binding protein 3 (PCBP3), transcript variant 1, mRNA [NM_020528] |
| A_33_P3251647 | 0,014974359 | up | 1,8740684 | PCBP3-OT1 | Homo sapiens cDNA FLJ44028 fis, clone TESTI4027516. [AK126016] |

| | | | | | |
|---|---|---|---|---|---|
| A_33_P3403887 | 0,04875271 | up | 1,5811639 | PCDH11X | protocadherin 11 X-linked [Source:HGNC Symbol;Acc:HGNC:8656] [ENST00000361724] |
| A_23_P161331 | 0,031913448 | up | 1,7783332 | PCDH15 | Homo sapiens protocadherin-related 15 (PCDH15), transcript variant C, mRNA [NM_033056] |
| A_33_P3258244 | 0,020784264 | up | 1,520716 | PCDH15 | Homo sapiens protocadherin-related 15 (PCDH15), transcript variant K, mRNA [NM_001142771] |
| A_24_P111147 | 0,028626699 | up | 1,5527307 | PCDH15 | protocadherin-related 15 [Source:HGNC Symbol;Acc:HGNC:14674] [ENST00000373955] |
| A_33_P3225760 | 0,004955962 | up | 1,5228286 | PCDH18 | Homo sapiens protocadherin 18 (PCDH18), transcript variant 2, mRNA [NM_001300828] |
| A_33_P3250348 | 0,009171628 | up | 1,8104873 | PCDH18 | Homo sapiens protocadherin 18 (PCDH18), transcript variant 1, mRNA [NM_019035] |
| A_24_P419039 | 0,004960642 | down | -2,0291567 | PCDH19 | Homo sapiens protocadherin 19 (PCDH19), transcript variant 2, mRNA [NM_020766] |
| A_23_P204885 | 0,028828025 | up | 1,7656273 | PCDH20 | Homo sapiens protocadherin 20 (PCDH20), mRNA [NM_022843] |
| A_23_P420236 | 0,00430319 | up | 1,5252738 | PCDH9 | Homo sapiens protocadherin 9 (PCDH9), transcript variant 1, mRNA [NM_203487] |
| A_33_P3258546 | 1,88E-05 | down | -3,6776645 | PCDHA5 | Homo sapiens protocadherin alpha 5 (PCDHA5), transcript variant 2, mRNA [NM_031501] |
| A_33_P3250695 | 0,012835939 | up | 2,3197448 | PCDHA8 | Homo sapiens protocadherin alpha 8 (PCDHA8), transcript variant 2, mRNA [NM_031856] |
| A_32_P932089 | 0,004047099 | up | 1,5281701 | PCDHB19P | Homo sapiens protocadherin beta 19 pseudogene (PCDHB19P), non-coding RNA [NR_001282] |
| A_24_P251962 | 0,005186366 | down | -1,8046378 | PCDHB2 | Homo sapiens protocadherin beta 2 (PCDHB2), mRNA [NM_018936] |
| A_23_P81262 | 5,64E-04 | down | -2,4929066 | PCDHB4 | Homo sapiens protocadherin beta 4 (PCDHB4), mRNA [NM_018938] |
| A_23_P41599 | 0,001876269 | down | -2,7657032 | PCDHB8 | Homo sapiens protocadherin beta 8 (PCDHB8), mRNA [NM_019120] |
| A_24_P308851 | 0,008575876 | down | -1,5936902 | PCDHGA7 | Homo sapiens protocadherin gamma subfamily A, 7 (PCDHGA7), transcript variant 2, mRNA [NM_032087] |
| A_23_P435444 | 0,001924096 | down | -1,5704226 | PCDHGA7 | Homo sapiens protocadherin gamma subfamily A, 7 (PCDHGA7), transcript variant 2, mRNA [NM_032087] |
| A_33_P3417611 | 4,27E-05 | down | -3,1261234 | PCDHGB1 | Homo sapiens protocadherin gamma subfamily B, 1 (PCDHGB1), transcript variant 2, mRNA [NM_032095] |
| A_33_P3410981 | 1,46E-04 | down | -1,8189934 | PCDHGB7 | Homo sapiens protocadherin gamma subfamily B, 7 (PCDHGB7), transcript variant 2, mRNA [NM_032101] |
| A_23_P303101 | 5,35E-04 | down | -3,676147 | PCDHGC4 | Homo sapiens protocadherin gamma subfamily C, 4 (PCDHGC4), transcript variant 2, mRNA [NM_032406] |
| A_23_P115703 | 0,001392821 | up | 1,6541017 | PCGF6 | Homo sapiens polycomb group ring finger 6 (PCGF6), transcript variant 1, mRNA [NM_001011663] |
| A_21_P0013977 | 8,71E-04 | up | 1,8891376 | PCID2 | PCI domain containing 2 [Source:HGNC Symbol;Acc:HGNC:25653] [ENST00000493650] |
| A_23_P210496 | 0,014724569 | down | -1,6505382 | PCIF1 | Homo sapiens PDX1 C-terminal inhibiting factor 1 (PCIF1), mRNA [NM_022104] |
| A_24_P247902 | 0,011761056 | down | -2,4332666 | PCLO | Homo sapiens piccolo presynaptic cytomatrix protein (PCLO), transcript variant 2, mRNA [NM_014510] |
| A_33_P3249529 | 7,27E-04 | down | -2,2273586 | PCNX | Homo sapiens pecanex homolog (Drosophila) (PCNX), mRNA [NM_014982] |
| A_23_P114585 | 0,023721797 | down | -2,0653741 | PCNXL2 | pecanex-like 2 (Drosophila) [Source:HGNC Symbol;Acc:HGNC:8736] [ENST00000324142] |
| A_21_P0000835 | 0,043082505 | up | 1,6880795 | PCOLCE-AS1 | Homo sapiens PCOLCE antisense RNA 1 (PCOLCE-AS1), long non-coding RNA [NR_038910] |
| A_24_P399500 | 0,014807835 | down | -1,5888704 | PCP2 | Homo sapiens Purkinje cell protein 2 (PCP2), transcript variant 1, mRNA [NM_174895] |
| A_23_P109322 | 0,013266861 | down | -2,1013622 | PCP4 | Homo sapiens Purkinje cell protein 4 (PCP4), mRNA [NM_006198] |
| A_33_P3358735 | 0,002008024 | up | 3,3392859 | PCSK5 | Homo sapiens proprotein convertase subtilisin/kexin type 5 (PCSK5), transcript variant 1, mRNA [NM_001190482] |
| A_21_P0011001 | 3,65E-04 | up | 1,8690726 | PCSK7 | Homo sapiens proprotein convertase subtilisin/kexin type 7 (PCSK7), mRNA [NM_004716] |
| A_21_P0013958 | 0,014385906 | down | -1,5298827 | PDCD6 | programmed cell death 6 [Source:HGNC Symbol;Acc:HGNC:8765] [ENST00000512466] |
| A_21_P0003089 | 0,01776953 | up | 1,5166916 | PDCD6 | Homo sapiens programmed cell death 6 (PDCD6), transcript variant 6, non-coding RNA [NR_073609] |
| A_33_P3258127 | 0,001475319 | down | -2,389518 | PDE10A | phosphodiesterase 10A [Source:HGNC Symbol;Acc:HGNC:8772] [ENST00000616273] |
| A_23_P356677 | 0,012655777 | up | 1,8258493 | PDE10A | Homo sapiens phosphodiesterase 10A (PDE10A), transcript variant 1, mRNA [NM_001130690] |
| A_23_P401106 | 0,010485435 | down | -2,174857 | PDE2A | Homo sapiens phosphodiesterase 2A, cGMP-stimulated (PDE2A), transcript variant 1, mRNA [NM_002599] |
| A_23_P208334 | 0,00983901 | up | 3,0731218 | PDE4A | Homo sapiens phosphodiesterase 4A, cAMP-specific (PDE4A), transcript variant 4, mRNA [NM_006202] |
| A_33_P3389658 | 0,03572454 | down | -1,835731 | PDE4D | Homo sapiens phosphodiesterase 4D, cAMP-specific (PDE4D), transcript variant 3, mRNA [NM_001165899] |
| A_33_P3240552 | 0,0343672 | up | 2,0277257 | PDE4D | phosphodiesterase 4D, cAMP-specific [Source:HGNC Symbol;Acc:HGNC:8783] [ENST00000509355] |
| A_33_P3372666 | 0,011251126 | down | -1,5109837 | PDGFA | Homo sapiens platelet-derived growth factor alpha polypeptide (PDGFA), transcript variant 2, mRNA [NM_033023] |
| A_24_P339944 | 0,048677154 | up | 2,206824 | PDGFB | Homo sapiens platelet-derived growth factor beta polypeptide (PDGFB), transcript variant 1, mRNA [NM_002608] |
| A_23_P250478 | 0,008366736 | down | -1,5788321 | PDK3 | Homo sapiens pyruvate dehydrogenase kinase, isozyme 3 (PDK3), transcript variant 2, mRNA [NM_005391] |
| A_33_P3419383 | 0,008829522 | up | 1,502299 | PDLIM3 | Homo sapiens PDZ and LIM domain 3 (PDLIM3), transcript variant 1, mRNA [NM_014476] |
| A_33_P3357964 | 0,013986516 | down | -1,7802666 | PDPK1 | Homo sapiens PRO0461 mRNA, complete cds. [AF111845] |
| A_24_P830690 | 5,89E-04 | up | 1,7382834 | PDPK1 | Homo sapiens 3-phosphoinositide dependent protein kinase 1 (PDPK1), transcript variant 1, mRNA [NM_002613] |
| A_33_P3319113 | 0,014818827 | down | -1,6750504 | PDPR | Homo sapiens pyruvate dehydrogenase phosphatase regulatory subunit (PDPR), mRNA [NM_017990] |
| A_33_P3311653 | 0,001040948 | up | 1,9118632 | PDS5A | PDS5, regulator of cohesion maintenance, homolog A (S. cerevisiae) [Source:HGNC Symbol;Acc:HGNC:29088] [ENST00000507766] |
| A_23_P161152 | 0,04840581 | up | 1,540339 | PDSS1 | Homo sapiens prenyl (decaprenyl) diphosphate synthase, subunit 1 (PDSS1), mRNA [NM_014317] |
| A_23_P134167 | 4,32E-04 | up | 1,7929859 | PDSS2 | Homo sapiens prenyl (decaprenyl) diphosphate synthase, subunit 2 (PDSS2), mRNA [NM_020381] |
| A_23_P393401 | 0,016259504 | up | 1,5272396 | PDXDC2P | Homo sapiens pyridoxal-dependent decarboxylase domain containing 2, pseudogene (PDXDC2P), non-coding RNA [NR_003610] |
| A_32_P216872 | 0,005942687 | up | 1,6885954 | PDXDC2P | Homo sapiens mRNA; cDNA DKFZp686B0962 (from clone DKFZp686B0962). [BX647358] |
| A_33_P3374035 | 0,030991394 | down | -2,1133242 | PDXK | Homo sapiens cDNA FLJ37311 fis, clone BRAMY2016771. [AK094630] |
| A_24_P279870 | 0,007958705 | up | 1,5233358 | PDYN | Homo sapiens prodynorphin (PDYN), transcript variant 1, mRNA [NM_024411] |
| A_23_P7402 | 0,006708093 | up | 2,3129182 | PDZD2 | Homo sapiens PDZ domain containing 2 (PDZD2), mRNA [NM_178140] |
| A_33_P3283515 | 0,027514 | up | 1,6639405 | PDZD2 | PDZ domain containing 2 [Source:HGNC Symbol;Acc:HGNC:18486] [ENST00000397559] |
| A_24_P265088 | 0,007923596 | down | -2,3056407 | PDZD4 | PDZ domain containing 4 [Source:HGNC Symbol;Acc:HGNC:21167] [ENST00000393758] |
| A_23_P412508 | 0,009846273 | up | 1,5751697 | PDZD9 | Homo sapiens PDZ domain containing 9 (PDZD9), transcript variant 1, mRNA [NM_173806] |
| A_23_P21618 | 0,012031486 | up | 2,1202831 | PDZRN3 | Homo sapiens PDZ domain containing ring finger 3 (PDZRN3), transcript variant 1, mRNA [NM_015009] |
| A_33_P3364211 | 0,027863368 | down | -1,9330724 | PDZRN3-AS1 | Homo sapiens cDNA FLJ44459 fis, clone UTERU2024656. [AK126423] |
| A_23_P252471 | 0,01806243 | up | 1,7748026 | PECAM1 | Homo sapiens platelet/endothelial cell adhesion molecule 1 (PECAM1), mRNA [NM_000442] |
| A_23_P82503 | 0,03747029 | down | -1,8863531 | PEG10 | Homo sapiens paternally expressed 10 (PEG10), transcript variant 1, mRNA [NM_001040152] |

| | | | | | |
|---|---|---|---|---|---|
| A_33_P3316928 | 0,034693643 | up | 1,5084666 | PELI1 | Homo sapiens pellino E3 ubiquitin protein ligase 1 (PELI1), mRNA [NM_020651] |
| A_23_P89589 | 3,76E-07 | up | 2,8826103 | PER1 | Homo sapiens period circadian clock 1 (PER1), mRNA [NM_002616] |
| A_24_P230948 | 0,04239668 | down | -1,7120575 | PER3 | Homo sapiens period circadian clock 3 (PER3), transcript variant 4, mRNA [NM_016831] |
| A_23_P501745 | 0,016275462 | down | -1,6327611 | PEX10 | Homo sapiens peroxisomal biogenesis factor 10 (PEX10), transcript variant 1, mRNA [NM_153818] |
| A_23_P402610 | 4,52E-06 | up | 3,0052128 | PFAS | Homo sapiens phosphoribosylformylglycinamidine synthase (PFAS), mRNA [NM_012393] |
| A_24_P413669 | 0,013960704 | up | 2,3087158 | PFKFB2 | Homo sapiens 6-phosphofructo-2-kinase/fructose-2,6-biphosphatase 2 (PFKFB2), transcript variant 2, mRNA [NM_001018053] |
| A_24_P261259 | 7,48E-05 | up | 2,1152768 | PFKFB3 | Homo sapiens 6-phosphofructo-2-kinase/fructose-2,6-biphosphatase 3 (PFKFB3), transcript variant 1, mRNA [NM_004566] |
| A_23_P29079 | 0,003608244 | down | -1,8552675 | PFKL | Homo sapiens phosphofructokinase, liver (PFKL), transcript variant 1, mRNA [NM_001002021] |
| A_33_P3413962 | 5,42E-04 | up | 2,0275688 | PFKP | Homo sapiens phosphofructokinase, platelet (PFKP), transcript variant 2, mRNA [NM_001242339] |
| A_23_P46928 | 0,008706077 | up | 1,5142171 | PFKP | Homo sapiens phosphofructokinase, platelet (PFKP), transcript variant 1, mRNA [NM_002627] |
| A_23_P350059 | 4,36E-05 | down | -2,3271964 | PFN1P2 | Homo sapiens profilin 1 pseudogene 2 (PFN1P2), non-coding RNA [NR_003242] |
| A_33_P3345743 | 6,16E-04 | up | 2,3182824 | PFN1P2 | Homo sapiens profilin 1 pseudogene 2 (PFN1P2), non-coding RNA [NR_003242] |
| A_23_P150549 | 0,007538144 | down | -2,4910462 | PGA3 | Homo sapiens pepsinogen 3, group I (pepsinogen A) (PGA3), mRNA [NM_001079807] |
| A_23_P79247 | 0,008119736 | down | -1,7394718 | PGAP1 | Homo sapiens post-GPI attachment to proteins 1 (PGAP1), mRNA [NM_024989] |
| A_23_P7965 | 0,002947436 | down | -2,1705506 | PGC | Homo sapiens progastricsin (pepsinogen C) (PGC), transcript variant 1, mRNA [NM_002630] |
| A_23_P208747 | 0,01340853 | up | 2,40533 | PGLYRP1 | Homo sapiens peptidoglycan recognition protein 1 (PGLYRP1), mRNA [NM_005091] |
| A_21_P0012083 | 0,004722489 | up | 1,83574 | PGM5P4-AS1 | Homo sapiens PGM5P4 antisense RNA 1 (PGM5P4-AS1), transcript variant 1, long non-coding RNA [NR_121185] |
| A_32_P3534 | 0,012823317 | down | -2,356194 | PGPEP1L | Homo sapiens pyroglutamyl-peptidase I-like (PGPEP1L), transcript variant 2, mRNA [NM_001167902] |
| A_24_P943566 | 0,027239794 | up | 2,0895507 | PHACTR1 | phosphatase and actin regulator 1 [Source:HGNC Symbol;Acc:HGNC:20990] [ENST00000379350] |
| A_23_P28834 | 3,16E-05 | up | 11,316884 | PHACTR3 | Homo sapiens phosphatase and actin regulator 3 (PHACTR3), transcript variant 1, mRNA [NM_080672] |
| A_33_P3289371 | 0,001038251 | up | 5,7130313 | PHACTR3 | phosphatase and actin regulator 3 [Source:HGNC Symbol;Acc:HGNC:15833] [ENST00000492611] |
| A_24_P119577 | 0,006047097 | up | 1,7302179 | PHACTR4 | Homo sapiens phosphatase and actin regulator 4 (PHACTR4), transcript variant 1, mRNA [NM_001048183] |
| A_23_P323094 | 4,07E-04 | up | 2,176607 | PHC1 | Homo sapiens polyhomeotic homolog 1 (Drosophila) (PHC1), mRNA [NM_004426] |
| A_23_P423864 | 0,004049486 | up | 1,7042192 | PHC2 | Homo sapiens polyhomeotic homolog 2 (Drosophila) (PHC2), transcript variant 1, mRNA [NM_198040] |
| A_23_P34510 | 0,041748386 | up | 1,549929 | PHC2 | Homo sapiens polyhomeotic homolog 2 (Drosophila) (PHC2), transcript variant 1, mRNA [NM_198040] |
| A_24_P535219 | 0,001341922 | up | 1,7995362 | PHF10 | PHD finger protein 10 [Source:HGNC Symbol;Acc:HGNC:18250] [ENST00000612128] |
| A_33_P3280666 | 0,031207943 | down | -1,5409517 | PHF21B | Homo sapiens PHD finger protein 21B (PHF21B), transcript variant 1, mRNA [NM_138415] |
| A_24_P5550 | 0,013216608 | up | 1,6276022 | PHF23 | Homo sapiens PHD finger protein 23 (PHF23), transcript variant 1, mRNA [NM_024297] |
| A_33_P3361925 | 0,001185814 | down | -1,9337939 | PHF8 | Homo sapiens PHD finger protein 8 (PHF8), transcript variant 4, mRNA [NM_001184898] |
| A_23_P85783 | 0,00733883 | down | -2,3864548 | PHGDH | Homo sapiens phosphoglycerate dehydrogenase (PHGDH), mRNA [NM_006623] |
| A_24_P238543 | 8,28E-04 | down | -1,7213258 | PHKA1 | Homo sapiens phosphorylase kinase, alpha 1 (muscle) (PHKA1), transcript variant 1, mRNA [NM_002637] |
| A_23_P159671 | 0,00226331 | up | 1,6186702 | PHKA2 | Homo sapiens phosphorylase kinase, alpha 2 (liver) (PHKA2), mRNA [NM_000292] |
| A_24_P296457 | 1,90E-04 | up | 2,1222544 | PHKG2 | Homo sapiens phosphorylase kinase, gamma 2 (testis) (PHKG2), transcript variant 1, mRNA [NM_000294] |
| A_33_P3311083 | 0,02253417 | up | 1,6500975 | PHLDB3 | pleckstrin homology-like domain, family B, member 3 [Source:HGNC Symbol;Acc:HGNC:30499] [ENST00000599242] |
| A_23_P89762 | 3,18E-04 | up | 2,2165082 | PHLPP1 | Homo sapiens PH domain and leucine rich repeat protein phosphatase 1 (PHLPP1), mRNA [NM_194449] |
| A_23_P418234 | 0,013928336 | up | 1,6650499 | PHLPP2 | Homo sapiens PH domain and leucine rich repeat protein phosphatase 2 (PHLPP2), transcript variant 1, mRNA [NM_015020] |
| A_33_P3353295 | 0,005745086 | up | 1,6120979 | PHOX2A | Homo sapiens paired-like homeobox 2a (PHOX2A), mRNA [NM_005169] |
| A_33_P3709173 | 0,005157729 | up | 1,5992016 | PHTF2 | Homo sapiens putative homeodomain transcription factor 2 (PHTF2), transcript variant 5, mRNA [NM_001127360] |
| A_21_P0012340 | 0,041691776 | up | 1,5147142 | PI4KA | Homo sapiens phosphatidylinositol 4-kinase, catalytic, alpha (PI4KA), mRNA [NM_058004] |
| A_24_P248185 | 9,65E-05 | down | -2,3683958 | PIANP | Homo sapiens PILR alpha associated neural protein (PIANP), transcript variant 1, mRNA [NM_153685] |
| A_24_P48627 | 0,017477155 | down | -2,518895 | PIAS4 | protein inhibitor of activated STAT, 4 [Source:HGNC Symbol;Acc:HGNC:17002] [ENST00000262971] |
| A_33_P3382157 | 0,002631512 | up | 1,7129724 | PICALM | Homo sapiens phosphatidylinositol binding clathrin assembly protein (PICALM), transcript variant 1, mRNA [NM_007166] |
| A_23_P130376 | 0,001480201 | down | -1,7383962 | PIEZO2 | Homo sapiens piezo-type mechanosensitive ion channel component 2 (PIEZO2), mRNA [NM_022068] |
| A_21_P0011651 | 0,021153457 | up | 2,3383312 | PIEZO2 | Homo sapiens piezo-type mechanosensitive ion channel component 2 (PIEZO2), mRNA [NM_022068] |
| A_33_P3351150 | 0,01559881 | up | 2,7126348 | PIEZO2 | Homo sapiens piezo-type mechanosensitive ion channel component 2 (PIEZO2), mRNA [NM_022068] |
| A_23_P424900 | 0,023848668 | down | -2,184422 | PIFO | Homo sapiens primary cilia formation (PIFO), transcript variant 1, mRNA [NM_181643] |
| A_23_P77174 | 0,005043936 | up | 1,6191792 | PIGB | Homo sapiens phosphatidylinositol glycan anchor biosynthesis, class B (PIGB), mRNA [NM_004855] |
| A_23_P136547 | 0,011616493 | up | 1,616075 | PIGG | Homo sapiens phosphatidylinositol glycan anchor biosynthesis, class G (PIGG), transcript variant 2, mRNA [NM_017733] |
| A_33_P3278058 | 0,025202228 | down | -2,7829816 | PIH1D2 | Homo sapiens PIH1 domain containing 2 (PIH1D2), transcript variant 1, mRNA [NM_138789] |
| A_33_P3419419 | 0,007618399 | down | -1,6212959 | PIK3C2B | phosphatidylinositol-4-phosphate 3-kinase, catalytic subunit type 2 beta [Source:HGNC Symbol;Acc:HGNC:8972] [ENST00000367184] |
| A_33_P3789056 | 0,001307792 | down | -1,6110413 | PIK3CD-AS1 | Homo sapiens PIK3CD antisense RNA 1 (PIK3CD-AS1), long non-coding RNA [NR_027045] |
| A_24_P29401 | 0,010532326 | up | 1,5958288 | PIK3R1 | Homo sapiens phosphoinositide-3-kinase, regulatory subunit 1 (alpha) (PIK3R1), transcript variant 1, mRNA [NM_181523] |
| A_24_P943957 | 4,65E-05 | up | 1,9155309 | PIKFYVE | Homo sapiens phosphoinositide kinase, FYVE finger containing (PIKFYVE), transcript variant 2, mRNA [NM_015040] |
| A_23_P345118 | 0,008579648 | up | 1,9835274 | PIM1 | Homo sapiens Pim-1 proto-oncogene, serine/threonine kinase (PIM1), transcript variant 1, mRNA [NM_002648] |
| A_23_P61398 | 0,010092692 | up | 1,5026766 | PIM3 | Homo sapiens Pim-3 proto-oncogene, serine/threonine kinase (PIM3), mRNA [NM_001001852] |
| A_23_P330262 | 0,002615545 | up | 1,8486665 | PIP5K1C | Homo sapiens phosphatidylinositol-4-phosphate 5-kinase, type I, gamma (PIP5K1C), transcript variant 2, mRNA [NM_012398] |
| A_23_P143662 | 2,76E-04 | up | 2,1444888 | PISD | Homo sapiens phosphatidylserine decarboxylase (PISD), mRNA [NM_014338] |
| A_21_P0000517 | 0,002071011 | up | 2,0599625 | PISRT1 | Homo sapiens polled intersex syndrome regulated transcript 1 (non-protein coding RNA) (PISRT1), long non-coding RNA [NR_027070] |
| A_23_P200073 | 0,036677163 | up | 1,540191 | PITHD1 | Homo sapiens PITH (C-terminal proteasome-interacting domain of thioredoxin-like) domain containing 1 (PITHD1), mRNA [NM_020362] |
| A_23_P401361 | 1,83E-04 | up | 1,9786284 | PITPNM2 | Homo sapiens phosphatidylinositol transfer protein, membrane-associated 2 (PITPNM2), transcript variant 1, mRNA [NM_020845] |
| A_33_P3342561 | 1,19E-05 | down | -2,7735965 | PITPNM3 | Homo sapiens PITPNM family member 3 (PITPNM3), transcript variant 1, mRNA [NM_031220] |

| | | | | | |
|---|---|---|---|---|---|
| A_33_P3240318 | 0,002864732 | down | -1,6037807 | PITX3 | Homo sapiens paired-like homeodomain 3 (PITX3), mRNA [NM_005029] |
| A_21_P0014687 | 0,007259573 | down | -1,5874139 | PKD1 | Homo sapiens polycystic kidney disease 1 (autosomal dominant) (PKD1), transcript variant 2, mRNA [NM_000296] |
| A_33_P3812815 | 6,62E-04 | down | -1,9907733 | PKD1 | Homo sapiens polycystic kidney disease 1 (autosomal dominant) (PKD1), transcript variant 2, mRNA [NM_000296] |
| A_21_P0011655 | 8,83E-04 | up | 1,7210083 | PKD1P6 | Homo sapiens polycystic kidney disease 1 (autosomal dominant) pseudogene 6 (PKD1P6), non-coding RNA [NR_123722] |
| A_33_P3247325 | 0,002702415 | up | 1,7831905 | PKD2L2 | Homo sapiens polycystic kidney disease 2-like 2 (PKD2L2), transcript variant 4, mRNA [NM_001300921] |
| A_33_P3387420 | 0,004623193 | up | 1,5654238 | PKHD1 | Homo sapiens polycystic kidney and hepatic disease 1 (autosomal recessive) (PKHD1), transcript variant 1, mRNA [NM_138694] |
| A_23_P402187 | 0,008243315 | up | 1,6691083 | PKHD1 | Homo sapiens polycystic kidney and hepatic disease 1 (autosomal recessive) (PKHD1), transcript variant 1, mRNA [NM_138694] |
| A_23_P31765 | 0,021783268 | down | -2,1397429 | PKIA | Homo sapiens protein kinase (cAMP-dependent, catalytic) inhibitor alpha (PKIA), transcript variant 1, mRNA [NM_006823] |
| A_33_P3310293 | 0,005859711 | down | -1,5357149 | PKIG | Homo sapiens protein kinase (cAMP-dependent, catalytic) inhibitor gamma (PKIG), transcript variant 1, mRNA [NM_181805] |
| A_24_P387869 | 0,002091981 | up | 1,5989832 | PKN2 | Homo sapiens protein kinase N2 (PKN2), mRNA [NM_006256] |
| A_33_P3242829 | 2,41E-04 | up | 2,6164134 | PKN2 | protein kinase N2 [Source:HGNC Symbol;Acc:HGNC:9406] [ENST00000370505] |
| A_33_P3220202 | 0,001581906 | up | 1,8978763 | PKN3 | Homo sapiens protein kinase N3 (PKN3), mRNA [NM_013355] |
| A_33_P3405728 | 0,024912672 | up | 1,5251138 | PKP2 | Homo sapiens plakophilin 2 (PKP2), transcript variant 2b, mRNA [NM_004572] |
| A_21_P0011456 | 0,005465929 | down | -1,5533979 | PLA2G10 | Homo sapiens phospholipase A2, group X (PLA2G10), mRNA [NM_003561] |
| A_23_P388150 | 0,006930857 | up | 2,5955443 | PLA2G12B | Homo sapiens phospholipase A2, group XIIB (PLA2G12B), mRNA [NM_032562] |
| A_33_P3350423 | 0,018960243 | up | 1,6337966 | PLA2G2E | Homo sapiens phospholipase A2, group IIE (PLA2G2E), mRNA [NM_014589] |
| A_33_P3241316 | 0,034178782 | down | -2,097993 | PLA2G4F | Homo sapiens phospholipase A2, group IVF (PLA2G4F), transcript variant 1, mRNA [NM_213600] |
| A_23_P103465 | 0,008002033 | down | -1,8436462 | PLA2G5 | Homo sapiens phospholipase A2, group V (PLA2G5), mRNA [NM_000929] |
| A_24_P78161 | 0,014255478 | up | 1,5105289 | PLAA | Homo sapiens phospholipase A2-activating protein (PLAA), mRNA [NM_001031689] |
| A_33_P3282162 | 0,001037008 | down | -2,8853543 | PLAC4 | Homo sapiens placenta-specific 4 (PLAC4), mRNA [NM_182832] |
| A_33_P3241989 | 0,015413621 | up | 1,7271894 | PLAC8 | Homo sapiens placenta-specific 8 (PLAC8), transcript variant 3, mRNA [NM_001130715] |
| A_33_P3252785 | 0,03483657 | down | -1,8490459 | PLAC9 | placenta-specific 9 [Source:HGNC Symbol;Acc:HGNC:19255] [ENST00000372263] |
| A_33_P3252781 | 0,006999389 | up | 1,5099521 | PLAC9 | Homo sapiens placenta-specific 9 (PLAC9), mRNA [NM_001012973] |
| A_33_P3335825 | 0,006159968 | up | 3,4049203 | PLCD4 | phospholipase C, delta 4 [Source:HGNC Symbol;Acc:HGNC:9062] [ENST00000483685] |
| A_23_P35617 | 0,04928663 | up | 2,0297616 | PLCE1 | Homo sapiens phospholipase C, epsilon 1 (PLCE1), transcript variant 1, mRNA [NM_016341] |
| A_33_P3327041 | 0,00337179 | up | 1,643299 | PLCE1-AS1 | Homo sapiens PLCE1 antisense RNA 1 (PLCE1-AS1), long non-coding RNA [NR_033969] |
| A_23_P254801 | 6,01E-04 | up | 2,3588884 | PLCG1 | Homo sapiens phospholipase C, gamma 1 (PLCG1), transcript variant 1, mRNA [NM_002660] |
| A_33_P3334180 | 9,84E-05 | down | -2,8763728 | PLCH2 | phospholipase C, eta 2 [Source:HGNC Symbol;Acc:HGNC:29037] [ENST00000473964] |
| A_33_P3214339 | 0,001968356 | down | -2,0089598 | PLCXD2 | Homo sapiens phosphatidylinositol-specific phospholipase C, X domain containing 2 (PLCXD2), transcript variant 2, mRNA [NM_153268] |
| A_23_P88222 | 0,005266823 | down | -2,2301295 | PLD4 | Homo sapiens phospholipase D family, member 4 (PLD4), mRNA [NM_138790] |
| A_23_P510 | 6,25E-04 | up | 3,4387665 | PLD5 | Homo sapiens phospholipase D family, member 5 (PLD5), transcript variant 1, mRNA [NM_152666] |
| A_21_P0010918 | 0,025708783 | up | 2,9363875 | PLD5 | Homo sapiens phospholipase D family, member 5 (PLD5), transcript variant 1, mRNA [NM_152666] |
| A_33_P3339860 | 0,010629199 | up | 1,77938 | PLEKHA2 | Homo sapiens pleckstrin homology domain containing, family A (phosphoinositide binding specific) member 2 (PLEKHA2), mRNA [NM_021623] |
| A_23_P218068 | 1,26E-04 | up | 1,9405653 | PLEKHA5 | Homo sapiens pleckstrin homology domain containing, family A member 5 (PLEKHA5), transcript variant 1, mRNA [NM_019012] |
| A_23_P431268 | 0,011251616 | up | 2,27729 | PLEKHA6 | Homo sapiens pleckstrin homology domain containing, family A member 6 (PLEKHA6), mRNA [NM_014935] |
| A_23_P28638 | 0,01534733 | up | 1,548485 | PLEKHB2 | Homo sapiens pleckstrin homology domain containing, family B (evectins) member 2 (PLEKHB2), transcript variant 2, mRNA [NM_017958] |
| A_33_P3353767 | 0,013383596 | up | 2,037659 | PLEKHD1 | Homo sapiens pleckstrin homology domain containing, family D (with coiled-coil domains) member 1 (PLEKHD1), mRNA [NM_001161498] |
| A_33_P3367471 | 0,004000416 | down | -1,748492 | PLEKHG1 | Homo sapiens pleckstrin homology domain containing, family G (with RhoGef domain) member 1 (PLEKHG1), mRNA [NM_001029884] |
| A_23_P323563 | 0,002078316 | up | 1,650355 | PLEKHG2 | Homo sapiens pleckstrin homology domain containing, family G (with RhoGef domain) member 2 (PLEKHG2), mRNA [NM_022835] |
| A_33_P3649715 | 0,012625907 | up | 1,5498322 | PLEKHG4B | Homo sapiens pleckstrin homology domain containing, family G (with RhoGef domain) member 4B (PLEKHG4B), mRNA [NM_052909] |
| A_23_P152727 | 0,009613545 | up | 1,5345246 | PLEKHM1 | Homo sapiens pleckstrin homology domain containing, family M (with RUN domain) member 1 (PLEKHM1), transcript variant 1, mRNA [NM_014798] |
| A_33_P3317048 | 0,004779528 | down | -1,9493797 | PLEKHM3 | Homo sapiens pleckstrin homology domain containing, family M, member 3 (PLEKHM3), mRNA [NM_001080475] |
| A_21_P0001706 | 0,012000951 | down | -1,6260571 | PLEKHO1 | pleckstrin homology domain containing, family O member 1 [Source:HGNC Symbol;Acc:HGNC:24310] [ENST00000502767] |
| A_33_P3412463 | 0,006222635 | down | -2,668515 | PLEKHS1 | Homo sapiens pleckstrin homology domain containing, family S member 1 (PLEKHS1), transcript variant 2, mRNA [NM_001193434] |
| A_24_P286951 | 0,007781419 | up | 3,3770964 | PLEKHS1 | Homo sapiens pleckstrin homology domain containing, family S member 1 (PLEKHS1), transcript variant 4, mRNA [NM_024889] |
| A_33_P3309832 | 0,032971818 | up | 1,8836011 | PLEKHS1 | Homo sapiens pleckstrin homology domain containing, family S member 1 (PLEKHS1), transcript variant 1, mRNA [NM_182601] |
| A_23_P26154 | 0,013356475 | up | 2,4942176 | PLIN1 | Homo sapiens perilipin 1 (PLIN1), transcript variant 1, mRNA [NM_002666] |
| A_21_P0014779 | 0,012827786 | up | 2,008604 | PLIN2 | perilipin 2 [Source:HGNC Symbol;Acc:HGNC:248] [ENST00000494753] |
| A_33_P3358824 | 0,03382074 | down | -2,5166667 | PLOD1 | Homo sapiens procollagen-lysine, 2-oxoglutarate 5-dioxygenase 1 (PLOD1), mRNA [NM_000302] |
| A_23_P211909 | 0,03317228 | up | 1,5309675 | PLS1 | Homo sapiens plastin 1 (PLS1), transcript variant 2, mRNA [NM_002670] |
| A_23_P250607 | 0,001490629 | up | 1,7490336 | PLS3 | Homo sapiens plastin 3 (PLS3), transcript variant 1, mRNA [NM_005032] |
| A_33_P3209885 | 0,033561006 | down | -1,6201117 | PLXDC1 | Homo sapiens plexin domain containing 1 (PLXDC1), mRNA [NM_020405] |
| A_23_P57667 | 0,0077576 | up | 1,6387016 | PLXNA1 | Homo sapiens plexin A1 (PLXNA1), mRNA [NM_032242] |
| A_33_P3413048 | 0,028804734 | up | 1,6144204 | PLXNA3 | Homo sapiens plexin A3 (PLXNA3), mRNA [NM_017514] |
| A_23_P414958 | 0,033463933 | up | 1,6636959 | PLXNC1 | Homo sapiens plexin C1 (PLXNC1), transcript variant 1, mRNA [NM_005761] |
| A_33_P3490572 | 0,003390943 | up | 1,5627593 | PM20D1 | Homo sapiens peptidase M20 domain containing 1 (PM20D1), mRNA [NM_152491] |
| A_32_P86118 | 0,012181427 | up | 1,5631951 | PM20D2 | Homo sapiens peptidase M20 domain containing 2 (PM20D2), mRNA [NM_001010853] |
| A_21_P0013389 | 0,01835113 | down | -1,6778427 | PMS2P9 | Homo sapiens postmeiotic segregation increased 2 pseudogene 9 (PMS2P9), non-coding RNA [NR_028058] |
| A_23_P377094 | 0,034863096 | up | 2,2095087 | PNMA3 | Homo sapiens paraneoplastic Ma antigen 3 (PNMA3), transcript variant 1, mRNA [NM_013364] |
| A_33_P3377851 | 9,90E-05 | down | -3,5596654 | PNMAL2 | Homo sapiens paraneoplastic Ma antigen family-like 2 (PNMAL2), mRNA [NM_020709] |
| A_24_P336853 | 0,012564305 | up | 1,5715804 | PNO1 | Homo sapiens partner of NOB1 homolog (S. cerevisiae) (PNO1), mRNA [NM_020143] |

| | | | | | |
|---|---|---|---|---|---|
| A_33_P3219870 | 0,007822102 | down | -1,6705767 | PNPLA2 | Homo sapiens patatin-like phospholipase domain containing 2 (PNPLA2), mRNA [NM_020376] |
| A_23_P17914 | 0,025899716 | up | 1,5280464 | PNPLA3 | Homo sapiens patatin-like phospholipase domain containing 3 (PNPLA3), mRNA [NM_025225] |
| A_33_P3559060 | 1,09E-04 | down | -3,2642949 | PNPLA5 | Homo sapiens patatin-like phospholipase domain containing 5 (PNPLA5), transcript variant 1, mRNA [NM_138814] |
| A_23_P212284 | 0,001113374 | down | -1,7656411 | POC1A | Homo sapiens POC1 centriolar protein A (POC1A), transcript variant 1, mRNA [NM_015426] |
| A_23_P368154 | 6,86E-04 | up | 2,2901745 | PODN | Homo sapiens podocan (PODN), transcript variant 1, mRNA [NM_153703] |
| A_23_P63219 | 0,001778208 | up | 1,5792227 | POGZ | Homo sapiens pogo transposable element with ZNF domain (POGZ), transcript variant 1, mRNA [NM_015100] |
| A_23_P500734 | 8,36E-05 | up | 2,0432944 | POGZ | Homo sapiens pogo transposable element with ZNF domain (POGZ), transcript variant 1, mRNA [NM_015100] |
| A_33_P3229177 | 0,003255644 | down | -1,5505521 | POLE4 | Homo sapiens polymerase (DNA-directed), epsilon 4, accessory subunit (POLE4), mRNA [NM_019896] |
| A_23_P26905 | 0,0107726 | up | 1,5457387 | POLG2 | Homo sapiens polymerase (DNA directed), gamma 2, accessory subunit (POLG2), mRNA [NM_007215] |
| A_23_P422718 | 9,05E-04 | down | -1,8322083 | POLH | Homo sapiens polymerase (DNA directed), eta (POLH), transcript variant 1, mRNA [NM_006502] |
| A_23_P306890 | 0,005746534 | up | 1,6262025 | POLI | Homo sapiens polymerase (DNA directed) iota (POLI), mRNA [NM_007195] |
| A_33_P3391097 | 0,005493749 | up | 2,3739965 | POLM | Homo sapiens cDNA FLJ35482 fis, clone SMINT2008133. [AK092801] |
| A_33_P3339109 | 0,003636692 | down | -1,6478926 | POLR1C | polymerase (RNA) I polypeptide C, 30kDa [Source:HGNC Symbol;Acc:HGNC:20194] [ENST00000512472] |
| A_33_P3383561 | 0,009334689 | up | 3,3181188 | POLR2F | polymerase (RNA) II (DNA directed) polypeptide F [Source:HGNC Symbol;Acc:HGNC:9193] [ENST00000443002] |
| A_33_P3396459 | 3,32E-07 | down | -3,3470168 | POLR2H | Homo sapiens polymerase (RNA) II (DNA directed) polypeptide H (POLR2H), transcript variant 5, mRNA [NM_001278714] |
| A_23_P152181 | 9,56E-04 | up | 1,6266087 | POLR3E | Homo sapiens polymerase (RNA) III (DNA directed) polypeptide E (80kD) (POLR3E), transcript variant 1, mRNA [NM_018119] |
| A_33_P3286041 | 4,18E-04 | up | 1,8272091 | POLR3E | Homo sapiens polymerase (RNA) III (DNA directed) polypeptide E (80kD) (POLR3E), transcript variant 1, mRNA [NM_018119] |
| A_33_P3354344 | 2,81E-04 | down | -3,0413911 | POM121L10P | Homo sapiens POM121 transmembrane nucleoporin-like 10, pseudogene (POM121L10P), non-coding RNA [NR_024593] |
| A_33_P3345036 | 0,045314338 | up | 1,9826753 | POMC | Homo sapiens proopiomelanocortin (POMC), transcript variant 1, mRNA [NM_001035256] |
| A_33_P3412233 | 0,010413661 | down | -1,6972959 | POMZP3 | Homo sapiens POM121 and ZP3 fusion (POMZP3), transcript variant 2, mRNA [NM_152992] |
| A_33_P3410895 | 0,018700404 | up | 1,595323 | POR | P450 (cytochrome) oxidoreductase [Source:HGNC Symbol;Acc:HGNC:9208] [ENST00000421059] |
| A_33_P3882624 | 5,26E-04 | down | -5,584958 | POT1 | Homo sapiens protection of telomeres 1 (POT1), transcript variant 1, mRNA [NM_015450] |
| A_24_P237804 | 0,023608912 | up | 1,6220411 | POTED | Homo sapiens POTE ankyrin domain family, member D (POTED), mRNA [NM_174981] |
| A_24_P825874 | 0,002766264 | up | 2,876811 | POTEI | Homo sapiens POTE ankyrin domain family, member I (POTEI), mRNA [NM_001277406] |
| A_33_P3835524 | 0,027530218 | down | -1,5526793 | POU2F2 | Homo sapiens POU class 2 homeobox 2 (POU2F2), transcript variant 3, mRNA [NM_001207026] |
| A_23_P114210 | 0,008985997 | down | -2,0641937 | POU3F4 | Homo sapiens POU class 3 homeobox 4 (POU3F4), mRNA [NM_000307] |
| A_24_P238118 | 2,36E-04 | down | -2,50418 | PP12613 | Homo sapiens uncharacterized LOC100192379 (PP12613), long non-coding RNA [NR_024365] |
| A_33_P3271241 | 0,04998883 | up | 1,5499318 | PPA1 | Homo sapiens pyrophosphatase (inorganic) 1 (PPA1), mRNA [NM_021129] |
| A_23_P161338 | 0,009250553 | up | 1,7846388 | PPA1 | Homo sapiens pyrophosphatase (inorganic) 1 (PPA1), mRNA [NM_021129] |
| A_19_P00807336 | 0,016940538 | up | 1,8278276 | PPA1 | Homo sapiens pyrophosphatase (inorganic) 1 (PPA1), mRNA [NM_021129] |
| A_33_P3219010 | 0,02114601 | down | -1,9349713 | PPA2 | pyrophosphatase (inorganic) 2 [Source:HGNC Symbol;Acc:HGNC:28883] [ENST00000505713] |
| A_33_P3239759 | 0,004494549 | up | 1,5150144 | PPAN-P2RY11 | Homo sapiens PPAN-P2RY11 readthrough (PPAN-P2RY11), transcript variant 2, mRNA [NM_001198690] |
| A_33_P3227079 | 0,012806513 | up | 2,0812268 | PPAPDC3 | phosphatidic acid phosphatase type 2 domain containing 3 [Source:HGNC Symbol;Acc:HGNC:28174] [ENST00000372261] |
| A_23_P18447 | 0,036270846 | up | 2,6246493 | PPARGC1A | Homo sapiens peroxisome proliferator-activated receptor gamma, coactivator 1 alpha (PPARGC1A), mRNA [NM_013261] |
| A_33_P3337009 | 0,024584478 | down | -1,5385798 | PPCS | Homo sapiens phosphopantothenoylcysteine synthetase (PPCS), transcript variant 4, mRNA [NM_001287507] |
| A_33_P3218797 | 0,022143884 | down | -1,9223186 | PPDPF | pancreatic progenitor cell differentiation and proliferation factor [Source:HGNC Symbol;Acc:HGNC:16142] [ENST00000370177] |
| A_23_P75516 | 4,03E-04 | up | 1,6696042 | PPFIA1 | Homo sapiens protein tyrosine phosphatase, receptor type, f polypeptide (PTPRF), interacting protein (liprin), alpha 1 (PPFIA1), transcript variant 2, mRNA [NM_003626] |
| A_21_P0013956 | 0,002213354 | down | -2,0344625 | PPFIA4 | protein tyrosine phosphatase, receptor type, f polypeptide (PTPRF), interacting protein (liprin), alpha 4 [Source:HGNC Symbol;Acc:HGNC:9248] [ENST00000601609] |
| A_23_P420692 | 0,028761214 | up | 1,8647918 | PPFIA4 | protein tyrosine phosphatase, receptor type, f polypeptide (PTPRF), interacting protein (liprin), alpha 4 [Source:HGNC Symbol;Acc:HGNC:9248] [ENST00000295706] |
| A_33_P3239122 | 0,002340429 | down | -1,80979 | PPIH | peptidylprolyl isomerase H (cyclophilin H) [Source:HGNC Symbol;Acc:HGNC:14651] [ENST00000372550] |
| A_33_P3239123 | 0,017964281 | down | -1,7453872 | PPIH | peptidylprolyl isomerase H (cyclophilin H) [Source:HGNC Symbol;Acc:HGNC:14651] [ENST00000372550] |
| A_23_P409553 | 1,90E-04 | up | 1,8715402 | PPM1A | Homo sapiens protein phosphatase, Mg2+/Mn2+ dependent, 1A (PPM1A), transcript variant 2, mRNA [NM_177951] |
| A_33_P3407746 | 0,001197049 | up | 1,9412636 | PPM1B | Homo sapiens protein phosphatase, Mg2+/Mn2+ dependent, 1B (PPM1B), transcript variant 2, mRNA [NM_177968] |
| A_24_P405298 | 0,004236244 | up | 1,8039216 | PPP1CB | Homo sapiens protein phosphatase 1, catalytic subunit, beta isozyme (PPP1CB), transcript variant 1, mRNA [NM_002709] |
| A_23_P204423 | 2,79E-04 | up | 1,9984902 | PPP1CC | Homo sapiens protein phosphatase 1, catalytic subunit, gamma isozyme (PPP1CC), transcript variant 1, mRNA [NM_002710] |
| A_33_P3257607 | 0,004009144 | down | -1,7324524 | PPP1R12B | Homo sapiens protein phosphatase 1, regulatory subunit 12B (PPP1R12B), transcript variant 7, mRNA [NM_001197131] |
| A_33_P3335682 | 1,62E-04 | down | -2,8168273 | PPP1R14A | Homo sapiens protein phosphatase 1, regulatory (inhibitor) subunit 14A (PPP1R14A), transcript variant 1, mRNA [NM_033256] |
| A_23_P45011 | 0,030808013 | up | 1,5255541 | PPP1R14C | Homo sapiens protein phosphatase 1, regulatory (inhibitor) subunit 14C (PPP1R14C), mRNA [NM_030949] |
| A_23_P352535 | 0,001242077 | down | -2,8062792 | PPP1R16B | Homo sapiens protein phosphatase 1, regulatory subunit 16B (PPP1R16B), transcript variant 1, mRNA [NM_015568] |
| A_23_P331479 | 0,015057822 | up | 1,7213936 | PPP1R18 | Homo sapiens protein phosphatase 1, regulatory subunit 18 (PPP1R18), transcript variant 1, mRNA [NM_133471] |
| A_24_P13285 | 0,007212185 | down | -2,9211314 | PPP1R1A | Homo sapiens protein phosphatase 1, regulatory (inhibitor) subunit 1A (PPP1R1A), mRNA [NM_006741] |
| A_33_P3215720 | 0,020530097 | down | -1,8140057 | PPP1R2 | Homo sapiens protein phosphatase 1, regulatory (inhibitor) subunit 2 (PPP1R2), transcript variant 1, mRNA [NM_001291504] |
| A_33_P3306964 | 0,007206157 | down | -1,666151 | PPP1R2 | Homo sapiens protein phosphatase 1, regulatory (inhibitor) subunit 2 (PPP1R2), transcript variant 2, mRNA [NM_006241] |
| A_24_P68819 | 0,001995967 | up | 1,8159274 | PPP1R21 | Homo sapiens protein phosphatase 1, regulatory subunit 21 (PPP1R21), transcript variant 2, mRNA [NM_152994] |
| A_23_P90895 | 0,014892142 | up | 1,6611626 | PPP1R21 | Homo sapiens protein phosphatase 1, regulatory subunit 21 (PPP1R21), transcript variant 2, mRNA [NM_152994] |
| A_23_P146497 | 2,73E-04 | up | 1,6345544 | PPP1R26 | Homo sapiens protein phosphatase 1, regulatory subunit 26 (PPP1R26), mRNA [NM_014811] |
| A_33_P3231771 | 0,005032578 | down | -2,3496358 | PPP1R2P3 | Homo sapiens protein phosphatase 1, regulatory (inhibitor) subunit 2 pseudogene 3 (PPP1R2P3), non-coding RNA [NR_038443] |
| A_33_P3344673 | 0,004274461 | up | 1,8455278 | PPP1R37 | Homo sapiens protein phosphatase 1, regulatory subunit 37 (PPP1R37), mRNA [NM_019121] |
| A_21_P0005852 | 0,019251255 | down | -1,8068202 | PPP1R42 | protein phosphatase 1, regulatory subunit 42 [Source:HGNC Symbol;Acc:HGNC:33732] [ENST00000521040] |
| A_24_P139773 | 0,002977596 | down | -1,5707839 | PPP1R8 | Homo sapiens protein phosphatase 1, regulatory subunit 8 (PPP1R8), transcript variant 2, mRNA [NM_138558] |
| A_23_P122041 | 0,023907878 | up | 1,6683453 | PPP2CA | Homo sapiens protein phosphatase 2, catalytic subunit, alpha isozyme (PPP2CA), mRNA [NM_002715] |

| | | | | | |
|---|---|---|---|---|---|
| A_23_P213620 | 0,013498493 | down | -2,5574756 | PPP2R2B | Homo sapiens protein phosphatase 2, regulatory subunit B, beta (PPP2R2B), transcript variant 10, mRNA [NM_001271948] |
| A_21_P0000166 | 0,00177455 | down | -2,5599337 | PPP5D1 | Homo sapiens PPP5 tetratricopeptide repeat domain containing 1 (PPP5D1), mRNA [NM_001205281] |
| A_24_P276628 | 4,84E-04 | down | -1,8983158 | PPT1 | Homo sapiens palmitoyl-protein thioesterase 1 (PPT1), transcript variant 1, mRNA [NM_000310] |
| A_23_P306933 | 0,005549007 | up | 1,5090388 | PPTC7 | Homo sapiens PTC7 protein phosphatase homolog (S. cerevisiae) (PPTC7), mRNA [NM_139283] |
| A_23_P207336 | 0,002166893 | down | -3,1298516 | PPY | Homo sapiens pancreatic polypeptide (PPY), mRNA [NM_002722] |
| A_33_P3245824 | 0,04817495 | up | 1,5524832 | PQLC1 | Homo sapiens PQ loop repeat containing 1 (PQLC1), transcript variant 1, mRNA [NM_025078] |
| A_33_P3253440 | 0,002096746 | down | -1,6953993 | PQLC3 | Homo sapiens PQ loop repeat containing 3 (PQLC3), transcript variant 1, mRNA [NM_152391] |
| A_33_P3308456 | 6,86E-06 | down | -4,3014083 | PRAC2 | Homo sapiens prostate cancer susceptibility candidate 2 (PRAC2), transcript variant 2, mRNA [NM_001282275] |
| A_33_P3344233 | 0,020328356 | down | -1,8347521 | PRAC2 | Homo sapiens prostate cancer susceptibility candidate 2 (PRAC2), transcript variant 1, mRNA [NM_001282276] |
| A_33_P3249349 | 7,76E-04 | down | -4,76397 | PRAME | preferentially expressed antigen in melanoma [Source:HGNC Symbol;Acc:HGNC:9336] [ENST00000406503] |
| A_33_P3249345 | 0,012997548 | up | 2,0449436 | PRAME | Homo sapiens preferentially expressed antigen in melanoma (PRAME), transcript variant 9, mRNA [NM_001291719] |
| A_33_P3303594 | 0,042875115 | up | 1,7719252 | PRAMEF11 | Homo sapiens PRAME family member 11 (PRAMEF11), mRNA [NM_001146344] |
| A_33_P3418710 | 0,034267582 | up | 2,3912597 | PRAMEF12 | Homo sapiens PRAME family member 12 (PRAMEF12), mRNA [NM_001080830] |
| A_33_P3418987 | 0,006561586 | up | 1,6342255 | PRAMEF14 | Homo sapiens PRAME family member 14 (PRAMEF14), mRNA [NM_001024661] |
| A_23_P97465 | 0,007289542 | up | 1,657194 | PRAMEF2 | Homo sapiens PRAME family member 2 (PRAMEF2), mRNA [NM_023014] |
| A_33_P3309911 | 3,63E-04 | down | -1,9711676 | PRAMEF5 | Homo sapiens PRAME family member 5 (PRAMEF5), mRNA [NM_001013407] |
| A_33_P3249449 | 0,046935804 | up | 1,5784236 | PRAMEF5 | Homo sapiens PRAME family member 5 (PRAMEF5), mRNA [NM_001013407] |
| A_33_P3332625 | 0,04309581 | down | -2,6421247 | PRB1 | Homo sapiens proline-rich protein BstNI subfamily 1 (PRB1), transcript variant 3, mRNA [NM_199354] |
| A_33_P3258593 | 0,024259994 | down | -1,920203 | PRB1 | Homo sapiens proline-rich protein BstNI subfamily 1 (PRB1), transcript variant 1, mRNA [NM_005039] |
| A_23_P116890 | 3,20E-04 | down | -2,2638621 | PRB3 | Homo sapiens proline-rich protein BstNI subfamily 3 (PRB3), mRNA [NM_006249] |
| A_33_P3342081 | 0,007330516 | up | 2,7763085 | PRDM1 | Homo sapiens PR domain containing 1, with ZNF domain (PRDM1), transcript variant 1, mRNA [NM_001198] |
| A_32_P228699 | 0,005859808 | up | 1,5085723 | PRDM10 | Homo sapiens PR domain containing 10 (PRDM10), transcript variant 2, mRNA [NM_199437] |
| A_24_P9833 | 0,0407953 | down | -2,0071588 | PRDM11 | Homo sapiens PR domain containing 11 (PRDM11), transcript variant 2, mRNA [NM_001256696] |
| A_33_P3214803 | 1,97E-05 | down | -3,2297122 | PRDM13 | Homo sapiens PR domain containing 13 (PRDM13), mRNA [NM_021620] |
| A_23_P123488 | 0,004823328 | up | 1,9725349 | PRDM14 | Homo sapiens PR domain containing 14 (PRDM14), mRNA [NM_024504] |
| A_33_P3242548 | 0,032455094 | down | -2,02058 | PRDM5 | Homo sapiens PR domain containing 5 (PRDM5), transcript variant 3, mRNA [NM_001300824] |
| A_32_P222684 | 0,01797048 | down | -1,8577124 | PRDM6 | Homo sapiens PR domain containing 6 (PRDM6), mRNA [NM_001136239] |
| A_23_P255328 | 0,010939207 | down | -2,4855416 | PRDM8 | Homo sapiens PR domain containing 8 (PRDM8), transcript variant 1, mRNA [NM_020226] |
| A_33_P3234882 | 0,005994462 | up | 1,5991429 | PRELP | Homo sapiens proline/arginine-rich end leucine-rich repeat protein (PRELP), transcript variant 1, mRNA [NM_002725] |
| A_23_P413641 | 0,040179744 | up | 2,151303 | PREX1 | Homo sapiens phosphatidylinositol-3,4,5-trisphosphate-dependent Rac exchange factor 1 (PREX1), mRNA [NM_020820] |
| A_33_P3394993 | 9,22E-08 | down | -11,281841 | PREX2 | Homo sapiens phosphatidylinositol-3,4,5-trisphosphate-dependent Rac exchange factor 2 (PREX2), transcript variant 2, mRNA [NM_025170] |
| A_23_P47466 | 0,004676359 | down | -2,159779 | PRG3 | Homo sapiens proteoglycan 3 (PRG3), mRNA [NM_006093] |
| A_23_P408285 | 0,014864335 | down | -1,7321731 | PRICKLE1 | Homo sapiens prickle homolog 1 (Drosophila) (PRICKLE1), transcript variant 1, mRNA [NM_153026] |
| A_23_P25019 | 0,00174487 | down | -1,7418295 | PRIM1 | Homo sapiens primase, DNA, polypeptide 1 (49kDa) (PRIM1), mRNA [NM_000946] |
| A_33_P3261982 | 2,54E-04 | up | 3,126055 | PRIM2 | Homo sapiens primase, DNA, polypeptide 2 (58kDa) (PRIM2), transcript variant 1, mRNA [NM_000947] |
| A_33_P3312212 | 0,002872198 | down | -1,5373639 | PRKAA1 | Homo sapiens protein kinase, AMP-activated, alpha 1 catalytic subunit (PRKAA1), transcript variant 2, mRNA [NM_206907] |
| A_32_P94160 | 0,013719295 | up | 1,8844142 | PRKAA2 | Homo sapiens protein kinase, AMP-activated, alpha 2 catalytic subunit (PRKAA2), mRNA [NM_006252] |
| A_23_P204503 | 0,004475147 | up | 1,7182993 | PRKAB1 | Homo sapiens protein kinase, AMP-activated, beta 1 non-catalytic subunit (PRKAB1), mRNA [NM_006253] |
| A_33_P3215317 | 4,38E-05 | down | -2,667875 | PRKACB | Homo sapiens protein kinase, cAMP-dependent, catalytic, beta (PRKACB), transcript variant 12, mRNA [NM_001300917] |
| A_33_P3772996 | 0,00551668 | down | -1,7463421 | PRKACB | Homo sapiens protein kinase, cAMP-dependent, catalytic, beta (PRKACB), transcript variant 7, mRNA [NM_001242860] |
| A_23_P71926 | 0,001087214 | down | -2,1778398 | PRKACG | Homo sapiens protein kinase, cAMP-dependent, catalytic, gamma (PRKACG), mRNA [NM_002732] |
| A_21_P0002740 | 0,006370881 | down | -1,7952816 | PRKAR2A-AS1 | Homo sapiens PRKAR2A antisense RNA 1 (PRKAR2A-AS1), transcript variant 1, long non-coding RNA [NR_109996] |
| A_23_P16189 | 0,001438308 | down | -1,9232069 | PRKCG | Homo sapiens protein kinase C, gamma (PRKCG), mRNA [NM_002739] |
| A_23_P205567 | 0,0270661 | up | 1,6504697 | PRKCH | Homo sapiens protein kinase C, eta (PRKCH), mRNA [NM_006255] |
| A_21_P0006826 | 0,043739267 | up | 2,0073116 | PRKCQ-AS1 | PRKCQ antisense RNA 1 [Source:HGNC Symbol;Acc:HGNC:44689] [ENST00000624974] |
| A_23_P106016 | 0,019406999 | down | -1,8192679 | PRKD1 | Homo sapiens protein kinase D1 (PRKD1), mRNA [NM_002742] |
| A_33_P3351087 | 0,010106999 | up | 1,5115802 | PRKG1 | protein kinase, cGMP-dependent, type I [Source:HGNC Symbol;Acc:HGNC:9414] [ENST00000373976] |
| A_24_P320759 | 0,012412357 | up | 1,5711564 | PRKRIP1 | Homo sapiens PRKR interacting protein 1 (IL11 inducible) (PRKRIP1), mRNA [NM_024653] |
| A_33_P3358893 | 2,16E-04 | down | -3,6980028 | PRM2 | Homo sapiens protamine 2 (PRM2), transcript variant 5, mRNA [NM_001286359] |
| A_33_P3324137 | 0,017092342 | up | 1,5691457 | PRO0628 | Homo sapiens, clone IMAGE:4399587, mRNA. [BC015320] |
| A_32_P206735 | 4,49E-04 | down | -2,5086744 | PRO1082 | Homo sapiens PRO1082 mRNA, complete cds. [AF116655] |
| A_33_P3538279 | 0,006534381 | up | 1,9008074 | PRO2852 | PREDICTED: Homo sapiens uncharacterized protein PRO2852 (PRO2852), misc_RNA [XR_158845] |
| A_33_P3368950 | 0,001732861 | down | -2,687346 | PROB1 | Homo sapiens proline-rich basic protein 1 (PROB1), mRNA [NM_001161546] |
| A_23_P165201 | 0,035483174 | down | -1,9079994 | PRODH2 | Homo sapiens proline dehydrogenase (oxidase) 2 (PRODH2), mRNA [NM_021232] |
| A_23_P412603 | 0,005127144 | down | -2,4754798 | PROKR2 | Homo sapiens prokineticin receptor 2 (PROKR2), mRNA [NM_144773] |
| A_33_P3394465 | 7,65E-04 | down | -2,2899082 | PROP1 | Homo sapiens PROP paired-like homeobox 1 (PROP1), mRNA [NM_006261] |
| A_21_P0012453 | 0,014893443 | down | -1,5107372 | PROS1 | Homo sapiens protein S (alpha) (PROS1), mRNA [NM_000313] |
| A_33_P3312182 | 0,007218325 | up | 1,6894222 | PROSER2 | Homo sapiens proline and serine rich 2 (PROSER2), mRNA [NM_153256] |
| A_24_P389608 | 0,003314322 | up | 1,9640638 | PROSER2 | Homo sapiens proline and serine rich 2 (PROSER2), mRNA [NM_153256] |
| A_23_P97573 | 0,006458786 | up | 1,5172998 | PRPF3 | Homo sapiens pre-mRNA processing factor 3 (PRPF3), mRNA [NM_004698] |
| A_24_P97001 | 0,012018967 | up | 1,5912381 | PRPF38A | Homo sapiens pre-mRNA processing factor 38A (PRPF38A), mRNA [NM_032864] |

| | | | | | |
|---|---|---|---|---|---|
| A_23_P163113 | 1,64E-04 | up | 1,777695 | PRPF39 | Homo sapiens pre-mRNA processing factor 39 (PRPF39), mRNA [NM_017922] |
| A_24_P941787 | 0,002848825 | up | 1,6263878 | PRPF4B | Homo sapiens pre-mRNA processing factor 4B (PRPF4B), mRNA [NM_003913] |
| A_24_P349466 | 4,11E-04 | down | -1,8044634 | PRR13 | Homo sapiens proline rich 13 (PRR13), transcript variant 3, mRNA [NM_001005354] |
| A_23_P118894 | 0,030321714 | up | 2,3577964 | PRR15L | Homo sapiens proline rich 15-like (PRR15L), mRNA [NM_024320] |
| A_32_P407293 | 0,00794215 | up | 3,1794348 | PRR18 | Homo sapiens proline rich 18 (PRR18), mRNA [NM_175922] |
| A_24_P178300 | 3,94E-04 | down | -5,955972 | PRR21 | Homo sapiens proline rich 21 (PRR21), mRNA [NM_001080835] |
| A_33_P3222769 | 9,60E-04 | down | -2,4473593 | PRR25 | Homo sapiens proline rich 25 (PRR25), mRNA [NM_001013638] |
| A_33_P3391039 | 0,00563498 | down | -2,4642854 | PRR27 | Homo sapiens proline rich 27 (PRR27), mRNA [NM_214711] |
| A_33_P3312642 | 0,019251851 | down | -1,9339738 | PRR32 | Homo sapiens proline rich 32 (PRR32), mRNA [NM_001122716] |
| A_24_P247273 | 0,02191441 | down | -1,5502107 | PRR33 | proline rich 33 [Source:HGNC Symbol;Acc:HGNC:35118] [ENST00000391480] |
| A_24_P147540 | 0,00196429 | down | -3,4805665 | PRR36 | Homo sapiens proline rich 36 (PRR36), transcript variant 1, mRNA [NM_001190467] |
| A_33_P3265855 | 0,031220503 | up | 1,614503 | PRR5-ARHGAP8 | Homo sapiens PRR5-ARHGAP8 readthrough (PRR5-ARHGAP8), mRNA [NM_181334] |
| A_23_P412409 | 1,85E-04 | up | 1,8376045 | PRRC2C | Homo sapiens proline-rich coiled-coil 2C (PRRC2C), mRNA [NM_015172] |
| A_23_P387537 | 0,004335125 | down | -2,3049247 | PRRG3 | Homo sapiens proline rich Gla (G-carboxyglutamic acid) 3 (transmembrane) (PRRG3), transcript variant 1, mRNA [NM_024082] |
| A_32_P192474 | 0,02048609 | down | -1,9156978 | PRRT1 | Homo sapiens proline-rich transmembrane protein 1 (PRRT1), mRNA [NM_030651] |
| A_24_P937405 | 0,009434659 | down | -1,6746392 | PRSS23 | Homo sapiens protease, serine, 23 (PRSS23), transcript variant 1, mRNA [NM_007173] |
| A_23_P350719 | 4,25E-04 | up | 3,0924425 | PRSS30P | Homo sapiens protease, serine, 30, pseudogene (PRSS30P), non-coding RNA [NR_026864] |
| A_24_P412734 | 0,009196385 | down | -4,3668303 | PRSS36 | Homo sapiens protease, serine, 36 (PRSS36), transcript variant 1, mRNA [NM_173502] |
| A_33_P3223898 | 0,010880706 | up | 3,306784 | PRSS45 | Homo sapiens protease, serine, 45 (PRSS45), mRNA [NM_199183] |
| A_33_P3340565 | 0,001512089 | up | 2,098737 | PRSS53 | Homo sapiens protease, serine, 53 (PRSS53), mRNA [NM_001039503] |
| A_33_P3345309 | 0,038809247 | down | -2,256835 | PRSS56 | Homo sapiens protease, serine, 56 (PRSS56), mRNA [NM_001195129] |
| A_33_P3380523 | 0,006615094 | down | -1,6366733 | PRTFDC1 | phosphoribosyl transferase domain containing 1 [Source:HGNC Symbol;Acc:HGNC:23333] [ENST00000376376] |
| A_24_P390909 | 0,006163849 | up | 2,0693228 | PRX | Homo sapiens periaxin (PRX), transcript variant 2, mRNA [NM_181882] |
| A_23_P11408 | 2,32E-04 | down | -2,6316955 | PRY2 | Homo sapiens PTPN13-like, Y-linked 2 (PRY2), mRNA [NM_001002758] |
| A_33_P3419108 | 0,008327827 | down | -1,7281331 | PRY2 | Homo sapiens PTPN13-like, Y-linked 2 (PRY2), mRNA [NM_001002758] |
| A_23_P12680 | 0,023089165 | up | 1,5306673 | PSAP | Homo sapiens prosaposin (PSAP), transcript variant 2, mRNA [NM_001042465] |
| A_33_P3233125 | 0,001158465 | down | -1,7514906 | PSD | Homo sapiens pleckstrin and Sec7 domain containing (PSD), transcript variant 1, mRNA [NM_002779] |
| A_33_P3312504 | 1,60E-05 | down | -2,4016175 | PSD4 | Homo sapiens pleckstrin and Sec7 domain containing 4 (PSD4), mRNA [NM_012455] |
| A_24_P22800 | 0,001246744 | down | -2,9244032 | PSG11 | Homo sapiens pregnancy specific beta-1-glycoprotein 11 (PSG11), transcript variant 1, mRNA [NM_002785] |
| A_23_P39304 | 0,002316486 | up | 1,7989802 | PSG9 | Homo sapiens pregnancy specific beta-1-glycoprotein 9 (PSG9), transcript variant 1, mRNA [NM_002784] |
| A_33_P3226610 | 0,026294341 | down | -1,7350887 | PSIP1 | Homo sapiens PC4 and SFRS1 interacting protein 1 (PSIP1), transcript variant 1, mRNA [NM_021144] |
| A_23_P383601 | 7,86E-04 | up | 1,6240243 | PSMA3-AS1 | Homo sapiens PSMA3 antisense RNA 1 (PSMA3-AS1), transcript variant 1, long non-coding RNA [NR_029434] |
| A_23_P111000 | 0,017467428 | down | -1,7186496 | PSMB9 | Homo sapiens proteasome (prosome, macropain) subunit, beta type, 9 (PSMB9), mRNA [NM_002800] |
| A_33_P3303687 | 0,015110462 | down | -1,5128661 | PSMC1 | Homo sapiens proteasome (prosome, macropain) 26S subunit, ATPase, 1 (PSMC1), mRNA [NM_002802] |
| A_24_P287941 | 0,005470315 | up | 1,5621456 | PSMC3IP | Homo sapiens PSMC3 interacting protein (PSMC3IP), transcript variant 1, mRNA [NM_013290] |
| A_33_P3335624 | 0,003581008 | down | -1,6806275 | PSMD10 | proteasome (prosome, macropain) 26S subunit, non-ATPase, 10 [Source:HGNC Symbol;Acc:HGNC:9555] [ENST00000338548] |
| A_33_P3305254 | 6,22E-04 | down | -1,6410416 | PSMD5 | Homo sapiens proteasome (prosome, macropain) 26S subunit, non-ATPase, 5 (PSMD5), transcript variant 1, mRNA [NM_005047] |
| A_33_P3305243 | 0,020218946 | up | 1,6627157 | PSMD5 | proteasome (prosome, macropain) 26S subunit, non-ATPase, 5 [Source:HGNC Symbol;Acc:HGNC:9563] [ENST00000373903] |
| A_23_P79628 | 0,015541367 | up | 1,5461434 | PSME4 | Homo sapiens proteasome (prosome, macropain) activator subunit 4 (PSME4), mRNA [NM_014614] |
| A_33_P3357653 | 7,40E-04 | down | -2,091413 | PSMG2 | proteasome (prosome, macropain) assembly chaperone 2 [Source:HGNC Symbol;Acc:HGNC:24929] [ENST00000585853] |
| A_33_P3565787 | 0,00816161 | down | -2,1922495 | PSMG3-AS1 | Homo sapiens PSMG3 antisense RNA 1 (head to head) (PSMG3-AS1), transcript variant 1, long non-coding RNA [NR_027329] |
| A_33_P3393537 | 0,019650662 | down | -1,6607398 | PTAFR | Homo sapiens platelet-activating factor receptor (PTAFR), transcript variant 2, mRNA [NM_001164722] |
| A_33_P3209915 | 0,019001298 | up | 1,6676233 | PTAR1 | Homo sapiens protein prenyltransferase alpha subunit repeat containing 1 (PTAR1), mRNA [NM_001099666] |
| A_24_P106910 | 0,03010083 | up | 1,6104153 | PTCH1 | Homo sapiens patched 1 (PTCH1), transcript variant 1a, mRNA [NM_001083602] |
| A_23_P355311 | 1,04E-04 | down | -4,881239 | PTCH2 | Homo sapiens patched 2 (PTCH2), transcript variant 1, mRNA [NM_003738] |
| A_33_P3348156 | 5,75E-08 | down | -6,1244626 | PTCH2 | patched 2 [Source:HGNC Symbol;Acc:HGNC:9586] [ENST00000438067] |
| A_32_P232411 | 0,044920914 | up | 1,5179249 | PTCHD2 | Homo sapiens patched domain containing 2 (PTCHD2), mRNA [NM_020780] |
| A_33_P3219811 | 4,02E-04 | down | -1,8142681 | PTGDS | prostaglandin D2 synthase 21kDa (brain) [Source:HGNC Symbol;Acc:HGNC:9592] [ENST00000371623] |
| A_23_P148047 | 0,010142119 | up | 1,6963592 | PTGER4 | Homo sapiens prostaglandin E receptor 4 (subtype EP4) (PTGER4), mRNA [NM_000958] |
| A_24_P135875 | 0,04670934 | down | -1,7586254 | PTGER4P2-CDK2AP2P2 | Homo sapiens PTGER4P2-CDK2AP2P2 readthrough transcribed pseudogene (PTGER4P2-CDK2AP2P2), non-coding RNA [NR_024496] |
| A_33_P3325257 | 7,85E-07 | down | -2,799685 | PTGER4P2-CDK2AP2P2 | Homo sapiens PTGER4P2-CDK2AP2P2 readthrough transcribed pseudogene (PTGER4P2-CDK2AP2P2), non-coding RNA [NR_024496] |
| A_24_P403417 | 0,001391525 | up | 2,639865 | PTGES | Homo sapiens prostaglandin E synthase (PTGES), mRNA [NM_004878] |
| A_33_P3358183 | 0,001996976 | down | -2,0597095 | PTGFR | Homo sapiens prostaglandin F receptor (FP) (PTGFR), transcript variant 1, mRNA [NM_000959] |
| A_33_P3376551 | 0,003325508 | up | 1,5064692 | PTGFR | Homo sapiens prostaglandin F receptor (FP) (PTGFR), transcript variant 2, mRNA [NM_001039585] |
| A_23_P357374 | 0,023432512 | down | -1,6341544 | PTH2 | Homo sapiens parathyroid hormone 2 (PTH2), mRNA [NM_178449] |
| A_23_P22096 | 1,95E-04 | up | 2,083669 | PTK2 | Homo sapiens protein tyrosine kinase 2 (PTK2), transcript variant 1, mRNA [NM_153831] |
| A_33_P3273000 | 0,019328482 | down | -1,63205 | PTK2B | Homo sapiens cDNA FLJ46514 fis, clone THYMU3032798, highly similar to Focal adhesion kinase 2 (EC 2.7.1.112). [AK128371] |
| A_33_P3351536 | 0,002852898 | up | 2,567587 | PTK2B | Homo sapiens protein tyrosine kinase 2 beta (PTK2B), transcript variant 1, mRNA [NM_173174] |
| A_33_P3333038 | 9,07E-04 | down | -2,529366 | PTMS | Homo sapiens parathymosin (PTMS), mRNA [NM_002824] |
| A_23_P23114 | 7,41E-06 | up | 2,190312 | PTP4A2 | Homo sapiens protein tyrosine phosphatase type IVA, member 2 (PTP4A2), transcript variant 1, mRNA [NM_080391] |
| A_21_P0012454 | 0,01580442 | down | -1,7574232 | PTPN11 | Homo sapiens protein tyrosine phosphatase, non-receptor type 11 (PTPN11), transcript variant 2, mRNA [NM_080601] |

| | | | | | |
|---|---|---|---|---|---|
| A_23_P8763 | 0,025346942 | up | 1,6491331 | PTPN12 | Homo sapiens protein tyrosine phosphatase, non-receptor type 12 (PTPN12), transcript variant 1, mRNA [NM_002835] |
| A_23_P18493 | 0,009678642 | up | 1,5633535 | PTPN13 | Homo sapiens protein tyrosine phosphatase, non-receptor type 13 (APO-1/CD95 (Fas)-associated phosphatase) (PTPN13), transcript variant 4, mRNA [NM_080685] |
| A_24_P2648 | 0,001180038 | down | -1,7926356 | PTPN14 | Homo sapiens protein tyrosine phosphatase, non-receptor type 14 (PTPN14), mRNA [NM_005401] |
| A_33_P3489222 | 0,014999097 | down | -2,3128688 | PTPN20B | Homo sapiens protein tyrosine phosphatase, non-receptor type 20B (PTPN20B), transcript variant 1, mRNA [NM_001042357] |
| A_23_P403898 | 4,27E-04 | up | 1,9669343 | PTPN3 | Homo sapiens protein tyrosine phosphatase, non-receptor type 3 (PTPN3), transcript variant 1, mRNA [NM_002829] |
| A_24_P409519 | 0,016523872 | down | -2,3263204 | PTPN5 | Homo sapiens protein tyrosine phosphatase, non-receptor type 5 (striatum-enriched) (PTPN5), transcript variant 1, mRNA [NM_006906] |
| A_21_P0005998 | 0,002613107 | up | 1,9997398 | PTPRD-AS2 | Homo sapiens PTPRD antisense RNA 2 (head to head) (PTPRD-AS2), long non-coding RNA [NR_110696] |
| A_33_P3276282 | 0,029978862 | up | 2,0947654 | PTPRF | protein tyrosine phosphatase, receptor type, F [Source:HGNC Symbol;Acc:HGNC:9670] [ENST00000617451] |
| A_21_P0000749 | 0,032937635 | up | 2,590119 | PTPRG-AS1 | Homo sapiens PTPRG antisense RNA 1 (PTPRG-AS1), transcript variant 1, long non-coding RNA [NR_038281] |
| A_24_P921321 | 1,97E-04 | up | 2,562148 | PTPRJ | Homo sapiens protein tyrosine phosphatase, receptor type, J (PTPRJ), transcript variant 1, mRNA [NM_002843] |
| A_23_P159255 | 0,022873826 | up | 1,5025264 | PTPRM | Homo sapiens protein tyrosine phosphatase, receptor type, M (PTPRM), transcript variant 2, mRNA [NM_002845] |
| A_32_P79434 | 0,029004099 | up | 2,5447445 | PTPRN2 | Homo sapiens protein tyrosine phosphatase, receptor type, N polypeptide 2 (PTPRN2), transcript variant 1, mRNA [NM_002847] |
| A_23_P60016 | 0,005232151 | down | -1,8027292 | PTTG3P | Homo sapiens pituitary tumor-transforming 3, pseudogene (PTTG3P), non-coding RNA [NR_002734] |
| A_23_P170820 | 5,69E-04 | up | 1,5989779 | PUM1 | Homo sapiens pumilio RNA-binding family member 1 (PUM1), transcript variant 1, mRNA [NM_001020658] |
| A_23_P10815 | 9,17E-04 | up | 1,5602882 | PUM1 | Homo sapiens pumilio RNA-binding family member 1 (PUM1), transcript variant 1, mRNA [NM_001020658] |
| A_24_P42446 | 0,00506156 | up | 1,747903 | PURG | purine-rich element binding protein G [Source:HGNC Symbol;Acc:HGNC:17930] [ENST00000339382] |
| A_23_P82478 | 0,004418466 | up | 1,5047832 | PUS7 | Homo sapiens pseudouridylate synthase 7 (putative) (PUS7), mRNA [NM_019042] |
| A_23_P208293 | 0,009430461 | up | 1,5246649 | PVRL2 | Homo sapiens poliovirus receptor-related 2 (herpesvirus entry mediator B) (PVRL2), transcript variant delta, mRNA [NM_001042724] |
| A_19_P00321075 | 0,00463438 | up | 1,5071967 | PVT1 | Homo sapiens Pvt1 oncogene (non-protein coding) (PVT1), long non-coding RNA [NR_003367] |
| A_33_P3357748 | 0,001441822 | up | 1,6041416 | PVT1 | Homo sapiens Pvt1 oncogene (non-protein coding) (PVT1), long non-coding RNA [NR_003367] |
| A_21_P0008707 | 0,003441147 | down | -2,9433699 | PWRN2 | Homo sapiens Prader-Willi region non-protein coding RNA 2 (PWRN2), long non-coding RNA [NR_026647] |
| A_24_P272290 | 0,001726498 | up | 1,7014121 | PXDC1 | Homo sapiens PX domain containing 1 (PXDC1), mRNA [NM_183373] |
| A_33_P3271455 | 0,01759384 | up | 1,9033346 | PXDN | Homo sapiens peroxidasin (PXDN), mRNA [NM_012293] |
| A_33_P3238166 | 0,049109433 | up | 1,6641766 | PXDN | Homo sapiens peroxidasin (PXDN), mRNA [NM_012293] |
| A_21_P0000840 | 0,026664685 | down | -2,0253441 | PXN-AS1 | Homo sapiens PXN antisense RNA 1 (PXN-AS1), long non-coding RNA [NR_038924] |
| A_23_P333022 | 0,017214615 | up | 1,7253644 | PXT1 | Homo sapiens peroxisomal, testis specific 1 (PXT1), mRNA [NM_152990] |
| A_23_P383278 | 0,00553417 | down | -1,5566671 | PYCRL | Homo sapiens pyrroline-5-carboxylate reductase-like (PYCRL), mRNA [NM_023078] |
| A_33_P3417313 | 0,02120772 | down | -1,9135177 | PYCRL | Homo sapiens pyrroline-5-carboxylate reductase-like (PYCRL), mRNA [NM_023078] |
| A_23_P405815 | 0,003467581 | up | 2,612355 | PYGM | Homo sapiens phosphorylase, glycogen, muscle (PYGM), transcript variant 1, mRNA [NM_005609] |
| A_33_P3289352 | 9,84E-04 | down | -1,773691 | PYGO2 | Homo sapiens pygopus family PHD finger 2 (PYGO2), mRNA [NM_138300] |
| A_24_P139604 | 0,011016646 | up | 2,5214603 | PYHIN1 | Homo sapiens pyrin and HIN domain family, member 1 (PYHIN1), transcript variant 4, mRNA [NM_198930] |
| A_24_P233078 | 5,93E-04 | down | -2,4294772 | PYY2 | Homo sapiens peptide YY, 2 (pseudogene) (PYY2), non-coding RNA [NR_003064] |
| A_24_P71468 | 0,044907473 | up | 1,5333086 | QPCT | Homo sapiens glutaminyl-peptide cyclotransferase (QPCT), mRNA [NM_012413] |
| A_23_P92467 | 0,007399319 | up | 1,5414495 | QRFPR | Homo sapiens pyroglutamylated RFamide peptide receptor (QRFPR), mRNA [NM_198179] |
| A_23_P395632 | 2,88E-05 | down | -4,635553 | R3HDML | Homo sapiens R3H domain containing-like (R3HDML), mRNA [NM_178491] |
| A_33_P3391240 | 0,001118115 | up | 2,1059177 | RAB19 | Homo sapiens RAB19, member RAS oncogene family (RAB19), mRNA [NM_001008749] |
| A_33_P3295803 | 0,006737907 | down | -1,6081345 | RAB2B | Homo sapiens RAB2B, member RAS oncogene family (RAB2B), transcript variant 1, mRNA [NM_032846] |
| A_24_P167338 | 0,016310861 | down | -2,0486846 | RAB30 | Homo sapiens RAB30, member RAS oncogene family (RAB30), transcript variant 2, mRNA [NM_014488] |
| A_23_P103011 | 0,001248569 | down | -2,1927872 | RAB36 | Homo sapiens RAB36, member RAS oncogene family (RAB36), mRNA [NM_004914] |
| A_33_P3314594 | 3,13E-06 | down | -2,4505167 | RAB37 | Homo sapiens RAB37, member RAS oncogene family (RAB37), transcript variant 3, mRNA [NM_175738] |
| A_33_P3665739 | 6,21E-04 | down | -2,974374 | RAB43 | AGENCOURT_6606420 NIH_MGC_106 Homo sapiens cDNA clone IMAGE:5483928 5', mRNA sequence [BM917410] |
| A_21_P0005115 | 4,02E-04 | down | -3,1531613 | RAB44 | Homo sapiens RAB44, member RAS oncogene family (RAB44), mRNA [NM_001257357] |
| A_23_P410998 | 0,001891117 | up | 1,7293711 | RAB5B | Homo sapiens RAB5B, member RAS oncogene family (RAB5B), transcript variant 1, mRNA [NM_002868] |
| A_33_P3223378 | 0,001194862 | up | 1,9908838 | RAB6C-AS1 | RAB6C antisense RNA 1 [Source:HGNC Symbol;Acc:HGNC:49278] [ENST00000624615] |
| A_24_P126139 | 0,032981273 | up | 1,6867243 | RAB9B | Homo sapiens RAB9B, member RAS oncogene family (RAB9B), mRNA [NM_016370] |
| A_33_P3278220 | 0,001509255 | down | -5,0602045 | RABEPK | Rab9 effector protein with kelch motifs [Source:HGNC Symbol;Acc:HGNC:16896] [ENST00000373544] |
| A_33_P3411991 | 0,003470286 | down | -1,5427945 | RABGAP1 | Homo sapiens RAB GTPase activating protein 1 (RABGAP1), mRNA [NM_012197] |
| A_24_P336705 | 0,011896983 | up | 1,5313325 | RABGGTB | Homo sapiens Rab geranylgeranyltransferase, beta subunit (RABGGTB), transcript variant 1, mRNA [NM_004582] |
| A_33_P3344039 | 0,001727948 | down | -1,5704852 | RABL2A | Homo sapiens RAB, member of RAS oncogene family-like 2A (RABL2A), transcript variant 2, mRNA [NM_007082] |
| A_33_P3333045 | 0,001593479 | down | -1,8495921 | RABL2A | RAB, member of RAS oncogene family-like 2A [Source:HGNC Symbol;Acc:HGNC:9799] [ENST00000467600] |
| A_33_P3220160 | 0,0441222 | down | -1,6896552 | RAC2 | Homo sapiens cDNA FLJ39605 fis, clone SKNSH2005981, weakly similar to RAS-RELATED C3 BOTULINUM TOXIN SUBSTRATE 2. [AK096924] |
| A_24_P97836 | 3,43E-04 | up | 2,1621776 | RAD17 | Homo sapiens RAD17 homolog (S. pombe) (RAD17), transcript variant 8, mRNA [NM_002873] |
| A_21_P0011373 | 0,046105567 | up | 1,5366485 | RAD51-AS1 | Homo sapiens RAD51 antisense RNA 1 (head to head) (RAD51-AS1), long non-coding RNA [NR_040058] |
| A_23_P207014 | 0,034443352 | down | -1,5384098 | RAD51C | Homo sapiens RAD51 paralog C (RAD51C), transcript variant 2, mRNA [NM_002876] |
| A_23_P94141 | 0,023680137 | down | -1,5278962 | RAD54B | Homo sapiens RAD54 homolog B (S. cerevisiae) (RAD54B), transcript variant 2, mRNA [NM_001205262] |
| A_33_P3361227 | 0,005746037 | up | 1,9154615 | RAD9B | RAD9 homolog B (S. pombe) [Source:HGNC Symbol;Acc:HGNC:21700] [ENST00000409461] |
| A_21_P0013163 | 0,002678124 | down | -1,6831219 | RAET1K | Homo sapiens retinoic acid early transcript 1K pseudogene (RAET1K), non-coding RNA [NR_024045] |
| A_23_P64525 | 0,047397956 | up | 1,6659983 | RAG2 | Homo sapiens recombination activating gene 2 (RAG2), transcript variant 1, mRNA [NM_000536] |
| A_23_P417148 | 7,33E-05 | up | 1,7668754 | RAI1 | Homo sapiens retinoic acid induced 1 (RAI1), mRNA [NM_030665] |
| A_23_P254165 | 1,71E-04 | down | -2,8871856 | RAI2 | Homo sapiens retinoic acid induced 2 (RAI2), transcript variant 2, mRNA [NM_021785] |
| A_23_P151565 | 0,002268979 | up | 1,5007187 | RALGAPA1 | Homo sapiens Ral GTPase activating protein, alpha subunit 1 (catalytic) (RALGAPA1), transcript variant 1, mRNA [NM_014990] |

| | | | | | |
|---|---|---|---|---|---|
| A_33_P3291998 | 0,017874971 | up | 1,6660522 | RALGPS2 | Ral GEF with PH domain and SH3 binding motif 2 [Source:HGNC Symbol;Acc:HGNC:30279] [ENST00000495034] |
| A_21_P0014397 | 1,21E-05 | down | -2,3207762 | RALY-AS1 | Homo sapiens RALY antisense RNA 1 (RALY-AS1), transcript variant 1, long non-coding RNA [NR_109885] |
| A_23_P50946 | 0,044186957 | down | -1,5200957 | RAMP1 | Homo sapiens receptor (G protein-coupled) activity modifying protein 1 (RAMP1), mRNA [NM_005855] |
| A_23_P111737 | 0,022973446 | down | -1,9051359 | RAMP3 | Homo sapiens receptor (G protein-coupled) activity modifying protein 3 (RAMP3), mRNA [NM_005856] |
| A_24_P81965 | 0,005475187 | up | 1,7093656 | RAP2A | Homo sapiens RAP2A, member of RAS oncogene family (RAP2A), mRNA [NM_021033] |
| A_33_P3414207 | 0,034336425 | down | -2,013535 | RAP2C-AS1 | Homo sapiens RAP2C antisense RNA 1 (RAP2C-AS1), long non-coding RNA [NR_110410] |
| A_23_P304237 | 0,01405064 | up | 1,528514 | RAPGEF1 | Homo sapiens Rap guanine nucleotide exchange factor (GEF) 1 (RAPGEF1), transcript variant 2, mRNA [NM_198679] |
| A_23_P133095 | 0,025099913 | up | 1,7896525 | RAPGEF2 | Homo sapiens Rap guanine nucleotide exchange factor (GEF) 2 (RAPGEF2), mRNA [NM_014247] |
| A_23_P151307 | 0,033655073 | up | 2,3663552 | RAPGEF3 | Homo sapiens Rap guanine nucleotide exchange factor (GEF) 3 (RAPGEF3), transcript variant 2, mRNA [NM_006105] |
| A_23_P215505 | 1,16E-04 | up | 2,5510957 | RAPGEF5 | Homo sapiens Rap guanine nucleotide exchange factor (GEF) 5 (RAPGEF5), mRNA [NM_012294] |
| A_23_P144999 | 5,14E-05 | up | 2,2620676 | RAPGEF6 | Homo sapiens Rap guanine nucleotide exchange factor (GEF) 6 (RAPGEF6), transcript variant 2, mRNA [NM_016340] |
| A_24_P924862 | 0,04157477 | up | 1,5708361 | RAPH1 | Homo sapiens Ras association (RalGDS/AF-6) and pleckstrin homology domains 1 (RAPH1), transcript variant 1, mRNA [NM_213589] |
| A_23_P134237 | 0,018045304 | up | 2,32104 | RARRES2 | Homo sapiens retinoic acid receptor responder (tazarotene induced) 2 (RARRES2), mRNA [NM_002889] |
| A_33_P3287972 | 0,043227836 | up | 1,5534444 | RASA2 | RAS p21 protein activator 2 [Source:HGNC Symbol;Acc:HGNC:9872] [ENST00000452898] |
| A_23_P502747 | 0,00864787 | up | 1,5678241 | RASAL2 | Homo sapiens RAS protein activator like 2 (RASAL2), transcript variant 2, mRNA [NM_170692] |
| A_33_P3364646 | 0,015305883 | up | 2,6078625 | RASGEF1B | Homo sapiens RasGEF domain family, member 1B (RASGEF1B), transcript variant 1, mRNA [NM_152545] |
| A_24_P383762 | 0,02570745 | down | -2,046402 | RASGEF1C | Homo sapiens RasGEF domain family, member 1C (RASGEF1C), mRNA [NM_175062] |
| A_21_P0004417 | 0,006168042 | up | 1,7392806 | RASGRF2-AS1 | Homo sapiens RASGRF2 antisense RNA 1 (RASGRF2-AS1), long non-coding RNA [NR_105015] |
| A_23_P124642 | 0,04494301 | down | -2,272042 | RASGRP1 | Homo sapiens RAS guanyl releasing protein 1 (calcium and DAG-regulated) (RASGRP1), transcript variant 1, mRNA [NM_005739] |
| A_33_P3209950 | 5,66E-06 | down | -4,3421087 | RASGRP2 | RAS guanyl releasing protein 2 (calcium and DAG-regulated) [Source:HGNC Symbol;Acc:HGNC:9879] [ENST00000377494] |
| A_33_P3365582 | 0,01566235 | up | 1,7554519 | RASGRP3 | Homo sapiens RAS guanyl releasing protein 3 (calcium and DAG-regulated) (RASGRP3), transcript variant 1, mRNA [NM_001139488] |
| A_23_P165007 | 0,00291106 | down | -2,3761446 | RASGRP4 | Homo sapiens RAS guanyl releasing protein 4 (RASGRP4), transcript variant a, mRNA [NM_170604] |
| A_23_P14124 | 0,007778268 | down | -1,8718473 | RASL11A | Homo sapiens RAS-like, family 11, member A (RASL11A), mRNA [NM_206827] |
| A_24_P150580 | 0,013452164 | up | 1,6629448 | RASL12 | Homo sapiens RAS-like, family 12 (RASL12), mRNA [NM_016563] |
| A_24_P288448 | 3,82E-05 | down | -4,077424 | RASSF2 | Ras association (RalGDS/AF-6) domain family member 2 [Source:HGNC Symbol;Acc:HGNC:9883] [ENST00000478553] |
| A_33_P3293381 | 3,24E-04 | down | -4,074582 | RASSF4 | Ras association (RalGDS/AF-6) domain family member 4 [Source:HGNC Symbol;Acc:HGNC:20793] [ENST00000428466] |
| A_24_P199251 | 2,14E-04 | down | -1,7630353 | RASSF9 | Homo sapiens Ras association (RalGDS/AF-6) domain family (N-terminal) member 9 (RASSF9), mRNA [NM_005447] |
| A_33_P3353622 | 0,030908665 | down | -1,5076305 | RBAK | Homo sapiens RB-associated KRAB zinc finger (RBAK), transcript variant 1, mRNA [NM_021163] |
| A_24_P72394 | 0,026535721 | up | 1,6691414 | RBAK | Homo sapiens RB-associated KRAB zinc finger (RBAK), transcript variant 1, mRNA [NM_021163] |
| A_33_P3232038 | 8,69E-04 | down | -2,0132763 | RBAK-RBAKDN | Homo sapiens RBAK-RBAKDN readthrough (RBAK-RBAKDN), mRNA [NM_001204513] |
| A_33_P3365047 | 1,16E-05 | down | -2,7483594 | RBBP9 | Homo sapiens retinoblastoma binding protein 9 (RBBP9), mRNA [NM_006606] |
| A_23_P166051 | 0,021750031 | down | -1,6322806 | RBCK1 | Homo sapiens RanBP-type and C3HC4-type zinc finger containing 1 (RBCK1), transcript variant 2, mRNA [NM_031229] |
| A_33_P3238315 | 0,009534211 | down | -2,3890512 | RBCK1 | Homo sapiens RanBP-type and C3HC4-type zinc finger containing 1 (RBCK1), transcript variant 1, mRNA [NM_006462] |
| A_24_P147242 | 0,008757206 | down | -2,6887069 | RBFOX1 | Homo sapiens RNA binding protein, fox-1 homolog (C. elegans) 1 (RBFOX1), transcript variant 3, mRNA [NM_145893] |
| A_21_P0013952 | 0,006171758 | down | -1,7266669 | RBFOX3 | Homo sapiens RNA binding protein, fox-1 homolog (C. elegans) 3 (RBFOX3), mRNA [NM_001082575] |
| A_24_P416159 | 4,85E-04 | up | 1,7074968 | RBL2 | Homo sapiens retinoblastoma-like 2 (RBL2), mRNA [NM_005611] |
| A_33_P3499102 | 0,043178346 | down | -1,6219267 | RBM10 | Homo sapiens cDNA FLJ40796 fis, clone TRACH2007754, highly similar to DXS8237E PROTEIN. [AK098115] |
| A_33_P3378935 | 0,030617002 | down | -2,2623653 | RBM14 | Homo sapiens mRNA for RNA binding motif protein 14 variant protein. [AB209007] |
| A_23_P161644 | 0,026974136 | up | 1,6930001 | RBM14 | Homo sapiens RNA binding motif protein 14 (RBM14), transcript variant 1, mRNA [NM_006328] |
| A_21_P0000020 | 0,007464056 | up | 1,5662596 | RBM19 | Homo sapiens RNA binding motif protein 19 (RBM19), transcript variant 1, mRNA [NM_001146699] |
| A_33_P3307163 | 0,013960026 | down | -1,7930838 | RBM20 | Homo sapiens RNA binding motif protein 20 (RBM20), mRNA [NM_001134363] |
| A_21_P0000141 | 0,00108032 | up | 1,7289416 | RBM4 | Homo sapiens RNA binding motif protein 4 (RBM4), transcript variant 3, mRNA [NM_001198844] |
| A_33_P3301346 | 2,52E-05 | down | -3,2401056 | RBM44 | RNA binding motif protein 44 [Source:HGNC Symbol;Acc:HGNC:24756] [ENST00000491996] |
| A_33_P3301351 | 0,010293387 | up | 1,7917999 | RBM44 | Homo sapiens RNA binding motif protein 44 (RBM44), mRNA [NM_001080504] |
| A_23_P132910 | 0,015629603 | up | 1,7826651 | RBM47 | Homo sapiens RNA binding motif protein 47 (RBM47), transcript variant 2, mRNA [NM_019027] |
| A_33_P3256532 | 4,40E-06 | down | -2,7307422 | RBM48 | Homo sapiens RNA binding motif protein 48 (RBM48), mRNA [NM_032120] |
| A_23_P18276 | 0,002310641 | up | 1,5069178 | RBM5 | Homo sapiens RNA binding motif protein 5 (RBM5), transcript variant 1, mRNA [NM_005778] |
| A_33_P3316323 | 0,01302472 | up | 1,5602902 | RBMS1 | Homo sapiens RNA binding motif, single stranded interacting protein 1 (RBMS1), transcript variant 1, mRNA [NM_016836] |
| A_32_P56392 | 0,012582959 | down | -1,6539619 | RBMX | Homo sapiens RNA binding motif protein, X-linked (RBMX), transcript variant 1, mRNA [NM_002139] |
| A_24_P357406 | 1,61E-04 | down | -3,2828984 | RBMY1B | Homo sapiens RNA binding motif protein, Y-linked, family 1, member B (RBMY1B), mRNA [NM_001006121] |
| A_33_P3389771 | 0,023934536 | up | 1,728947 | RBMY1B | Homo sapiens RNA binding motif protein, Y-linked, family 1, member B (RBMY1B), mRNA [NM_001006121] |
| A_21_P0013892 | 0,016188273 | up | 1,5743178 | RBMY1B | Homo sapiens RNA binding motif protein, Y-linked, family 1, member B (RBMY1B), mRNA [NM_001006121] |
| A_23_P171397 | 0,008144339 | up | 1,6301136 | RBMY1B | Homo sapiens RNA binding motif protein, Y-linked, family 1, member B (RBMY1B), mRNA [NM_001006121] |
| A_23_P257649 | 0,003163375 | down | -2,151681 | RBP1 | Homo sapiens retinol binding protein 1, cellular (RBP1), transcript variant 1, mRNA [NM_002899] |
| A_21_P0013567 | 2,63E-04 | up | 1,6198841 | RBPJ | Homo sapiens recombination signal binding protein for immunoglobulin kappa J region (RBPJ), transcript variant 4, mRNA [NM_203284] |
| A_23_P71316 | 0,007061335 | up | 1,6563023 | RBPMS | Homo sapiens RNA binding protein with multiple splicing (RBPMS), transcript variant 3, mRNA [NM_001008712] |
| A_23_P100056 | 0,010663555 | up | 1,7511945 | RBPMS2 | Homo sapiens RNA binding protein with multiple splicing 2 (RBPMS2), mRNA [NM_194272] |
| A_33_P3263841 | 0,003475038 | up | 1,595424 | RCHY1 | Homo sapiens ring finger and CHY zinc finger domain containing 1, E3 ubiquitin protein ligase (RCHY1), transcript variant 9, mRNA [NM_001278539] |
| A_23_P107132 | 0,00531427 | up | 2,1907098 | RCVRN | Homo sapiens recoverin (RCVRN), mRNA [NM_002903] |
| A_33_P3373955 | 0,009006939 | up | 1,5567517 | RD3 | Homo sapiens retinal degeneration 3 (RD3), transcript variant 1, mRNA [NM_183059] |
| A_32_P25050 | 1,07E-04 | up | 2,3487897 | RDH10 | Homo sapiens retinol dehydrogenase 10 (all-trans) (RDH10), mRNA [NM_172037] |

| | | | | | |
|---|---|---|---|---|---|
| A_23_P25684 | 0,001715773 | down | -1,9023782 | RDH11 | Homo sapiens retinol dehydrogenase 11 (all-trans/9-cis/11-cis) (RDH11), transcript variant 1, mRNA [NM_016026] |
| A_23_P50291 | 0,013639935 | down | -1,8455321 | RDH8 | Homo sapiens retinol dehydrogenase 8 (all-trans) (RDH8), mRNA [NM_015725] |
| A_23_P203023 | 6,27E-04 | up | 1,6598053 | RDX | Homo sapiens radixin (RDX), transcript variant 3, mRNA [NM_002906] |
| A_33_P3278941 | 4,34E-04 | up | 2,2279098 | REC8 | Homo sapiens REC8 meiotic recombination protein (REC8), transcript variant 2, mRNA [NM_001048205] |
| A_23_P96285 | 0,002100263 | down | -2,82486 | REEP1 | Homo sapiens receptor accessory protein 1 (REEP1), transcript variant 2, mRNA [NM_022912] |
| A_33_P3369834 | 0,002428134 | down | -2,056249 | REEP6 | Homo sapiens receptor accessory protein 6 (REEP6), mRNA [NM_138393] |
| A_23_P90743 | 0,003791076 | down | -1,8567564 | REG1A | Homo sapiens regenerating islet-derived 1 alpha (REG1A), mRNA [NM_002909] |
| A_23_P389500 | 0,012484595 | down | -2,051545 | REG1B | Homo sapiens regenerating islet-derived 1 beta (REG1B), mRNA [NM_006507] |
| A_23_P56938 | 0,004576222 | up | 1,5793661 | REL | Homo sapiens v-rel avian reticuloendotheliosis viral oncogene homolog (REL), transcript variant 1, mRNA [NM_002908] |
| A_23_P104689 | 0,009092363 | up | 1,6518216 | RELA | Homo sapiens v-rel avian reticuloendotheliosis viral oncogene homolog A (RELA), transcript variant 1, mRNA [NM_021975] |
| A_24_P309095 | 0,005914027 | up | 1,6514089 | RELN | Homo sapiens reelin (RELN), transcript variant 1, mRNA [NM_005045] |
| A_23_P28948 | 0,04325109 | up | 1,8310893 | REM1 | Homo sapiens RAS (RAD and GEM)-like GTP-binding 1 (REM1), mRNA [NM_014012] |
| A_33_P3305487 | 0,00113792 | up | 1,8648372 | REM2 | Homo sapiens RAS (RAD and GEM)-like GTP binding 2 (REM2), mRNA [NM_173527] |
| A_23_P137157 | 5,89E-04 | down | -3,9303076 | RENBP | Homo sapiens renin binding protein (RENBP), mRNA [NM_002910] |
| A_24_P342591 | 0,005684442 | up | 1,5617958 | RERE | Homo sapiens arginine-glutamic acid dipeptide (RE) repeats (RERE), transcript variant 1, mRNA [NM_012102] |
| A_24_P321525 | 0,015343121 | down | -2,7631009 | RERG | Homo sapiens RAS-like, estrogen-regulated, growth inhibitor (RERG), transcript variant 1, mRNA [NM_032918] |
| A_23_P92196 | 1,18E-06 | down | -4,9953637 | RETNLB | Homo sapiens resistin like beta (RETNLB), mRNA [NM_032579] |
| A_33_P3312682 | 0,025760412 | down | -1,5892729 | REXO1 | Homo sapiens REX1, RNA exonuclease 1 homolog (S. cerevisiae) (REXO1), mRNA [NM_020695] |
| A_23_P18465 | 0,002443944 | up | 1,5012732 | RFC1 | Homo sapiens replication factor C (activator 1) 1, 145kDa (RFC1), transcript variant 1, mRNA [NM_002913] |
| A_33_P3375790 | 6,27E-05 | down | -3,029307 | RFPL4AL1 | Homo sapiens ret finger protein-like 4A-like 1 (RFPL4AL1), mRNA [NM_001277397] |
| A_33_P3366127 | 0,020132273 | up | 1,5307356 | RFPL4AL1 | Homo sapiens ret finger protein-like 4A-like 1 (RFPL4AL1), mRNA [NM_001277397] |
| A_33_P3209356 | 0,002484388 | up | 1,5063716 | RFWD2 | Homo sapiens ring finger and WD repeat domain 2, E3 ubiquitin protein ligase (RFWD2), transcript variant 1, mRNA [NM_022457] |
| A_33_P3267460 | 5,58E-04 | down | -2,0930707 | RFX3 | Homo sapiens regulatory factor X, 3 (influences HLA class II expression) (RFX3), transcript variant 4, mRNA [NM_001282117] |
| A_21_P0013689 | 0,0421808 | down | -1,586674 | RFX3-AS1 | Homo sapiens RFX3 antisense RNA 1 (RFX3-AS1), long non-coding RNA [NR_121586] |
| A_23_P76332 | 0,008245667 | up | 1,5964154 | RFX4 | Homo sapiens regulatory factor X, 4 (influences HLA class II expression) (RFX4), transcript variant 3, mRNA [NM_213594] |
| A_33_P3281741 | 0,02792349 | down | -2,1476052 | RFX8 | Homo sapiens RFX family member 8, lacking RFX DNA binding domain (RFX8), mRNA [NM_001145664] |
| A_19_P00319745 | 0,037639253 | down | -2,1801445 | RFX8 | Homo sapiens RFX family member 8, lacking RFX DNA binding domain (RFX8), mRNA [NM_001145664] |
| A_19_P00317318 | 0,018827833 | down | -2,5100098 | RFX8 | Homo sapiens RFX family member 8, lacking RFX DNA binding domain (RFX8), mRNA [NM_001145664] |
| A_33_P3240532 | 2,47E-04 | down | -3,523404 | RGL1 | Homo sapiens ral guanine nucleotide dissociation stimulator-like 1 (RGL1), transcript variant 1, mRNA [NM_015149] |
| A_23_P114423 | 0,003127139 | up | 2,5771427 | RGN | Homo sapiens regucalcin (RGN), transcript variant 2, mRNA [NM_152869] |
| A_33_P3326914 | 0,028714042 | down | -1,6059132 | RGP1 | Homo sapiens RGP1 retrograde golgi transport homolog (S. cerevisiae) (RGP1), mRNA [NM_001080496] |
| A_21_P0002089 | 0,02962188 | down | -2,0340915 | RGPD4-AS1 | RGPD4 antisense RNA 1 (head to head) [Source:HGNC Symbol;Acc:HGNC:49273] [ENST00000417284] |
| A_23_P218637 | 0,007867793 | up | 1,6798602 | RGPD5 | Homo sapiens RANBP2-like and GRIP domain containing 5 (RGPD5), transcript variant 1, mRNA [NM_005054] |
| A_24_P289273 | 5,38E-04 | down | -2,7538824 | RGR | retinal G protein coupled receptor [Source:HGNC Symbol;Acc:HGNC:9990] [ENST00000372092] |
| A_23_P219197 | 8,43E-04 | down | -1,9183941 | RGS3 | Homo sapiens regulator of G-protein signaling 3 (RGS3), transcript variant 4, mRNA [NM_134427] |
| A_33_P3212022 | 0,003736527 | up | 2,1705914 | RGS6 | Homo sapiens regulator of G-protein signaling 6 (RGS6), transcript variant 4, mRNA [NM_001204418] |
| A_32_P355943 | 0,008913815 | up | 1,5919659 | RGSL1 | Homo sapiens regulator of G-protein signaling like 1 (RGSL1), mRNA [NM_001137669] |
| A_33_P3330731 | 0,014096583 | down | -1,5629982 | RHBDD3 | rhomboid domain containing 3 [Source:HGNC Symbol;Acc:HGNC:1308] [ENST00000406335] |
| A_23_P329870 | 0,001003534 | up | 1,6771039 | RHBDF2 | Homo sapiens rhomboid 5 homolog 2 (Drosophila) (RHBDF2), transcript variant 1, mRNA [NM_024599] |
| A_23_P26468 | 0,001100711 | down | -2,1971986 | RHBDL1 | Homo sapiens rhomboid, veinlet-like 1 (Drosophila) (RHBDL1), transcript variant 1, mRNA [NM_001278720] |
| A_33_P3364661 | 0,00169643 | down | -1,9726269 | RHOA | ras homolog family member A [Source:HGNC Symbol;Acc:HGNC:667] [ENST00000265538] |
| A_24_P104119 | 0,005296737 | up | 1,8769523 | RHOF | Homo sapiens ras homolog family member F (in filopodia) (RHOF), mRNA [NM_019034] |
| A_23_P58132 | 0,023632947 | down | -2,030157 | RHOH | Homo sapiens ras homolog family member H (RHOH), transcript variant 6, mRNA [NM_004310] |
| A_32_P49844 | 0,013334809 | down | -1,5265878 | RHOQ | Homo sapiens ras homolog family member Q (RHOQ), mRNA [NM_012249] |
| A_33_P3222962 | 0,008586072 | up | 2,8772078 | RIBC1 | Homo sapiens RIB43A domain with coiled-coils 1 (RIBC1), transcript variant 2, mRNA [NM_144968] |
| A_32_P193322 | 1,65E-04 | up | 1,9907811 | RICTOR | Homo sapiens RPTOR independent companion of MTOR, complex 2 (RICTOR), transcript variant 1, mRNA [NM_152756] |
| A_32_P334325 | 0,015734648 | down | -1,517057 | RIMBP2 | Homo sapiens RIMS binding protein 2 (RIMBP2), mRNA [NM_015347] |
| A_23_P342585 | 0,013035848 | up | 1,5378355 | RIMS1 | regulating synaptic membrane exocytosis 1 [Source:HGNC Symbol;Acc:HGNC:17282] [ENST00000370419] |
| A_32_P199901 | 0,005005373 | up | 1,5814513 | RIPPLY1 | Homo sapiens ripply transcriptional repressor 1 (RIPPLY1), transcript variant 1, mRNA [NM_138382] |
| A_33_P3416588 | 0,038023844 | down | -1,8489927 | RIT2 | Homo sapiens Ras-like without CAAX 2 (RIT2), transcript variant 1, mRNA [NM_002930] |
| A_23_P163361 | 2,64E-04 | down | -4,6040835 | RLBP1 | Homo sapiens retinaldehyde binding protein 1 (RLBP1), mRNA [NM_000326] |
| A_33_P3236591 | 0,001729126 | down | -1,6308081 | RLTPR | Homo sapiens RGD motif, leucine rich repeats, tropomodulin domain and proline-rich containing (RLTPR), mRNA [NM_001013838] |
| A_33_P3401284 | 0,005443686 | down | -2,4584398 | RMRP | Homo sapiens RNA component of mitochondrial RNA processing endoribonuclease (RMRP), RNase MRP RNA [NR_003051] |
| A_33_P3318791 | 0,034196537 | down | -1,7561986 | RMST | Homo sapiens rhabdomyosarcoma 2 associated transcript (non-protein coding) (RMST), long non-coding RNA [NR_024037] |
| A_33_P3336632 | 0,002288778 | down | -1,8485851 | RNA28S5 | Homo sapiens RNA, 28S ribosomal 5 (RNA28S5), ribosomal RNA [NR_003287] |
| A_33_P3261937 | 9,49E-04 | down | -2,8908558 | RNASE13 | Homo sapiens ribonuclease, RNase A family, 13 (non-active) (RNASE13), mRNA [NM_001012264] |
| A_23_P376124 | 0,002467793 | up | 2,3158414 | RNASE8 | Homo sapiens ribonuclease, RNase A family, 8 (RNASE8), mRNA [NM_138331] |
| A_33_P3358977 | 1,66E-04 | up | 1,7965621 | RNASEH2C | Homo sapiens ribonuclease H2, subunit C (RNASEH2C), mRNA [NM_032193] |
| A_23_P56709 | 3,12E-05 | up | 2,1132991 | RNF103 | Homo sapiens ring finger protein 103 (RNF103), transcript variant 1, mRNA [NM_005667] |
| A_23_P107116 | 0,01365763 | up | 1,6479564 | RNF112 | Homo sapiens ring finger protein 112 (RNF112), mRNA [NM_007148] |
| A_23_P416112 | 4,87E-04 | down | -1,8545179 | RNF168 | Homo sapiens ring finger protein 168, E3 ubiquitin protein ligase (RNF168), mRNA [NM_152617] |

| | | | | | |
|---|---|---|---|---|---|
| A_23_P350172 | 0,022705125 | up | 1,5182712 | RNF169 | Homo sapiens ring finger protein 169 (RNF169), mRNA [NM_001098638] |
| A_24_P23995 | 0,040603496 | down | -1,6703297 | RNF187 | Homo sapiens ring finger protein 187 (RNF187), mRNA [NM_001010858] |
| A_33_P3316978 | 0,02572365 | up | 1,8804259 | RNF207 | Homo sapiens ring finger protein 207 (RNF207), mRNA [NM_207396] |
| A_33_P3304538 | 0,017535655 | up | 2,1021073 | RNF207 | Homo sapiens ring finger protein 207 (RNF207), mRNA [NM_207396] |
| A_33_P3304533 | 0,03296044 | up | 1,93341 | RNF207 | ring finger protein 207 [Source:HGNC Symbol;Acc:HGNC:32947] [ENST00000466994] |
| A_33_P3318444 | 3,03E-04 | down | -2,2894619 | RNF222 | Homo sapiens ring finger protein 222 (RNF222), mRNA [NM_001146684] |
| A_24_P314931 | 0,048437648 | up | 2,2755525 | RNF39 | Homo sapiens ring finger protein 39 (RNF39), transcript variant 1, mRNA [NM_025236] |
| A_23_P213592 | 9,91E-05 | up | 1,8533226 | RNF44 | Homo sapiens ring finger protein 44 (RNF44), mRNA [NM_014901] |
| A_24_P260699 | 0,008258455 | up | 1,6887281 | RNF8 | Homo sapiens ring finger protein 8, E3 ubiquitin protein ligase (RNF8), transcript variant 1, mRNA [NM_003958] |
| A_33_P3230103 | 0,025288569 | down | -1,8244199 | RNFT2 | Homo sapiens ring finger protein, transmembrane 2 (RNFT2), transcript variant 2, mRNA [NM_032814] |
| A_23_P80503 | 0,01066331 | down | -1,7480675 | ROBO1 | Homo sapiens roundabout, axon guidance receptor, homolog 1 (Drosophila) (ROBO1), transcript variant 2, mRNA [NM_133631] |
| A_33_P3278877 | 0,001325925 | down | -3,667594 | ROBO2 | Homo sapiens roundabout, axon guidance receptor, homolog 2 (Drosophila) (ROBO2), transcript variant 5, mRNA [NM_001290065] |
| A_24_P538403 | 0,001914673 | up | 1,5680584 | ROCK1 | Homo sapiens Rho-associated, coiled-coil containing protein kinase 1 (ROCK1), mRNA [NM_005406] |
| A_23_P100499 | 0,008835088 | down | -1,7705923 | ROGDI | Homo sapiens rogdi homolog (Drosophila) (ROGDI), transcript variant 1, mRNA [NM_024589] |
| A_32_P184464 | 0,001026554 | down | -4,3947144 | ROPN1 | Homo sapiens rhophilin associated tail protein 1 (ROPN1), mRNA [NM_017578] |
| A_23_P121885 | 4,78E-06 | down | -2,8940463 | ROPN1L | Homo sapiens rhophilin associated tail protein 1-like (ROPN1L), transcript variant 1, mRNA [NM_031916] |
| A_19_P00322190 | 0,006460912 | down | -1,5520355 | RP9P | Homo sapiens retinitis pigmentosa 9 pseudogene (RP9P), non-coding RNA [NR_003500] |
| A_33_P3214597 | 0,002991322 | down | -1,6324142 | RP9P | Homo sapiens retinitis pigmentosa 9 pseudogene (RP9P), non-coding RNA [NR_003500] |
| A_33_P3312802 | 5,42E-04 | down | -1,7872787 | RPL13AP17 | Homo sapiens ribosomal protein L13a pseudogene 17 (RPL13AP17), non-coding RNA [NR_003680] |
| A_24_P849801 | 0,008497959 | up | 1,5489717 | RPL22 | Homo sapiens ribosomal protein L22 (RPL22), mRNA [NM_000983] |
| A_33_P3257943 | 0,027525201 | down | -1,5476283 | RPL26 | Homo sapiens ribosomal protein L26 (RPL26), mRNA [NM_000987] |
| A_21_P0011094 | 0,012183568 | down | -2,2106445 | RPL31 | Homo sapiens ribosomal protein L31 (RPL31), transcript variant 1, mRNA [NM_000993] |
| A_33_P3278137 | 0,045787133 | up | 2,1648536 | RPL31P11 | Homo sapiens ribosomal protein L31 pseudogene 11 (RPL31P11), non-coding RNA [NR_002595] |
| A_21_P0012460 | 0,008111614 | up | 1,6069053 | RPL32P3 | Homo sapiens ribosomal protein L32 pseudogene 3 (RPL32P3), non-coding RNA [NR_003111] |
| A_24_P7629 | 0,001120603 | up | 2,6260662 | RPL32P3 | Homo sapiens ribosomal protein L32 pseudogene 3 (RPL32P3), non-coding RNA [NR_003111] |
| A_33_P3255647 | 0,004308448 | down | -1,8121257 | RPL36A-HNRNPH2 | Homo sapiens RPL36A-HNRNPH2 readthrough (RPL36A-HNRNPH2), transcript variant 1, mRNA [NM_001199973] |
| A_33_P3587611 | 1,38E-04 | up | 2,4769306 | RPL36AP33 | Homo sapiens cDNA clone MGC:23914 IMAGE:4769647, complete cds. [BC020828] |
| A_23_P118095 | 0,032312077 | down | -1,883266 | RPL3L | Homo sapiens ribosomal protein L3-like (RPL3L), mRNA [NM_005061] |
| A_33_P3291349 | 0,015244132 | up | 1,6385138 | RPRD1A | Homo sapiens regulation of nuclear pre-mRNA domain containing 1A (RPRD1A), transcript variant 4, mRNA [NM_001303413] |
| A_24_P250499 | 8,23E-05 | up | 1,9890655 | RPRD1B | Homo sapiens regulation of nuclear pre-mRNA domain containing 1B (RPRD1B), mRNA [NM_021215] |
| A_32_P203013 | 0,005437937 | down | -2,5153086 | RPS10P7 | Homo sapiens ribosomal protein S10 pseudogene 7 (RPS10P7), non-coding RNA [NR_026667] |
| A_23_P82128 | 0,009298298 | down | -1,5059098 | RPS12 | Homo sapiens ribosomal protein S12 (RPS12), mRNA [NM_001016] |
| A_33_P3357049 | 4,27E-04 | up | 2,3815944 | RPS15AP10 | Homo sapiens ribosomal protein S15a pseudogene 10 (RPS15AP10), non-coding RNA [NR_026768] |
| A_21_P0010964 | 0,004251708 | down | -1,53368 | RPS2 | Homo sapiens ribosomal protein S2 (RPS2), mRNA [NM_002952] |
| A_33_P3345414 | 0,009726422 | up | 1,9653456 | RPS4X | ribosomal protein S4, X-linked [Source:HGNC Symbol;Acc:HGNC:10424] [ENST00000373626] |
| A_24_P497226 | 0,007107258 | up | 1,5296128 | RPS6KB1 | Homo sapiens ribosomal protein S6 kinase, 70kDa, polypeptide 1 (RPS6KB1), transcript variant 1, mRNA [NM_003161] |
| A_24_P349743 | 0,007047434 | down | -1,7184106 | RPSAP58 | Homo sapiens ribosomal protein SA pseudogene 58 (RPSAP58), non-coding RNA [NR_003662] |
| A_33_P3220728 | 6,65E-04 | up | 1,7966636 | RREB1 | Homo sapiens ras responsive element binding protein 1 (RREB1), transcript variant 1, mRNA [NM_001003699] |
| A_24_P382017 | 4,46E-04 | up | 1,8120234 | RRN3 | Homo sapiens RRN3 RNA polymerase I transcription factor homolog (S. cerevisiae) (RRN3), transcript variant 1, mRNA [NM_018427] |
| A_21_P0006626 | 0,03365857 | up | 1,8245717 | RRP12 | ribosomal RNA processing 12 homolog (S. cerevisiae) [Source:HGNC Symbol;Acc:HGNC:29100] [ENST00000621606] |
| A_32_P221822 | 0,029378207 | up | 1,6278316 | RRP15 | Homo sapiens ribosomal RNA processing 15 homolog (S. cerevisiae) (RRP15), mRNA [NM_016052] |
| A_24_P242036 | 2,90E-04 | down | -1,742992 | RRP7B | Homo sapiens ribosomal RNA processing 7 homolog B (S. cerevisiae) (RRP7B), non-coding RNA [NR_002184] |
| A_24_P365679 | 0,02222675 | up | 1,7975917 | RRP7B | ribosomal RNA processing 7 homolog B (S. cerevisiae) [Source:HGNC Symbol;Acc:HGNC:30454] [ENST00000357802] |
| A_33_P3888365 | 0,001217174 | up | 1,5070258 | RSBN1 | Homo sapiens round spermatid basic protein 1 (RSBN1), mRNA [NM_018364] |
| A_23_P139388 | 0,006957973 | up | 1,509701 | RSF1 | Homo sapiens remodeling and spacing factor 1 (RSF1), mRNA [NM_016578] |
| A_33_P3294654 | 0,0444919 | down | -1,7498479 | RSG1 | Homo sapiens REM2 and RAB-like small GTPase 1 (RSG1), mRNA [NM_030907] |
| A_23_P362736 | 0,025669498 | up | 2,3711314 | RSPH10B | Homo sapiens radial spoke head 10 homolog B (Chlamydomonas) (RSPH10B), mRNA [NM_173565] |
| A_23_P67414 | 0,003329935 | up | 2,6319323 | RSPH6A | Homo sapiens radial spoke head 6 homolog A (Chlamydomonas) (RSPH6A), mRNA [NM_030785] |
| A_33_P3248749 | 0,002641808 | down | -1,927675 | RSPH9 | radial spoke head 9 homolog (Chlamydomonas) [Source:HGNC Symbol;Acc:HGNC:21057] [ENST00000372165] |
| A_24_P106542 | 1,71E-06 | down | -4,568058 | RSPO3 | Homo sapiens R-spondin 3 (RSPO3), mRNA [NM_032784] |
| A_23_P53267 | 0,003116291 | up | 1,5050066 | RSRC2 | Homo sapiens arginine/serine-rich coiled-coil 2 (RSRC2), transcript variant 1, mRNA [NM_023012] |
| A_24_P390403 | 2,35E-04 | up | 1,7353557 | RTF1 | Homo sapiens Rtf1, Paf1/RNA polymerase II complex component, homolog (S. cerevisiae) (RTF1), mRNA [NM_015138] |
| A_33_P3362371 | 0,011861797 | down | -1,6759728 | RTN3 | Homo sapiens reticulon 3 (RTN3), transcript variant 7, mRNA [NM_001265591] |
| A_23_P432056 | 0,017068157 | down | -2,6403542 | RTN4RL1 | Homo sapiens reticulon 4 receptor-like 1 (RTN4RL1), mRNA [NM_178568] |
| A_32_P528967 | 0,010179764 | down | -2,4457114 | RTP1 | Homo sapiens receptor (chemosensory) transporter protein 1 (RTP1), mRNA [NM_153708] |
| A_33_P3341841 | 2,04E-04 | up | 3,191923 | RTP2 | Homo sapiens receptor (chemosensory) transporter protein 2 (RTP2), mRNA [NM_001004312] |
| A_24_P197284 | 0,02976383 | down | -2,0681543 | RTP5 | Homo sapiens receptor (chemosensory) transporter protein 5 (putative) (RTP5), mRNA [NM_173821] |
| A_24_P941708 | 0,00716175 | up | 1,650449 | RUFY2 | Homo sapiens RUN and FYVE domain containing 2 (RUFY2), transcript variant 2, mRNA [NM_001042417] |
| A_24_P221575 | 8,47E-04 | down | -2,3704712 | RUFY3 | Homo sapiens RUN and FYVE domain containing 3 (RUFY3), transcript variant 1, mRNA [NM_001037442] |
| A_24_P91094 | 0,02140403 | down | -2,134461 | RUNDC3A | Homo sapiens RUN domain containing 3A (RUNDC3A), transcript variant 2, mRNA [NM_006695] |
| A_33_P3418917 | 2,28E-04 | up | 1,7168255 | RUSC1-AS1 | Homo sapiens RUSC1 antisense RNA 1 (RUSC1-AS1), mRNA [NM_001039517] |

| | | | | | |
|---|---|---|---|---|---|
| A_23_P363365 | 0,019214367 | down | -2,404756 | RXFP3 | Homo sapiens relaxin/insulin-like family peptide receptor 3 (RXFP3), mRNA [NM_016568] |
| A_24_P154142 | 0,004279739 | down | -2,6214268 | RXFP4 | Homo sapiens relaxin/insulin-like family peptide receptor 4 (RXFP4), mRNA [NM_181885] |
| A_23_P423197 | 0,00364023 | up | 1,6919554 | RXRA | Homo sapiens retinoid X receptor, alpha (RXRA), transcript variant 1, mRNA [NM_002957] |
| A_23_P201711 | 0,020470437 | down | -1,5268294 | S100A6 | Homo sapiens S100 calcium binding protein A6 (S100A6), mRNA [NM_014624] |
| A_24_P335092 | 0,001784169 | up | 3,75498 | SAA1 | Homo sapiens serum amyloid A1 (SAA1), transcript variant 1, mRNA [NM_000331] |
| A_33_P3408913 | 0,015974768 | up | 2,7418876 | SAA2 | Homo sapiens serum amyloid A2 (SAA2), transcript variant 2, mRNA [NM_001127380] |
| A_33_P3408918 | 0,020361034 | up | 2,394654 | SAA2 | Homo sapiens serum amyloid A2 (SAA2), transcript variant 1, mRNA [NM_030754] |
| A_23_P87238 | 0,018635644 | up | 2,870104 | SAA4 | Homo sapiens serum amyloid A4, constitutive (SAA4), mRNA [NM_006512] |
| A_21_P0014499 | 1,32E-04 | down | -5,415231 | SACS-AS1 | Homo sapiens SACS antisense RNA 1 (SACS-AS1), long non-coding RNA [NR_103450] |
| A_32_P130788 | 0,016074648 | up | 2,0872233 | SAMD13 | Homo sapiens sterile alpha motif domain containing 13 (SAMD13), transcript variant 1, mRNA [NM_001010971] |
| A_24_P175187 | 0,007095725 | up | 1,8445222 | SAMD9 | Homo sapiens sterile alpha motif domain containing 9 (SAMD9), transcript variant 1, mRNA [NM_017654] |
| A_33_P3406281 | 0,001567317 | down | -1,5747497 | SAMHD1 | Homo sapiens SAM domain and HD domain 1 (SAMHD1), mRNA [NM_015474] |
| A_24_P191971 | 0,002663335 | up | 1,9733967 | SAP30L | Homo sapiens SAP30-like (SAP30L), transcript variant 1, mRNA [NM_024632] |
| A_24_P115507 | 7,57E-04 | down | -2,0262835 | SARDH | Homo sapiens sarcosine dehydrogenase (SARDH), transcript variant 1, mRNA [NM_007101] |
| A_24_P35400 | 0,012271836 | up | 1,6563976 | SARDH | Homo sapiens sarcosine dehydrogenase (SARDH), transcript variant 1, mRNA [NM_007101] |
| A_24_P658584 | 0,002428395 | up | 2,5485418 | SASH1 | Homo sapiens SAM and SH3 domain containing 1 (SASH1), mRNA [NM_015278] |
| A_33_P3209279 | 0,010276693 | up | 1,5724483 | SASH1 | Homo sapiens SAM and SH3 domain containing 1 (SASH1), mRNA [NM_015278] |
| A_33_P3209283 | 0,01222463 | up | 2,4702744 | SASH1 | SAM and SH3 domain containing 1 [Source:HGNC Symbol;Acc:HGNC:19182] [ENST00000367469] |
| A_24_P349151 | 0,026360476 | up | 1,5263638 | SASS6 | Homo sapiens spindle assembly 6 homolog (C. elegans) (SASS6), mRNA [NM_194292] |
| A_33_P3223602 | 0,004975528 | up | 1,509354 | SATL1 | Homo sapiens spermidine/spermine N1-acetyl transferase-like 1 (SATL1), mRNA [NM_001012980] |
| A_23_P142031 | 9,08E-04 | down | -1,5599358 | SBK2 | Homo sapiens SH3 domain binding kinase family, member 2 (SBK2), mRNA [NM_001101401] |
| A_33_P3237744 | 0,012145695 | down | -2,1139839 | SBK3 | Homo sapiens SH3 domain binding kinase family, member 3 (SBK3), mRNA [NM_001199824] |
| A_33_P3298577 | 0,010685275 | down | -1,6371659 | SBNO1 | Homo sapiens strawberry notch homolog 1 (Drosophila) (SBNO1), transcript variant 1, mRNA [NM_001167856] |
| A_24_P79153 | 0,008856625 | up | 1,5400203 | SCAMP4 | Homo sapiens secretory carrier membrane protein 4 (SCAMP4), mRNA [NM_079834] |
| A_33_P3259587 | 0,012732944 | down | -2,0280046 | SCAND2P | Homo sapiens SCAN domain containing 2 pseudogene (SCAND2P), transcript variant 2, non-coding RNA [NR_003654] |
| A_23_P205875 | 0,011283192 | down | -1,5718697 | SCAND2P | Homo sapiens SCAN domain containing 2 pseudogene (SCAND2P), transcript variant 1, non-coding RNA [NR_004859] |
| A_24_P162244 | 0,0343151 | up | 1,5483806 | SCAPER | S-phase cyclin A-associated protein in the ER [Source:HGNC Symbol;Acc:HGNC:13081] [ENST00000303521] |
| A_23_P163408 | 0,004678111 | up | 1,5491894 | SCAPER | Homo sapiens S-phase cyclin A-associated protein in the ER (SCAPER), transcript variant 1, mRNA [NM_020843] |
| A_23_P15414 | 0,003371753 | down | -1,7558266 | SCARF1 | Homo sapiens scavenger receptor class F, member 1 (SCARF1), transcript variant 1, mRNA [NM_003693] |
| A_33_P3832341 | 0,002615244 | up | 1,6130958 | SCARNA22 | AGENCOURT_10609207 NIH_MGC_126 Homo sapiens cDNA clone IMAGE:6727460 5', mRNA sequence [BU955376] |
| A_21_P0000349 | 0,003380718 | up | 1,7191432 | SCARNA3 | Homo sapiens small Cajal body-specific RNA 3 (SCARNA3), guide RNA [NR_002998] |
| A_33_P3460485 | 0,001448317 | up | 2,7967365 | SCARNA4 | MR2-CI0186-291100-008-f04 CI0186 Homo sapiens cDNA, mRNA sequence [BF814437] |
| A_33_P3274691 | 0,020779708 | down | -2,263404 | SCART1 | scavenger receptor protein family member [Source:EntrezGene;Acc:619207] [ENST00000482993] |
| A_33_P3247204 | 0,002139085 | down | -2,3030558 | SCART1 | scavenger receptor protein family member [Source:EntrezGene;Acc:619207] [ENST00000475114] |
| A_24_P385134 | 0,010877753 | up | 1,6940075 | SCD5 | Homo sapiens stearoyl-CoA desaturase 5 (SCD5), transcript variant 1, mRNA [NM_001037582] |
| A_24_P88696 | 0,004804327 | up | 1,8996855 | SCG2 | Homo sapiens secretogranin II (SCG2), mRNA [NM_003469] |
| A_33_P3340869 | 8,05E-04 | down | -2,3077233 | SCGB1B2P | Homo sapiens secretoglobin, family 1B, member 2, pseudogene (SCGB1B2P), non-coding RNA [NR_027620] |
| A_33_P3228014 | 0,004605068 | up | 1,5544705 | SCGB1D4 | Homo sapiens secretoglobin, family 1D, member 4 (SCGB1D4), mRNA [NM_206998] |
| A_32_P191860 | 0,001704654 | down | -2,0999014 | SCGB2B2 | Homo sapiens secretoglobin, family 2B, member 2 (SCGB2B2), mRNA [NM_001025591] |
| A_33_P3210379 | 0,030095743 | down | -1,5758264 | SCGB3A1 | Homo sapiens secretoglobin, family 3A, member 1 (SCGB3A1), mRNA [NM_052863] |
| A_23_P145006 | 0,008274924 | down | -1,7500328 | SCGB3A2 | Homo sapiens secretoglobin, family 3A, member 2 (SCGB3A2), mRNA [NM_054023] |
| A_23_P251412 | 1,22E-04 | down | -3,1759095 | SCGN | Homo sapiens secretagogin, EF-hand calcium binding protein (SCGN), mRNA [NM_006998] |
| A_33_P3332126 | 7,31E-04 | down | -1,6246885 | SCLY | selenocysteine lyase [Source:HGNC Symbol;Acc:HGNC:18161] [ENST00000409736] |
| A_33_P3319987 | 0,003056902 | up | 1,6238405 | SCML4 | Homo sapiens sex comb on midleg-like 4 (Drosophila) (SCML4), transcript variant 1, mRNA [NM_198081] |
| A_32_P187571 | 0,001238136 | down | -3,9577546 | SCN2B | Homo sapiens sodium channel, voltage gated, type II beta subunit (SCN2B), mRNA [NM_004588] |
| A_23_P67896 | 0,022746904 | down | -1,7484857 | SCN3A | Homo sapiens sodium channel, voltage gated, type III alpha subunit (SCN3A), transcript variant 1, mRNA [NM_006922] |
| A_24_P270999 | 5,35E-06 | down | -3,1011395 | SCN4A | Homo sapiens sodium channel, voltage gated, type IV alpha subunit (SCN4A), mRNA [NM_000334] |
| A_33_P3391455 | 0,028006103 | down | -1,6212206 | SCN5A | Homo sapiens sodium channel, voltage gated, type V alpha subunit (SCN5A), transcript variant 6, mRNA [NM_001160161] |
| A_33_P3408962 | 0,017416304 | down | -1,6627618 | SCN8A | Homo sapiens sodium channel, voltage gated, type VIII alpha subunit (SCN8A), transcript variant 1, mRNA [NM_014191] |
| A_24_P3005 | 0,014355835 | down | -3,53593 | SCN9A | Homo sapiens sodium channel, voltage gated, type IX alpha subunit (SCN9A), mRNA [NM_002977] |
| A_23_P128323 | 0,003147691 | up | 2,2939777 | SCNN1A | Homo sapiens sodium channel, non voltage gated 1 alpha subunit (SCNN1A), transcript variant 1, mRNA [NM_001038] |
| A_23_P46412 | 0,027054822 | down | -1,6049262 | SCNN1D | Homo sapiens sodium channel, non voltage gated 1 delta subunit (SCNN1D), transcript variant 1, mRNA [NM_001130413] |
| A_23_P206626 | 1,36E-05 | up | 4,2348213 | SCNN1G | Homo sapiens sodium channel, non voltage gated 1 gamma subunit (SCNN1G), mRNA [NM_001039] |
| A_24_P46334 | 8,45E-04 | down | -1,7333561 | SCOC | Homo sapiens short coiled-coil protein (SCOC), transcript variant 4, mRNA [NM_032547] |
| A_33_P3285893 | 0,04730037 | down | -1,5847619 | SCRN1 | Homo sapiens secernin 1 (SCRN1), transcript variant 1, mRNA [NM_001145513] |
| A_33_P3235137 | 0,003217722 | up | 1,5633991 | SCRT1 | Homo sapiens scratch family zinc finger 1 (SCRT1), mRNA [NM_031309] |
| A_33_P3252974 | 0,008450981 | up | 1,5897673 | SCRT2 | Homo sapiens scratch family zinc finger 2 (SCRT2), mRNA [NM_033129] |
| A_33_P3417487 | 0,007327869 | down | -2,2266247 | SCUBE1 | signal peptide, CUB domain, EGF-like 1 [Source:HGNC Symbol;Acc:HGNC:13441] [ENST00000290460] |
| A_24_P943894 | 0,042127144 | down | -3,035487 | SCUBE3 | Homo sapiens signal peptide, CUB domain, EGF-like 3 (SCUBE3), transcript variant 1, mRNA [NM_152753] |
| A_33_P3333712 | 0,006511398 | up | 1,7037336 | SCYL2 | Homo sapiens SCY1-like 2 (S. cerevisiae) (SCYL2), mRNA [NM_017988] |
| A_33_P3241884 | 0,001296546 | down | -2,3102038 | SDC3 | Homo sapiens syndecan 3 (SDC3), mRNA [NM_014654] |

| | | | | | |
|---|---|---|---|---|---|
| A_21_P0000891 | 0,003087163 | down | -2,2557697 | SDCBP2-AS1 | Homo sapiens SDCBP2 antisense RNA 1 (SDCBP2-AS1), transcript variant 1, long non-coding RNA [NR_040047] |
| A_23_P6344 | 0,008901294 | down | -2,300249 | SDF2L1 | Homo sapiens stromal cell-derived factor 2-like 1 (SDF2L1), mRNA [NM_022044] |
| A_32_P67259 | 4,66E-04 | up | 1,5742977 | SDHA | Homo sapiens succinate dehydrogenase complex, subunit A, flavoprotein (Fp) (SDHA), transcript variant 1, mRNA [NM_004168] |
| A_23_P2645 | 0,024030855 | down | -1,6327813 | SDS | Homo sapiens serine dehydratase (SDS), mRNA [NM_006843] |
| A_33_P3227472 | 9,74E-04 | down | -2,3618214 | SDSL | Homo sapiens serine dehydratase-like (SDSL), mRNA [NM_138432] |
| A_33_P3309506 | 6,68E-04 | down | -2,922057 | SEBOX | Homo sapiens SEBOX homeobox (SEBOX), mRNA [NM_001080837] |
| A_19_P00320158 | 0,005382252 | down | -1,7128203 | SEC14L1 | Homo sapiens SEC14-like 1 (S. cerevisiae) (SEC14L1), transcript variant 6, mRNA [NM_001204408] |
| A_33_P3275751 | 0,006275934 | up | 1,6549608 | SEC14L1 | Homo sapiens SEC14-like 1 (S. cerevisiae) (SEC14L1), transcript variant 7, mRNA [NM_001204410] |
| A_33_P3286754 | 0,014752342 | up | 2,2516189 | SEC14L2 | Homo sapiens SEC14-like 2 (S. cerevisiae) (SEC14L2), transcript variant 2, mRNA [NM_033382] |
| A_33_P3418218 | 0,007625056 | up | 2,827233 | SEC16B | Homo sapiens SEC16 homolog B (S. cerevisiae) (SEC16B), mRNA [NM_033127] |
| A_33_P3548860 | 0,015324292 | up | 2,795894 | SEC1P | Homo sapiens secretory blood group 1, pseudogene (SEC1P), non-coding RNA [NR_004401] |
| A_23_P394605 | 0,001062674 | up | 1,7801322 | SEC24A | Homo sapiens SEC24 family member A (SEC24A), transcript variant 1, mRNA [NM_021982] |
| A_33_P3331085 | 0,0090387 | up | 1,521197 | SEC24A | Homo sapiens SEC24 family member A (SEC24A), transcript variant 1, mRNA [NM_021982] |
| A_33_P3331080 | 0,04827163 | up | 1,5309978 | SEC24A | Homo sapiens SEC24 family member A (SEC24A), transcript variant 2, mRNA [NM_001252231] |
| A_23_P69683 | 6,01E-05 | up | 1,9504158 | SEC24B | Homo sapiens SEC24 family member B (SEC24B), transcript variant 1, mRNA [NM_006323] |
| A_24_P82135 | 0,036719617 | up | 1,6079967 | SECISBP2L | Homo sapiens SECIS binding protein 2-like (SECISBP2L), transcript variant 2, mRNA [NM_014701] |
| A_24_P48204 | 0,004966251 | up | 2,7164092 | SECTM1 | Homo sapiens secreted and transmembrane 1 (SECTM1), mRNA [NM_003004] |
| A_33_P3275968 | 0,049908124 | down | -1,9121857 | SEL1L | Homo sapiens sel-1 suppressor of lin-12-like (C. elegans) (SEL1L), transcript variant 1, mRNA [NM_005065] |
| A_33_P3341796 | 0,011339321 | up | 1,56853 | SEL1L2 | Homo sapiens sel-1 suppressor of lin-12-like 2 (C. elegans) (SEL1L2), transcript variant 2, non-coding RNA [NR_073206] |
| A_23_P6413 | 0,007122965 | down | -1,6397164 | SELM | Homo sapiens selenoprotein M (SELM), mRNA [NM_080430] |
| A_33_P3339100 | 0,007566612 | down | -1,8293401 | SELP | Homo sapiens selectin P (granule membrane protein 140kDa, antigen CD62) (SELP), mRNA [NM_003005] |
| A_33_P3290667 | 0,003617296 | up | 1,9427953 | SELT | Homo sapiens selenoprotein T (SELT), mRNA [NM_016275] |
| A_33_P3212500 | 0,023072535 | down | -2,7920227 | SEMA3F-AS1 | Homo sapiens cDNA FLJ46043 fis, clone SPLEN2042051. [AK127934] |
| A_24_P261169 | 0,016342336 | up | 2,7707102 | SEMA4D | Homo sapiens sema domain, immunoglobulin domain (Ig), transmembrane domain (TM) and short cytoplasmic domain, (semaphorin) 4D (SEMA4D), transcript variant 1, mRNA [NM_00637 |
| A_33_P3697530 | 0,004191086 | up | 2,1787066 | SEMA4D | Homo sapiens sema domain, immunoglobulin domain (Ig), transmembrane domain (TM) and short cytoplasmic domain, (semaphorin) 4D (SEMA4D), transcript variant 2, mRNA [NM_00114 |
| A_23_P121374 | 0,04044178 | up | 1,8364282 | SEMA5B | Homo sapiens sema domain, seven thrombospondin repeats (type 1 and type 1-like), transmembrane domain (TM) and short cytoplasmic domain, (semaphorin) 5B (SEMA5B), transcript va |
| A_24_P213478 | 0,04866794 | up | 3,3118303 | SEMA6A | Homo sapiens sema domain, transmembrane domain (TM), and cytoplasmic domain, (semaphorin) 6A (SEMA6A), transcript variant 2, mRNA [NM_020796] |
| A_33_P3263651 | 4,41E-04 | down | -2,652971 | SEMA6B | Homo sapiens sema domain, transmembrane domain (TM), and cytoplasmic domain, (semaphorin) 6B (SEMA6B), mRNA [NM_032108] |
| A_33_P3258265 | 0,009179452 | up | 1,5066835 | SEMA6C | Homo sapiens sema domain, transmembrane domain (TM), and cytoplasmic domain, (semaphorin) 6C (SEMA6C), transcript variant 2, mRNA [NM_030913] |
| A_23_P164000 | 0,011679118 | up | 1,5801141 | SENP3 | Homo sapiens SUMO1/sentrin/SMT3 specific peptidase 3 (SENP3), mRNA [NM_015670] |
| A_23_P211572 | 0,0383062 | up | 2,549837 | SEP3 | Homo sapiens septin 3 (SEPT3), transcript variant A, mRNA [NM_145733] |
| A_33_P3371425 | 6,67E-04 | down | -1,9801669 | SEPN1 | Homo sapiens selenoprotein N, 1 (SEPN1), transcript variant 1, mRNA [NM_020451] |
| A_23_P121926 | 0,044758007 | up | 1,6688346 | SEPP1 | Homo sapiens selenoprotein P, plasma, 1 (SEPP1), transcript variant 1, mRNA [NM_005410] |
| A_19_P00801735 | 1,61E-04 | down | -2,2395895 | SEPT7-AS1 | SEPT7 antisense RNA 1 (head to head) [Source:HGNC Symbol;Acc:HGNC:51153] [ENST00000437235] |
| A_19_P00318803 | 0,006127265 | down | -1,811515 | SEPT7-AS1 | Homo sapiens SEPT7 antisense RNA 1 (head to head) (SEPT7-AS1), transcript variant 2, long non-coding RNA [NR_120512] |
| A_33_P3418576 | 0,022532647 | up | 1,5294709 | SERAC1 | Homo sapiens serine active site containing 1 (SERAC1), transcript variant 2, non-coding RNA [NR_073096] |
| A_23_P120953 | 0,002958499 | down | -1,5509462 | SERHL2 | Homo sapiens serine hydrolase-like 2 (SERHL2), transcript variant 1, mRNA [NM_014509] |
| A_24_P340696 | 0,012519699 | down | -1,8857526 | SERHL2 | Homo sapiens serine hydrolase-like 2 (SERHL2), transcript variant 1, mRNA [NM_014509] |
| A_33_P3403936 | 0,012878682 | down | -1,9594 | SERINC4 | Homo sapiens serine incorporator 4 (SERINC4), transcript variant 1, mRNA [NM_001258031] |
| A_23_P423457 | 0,004363694 | down | -1,6536953 | SERINC5 | Homo sapiens serine incorporator 5 (SERINC5), transcript variant 2, mRNA [NM_178276] |
| A_33_P3262470 | 3,49E-04 | down | -1,9260278 | SERP2 | stress-associated endoplasmic reticulum protein family member 2 [Source:HGNC Symbol;Acc:HGNC:20607] [ENST00000493476] |
| A_33_P3262475 | 0,027158674 | up | 2,238935 | SERP2 | stress-associated endoplasmic reticulum protein family member 2 [Source:HGNC Symbol;Acc:HGNC:20607] [ENST00000474333] |
| A_23_P128759 | 0,002204092 | up | 3,199516 | SERPINA10 | Homo sapiens serpin peptidase inhibitor, clade A (alpha-1 antiproteinase, antitrypsin), member 10 (SERPINA10), transcript variant 1, mRNA [NM_016186] |
| A_33_P3399788 | 0,005280896 | up | 2,8176792 | SERPINA3 | Homo sapiens serpin peptidase inhibitor, clade A (alpha-1 antiproteinase, antitrypsin), member 3 (SERPINA3), mRNA [NM_001085] |
| A_23_P2920 | 0,00475104 | up | 3,35018 | SERPINA3 | Homo sapiens serpin peptidase inhibitor, clade A (alpha-1 antiproteinase, antitrypsin), member 3 (SERPINA3), mRNA [NM_001085] |
| A_23_P162918 | 0,009459312 | up | 2,555194 | SERPINA3 | Homo sapiens serpin peptidase inhibitor, clade A (alpha-1 antiproteinase, antitrypsin), member 3 (SERPINA3), mRNA [NM_001085] |
| A_24_P321766 | 0,007945741 | up | 1,561504 | SERPINA5 | Homo sapiens serpin peptidase inhibitor, clade A (alpha-1 antiproteinase, antitrypsin), member 5 (SERPINA5), mRNA [NM_000624] |
| A_23_P162861 | 0,001057892 | up | 1,8747435 | SERPINE3 | Homo sapiens serpin peptidase inhibitor, clade E (nexin, plasminogen activator inhibitor type 1), member 3 (SERPINE3), mRNA [NM_001101320] |
| A_33_P3413993 | 0,032133173 | down | -1,6603845 | SERPING1 | serpin peptidase inhibitor, clade G (C1 inhibitor), member 1 [Source:HGNC Symbol;Acc:HGNC:1228] [ENST00000405496] |
| A_23_P141960 | 0,001749096 | up | 1,5310022 | SERTAD3 | Homo sapiens SERTA domain containing 3 (SERTAD3), transcript variant 2, mRNA [NM_203344] |
| A_23_P318904 | 4,77E-04 | up | 2,1115363 | SERTAD4 | Homo sapiens SERTA domain containing 4 (SERTAD4), mRNA [NM_019605] |
| A_23_P361448 | 0,018529283 | down | -1,7979342 | SESN3 | Homo sapiens sestrin 3 (SESN3), transcript variant 1, mRNA [NM_144665] |
| A_33_P3378047 | 0,009061265 | up | 1,5662044 | SESTD1 | Homo sapiens SEC14 and spectrin domains 1 (SESTD1), mRNA [NM_178123] |
| A_24_P917866 | 0,030245619 | down | -2,6354187 | SET | Homo sapiens SET nuclear proto-oncogene (SET), transcript variant 2, mRNA [NM_003011] |
| A_23_P319895 | 2,07E-04 | up | 2,3630521 | SETD1B | Homo sapiens SET domain containing 1B (SETD1B), mRNA [NM_015048] |
| A_23_P44760 | 2,03E-04 | up | 1,8022878 | SETD2 | Homo sapiens SET domain containing 2 (SETD2), mRNA [NM_014159] |
| A_33_P3315355 | 0,011300369 | up | 1,6037974 | SETD6 | Homo sapiens SET domain containing 6 (SETD6), transcript variant 1, mRNA [NM_001160305] |
| A_32_P191527 | 0,002224782 | up | 1,5663991 | SETD8 | Homo sapiens SET domain containing (lysine methyltransferase) 8 (SETD8), mRNA [NM_020382] |
| A_32_P191859 | 2,74E-05 | up | 2,059747 | SETD8 | Homo sapiens SET domain containing (lysine methyltransferase) 8 (SETD8), mRNA [NM_020382] |
| A_33_P3353916 | 0,015137932 | down | -1,6837811 | SETMAR | Homo sapiens SET domain and mariner transposase fusion gene (SETMAR), transcript variant 4, mRNA [NM_001276325] |
| A_24_P235783 | 0,001490867 | up | 1,6836509 | SF1 | splicing factor 1 [Source:HGNC Symbol;Acc:HGNC:12950] [ENST00000463343] |

| | | | | | |
|---|---|---|---|---|---|
| A_23_P135914 | 0,001693261 | up | 1,5202764 | SF3B3 | Homo sapiens splicing factor 3b, subunit 3, 130kDa (SF3B3), mRNA [NM_012426] |
| A_33_P3474175 | 5,07E-04 | down | -3,1075985 | SFTA1P | AGENCOURT_10018270 NIH_MGC_142 Homo sapiens cDNA clone IMAGE:6495000 5', mRNA sequence [BU601128] |
| A_33_P3344574 | 2,55E-04 | down | -2,2237048 | SFTPA2 | Homo sapiens surfactant protein A2 (SFTPA2), mRNA [NM_001098668] |
| A_33_P3268919 | 0,008836153 | up | 1,6171151 | SFTPA2 | Homo sapiens surfactant protein A2 (SFTPA2), mRNA [NM_001098668] |
| A_33_P3275873 | 0,001297309 | up | 2,5635893 | SFTPB | Homo sapiens surfactant protein B (SFTPB), transcript variant 1, mRNA [NM_000542] |
| A_33_P3292896 | 0,004012996 | down | -1,7495035 | SFXN5 | Homo sapiens sideroflexin 5 (SFXN5), mRNA [NM_144579] |
| A_23_P62881 | 0,005706027 | down | -2,4018383 | SGIP1 | Homo sapiens SH3-domain GRB2-like (endophilin) interacting protein 1 (SGIP1), mRNA [NM_032291] |
| A_23_P19673 | 0,010832625 | up | 1,7149177 | SGK1 | Homo sapiens serum/glucocorticoid regulated kinase 1 (SGK1), transcript variant 1, mRNA [NM_005627] |
| A_23_P415633 | 0,040135864 | up | 1,5395622 | SGK494 | Homo sapiens uncharacterized serine/threonine-protein kinase SgK494 (SGK494), mRNA [NM_001174103] |
| A_21_P0010830 | 0,003583453 | down | -2,195242 | SGMS1-AS1 | Homo sapiens SGMS1 antisense RNA 1 (SGMS1-AS1), long non-coding RNA [NR_126411] |
| A_33_P3283619 | 0,029520031 | up | 1,618403 | SH2D1A | Homo sapiens SH2 domain containing 1A (SH2D1A), transcript variant 2, mRNA [NM_001114937] |
| A_33_P3283616 | 0,014358903 | up | 1,5350711 | SH2D1A | Homo sapiens SH2 domain containing 1A (SH2D1A), transcript variant 1, mRNA [NM_002351] |
| A_33_P3254320 | 0,012193667 | up | 1,6209673 | SH2D3A | Homo sapiens SH2 domain containing 3A (SH2D3A), mRNA [NM_005490] |
| A_33_P3357199 | 0,022010054 | down | -1,7025766 | SH2D6 | Homo sapiens SH2 domain containing 6 (SH2D6), mRNA [NM_198482] |
| A_33_P3313730 | 4,40E-05 | up | 2,2420454 | SH3BP2 | Homo sapiens SH3-domain binding protein 2 (SH3BP2), transcript variant 4, mRNA [NM_001145855] |
| A_32_P24585 | 0,00704676 | up | 1,5628643 | SH3PXD2B | Homo sapiens SH3 and PX domains 2B (SH3PXD2B), mRNA [NM_001017995] |
| A_21_P0000575 | 0,023590425 | up | 1,5994538 | SH3RF3-AS1 | Homo sapiens SH3RF3 antisense RNA 1 (SH3RF3-AS1), long non-coding RNA [NR_029193] |
| A_33_P3399443 | 0,019579383 | down | -2,7115078 | SH3TC1 | SH3 domain and tetratricopeptide repeats 1 [Source:HGNC Symbol;Acc:HGNC:26009] [ENST00000502350] |
| A_23_P41390 | 6,43E-04 | up | 1,8494447 | SH3TC1 | Homo sapiens SH3 domain and tetratricopeptide repeats 1 (SH3TC1), mRNA [NM_018986] |
| A_33_P3399448 | 1,05E-04 | up | 3,1648664 | SH3TC1 | Homo sapiens SH3 domain and tetratricopeptide repeats 1, mRNA (cDNA clone IMAGE:6291103), complete cds. [BC068094] |
| A_32_P305888 | 1,34E-04 | up | 2,155385 | SH3TC2 | Homo sapiens SH3 domain and tetratricopeptide repeats 2 (SH3TC2), mRNA [NM_024577] |
| A_33_P3244249 | 0,011738223 | up | 2,5555139 | SH3TC2 | SH3 domain and tetratricopeptide repeats 2 [Source:HGNC Symbol;Acc:HGNC:29427] [ENST00000513604] |
| A_23_P208636 | 4,76E-05 | down | -3,1542659 | SHANK1 | Homo sapiens SH3 and multiple ankyrin repeat domains 1 (SHANK1), mRNA [NM_016148] |
| A_23_P424727 | 9,57E-04 | down | -2,3812509 | SHANK2 | SH3 and multiple ankyrin repeat domains 2 [Source:HGNC Symbol;Acc:HGNC:14295] [ENST00000409530] |
| A_21_P0000018 | 6,18E-04 | down | -3,828885 | SHBG | Homo sapiens sex hormone-binding globulin (SHBG), transcript variant 4, mRNA [NM_001146281] |
| A_23_P142255 | 0,001616521 | down | -1,7924905 | SHD | Homo sapiens Src homology 2 domain containing transforming protein D (SHD), mRNA [NM_020209] |
| A_23_P111657 | 0,001662006 | down | -2,3912683 | SHH | Homo sapiens sonic hedgehog (SHH), mRNA [NM_000193] |
| A_23_P115573 | 4,24E-06 | down | -2,5952423 | SHISA4 | Homo sapiens shisa family member 4 (SHISA4), transcript variant 1, mRNA [NM_198149] |
| A_24_P161581 | 2,80E-05 | down | -2,5467992 | SHISA7 | Homo sapiens shisa family member 7 (SHISA7), mRNA [NM_001145176] |
| A_23_P390443 | 0,0108852 | down | -2,3838296 | SHISA7 | Homo sapiens shisa family member 7 (SHISA7), mRNA [NM_001145176] |
| A_33_P3222932 | 5,12E-05 | down | -3,0041356 | SHISA8 | Homo sapiens shisa family member 8 (SHISA8), mRNA [NM_001207020] |
| A_23_P22761 | 1,62E-06 | down | -6,516969 | SHOX | Homo sapiens short stature homeobox (SHOX), transcript variant 2, mRNA [NM_006883] |
| A_23_P337790 | 9,63E-04 | up | 1,5484964 | SHPRH | Homo sapiens SNF2 histone linker PHD RING helicase, E3 ubiquitin protein ligase (SHPRH), transcript variant 1, mRNA [NM_001042683] |
| A_33_P3236133 | 0,003592285 | down | -2,11902 | SHQ1 | Homo sapiens SHQ1, H/ACA ribonucleoprotein assembly factor (SHQ1), mRNA [NM_018130] |
| A_23_P217755 | 0,001245775 | up | 1,6016028 | SHROOM2 | Homo sapiens shroom family member 2 (SHROOM2), mRNA [NM_001649] |
| A_33_P3417339 | 0,026200853 | up | 1,585902 | SHROOM3 | Homo sapiens shroom family member 3 (SHROOM3), mRNA [NM_020859] |
| A_33_P3336700 | 0,004918368 | up | 1,7366697 | SHROOM3 | Homo sapiens shroom family member 3 (SHROOM3), mRNA [NM_020859] |
| A_33_P3382493 | 0,006574431 | down | -2,0962715 | SIGLEC14 | Homo sapiens sialic acid binding Ig-like lectin 14 (SIGLEC14), mRNA [NM_001098612] |
| A_23_P50146 | 0,002656308 | down | -2,0516112 | SIGLEC15 | Homo sapiens sialic acid binding Ig-like lectin 15 (SIGLEC15), mRNA [NM_213602] |
| A_23_P4592 | 0,001608975 | down | -2,6801093 | SIGLEC6 | Homo sapiens sialic acid binding Ig-like lectin 6 (SIGLEC6), transcript variant 1, mRNA [NM_001245] |
| A_23_P132121 | 0,022151582 | up | 1,8144488 | SIK1 | Homo sapiens salt-inducible kinase 1 (SIK1), mRNA [NM_173354] |
| A_24_P329924 | 0,006482645 | up | 1,5371883 | SIK3 | Homo sapiens SIK family kinase 3 (SIK3), transcript variant 1, mRNA [NM_025164] |
| A_33_P3341586 | 0,014554692 | down | -1,7293783 | SIL1 | Homo sapiens SIL1 nucleotide exchange factor (SIL1), transcript variant 1, mRNA [NM_001037633] |
| A_23_P214743 | 0,002351229 | down | -3,2233477 | SIM1 | Homo sapiens single-minded family bHLH transcription factor 1 (SIM1), mRNA [NM_005068] |
| A_33_P3225230 | 0,00424071 | up | 1,5198897 | SIN3B | Homo sapiens SIN3 transcription regulator family member B (SIN3B), transcript variant 1, mRNA [NM_015260] |
| A_23_P17456 | 1,52E-04 | down | -3,4818482 | SIRPB1 | Homo sapiens signal-regulatory protein beta 1 (SIRPB1), transcript variant 1, mRNA [NM_006065] |
| A_33_P3356577 | 0,002880717 | down | -2,9005287 | SIRPB1 | signal-regulatory protein beta 1 [Source:HGNC Symbol;Acc:HGNC:15928] [ENST00000381596] |
| A_33_P3248072 | 0,010667515 | down | -1,5954688 | SIRPB1 | Homo sapiens signal-regulatory protein beta 1 (SIRPB1), transcript variant 3, mRNA [NM_001135844] |
| A_33_P3272260 | 8,88E-04 | up | 1,6981682 | SIRPB2 | Homo sapiens signal-regulatory protein beta 2 (SIRPB2), transcript variant 1, mRNA [NM_001122962] |
| A_23_P307955 | 0,02712265 | up | 1,7401056 | SIRT5 | Homo sapiens sirtuin 5 (SIRT5), transcript variant 2, mRNA [NM_031244] |
| A_24_P34611 | 3,03E-04 | down | -1,9670649 | SIX3 | Homo sapiens SIX homeobox 3 (SIX3), mRNA [NM_005413] |
| A_23_P106117 | 0,040168803 | down | -1,8610141 | SIX6 | Homo sapiens SIX homeobox 6 (SIX6), mRNA [NM_007374] |
| A_24_P322354 | 0,021019215 | up | 1,7241764 | SKA1 | Homo sapiens spindle and kinetochore associated complex subunit 1 (SKA1), transcript variant 1, mRNA [NM_001039535] |
| A_33_P3421331 | 0,0017447 | up | 3,0815234 | SKOR2 | Homo sapiens SKI family transcriptional corepressor 2 (SKOR2), transcript variant 1, mRNA [NM_001278063] |
| A_23_P156310 | 0,012596509 | down | -1,632657 | SKP2 | Homo sapiens S-phase kinase-associated protein 2, E3 ubiquitin protein ligase (SKP2), transcript variant 2, mRNA [NM_032637] |
| A_23_P62647 | 0,026625637 | down | -1,5110255 | SLAMF1 | Homo sapiens signaling lymphocytic activation molecule family member 1 (SLAMF1), transcript variant 1, mRNA [NM_003037] |
| A_24_P45451 | 0,006203731 | up | 1,8502245 | SLAMF6 | Homo sapiens SLAM family member 6 (SLAMF6), transcript variant 2, mRNA [NM_052931] |
| A_24_P353638 | 0,036979973 | down | -1,6753854 | SLAMF7 | Homo sapiens SLAM family member 7 (SLAMF7), transcript variant 1, mRNA [NM_021181] |
| A_23_P200138 | 0,040580712 | down | -1,9198127 | SLAMF8 | Homo sapiens SLAM family member 8 (SLAMF8), mRNA [NM_020125] |
| A_33_P3323048 | 0,013185269 | down | -1,7119234 | SLC10A7 | Homo sapiens solute carrier family 10, member 7 (SLC10A7), transcript variant 2, mRNA [NM_001029998] |
| A_33_P3357516 | 0,001148378 | down | -2,4897714 | SLC10A7 | Homo sapiens solute carrier family 10, member 7 (SLC10A7), transcript variant 3, mRNA [NM_032128] |

| | | | | | |
|---|---|---|---|---|---|
| A_33_P3702055 | 0,001109539 | down | -1,8588471 | SLC11A1 | Homo sapiens solute carrier family 11 (proton-coupled divalent metal ion transporter), member 1 (SLC11A1), mRNA [NM_000578] |
| A_33_P3212112 | 0,003545 | up | 1,6816386 | SLC12A6 | Homo sapiens solute carrier family 12 (potassium/chloride transporter), member 6 (SLC12A6), transcript variant 5, mRNA [NM_001042496] |
| A_33_P3357530 | 0,02232672 | up | 1,6010079 | SLC12A7 | Homo sapiens solute carrier family 12 (potassium/chloride transporter), member 7 (SLC12A7), mRNA [NM_006598] |
| A_33_P3298861 | 0,002366021 | up | 1,7623019 | SLC12A8 | Homo sapiens solute carrier family 12, member 8 (SLC12A8), transcript variant 1, mRNA [NM_024628] |
| A_33_P3210399 | 0,015078853 | down | -1,7171017 | SLC14A1 | Homo sapiens solute carrier family 14 (urea transporter), member 1 (Kidd blood group) (SLC14A1), transcript variant 4, mRNA [NM_001146037] |
| A_24_P926507 | 0,006828318 | up | 2,66991 | SLC14A1 | Homo sapiens solute carrier family 14 (urea transporter), member 1 (Kidd blood group) (SLC14A1), transcript variant 4, mRNA [NM_001146037] |
| A_23_P27353 | 0,001830024 | down | -3,9389603 | SLC14A2 | Homo sapiens solute carrier family 14 (urea transporter), member 2 (SLC14A2), transcript variant 1, mRNA [NM_007163] |
| A_24_P13311 | 0,008073097 | up | 1,5530165 | SLC15A4 | Homo sapiens solute carrier family 15 (oligopeptide transporter), member 4 (SLC15A4), mRNA [NM_145648] |
| A_21_P0001002 | 0,04539248 | up | 1,6861295 | SLC16A1-AS1 | Homo sapiens SLC16A1 antisense RNA 1 (SLC16A1-AS1), transcript variant 2, long non-coding RNA [NR_103744] |
| A_23_P363313 | 0,021144677 | down | -1,701725 | SLC16A11 | Homo sapiens solute carrier family 16, member 11 (SLC16A11), mRNA [NM_153357] |
| A_33_P3227920 | 0,004983796 | down | -2,0600548 | SLC16A4 | Homo sapiens solute carrier family 16, member 4 (SLC16A4), transcript variant 1, mRNA [NM_004696] |
| A_32_P130630 | 0,04545641 | up | 2,1444921 | SLC16A7 | Homo sapiens solute carrier family 16 (monocarboxylate transporter), member 7 (SLC16A7), transcript variant 3, mRNA [NM_004731] |
| A_33_P3353365 | 1,31E-05 | down | -5,0477033 | SLC17A4 | solute carrier family 17, member 4 [Source:HGNC Symbol;Acc:HGNC:10932] [ENST00000397076] |
| A_33_P3379926 | 0,012111228 | up | 1,6135839 | SLC17A4 | Homo sapiens solute carrier family 17, member 4 (SLC17A4), transcript variant 1, mRNA [NM_005495] |
| A_33_P3403162 | 0,008806555 | down | -1,7370644 | SLC18A1 | Homo sapiens solute carrier family 18 (vesicular monoamine transporter), member 1 (SLC18A1), transcript variant 4, mRNA [NM_001142325] |
| A_23_P160466 | 0,002230071 | up | 1,6464821 | SLC19A2 | Homo sapiens solute carrier family 19 (thiamine transporter), member 2 (SLC19A2), mRNA [NM_006996] |
| A_23_P325562 | 0,015175088 | down | -1,5034018 | SLC1A7 | Homo sapiens solute carrier family 1 (glutamate transporter), member 7 (SLC1A7), transcript variant 4, mRNA [NM_006671] |
| A_33_P3386835 | 0,00521884 | up | 1,653389 | SLC20A2 | solute carrier family 20 (phosphate transporter), member 2 [Source:HGNC Symbol;Acc:HGNC:10947] [ENST00000523340] |
| A_23_P161968 | 0,006746225 | down | -2,3777614 | SLC22A10 | Homo sapiens solute carrier family 22, member 10 (SLC22A10), mRNA [NM_001039752] |
| A_23_P47199 | 0,0079073 | down | -2,857017 | SLC22A11 | Homo sapiens solute carrier family 22 (organic anion/urate transporter), member 11 (SLC22A11), mRNA [NM_018484] |
| A_23_P144123 | 0,01600522 | up | 1,8993189 | SLC22A13 | Homo sapiens solute carrier family 22 (organic anion/urate transporter), member 13 (SLC22A13), mRNA [NM_004256] |
| A_33_P3361412 | 0,001091631 | down | -1,6954598 | SLC22A15 | Homo sapiens solute carrier family 22, member 15 (SLC22A15), mRNA [NM_018420] |
| A_23_P42004 | 0,02553769 | up | 2,0650787 | SLC22A16 | Homo sapiens solute carrier family 22 (organic cation/carnitine transporter), member 16 (SLC22A16), mRNA [NM_033125] |
| A_23_P111395 | 6,49E-07 | down | -4,483563 | SLC22A2 | Homo sapiens solute carrier family 22 (organic cation transporter), member 2 (SLC22A2), mRNA [NM_003058] |
| A_24_P350228 | 0,015793215 | down | -1,5329905 | SLC22A23 | Homo sapiens solute carrier family 22, member 23 (SLC22A23), transcript variant 2, mRNA [NM_021945] |
| A_21_P0011492 | 0,01663163 | down | -1,6219758 | SLC22A31 | PREDICTED: Homo sapiens solute carrier family 22, member 31 (SLC22A31), transcript variant X3, mRNA [XM_006721147] |
| A_33_P3294603 | 0,001134546 | down | -1,8143308 | SLC22A31 | Homo sapiens solute carrier family 22, member 31 (SLC22A31), mRNA [NM_001242757] |
| A_24_P174755 | 0,045434557 | up | 1,6351558 | SLC22A5 | Homo sapiens solute carrier family 22 (organic cation/carnitine transporter), member 5 (SLC22A5), mRNA [NM_003060] |
| A_33_P3367171 | 0,015672782 | down | -2,239598 | SLC22A8 | solute carrier family 22 (organic anion transporter), member 8 [Source:HGNC Symbol;Acc:HGNC:10972] [ENST00000451262] |
| A_24_P254278 | 2,32E-04 | up | 1,7519188 | SLC23A2 | Homo sapiens solute carrier family 23 (ascorbic acid transporter), member 2 (SLC23A2), transcript variant 2, mRNA [NM_203327] |
| A_23_P28246 | 0,006720792 | up | 1,6342474 | SLC23A3 | Homo sapiens solute carrier family 23, member 3 (SLC23A3), transcript variant 1, mRNA [NM_144712] |
| A_23_P503010 | 0,008530051 | down | -2,180445 | SLC24A4 | Homo sapiens solute carrier family 24 (sodium/potassium/calcium exchanger), member 4 (SLC24A4), transcript variant 1, mRNA [NM_153646] |
| A_33_P3387901 | 0,001848083 | down | -1,9712765 | SLC25A19 | Homo sapiens solute carrier family 25 (mitochondrial thiamine pyrophosphate carrier), member 19 (SLC25A19), transcript variant 1, mRNA [NM_001126121] |
| A_21_P0000084 | 0,021580437 | up | 1,7123264 | SLC25A21 | Homo sapiens solute carrier family 25 (mitochondrial oxoadipate carrier), member 21 (SLC25A21), transcript variant 1, mRNA [NM_030631] |
| A_21_P0006396 | 4,86E-05 | down | -2,3872218 | SLC25A25-AS1 | Homo sapiens SLC25A25 antisense RNA 1 (SLC25A25-AS1), long non-coding RNA [NR_033374] |
| A_24_P246841 | 3,46E-05 | down | -3,9072227 | SLC25A27 | Homo sapiens solute carrier family 25, member 27 (SLC25A27), transcript variant 1, mRNA [NM_004277] |
| A_23_P77048 | 0,040437292 | down | -1,5081564 | SLC25A29 | Homo sapiens solute carrier family 25 (mitochondrial carnitine/acylcarnitine carrier), member 29 (SLC25A29), transcript variant 1, mRNA [NM_001039355] |
| A_24_P64100 | 0,02769919 | up | 1,8741988 | SLC25A37 | solute carrier family 25 (mitochondrial iron transporter), member 37 [Source:HGNC Symbol;Acc:HGNC:29786] [ENST00000519973] |
| A_33_P3343210 | 0,027331894 | up | 1,5988423 | SLC25A37 | Homo sapiens solute carrier family 25 (mitochondrial iron transporter), member 37 (SLC25A37), mRNA [NM_016612] |
| A_33_P3309365 | 0,005677377 | down | -2,2763224 | SLC25A3P1 | Homo sapiens solute carrier family 25 (mitochondrial carrier; phosphate carrier), member 3 pseudogene 1 (SLC25A3P1), non-coding RNA [NR_002314] |
| A_23_P64070 | 0,02942348 | down | -1,8379774 | SLC25A45 | Homo sapiens solute carrier family 25, member 45 (SLC25A45), transcript variant 1, mRNA [NM_182556] |
| A_23_P424051 | 0,031716976 | up | 2,054992 | SLC25A48 | Homo sapiens solute carrier family 25, member 48 (SLC25A48), mRNA [NM_145282] |
| A_33_P3218178 | 0,00113732 | down | -3,5651734 | SLC25A53 | Homo sapiens solute carrier family 25, member 53 (SLC25A53), mRNA [NM_001012755] |
| A_23_P80954 | 0,013982988 | down | -1,7716699 | SLC26A1 | Homo sapiens solute carrier family 26 (anion exchanger), member 1 (SLC26A1), transcript variant 1, mRNA [NM_022042] |
| A_23_P123228 | 0,002864762 | down | -1,8581364 | SLC26A3 | Homo sapiens solute carrier family 26 (anion exchanger), member 3 (SLC26A3), mRNA [NM_000111] |
| A_23_P30950 | 0,007154253 | up | 1,8764818 | SLC26A8 | Homo sapiens solute carrier family 26 (anion exchanger), member 8 (SLC26A8), transcript variant 1, mRNA [NM_052961] |
| A_24_P179816 | 0,004392616 | down | -1,8160311 | SLC27A3 | Homo sapiens solute carrier family 27 (fatty acid transporter), member 3 (SLC27A3), mRNA [NM_024330] |
| A_23_P168551 | 0,003313331 | down | -1,8532997 | SLC29A4 | Homo sapiens solute carrier family 29 (equilibrative nucleoside transporter), member 4 (SLC29A4), transcript variant 1, mRNA [NM_001040661] |
| A_33_P3224190 | 0,009510842 | down | -2,141082 | SLC29A4 | Homo sapiens solute carrier family 29 (equilibrative nucleoside transporter), member 4 (SLC29A4), transcript variant 1, mRNA [NM_001040661] |
| A_33_P3694746 | 0,03243735 | down | -2,3276434 | SLC2A1-AS1 | SLC2A1 antisense RNA 1 [Source:HGNC Symbol;Acc:HGNC:44187] [ENST00000416689] |
| A_33_P3354514 | 0,007108916 | up | 1,8194699 | SLC2A13 | Homo sapiens solute carrier family 2 (facilitated glucose transporter), member 13, mRNA (cDNA clone MGC:48624 IMAGE:5272386), complete cds. [BC047507] |
| A_24_P111054 | 8,56E-04 | down | -3,444315 | SLC2A5 | solute carrier family 2 (facilitated glucose/fructose transporter), member 5 [Source:HGNC Symbol;Acc:HGNC:11010] [ENST00000377414] |
| A_23_P160159 | 0,017002795 | up | 1,6233834 | SLC2A5 | Homo sapiens solute carrier family 2 (facilitated glucose/fructose transporter), member 5 (SLC2A5), transcript variant 1, mRNA [NM_003039] |
| A_33_P3238425 | 0,021308208 | up | 2,2451806 | SLC2A7 | Homo sapiens solute carrier family 2 (facilitated glucose transporter), member 7 (SLC2A7), mRNA [NM_207420] |
| A_23_P213014 | 0,010669073 | down | -1,5450764 | SLC2A9 | Homo sapiens solute carrier family 2 (facilitated glucose transporter), member 9 (SLC2A9), transcript variant 2, mRNA [NM_001001290] |
| A_23_P302568 | 0,027633887 | down | -2,7462046 | SLC30A3 | Homo sapiens solute carrier family 30 (zinc transporter), member 3 (SLC30A3), mRNA [NM_003459] |
| A_24_P342807 | 0,042129297 | down | -2,2207477 | SLC30A6 | Homo sapiens solute carrier family 30 (zinc transporter), member 6 (SLC30A6), transcript variant 2, mRNA [NM_017964] |
| A_23_P22915 | 0,04883822 | down | -1,5949436 | SLC30A7 | Homo sapiens solute carrier family 30 (zinc transporter), member 7 (SLC30A7), transcript variant 1, mRNA [NM_133496] |
| A_24_P563545 | 0,01612536 | up | 1,5294812 | SLC30A7 | Homo sapiens solute carrier family 30 (zinc transporter), member 7 (SLC30A7), transcript variant 1, mRNA [NM_133496] |
| A_33_P3283813 | 2,21E-06 | down | -3,4574656 | SLC30A8 | Homo sapiens solute carrier family 30 (zinc transporter), member 8 (SLC30A8), transcript variant 4, mRNA [NM_001172813] |
| A_23_P217109 | 0,048782542 | up | 1,5038419 | SLC31A2 | Homo sapiens solute carrier family 31 (copper transporter), member 2 (SLC31A2), mRNA [NM_001860] |

| | | | | | |
|---|---|---|---|---|---|
| A_23_P307382 | 8,71E-04 | down | -2,5674372 | SLC32A1 | Homo sapiens solute carrier family 32 (GABA vesicular transporter), member 1 (SLC32A1), mRNA [NM_080552] |
| A_23_P58729 | 0,02645983 | down | -1,6256578 | SLC34A1 | Homo sapiens solute carrier family 34 (type II sodium/phosphate cotransporter), member 1 (SLC34A1), transcript variant 1, mRNA [NM_003052] |
| A_33_P3297468 | 0,002485762 | down | -2,1130145 | SLC34A3 | Homo sapiens solute carrier family 34 (type II sodium/phosphate cotransporter), member 3 (SLC34A3), transcript variant 2, mRNA [NM_001177317] |
| A_23_P255057 | 1,01E-04 | down | -1,980482 | SLC35A5 | Homo sapiens solute carrier family 35, member A5 (SLC35A5), mRNA [NM_017945] |
| A_33_P3358799 | 0,003189898 | down | -1,6214565 | SLC35C2 | Homo sapiens solute carrier family 35 (GDP-fucose transporter), member C2 (SLC35C2), transcript variant 7, mRNA [NM_001281460] |
| A_21_P0007577 | 0,002038199 | up | 2,1795626 | SLC35E3 | solute carrier family 35, member E3 [Source:HGNC Symbol;Acc:HGNC:20864] [ENST00000399333] |
| A_33_P3368506 | 0,010172435 | down | -2,1061807 | SLC38A8 | Homo sapiens solute carrier family 38, member 8 (SLC38A8), mRNA [NM_001080442] |
| A_24_P383356 | 0,02345179 | down | -2,0739799 | SLC39A14 | Homo sapiens solute carrier family 39 (zinc transporter), member 14 (SLC39A14), transcript variant 2, mRNA [NM_015359] |
| A_33_P3418421 | 0,004413955 | up | 2,6601036 | SLC3A1 | Homo sapiens SLC3A1 variant F (SLC3A1) mRNA, complete cds, alternatively spliced. [DQ023516] |
| A_33_P3418417 | 0,003290925 | up | 1,515435 | SLC3A1 | Homo sapiens SLC3A1 variant C (SLC3A1) mRNA, complete cds, alternatively spliced. [DQ023513] |
| A_23_P86195 | 0,034199588 | up | 1,9195367 | SLC44A3 | Homo sapiens solute carrier family 44, member 3 (SLC44A3), transcript variant 2, mRNA [NM_152369] |
| A_32_P223059 | 0,011301065 | up | 1,5945722 | SLC45A1 | Homo sapiens solute carrier family 45, member 1 (SLC45A1), mRNA [NM_001080397] |
| A_33_P3224819 | 0,028564773 | up | 1,6512384 | SLC45A4 | Homo sapiens solute carrier family 45, member 4 (SLC45A4), transcript variant 1, mRNA [NM_001286646] |
| A_24_P142503 | 0,005354099 | down | -2,0312164 | SLC47A1 | Homo sapiens solute carrier family 47 (multidrug and toxin extrusion), member 1 (SLC47A1), mRNA [NM_018242] |
| A_33_P3403399 | 0,02070439 | up | 1,8269117 | SLC47A1 | Homo sapiens solute carrier family 47 (multidrug and toxin extrusion), member 1 (SLC47A1), mRNA [NM_018242] |
| A_23_P77980 | 0,044439267 | down | -1,510532 | SLC4A1 | Homo sapiens solute carrier family 4 (anion exchanger), member 1 (Diego blood group) (SLC4A1), mRNA [NM_000342] |
| A_24_P314786 | 0,041899208 | up | 2,5290847 | SLC4A10 | Homo sapiens solute carrier family 4, sodium bicarbonate transporter, member 10 (SLC4A10), transcript variant 2, mRNA [NM_022058] |
| A_24_P75680 | 7,20E-06 | down | -2,9817424 | SLC4A8 | Homo sapiens solute carrier family 4, sodium bicarbonate cotransporter, member 8 (SLC4A8), transcript variant 4, mRNA [NM_001258402] |
| A_23_P15692 | 0,013870902 | down | -1,9000815 | SLC52A1 | Homo sapiens solute carrier family 52 (riboflavin transporter), member 1 (SLC52A1), transcript variant 2, mRNA [NM_017986] |
| A_24_P934387 | 8,07E-04 | up | 1,8745338 | SLC5A12 | Homo sapiens solute carrier family 5 (sodium/monocarboxylate cotransporter), member 12 (SLC5A12), mRNA [NM_178498] |
| A_33_P3299339 | 0,005541632 | down | -1,8071553 | SLC5A7 | Homo sapiens solute carrier family 5 (sodium/choline cotransporter), member 7 (SLC5A7), mRNA [NM_021815] |
| A_24_P365721 | 0,031639114 | up | 1,9860376 | SLC6A14 | Homo sapiens solute carrier family 6 (amino acid transporter), member 14 (SLC6A14), mRNA [NM_007231] |
| A_24_P820037 | 0,011228663 | down | -3,8411095 | SLC6A17 | Homo sapiens solute carrier family 6 (neutral amino acid transporter), member 17 (SLC6A17), mRNA [NM_001010898] |
| A_33_P3259022 | 0,002609086 | down | -2,0634558 | SLC6A2 | Homo sapiens solute carrier family 6 (neurotransmitter transporter), member 2 (SLC6A2), transcript variant 1, mRNA [NM_001172504] |
| A_23_P152995 | 0,002661013 | down | -2,4936614 | SLC6A4 | Homo sapiens solute carrier family 6 (neurotransmitter transporter), member 4 (SLC6A4), mRNA [NM_001045] |
| A_23_P116624 | 1,98E-05 | down | -2,5294485 | SLC6A5 | Homo sapiens solute carrier family 6 (neurotransmitter transporter), member 5 (SLC6A5), mRNA [NM_004211] |
| A_24_P359191 | 2,14E-05 | down | -2,7234173 | SLC6A6 | Homo sapiens solute carrier family 6 (neurotransmitter transporter), member 6 (SLC6A6), transcript variant 1, mRNA [NM_003043] |
| A_33_P3316539 | 0,027641656 | up | 3,948647 | SLC7A2 | Homo sapiens solute carrier family 7 (cationic amino acid transporter, y+ system), member 2 (SLC7A2), transcript variant 2, mRNA [NM_001008539] |
| A_33_P3310301 | 0,001342012 | down | -2,880075 | SLC7A8 | Homo sapiens solute carrier family 7 (amino acid transporter light chain, L system), member 8 (SLC7A8), transcript variant 1, mRNA [NM_012244] |
| A_33_P3262156 | 0,004728272 | down | -2,2237327 | SLC8A1 | Homo sapiens solute carrier family 8 (sodium/calcium exchanger), member 1 (SLC8A1), transcript variant A, mRNA [NM_021097] |
| A_33_P3267059 | 0,04659394 | up | 1,8846856 | SLC9A1 | solute carrier family 9, subfamily A (NHE1, cation proton antiporter 1), member 1 [Source:HGNC Symbol;Acc:HGNC:11071] [ENST00000374084] |
| A_21_P0011428 | 0,007678522 | down | -3,0934849 | SLC9B1 | Homo sapiens solute carrier family 9, subfamily B (NHA1, cation proton antiporter 1), member 1 (SLC9B1), transcript variant 2, mRNA [NM_001100874] |
| A_32_P69802 | 0,004670263 | up | 1,856972 | SLCO1A2 | Homo sapiens solute carrier organic anion transporter family, member 1A2 (SLCO1A2), transcript variant 1, mRNA [NM_134431] |
| A_23_P116868 | 0,009805758 | up | 1,9067334 | SLCO1C1 | Homo sapiens solute carrier organic anion transporter family, member 1C1 (SLCO1C1), transcript variant 2, mRNA [NM_017435] |
| A_23_P150768 | 0,022071289 | up | 1,5862137 | SLCO2B1 | Homo sapiens solute carrier organic anion transporter family, member 2B1 (SLCO2B1), transcript variant 1, mRNA [NM_007256] |
| A_23_P5903 | 0,030531606 | up | 2,5411322 | SLCO4A1 | Homo sapiens solute carrier organic anion transporter family, member 4A1 (SLCO4A1), mRNA [NM_016354] |
| A_33_P3381235 | 0,03067456 | down | -1,7796967 | SLCO4A1-AS1 | Homo sapiens SLCO4A1 antisense RNA 1 (SLCO4A1-AS1), long non-coding RNA [NR_024470] |
| A_23_P81484 | 0,001768669 | down | -2,3279634 | SLCO4C1 | Homo sapiens PRO2176 mRNA, complete cds. [AF119865] |
| A_23_P135669 | 0,006638752 | down | -2,314436 | SLCO5A1 | Homo sapiens solute carrier organic anion transporter family, member 5A1 (SLCO5A1), transcript variant 1, mRNA [NM_030958] |
| A_33_P3209522 | 0,009198317 | down | -1,5086755 | SLFN5 | Homo sapiens schlafen family member 5 (SLFN5), mRNA [NM_144975] |
| A_21_P0008099 | 3,32E-04 | down | -3,884536 | SLITRK5 | SLIT and NTRK-like family, member 5 [Source:HGNC Symbol;Acc:HGNC:20295] [ENST00000325089] |
| A_33_P3230189 | 4,91E-06 | down | -4,519842 | SLITRK6 | Homo sapiens SLIT and NTRK-like family, member 6 (SLITRK6), mRNA [NM_032229] |
| A_24_P146670 | 0,018891655 | up | 1,7328757 | SLK | Homo sapiens STE20-like kinase (SLK), mRNA [NM_014720] |
| A_33_P3243399 | 4,64E-04 | up | 1,8441696 | SLMAP | sarcolemma associated protein [Source:HGNC Symbol;Acc:HGNC:16643] [ENST00000467901] |
| A_33_P3243394 | 0,0323867 | up | 1,5183936 | SLMAP | sarcolemma associated protein [Source:HGNC Symbol;Acc:HGNC:16643] [ENST00000383718] |
| A_24_P288424 | 0,034604963 | up | 1,5011522 | SLMO2 | Homo sapiens slowmo homolog 2 (Drosophila) (SLMO2), transcript variant 1, mRNA [NM_016045] |
| A_33_P3287892 | 5,62E-04 | up | 2,7001781 | SLTM | SAFB-like, transcription modulator [Source:HGNC Symbol;Acc:HGNC:20709] [ENST00000480144] |
| A_24_P174824 | 0,010369504 | up | 1,5047495 | SLU7 | Homo sapiens SLU7 splicing factor homolog (S. cerevisiae) (SLU7), mRNA [NM_006425] |
| A_33_P3382281 | 0,017420491 | down | -1,9741542 | SLX4IP | SLX4 interacting protein [Source:HGNC Symbol;Acc:HGNC:16225] [ENST00000488816] |
| A_23_P48936 | 0,010684421 | up | 2,0509565 | SMAD3 | Homo sapiens SMAD family member 3 (SMAD3), transcript variant 1, mRNA [NM_005902] |
| A_23_P55518 | 0,03445947 | up | 1,5518922 | SMAD7 | Homo sapiens SMAD family member 7 (SMAD7), transcript variant 1, mRNA [NM_005904] |
| A_23_P406330 | 9,49E-04 | up | 2,0307677 | SMAP2 | Homo sapiens small ArfGAP2 (SMAP2), transcript variant 1, mRNA [NM_022733] |
| A_23_P252764 | 0,009426246 | down | -1,5202755 | SMARCA2 | Homo sapiens SWI/SNF related, matrix associated, actin dependent regulator of chromatin, subfamily a, member 2 (SMARCA2), transcript variant 2, mRNA [NM_139045] |
| A_21_P0014445 | 0,001637447 | down | -1,9633157 | SMARCA5-AS1 | Homo sapiens SMARCA5 antisense RNA 1 (SMARCA5-AS1), long non-coding RNA [NR_104027] |
| A_23_P66487 | 0,006530827 | up | 1,5353471 | SMARCD2 | Homo sapiens SWI/SNF related, matrix associated, actin dependent regulator of chromatin, subfamily d, member 2 (SMARCD2), mRNA [NM_001098426] |
| A_32_P126609 | 0,001883346 | up | 1,779294 | SMC5 | Homo sapiens structural maintenance of chromosomes 5 (SMC5), mRNA [NM_015110] |
| A_33_P3877739 | 0,01878333 | down | -2,0573351 | SMCR2 | ag52h12.x5 Gessler Wilms tumor Homo sapiens cDNA clone IMAGE:1126631 3', mRNA sequence [AI821758] |
| A_33_P3294679 | 0,002908388 | up | 2,527971 | SMCR5 | Homo sapiens Smith-Magenis syndrome chromosome region, candidate 5 (non-protein coding) (SMCR5), long non-coding RNA [NR_024007] |
| A_33_P3877728 | 0,013347329 | up | 1,6442406 | SMCR6 | Homo sapiens mRNA; cDNA DKFZp686J19223 (from clone DKFZp686J19223). [BX647090] |
| A_23_P300124 | 0,001975502 | down | -1,6241792 | SMCR8 | Homo sapiens Smith-Magenis syndrome chromosome region, candidate 8 (SMCR8), mRNA [NM_144775] |
| A_23_P218158 | 5,27E-04 | up | 1,56849 | SMEK1 | Homo sapiens SMEK homolog 1, suppressor of mek1 (Dictyostelium) (SMEK1), transcript variant 3, mRNA [NM_001284280] |

| | | | | | |
|---|---|---|---|---|---|
| A_33_P3391905 | 0,012787901 | up | 1,5408386 | SMEK3P | Homo sapiens cDNA FLJ16643 fis, clone TESTI4029528. [AK131475] |
| A_24_P244952 | 7,07E-06 | up | 2,121456 | SMG1 | Homo sapiens SMG1 phosphatidylinositol 3-kinase-related kinase (SMG1), mRNA [NM_015092] |
| A_33_P3326349 | 0,00239346 | down | -1,6047401 | SMG5 | Homo sapiens SMG5 nonsense mediated mRNA decay factor (SMG5), mRNA [NM_015327] |
| A_33_P3356031 | 0,016013835 | up | 1,6145606 | SMG7 | SMG7 nonsense mediated mRNA decay factor [Source:HGNC Symbol;Acc:HGNC:16792] [ENST00000367538] |
| A_21_P0000896 | 0,007021104 | up | 2,0751615 | SMG7-AS1 | Homo sapiens SMG7 antisense RNA 1 (SMG7-AS1), long non-coding RNA [NR_040063] |
| A_21_P0011718 | 0,021553854 | up | 1,6233528 | SMIM17 | Homo sapiens small integral membrane protein 17 (SMIM17), mRNA [NM_001193628] |
| A_21_P0000170 | 0,042239547 | up | 1,5834745 | SMIM18 | Homo sapiens small integral membrane protein 18 (SMIM18), mRNA [NM_001206847] |
| A_23_P151361 | 0,02445642 | down | -2,1034245 | SMIM2 | Homo sapiens small integral membrane protein 2 (SMIM2), mRNA [NM_024058] |
| A_19_P00810806 | 1,37E-04 | down | -2,4049666 | SMIM2-AS1 | Homo sapiens SMIM2 antisense RNA 1 (SMIM2-AS1), transcript variant 1, long non-coding RNA [NR_104064] |
| A_19_P00319971 | 0,019625295 | down | -1,5658339 | SMIM2-AS1 | Homo sapiens SMIM2 antisense RNA 1 (SMIM2-AS1), transcript variant 1, long non-coding RNA [NR_104064] |
| A_21_P0000125 | 0,004087763 | up | 1,7251751 | SMKR1 | Homo sapiens small lysine-rich protein 1 (SMKR1), mRNA [NM_001195243] |
| A_24_P463929 | 0,014470084 | up | 1,5228747 | SMLR1 | Homo sapiens small leucine-rich protein 1 (SMLR1), mRNA [NM_001195597] |
| A_19_P00319854 | 0,028193438 | down | -1,5991454 | SNAI3-AS1 | Homo sapiens SNAI3 antisense RNA 1 (SNAI3-AS1), transcript variant 2, long non-coding RNA [NR_024399] |
| A_33_P3338693 | 0,003532762 | down | -2,9789348 | SNAP25 | Homo sapiens synaptosomal-associated protein, 25kDa (SNAP25), transcript variant 1, mRNA [NM_003081] |
| A_33_P3316118 | 8,18E-04 | down | -2,6062465 | SNAP91 | Homo sapiens synaptosomal-associated protein, 91kDa (SNAP91), transcript variant 6, mRNA [NM_001256717] |
| A_33_P3335746 | 0,047957446 | up | 2,0194252 | SNAPC5 | Homo sapiens small nuclear RNA activating complex, polypeptide 5, 19kDa (SNAPC5), mRNA [NM_006049] |
| A_33_P3786807 | 0,003366749 | down | -3,0502307 | SNAR-E | UI-H-DF1-auh-i-18-0-UI.s1 NCI_CGAP_DF1 Homo sapiens cDNA clone UI-H-DF1-auh-i-18-0-UI 3', mRNA sequence [CA436475] |
| A_33_P3542886 | 0,026648767 | up | 1,6260659 | SNAR-G1 | AGENCOURT_10224340 NIH_MGC_141 Homo sapiens cDNA clone IMAGE:6565454 5', mRNA sequence [BU536871] |
| A_21_P0000512 | 0,002967795 | down | -2,19212 | SNAR-I | Homo sapiens small ILF3/NF90-associated RNA I (SNAR-I), small nuclear RNA [NR_024343] |
| A_23_P29939 | 0,012662336 | down | -1,7152753 | SNCA | Homo sapiens synuclein, alpha (non A4 component of amyloid precursor) (SNCA), transcript variant 4, mRNA [NM_007308] |
| A_24_P389285 | 0,03252091 | down | -1,8939469 | SND1-IT1 | Homo sapiens SND1 intronic transcript 1 (non-protein coding) (SND1-IT1), long non-coding RNA [NR_027330] |
| A_33_P3507562 | 0,048725072 | down | -1,9155712 | SNHG1 | AGENCOURT_6573317 NIH_MGC_124 Homo sapiens cDNA clone IMAGE:5732165 5', mRNA sequence [BM548627] |
| A_23_P74581 | 0,003234041 | up | 1,530148 | SNHG12 | Homo sapiens small nucleolar RNA host gene 12 (non-protein coding) (SNHG12), long non-coding RNA [NR_024127] |
| A_19_P00807665 | 0,021916734 | up | 1,6464124 | SNHG18 | Homo sapiens small nucleolar RNA host gene 18 (non-protein coding) (SNHG18), long non-coding RNA [NR_045196] |
| A_32_P8529 | 9,41E-04 | up | 2,0456448 | SNHG20 | Homo sapiens small nucleolar RNA host gene 20 (non-protein coding) (SNHG20), long non-coding RNA [NR_027058] |
| A_19_P00810899 | 9,17E-04 | up | 2,0027947 | SNHG20 | Homo sapiens small nucleolar RNA host gene 20 (non-protein coding) (SNHG20), long non-coding RNA [NR_027058] |
| A_19_P00320090 | 0,001525283 | up | 1,8496276 | SNHG20 | Homo sapiens small nucleolar RNA host gene 20 (non-protein coding) (SNHG20), long non-coding RNA [NR_027058] |
| A_19_P00316919 | 0,002560078 | up | 1,7003634 | SNHG20 | Homo sapiens small nucleolar RNA host gene 20 (non-protein coding) (SNHG20), long non-coding RNA [NR_027058] |
| A_21_P0014623 | 0,036497187 | up | 1,5703735 | SNHG21 | Homo sapiens small nucleolar RNA host gene 21 (non-protein coding) (SNHG21), transcript variant 1, long non-coding RNA [NR_110096] |
| A_33_P3280945 | 0,013466674 | up | 1,8244015 | SNHG3 | Homo sapiens small nucleolar RNA host gene 3 (non-protein coding) (SNHG3), transcript variant 1, long non-coding RNA [NR_036473] |
| A_24_P276531 | 0,001915744 | up | 1,6597381 | SNIP1 | Homo sapiens Smad nuclear interacting protein 1 (SNIP1), mRNA [NM_024700] |
| A_33_P3563369 | 0,010538275 | up | 1,744467 | SNORA10 | CM1-HT1146-090101-713-g10 HT1146 Homo sapiens cDNA, mRNA sequence [BG987537] |
| A_21_P0000309 | 0,010939649 | down | -1,7752984 | SNORA11 | Homo sapiens small nucleolar RNA, H/ACA box 11 (SNORA11), small nucleolar RNA [NR_002953] |
| A_21_P0000313 | 0,002539149 | down | -2,2453315 | SNORA15 | Homo sapiens small nucleolar RNA, H/ACA box 15 (SNORA15), small nucleolar RNA [NR_002957] |
| A_21_P0000494 | 0,044834394 | down | -2,3159316 | SNORA16B | Homo sapiens small nucleolar RNA, H/ACA box 16B (SNORA16B), small nucleolar RNA [NR_004389] |
| A_33_P3892710 | 0,024733124 | down | -1,7523577 | SNORA22 | DKFZp686E1139_r1 686 (synonym: hlcc3) Homo sapiens cDNA clone DKFZp686E1139 5', mRNA sequence [AL706006] |
| A_21_P0000321 | 0,020935867 | up | 1,5061066 | SNORA31 | Homo sapiens small nucleolar RNA, H/ACA box 31 (SNORA31), small nucleolar RNA [NR_002967] |
| A_33_P3650353 | 0,002011514 | up | 2,5030046 | SNORA33 | wm38b12.x1 NCI_CGAP_Ut4 Homo sapiens cDNA clone IMAGE:2438207 3' similar to gb:X53505 40S RIBOSOMAL PROTEIN S12 (HUMAN);, mRNA sequence [AI887274] |
| A_21_P0000322 | 0,048445106 | down | -1,6067711 | SNORA34 | Homo sapiens small nucleolar RNA, H/ACA box 34 (SNORA34), small nucleolar RNA [NR_002968] |
| A_21_P0000273 | 0,036137193 | up | 1,5086809 | SNORA41 | Homo sapiens small nucleolar RNA, H/ACA box 41 (SNORA41), small nucleolar RNA [NR_002590] |
| A_21_P0000331 | 0,012142163 | up | 1,5566672 | SNORA44 | Homo sapiens small nucleolar RNA, H/ACA box 44 (SNORA44), small nucleolar RNA [NR_002976] |
| A_21_P0000357 | 9,16E-04 | down | -1,7703443 | SNORA47 | Homo sapiens small nucleolar RNA, H/ACA box 47 (SNORA47), small nucleolar RNA [NR_003014] |
| A_21_P0000334 | 0,016384328 | down | -1,6220205 | SNORA49 | Homo sapiens small nucleolar RNA, H/ACA box 49 (SNORA49), small nucleolar RNA [NR_002979] |
| A_21_P0000269 | 0,016314311 | up | 1,5530452 | SNORA52 | Homo sapiens small nucleolar RNA, H/ACA box 52 (SNORA52), small nucleolar RNA [NR_002585] |
| A_33_P3462692 | 0,031632207 | up | 2,3233082 | SNORA58 | hz67c08.x1 NCI_CGAP_Lu24 Homo sapiens cDNA clone IMAGE:3213038 3', mRNA sequence [BE467637] |
| A_33_P3419458 | 0,001175921 | up | 1,7382413 | SNORA59B | Homo sapiens small nucleolar RNA, H/ACA box 59B (SNORA59B), small nucleolar RNA [NR_003022] |
| A_21_P0000342 | 0,039600268 | down | -1,7485112 | SNORA5B | Homo sapiens small nucleolar RNA, H/ACA box 5B (SNORA5B), small nucleolar RNA [NR_002990] |
| A_33_P3465703 | 4,80E-04 | down | -4,2223706 | SNORA60 | 601888042F1 NIH_MGC_17 Homo sapiens cDNA clone IMAGE:4121819 5', mRNA sequence [BF304636] |
| A_21_P0000340 | 0,005858892 | down | -2,1300526 | SNORA60 | Homo sapiens small nucleolar RNA, H/ACA box 60 (SNORA60), small nucleolar RNA [NR_002986] |
| A_21_P0000341 | 0,006828111 | up | 1,7214358 | SNORA61 | Homo sapiens small nucleolar RNA, H/ACA box 61 (SNORA61), small nucleolar RNA [NR_002987] |
| A_21_P0000270 | 0,008712771 | down | -1,7762682 | SNORA63 | Homo sapiens small nucleolar RNA, H/ACA box 63 (SNORA63), small nucleolar RNA [NR_002586] |
| A_21_P0000236 | 0,006464346 | up | 1,6503971 | SNORA64 | Homo sapiens small nucleolar RNA, H/ACA box 64 (SNORA64), small nucleolar RNA [NR_002326] |
| A_21_P0000251 | 0,015205678 | up | 1,5664947 | SNORA65 | Homo sapiens small nucleolar RNA, H/ACA box 65 (SNORA65), small nucleolar RNA [NR_002449] |
| A_21_P0000295 | 0,006032473 | up | 1,5848244 | SNORA67 | Homo sapiens small nucleolar RNA, H/ACA box 67 (SNORA67), small nucleolar RNA [NR_002912] |
| A_21_P0000212 | 5,82E-04 | up | 1,8266126 | SNORA68 | Homo sapiens small nucleolar RNA, H/ACA box 68 (SNORA68), small nucleolar RNA [NR_000012] |
| A_21_P0000475 | 0,043606266 | up | 1,5940142 | SNORA70B | Homo sapiens small nucleolar RNA, H/ACA box 70B (SNORA70B), small nucleolar RNA [NR_003707] |
| A_21_P0000593 | 0,015089021 | up | 1,6602123 | SNORA70D | Homo sapiens small nucleolar RNA, H/ACA box 70D (SNORA70D), small nucleolar RNA [NR_033337] |
| A_21_P0000591 | 0,011378228 | up | 1,5358397 | SNORA70F | Homo sapiens small nucleolar RNA, H/ACA box 70F (SNORA70F), small nucleolar RNA [NR_033309] |
| A_21_P0000592 | 0,039196156 | up | 1,5337374 | SNORA70G | Homo sapiens small nucleolar RNA, H/ACA box 70G (SNORA70G), small nucleolar RNA [NR_033335] |
| A_33_P3495962 | 0,009419474 | down | -2,0101612 | SNORA71A | AGENCOURT_6611325 NIH_MGC_106 Homo sapiens cDNA clone IMAGE:5485440 5', mRNA sequence [BM918074] |
| A_21_P0000327 | 0,003137789 | down | -2,5250878 | SNORA71E | Homo sapiens small nucleolar RNA, H/ACA box 71E (SNORA71E), small nucleolar RNA [NR_002972] |

| | | | | | |
|---|---|---|---|---|---|
| A_33_P3327519 | 0,013498295 | down | -1,8012974 | SNORA74A | Homo sapiens small nucleolar RNA, H/ACA box 74A (SNORA74A), small nucleolar RNA [NR_002915] |
| A_33_P3876591 | 0,04042346 | down | -1,8173707 | SNORA76C | AGENCOURT_13972169 NIH_MGC_173 Homo sapiens cDNA 5', mRNA sequence [CD050382] |
| A_33_P3579391 | 0,024037624 | up | 1,7647849 | SNORA77 | DB361496 MAMMA1 Homo sapiens cDNA clone MAMMA1001810 3', mRNA sequence [DB361496] |
| A_21_P0000308 | 0,005737074 | down | -1,6070489 | SNORA9 | Homo sapiens small nucleolar RNA, H/ACA box 9 (SNORA9), small nucleolar RNA [NR_002952] |
| A_21_P0000463 | 0,001129832 | down | -2,49153 | SNORD105B | Homo sapiens small nucleolar RNA, C/D box 105B (SNORD105B), small nucleolar RNA [NR_003688] |
| A_21_P0000366 | 0,012617496 | up | 1,5168216 | SNORD11 | Homo sapiens small nucleolar RNA, C/D box 11 (SNORD11), small nucleolar RNA [NR_003031] |
| A_21_P0000415 | 0,009631189 | up | 1,5934927 | SNORD114-18 | Homo sapiens small nucleolar RNA, C/D box 114-18 (SNORD114-18), small nucleolar RNA [NR_003211] |
| A_21_P0000456 | 0,001632496 | up | 1,8205398 | SNORD115-48 | Homo sapiens small nucleolar RNA, C/D box 115-48 (SNORD115-48), small nucleolar RNA [NR_003362] |
| A_21_P0000447 | 0,003665484 | down | -3,2352173 | SNORD116-11 | Homo sapiens small nucleolar RNA, C/D box 116-11 (SNORD116-11), small nucleolar RNA [NR_003326] |
| A_21_P0000448 | 0,018371584 | down | -2,4427204 | SNORD116-12 | Homo sapiens small nucleolar RNA, C/D box 116-12 (SNORD116-12), small nucleolar RNA [NR_003327] |
| A_21_P0000445 | 0,043550096 | down | -1,7331018 | SNORD116-6 | Homo sapiens small nucleolar RNA, C/D box 116-6 (SNORD116-6), small nucleolar RNA [NR_003321] |
| A_21_P0000398 | 0,007444821 | down | -1,9945382 | SNORD117 | Homo sapiens small nucleolar RNA, C/D box 117 (SNORD117), small nucleolar RNA [NR_003140] |
| A_21_P0000459 | 0,014022734 | up | 1,6801827 | SNORD119 | Homo sapiens small nucleolar RNA, C/D box 119 (SNORD119), small nucleolar RNA [NR_003684] |
| A_21_P0000460 | 0,003709252 | down | -1,9065627 | SNORD121A | Homo sapiens small nucleolar RNA, C/D box 121A (SNORD121A), small nucleolar RNA [NR_003685] |
| A_33_P3578325 | 0,009560458 | down | -1,9017425 | SNORD15A | Homo sapiens small nucleolar RNA, C/D box 15A (SNORD15A), small nucleolar RNA [NR_000005] |
| A_21_P0000247 | 0,03509152 | up | 1,5110737 | SNORD18C | Homo sapiens small nucleolar RNA, C/D box 18C (SNORD18C), small nucleolar RNA [NR_002443] |
| A_21_P0000496 | 0,02720046 | up | 1,6064279 | SNORD1A | Homo sapiens small nucleolar RNA, C/D box 1A (SNORD1A), small nucleolar RNA [NR_004395] |
| A_21_P0000208 | 0,017108412 | up | 1,5037471 | SNORD22 | Homo sapiens small nucleolar RNA, C/D box 22 (SNORD22), small nucleolar RNA [NR_000008] |
| A_21_P0000217 | 8,66E-04 | up | 2,4410224 | SNORD36B | Homo sapiens small nucleolar RNA, C/D box 36B (SNORD36B), small nucleolar RNA [NR_000017] |
| A_21_P0000216 | 0,001096856 | up | 1,8858453 | SNORD36C | Homo sapiens small nucleolar RNA, C/D box 36C (SNORD36C), small nucleolar RNA [NR_000016] |
| A_21_P0000277 | 0,042853054 | down | -1,6330094 | SNORD37 | Homo sapiens small nucleolar RNA, C/D box 37 (SNORD37), small nucleolar RNA [NR_002602] |
| A_21_P0000213 | 0,013207308 | up | 1,6422514 | SNORD42B | Homo sapiens small nucleolar RNA, C/D box 42B (SNORD42B), small nucleolar RNA [NR_000013] |
| A_21_P0000287 | 0,00260944 | up | 1,5744709 | SNORD47 | Homo sapiens small nucleolar RNA, C/D box 47 (SNORD47), small nucleolar RNA [NR_002746] |
| A_21_P0000210 | 0,015711462 | up | 1,6161708 | SNORD4A | Homo sapiens small nucleolar RNA, C/D box 4A (SNORD4A), small nucleolar RNA [NR_000010] |
| A_21_P0000282 | 0,034710076 | up | 1,5615475 | SNORD56 | Homo sapiens small nucleolar RNA, C/D box 56 (SNORD56), small nucleolar RNA [NR_002739] |
| A_21_P0000297 | 2,87E-05 | down | -3,424353 | SNORD62A | Homo sapiens small nucleolar RNA, C/D box 62A (SNORD62A), small nucleolar RNA [NR_002914] |
| A_21_P0000378 | 0,012299744 | down | -1,5496659 | SNORD66 | Homo sapiens small nucleolar RNA, C/D box 66 (SNORD66), small nucleolar RNA [NR_003055] |
| A_21_P0000371 | 0,01607081 | up | 1,7449336 | SNORD7 | Homo sapiens small nucleolar RNA, C/D box 7 (SNORD7), small nucleolar RNA [NR_003037] |
| A_33_P3843358 | 0,012032601 | up | 1,7385544 | SNORD8 | ax47d11.x1 Proliferating Human Erythroid Cells (LCB:ax library) Homo sapiens cDNA clone ax47d11 random, mRNA sequence [BG944179] |
| A_21_P0000384 | 0,016472233 | down | -2,0231233 | SNORD85 | Homo sapiens small nucleolar RNA, C/D box 85 (SNORD85), small nucleolar RNA [NR_003066] |
| A_21_P0000500 | 0,012820842 | up | 1,5940045 | SNORD86 | Homo sapiens small nucleolar RNA, C/D box 86 (SNORD86), small nucleolar RNA [NR_004399] |
| A_21_P0000501 | 0,015770653 | down | -1,5651679 | SNORD97 | Homo sapiens small nucleolar RNA, C/D box 97 (SNORD97), small nucleolar RNA [NR_004403] |
| A_21_P0000392 | 0,020515464 | down | -1,502568 | SNORD98 | Homo sapiens small nucleolar RNA, C/D box 98 (SNORD98), small nucleolar RNA [NR_003076] |
| A_21_P0000393 | 0,03309451 | up | 1,5095688 | SNORD99 | Homo sapiens small nucleolar RNA, C/D box 99 (SNORD99), small nucleolar RNA [NR_003077] |
| A_23_P211985 | 6,86E-04 | up | 1,9152808 | SNRK | Homo sapiens SNF related kinase (SNRK), transcript variant 1, mRNA [NM_017719] |
| A_23_P159101 | 0,001891439 | up | 1,5801884 | SNRNP35 | Homo sapiens small nuclear ribonucleoprotein 35kDa (U11/U12) (SNRNP35), transcript variant 2, mRNA [NM_022717] |
| A_33_P3384900 | 2,50E-05 | down | -2,457822 | SNRPD2P2 | Homo sapiens small nuclear ribonucleoprotein D2 pseudogene 2 (SNRPD2P2), non-coding RNA [NR_033826] |
| A_33_P3314511 | 0,006940991 | down | -2,4550972 | SNRPD3 | small nuclear ribonucleoprotein D3 polypeptide 18kDa [Source:HGNC Symbol;Acc:HGNC:11160] [ENST00000402849] |
| A_24_P27412 | 0,03834043 | up | 1,5068727 | SNUPN | Homo sapiens snurportin 1 (SNUPN), transcript variant 1, mRNA [NM_005701] |
| A_33_P3268716 | 0,009444252 | up | 1,5913441 | SNX1 | Homo sapiens sorting nexin 1 (SNX1), transcript variant 1, mRNA [NM_003099] |
| A_33_P3289536 | 0,017850125 | up | 1,917376 | SNX10 | Homo sapiens sorting nexin 10 (SNX10), transcript variant 1, mRNA [NM_001199835] |
| A_33_P3250128 | 1,45E-04 | down | -2,1514487 | SNX18 | Homo sapiens sorting nexin 18 (SNX18), transcript variant 2, mRNA [NM_052870] |
| A_24_P117177 | 0,01659635 | down | -1,6674821 | SNX21 | Homo sapiens sorting nexin family member 21 (SNX21), transcript variant 2, mRNA [NM_152897] |
| A_23_P154585 | 0,00464523 | down | -1,5175135 | SNX21 | Homo sapiens sorting nexin family member 21 (SNX21), transcript variant 4, mRNA [NM_001042633] |
| A_33_P3212172 | 0,020338247 | up | 2,3909264 | SNX22 | Homo sapiens sorting nexin 22 (SNX22), transcript variant 1, mRNA [NM_024798] |
| A_32_P107002 | 5,98E-05 | up | 2,1039908 | SNX29 | Homo sapiens sorting nexin 29 (SNX29), mRNA [NM_032167] |
| A_33_P3292679 | 0,002123101 | up | 1,5200515 | SNX6 | Homo sapiens sorting nexin 6 (SNX6), transcript variant 1, mRNA [NM_021249] |
| A_19_P00807670 | 0,0154589 | down | -1,6005719 | SOCS2-AS1 | Homo sapiens SOCS2 antisense RNA 1 (SOCS2-AS1), long non-coding RNA [NR_038263] |
| A_19_P00802794 | 0,025202228 | down | -1,9329783 | SOCS2-AS1 | Homo sapiens SOCS2 antisense RNA 1 (SOCS2-AS1), long non-coding RNA [NR_038263] |
| A_23_P207058 | 0,009831551 | up | 1,8462876 | SOCS3 | Homo sapiens suppressor of cytokine signaling 3 (SOCS3), mRNA [NM_003955] |
| A_24_P367211 | 1,72E-04 | down | -1,9877064 | SOCS7 | Homo sapiens suppressor of cytokine signaling 7 (SOCS7), mRNA [NM_014598] |
| A_24_P307964 | 0,017352385 | up | 2,838105 | SOHLH1 | Homo sapiens spermatogenesis and oogenesis specific basic helix-loop-helix 1 (SOHLH1), transcript variant 2, mRNA [NM_001012415] |
| A_24_P67142 | 0,037885543 | down | -1,5337447 | SON | Homo sapiens SON DNA binding protein (SON), transcript variant b, mRNA [NM_032195] |
| A_23_P402733 | 0,001443486 | up | 1,5882757 | SON | Homo sapiens SON DNA binding protein (SON), transcript variant b, mRNA [NM_032195] |
| A_33_P3313314 | 0,00406494 | down | -2,8300092 | SORBS1 | Homo sapiens sorbin and SH3 domain containing 1 (SORBS1), transcript variant 3, mRNA [NM_001034954] |
| A_33_P3409625 | 0,005571186 | down | -1,5338488 | SORBS3 | Homo sapiens sorbin and SH3 domain containing 3 (SORBS3), transcript variant 1, mRNA [NM_005775] |
| A_33_P3420889 | 0,026719104 | down | -1,7619413 | SORCS3 | Homo sapiens sortilin-related VPS10 domain containing receptor 3 (SORCS3), mRNA [NM_014978] |
| A_33_P3407549 | 0,013960474 | down | -2,0702763 | SOWAHA | Homo sapiens sosondowah ankyrin repeat domain family member A (SOWAHA), mRNA [NM_175873] |
| A_23_P304110 | 0,015957905 | up | 2,4265115 | SOWAHA | Homo sapiens sosondowah ankyrin repeat domain family member A (SOWAHA), mRNA [NM_175873] |
| A_23_P143694 | 0,010440017 | down | -2,090848 | SOX10 | Homo sapiens SRY (sex determining region Y)-box 10 (SOX10), mRNA [NM_006941] |
| A_33_P3325110 | 0,001252482 | down | -2,411466 | SOX13 | Homo sapiens SRY (sex determining region Y)-box 13 (SOX13), mRNA [NM_005686] |

| | | | | | |
|---|---|---|---|---|---|
| A_23_P80662 | 0,001957604 | up | 3,1804535 | SOX14 | Homo sapiens SRY (sex determining region Y)-box 14 (SOX14), mRNA [NM_004189] |
| A_23_P85218 | 0,00244925 | down | -8,071605 | SOX3 | Homo sapiens SRY (sex determining region Y)-box 3 (SOX3), mRNA [NM_005634] |
| A_33_P3233916 | 0,008451398 | down | -2,183581 | SOX6 | Homo sapiens SRY (sex determining region Y)-box 6 (SOX6), transcript variant 1, mRNA [NM_017508] |
| A_33_P3228325 | 0,009539026 | up | 1,5451137 | SP100 | Homo sapiens SP100 nuclear antigen (SP100), transcript variant 1, mRNA [NM_001080391] |
| A_23_P381449 | 0,002673548 | up | 1,7262025 | SP2 | Homo sapiens Sp2 transcription factor (SP2), mRNA [NM_003110] |
| A_33_P3352664 | 0,001032771 | down | -2,2815936 | SP7 | Homo sapiens Sp7 transcription factor (SP7), transcript variant 3, mRNA [NM_001300837] |
| A_33_P3250148 | 0,039825235 | down | -1,5433004 | SP8 | Homo sapiens Sp8 transcription factor (SP8), transcript variant 2, mRNA [NM_198956] |
| A_23_P30649 | 0,002535965 | down | -2,330642 | SPACA1 | Homo sapiens sperm acrosome associated 1 (SPACA1), mRNA [NM_030960] |
| A_21_P0000880 | 0,022133902 | down | -2,1848845 | SPAG5-AS1 | Homo sapiens SPAG5 antisense RNA 1 (SPAG5-AS1), long non-coding RNA [NR_040012] |
| A_24_P365025 | 0,002424178 | up | 1,713438 | SPAG9 | Homo sapiens sperm associated antigen 9 (SPAG9), transcript variant 3, mRNA [NM_003971] |
| A_33_P3377609 | 0,033370137 | down | -1,9062088 | SPAM1 | Homo sapiens sperm adhesion molecule 1 (PH-20 hyaluronidase, zona pellucida binding) (SPAM1), transcript variant 3, mRNA [NM_001174044] |
| A_24_P305784 | 0,014370482 | up | 2,531941 | SPANXB1 | Homo sapiens SPANX family, member B1 (SPANXB1), mRNA [NM_032461] |
| A_33_P3276653 | 0,003191604 | up | 1,7886204 | SPANXN1 | Homo sapiens SPANX family, member N1 (SPANXN1), mRNA [NM_001009614] |
| A_24_P91830 | 0,04599167 | up | 1,9761709 | SPANXN3 | Homo sapiens SPANX family, member N3 (SPANXN3), mRNA [NM_001009609] |
| A_23_P103897 | 0,01474256 | down | -1,6414444 | SPATA1 | spermatogenesis associated 1 [Source:HGNC Symbol;Acc:HGNC:14682] [ENST00000431031] |
| A_33_P3298111 | 0,03585235 | up | 2,3773272 | SPATA13 | spermatogenesis associated 13 [Source:HGNC Symbol;Acc:HGNC:23222] [ENST00000466831] |
| A_23_P210554 | 0,001347662 | up | 1,7344328 | SPATA2 | Homo sapiens spermatogenesis associated 2 (SPATA2), transcript variant 1, mRNA [NM_006038] |
| A_33_P3216438 | 0,017247561 | down | -2,4187534 | SPATA21 | Homo sapiens spermatogenesis associated 21 (SPATA21), mRNA [NM_198546] |
| A_21_P0000078 | 0,03500633 | up | 2,075116 | SPATA22 | Homo sapiens spermatogenesis associated 22 (SPATA22), transcript variant 1, mRNA [NM_001170699] |
| A_21_P0002142 | 0,002831261 | down | -2,7909002 | SPATA3-AS1 | SPATA3 antisense RNA 1 (head to head) [Source:HGNC Symbol;Acc:HGNC:28013] [ENST00000426904] |
| A_21_P0013668 | 0,011819395 | down | -1,6053566 | SPATA31C2 | Homo sapiens SPATA31 subfamily C, member 2 (SPATA31C2), mRNA [NM_001166137] |
| A_32_P70056 | 0,017306114 | up | 1,6965684 | SPATA42 | Homo sapiens spermatogenesis associated 42 (non-protein coding) (SPATA42), transcript variant 2, long non-coding RNA [NR_049777] |
| A_33_P3321919 | 8,86E-05 | up | 2,7889643 | SPATA45 | Homo sapiens spermatogenesis associated 45 (SPATA45), mRNA [NM_001024601] |
| A_33_P3268224 | 0,035701726 | up | 2,0021043 | SPATA5 | Homo sapiens spermatogenesis associated 5 (SPATA5), mRNA [NM_145207] |
| A_24_P922252 | 0,007018658 | up | 2,316299 | SPATA5L1 | Homo sapiens cDNA FLJ12286 fis, clone MAMMA1001768, weakly similar to CELL DIVISION CYCLE PROTEIN 48 HOMOLOG MJ1156. [AK022348] |
| A_33_P3254073 | 0,042611912 | down | -1,9304503 | SPATA6L | spermatogenesis associated 6-like [Source:HGNC Symbol;Acc:HGNC:25472] [ENST00000471669] |
| A_32_P102935 | 0,037747014 | up | 1,6198094 | SPDYA | Homo sapiens speedy/RINGO cell cycle regulator family member A (SPDYA), transcript variant 2, mRNA [NM_001008779] |
| A_33_P3322945 | 0,010294055 | down | -1,5167365 | SPDYC | Homo sapiens speedy/RINGO cell cycle regulator family member C (SPDYC), mRNA [NM_001008778] |
| A_23_P425967 | 0,01187239 | down | -2,5839787 | SPDYE1 | Homo sapiens speedy/RINGO cell cycle regulator family member E1 (SPDYE1), mRNA [NM_175064] |
| A_24_P255986 | 0,009197988 | down | -2,3075967 | SPDYE4 | Homo sapiens speedy/RINGO cell cycle regulator family member E4 (SPDYE4), mRNA [NM_001128076] |
| A_32_P202182 | 0,03537796 | up | 1,782816 | SPDYE8P | Homo sapiens speedy/RINGO cell cycle regulator family member E8, pseudogene (SPDYE8P), non-coding RNA [NR_003664] |
| A_23_P12053 | 1,11E-05 | up | 2,0489528 | SPEN | Homo sapiens spen family transcriptional repressor (SPEN), mRNA [NM_015001] |
| A_19_P00806499 | 1,19E-05 | down | -4,1503005 | SPG20-AS1 | Homo sapiens SPG20 antisense RNA 1 (SPG20-AS1), transcript variant 1, long non-coding RNA [NR_045180] |
| A_21_P0008998 | 0,04221182 | up | 1,5715525 | SPG7 | spastic paraplegia 7 (pure and complicated autosomal recessive) [Source:HGNC Symbol;Acc:HGNC:11237] [ENST00000568509] |
| A_21_P0009117 | 0,008526453 | up | 1,7833046 | SPG7 | spastic paraplegia 7 (pure and complicated autosomal recessive) [Source:HGNC Symbol;Acc:HGNC:11237] [ENST00000568509] |
| A_24_P5305 | 0,019773852 | up | 1,5983998 | SPG7 | Homo sapiens spastic paraplegia 7 (pure and complicated autosomal recessive) (SPG7), transcript variant 1, mRNA [NM_003119] |
| A_23_P433990 | 0,026564928 | up | 1,6278061 | SPG7 | Homo sapiens spastic paraplegia 7 (pure and complicated autosomal recessive) (SPG7), transcript variant 1, mRNA [NM_003119] |
| A_33_P3489646 | 0,03594257 | down | -1,6157703 | SPI1 | Homo sapiens Spi-1 proto-oncogene (SPI1), transcript variant 1, mRNA [NM_001080547] |
| A_33_P3388006 | 0,030238435 | up | 1,5969714 | SPICE1 | Homo sapiens spindle and centriole associated protein 1 (SPICE1), mRNA [NM_144718] |
| A_21_P0014029 | 0,026568804 | down | -1,6992115 | SPIDR | scaffolding protein involved in DNA repair [Source:HGNC Symbol;Acc:HGNC:28971] [ENST00000521798] |
| A_24_P143686 | 0,001476954 | up | 1,5441651 | SPIDR | Homo sapiens scaffolding protein involved in DNA repair (SPIDR), transcript variant 1, mRNA [NM_001080394] |
| A_24_P494454 | 0,011750751 | up | 1,6786367 | SPIN3 | Homo sapiens spindlin family, member 3 (SPIN3), transcript variant 1, mRNA [NM_001010862] |
| A_33_P3381836 | 0,006196491 | down | -2,4531546 | SPINK14 | Homo sapiens serine peptidase inhibitor, Kazal type 14 (putative) (SPINK14), mRNA [NM_001001325] |
| A_23_P155688 | 0,012787063 | up | 1,5551062 | SPINK2 | Homo sapiens serine peptidase inhibitor, Kazal type 2 (acrosin-trypsin inhibitor) (SPINK2), transcript variant 2, mRNA [NM_021114] |
| A_33_P3424302 | 0,045472402 | up | 1,5086557 | SPINT4 | Homo sapiens serine peptidase inhibitor, Kunitz type 4 (SPINT4), mRNA [NM_178455] |
| A_33_P3300172 | 1,13E-04 | down | -2,9129062 | SPON2 | Homo sapiens cDNA FLJ42615 fis, clone BRACE3014231. [AK124606] |
| A_24_P107859 | 0,010338904 | up | 1,7688456 | SPRED1 | Homo sapiens sprouty-related, EVH1 domain containing 1 (SPRED1), mRNA [NM_152594] |
| A_32_P225854 | 0,001193612 | up | 1,5619863 | SPRED2 | Homo sapiens sprouty-related, EVH1 domain containing 2 (SPRED2), transcript variant 1, mRNA [NM_181784] |
| A_33_P3401981 | 0,001867043 | up | 2,9933386 | SPRED3 | Homo sapiens sprouty-related, EVH1 domain containing 3 (SPRED3), transcript variant 1, mRNA [NM_001042522] |
| A_24_P576506 | 0,004625077 | down | -2,2741063 | SPRN | Homo sapiens shadow of prion protein homolog (zebrafish) (SPRN), mRNA [NM_001012508] |
| A_23_P144476 | 0,010696892 | up | 1,925552 | SPRY1 | Homo sapiens sprouty homolog 1, antagonist of FGF signaling (Drosophila) (SPRY1), transcript variant 2, mRNA [NM_199327] |
| A_24_P204819 | 0,025078906 | down | -1,796512 | SPRY3 | Homo sapiens sprouty homolog 3 (Drosophila) (SPRY3), mRNA [NM_005840] |
| A_33_P3338423 | 0,026434213 | down | -1,721988 | SPRY4 | Homo sapiens sprouty homolog 4 (Drosophila) (SPRY4), transcript variant 3, mRNA [NM_001293289] |
| A_23_P368718 | 0,014416411 | up | 1,5694852 | SPRYD7 | SPRY domain containing 7 [Source:HGNC Symbol;Acc:HGNC:14297] [ENST00000613924] |
| A_24_P96961 | 0,01789821 | up | 1,5177591 | SPSB1 | Homo sapiens splA/ryanodine receptor domain and SOCS box containing 1 (SPSB1), mRNA [NM_025106] |
| A_23_P258381 | 0,01269423 | down | -2,1155863 | SPSB4 | splA/ryanodine receptor domain and SOCS box containing 4 [Source:HGNC Symbol;Acc:HGNC:30630] [ENST00000508126] |
| A_23_P20832 | 0,014934784 | up | 1,6108845 | SPTAN1 | Homo sapiens spectrin, alpha, non-erythrocytic 1 (SPTAN1), transcript variant 2, mRNA [NM_003127] |
| A_23_P65506 | 0,044793073 | down | -1,6646454 | SPTB | Homo sapiens spectrin, beta, erythrocytic (SPTB), transcript variant 2, mRNA [NM_000347] |
| A_24_P293114 | 0,020756127 | down | -1,735082 | SPTBN4 | Homo sapiens spectrin, beta, non-erythrocytic 4 (SPTBN4), transcript variant sigma1, mRNA [NM_020971] |
| A_24_P150486 | 4,15E-04 | up | 1,756872 | SPTLC2 | Homo sapiens serine palmitoyltransferase, long chain base subunit 2 (SPTLC2), mRNA [NM_004863] |
| A_33_P3305617 | 0,001885898 | down | -2,1076288 | SPTSSB | serine palmitoyltransferase, small subunit B [Source:HGNC Symbol;Acc:HGNC:24045] [ENST00000617024] |

| | | | | | |
|---|---|---|---|---|---|
| A_23_P308603 | 4,99E-04 | up | 2,3055863 | SRC | Homo sapiens SRC proto-oncogene, non-receptor tyrosine kinase (SRC), transcript variant 1, mRNA [NM_005417] |
| A_24_P944991 | 0,012150929 | down | -1,52653 | SREK1IP1 | Homo sapiens SREK1-interacting protein 1 (SREK1IP1), mRNA [NM_173829] |
| A_24_P337657 | 0,017482813 | up | 2,1334105 | SRF | Homo sapiens serum response factor (c-fos serum response element-binding transcription factor) (SRF), transcript variant 1, mRNA [NM_003131] |
| A_33_P3213772 | 0,016628768 | up | 1,553939 | SRGAP2 | Homo sapiens SLIT-ROBO Rho GTPase activating protein 2 (SRGAP2), transcript variant 4, mRNA [NM_001300952] |
| A_33_P3368540 | 4,37E-05 | down | -2,333685 | SRGAP3 | SLIT-ROBO Rho GTPase activating protein 3 [Source:HGNC Symbol;Acc:HGNC:19744] [ENST00000490889] |
| A_23_P358410 | 0,005668055 | up | 2,4407232 | SRGAP3 | Homo sapiens SLIT-ROBO Rho GTPase activating protein 3 (SRGAP3), transcript variant 1, mRNA [NM_014850] |
| A_33_P3236798 | 0,037908956 | down | -2,0868375 | SRMS | Homo sapiens src-related kinase lacking C-terminal regulatory tyrosine and N-terminal myristylation sites (SRMS), mRNA [NM_080823] |
| A_21_P0000895 | 0,020874841 | up | 1,5700223 | SRP14-AS1 | Homo sapiens SRP14 antisense RNA1 (head to head) (SRP14-AS1), transcript variant 4, long non-coding RNA [NR_040062] |
| A_23_P162945 | 0,019539094 | up | 1,5316727 | SRP54 | Homo sapiens signal recognition particle 54kDa (SRP54), transcript variant 1, mRNA [NM_003136] |
| A_24_P11131 | 7,43E-05 | up | 1,8887756 | SRRM1 | Homo sapiens serine/arginine repetitive matrix 1 (SRRM1), transcript variant 2, mRNA [NM_005839] |
| A_33_P3324755 | 2,47E-04 | down | -2,760445 | SRRM2-AS1 | Homo sapiens SRRM2 antisense RNA 1 (SRRM2-AS1), transcript variant 1, long non-coding RNA [NR_027274] |
| A_23_P42768 | 0,013918295 | down | -1,765268 | SRRM3 | Homo sapiens serine/arginine repetitive matrix 3 (SRRM3), transcript variant 2, mRNA [NM_001110199] |
| A_23_P331700 | 0,008796485 | up | 2,2135015 | SRRM3 | Homo sapiens serine/arginine repetitive matrix 3 (SRRM3), transcript variant 1, mRNA [NM_001291831] |
| A_23_P111481 | 0,001834746 | up | 1,6834066 | SRRT | Homo sapiens serrate, RNA effector molecule (SRRT), transcript variant 1, mRNA [NM_015908] |
| A_23_P111487 | 0,003303443 | up | 1,6821806 | SRRT | Homo sapiens serrate, RNA effector molecule (SRRT), transcript variant 4, mRNA [NM_001128853] |
| A_23_P352291 | 0,017436704 | up | 1,5974357 | SRSF10 | Homo sapiens serine/arginine-rich splicing factor 10 (SRSF10), transcript variant 2, mRNA [NM_054016] |
| A_23_P77776 | 9,89E-05 | up | 1,989498 | SRSF2 | Homo sapiens serine/arginine-rich splicing factor 2 (SRSF2), transcript variant 1, mRNA [NM_003016] |
| A_23_P423926 | 0,001521755 | up | 1,9072506 | SS18L1 | Homo sapiens synovial sarcoma translocation gene on chromosome 18-like 1 (SS18L1), transcript variant 1, mRNA [NM_198935] |
| A_33_P3393931 | 0,02454142 | down | -2,4707692 | SSBP3-AS1 | Homo sapiens SSBP3 antisense RNA 1 (SSBP3-AS1), long non-coding RNA [NR_103541] |
| A_21_P0000127 | 0,027079117 | up | 2,2916749 | SSC5D | Homo sapiens scavenger receptor cysteine rich family, 5 domains (SSC5D), transcript variant 2, mRNA [NM_001195267] |
| A_23_P102060 | 0,041046876 | up | 1,5175989 | SSFA2 | Homo sapiens sperm specific antigen 2 (SSFA2), transcript variant 2, mRNA [NM_006751] |
| A_24_P332647 | 0,009371382 | up | 1,7682351 | SSH1 | Homo sapiens slingshot protein phosphatase 1 (SSH1), transcript variant 3, mRNA [NM_001161331] |
| A_33_P3367917 | 0,004884202 | up | 1,7391522 | SSH2 | Homo sapiens slingshot protein phosphatase 2 (SSH2), transcript variant 3, mRNA [NM_001282130] |
| A_33_P3363168 | 0,005870757 | up | 1,5802524 | SSH2 | Homo sapiens slingshot protein phosphatase 2 (SSH2), transcript variant 1, mRNA [NM_001282129] |
| A_33_P3277183 | 0,035661407 | up | 2,0261858 | SSPO | SCO-spondin [Source:HGNC Symbol;Acc:HGNC:21998] [ENST00000475488] |
| A_33_P3352467 | 0,009910471 | up | 2,726636 | SSTR2 | Homo sapiens somatostatin receptor 2 (SSTR2), mRNA [NM_001050] |
| A_23_P68910 | 0,04933239 | down | -1,5359924 | SSTR3 | Homo sapiens somatostatin receptor 3 (SSTR3), transcript variant 1, mRNA [NM_001051] |
| A_33_P3414267 | 1,78E-04 | down | -3,9464931 | SSTR4 | Homo sapiens somatostatin receptor 4 (SSTR4), mRNA [NM_001052] |
| A_23_P77545 | 0,018193105 | down | -1,9678162 | SSTR5 | Homo sapiens somatostatin receptor 5 (SSTR5), transcript variant 1, mRNA [NM_001053] |
| A_24_P256243 | 0,00131506 | down | -1,9598496 | SSX2 | Homo sapiens synovial sarcoma, X breakpoint 2 (SSX2), transcript variant 2, mRNA [NM_175698] |
| A_23_P252023 | 4,87E-04 | up | 3,698383 | SSX2 | Homo sapiens synovial sarcoma, X breakpoint 2 (SSX2), transcript variant 2, mRNA [NM_175698] |
| A_23_P31858 | 0,013593148 | down | -2,1409123 | ST18 | Homo sapiens suppression of tumorigenicity 18, zinc finger (ST18), mRNA [NM_014682] |
| A_24_P268123 | 0,009517816 | up | 1,6738088 | ST3GAL3 | Homo sapiens ST3 beta-galactoside alpha-2,3-sialyltransferase 3 (ST3GAL3), transcript variant 1, mRNA [NM_174963] |
| A_33_P3405966 | 1,20E-04 | up | 2,5471005 | ST3GAL3 | Homo sapiens ST3 beta-galactoside alpha-2,3-sialyltransferase 3 (ST3GAL3), transcript variant 1, mRNA [NM_174963] |
| A_23_P250800 | 0,037591886 | up | 1,5630652 | ST3GAL6 | Homo sapiens ST3 beta-galactoside alpha-2,3-sialyltransferase 6 (ST3GAL6), transcript variant 1, mRNA [NM_006100] |
| A_24_P388528 | 0,001458094 | up | 1,7945207 | ST6GAL1 | Homo sapiens ST6 beta-galactosamide alpha-2,6-sialyltranferase 1 (ST6GAL1), transcript variant 1, mRNA [NM_173216] |
| A_23_P94736 | 0,003538807 | down | -1,5662678 | ST6GALNAC4 | Homo sapiens ST6 (alpha-N-acetyl-neuraminyl-2,3-beta-galactosyl-1,3)-N-acetylgalactosaminide alpha-2,6-sialyltransferase 4 (ST6GALNAC4), transcript variant 1, mRNA [NM_175039] |
| A_23_P33093 | 0,04625036 | down | -1,8184602 | ST6GALNAC5 | Homo sapiens ST6 (alpha-N-acetyl-neuraminyl-2,3-beta-galactosyl-1,3)-N-acetylgalactosaminide alpha-2,6-sialyltransferase 5 (ST6GALNAC5), mRNA [NM_030965] |
| A_33_P3380751 | 0,001412595 | down | -1,6573055 | ST8SIA1 | Homo sapiens ST8 alpha-N-acetyl-neuraminide alpha-2,8-sialyltransferase 1 (ST8SIA1), mRNA [NM_003034] |
| A_33_P3291484 | 0,044667147 | up | 1,5696695 | ST8SIA4 | Homo sapiens ST8 alpha-N-acetyl-neuraminide alpha-2,8-sialyltransferase 4 (ST8SIA4), transcript variant 1, mRNA [NM_005668] |
| A_23_P162607 | 0,004162413 | down | -2,983613 | STAB2 | Homo sapiens stabilin 2 (STAB2), mRNA [NM_017564] |
| A_32_P152195 | 0,016831234 | down | -1,7888857 | STAC2 | Homo sapiens SH3 and cysteine rich domain 2 (STAC2), mRNA [NM_198993] |
| A_33_P3220663 | 0,002778671 | up | 1,6144528 | STAC3 | Homo sapiens SH3 and cysteine rich domain 3 (STAC3), transcript variant 1, mRNA [NM_145064] |
| A_33_P3304304 | 3,21E-04 | up | 2,5917065 | STAG2 | stromal antigen 2 [Source:HGNC Symbol;Acc:HGNC:11355] [ENST00000394478] |
| A_24_P374962 | 0,00277838 | up | 1,5480841 | STAG3L2 | Homo sapiens stromal antigen 3-like 2 (pseudogene) (STAG3L2), non-coding RNA [NR_040584] |
| A_21_P0011319 | 0,008284524 | down | -2,4625785 | STARD9 | Homo sapiens StAR-related lipid transfer (START) domain containing 9 (STARD9), mRNA [NM_020759] |
| A_21_P0011322 | 0,03867572 | up | 1,9692532 | STARD9 | Homo sapiens StAR-related lipid transfer (START) domain containing 9 (STARD9), mRNA [NM_020759] |
| A_23_P100795 | 0,003314072 | up | 1,6543717 | STAT3 | Homo sapiens signal transducer and activator of transcription 3 (acute-phase response factor) (STAT3), transcript variant 3, mRNA [NM_213662] |
| A_24_P116805 | 0,019763608 | up | 1,5172842 | STAT3 | Homo sapiens signal transducer and activator of transcription 3 (acute-phase response factor) (STAT3), transcript variant 3, mRNA [NM_213662] |
| A_23_P207367 | 0,022036076 | up | 1,5080404 | STAT5A | Homo sapiens signal transducer and activator of transcription 5A (STAT5A), transcript variant 2, mRNA [NM_003152] |
| A_33_P3380867 | 7,87E-04 | up | 1,6934508 | STAT5B | Homo sapiens signal transducer and activator of transcription 5B (STAT5B), mRNA [NM_012448] |
| A_33_P3265783 | 0,009251297 | up | 2,3780766 | STATH | Homo sapiens statherin (STATH), transcript variant 1, mRNA [NM_003154] |
| A_33_P3371650 | 0,001019261 | up | 2,8847733 | STK17B | Homo sapiens serine/threonine kinase 17b (apoptosis-inducing), mRNA (cDNA clone IMAGE:6280720), partial cds. [BC052561] |
| A_23_P253177 | 0,0302884 | up | 1,692294 | STK24 | Homo sapiens serine/threonine kinase 24 (STK24), transcript variant 2, mRNA [NM_001032296] |
| A_21_P0000774 | 0,003525023 | down | -2,1243298 | STK4-AS1 | Homo sapiens STK4 antisense RNA 1 (head to head) (STK4-AS1), long non-coding RNA [NR_038341] |
| A_33_P3632866 | 0,024707682 | down | -1,7073007 | STKLD1 | Homo sapiens serine/threonine kinase-like domain containing 1 (STKLD1), transcript variant 1, mRNA [NM_153710] |
| A_19_P00320974 | 0,008538269 | down | -2,3183875 | STMN1 | Homo sapiens stathmin 1 (STMN1), transcript variant 4, mRNA [NM_001145454] |
| A_21_P0013125 | 0,04575318 | up | 1,8549829 | STMND1 | Homo sapiens stathmin domain containing 1 (STMND1), mRNA [NM_001190766] |
| A_23_P302005 | 0,03479545 | up | 1,9865717 | STON1 | Homo sapiens stonin 1 (STON1), transcript variant 2, mRNA [NM_006873] |
| A_33_P3307495 | 0,010076329 | up | 4,406939 | STRA6 | Homo sapiens stimulated by retinoic acid 6 (STRA6), transcript variant 8, mRNA [NM_001199042] |
| A_21_P0011374 | 0,01429278 | down | -1,5925373 | STRC | Homo sapiens stereocilin (STRC), mRNA [NM_153700] |

| | | | | | |
|---|---|---|---|---|---|
| A_33_P3270519 | 0,024414364 | down | -1,5304848 | STRIP1 | Homo sapiens striatin interacting protein 1 (STRIP1), transcript variant 1, mRNA [NM_033088] |
| A_19_P00316199 | 0,001771349 | up | 1,7783793 | STRN | Homo sapiens striatin, calmodulin binding protein (STRN), mRNA [NM_003162] |
| A_19_P00316200 | 0,001438353 | up | 1,6647763 | STRN | Homo sapiens striatin, calmodulin binding protein (STRN), mRNA [NM_003162] |
| A_23_P392076 | 0,003681422 | up | 1,5448192 | STRN3 | Homo sapiens striatin, calmodulin binding protein 3 (STRN3), transcript variant 2, mRNA [NM_014574] |
| A_23_P168556 | 0,017168421 | up | 1,9830005 | STX1A | Homo sapiens syntaxin 1A (brain) (STX1A), transcript variant 1, mRNA [NM_004603] |
| A_33_P3326382 | 0,003554973 | down | -2,5165343 | STX1B | syntaxin 1B [Source:HGNC Symbol;Acc:HGNC:18539] [ENST00000215095] |
| A_23_P256375 | 0,00254102 | up | 1,5247668 | STX4 | Homo sapiens syntaxin 4 (STX4), transcript variant 3, mRNA [NM_004604] |
| A_23_P375566 | 0,006420092 | down | -1,6829679 | STXBP4 | Homo sapiens syntaxin binding protein 4 (STXBP4), mRNA [NM_178509] |
| A_32_P95462 | 0,002737095 | down | -2,1880145 | STXBP5L | Homo sapiens syntaxin binding protein 5-like (STXBP5L), mRNA [NM_014980] |
| A_23_P205713 | 6,08E-04 | down | -2,1435587 | STXBP6 | Homo sapiens syntaxin binding protein 6 (amisyn) (STXBP6), mRNA [NM_014178] |
| A_24_P10214 | 0,020628765 | up | 1,7991759 | STXBP6 | Homo sapiens syntaxin binding protein 6 (amisyn) (STXBP6), mRNA [NM_014178] |
| A_21_P0000093 | 0,04029708 | down | -1,5888788 | SUCLG2 | Homo sapiens succinate-CoA ligase, GDP-forming, beta subunit (SUCLG2), transcript variant 1, mRNA [NM_001177599] |
| A_24_P149902 | 0,02631148 | down | -1,702542 | SUFU | Homo sapiens suppressor of fused homolog (Drosophila) (SUFU), transcript variant 1, mRNA [NM_016169] |
| A_23_P425304 | 0,01276979 | up | 1,5624954 | SUFU | Homo sapiens suppressor of fused homolog (Drosophila) (SUFU), transcript variant 1, mRNA [NM_016169] |
| A_23_P145711 | 0,03995122 | down | -2,1568646 | SUGCT | Homo sapiens succinyl-CoA:glutarate-CoA transferase (SUGCT), transcript variant 4, mRNA [NM_024728] |
| A_23_P16562 | 0,004525502 | up | 1,9305263 | SUGP2 | Homo sapiens SURP and G patch domain containing 2 (SUGP2), transcript variant 1, mRNA [NM_001017392] |
| A_23_P106773 | 0,039838556 | down | -1,576894 | SULT1A2 | Homo sapiens sulfotransferase family, cytosolic, 1A, phenol-preferring, member 2 (SULT1A2), transcript variant 2, mRNA [NM_177528] |
| A_23_P51002 | 0,007016542 | up | 2,4616885 | SULT1C2 | Homo sapiens sulfotransferase family, cytosolic, 1C, member 2 (SULT1C2), transcript variant 2, mRNA [NM_176825] |
| A_33_P3391220 | 0,016811999 | down | -2,0781744 | SULT1C2P1 | Homo sapiens sulfotransferase family, cytosolic, 1C, member 2 pseudogene 1 (SULT1C2P1), non-coding RNA [NR_037191] |
| A_33_P3315564 | 0,004130895 | up | 3,4209287 | SULT1C3 | Homo sapiens sulfotransferase family, cytosolic, 1C, member 3 (SULT1C3), mRNA [NM_001008743] |
| A_23_P147070 | 0,006057499 | up | 1,9089094 | SUN1 | Sad1 and UNC84 domain containing 1 [Source:HGNC Symbol;Acc:HGNC:18587] [ENST00000340926] |
| A_23_P109229 | 0,032642856 | down | -2,6387162 | SUN5 | Homo sapiens Sad1 and UNC84 domain containing 5 (SUN5), mRNA [NM_080675] |
| A_23_P140050 | 5,35E-04 | down | -2,064423 | SUPT20H | Homo sapiens suppressor of Ty 20 homolog (S. cerevisiae) (SUPT20H), transcript variant 2, mRNA [NM_017569] |
| A_33_P3384284 | 2,26E-04 | down | -2,7224739 | SUPT20HL1 | Homo sapiens suppressor of Ty 20 homolog (S. cerevisiae)-like 1 (SUPT20HL1), mRNA [NM_001136234] |
| A_33_P3347152 | 0,002022004 | down | -1,9127482 | SUPT3H | suppressor of Ty 3 homolog (S. cerevisiae) [Source:HGNC Symbol;Acc:HGNC:11466] [ENST00000371458] |
| A_23_P210091 | 2,51E-04 | up | 1,6553069 | SUPT7L | Homo sapiens suppressor of Ty 7 (S. cerevisiae)-like (SUPT7L), transcript variant 1, mRNA [NM_014860] |
| A_33_P3394793 | 0,019623002 | up | 2,6560774 | SUSD2 | Homo sapiens sushi domain containing 2 (SUSD2), mRNA [NM_019601] |
| A_33_P3327663 | 2,38E-05 | down | -3,057104 | SUSD4 | sushi domain containing 4 [Source:HGNC Symbol;Acc:HGNC:25470] [ENST00000342943] |
| A_33_P3327657 | 0,001848582 | up | 2,1881092 | SUSD4 | sushi domain containing 4 [Source:HGNC Symbol;Acc:HGNC:25470] [ENST00000342943] |
| A_24_P259607 | 0,030329175 | up | 1,7629316 | SUSD4 | Homo sapiens sushi domain containing 4 (SUSD4), transcript variant 1, mRNA [NM_017982] |
| A_24_P238131 | 0,009549734 | up | 1,8852487 | SUSD5 | Homo sapiens sushi domain containing 5 (SUSD5), mRNA [NM_015551] |
| A_23_P371787 | 0,002571608 | up | 1,7613299 | SUSD6 | Homo sapiens sushi domain containing 6 (SUSD6), mRNA [NM_014734] |
| A_23_P23728 | 0,023567552 | down | -1,5253055 | SV2A | Homo sapiens synaptic vesicle glycoprotein 2A (SV2A), transcript variant 1, mRNA [NM_014849] |
| A_23_P35349 | 6,29E-05 | up | 2,0897424 | SVIL | Homo sapiens supervillin (SVIL), transcript variant 2, mRNA [NM_021738] |
| A_33_P3303464 | 0,004717279 | down | -1,7188963 | SWSAP1 | Homo sapiens SWIM-type zinc finger 7 associated protein 1 (SWSAP1), mRNA [NM_175871] |
| A_23_P115805 | 0,017703062 | up | 2,1787155 | SYCE1 | Homo sapiens synaptonemal complex central element protein 1 (SYCE1), transcript variant 4, mRNA [NM_001143764] |
| A_33_P3314673 | 0,009080916 | down | -2,198776 | SYCE1L | Homo sapiens synaptonemal complex central element protein 1-like (SYCE1L), mRNA [NM_001129979] |
| A_23_P210675 | 5,19E-04 | up | 2,709709 | SYCP2 | Homo sapiens synaptonemal complex protein 2 (SYCP2), mRNA [NM_014258] |
| A_23_P33847 | 2,17E-05 | down | -4,2669168 | SYN1 | Homo sapiens synapsin I (SYN1), transcript variant Ia, mRNA [NM_006950] |
| A_33_P3317613 | 0,049271774 | down | -1,8612275 | SYN2 | Homo sapiens synapsin II (SYN2), transcript variant IIa, mRNA [NM_133625] |
| A_23_P500861 | 0,021121373 | down | -2,161066 | SYNE1 | Homo sapiens spectrin repeat containing, nuclear envelope 1 (SYNE1), transcript variant 1, mRNA [NM_182961] |
| A_33_P3335920 | 2,87E-05 | down | -3,542771 | SYNE1 | Homo sapiens spectrin repeat containing, nuclear envelope 1 (SYNE1), transcript variant 1, mRNA [NM_182961] |
| A_21_P0014520 | 6,70E-06 | down | -4,0381513 | SYNE1-AS1 | Homo sapiens SYNE1 antisense RNA 1 (SYNE1-AS1), long non-coding RNA [NR_120501] |
| A_33_P3210278 | 0,007956684 | up | 1,5031699 | SYNE2 | Homo sapiens spectrin repeat containing, nuclear envelope 2 (SYNE2), transcript variant 5, mRNA [NM_182914] |
| A_33_P3324839 | 0,019012272 | down | -1,5145072 | SYNGAP1 | Homo sapiens synaptic Ras GTPase activating protein 1 (SYNGAP1), mRNA [NM_006772] |
| A_23_P164927 | 2,03E-05 | down | -3,6661375 | SYNGR4 | Homo sapiens synaptogyrin 4 (SYNGR4), mRNA [NM_012451] |
| A_33_P3354990 | 0,007211428 | down | -1,5309813 | SYNJ2 | synaptojanin 2 [Source:HGNC Symbol;Acc:HGNC:11504] [ENST00000449320] |
| A_23_P80718 | 0,002882583 | up | 1,5769912 | SYNPR | Homo sapiens synaptoporin (SYNPR), transcript variant 2, mRNA [NM_144642] |
| A_33_P3307486 | 0,00723893 | down | -1,5220464 | SYNRG | Homo sapiens synergin, gamma (SYNRG), transcript variant 7, mRNA [NM_001163547] |
| A_21_P0000004 | 0,011233006 | up | 1,5811169 | SYS1 | Homo sapiens Sys1 golgi trafficking protein (SYS1), transcript variant 2, mRNA [NM_001099791] |
| A_33_P3340525 | 0,008706681 | up | 1,5677933 | SYT10 | Homo sapiens synaptotagmin X (SYT10), mRNA [NM_198992] |
| A_33_P3353979 | 0,001603556 | down | -2,3321605 | SYT15 | Homo sapiens synaptotagmin XV (SYT15), transcript variant b, mRNA [NM_181519] |
| A_23_P163697 | 0,027520424 | up | 1,7787806 | SYT17 | Homo sapiens synaptotagmin XVII (SYT17), mRNA [NM_016524] |
| A_23_P55917 | 1,05E-05 | down | -5,1080933 | SYT3 | Homo sapiens synaptotagmin III (SYT3), transcript variant 1, mRNA [NM_032298] |
| A_33_P3235322 | 0,001129445 | down | -1,776362 | SYTL1 | Homo sapiens synaptotagmin-like 1 (SYTL1), transcript variant 2, mRNA [NM_032872] |
| A_23_P53193 | 0,03945926 | up | 1,7809179 | SYTL2 | Homo sapiens synaptotagmin-like 2 (SYTL2), transcript variant a, mRNA [NM_032943] |
| A_33_P3266730 | 0,047673915 | down | -1,8205093 | SYTL4 | Homo sapiens synaptotagmin-like 4 (SYTL4), transcript variant 1, mRNA [NM_080737] |
| A_33_P3373134 | 0,039410558 | down | -1,6901146 | SZT2 | Homo sapiens seizure threshold 2 homolog (mouse) (SZT2), mRNA [NM_015284] |
| A_33_P3373139 | 0,01874433 | up | 1,5288887 | SZT2 | seizure threshold 2 homolog (mouse) [Source:HGNC Symbol;Acc:HGNC:29040] [ENST00000470139] |
| A_23_P70733 | 0,0179462 | down | -3,5213706 | TAAR2 | Homo sapiens trace amine associated receptor 2 (TAAR2), transcript variant 1, mRNA [NM_001033080] |
| A_23_P255968 | 2,39E-04 | down | -3,118682 | TAAR5 | Homo sapiens trace amine associated receptor 5 (TAAR5), mRNA [NM_003967] |

| | | | | | |
|---|---|---|---|---|---|
| A_23_P168266 | 0,003970861 | up | 2,0324726 | TAAR9 | Homo sapiens trace amine associated receptor 9 (gene/pseudogene) (TAAR9), mRNA [NM_175057] |
| A_32_P162150 | 0,021909224 | up | 1,575039 | TAB3 | Homo sapiens TGF-beta activated kinase 1/MAP3K7 binding protein 3 (TAB3), mRNA [NM_152787] |
| A_33_P3258046 | 0,009542847 | down | -2,2829373 | TAC4 | Homo sapiens tachykinin 4 (hemokinin) (TAC4), transcript variant alpha, mRNA [NM_170685] |
| A_33_P3253877 | 0,03504794 | up | 1,7756906 | TACC1 | transforming, acidic coiled-coil containing protein 1 [Source:HGNC Symbol;Acc:HGNC:11522] [ENST00000518415] |
| A_24_P98249 | 0,00174088 | up | 1,5538617 | TACC1 | Homo sapiens transforming, acidic coiled-coil containing protein 1 (TACC1), transcript variant 1, mRNA [NM_006283] |
| A_23_P35148 | 0,004193523 | down | -1,6253346 | TAF13 | Homo sapiens TAF13 RNA polymerase II, TATA box binding protein (TBP)-associated factor, 18kDa (TAF13), mRNA [NM_005645] |
| A_33_P3257457 | 0,016303074 | down | -1,616389 | TAF1L | Homo sapiens TAF1 RNA polymerase II, TATA box binding protein (TBP)-associated factor, 210kDa-like (TAF1L), mRNA [NM_153809] |
| A_23_P80062 | 7,72E-05 | up | 2,0873044 | TAF4 | Homo sapiens TAF4 RNA polymerase II, TATA box binding protein (TBP)-associated factor, 135kDa (TAF4), mRNA [NM_003185] |
| A_33_P3242323 | 0,038226143 | down | -1,8742262 | TAF5L | TAF5-like RNA polymerase II, p300/CBP-associated factor (PCAF)-associated factor, 65kDa [Source:HGNC Symbol;Acc:HGNC:17304] [ENST00000366674] |
| A_24_P376441 | 0,004624005 | up | 1,756564 | TAF5L | Homo sapiens TAF5-like RNA polymerase II, p300/CBP-associated factor (PCAF)-associated factor, 65kDa (TAF5L), transcript variant 2, mRNA [NM_001025247] |
| A_23_P35309 | 0,002954134 | up | 1,7182579 | TAF5L | Homo sapiens TAF5-like RNA polymerase II, p300/CBP-associated factor (PCAF)-associated factor, 65kDa (TAF5L), transcript variant 1, mRNA [NM_014409] |
| A_23_P217621 | 0,01646985 | down | -2,1333218 | TAF7L | Homo sapiens TAF7-like RNA polymerase II, TATA box binding protein (TBP)-associated factor, 50kDa (TAF7L), transcript variant 1, mRNA [NM_024885] |
| A_24_P354724 | 0,014145938 | up | 1,6629279 | TAGAP | Homo sapiens T-cell activation RhoGTPase activating protein (TAGAP), transcript variant 2, mRNA [NM_054114] |
| A_23_P63371 | 0,001195285 | up | 2,6366453 | TAL1 | Homo sapiens T-cell acute lymphocytic leukemia 1 (TAL1), transcript variant 1, mRNA [NM_003189] |
| A_24_P942068 | 0,005337815 | up | 1,8964957 | TANC2 | Homo sapiens tetratricopeptide repeat, ankyrin repeat and coiled-coil containing 2 (TANC2), mRNA [NM_025185] |
| A_33_P3369452 | 0,001677061 | up | 1,5293593 | TANGO2 | Homo sapiens transport and golgi organization 2 homolog (Drosophila) (TANGO2), transcript variant 1, mRNA [NM_001283106] |
| A_23_P52058 | 0,009608224 | up | 1,5336201 | TARBP1 | Homo sapiens TAR (HIV-1) RNA binding protein 1 (TARBP1), mRNA [NM_005646] |
| A_21_P0011146 | 0,001075666 | up | 1,7323678 | TARDBP | Homo sapiens TAR DNA binding protein (TARDBP), mRNA [NM_007375] |
| A_23_P403955 | 0,004627589 | up | 2,1084597 | TARDBP | Homo sapiens TAR DNA binding protein (TARDBP), mRNA [NM_007375] |
| A_33_P3286973 | 2,13E-05 | down | -2,5789876 | TAS1R3 | taste receptor, type 1, member 3 [Source:HGNC Symbol;Acc:HGNC:15661] [ENST00000339381] |
| A_33_P3260989 | 0,04753429 | up | 1,7404476 | TAS1R3 | Homo sapiens taste receptor, type 1, member 3 (TAS1R3), mRNA [NM_152228] |
| A_23_P99080 | 0,001353492 | down | -2,7908025 | TAS2R13 | Homo sapiens taste receptor, type 2, member 13 (TAS2R13), mRNA [NM_023920] |
| A_23_P59825 | 0,003809263 | up | 1,507686 | TAS2R16 | Homo sapiens taste receptor, type 2, member 16 (TAS2R16), mRNA [NM_016945] |
| A_23_P340308 | 0,045174688 | down | -1,6742015 | TAS2R19 | Homo sapiens taste receptor, type 2, member 19 (TAS2R19), mRNA [NM_176888] |
| A_23_P416191 | 0,037701726 | down | -1,7293552 | TAS2R31 | Homo sapiens taste receptor, type 2, member 31 (TAS2R31), mRNA [NM_176885] |
| A_32_P309662 | 0,048940383 | down | -1,884459 | TAS2R39 | Homo sapiens taste receptor, type 2, member 39 (TAS2R39), mRNA [NM_176881] |
| A_24_P14902 | 0,002780172 | up | 1,6159124 | TAS2R42 | Homo sapiens taste receptor, type 2, member 42 (TAS2R42), mRNA [NM_181429] |
| A_23_P367013 | 1,56E-04 | down | -4,764367 | TAS2R60 | Homo sapiens taste receptor, type 2, member 60 (TAS2R60), mRNA [NM_177437] |
| A_23_P336506 | 5,16E-04 | down | -5,0180306 | TAS2R9 | Homo sapiens taste receptor, type 2, member 9 (TAS2R9), mRNA [NM_023917] |
| A_24_P330633 | 0,039774437 | down | -1,5567812 | TAT | Homo sapiens tyrosine aminotransferase (TAT), mRNA [NM_000353] |
| A_23_P12858 | 0,005354411 | down | -2,0258272 | TBATA | Homo sapiens thymus, brain and testes associated (TBATA), mRNA [NM_152710] |
| A_33_P3370812 | 0,024417816 | down | -1,5537984 | TBC1D1 | Homo sapiens TBC1 (tre-2/USP6, BUB2, cdc16) domain family, member 1, mRNA (cDNA clone IMAGE:5211948), with apparent retained intron. [BC028196] |
| A_23_P24215 | 8,95E-04 | up | 1,6316078 | TBC1D12 | Homo sapiens TBC1 domain family, member 12 (TBC1D12), mRNA [NM_015188] |
| A_23_P58983 | 0,004236145 | up | 1,5933094 | TBC1D22B | Homo sapiens TBC1 domain family, member 22B (TBC1D22B), transcript variant 1, mRNA [NM_017772] |
| A_23_P391637 | 0,010068996 | up | 1,5152237 | TBC1D24 | Homo sapiens TBC1 domain family, member 24 (TBC1D24), transcript variant 2, mRNA [NM_020705] |
| A_33_P3267230 | 0,001090529 | up | 2,024126 | TBC1D3B | Homo sapiens TBC1 domain family, member 3B (TBC1D3B), mRNA [NM_001001417] |
| A_33_P3344991 | 7,45E-07 | down | -2,7088768 | TBC1D3L | Homo sapiens cDNA FLJ16441 fis, clone BRAMY3003205, highly similar to TBC1 domain family member 3. [AK122833] |
| A_21_P0014921 | 5,53E-05 | up | 2,2779322 | TBC1D3P1-DHX40P1 | Homo sapiens TBC1D3P1-DHX40P1 readthrough transcribed pseudogene (TBC1D3P1-DHX40P1), non-coding RNA [NR_002924] |
| A_21_P0011619 | 0,005528416 | up | 1,6338001 | TBC1D3P5 | Homo sapiens TBC1 domain family, member 3 pseudogene 5 (TBC1D3P5), non-coding RNA [NR_033892] |
| A_23_P111273 | 0,007258558 | down | -1,5076747 | TBC1D7 | Homo sapiens TBC1 domain family, member 7 (TBC1D7), transcript variant 1, mRNA [NM_016495] |
| A_33_P3312194 | 0,024062831 | up | 1,5189587 | TBCB | tubulin folding cofactor B [Source:HGNC Symbol;Acc:HGNC:1989] [ENST00000392178] |
| A_23_P121345 | 4,68E-04 | up | 1,6565269 | TBCCD1 | Homo sapiens TBCC domain containing 1 (TBCCD1), transcript variant 2, mRNA [NM_018138] |
| A_23_P212552 | 7,51E-05 | up | 1,9847479 | TBL1XR1 | Homo sapiens transducin (beta)-like 1 X-linked receptor 1 (TBL1XR1), mRNA [NM_024665] |
| A_23_P5685 | 0,003378721 | up | 1,5668216 | TBR1 | Homo sapiens T-box, brain, 1 (TBR1), mRNA [NM_006593] |
| A_23_P40527 | 0,008303398 | up | 1,5918165 | TBX1 | Homo sapiens T-box 1 (TBX1), transcript variant A, mRNA [NM_080646] |
| A_32_P538017 | 0,03121254 | down | -1,5202376 | TBX10 | Homo sapiens T-box 10 (TBX10), mRNA [NM_005995] |
| A_23_P137705 | 0,003956246 | up | 1,6109941 | TBX19 | Homo sapiens T-box 19 (TBX19), mRNA [NM_005149] |
| A_33_P3307886 | 0,03623908 | down | -1,5560447 | TBXA2R | Homo sapiens thromboxane A2 receptor (TBXA2R), transcript variant a, mRNA [NM_001060] |
| A_24_P274814 | 0,001707639 | down | -2,4079053 | TBXAS1 | Homo sapiens thromboxane A synthase 1 (platelet) (TBXAS1), transcript variant 2, mRNA [NM_030984] |
| A_32_P214925 | 0,013152985 | down | -2,7147574 | TCAF2 | Homo sapiens family with sequence similarity 115, member C (FAM115C), transcript variant 2, mRNA [NM_173678] |
| A_24_P237912 | 0,010329962 | up | 2,4790177 | TCAF2 | Homo sapiens family with sequence similarity 115, member C (FAM115C), transcript variant 3, mRNA [NM_001130026] |
| A_23_P107051 | 0,008358733 | up | 1,5887583 | TCAP | Homo sapiens titin-cap (TCAP), mRNA [NM_003673] |
| A_24_P126651 | 0,006246409 | down | -2,0935302 | TCEA1 | Homo sapiens transcription elongation factor A (SII), 1 (TCEA1), transcript variant 1, mRNA [NM_006756] |
| A_33_P3400057 | 4,21E-04 | down | -2,321735 | TCEANC | Homo sapiens transcription elongation factor A (SII) N-terminal and central domain containing (TCEANC), transcript variant 2, mRNA [NM_001297563] |
| A_23_P74097 | 0,001666558 | up | 1,6200799 | TCEB3 | Homo sapiens transcription elongation factor B (SIII), polypeptide 3 (110kDa, elongin A) (TCEB3), mRNA [NM_003198] |
| A_24_P122403 | 0,002662758 | up | 1,5388453 | TCEB3 | Homo sapiens transcription elongation factor B (SIII), polypeptide 3 (110kDa, elongin A) (TCEB3), mRNA [NM_003198] |
| A_23_P133365 | 0,005858981 | up | 1,5510819 | TCERG1 | Homo sapiens transcription elongation regulator 1 (TCERG1), transcript variant 1, mRNA [NM_006706] |
| A_23_P357717 | 0,012369526 | down | -2,0700738 | TCL1A | Homo sapiens T-cell leukemia/lymphoma 1A (TCL1A), transcript variant 1, mRNA [NM_021966] |
| A_23_P70598 | 0,001152422 | up | 2,0072594 | TCP11 | Homo sapiens t-complex 11, testis-specific (TCP11), transcript variant 2, mRNA [NM_018679] |
| A_33_P3375185 | 0,001623307 | down | -1,8151797 | TCTEX1D4 | Homo sapiens Tctex1 domain containing 4 (TCTEX1D4), mRNA [NM_001013632] |
| A_23_P80974 | 0,006150122 | up | 1,5368067 | TDO2 | Homo sapiens tryptophan 2,3-dioxygenase (TDO2), mRNA [NM_005651] |

| | | | | | |
|---|---|---|---|---|---|
| A_33_P3373329 | 0,015174741 | down | -1,6731936 | TDRD1 | Homo sapiens tudor domain containing 1 (TDRD1), mRNA [NM_198795] |
| A_33_P3267248 | 0,00712747 | up | 1,6702461 | TDRD10 | Homo sapiens tudor domain containing 10 (TDRD10), transcript variant 2, mRNA [NM_182499] |
| A_33_P3232960 | 0,003624465 | up | 1,5095251 | TDRD6 | Homo sapiens tudor domain containing 6 (TDRD6), transcript variant 1, mRNA [NM_001010870] |
| A_19_P00316714 | 0,02462068 | down | -2,1763072 | TDRG1 | Homo sapiens testis development related 1 (non-protein coding) (TDRG1), long non-coding RNA [NR_024015] |
| A_23_P82000 | 8,40E-04 | up | 1,7618442 | TEAD3 | Homo sapiens TEA domain family member 3 (TEAD3), mRNA [NM_003214] |
| A_33_P3305958 | 7,41E-04 | down | -1,8166286 | TECPR2 | Homo sapiens tectonin beta-propeller repeat containing 2 (TECPR2), transcript variant 2, mRNA [NM_001172631] |
| A_23_P207245 | 0,011572216 | up | 2,2441416 | TEKT3 | Homo sapiens tektin 3 (TEKT3), mRNA [NM_031898] |
| A_33_P3283863 | 0,002991174 | down | -2,1305807 | TENM1 | Homo sapiens teneurin transmembrane protein 1 (TENM1), transcript variant 1, mRNA [NM_001163278] |
| A_23_P76538 | 3,16E-04 | down | -3,702577 | TESC | Homo sapiens tescalcin (TESC), transcript variant 1, mRNA [NM_017899] |
| A_33_P3327265 | 2,32E-04 | down | -4,9827123 | TESPA1 | Homo sapiens thymocyte expressed, positive selection associated 1 (TESPA1), transcript variant 1, mRNA [NM_001136030] |
| A_19_P00806490 | 0,005431165 | down | -2,1822546 | TET1 | Homo sapiens tet methylcytosine dioxygenase 1 (TET1), mRNA [NM_030625] |
| A_33_P3331335 | 0,001307932 | up | 2,3462172 | TET3 | Homo sapiens tet methylcytosine dioxygenase 3 (TET3), mRNA [NM_001287491] |
| A_23_P73637 | 0,013376932 | down | -1,5638967 | TEX13B | Homo sapiens testis expressed 13B (TEX13B), mRNA [NM_031273] |
| A_33_P3261695 | 0,027087625 | up | 1,9602823 | TEX22 | Homo sapiens testis expressed 22 (TEX22), mRNA [NM_001195082] |
| A_21_P0000752 | 0,03841041 | up | 2,740108 | TEX26-AS1 | Homo sapiens TEX26 antisense RNA 1 (TEX26-AS1), transcript variant 2, long non-coding RNA [NR_038288] |
| A_24_P56330 | 0,003868824 | up | 1,5797911 | TEX29 | Homo sapiens testis expressed 29 (TEX29), transcript variant 2, mRNA [NM_152324] |
| A_32_P97862 | 0,033858668 | up | 2,161101 | TEX38 | Homo sapiens testis expressed 38 (TEX38), transcript variant 1, mRNA [NM_001145474] |
| A_21_P0012026 | 0,001491311 | down | -2,5579803 | TEX41 | testis expressed 41 (non-protein coding) [Source:HGNC Symbol;Acc:HGNC:48667] [ENST00000432608] |
| A_33_P3361546 | 0,03297328 | up | 1,5392259 | TFAP2A | Homo sapiens transcription factor AP-2 alpha (activating enhancer binding protein 2 alpha) (TFAP2A), transcript variant 2, mRNA [NM_001032280] |
| A_23_P386973 | 0,002409866 | up | 1,7987922 | TFAP2D | Homo sapiens transcription factor AP-2 delta (activating enhancer binding protein 2 delta) (TFAP2D), mRNA [NM_172238] |
| A_23_P141092 | 0,002279019 | down | -1,9685519 | TFAP4 | Homo sapiens transcription factor AP-4 (activating enhancer binding protein 4) (TFAP4), mRNA [NM_003223] |
| A_23_P5301 | 0,001109305 | up | 2,7775645 | TFCP2L1 | Homo sapiens transcription factor CP2-like 1 (TFCP2L1), mRNA [NM_014553] |
| A_33_P3224710 | 0,036867216 | down | -1,8219358 | TFEC | Homo sapiens transcription factor EC (TFEC), transcript variant 1, mRNA [NM_012252] |
| A_32_P184394 | 0,049562704 | up | 1,5260336 | TFEC | Homo sapiens transcription factor EC (TFEC), transcript variant 1, mRNA [NM_012252] |
| A_23_P57364 | 0,004548416 | down | -1,9760828 | TFF2 | Homo sapiens trefoil factor 2 (TFF2), mRNA [NM_005423] |
| A_33_P3334305 | 0,011367063 | down | -2,5334983 | TFF3 | Homo sapiens trefoil factor 3 (intestinal) (TFF3), mRNA [NM_003226] |
| A_23_P377291 | 0,002573235 | up | 2,3051026 | TGFA | Homo sapiens transforming growth factor, alpha (TGFA), transcript variant 1, mRNA [NM_003236] |
| A_33_P3313825 | 0,02549217 | up | 1,7617736 | TGFBR2 | PREDICTED: Homo sapiens transforming growth factor, beta receptor II (70/80kDa) (TGFBR2), transcript variant X1, mRNA [XM_006713316] |
| A_23_P211957 | 0,00181537 | up | 2,3267586 | TGFBR2 | Homo sapiens transforming growth factor, beta receptor II (70/80kDa) (TGFBR2), transcript variant 1, mRNA [NM_001024847] |
| A_33_P3311717 | 1,02E-05 | down | -2,9635978 | TGIF1 | Homo sapiens TGFB-induced factor homeobox 1 (TGIF1), transcript variant 5, mRNA [NM_173209] |
| A_23_P153197 | 8,17E-04 | up | 1,7488767 | TGIF1 | Homo sapiens TGFB-induced factor homeobox 1 (TGIF1), transcript variant 1, mRNA [NM_170695] |
| A_23_P41241 | 0,003828574 | up | 1,5058489 | TGM4 | Homo sapiens transglutaminase 4 (TGM4), mRNA [NM_003241] |
| A_33_P3210463 | 1,90E-08 | down | -3,9146557 | TGM6 | Homo sapiens transglutaminase 6 (TGM6), transcript variant 1, mRNA [NM_198994] |
| A_24_P413988 | 0,00339076 | up | 1,5886073 | TGOLN2 | Homo sapiens trans-golgi network protein 2 (TGOLN2), transcript variant 1, mRNA [NM_006464] |
| A_24_P101786 | 0,004257472 | down | -1,8038639 | THADA | Homo sapiens thyroid adenoma associated (THADA), transcript variant 5, mRNA [NM_001271644] |
| A_24_P928510 | 0,04481491 | down | -1,7938004 | THEM4 | Homo sapiens thioesterase superfamily member 4 (THEM4), mRNA [NM_053055] |
| A_24_P85775 | 0,025469813 | up | 1,6504298 | THEMIS2 | Homo sapiens thymocyte selection associated family member 2 (THEMIS2), transcript variant 2, mRNA [NM_001039477] |
| A_24_P377124 | 0,009573123 | down | -2,5390458 | THPO | Homo sapiens thrombopoietin (THPO), transcript variant 1, mRNA [NM_000460] |
| A_33_P3409974 | 0,002099822 | up | 2,5549796 | THRB | Homo sapiens thyroid hormone receptor, beta (THRB), transcript variant 4, mRNA [NM_001252634] |
| A_23_P363954 | 0,029226936 | up | 1,7492441 | THRSP | Homo sapiens thyroid hormone responsive (THRSP), mRNA [NM_003251] |
| A_24_P405430 | 0,013205801 | up | 1,5328708 | TIA1 | Homo sapiens TIA1 cytotoxic granule-associated RNA binding protein (TIA1), transcript variant 2, mRNA [NM_022173] |
| A_23_P90311 | 0,019222543 | up | 1,604598 | TICAM1 | Homo sapiens toll-like receptor adaptor molecule 1 (TICAM1), mRNA [NM_182919] |
| A_23_P129577 | 0,014836741 | up | 1,8727757 | TIGD7 | Homo sapiens tigger transposable element derived 7 (TIGD7), mRNA [NM_033208] |
| A_33_P3235213 | 0,0298507 | down | -1,948343 | TIGIT | Homo sapiens T cell immunoreceptor with Ig and ITIM domains (TIGIT), mRNA [NM_173799] |
| A_21_P0000013 | 6,13E-04 | down | -1,6694846 | TIMM8A | Homo sapiens translocase of inner mitochondrial membrane 8 homolog A (yeast) (TIMM8A), transcript variant 2, mRNA [NM_001145951] |
| A_33_P3421656 | 0,006840956 | up | 1,8606685 | TINAG | Homo sapiens tubulointerstitial nephritis antigen (TINAG), mRNA [NM_014464] |
| A_23_P397856 | 0,007624481 | down | -1,6988524 | TIRAP | Homo sapiens toll-interleukin 1 receptor (TIR) domain containing adaptor protein (TIRAP), transcript variant 3, mRNA [NM_001039661] |
| A_23_P202905 | 0,018236134 | up | 1,5477827 | TIRAP | Homo sapiens toll-interleukin 1 receptor (TIR) domain containing adaptor protein (TIRAP), transcript variant 2, mRNA [NM_148910] |
| A_23_P81811 | 0,006519184 | up | 1,69467 | TJAP1 | Homo sapiens tight junction associated protein 1 (peripheral) (TJAP1), transcript variant 4, mRNA [NM_080604] |
| A_24_P193435 | 0,003998066 | up | 1,6972336 | TJP1 | Homo sapiens tight junction protein 1 (TJP1), transcript variant 1, mRNA [NM_003257] |
| A_33_P3215529 | 6,39E-04 | down | -2,193177 | TJP2 | tight junction protein 2 [Source:HGNC Symbol;Acc:HGNC:11828] [ENST00000413932] |
| A_23_P9293 | 0,001013818 | up | 2,1244488 | TJP2 | Homo sapiens tight junction protein 2 (TJP2), transcript variant 1, mRNA [NM_004817] |
| A_24_P201153 | 0,027023602 | up | 1,7102287 | TJP2 | Homo sapiens tight junction protein 2 (TJP2), transcript variant 2, mRNA [NM_201629] |
| A_23_P259901 | 0,004215826 | down | -2,026023 | TKTL1 | Homo sapiens transketolase-like 1 (TKTL1), transcript variant 1, mRNA [NM_012253] |
| A_33_P3345210 | 0,01487583 | down | -1,7977148 | TLCD2 | Homo sapiens TLC domain containing 2 (TLCD2), mRNA [NM_001164407] |
| A_33_P3217731 | 0,001285348 | up | 1,8934226 | TLE1 | transducin-like enhancer of split 1 (E(sp1) homolog, Drosophila) [Source:HGNC Symbol;Acc:HGNC:11837] [ENST00000376484] |
| A_23_P342934 | 7,65E-05 | up | 2,0191696 | TLE3 | Homo sapiens transducin-like enhancer of split 3 (TLE3), transcript variant 1, mRNA [NM_005078] |
| A_21_P0000156 | 0,010980592 | down | -2,4066725 | TLL1 | Homo sapiens tolloid-like 1 (TLL1), transcript variant 2, mRNA [NM_001204760] |
| A_33_P3264815 | 8,16E-04 | down | -1,676049 | TLN2 | Homo sapiens talin 2 (TLN2), mRNA [NM_015059] |
| A_23_P92499 | 3,17E-04 | up | 6,9574194 | TLR2 | Homo sapiens toll-like receptor 2 (TLR2), mRNA [NM_003264] |
| A_24_P69538 | 0,041333888 | up | 1,9853272 | TLR4 | Homo sapiens toll-like receptor 4 (TLR4), transcript variant 1, mRNA [NM_138554] |

| | | | | | |
|---|---|---|---|---|---|
| A_33_P3263423 | 0,002148818 | down | -1,6689425 | TLR5 | Homo sapiens toll-like receptor 5 (TLR5), mRNA [NM_003268] |
| A_23_P73837 | 0,008896939 | up | 1,7144734 | TLR8 | Homo sapiens toll-like receptor 8 (TLR8), transcript variant 1, mRNA [NM_016610] |
| A_33_P3380807 | 0,03491465 | down | -2,0183103 | TLR9 | Homo sapiens toll-like receptor 9 (TLR9), mRNA [NM_017442] |
| A_23_P77117 | 0,04764301 | down | -1,6495701 | TM6SF1 | Homo sapiens transmembrane 6 superfamily member 1 (TM6SF1), transcript variant 1, mRNA [NM_023003] |
| A_23_P142403 | 8,10E-04 | down | -2,7095315 | TM6SF2 | Homo sapiens transmembrane 6 superfamily member 2 (TM6SF2), mRNA [NM_001001524] |
| A_33_P3308307 | 0,001983899 | up | 2,0538886 | TMC1 | Homo sapiens transmembrane channel-like 1 (TMC1), mRNA [NM_138691] |
| A_24_P305933 | 0,048089523 | up | 1,5275813 | TMCC3 | Homo sapiens transmembrane and coiled-coil domain family 3 (TMCC3), transcript variant 1, mRNA [NM_020698] |
| A_23_P355067 | 0,023844171 | up | 1,5109117 | TMCO1 | Homo sapiens transmembrane and coiled-coil domains 1 (TMCO1), transcript variant 1, mRNA [NM_019026] |
| A_32_P87649 | 0,045854766 | down | -1,9546345 | TMCO2 | Homo sapiens transmembrane and coiled-coil domains 2 (TMCO2), mRNA [NM_001008740] |
| A_23_P158277 | 0,023869367 | up | 1,6801027 | TMCO4 | Homo sapiens transmembrane and coiled-coil domains 4 (TMCO4), mRNA [NM_181719] |
| A_33_P3350858 | 0,013168083 | up | 1,8672395 | TMCO5B | Homo sapiens transmembrane and coiled-coil domains 5B, pseudogene (TMCO5B), non-coding RNA [NR_046005] |
| A_21_P0012765 | 7,78E-04 | up | 1,869087 | TMED11P | Homo sapiens transmembrane emp24 protein transport domain containing 11, pseudogene (TMED11P), non-coding RNA [NR_033768] |
| A_33_P3327270 | 0,004760842 | down | -2,3287694 | TMED7-TICAM2 | Homo sapiens TMED7-TICAM2 readthrough (TMED7-TICAM2), transcript variant 2, mRNA [NM_001164469] |
| A_23_P125383 | 0,025502855 | up | 2,4258204 | TMEFF2 | Homo sapiens transmembrane protein with EGF-like and two follistatin-like domains 2 (TMEFF2), mRNA [NM_016192] |
| A_23_P349966 | 0,00828659 | up | 1,5732754 | TMEM130 | Homo sapiens transmembrane protein 130 (TMEM130), transcript variant 2, mRNA [NM_152913] |
| A_32_P153812 | 0,007699005 | up | 2,5732741 | TMEM132B | Homo sapiens transmembrane protein 132B (TMEM132B), transcript variant 1, mRNA [NM_052907] |
| A_24_P6626 | 0,019462455 | up | 1,8850728 | TMEM132B | Homo sapiens transmembrane protein 132B (TMEM132B), transcript variant 1, mRNA [NM_052907] |
| A_24_P127075 | 4,43E-04 | down | -1,8790023 | TMEM135 | Homo sapiens transmembrane protein 135 (TMEM135), transcript variant 1, mRNA [NM_022918] |
| A_24_P372134 | 0,00169985 | down | -2,6296895 | TMEM140 | Homo sapiens transmembrane protein 140 (TMEM140), mRNA [NM_018295] |
| A_33_P3257279 | 0,003539773 | down | -1,848721 | TMEM145 | Homo sapiens transmembrane protein 145 (TMEM145), mRNA [NM_173633] |
| A_21_P0000784 | 0,027640052 | up | 1,5818982 | TMEM147-AS1 | Homo sapiens TMEM147 antisense RNA 1 (TMEM147-AS1), long non-coding RNA [NR_038396] |
| A_23_P354217 | 0,002540597 | down | -2,0574203 | TMEM151A | Homo sapiens transmembrane protein 151A (TMEM151A), mRNA [NM_153266] |
| A_23_P369899 | 0,025525356 | up | 2,1845257 | TMEM158 | Homo sapiens transmembrane protein 158 (gene/pseudogene) (TMEM158), mRNA [NM_015444] |
| A_23_P344594 | 0,008980894 | down | -2,6200573 | TMEM174 | Homo sapiens transmembrane protein 174 (TMEM174), mRNA [NM_153217] |
| A_33_P3249716 | 0,006870549 | down | -1,5627234 | TMEM178A | transmembrane protein 178A [Source:HGNC Symbol;Acc:HGNC:28517] [ENST00000437068] |
| A_24_P93206 | 0,030346766 | down | -2,3288531 | TMEM179 | Homo sapiens transmembrane protein 179 (TMEM179), transcript variant 2, mRNA [NM_001286390] |
| A_23_P331943 | 2,36E-04 | up | 1,7875054 | TMEM181 | Homo sapiens transmembrane protein 181 (TMEM181), mRNA [NM_020823] |
| A_23_P18739 | 3,87E-05 | up | 2,4748974 | TMEM184C | Homo sapiens transmembrane protein 184C (TMEM184C), mRNA [NM_018241] |
| A_33_P3259801 | 0,003059276 | up | 1,6699651 | TMEM189 | Homo sapiens transmembrane protein 189 (TMEM189), transcript variant 1, mRNA [NM_199129] |
| A_24_P126628 | 2,54E-04 | up | 2,0323074 | TMEM194A | Homo sapiens transmembrane protein 194A (TMEM194A), transcript variant 2, mRNA [NM_015257] |
| A_33_P3389261 | 0,013637642 | down | -1,9081903 | TMEM194B | Homo sapiens transmembrane protein 194B (TMEM194B), mRNA [NM_001142645] |
| A_23_P94552 | 0,00720488 | up | 1,5140795 | TMEM2 | Homo sapiens transmembrane protein 2 (TMEM2), transcript variant 1, mRNA [NM_013390] |
| A_24_P933011 | 1,52E-04 | down | -2,0849667 | TMEM200C | Homo sapiens transmembrane protein 200C (TMEM200C), mRNA [NM_001080209] |
| A_33_P3239417 | 0,03114558 | up | 2,1724145 | TMEM200C | Homo sapiens transmembrane protein 200C (TMEM200C), mRNA [NM_001080209] |
| A_24_P522631 | 0,04556544 | down | -1,8878886 | TMEM201 | Homo sapiens transmembrane protein 201 (TMEM201), transcript variant 1, mRNA [NM_001130924] |
| A_23_P26062 | 0,038354557 | up | 2,1736763 | TMEM202 | Homo sapiens transmembrane protein 202 (TMEM202), mRNA [NM_001080462] |
| A_33_P3270996 | 0,005080934 | up | 1,5025216 | TMEM207 | Homo sapiens transmembrane protein 207 (TMEM207), mRNA [NM_207316] |
| A_33_P3233784 | 0,016025638 | down | -3,2363856 | TMEM211 | Homo sapiens transmembrane protein 211 (TMEM211), mRNA [NM_001001663] |
| A_24_P706953 | 0,004520026 | up | 1,5907991 | TMEM213 | Homo sapiens transmembrane protein 213 (TMEM213), mRNA [NM_001085429] |
| A_33_P3266639 | 0,011555171 | up | 1,545429 | TMEM220 | Homo sapiens transmembrane protein 220 (TMEM220), mRNA [NM_001004313] |
| A_33_P3411612 | 0,008939663 | down | -3,4461539 | TMEM221 | Homo sapiens transmembrane protein 221 (TMEM221), mRNA [NM_001190844] |
| A_24_P381975 | 6,25E-04 | down | -2,2215624 | TMEM231 | Homo sapiens transmembrane protein 231 (TMEM231), transcript variant 1, mRNA [NM_001077416] |
| A_32_P489986 | 0,004345575 | down | -2,3774643 | TMEM232 | Homo sapiens transmembrane protein 232 (TMEM232), mRNA [NM_001039763] |
| A_33_P3236558 | 0,004936373 | down | -1,5385344 | TMEM234 | Homo sapiens transmembrane protein 234 (TMEM234), mRNA [NM_019118] |
| A_33_P3362153 | 0,035983607 | down | -1,659743 | TMEM238 | Homo sapiens transmembrane protein 238 (TMEM238), mRNA [NM_001190764] |
| A_33_P3219459 | 0,001092172 | down | -1,9866561 | TMEM240 | Homo sapiens transmembrane protein 240 (TMEM240), mRNA [NM_001114748] |
| A_33_P3282597 | 0,016761415 | up | 1,6388777 | TMEM241 | transmembrane protein 241 [Source:HGNC Symbol;Acc:HGNC:31723] [ENST00000580025] |
| A_24_P132633 | 4,68E-04 | down | -2,5473466 | TMEM252 | Homo sapiens transmembrane protein 252 (TMEM252), mRNA [NM_153237] |
| A_33_P3211327 | 0,001855039 | down | -2,7316775 | TMEM262 | transmembrane protein 262 [Source:HGNC Symbol;Acc:HGNC:49389] [ENST00000334821] |
| A_21_P0000560 | 0,040213693 | down | -1,5650579 | TMEM30C | Homo sapiens transmembrane protein 30C (TMEM30C), transcript variant 2, non-coding RNA [NR_028358] |
| A_33_P3225357 | 0,004249297 | up | 1,578592 | TMEM30C | Homo sapiens transmembrane protein 30C (TMEM30C), transcript variant 1, non-coding RNA [NR_028357] |
| A_33_P3423270 | 0,011475662 | up | 1,5278215 | TMEM40 | Homo sapiens transmembrane protein 40 (TMEM40), transcript variant 1, mRNA [NM_001284406] |
| A_33_P3327567 | 0,031175122 | up | 1,5246078 | TMEM41A | transmembrane protein 41A [Source:HGNC Symbol;Acc:HGNC:30544] [ENST00000467061] |
| A_21_P0014585 | 0,002921852 | down | -2,284614 | TMEM44-AS1 | Homo sapiens TMEM44 antisense RNA 1 (TMEM44-AS1), transcript variant 1, long non-coding RNA [NR_047573] |
| A_24_P305623 | 0,004671027 | down | -1,6904352 | TMEM50B | Homo sapiens transmembrane protein 50B (TMEM50B), transcript variant 1, mRNA [NM_006134] |
| A_23_P318938 | 2,13E-04 | down | -2,6094017 | TMEM52B | Homo sapiens transmembrane protein 52B (TMEM52B), transcript variant 1, mRNA [NM_153022] |
| A_33_P3332393 | 0,003235944 | down | -1,7774554 | TMEM53 | transmembrane protein 53 [Source:HGNC Symbol;Acc:HGNC:26186] [ENST00000372244] |
| A_23_P422083 | 0,001214894 | down | -1,9114054 | TMEM55A | Homo sapiens transmembrane protein 55A (TMEM55A), mRNA [NM_018710] |
| A_33_P3332284 | 0,00651513 | up | 1,6417179 | TMEM59 | transmembrane protein 59 [Source:HGNC Symbol;Acc:HGNC:1239] [ENST00000371337] |
| A_23_P167856 | 0,044093177 | down | -1,6331154 | TMEM63B | Homo sapiens transmembrane protein 63B (TMEM63B), mRNA [NM_018426] |
| A_33_P3406661 | 0,022777172 | up | 2,0687163 | TMEM63C | Homo sapiens transmembrane protein 63C (TMEM63C), mRNA [NM_020431] |

| | | | | | |
|---|---|---|---|---|---|
| A_24_P171058 | 0,006507585 | up | 1,5541812 | TMEM64 | Homo sapiens transmembrane protein 64 (TMEM64), transcript variant 1, mRNA [NM_001008495] |
| A_24_P704446 | 0,010996197 | down | -1,9674433 | TMEM72 | Homo sapiens transmembrane protein 72 (TMEM72), mRNA [NM_001123376] |
| A_33_P3722782 | 0,004140231 | up | 2,1367366 | TMEM72-AS1 | Homo sapiens TMEM72 antisense RNA 1 (TMEM72-AS1), long non-coding RNA [NR_033842] |
| A_24_P341344 | 0,004393045 | down | -2,2883487 | TMEM75 | Homo sapiens transmembrane protein 75 (TMEM75), long non-coding RNA [NR_103558] |
| A_23_P46690 | 0,029625282 | up | 1,5715126 | TMEM81 | Homo sapiens transmembrane protein 81 (TMEM81), mRNA [NM_203376] |
| A_33_P3326553 | 0,006993435 | down | -1,7355269 | TMEM82 | transmembrane protein 82 [Source:HGNC Symbol;Acc:HGNC:32350] [ENST00000375782] |
| A_24_P65098 | 0,019066507 | up | 1,7904749 | TMEM87A | Homo sapiens transmembrane protein 87A (TMEM87A), transcript variant 2, mRNA [NM_001110503] |
| A_24_P317827 | 0,006681121 | down | -1,6428399 | TMEM8B | Homo sapiens transmembrane protein 8B (TMEM8B), transcript variant 1, mRNA [NM_001042590] |
| A_33_P3277611 | 4,34E-04 | down | -3,7270422 | TMEM8C | Homo sapiens transmembrane protein 8C (TMEM8C), mRNA [NM_001080483] |
| A_24_P307289 | 0,002629065 | down | -2,6153393 | TMEM95 | Homo sapiens transmembrane protein 95 (TMEM95), mRNA [NM_198154] |
| A_33_P3346267 | 0,018056259 | up | 1,7210217 | TMIGD1 | Homo sapiens transmembrane and immunoglobulin domain containing 1 (TMIGD1), mRNA [NM_206832] |
| A_33_P3412613 | 4,39E-05 | down | -2,155985 | TMPO | Homo sapiens thymopoietin (TMPO), transcript variant 2, mRNA [NM_001032283] |
| A_21_P0012777 | 0,033450183 | up | 2,2758837 | TMPRSS11BNL | Homo sapiens TMPRSS11B N-terminal like, pseudogene (TMPRSS11BNL), non-coding RNA [NR_104048] |
| A_21_P0000760 | 3,83E-04 | down | -2,4461968 | TMPRSS4-AS1 | Homo sapiens TMPRSS4 antisense RNA 1 (TMPRSS4-AS1), long non-coding RNA [NR_038318] |
| A_23_P211493 | 0,001073249 | down | -2,1346393 | TMPRSS6 | Homo sapiens transmembrane protease, serine 6 (TMPRSS6), transcript variant 2, mRNA [NM_153609] |
| A_33_P3814473 | 0,003317247 | up | 2,0750904 | TMSB4XP1 | 601859229F1 NIH_MGC_58 Homo sapiens cDNA clone IMAGE:4069735 5', mRNA sequence [BF248073] |
| A_23_P171388 | 0,002191657 | down | -2,1227064 | TMSB4Y | Homo sapiens thymosin beta 4, Y-linked (TMSB4Y), mRNA [NM_004202] |
| A_33_P3249364 | 0,035291966 | down | -1,6209273 | TMTC1 | transmembrane and tetratricopeptide repeat containing 1 [Source:HGNC Symbol;Acc:HGNC:24099] [ENST00000553189] |
| A_32_P2452 | 0,006993477 | up | 2,3308442 | TMTC1 | Homo sapiens transmembrane and tetratricopeptide repeat containing 1 (TMTC1), transcript variant 2, mRNA [NM_175861] |
| A_23_P203957 | 0,00553861 | up | 2,5570455 | TMTC1 | Homo sapiens transmembrane and tetratricopeptide repeat containing 1 (TMTC1), transcript variant 2, mRNA [NM_175861] |
| A_24_P944222 | 0,029569123 | down | -1,6858131 | TMTC3 | Homo sapiens transmembrane and tetratricopeptide repeat containing 3 (TMTC3), mRNA [NM_181783] |
| A_33_P3304688 | 0,007362048 | down | -2,0754933 | TNAP | Homo sapiens NIK-associated protein mRNA, complete cds. [AF463496] |
| A_23_P421423 | 0,027894204 | up | 1,5095543 | TNFAIP2 | Homo sapiens tumor necrosis factor, alpha-induced protein 2 (TNFAIP2), mRNA [NM_006291] |
| A_24_P157926 | 0,002374796 | up | 2,8410902 | TNFAIP3 | Homo sapiens tumor necrosis factor, alpha-induced protein 3 (TNFAIP3), transcript variant 3, mRNA [NM_006290] |
| A_23_P46356 | 0,008898825 | up | 1,8801632 | TNFAIP8L2 | Homo sapiens tumor necrosis factor, alpha-induced protein 8-like 2 (TNFAIP8L2), mRNA [NM_024575] |
| A_23_P255653 | 5,09E-04 | up | 1,7924916 | TNFRSF10A | Homo sapiens tumor necrosis factor receptor superfamily, member 10a (TNFRSF10A), mRNA [NM_003844] |
| A_23_P91764 | 0,001150632 | down | -2,0252533 | TNFRSF13C | Homo sapiens tumor necrosis factor receptor superfamily, member 13C (TNFRSF13C), mRNA [NM_052945] |
| A_33_P3286302 | 0,009436668 | down | -1,5180855 | TNFRSF14 | tumor necrosis factor receptor superfamily, member 14 [Source:HGNC Symbol;Acc:HGNC:11912] [ENST00000463835] |
| A_33_P3248962 | 0,021524772 | up | 2,4108338 | TNFRSF14 | Homo sapiens tumor necrosis factor receptor superfamily, member 14 (TNFRSF14), transcript variant 1, mRNA [NM_003820] |
| A_24_P411121 | 0,037561987 | down | -1,9422615 | TNFRSF18 | Homo sapiens tumor necrosis factor receptor superfamily, member 18 (TNFRSF18), transcript variant 2, mRNA [NM_148901] |
| A_23_P126844 | 0,02995942 | up | 1,5412451 | TNFRSF25 | Homo sapiens tumor necrosis factor receptor superfamily, member 25 (TNFRSF25), transcript variant 1, mRNA [NM_148965] |
| A_33_P3286157 | 0,004830378 | down | -2,1540418 | TNFRSF4 | Homo sapiens tumor necrosis factor receptor superfamily, member 4 (TNFRSF4), mRNA [NM_003327] |
| A_23_P218646 | 0,018698787 | up | 1,9780052 | TNFRSF6B | Homo sapiens tumor necrosis factor receptor superfamily, member 6b, decoy (TNFRSF6B), mRNA [NM_003823] |
| A_23_P115444 | 0,004894888 | up | 1,6306903 | TNFSF18 | Homo sapiens tumor necrosis factor (ligand) superfamily, member 18 (TNFSF18), mRNA [NM_005092] |
| A_23_P386478 | 0,025265278 | up | 1,9329857 | TNIP3 | Homo sapiens TNFAIP3 interacting protein 3 (TNIP3), transcript variant 1, mRNA [NM_024873] |
| A_33_P3365732 | 0,005786107 | down | -1,573599 | TNKS1BP1 | Homo sapiens tankyrase 1 binding protein 1, 182kDa (TNKS1BP1), mRNA [NM_033396] |
| A_23_P125588 | 0,0490513 | up | 1,54665 | TNMD | Homo sapiens tenomodulin (TNMD), mRNA [NM_022144] |
| A_23_P24784 | 0,029587021 | up | 2,1108358 | TNNI2 | Homo sapiens troponin I type 2 (skeletal, fast) (TNNI2), transcript variant 1, mRNA [NM_003282] |
| A_23_P67453 | 0,023669345 | down | -1,8173149 | TNNI3 | Homo sapiens troponin I type 3 (cardiac) (TNNI3), mRNA [NM_000363] |
| A_23_P165504 | 0,039379798 | up | 2,2714815 | TNP1 | Homo sapiens transition protein 1 (during histone to protamine replacement) (TNP1), mRNA [NM_003284] |
| A_24_P260440 | 5,60E-05 | up | 2,180152 | TNPO1 | Homo sapiens transportin 1 (TNPO1), transcript variant 1, mRNA [NM_002270] |
| A_23_P377434 | 0,021940421 | up | 1,530816 | TNRC18 | Homo sapiens trinucleotide repeat containing 18 (TNRC18), mRNA [NM_001080495] |
| A_33_P3570228 | 9,14E-04 | down | -1,8805723 | TNRC18P1 | Homo sapiens TNRC18P1 (TNRC18P1), non-coding RNA [NR_077215] |
| A_23_P349310 | 1,15E-04 | up | 1,7653097 | TNRC6A | Homo sapiens trinucleotide repeat containing 6A (TNRC6A), mRNA [NM_014494] |
| A_33_P3302423 | 0,044048876 | up | 1,766496 | TNRC6C | Homo sapiens trinucleotide repeat containing 6C (TNRC6C), transcript variant 1, mRNA [NM_001142640] |
| A_23_P207850 | 0,007030803 | up | 1,9151545 | TNS4 | Homo sapiens tensin 4 (TNS4), mRNA [NM_032865] |
| A_24_P307135 | 1,96E-04 | down | -2,0157833 | TNXB | Homo sapiens tenascin XB (TNXB), transcript variant XB, mRNA [NM_019105] |
| A_23_P164179 | 3,94E-04 | up | 1,9493697 | TOB1 | Homo sapiens transducer of ERBB2, 1 (TOB1), transcript variant 1, mRNA [NM_005749] |
| A_32_P179676 | 8,87E-04 | up | 1,7608206 | TOB2 | Homo sapiens transducer of ERBB2, 2 (TOB2), mRNA [NM_016272] |
| A_33_P3348011 | 0,003174414 | down | -2,6672611 | TOB2P1 | Homo sapiens transducer of ERBB2, 2 pseudogene 1 (TOB2P1), non-coding RNA [NR_002936] |
| A_33_P337155 | 0,012419402 | down | -1,567834 | TOM1L2 | Homo sapiens target of myb1-like 2 (chicken) (TOM1L2), transcript variant 3, mRNA [NM_001082968] |
| A_23_P141180 | 0,008375721 | up | 1,5733606 | TOM1L2 | Homo sapiens target of myb1-like 2 (chicken) (TOM1L2), transcript variant 3, mRNA [NM_001082968] |
| A_24_P393838 | 0,004678244 | up | 1,5692942 | TOMM20 | Homo sapiens translocase of outer mitochondrial membrane 20 homolog (yeast) (TOMM20), mRNA [NM_014765] |
| A_24_P117942 | 4,98E-06 | down | -4,4766316 | TOMM20L | Homo sapiens translocase of outer mitochondrial membrane 20 homolog (yeast)-like (TOMM20L), mRNA [NM_207377] |
| A_24_P1054 | 0,002303528 | down | -1,7970877 | TONSL | Homo sapiens tonsoku-like, DNA repair protein (TONSL), mRNA [NM_013432] |
| A_24_P374382 | 0,005223304 | up | 2,0220137 | TOP1P2 | Homo sapiens topoisomerase (DNA) I pseudogene 2 (TOP1P2), non-coding RNA [NR_001283] |
| A_33_P3312366 | 0,014641984 | up | 1,5618618 | TOP3B | Homo sapiens topoisomerase (DNA) III beta (TOP3B), transcript variant 1, mRNA [NM_003935] |
| A_33_P3275835 | 0,042101268 | down | -1,5249183 | TOR2A | Homo sapiens torsin family 2, member A (TOR2A), transcript variant 3, mRNA [NM_001134430] |
| A_33_P3247403 | 8,56E-06 | down | -4,1901817 | TOR3A | Homo sapiens torsin family 3, member A (TOR3A), mRNA [NM_022371] |
| A_23_P147109 | 0,004592277 | up | 1,9116986 | TOR4A | Homo sapiens torsin family 4, member A (TOR4A), mRNA [NM_017723] |
| A_23_P12526 | 6,79E-04 | up | 1,8588327 | TP53BP2 | Homo sapiens tumor protein p53 binding protein 2 (TP53BP2), transcript variant 2, mRNA [NM_005426] |

| | | | | | |
|---|---|---|---|---|---|
| A_33_P3286846 | 0,001826862 | down | -3,2133377 | TP53TG3C | Homo sapiens TP53 target 3C (TP53TG3C), transcript variant 1, mRNA [NM_001205259] |
| A_24_P273756 | 0,001521351 | down | -1,6682527 | TP63 | Homo sapiens tumor protein p63 (TP63), transcript variant 1, mRNA [NM_003722] |
| A_24_P413470 | 0,004955743 | down | -2,1662354 | TP73 | Homo sapiens tumor protein p73 (TP73), transcript variant 1, mRNA [NM_005427] |
| A_33_P3296831 | 0,005377436 | down | -1,6416179 | TP73 | Homo sapiens tumor protein p73 (TP73), transcript variant 10, mRNA [NM_001204186] |
| A_23_P131771 | 0,006516386 | up | 1,6746222 | TPD52L2 | Homo sapiens tumor protein D52-like 2 (TPD52L2), transcript variant 1, mRNA [NM_199360] |
| A_23_P150595 | 0,017153978 | up | 1,7113118 | TPH1 | Homo sapiens tryptophan hydroxylase 1 (TPH1), mRNA [NM_004179] |
| A_33_P3346193 | 1,45E-04 | down | -2,5340145 | TPM3 | Homo sapiens tropomyosin 3 (TPM3), transcript variant Tpm3.2, mRNA [NM_001043351] |
| A_33_P3405848 | 0,03717553 | down | -1,7990907 | TPO | Homo sapiens thyroid peroxidase (TPO), transcript variant 2, mRNA [NM_175719] |
| A_33_P3405839 | 0,025762493 | down | -2,0435247 | TPO | Homo sapiens thyroid peroxidase (TPO), transcript variant 5, mRNA [NM_175722] |
| A_33_P3324454 | 0,003751222 | down | -1,8295172 | TPP1 | Homo sapiens tripeptidyl peptidase I (TPP1), mRNA [NM_000391] |
| A_23_P26294 | 4,71E-05 | down | -3,6269126 | TPSG1 | Homo sapiens tryptase gamma 1 (TPSG1), mRNA [NM_012467] |
| A_33_P3247392 | 0,005679519 | up | 1,6498615 | TPTE | Homo sapiens transmembrane phosphatase with tensin homology (TPTE), transcript variant 1, mRNA [NM_199261] |
| A_21_P0011133 | 0,001047304 | down | -2,943495 | TPTE2 | Homo sapiens transmembrane phosphoinositide 3-phosphatase and tensin homolog 2 (TPTE2), transcript variant 3, mRNA [NM_199254] |
| A_24_P118052 | 0,02178078 | up | 2,2166007 | TPTE2 | Homo sapiens transmembrane phosphoinositide 3-phosphatase and tensin homolog 2 (TPTE2), transcript variant 3, mRNA [NM_199254] |
| A_21_P0012240 | 0,017024474 | up | 1,5177863 | TPTEP1 | Homo sapiens transmembrane phosphatase with tensin homology pseudogene 1 (TPTEP1), non-coding RNA [NR_001591] |
| A_21_P0000637 | 4,37E-04 | down | -1,8235024 | TRAF3IP2-AS1 | Homo sapiens TRAF3IP2 antisense RNA 1 (TRAF3IP2-AS1), transcript variant 4, long non-coding RNA [NR_034111] |
| A_23_P75921 | 0,011485987 | up | 1,5575202 | TRAF6 | Homo sapiens TNF receptor-associated factor 6, E3 ubiquitin protein ligase (TRAF6), transcript variant 1, mRNA [NM_145803] |
| A_24_P221960 | 0,004241401 | up | 1,5536567 | TRAM1 | Homo sapiens clone FLB4941 PRO1292 mRNA, complete cds. [AF130055] |
| A_23_P18518 | 0,020910984 | down | -1,7587408 | TRAM1L1 | Homo sapiens translocation associated membrane protein 1-like 1 (TRAM1L1), mRNA [NM_152402] |
| A_21_P0012437 | 0,014673613 | down | -2,0315356 | TRANK1 | Homo sapiens tetratricopeptide repeat and ankyrin repeat containing 1 (TRANK1), mRNA [NM_014831] |
| A_21_P0012439 | 0,030678118 | down | -2,3466284 | TRANK1 | Homo sapiens tetratricopeptide repeat and ankyrin repeat containing 1 (TRANK1), mRNA [NM_014831] |
| A_33_P3312534 | 0,004241541 | up | 1,7213973 | TRAPPC10 | trafficking protein particle complex 10 [Source:HGNC Symbol;Acc:HGNC:11868] [ENST00000459741] |
| A_23_P317683 | 0,001508911 | up | 1,6215832 | TRAPPC10 | Homo sapiens trafficking protein particle complex 10 (TRAPPC10), mRNA [NM_003274] |
| A_24_P390928 | 8,41E-04 | down | -1,7741452 | TRAPPC6A | Homo sapiens trafficking protein particle complex 6A (TRAPPC6A), transcript variant 1, mRNA [NM_024108] |
| A_33_P3248799 | 0,005196196 | up | 1,5957714 | TRDN | Homo sapiens triadin (TRDN), transcript variant 5, mRNA [NM_001256022] |
| A_24_P214858 | 0,023271663 | down | -2,015562 | TREML2 | Homo sapiens triggering receptor expressed on myeloid cells-like 2 (TREML2), mRNA [NM_024807] |
| A_23_P366983 | 0,015014595 | up | 1,6568924 | TRHDE | Homo sapiens thyrotropin-releasing hormone degrading enzyme (TRHDE), mRNA [NM_013381] |
| A_21_P0007724 | 6,75E-04 | down | -3,1847854 | TRHDE-AS1 | Homo sapiens TRHDE antisense RNA 1 (TRHDE-AS1), transcript variant 1, long non-coding RNA [NR_026837] |
| A_23_P322008 | 4,00E-06 | down | -5,4685993 | TRHR | Homo sapiens thyrotropin-releasing hormone receptor (TRHR), mRNA [NM_003301] |
| A_24_P396753 | 0,033382367 | up | 1,723587 | TRIB2 | Homo sapiens tribbles pseudokinase 2 (TRIB2), transcript variant 1, mRNA [NM_021643] |
| A_24_P183664 | 0,045273438 | down | -1,6765625 | TRIL | Homo sapiens TLR4 interactor with leucine-rich repeats (TRIL), mRNA [NM_014817] |
| A_23_P420831 | 0,01003106 | down | -2,133173 | TRIM10 | Homo sapiens tripartite motif containing 10 (TRIM10), transcript variant 2, mRNA [NM_052828] |
| A_33_P3220182 | 0,030000616 | up | 2,1528053 | TRIM15 | Homo sapiens tripartite motif containing 15 (TRIM15), mRNA [NM_033229] |
| A_33_P3416687 | 8,76E-04 | down | -1,6415433 | TRIM16 | tripartite motif containing 16 [Source:HGNC Symbol;Acc:HGNC:17241] [ENST00000582708] |
| A_23_P129903 | 0,043309964 | up | 1,8024665 | TRIM16L | Homo sapiens tripartite motif containing 16-like (TRIM16L), mRNA [NM_001037330] |
| A_24_P208998 | 0,015597605 | down | -2,2018561 | TRIM23 | Homo sapiens tripartite motif containing 23 (TRIM23), transcript variant alpha, mRNA [NM_001656] |
| A_33_P3554318 | 0,015171251 | down | -1,7237769 | TRIM3 | Homo sapiens tripartite motif containing 3 (TRIM3), transcript variant 1, mRNA [NM_006458] |
| A_23_P21230 | 0,014120425 | up | 1,5009241 | TRIM37 | Homo sapiens tripartite motif containing 37 (TRIM37), transcript variant 1, mRNA [NM_015294] |
| A_23_P402778 | 0,047338862 | up | 2,4589398 | TRIM40 | Homo sapiens tripartite motif containing 40 (TRIM40), transcript variant 2, mRNA [NM_138700] |
| A_33_P3474250 | 0,036595564 | down | -1,875239 | TRIM44 | tripartite motif containing 44 [Source:HGNC Symbol;Acc:HGNC:19016] [ENST00000299413] |
| A_21_P0011007 | 0,0492507 | up | 2,0068953 | TRIM49B | Homo sapiens tripartite motif containing 49B (TRIM49B), mRNA [NM_001206626] |
| A_33_P3374970 | 0,007084501 | up | 1,590926 | TRIM49D1 | Homo sapiens tripartite motif containing 49D1 (TRIM49D1), mRNA [NM_001206627] |
| A_23_P150483 | 0,027224904 | up | 1,675102 | TRIM51 | Homo sapiens tripartite motif-containing 51 (TRIM51), mRNA [NM_032681] |
| A_33_P3374971 | 0,003119896 | down | -2,6283765 | TRIM53AP | Homo sapiens tripartite motif containing 53A, pseudogene (TRIM53AP), non-coding RNA [NR_028346] |
| A_33_P3271594 | 0,001579506 | down | -2,9134753 | TRIM54 | Homo sapiens tripartite motif containing 54 (TRIM54), transcript variant 1, mRNA [NM_032546] |
| A_23_P123402 | 0,031334672 | up | 2,1835043 | TRIM55 | Homo sapiens tripartite motif containing 55 (TRIM55), transcript variant 3, mRNA [NM_184086] |
| A_33_P3249414 | 0,004316633 | down | -2,6341598 | TRIM58 | Homo sapiens tripartite motif containing 58 (TRIM58), mRNA [NM_015431] |
| A_33_P3220247 | 3,73E-06 | down | -6,03542 | TRIM6 | Homo sapiens tripartite motif containing 6 (TRIM6), transcript variant 1, mRNA [NM_001003818] |
| A_24_P213120 | 0,01070547 | up | 1,6140071 | TRIM64 | Homo sapiens tripartite motif containing 64 (TRIM64), mRNA [NM_001136486] |
| A_19_P00804979 | 0,001159481 | down | -2,5883381 | TRIM66 | PREDICTED: Homo sapiens tripartite motif containing 66 (TRIM66), transcript variant X1, mRNA [XM_006718397] |
| A_33_P3383994 | 0,001695942 | down | -2,4246817 | TRIM67 | Homo sapiens tripartite motif containing 67 (TRIM67), transcript variant 1, mRNA [NM_001004342] |
| A_23_P48826 | 0,001364796 | down | -1,9031575 | TRIM69 | Homo sapiens tripartite motif containing 69 (TRIM69), transcript variant a, mRNA [NM_182985] |
| A_24_P369656 | 0,013315111 | up | 1,5092518 | TRIM9 | Homo sapiens tripartite motif containing 9 (TRIM9), transcript variant 2, mRNA [NM_052978] |
| A_33_P3415500 | 0,004081265 | up | 1,9143429 | TRIO | trio Rho guanine nucleotide exchange factor [Source:HGNC Symbol;Acc:HGNC:12303] [ENST00000620511] |
| A_33_P3325753 | 0,002644982 | up | 1,8699697 | TRIOBP | Homo sapiens TRIO and F-actin binding protein (TRIOBP), transcript variant 6, mRNA [NM_001039141] |
| A_23_P50349 | 0,006728314 | up | 1,6539199 | TRIP10 | Homo sapiens thyroid hormone receptor interactor 10 (TRIP10), transcript variant 2, mRNA [NM_004240] |
| A_33_P3281363 | 0,001101003 | up | 1,7036574 | TRIP12 | Homo sapiens thyroid hormone receptor interactor 12, mRNA (cDNA clone IMAGE:5742743), with apparent retained intron. [BC037956] |
| A_23_P147984 | 0,001986707 | up | 1,5636333 | TRIP12 | Homo sapiens thyroid hormone receptor interactor 12 (TRIP12), transcript variant 3, mRNA [NM_004238] |
| A_33_P3370310 | 0,003721422 | down | -1,7966144 | TRIQK | Homo sapiens triple QxxK/R motif containing (TRIQK), transcript variant 1, mRNA [NM_001171796] |
| A_33_P3214204 | 0,004737932 | up | 2,3468478 | TRIQK | Homo sapiens clone FLB3344 PRO0845 mRNA, complete cds. [AF130048] |
| A_23_P16683 | 0,017194174 | up | 1,5172882 | TRMT1 | Homo sapiens tRNA methyltransferase 1 homolog (S. cerevisiae) (TRMT1), transcript variant 1, mRNA [NM_017722] |

| | | | | | |
|---|---|---|---|---|---|
| A_23_P400459 | 0,016816825 | up | 1,6044319 | TRMT10B | Homo sapiens tRNA methyltransferase 10 homolog B (S. cerevisiae) (TRMT10B), transcript variant 1, mRNA [NM_144964] |
| A_33_P3225567 | 6,27E-04 | down | -2,1963825 | TRMT2B | Homo sapiens tRNA methyltransferase 2 homolog B (S. cerevisiae) (TRMT2B), transcript variant 1, mRNA [NM_024917] |
| A_33_P3238976 | 8,97E-04 | down | -1,5664033 | TRMT5 | Homo sapiens tRNA methyltransferase 5 (TRMT5), mRNA [NM_020810] |
| A_24_P373126 | 1,05E-04 | down | -2,2577674 | TRMT61A | Homo sapiens tRNA methyltransferase 61A (TRMT61A), mRNA [NM_152307] |
| A_24_P734953 | 9,11E-06 | up | 3,6274762 | TRNP1 | Homo sapiens TMF1-regulated nuclear protein 1 (TRNP1), mRNA [NM_001013642] |
| A_33_P3237874 | 0,037012126 | down | -1,7043241 | TROAP | Homo sapiens trophinin associated protein (TROAP), transcript variant 2, mRNA [NM_001100620] |
| A_33_P3250394 | 0,001867106 | down | -1,7140377 | TRPC2 | transient receptor potential cation channel, subfamily C, member 2, pseudogene [Source:HGNC Symbol;Acc:HGNC:12334] [ENST00000451043] |
| A_23_P105873 | 0,046042353 | up | 1,5677065 | TRPC4 | Homo sapiens transient receptor potential cation channel, subfamily C, member 4 (TRPC4), transcript variant alpha, mRNA [NM_016179] |
| A_33_P3337627 | 0,004666292 | down | -2,5219722 | TRPC6 | Homo sapiens transient receptor potential cation channel, subfamily C, member 6 (TRPC6), mRNA [NM_004621] |
| A_33_P3337632 | 0,001983322 | up | 2,7272894 | TRPC7 | Homo sapiens transient receptor potential cation channel, subfamily C, member 7 (TRPC7), transcript variant 1, mRNA [NM_020389] |
| A_23_P114061 | 0,047582816 | down | -1,5848554 | TRPM3 | Homo sapiens transient receptor potential cation channel, subfamily M, member 3 (TRPM3), transcript variant 9, mRNA [NM_001007471] |
| A_23_P87279 | 0,001039487 | down | -4,6887093 | TRPM5 | Homo sapiens transient receptor potential cation channel, subfamily M, member 5 (TRPM5), mRNA [NM_014555] |
| A_24_P32692 | 0,017477417 | up | 2,5492985 | TRPM6 | Homo sapiens transient receptor potential cation channel, subfamily M, member 6 (TRPM6), transcript variant a, mRNA [NM_017662] |
| A_33_P3317327 | 0,016293019 | up | 2,8130136 | TRPM8 | transient receptor potential cation channel, subfamily M, member 8 [Source:HGNC Symbol;Acc:HGNC:17961] [ENST00000355722] |
| A_23_P94053 | 0,004264571 | up | 1,5132822 | TRRAP | Homo sapiens transformation/transcription domain-associated protein (TRRAP), transcript variant 2, mRNA [NM_003496] |
| A_32_P41553 | 0,011646858 | down | -1,5654534 | TSACC | Homo sapiens TSSK6 activating co-chaperone (TSACC), mRNA [NM_144627] |
| A_33_P3281686 | 0,005496321 | down | -1,6911077 | TSC2 | Homo sapiens tuberous sclerosis 2 (TSC2), transcript variant 4, mRNA [NM_001077183] |
| A_23_P92230 | 0,002020946 | up | 2,097018 | TSC22D2 | Homo sapiens TSC22 domain family, member 2 (TSC22D2), transcript variant 1, mRNA [NM_014779] |
| A_21_P0003042 | 0,01859077 | up | 2,0143878 | TSC22D2 | TSC22 domain family, member 2 [Source:HGNC Symbol;Acc:HGNC:29095] [ENST00000361875] |
| A_23_P12128 | 0,007648613 | down | -1,759504 | TSHB | Homo sapiens thyroid stimulating hormone, beta (TSHB), transcript variant 1, mRNA [NM_000549] |
| A_33_P3247753 | 0,014831217 | down | -1,5778465 | TSIX | Homo sapiens TSIX transcript, XIST antisense RNA (TSIX), antisense RNA [NR_003255] |
| A_33_P3297126 | 0,008393668 | down | -2,0750074 | TSNARE1 | Homo sapiens t-SNARE domain containing 1 (TSNARE1), transcript variant 1, mRNA [NM_145003] |
| A_33_P3291567 | 0,001669911 | down | -2,6131365 | TSNARE1 | Homo sapiens t-SNARE domain containing 1 (TSNARE1), transcript variant 1, mRNA [NM_145003] |
| A_24_P213643 | 3,04E-04 | down | -2,9417005 | TSPAN10 | Homo sapiens tetraspanin 10 (TSPAN10), transcript variant 2, mRNA [NM_031945] |
| A_24_P135483 | 0,004494168 | up | 1,7898215 | TSPAN14 | Homo sapiens tetraspanin 14 (TSPAN14), transcript variant 1, mRNA [NM_030927] |
| A_24_P98613 | 0,001118244 | up | 1,6642308 | TSPAN14 | Homo sapiens tetraspanin 14 (TSPAN14), transcript variant 1, mRNA [NM_030927] |
| A_23_P142070 | 0,042859383 | up | 2,1588383 | TSPAN16 | Homo sapiens tetraspanin 16 (TSPAN16), transcript variant 1, mRNA [NM_012466] |
| A_24_P13083 | 0,008062377 | up | 2,6586716 | TSPAN18 | Homo sapiens tetraspanin 18 (TSPAN18), mRNA [NM_130783] |
| A_23_P316741 | 0,02459322 | up | 1,6268053 | TSPAN4 | Homo sapiens tetraspanin 4 (TSPAN4), transcript variant 1, mRNA [NM_001025237] |
| A_33_P3234222 | 0,001329873 | down | -1,7440497 | TSPO2 | Homo sapiens translocator protein 2 (TSPO2), transcript variant 1, mRNA [NM_001010873] |
| A_33_P3353846 | 0,014104866 | down | -2,2450576 | TSPO2 | Homo sapiens translocator protein 2 (TSPO2), transcript variant 1, mRNA [NM_001010873] |
| A_23_P110882 | 0,005459722 | up | 1,5258228 | TSPYL4 | Homo sapiens TSPY-like 4 (TSPYL4), mRNA [NM_021648] |
| A_23_P91035 | 0,038442124 | up | 1,8312553 | TSPYL6 | Homo sapiens TSPY-like 6 (TSPYL6), mRNA [NM_001003937] |
| A_21_P0014764 | 0,039688963 | up | 2,8415434 | TSRM | Homo sapiens zinc finger domain-related protein TSRM (TSRM) mRNA, partial cds. [AY742823] |
| A_23_P108221 | 0,035734188 | down | -1,537267 | TSSK6 | Homo sapiens testis-specific serine kinase 6 (TSSK6), mRNA [NM_032037] |
| A_33_P3392374 | 0,010447417 | up | 2,6641047 | TTBK1 | Homo sapiens tau tubulin kinase 1 (TTBK1), mRNA [NM_032538] |
| A_23_P103864 | 6,26E-04 | up | 1,7566435 | TTC13 | Homo sapiens tetratricopeptide repeat domain 13 (TTC13), transcript variant 1, mRNA [NM_024525] |
| A_24_P11436 | 0,042666562 | down | -1,5908494 | TTC22 | Homo sapiens tetratricopeptide repeat domain 22 (TTC22), transcript variant 2, mRNA [NM_017904] |
| A_23_P390726 | 0,010854059 | down | -1,716021 | TTC23L | Homo sapiens tetratricopeptide repeat domain 23-like (TTC23L), mRNA [NM_144725] |
| A_33_P3297345 | 0,016989125 | down | -2,236209 | TTC24 | Homo sapiens tetratricopeptide repeat domain 24 (TTC24), mRNA [NM_001105669] |
| A_24_P17453 | 0,002689734 | down | -3,0651875 | TTC30A | Homo sapiens tetratricopeptide repeat domain 30A (TTC30A), mRNA [NM_152275] |
| A_21_P0000586 | 0,002894922 | down | -2,3651474 | TTC3P1 | Homo sapiens tetratricopeptide repeat domain 3 pseudogene 1 (TTC3P1), non-coding RNA [NR_030737] |
| A_24_P337000 | 0,003932091 | up | 1,6846331 | TTC7A | Homo sapiens tetratricopeptide repeat domain 7A (TTC7A), transcript variant 2, mRNA [NM_020458] |
| A_24_P273742 | 0,028610297 | up | 2,2197688 | TTC9 | Homo sapiens tetratricopeptide repeat domain 9 (TTC9), mRNA [NM_015351] |
| A_33_P3293593 | 0,021189176 | down | -2,2098453 | TTC9B | Homo sapiens tetratricopeptide repeat domain 9B (TTC9B), mRNA [NM_152479] |
| A_33_P3324655 | 0,021874966 | up | 1,5428953 | TTI2 | Homo sapiens TELO2 interacting protein 2 (TTI2), transcript variant 2, mRNA [NM_025115] |
| A_24_P165545 | 0,012925072 | up | 2,635343 | TTLL10 | Homo sapiens tubulin tyrosine ligase-like family member 10 (TTLL10), transcript variant 2, mRNA [NM_153254] |
| A_33_P3237250 | 6,41E-04 | down | -2,1258204 | TTLL13P | Homo sapiens tubulin tyrosine ligase-like family member 13, pseudogene (TTLL13P), non-coding RNA [NR_104604] |
| A_23_P7901 | 0,002748334 | up | 1,7775527 | TTLL2 | Homo sapiens tubulin tyrosine ligase-like family member 2 (TTLL2), mRNA [NM_031949] |
| A_33_P3263387 | 0,00381175 | down | -1,5321544 | TTLL4 | Homo sapiens tubulin tyrosine ligase-like family member 4 (TTLL4), mRNA [NM_014640] |
| A_33_P3410060 | 0,003339526 | up | 1,5238185 | TTLL6 | tubulin tyrosine ligase-like family, member 6 [Source:HGNC Symbol;Acc:HGNC:26664] [ENST00000376681] |
| A_21_P0014021 | 4,84E-04 | down | -3,3383427 | TTLL8 | PREDICTED: Homo sapiens tubulin tyrosine ligase-like family, member 8 (TTLL8), mRNA [XM_003403494] |
| A_33_P3398000 | 0,001147532 | down | -1,7442329 | TTN-AS1 | Homo sapiens TTN antisense RNA 1 (TTN-AS1), transcript variant 1, long non-coding RNA [NR_038272] |
| A_24_P323131 | 0,008290255 | up | 1,7419257 | TTTY1 | Homo sapiens testis-specific transcript, Y-linked 1 (non-protein coding) (TTTY1), long non-coding RNA [NR_001538] |
| A_23_P160017 | 0,00703573 | up | 3,260865 | TTTY11 | Homo sapiens testis-specific transcript, Y-linked 11 (non-protein coding) (TTTY11), long non-coding RNA [NR_001548] |
| A_23_P11397 | 0,001886736 | down | -2,099413 | TTTY13 | Homo sapiens testis-specific transcript, Y-linked 13 (non-protein coding) (TTTY13), long non-coding RNA [NR_001537] |
| A_21_P0013906 | 0,005851587 | up | 1,6556604 | TTTY14 | testis-specific transcript, Y-linked 14 (non-protein coding) [Source:HGNC Symbol;Acc:HGNC:18495] [ENST00000447937] |
| A_33_P3349495 | 0,017719604 | down | -2,217408 | TTTY16 | Homo sapiens testis-specific transcript, Y-linked 16 (non-protein coding) (TTTY16), long non-coding RNA [NR_001552] |
| A_24_P186114 | 0,005110478 | up | 1,9961617 | TTTY21 | Homo sapiens testis-specific transcript, Y-linked 21 (non-protein coding) (TTTY21), long non-coding RNA [NR_001535] |
| A_23_P34209 | 0,034288146 | up | 1,9781096 | TTTY5 | Homo sapiens testis-specific transcript, Y-linked 5 (non-protein coding) (TTTY5), long non-coding RNA [NR_001541] |
| A_32_P737629 | 0,007357933 | up | 1,5271895 | TTTY7 | Homo sapiens testis-specific transcript, Y-linked 7 (non-protein coding) (TTTY7), long non-coding RNA [NR_001534] |

| | | | | | |
|---|---|---|---|---|---|
| A_23_P45592 | 0,012408071 | up | 1,7974554 | TTTY8 | Homo sapiens testis-specific transcript, Y-linked 8 (non-protein coding) (TTTY8), long non-coding RNA [NR_001533] |
| A_23_P500892 | 0,03667949 | up | 1,5189434 | TUB | Homo sapiens tubby bipartite transcription factor (TUB), transcript variant 1, mRNA [NM_003320] |
| A_23_P128598 | 0,013723782 | down | -1,6272969 | TUBA3C | Homo sapiens tubulin, alpha 3c (TUBA3C), mRNA [NM_006001] |
| A_23_P56736 | 0,002231384 | down | -2,3615854 | TUBA3D | Homo sapiens tubulin, alpha 3d (TUBA3D), mRNA [NM_080386] |
| A_24_P228550 | 0,014226709 | up | 2,1233552 | TUBB1 | Homo sapiens tubulin, beta 1 class VI (TUBB1), mRNA [NM_030773] |
| A_23_P113656 | 0,007158907 | down | -3,7590454 | TUBB4A | Homo sapiens tubulin, beta 4A class IVa (TUBB4A), transcript variant 3, mRNA [NM_006087] |
| A_23_P26895 | 0,040667567 | up | 1,527745 | TUBD1 | Homo sapiens tubulin, delta 1 (TUBD1), transcript variant 1, mRNA [NM_016261] |
| A_23_P382045 | 0,009456533 | up | 1,6167277 | TULP4 | Homo sapiens tubby like protein 4 (TULP4), transcript variant 1, mRNA [NM_020245] |
| A_33_P3416171 | 0,010030719 | down | -1,9001739 | TUNAR | TCL1 upstream neural differentiation-associated RNA [Source:HGNC Symbol;Acc:HGNC:44088] [ENST00000554321] |
| A_32_P133884 | 0,011149406 | up | 1,5486851 | TUSC1 | Homo sapiens tumor suppressor candidate 1 (TUSC1), mRNA [NM_001004125] |
| A_33_P3354574 | 0,002576389 | down | -1,6873969 | TUSC8 | tumor suppressor candidate 8 (non-protein coding) [Source:HGNC Symbol;Acc:HGNC:49111] [ENST00000607312] |
| A_24_P262321 | 0,006148286 | down | -2,1817665 | TVP23C-CDRT4 | Homo sapiens TVP23C-CDRT4 readthrough (TVP23C-CDRT4), transcript variant 1, mRNA [NM_001204478] |
| A_19_P00322910 | 0,003467158 | down | -3,3201532 | TWIST2 | Homo sapiens twist family bHLH transcription factor 2 (TWIST2), transcript variant 2, mRNA [NM_057179] |
| A_24_P202497 | 0,005055445 | down | -2,0545456 | TWSG1 | Homo sapiens twisted gastrulation BMP signaling modulator 1 (TWSG1), mRNA [NM_020648] |
| A_33_P3270451 | 0,007465159 | down | -1,6159441 | TXNDC5 | Homo sapiens thioredoxin domain containing 5 (endoplasmic reticulum) (TXNDC5), transcript variant 1, mRNA [NM_030810] |
| A_33_P3278275 | 0,004950892 | up | 1,882938 | TXNDC8 | thioredoxin domain containing 8 (spermatozoa) [Source:HGNC Symbol;Acc:HGNC:31454] [ENST00000374511] |
| A_23_P97700 | 0,004463571 | up | 2,597031 | TXNIP | Homo sapiens thioredoxin interacting protein (TXNIP), mRNA [NM_006472] |
| A_33_P3344956 | 0,013020637 | down | -4,954335 | TXNL1 | thioredoxin-like 1 [Source:HGNC Symbol;Acc:HGNC:12436] [ENST00000587807] |
| A_33_P3351120 | 0,014809366 | down | -1,555605 | TXNRD1 | Homo sapiens thioredoxin reductase 1 (TXNRD1), transcript variant 6, mRNA [NM_001261445] |
| A_33_P3374504 | 0,017620482 | down | -1,5618143 | TXNRD2 | Homo sapiens thioredoxin reductase 2 (TXNRD2), transcript variant 2, mRNA [NM_001282512] |
| A_33_P3255949 | 3,06E-05 | down | -3,012885 | TYMP | Homo sapiens thymidine phosphorylase (TYMP), transcript variant 3, mRNA [NM_001113756] |
| A_33_P3409062 | 0,011818289 | down | -1,846496 | TYROBP | Homo sapiens TYRO protein tyrosine kinase binding protein (TYROBP), transcript variant 1, mRNA [NM_003332] |
| A_23_P94403 | 0,006477221 | up | 1,5475277 | TYRP1 | Homo sapiens tyrosinase-related protein 1 (TYRP1), mRNA [NM_000550] |
| A_24_P943106 | 0,003582841 | up | 1,601863 | U2SURP | Homo sapiens U2 snRNP-associated SURP domain containing (U2SURP), mRNA [NM_001080415] |
| A_23_P160460 | 1,91E-04 | up | 1,6751441 | UAP1 | Homo sapiens UDP-N-acetylglucosamine pyrophosphorylase 1 (UAP1), mRNA [NM_003115] |
| A_19_P00319376 | 7,56E-04 | down | -1,8988212 | UBA6-AS1 | Homo sapiens UBA6 antisense RNA 1 (head to head) (UBA6-AS1), long non-coding RNA [NR_015439] |
| A_19_P00321263 | 0,042639904 | down | -1,5306804 | UBA6-AS1 | Homo sapiens UBA6 antisense RNA 1 (head to head) (UBA6-AS1), long non-coding RNA [NR_015439] |
| A_21_P0000659 | 0,026447304 | down | -1,7114892 | UBAC2-AS1 | Homo sapiens UBAC2 antisense RNA 1 (UBAC2-AS1), transcript variant 1, long non-coding RNA [NR_036531] |
| A_23_P123866 | 0,004610841 | up | 1,5552936 | UBAP1 | Homo sapiens ubiquitin associated protein 1 (UBAP1), transcript variant 1, mRNA [NM_016525] |
| A_23_P60472 | 0,001226355 | up | 1,6082758 | UBAP2 | Homo sapiens ubiquitin associated protein 2 (UBAP2), transcript variant 1, mRNA [NM_018449] |
| A_33_P3385909 | 0,017131442 | up | 2,1340492 | UBASH3A | Homo sapiens ubiquitin associated and SH3 domain containing A (UBASH3A), transcript variant 3, mRNA [NM_001243467] |
| A_23_P58293 | 0,007146007 | up | 2,0103192 | UBE2D3 | Homo sapiens ubiquitin-conjugating enzyme E2D 3 (UBE2D3), transcript variant 2, mRNA [NM_181886] |
| A_23_P367071 | 3,09E-07 | down | -6,9892755 | UBE2DNL | Homo sapiens ubiquitin-conjugating enzyme E2D N-terminal like (pseudogene) (UBE2DNL), non-coding RNA [NR_024062] |
| A_19_P00323103 | 0,007473184 | up | 1,5065261 | UBE2E3 | ubiquitin-conjugating enzyme E2E 3 [Source:HGNC Symbol;Acc:HGNC:12479] [ENST00000602837] |
| A_23_P145584 | 0,02731575 | up | 1,7043353 | UBE2H | Homo sapiens ubiquitin-conjugating enzyme E2H (UBE2H), transcript variant 1, mRNA [NM_003344] |
| A_33_P3341234 | 0,001433521 | up | 1,936827 | UBE2H | Homo sapiens ubiquitin-conjugating enzyme E2H (UBE2H), transcript variant 1, mRNA [NM_003344] |
| A_23_P141847 | 0,005339312 | up | 1,5948155 | UBE2M | Homo sapiens ubiquitin-conjugating enzyme E2M (UBE2M), mRNA [NM_003969] |
| A_23_P116829 | 0,007356992 | up | 1,8320801 | UBE2N | Homo sapiens ubiquitin-conjugating enzyme E2N (UBE2N), mRNA [NM_003348] |
| A_33_P3342992 | 0,031191131 | up | 1,6651679 | UBE2Q2P2 | Homo sapiens ubiquitin-conjugating enzyme E2Q family member 2 pseudogene 2 (UBE2Q2P2), non-coding RNA [NR_004847] |
| A_24_P479551 | 0,022137523 | up | 1,6183347 | UBE2QL1 | Homo sapiens ubiquitin-conjugating enzyme E2Q family-like 1 (UBE2QL1), mRNA [NM_001145161] |
| A_33_P3333820 | 0,005024214 | up | 1,6990418 | UBE2R2 | Homo sapiens ubiquitin-conjugating enzyme E2R 2 (UBE2R2), mRNA [NM_017811] |
| A_24_P207150 | 0,001401988 | up | 1,5912294 | UBE3A | Homo sapiens ubiquitin protein ligase E3A (UBE3A), transcript variant 3, mRNA [NM_130839] |
| A_33_P3379581 | 0,017841863 | up | 2,337463 | UBR2 | Homo sapiens ubiquitin protein ligase E3 component n-recognin 2 (UBR2), transcript variant 2, mRNA [NM_001184801] |
| A_32_P159400 | 0,012663381 | down | -1,5615935 | UBR5 | Homo sapiens ubiquitin protein ligase E3 component n-recognin 5 (UBR5), transcript variant 1, mRNA [NM_015902] |
| A_33_P3220565 | 0,014146178 | up | 1,5286041 | UBTD2 | Homo sapiens ubiquitin domain containing 2 (UBTD2), mRNA [NM_152277] |
| A_33_P3292588 | 0,007643314 | up | 2,1416574 | UBTFL1 | Homo sapiens upstream binding transcription factor, RNA polymerase I-like 1 (UBTFL1), mRNA [NM_001143975] |
| A_33_P3303982 | 0,001065236 | up | 2,4934702 | UBTFL1 | Homo sapiens upstream binding transcription factor, RNA polymerase I-like 1 (UBTFL1), mRNA [NM_001143975] |
| A_33_P3394031 | 8,45E-04 | down | -2,6057696 | UBXN11 | Homo sapiens UBX domain protein 11 (UBXN11), transcript variant 2, mRNA [NM_183008] |
| A_33_P3248272 | 0,003770055 | up | 1,5690677 | UBXN8 | Homo sapiens UBX domain protein 8 (UBXN8), transcript variant 1, mRNA [NM_005671] |
| A_24_P292470 | 5,48E-05 | down | -2,8343463 | UCP3 | Homo sapiens uncoupling protein 3 (mitochondrial, proton carrier) (UCP3), transcript variant short, mRNA [NM_022803] |
| A_23_P313389 | 0,023244418 | up | 1,6035728 | UGCG | Homo sapiens UDP-glucose ceramide glucosyltransferase (UGCG), mRNA [NM_003358] |
| A_33_P3396607 | 0,011649787 | down | -1,6122608 | UGDH | Homo sapiens UDP-glucose 6-dehydrogenase (UGDH), transcript variant 1, mRNA [NM_003359] |
| A_24_P44341 | 0,04534711 | down | -1,5584372 | UGGT2 | Homo sapiens UDP-glucose glycoprotein glucosyltransferase 2 (UGGT2), mRNA [NM_020121] |
| A_24_P334378 | 0,009087764 | up | 1,7854121 | UGT2A3 | Homo sapiens UDP glucuronosyltransferase 2 family, polypeptide A3 (UGT2A3), transcript variant 1, mRNA [NM_024743] |
| A_21_P0012591 | 0,003437565 | down | -3,3251035 | UGT2B11 | Homo sapiens UDP glucuronosyltransferase 2 family, polypeptide B11 (UGT2B11), mRNA [NM_001073] |
| A_33_P3424872 | 7,99E-04 | up | 2,9847493 | UGT2B28 | UDP glucuronosyltransferase 2 family, polypeptide B28 [Source:HGNC Symbol;Acc:HGNC:13479] [ENST00000612433] |
| A_23_P122144 | 1,86E-05 | down | -2,5317526 | UGT3A1 | Homo sapiens UDP glycosyltransferase 3 family, polypeptide A1 (UGT3A1), transcript variant 1, mRNA [NM_152404] |
| A_23_P208880 | 0,005669687 | up | 1,7820604 | UHRF1 | Homo sapiens ubiquitin-like with PHD and ring finger domains 1 (UHRF1), transcript variant 2, mRNA [NM_013282] |
| A_24_P303815 | 0,0045696 | up | 1,8270242 | UHRF2 | Homo sapiens ubiquitin-like with PHD and ring finger domains 2, E3 ubiquitin protein ligase (UHRF2), transcript variant 1, mRNA [NM_152896] |
| A_33_P3422802 | 0,024498425 | down | -2,1890485 | ULBP1 | Homo sapiens UL16 binding protein 1 (ULBP1), mRNA [NM_025218] |
| A_23_P134100 | 3,60E-04 | down | -2,2354958 | ULBP3 | Homo sapiens UL16 binding protein 3 (ULBP3), mRNA [NM_024518] |

| | | | | | |
|---|---|---|---|---|---|
| A_24_P932594 | 0,017401619 | up | 2,071302 | ULK2 | Homo sapiens unc-51 like autophagy activating kinase 2 (ULK2), transcript variant 1, mRNA [NM_014683] |
| A_23_P70998 | 0,002805721 | down | -2,147136 | UMAD1 | Homo sapiens UBAP1-MVB12-associated (UMA) domain containing 1 (UMAD1), transcript variant 3, mRNA [NM_001302350] |
| A_33_P3271065 | 0,045505 | down | -2,179497 | UMODL1 | Homo sapiens uromodulin-like 1 (UMODL1), transcript variant 3, mRNA [NM_001199527] |
| A_33_P3355732 | 0,007455724 | down | -2,297192 | UMODL1 | uromodulin-like 1 [Source:HGNC Symbol;Acc:HGNC:12560] [ENST00000491559] |
| A_33_P3274478 | 5,76E-04 | down | -1,7238563 | UNC119B | Homo sapiens unc-119 homolog B (C. elegans) (UNC119B), mRNA [NM_001080533] |
| A_24_P472007 | 0,048384152 | down | -1,9870551 | UNC13A | Homo sapiens unc-13 homolog A (C. elegans) (UNC13A), mRNA [NM_001080421] |
| A_33_P3318926 | 1,26E-04 | down | -2,0021248 | UNC13B | Homo sapiens unc-13 homolog B (C. elegans) (UNC13B), mRNA [NM_006377] |
| A_33_P3379001 | 0,002796473 | down | -2,3254333 | UNC5A | Homo sapiens unc-5 homolog A (C. elegans) (UNC5A), mRNA [NM_133369] |
| A_33_P3374299 | 0,003661971 | up | 1,7649646 | UNC5CL | Homo sapiens unc-5 homolog C (C. elegans)-like (UNC5CL), mRNA [NM_173561] |
| A_23_P350808 | 0,019746076 | up | 1,6176647 | UNC80 | unc-80 homolog (C. elegans) [Source:HGNC Symbol;Acc:HGNC:26582] [ENST00000489023] |
| A_33_P3411388 | 2,14E-05 | up | 3,1173885 | UNCX | Homo sapiens UNC homeobox (UNCX), mRNA [NM_001080461] |
| A_23_P100141 | 0,003387842 | up | 1,6115358 | UNKL | Homo sapiens unkempt family zinc finger-like (UNKL), transcript variant 1, mRNA [NM_001193388] |
| A_24_P194154 | 1,94E-04 | up | 1,6923257 | UPF1 | Homo sapiens UPF1 regulator of nonsense transcripts homolog (yeast) (UPF1), transcript variant 2, mRNA [NM_002911] |
| A_24_P65910 | 0,002141687 | up | 1,9697706 | UQCC1 | Homo sapiens ubiquinol-cytochrome c reductase complex assembly factor 1 (UQCC1), transcript variant 1, mRNA [NM_018244] |
| A_24_P66001 | 0,017661117 | down | -1,7027557 | UQCR10 | Homo sapiens ubiquinol-cytochrome c reductase, complex III subunit X (UQCR10), transcript variant 2, mRNA [NM_001003684] |
| A_23_P118002 | 0,004981527 | up | 1,5581342 | UQCRC2 | Homo sapiens ubiquinol-cytochrome c reductase core protein II (UQCRC2), mRNA [NM_003366] |
| A_23_P74914 | 7,17E-04 | up | 1,9650215 | URB2 | Homo sapiens URB2 ribosome biogenesis 2 homolog (S. cerevisiae) (URB2), mRNA [NM_014777] |
| A_33_P3387666 | 0,018465133 | up | 1,865817 | URM1 | Homo sapiens ubiquitin related modifier 1 (URM1), transcript variant 3, mRNA [NM_001265582] |
| A_24_P366859 | 1,41E-05 | down | -6,901264 | USHBP1 | Homo sapiens Usher syndrome 1C binding protein 1 (USHBP1), transcript variant 1, mRNA [NM_031941] |
| A_33_P3286980 | 0,015309117 | up | 1,9258333 | USP17L1 | Homo sapiens ubiquitin specific peptidase 17-like family member 1 (USP17L1), mRNA [NM_001256873] |
| A_24_P272917 | 0,003182658 | down | -1,7360724 | USP17L25 | Homo sapiens ubiquitin specific peptidase 17-like family member 25 (USP17L25), mRNA [NM_001242326] |
| A_33_P3336242 | 6,47E-04 | down | -2,2228823 | USP17L7 | Homo sapiens ubiquitin specific peptidase 17-like family member 7 (USP17L7), mRNA [NM_001256869] |
| A_33_P3342153 | 0,027630147 | up | 2,6452026 | USP17L8 | Homo sapiens ubiquitin specific peptidase 17-like family member 8 (USP17L8), mRNA [NM_001256872] |
| A_23_P126689 | 0,004632648 | up | 1,5014036 | USP21 | Homo sapiens ubiquitin specific peptidase 21 (USP21), transcript variant 3, mRNA [NM_001014443] |
| A_23_P422540 | 0,001872336 | up | 1,7214036 | USP22 | Homo sapiens ubiquitin specific peptidase 22 (USP22), mRNA [NM_015276] |
| A_24_P58122 | 1,69E-04 | up | 1,9033366 | USP24 | Homo sapiens ubiquitin specific peptidase 24 (USP24), mRNA [NM_015306] |
| A_32_P155826 | 0,001346855 | down | -2,3668566 | USP27X-AS1 | Homo sapiens USP27X antisense RNA 1 (head to head) (USP27X-AS1), long non-coding RNA [NR_026742] |
| A_21_P0000632 | 0,00421454 | down | -2,1433976 | USP3-AS1 | Homo sapiens USP3 antisense RNA 1 (USP3-AS1), long non-coding RNA [NR_034080] |
| A_24_P390583 | 0,03107343 | up | 1,6285976 | USP31 | Homo sapiens ubiquitin specific peptidase 31 (USP31), mRNA [NM_020718] |
| A_24_P126417 | 4,70E-05 | up | 1,8979473 | USP34 | Homo sapiens ubiquitin specific peptidase 34 (USP34), mRNA [NM_014709] |
| A_23_P116602 | 0,016516466 | up | 1,5119859 | USP35 | Homo sapiens ubiquitin specific peptidase 35 (USP35), mRNA [NM_020798] |
| A_24_P310009 | 0,008582053 | up | 1,6179249 | USP43 | Homo sapiens ubiquitin specific peptidase 43 (USP43), transcript variant 1, mRNA [NM_153210] |
| A_33_P3862375 | 0,003922578 | up | 1,6951578 | USP45 | Homo sapiens ubiquitin specific peptidase 45 (USP45), mRNA [NM_001080481] |
| A_24_P377225 | 0,001081587 | up | 1,7870636 | USP46 | Homo sapiens ubiquitin specific peptidase 46 (USP46), transcript variant 1, mRNA [NM_022832] |
| A_33_P3612740 | 0,008332185 | up | 1,5452555 | USP48 | Homo sapiens ubiquitin specific peptidase 48 (USP48), transcript variant 2, mRNA [NM_001032730] |
| A_23_P331770 | 0,002900086 | up | 1,9706173 | USP49 | Homo sapiens ubiquitin specific peptidase 49 (USP49), transcript variant 2, mRNA [NM_018561] |
| A_33_P3301915 | 0,017260931 | up | 2,8908288 | USP51 | Homo sapiens ubiquitin specific peptidase 51 (USP51), mRNA [NM_201286] |
| A_24_P137522 | 0,026947761 | up | 1,6993978 | USP53 | Homo sapiens ubiquitin specific peptidase 53 (USP53), mRNA [NM_019050] |
| A_32_P128701 | 0,007937664 | up | 1,6993574 | USP53 | Homo sapiens ubiquitin specific peptidase 53 (USP53), mRNA [NM_019050] |
| A_23_P129058 | 6,21E-04 | up | 1,650029 | USP8 | Homo sapiens ubiquitin specific peptidase 8 (USP8), transcript variant 1, mRNA [NM_005154] |
| A_33_P3294217 | 0,001803332 | down | -1,842335 | UTF1 | Homo sapiens undifferentiated embryonic cell transcription factor 1 (UTF1), mRNA [NM_003577] |
| A_23_P205216 | 0,009640954 | up | 1,5163854 | UTP14A | Homo sapiens UTP14, U3 small nucleolar ribonucleoprotein, homolog A (yeast) (UTP14A), transcript variant 1, mRNA [NM_006649] |
| A_33_P3234864 | 0,04249241 | down | -1,7918518 | UTRN | Homo sapiens utrophin (UTRN), mRNA [NM_007124] |
| A_33_P3234855 | 0,0287781 | down | -1,8489207 | UTRN | Homo sapiens utrophin (UTRN), mRNA [NM_007124] |
| A_32_P225659 | 0,022013497 | up | 1,682456 | UTS2B | Homo sapiens urotensin 2B (UTS2B), mRNA [NM_198152] |
| A_23_P146885 | 0,016075369 | down | -2,2969403 | UTS2R | Homo sapiens urotensin 2 receptor (UTS2R), mRNA [NM_018949] |
| A_33_P3287113 | 5,30E-04 | down | -1,916318 | UVSSA | UV-stimulated scaffold protein A [Source:HGNC Symbol;Acc:HGNC:29304] [ENST00000503548] |
| A_23_P164341 | 0,006234164 | down | -2,099671 | VAMP2 | Homo sapiens vesicle-associated membrane protein 2 (synaptobrevin 2) (VAMP2), mRNA [NM_014232] |
| A_23_P129695 | 0,016147824 | up | 2,0484507 | VASN | Homo sapiens vasorin (VASN), mRNA [NM_138440] |
| A_33_P3267562 | 0,002788119 | down | -2,4548776 | VAX2 | Homo sapiens ventral anterior homeobox 2 (VAX2), mRNA [NM_012476] |
| A_24_P47182 | 9,72E-04 | up | 1,8780044 | VCL | Homo sapiens vinculin (VCL), transcript variant 1, mRNA [NM_014000] |
| A_23_P70398 | 0,02748651 | down | -1,9479349 | VEGFA | Homo sapiens vascular endothelial growth factor A (VEGFA), transcript variant 6, mRNA [NM_001025370] |
| A_23_P380208 | 0,015180294 | up | 2,3050184 | VEPH1 | Homo sapiens ventricular zone expressed PH domain-containing 1 (VEPH1), transcript variant 1, mRNA [NM_024621] |
| A_23_P132763 | 0,031459384 | up | 2,2169728 | VGLL3 | Homo sapiens vestigial-like family member 3 (VGLL3), mRNA [NM_016206] |
| A_23_P63096 | 0,001576257 | down | -2,0122786 | VHLL | Homo sapiens von Hippel-Lindau tumor suppressor-like (VHLL), mRNA [NM_001004319] |
| A_21_P0014785 | 0,009254421 | down | -1,8108398 | VIM-AS1 | Homo sapiens VIM antisense RNA 1 (VIM-AS1), transcript variant 1, long non-coding RNA [NR_108061] |
| A_23_P145978 | 0,001884015 | down | -2,4131382 | VIPR2 | Homo sapiens vasoactive intestinal peptide receptor 2 (VIPR2), mRNA [NM_003382] |
| A_32_P153071 | 9,84E-04 | down | -3,0681288 | VIPR2 | Homo sapiens vasoactive intestinal peptide receptor 2 (VIPR2), mRNA [NM_003382] |
| A_33_P3284925 | 0,009023916 | down | -1,6119398 | VLDLR | Homo sapiens very low density lipoprotein receptor (VLDLR), transcript variant 2, mRNA [NM_001018056] |
| A_23_P303238 | 0,028645065 | down | -1,5161362 | VN1R5 | Homo sapiens vomeronasal 1 receptor 5 (gene/pseudogene) (VN1R5), mRNA [NM_173858] |
| A_23_P212595 | 0,008827776 | up | 1,5807784 | VPRBP | Homo sapiens Vpr (HIV-1) binding protein (VPRBP), transcript variant 1, mRNA [NM_014703] |

| | | | | | |
|---|---|---|---|---|---|
| A_33_P3401990 | 0,021329291 | down | -1,6188238 | VPREB3 | Homo sapiens pre-B lymphocyte 3 (VPREB3), mRNA [NM_013378] |
| A_23_P206228 | 0,009200818 | up | 1,641588 | VPS13C | Homo sapiens vacuolar protein sorting 13 homolog C (S. cerevisiae) (VPS13C), transcript variant 2A, mRNA [NM_020821] |
| A_23_P23719 | 0,00786595 | up | 1,5701491 | VPS45 | Homo sapiens vacuolar protein sorting 45 homolog (S. cerevisiae) (VPS45), transcript variant 1, mRNA [NM_007259] |
| A_33_P3394352 | 0,004167734 | down | -1,8696234 | VPS53 | vacuolar protein sorting 53 homolog (S. cerevisiae) [Source:HGNC Symbol;Acc:HGNC:25608] [ENST00000541903] |
| A_24_P147398 | 0,016786974 | down | -2,3346572 | VPS53 | Homo sapiens vacuolar protein sorting 53 homolog (S. cerevisiae) (VPS53), transcript variant 2, mRNA [NM_018289] |
| A_24_P279489 | 0,009092806 | up | 1,9188685 | VPS53 | Homo sapiens vacuolar protein sorting 53 homolog (S. cerevisiae) (VPS53), transcript variant 1, mRNA [NM_001128159] |
| A_21_P0007894 | 3,74E-04 | down | -2,6905212 | VSIG10 | V-set and immunoglobulin domain containing 10 [Source:HGNC Symbol;Acc:HGNC:26078] [ENST00000321694] |
| A_21_P0007770 | 0,006290621 | down | -2,3764155 | VSIG10 | V-set and immunoglobulin domain containing 10 [Source:HGNC Symbol;Acc:HGNC:26078] [ENST00000321694] |
| A_33_P3250133 | 0,003801405 | up | 1,8579229 | VSIG10 | Homo sapiens V-set and immunoglobulin domain containing 10 (VSIG10), mRNA [NM_019086] |
| A_23_P72059 | 0,001493154 | up | 2,0955324 | VSIG10 | Homo sapiens V-set and immunoglobulin domain containing 10 (VSIG10), mRNA [NM_019086] |
| A_23_P217269 | 0,003074621 | down | -2,2050657 | VSIG4 | Homo sapiens V-set and immunoglobulin domain containing 4 (VSIG4), transcript variant 1, mRNA [NM_007268] |
| A_23_P209978 | 0,035109587 | down | -1,7741818 | VSNL1 | Homo sapiens visinin-like 1 (VSNL1), mRNA [NM_003385] |
| A_33_P3564487 | 0,025611885 | up | 1,877773 | VSTM2A-OT1 | Homo sapiens VSTM2A overlapping transcript 1 (VSTM2A-OT1), long non-coding RNA [NR_038994] |
| A_23_P79803 | 0,002155611 | up | 4,9266686 | VSTM2L | Homo sapiens V-set and transmembrane domain containing 2 like (VSTM2L), mRNA [NM_080607] |
| A_24_P941038 | 0,004013105 | up | 1,5043513 | VSTM4 | Homo sapiens V-set and transmembrane domain containing 4 (VSTM4), transcript variant 1, mRNA [NM_001031746] |
| A_33_P3233189 | 0,045907967 | up | 2,0857804 | VSTM5 | Homo sapiens V-set and transmembrane domain containing 5 (VSTM5), mRNA [NM_001144871] |
| A_33_P3342295 | 2,34E-04 | down | -2,1988962 | VSX1 | Homo sapiens visual system homeobox 1 (VSX1), transcript variant 2, mRNA [NM_199425] |
| A_33_P3305105 | 7,74E-04 | down | -1,99174 | VWA5A | Homo sapiens von Willebrand factor A domain containing 5A (VWA5A), transcript variant 2, mRNA [NM_198315] |
| A_33_P3405434 | 0,006484724 | down | -2,2374501 | VWA5B1 | von Willebrand factor A domain containing 5B1 [Source:HGNC Symbol;Acc:HGNC:26538] [ENST00000467486] |
| A_23_P42288 | 0,001980035 | down | -2,5435429 | VWA7 | Homo sapiens von Willebrand factor A domain containing 7 (VWA7), mRNA [NM_025258] |
| A_33_P3211238 | 2,76E-05 | down | -2,057008 | VWCE | Homo sapiens von Willebrand factor C and EGF domains (VWCE), mRNA [NM_152718] |
| A_21_P0012280 | 0,005522945 | down | -1,8464898 | VWF | Homo sapiens von Willebrand factor (VWF), mRNA [NM_000552] |
| A_23_P342185 | 7,64E-04 | up | 1,6386156 | WAPAL | Homo sapiens wings apart-like homolog (Drosophila) (WAPAL), mRNA [NM_015045] |
| A_23_P65651 | 0,031222614 | down | -1,8876302 | WARS | Homo sapiens tryptophanyl-tRNA synthetase (WARS), transcript variant 1, mRNA [NM_004184] |
| A_23_P96331 | 3,82E-05 | down | -5,3056407 | WAS | Homo sapiens Wiskott-Aldrich syndrome (WAS), mRNA [NM_000377] |
| A_33_P3243812 | 2,72E-04 | down | -1,7595102 | WASF1 | Homo sapiens WAS protein family, member 1 (WASF1), transcript variant 1, mRNA [NM_003931] |
| A_33_P3337259 | 0,00258696 | down | -1,708374 | WASH5P | Homo sapiens WAS protein family homolog 5 pseudogene (WASH5P), non-coding RNA [NR_033266] |
| A_23_P410128 | 3,70E-04 | up | 2,2463613 | WBP2NL | Homo sapiens WBP2 N-terminal like (WBP2NL), mRNA [NM_152613] |
| A_33_P3304878 | 0,018216906 | down | -2,722079 | WDFY4 | Homo sapiens WDFY family member 4 (WDFY4), mRNA [NM_020945] |
| A_32_P30905 | 0,048917517 | down | -1,7277943 | WDFY4 | Homo sapiens WDFY family member 4 (WDFY4), mRNA [NM_020945] |
| A_23_P90565 | 0,006377874 | down | -1,8791764 | WDPCP | Homo sapiens WD repeat containing planar cell polarity effector (WDPCP), transcript variant 2, mRNA [NM_015910] |
| A_21_P0011923 | 0,004116711 | down | -2,6893632 | WDPCP | WD repeat containing planar cell polarity effector [Source:HGNC Symbol;Acc:HGNC:28027] [ENST00000467687] |
| A_33_P3376404 | 0,011735487 | up | 1,5299648 | WDR13 | Homo sapiens WD repeat domain 13 (WDR13), transcript variant 1, mRNA [NM_017883] |
| A_33_P3375736 | 0,00537794 | up | 1,9962615 | WDR17 | Homo sapiens WD repeat domain 17 (WDR17), transcript variant 1, mRNA [NM_170710] |
| A_24_P184769 | 0,001114525 | up | 1,6736794 | WDR20 | Homo sapiens WD repeat domain 20 (WDR20), transcript variant 1, mRNA [NM_181291] |
| A_23_P84202 | 4,50E-05 | down | -3,5230105 | WDR25 | Homo sapiens WD repeat domain 25 (WDR25), transcript variant 1, mRNA [NM_024515] |
| A_24_P36868 | 0,004081666 | up | 1,7304237 | WDR26 | Homo sapiens WD repeat domain 26 (WDR26), transcript variant 1, mRNA [NM_025160] |
| A_32_P449380 | 0,039086983 | down | -1,5353272 | WDR38 | Homo sapiens WD repeat domain 38 (WDR38), transcript variant 2, mRNA [NM_001045476] |
| A_21_P0011593 | 0,002610374 | up | 1,7681557 | WDR55 | WD repeat domain 55 [Source:HGNC Symbol;Acc:HGNC:25971] [ENST00000358337] |
| A_23_P206856 | 9,04E-04 | up | 1,5938821 | WDR59 | Homo sapiens WD repeat domain 59 (WDR59), mRNA [NM_030581] |
| A_23_P157361 | 0,013334332 | up | 1,7446786 | WDR60 | Homo sapiens WD repeat domain 60 (WDR60), mRNA [NM_018051] |
| A_23_P422732 | 0,03216381 | down | -2,0809667 | WDR63 | Homo sapiens WD repeat domain 63 (WDR63), transcript variant 1, mRNA [NM_145172] |
| A_33_P3388357 | 0,041100085 | down | -1,7873135 | WDR87 | Homo sapiens WD repeat domain 87 (WDR87), transcript variant 1, mRNA [NM_001291088] |
| A_23_P374216 | 0,013736706 | up | 1,5012302 | WFDC11 | Homo sapiens WAP four-disulfide core domain 11 (WFDC11), mRNA [NM_147197] |
| A_24_P250507 | 0,032359894 | up | 1,5343962 | WFDC13 | Homo sapiens WAP four-disulfide core domain 13 (WFDC13), mRNA [NM_172005] |
| A_24_P307827 | 0,002068951 | up | 1,6337994 | WHAMM | Homo sapiens WAS protein homolog associated with actin, golgi membranes and microtubules (WHAMM), mRNA [NM_001080435] |
| A_33_P3423631 | 5,13E-04 | down | -1,8145674 | WHSC1 | Homo sapiens Wolf-Hirschhorn syndrome candidate 1 (WHSC1), transcript variant 8, mRNA [NM_007331] |
| A_23_P389919 | 0,001407249 | up | 1,9050585 | WHSC1 | Homo sapiens Wolf-Hirschhorn syndrome candidate 1 (WHSC1), transcript variant 1, mRNA [NM_133330] |
| A_23_P136325 | 0,0024706 | up | 1,5892291 | WIPF2 | Homo sapiens WAS/WASL interacting protein family, member 2 (WIPF2), mRNA [NM_133264] |
| A_33_P3274299 | 0,046321683 | up | 1,5223137 | WIPF3 | WAS/WASL interacting protein family, member 3 [Source:HGNC Symbol;Acc:HGNC:22004] [ENST00000409123] |
| A_24_P769359 | 5,43E-05 | up | 1,9007136 | WNK1 | Homo sapiens WNK lysine deficient protein kinase 1 (WNK1), transcript variant 4, mRNA [NM_001184985] |
| A_23_P102117 | 0,04318896 | down | -1,8718952 | WNT10A | Homo sapiens wingless-type MMTV integration site family, member 10A (WNT10A), mRNA [NM_025216] |
| A_23_P162322 | 0,001280028 | down | -3,2741096 | WNT10B | Homo sapiens wingless-type MMTV integration site family, member 10B (WNT10B), mRNA [NM_003394] |
| A_33_P3417502 | 8,24E-04 | down | -2,4141884 | WNT3A | Homo sapiens wingless-type MMTV integration site family, member 3A (WNT3A), mRNA [NM_033131] |
| A_33_P3321070 | 0,025135316 | down | -1,9900742 | WNT4 | wingless-type MMTV integration site family, member 4 [Source:HGNC Symbol;Acc:HGNC:12783] [ENST00000415567] |
| A_33_P3384058 | 0,03934742 | up | 2,2758944 | WNT9A | Homo sapiens wingless-type MMTV integration site family, member 9A (WNT9A), mRNA [NM_003395] |
| A_23_P55064 | 0,002793468 | up | 1,9664865 | WRAP53 | Homo sapiens WD repeat containing, antisense to TP53 (WRAP53), transcript variant 1, mRNA [NM_018081] |
| A_24_P192197 | 0,001866956 | up | 1,5407037 | WRNIP1 | Homo sapiens Werner helicase interacting protein 1 (WRNIP1), transcript variant 2, mRNA [NM_130395] |
| A_23_P4353 | 0,04696486 | up | 1,8487909 | WSB1 | Homo sapiens WD repeat and SOCS box containing 1 (WSB1), transcript variant 1, mRNA [NM_015626] |
| A_23_P81392 | 0,00825232 | up | 1,6719011 | WWC1 | Homo sapiens WW and C2 domain containing 1 (WWC1), transcript variant 3, mRNA [NM_015238] |
| A_23_P71972 | 3,68E-04 | down | -2,2378073 | WWOX | Homo sapiens WW domain containing oxidoreductase (WWOX), transcript variant 1, mRNA [NM_016373] |

| | | | | | |
|---|---|---|---|---|---|
| A_23_P142013 | 0,001076677 | down | -1,7188562 | XAB2 | Homo sapiens XPA binding protein 2 (XAB2), mRNA [NM_020196] |
| A_33_P3352304 | 0,006724028 | up | 2,8593209 | XAGE-4 | PREDICTED: Homo sapiens XAGE-4 protein (XAGE-4), misc_RNA [XR_430517] |
| A_24_P315014 | 0,034123838 | up | 2,3662803 | XAGE-4 | PREDICTED: Homo sapiens XAGE-4 protein (XAGE-4), misc_RNA [XR_430517] |
| A_24_P288779 | 0,042096756 | up | 1,8885725 | XAGE5 | Homo sapiens X antigen family, member 5 (XAGE5), mRNA [NM_130775] |
| A_33_P3270863 | 7,03E-04 | up | 2,5602431 | XDH | Homo sapiens xanthine dehydrogenase (XDH), mRNA [NM_000379] |
| A_33_P3304948 | 0,007767445 | up | 1,5400206 | XKR7 | Homo sapiens XK, Kell blood group complex subunit-related family, member 7 (XKR7), mRNA [NM_001011718] |
| A_33_P3225298 | 1,30E-04 | down | -2,3436878 | XKR9 | Homo sapiens XK, Kell blood group complex subunit-related family, member 9 (XKR9), transcript variant 2, mRNA [NM_001287258] |
| A_23_P331253 | 0,001044964 | up | 1,6269107 | XPNPEP1 | Homo sapiens X-prolyl aminopeptidase (aminopeptidase P) 1, soluble (XPNPEP1), transcript variant 1, mRNA [NM_020383] |
| A_24_P333716 | 4,16E-04 | up | 1,6834756 | XPO4 | Homo sapiens exportin 4 (XPO4), mRNA [NM_022459] |
| A_33_P3363515 | 4,29E-04 | down | -1,6408669 | XRCC3 | X-ray repair complementing defective repair in Chinese hamster cells 3 [Source:HGNC Symbol;Acc:HGNC:12830] [ENST00000557439] |
| A_33_P3377549 | 0,004247702 | up | 1,5251852 | XRCC5 | X-ray repair complementing defective repair in Chinese hamster cells 5 (double-strand-break rejoining) [Source:HGNC Symbol;Acc:HGNC:12833] [ENST00000392133] |
| A_23_P319792 | 0,002936522 | up | 1,5863557 | XRN1 | Homo sapiens 5'-3' exoribonuclease 1 (XRN1), transcript variant 1, mRNA [NM_019001] |
| A_33_P3422701 | 0,003154572 | down | -1,5329261 | YEATS2 | Homo sapiens cDNA FLJ33401 fis, clone BRACE2009886. [AK090720] |
| A_32_P178945 | 0,006584567 | up | 2,200248 | YOD1 | Homo sapiens YOD1 deubiquitinase (YOD1), transcript variant 1, mRNA [NM_018566] |
| A_24_P787947 | 0,04532634 | up | 1,5965841 | YPEL2 | Homo sapiens yippee-like 2 (Drosophila) (YPEL2), mRNA [NM_001005404] |
| A_23_P15108 | 9,51E-05 | up | 2,123709 | YPEL3 | Homo sapiens yippee-like 3 (Drosophila) (YPEL3), transcript variant 1, mRNA [NM_031477] |
| A_24_P21770 | 2,33E-05 | down | -4,654021 | YPEL4 | Homo sapiens yippee-like 4 (Drosophila) (YPEL4), mRNA [NM_145008] |
| A_33_P3407105 | 5,68E-04 | down | -1,7463429 | YTHDF1 | Homo sapiens YTH N(6)-methyladenosine RNA binding protein 1 (YTHDF1), mRNA [NM_017798] |
| A_33_P3283237 | 0,021721639 | down | -1,6232886 | YY2 | Homo sapiens YY2 transcription factor (YY2), mRNA [NM_206923] |
| A_23_P316064 | 0,001397128 | up | 1,5935904 | ZACN | Homo sapiens zinc activated ligand-gated ion channel (ZACN), mRNA [NM_180990] |
| A_24_P22746 | 0,002058693 | up | 1,5090033 | ZADH2 | Homo sapiens zinc binding alcohol dehydrogenase domain containing 2 (ZADH2), mRNA [NM_175907] |
| A_23_P366394 | 0,02046339 | up | 1,6083752 | ZAK | Homo sapiens sterile alpha motif and leucine zipper containing kinase AZK (ZAK), transcript variant 1, mRNA [NM_016653] |
| A_23_P82286 | 1,64E-04 | down | -2,5443673 | ZAN | Homo sapiens zonadhesin (gene/pseudogene) (ZAN), transcript variant 6, mRNA [NM_173059] |
| A_33_P3313476 | 6,19E-04 | down | -3,6854281 | ZAR1L | Homo sapiens zygote arrest 1-like (ZAR1L), mRNA [NM_001136571] |
| A_23_P217507 | 0,031632617 | up | 1,5407128 | ZBED1 | Homo sapiens zinc finger, BED-type containing 1 (ZBED1), transcript variant 2, mRNA [NM_004729] |
| A_23_P93032 | 0,04962405 | down | -2,3807964 | ZBED3 | Homo sapiens zinc finger, BED-type containing 3 (ZBED3), mRNA [NM_032367] |
| A_23_P155147 | 2,10E-05 | up | 2,1566644 | ZBED4 | Homo sapiens zinc finger, BED-type containing 4 (ZBED4), mRNA [NM_014838] |
| A_33_P3357609 | 0,007723198 | up | 1,6784288 | ZBP1 | Homo sapiens Z-DNA binding protein 1 (ZBP1), transcript variant 4, mRNA [NM_001160419] |
| A_23_P40866 | 0,001713608 | up | 1,8603411 | ZBTB20 | Homo sapiens zinc finger and BTB domain containing 20 (ZBTB20), transcript variant 2, mRNA [NM_015642] |
| A_21_P0000867 | 0,020338764 | down | -2,1337094 | ZBTB20-AS1 | Homo sapiens ZBTB20 antisense RNA 1 (ZBTB20-AS1), long non-coding RNA [NR_038993] |
| A_33_P3295415 | 0,002001245 | down | -1,8396639 | ZBTB3 | Homo sapiens zinc finger and BTB domain containing 3 (ZBTB3), mRNA [NM_024784] |
| A_33_P3289167 | 0,018526398 | down | -1,6228967 | ZBTB32 | Homo sapiens zinc finger and BTB domain containing 32 (ZBTB32), mRNA [NM_014383] |
| A_23_P131024 | 0,031047247 | up | 3,0692859 | ZBTB32 | Homo sapiens zinc finger and BTB domain containing 32 (ZBTB32), mRNA [NM_014383] |
| A_24_P314351 | 4,96E-05 | up | 1,7976699 | ZBTB33 | Homo sapiens zinc finger and BTB domain containing 33 (ZBTB33), transcript variant 2, mRNA [NM_006777] |
| A_32_P194779 | 4,52E-04 | up | 1,9964631 | ZBTB34 | Homo sapiens zinc finger and BTB domain containing 34 (ZBTB34), mRNA [NM_001099270] |
| A_23_P100654 | 1,67E-04 | up | 1,7856913 | ZBTB4 | Homo sapiens zinc finger and BTB domain containing 4 (ZBTB4), transcript variant 1, mRNA [NM_020899] |
| A_33_P3349414 | 0,001235643 | up | 1,7212595 | ZBTB43 | Homo sapiens zinc finger and BTB domain containing 43 (ZBTB43), transcript variant 1, mRNA [NM_014007] |
| A_24_P592012 | 0,016507382 | up | 1,8579785 | ZBTB46 | Homo sapiens zinc finger and BTB domain containing 46 (ZBTB46), mRNA [NM_025224] |
| A_33_P3266674 | 0,027253147 | up | 1,5977225 | ZBTB46 | Homo sapiens zinc finger and BTB domain containing 46 (ZBTB46), mRNA [NM_025224] |
| A_33_P3217307 | 0,047928065 | down | -1,6813438 | ZBTB8OS | Homo sapiens archease-like protein isoform ABAC (ARCH) mRNA, complete cds; alternatively spliced. [AY158005] |
| A_24_P397903 | 0,005106407 | down | -2,2855434 | ZC2HC1A | Homo sapiens zinc finger, C2HC-type containing 1A (ZC2HC1A), mRNA [NM_016010] |
| A_33_P3307820 | 0,001592618 | down | -3,079182 | ZC2HC1B | Homo sapiens zinc finger, C2HC-type containing 1B (ZC2HC1B), mRNA [NM_001013623] |
| A_32_P130265 | 0,002936289 | up | 1,5164685 | ZC3H11A | Homo sapiens zinc finger CCCH-type containing 11A (ZC3H11A), mRNA [NM_014827] |
| A_23_P58967 | 0,001101257 | up | 1,5498068 | ZC3H11A | Homo sapiens zinc finger CCCH-type containing 11A (ZC3H11A), mRNA [NM_014827] |
| A_33_P3214720 | 0,025693426 | up | 1,5713482 | ZC3H12A | Homo sapiens zinc finger CCCH-type containing 12A (ZC3H12A), mRNA [NM_025079] |
| A_33_P3419399 | 1,17E-04 | down | -2,4607363 | ZC3H12D | Homo sapiens zinc finger CCCH-type containing 12D (ZC3H12D), mRNA [NM_207360] |
| A_24_P241330 | 6,58E-04 | up | 1,5652162 | ZC3H14 | Homo sapiens zinc finger CCCH-type containing 14 (ZC3H14), transcript variant 1, mRNA [NM_024824] |
| A_24_P220618 | 0,00206191 | up | 1,522662 | ZC3H4 | Homo sapiens zinc finger CCCH-type containing 4 (ZC3H4), mRNA [NM_015168] |
| A_23_P218807 | 0,022151593 | up | 1,959699 | ZC3H7B | Homo sapiens zinc finger CCCH-type containing 7B (ZC3H7B), mRNA [NM_017590] |
| A_23_P31532 | 4,96E-05 | down | -3,0038102 | ZC3HAV1L | Homo sapiens zinc finger CCCH-type, antiviral 1-like (ZC3HAV1L), mRNA [NM_080660] |
| A_33_P3214432 | 0,009547078 | down | -1,5655495 | ZC3HAV1L | Homo sapiens zinc finger CCCH-type, antiviral 1-like (ZC3HAV1L), mRNA [NM_080660] |
| A_23_P34433 | 0,006382509 | up | 1,7350188 | ZCCHC11 | Homo sapiens zinc finger, CCHC domain containing 11 (ZCCHC11), transcript variant 1, mRNA [NM_001009881] |
| A_32_P134290 | 8,95E-05 | up | 1,9441653 | ZCCHC2 | Homo sapiens zinc finger, CCHC domain containing 2 (ZCCHC2), transcript variant 1, mRNA [NM_017742] |
| A_33_P3210228 | 2,33E-04 | up | 3,362748 | ZCCHC6 | zinc finger, CCHC domain containing 6 [Source:HGNC Symbol;Acc:HGNC:25817] [ENST00000375947] |
| A_21_P0013817 | 0,002456426 | down | -1,7210193 | ZCRB1 | Homo sapiens zinc finger CCHC-type and RNA binding motif 1 (ZCRB1), mRNA [NM_033114] |
| A_33_P3404531 | 0,014665444 | down | -1,7070621 | ZCWPW1 | Homo sapiens zinc finger, CW type with PWWP domain 1 (ZCWPW1), transcript variant 2, mRNA [NM_001258008] |
| A_21_P0012878 | 0,018345289 | up | 2,0727983 | ZDHHC11 | Homo sapiens zinc finger, DHHC-type containing 11 (ZDHHC11), mRNA [NM_024786] |
| A_23_P319633 | 0,004432304 | up | 1,5475016 | ZDHHC15 | Homo sapiens zinc finger, DHHC-type containing 15 (ZDHHC15), transcript variant 1, mRNA [NM_144969] |
| A_23_P314789 | 5,16E-04 | down | -3,3098562 | ZDHHC19 | Homo sapiens zinc finger, DHHC-type containing 19 (ZDHHC19), mRNA [NM_001039617] |
| A_23_P355447 | 0,012964112 | up | 1,6627517 | ZDHHC22 | Homo sapiens zinc finger, DHHC-type containing 22 (ZDHHC22), mRNA [NM_174976] |
| A_23_P161183 | 5,27E-04 | up | 1,8322465 | ZDHHC6 | Homo sapiens zinc finger, DHHC-type containing 6 (ZDHHC6), transcript variant 1, mRNA [NM_022494] |

| | | | | | |
|---|---|---|---|---|---|
| A_23_P148057 | 7,73E-04 | up | 1,6142142 | ZDHHC9 | Homo sapiens zinc finger, DHHC-type containing 9 (ZDHHC9), transcript variant 1, mRNA [NM_016032] |
| A_33_P3301782 | 0,002815629 | up | 1,956444 | ZEB1-AS1 | Homo sapiens ZEB1 antisense RNA 1 (ZEB1-AS1), long non-coding RNA [NR_024284] |
| A_23_P1170 | 0,013115049 | down | -1,8267266 | ZFAND4 | Homo sapiens zinc finger, AN1-type domain 4 (ZFAND4), transcript variant 1, mRNA [NM_174890] |
| A_33_P3301316 | 0,017113252 | down | -1,7618134 | ZFAND4 | Homo sapiens zinc finger, AN1-type domain 4 (ZFAND4), transcript variant 3, mRNA [NM_001282905] |
| A_23_P150950 | 2,95E-04 | up | 1,8891126 | ZFC3H1 | Homo sapiens zinc finger, C3H1-type containing (ZFC3H1), mRNA [NM_144982] |
| A_23_P3856 | 0,005283977 | up | 1,6469296 | ZFP1 | Homo sapiens ZFP1 zinc finger protein (ZFP1), mRNA [NM_153688] |
| A_24_P151106 | 0,040223103 | up | 2,0224743 | ZFP28 | Homo sapiens ZFP28 zinc finger protein (ZFP28), mRNA [NM_020828] |
| A_23_P39237 | 0,012622566 | up | 2,032421 | ZFP36 | Homo sapiens ZFP36 ring finger protein (ZFP36), mRNA [NM_003407] |
| A_23_P101960 | 1,32E-04 | up | 2,2432005 | ZFP36L2 | Homo sapiens ZFP36 ring finger protein-like 2 (ZFP36L2), mRNA [NM_006887] |
| A_23_P203344 | 0,004655898 | up | 1,5532017 | ZFP91 | Homo sapiens ZFP91 zinc finger protein (ZFP91), transcript variant 1, mRNA [NM_053023] |
| A_33_P3329664 | 1,17E-05 | down | -5,4850106 | ZFP92 | Homo sapiens ZFP92 zinc finger protein (ZFP92), mRNA [NM_001136273] |
| A_24_P32118 | 0,001080765 | up | 1,6431007 | ZFYVE26 | Homo sapiens zinc finger, FYVE domain containing 26 (ZFYVE26), mRNA [NM_015346] |
| A_33_P3278560 | 3,11E-04 | down | -1,6644646 | ZIK1 | Homo sapiens zinc finger protein interacting with K protein 1 (ZIK1), mRNA [NM_001010879] |
| A_23_P78479 | 0,00115439 | down | -2,7361388 | ZIM2 | Homo sapiens zinc finger, imprinted 2 (ZIM2), transcript variant 1, mRNA [NM_015363] |
| A_23_P259663 | 0,047861103 | up | 1,5446947 | ZKSCAN5 | Homo sapiens zinc finger with KRAB and SCAN domains 5 (ZKSCAN5), transcript variant 1, mRNA [NM_014569] |
| A_23_P125748 | 0,002345292 | down | -2,362673 | ZMAT1 | Homo sapiens zinc finger, matrin-type 1 (ZMAT1), transcript variant 2, mRNA [NM_001282400] |
| A_33_P3216133 | 0,029354028 | down | -3,2312822 | ZMAT4 | Homo sapiens zinc finger, matrin-type 4 (ZMAT4), transcript variant 1, mRNA [NM_024645] |
| A_23_P138435 | 0,031255275 | up | 1,6037539 | ZMIZ1 | Homo sapiens zinc finger, MIZ-type containing 1 (ZMIZ1), mRNA [NM_020338] |
| A_24_P29686 | 7,50E-04 | up | 1,8061341 | ZMIZ2 | Homo sapiens zinc finger, MIZ-type containing 2 (ZMIZ2), transcript variant 1, mRNA [NM_031449] |
| A_23_P161091 | 0,006157749 | up | 1,692592 | ZMYM1 | Homo sapiens zinc finger, MYM-type 1 (ZMYM1), transcript variant 5, mRNA [NM_024772] |
| A_23_P99405 | 6,66E-05 | up | 2,0128722 | ZMYM2 | Homo sapiens zinc finger, MYM-type 2 (ZMYM2), transcript variant 1, mRNA [NM_003453] |
| A_24_P237757 | 3,18E-04 | up | 1,902595 | ZMYM2 | zinc finger, MYM-type 2 [Source:HGNC Symbol;Acc:HGNC:12989] [ENST00000382881] |
| A_24_P25326 | 0,00150186 | up | 1,6642265 | ZMYM6 | Homo sapiens zinc finger, MYM-type 6 (ZMYM6), mRNA [NM_007167] |
| A_23_P75083 | 5,56E-04 | down | -1,8222739 | ZMYND11 | Homo sapiens zinc finger, MYND-type containing 11 (ZMYND11), transcript variant 2, mRNA [NM_212479] |
| A_23_P369666 | 1,19E-04 | up | 2,2849078 | ZMYND8 | zinc finger, MYND-type containing 8 [Source:HGNC Symbol;Acc:HGNC:9397] [ENST00000468376] |
| A_23_P45087 | 0,03183994 | up | 1,5495541 | ZNF107 | Homo sapiens zinc finger protein 107 (ZNF107), transcript variant 1, mRNA [NM_016220] |
| A_23_P101811 | 0,001648302 | up | 1,5837448 | ZNF14 | Homo sapiens zinc finger protein 14 (ZNF14), mRNA [NM_021030] |
| A_24_P241792 | 0,009821941 | up | 1,5391015 | ZNF160 | Homo sapiens zinc finger protein 160 (ZNF160), transcript variant 2, mRNA [NM_198893] |
| A_33_P3421363 | 0,00155421 | down | -1,5181446 | ZNF169 | Homo sapiens zinc finger protein 169 (ZNF169), transcript variant 2, mRNA [NM_194320] |
| A_23_P156620 | 0,002759121 | up | 1,554876 | ZNF184 | Homo sapiens zinc finger protein 184 (ZNF184), mRNA [NM_007149] |
| A_23_P415558 | 0,008701056 | up | 1,6448098 | ZNF212 | Homo sapiens zinc finger protein 212 (ZNF212), mRNA [NM_012256] |
| A_23_P210608 | 9,27E-04 | up | 1,6610337 | ZNF217 | Homo sapiens zinc finger protein 217 (ZNF217), mRNA [NM_006526] |
| A_24_P113287 | 0,007657573 | up | 1,6220276 | ZNF229 | Homo sapiens zinc finger protein 229 (ZNF229), transcript variant 1, mRNA [NM_014518] |
| A_23_P153286 | 0,008246581 | up | 1,595224 | ZNF234 | Homo sapiens zinc finger protein 234 (ZNF234), transcript variant 1, mRNA [NM_006630] |
| A_33_P3360912 | 8,01E-04 | down | -2,222186 | ZNF236 | zinc finger protein 236 [Source:HGNC Symbol;Acc:HGNC:13028] [ENST00000543926] |
| A_23_P35343 | 0,040166385 | up | 1,5952938 | ZNF248 | Homo sapiens zinc finger protein 248 (ZNF248), transcript variant 1, mRNA [NM_021045] |
| A_24_P49517 | 0,005189719 | up | 1,6091839 | ZNF251 | Homo sapiens zinc finger protein 251 (ZNF251), mRNA [NM_138367] |
| A_33_P3412349 | 0,008703049 | down | -1,990491 | ZNF268 | Homo sapiens zinc finger protein 268 (ZNF268), transcript variant 3, mRNA [NM_152943] |
| A_33_P3340682 | 0,010270871 | up | 1,5638185 | ZNF276 | Homo sapiens zinc finger protein 276 (ZNF276), transcript variant a, mRNA [NM_001113525] |
| A_24_P282043 | 0,005437287 | up | 2,49193 | ZNF28 | Homo sapiens zinc finger protein 28 (ZNF28), transcript variant 1, mRNA [NM_006969] |
| A_23_P154972 | 0,013814126 | down | -2,225794 | ZNF280A | Homo sapiens zinc finger protein 280A (ZNF280A), mRNA [NM_080740] |
| A_23_P311087 | 0,0020305 | up | 1,7534053 | ZNF281 | Homo sapiens zinc finger protein 281 (ZNF281), transcript variant 1, mRNA [NM_012482] |
| A_23_P311358 | 0,01359433 | up | 1,5087961 | ZNF282 | zinc finger protein 282 [Source:HGNC Symbol;Acc:HGNC:13076] [ENST00000610704] |
| A_33_P3221432 | 0,004107908 | up | 1,9055258 | ZNF284 | Homo sapiens zinc finger protein 284 (ZNF284), mRNA [NM_001037813] |
| A_32_P129968 | 3,43E-04 | up | 2,0094275 | ZNF284 | zinc finger protein 284 [Source:HGNC Symbol;Acc:HGNC:13078] [ENST00000421176] |
| A_33_P3237517 | 0,007953232 | up | 1,7553753 | ZNF292 | zinc finger protein 292 [Source:HGNC Symbol;Acc:HGNC:18410] [ENST00000369578] |
| A_23_P208288 | 0,001252858 | up | 1,5484003 | ZNF304 | Homo sapiens zinc finger protein 304 (ZNF304), transcript variant 2, mRNA [NM_020657] |
| A_33_P3343442 | 9,23E-04 | down | -1,8831791 | ZNF316 | Homo sapiens zinc finger protein 316 (ZNF316), mRNA [NM_001278559] |
| A_33_P3353185 | 0,015497495 | up | 1,6351055 | ZNF316 | Homo sapiens zinc finger protein 316 (ZNF316), mRNA [NM_001278559] |
| A_33_P3292060 | 0,010719216 | up | 1,879416 | ZNF331 | Homo sapiens zinc finger protein 331 (ZNF331), transcript variant 1, mRNA [NM_018555] |
| A_33_P3379512 | 3,14E-05 | down | -2,2249603 | ZNF333 | Homo sapiens zinc finger protein 333 (ZNF333), transcript variant 2, mRNA [NM_001300912] |
| A_33_P3333805 | 0,003363235 | up | 1,8525565 | ZNF337 | Homo sapiens zinc finger protein 337 (ZNF337), transcript variant 1, mRNA [NM_001290261] |
| A_24_P318939 | 0,010021959 | up | 1,7125903 | ZNF337 | Homo sapiens zinc finger protein 337 (ZNF337), transcript variant 2, mRNA [NM_015655] |
| A_21_P0010866 | 0,028965062 | up | 1,5151695 | ZNF37A | zinc finger protein 37A [Source:HGNC Symbol;Acc:HGNC:13102] [ENST00000361085] |
| A_33_P3331031 | 0,014681691 | down | -1,9134878 | ZNF385C | zinc finger protein 385C [Source:HGNC Symbol;Acc:HGNC:33722] [ENST00000453355] |
| A_21_P0011589 | 0,002191905 | down | -2,5513875 | ZNF385C | Homo sapiens zinc finger protein 385C (ZNF385C), mRNA [NM_001242704] |
| A_21_P0011588 | 0,019014973 | up | 2,1412284 | ZNF385C | Homo sapiens zinc finger protein 385C (ZNF385C), mRNA [NM_001242704] |
| A_21_P0013971 | 0,037075818 | down | -1,8197136 | ZNF385D | zinc finger protein 385D [Source:HGNC Symbol;Acc:HGNC:26191] [ENST00000497570] |
| A_24_P393565 | 9,62E-06 | down | -2,911078 | ZNF396 | zinc finger protein 396 [Source:HGNC Symbol;Acc:HGNC:18824] [ENST00000589332] |
| A_32_P151875 | 0,012517402 | up | 1,5198244 | ZNF398 | Homo sapiens zinc finger protein 398 (ZNF398), transcript variant 2, mRNA [NM_020781] |
| A_21_P0009769 | 7,61E-04 | down | -2,394029 | ZNF404 | zinc finger protein 404 [Source:HGNC Symbol;Acc:HGNC:19417] [ENST00000588094] |

| | | | | | |
|---|---|---|---|---|---|
| A_33_P3340385 | 0,006008596 | up | 1,5007534 | ZNF43 | Homo sapiens zinc finger protein 43 (ZNF43), transcript variant 1, mRNA [NM_001256648] |
| A_23_P56298 | 0,001550232 | up | 1,6654925 | ZNF430 | Homo sapiens zinc finger protein 430 (ZNF430), transcript variant 1, mRNA [NM_025189] |
| A_33_P3327842 | 0,012248652 | down | -1,7173463 | ZNF436 | Homo sapiens zinc finger protein 436 (ZNF436), transcript variant 1, mRNA [NM_001077195] |
| A_21_P0006983 | 0,004283102 | up | 1,6180395 | ZNF438 | zinc finger protein 438 [Source:HGNC Symbol;Acc:HGNC:21029] [ENST00000609683] |
| A_33_P3278774 | 0,031069675 | up | 1,5957682 | ZNF44 | zinc finger protein 44 [Source:HGNC Symbol;Acc:HGNC:13110] [ENST00000483826] |
| A_23_P316582 | 0,003018955 | up | 2,192878 | ZNF441 | Homo sapiens zinc finger protein 441 (ZNF441), mRNA [NM_152355] |
| A_24_P218074 | 8,19E-06 | down | -3,6372604 | ZNF467 | Homo sapiens zinc finger protein 467 (ZNF467), mRNA [NM_207336] |
| A_24_P16249 | 0,02886016 | down | -1,9525875 | ZNF479 | Homo sapiens zinc finger protein 479 (ZNF479), mRNA [NM_033273] |
| A_23_P319050 | 0,010992566 | up | 1,8985858 | ZNF493 | Homo sapiens zinc finger protein 493 (ZNF493), transcript variant 2, mRNA [NM_145326] |
| A_24_P376787 | 0,039462384 | down | -1,5595059 | ZNF496 | Homo sapiens zinc finger protein 496 (ZNF496), mRNA [NM_032752] |
| A_33_P3239143 | 9,78E-05 | down | -3,0111766 | ZNF497 | Homo sapiens zinc finger protein 497 (ZNF497), transcript variant 1, mRNA [NM_198458] |
| A_33_P3262495 | 0,003131289 | up | 1,5575842 | ZNF503 | Homo sapiens zinc finger protein 503 (ZNF503), transcript variant 1, mRNA [NM_032772] |
| A_23_P391164 | 0,020304194 | up | 1,5296849 | ZNF512 | Homo sapiens zinc finger protein 512 (ZNF512), transcript variant 1, mRNA [NM_032434] |
| A_24_P88775 | 0,002578368 | up | 1,7500819 | ZNF514 | Homo sapiens zinc finger protein 514 (ZNF514), mRNA [NM_032788] |
| A_21_P0014669 | 0,036021646 | up | 1,7896283 | ZNF518A | zinc finger protein 518A [Source:HGNC Symbol;Acc:HGNC:29009] [ENST00000563195] |
| A_21_P0010834 | 0,005084815 | down | -1,5858958 | ZNF532 | Homo sapiens zinc finger protein 532 (ZNF532), mRNA [NM_018181] |
| A_23_P399146 | 0,007310599 | up | 1,5409237 | ZNF549 | Homo sapiens zinc finger protein 549 (ZNF549), transcript variant 2, mRNA [NM_153263] |
| A_24_P68019 | 0,001318917 | up | 1,8666247 | ZNF551 | Homo sapiens zinc finger protein 551 (ZNF551), transcript variant 1, mRNA [NM_138347] |
| A_23_P39454 | 0,033834122 | up | 2,079873 | ZNF556 | Homo sapiens zinc finger protein 556 (ZNF556), transcript variant 1, mRNA [NM_024967] |
| A_24_P284584 | 0,024975754 | up | 1,6601096 | ZNF559 | Homo sapiens zinc finger protein 559 (ZNF559), transcript variant 2, mRNA [NM_032497] |
| A_23_P306956 | 0,04125525 | down | -1,6411555 | ZNF566 | Homo sapiens zinc finger protein 566 (ZNF566), transcript variant 3, mRNA [NM_032838] |
| A_33_P3345126 | 0,004057325 | down | -1,9128958 | ZNF568 | Homo sapiens zinc finger protein 568 (ZNF568), transcript variant 1, mRNA [NM_198539] |
| A_23_P339079 | 0,001056467 | up | 1,5447397 | ZNF573 | Homo sapiens zinc finger protein 573 (ZNF573), transcript variant 1, mRNA [NM_152360] |
| A_33_P3248035 | 0,04345271 | up | 1,6893604 | ZNF577 | zinc finger protein 577 [Source:HGNC Symbol;Acc:HGNC:28673] [ENST00000588129] |
| A_33_P3345132 | 0,007508155 | up | 2,1124203 | ZNF578 | Homo sapiens zinc finger protein 578 (ZNF578), mRNA [NM_001099694] |
| A_23_P101281 | 0,002846152 | up | 1,9390533 | ZNF587 | Homo sapiens zinc finger protein 587 (ZNF587), transcript variant 1, mRNA [NM_032828] |
| A_21_P0000157 | 0,013018698 | up | 1,5504974 | ZNF587B | Homo sapiens zinc finger protein 587B (ZNF587B), mRNA [NM_001204818] |
| A_33_P3396120 | 0,005869282 | down | -1,6969029 | ZNF594 | Homo sapiens zinc finger protein 594 (ZNF594), mRNA [NM_032530] |
| A_32_P187663 | 0,009927182 | down | -1,6942936 | ZNF596 | Homo sapiens zinc finger protein 596 (ZNF596), transcript variant 1, mRNA [NM_001042416] |
| A_24_P378402 | 0,00285356 | down | -1,9799507 | ZNF597 | Homo sapiens zinc finger protein 597 (ZNF597), mRNA [NM_152457] |
| A_21_P0014663 | 0,008045577 | down | -1,6548572 | ZNF605 | Homo sapiens zinc finger protein 605 (ZNF605), transcript variant 1, mRNA [NM_183238] |
| A_23_P355993 | 2,42E-05 | down | -1,936438 | ZNF614 | Homo sapiens zinc finger protein 614 (ZNF614), mRNA [NM_025040] |
| A_33_P3245784 | 0,039974395 | down | -1,5100654 | ZNF620 | Homo sapiens zinc finger protein 620 (ZNF620), transcript variant 1, mRNA [NM_175888] |
| A_33_P3209030 | 0,0172236 | up | 1,5053298 | ZNF623 | Homo sapiens zinc finger protein 623 (ZNF623), transcript variant 2, mRNA [NM_001082480] |
| A_24_P419177 | 0,00198491 | down | -1,6727167 | ZNF638 | Homo sapiens zinc finger protein 638 (ZNF638), transcript variant 1, mRNA [NM_014497] |
| A_33_P3239790 | 0,02109691 | down | -1,6541026 | ZNF646 | Homo sapiens zinc finger protein 646 (ZNF646), mRNA [NM_014699] |
| A_21_P0014372 | 0,024754934 | down | -1,956557 | ZNF649-AS1 | Homo sapiens ZNF649 antisense RNA 1 (ZNF649-AS1), long non-coding RNA [NR_110733] |
| A_32_P79591 | 0,011475936 | down | -2,3474417 | ZNF665 | Homo sapiens zinc finger protein 665 (ZNF665), mRNA [NM_024733] |
| A_23_P101623 | 0,006858968 | up | 1,6665347 | ZNF667 | Homo sapiens zinc finger protein 667 (ZNF667), transcript variant 1, mRNA [NM_022103] |
| A_23_P86386 | 0,005405157 | up | 1,7329061 | ZNF669 | Homo sapiens zinc finger protein 669 (ZNF669), transcript variant 1, mRNA [NM_024804] |
| A_23_P74981 | 0,001611604 | down | -1,7609196 | ZNF670 | Homo sapiens zinc finger protein 670 (ZNF670), transcript variant 1, mRNA [NM_033213] |
| A_24_P256654 | 0,003882135 | up | 1,8364304 | ZNF674-AS1 | Homo sapiens ZNF674 antisense RNA 1 (head to head) (ZNF674-AS1), long non-coding RNA [NR_015378] |
| A_33_P3231700 | 0,011005186 | up | 1,9938086 | ZNF677 | Homo sapiens mRNA; cDNA DKFZp686M13108 (from clone DKFZp686M13108). [BX648243] |
| A_24_P944588 | 0,006556612 | up | 1,5473949 | ZNF682 | Homo sapiens zinc finger protein 682 (ZNF682), transcript variant 1, mRNA [NM_033196] |
| A_33_P3208970 | 0,041954774 | down | -1,8834369 | ZNF683 | Homo sapiens zinc finger protein 683 (ZNF683), transcript variant 1, mRNA [NM_001114759] |
| A_23_P35316 | 0,007658263 | down | -1,819974 | ZNF695 | Homo sapiens zinc finger protein 695 (ZNF695), transcript variant 1, mRNA [NM_020394] |
| A_23_P130444 | 0,009580362 | up | 1,5431291 | ZNF701 | Homo sapiens zinc finger protein 701 (ZNF701), transcript variant 2, mRNA [NM_018260] |
| A_33_P3316169 | 0,035464313 | up | 1,7066997 | ZNF705E | Homo sapiens zinc finger protein 705E (ZNF705E), mRNA [NM_001278713] |
| A_33_P3275174 | 0,022207825 | down | -1,8868585 | ZNF705G | Homo sapiens zinc finger protein 705G (ZNF705G), mRNA [NM_001164457] |
| A_23_P345674 | 0,003397771 | up | 1,6243421 | ZNF71 | Homo sapiens zinc finger protein 71 (ZNF71), mRNA [NM_021216] |
| A_33_P3408117 | 0,007869578 | down | -2,3195586 | ZNF717 | Homo sapiens zinc finger protein 717 (ZNF717), transcript variant 1, mRNA [NM_001128223] |
| A_21_P0011427 | 0,004032261 | up | 1,8847568 | ZNF720 | PREDICTED: Homo sapiens zinc finger protein 720 (ZNF720), transcript variant X1, mRNA [XM_005255122] |
| A_21_P0011683 | 0,025183277 | up | 2,3293517 | ZNF729 | Homo sapiens zinc finger protein 729 (ZNF729), mRNA [NM_001242680] |
| A_33_P3388067 | 0,048150588 | down | -1,7784135 | ZNF735 | Homo sapiens zinc finger protein 735 (ZNF735), mRNA [NM_001159524] |
| A_23_P28042 | 0,036272682 | down | -1,7023242 | ZNF737 | Homo sapiens zinc finger protein 737 (ZNF737), mRNA [NM_001159293] |
| A_19_P00811828 | 0,006277138 | up | 2,215944 | ZNF737 | Homo sapiens zinc finger protein 737 (ZNF737), mRNA [NM_001159293] |
| A_24_P206344 | 1,16E-04 | down | -1,7848634 | ZNF746 | Homo sapiens zinc finger protein 746 (ZNF746), transcript variant 2, mRNA [NM_152557] |
| A_24_P941487 | 0,004028323 | up | 1,5936552 | ZNF761 | Homo sapiens zinc finger protein 761 (ZNF761), transcript variant 1, mRNA [NM_001008401] |
| A_33_P3289422 | 5,38E-04 | up | 1,7090048 | ZNF765 | Homo sapiens zinc finger protein 765 (ZNF765), mRNA [NM_001040185] |
| A_33_P3267081 | 0,010437447 | down | -1,6109357 | ZNF771 | Homo sapiens zinc finger protein 771 (ZNF771), transcript variant 1, mRNA [NM_016643] |
| A_23_P170581 | 0,008682855 | up | 1,5195721 | ZNF778 | zinc finger protein 778 [Source:HGNC Symbol;Acc:HGNC:26479] [ENST00000620195] |

| | | | | | |
|---|---|---|---|---|---|
| A_23_P365418 | 0,002797879 | up | 1,760648 | ZNF783 | Homo sapiens zinc finger family member 783 (ZNF783), mRNA [NM_001195220] |
| A_23_P157022 | 1,22E-04 | up | 2,0179043 | ZNF786 | Homo sapiens zinc finger protein 786 (ZNF786), mRNA [NM_152411] |
| A_33_P3261927 | 0,017966352 | up | 1,6794516 | ZNF789 | Homo sapiens zinc finger protein 789 (ZNF789), transcript variant 1, mRNA [NM_213603] |
| A_33_P3311770 | 0,003019964 | up | 1,94538 | ZNF789 | Homo sapiens zinc finger protein 789 (ZNF789), transcript variant 1, mRNA [NM_213603] |
| A_21_P0000184 | 0,037386067 | down | -1,6586441 | ZNF790 | Homo sapiens zinc finger protein 790 (ZNF790), transcript variant 4, mRNA [NM_001242802] |
| A_23_P50418 | 5,99E-04 | up | 1,6197184 | ZNF791 | Homo sapiens zinc finger protein 791 (ZNF791), mRNA [NM_153358] |
| A_33_P3399864 | 0,011074869 | down | -2,3927116 | ZNF792 | Homo sapiens zinc finger protein 792 (ZNF792), mRNA [NM_175872] |
| A_23_P256526 | 0,024133723 | down | -2,051116 | ZNF80 | Homo sapiens zinc finger protein 80 (ZNF80), mRNA [NM_007136] |
| A_19_P00812349 | 0,004448604 | up | 1,6791031 | ZNF805 | Homo sapiens zinc finger protein 805 (ZNF805), transcript variant 1, mRNA [NM_001023563] |
| A_19_P00315963 | 0,007330078 | up | 1,7074558 | ZNF805 | Homo sapiens zinc finger protein 805 (ZNF805), transcript variant 1, mRNA [NM_001023563] |
| A_21_P0011739 | 5,39E-05 | down | -1,7915095 | ZNF807 | zinc finger protein 807 [Source:HGNC Symbol;Acc:HGNC:33229] [ENST00000328088] |
| A_33_P3210860 | 0,013527683 | down | -2,073493 | ZNF808 | Homo sapiens zinc finger protein 808 (ZNF808), mRNA [NM_001039886] |
| A_33_P3273298 | 0,001567165 | down | -1,5696726 | ZNF813 | Homo sapiens zinc finger protein 813 (ZNF813), mRNA [NM_001004301] |
| A_33_P3247848 | 0,001648722 | up | 1,6462917 | ZNF814 | Homo sapiens zinc finger protein 814 (ZNF814), mRNA [NM_001144989] |
| A_33_P3220113 | 1,48E-04 | up | 2,0116518 | ZNF816-ZNF321P | Homo sapiens ZNF816-ZNF321P readthrough (ZNF816-ZNF321P), mRNA [NM_001202473] |
| A_23_P105276 | 0,003575847 | up | 1,5394777 | ZNF84 | Homo sapiens zinc finger protein 84 (ZNF84), transcript variant 1, mRNA [NM_003428] |
| A_24_P161144 | 1,15E-04 | down | -2,337286 | ZNF843 | Homo sapiens zinc finger protein 843 (ZNF843), mRNA [NM_001136509] |
| A_21_P0012116 | 0,004108244 | down | -1,6700088 | ZNF850 | Homo sapiens zinc finger protein 850 (ZNF850), transcript variant 1, mRNA [NM_001193552] |
| A_24_P333421 | 0,002214947 | up | 1,6563462 | ZNF862 | Homo sapiens zinc finger protein 862 (ZNF862), mRNA [NM_001099220] |
| A_24_P323967 | 0,006877268 | down | -1,8901371 | ZNF880 | Homo sapiens zinc finger protein 880 (ZNF880), mRNA [NM_001145434] |
| A_21_P0011108 | 0,012927027 | down | -2,0651278 | ZNF891 | Homo sapiens zinc finger protein 891 (ZNF891), mRNA [NM_001277291] |
| A_19_P00801042 | 2,23E-04 | down | -2,4501977 | ZNF90 | zinc finger protein 90 [Source:HGNC Symbol;Acc:HGNC:13165] [ENST00000469078] |
| A_24_P358619 | 0,016196823 | down | -1,5503647 | ZNF99 | Homo sapiens zinc finger protein 99 (ZNF99), mRNA [NM_001080409] |
| A_33_P3243248 | 0,005519266 | down | -3,1779187 | ZNFX1 | zinc finger, NFX1-type containing 1 [Source:HGNC Symbol;Acc:HGNC:29271] [ENST00000469991] |
| A_33_P3304516 | 0,017013807 | up | 1,5092747 | ZNFX1 | Homo sapiens zinc finger, NFX1-type containing 1 (ZNFX1), mRNA [NM_021035] |
| A_33_P3343976 | 0,006749864 | down | -1,952113 | ZNRD1-AS1 | Homo sapiens ZNRD1 antisense RNA 1 (ZNRD1-AS1), long non-coding RNA [NR_026751] |
| A_21_P0000536 | 0,007522089 | down | -1,7395022 | ZNRF2 | Homo sapiens zinc and ring finger 2, E3 ubiquitin protein ligase (ZNRF2), mRNA [NM_147128] |
| A_23_P427502 | 0,008958895 | up | 1,561115 | ZNRF3 | Homo sapiens zinc and ring finger 3 (ZNRF3), transcript variant 1, mRNA [NM_001206998] |
| A_23_P1912 | 0,027355542 | up | 1,8704859 | ZP1 | Homo sapiens zona pellucida glycoprotein 1 (sperm receptor) (ZP1), mRNA [NM_207341] |
| A_33_P3398096 | 0,034812048 | down | -1,7620234 | ZPLD1 | Homo sapiens zona pellucida-like domain containing 1 (ZPLD1), mRNA [NM_175056] |
| A_33_P3261565 | 0,00196021 | up | 1,7436241 | ZRANB1 | Homo sapiens zinc finger, RAN-binding domain containing 1 (ZRANB1), mRNA [NM_017580] |
| A_23_P322395 | 0,003192166 | down | -3,8170924 | ZSCAN1 | Homo sapiens zinc finger and SCAN domain containing 1 (ZSCAN1), mRNA [NM_182572] |
| A_21_P0000526 | 6,58E-04 | down | -2,253439 | ZSCAN18 | Homo sapiens zinc finger and SCAN domain containing 18 (ZSCAN18), transcript variant 5, non-coding RNA [NR_027135] |
| A_23_P23102 | 0,023017745 | up | 1,5080118 | ZSCAN20 | Homo sapiens zinc finger and SCAN domain containing 20 (ZSCAN20), mRNA [NM_145238] |
| A_32_P196115 | 0,029359324 | up | 1,5416459 | ZSCAN25 | Homo sapiens zinc finger and SCAN domain containing 25 (ZSCAN25), mRNA [NM_145115] |
| A_33_P3302741 | 0,002714026 | down | -1,7438822 | ZSCAN30 | zinc finger and SCAN domain containing 30 [Source:HGNC Symbol;Acc:HGNC:33517] [ENST00000589178] |
| A_23_P383118 | 0,007561164 | down | -1,8940718 | ZSWIM5 | Homo sapiens zinc finger, SWIM-type containing 5 (ZSWIM5), mRNA [NM_020883] |
| A_19_P00322299 | 0,039448485 | down | -1,5543164 | ZSWIM6 | Homo sapiens zinc finger, SWIM-type containing 6 (ZSWIM6), mRNA [NM_020928] |
| A_21_P0000778 | 0,01967905 | up | 1,668465 | ZSWIM8-AS1 | Homo sapiens ZSWIM8 antisense RNA 1 (ZSWIM8-AS1), long non-coding RNA [NR_038357] |
| A_23_P63789 | 0,012706506 | up | 1,5533699 | ZWINT | Homo sapiens ZW10 interacting kinetochore protein (ZWINT), transcript variant 2, mRNA [NM_032997] |
| A_24_P282108 | 0,001146444 | up | 1,5750476 | ZZEF1 | Homo sapiens zinc finger, ZZ-type with EF-hand domain 1 (ZZEF1), mRNA [NM_015113] |
| A_33_P3319026 | 4,57E-04 | up | 1,5878677 | ZZZ3 | Homo sapiens zinc finger, ZZ-type containing 3 (ZZZ3), mRNA [NM_015534] |
