## Supplementary material for "Early Reduction of SARS-CoV-2 Replication in Bronchial Epithelium by Kinin B_2_ Receptor Antagonism": Supl. Table 6

**Supplemental Table S6.** DEGs of antiviral epithelial response comparing SARS-CoV-2 versus medium

| ProbeName | p ([SARS-CoV- | Regulatic | FC ([SARS-C | GeneSymbol | Description |
|---|---|---|---|---|---|
| A_32_P9543 | 3,84E-05 | up | 9,134873 | APOBEC3A | Homo sapiens apolipoprotein B mRNA editing enzyme, catalytic polypeptide-like 3A (APOBEC3A), transcript variant 1, mRNA [NM_145699] |
| A_23_P409438 | 3,40E-04 | up | 3,081694 | IFNL2 | Homo sapiens interferon, lambda 2 (IFNL2), mRNA [NM_172138] |
| A_23_P373619 | 0,001634764 | up | 4,4576945 | IFNL3 | Homo sapiens interferon, lambda 3 (IFNL3), mRNA [NM_172139] |
| A_23_P71774 | 3,45E-06 | up | 5,655095 | IFNB1 | Homo sapiens interferon, beta 1, fibroblast (IFNB1), mRNA [NM_002176] |
| A_23_P120931 | 0,045707267 | down | -1,5221866 | APOBEC3C | Homo sapiens apolipoprotein B mRNA editing enzyme, catalytic polypeptide-like 3C (APOBEC3C), mRNA [NM_014508] |
| A_23_P357101 | 0,043491352 | down | -1,5244867 | APOBEC3F | Homo sapiens apolipoprotein B mRNA editing enzyme, catalytic polypeptide-like 3F (APOBEC3F), transcript variant 1, mRNA [NM_145298] |
| A_33_P3345086 | 0,030169737 | up | 1,61869 | IFNA7 | Homo sapiens interferon, alpha 7 (IFNA7), mRNA [NM_021057] |
| A_23_P302060 | 0,01776837 | up | 1,8613136 | IFNE | Homo sapiens interferon, epsilon (IFNE), mRNA [NM_176891] |
| A_23_P337800 | 0,001176448 | up | 2,691211 | IFNL1 | Homo sapiens interferon, lambda 1 (IFNL1), mRNA [NM_172140] |
| A_23_P139786 | 8,80E-04 | up | 3,6718917 | OASL | Homo sapiens 2'-5'-oligoadenylate synthetase-like (OASL), transcript variant 1, mRNA [NM_003733] |
| A_24_P66027 | 0,00480432 | up | 1,6422455 | APOBEC3B | Homo sapiens apolipoprotein B mRNA editing enzyme, catalytic polypeptide-like 3B (APOBEC3B), transcript variant 1, mRNA [NM_004900] |
| A_23_P68155 | 0,019437067 | up | 1,6737528 | IFIH1 | Homo sapiens interferon induced with helicase C domain 1 (IFIH1), mRNA [NM_022168] |
| A_33_P3418170 | 0,020984236 | up | 1,7916337 | DDX58 | Homo sapiens DEAD (Asp-Glu-Ala-Asp) box polypeptide 58 (DDX58), mRNA [NM_014314] |
