## Supplementary material for "Early Reduction of SARS-CoV-2 Replication in Bronchial Epithelium by Kinin B_2_ Receptor Antagonism": Supl. Table 7

**Supplemental Table S7.** DEGs of antiviral epithelial response comparing SARS-CoV-2 versus medium

| ProbeName | p ([SARS-Cov-: | Regulatic | FC ([SARS-C | GeneSymbol | Description |
|---|---|---|---|---|---|
| A_23_P121533 | 2,38E-05 | down | -2,8165832 | SPON2 | Homo sapiens spondin 2, extracellular matrix protein (SPON2), transcript variant 1, mRNA [NM_012445] |
| A_23_P150053 | 0,001929764 | down | -2,1835153 | ACTA2 | Homo sapiens actin, alpha 2, smooth muscle, aorta (ACTA2), transcript variant 2, mRNA [NM_001613] |
| A_23_P165840 | 0,030665506 | down | -2,0057435 | ODC1 | Homo sapiens ornithine decarboxylase 1 (ODC1), transcript variant 1, mRNA [NM_002539] |
| A_32_P108474 | 0,008637361 | down | -1,5648061 | ABCE1 | Homo sapiens ATP-binding cassette, sub-family E (OABP), member 1 (ABCE1), transcript variant 1, mRNA [NM_002940] |
| A_24_P211151 | 0,003254394 | down | -1,92949 | EXOSC5 | Homo sapiens exosome component 5 (EXOSC5), mRNA [NM_020158] |
| A_24_P225961 | 0,003420366 | down | -1,6345493 | DAG1 | Homo sapiens dystroglycan 1 (dystrophin-associated glycoprotein 1) (DAG1), transcript variant 2, mRNA [NM_004393] |
| A_23_P55998 | 0,041465864 | down | -1,8013846 | SLC1A5 | Homo sapiens solute carrier family 1 (neutral amino acid transporter), member 5 (SLC1A5), transcript variant 1, mRNA [NM_005628] |
| A_23_P120931 | 0,045707267 | down | -1,5221866 | APOBEC3C | Homo sapiens apolipoprotein B mRNA editing enzyme, catalytic polypeptide-like 3C (APOBEC3C), mRNA [NM_014508] |
| A_23_P357101 | 0,043491352 | down | -1,5244867 | APOBEC3F | Homo sapiens apolipoprotein B mRNA editing enzyme, catalytic polypeptide-like 3F (APOBEC3F), transcript variant 1, mRNA [NM_145298] |
| A_23_P211207 | 0,005282774 | down | -1,6142308 | ADARB1 | Homo sapiens adenosine deaminase, RNA-specific, B1 (ADARB1), transcript variant 1, mRNA [NM_001112] |
| A_23_P91019 | 0,010147429 | down | -1,5265458 | PRKRA | Homo sapiens protein kinase, interferon-inducible double stranded RNA dependent activator (PRKRA), transcript variant 1, mRNA [NM_003690] |
| A_33_P3317523 | 0,04932188 | down | -1,6132929 | STMN1 | Homo sapiens stathmin 1 (STMN1), transcript variant 1, mRNA [NM_203401] |
| A_32_P9543 | 3,84E-05 | up | 9,134873 | APOBEC3A | Homo sapiens apolipoprotein B mRNA editing enzyme, catalytic polypeptide-like 3A (APOBEC3A), transcript variant 1, mRNA [NM_145699] |
| A_23_P409438 | 3,40E-04 | up | 3,081694 | IFNL2 | Homo sapiens interferon, lambda 2 (IFNL2), mRNA [NM_172138] |
| A_23_P373619 | 0,001634764 | up | 4,4576945 | IFNL3 | Homo sapiens interferon, lambda 3 (IFNL3), mRNA [NM_172139] |
| A_23_P71774 | 3,45E-06 | up | 5,655095 | IFNB1 | Homo sapiens interferon, beta 1, fibroblast (IFNB1), mRNA [NM_002176] |
| A_33_P3351298 | 0,032510124 | up | 1,6358182 | CHRM2 | cholinergic receptor, muscarinic 2 [Source:HGNC Symbol;Acc:HGNC:1951] [ENST00000480591] |
| A_33_P3226420 | 0,021980299 | up | 1,9967993 | RSAD2 | radical S-adenosyl methionine domain containing 2 [Source:HGNC Symbol;Acc:HGNC:30908] [ENST00000474872] |
| A_33_P3345086 | 0,030169737 | up | 1,61869 | IFNA7 | Homo sapiens interferon, alpha 7 (IFNA7), mRNA [NM_021057] |
| A_23_P302060 | 0,01776837 | up | 1,8613136 | IFNE | Homo sapiens interferon, epsilon (IFNE), mRNA [NM_176891] |
| A_33_P3340639 | 0,008225714 | up | 1,7341815 | GPAM | glycerol-3-phosphate acyltransferase, mitochondrial [Source:HGNC Symbol;Acc:HGNC:24865] [ENST00000369425] |
| A_23_P69109 | 0,001251698 | up | 1,6378775 | PLSCR1 | Homo sapiens phospholipid scramblase 1 (PLSCR1), mRNA [NM_021105] |
| A_33_P3418170 | 0,020984236 | up | 1,7916337 | DDX58 | Homo sapiens DEAD (Asp-Glu-Ala-Asp) box polypeptide 58 (DDX58), mRNA [NM_014314] |
| A_23_P68155 | 0,019437067 | up | 1,6737528 | IFIH1 | Homo sapiens interferon induced with helicase C domain 1 (IFIH1), mRNA [NM_022168] |
| A_23_P500433 | 0,012565249 | up | 1,6471444 | CARD9 | Homo sapiens caspase recruitment domain family, member 9 (CARD9), transcript variant 1, mRNA [NM_052813] |
| A_23_P69329 | 0,03443829 | up | 2,0992048 | HYAL1 | Homo sapiens hyaluronoglucosaminidase 1 (HYAL1), transcript variant 8, mRNA [NM_153281] |
| A_24_P66027 | 0,00480432 | up | 1,6422455 | APOBEC3B | Homo sapiens apolipoprotein B mRNA editing enzyme, catalytic polypeptide-like 3B (APOBEC3B), transcript variant 1, mRNA [NM_004900] |
| A_24_P178503 | 0,04903726 | up | 3,058975 | ABCC9 | Homo sapiens ATP-binding cassette, sub-family C (CFTR/MRP), member 9 (ABCC9), transcript variant SUR2A, mRNA [NM_005691] |
| A_23_P76078 | 0,045698304 | up | 1,8592229 | IL23A | Homo sapiens interleukin 23, alpha subunit p19 (IL23A), mRNA [NM_016584] |
| A_23_P61371 | 0,039991934 | up | 1,7341859 | TMEM173 | Homo sapiens transmembrane protein 173 (TMEM173), transcript variant 1, mRNA [NM_198282] |
| A_24_P53215 | 0,049799215 | up | 1,6423753 | UNC13D | Homo sapiens unc-13 homolog D (C. elegans) (UNC13D), mRNA [NM_199242] |
| A_33_P3462960 | 0,015330092 | up | 1,6635326 | DNAJC3 | Homo sapiens DnaJ (Hsp40) homolog, subfamily C, member 3 (DNAJC3), mRNA [NM_006260] |
| A_23_P4662 | 0,009302298 | up | 1,5956694 | BCL3 | Homo sapiens B-cell CLL/lymphoma 3 (BCL3), mRNA [NM_005178] |
| A_24_P126060 | 0,03264237 | up | 1,5573117 | DDX3X | Homo sapiens DEAD (Asp-Glu-Ala-Asp) box helicase 3, X-linked (DDX3X), transcript variant 1, mRNA [NM_001356] |
| A_23_P32404 | 0,003435594 | up | 1,610271 | ISG20 | Homo sapiens interferon stimulated exonuclease gene 20kDa (ISG20), transcript variant 1, mRNA [NM_002201] |
| A_24_P139191 | 6,40E-04 | up | 1,7508626 | ITCH | Homo sapiens itchy E3 ubiquitin protein ligase (ITCH), transcript variant 2, mRNA [NM_031483] |
| A_23_P153320 | 0,007720421 | up | 2,796725 | ICAM1 | Homo sapiens intercellular adhesion molecule 1 (ICAM1), mRNA [NM_000201] |
| A_23_P376488 | 0,002987986 | up | 4,1591763 | TNF | Homo sapiens tumor necrosis factor (TNF), mRNA [NM_000594] |
| A_23_P139786 | 8,80E-04 | up | 3,6718917 | OASL | Homo sapiens 2'-5'-oligoadenylate synthetase-like (OASL), transcript variant 1, mRNA [NM_003733] |
| A_23_P169437 | 0,004435565 | up | 2,1921763 | LCN2 | Homo sapiens lipocalin 2 (LCN2), mRNA [NM_005564] |
| A_23_P162300 | 0,019243024 | up | 2,4734468 | IRAK3 | Homo sapiens interleukin-1 receptor-associated kinase 3 (IRAK3), transcript variant 1, mRNA [NM_007199] |

| | | | | | |
|---|---|---|---|---|---|
| A_23_P337800 | 0,001176448 | up | 2,691211 | IFNL1 | Homo sapiens interferon, lambda 1 (IFNL1), mRNA [NM_172140] |
| A_23_P55632 | 0,003332381 | up | 3,7865973 | SERPINB3 | Homo sapiens serpin peptidase inhibitor, clade B (ovalbumin), member 3 (SERPINB3), mRNA [NM_006919] |
