## Supplementary material for "Early Reduction of SARS-CoV-2 Replication in Bronchial Epithelium by Kinin B_2_ Receptor Antagonism": Supl. Table 8

**Supplemental Table S8**. Gene expression of members of the acute-phase response comparing SARS-CoV-2 versus medium

| ProbeName | p ([SARS-Cov-: | Regulatic | FC ([SARS-C | GeneSymbol | Description |
|---|---|---|---|---|---|
| A_32_P87013 | 2,51E-07 | up | 11,469553 | CXCL8 | Homo sapiens chemokine (C-X-C motif) ligand 8 (CXCL8), mRNA [NM_000584] |
| A_23_P71037 | 0,101391874 | up | 2,4503098 | IL6 | Homo sapiens interleukin 6 (IL6), mRNA [NM_000600] |
| A_23_P133408 | 0,26144698 | up | 2,5167227 | CSF2 | Homo sapiens colony stimulating factor 2 (granulocyte-macrophage) (CSF2), mRNA [NM_000758] |
| A_23_P376488 | 0,002987986 | up | 4,1591763 | TNF | Homo sapiens tumor necrosis factor (TNF), mRNA [NM_000594] |
| A_33_P3316273 | 0,14624505 | up | 1,8288513 | CCL3 | Homo sapiens chemokine (C-C motif) ligand 3 (CCL3), mRNA [NM_002983] |
| A_23_P72096 | 0,07817248 | up | 1,391977 | IL1A | Homo sapiens interleukin 1, alpha (IL1A), mRNA [NM_000575] |
| A_23_P126735 | 0,18970466 | up | 1,4730885 | IL10 | Homo sapiens interleukin 10 (IL10), mRNA [NM_000572] |
| A_23_P89431 | 0,29730952 | up | 1,366798 | CCL2 | Homo sapiens chemokine (C-C motif) ligand 2 (CCL2), mRNA [NM_002982] |
| A_33_P3407013 | 0,38391533 | up | 1,3638464 | IL6 | PREDICTED: Homo sapiens interleukin 6 (interferon, beta 2) (IL6), transcript variant X1, mRNA [XM_005249745] |
| A_24_P303091 | 0,3348003 | up | 1,6963456 | CXCL10 | Homo sapiens chemokine (C-X-C motif) ligand 10 (CXCL10), mRNA [NM_001565] |
| A_23_P152838 | 0,4901788 | up | 1,369833 | CCL5 | Homo sapiens chemokine (C-C motif) ligand 5 (CCL5), transcript variant 1, mRNA [NM_002985] |
| A_33_P3343175 | 0,64367074 | up | 1,3233948 | CXCL10 | Homo sapiens chemokine (C-X-C motif) ligand 10 (CXCL10), mRNA [NM_001565] |
