## Supplementary material for "Early Reduction of SARS-CoV-2 Replication in Bronchial Epithelium by Kinin B_2_ Receptor Antagonism": Supl. Table 9

**Supplemental Table S9.** Gene expression of members of RAS and KKS comparing SARS-CoV-2 versus medium

| ProbeName | p ([SARS-Co\ | Regulation ([S | FC ([SARS-C | GeneSymbol | Description |
|---|---|---|---|---|---|
| A_24_P416645 | 0,15024692 | up | 2,9355443 | KLK13 | Homo sapiens kallikrein-related peptidase 13 (KLK13), mRNA [NM_015596] |
| A_24_P333697 | 0,12090711 | up | 2,249657 | KLK13 | Homo sapiens kallikrein-related peptidase 13 (KLK13), mRNA [NM_015596] |
| A_24_P236935 | 0,00131997 | up | 2,2306533 | KLK6 | Homo sapiens kallikrein-related peptidase 6 (KLK6), transcript variant B, mRNA [NM_001012964] |
| A_33_P3235940 | 2,14E-04 | up | 2,5737338 | KLK6 | Homo sapiens kallikrein-related peptidase 6 (KLK6), transcript variant B, mRNA [NM_001012964] |
| A_23_P101131 | 0,6882029 | up | 1,0837805 | GRP | Homo sapiens gastrin-releasing peptide (GRP), transcript variant 1, mRNA [NM_002091] |
| A_33_P3259708 | 0,73542434 | up | 1,1080414 | CMA1 | Homo sapiens chymase 1, mast cell (CMA1), mRNA [NM_001836] |
| A_23_P4808 | 0,02388008 | up | 1,7446251 | PTGER1 | Homo sapiens prostaglandin E receptor 1 (subtype EP1), 42kDa (PTGER1), mRNA [NM_000955] |
| A_23_P34637 | 0,14624354 | up | 1,4134759 | REN | Homo sapiens renin (REN), mRNA [NM_000537] |
| A_23_P304897 | 0,04153157 | up | 1,4415972 | BDKRB2 | Homo sapiens bradykinin receptor B2 (BDKRB2), mRNA [NM_000623] |
| A_23_P128744 | 0,27854535 | up | 1,213907 | BDKRB1 | Homo sapiens bradykinin receptor B1 (BDKRB1), mRNA [NM_000710] |
| A_23_P16252 | 0,5514491 | up | 1,2131916 | KLK1 | Homo sapiens kallikrein 1 (KLK1), mRNA [NM_002257] |
| A_33_P3245183 | 0,07927634 | up | 1,3502693 | HRH1 | Homo sapiens histamine receptor H1 (HRH1), transcript variant 1, mRNA [NM_001098213] |
| A_33_P3413989 | 0,5090957 | up | 1,2881964 | SERPING1 | Homo sapiens serpin peptidase inhibitor, clade G (C1 inhibitor), member 1 (SERPING1), transcript variant 1, mRNA [NM_000062] |
| A_33_P3296479 | 0,74465394 | down | -1,0623248 | APP | Homo sapiens amyloid beta (A4) precursor protein (APP), transcript variant 10, mRNA [NM_001204303] |
| A_23_P11353 | 0,5416322 | down | -1,0638071 | ATP6AP2 | Homo sapiens ATPase, H+ transporting, lysosomal accessory protein 2 (ATP6AP2), mRNA [NM_005765] |
| A_33_P3290707 | 0,7751802 | up | 1,0692633 | MME | Homo sapiens membrane metallo-endopeptidase (MME), transcript variant 2b, mRNA [NM_007289] |
| A_23_P252981 | 0,16595131 | up | 1,5016031 | ACE2 | Homo sapiens angiotensin I converting enzyme 2 (ACE2), mRNA [NM_021804] |
| A_21_P0000025 | 0,5958493 | up | 1,1578906 | NOS3 | Homo sapiens nitric oxide synthase 3 (endothelial cell) (NOS3), transcript variant 2, mRNA [NM_001160109] |
| A_33_P3305790 | 0,6480025 | up | 1,0676521 | NOS3 | Homo sapiens nitric oxide synthase 3 (endothelial cell) (NOS3), transcript variant 1, mRNA [NM_000603] |
| A_33_P3413987 | 0,43816745 | up | 1,2068092 | SERPING1 | Homo sapiens serpin peptidase inhibitor, clade G (C1 inhibitor), member 1 (SERPING1), transcript variant 1, mRNA [NM_000062] |
| A_33_P3229288 | 0,6186387 | up | 1,0869983 | ACE | Homo sapiens angiotensin I converting enzyme (ACE), transcript variant 1, mRNA [NM_000789] |
| A_33_P3215838 | 0,5450845 | down | -1,1113249 | KLK3 | kallikrein-related peptidase 3 [Source:HGNC Symbol;Acc:HGNC:6364] [ENST00000595151] |
| A_23_P98147 | 0,765945 | up | 1,0595655 | CPN1 | Homo sapiens carboxypeptidase N, polypeptide 1 (CPN1), mRNA [NM_001308] |
| A_23_P252236 | 0,16494894 | up | 1,4089898 | KLKB1 | Homo sapiens kallikrein B, plasma (Fletcher factor) 1 (KLKB1), mRNA [NM_000892] |
| A_23_P139123 | 0,2989573 | down | -1,6766678 | SERPING1 | Homo sapiens serpin peptidase inhibitor, clade G (C1 inhibitor), member 1 (SERPING1), transcript variant 1, mRNA [NM_000062] |
| A_23_P115261 | 0,46579316 | down | -1,2198637 | AGT | Homo sapiens angiotensinogen (serpin peptidase inhibitor, clade A, member 8) (AGT), mRNA [NM_000029] |
| A_23_P148047 | 1 | down | -1,7319175 | PTGER4 | Homo sapiens prostaglandin E receptor 4 (subtype EP4) (PTGER4), mRNA [NM_000958] |
| A_33_P3233871 | 5,25E-04 | down | -1,863506 | F12 | Homo sapiens coagulation factor XII (Hageman factor) (F12), mRNA [NM_000505] |
| A_23_P127964 | 0,10364446 | down | -1,3418982 | PRCP | Homo sapiens prolylcarboxypeptidase (angiotensinase C) (PRCP), transcript variant 2, mRNA [NM_199418] |
| A_23_P37910 | 0,4741085 | up | 1,1568551 | MAPK3 | Homo sapiens mitogen-activated protein kinase 3 (MAPK3), transcript variant 1, mRNA [NM_002746] |
| A_24_P260101 | 1 | down | -1,1484418 | MME | Homo sapiens membrane metallo-endopeptidase (MME), transcript variant 2b, mRNA [NM_007289] |
| A_33_P3370094 | 0,9790554 | up | 1,0063342 | MME | Homo sapiens membrane metallo-endopeptidase (MME), transcript variant 2b, mRNA [NM_007289] |
| A_33_P3508822 | 0,786705 | down | -1,0665829 | APP | Homo sapiens amyloid beta (A4) precursor protein (APP), transcript variant 1, mRNA [NM_000484] |
| A_23_P214821 | 0,2440996 | up | 1,4510032 | EDN1 | Homo sapiens endothelin 1 (EDN1), transcript variant 1, mRNA [NM_001955] |
| A_33_P3258392 | 0,40472084 | up | 1,2779604 | EDN1 | Homo sapiens endothelin 1 (EDN1), transcript variant 1, mRNA [NM_001955] |
| A_33_P3265749 | 0,83497095 | up | 1,0902357 | PTGER3 | Homo sapiens prostaglandin E receptor 3 (subtype EP3) (PTGER3), transcript variant 7, mRNA [NM_198717] |
| A_23_P151778 | 0,27874944 | up | 1,5235522 | CMA1 | Homo sapiens chymase 1, mast cell (CMA1), mRNA [NM_001836] |
| A_33_P3413993 | 0,30919817 | up | 1,228207 | SERPING1 | serpin peptidase inhibitor, clade G (C1 inhibitor), member 1 [Source:HGNC Symbol;Acc:HGNC:1228] [ENST00000405496] |
| A_33_P3265744 | 0,44538012 | up | 1,14883 | PTGER3 | Homo sapiens prostaglandin E receptor 3 (subtype EP3) (PTGER3), transcript variant 5, mRNA [NM_198715] |
| A_23_P151710 | 0,7410629 | down | -1,0890169 | PTGER2 | Homo sapiens prostaglandin E receptor 2 (subtype EP2), 53kDa (PTGER2), mRNA [NM_000956] |
| A_23_P212258 | 0,5087776 | up | 1,2136151 | KNG1 | Homo sapiens kininogen 1 (KNG1), transcript variant 2, mRNA [NM_000893] |

| | | | | | |
|---|---|---|---|---|---|
| A_23_P25720 | 0,46624374 | up | 1,2555822 | SERPINA4 | Homo sapiens serpin peptidase inhibitor, clade A (alpha-1 antiproteinase, antitrypsin), member 4 (SERPINA4), transcript variant 3, mRNA [ |
| A_33_P3376493 | 0,16948764 | up | 1,5416175 | AGTR2 | Homo sapiens angiotensin II receptor, type 2 (AGTR2), mRNA [NM_000686] |
| A_23_P166616 | 0,13536854 | up | 1,3171386 | AGTR1 | Homo sapiens angiotensin II receptor, type 1 (AGTR1), transcript variant 4, mRNA [NM_031850] |
| A_33_P3411338 | 0,03315813 | up | 1,5271437 | ENPEP | Homo sapiens glutamyl aminopeptidase (aminopeptidase A) (ENPEP), mRNA [NM_001977] |
| A_23_P62309 | 0,12392495 | up | 1,378373 | AGTR2 | Homo sapiens angiotensin II receptor, type 2 (AGTR2), mRNA [NM_000686] |
| A_33_P3215843 | 0,20070823 | up | 1,2512552 | KLK3 | Homo sapiens kallikrein-related peptidase 3 (KLK3), transcript variant 3, mRNA [NM_001030047] |
| A_33_P3265739 | 0,368766 | up | 1,2127178 | PTGER3 | Homo sapiens prostaglandin E receptor 3 (subtype EP3) (PTGER3), transcript variant 9, mRNA [NM_198719] |
| A_23_P103328 | 0,15274443 | up | 1,2889884 | PTGER3 | Homo sapiens prostaglandin E receptor 3 (subtype EP3) (PTGER3), transcript variant 4, mRNA [NM_198714] |
| A_24_P38290 | 0,05213348 | up | 1,4935644 | TAC1 | Homo sapiens tachykinin, precursor 1 (TAC1), transcript variant beta, mRNA [NM_003182] |
| A_33_P3251065 | 0,56487787 | up | 1,1569742 | KLK13 | kallikrein-related peptidase 13 [Source:HGNC Symbol;Acc:HGNC:6361] [ENST00000602090] |
