## Supplementary material for "Early Reduction of SARS-CoV-2 Replication in Bronchial Epithelium by Kinin B_2_ Receptor Antagonism": Supl. Table 10

**Supplemental Table S10.** Differentially expressed interleukins comparing SARS-CoV-2 versus medium

| ProbeName | p ([SARS-CoV- | Regulatic | FC ([SARS-C | GeneSymbol | Description |
|---|---|---|---|---|---|
| A_33_P3211608 | 3,97E-02 | up | 2,974933 | IL1RL1 | Homo sapiens interleukin 1 receptor-like 1 (IL1RL1), transcript variant 4, non-coding RNA [NR_104167] |
| A_23_P17053 | 6,39262E-05 | up | 6,132556 | IL36G | Homo sapiens interleukin 36, gamma (IL36G), transcript variant 1, mRNA [NM_019618] |
| A_33_P3339625 | 1,17303E-05 | up | 16,473091 | IL17C | Homo sapiens interleukin 17C (IL17C), mRNA [NM_013278] |
| A_33_P3243230 | 1,76904E-07 | up | 16,647413 | | HSINTLK8M interleukin 8 {Homo sapiens} (exp=-1; wgp=0; cg=0), partial (97%) [THC2544321] |
| A_32_P223777 | 0,00139653 | up | 1,9878159 | IL6ST | Homo sapiens interleukin 6 signal transducer (IL6ST), transcript variant 1, mRNA [NM_002184] |
| A_23_P76078 | 0,045698304 | up | 1,8592229 | IL23A | Homo sapiens interleukin 23, alpha subunit p19 (IL23A), mRNA [NM_016584] |
| A_23_P336554 | 0,00221631 | up | 1,7976834 | IL1RAP | Homo sapiens interleukin 1 receptor accessory protein (IL1RAP), transcript variant 2, mRNA [NM_134470] |
| A_33_P3251876 | 0,03411692 | up | 1,5917065 | IL18R1 | Homo sapiens interleukin 18 receptor 1 (IL18R1), transcript variant 1, mRNA [NM_003855] |
| A_33_P3211666 | 0,026903022 | up | 1,5705488 | IL18R1 | Homo sapiens interleukin 18 receptor 1 (IL18R1), transcript variant 1, mRNA [NM_003855] |
| A_23_P90925 | 0,004416244 | up | 2,695789 | IL36B | Homo sapiens interleukin 36, beta (IL36B), transcript variant 2, mRNA [NM_173178] |
| A_24_P68783 | 0,000239245 | up | 2,834167 | IL36RN | Homo sapiens interleukin 36 receptor antagonist (IL36RN), transcript variant 1, mRNA [NM_012275] |
| A_23_P162300 | 0,019243024 | up | 2,4734468 | IRAK3 | Homo sapiens interleukin-1 receptor-associated kinase 3 (IRAK3), transcript variant 1, mRNA [NM_007199] |
| A_33_P3352970 | 7,91E-05 | up | 2,3658743 | IRAK2 | Homo sapiens interleukin-1 receptor-associated kinase 2 (IRAK2), mRNA [NM_001570] |
| A_33_P3396389 | 1,43E-03 | up | 2,20656 | IL1R1 | Homo sapiens interleukin 1 receptor, type I (IL1R1), transcript variant 1, mRNA [NM_000877] |
| A_33_P3246833 | 0,000372491 | up | 2,0205843 | IL1RN | Homo sapiens interleukin 1 receptor antagonist (IL1RN), transcript variant 4, mRNA [NM_173843] |
| A_33_P3246829 | 3,02E-04 | up | 2,0667813 | IL1RN | Homo sapiens interleukin 1 receptor antagonist (IL1RN), transcript variant 4, mRNA [NM_173843] |
| A_33_P3395947 | 0,049509153 | down | -1,6004809 | IL4R | Homo sapiens interleukin 4 receptor (IL4R), transcript variant 4, mRNA [NM_001257407] |
| A_33_P3243887 | 0,041839596 | down | -1,7166973 | IL11 | Homo sapiens interleukin 11 (IL11), transcript variant 1, mRNA [NM_000641] |
| A_24_P203000 | 0,016171468 | down | -2,173952 | IL2RB | Homo sapiens interleukin 2 receptor, beta (IL2RB), mRNA [NM_000878] |
| A_23_P345692 | 0,001358746 | down | -3,1651986 | IL17D | Homo sapiens interleukin 17D (IL17D), mRNA [NM_138284] |
| A_33_P3219745 | 0,03622351 | down | -1,9250232 | ILF2 | interleukin enhancer binding factor 2 [Source:HGNC Symbol;Acc:HGNC:6037] [ENST00000368681] |
| A_23_P61057 | 0,04433362 | down | -1,8543203 | IL16 | Homo sapiens interleukin 16 (IL16), transcript variant 1, mRNA [NM_004513] |
| A_23_P35092 | 0,011103406 | up | 1,8246565 | IL19 | Homo sapiens interleukin 19 (IL19), transcript variant 1, mRNA [NM_153758] |
| A_23_P46482 | 0,007016526 | down | -2,929152 | IL20 | Homo sapiens interleukin 20 (IL20), mRNA [NM_018724] |
| A_23_P104798 | 0,013057214 | down | -1,5936793 | IL18 | Homo sapiens interleukin 18 (IL18), transcript variant 1, mRNA [NM_001562] |
| A_33_P3288844 | 0,035492603 | down | -1,7655479 | IL6R | Homo sapiens interleukin 6 receptor (IL6R), transcript variant 1, mRNA [NM_000565] |
| A_21_P0000171 | 0,004423897 | down | -1,7600982 | IL6R | Homo sapiens interleukin 6 receptor (IL6R), transcript variant 3, mRNA [NM_001206866] |
| A_23_P32253 | 0,007869842 | down | -1,5080216 | NFIL3 | Homo sapiens nuclear factor, interleukin 3 regulated (NFIL3), transcript variant 3, mRNA [NM_005384] |
| A_33_P3399267 | 0,0328938 | down | -1,7424508 | IL15RA | Homo sapiens interleukin 15 receptor, alpha (IL15RA), transcript variant 2, mRNA [NM_172200] |
