## Supplementary material for "Early Reduction of SARS-CoV-2 Replication in Bronchial Epithelium by Kinin B_2_ Receptor Antagonism": Supl. Table 11

**Supplemental Table S11.** Potential viral entry receptor DEGs comparing SARS-CoV-2 versus medium

| ProbeName | p ([SARS-Co\ | Regulatic | FC ([SARS-( | GeneSymbol | Description |
|---|---|---|---|---|---|
| A_24_P186539 | 0,04956434 | up | 1,6313815 | CD209 | Homo sapiens CD209 molecule (CD209), transcript variant 1, mRNA [NM_021155] |
| A_33_P3374678 | 9,62E-03 | up | 2,0423372 | TMPRSS11D | Homo sapiens transmembrane protease, serine 11D (TMPRSS11D), mRNA [NM_004262] |
| A_33_P3217845 | 4,27E-03 | up | 3,431767 | TMPRSS11A | Homo sapiens transmembrane protease, serine 11A (TMPRSS11A), transcript variant 1, mRNA [NM_182606] |
| A_24_P109101 | 3,59E-02 | up | 3,0592778 | TMPRSS11A | Homo sapiens transmembrane protease, serine 11A (TMPRSS11A), transcript variant 1, mRNA [NM_182606] |
