## Supplementary material for "Early Reduction of SARS-CoV-2 Replication in Bronchial Epithelium by Kinin B_2_ Receptor Antagonism": Supl. Table 12

**Supplemental Table S12.** Toll-like receptor (TLR) DEGs comparing SARS-CoV-2 versus medium

| ProbeName | p ([SARS-CoV- | Regulatic | FC ([SARS-( | GeneS | Description |
|---|---|---|---|---|---|
| A_23_P92499 | 0,022723636 | up | 3,1690073 | TLR2 | Homo sapiens toll-like receptor 2 (TLR2), mRNA [NM_003264] |
| A_23_P10873 | 0,02445174 | up | 1,9784745 | TLR1 | Homo sapiens toll-like receptor 1 (TLR1), mRNA [NM_003263] |
| A_33_P326342 | 0,005945041 | up | 1,5899574 | TLR5 | Homo sapiens toll-like receptor 5 (TLR5), mRNA [NM_003268] |
