## Supplementary material for "Early Reduction of SARS-CoV-2 Replication in Bronchial Epithelium by Kinin B_2_ Receptor Antagonism": Supl. Table 13

**Supplemental Table S13.** Chemokine DEGs comparing SARS-CoV-2 versus medium

| ProbeName | p ([Cov2] Vs [M | Regulatic | FC ([Cov2] V | GeneSym | Description |
|---|---|---|---|---|---|
| A_23_P121596 | 0,002959288 | up | 2,6544087 | PPBP | Homo sapiens pro-platelet basic protein (chemokine (C-X-C motif) ligand 7) (PPBP), mRNA [NM_002704] |
| A_23_P50269 | 0,018069379 | up | 2,2989175 | CXCL17 | Homo sapiens chemokine (C-X-C motif) ligand 17 (CXCL17), mRNA [NM_198477] |
| A_24_P183150 | 0,00165627 | up | 2,7300987 | CXCL3 | Homo sapiens chemokine (C-X-C motif) ligand 3 (CXCL3), mRNA [NM_002090] |
| A_24_P257416 | 1,25E-04 | up | 2,7464068 | CXCL2 | Homo sapiens chemokine (C-X-C motif) ligand 2 (CXCL2), mRNA [NM_002089] |
| A_23_P315364 | 0,006391583 | up | 2,3417945 | CXCL2 | Homo sapiens chemokine (C-X-C motif) ligand 2 (CXCL2), mRNA [NM_002089] |
| A_23_P69012 | 0,012230188 | up | 2,2441964 | CCR8 | Homo sapiens chemokine (C-C motif) receptor 8 (CCR8), mRNA [NM_005201] |
| A_23_P67932 | 0,03768801 | up | 1,577224 | CXCR1 | Homo sapiens chemokine (C-X-C motif) receptor 1 (CXCR1), mRNA [NM_000634] |
| A_23_P211699 | 0,006295188 | up | 2,1925251 | | CCR8_HUMAN (P51685) C-C chemokine receptor type 8 (C-C CKR-8) (CC-CKR-8) (CCR-8) (GPR-CY6) (GPRCY6) (Chemokine receptor-like 1) (CKR-L1) (TER1) (CMKBRL2) (CC-chemokine receptor CHEMR1) (CDw198 antigen), complete [THC2477815] |
| A_33_P3354607 | 0,001333996 | up | 2,4903448 | CCL4L2 | Homo sapiens chemokine (C-C motif) ligand 4-like 2 (CCL4L2), transcript variant CCL4L2b2, mRNA [NM_001291470] |
| A_23_P17065 | 2,48E-05 | up | 7,191096 | CCL20 | Homo sapiens chemokine (C-C motif) ligand 20 (CCL20), transcript variant 1, mRNA [NM_004591] |
| A_32_P87013 | 2,51E-07 | up | 11,469553 | CXCL8 | Homo sapiens chemokine (C-X-C motif) ligand 8 (CXCL8), mRNA [NM_000584] |
| A_33_P3330264 | 5,94E-05 | up | 4,943028 | CXCL1 | Homo sapiens chemokine (C-X-C motif) ligand 1 (melanoma growth stimulating activity, alpha) (CXCL1), transcript variant 1, mRNA [NM_001511] |
| A_23_P7144 | 1,86E-05 | up | 6,293819 | CXCL1 | Homo sapiens chemokine (C-X-C motif) ligand 1 (melanoma growth stimulating activity, alpha) (CXCL1), transcript variant 1, mRNA [NM_001511] |
