## Supplementary material for "Early Reduction of SARS-CoV-2 Replication in Bronchial Epithelium by Kinin B_2_ Receptor Antagonism": Supl. Table 14

**Supplemental Table S14.** Gene expression of potential viral entry receptors comparing SARS-CoV-2 versus medium

| ProbeName | p ([SARS-CoV-2 | Regulatic | FC ([SARS-( | GeneSymbol | Description |
|---|---|---|---|---|---|
| A_33_P3272493 | 0,41989717 | up | 13.300.666 | CD209 | Homo sapiens CD209 molecule (CD209), transcript variant 4, mRNA [NM_001144897] |
| A_24_P246196 | 0,16616632 | up | 14.499.686 | CLEC4M | Homo sapiens C-type lectin domain family 4, member M (CLEC4M), transcript variant 3, mRNA [NM_0011449 |
| A_23_P29067 | 0,1234412 | up | 13.455.415 | TMPRSS2 | Homo sapiens transmembrane protease, serine 2 (TMPRSS2), transcript variant 2, mRNA [NM_005656] |
| A_33_P3287223 | 0,5507992 | up | 14.774.978 | DPP4 | Homo sapiens dipeptidyl-peptidase 4 (DPP4), mRNA [NM_001935] |
| A_24_P186539 | 0,04956434 | up | 16.313.815 | CD209 | Homo sapiens CD209 molecule (CD209), transcript variant 1, mRNA [NM_021155] |
| A_23_P143120 | 0,03504883 | up | 12.945.154 | ADAM17 | Homo sapiens ADAM metallopeptidase domain 17 (ADAM17), mRNA [NM_003183] |
| A_33_P3243857 | 0,22771475 | up | 11.777.034 | ADAM10 | Homo sapiens ADAM metallopeptidase domain 10 (ADAM10), mRNA [NM_001110] |
| A_23_P48886 | 0,17386465 | up | 12.107.117 | ADAM10 | Homo sapiens ADAM metallopeptidase domain 10 (ADAM10), mRNA [NM_001110] |
| A_24_P305345 | 0,60484207 | up | 10.998.168 | CD209 | Homo sapiens CD209 molecule (CD209), transcript variant 1, mRNA [NM_021155] |
| A_23_P252981 | 0,16595131 | up | 15.016.031 | ACE2 | Homo sapiens angiotensin I converting enzyme 2 (ACE2), mRNA [NM_021804] |
| A_23_P208482 | 0,14751089 | up | 14.619.298 | CLEC4M | Homo sapiens C-type lectin domain family 4, member M (CLEC4M), transcript variant 2, mRNA [NM_0011449 |
| A_33_P3318097 | 0,37764555 | up | 17.235.893 | TMPRSS11B | Homo sapiens transmembrane protease, serine 11B (TMPRSS11B), mRNA [NM_182502] |
| A_23_P18751 | 0,2665045 | up | 1.731.451 | TMPRSS11E | Homo sapiens transmembrane protease, serine 11E (TMPRSS11E), mRNA [NM_014058] |
| A_33_P3374678 | 0,009622896 | up | 20.423.372 | TMPRSS11D | Homo sapiens transmembrane protease, serine 11D (TMPRSS11D), mRNA [NM_004262] |
| A_33_P3217845 | 0,004265649 | up | 3.431.767 | TMPRSS11A | Homo sapiens transmembrane protease, serine 11A (TMPRSS11A), transcript variant 1, mRNA [NM_182606] |
| A_24_P109101 | 0,035911784 | up | 30.592.778 | TMPRSS11A | Homo sapiens transmembrane protease, serine 11A (TMPRSS11A), transcript variant 1, mRNA [NM_182606] |
