## Supplementary material for "Early Reduction of SARS-CoV-2 Replication in Bronchial Epithelium by Kinin B_2_ Receptor Antagonism": Supl. Table 15

**Supplemental Table S15.** Membrane-bound receptor DEGs comparing SARS-CoV-2 + B2R antagonist versus SARS-CoV-2

| ProbeName | p ([SARS-Co\ | Regulation ([S | FC ([SARS-C | GeneSymbol | Description |
|---|---|---|---|---|---|
| A_33_P3258206 | 0,04418962 | down | -1,780782 | OR6N2 | Homo sapiens olfactory receptor, family 6, subfamily N, member 2 (OR6N2), mRNA [NM_001005278] |
| A_33_P3305388 | 0,00808579 | down | -1,750339 | OR8K5 | Homo sapiens olfactory receptor, family 8, subfamily K, member 5 (OR8K5), mRNA [NM_001004058] |
| A_33_P3419551 | 0,0297491 | down | -1,683478 | OR5K1 | Homo sapiens olfactory receptor, family 5, subfamily K, member 1 (OR5K1), mRNA [NM_001004736] |
| A_33_P3381097 | 0,03317082 | down | -1,9448466 | OR10V1 | Homo sapiens olfactory receptor, family 10, subfamily V, member 1 (OR10V1), mRNA [NM_001005324] |
| A_33_P3375766 | 0,01975872 | down | -1,9204632 | KCNC2 | Homo sapiens potassium channel, voltage gated Shaw related subfamily C, member 2 (KCNC2), transcript variant 3, mRNA [NM_153748] |
| A_23_P26062 | 0,04506961 | down | -1,85466 | TMEM202 | Homo sapiens transmembrane protein 202 (TMEM202), mRNA [NM_001080462] |
| A_23_P23292 | 0,01135998 | down | -1,8885309 | RXRG | Homo sapiens retinoid X receptor, gamma (RXRG), transcript variant 1, mRNA [NM_006917] |
| A_24_P229025 | 0,03108848 | down | -2,0338626 | GRIA3 | Homo sapiens glutamate receptor, ionotropic, AMPA 3 (GRIA3), transcript variant 3, mRNA [NM_001256743] |
| A_33_P3394699 | 0,01182187 | down | -1,8345557 | SCN2B | sodium channel, voltage-gated, type II, beta subunit [Source:HGNC Symbol;Acc:HGNC:10589] [ENST00000278947] |
| A_33_P3218975 | 0,04259694 | down | -1,5564153 | ENTPD1 | Homo sapiens ectonucleoside triphosphate diphosphohydrolase 1 (ENTPD1), transcript variant 1, mRNA [NM_001776] |
| A_23_P103328 | 0,0147997 | down | -1,5368288 | PTGER3 | Homo sapiens prostaglandin E receptor 3 (subtype EP3) (PTGER3), transcript variant 4, mRNA [NM_198714] |
| A_33_P3359383 | 0,0329296 | down | -2,2303607 | OR10H5 | Homo sapiens olfactory receptor, family 10, subfamily H, member 5 (OR10H5), mRNA [NM_001004466] |
| A_33_P3355856 | 0,00441775 | down | -1,965058 | | olfactory receptor, family 51, subfamily J, member 1 (gene/pseudogene) [Source:HGNC Symbol;Acc:HGNC:14856] [ENST00000332043] |
| A_33_P3330418 | 0,02829296 | down | -2,0804675 | OR7A5 | Homo sapiens olfactory receptor, family 7, subfamily A, member 5 (OR7A5), mRNA [NM_017506] |
| A_21_P0000167 | 0,04515371 | down | -1,5759147 | SORCS1 | Homo sapiens sortilin-related VPS10 domain containing receptor 1 (SORCS1), transcript variant 5, mRNA [NM_001206571] |
| A_33_P3252583 | 0,03593494 | down | -1,714341 | OR9I1 | Homo sapiens olfactory receptor, family 9, subfamily I, member 1 (OR9I1), mRNA [NM_001005211] |
| A_23_P406782 | 0,0197992 | down | -1,7807523 | HPN | Homo sapiens hepsin (HPN), transcript variant 1, mRNA [NM_182983] |
| A_24_P170439 | 0,02440206 | down | -1,7213168 | TREML3P | Homo sapiens triggering receptor expressed on myeloid cells-like 3, pseudogene (TREML3P), non-coding RNA [NR_027256] |
| A_23_P89132 | 0,02538322 | down | -1,9572493 | ZP2 | Homo sapiens zona pellucida glycoprotein 2 (sperm receptor) (ZP2), transcript variant 1, mRNA [NM_003460] |
| A_21_P0000170 | 0,02363338 | down | -1,593824 | SMIM18 | Homo sapiens small integral membrane protein 18 (SMIM18), mRNA [NM_001206847] |
| A_33_P3419562 | 0,01892465 | down | -1,7824667 | OR5K2 | Homo sapiens olfactory receptor, family 5, subfamily K, member 2 (OR5K2), mRNA [NM_001004737] |
| A_24_P178819 | 0,03222796 | down | -1,505357 | | T cell receptor alpha variable 12-2 [Source:HGNC Symbol;Acc:HGNC:12106] [ENST00000390437] |
| A_23_P109913 | 0,02610222 | down | -1,5543842 | CXCR6 | Homo sapiens chemokine (C-X-C motif) receptor 6 (CXCR6), mRNA [NM_006564] |
| A_33_P3330886 | 0,0191175 | down | -1,5555087 | OR6C65 | Homo sapiens olfactory receptor, family 6, subfamily C, member 65 (OR6C65), mRNA [NM_001005518] |
| A_33_P3422595 | 0,00383447 | down | -2,0085564 | | T cell receptor gamma variable 8 [Source:HGNC Symbol;Acc:HGNC:12294] [ENST00000390343] |
| A_33_P3416347 | 0,02879058 | down | -1,8018223 | | T cell receptor beta variable 23-1 (non-functional) [Source:HGNC Symbol;Acc:HGNC:12201] [ENST00000390396] |
| A_24_P655458 | 0,04344975 | down | -1,666827 | TREML4 | Homo sapiens triggering receptor expressed on myeloid cells-like 4 (TREML4), mRNA [NM_198153] |
| A_33_P3284621 | 0,03216589 | down | -1,7418137 | DDR2 | discoidin domain receptor tyrosine kinase 2 [Source:HGNC Symbol;Acc:HGNC:2731] [ENST00000367922] |
| A_33_P3408782 | 0,04539359 | down | -1,774031 | EPHA6 | Homo sapiens EPH receptor A6 (EPHA6), transcript variant 1, mRNA [NM_001080448] |
| A_23_P355377 | 0,02646404 | down | -1,9830261 | SLC12A5 | Homo sapiens solute carrier family 12 (potassium/chloride transporter), member 5 (SLC12A5), transcript variant 2, mRNA [NM_020708] |
| A_24_P118052 | 0,03708456 | down | -2,0904982 | TPTE2 | Homo sapiens transmembrane phosphoinositide 3-phosphatase and tensin homolog 2 (TPTE2), transcript variant 3, mRNA [NM_199254] |
| A_33_P3224535 | 0,02496846 | down | -1,6096613 | OR5D18 | Homo sapiens olfactory receptor, family 5, subfamily D, member 18 (OR5D18), mRNA [NM_001001952] |
| A_24_P31627 | 0,01857773 | down | -1,5557013 | KCNB1 | Homo sapiens potassium channel, voltage gated Shab related subfamily B, member 1 (KCNB1), mRNA [NM_004975] |
| A_33_P3392927 | 0,04515215 | down | -1,8602587 | | olfactory receptor, family 5, subfamily M, member 2 pseudogene [Source:HGNC Symbol;Acc:HGNC:14803] [ENST00000529003] |
| A_21_P0010873 | 0,01194135 | down | -1,8027905 | ANTXRLP1 | Homo sapiens anthrax toxin receptor-like pseudogene 1 (ANTXRLP1), transcript variant 2, non-coding RNA [NR_103828] |
| A_33_P3420605 | 0,03833315 | down | -2,3175163 | OR4D1 | Homo sapiens olfactory receptor, family 4, subfamily D, member 1 (OR4D1), mRNA [NM_012374] |
| A_21_P0010808 | 0,01438448 | down | -1,7226753 | MALRD1 | Homo sapiens MAM and LDL receptor class A domain containing 1 (MALRD1), mRNA [NM_001142308] |
| A_23_P21990 | 0,02143563 | down | -2,3075705 | SLC23A1 | Homo sapiens solute carrier family 23 (ascorbic acid transporter), member 1 (SLC23A1), transcript variant 2, mRNA [NM_152685] |
| A_33_P3264416 | 0,00374799 | down | -2,9500098 | | T cell receptor beta variable 18 [Source:HGNC Symbol;Acc:HGNC:12193] [ENST00000611520] |
| A_23_P205900 | 0,00460241 | down | -2,3870409 | NTRK3 | Homo sapiens neurotrophic tyrosine kinase, receptor, type 3 (NTRK3), transcript variant 1, mRNA [NM_001012338] |
| A_33_P3363725 | 0,00646022 | down | -1,9825865 | OR52N5 | Homo sapiens olfactory receptor, family 52, subfamily N, member 5 (OR52N5), mRNA [NM_001001922] |
| A_33_P3385696 | 0,01805648 | down | -1,9870719 | OR52D1 | Homo sapiens olfactory receptor, family 52, subfamily D, member 1 (OR52D1), mRNA [NM_001005163] |

| | | | | | |
|---|---|---|---|---|---|
| A_33_P3376493 | 0,03345821 | down | -1,8630021 | AGTR2 | Homo sapiens angiotensin II receptor, type 2 (AGTR2), mRNA [NM_000686] |
| A_33_P3396244 | 0,03147338 | down | -1,7422631 | NPY2R | Homo sapiens neuropeptide Y receptor Y2 (NPY2R), mRNA [NM_000910] |
| A_33_P3403399 | 0,03717863 | down | -1,6168706 | SLC47A1 | Homo sapiens solute carrier family 47 (multidrug and toxin extrusion), member 1 (SLC47A1), mRNA [NM_018242] |
| A_33_P3383988 | 0,02580289 | down | -1,7756995 | | T cell receptor alpha variable 9-2 [Source:HGNC Symbol;Acc:HGNC:12154] [ENST00000390441] |
| A_33_P3299329 | 0,02906759 | down | -2,0721483 | | H.sapiens mRNA for T-cell receptor IGR b 18 Vbeta 7. [A25969] |
| A_24_P24770 | 0,01016718 | down | -2,4188416 | SLC9C1 | Homo sapiens solute carrier family 9, subfamily C (Na+-transporting carboxylic acid decarboxylase), member 1 (SLC9C1), mRNA [NM_183061 |
| A_33_P3364811 | 0,00983733 | down | -2,0276864 | PTPRC | Homo sapiens protein tyrosine phosphatase, receptor type, C (PTPRC), transcript variant 1, mRNA [NM_002838] |
| A_33_P3226060 | 0,00557939 | down | -2,483791 | TMEM178B | Homo sapiens transmembrane protein 178B (TMEM178B), mRNA [NM_001195278] |
| A_23_P257962 | 0,01842982 | down | -1,8486652 | GRIN1 | Homo sapiens glutamate receptor, ionotropic, N-methyl D-aspartate 1 (GRIN1), transcript variant GluN1-1a, mRNA [NM_007327] |
| A_23_P413760 | 0,00141387 | down | -2,0794134 | P2RX5 | Homo sapiens purinergic receptor P2X, ligand gated ion channel, 5 (P2RX5), transcript variant 2, mRNA [NM_175080] |
| A_33_P3244215 | 0,01501591 | down | -2,1482882 | OR13C5 | Homo sapiens olfactory receptor, family 13, subfamily C, member 5 (OR13C5), mRNA [NM_001004482] |
| A_23_P13195 | 0,03000051 | down | -1,9745032 | OR8G1 | Homo sapiens olfactory receptor, family 8, subfamily G, member 1 (gene/pseudogene) (OR8G1), transcript variant 1, mRNA [NM_001002905] |
| A_33_P3340014 | 0,00952162 | down | -1,9314481 | TRO | Homo sapiens trophinin (TRO), transcript variant 3, mRNA [NM_016157] |
| A_23_P168403 | 0,01474765 | down | -1,5590942 | KCNH2 | Homo sapiens potassium voltage-gated channel, subfamily H (eag-related), member 2 (KCNH2), transcript variant 1, mRNA [NM_000238] |
| A_32_P60065 | 0,04420897 | down | -1,7265391 | F2RL2 | Homo sapiens coagulation factor II (thrombin) receptor-like 2 (F2RL2), transcript variant 1, mRNA [NM_004101] |
| A_33_P3343250 | 0,00175025 | down | -2,2864676 | GPR1 | Homo sapiens G protein-coupled receptor 1 (GPR1), transcript variant 4, mRNA [NM_001261453] |
| A_33_P3215635 | 0,01404406 | down | -1,6950067 | CHRNB2 | Homo sapiens cholinergic receptor, nicotinic, beta 2 (neuronal) (CHRNB2), mRNA [NM_000748] |
| A_33_P3261373 | 4,48E-05 | down | -3,8485372 | | olfactory receptor, family 4, subfamily C, member 5 (gene/pseudogene) [Source:HGNC Symbol;Acc:HGNC:14702] [ENST00000319813] |
| A_33_P3265709 | 0,03728425 | down | -2,1869175 | OR2Y1 | Homo sapiens olfactory receptor, family 2, subfamily Y, member 1 (OR2Y1), mRNA [NM_001001657] |
| A_24_P944964 | 0,01974662 | down | -2,1319976 | GP5 | Homo sapiens glycoprotein V (platelet) (GP5), mRNA [NM_004488] |
| A_24_P381844 | 0,00901695 | down | -1,9644018 | GRIN2A | Homo sapiens glutamate receptor, ionotropic, N-methyl D-aspartate 2A (GRIN2A), transcript variant 2, mRNA [NM_000833] |
| A_33_P3332576 | 0,00658593 | down | -2,063662 | GPR151 | Homo sapiens G protein-coupled receptor 151 (GPR151), mRNA [NM_194251] |
| A_23_P310410 | 0,01858192 | down | -1,6482953 | CD1E | Homo sapiens CD1e molecule (CD1E), transcript variant 2, mRNA [NM_001042583] |
| A_24_P945096 | 0,04818654 | down | -1,8050053 | CACNA1I | Homo sapiens calcium channel, voltage-dependent, T type, alpha 1I subunit (CACNA1I), transcript variant 1, mRNA [NM_021096] |
| A_33_P3405769 | 6,09E-04 | down | -2,5182621 | TARM1 | Homo sapiens T cell-interacting, activating receptor on myeloid cells 1 (TARM1), mRNA [NM_001135686] |
| A_33_P3323535 | 0,01457425 | down | -2,0621862 | | T cell receptor alpha variable 24 [Source:HGNC Symbol;Acc:HGNC:12121] [ENST00000390453] |
| A_33_P3413558 | 0,03426481 | down | -2,0173213 | CD226 | CD226 molecule [Source:HGNC Symbol;Acc:HGNC:16961] [ENST00000280200] |
| A_33_P3241021 | 0,01925859 | down | -2,1987698 | CD69 | CD69 molecule [Source:HGNC Symbol;Acc:HGNC:1694] [ENST00000416624] |
| A_33_P3326271 | 0,01507 | down | -2,1182187 | OR13C2 | Homo sapiens olfactory receptor, family 13, subfamily C, member 2 (OR13C2), mRNA [NM_001004481] |
| A_33_P3280646 | 0,00661566 | down | -2,2763705 | CASR | Homo sapiens calcium-sensing receptor (CASR), transcript variant 1, mRNA [NM_001178065] |
| A_23_P200728 | 0,00913424 | down | -2,0077603 | FCGR3A | Homo sapiens Fc fragment of IgG, low affinity IIIa, receptor (CD16a) (FCGR3A), transcript variant 1, mRNA [NM_000569] |
| A_33_P3224882 | 0,03538781 | down | -1,5173547 | | T cell receptor gamma variable 9 [Source:HGNC Symbol;Acc:HGNC:12295] [ENST00000444775] |
| A_33_P3238579 | 0,01659678 | down | -1,5561819 | | T cell receptor beta variable 11-3 [Source:HGNC Symbol;Acc:HGNC:12182] [ENST00000611787] |
| A_33_P3313215 | 0,0329466 | down | -1,5020438 | LRIT2 | Homo sapiens leucine-rich repeat, immunoglobulin-like and transmembrane domains 2 (LRIT2), transcript variant 1, mRNA [NM_001284223] |
| A_24_P319113 | 0,01615628 | down | -1,7449347 | P2RX7 | Homo sapiens purinergic receptor P2X, ligand-gated ion channel, 7 (P2RX7), transcript variant 1, mRNA [NM_002562] |
| A_33_P3273679 | 0,01952371 | down | -1,5049766 | NRXN1 | Homo sapiens neurexin 1, mRNA (cDNA clone IMAGE:4815048), complete cds. [BC046631] |
| A_33_P3266769 | 0,02908384 | down | -1,6246543 | LHCGR | luteinizing hormone/choriogonadotropin receptor [Source:HGNC Symbol;Acc:HGNC:6585] [ENST00000477576] |
| A_23_P95536 | 0,01830276 | down | -2,1320918 | ADAM29 | Homo sapiens ADAM metallopeptidase domain 29 (ADAM29), transcript variant 1, mRNA [NM_014269] |
| A_33_P3346529 | 0,03544362 | down | -2,147902 | OR2W5 | Homo sapiens olfactory receptor, family 2, subfamily W, member 5 (gene/pseudogene) (OR2W5), mRNA [NM_001004698] |
| A_33_P3414192 | 0,0207025 | down | -2,2150536 | SORCS1 | Homo sapiens sortilin-related VPS10 domain containing receptor 1 (SORCS1), transcript variant 6, mRNA [NM_001206572] |
| A_33_P3278013 | 0,03389755 | down | -2,0211592 | CD2 | CD2 molecule [Source:HGNC Symbol;Acc:HGNC:1639] [ENST00000369477] |
| A_23_P417173 | 0,0386335 | down | -1,7254565 | KCNA5 | Homo sapiens potassium channel, voltage gated shaker related subfamily A, member 5 (KCNA5), mRNA [NM_002234] |
| A_23_P23829 | 0,00684698 | down | -2,085793 | CD34 | Homo sapiens CD34 molecule (CD34), transcript variant 2, mRNA [NM_001773] |
| A_33_P3342056 | 0,00291116 | down | -2,5372047 | TIGIT | Homo sapiens T cell immunoreceptor with Ig and ITIM domains (TIGIT), mRNA [NM_173799] |
| A_23_P31996 | 0,02925612 | down | -1,7108716 | SLC46A2 | Homo sapiens solute carrier family 46, member 2 (SLC46A2), mRNA [NM_033051] |
| A_24_P254833 | 0,01048419 | down | -1,5933747 | | T cell receptor beta variable 7-9 [Source:HGNC Symbol;Acc:HGNC:12243] [ENST00000612787] |

| | | | | | |
|---|---|---|---|---|---|
| A_23_P64879 | 0,04161025 | down | -1,5252799 | KCNJ8 | Homo sapiens potassium channel, inwardly rectifying subfamily J, member 8 (KCNJ8), mRNA [NM_004982] |
| A_33_P3398951 | 0,02968554 | down | -1,5072616 | | T cell receptor delta variable 1 [Source:HGNC Symbol;Acc:HGNC:12262] [ENST00000390452] |
| A_33_P3270104 | 0,02106279 | down | -1,9156722 | FAM26D | Homo sapiens family with sequence similarity 26, member D (FAM26D), transcript variant 1, mRNA [NM_001256887] |
| A_24_P299769 | 0,03918802 | down | -1,680728 | OR1C1 | Homo sapiens olfactory receptor, family 1, subfamily C, member 1 (OR1C1), mRNA [NM_012353] |
| A_23_P705 | 0,02073175 | down | -1,5605893 | OR10R2 | Homo sapiens olfactory receptor, family 10, subfamily R, member 2 (OR10R2), mRNA [NM_001004472] |
| A_23_P128023 | 0,02571472 | down | -1,6901866 | OR51B4 | Homo sapiens olfactory receptor, family 51, subfamily B, member 4 (OR51B4), mRNA [NM_033179] |
| A_33_P3222124 | 0,04038541 | down | -1,528293 | OR6B3 | Homo sapiens olfactory receptor, family 6, subfamily B, member 3 (OR6B3), mRNA [NM_173351] |
| A_23_P129225 | 0,04824073 | down | -1,5774928 | TRPM1 | Homo sapiens transient receptor potential cation channel, subfamily M, member 1 (TRPM1), transcript variant 2, mRNA [NM_002420] |
| A_24_P97405 | 0,04106878 | down | -1,8425514 | CCRL2 | Homo sapiens chemokine (C-C motif) receptor-like 2 (CCRL2), transcript variant 1, mRNA [NM_003965] |
| A_23_P337867 | 0,01912973 | down | -1,8654202 | TAS2R41 | Homo sapiens taste receptor, type 2, member 41 (TAS2R41), mRNA [NM_176883] |
| A_23_P38634 | 0,04309718 | down | -1,5717353 | OR1G1 | Homo sapiens olfactory receptor, family 1, subfamily G, member 1 (OR1G1), mRNA [NM_003555] |
| A_33_P3406836 | 0,00964598 | down | -1,7121874 | HTR1F | Homo sapiens 5-hydroxytryptamine (serotonin) receptor 1F, G protein-coupled (HTR1F), mRNA [NM_000866] |
| A_33_P3416757 | 0,03221973 | down | -1,5744605 | PRLR | Homo sapiens prolactin receptor (PRLR), transcript variant 2, mRNA [NM_001204315] |
| A_23_P24294 | 0,04997865 | down | -1,6079974 | SLC17A6 | Homo sapiens solute carrier family 17 (vesicular glutamate transporter), member 6 (SLC17A6), mRNA [NM_020346] |
| A_33_P3270252 | 0,0268541 | down | -1,6455377 | OR1B1 | Homo sapiens olfactory receptor, family 1, subfamily B, member 1 (gene/pseudogene) (OR1B1), mRNA [NM_001004450] |
| A_33_P3352827 | 0,04088375 | down | -1,5143107 | SLAMF1 | Homo sapiens signaling lymphocytic activation molecule family member 1 (SLAMF1), transcript variant 1, mRNA [NM_003037] |
| A_23_P167121 | 0,02155313 | down | -1,5914401 | GABRA2 | Homo sapiens gamma-aminobutyric acid (GABA) A receptor, alpha 2 (GABRA2), transcript variant 1, mRNA [NM_000807] |
| A_23_P65629 | 0,03001706 | down | -1,5310405 | KCNK10 | Homo sapiens potassium channel, two pore domain subfamily K, member 10 (KCNK10), transcript variant 1, mRNA [NM_021161] |
| A_24_P276576 | 0,04108823 | down | -1,506987 | FCRLA | Homo sapiens Fc receptor-like A (FCRLA), transcript variant 2, mRNA [NM_032738] |
| A_24_P304311 | 0,02470366 | down | -1,5111704 | SLC22A8 | Homo sapiens solute carrier family 22 (organic anion transporter), member 8 (SLC22A8), transcript variant 1, mRNA [NM_004254] |
| A_23_P209055 | 0,03686409 | down | -1,6131264 | CD22 | Homo sapiens CD22 molecule (CD22), transcript variant 1, mRNA [NM_001771] |
| A_23_P157027 | 0,02544632 | down | -1,5663916 | VSTM2A | Homo sapiens V-set and transmembrane domain containing 2A (VSTM2A), transcript variant 1, mRNA [NM_182546] |
| A_33_P3336715 | 0,03190766 | down | -1,5230186 | GABRB2 | Homo sapiens gamma-aminobutyric acid (GABA) A receptor, beta 2 (GABRB2), transcript variant 1, mRNA [NM_021911] |
| A_23_P382240 | 0,03837828 | down | -1,5911989 | TMEM26 | Homo sapiens transmembrane protein 26 (TMEM26), mRNA [NM_178505] |
| A_33_P3349751 | 0,01031384 | down | -1,5279223 | OR4K5 | Homo sapiens olfactory receptor, family 4, subfamily K, member 5 (OR4K5), mRNA [NM_001005483] |
| A_23_P94902 | 0,02063369 | down | -1,8646433 | KCTD8 | Homo sapiens potassium channel tetramerization domain containing 8 (KCTD8), mRNA [NM_198353] |
| A_33_P3409225 | 0,01707313 | down | -1,9776696 | | T cell receptor beta variable 5-3 (non-functional) [Source:HGNC Symbol;Acc:HGNC:12220] [ENST00000390362] |
| A_33_P3318631 | 0,04450053 | down | -1,5870581 | OR5H2 | Homo sapiens olfactory receptor, family 5, subfamily H, member 2 (OR5H2), mRNA [NM_001005482] |
| A_33_P3400700 | 0,03422651 | down | -1,6804116 | SLC26A5 | Homo sapiens solute carrier family 26 (anion exchanger), member 5 (SLC26A5), transcript variant d, mRNA [NM_206885] |
| A_23_P306610 | 0,02637717 | down | -1,7250291 | MUC17 | Homo sapiens mucin 17, cell surface associated (MUC17), mRNA [NM_001040105] |
| A_23_P304509 | 0,03105481 | down | -1,7640876 | VSTM4 | Homo sapiens V-set and transmembrane domain containing 4 (VSTM4), transcript variant 2, mRNA [NM_144984] |
| A_32_P485915 | 0,02388218 | down | -1,8074466 | SLC9C2 | Homo sapiens solute carrier family 9, member C2 (putative) (SLC9C2), mRNA [NM_178527] |
| A_23_P42189 | 0,03699989 | down | -1,5042356 | SLC17A1 | Homo sapiens solute carrier family 17 (organic anion transporter), member 1 (SLC17A1), mRNA [NM_005074] |
| A_24_P98006 | 0,01291829 | down | -1,5489992 | MCHR2 | Homo sapiens melanin-concentrating hormone receptor 2 (MCHR2), transcript variant 1, mRNA [NM_001040179] |
| A_32_P169114 | 0,03321869 | down | -1,513263 | GRIN2A | Homo sapiens glutamate receptor, ionotropic, N-methyl D-aspartate 2A (GRIN2A), transcript variant 1, mRNA [NM_001134407] |
| A_23_P58464 | 0,03433973 | down | -1,567382 | PCDHB6 | Homo sapiens protocadherin beta 6 (PCDHB6), transcript variant 1, mRNA [NM_018939] |
| A_23_P59783 | 0,01958949 | down | -1,591302 | OR2A14 | Homo sapiens olfactory receptor, family 2, subfamily A, member 14 (OR2A14), mRNA [NM_001001659] |
| A_23_P62309 | 0,02713982 | down | -1,5687008 | AGTR2 | Homo sapiens angiotensin II receptor, type 2 (AGTR2), mRNA [NM_000686] |
| A_24_P945113 | 0,0275925 | down | -1,5558444 | ACVRL1 | Homo sapiens activin A receptor type II-like 1 (ACVRL1), transcript variant 1, mRNA [NM_000020] |
| A_21_P0000762 | 0,02503607 | down | -1,5392176 | OR51B5 | Homo sapiens olfactory receptor, family 51, subfamily B, member 5 (OR51B5), transcript variant 2, non-coding RNA [NR_038321] |
| A_23_P311895 | 0,01512973 | down | -1,6527369 | CLIC5 | Homo sapiens chloride intracellular channel 5 (CLIC5), transcript variant 2, mRNA [NM_016929] |
| A_24_P242581 | 0,0472428 | down | -1,5385278 | SLC5A9 | Homo sapiens solute carrier family 5 (sodium/sugar cotransporter), member 9 (SLC5A9), transcript variant 2, mRNA [NM_001011547] |
| A_23_P415706 | 0,03804463 | down | -1,501283 | GPR133 | Homo sapiens G protein-coupled receptor 133 (GPR133), mRNA [NM_198827] |
| A_33_P3386203 | 0,01391168 | down | -2,0066903 | OR2T10 | Homo sapiens olfactory receptor, family 2, subfamily T, member 10 (OR2T10), mRNA [NM_001004693] |
| A_33_P3285275 | 0,04207528 | down | -1,8045393 | CACNB2 | calcium channel, voltage-dependent, beta 2 subunit [Source:HGNC Symbol;Acc:HGNC:1402] [ENST00000498816] |
| A_23_P85039 | 0,00650566 | down | -1,6968335 | IRS4 | Homo sapiens insulin receptor substrate 4 (IRS4), mRNA [NM_003604] |

| | | | | | |
|---|---|---|---|---|---|
| A_33_P3335506 | 0,02240652 | down | -1,5982019 | FCRL5 | Fc receptor-like 5 [Source:HGNC Symbol;Acc:HGNC:18508] [ENST00000368190] |
| A_33_P3236813 | 0,03666383 | down | -1,6711134 | GPR19 | Homo sapiens G protein-coupled receptor 19 (GPR19), mRNA [NM_006143] |
| A_21_P0010561 | 0,03149213 | down | -1,549648 | FCGR1B | Homo sapiens Fc fragment of IgG, high affinity Ib, receptor (CD64) (FCGR1B), transcript variant 3, mRNA [NM_001244910] |
| A_33_P3344911 | 0,04665769 | down | -1,65717 | PTPRQ | Homo sapiens protein tyrosine phosphatase, receptor type, Q (PTPRQ), mRNA [NM_001145026] |
| A_23_P359746 | 0,02949573 | down | -1,6928108 | TAS2R38 | Homo sapiens taste receptor, type 2, member 38 (TAS2R38), mRNA [NM_176817] |
| A_24_P116710 | 0,02468533 | down | -1,5919541 | RAMP2 | Homo sapiens receptor (G protein-coupled) activity modifying protein 2 (RAMP2), mRNA [NM_005854] |
| A_33_P3222469 | 0,04641605 | down | -1,5104042 | OR4K13 | Homo sapiens olfactory receptor, family 4, subfamily K, member 13 (OR4K13), mRNA [NM_001004714] |
| A_33_P3257428 | 0,02163425 | down | -1,5667188 | | T cell receptor beta variable 7-7 [Source:HGNC Symbol;Acc:HGNC:12241] [ENST00000390377] |
| A_23_P18342 | 0,03890482 | down | -1,6254228 | EPHA6 | Homo sapiens EPH receptor A6 (EPHA6), transcript variant 3, mRNA [NM_001278300] |
| A_33_P3397599 | 0,02240254 | down | -1,5443267 | LILRA6 | Homo sapiens leukocyte immunoglobulin-like receptor, subfamily A (with TM domain), member 6 (LILRA6), transcript variant 2, non-coding RN |
| A_33_P3368776 | 0,02910659 | down | -1,5082635 | TMIGD1 | Homo sapiens transmembrane and immunoglobulin domain containing 1 (TMIGD1), mRNA [NM_206832] |
| A_33_P3363341 | 0,02262146 | down | -1,5413668 | TMEM212 | Homo sapiens transmembrane protein 212 (TMEM212), mRNA [NM_001164436] |
| A_23_P430747 | 0,01508784 | down | -1,5232787 | TMEM257 | Homo sapiens transmembrane protein 257 (TMEM257), mRNA [NM_004709] |
| A_23_P24948 | 0,01242467 | down | -1,6533204 | KCNE3 | Homo sapiens potassium channel, voltage gated subfamily E regulatory beta subunit 3 (KCNE3), mRNA [NM_005472] |
| A_33_P3385716 | 0,02717383 | down | -1,5179464 | OR52K1 | Homo sapiens olfactory receptor, family 52, subfamily K, member 1 (OR52K1), mRNA [NM_001005171] |
| A_33_P3321263 | 0,00919623 | down | -1,5800287 | | olfactory receptor, family 4, subfamily G, member 2 pseudogene [Source:HGNC Symbol;Acc:HGNC:8303] [ENST00000328113] |
| A_24_P405705 | 0,02976372 | down | -1,5351261 | SLC2A2 | Homo sapiens solute carrier family 2 (facilitated glucose transporter), member 2 (SLC2A2), transcript variant 1, mRNA [NM_000340] |
| A_24_P319374 | 0,01225101 | down | -1,620831 | GPA33 | Homo sapiens glycoprotein A33 (transmembrane) (GPA33), mRNA [NM_005814] |
| A_23_P1863 | 0,00435315 | down | -1,633698 | OR5AK2 | Homo sapiens olfactory receptor, family 5, subfamily AK, member 2 (OR5AK2), mRNA [NM_001005323] |
| A_33_P3359483 | 0,03086149 | down | -1,6683838 | OR5AC2 | Homo sapiens olfactory receptor, family 5, subfamily AC, member 2 (OR5AC2), mRNA [NM_054106] |
| A_23_P151264 | 0,02073521 | down | -1,7591082 | GRIN2B | Homo sapiens glutamate receptor, ionotropic, N-methyl D-aspartate 2B (GRIN2B), mRNA [NM_000834] |
| A_23_P115200 | 0,01340452 | down | -2,1123626 | FCRL4 | Homo sapiens Fc receptor-like 4 (FCRL4), mRNA [NM_031282] |
| A_23_P43369 | 0,00974178 | down | -1,8951069 | SIT1 | Homo sapiens signaling threshold regulating transmembrane adaptor 1 (SIT1), mRNA [NM_014450] |
| A_23_P308839 | 0,03741295 | down | -1,7640392 | TMEM132D | Homo sapiens transmembrane protein 132D (TMEM132D), mRNA [NM_133448] |
| A_33_P3400239 | 0,01600916 | down | -2,0678427 | SYNDIG1 | PREDICTED: Homo sapiens synapse differentiation inducing 1 (SYNDIG1), transcript variant X1, mRNA [XM_006723626] |
| A_24_P295465 | 0,04664632 | down | -1,7114917 | LRRTM3 | Homo sapiens leucine rich repeat transmembrane neuronal 3 (LRRTM3), transcript variant 1, mRNA [NM_178011] |
| A_33_P3254246 | 0,02412851 | down | -1,9104643 | OR7C1 | Homo sapiens olfactory receptor, family 7, subfamily C, member 1 (OR7C1), mRNA [NM_198944] |
| A_33_P3334615 | 0,02243861 | down | -1,8690815 | CNGA2 | Homo sapiens cyclic nucleotide gated channel alpha 2 (CNGA2), mRNA [NM_005140] |
| A_33_P3349746 | 0,04382502 | down | -1,5753626 | OR4K2 | Homo sapiens olfactory receptor, family 4, subfamily K, member 2 (OR4K2), mRNA [NM_001005501] |
| A_33_P3330881 | 0,04344924 | down | -1,5133283 | OR6C75 | Homo sapiens olfactory receptor, family 6, subfamily C, member 75 (OR6C75), mRNA [NM_001005497] |
| A_33_P3368991 | 0,02198167 | down | -1,5019486 | MPZ | Homo sapiens myelin protein zero (MPZ), mRNA [NM_000530] |
| A_23_P12392 | 0,02155351 | down | -1,5038058 | PTPRC | Homo sapiens protein tyrosine phosphatase, receptor type, C (PTPRC), transcript variant 4, non-coding RNA [NR_052021] |
| A_23_P167468 | 0,0266862 | down | -1,6078331 | PRLR | Homo sapiens prolactin receptor (PRLR), transcript variant 1, mRNA [NM_000949] |
| A_23_P106362 | 0,02455532 | down | -1,551873 | AQP9 | Homo sapiens aquaporin 9 (AQP9), mRNA [NM_020980] |
| A_23_P41847 | 0,03045766 | down | -1,6563475 | GABRA6 | Homo sapiens gamma-aminobutyric acid (GABA) A receptor, alpha 6 (GABRA6), mRNA [NM_000811] |
| A_33_P3357853 | 0,03612573 | down | -1,5698785 | SLC25A48 | solute carrier family 25, member 48 [Source:HGNC Symbol;Acc:HGNC:30451] [ENST00000274513] |
| A_23_P114008 | 0,02951521 | down | -1,514669 | TM4SF20 | Homo sapiens transmembrane 4 L six family member 20 (TM4SF20), mRNA [NM_024795] |
| A_33_P3211488 | 0,02943458 | down | -1,640576 | OR6K3 | Homo sapiens olfactory receptor, family 6, subfamily K, member 3 (OR6K3), mRNA [NM_001005327] |
| A_21_P0011718 | 0,02753268 | down | -1,5028747 | SMIM17 | Homo sapiens small integral membrane protein 17 (SMIM17), mRNA [NM_001193628] |
| A_21_P0000178 | 0,02854586 | down | -1,6577682 | TREM1 | Homo sapiens triggering receptor expressed on myeloid cells 1 (TREM1), transcript variant 2, mRNA [NM_001242589] |
| A_24_P930111 | 0,01632282 | down | -1,7870566 | SLC4A10 | Homo sapiens solute carrier family 4, sodium bicarbonate transporter, member 10 (SLC4A10), transcript variant 2, mRNA [NM_022058] |
| A_33_P3362652 | 0,03069491 | down | -1,8591263 | OR5H14 | Homo sapiens olfactory receptor, family 5, subfamily H, member 14 (OR5H14), mRNA [NM_001005514] |
| A_24_P223018 | 0,04327273 | down | -1,6534114 | SCN11A | Homo sapiens sodium channel, voltage gated, type XI alpha subunit (SCN11A), transcript variant 1, mRNA [NM_014139] |
| A_21_P0010946 | 0,00758381 | down | -1,7793748 | ANTXRLP1 | Homo sapiens anthrax toxin receptor-like pseudogene 1 (ANTXRLP1), transcript variant 2, non-coding RNA [NR_103828] |
| A_23_P16630 | 0,01547165 | down | -1,6853212 | OR7A5 | Homo sapiens olfactory receptor, family 7, subfamily A, member 5 (OR7A5), mRNA [NM_017506] |
| A_33_P3780983 | 0,02941013 | down | -1,6129428 | SLC34A3 | Homo sapiens solute carrier family 34 (type II sodium/phosphate cotransporter), member 3 (SLC34A3), transcript variant 2, mRNA [NM_00117 |

| | | | | | |
|---|---|---|---|---|---|
| A_24_P941896 | 0,01164032 | down | -1,7608724 | GRID1 | Homo sapiens glutamate receptor, ionotropic, delta 1 (GRID1), mRNA [NM_017551] |
| A_33_P3218980 | 0,00842023 | down | -1,7830722 | ENTPD1 | Homo sapiens ectonucleoside triphosphate diphosphohydrolase 1 (ENTPD1), transcript variant 1, mRNA [NM_001776] |
| A_33_P3379190 | 0,03627465 | down | -1,5157654 | OR6C4 | Homo sapiens olfactory receptor, family 6, subfamily C, member 4 (OR6C4), mRNA [NM_001005494] |
| A_23_P433586 | 0,03362111 | down | -1,6894854 | HTR2C | 5-hydroxytryptamine (serotonin) receptor 2C, G protein-coupled [Source:HGNC Symbol;Acc:HGNC:5295] [ENST00000276198] |
| A_23_P153155 | 0,02166423 | down | -1,6480209 | GALR1 | Homo sapiens galanin receptor 1 (GALR1), mRNA [NM_001480] |
| A_23_P367013 | 0,03421049 | down | -1,915556 | TAS2R60 | Homo sapiens taste receptor, type 2, member 60 (TAS2R60), mRNA [NM_177437] |
| A_32_P812268 | 0,00453953 | down | -1,9035704 | CALHM1 | Homo sapiens calcium homeostasis modulator 1 (CALHM1), mRNA [NM_001001412] |
| A_33_P3651948 | 0,00263292 | down | -2,4755828 | NEO1 | Homo sapiens neogenin 1 (NEO1), transcript variant 1, mRNA [NM_002499] |
| A_23_P50039 | 0,02453035 | down | -2,340947 | MC5R | Homo sapiens melanocortin 5 receptor (MC5R), mRNA [NM_005913] |
| A_23_P313278 | 0,03299773 | down | -2,2489772 | MUC3A | Homo sapiens MUC3A mRNA for intestinal mucin, partial cds. [AB038784] |
| A_23_P75707 | 0,01577365 | down | -2,4640841 | OR8H1 | Homo sapiens olfactory receptor, family 8, subfamily H, member 1 (OR8H1), mRNA [NM_001005199] |
| A_21_P0010681 | 0,01026646 | down | -1,7027928 | | olfactory receptor, family 6, subfamily R, member 1 pseudogene [Source:HGNC Symbol;Acc:HGNC:15037] [ENST00000431838] |
| A_23_P6943 | 0,0469381 | down | -2,3869176 | GPR15 | Homo sapiens G protein-coupled receptor 15 (GPR15), mRNA [NM_005290] |
| A_33_P3806676 | 0,02603149 | down | -2,4054852 | | T cell receptor beta variable 28 [Source:HGNC Symbol;Acc:HGNC:12209] [ENST00000390400] |
| A_32_P187571 | 0,03643743 | down | -1,8836282 | SCN2B | Homo sapiens sodium channel, voltage gated, type II beta subunit (SCN2B), mRNA [NM_004588] |
| A_33_P3258546 | 0,00421202 | down | -2,5884864 | PCDHA5 | Homo sapiens protocadherin alpha 5 (PCDHA5), transcript variant 2, mRNA [NM_031501] |
| A_33_P3355230 | 0,00818525 | down | -2,241749 | LAIR1 | Homo sapiens leukocyte-associated immunoglobulin-like receptor 1 (LAIR1), transcript variant a, mRNA [NM_002287] |
| A_23_P391857 | 1,56E-04 | down | -3,0335839 | ESRRB | Homo sapiens estrogen-related receptor beta (ESRRB), mRNA [NM_004452] |
| A_23_P347541 | 0,00443829 | down | -2,5857582 | GRIN3A | Homo sapiens glutamate receptor, ionotropic, N-methyl-D-aspartate 3A (GRIN3A), mRNA [NM_133445] |
| A_33_P3277611 | 0,0011636 | down | -1,8976617 | TMEM8C | Homo sapiens transmembrane protein 8C (TMEM8C), mRNA [NM_001080483] |
| A_23_P377882 | 0,02940033 | down | -2,4876564 | KCNH2 | Homo sapiens potassium voltage-gated channel, subfamily H (eag-related), member 2 (KCNH2), transcript variant 2, mRNA [NM_172056] |
| A_23_P79015 | 0,02266739 | down | -2,3576698 | SCN1B | Homo sapiens sodium channel, voltage gated, type I beta subunit (SCN1B), transcript variant b, mRNA [NM_199037] |
| A_33_P3423320 | 7,87E-04 | down | -3,9208627 | | olfactory receptor, family 8, subfamily J, member 2 (gene/pseudogene) [Source:HGNC Symbol;Acc:HGNC:15311] [ENST00000533152] |
| A_23_P14667 | 0,02109362 | down | -2,6855488 | SLC28A1 | Homo sapiens solute carrier family 28 (concentrative nucleoside transporter), member 1 (SLC28A1), transcript variant 1, mRNA [NM_004213] |
| A_33_P3290739 | 0,03496353 | down | -2,2976596 | | T cell receptor alpha variable 23/delta variable 6 [Source:HGNC Symbol;Acc:HGNC:12120] [ENST00000390451] |
| A_23_P26522 | 0,01826032 | down | -2,4486797 | AQP8 | Homo sapiens aquaporin 8 (AQP8), mRNA [NM_001169] |
| A_21_P0010506 | 0,02378679 | down | -1,8435849 | TNFRSF14 | Homo sapiens tumor necrosis factor receptor superfamily, member 14 (TNFRSF14), transcript variant 1, mRNA [NM_003820] |
| A_33_P3332955 | 0,02147174 | down | -2,0002813 | CLEC1B | Homo sapiens C-type lectin domain family 1, member B (CLEC1B), transcript variant 1, mRNA [NM_016509] |
| A_23_P255695 | 0,02983506 | down | -2,376025 | SLC17A3 | Homo sapiens solute carrier family 17 (organic anion transporter), member 3 (SLC17A3), transcript variant 2, mRNA [NM_006632] |
| A_33_P3415012 | 0,01891622 | down | -2,1078854 | KCNA6 | Homo sapiens potassium channel, voltage gated shaker related subfamily A, member 6 (KCNA6), mRNA [NM_002235] |
| A_24_P281439 | 0,02988154 | down | -1,895704 | OR2T5 | Homo sapiens olfactory receptor, family 2, subfamily T, member 5 (OR2T5), mRNA [NM_001004697] |
| A_33_P3302777 | 0,04969193 | down | -2,275277 | DCLK1 | Homo sapiens doublecortin-like kinase 1 (DCLK1), transcript variant 1, mRNA [NM_004734] |
| A_21_P0013981 | 0,02963747 | down | -1,7642552 | CHRM3 | cholinergic receptor, muscarinic 3 [Source:HGNC Symbol;Acc:HGNC:1952] [ENST00000481779] |
| A_33_P3290792 | 0,02446973 | down | -2,045246 | OR10G9 | Homo sapiens olfactory receptor, family 10, subfamily G, member 9 (OR10G9), mRNA [NM_001001953] |
| A_23_P313542 | 0,03216801 | down | -2,166786 | GRK1 | Homo sapiens G protein-coupled receptor kinase 1 (GRK1), mRNA [NM_002929] |
| A_24_P117147 | 0,005055 | down | -2,8706326 | KIR3DL1 | Homo sapiens killer cell immunoglobulin-like receptor, three domains, long cytoplasmic tail, 1 (KIR3DL1), mRNA [NM_013289] |
| A_23_P218058 | 0,02380868 | down | -2,2714703 | KLRC4 | Homo sapiens killer cell lectin-like receptor subfamily C, member 4 (KLRC4), mRNA [NM_013431] |
| A_23_P387537 | 0,00370691 | down | -2,592161 | PRRG3 | Homo sapiens proline rich Gla (G-carboxyglutamic acid) 3 (transmembrane) (PRRG3), transcript variant 1, mRNA [NM_024082] |
| A_33_P3300610 | 0,02289296 | down | -1,8640034 | OR7G2 | Homo sapiens olfactory receptor, family 7, subfamily G, member 2 (OR7G2), mRNA [NM_001005193] |
| A_23_P139654 | 0,00836091 | down | -2,1316981 | KLRC1 | Homo sapiens killer cell lectin-like receptor subfamily C, member 1 (KLRC1), transcript variant 2, mRNA [NM_007328] |
| A_23_P310931 | 0,04372076 | down | -2,2412853 | CNR2 | Homo sapiens cannabinoid receptor 2 (macrophage) (CNR2), mRNA [NM_001841] |
| A_23_P135226 | 6,52E-04 | down | -2,69251 | OR1N2 | Homo sapiens olfactory receptor, family 1, subfamily N, member 2 (OR1N2), mRNA [NM_001004457] |
| A_23_P85453 | 0,00486417 | down | -3,3950548 | CD244 | Homo sapiens CD244 molecule, natural killer cell receptor 2B4 (CD244), transcript variant 1, mRNA [NM_016382] |
| A_23_P111978 | 0,02651563 | down | -2,6335762 | KCNK9 | Homo sapiens potassium channel, two pore domain subfamily K, member 9 (KCNK9), transcript variant 1, mRNA [NM_001282534] |
| A_33_P3243008 | 0,00379402 | down | -2,5622954 | KCNU1 | Homo sapiens potassium channel, subfamily U, member 1 (KCNU1), mRNA [NM_001031836] |
| A_23_P71649 | 0,04060037 | down | -1,913628 | MUSK | Homo sapiens muscle, skeletal, receptor tyrosine kinase (MUSK), transcript variant 1, mRNA [NM_005592] |

| | | | | | |
|---|---|---|---|---|---|
| A_33_P3360296 | 0,03580659 | down | -1,9404445 | ANTXRL | Homo sapiens anthrax toxin receptor-like (ANTXRL), mRNA [NM_001278688] |
| A_32_P183765 | 0,04826975 | down | -2,016431 | ERBB4 | Homo sapiens erb-b2 receptor tyrosine kinase 4 (ERBB4), transcript variant JM-a/CVT-1, mRNA [NM_005235] |
| A_23_P254688 | 0,02878435 | down | -2,3979728 | TMEM108 | Homo sapiens transmembrane protein 108 (TMEM108), transcript variant 1, mRNA [NM_023943] |
| A_33_P3280106 | 0,03306894 | down | -3,169259 | CACNA1B | Homo sapiens calcium channel, voltage-dependent, N type, alpha 1B subunit (CACNA1B), transcript variant 2, mRNA [NM_001243812] |
| A_33_P3210399 | 0,03803759 | down | -1,8543372 | SLC14A1 | Homo sapiens solute carrier family 14 (urea transporter), member 1 (Kidd blood group) (SLC14A1), transcript variant 4, mRNA [NM_00114603 |
| A_23_P126528 | 0,02976564 | down | -2,3561606 | KCNA10 | Homo sapiens potassium channel, voltage gated shaker related subfamily A, member 10 (KCNA10), mRNA [NM_005549] |
| A_33_P3235213 | 0,0153642 | down | -2,5404625 | TIGIT | Homo sapiens T cell immunoreceptor with Ig and ITIM domains (TIGIT), mRNA [NM_173799] |
| A_23_P85963 | 0,00784953 | down | -2,8825476 | OR6Y1 | Homo sapiens olfactory receptor, family 6, subfamily Y, member 1 (OR6Y1), mRNA [NM_001005189] |
| A_33_P3260116 | 9,77E-04 | down | -2,3042867 | | T cell receptor alpha variable 26-2 [Source:HGNC Symbol;Acc:HGNC:12124] [ENST00000390460] |
| A_23_P303238 | 0,00824512 | down | -2,2485015 | VN1R5 | Homo sapiens vomeronasal 1 receptor 5 (gene/pseudogene) (VN1R5), mRNA [NM_173858] |
| A_33_P3395905 | 0,04373019 | down | -2,0588725 | | T cell receptor alpha variable 12-1 [Source:HGNC Symbol;Acc:HGNC:12105] [ENST00000390433] |
| A_33_P3295313 | 2,88E-04 | down | -3,3100839 | MS4A2 | Homo sapiens membrane-spanning 4-domains, subfamily A, member 2 (MS4A2), transcript variant 1, mRNA [NM_000139] |
| A_33_P3251024 | 0,00386581 | down | -2,1450882 | LRRTM2 | Homo sapiens leucine rich repeat transmembrane neuronal 2 (LRRTM2), mRNA [NM_015564] |
| A_23_P214727 | 0,03161659 | down | -1,8933461 | GPR63 | Homo sapiens G protein-coupled receptor 63 (GPR63), transcript variant 2, mRNA [NM_030784] |
| A_23_P123424 | 0,03585705 | down | -1,8078041 | CHRNB3 | Homo sapiens cholinergic receptor, nicotinic, beta 3 (neuronal) (CHRNB3), mRNA [NM_000749] |
| A_23_P501193 | 0,03600597 | down | -1,5049622 | KCNJ16 | Homo sapiens potassium channel, inwardly rectifying subfamily J, member 16 (KCNJ16), transcript variant 2, mRNA [NM_170741] |
| A_24_P142503 | 0,00630497 | down | -2,2221107 | SLC47A1 | Homo sapiens solute carrier family 47 (multidrug and toxin extrusion), member 1 (SLC47A1), mRNA [NM_018242] |
| A_23_P256641 | 0,027132 | down | -1,7334518 | KCNE5 | Homo sapiens potassium channel, voltage gated subfamily E regulatory beta subunit 5 (KCNE5), mRNA [NM_012282] |
| A_33_P3234020 | 0,00211696 | down | -2,629176 | IGDCC3 | Homo sapiens immunoglobulin superfamily, DCC subclass, member 3 (IGDCC3), mRNA [NM_004884] |
| A_32_P310503 | 0,04494036 | down | -2,1407912 | OR2M2 | Homo sapiens olfactory receptor, family 2, subfamily M, member 2 (OR2M2), mRNA [NM_001004688] |
| A_23_P43350 | 0,01820944 | down | -2,549153 | MLANA | Homo sapiens melan-A (MLANA), mRNA [NM_005511] |
| A_33_P3240867 | 0,01873725 | down | -2,384537 | GRK1 | Homo sapiens G protein-coupled receptor kinase 1 (GRK1), mRNA [NM_002929] |
| A_33_P3249229 | 0,02840515 | down | -2,1856308 | OR3A4P | Homo sapiens olfactory receptor, family 3, subfamily A, member 4 pseudogene (OR3A4P), non-coding RNA [NR_024128] |
| A_23_P75867 | 0,02042555 | down | -2,7280943 | OR10A4 | Homo sapiens olfactory receptor, family 10, subfamily A, member 4 (OR10A4), mRNA [NM_207186] |
| A_23_P13271 | 0,01469271 | down | -2,5435812 | | olfactory receptor, family 5, subfamily AK, member 3 pseudogene [Source:HGNC Symbol;Acc:HGNC:15252] [ENST00000527486] |
| A_24_P97687 | 0,01981213 | down | -2,5974538 | HTR1A | Homo sapiens 5-hydroxytryptamine (serotonin) receptor 1A, G protein-coupled (HTR1A), mRNA [NM_000524] |
| A_24_P289648 | 0,01360577 | down | -2,7294364 | HFE | Homo sapiens hemochromatosis (HFE), transcript variant 11, mRNA [NM_139011] |
| A_33_P3384958 | 0,04612382 | down | -1,7408122 | LPPR4 | Homo sapiens lipid phosphate phosphatase-related protein type 4 (LPPR4), transcript variant 1, mRNA [NM_014839] |
| A_23_P13479 | 0,02303714 | down | -1,939346 | OR4S1 | Homo sapiens olfactory receptor, family 4, subfamily S, member 1 (OR4S1), mRNA [NM_001004725] |
| A_24_P52293 | 0,0104476 | down | -2,7451172 | OR2A25 | Homo sapiens olfactory receptor, family 2, subfamily A, member 25 (OR2A25), mRNA [NM_001004488] |
| A_33_P3225046 | 0,03421618 | down | -2,5512295 | CD34 | Homo sapiens CD34 molecule (CD34), transcript variant 1, mRNA [NM_001025109] |
| A_33_P3383246 | 3,16E-04 | down | -2,1851623 | OR52R1 | Homo sapiens olfactory receptor, family 52, subfamily R, member 1 (gene/pseudogene) (OR52R1), mRNA [NM_001005177] |
| A_23_P81683 | 0,00596197 | down | -2,900122 | GPRC6A | Homo sapiens G protein-coupled receptor, class C, group 6, member A (GPRC6A), transcript variant 1, mRNA [NM_148963] |
| A_23_P127662 | 0,00735194 | down | -2,6807091 | OR8D1 | Homo sapiens olfactory receptor, family 8, subfamily D, member 1 (OR8D1), mRNA [NM_001002917] |
| A_23_P12746 | 0,01751062 | down | -2,5118883 | MRC1 | Homo sapiens mannose receptor, C type 1 (MRC1), mRNA [NM_002438] |
| A_33_P3263651 | 0,00737022 | down | -2,925967 | SEMA6B | Homo sapiens sema domain, transmembrane domain (TM), and cytoplasmic domain, (semaphorin) 6B (SEMA6B), mRNA [NM_032108] |
| A_23_P34424 | 0,01256995 | down | -3,5274222 | KCNQ4 | Homo sapiens potassium channel, voltage gated KQT-like subfamily Q, member 4 (KCNQ4), transcript variant 1, mRNA [NM_004700] |
| A_32_P87649 | 0,0370746 | down | -2,2158432 | TMCO2 | Homo sapiens transmembrane and coiled-coil domains 2 (TMCO2), mRNA [NM_001008740] |
| A_33_P3422248 | 0,00286837 | down | -1,7062064 | TMEM200C | transmembrane protein 200C [Source:HGNC Symbol;Acc:HGNC:37208] [ENST00000581347] |
| A_33_P3330074 | 0,01487756 | down | -1,6581982 | TRPC2 | transient receptor potential cation channel, subfamily C, member 2, pseudogene [Source:HGNC Symbol;Acc:HGNC:12334] [ENST0000045104 |
| A_23_P252082 | 0,00600484 | down | -1,7340733 | TMEM176A | Homo sapiens transmembrane protein 176A (TMEM176A), mRNA [NM_018487] |
| A_33_P3222203 | 0,04621681 | down | -1,5790596 | OXER1 | Homo sapiens oxoeicosanoid (OXE) receptor 1 (OXER1), mRNA [NM_148962] |
| A_33_P3228837 | 0,00780649 | down | -1,7269272 | CD8A | Homo sapiens CD8a molecule (CD8A), transcript variant 3, mRNA [NM_001145873] |
| A_33_P3325661 | 0,01025367 | down | -1,6960107 | | T cell receptor beta variable 24-1 [Source:HGNC Symbol;Acc:HGNC:12203] [ENST00000390397] |
| A_33_P3562537 | 0,00978484 | down | -1,727199 | RET | Homo sapiens ret proto-oncogene (RET), transcript variant 4, mRNA [NM_020630] |
| A_21_P0012333 | 0,00252424 | down | -1,7794586 | KCNMB3 | Homo sapiens potassium channel subfamily M regulatory beta subunit 3 (KCNMB3), transcript variant 2, mRNA [NM_171829] |

| | | | | | |
|---|---|---|---|---|---|
| A_33_P3342156 | 0,00112467 | down | -3,4905586 | | T cell receptor alpha variable 36/delta variable 7 [Source:HGNC Symbol;Acc:HGNC:12135] [ENST00000390463] |
| A_24_P7965 | 0,01767049 | down | -3,0306885 | ESRRG | Homo sapiens estrogen-related receptor gamma (ESRRG), transcript variant 2, mRNA [NM_206594] |
| A_23_P168993 | 0,003327 | down | -3,0895123 | ADRB3 | Homo sapiens adrenoceptor beta 3 (ADRB3), mRNA [NM_000025] |
| A_33_P3370029 | 0,04458654 | down | -2,6767948 | | T cell receptor gamma variable 11 (non-functional) [Source:HGNC Symbol;Acc:HGNC:12286] [ENST00000390340] |
| A_33_P3260659 | 0,00482139 | down | -2,3211706 | OR2T1 | Homo sapiens olfactory receptor, family 2, subfamily T, member 1 (OR2T1), mRNA [NM_030904] |
| A_23_P15832 | 2,04E-04 | down | -3,2454913 | OR1D5 | Homo sapiens olfactory receptor, family 1, subfamily D, member 5 (OR1D5), mRNA [NM_014566] |
| A_33_P3249976 | 0,00308756 | down | -2,360399 | JAM2 | Homo sapiens junctional adhesion molecule 2 (JAM2), transcript variant 1, mRNA [NM_021219] |
| A_33_P3260684 | 0,00407762 | down | -2,4415846 | OR2T6 | Homo sapiens olfactory receptor, family 2, subfamily T, member 6 (OR2T6), mRNA [NM_001005471] |
| A_33_P3417113 | 0,01487972 | down | -2,569186 | NPSR1 | Homo sapiens neuropeptide S receptor 1 (NPSR1), transcript variant 4, mRNA [NM_001300934] |
| A_33_P3341442 | 0,02313304 | down | -2,068512 | CSF2RB | Homo sapiens colony stimulating factor 2 receptor, beta, low-affinity (granulocyte-macrophage) (CSF2RB), mRNA [NM_000395] |
| A_23_P58729 | 0,00943444 | down | -2,8295608 | SLC34A1 | Homo sapiens solute carrier family 34 (type II sodium/phosphate cotransporter), member 1 (SLC34A1), transcript variant 1, mRNA [NM_003052 |
| A_21_P0011331 | 0,02811525 | down | -2,1181018 | CSPG4 | Homo sapiens chondroitin sulfate proteoglycan 4 (CSPG4), mRNA [NM_001897] |
| A_23_P385199 | 0,00241181 | down | -1,9690264 | EPHA10 | Homo sapiens EPH receptor A10 (EPHA10), transcript variant 2, mRNA [NM_173641] |
| A_24_P350759 | 0,04573663 | down | -2,0317166 | SLC1A2 | Homo sapiens solute carrier family 1 (glial high affinity glutamate transporter), member 2 (SLC1A2), transcript variant 1, mRNA [NM_004171] |
| A_23_P434289 | 0,01099968 | down | -1,8822633 | GPR62 | Homo sapiens G protein-coupled receptor 62 (GPR62), mRNA [NM_080865] |
| A_33_P3321462 | 0,03432452 | down | -2,1516585 | OR5B21 | Homo sapiens olfactory receptor, family 5, subfamily B, member 21 (OR5B21), mRNA [NM_001005218] |
| A_32_P486693 | 0,04066332 | down | -1,8055965 | NRIP3 | Homo sapiens nuclear receptor interacting protein 3 (NRIP3), mRNA [NM_020645] |
| A_33_P3278789 | 0,04292123 | down | -2,1384544 | KCNK16 | Homo sapiens potassium channel, two pore domain subfamily K, member 16 (KCNK16), transcript variant 3, mRNA [NM_001135106] |
| A_23_P402936 | 0,03402005 | down | -2,4539344 | PPFIA2 | Homo sapiens protein tyrosine phosphatase, receptor type, f polypeptide (PTPRF), interacting protein (liprin), alpha 2 (PPFIA2), transcript variar |
| A_33_P3259693 | 0,00707328 | down | -4,365351 | OR6P1 | Homo sapiens olfactory receptor, family 6, subfamily P, member 1 (OR6P1), mRNA [NM_001160325] |
| A_24_P212234 | 0,01227298 | down | -2,2397532 | SLC6A18 | Homo sapiens solute carrier family 6 (neutral amino acid transporter), member 18 (SLC6A18), mRNA [NM_182632] |
| A_23_P315991 | 2,55E-04 | down | -4,965023 | OR10A5 | Homo sapiens olfactory receptor, family 10, subfamily A, member 5 (OR10A5), mRNA [NM_178168] |
| A_23_P319423 | 0,00103116 | down | -3,493477 | KCNK5 | Homo sapiens potassium channel, two pore domain subfamily K, member 5 (KCNK5), mRNA [NM_003740] |
| A_33_P3265309 | 0,02638436 | down | -1,658292 | CEACAM4 | Homo sapiens carcinoembryonic antigen-related cell adhesion molecule 4 (CEACAM4), mRNA [NM_001817] |
| A_23_P432056 | 0,00145678 | down | -1,9153086 | RTN4RL1 | Homo sapiens reticulon 4 receptor-like 1 (RTN4RL1), mRNA [NM_178568] |
| A_23_P416191 | 5,99E-04 | down | -2,2355773 | TAS2R31 | Homo sapiens taste receptor, type 2, member 31 (TAS2R31), mRNA [NM_176885] |
| A_33_P3232120 | 6,77E-04 | down | -2,7809799 | | NMDA receptor synaptonuclear signaling and neuronal migration factor [Source:HGNC Symbol;Acc:HGNC:29843] [ENST00000371468] |
| A_33_P3262069 | 0,00572064 | down | -2,7597675 | OR2AG2 | Homo sapiens olfactory receptor, family 2, subfamily AG, member 2 (OR2AG2), mRNA [NM_001004490] |
| A_23_P156824 | 8,35E-06 | down | -3,658279 | HTR1B | Homo sapiens 5-hydroxytryptamine (serotonin) receptor 1B, G protein-coupled (HTR1B), mRNA [NM_000863] |
| A_23_P24676 | 0,01071806 | down | -2,850212 | OR8U1 | Homo sapiens olfactory receptor, family 8, subfamily U, member 1 (OR8U1), mRNA [NM_001005204] |
| A_23_P162607 | 6,94E-04 | down | -3,8409832 | STAB2 | Homo sapiens stabilin 2 (STAB2), mRNA [NM_017564] |
| A_23_P27107 | 0,00357729 | down | -3,6172636 | TM4SF5 | Homo sapiens transmembrane 4 L six family member 5 (TM4SF5), mRNA [NM_003963] |
| A_33_P3345016 | 0,00400523 | down | -2,5235734 | HTR6 | Homo sapiens 5-hydroxytryptamine (serotonin) receptor 6, G protein-coupled (HTR6), mRNA [NM_000871] |
| A_24_P350622 | 0,02168956 | down | -2,5401216 | KIR2DL4 | Homo sapiens killer cell immunoglobulin-like receptor, two domains, long cytoplasmic tail, 4 (KIR2DL4), transcript variant 1, mRNA [NM_00225 |
| A_23_P59772 | 0,00263268 | down | -2,0110087 | CLCN1 | Homo sapiens chloride channel, voltage-sensitive 1 (CLCN1), transcript variant 1, mRNA [NM_000083] |
| A_33_P3390637 | 0,00171263 | down | -2,0518823 | SLC23A3 | Homo sapiens solute carrier family 23, member 3 (SLC23A3), transcript variant 1, mRNA [NM_144712] |
| A_33_P3314151 | 5,65E-04 | down | -2,5952857 | CRACR2A | Homo sapiens calcium release activated channel regulator 2A (CRACR2A), transcript variant 1, mRNA [NM_001144958] |
| A_23_P51761 | 0,00821699 | down | -5,356401 | OR6K2 | Homo sapiens olfactory receptor, family 6, subfamily K, member 2 (OR6K2), mRNA [NM_001005279] |
| A_33_P3266489 | 0,00496559 | down | -2,2513986 | OR13H1 | Homo sapiens olfactory receptor, family 13, subfamily H, member 1 (OR13H1), mRNA [NM_001004486] |
| A_33_P3344423 | 0,03575051 | down | -2,0579205 | NCR3 | Homo sapiens natural cytotoxicity triggering receptor 3 (NCR3), transcript variant 3, mRNA [NM_001145467] |
| A_23_P151361 | 0,02803455 | down | -2,04211 | SMIM2 | Homo sapiens small integral membrane protein 2 (SMIM2), mRNA [NM_024058] |
| A_33_P3358943 | 0,01544227 | up | 1,8035592 | GRM2 | Homo sapiens glutamate receptor, metabotropic 2 (GRM2), transcript variant 1, mRNA [NM_000839] |
| A_23_P121480 | 0,02160561 | up | 1,6621829 | CD200 | Homo sapiens CD200 molecule (CD200), transcript variant 2, mRNA [NM_001004196] |
| A_33_P3295098 | 0,04833084 | up | 1,7559806 | ESYT3 | extended synaptotagmin-like protein 3 [Source:HGNC Symbol;Acc:HGNC:24295] [ENST00000289135] |
| A_23_P16469 | 0,03658775 | up | 2,0255094 | PLAUR | Homo sapiens plasminogen activator, urokinase receptor (PLAUR), transcript variant 3, mRNA [NM_001005377] |
| A_23_P98616 | 0,00797538 | up | 1,9063076 | SLC22A6 | Homo sapiens solute carrier family 22 (organic anion transporter), member 6 (SLC22A6), transcript variant 3, mRNA [NM_153277] |

| | | | | | |
|---|---|---|---|---|---|
| A_33_P3377256 | 0,00115396 | up | 2,0249357 | OGFRP1 | Homo sapiens opioid growth factor receptor pseudogene 1 (OGFRP1), non-coding RNA [NR_036498] |
| A_33_P3303291 | 0,0032364 | up | 1,6796671 | RIPK3 | Homo sapiens receptor-interacting serine-threonine kinase 3 (RIPK3), mRNA [NM_006871] |
| A_23_P19428 | 0,02121763 | up | 1,666443 | OR2J2 | Homo sapiens olfactory receptor, family 2, subfamily J, member 2 (OR2J2), mRNA [NM_030905] |
| A_23_P93302 | 0,02325907 | up | 1,5119251 | GABBR1 | Homo sapiens gamma-aminobutyric acid (GABA) B receptor, 1 (GABBR1), transcript variant 1, mRNA [NM_001470] |
| A_23_P94103 | 0,03416655 | up | 1,9073746 | SCARA5 | Homo sapiens scavenger receptor class A, member 5 (SCARA5), mRNA [NM_173833] |
| A_23_P92650 | 0,00807711 | down | -2,16979 | SLC25A2 | Homo sapiens solute carrier family 25 (mitochondrial carrier; ornithine transporter) member 2 (SLC25A2), mRNA [NM_031947] |
| A_33_P3261957 | 0,04173563 | down | -1,9484638 | CALCRL | Homo sapiens calcitonin receptor-like (CALCRL), transcript variant 1, mRNA [NM_005795] |
| A_24_P792124 | 0,00670627 | down | -2,949434 | SCN9A | Homo sapiens sodium channel, voltage gated, type IX alpha subunit (SCN9A), mRNA [NM_002977] |
| A_33_P3410274 | 0,0021756 | down | -2,7136688 | | transient receptor potential cation channel, subfamily C, member 6 pseudogene [Source:HGNC Symbol;Acc:HGNC:23270] [ENST0000042123 |
| A_33_P3375541 | 0,00132988 | down | -2,4598093 | CD3D | Homo sapiens CD3d molecule, delta (CD3-TCR complex) (CD3D), transcript variant 1, mRNA [NM_000732] |
| A_33_P3400217 | 0,02282586 | down | -2,310932 | SLC4A1 | Homo sapiens solute carrier family 4 (anion exchanger), member 1 (Diego blood group) (SLC4A1), mRNA [NM_000342] |
| A_23_P318938 | 0,02817906 | down | -2,0037775 | TMEM52B | Homo sapiens transmembrane protein 52B (TMEM52B), transcript variant 1, mRNA [NM_153022] |
| A_33_P3335511 | 0,02132568 | down | -1,8054705 | FCRL5 | Homo sapiens Fc receptor-like 5 (FCRL5), transcript variant 2, mRNA [NM_001195388] |
| A_23_P46412 | 0,00406527 | down | -1,7652537 | SCNN1D | Homo sapiens sodium channel, non voltage gated 1 delta subunit (SCNN1D), transcript variant 1, mRNA [NM_001130413] |
| A_23_P56604 | 0,02239883 | down | -1,5287845 | IL1RL2 | Homo sapiens interleukin 1 receptor-like 2 (IL1RL2), mRNA [NM_003854] |
| A_33_P3421028 | 0,01785833 | down | -1,6955128 | ROS1 | ROS proto-oncogene 1 , receptor tyrosine kinase [Source:HGNC Symbol;Acc:HGNC:10261] [ENST00000403284] |
| A_32_P489986 | 0,02345983 | down | -2,1372492 | TMEM232 | Homo sapiens transmembrane protein 232 (TMEM232), mRNA [NM_001039763] |
| A_23_P2431 | 7,59E-04 | down | -1,9494691 | C3AR1 | Homo sapiens complement component 3a receptor 1 (C3AR1), mRNA [NM_004054] |
| A_23_P373031 | 9,21E-04 | down | -2,0299516 | CACNA1C | Homo sapiens calcium channel, voltage-dependent, L type, alpha 1C subunit (CACNA1C), transcript variant 18, mRNA [NM_000719] |
| A_23_P161968 | 0,01299055 | down | -2,0607798 | SLC22A10 | Homo sapiens solute carrier family 22, member 10 (SLC22A10), mRNA [NM_001039752] |
| A_24_P383478 | 0,02022964 | down | -1,5490509 | ESR1 | Homo sapiens estrogen receptor 1 (ESR1), transcript variant 1, mRNA [NM_000125] |
| A_33_P3286157 | 0,02185078 | down | -1,5149355 | TNFRSF4 | Homo sapiens tumor necrosis factor receptor superfamily, member 4 (TNFRSF4), mRNA [NM_003327] |
| A_23_P42241 | 0,01187852 | down | -1,8668864 | OR5V1 | Homo sapiens olfactory receptor, family 5, subfamily V, member 1 (OR5V1), mRNA [NM_030876] |
| A_33_P3356577 | 0,02376668 | down | -1,943252 | SIRPB1 | signal-regulatory protein beta 1 [Source:HGNC Symbol;Acc:HGNC:15928] [ENST00000381596] |
| A_23_P90497 | 0,0304226 | down | -1,6590503 | LILRA4 | Homo sapiens leukocyte immunoglobulin-like receptor, subfamily A (with TM domain), member 4 (LILRA4), mRNA [NM_012276] |
| A_23_P363313 | 0,03322676 | down | -1,6353767 | SLC16A11 | Homo sapiens solute carrier family 16, member 11 (SLC16A11), mRNA [NM_153357] |
| A_23_P119042 | 0,03121998 | down | -1,8689405 | NKG7 | Homo sapiens natural killer cell granule protein 7 (NKG7), mRNA [NM_005601] |
| A_33_P3359647 | 0,00710027 | down | -1,9313471 | OPRL1 | Homo sapiens opiate receptor-like 1 (OPRL1), transcript variant 1, mRNA [NM_182647] |
| A_33_P3401701 | 0,04848501 | down | -1,7141898 | EDNRA | Homo sapiens endothelin receptor type A (EDNRA), transcript variant 1, mRNA [NM_001957] |
| A_33_P3279276 | 0,04257939 | down | -2,21045 | GRIP2 | Homo sapiens glutamate receptor interacting protein 2 (GRIP2), mRNA [NM_001080423] |
| A_23_P111395 | 0,02838425 | down | -1,834713 | SLC22A2 | Homo sapiens solute carrier family 22 (organic cation transporter), member 2 (SLC22A2), mRNA [NM_003058] |
| A_23_P17456 | 0,01355448 | down | -1,6302507 | SIRPB1 | Homo sapiens signal-regulatory protein beta 1 (SIRPB1), transcript variant 1, mRNA [NM_006065] |
| A_23_P92754 | 0,00185557 | down | -1,6196762 | FGFR4 | Homo sapiens fibroblast growth factor receptor 4 (FGFR4), transcript variant 3, mRNA [NM_213647] |
| A_33_P3230189 | 0,0163531 | down | -1,5824893 | SLITRK6 | Homo sapiens SLIT and NTRK-like family, member 6 (SLITRK6), mRNA [NM_032229] |
| A_33_P3273906 | 0,04627171 | down | -1,9127558 | OR10G4 | Homo sapiens olfactory receptor, family 10, subfamily G, member 4 (OR10G4), mRNA [NM_001004462] |
| A_33_P3286923 | 0,04049462 | up | 1,6092532 | TMPRSS5 | Homo sapiens transmembrane protease, serine 5 (TMPRSS5), transcript variant 4, mRNA [NM_001288751] |
| A_33_P3337500 | 0,04931976 | up | 1,5241963 | | T cell receptor alpha variable 22 [Source:HGNC Symbol;Acc:HGNC:12119] [ENST00000390450] |
| A_23_P8497 | 0,0339932 | up | 1,8711636 | GHRHR | Homo sapiens growth hormone releasing hormone receptor (GHRHR), mRNA [NM_000823] |
