## Supplementary material for "Early Reduction of SARS-CoV-2 Replication in Bronchial Epithelium by Kinin B_2_ Receptor Antagonism": Supl. Table 16

**Supplemental Table S16.** Membrane-bound receptor DEGs comparing SARS-CoV-2 + B2R antagonist versus SARS-CoV-2 (P <= 0.05; FC >=2.5)

| ProbeName | p ([SARS-Co\ | Regulation ([S | FC ([SARS-C | GeneSymbol | Description |
|---|---|---|---|---|---|
| A_33_P3259 | 0,00707328 | down | -4,365351 | OR6P1 | Homo sapiens olfactory receptor, family 6, subfamily P, member 1 (OR6P1), mRNA [NM_001160325] |
| A_23_P3159 | 2,55E-04 | down | -4,965023 | OR10A5 | Homo sapiens olfactory receptor, family 10, subfamily A, member 5 (OR10A5), mRNA [NM_178168] |
| A_23_P3194 | 0,00103116 | down | -3,493477 | KCNK5 | Homo sapiens potassium channel, two pore domain subfamily K, member 5 (KCNK5), mRNA [NM_003740] |
| A_23_P1583 | 2,04E-04 | down | -3,2454913 | OR1D5 | Homo sapiens olfactory receptor, family 1, subfamily D, member 5 (OR1D5), mRNA [NM_014566] |
| A_23_P1689 | 0,003327 | down | -3,0895123 | ADRB3 | Homo sapiens adrenoceptor beta 3 (ADRB3), mRNA [NM_000025] |
| A_33_P3262 | 0,00572064 | down | -2,7597675 | OR2AG2 | Homo sapiens olfactory receptor, family 2, subfamily AG, member 2 (OR2AG2), mRNA [NM_001004490] |
| A_24_P7965 | 0,01767049 | down | -3,0306885 | ESRRG | Homo sapiens estrogen-related receptor gamma (ESRRG), transcript variant 2, mRNA [NM_206594] |
| A_23_P1568 | 8,35E-06 | down | -3,658279 | HTR1B | Homo sapiens 5-hydroxytryptamine (serotonin) receptor 1B, G protein-coupled (HTR1B), mRNA [NM_000863] |
| A_23_P2467 | 0,01071806 | down | -2,850212 | OR8U1 | Homo sapiens olfactory receptor, family 8, subfamily U, member 1 (OR8U1), mRNA [NM_001005204] |
| A_23_P1626 | 6,94E-04 | down | -3,8409832 | STAB2 | Homo sapiens stabilin 2 (STAB2), mRNA [NM_017564] |
| A_23_P5176 | 0,00821699 | down | -5,356401 | OR6K2 | Homo sapiens olfactory receptor, family 6, subfamily K, member 2 (OR6K2), mRNA [NM_001005279] |
| A_23_P2710 | 0,00357729 | down | -3,6172636 | TM4SF5 | Homo sapiens transmembrane 4 L six family member 5 (TM4SF5), mRNA [NM_003963] |
| A_33_P3314 | 5,65E-04 | down | -2,5952857 | CRACR2A | Homo sapiens calcium release activated channel regulator 2A (CRACR2A), transcript variant 1, mRNA [NM_001144958] |
| A_33_P3345 | 0,00400523 | down | -2,5235734 | HTR6 | Homo sapiens 5-hydroxytryptamine (serotonin) receptor 6, G protein-coupled (HTR6), mRNA [NM_000871] |
| A_24_P3506 | 0,02168956 | down | -2,5401216 | KIR2DL4 | Homo sapiens killer cell immunoglobulin-like receptor, two domains, long cytoplasmic tail, 4 (KIR2DL4), transcript variant 1, mRNA [NM_002255] |
| A_24_P7921 | 0,00670627 | down | -2,949434 | SCN9A | Homo sapiens sodium channel, voltage gated, type IX alpha subunit (SCN9A), mRNA [NM_002977] |
| A_33_P3280 | 0,03306894 | down | -3,169259 | CACNA1B | Homo sapiens calcium channel, voltage-dependent, N type, alpha 1B subunit (CACNA1B), transcript variant 2, mRNA [NM_001243812] |
| A_33_P3243 | 0,00379402 | down | -2,5622954 | KCNU1 | Homo sapiens potassium channel, subfamily U, member 1 (KCNU1), mRNA [NM_001031836] |
| A_33_P3405 | 6,09E-04 | down | -2,5182621 | TARM1 | Homo sapiens T cell-interacting, activating receptor on myeloid cells 1 (TARM1), mRNA [NM_001135686] |
| A_23_P5872 | 0,00943444 | down | -2,8295608 | SLC34A1 | Homo sapiens solute carrier family 34 (type II sodium/phosphate cotransporter), member 1 (SLC34A1), transcript variant 1, mRNA [NM_003052] |
| A_33_P3295 | 2,88E-04 | down | -3,3100839 | MS4A2 | Homo sapiens membrane-spanning 4-domains, subfamily A, member 2 (MS4A2), transcript variant 1, mRNA [NM_000139] |
| A_23_P8596 | 0,00784953 | down | -2,8825476 | OR6Y1 | Homo sapiens olfactory receptor, family 6, subfamily Y, member 1 (OR6Y1), mRNA [NM_001005189] |
| A_33_P3342 | 0,00291116 | down | -2,5372047 | TIGIT | Homo sapiens T cell immunoreceptor with Ig and ITIM domains (TIGIT), mRNA [NM_173799] |
| A_33_P3235 | 0,0153642 | down | -2,5404625 | TIGIT | Homo sapiens T cell immunoreceptor with Ig and ITIM domains (TIGIT), mRNA [NM_173799] |
| A_23_P1274 | 0,01751062 | down | -2,5118883 | MRC1 | Homo sapiens mannose receptor, C type 1 (MRC1), mRNA [NM_002438] |
| A_23_P8168 | 0,00596197 | down | -2,900122 | GPRC6A | Homo sapiens G protein-coupled receptor, class C, group 6, member A (GPRC6A), transcript variant 1, mRNA [NM_148963] |
| A_33_P3417 | 0,01487972 | down | -2,569186 | NPSR1 | Homo sapiens neuropeptide S receptor 1 (NPSR1), transcript variant 4, mRNA [NM_001300934] |
| A_33_P3234 | 0,00211696 | down | -2,629176 | IGDCC3 | Homo sapiens immunoglobulin superfamily, DCC subclass, member 3 (IGDCC3), mRNA [NM_004884] |
| A_24_P5229 | 0,0104476 | down | -2,7451172 | OR2A25 | Homo sapiens olfactory receptor, family 2, subfamily A, member 25 (OR2A25), mRNA [NM_001004488] |
| A_33_P3225 | 0,03421618 | down | -2,5512295 | CD34 | Homo sapiens CD34 molecule (CD34), transcript variant 1, mRNA [NM_001025109] |
| A_23_P1276 | 0,00735194 | down | -2,6807091 | OR8D1 | Homo sapiens olfactory receptor, family 8, subfamily D, member 1 (OR8D1), mRNA [NM_001002917] |
| A_23_P4335 | 0,01820944 | down | -2,549153 | MLANA | Homo sapiens melan-A (MLANA), mRNA [NM_005511] |
| A_23_P3918 | 1,56E-04 | down | -3,0335839 | ESRRB | Homo sapiens estrogen-related receptor beta (ESRRB), mRNA [NM_004452] |
| A_24_P9768 | 0,01981213 | down | -2,5974538 | HTR1A | Homo sapiens 5-hydroxytryptamine (serotonin) receptor 1A, G protein-coupled (HTR1A), mRNA [NM_000524] |
| A_23_P7586 | 0,02042555 | down | -2,7280943 | OR10A4 | Homo sapiens olfactory receptor, family 10, subfamily A, member 4 (OR10A4), mRNA [NM_207186] |
| A_24_P2896 | 0,01360577 | down | -2,7294364 | HFE | Homo sapiens hemochromatosis (HFE), transcript variant 11, mRNA [NM_139011] |
| A_23_P3442 | 0,01256995 | down | -3,5274222 | KCNQ4 | Homo sapiens potassium channel, voltage gated KQT-like subfamily Q, member 4 (KCNQ4), transcript variant 1, mRNA [NM_004700] |
| A_24_P1171 | 0,005055 | down | -2,8706326 | KIR3DL1 | Homo sapiens killer cell immunoglobulin-like receptor, three domains, long cytoplasmic tail, 1 (KIR3DL1), mRNA [NM_013289] |
| A_33_P3258 | 0,00421202 | down | -2,5884864 | PCDHA5 | Homo sapiens protocadherin alpha 5 (PCDHA5), transcript variant 2, mRNA [NM_031501] |
| A_23_P3475 | 0,00443829 | down | -2,5857582 | GRIN3A | Homo sapiens glutamate receptor, ionotropic, N-methyl-D-aspartate 3A (GRIN3A), mRNA [NM_133445] |
| A_33_P3263 | 0,00737022 | down | -2,925967 | SEMA6B | Homo sapiens sema domain, transmembrane domain (TM), and cytoplasmic domain, (semaphorin) 6B (SEMA6B), mRNA [NM_032108] |
| A_23_P3875 | 0,00370691 | down | -2,592161 | PRRG3 | Homo sapiens proline rich Gla (G-carboxyglutamic acid) 3 (transmembrane) (PRRG3), transcript variant 1, mRNA [NM_024082] |
| A_23_P1119 | 0,02651563 | down | -2,6335762 | KCNK9 | Homo sapiens potassium channel, two pore domain subfamily K, member 9 (KCNK9), transcript variant 1, mRNA [NM_001282534] |
| A_23_P1466 | 0,02109362 | down | -2,6855488 | SLC28A1 | Homo sapiens solute carrier family 28 (concentrative nucleoside transporter), member 1 (SLC28A1), transcript variant 1, mRNA [NM_004213] |
| A_23_P1352 | 6,52E-04 | down | -2,69251 | OR1N2 | Homo sapiens olfactory receptor, family 1, subfamily N, member 2 (OR1N2), mRNA [NM_001004457] |
| A_23_P8545 | 0,00486417 | down | -3,3950548 | CD244 | Homo sapiens CD244 molecule, natural killer cell receptor 2B4 (CD244), transcript variant 1, mRNA [NM_016382] |
