## Supplementary material for "Early Reduction of SARS-CoV-2 Replication in Bronchial Epithelium by Kinin B_2_ Receptor Antagonism": Supl. Table 17

**Supplemental Table S17.** Interactions output of String network analysis of membrane-bound receptor DEGs comparing SARS-CoV-2 + B2R antagonist versus SARS-CoV-2

| #node1 | node2 | node1_string_id | node2_string_id | neight | gene_ | phylog | homolo | coexpre | experime | databa | automa | combined_score |
|---|---|---|---|---|---|---|---|---|---|---|---|---|
| ADRB3 | RAMP2 | 9606.ENSP00000343782 | 9606.ENSP00000253796 | 0 | 0 | 0 | 0 | 0 | 0 | 0,9 | 0,055 | 0,901 |
| ADRB3 | GPR15 | 9606.ENSP00000343782 | 9606.ENSP00000284311 | 0 | 0 | 0 | 0,579 | 0 | 0 | 0,9 | 0,05 | 0,9 |
| ADRB3 | HTR6 | 9606.ENSP00000343782 | 9606.ENSP00000289753 | 0 | 0 | 0 | 0,774 | 0 | 0 | 0,9 | 0,146 | 0,902 |
| ADRB3 | LHCGR | 9606.ENSP00000343782 | 9606.ENSP00000294954 | 0 | 0 | 0 | 0 | 0,044 | 0 | 0,9 | 0,153 | 0,911 |
| ADRB3 | MC5R | 9606.ENSP00000343782 | 9606.ENSP00000318077 | 0 | 0 | 0 | 0,602 | 0 | 0 | 0,9 | 0,167 | 0,905 |
| ADRB3 | GHRHR | 9606.ENSP00000343782 | 9606.ENSP00000320180 | 0 | 0 | 0 | 0 | 0 | 0 | 0,9 | 0,111 | 0,907 |
| ADRB3 | NPSR1 | 9606.ENSP00000343782 | 9606.ENSP00000370950 | 0 | 0 | 0 | 0,584 | 0 | 0 | 0,9 | 0,117 | 0,903 |
| ADRB3 | CALCRL | 9606.ENSP00000343782 | 9606.ENSP00000386972 | 0 | 0 | 0 | 0 | 0 | 0 | 0,9 | 0,171 | 0,913 |
| AGTR2 | HTR2C | 9606.ENSP00000360973 | 9606.ENSP00000276198 | 0 | 0 | 0 | 0,572 | 0 | 0 | 0,9 | 0,158 | 0,905 |
| AGTR2 | MCHR2 | 9606.ENSP00000360973 | 9606.ENSP00000281806 | 0 | 0 | 0 | 0,657 | 0 | 0 | 0,9 | 0,073 | 0,901 |
| AGTR2 | F2RL2 | 9606.ENSP00000360973 | 9606.ENSP00000296641 | 0 | 0 | 0 | 0,635 | 0 | 0 | 0,9 | 0,08 | 0,901 |
| AGTR2 | GALR1 | 9606.ENSP00000360973 | 9606.ENSP00000299727 | 0 | 0 | 0 | 0,685 | 0 | 0 | 0,9 | 0,094 | 0,901 |
| AGTR2 | C3AR1 | 9606.ENSP00000360973 | 9606.ENSP00000302079 | 0 | 0 | 0 | 0,595 | 0 | 0 | 0,9 | 0 | 0,9 |
| AGTR2 | EDNRA | 9606.ENSP00000360973 | 9606.ENSP00000315011 | 0 | 0 | 0 | 0,577 | 0,061 | 0 | 0,9 | 0,378 | 0,916 |
| AGTR2 | HTR1A | 9606.ENSP00000360973 | 9606.ENSP00000316244 | 0 | 0 | 0 | 0,572 | 0 | 0 | 0,9 | 0,09 | 0,902 |
| AGTR2 | HTR1F | 9606.ENSP00000360973 | 9606.ENSP00000322924 | 0 | 0 | 0 | 0,589 | 0 | 0 | 0,9 | 0,162 | 0,905 |
| AGTR2 | TAS2R60 | 9606.ENSP00000360973 | 9606.ENSP00000327724 | 0 | 0 | 0 | 0 | 0 | 0 | 0,9 | 0 | 0,9 |
| AGTR2 | NPY2R | 9606.ENSP00000360973 | 9606.ENSP00000332591 | 0 | 0 | 0 | 0,632 | 0,06 | 0 | 0,9 | 0,141 | 0,905 |
| AGTR2 | OPRL1 | 9606.ENSP00000360973 | 9606.ENSP00000336764 | 0 | 0 | 0 | 0,697 | 0 | 0 | 0,9 | 0,211 | 0,905 |
| AGTR2 | PTGER3 | 9606.ENSP00000360973 | 9606.ENSP00000349003 | 0 | 0 | 0 | 0 | 0 | 0 | 0,9 | 0,064 | 0,902 |
| AGTR2 | HTR1B | 9606.ENSP00000360973 | 9606.ENSP00000358963 | 0 | 0 | 0 | 0,587 | 0 | 0 | 0,9 | 0,113 | 0,903 |
| AGTR2 | OXER1 | 9606.ENSP00000360973 | 9606.ENSP00000367930 | 0 | 0 | 0 | 0,625 | 0 | 0 | 0,9 | 0 | 0,9 |
| AGTR2 | TAS2R31 | 9606.ENSP00000360973 | 9606.ENSP00000375093 | 0 | 0 | 0 | 0 | 0 | 0 | 0,9 | 0 | 0,9 |
| AGTR2 | TAS2R41 | 9606.ENSP00000360973 | 9606.ENSP00000386201 | 0 | 0 | 0 | 0 | 0 | 0 | 0,9 | 0 | 0,9 |
| AGTR2 | TAS2R38 | 9606.ENSP00000360973 | 9606.ENSP00000448219 | 0 | 0 | 0 | 0 | 0 | 0 | 0,9 | 0 | 0,9 |
| AGTR2 | CXCR6 | 9606.ENSP00000360973 | 9606.ENSP00000395704 | 0 | 0 | 0 | 0,689 | 0 | 0 | 0,9 | 0 | 0,9 |
| AGTR2 | GRM2 | 9606.ENSP00000360973 | 9606.ENSP00000378492 | 0 | 0 | 0 | 0 | 0 | 0 | 0,9 | 0,052 | 0,901 |
| AGTR2 | GABBR1 | 9606.ENSP00000360973 | 9606.ENSP00000366233 | 0 | 0 | 0 | 0 | 0 | 0 | 0,9 | 0,073 | 0,903 |
| AGTR2 | NPSR1 | 9606.ENSP00000360973 | 9606.ENSP00000370950 | 0 | 0 | 0 | 0,6 | 0 | 0 | 0,9 | 0,137 | 0,904 |
| AGTR2 | CASR | 9606.ENSP00000360973 | 9606.ENSP00000420194 | 0 | 0 | 0 | 0 | 0 | 0 | 0,9 | 0,151 | 0,911 |
| AGTR2 | CNR2 | 9606.ENSP00000360973 | 9606.ENSP00000363596 | 0 | 0 | 0 | 0 | 0 | 0 | 0,9 | 0,162 | 0,912 |
| C3AR1 | MCHR2 | 9606.ENSP00000302079 | 9606.ENSP00000281806 | 0 | 0 | 0 | 0,611 | 0 | 0 | 0,9 | 0,314 | 0,911 |
| C3AR1 | GALR1 | 9606.ENSP00000302079 | 9606.ENSP00000299727 | 0 | 0 | 0 | 0,591 | 0,061 | 0 | 0,9 | 0,227 | 0,909 |
| C3AR1 | HTR1A | 9606.ENSP00000302079 | 9606.ENSP00000316244 | 0 | 0 | 0 | 0,561 | 0 | 0 | 0,9 | 0 | 0,9 |
| C3AR1 | TARM1 | 9606.ENSP00000302079 | 9606.ENSP00000439454 | 0 | 0 | 0 | 0 | 0 | 0 | 0,9 | 0 | 0,9 |
| C3AR1 | TAS2R31 | 9606.ENSP00000302079 | 9606.ENSP00000375093 | 0 | 0 | 0 | 0 | 0 | 0 | 0,9 | 0,049 | 0,9 |
| C3AR1 | GABBR1 | 9606.ENSP00000302079 | 9606.ENSP00000366233 | 0 | 0 | 0 | 0 | 0 | 0 | 0,9 | 0 | 0,9 |
| C3AR1 | CASR | 9606.ENSP00000302079 | 9606.ENSP00000420194 | 0 | 0 | 0 | 0 | 0 | 0 | 0,9 | 0 | 0,9 |
| C3AR1 | HTR1B | 9606.ENSP00000302079 | 9606.ENSP00000358963 | 0 | 0 | 0 | 0 | 0 | 0 | 0,9 | 0 | 0,9 |
| C3AR1 | TAS2R41 | 9606.ENSP00000302079 | 9606.ENSP00000386201 | 0 | 0 | 0 | 0 | 0 | 0 | 0,9 | 0 | 0,9 |
| C3AR1 | TAS2R38 | 9606.ENSP00000302079 | 9606.ENSP00000448219 | 0 | 0 | 0 | 0 | 0 | 0 | 0,9 | 0 | 0,9 |
| C3AR1 | NPY2R | 9606.ENSP00000302079 | 9606.ENSP00000332591 | 0 | 0 | 0 | 0,582 | 0 | 0 | 0,9 | 0 | 0,9 |
| C3AR1 | TAS2R60 | 9606.ENSP00000302079 | 9606.ENSP00000327724 | 0 | 0 | 0 | 0 | 0 | 0 | 0,9 | 0 | 0,9 |
| C3AR1 | GRM2 | 9606.ENSP00000302079 | 9606.ENSP00000378492 | 0 | 0 | 0 | 0 | 0 | 0 | 0,9 | 0,05 | 0,9 |
| C3AR1 | CNR2 | 9606.ENSP00000302079 | 9606.ENSP00000363596 | 0 | 0 | 0 | 0 | 0,061 | 0 | 0,9 | 0 | 0,902 |
| C3AR1 | OPRL1 | 9606.ENSP00000302079 | 9606.ENSP00000336764 | 0 | 0 | 0 | 0,623 | 0,076 | 0 | 0,9 | 0,045 | 0,903 |
| C3AR1 | OXER1 | 9606.ENSP00000302079 | 9606.ENSP00000367930 | 0 | 0 | 0 | 0,582 | 0,096 | 0 | 0,9 | 0 | 0,905 |
| C3AR1 | PLAUR | 9606.ENSP00000302079 | 9606.ENSP00000339328 | 0 | 0 | 0 | 0 | 0,088 | 0 | 0,9 | 0,047 | 0,905 |
| C3AR1 | HTR1F | 9606.ENSP00000302079 | 9606.ENSP00000322924 | 0 | 0 | 0 | 0 | 0 | 0 | 0,9 | 0,118 | 0,908 |
| C3AR1 | PTGER3 | 9606.ENSP00000302079 | 9606.ENSP00000349003 | 0 | 0 | 0 | 0 | 0 | 0 | 0,9 | 0,126 | 0,908 |
| C3AR1 | CXCR6 | 9606.ENSP00000302079 | 9606.ENSP00000395704 | 0 | 0 | 0 | 0,596 | 0,082 | 0 | 0,9 | 0,177 | 0,909 |
| C3AR1 | LAIR1 | 9606.ENSP00000302079 | 9606.ENSP00000375622 | 0 | 0 | 0 | 0 | 0,16 | 0 | 0,9 | 0,062 | 0,914 |
| CACNA1C | CACNB2 | 9606.ENSP00000266376 | 9606.ENSP00000320025 | 0 | 0 | 0 | 0 | 0,086 | 0 | 0,9 | 0,807 | 0,98 |
| CALCRL | RAMP2 | 9606.ENSP00000386972 | 9606.ENSP00000253796 | 0 | 0 | 0 | 0 | 0,063 | 0 | 0,9 | 0,955 | 0,995 |
| CALCRL | GPR15 | 9606.ENSP00000386972 | 9606.ENSP00000284311 | 0 | 0 | 0 | 0 | 0 | 0 | 0,9 | 0,041 | 0,9 |
| CALCRL | HTR6 | 9606.ENSP00000386972 | 9606.ENSP00000289753 | 0 | 0 | 0 | 0 | 0 | 0 | 0,9 | 0 | 0,9 |
| CALCRL | LHCGR | 9606.ENSP00000386972 | 9606.ENSP00000294954 | 0 | 0 | 0 | 0 | 0,064 | 0 | 0,9 | 0,174 | 0,916 |
| CALCRL | MC5R | 9606.ENSP00000386972 | 9606.ENSP00000318077 | 0 | 0 | 0 | 0 | 0 | 0 | 0,9 | 0,166 | 0,913 |
| CALCRL | GHRHR | 9606.ENSP00000386972 | 9606.ENSP00000320180 | 0 | 0 | 0 | 0,695 | 0 | 0 | 0,9 | 0,368 | 0,91 |
| CALCRL | NPSR1 | 9606.ENSP00000386972 | 9606.ENSP00000370950 | 0 | 0 | 0 | 0 | 0 | 0 | 0,9 | 0,333 | 0,93 |
| CASR | CHRM3 | 9606.ENSP00000420194 | 9606.ENSP00000255380 | 0 | 0 | 0 | 0 | 0 | 0 | 0,9 | 0,125 | 0,908 |
| CASR | HTR2C | 9606.ENSP00000420194 | 9606.ENSP00000276198 | 0 | 0 | 0 | 0 | 0,059 | 0 | 0,9 | 0,099 | 0,907 |
| CASR | MCHR2 | 9606.ENSP00000420194 | 9606.ENSP00000281806 | 0 | 0 | 0 | 0 | 0,055 | 0 | 0,9 | 0,049 | 0,902 |
| CASR | F2RL2 | 9606.ENSP00000420194 | 9606.ENSP00000296641 | 0 | 0 | 0 | 0 | 0 | 0 | 0,9 | 0 | 0,9 |
| CASR | GALR1 | 9606.ENSP00000420194 | 9606.ENSP00000299727 | 0 | 0 | 0 | 0 | 0,061 | 0 | 0,9 | 0,094 | 0,907 |
| CASR | GPRC6A | 9606.ENSP00000420194 | 9606.ENSP00000309493 | 0 | 0 | 0 | 0,83 | 0 | 0 | 0,9 | 0,861 | 0,914 |
| CASR | EDNRA | 9606.ENSP00000420194 | 9606.ENSP00000315011 | 0 | 0 | 0 | 0 | 0 | 0 | 0,9 | 0,127 | 0,908 |
| CASR | HTR1A | 9606.ENSP00000420194 | 9606.ENSP00000316244 | 0 | 0 | 0 | 0 | 0 | 0 | 0,9 | 0,123 | 0,908 |
| CASR | HTR1F | 9606.ENSP00000420194 | 9606.ENSP00000322924 | 0 | 0 | 0 | 0 | 0 | 0 | 0,9 | 0,043 | 0,9 |
| CASR | TAS2R60 | 9606.ENSP00000420194 | 9606.ENSP00000327724 | 0 | 0 | 0 | 0 | 0 | 0 | 0,9 | 0,324 | 0,929 |
| CASR | NPY2R | 9606.ENSP00000420194 | 9606.ENSP00000332591 | 0 | 0 | 0 | 0 | 0,049 | 0 | 0,9 | 0,162 | 0,913 |
| CASR | OPRL1 | 9606.ENSP00000420194 | 9606.ENSP00000336764 | 0 | 0 | 0 | 0 | 0,055 | 0 | 0,9 | 0,121 | 0,909 |
| CASR | PTGER3 | 9606.ENSP00000420194 | 9606.ENSP00000349003 | 0 | 0 | 0 | 0 | 0 | 0 | 0,9 | 0,063 | 0,902 |
| CASR | HTR1B | 9606.ENSP00000420194 | 9606.ENSP00000358963 | 0 | 0 | 0 | 0 | 0 | 0 | 0,9 | 0,093 | 0,905 |
| CASR | CNR2 | 9606.ENSP00000420194 | 9606.ENSP00000363596 | 0 | 0 | 0 | 0 | 0 | 0 | 0,9 | 0,175 | 0,913 |

| | | | | | | | | | | | | |
|---|---|---|---|---|---|---|---|---|---|---|---|---|
| CASR | GABBR1 | 9606.ENSP00000420194 | 9606.ENSP00000366233 | 0 | 0 | 0 | 0 | 0,049 | 0 | 0,9 | 0,473 | 0,945 |
| CASR | OXER1 | 9606.ENSP00000420194 | 9606.ENSP00000367930 | 0 | 0 | 0 | 0 | 0 | 0 | 0,9 | 0,067 | 0,902 |
| CASR | NPSR1 | 9606.ENSP00000420194 | 9606.ENSP00000370950 | 0 | 0 | 0 | 0 | 0 | 0 | 0,9 | 0,68 | 0,966 |
| CASR | TAS2R31 | 9606.ENSP00000420194 | 9606.ENSP00000375093 | 0 | 0 | 0 | 0 | 0 | 0 | 0,9 | 0,337 | 0,93 |
| CASR | GRM2 | 9606.ENSP00000420194 | 9606.ENSP00000378492 | 0 | 0 | 0 | 0,801 | 0 | 0 | 0,9 | 0,297 | 0,905 |
| CASR | TAS2R41 | 9606.ENSP00000420194 | 9606.ENSP00000386201 | 0 | 0 | 0 | 0 | 0 | 0 | 0,9 | 0,309 | 0,928 |
| CASR | CXCR6 | 9606.ENSP00000420194 | 9606.ENSP00000395704 | 0 | 0 | 0 | 0 | 0 | 0 | 0,9 | 0 | 0,9 |
| CASR | TAS2R38 | 9606.ENSP00000420194 | 9606.ENSP00000448219 | 0 | 0 | 0 | 0 | 0 | 0 | 0,9 | 0,501 | 0,948 |
| CD2 | FCGR3A | 9606.ENSP00000358490 | 9606.ENSP00000356946 | 0 | 0 | 0 | 0 | 0,337 | 0 | 0,9 | 0,352 | 0,953 |
| CD22 | PTPRC | 9606.ENSP00000085219 | 9606.ENSP00000411355 | 0 | 0 | 0 | 0 | 0,182 | 0 | 0,8 | 0,816 | 0,967 |
| CD34 | PTPRC | 9606.ENSP00000310036 | 9606.ENSP00000411355 | 0 | 0 | 0 | 0 | 0 | 0 | 0 | 0,925 | 0,925 |
| CD3D | PTPRC | 9606.ENSP00000300692 | 9606.ENSP00000411355 | 0 | 0 | 0 | 0 | 0,302 | 0 | 0,9 | 0,33 | 0,949 |
| CD3D | CD8A | 9606.ENSP00000300692 | 9606.ENSP00000386559 | 0 | 0 | 0 | 0 | 0,383 | 0 | 0,9 | 0,389 | 0,959 |
| CHRM3 | NPSR1 | 9606.ENSP00000255380 | 9606.ENSP00000370950 | 0 | 0 | 0 | 0,575 | 0 | 0 | 0,9 | 0 | 0,9 |
| CHRM3 | F2RL2 | 9606.ENSP00000255380 | 9606.ENSP00000296641 | 0 | 0 | 0 | 0 | 0 | 0 | 0,9 | 0,056 | 0,901 |
| CHRM3 | GPRC6A | 9606.ENSP00000255380 | 9606.ENSP00000309493 | 0 | 0 | 0 | 0 | 0 | 0 | 0,9 | 0,062 | 0,902 |
| CHRM3 | MCHR2 | 9606.ENSP00000255380 | 9606.ENSP00000281806 | 0 | 0 | 0 | 0,577 | 0,063 | 0 | 0,9 | 0,063 | 0,903 |
| CHRM3 | EDNRA | 9606.ENSP00000255380 | 9606.ENSP00000315011 | 0 | 0 | 0 | 0 | 0 | 0 | 0,9 | 0,111 | 0,907 |
| CHRM3 | HTR2C | 9606.ENSP00000255380 | 9606.ENSP00000276198 | 0 | 0 | 0 | 0,625 | 0,098 | 0 | 0,9 | 0,318 | 0,916 |
| CHRNB2 | CHRNB3 | 9606.ENSP00000357461 | 9606.ENSP00000289957 | 0 | 0 | 0 | 0,914 | 0,085 | 0 | 0,9 | 0,801 | 0,911 |
| CNR2 | MCHR2 | 9606.ENSP00000363596 | 9606.ENSP00000281806 | 0 | 0 | 0 | 0 | 0 | 0 | 0,9 | 0,065 | 0,902 |
| CNR2 | GALR1 | 9606.ENSP00000363596 | 9606.ENSP00000299727 | 0 | 0 | 0 | 0 | 0,089 | 0 | 0,9 | 0,188 | 0,919 |
| CNR2 | HTR1A | 9606.ENSP00000363596 | 9606.ENSP00000316244 | 0 | 0 | 0 | 0 | 0 | 0 | 0,9 | 0,429 | 0,94 |
| CNR2 | HTR1F | 9606.ENSP00000363596 | 9606.ENSP00000322924 | 0 | 0 | 0 | 0 | 0,062 | 0 | 0,9 | 0,108 | 0,909 |
| CNR2 | TAS2R60 | 9606.ENSP00000363596 | 9606.ENSP00000327724 | 0 | 0 | 0 | 0 | 0 | 0 | 0,9 | 0,046 | 0,9 |
| CNR2 | NPY2R | 9606.ENSP00000363596 | 9606.ENSP00000332591 | 0 | 0 | 0 | 0 | 0 | 0 | 0,9 | 0,116 | 0,907 |
| CNR2 | OPRL1 | 9606.ENSP00000363596 | 9606.ENSP00000336764 | 0 | 0 | 0 | 0 | 0 | 0 | 0,9 | 0,652 | 0,963 |
| CNR2 | PTGER3 | 9606.ENSP00000363596 | 9606.ENSP00000349003 | 0 | 0 | 0 | 0 | 0 | 0 | 0,9 | 0,14 | 0,91 |
| CNR2 | HTR1B | 9606.ENSP00000363596 | 9606.ENSP00000358963 | 0 | 0 | 0 | 0,582 | 0 | 0 | 0,9 | 0,167 | 0,905 |
| CNR2 | TAS2R31 | 9606.ENSP00000363596 | 9606.ENSP00000375093 | 0 | 0 | 0 | 0 | 0 | 0 | 0,9 | 0 | 0,9 |
| CNR2 | TAS2R41 | 9606.ENSP00000363596 | 9606.ENSP00000386201 | 0 | 0 | 0 | 0 | 0 | 0 | 0,9 | 0,047 | 0,9 |
| CNR2 | TAS2R38 | 9606.ENSP00000363596 | 9606.ENSP00000448219 | 0 | 0 | 0 | 0 | 0 | 0 | 0,9 | 0 | 0,9 |
| CNR2 | OXER1 | 9606.ENSP00000363596 | 9606.ENSP00000367930 | 0 | 0 | 0 | 0 | 0 | 0 | 0,9 | 0,056 | 0,901 |
| CNR2 | CXCR6 | 9606.ENSP00000363596 | 9606.ENSP00000395704 | 0 | 0 | 0 | 0 | 0,061 | 0 | 0,9 | 0,091 | 0,907 |
| CNR2 | GRM2 | 9606.ENSP00000363596 | 9606.ENSP00000378492 | 0 | 0 | 0 | 0 | 0 | 0 | 0,9 | 0,229 | 0,919 |
| CNR2 | GABBR1 | 9606.ENSP00000363596 | 9606.ENSP00000366233 | 0 | 0 | 0 | 0 | 0 | 0 | 0,9 | 0,358 | 0,933 |
| CXCR6 | MCHR2 | 9606.ENSP00000395704 | 9606.ENSP00000281806 | 0 | 0 | 0 | 0,648 | 0 | 0 | 0,9 | 0 | 0,9 |
| CXCR6 | GALR1 | 9606.ENSP00000395704 | 9606.ENSP00000299727 | 0 | 0 | 0 | 0,647 | 0 | 0 | 0,9 | 0 | 0,9 |
| CXCR6 | HTR1A | 9606.ENSP00000395704 | 9606.ENSP00000316244 | 0 | 0 | 0 | 0,582 | 0 | 0 | 0,9 | 0 | 0,9 |
| CXCR6 | HTR1F | 9606.ENSP00000395704 | 9606.ENSP00000322924 | 0 | 0 | 0 | 0,578 | 0 | 0 | 0,9 | 0 | 0,9 |
| CXCR6 | TAS2R60 | 9606.ENSP00000395704 | 9606.ENSP00000327724 | 0 | 0 | 0 | 0 | 0 | 0 | 0,9 | 0 | 0,9 |
| CXCR6 | NPY2R | 9606.ENSP00000395704 | 9606.ENSP00000332591 | 0 | 0 | 0 | 0,606 | 0,062 | 0 | 0,9 | 0 | 0,902 |
| CXCR6 | OPRL1 | 9606.ENSP00000395704 | 9606.ENSP00000336764 | 0 | 0 | 0 | 0,702 | 0 | 0 | 0,9 | 0 | 0,9 |
| CXCR6 | PTGER3 | 9606.ENSP00000395704 | 9606.ENSP00000349003 | 0 | 0 | 0 | 0 | 0 | 0 | 0,9 | 0,07 | 0,903 |
| CXCR6 | HTR1B | 9606.ENSP00000395704 | 9606.ENSP00000358963 | 0 | 0 | 0 | 0 | 0 | 0 | 0,9 | 0 | 0,9 |
| CXCR6 | GABBR1 | 9606.ENSP00000395704 | 9606.ENSP00000366233 | 0 | 0 | 0 | 0 | 0 | 0 | 0,9 | 0 | 0,9 |
| CXCR6 | OXER1 | 9606.ENSP00000395704 | 9606.ENSP00000367930 | 0 | 0 | 0 | 0,612 | 0 | 0 | 0,9 | 0 | 0,9 |
| CXCR6 | TAS2R31 | 9606.ENSP00000395704 | 9606.ENSP00000375093 | 0 | 0 | 0 | 0 | 0 | 0 | 0,9 | 0 | 0,9 |
| CXCR6 | GRM2 | 9606.ENSP00000395704 | 9606.ENSP00000378492 | 0 | 0 | 0 | 0 | 0 | 0 | 0,9 | 0 | 0,9 |
| CXCR6 | TAS2R41 | 9606.ENSP00000395704 | 9606.ENSP00000386201 | 0 | 0 | 0 | 0 | 0 | 0 | 0,9 | 0 | 0,9 |
| CXCR6 | TAS2R38 | 9606.ENSP00000395704 | 9606.ENSP00000448219 | 0 | 0 | 0 | 0 | 0 | 0 | 0,9 | 0 | 0,9 |
| EDNRA | HTR2C | 9606.ENSP00000315011 | 9606.ENSP00000276198 | 0 | 0 | 0 | 0 | 0 | 0 | 0,9 | 0,108 | 0,906 |
| EDNRA | MCHR2 | 9606.ENSP00000315011 | 9606.ENSP00000281806 | 0 | 0 | 0 | 0,58 | 0 | 0 | 0,9 | 0,082 | 0,901 |
| EDNRA | F2RL2 | 9606.ENSP00000315011 | 9606.ENSP00000296641 | 0 | 0 | 0 | 0 | 0,061 | 0 | 0,9 | 0,086 | 0,906 |
| EDNRA | GPRC6A | 9606.ENSP00000315011 | 9606.ENSP00000309493 | 0 | 0 | 0 | 0 | 0 | 0 | 0,9 | 0,082 | 0,904 |
| EDNRA | NPSR1 | 9606.ENSP00000315011 | 9606.ENSP00000370950 | 0 | 0 | 0 | 0 | 0 | 0 | 0,9 | 0,125 | 0,908 |
| EPHA10 | EPHA6 | 9606.ENSP00000362139 | 9606.ENSP00000374323 | 0 | 0 | 0 | 0,929 | 0,08 | 0 | 0,9 | 0,313 | 0,905 |
| ERBB4 | PRLR | 9606.ENSP00000342235 | 9606.ENSP00000482954 | 0 | 0 | 0 | 0 | 0 | 0 | 0,9 | 0,304 | 0,927 |
| ERBB4 | GRIN2B | 9606.ENSP00000342235 | 9606.ENSP00000477455 | 0 | 0 | 0 | 0 | 0,121 | 0 | 0,9 | 0,398 | 0,942 |
| F2RL2 | HTR2C | 9606.ENSP00000296641 | 9606.ENSP00000276198 | 0 | 0 | 0 | 0 | 0 | 0 | 0,9 | 0,35 | 0,932 |
| F2RL2 | MCHR2 | 9606.ENSP00000296641 | 9606.ENSP00000281806 | 0 | 0 | 0 | 0,648 | 0 | 0 | 0,9 | 0,06 | 0,9 |
| F2RL2 | GPRC6A | 9606.ENSP00000296641 | 9606.ENSP00000309493 | 0 | 0 | 0 | 0 | 0 | 0 | 0,9 | 0 | 0,9 |
| F2RL2 | NPSR1 | 9606.ENSP00000296641 | 9606.ENSP00000370950 | 0 | 0 | 0 | 0,601 | 0 | 0 | 0,9 | 0 | 0,9 |
| FCGR1B | MRC1 | 9606.ENSP00000358391 | 9606.ENSP00000455897 | 0 | 0 | 0 | 0 | 0,124 | 0 | 0,9 | 0,115 | 0,915 |
| GABBR1 | MCHR2 | 9606.ENSP00000366233 | 9606.ENSP00000281806 | 0 | 0 | 0 | 0 | 0,063 | 0 | 0,9 | 0,103 | 0,908 |
| GABBR1 | GALR1 | 9606.ENSP00000366233 | 9606.ENSP00000299727 | 0 | 0 | 0 | 0 | 0,061 | 0 | 0,9 | 0,118 | 0,91 |
| GABBR1 | HTR1A | 9606.ENSP00000366233 | 9606.ENSP00000316244 | 0 | 0 | 0 | 0 | 0,108 | 0 | 0,9 | 0,638 | 0,964 |
| GABBR1 | HTR1F | 9606.ENSP00000366233 | 9606.ENSP00000322924 | 0 | 0 | 0 | 0 | 0,062 | 0 | 0,9 | 0,185 | 0,916 |
| GABBR1 | TAS2R60 | 9606.ENSP00000366233 | 9606.ENSP00000327724 | 0 | 0 | 0 | 0 | 0 | 0 | 0,9 | 0 | 0,9 |
| GABBR1 | NPY2R | 9606.ENSP00000366233 | 9606.ENSP00000332591 | 0 | 0 | 0 | 0 | 0,088 | 0 | 0,9 | 0,267 | 0,927 |
| GABBR1 | OPRL1 | 9606.ENSP00000366233 | 9606.ENSP00000336764 | 0 | 0 | 0 | 0 | 0,098 | 0 | 0,9 | 0,583 | 0,959 |
| GABBR1 | PTGER3 | 9606.ENSP00000366233 | 9606.ENSP00000349003 | 0 | 0 | 0 | 0 | 0 | 0 | 0,9 | 0,131 | 0,909 |
| GABBR1 | HTR1B | 9606.ENSP00000366233 | 9606.ENSP00000358963 | 0 | 0 | 0 | 0 | 0,082 | 0 | 0,9 | 0,297 | 0,929 |
| GABBR1 | TAS2R31 | 9606.ENSP00000366233 | 9606.ENSP00000375093 | 0 | 0 | 0 | 0 | 0 | 0 | 0,9 | 0 | 0,9 |
| GABBR1 | TAS2R41 | 9606.ENSP00000366233 | 9606.ENSP00000386201 | 0 | 0 | 0 | 0 | 0 | 0 | 0,9 | 0 | 0,9 |
| GABBR1 | TAS2R38 | 9606.ENSP00000366233 | 9606.ENSP00000448219 | 0 | 0 | 0 | 0 | 0 | 0 | 0,9 | 0,041 | 0,9 |
| GABBR1 | OXER1 | 9606.ENSP00000366233 | 9606.ENSP00000367930 | 0 | 0 | 0 | 0 | 0 | 0 | 0,9 | 0,056 | 0,901 |
| GABBR1 | KCNJ16 | 9606.ENSP00000366233 | 9606.ENSP00000465295 | 0 | 0 | 0 | 0 | 0,051 | 0 | 0,9 | 0 | 0,901 |
| GABBR1 | GRM2 | 9606.ENSP00000366233 | 9606.ENSP00000378492 | 0 | 0 | 0 | 0,545 | 0,108 | 0 | 0,9 | 0,574 | 0,93 |

| | | | | | | | | | | | | |
|---|---|---|---|---|---|---|---|---|---|---|---|---|
| GABRA2 | GABRA6 | 9606.ENSP00000421828 | 9606.ENSP00000274545 | 0 | 0 | 0 | 0,954 | 0,158 | 0 | 0,9 | 0,702 | 0,915 |
| GABRA2 | GABRB2 | 9606.ENSP00000421828 | 9606.ENSP00000274547 | 0 | 0 | 0 | 0,847 | 0,176 | 0 | 0,9 | 0,72 | 0,923 |
| GABRA6 | GABRB2 | 9606.ENSP00000274545 | 9606.ENSP00000274547 | 0 | 0 | 0 | 0,832 | 0,458 | 0 | 0,9 | 0,783 | 0,95 |
| GALR1 | MCHR2 | 9606.ENSP00000299727 | 9606.ENSP00000281806 | 0 | 0 | 0 | 0,672 | 0 | 0 | 0,9 | 0,508 | 0,916 |
| GALR1 | TAS2R31 | 9606.ENSP00000299727 | 9606.ENSP00000375093 | 0 | 0 | 0 | 0 | 0 | 0 | 0,9 | 0 | 0,9 |
| GALR1 | PTGER3 | 9606.ENSP00000299727 | 9606.ENSP00000349003 | 0 | 0 | 0 | 0 | 0 | 0 | 0,9 | 0,041 | 0,9 |
| GALR1 | TAS2R41 | 9606.ENSP00000299727 | 9606.ENSP00000386201 | 0 | 0 | 0 | 0 | 0 | 0 | 0,9 | 0 | 0,9 |
| GALR1 | TAS2R38 | 9606.ENSP00000299727 | 9606.ENSP00000448219 | 0 | 0 | 0 | 0 | 0 | 0 | 0,9 | 0 | 0,9 |
| GALR1 | TAS2R60 | 9606.ENSP00000299727 | 9606.ENSP00000327724 | 0 | 0 | 0 | 0 | 0 | 0 | 0,9 | 0 | 0,9 |
| GALR1 | OXER1 | 9606.ENSP00000299727 | 9606.ENSP00000367930 | 0 | 0 | 0 | 0,59 | 0 | 0 | 0,9 | 0,167 | 0,905 |
| GALR1 | HTR1F | 9606.ENSP00000299727 | 9606.ENSP00000322924 | 0 | 0 | 0 | 0,586 | 0,062 | 0 | 0,9 | 0,145 | 0,906 |
| GALR1 | HTR1B | 9606.ENSP00000299727 | 9606.ENSP00000358963 | 0 | 0 | 0 | 0,596 | 0,061 | 0 | 0,9 | 0,143 | 0,906 |
| GALR1 | OPRL1 | 9606.ENSP00000299727 | 9606.ENSP00000336764 | 0 | 0 | 0 | 0,777 | 0,063 | 0 | 0,9 | 0,399 | 0,91 |
| GALR1 | NPY2R | 9606.ENSP00000299727 | 9606.ENSP00000332591 | 0 | 0 | 0 | 0,751 | 0,063 | 0 | 0,9 | 0,442 | 0,912 |
| GALR1 | GRM2 | 9606.ENSP00000299727 | 9606.ENSP00000378492 | 0 | 0 | 0 | 0 | 0,061 | 0 | 0,9 | 0,165 | 0,914 |
| GALR1 | HTR1A | 9606.ENSP00000299727 | 9606.ENSP00000316244 | 0 | 0 | 0 | 0,587 | 0,062 | 0 | 0,9 | 0,473 | 0,92 |
| GHRHR | RAMP2 | 9606.ENSP00000320180 | 9606.ENSP00000253796 | 0 | 0 | 0 | 0 | 0 | 0 | 0,9 | 0,108 | 0,907 |
| GHRHR | GPR15 | 9606.ENSP00000320180 | 9606.ENSP00000284311 | 0 | 0 | 0 | 0 | 0 | 0 | 0,9 | 0,05 | 0,9 |
| GHRHR | HTR6 | 9606.ENSP00000320180 | 9606.ENSP00000289753 | 0 | 0 | 0 | 0 | 0 | 0 | 0,9 | 0 | 0,9 |
| GHRHR | LHCGR | 9606.ENSP00000320180 | 9606.ENSP00000294954 | 0 | 0 | 0 | 0 | 0,061 | 0 | 0,9 | 0,166 | 0,914 |
| GHRHR | MC5R | 9606.ENSP00000320180 | 9606.ENSP00000318077 | 0 | 0 | 0 | 0 | 0 | 0 | 0,9 | 0,058 | 0,901 |
| GHRHR | NPSR1 | 9606.ENSP00000320180 | 9606.ENSP00000370950 | 0 | 0 | 0 | 0 | 0 | 0 | 0,9 | 0,137 | 0,91 |
| GPR15 | RAMP2 | 9606.ENSP00000284311 | 9606.ENSP00000253796 | 0 | 0 | 0 | 0 | 0 | 0 | 0,9 | 0 | 0,9 |
| GPR15 | HTR6 | 9606.ENSP00000284311 | 9606.ENSP00000289753 | 0 | 0 | 0 | 0 | 0 | 0 | 0,9 | 0 | 0,9 |
| GPR15 | NPSR1 | 9606.ENSP00000284311 | 9606.ENSP00000370950 | 0 | 0 | 0 | 0,594 | 0 | 0 | 0,9 | 0,041 | 0,9 |
| GPR15 | MC5R | 9606.ENSP00000284311 | 9606.ENSP00000318077 | 0 | 0 | 0 | 0 | 0 | 0 | 0,9 | 0 | 0,9 |
| GPR15 | LHCGR | 9606.ENSP00000284311 | 9606.ENSP00000294954 | 0 | 0 | 0 | 0 | 0 | 0 | 0,9 | 0,067 | 0,902 |
| GPRC6A | HTR2C | 9606.ENSP00000309493 | 9606.ENSP00000276198 | 0 | 0 | 0 | 0 | 0 | 0 | 0,9 | 0,098 | 0,905 |
| GPRC6A | MCHR2 | 9606.ENSP00000309493 | 9606.ENSP00000281806 | 0 | 0 | 0 | 0 | 0 | 0 | 0,9 | 0,074 | 0,903 |
| GPRC6A | GRM2 | 9606.ENSP00000309493 | 9606.ENSP00000378492 | 0 | 0 | 0 | 0,706 | 0,062 | 0 | 0,9 | 0,195 | 0,906 |
| GPRC6A | NPSR1 | 9606.ENSP00000309493 | 9606.ENSP00000370950 | 0 | 0 | 0 | 0 | 0 | 0 | 0,9 | 0,307 | 0,927 |
| GRIA3 | GRIN1 | 9606.ENSP00000481554 | 9606.ENSP00000360608 | 0 | 0 | 0 | 0,639 | 0,159 | 0 | 0,9 | 0,77 | 0,936 |
| GRIA3 | GRIN2A | 9606.ENSP00000481554 | 9606.ENSP00000379818 | 0 | 0 | 0 | 0,603 | 0,164 | 0 | 0,9 | 0,712 | 0,937 |
| GRIA3 | GRIN2B | 9606.ENSP00000481554 | 9606.ENSP00000477455 | 0 | 0 | 0 | 0,605 | 0,151 | 0 | 0,9 | 0,712 | 0,935 |
| GRIA3 | GRIP2 | 9606.ENSP00000481554 | 9606.ENSP00000480660 | 0 | 0 | 0 | 0 | 0,064 | 0 | 0,9 | 0,346 | 0,933 |
| GRIN1 | GRIN3A | 9606.ENSP00000360608 | 9606.ENSP00000355155 | 0 | 0 | 0 | 0,662 | 0,098 | 0 | 0,9 | 0,815 | 0,931 |
| GRIN1 | GRIN2A | 9606.ENSP00000360608 | 9606.ENSP00000379818 | 0 | 0 | 0 | 0,687 | 0,266 | 0 | 0,9 | 0,96 | 0,946 |
| GRIN1 | GRIN2B | 9606.ENSP00000360608 | 9606.ENSP00000477455 | 0 | 0 | 0 | 0,697 | 0,267 | 0 | 0,9 | 0,935 | 0,945 |
| GRIN2A | GRIN3A | 9606.ENSP00000379818 | 9606.ENSP00000355155 | 0 | 0 | 0 | 0,658 | 0,063 | 0 | 0,9 | 0,861 | 0,93 |
| GRIN2A | GRIN2B | 9606.ENSP00000379818 | 9606.ENSP00000477455 | 0 | 0 | 0 | 0,945 | 0,175 | 0 | 0,9 | 0,961 | 0,918 |
| GRIN2B | GRIN3A | 9606.ENSP00000477455 | 9606.ENSP00000355155 | 0 | 0 | 0 | 0,661 | 0,063 | 0 | 0,9 | 0,876 | 0,931 |
| GRM2 | MCHR2 | 9606.ENSP00000378492 | 9606.ENSP00000281806 | 0 | 0 | 0 | 0 | 0,085 | 0 | 0,9 | 0,079 | 0,908 |
| GRM2 | HTR1A | 9606.ENSP00000378492 | 9606.ENSP00000316244 | 0 | 0 | 0 | 0 | 0,117 | 0 | 0,9 | 0,577 | 0,959 |
| GRM2 | HTR1F | 9606.ENSP00000378492 | 9606.ENSP00000322924 | 0 | 0 | 0 | 0 | 0,061 | 0 | 0,9 | 0,164 | 0,914 |
| GRM2 | TAS2R60 | 9606.ENSP00000378492 | 9606.ENSP00000327724 | 0 | 0 | 0 | 0 | 0 | 0 | 0,9 | 0,061 | 0,902 |
| GRM2 | NPY2R | 9606.ENSP00000378492 | 9606.ENSP00000332591 | 0 | 0 | 0 | 0 | 0,072 | 0 | 0,9 | 0,214 | 0,92 |
| GRM2 | OPRL1 | 9606.ENSP00000378492 | 9606.ENSP00000336764 | 0 | 0 | 0 | 0 | 0,086 | 0 | 0,9 | 0,44 | 0,944 |
| GRM2 | PTGER3 | 9606.ENSP00000378492 | 9606.ENSP00000349003 | 0 | 0 | 0 | 0 | 0 | 0 | 0,9 | 0,093 | 0,905 |
| GRM2 | HTR1B | 9606.ENSP00000378492 | 9606.ENSP00000358963 | 0 | 0 | 0 | 0 | 0,062 | 0 | 0,9 | 0,313 | 0,929 |
| GRM2 | OXER1 | 9606.ENSP00000378492 | 9606.ENSP00000367930 | 0 | 0 | 0 | 0 | 0 | 0 | 0,9 | 0,059 | 0,901 |
| GRM2 | TAS2R31 | 9606.ENSP00000378492 | 9606.ENSP00000375093 | 0 | 0 | 0 | 0 | 0 | 0 | 0,9 | 0,042 | 0,9 |
| GRM2 | TAS2R41 | 9606.ENSP00000378492 | 9606.ENSP00000386201 | 0 | 0 | 0 | 0 | 0 | 0 | 0,9 | 0,058 | 0,901 |
| GRM2 | TAS2R38 | 9606.ENSP00000378492 | 9606.ENSP00000448219 | 0 | 0 | 0 | 0 | 0 | 0 | 0,9 | 0,095 | 0,905 |
| HTR1A | MCHR2 | 9606.ENSP00000316244 | 9606.ENSP00000281806 | 0 | 0 | 0 | 0,572 | 0,111 | 0 | 0,9 | 0,164 | 0,912 |
| HTR1A | OXER1 | 9606.ENSP00000316244 | 9606.ENSP00000367930 | 0 | 0 | 0 | 0 | 0 | 0 | 0,9 | 0 | 0,9 |
| HTR1A | TAS2R31 | 9606.ENSP00000316244 | 9606.ENSP00000375093 | 0 | 0 | 0 | 0 | 0 | 0 | 0,9 | 0 | 0,9 |
| HTR1A | TAS2R38 | 9606.ENSP00000316244 | 9606.ENSP00000448219 | 0 | 0 | 0 | 0 | 0 | 0 | 0,9 | 0 | 0,9 |
| HTR1A | TAS2R60 | 9606.ENSP00000316244 | 9606.ENSP00000327724 | 0 | 0 | 0 | 0 | 0 | 0 | 0,9 | 0 | 0,9 |
| HTR1A | TAS2R41 | 9606.ENSP00000316244 | 9606.ENSP00000386201 | 0 | 0 | 0 | 0 | 0 | 0 | 0,9 | 0,086 | 0,904 |
| HTR1A | PTGER3 | 9606.ENSP00000316244 | 9606.ENSP00000349003 | 0 | 0 | 0 | 0 | 0 | 0 | 0,9 | 0,118 | 0,908 |
| HTR1A | HTR1F | 9606.ENSP00000316244 | 9606.ENSP00000322924 | 0 | 0 | 0 | 0,896 | 0,062 | 0 | 0,9 | 0,748 | 0,909 |
| HTR1A | NPY2R | 9606.ENSP00000316244 | 9606.ENSP00000332591 | 0 | 0 | 0 | 0,597 | 0,171 | 0 | 0,9 | 0,348 | 0,924 |
| HTR1A | OPRL1 | 9606.ENSP00000316244 | 9606.ENSP00000336764 | 0 | 0 | 0 | 0,589 | 0,087 | 0 | 0,9 | 0,588 | 0,927 |
| HTR1A | HTR1B | 9606.ENSP00000316244 | 9606.ENSP00000358963 | 0 | 0 | 0 | 0,904 | 0,097 | 0 | 0,9 | 0,938 | 0,914 |
| HTR1B | MCHR2 | 9606.ENSP00000358963 | 9606.ENSP00000281806 | 0 | 0 | 0 | 0,594 | 0,062 | 0 | 0,9 | 0 | 0,902 |
| HTR1B | HTR1F | 9606.ENSP00000358963 | 9606.ENSP00000322924 | 0 | 0 | 0 | 0,932 | 0,061 | 0 | 0,9 | 0,842 | 0,907 |
| HTR1B | TAS2R60 | 9606.ENSP00000358963 | 9606.ENSP00000327724 | 0 | 0 | 0 | 0 | 0 | 0 | 0,9 | 0 | 0,9 |
| HTR1B | NPY2R | 9606.ENSP00000358963 | 9606.ENSP00000332591 | 0 | 0 | 0 | 0,616 | 0,095 | 0 | 0,9 | 0,144 | 0,909 |
| HTR1B | OPRL1 | 9606.ENSP00000358963 | 9606.ENSP00000336764 | 0 | 0 | 0 | 0,622 | 0,062 | 0 | 0,9 | 0,472 | 0,918 |
| HTR1B | PTGER3 | 9606.ENSP00000358963 | 9606.ENSP00000349003 | 0 | 0 | 0 | 0 | 0 | 0 | 0,9 | 0,064 | 0,902 |
| HTR1B | OXER1 | 9606.ENSP00000358963 | 9606.ENSP00000367930 | 0 | 0 | 0 | 0 | 0 | 0 | 0,9 | 0 | 0,9 |
| HTR1B | TAS2R31 | 9606.ENSP00000358963 | 9606.ENSP00000375093 | 0 | 0 | 0 | 0 | 0 | 0 | 0,9 | 0 | 0,9 |
| HTR1B | TAS2R38 | 9606.ENSP00000358963 | 9606.ENSP00000448219 | 0 | 0 | 0 | 0 | 0 | 0 | 0,9 | 0 | 0,9 |
| HTR1B | TAS2R41 | 9606.ENSP00000358963 | 9606.ENSP00000386201 | 0 | 0 | 0 | 0 | 0 | 0 | 0,9 | 0,09 | 0,905 |
| HTR1F | MCHR2 | 9606.ENSP00000322924 | 9606.ENSP00000281806 | 0 | 0 | 0 | 0,598 | 0 | 0 | 0,9 | 0,221 | 0,907 |
| HTR1F | TAS2R41 | 9606.ENSP00000322924 | 9606.ENSP00000386201 | 0 | 0 | 0 | 0 | 0 | 0 | 0,9 | 0 | 0,9 |
| HTR1F | TAS2R38 | 9606.ENSP00000322924 | 9606.ENSP00000448219 | 0 | 0 | 0 | 0 | 0 | 0 | 0,9 | 0 | 0,9 |
| HTR1F | TAS2R31 | 9606.ENSP00000322924 | 9606.ENSP00000375093 | 0 | 0 | 0 | 0 | 0 | 0 | 0,9 | 0 | 0,9 |
| HTR1F | TAS2R60 | 9606.ENSP00000322924 | 9606.ENSP00000327724 | 0 | 0 | 0 | 0 | 0 | 0 | 0,9 | 0 | 0,9 |

| | | | | | | | | | | | | |
|---|---|---|---|---|---|---|---|---|---|---|---|---|
| HTR1F | PTGER3 | 9606.ENSP00000322924 | 9606.ENSP00000349003 | 0 | 0 | 0 | 0 | 0 | 0 | 0,9 | 0,058 | 0,901 |
| HTR1F | NPY2R | 9606.ENSP00000322924 | 9606.ENSP00000332591 | 0 | 0 | 0 | 0,6 | 0,062 | 0 | 0,9 | 0 | 0,902 |
| HTR1F | OPRL1 | 9606.ENSP00000322924 | 9606.ENSP00000336764 | 0 | 0 | 0 | 0,588 | 0,062 | 0 | 0,9 | 0,169 | 0,907 |
| HTR1F | OXER1 | 9606.ENSP00000322924 | 9606.ENSP00000367930 | 0 | 0 | 0 | 0 | 0 | 0 | 0,9 | 0,164 | 0,912 |
| HTR2C | NPSR1 | 9606.ENSP00000276198 | 9606.ENSP00000370950 | 0 | 0 | 0 | 0,577 | 0,062 | 0 | 0,9 | 0,164 | 0,907 |
| HTR2C | MCHR2 | 9606.ENSP00000276198 | 9606.ENSP00000281806 | 0 | 0 | 0 | 0,582 | 0,085 | 0 | 0,9 | 0,192 | 0,91 |
| HTR6 | RAMP2 | 9606.ENSP00000289753 | 9606.ENSP00000253796 | 0 | 0 | 0 | 0 | 0 | 0 | 0,9 | 0 | 0,9 |
| HTR6 | NPSR1 | 9606.ENSP00000289753 | 9606.ENSP00000370950 | 0 | 0 | 0 | 0 | 0,061 | 0 | 0,9 | 0 | 0,902 |
| HTR6 | MC5R | 9606.ENSP00000289753 | 9606.ENSP00000318077 | 0 | 0 | 0 | 0,597 | 0 | 0 | 0,9 | 0,138 | 0,904 |
| HTR6 | LHCGR | 9606.ENSP00000289753 | 9606.ENSP00000294954 | 0 | 0 | 0 | 0 | 0,044 | 0 | 0,9 | 0,1 | 0,906 |
| KCNE1L | KCNH2 | 9606.ENSP00000361173 | 9606.ENSP00000262186 | 0 | 0 | 0 | 0 | 0,061 | 0 | 0,9 | 0,625 | 0,961 |
| KCNE3 | KCNH2 | 9606.ENSP00000310557 | 9606.ENSP00000262186 | 0 | 0 | 0 | 0 | 0 | 0 | 0,9 | 0,685 | 0,967 |
| KIR3DL1 | KLRC1 | 9606.ENSP00000375608 | 9606.ENSP00000438038 | 0 | 0 | 0 | 0 | 0,072 | 0 | 0,8 | 0,843 | 0,968 |
| KLRC1 | KLRC4 | 9606.ENSP00000438038 | 9606.ENSP00000310216 | 0 | 0 | 0 | 0,956 | 0,137 | 0 | 0,9 | 0,745 | 0,912 |
| LAIR1 | PLAUR | 9606.ENSP00000375622 | 9606.ENSP00000339328 | 0 | 0 | 0 | 0 | 0 | 0 | 0,9 | 0,049 | 0,9 |
| LAIR1 | TARM1 | 9606.ENSP00000375622 | 9606.ENSP00000439454 | 0 | 0 | 0 | 0,616 | 0,061 | 0 | 0,9 | 0 | 0,902 |
| LHCGR | RAMP2 | 9606.ENSP00000294954 | 9606.ENSP00000253796 | 0 | 0 | 0 | 0 | 0 | 0 | 0,9 | 0,047 | 0,9 |
| LHCGR | MC5R | 9606.ENSP00000294954 | 9606.ENSP00000318077 | 0 | 0 | 0 | 0 | 0,044 | 0 | 0,9 | 0,071 | 0,903 |
| LHCGR | NPSR1 | 9606.ENSP00000294954 | 9606.ENSP00000370950 | 0 | 0 | 0 | 0 | 0,044 | 0 | 0,9 | 0,178 | 0,914 |
| LRRTM2 | NRXN1 | 9606.ENSP00000274711 | 9606.ENSP00000385142 | 0 | 0 | 0 | 0 | 0,16 | 0 | 0,9 | 0,84 | 0,985 |
| LRRTM3 | NRXN1 | 9606.ENSP00000355187 | 9606.ENSP00000385142 | 0 | 0 | 0 | 0 | 0,3 | 0 | 0,9 | 0,673 | 0,975 |
| MC5R | RAMP2 | 9606.ENSP00000318077 | 9606.ENSP00000253796 | 0 | 0 | 0 | 0 | 0 | 0 | 0,9 | 0,056 | 0,901 |
| MC5R | NPSR1 | 9606.ENSP00000318077 | 9606.ENSP00000370950 | 0 | 0 | 0 | 0 | 0 | 0 | 0,9 | 0,256 | 0,922 |
| MCHR2 | TAS2R31 | 9606.ENSP00000281806 | 9606.ENSP00000375093 | 0 | 0 | 0 | 0 | 0 | 0 | 0,9 | 0 | 0,9 |
| MCHR2 | PTGER3 | 9606.ENSP00000281806 | 9606.ENSP00000349003 | 0 | 0 | 0 | 0 | 0 | 0 | 0,9 | 0 | 0,9 |
| MCHR2 | TAS2R41 | 9606.ENSP00000281806 | 9606.ENSP00000386201 | 0 | 0 | 0 | 0 | 0 | 0 | 0,9 | 0 | 0,9 |
| MCHR2 | TAS2R60 | 9606.ENSP00000281806 | 9606.ENSP00000327724 | 0 | 0 | 0 | 0 | 0 | 0 | 0,9 | 0 | 0,9 |
| MCHR2 | TAS2R38 | 9606.ENSP00000281806 | 9606.ENSP00000448219 | 0 | 0 | 0 | 0 | 0 | 0 | 0,9 | 0,056 | 0,901 |
| MCHR2 | OPRL1 | 9606.ENSP00000281806 | 9606.ENSP00000336764 | 0 | 0 | 0 | 0,723 | 0 | 0 | 0,9 | 0,161 | 0,903 |
| MCHR2 | NPSR1 | 9606.ENSP00000281806 | 9606.ENSP00000370950 | 0 | 0 | 0 | 0,608 | 0 | 0 | 0,9 | 0,17 | 0,905 |
| MCHR2 | NPY2R | 9606.ENSP00000281806 | 9606.ENSP00000332591 | 0 | 0 | 0 | 0,628 | 0,07 | 0 | 0,9 | 0,198 | 0,908 |
| MCHR2 | OXER1 | 9606.ENSP00000281806 | 9606.ENSP00000367930 | 0 | 0 | 0 | 0,601 | 0 | 0 | 0,9 | 0,368 | 0,913 |
| MUC17 | MUC3A | 9606.ENSP00000302716 | 9606.ENSP00000368771 | 0 | 0 | 0 | 0,552 | 0,139 | 0 | 0,9 | 0,789 | 0,941 |
| NPSR1 | RAMP2 | 9606.ENSP00000370950 | 9606.ENSP00000253796 | 0 | 0 | 0 | 0 | 0 | 0 | 0,9 | 0,041 | 0,9 |
| NPY2R | TAS2R60 | 9606.ENSP00000332591 | 9606.ENSP00000327724 | 0 | 0 | 0 | 0 | 0 | 0 | 0,9 | 0 | 0,9 |
| NPY2R | TAS2R31 | 9606.ENSP00000332591 | 9606.ENSP00000375093 | 0 | 0 | 0 | 0 | 0 | 0 | 0,9 | 0 | 0,9 |
| NPY2R | PTGER3 | 9606.ENSP00000332591 | 9606.ENSP00000349003 | 0 | 0 | 0 | 0 | 0 | 0 | 0,9 | 0 | 0,9 |
| NPY2R | OXER1 | 9606.ENSP00000332591 | 9606.ENSP00000367930 | 0 | 0 | 0 | 0,576 | 0 | 0 | 0,9 | 0,126 | 0,903 |
| NPY2R | TAS2R41 | 9606.ENSP00000332591 | 9606.ENSP00000386201 | 0 | 0 | 0 | 0 | 0 | 0 | 0,9 | 0,103 | 0,906 |
| NPY2R | TAS2R38 | 9606.ENSP00000332591 | 9606.ENSP00000448219 | 0 | 0 | 0 | 0 | 0 | 0 | 0,9 | 0,103 | 0,906 |
| NPY2R | OPRL1 | 9606.ENSP00000332591 | 9606.ENSP00000336764 | 0 | 0 | 0 | 0,673 | 0,077 | 0 | 0,9 | 0,421 | 0,916 |
| OPRL1 | TAS2R60 | 9606.ENSP00000336764 | 9606.ENSP00000327724 | 0 | 0 | 0 | 0 | 0 | 0 | 0,9 | 0 | 0,9 |
| OPRL1 | TAS2R31 | 9606.ENSP00000336764 | 9606.ENSP00000375093 | 0 | 0 | 0 | 0 | 0 | 0 | 0,9 | 0 | 0,9 |
| OPRL1 | TAS2R41 | 9606.ENSP00000336764 | 9606.ENSP00000386201 | 0 | 0 | 0 | 0 | 0 | 0 | 0,9 | 0 | 0,9 |
| OPRL1 | TAS2R38 | 9606.ENSP00000336764 | 9606.ENSP00000448219 | 0 | 0 | 0 | 0 | 0 | 0 | 0,9 | 0 | 0,9 |
| OPRL1 | OXER1 | 9606.ENSP00000336764 | 9606.ENSP00000367930 | 0 | 0 | 0 | 0,604 | 0,076 | 0 | 0,9 | 0,069 | 0,904 |
| OPRL1 | PTGER3 | 9606.ENSP00000336764 | 9606.ENSP00000349003 | 0 | 0 | 0 | 0 | 0 | 0 | 0,9 | 0,232 | 0,919 |
| OXER1 | TAS2R60 | 9606.ENSP00000367930 | 9606.ENSP00000327724 | 0 | 0 | 0 | 0 | 0 | 0 | 0,9 | 0 | 0,9 |
| OXER1 | PTGER3 | 9606.ENSP00000367930 | 9606.ENSP00000349003 | 0 | 0 | 0 | 0 | 0 | 0 | 0,9 | 0,065 | 0,902 |
| OXER1 | TAS2R31 | 9606.ENSP00000367930 | 9606.ENSP00000375093 | 0 | 0 | 0 | 0 | 0 | 0 | 0,9 | 0 | 0,9 |
| OXER1 | TAS2R41 | 9606.ENSP00000367930 | 9606.ENSP00000386201 | 0 | 0 | 0 | 0 | 0 | 0 | 0,9 | 0 | 0,9 |
| OXER1 | TAS2R38 | 9606.ENSP00000367930 | 9606.ENSP00000448219 | 0 | 0 | 0 | 0 | 0 | 0 | 0,9 | 0 | 0,9 |
| P2RX5 | P2RX7 | 9606.ENSP00000225328 | 9606.ENSP00000330696 | 0 | 0 | 0 | 0,815 | 0,063 | 0 | 0,9 | 0,819 | 0,916 |
| PLAUR | TARM1 | 9606.ENSP00000339328 | 9606.ENSP00000439454 | 0 | 0 | 0 | 0 | 0 | 0 | 0,9 | 0 | 0,9 |
| PTGER3 | TAS2R60 | 9606.ENSP00000349003 | 9606.ENSP00000327724 | 0 | 0 | 0 | 0 | 0 | 0 | 0,9 | 0,049 | 0,9 |
| PTGER3 | TAS2R31 | 9606.ENSP00000349003 | 9606.ENSP00000375093 | 0 | 0 | 0 | 0 | 0 | 0 | 0,9 | 0 | 0,9 |
| PTGER3 | TAS2R38 | 9606.ENSP00000349003 | 9606.ENSP00000448219 | 0 | 0 | 0 | 0 | 0 | 0 | 0,9 | 0,042 | 0,9 |
| PTGER3 | TAS2R41 | 9606.ENSP00000349003 | 9606.ENSP00000386201 | 0 | 0 | 0 | 0 | 0 | 0 | 0,9 | 0,145 | 0,91 |
| SCN11A | SCN2B | 9606.ENSP00000307599 | 9606.ENSP00000278947 | 0 | 0 | 0 | 0 | 0 | 0 | 0,9 | 0,428 | 0,94 |
| SCN11A | SCN1B | 9606.ENSP00000307599 | 9606.ENSP00000396915 | 0 | 0 | 0 | 0 | 0 | 0 | 0,9 | 0,558 | 0,953 |
| SCN1B | SCN2B | 9606.ENSP00000396915 | 9606.ENSP00000278947 | 0 | 0 | 0 | 0 | 0,14 | 0 | 0,72 | 0,887 | 0,97 |
| SCN1B | SCN9A | 9606.ENSP00000396915 | 9606.ENSP00000386306 | 0 | 0 | 0 | 0 | 0 | 0 | 0,9 | 0,757 | 0,974 |
| SCN2B | SCN9A | 9606.ENSP00000278947 | 9606.ENSP00000386306 | 0 | 0 | 0 | 0 | 0,062 | 0 | 0,9 | 0,59 | 0,958 |
| TAS2R31 | TAS2R60 | 9606.ENSP00000375093 | 9606.ENSP00000327724 | 0 | 0 | 0 | 0,653 | 0 | 0 | 0,9 | 0,6 | 0,92 |
| TAS2R31 | TAS2R41 | 9606.ENSP00000375093 | 9606.ENSP00000386201 | 0 | 0 | 0 | 0,739 | 0 | 0 | 0,9 | 0,587 | 0,914 |
| TAS2R31 | TAS2R38 | 9606.ENSP00000375093 | 9606.ENSP00000448219 | 0 | 0 | 0 | 0,688 | 0,061 | 0 | 0,9 | 0,8 | 0,926 |
| TAS2R38 | TAS2R60 | 9606.ENSP00000448219 | 9606.ENSP00000327724 | 0 | 0 | 0 | 0,619 | 0,049 | 0 | 0,9 | 0,656 | 0,925 |
| TAS2R38 | TAS2R41 | 9606.ENSP00000448219 | 9606.ENSP00000386201 | 0 | 0 | 0 | 0,693 | 0,061 | 0 | 0,9 | 0,601 | 0,919 |
| TAS2R41 | TAS2R60 | 9606.ENSP00000386201 | 9606.ENSP00000327724 | 0 | 0 | 0 | 0,892 | 0,141 | 0 | 0,9 | 0,736 | 0,917 |
