## Supplementary material for "Early Reduction of SARS-CoV-2 Replication in Bronchial Epithelium by Kinin B_2_ Receptor Antagonism": Supl. Table 18

**Supplemental Table S18.** Cellular process enrichment analysis output of String network analysis of membrane-bound receptor DEGs comparing SARS-CoV-2 + B2R antagonist versus SARS-CoV-2

| #term ID | term description | observed | backgrou | streng | false discove | matching proteins in your network (labels) |
|---|---|---|---|---|---|---|
| GO:0007186 | G protein-coupled r | 111 | 1247 | 0,76 | 6,77E-49 | GPR63,OR7C1,RAMP2,CHRM3,GPR133,SORCS1,GABRA6,GABRB2,HTR2C,MCHR2,GPR15,HTR6,LHCGR,F2RL2,OR4K2,OR10A5,GALR1,OR51B5,C3AR1,OR10V1,OR52K1,OR9I1,OR1B1,OR7G2,OR8U1,OR6Y1,GPR151,GPRC6A,OR10H5,OR2Y1,EDNRA,HTR1A,OR7A5,MC5R,GPR62,OR4K13,OR4K5,GHRHR,OR4S1,OR6B3,OR5AK2,OR52N5,HTR1F,OR8H1,OR8K5,OR10G4,OR52D1,TAS2R60,OR2T10,OR1G1,NPY2R,OR6P1,GRK1,OR5D18,OPRL1,OR13H1,OR2AG2,ADRB3,OR6N2,OR5H2,OR2T6,OR52R1,PTGER3,CCRL2,OR8D1,OR5AC2,OR6K2,OR2M2,OR5B21,OR2T5,OR2T1,OR6K3,OR10R2,HTR1B,AGTR2,OR1N2,CNR2,OR13C5,OR10G9,OR4D1,GABBR1,OR5V1,OR2J2,OXER1,OR6C65,OR6C75,OR10A4,OR51B4,NPSR1,OR5K1,TAS2R31,OR6C4,GRM2,OR2A14,OR1C1,OR2A25,TAS2R41,CALCRL,OR5K2,CXCR6,OR5H14,ESR1,CASR,GABRA2,TRPM1,OR13C2,GPR19,TAS2R38,OR1D5,OR8G1,GPR1 |
| GO:0003008 | system process | 126 | 1827 | 0,65 | 1,05E-45 | CLIC5,AQP9,P2RX5,KCNJ8,OR7C1,KCNA5,RAMP2,CHRM3,KCNH2,HPN,KCNQ4,CACNA1C,GABRB2,SCN2B,CHRNB3,LHCGR,OR4K2,OR10A5,OR51B5,C3AR1,OR10V1,OR52K1,OR9I1,OR1B1,OR7G2,OR8U1,SLC26A5,OR6Y1,SCN11A,CD34,OR10H5,OR2Y1,KCNK10,EDNRA,HTR1A,OR7A5,OR4K13,KCNMB3,OR4K5,CACNB2,OR4S1,OR6B3,OR5AK2,OR52N5,SLC2A2,OR8H1,OR8K5,OR10G4,OR52D1,TAS2R60,CNGA2,OR2T10,CALHM1,GRID1,P2RX7,OR1G1,IGDCC3,OR6P1,GRK1,OR5D18,OPRL1,CLCN1,OR13H1,OR2AG2,ADRB3,OR6N2,OR5H2,OR2T6,OR52R1,PTGER3,OR8D1,OR5AC2,KCNK5,OR6K2,OR2M2,OR5B21,GRIN3A,OR2T5,OR2T1,OR6K3,OR10R2,CHRNB2,HTR1B,CACNA1B,GRIN1,AGTR2,KCNE1L,OR1N2,MUSK,CNR2,OR13C5,OR10G9,OR4D1,OR5V1,OR2J2,SLC22A6,SCNN1D,OR6C65,OR6C75,OR10A4,OR51B4,OR5K1,ACVRL1,TAS2R31,OR6C4,GRIN2A,SLITRK6,CACNA1I,NRXN1,OR2A14,OR1C1,OR2A25,TAS2R41,SCN9A,SLC12A5,OR5K2,SCN1B,OR5H14,CASR,TRPM1,OR13C2,TAS2R38,OR1D5,OR8G1,GRIN2B,GRIA3 |
| GO:0023052 | signaling | 201 | 5108 | 0,41 | 5,23E-45 | RIPK3,P2RX5,CD69,GPR63,PCDHB6,TREM1,OR7C1,KCNA5,RAMP2,CHRM3,DCLK1,SIT1,GPR133,KCNH2,HPN,SORCS1,IL1RL2,CACNA1C,GABRA6,GABRB2,HTR2C,SLC1A2,MS4A2,SCN2B,CD226,MCHR2,GPR15,HTR6,CHRNB3,LILRA4,FGFR4,LHCGR,F2RL2,CLEC1B,OR4K2,OR10A5,GALR1,CD3D,OR51B5,C3AR1,OR10V1,OR52K1,OR9I1,MUC17,OR1B1,OR7G2,OR8U1,OR6Y1,SLAMF1,SCN11A,GPR151,GPRC6A,CD34,KCNE3,OR10H5,OR2Y1,CSPG4,KCNK10,EDNRA,HTR1A,OR7A5,MC5R,GPR62,OR4K13,KCNMB3,OR4K5,CACNB2,GHRHR,OR4S1,OR6B3,OR5AK2,OR52N5,HTR1F,OR8H1,OR8K5,OR10G4,OR52D1,TAS2R60,OR2T10,GRID1,RTN4RL1,P2RX7,OR1G1,NPY2R,OR6P1,GRK1,OR5D18,OPRL1,PLAUR,KIR2DL4,CLCN1,OR13H1,NEO1,NCR3,ERBB4,OR2AG2,ADRB3,OR6N2,OR5H2,RET,TNFRSF14,OR2T6,OR52R1,PTGER3,CCRL2,OR8D1,OR5AC2,OR6K2,OR2M2,RXRG,OR5B21,NTRK3,GRIN3A,LRRTM3,OR2T5,OR2T1,ESRRG,SLC22A2,GPA33,DDR2,FCGR3A,CD244,OR6K3,OR10R2,CHRNB2,ROS1,FCGR1B,CD2,HTR1B,LPPR4,CACNA1B,GRIN1,KCNB1,AGTR2,KCNE1L,IRS4,EPHA10,OR1N2,MUSK,CNR2,OR13C5,OR10G9,OR4D1,GABBR1,OR5V1,OR2J2,OXER1,TNFRSF4,MUC3A,OR6C65,OR6C75,OR10A4,OR51B4,NPSR1,SIRPB1,OR5K1,ACVRL1,EPHA6,TAS2R31,KIR3DL1,OR6C4,GRM2,GRIN2A,CSF2RB,CACNA1I,NRXN1,OR2A14,OR1C1,OR2A25,TAS2R41,SCN9A,CD8A,CALCRL,SLC12A5,OR5K2,CXCR6,SCN1B,OR5H14,ESR1,PTPRC,HFE,CASR,GABRA2,ESRRB,MPZ,PCDHA5,TRPM1,KLRC1,OR13C2,GPR19,TAS2R38,KCNC2,MRC1,OR1D5,ZP2,OR8G1,GRIN2B,GPR1,ANTXRL,GRIA3,PRLR |
| GO:0007154 | cell communication | 200 | 5219 | 0,39 | 6,64E-43 | RIPK3,P2RX5,CD69,GPR63,PCDHB6,TREM1,OR7C1,KCNA5,RAMP2,CHRM3,DCLK1,SIT1,GPR133,KCNH2,HPN,SORCS1,IL1RL2,CACNA1C,GABRA6,GABRB2,HTR2C,SLC1A2,MS4A2,SCN2B,CD226,MCHR2,GPR15,HTR6,CHRNB3,LILRA4,FGFR4,LHCGR,F2RL2,CLEC1B,OR4K2,OR10A5,GALR1,CD3D,OR51B5,C3AR1,OR10V1,OR52K1,OR9I1,MUC17,OR1B1,OR7G2,OR8U1,OR6Y1,SLAMF1,SCN11A,GPR151,GPRC6A,CD34,OR10H5,OR2Y1,CSPG4,KCNK10,EDNRA,HTR1A,OR7A5,MC5R,GPR62,OR4K13,KCNMB3,OR4K5,CACNB2,GHRHR,OR4S1,OR6B3,OR5AK2,OR52N5,HTR1F,OR8H1,OR8K5,OR10G4,OR52D1,TAS2R60,OR2T10,GRID1,RTN4RL1,P2RX7,OR1G1,NPY2R,OR6P1,GRK1,OR5D18,OPRL1,PLAUR,KIR2DL4,CLCN1,OR13H1,NEO1,NCR3,ERBB4,OR2AG2,ADRB3,OR6N2,OR5H2,RET,TNFRSF14,OR2T6,OR52R1,PTGER3,CCRL2,OR8D1,OR5AC2,OR6K2,OR2M2,RXRG,OR5B21,NTRK3,GRIN3A,LRRTM3,OR2T5,OR2T1,ESRRG,SLC22A2,GPA33,DDR2,FCGR3A,CD244,OR6K3,OR10R2,CHRNB2,ROS1,FCGR1B,CD2,HTR1B,LPPR4,CACNA1B,GRIN1,KCNB1,AGTR2,KCNE1L,IRS4,EPHA10,OR1N2,MUSK,CNR2,OR13C5,OR10G9,OR4D1,GABBR1,OR5V1,OR2J2,OXER1,TNFRSF4,MUC3A,OR6C65,OR6C75,OR10A4,OR51B4,NPSR1,SIRPB1,OR5K1,ACVRL1,EPHA6,TAS2R31,KIR3DL1,OR6C4,GRM2,GRIN2A,CSF2RB,CACNA1I,NRXN1,OR2A14,OR1C1,OR2A25,TAS2R41,SCN9A,CD8A,CALCRL,SLC12A5,OR5K2,CXCR6,SCN1B,OR5H14,ESR1,PTPRC,HFE,CASR,GABRA2,ESRRB,MPZ,PCDHA5,TRPM1,KLRC1,OR13C2,GPR19,TAS2R38,KCNC2,MRC1,OR1D5,ZP2,OR8G1,GRIN2B,GPR1,ANTXRL,GRIA3,PRLR |
| GO:0050877 | nervous system pro | 104 | 1271 | 0,72 | 1,01E-42 | CLIC5,P2RX5,OR7C1,HPN,KCNQ4,GABRB2,CHRNB3,LHCGR,OR4K2,OR10A5,OR51B5,OR10V1,OR52K1,OR9I1,OR1B1,OR7G2,OR8U1,SLC26A5,OR6Y1,SCN11A,OR10H5,OR2Y1,KCNK10,OR7A5,OR4K13,KCNMB3,OR4K5,CACNB2,OR4S1,OR6B3,OR5AK2,OR52N5,OR8H1,OR8K5,OR10G4,OR52D1,TAS2R60,CNGA2,OR2T10,CALHM1,GRID1,P2RX7,OR1G1,IGDCC3,OR6P1,GRK1,OR5D18,OPRL1,CLCN1,OR13H1,OR2AG2,OR6N2,OR5H2,OR2T6,OR52R1,OR8D1,OR5AC2,OR6K2,OR2M2,OR5B21,GRIN3A,OR2T5,OR2T1,OR6K3,OR10R2,CHRNB2,GRIN1,AGTR2,OR1N2,MUSK,CNR2,OR13C5,OR10G9,OR4D1,OR5V1,OR2J2,SCNN1D,OR6C65,OR6C75,OR10A4,OR51B4,OR5K1,TAS2R31,OR6C4,GRIN2A,SLITRK6,CACNA1I,NRXN1,OR2A14,OR1C1,OR2A25,TAS2R41,SCN9A,SLC12A5,OR5K2,SCN1B,OR5H14,TRPM1,OR13C2,TAS2R38,OR1D5,OR8G1,GRIN2B,GRIA3 |
| GO:0007606 | sensory perception | 70 | 487 | 0,97 | 2,54E-41 | OR7C1,OR4K2,OR10A5,OR51B5,OR10V1,OR52K1,OR9I1,OR1B1,OR7G2,OR8U1,OR6Y1,OR10H5,OR2Y1,OR7A5,OR4K13,OR4K5,OR4S1,OR6B3,OR5AK2,OR52N5,OR8H1,OR8K5,OR10G4,OR52D1,TAS2R60,CNGA2,OR2T10,CALHM1,OR1G1,OR6P1,OR5D18,OR13H1,OR2AG2,OR6N2,OR5H2,OR2T6,OR52R1,OR8D1,OR5AC2,OR6K2,OR2M2,OR5B21,OR2T5,OR2T1,OR6K3,OR10R2,OR1N2,OR13C5,OR10G9,OR4D1,OR5V1,OR2J2,SCNN1D,OR6C65,OR6C75,OR10A4,OR51B4,OR5K1,TAS2R31,OR6C4,OR2A14,OR1C1,OR2A25,TAS2R41,OR5K2,OR5H14,OR13C2,TAS2R38,OR1D5,OR8G1 |
| GO:0050911 | detection of chemica | 63 | 385 | 1,03 | 7,18E-40 | OR7C1,OR4K2,OR10A5,OR51B5,OR10V1,OR52K1,OR9I1,OR1B1,OR7G2,OR8U1,OR6Y1,OR10H5,OR2Y1,OR7A5,OR4K13,OR4K5,OR4S1,OR6B3,OR5AK2,OR52N5,OR8H1,OR8K5,OR10G4,OR52D1,OR2T10,OR1G1,OR6P1,OR5D18,OR13H1,OR2AG2,OR6N2,OR5H2,OR2T6,OR52R1,OR8D1,OR5AC2,OR6K2,OR2M2,OR5B21,OR2T5,OR2T1,OR6K3,OR10R2,OR1N2,OR13C5,OR10G9,OR4D1,OR5V1,OR2J2,OR6C65,OR6C75,OR10A4,OR51B4,OR5K1,OR6C4,OR2A14,OR1C1,OR2A25,OR5K2,OR5H14,OR13C2,OR1D5,OR8G1 |
| GO:0007608 | sensory perception | 64 | 413 | 1 | 2,36E-39 | OR7C1,OR4K2,OR10A5,OR51B5,OR10V1,OR52K1,OR9I1,OR1B1,OR7G2,OR8U1,OR6Y1,OR10H5,OR2Y1,OR7A5,OR4K13,OR4K5,OR4S1,OR6B3,OR5AK2,OR52N5,OR8H1,OR8K5,OR10G4,OR52D1,CNGA2,OR2T10,OR1G1,OR6P1,OR5D18,OR13H1,OR2AG2,OR6N2,OR5H2,OR2T6,OR52R1,OR8D1,OR5AC2,OR6K2,OR2M2,OR5B21,OR2T5,OR2T1,OR6K3,OR10R2,OR1N2,OR13C5,OR10G9,OR4D1,OR5V1,OR2J2,OR6C65,OR6C75,OR10A4,OR51B4,OR5K1,OR6C4,OR2A14,OR1C1,OR2A25,OR5K2,OR5H14,OR13C2,OR1D5,OR8G1 |
| GO:0009593 | detection of chemica | 67 | 469 | 0,97 | 2,36E-39 | OR7C1,OR4K2,OR10A5,OR51B5,OR10V1,OR52K1,OR9I1,OR1B1,OR7G2,OR8U1,OR6Y1,OR10H5,OR2Y1,OR7A5,OR4K13,KCNMB3,OR4K5,OR4S1,OR6B3,OR5AK2,OR52N5,OR8H1,OR8K5,OR10G4,OR52D1,OR2T10,OR1G1,OR6P1,OR5D18,OR13H1,OR2AG2,OR6N2,OR5H2,OR2T6,OR52R1,OR8D1,OR5AC2,OR6K2,OR2M2,OR5B21,OR2T5,OR2T1,OR6K3,OR10R2,OR1N2,OR13C5,OR10G9,OR4D1,OR5V1,OR2J2,OR6C65,OR6C75,OR10A4,OR51B4,OR5K1,TAS2R31,OR6C4,OR2A14,OR1C1,OR2A25,OR5K2,OR5H14,CASR,OR13C2,TAS2R38,OR1D5,OR8G1 |
| GO:0050907 | detection of chemica | 65 | 431 | 0,99 | 2,36E-39 | OR7C1,OR4K2,OR10A5,OR51B5,OR10V1,OR52K1,OR9I1,OR1B1,OR7G2,OR8U1,OR6Y1,OR10H5,OR2Y1,OR7A5,OR4K13,OR4K5,OR4S1,OR6B3,OR5AK2,OR52N5,OR8H1,OR8K5,OR10G4,OR52D1,OR2T10,OR1G1,OR6P1,OR5D18,OR13H1,OR2AG2,OR6N2,OR5H2,OR2T6,OR52R1,OR8D1,OR5AC2,OR6K2,OR2M2,OR5B21,OR2T5,OR2T1,OR6K3,OR10R2,OR1N2,OR13C5,OR10G9,OR4D1,OR5V1,OR2J2,OR6C65,OR6C75,OR10A4,OR51B4,OR5K1,TAS2R31,OR6C4,OR2A14,OR1C1,OR2A25,OR5K2,OR5H14,OR13C2,TAS2R38,OR1D5,OR8G1 |

| | | | | | | |
|---|---|---|---|---|---|---|
| GO:0007165 | signal transduction | 185 | 4738 | 0,4 | 2,93E-39 | RIPK3,P2RX5,CD69,GPR63,TREM1,OR7C1,KCNA5,RAMP2,CHRM3,DCLK1,SIT1,GPR133,KCNH2,HPN,SORCS1,IL1RL2,CACNA1C,GABRA6,GABRB2,HTR2C,MS4A2,CD226,MCHR2,GPR15,HTR6,CHRNB3,LILRA4,FGFR4,LHCGR,F2RL2,CLEC1B,OR4K2,OR10A5,GALR1,CD3D,OR51B5,C3AR1,OR10V1,OR52K1,OR9I1,MUC17,OR1B1,OR7G2,OR8U1,OR6Y1,SLAMF1,GPR151,GPRC6A,CD34,OR10H5,OR2Y1,CSPG4,KCNK10,EDNRA,HTR1A,OR7A5,MC5R,GPR62,OR4K13,OR4K5,GHRHR,OR4S1,OR6B3,OR5AK2,OR52N5,HTR1F,OR8H1,OR8K5,OR10G4,OR52D1,TAS2R60,OR2T10,GRID1,RTN4RL1,P2RX7,OR1G1,NPY2R,OR6P1,GRK1,OR5D18,OPRL1,PLAUR,KIR2DL4,OR13H1,NEO1,NCR3,ERBB4,OR2AG2,ADRB3,OR6N2,OR5H2,RET,TNFRSF14,OR2T6,OR52R1,PTGER3,CCRL2,OR8D1,OR5AC2,OR6K2,OR2M2,RXRG,OR5B21,NTRK3,GRIN3A,LRRTM3,OR2T5,OR2T1,ESRRG,GPA33,DDR2,FCGR3A,CD244,OR6K3,OR10R2,CHRNB2,ROS1,FCGR1B,CD2,HTR1B,LPPR4,GRIN1,KCNB1,AGTR2,IRS4,EPHA10,OR1N2,MUSK,CNR2,OR13C5,OR10G9,OR4D1,GABBR1,OR5V1,OR2J2,OXER1,TNFRSF4,MUC3A,OR6C65,OR6C75,OR10A4,OR51B4,NPSR1,SIRPB1,OR5K1,ACVRL1,EPHA6,TAS2R31,KIR3DL1,OR6C4,GRM2,GRIN2A,CSF2RB,CACNA1I,NRXN1,OR2A14,OR1C1,OR2A25,TAS2R41,CD8A,CALCRL,OR5K2,CXCR6,OR5H14,ESR1,PTPRC,HFE,CASR,GABRA2,ESRRB,TRPM1,KLRC1,OR13C2,GPR19,TAS2R38,KCNC2,MRC1,OR1D5,ZP2,OR8G1,GRIN2B,GPR1,ANTXRL,GRIA3,PRLR |
| GO:0007600 | sensory perception | 86 | 901 | 0,79 | 2,93E-39 | CLIC5,OR7C1,HPN,KCNQ4,GABRB2,OR4K2,OR10A5,OR51B5,OR10V1,OR52K1,OR9I1,OR1B1,OR7G2,OR8U1,SLC26A5,OR6Y1,OR10H5,OR2Y1,OR7A5,OR4K13,OR4K5,CACNB2,OR4S1,OR6B3,OR5AK2,OR52N5,OR8H1,OR8K5,OR10G4,OR52D1,TAS2R60,CNGA2,OR2T10,CALHM1,P2RX7,OR1G1,OR6P1,GRK1,OR5D18,OPRL1,OR13H1,OR2AG2,OR6N2,OR5H2,OR2T6,OR52R1,OR8D1,OR5AC2,OR6K2,OR2M2,OR5B21,OR2T5,OR2T1,OR6K3,OR10R2,CHRNB2,GRIN1,OR1N2,CNR2,OR13C5,OR10G9,OR4D1,OR5V1,OR2J2,SCNN1D,OR6C65,OR6C75,OR10A4,OR51B4,OR5K1,TAS2R31,OR6C4,GRIN2A,SLITRK6,OR2A14,OR1C1,OR2A25,TAS2R41,SCN9A,OR5K2,OR5H14,TRPM1,OR13C2,TAS2R38,OR1D5,OR8G1 |
| GO:0050906 | detection of stimulu | 66 | 484 | 0,95 | 8,90E-38 | OR7C1,HPN,OR4K2,OR10A5,OR51B5,OR10V1,OR52K1,OR9I1,OR1B1,OR7G2,OR8U1,OR6Y1,OR10H5,OR2Y1,OR7A5,OR4K13,OR4K5,OR4S1,OR6B3,OR5AK2,OR52N5,OR8H1,OR8K5,OR10G4,OR52D1,OR2T10,OR1G1,OR6P1,OR5D18,OR13H1,OR2AG2,OR6N2,OR5H2,OR2T6,OR52R1,OR8D1,OR5AC2,OR6K2,OR2M2,OR5B21,OR2T5,OR2T1,OR6K3,OR10R2,OR1N2,OR13C5,OR10G9,OR4D1,OR5V1,OR2J2,OR6C65,OR6C75,OR10A4,OR51B4,OR5K1,TAS2R31,OR6C4,OR2A14,OR1C1,OR2A25,OR5K2,OR5H14,OR13C2,TAS2R38,OR1D5,OR8G1 |
| GO:0051606 | detection of stimulu | 69 | 638 | 0,85 | 7,81E-34 | OR7C1,HPN,OR4K2,OR10A5,OR51B5,OR10V1,OR52K1,OR9I1,OR1B1,OR7G2,OR8U1,OR6Y1,OR10H5,OR2Y1,OR7A5,OR4K13,KCNMB3,OR4K5,OR4S1,OR6B3,OR5AK2,OR52N5,OR8H1,OR8K5,OR10G4,OR52D1,OR2T10,OR1G1,OR6P1,GRK1,OR5D18,OR13H1,OR2AG2,OR6N2,OR5H2,OR2T6,OR52R1,OR8D1,OR5AC2,OR6K2,OR2M2,OR5B21,OR2T5,OR2T1,OR6K3,OR10R2,OR1N2,OR13C5,OR10G9,OR4D1,OR5V1,OR2J2,OR6C65,OR6C75,OR10A4,OR51B4,OR5K1,TAS2R31,OR6C4,OR2A14,OR1C1,OR2A25,OR5K2,OR5H14,CASR,OR13C2,TAS2R38,OR1D5,OR8G1 |
| GO:0050896 | response to stimulu | 227 | 7824 | 0,27 | 1,55E-32 | RIPK3,AQP8,AQP9,P2RX5,CD69,GPR63,SLC25A2,KCNJ8,TREM1,OR7C1,KCNA5,RAMP2,CHRM3,DCLK1,SIT1,GPR133,KCNH2,HPN,SORCS1,SLC17A6,IL1RL2,CACNA1C,SLC47A1,FCRL4,GABRA6,GABRB2,HTR2C,SLC1A2,MS4A2,SCN2B,CD226,MCHR2,GPR15,HTR6,CHRNB3,LILRA4,FGFR4,LHCGR,F2RL2,CLEC1B,OR4K2,OR10A5,GALR1,CD3D,OR51B5,C3AR1,OR10V1,OR52K1,OR9I1,MUC17,SLC23A1,OR1B1,OR7G2,OR8U1,SLC26A5,OR6Y1,SLAMF1,SCN11A,GPR151,GPRC6A,CD34,KLRC4,OR10H5,OR2Y1,CSPG4,KCNK10,EDNRA,HTR1A,OR7A5,MC5R,GPR62,OR4K13,KCNMB3,OR4K5,GHRHR,SLC34A1,OR4S1,OR6B3,OR5AK2,OR52N5,HTR1F,OR8H1,OR8K5,TMEM108,OR10G4,OR52D1,TAS2R60,CNGA2,OR2T10,CALHM1,GRID1,RTN4RL1,P2RX7,OR1G1,NPY2R,OR6P1,GRK1,OR5D18,OPRL1,SLC22A8,PLAUR,KIR2DL4,OR13H1,NEO1,NCR3,ERBB4,TREML4,OR2AG2,ADRB3,OR6N2,SCARA5,OR5H2,RET,TNFRSF14,OR2T6,OR52R1,PTGER3,CCRL2,OR8D1,OR5AC2,OR6K2,OR2M2,RXRG,OR5B21,NTRK3,GRIN3A,LRRTM3,OR2T5,OR2T1,ESRRG,SLC22A2,GPA33,DDR2,FCGR3A,CD244,OR6K3,OR10R2,CD1E,CHRNB2,ROS1,FCGR1B,CD2,HTR1B,LPPR4,ENTPD1,CACNA1B,GRIN1,KCNB1,AGTR2,IRS4,EPHA10,OR1N2,MUSK,CNR2,OR13C5,OR10G9,OR4D1,SYNDIG1,GABBR1,OR5V1,OR2J2,SLC22A6,OXER1,SCNN1D,TNFRSF4,MUC3A,OR6C65,OR6C75,OR10A4,OR51B4,NPSR1,SIRPB1,OR5K1,STAB2,ACVRL1,EPHA6,TAS2R31,KIR3DL1,LAIR1,OR6C4,GRM2,LILRA6,GRIN2A,SLC17A3,SLITRK6,GP5,CSF2RB,VSTM2A,CACNA1I,NRXN1,OR2A14,OR1C1,OR2A25,TAS2R41,SCN9A,CD8A,CALCRL,SLC12A5,SLC4A10,OR5K2,CXCR6,SCN1B,OR5H14,ESR1,CRACR2A,PTPRC,HFE,TMEM8C,CASR,GABRA2,ESRRB,TRPM1,KLRC1,OR13C2,TARM1,GPR19,TAS2R38,KCNC2,MRC1,OR1D5,ZP2,OR8G1,GRIN2B,GPR1,ANTXRL,GRIA3,PRLR |
| GO:0034220 | ion transmembrane | 76 | 995 | 0,69 | 2,97E-28 | CLIC5,AQP8,AQP9,P2RX5,SLC5A9,SLC25A2,KCNJ8,SLC17A1,KCNA5,KCNH2,SLC4A1,HPN,KCNQ4,SLC17A6,CACNA1C,SLC47A1,GABRA6,GABRB2,HTR2C,SLC1A2,SCN2B,KCNA6,CHRNB3,SLC23A1,SLC26A5,SLC9C1,SCN11A,SLC16A11,KCNE3,KCNK10,KCNMB3,CACNB2,SLC34A1,SLC6A18,SLC2A2,SLC22A10,CNGA2,CALHM1,GRID1,P2RX7,SLC22A8,CLCN1,SCARA5,KCNK5,GRIN3A,SLC22A2,SLC9C2,CHRNB2,KCNA10,CACNA1B,GRIN1,KCNB1,KCNE1L,SLC22A6,SCNN1D,SLC28A1,GRIN2A,SLC17A3,KCNU1,CACNA1I,FAM26D,SCN9A,SLC12A5,KCNK16,SLC4A10,SCN1B,PTPRC,HFE,GABRA2,KCNK9,TRPM1,SLC34A3,KCNC2,KCNJ16,GRIN2B,GRIA3 |
| GO:0051716 | cellular response to | 192 | 6212 | 0,3 | 4,44E-27 | RIPK3,AQP8,AQP9,P2RX5,CD69,GPR63,TREM1,OR7C1,KCNA5,RAMP2,CHRM3,DCLK1,SIT1,GPR133,KCNH2,HPN,SORCS1,IL1RL2,CACNA1C,GABRA6,GABRB2,HTR2C,SLC1A2,MS4A2,CD226,MCHR2,GPR15,HTR6,CHRNB3,LILRA4,FGFR4,LHCGR,F2RL2,CLEC1B,OR4K2,OR10A5,GALR1,CD3D,OR51B5,C3AR1,OR10V1,OR52K1,OR9I1,MUC17,OR1B1,OR7G2,OR8U1,OR6Y1,SLAMF1,GPR151,GPRC6A,CD34,OR10H5,OR2Y1,CSPG4,KCNK10,EDNRA,HTR1A,OR7A5,MC5R,GPR62,OR4K13,OR4K5,GHRHR,SLC34A1,OR4S1,OR6B3,OR5AK2,OR52N5,HTR1F,OR8H1,OR8K5,TMEM108,OR10G4,OR52D1,TAS2R60,OR2T10,GRID1,RTN4RL1,P2RX7,OR1G1,NPY2R,OR6P1,GRK1,OR5D18,OPRL1,PLAUR,KIR2DL4,OR13H1,NEO1,NCR3,ERBB4,OR2AG2,ADRB3,OR6N2,SCARA5,OR5H2,RET,TNFRSF14,OR2T6,OR52R1,PTGER3,CCRL2,OR8D1,OR5AC2,OR6K2,OR2M2,RXRG,OR5B21,NTRK3,GRIN3A,LRRTM3,OR2T5,OR2T1,ESRRG,GPA33,DDR2,FCGR3A,CD244,OR6K3,OR10R2,CHRNB2,ROS1,FCGR1B,CD2,HTR1B,LPPR4,GRIN1,KCNB1,AGTR2,IRS4,EPHA10,OR1N2,MUSK,CNR2,OR13C5,OR10G9,OR4D1,GABBR1,OR5V1,OR2J2,OXER1,TNFRSF4,MUC3A,OR6C65,OR6C75,OR10A4,OR51B4,NPSR1,SIRPB1,OR5K1,ACVRL1,EPHA6,TAS2R31,KIR3DL1,OR6C4,GRM2,GRIN2A,CSF2RB,VSTM2A,CACNA1I,NRXN1,OR2A14,OR1C1,OR2A25,TAS2R41,CD8A,CALCRL,OR5K2,CXCR6,OR5H14,ESR1,PTPRC,HFE,CASR,GABRA2,ESRRB,TRPM1,KLRC1,OR13C2,GPR19,TAS2R38,KCNC2,MRC1,OR1D5,ZP2,OR8G1,GRIN2B,GPR1,ANTXRL,GRIA3,PRLR |
| GO:0055085 | transmembrane trar | 81 | 1235 | 0,63 | 3,75E-26 | CLIC5,AQP8,AQP9,P2RX5,SLC5A9,SLC25A2,KCNJ8,SLC17A1,KCNA5,KCNH2,SLC4A1,HPN,KCNQ4,SLC17A6,CACNA1C,SLC47A1,GABRA6,GABRB2,HTR2C,SLC1A2,SCN2B,KCNA6,CHRNB3,SLC23A1,SLC26A5,SLC9C1,SCN11A,SLC16A11,KCNE3,KCNK10,EDNRA,KCNMB3,CACNB2,SLC34A1,SLC6A18,SLC2A2,SLC22A10,CNGA2,CALHM1,GRID1,P2RX7,SLC22A8,CLCN1,SCARA5,KCNK5,GRIN3A,SLC22A2,SLC9C2,CHRNB2,KCNA10,CACNA1B,GRIN1,KCNB1,KCNE1L,SLC46A2,SLC22A6,SCNN1D,SLC28A1,GRIN2A,SLC17A3,KCNU1,CACNA1I,FAM26D,SCN9A,SLC12A5,SLC14A1,KCNK16,SLC4A10,SCN1B,SLC23A3,PTPRC,SLC25A48,HFE,GABRA2,KCNK9,TRPM1,SLC34A3,KCNC2,KCNJ16,GRIN2B,GRIA3 |
| GO:0098655 | cation transmembra | 63 | 720 | 0,75 | 6,15E-26 | P2RX5,SLC5A9,SLC25A2,KCNJ8,SLC17A1,KCNA5,KCNH2,SLC4A1,HPN,KCNQ4,CACNA1C,SLC47A1,HTR2C,SLC1A2,SCN2B,KCNA6,CHRNB3,SLC23A1,SLC9C1,SCN11A,KCNE3,KCNK10,KCNMB3,CACNB2,SLC34A1,SLC6A18,SLC22A10,CNGA2,CALHM1,P2RX7,SCARA5,KCNK5,GRIN3A,SLC22A2,SLC9C2,CHRNB2,KCNA10,CACNA1B,GRIN1,KCNB1,KCNE1L,SLC22A6,SCNN1D,SLC28A1,GRIN2A,SLC17A3,KCNU1,CACNA1I,FAM26D,SCN9A,SLC12A5,KCNK16,SLC4A10,SCN1B,PTPRC,HFE,KCNK9,TRPM1,SLC34A3,KCNC2,KCNJ16,GRIN2B,GRIA3 |
| GO:0006812 | cation transport | 68 | 866 | 0,71 | 1,16E-25 | P2RX5,SLC5A9,SLC25A2,KCNJ8,SLC17A1,KCNA5,RAMP2,KCNH2,SLC4A1,HPN,KCNQ4,SLC17A6,CACNA1C,SLC47A1,HTR2C,SLC1A2,SCN2B,KCNA6,CHRNB3,SLC23A1,SLC9C1,SCN11A,KCNE3,KCNK10,KCNMB3,CACNB2,SLC34A1,SLC6A18,SLC22A10,CNGA2,CALHM1,P2RX7,SCARA5,KCNK5,GRIN3A,SLC22A2,SLC9C2,CHRNB2,KCNA10,CACNA1B,GRIN1,KCNB1,KCNE1L,SLC22A6,SCNN1D,SLC28A1,GRIN2A,SLC17A3,KCNU1,CACNA1I,FAM26D,SCN9A,CALCRL,SLC12A5,KCNK16,SLC4A10,SCN1B,CRACR2A,PTPRC,HFE,CASR,KCNK9,TRPM1,SLC34A3,KCNC2,KCNJ16,GRIN2B,GRIA3 |
| GO:0006811 | ion transport | 81 | 1292 | 0,61 | 5,66E-25 | CLIC5,AQP8,AQP9,P2RX5,SLC5A9,SLC25A2,KCNJ8,SLC17A1,KCNA5,RAMP2,KCNH2,SLC4A1,HPN,KCNQ4,SLC17A6,CACNA1C,SLC47A1,GABRA6,GABRB2,HTR2C,SLC1A2,SCN2B,KCNA6,CHRNB3,SLC23A1,SLC26A5,SLC9C1,SCN11A,SLC16A11,KCNE3,KCNK10,KCNMB3,CACNB2,SLC34A1,SLC6A18,SLC2A2,SLC22A10,CNGA2,CALHM1,GRID1,P2RX7,SLC22A8,CLCN1,SCARA5,KCNK5,GRIN3A,SLC22A2,SLC9C2,CHRNB2,KCNA10,CACNA1B,GRIN1,KCNB1,KCNE1L,SLC22A6,SCNN1D,SLC28A1,GRM2,GRIN2A,SLC17A3,KCNU1,CACNA1I,FAM26D,SCN9A,CALCRL,SLC12A5,KCNK16,SLC4A10,SCN1B,CRACR2A,PTPRC,HFE,CASR,GABRA2,KCNK9,TRPM1,SLC34A3,KCNC2,KCNJ16,GRIN2B,GRIA3 |

| | | | | | | |
|---|---|---|---|---|---|---|
| GO:0030001 | metal ion transport | 59 | 664 | 0,76 | 1,67E-24 | SLC5A9,KCNJ8,SLC17A1,KCNA5,RAMP2,KCNH2,SLC4A1,HPN,KCNQ4,SLC17A6,CACNA1C,HTR2C,SLC1A2,SCN2B,KCNA6,SLC23A1,SLC9C1,SCN11A,KCNE3,KCNK10,KCNMB3,CACNB2,SLC34A1,SLC6A18,CNGA2,CALHM1,P2RX7,SCARA5,KCNK5,GRIN3A,SLC9C2,CHRNB2,KCNA10,CACNA1B,GRIN1,KCNB1,KCNE1L,SCNN1D,SLC28A1,GRIN2A,SLC17A3,KCNU1,CACNA1I,SCN9A,CALCRL,SLC12A5,KCNK16,SLC4A10,SCN1B,CRACR2A,PTPRC,HFE,CASR,KCNK9,TRPM1,SLC34A3,KCNC2,KCNJ16,GRIN2B |
| GO:0098660 | inorganic ion transm | 60 | 707 | 0,74 | 5,24E-24 | CLIC5,SLC5A9,KCNJ8,SLC17A1,KCNA5,KCNH2,SLC4A1,HPN,KCNQ4,CACNA1C,SLC47A1,GABRA6,GABRB2,HTR2C,SLC1A2,SCN2B,KCNA6,SLC23A1,SLC26A5,SLC9C1,SCN11A,KCNE3,KCNK10,KCNMB3,CACNB2,SLC34A1,SLC6A18,CNGA2,CALHM1,P2RX7,CLCN1,SCARA5,KCNK5,GRIN3A,SLC9C2,KCNA10,CACNA1B,GRIN1,KCNB1,KCNE1L,SCNN1D,SLC28A1,GRIN2A,SLC17A3,KCNU1,CACNA1I,SCN9A,SLC12A5,KCNK16,SLC4A10,SCN1B,PTPRC,HFE,GABRA2,KCNK9,TRPM1,SLC34A3,KCNC2,KCNJ16,GRIN2B |
| GO:0098662 | inorganic cation trar | 54 | 618 | 0,75 | 5,97E-22 | SLC5A9,KCNJ8,SLC17A1,KCNA5,KCNH2,SLC4A1,HPN,KCNQ4,CACNA1C,SLC47A1,HTR2C,SLC1A2,SCN2B,KCNA6,SLC23A1,SLC9C1,SCN11A,KCNE3,KCNK10,KCNMB3,CACNB2,SLC34A1,SLC6A18,CNGA2,CALHM1,P2RX7,SCARA5,KCNK5,GRIN3A,SLC9C2,KCNA10,CACNA1B,GRIN1,KCNB1,KCNE1L,SCNN1D,SLC28A1,GRIN2A,SLC17A3,KCNU1,CACNA1I,SCN9A,SLC12A5,KCNK16,SLC4A10,SCN1B,PTPRC,HFE,KCNK9,TRPM1,SLC34A3,KCNC2,KCNJ16,GRIN2B |
| GO:0042221 | response to chemica | 142 | 4153 | 0,35 | 6,79E-21 | AQP8,AQP9,P2RX5,CD69,SLC25A2,KCNJ8,OR7C1,KCNA5,RAMP2,CHRM3,KCNH2,HPN,SLC17A6,IL1RL2,SLC47A1,GABRB2,HTR2C,SLC1A2,SCN2B,CHRNB3,FGFR4,LHCGR,OR4K2,OR10A5,OR51B5,C3AR1,OR10V1,OR52K1,OR9I1,SLC23A1,OR1B1,OR7G2,OR8U1,SLC26A5,OR6Y1,SCN11A,GPRC6A,OR10H5,OR2Y1,OR7A5,OR4K13,KCNMB3,OR4K5,GHRHR,SLC34A1,OR4S1,OR6B3,OR5AK2,OR52N5,OR8H1,OR8K5,TMEM108,OR10G4,OR52D1,OR2T10,CALHM1,RTN4RL1,P2RX7,OR1G1,OR6P1,OR5D18,OPRL1,SLC22A8,PLAUR,OR13H1,NEO1,ERBB4,OR2AG2,ADRB3,OR6N2,OR5H2,RET,TNFRSF14,OR2T6,OR52R1,CCRL2,OR8D1,OR5AC2,OR6K2,OR2M2,RXRG,OR5B21,NTRK3,GRIN3A,LRRTM3,OR2T5,OR2T1,ESRRG,SLC22A2,OR6K3,OR10R2,CHRNB2,FCGR1B,HTR1B,GRIN1,KCNB1,AGTR2,IRS4,OR1N2,CNR2,OR13C5,OR10G9,OR4D1,OR5V1,OR2J2,SLC22A6,TNFRSF4,OR6C65,OR6C75,OR10A4,OR51B4,OR5K1,ACVRL1,TAS2R31,OR6C4,GRIN2A,SLC17A3,CSF2RB,VSTM2A,NRXN1,OR2A14,OR1C1,OR2A25,SCN9A,CALCRL,SLC12A5,OR5K2,CXCR6,SCN1B,OR5H14,ESR1,HFE,CASR,ESRRB,OR13C2,TAS2R38,KCNC2,MRC1,OR1D5,OR8G1,GRIN2B,PRLR |
| GO:0015672 | monovalent inorgan | 41 | 437 | 0,78 | 2,57E-17 | SLC5A9,KCNJ8,SLC17A1,KCNA5,KCNH2,SLC4A1,HPN,KCNQ4,SLC17A6,SLC47A1,SLC1A2,SCN2B,KCNA6,SLC23A1,SLC9C1,SCN11A,KCNE3,KCNK10,KCNMB3,SLC34A1,SLC6A18,CNGA2,KCNK5,SLC9C2,KCNA10,KCNB1,KCNE1L,SCNN1D,SLC28A1,SLC17A3,KCNU1,CACNA1I,SCN9A,SLC12A5,KCNK16,SLC4A10,SCN1B,KCNK9,SLC34A3,KCNC2,KCNJ16 |
| GO:0032501 | multicellular organis | 175 | 6507 | 0,24 | 3,01E-16 | TRO,CLIC5,RIPK3,AQP9,P2RX5,PCDHB6,KCNJ8,OR7C1,KCNA5,RAMP2,CHRM3,DCLK1,KCNH2,HPN,KCNQ4,CACNA1C,GABRB2,HTR2C,SLC1A2,SCN2B,CD226,HTR6,CHRNB3,LHCGR,F2RL2,CLEC1B,OR4K2,OR10A5,CD3D,OR51B5,C3AR1,OR10V1,OR52K1,OR9I1,SLC23A1,OR1B1,OR7G2,OR8U1,SLC26A5,OR6Y1,SLC9C1,SCN11A,CD34,KCNE3,OR10H5,OR2Y1,CSPG4,KCNK10,EDNRA,HTR1A,OR7A5,OR4K13,KCNMB3,OR4K5,CACNB2,GHRHR,SLC34A1,OR4S1,OR6B3,OR5AK2,OR52N5,SLC2A2,OR8H1,OR8K5,TMEM108,OR10G4,OR52D1,TAS2R60,CNGA2,OR2T10,CALHM1,GRID1,RTN4RL1,P2RX7,OR1G1,NPY2R,IGDCC3,OR6P1,GRK1,OR5D18,OPRL1,PLAUR,CLCN1,OR13H1,NEO1,ERBB4,OR2AG2,ADRB3,OR6N2,OR5H2,RET,TNFRSF14,OR2T6,OR52R1,PTGER3,OR8D1,OR5AC2,KCNK5,OR6K2,OR2M2,OR5B21,NTRK3,GRIN3A,OR2T5,OR2T1,DDR2,OR6K3,OR10R2,CHRNB2,ROS1,HTR1B,LPPR4,ENTPD1,CACNA1B,GRIN1,AGTR2,KCNE1L,OR1N2,SLC46A2,MUSK,CNR2,OR13C5,OR10G9,OR4D1,OR5V1,OR2J2,SLC22A6,SCNN1D,TNFRSF4,OR6C65,OR6C75,OR10A4,OR51B4,OR5K1,STAB2,ACVRL1,TAS2R31,OR6C4,GRIN2A,SLITRK6,GP5,CSF2RB,CACNA1I,NRXN1,OR2A14,OR1C1,OR2A25,TAS2R41,SCN9A,CD8A,CALCRL,SLC12A5,SLC4A10,OR5K2,SCN1B,OR5H14,ESR1,PTPRC,HFE,TMEM8C,CASR,ESRRB,MPZ,PCDHA5,TRPM1,OR13C2,TAS2R38,KCNC2,OR1D5,SEMA6B,OR8G1,GRIN2B,GRIA3,PRLR,ADAM29 |
| GO:0042391 | regulation of membr | 37 | 408 | 0,77 | 4,34E-15 | P2RX5,KCNJ8,KCNA5,KCNH2,CACNA1C,SCN2B,CHRNB3,SLC26A5,SCN11A,KCNE3,KCNK10,KCNMB3,CACNB2,SLC34A1,TMEM108,CNGA2,GRID1,P2RX7,NPY2R,CLCN1,KCNK5,GRIN3A,CHRNB2,CACNA1B,GRIN1,KCNB1,KCNE1L,CNR2,GRIN2A,CACNA1I,NRXN1,SCN9A,KCNK16,SCN1B,KCNK9,GRIN2B,GRIA3 |
| GO:0043269 | regulation of ion trar | 43 | 618 | 0,65 | 7,91E-14 | CLIC5,P2RX5,KCNJ8,KCNA5,KCNH2,KCNQ4,CACNA1C,SCN2B,KCNA6,SLC26A5,SCN11A,KCNE3,KCNK10,HTR1A,CACNB2,SLC34A1,CALHM1,P2RX7,NPY2R,OPRL1,CLCN1,KCNK5,CHRNB2,ROS1,KCNA10,HTR1B,CACNA1B,GRIN1,KCNB1,AGTR2,KCNE1L,NPSR1,KCNU1,CACNA1I,NRXN1,SCN9A,KCNK16,SCN1B,CRACR2A,HFE,CASR,KCNC2,KCNJ16 |
| GO:0065007 | biological regulatior | 243 | 11740 | 0,13 | 3,52E-12 | CLIC5,RIPK3,AQP9,P2RX5,CD69,GPR63,KCNJ8,TREM1,OR7C1,KCNA5,RAMP2,CHRM3,DCLK1,SIT1,GPR133,KCNH2,SLC4A1,HPN,KCNQ4,SORCS1,IL1RL2,CACNA1C,GABRA6,GABRB2,LRRTM2,HTR2C,SLC1A2,MS4A2,SCN2B,CD226,KCNA6,MCHR2,GPR15,HTR6,CHRNB3,LILRA4,FGFR4,LHCGR,F2RL2,CLEC1B,OR4K2,OR10A5,GALR1,CD3D,OR51B5,C3AR1,OR10V1,OR52K1,OR9I1,MUC17,TM4SF20,OR1B1,OR7G2,OR8U1,SLC26A5,OR6Y1,SLAMF1,SLC9C1,SCN11A,GPR151,GPRC6A,CD34,KCNE3,OR10H5,OR2Y1,CSPG4,KCNK10,EDNRA,HTR1A,OR7A5,MC5R,GPR62,OR4K13,KCNMB3,OR4K5,CACNB2,GHRHR,SLC34A1,OR4S1,OR6B3,OR5AK2,OR52N5,HTR1F,SLC2A2,OR8H1,OR8K5,TMEM108,OR10G4,OR52D1,TAS2R60,CNGA2,OR2T10,CALHM1,GRID1,RTN4RL1,P2RX7,OR1G1,TMIGD1,NPY2R,OR6P1,GRK1,OR5D18,OPRL1,PLAUR,KIR2DL4,CLCN1,OR13H1,NEO1,NCR3,ERBB4,TREML4,OR2AG2,ADRB3,OR6N2,SCARA5,OR5H2,RET,TNFRSF14,OR2T6,OR52R1,PTGER3,CCRL2,OR8D1,OR5AC2,KCNK5,OR6K2,OR2M2,RXRG,KCTD8,OR5B21,NTRK3,GRIN3A,LRRTM3,OR2T5,OR2T1,ESRRG,SLC22A2,SLC9C2,GPA33,DDR2,FCGR3A,CD244,OR6K3,OR10R2,CHRNB2,ROS1,FCGR1B,CD2,KCNA10,HTR1B,LPPR4,ENTPD1,CACNA1B,GRIN1,KCNB1,AGTR2,KCNE1L,IRS4,EPHA10,OR1N2,SLC46A2,MUSK,CNR2,OR13C5,OR10G9,OR4D1,SYNDIG1,GABBR1,OR5V1,OR2J2,OXER1,TNFRSF4,MUC3A,OR6C65,OR6C75,OR10A4,OR51B4,NPSR1,SIRPB1,OR5K1,ACVRL1,EPHA6,TAS2R31,KIR3DL1,LAIR1,OR6C4,GRM2,GRIN2A,KCNU1,SLITRK6,JAM2,GP5,CSF2RB,VSTM2A,CACNA1I,NRXN1,OR2A14,OR1C1,OR2A25,TAS2R41,SCN9A,CD8A,CALCRL,SLC12A5,KCNK16,SLC4A10,OR5K2,CXCR6,SCN1B,OR5H14,ESR1,TMEM132D,CRACR2A,PTPRC,MALRD1,HFE,TMEM176A,TIGIT,CASR,CD200,GABRA2,ESRRB,KCNK9,MPZ,TRPM1,KLRC1,OR13C2,TARM1,GPR19,SLC34A3,TAS2R38,KCNC2,MRC1,OR1D5,ZP2,KCNJ16,OR8G1,GRIN2B,GPR1,ANTXRL,GRIA3,PTPRQ,PRLR |
| GO:0007268 | chemical synaptic tr | 32 | 402 | 0,71 | 1,71E-11 | P2RX5,PCDHB6,CHRM3,GABRB2,HTR2C,SLC1A2,SCN2B,HTR6,CHRNB3,KCNMB3,CACNB2,HTR1F,GRID1,P2RX7,NPY2R,OPRL1,GRIN3A,SLC22A2,CHRNB2,HTR1B,CACNA1B,GRIN1,GRM2,GRIN2A,NRXN1,SLC12A5,SCN1B,MPZ,GPR19,GRIN2B,GPR1,GRIA3 |
| GO:0071805 | potassium ion transı | 22 | 169 | 0,93 | 1,71E-11 | KCNJ8,KCNA5,KCNH2,HPN,KCNQ4,KCNA6,SLC9C1,KCNE3,KCNK10,KCNMB3,CNGA2,KCNK5,SLC9C2,KCNA10,KCNB1,KCNE1L,KCNU1,SLC12A5,KCNK16,KCNK9,KCNC2,KCNJ16 |
| GO:0034765 | regulation of ion trar | 33 | 434 | 0,69 | 2,09E-11 | CLIC5,KCNJ8,KCNA5,KCNH2,KCNQ4,CACNA1C,SCN2B,KCNA6,SLC26A5,SCN11A,KCNE3,KCNK10,CACNB2,SLC34A1,CALHM1,P2RX7,OPRL1,CLCN1,KCNK5,KCNA10,CACNA1B,KCNB1,KCNE1L,NPSR1,KCNU1,CACNA1I,NRXN1,SCN9A,KCNK16,SCN1B,CRACR2A,KCNC2,KCNJ16 |
| GO:0007187 | G protein-coupled re | 23 | 206 | 0,86 | 7,66E-11 | RAMP2,CHRM3,GPR133,HTR6,LHCGR,GALR1,EDNRA,HTR1A,MC5R,GHRHR,HTR1F,NPY2R,OPRL1,ADRB3,HTR1B,AGTR2,CNR2,GABBR1,OXER1,GRM2,CALCRL,CASR,GPR1 |
| GO:0034762 | regulation of transm | 35 | 524 | 0,64 | 1,10E-10 | CLIC5,KCNJ8,KCNA5,KCNH2,KCNQ4,CACNA1C,SLC1A2,SCN2B,KCNA6,SLC26A5,SCN11A,KCNE3,KCNK10,EDNRA,CACNB2,SLC34A1,CALHM1,P2RX7,OPRL1,CLCN1,KCNK5,KCNA10,CACNA1B,KCNB1,KCNE1L,NPSR1,KCNU1,CACNA1I,NRXN1,SCN9A,KCNK16,SCN1B,CRACR2A,KCNC2,KCNJ16 |
| GO:0050789 | regulation of biologi | 229 | 11116 | 0,13 | 4,92E-10 | CLIC5,RIPK3,P2RX5,CD69,GPR63,KCNJ8,TREM1,OR7C1,KCNA5,RAMP2,CHRM3,DCLK1,SIT1,GPR133,KCNH2,HPN,KCNQ4,SORCS1,IL1RL2,CACNA1C,GABRA6,GABRB2,LRRTM2,HTR2C,SLC1A2,MS4A2,SCN2B,CD226,KCNA6,MCHR2,GPR15,HTR6,CHRNB3,LILRA4,FGFR4,LHCGR,F2RL2,CLEC1B,OR4K2,OR10A5,GALR1,CD3D,OR51B5,C3AR1,OR10V1,OR52K1,OR9I1,MUC17,TM4SF20,OR1B1,OR7G2,OR8U1,SLC26A5,OR6Y1,SLAMF1,SCN11A,GPR151,GPRC6A,CD34,KCNE3,OR10H5,OR2Y1,CSPG4,KCNK10,EDNRA,HTR1A,OR7A5,MC5R,GPR62,OR4K13,OR4K5,CACNB2,GHRHR,SLC34A1,OR4S1,OR6B3,OR5AK2,OR52N5,HTR1F,SLC2A2,OR8H1,OR8K5,TMEM108,OR10G4,OR52D1,TAS2R60,OR2T10,CALHM1,GRID1,RTN4RL1,P2RX7,OR1G1,TMIGD1,NPY2R,OR6P1,GRK1,OR5D18,OPRL1,PLAUR,KIR2DL4,CLCN1,OR13H1,NEO1,NCR3,ERBB4,TREML4,OR2AG2,ADRB3,OR6N2,OR5H2,RET,TNFRSF14,OR2T6,OR52R1,PTGER3,CCRL2,OR8D1,OR5AC2,KCNK5,OR6K2,OR2M2,RXRG,KCTD8,OR5B21,NTRK3,GRIN3A,LRRTM3,OR2T5,OR2T1,ESRRG,GPA33,DDR2,FCGR3A,CD244,OR6K3,OR10R2,CHRNB2,ROS1,FCGR1B,CD2,KCNA10,HTR1B,LPPR4,CACNA1B,GRIN1,KCNB1,AGTR2,KCNE1L,IRS4,EPHA10,OR1N2,SLC46A2,MUSK,CNR2,OR13C5,OR10G9,OR4D1,SYNDIG1,GABBR1,OR5V1,OR2J2,OXER1,TNFRSF4,MUC3A,OR6C65,OR6C75,OR10A4,OR51B4,NPSR1,SIRPB1,OR5K1,ACVRL1,EPHA6,TAS2R31,KIR3DL1,LAIR1,OR6C4,GRM2,GRIN2A,KCNU1,SLITRK6,JAM2,CSF2RB,VSTM2A,CACNA1I,NRXN1,OR2A14,OR1C1,OR2A25,TAS2R41,SCN9A,CD8A,CALCRL,KCNK16,OR5K2,CXCR6,SCN1B,OR5H14,ESR1,TMEM132D,CRACR2A,PTPRC,MALRD1,HFE,TMEM176A,TIGIT,CASR,CD200,GABRA2,ESRRB,MPZ,TRPM1,KLRC1,OR13C2,TARM1,GPR19,TAS2R38,KCNC2,MRC1,OR1D5,ZP2,KCNJ16,OR8G1,GRIN2B,GPR1,ANTXRL,GRIA3,PTPRQ,PRLR |

| | | | | | |
|---|---|---|---|---|---|
| GO:0035725 | sodium ion transme | 19 | 160 | 0,89 | 2,72E-09 | SLC5A9,SLC17A1,SLC4A1,SLC1A2,SCN2B,SLC23A1,SLC9C1,SCN11A,SLC34A1,SLC6A18,SLC9C2,SCNN1D,SLC28A1,SLC17A3,CACNA1I,SCN9A,SLC4A10,SCN1B,SLC34A3 |
| GO:0001508 | action potential | 16 | 104 | 1 | 3,07E-09 | KCNJ8,KCNA5,KCNH2,CACNA1C,SCN2B,SCN11A,KCNMB3,CACNB2,CLCN1,CHRNB2,CACNA1B,KCNB1,KCNE1L,CACNA1I,SCN9A,SCN1B |
| GO:0006814 | sodium ion transpor | 20 | 189 | 0,84 | 5,20E-09 | SLC5A9,SLC17A1,SLC4A1,SLC17A6,SLC1A2,SCN2B,SLC23A1,SLC9C1,SCN11A,SLC34A1,SLC6A18,SLC9C2,SCNN1D,SLC28A1,SLC17A3,CACNA1I,SCN9A,SLC4A10,SCN1B,SLC34A3 |
| GO:0007188 | adenylate cyclase-n | 19 | 183 | 0,83 | 2,06E-08 | RAMP2,CHRM3,GPR133,LHCGR,GALR1,EDNRA,HTR1A,MC5R,GHRHR,HTR1F,NPY2R,OPRL1,ADRB3,HTR1B,GABBR1,OXER1,GRM2,CALCRL,CASR |
| GO:0098656 | anion transmembra | 25 | 353 | 0,66 | 5,85E-08 | CLIC5,AQP9,SLC25A2,SLC17A1,SLC4A1,SLC17A6,GABRA6,GABRB2,SLC1A2,SLC23A1,SLC26A5,SLC16A11,SLC34A1,SLC6A18,SLC2A2,SLC22A10,SLC22A8,CLCN1,SLC22A2,SLC22A6,SLC17A3,SLC12A5,SLC4A10,GABRA2,SLC34A3 |
| GO:0006810 | transport | 110 | 4130 | 0,24 | 7,22E-08 | CLIC5,AQP8,AQP9,CEACAM4,P2RX5,SLC5A9,SLC25A2,KCNJ8,SLC17A1,KCNA5,RAMP2,CHRM3,DCLK1,KCNH2,SLC4A1,HPN,KCNQ4,SLC17A6,CACNA1C,SLC47A1,GABRA6,GABRB2,HTR2C,SLC1A2,SCN2B,KCNA6,CHRNB3,TMPRSS5,C3AR1,SLC23A1,SLC26A5,SLAMF1,SLC9C1,SCN11A,SLC16A11,KCNE3,KCNK10,EDNRA,KCNMB3,CACNB2,GHRHR,SLC34A1,SLC6A18,SLC2A2,TMEM108,SLC22A10,CNGA2,CALHM1,GRID1,P2RX7,NPY2R,SLC22A8,PLAUR,CLCN1,ERBB4,SCARA5,KCNK5,GRIN3A,SLC22A2,SLC9C2,FCGR3A,CHRNB2,FCGR1B,KCNA10,HTR1B,CACNA1B,GRIN1,KCNB1,AGTR2,KCNE1L,SLC46A2,SYNDIG1,SLC22A6,SCNN1D,SIRPB1,STAB2,ESYT3,LAIR1,SLC28A1,GRM2,GRIN2A,SLC17A3,KCNU1,CACNA1I,NRXN1,FAM26D,SCN9A,CALCRL,SLC12A5,SLC14A1,KCNK16,SLC4A10,SCN1B,SLC23A3,CRACR2A,PTPRC,SLC25A48,HFE,CASR,GABRA2,KCNK9,TRPM1,TARM1,SLC34A3,KCNC2,MRC1,KCNJ16,GRIN2B,GRIA3,PRLR |
| GO:0050801 | ion homeostasis | 35 | 708 | 0,51 | 2,12E-07 | P2RX5,KCNA5,KCNH2,SLC4A1,CACNA1C,HTR2C,SCN2B,FGFR4,F2RL2,GALR1,C3AR1,SLC26A5,SLC9C1,EDNRA,SLC34A1,P2RX7,NPY2R,OPRL1,NEO1,SCARA5,SLC9C2,GRIN1,AGTR2,NPSR1,GRIN2A,SLC12A5,SLC4A10,SCN1B,ESR1,PTPRC,HFE,CASR,TRPM1,SLC34A3,GRIN2B |
| GO:0098771 | inorganic ion homeo | 33 | 643 | 0,52 | 2,53E-07 | P2RX5,KCNA5,KCNH2,SLC4A1,CACNA1C,HTR2C,FGFR4,F2RL2,GALR1,C3AR1,SLC26A5,SLC9C1,EDNRA,SLC34A1,P2RX7,NPY2R,OPRL1,NEO1,SCARA5,SLC9C2,GRIN1,AGTR2,NPSR1,GRIN2A,SLC12A5,SLC4A10,ESR1,PTPRC,HFE,CASR,TRPM1,SLC34A3,GRIN2B |
| GO:0065008 | regulation of biologi | 96 | 3559 | 0,24 | 7,62E-07 | RIPK3,AQP9,P2RX5,KCNJ8,KCNA5,RAMP2,CHRM3,SIT1,KCNH2,SLC4A1,HPN,CACNA1C,LRRTM2,HTR2C,SLC1A2,SCN2B,CHRNB3,FGFR4,F2RL2,CLEC1B,GALR1,C3AR1,MUC17,SLC26A5,SLC9C1,SCN11A,CD34,KCNE3,KCNK10,EDNRA,HTR1A,KCNMB3,CACNB2,GHRHR,SLC34A1,SLC2A2,TMEM108,CNGA2,GRID1,P2RX7,TMIGD1,NPY2R,OPRL1,PLAUR,CLCN1,NEO1,ERBB4,ADRB3,SCARA5,RET,PTGER3,KCNK5,NTRK3,GRIN3A,LRRTM3,SLC22A2,SLC9C2,CHRNB2,HTR1B,ENTPD1,CACNA1B,GRIN1,KCNB1,AGTR2,KCNE1L,SLC46A2,MUSK,CNR2,SYNDIG1,NPSR1,ACVRL1,GRIN2A,SLITRK6,GP5,VSTM2A,CACNA1I,NRXN1,SCN9A,SLC12A5,KCNK16,SLC4A10,SCN1B,ESR1,PTPRC,MALRD1,HFE,CASR,GABRA2,ESRRB,KCNK9,TRPM1,SLC34A3,KCNC2,GRIN2B,GRIA3,PRLR |
| GO:0050794 | regulation of cellula | 210 | 10484 | 0,11 | 8,87E-07 | RIPK3,P2RX5,CD69,GPR63,TREM1,OR7C1,KCNA5,RAMP2,CHRM3,DCLK1,SIT1,GPR133,KCNH2,HPN,SORCS1,IL1RL2,CACNA1C,GABRA6,GABRB2,LRRTM2,HTR2C,MS4A2,SCN2B,CD226,MCHR2,GPR15,HTR6,CHRNB3,LILRA4,FGFR4,LHCGR,F2RL2,CLEC1B,OR4K2,OR10A5,GALR1,CD3D,OR51B5,C3AR1,OR10V1,OR52K1,OR9I1,MUC17,TM4SF20,OR1B1,OR7G2,OR8U1,SLC26A5,OR6Y1,SLAMF1,GPR151,GPRC6A,CD34,KCNE3,OR10H5,OR2Y1,CSPG4,KCNK10,EDNRA,HTR1A,OR7A5,MC5R,GPR62,OR4K13,OR4K5,GHRHR,OR4S1,OR6B3,OR5AK2,OR52N5,HTR1F,SLC2A2,OR8H1,OR8K5,TMEM108,OR10G4,OR52D1,TAS2R60,OR2T10,GRID1,RTN4RL1,P2RX7,OR1G1,TMIGD1,NPY2R,OR6P1,GRK1,OR5D18,OPRL1,PLAUR,KIR2DL4,OR13H1,NEO1,NCR3,ERBB4,TREML4,OR2AG2,ADRB3,OR6N2,OR5H2,RET,TNFRSF14,OR2T6,OR52R1,PTGER3,CCRL2,OR8D1,OR5AC2,OR6K2,OR2M2,RXRG,KCTD8,OR5B21,NTRK3,GRIN3A,LRRTM3,OR2T5,OR2T1,ESRRG,GPA33,DDR2,FCGR3A,CD244,OR6K3,OR10R2,CHRNB2,ROS1,FCGR1B,CD2,HTR1B,LPPR4,GRIN1,KCNB1,AGTR2,KCNE1L,IRS4,EPHA10,OR1N2,SLC46A2,MUSK,CNR2,OR13C5,OR10G9,OR4D1,SYNDIG1,GABBR1,OR5V1,OR2J2,OXER1,TNFRSF4,MUC3A,OR6C65,OR6C75,OR10A4,OR51B4,NPSR1,SIRPB1,OR5K1,ACVRL1,EPHA6,TAS2R31,KIR3DL1,OR6C4,GRM2,GRIN2A,SLITRK6,JAM2,CSF2RB,VSTM2A,CACNA1I,NRXN1,OR2A14,OR1C1,OR2A25,TAS2R41,CD8A,CALCRL,OR5K2,CXCR6,SCN1B,OR5H14,ESR1,TMEM132D,PTPRC,MALRD1,HFE,TMEM176A,TIGIT,CASR,CD200,GABRA2,ESRRB,MPZ,TRPM1,KLRC1,OR13C2,TARM1,GPR19,TAS2R38,KCNC2,MRC1,OR1D5,ZP2,OR8G1,GRIN2B,GPR1,ANTXRL,GRIA3,PTPRQ,PRLR |
| GO:0051899 | membrane depolari | 11 | 64 | 1,05 | 1,04E-06 | KCNH2,CACNA1C,SCN2B,SCN11A,CACNB2,P2RX7,CHRNB2,CACNA1B,CACNA1I,SCN9A,SCN1B |
| GO:0006873 | cellular ion homeos | 30 | 584 | 0,52 | 1,20E-06 | P2RX5,KCNA5,SLC4A1,CACNA1C,HTR2C,F2RL2,GALR1,C3AR1,SLC26A5,SLC9C1,EDNRA,SLC34A1,P2RX7,NPY2R,OPRL1,SCARA5,SLC9C2,GRIN1,AGTR2,NPSR1,GRIN2A,SLC12A5,SLC4A10,ESR1,PTPRC,HFE,CASR,TRPM1,SLC34A3,GRIN2B |
| GO:0015698 | inorganic anion tran | 16 | 167 | 0,79 | 1,25E-06 | CLIC5,SLC17A1,SLC4A1,GABRA6,GABRB2,SLC26A5,SLC34A1,SLC22A10,SLC22A8,CLCN1,SLC22A6,SLC17A3,SLC12A5,SLC4A10,GABRA2,SLC34A3 |
| GO:0006820 | anion transport | 28 | 524 | 0,54 | 1,61E-06 | CLIC5,AQP9,SLC25A2,SLC17A1,SLC4A1,SLC17A6,GABRA6,GABRB2,SLC1A2,SLC23A1,SLC26A5,SLC16A11,SLC34A1,SLC6A18,SLC2A2,SLC22A10,CALHM1,P2RX7,SLC22A8,CLCN1,SLC22A2,SLC22A6,GRM2,SLC17A3,SLC12A5,SLC4A10,GABRA2,SLC34A3 |
| GO:0055082 | cellular chemical ho | 32 | 665 | 0,49 | 1,62E-06 | P2RX5,KCNA5,SLC4A1,CACNA1C,HTR2C,F2RL2,GALR1,C3AR1,SLC26A5,SLC9C1,EDNRA,GHRHR,SLC34A1,P2RX7,NPY2R,OPRL1,SCARA5,SLC9C2,GRIN1,KCNB1,AGTR2,NPSR1,GRIN2A,SLC12A5,SLC4A10,ESR1,PTPRC,HFE,CASR,TRPM1,SLC34A3,GRIN2B |
| GO:0051179 | localization | 125 | 5233 | 0,19 | 1,86E-06 | CLIC5,AQP8,AQP9,CEACAM4,P2RX5,SLC5A9,SLC25A2,KCNJ8,SLC17A1,TREM1,KCNA5,RAMP2,CHRM3,DCLK1,KCNH2,SLC4A1,HPN,KCNQ4,SLC17A6,CACNA1C,SLC47A1,GABRA6,GABRB2,HTR2C,SLC1A2,SCN2B,KCNA6,GPR15,HTR6,CHRNB3,FGFR4,TMPRSS5,C3AR1,SLC23A1,SLC26A5,SLAMF1,SLC9C1,SCN11A,CD34,SLC16A11,KCNE3,CSPG4,KCNK10,EDNRA,KCNMB3,CACNB2,GHRHR,SLC34A1,SLC6A18,SLC2A2,TMEM108,SLC22A10,CNGA2,CALHM1,GRID1,P2RX7,NPY2R,SLC22A8,PLAUR,CLCN1,ERBB4,SCARA5,RET,KCNK5,NTRK3,GRIN3A,SLC22A2,SLC9C2,FCGR3A,CD244,CHRNB2,FCGR1B,CD2,KCNA10,HTR1B,CACNA1B,GRIN1,KCNB1,AGTR2,KCNE1L,SLC46A2,MUSK,CNR2,SYNDIG1,SLC22A6,SCNN1D,SIRPB1,STAB2,ACVRL1,ESYT3,LAIR1,SLC28A1,GRM2,GRIN2A,SLC17A3,KCNU1,JAM2,CACNA1I,NRXN1,FAM26D,SCN9A,CALCRL,SLC12A5,SLC14A1,KCNK16,SLC4A10,SCN1B,ESR1,SLC23A3,CRACR2A,PTPRC,SLC25A48,HFE,CASR,GABRA2,KCNK9,TRPM1,TARM1,SLC34A3,KCNC2,MRC1,KCNJ16,GRIN2B,GRIA3,PRLR |
| GO:0035637 | multicellular organis | 13 | 110 | 0,88 | 2,67E-06 | KCNA5,KCNH2,CACNA1C,SCN2B,SCN11A,KCNE3,KCNMB3,CACNB2,CLCN1,KCNE1L,CACNA1I,SCN9A,SCN1B |
| GO:0086010 | membrane depolari | 9 | 42 | 1,14 | 3,57E-06 | KCNH2,CACNA1C,SCN2B,SCN11A,CACNB2,CACNA1B,CACNA1I,SCN9A,SCN1B |
| GO:0007215 | glutamate receptor s | 9 | 43 | 1,13 | 4,18E-06 | GRID1,GRIN3A,GRIN1,KCNB1,GRM2,GRIN2A,TRPM1,GRIN2B,GRIA3 |
| GO:0006816 | calcium ion transpo | 18 | 242 | 0,68 | 5,39E-06 | RAMP2,CACNA1C,HTR2C,CACNB2,CALHM1,P2RX7,GRIN3A,CHRNB2,CACNA1B,GRIN1,GRIN2A,CACNA1I,CALCRL,CRACR2A,PTPRC,CASR,TRPM1,GRIN2B |
| GO:0048878 | chemical homeosta | 39 | 995 | 0,4 | 8,06E-06 | AQP9,P2RX5,KCNA5,KCNH2,SLC4A1,CACNA1C,HTR2C,SCN2B,FGFR4,F2RL2,GALR1,C3AR1,SLC26A5,SLC9C1,EDNRA,GHRHR,SLC34A1,P2RX7,NPY2R,OPRL1,NEO1,SCARA5,SLC9C2,GRIN1,KCNB1,AGTR2,NPSR1,GRIN2A,SLC12A5,SLC4A10,SCN1B,ESR1,PTPRC,MALRD1,HFE,CASR,TRPM1,SLC34A3,GRIN2B |
| GO:0007193 | adenylate cyclase-i | 11 | 85 | 0,92 | 1,17E-05 | CHRM3,EDNRA,HTR1A,HTR1F,NPY2R,OPRL1,HTR1B,GABBR1,OXER1,GRM2,CASR |
| GO:0086002 | cardiac muscle cell | 8 | 36 | 1,16 | 1,41E-05 | KCNJ8,KCNA5,KCNH2,CACNA1C,SCN2B,CACNB2,KCNE1L,SCN1B |
| GO:0055080 | cation homeostasis | 29 | 629 | 0,48 | 1,52E-05 | P2RX5,KCNA5,KCNH2,SLC4A1,CACNA1C,HTR2C,F2RL2,GALR1,C3AR1,SLC26A5,SLC9C1,EDNRA,P2RX7,NPY2R,OPRL1,NEO1,SCARA5,SLC9C2,GRIN1,AGTR2,NPSR1,GRIN2A,SLC4A10,ESR1,PTPRC,HFE,CASR,TRPM1,GRIN2B |
| GO:0051049 | regulation of transp | 55 | 1732 | 0,31 | 1,57E-05 | CLIC5,P2RX5,KCNJ8,KCNA5,KCNH2,KCNQ4,CACNA1C,LRRTM2,HTR2C,SLC1A2,SCN2B,KCNA6,GALR1,SLC26A5,SCN11A,CD34,KCNE3,KCNK10,EDNRA,HTR1A,CACNB2,GHRHR,SLC34A1,SLC2A2,CALHM1,P2RX7,NPY2R,OPRL1,CLCN1,PTGER3,KCNK5,CD244,CHRNB2,ROS1,CD2,KCNA10,HTR1B,CACNA1B,GRIN1,KCNB1,AGTR2,KCNE1L,TNFRSF4,NPSR1,KCNU1,CACNA1I,NRXN1,SCN9A,KCNK16,SCN1B,CRACR2A,HFE,CASR,KCNC2,KCNJ16 |
| GO:0086091 | regulation of heart r | 8 | 37 | 1,15 | 1,61E-05 | KCNA5,KCNH2,CACNA1C,SCN2B,KCNE3,CACNB2,KCNE1L,SCN1B |
| GO:0007267 | cell-cell signaling | 40 | 1073 | 0,38 | 1,76E-05 | P2RX5,PCDHB6,KCNA5,CHRM3,CACNA1C,GABRB2,HTR2C,SLC1A2,SCN2B,HTR6,CHRNB3,CD34,KCNMB3,CACNB2,GHRHR,HTR1F,GRID1,P2RX7,NPY2R,OPRL1,ADRB3,GRIN3A,SLC22A2,CHRNB2,HTR1B,CACNA1B,GRIN1,AGTR2,KCNE1L,GRM2,GRIN2A,NRXN1,SLC12A5,SCN1B,MPZ,PCDHA5,GPR19,GRIN2B,GPR1,GRIA3 |
| GO:0019932 | second-messenger- | 18 | 266 | 0,64 | 1,77E-05 | RAMP2,GPR133,CACNA1C,LHCGR,GALR1,GPRC6A,MC5R,GPR62,GHRHR,P2RX7,NPY2R,ADRB3,GRIN1,AGTR2,GRIN2A,CALCRL,KCNC2,GRIN2B |
| GO:0030003 | cellular cation home | 27 | 570 | 0,49 | 2,20E-05 | P2RX5,KCNA5,SLC4A1,CACNA1C,HTR2C,F2RL2,GALR1,C3AR1,SLC26A5,SLC9C1,EDNRA,P2RX7,NPY2R,OPRL1,SCARA5,SLC9C2,GRIN1,AGTR2,NPSR1,GRIN2A,SLC4A10,ESR1,PTPRC,HFE,CASR,TRPM1,GRIN2B |
| GO:0019725 | cellular homeostasi | 33 | 806 | 0,42 | 2,69E-05 | P2RX5,KCNA5,SLC4A1,CACNA1C,HTR2C,F2RL2,GALR1,C3AR1,MUC17,SLC26A5,SLC9C1,EDNRA,GHRHR,SLC34A1,P2RX7,NPY2R,OPRL1,SCARA5,SLC9C2,GRIN1,KCNB1,AGTR2,NPSR1,GRIN2A,SLC12A5,SLC4A10,ESR1,PTPRC,HFE,CASR,TRPM1,SLC34A3,GRIN2B |

| | | | | | | |
|---|---|---|---|---|---|---|
| GO:0008015 | blood circulation | 21 | 373 | 0,56 | 3,13E-05 | KCNJ8,KCNA5,RAMP2,CHRM3,KCNH2,CACNA1C,SCN2B,C3AR1,CD34,EDNRA,HTR1A,CACNB2,OPRL1,ADRB3,HTR1B,CACNA1B,AGTR2,KCNE1L,ACVRL1,SCN1B,CASR |
| GO:0060079 | excitatory postsynap | 10 | 76 | 0,93 | 3,13E-05 | P2RX5,CHRNB3,GRID1,P2RX7,GRIN3A,CHRNB2,GRIN1,GRIN2A,GRIN2B,GRIA3 |
| GO:0007204 | positive regulation c | 17 | 251 | 0,64 | 3,41E-05 | P2RX5,CACNA1C,HTR2C,F2RL2,GALR1,C3AR1,EDNRA,P2RX7,NPY2R,OPRL1,GRIN1,NPSR1,GRIN2A,ESR1,PTPRC,TRPM1,GRIN2B |
| GO:0051480 | regulation of cytosol | 18 | 289 | 0,61 | 4,85E-05 | P2RX5,KCNA5,CACNA1C,HTR2C,F2RL2,GALR1,C3AR1,EDNRA,P2RX7,NPY2R,OPRL1,GRIN1,NPSR1,GRIN2A,ESR1,PTPRC,TRPM1,GRIN2B |
| GO:0098661 | inorganic anion tran | 11 | 111 | 0,81 | 9,95E-05 | CLIC5,SLC17A1,SLC4A1,GABRA6,GABRB2,SLC26A5,SLC34A1,CLCN1,SLC12A5,GABRA2,SLC34A3 |
| GO:0098664 | G protein-coupled s | 6 | 22 | 1,25 | 0,00013 | HTR2C,HTR6,OR10H5,HTR1A,HTR1F,HTR1B |
| GO:0035235 | ionotropic glutamate | 6 | 25 | 1,19 | 0,00023 | GRID1,GRIN3A,GRIN1,GRIN2A,GRIN2B,GRIA3 |
| GO:0042592 | homeostatic proces | 46 | 1491 | 0,3 | 0,00027 | RIPK3,AQP9,P2RX5,KCNA5,SIT1,KCNH2,SLC4A1,CACNA1C,HTR2C,SCN2B,FGFR4,F2RL2,GALR1,C3AR1,MUC17,SLC26A5,SLC9C1,CD34,EDNRA,GHRHR,SLC34A1,P2RX7,NPY2R,OPRL1,NEO1,ADRB3,SCARA5,SLC9C2,GRIN1,KCNB1,AGTR2,SLC46A2,NPSR1,GRIN2A,SLC12A5,SLC4A10,SCN1B,ESR1,PTPRC,MALRD1,HFE,CASR,ESRRB,TRPM1,SLC34A3,GRIN2B |
| GO:0044057 | regulation of system | 23 | 516 | 0,46 | 0,00034 | KCNA5,CHRM3,KCNH2,CACNA1C,HTR2C,SCN2B,GALR1,SCN11A,KCNE3,CACNB2,TMEM108,NPY2R,OPRL1,PTGER3,CACNA1B,GRIN1,AGTR2,KCNE1L,GRIN2A,NRXN1,CALCRL,SCN1B,CASR |
| GO:0055065 | metal ion homeosta | 24 | 555 | 0,45 | 0,00034 | P2RX5,KCNA5,KCNH2,CACNA1C,HTR2C,F2RL2,GALR1,C3AR1,EDNRA,P2RX7,NPY2R,OPRL1,NEO1,SCARA5,GRIN1,AGTR2,NPSR1,GRIN2A,ESR1,PTPRC,HFE,CASR,TRPM1,GRIN2B |
| GO:0015711 | organic anion trans | 20 | 414 | 0,5 | 0,00042 | AQP9,SLC25A2,SLC17A1,SLC4A1,SLC17A6,SLC1A2,SLC23A1,SLC26A5,SLC16A11,SLC6A18,SLC2A2,SLC22A10,CALHM1,P2RX7,SLC22A8,SLC22A2,SLC22A6,GRM2,SLC17A3,SLC4A10 |
| GO:0007200 | phospholipase C-ac | 10 | 109 | 0,77 | 0,00047 | CHRM3,HTR2C,MCHR2,LHCGR,F2RL2,C3AR1,OPRL1,PTGER3,ESR1,CASR |
| GO:0032879 | regulation of localiz | 66 | 2524 | 0,23 | 0,00047 | CLIC5,RIPK3,P2RX5,KCNJ8,KCNA5,DCLK1,KCNH2,KCNQ4,CACNA1C,LRRTM2,HTR2C,SLC1A2,SCN2B,KCNA6,GALR1,C3AR1,SLC26A5,SCN11A,CD34,KCNE3,KCNK10,EDNRA,HTR1A,CACNB2,GHRHR,SLC34A1,SLC2A2,CALHM1,P2RX7,TMIGD1,NPY2R,OPRL1,CLCN1,ERBB4,RET,PTGER3,KCNK5,NTRK3,DDR2,CD244,CHRNB2,ROS1,CD2,KCNA10,HTR1B,CACNA1B,GRIN1,KCNB1,AGTR2,KCNE1L,TNFRSF4,NPSR1,ACVRL1,KCNU1,VSTM2A,CACNA1I,NRXN1,SCN9A,KCNK16,SCN1B,CRACR2A,PTPRC,HFE,CASR,KCNC2,KCNJ16 |
| GO:0070588 | calcium ion transme | 13 | 189 | 0,65 | 0,00047 | CACNA1C,HTR2C,CACNB2,CALHM1,P2RX7,GRIN3A,CACNA1B,GRIN1,GRIN2A,CACNA1I,PTPRC,TRPM1,GRIN2B |
| GO:0051963 | regulation of synaps | 9 | 86 | 0,83 | 0,00048 | LRRTM2,NTRK3,LRRTM3,CHRNB2,GRIN1,MUSK,SYNDIG1,SLITRK6,NRXN1 |
| GO:0060402 | calcium ion transpo | 8 | 65 | 0,9 | 0,00049 | CACNA1C,HTR2C,P2RX7,GRIN1,GRIN2A,PTPRC,TRPM1,GRIN2B |
| GO:0042493 | response to drug | 32 | 900 | 0,36 | 0,0005 | P2RX5,CD69,SLC25A2,KCNA5,RAMP2,CHRM3,KCNH2,SLC17A6,SLC47A1,HTR2C,SLC1A2,CHRNB3,SLC26A5,SCN11A,SLC34A1,CALHM1,P2RX7,RET,NTRK3,GRIN3A,SLC22A2,CHRNB2,HTR1B,GRIN1,AGTR2,CNR2,SLC22A6,GRIN2A,SLC17A3,SLC12A5,KCNC2,GRIN2B |
| GO:0043270 | positive regulation c | 15 | 252 | 0,59 | 0,0005 | P2RX5,KCNH2,KCNE3,CACNB2,SLC34A1,P2RX7,NPY2R,CHRNB2,GRIN1,KCNE1L,NPSR1,SCN1B,CRACR2A,CASR,KCNC2 |
| GO:0006874 | cellular calcium ion | 19 | 388 | 0,5 | 0,00053 | P2RX5,KCNA5,CACNA1C,HTR2C,F2RL2,GALR1,C3AR1,EDNRA,P2RX7,NPY2R,OPRL1,GRIN1,NPSR1,GRIN2A,ESR1,PTPRC,CASR,TRPM1,GRIN2B |
| GO:0006875 | cellular metal ion hc | 22 | 499 | 0,46 | 0,00054 | P2RX5,KCNA5,CACNA1C,HTR2C,F2RL2,GALR1,C3AR1,EDNRA,P2RX7,NPY2R,OPRL1,SCARA5,GRIN1,AGTR2,NPSR1,GRIN2A,ESR1,PTPRC,HFE,CASR,TRPM1,GRIN2B |
| GO:0007610 | behavior | 23 | 541 | 0,44 | 0,0006 | HTR2C,SLC1A2,KCNK10,HTR1A,GRID1,NPY2R,OPRL1,ADRB3,CHRNB2,HTR1B,CACNA1B,GRIN1,AGTR2,MUSK,GRIN2A,SLITRK6,NRXN1,SCN9A,SLC12A5,SLC4A10,CASR,GPR19,GRIN2B |
| GO:0010959 | regulation of metal i | 18 | 360 | 0,51 | 0,00066 | P2RX5,KCNA5,KCNH2,CACNA1C,SCN2B,KCNE3,CACNB2,P2RX7,OPRL1,CACNA1B,GRIN1,KCNE1L,NPSR1,SCN1B,CRACR2A,HFE,CASR,KCNC2 |
| GO:0002027 | regulation of heart r | 9 | 95 | 0,79 | 0,0009 | KCNA5,KCNH2,CACNA1C,SCN2B,KCNE3,CACNB2,AGTR2,KCNE1L,SCN1B |
| GO:0006821 | chloride transport | 9 | 101 | 0,76 | 0,0014 | CLIC5,SLC4A1,GABRA6,GABRB2,SLC26A5,CLCN1,SLC12A5,SLC4A10,GABRA2 |
| GO:0015747 | urate transport | 4 | 11 | 1,37 | 0,0018 | SLC17A1,SLC22A10,SLC22A6,SLC17A3 |
| GO:0099623 | regulation of cardia | 5 | 24 | 1,13 | 0,0019 | KCNA5,KCNH2,KCNE3,KCNE1L,SCN1B |
| GO:0007166 | cell surface recepto | 57 | 2198 | 0,23 | 0,0021 | P2RX5,KCNA5,GPR133,HPN,IL1RL2,MS4A2,CHRNB3,LILRA4,FGFR4,CLEC1B,CD3D,C3AR1,MUC17,CSPG4,GHRHR,GRID1,RTN4RL1,P2RX7,PLAUR,NCR3,ERBB4,RET,TNFRSF14,CCRL2,NTRK3,GRIN3A,LRRTM3,DDR2,FCGR3A,CHRNB2,ROS1,FCGR1B,CD2,GRIN1,KCNB1,AGTR2,IRS4,EPHA10,MUSK,TNFRSF4,MUC3A,SIRPB1,ACVRL1,EPHA6,GRM2,GRIN2A,CSF2RB,CD8A,CALCRL,CXCR6,PTPRC,HFE,TRPM1,KLRC1,GRIN2B,GRIA3,PRLR |
| GO:0007189 | adenylate cyclase-a | 8 | 88 | 0,77 | 0,003 | RAMP2,GPR133,LHCGR,GALR1,MC5R,GHRHR,ADRB3,CALCRL |
| GO:0001964 | startle response | 5 | 27 | 1,08 | 0,0031 | GRIN3A,GRIN1,GRIN2A,SLITRK6,NRXN1 |
| GO:0086019 | cell-cell signaling in | 5 | 27 | 1,08 | 0,0031 | KCNA5,CACNA1C,CACNB2,KCNE1L,SCN1B |
| GO:0019935 | cyclic-nucleotide-me | 9 | 115 | 0,71 | 0,0032 | RAMP2,GPR133,LHCGR,GALR1,MC5R,GHRHR,ADRB3,CALCRL,KCNC2 |
| GO:0086014 | atrial cardiac muscle | 4 | 14 | 1,27 | 0,0036 | KCNA5,CACNA1C,CACNB2,KCNE1L |
| GO:0050795 | regulation of behavi | 7 | 68 | 0,82 | 0,0037 | HTR1A,GHRHR,NPY2R,OPRL1,CHRNB2,HTR1B,NRXN1 |
| GO:0006936 | muscle contraction | 13 | 244 | 0,54 | 0,0042 | KCNJ8,KCNA5,CHRM3,KCNH2,CACNA1C,SCN2B,EDNRA,CACNB2,CLCN1,PTGER3,CHRNB2,KCNE1L,SCN1B |
| GO:0007631 | feeding behavior | 8 | 94 | 0,74 | 0,0042 | HTR2C,NPY2R,OPRL1,ADRB3,CHRNB2,HTR1B,GRIN1,GPR19 |
| GO:1902476 | chloride transmemb | 8 | 94 | 0,74 | 0,0042 | CLIC5,SLC4A1,GABRA6,GABRB2,SLC26A5,CLCN1,SLC12A5,GABRA2 |
| GO:0034219 | carbohydrate transn | 6 | 50 | 0,89 | 0,005 | AQP9,SLC5A9,SLC23A1,SLC26A5,EDNRA,SLC2A2 |
| GO:0051259 | protein complex olig | 20 | 512 | 0,4 | 0,005 | RIPK3,KCNA5,SLC1A2,KCNA6,CD3D,SLC26A5,SLC34A1,CALHM1,P2RX7,SCARA5,RXRG,KCTD8,CHRNB2,KCNA10,GRIN1,KCNB1,SLC22A6,TRPM1,KCNC2,GRIN2B |
| GO:0030322 | stabilization of mem | 4 | 16 | 1,21 | 0,0051 | KCNK10,KCNK5,KCNK16,KCNK9 |
| GO:0086013 | membrane repolariz | 4 | 16 | 1,21 | 0,0051 | KCNJ8,KCNA5,KCNH2,KCNE1L |
| GO:0019233 | sensory perception | 7 | 75 | 0,78 | 0,0059 | P2RX7,OPRL1,CHRNB2,GRIN1,CNR2,GRIN2A,SCN9A |
| GO:0003012 | muscle system proc | 14 | 291 | 0,49 | 0,006 | KCNJ8,KCNA5,CHRM3,KCNH2,CACNA1C,SCN2B,EDNRA,CACNB2,CLCN1,PTGER3,CHRNB2,AGTR2,KCNE1L,SCN1B |
| GO:0006817 | phosphate ion trans | 4 | 17 | 1,18 | 0,006 | SLC17A1,SLC34A1,SLC17A3,SLC34A3 |
| GO:0086012 | membrane depolari | 4 | 17 | 1,18 | 0,006 | CACNA1C,SCN2B,CACNB2,SCN1B |
| GO:0019226 | transmission of nerv | 6 | 53 | 0,87 | 0,0061 | SCN11A,KCNMB3,CLCN1,CACNA1I,SCN9A,SCN1B |
| GO:0050807 | regulation of synaps | 10 | 161 | 0,6 | 0,0067 | LRRTM2,NTRK3,LRRTM3,CHRNB2,GRIN1,MUSK,SYNDIG1,SLITRK6,NRXN1,GRIN2B |
| GO:0046883 | regulation of hormo | 13 | 261 | 0,51 | 0,0069 | KCNA5,CACNA1C,HTR2C,GALR1,HTR1A,GHRHR,SLC2A2,KCNB1,AGTR2,NRXN1,HFE,CASR,KCNC2 |
| GO:0018108 | peptidyl-tyrosine ph | 11 | 195 | 0,56 | 0,0073 | FGFR4,ERBB4,RET,NTRK3,DDR2,ROS1,EPHA10,MUSK,EPHA6,CSF2RB,PRLR |
| GO:0055083 | monovalent inorgan | 4 | 19 | 1,13 | 0,0081 | FGFR4,SLC34A1,SLC12A5,SLC34A3 |
| GO:0044341 | sodium-dependent | 3 | 7 | 1,44 | 0,0082 | SLC17A1,SLC34A1,SLC34A3 |
| GO:0045188 | regulation of circadi | 3 | 7 | 1,44 | 0,0082 | GHRHR,NPY2R,CHRNB2 |
| GO:0098739 | import across plasm | 7 | 82 | 0,74 | 0,0087 | KCNJ8,SLC1A2,SLC9C1,SLC34A1,SLC9C2,HFE,KCNJ16 |
| GO:0097553 | calcium ion transme | 6 | 58 | 0,83 | 0,0088 | HTR2C,P2RX7,GRIN1,GRIN2A,PTPRC,GRIN2B |
| GO:0060307 | regulation of ventric | 4 | 20 | 1,11 | 0,0091 | KCNH2,KCNE3,KCNE1L,SCN1B |
| GO:0007218 | neuropeptide signal | 8 | 110 | 0,67 | 0,0092 | SORCS1,MCHR2,GALR1,NPY2R,OPRL1,NPSR1,GPR19,GPR1 |
| GO:0031644 | regulation of neurol | 8 | 112 | 0,67 | 0,0101 | HTR2C,SCN11A,TMEM108,NPY2R,OPRL1,GRIN1,GRIN2A,NRXN1 |
| GO:0051260 | protein homooligom | 14 | 312 | 0,46 | 0,0101 | RIPK3,KCNA5,SLC1A2,KCNA6,CD3D,SLC34A1,CALHM1,SCARA5,RXRG,KCTD8,KCNA10,KCNB1,SLC22A6,KCNC2 |
| GO:0034767 | positive regulation c | 9 | 142 | 0,61 | 0,0103 | KCNH2,KCNE3,CACNB2,SLC34A1,P2RX7,KCNE1L,NPSR1,CRACR2A,KCNC2 |
| GO:0015749 | monosaccharide tra | 5 | 39 | 0,92 | 0,0104 | SLC5A9,SLC23A1,SLC26A5,EDNRA,SLC2A2 |
| GO:0045759 | negative regulation | 3 | 8 | 1,39 | 0,0104 | KCNE3,CHRNB2,CNR2 |
| GO:0098976 | excitatory chemical | 3 | 8 | 1,39 | 0,0104 | GRIN1,GRIN2A,GRIN2B |
| GO:0051046 | regulation of secreti | 24 | 728 | 0,33 | 0,0105 | KCNA5,CACNA1C,HTR2C,GALR1,CD34,HTR1A,GHRHR,SLC2A2,P2RX7,NPY2R,OPRL1,PTGER3,CD244,CHRNB2,CD2,HTR1B,KCNB1,AGTR2,TNFRSF4,CACNA1I,NRXN1,HFE,CASR,KCNC2 |
| GO:0051928 | positive regulation c | 8 | 115 | 0,65 | 0,0115 | P2RX5,KCNE3,CACNB2,P2RX7,GRIN1,NPSR1,CRACR2A,CASR |
| GO:0015851 | nucleobase transpo | 3 | 9 | 1,33 | 0,0133 | AQP9,SLC23A1,SLC28A1 |
| GO:0097623 | potassium ion expo | 3 | 9 | 1,33 | 0,0133 | KCNA5,KCNH2,KCNE1L |
| GO:1903522 | regulation of blood | 12 | 249 | 0,49 | 0,0133 | KCNA5,CHRM3,KCNH2,CACNA1C,SCN2B,KCNE3,CACNB2,CACNA1B,AGTR2,KCNE1L,SCN1B,CASR |
| GO:0051965 | positive regulation c | 6 | 65 | 0,78 | 0,0138 | LRRTM2,NTRK3,LRRTM3,SYNDIG1,SLITRK6,NRXN1 |
| GO:0055067 | monovalent inorgan | 8 | 119 | 0,64 | 0,0138 | KCNA5,KCNH2,SLC4A1,SLC26A5,SLC9C1,SLC9C2,AGTR2,SLC4A10 |
| GO:0007214 | gamma-aminobutyri | 4 | 24 | 1,03 | 0,0147 | GABRA6,GABRB2,GABBR1,GABRA2 |
| GO:0050909 | sensory perception | 6 | 66 | 0,77 | 0,0147 | TAS2R60,CALHM1,SCNN1D,TAS2R31,TAS2R41,TAS2R38 |

| | | | | | |
|---|---|---|---|---|---|
| GO:0098659 | inorganic cation imp | 6 | 66 | 0,77 | 0,0147 | KCNJ8,SLC9C1,SLC34A1,SLC9C2,HFE,KCNJ16 |
| GO:0033555 | multicellular organis | 6 | 67 | 0,76 | 0,0155 | HTR2C,HTR1A,NPY2R,RET,CACNA1B,SCN9A |
| GO:0051050 | positive regulation c | 27 | 892 | 0,29 | 0,0155 | P2RX5,KCNH2,SLC1A2,GALR1,CD34,KCNE3,CACNB2,GHRHR,SLC34A1,P2RX7,NPY2R,OPRL1,CD244,CHRNB2,CD2,GRIN1,KCNB1,AGTR2,KCNE1L,TNFRSF4,NPSR1,CACNA1I,SCN1B,CRACR2A,HFE,CASR,KCNC2 |
| GO:0002791 | regulation of peptide | 17 | 452 | 0,39 | 0,0161 | KCNA5,CACNA1C,HTR2C,CD34,GHRHR,SLC2A2,P2RX7,NPY2R,CD244,CD2,KCNB1,AGTR2,TNFRSF4,NRXN1,HFE,CASR,KCNC2 |
| GO:0001914 | regulation of T cell r | 4 | 25 | 1,02 | 0,0165 | RIPK3,P2RX7,NCR3,PTPRC |
| GO:0007190 | activation of adenyla | 4 | 25 | 1,02 | 0,0165 | LHCGR,EDNRA,GHRHR,ADRB3 |
| GO:0090102 | cochlea developme | 5 | 45 | 0,86 | 0,0168 | HPN,GABRB2,SLC26A5,NTRK3,SLITRK6 |
| GO:1903530 | regulation of secreti | 22 | 672 | 0,33 | 0,0174 | KCNA5,CACNA1C,HTR2C,GALR1,CD34,HTR1A,GHRHR,SLC2A2,P2RX7,NPY2R,CD244,CHRNB2,CD2,HTR1B,KCNB1,AGTR2,TNFRSF4,CACNA1I,NRXN1,HFE,CASR,KCNC2 |
| GO:0002682 | regulation of immun | 37 | 1391 | 0,24 | 0,0181 | RIPK3,TREM1,SIT1,IL1RL2,MS4A2,CD226,LILRA4,LHCGR,CD3D,C3AR1,MUC17,CD34,P2RX7,KIR2DL4,NCR3,TREML4,TNFRSF14,FCGR3A,CD244,CHRNB2,FCGR1B,CD2,SLC46A2,CNR2,TNFRSF4,MUC3A,KIR3DL1,LAIR1,CD8A,ESR1,PTPRC,HFE,TMEM176A,TIGIT,CD200,KLRC1,TARM1 |
| GO:0009987 | cellular process | 249 | 14652 | 0,04 | 0,0202 | CLIC5,RIPK3,AQP8,AQP9,P2RX5,CD69,GPR63,PCDHB6,SLC25A2,KCNJ8,SLC17A1,TREM1,OR7C1,KCNA5,RAMP2,CHRM3,DCLK1,SIT1,GPR133,KCNH2,SLC4A1,HPN,KCNQ4,SORCS1,IL1RL2,CACNA1C,GABRA6,GABRB2,LRRTM2,HTR2C,SLC1A2,MS4A2,SCN2B,CD226,KCNA6,MCHR2,GPR15,HTR6,CHRNB3,LILRA4,FGFR4,LHCGR,F2RL2,CLEC1B,OR4K2,OR10A5,GALR1,CD3D,OR51B5,C3AR1,OR10V1,OR52K1,OR9I1,MUC17,SLC23A1,OR1B1,OR7G2,OR8U1,SLC26A5,OR6Y1,SLAMF1,SLC9C1,SCN11A,GPR151,GPRC6A,CD34,KCNE3,OR10H5,OR2Y1,CSPG4,KCNK10,EDNRA,HTR1A,OR7A5,MC5R,GPR62,OR4K13,KCNMB3,OR4K5,CACNB2,GHRHR,SLC34A1,OR4S1,OR6B3,OR5AK2,OR52N5,HTR1F,OR8H1,OR8K5,TMEM108,OR10G4,OR52D1,TAS2R60,CNGA2,OR2T10,CALHM1,GRID1,RTN4RL1,P2RX7,OR1G1,NPY2R,OR6P1,GRK1,OR5D18,OPRL1,PLAUR,KIR2DL4,CLCN1,OR13H1,NEO1,NCR3,ERBB4,OR2AG2,ADRB3,OR6N2,SCARA5,OR5H2,RET,TNFRSF14,OR2T6,OR52R1,PTGER3,CCRL2,OR8D1,OR5AC2,KCNK5,OR6K2,OR2M2,RXRG,KCTD8,OR5B21,NTRK3,GRIN3A,LRRTM3,OR2T5,OR2T1,ESRRG,SLC22A2,SLC9C2,GPA33,DDR2,FCRLA,FCGR3A,CD244,OR6K3,OR10R2,CHRNB2,ROS1,FCGR1B,CD2,KCNA10,HTR1B,LPPR4,ENTPD1,CACNA1B,GRIN1,KCNB1,AGTR2,KCNE1L,IRS4,EPHA10,OR1N2,MUSK,CNR2,OR13C5,OR10G9,OR4D1,GABBR1,OR5V1,OR2J2,SLC22A6,OXER1,TNFRSF4,MUC3A,OR6C65,OR6C75,OR10A4,OR51B4,NPSR1,SIRPB1,OR5K1,STAB2,ACVRL1,ESYT3,EPHA6,TAS2R31,KIR3DL1,LAIR1,OR6C4,SLC28A1,GRM2,GRIN2A,SLC17A3,KCNU1,TPTE2,SLITRK6,JAM2,GP5,CSF2RB,VSTM2A,CACNA1I,NRXN1,OR2A14,OR1C1,OR2A25,TAS2R41,SCN9A,CD8A,CALCRL,SLC12A5,KCNK16,SLC4A10,OR5K2,CXCR6,SCN1B,OR5H14,ESR1,CRACR2A,PTPRC,HFE,TIGIT,TMEM8C,CASR,CD200,GABRA2,ESRRB,KCNK9,MPZ,PCDHA5,TRPM1,KLRC1,OR13C2,TARM1,GPR19,SLC34A3,TAS2R38,KCNC2,MRC1,OR1D5,ZP2,KCNJ16,SEMA6B,OR8G1,GRIN2B,GPR1,ANTXRL,GRIA3,PTPRQ,PRLR |
| GO:0034764 | positive regulation c | 10 | 194 | 0,52 | 0,0203 | KCNH2,SLC1A2,KCNE3,CACNB2,SLC34A1,P2RX7,KCNE1L,NPSR1,CRACR2A,KCNC2 |
| GO:0035296 | regulation of tube di | 8 | 129 | 0,6 | 0,0205 | KCNA5,CHRM3,EDNRA,HTR1A,ADRB3,HTR1B,AGTR2,CASR |
| GO:0098900 | regulation of action | 5 | 48 | 0,83 | 0,0211 | CACNA1C,KCNE3,CHRNB2,KCNB1,CNR2 |
| GO:0003018 | vascular process in | 9 | 162 | 0,56 | 0,0212 | KCNA5,RAMP2,CHRM3,EDNRA,HTR1A,ADRB3,HTR1B,AGTR2,CASR |
| GO:0050433 | regulation of catech | 5 | 49 | 0,82 | 0,0227 | NPY2R,CHRNB2,HTR1B,KCNB1,AGTR2 |
| GO:1903765 | negative regulation | 2 | 2 | 1,81 | 0,0227 | KCNH2,KCNE3 |
| GO:0033605 | positive regulation c | 3 | 12 | 1,21 | 0,023 | NPY2R,CHRNB2,KCNB1 |
| GO:0051924 | regulation of calciur | 11 | 236 | 0,48 | 0,0243 | P2RX5,CACNA1C,KCNE3,CACNB2,P2RX7,OPRL1,CACNA1B,GRIN1,NPSR1,CRACR2A,CASR |
| GO:0008016 | regulation of heart c | 10 | 201 | 0,51 | 0,0248 | KCNA5,KCNH2,CACNA1C,SCN2B,KCNE3,CACNB2,CACNA1B,AGTR2,KCNE1L,SCN1B |
| GO:0045471 | response to ethanol | 8 | 134 | 0,59 | 0,0248 | NTRK3,GRIN3A,CHRNB2,HTR1B,GRIN1,GRIN2A,KCNC2,GRIN2B |
| GO:0051047 | positive regulation c | 15 | 393 | 0,39 | 0,0251 | GALR1,CD34,GHRHR,P2RX7,NPY2R,OPRL1,CD244,CHRNB2,CD2,KCNB1,AGTR2,TNFRSF4,CACNA1I,HFE,CASR |
| GO:0098657 | import into cell | 20 | 609 | 0,33 | 0,0261 | CEACAM4,KCNJ8,RAMP2,SLC1A2,TMPRSS5,SLAMF1,SLC9C1,SLC34A1,TMEM108,P2RX7,SCARA5,SLC9C2,FCGR3A,FCGR1B,HTR1B,STAB2,CALCRL,HFE,MRC1,KCNJ16 |
| GO:1901700 | response to oxygen | 37 | 1427 | 0,23 | 0,0261 | AQP8,AQP9,P2RX5,KCNJ8,KCNA5,RAMP2,CHRM3,SLC1A2,LHCGR,SLC26A5,GPRC6A,GHRHR,SLC34A1,P2RX7,OPRL1,RET,TNFRSF14,NTRK3,GRIN3A,CHRNB2,HTR1B,GRIN1,KCNB1,AGTR2,IRS4,CNR2,SLC22A6,TNFRSF4,GRIN2A,CSF2RB,CALCRL,ESR1,CASR,KCNC2,MRC1,GRIN2B,PRLR |
| GO:0030320 | cellular monovalent | 3 | 13 | 1,17 | 0,0269 | SLC34A1,SLC12A5,SLC34A3 |
| GO:0043267 | negative regulation | 4 | 30 | 0,94 | 0,0269 | KCNA5,KCNH2,KCNE3,KCNE1L |
| GO:0090276 | regulation of peptide | 10 | 204 | 0,5 | 0,0269 | KCNA5,CACNA1C,HTR2C,GHRHR,SLC2A2,KCNB1,NRXN1,HFE,CASR,KCNC2 |
| GO:0097746 | regulation of blood | 8 | 137 | 0,58 | 0,027 | KCNA5,CHRM3,EDNRA,HTR1A,ADRB3,HTR1B,AGTR2,CASR |
| GO:0002250 | adaptive immune re | 12 | 280 | 0,44 | 0,0281 | SIT1,FCRL4,CD3D,SLAMF1,CD244,CD1E,FCGR1B,LAIR1,LILRA6,CD8A,CRACR2A,TARM1 |
| GO:0019228 | neuronal action pote | 4 | 31 | 0,92 | 0,0293 | SCN11A,KCNMB3,CACNA1I,SCN9A |
| GO:0007157 | heterophilic cell-cell | 5 | 54 | 0,78 | 0,031 | CD226,NCR3,NRXN1,TIGIT,CD200 |
| GO:0055062 | phosphate ion home | 3 | 14 | 1,14 | 0,031 | FGFR4,SLC34A1,SLC34A3 |
| GO:0060134 | prepulse inhibition | 3 | 14 | 1,14 | 0,031 | GRIN3A,GRIN1,NRXN1 |
| GO:1903532 | positive regulation c | 14 | 364 | 0,4 | 0,031 | GALR1,CD34,GHRHR,P2RX7,NPY2R,CD244,CHRNB2,CD2,KCNB1,AGTR2,TNFRSF4,CACNA1I,HFE,CASR |
| GO:0051482 | positive regulation c | 4 | 32 | 0,91 | 0,0315 | HTR2C,F2RL2,C3AR1,OPRL1 |
| GO:0043271 | negative regulation | 8 | 143 | 0,56 | 0,033 | KCNA5,KCNH2,SLC26A5,KCNE3,OPRL1,HTR1B,AGTR2,KCNE1L |
| GO:0015855 | pyrimidine nucleoba | 2 | 3 | 1,64 | 0,0332 | AQP9,SLC28A1 |
| GO:0034155 | regulation of toll-like | 2 | 3 | 1,64 | 0,0332 | LILRA4,TREML4 |
| GO:0046010 | positive regulation c | 2 | 3 | 1,64 | 0,0332 | GHRHR,NPY2R |
| GO:0051952 | regulation of amine | 6 | 82 | 0,68 | 0,0332 | P2RX7,NPY2R,CHRNB2,HTR1B,KCNB1,AGTR2 |
| GO:0098912 | membrane depolari | 2 | 3 | 1,64 | 0,0332 | CACNA1C,CACNB2 |
| GO:0043029 | T cell homeostasis | 4 | 33 | 0,9 | 0,0338 | RIPK3,SIT1,P2RX7,SLC46A2 |
| GO:0035435 | phosphate ion trans | 3 | 15 | 1,11 | 0,0346 | SLC17A1,SLC34A1,SLC34A3 |
| GO:0098719 | sodium ion import a | 3 | 15 | 1,11 | 0,0346 | SLC9C1,SLC34A1,SLC9C2 |
| GO:1904062 | regulation of cation | 12 | 294 | 0,42 | 0,0377 | KCNH2,CACNA1C,SCN2B,KCNE3,CACNB2,OPRL1,KCNE1L,NPSR1,NRXN1,SCN1B,CRACR2A,KCNC2 |
| GO:0002709 | regulation of T cell r | 5 | 58 | 0,75 | 0,0383 | RIPK3,P2RX7,NCR3,PTPRC,HFE |
| GO:0051954 | positive regulation c | 4 | 35 | 0,87 | 0,0401 | P2RX7,NPY2R,CHRNB2,KCNB1 |
| GO:0001910 | regulation of leukoc | 5 | 60 | 0,73 | 0,0435 | RIPK3,CD226,P2RX7,NCR3,PTPRC |
| GO:0006835 | dicarboxylic acid tra | 5 | 60 | 0,73 | 0,0435 | SLC17A6,SLC1A2,SLC26A5,SLC22A6,GRM2 |
| GO:0045761 | regulation of adenyl | 4 | 36 | 0,86 | 0,0435 | LHCGR,GALR1,GABBR1,GRM2 |
| GO:0071242 | cellular response to | 5 | 60 | 0,73 | 0,0435 | CHRM3,GABRB2,SLC34A1,CHRNB2,KCNC2 |
| GO:0009636 | response to toxic su | 16 | 468 | 0,35 | 0,0438 | KCNA5,SCN2B,CHRNB3,SLC23A1,SLC22A8,NTRK3,GRIN3A,CHRNB2,HTR1B,GRIN1,CNR2,GRIN2A,SCN9A,SCN1B,KCNC2,GRIN2B |
| GO:0006952 | defense response | 32 | 1234 | 0,23 | 0,0458 | KCNJ8,TREM1,IL1RL2,HTR2C,MS4A2,LILRA4,CLEC1B,C3AR1,SLAMF1,KLRC4,HTR1A,P2RX7,NPY2R,KIR2DL4,NCR3,TREML4,TNFRSF14,CCRL2,CD244,FCGR1B,AGTR2,CNR2,TNFRSF4,SIRPB1,STAB2,KIR3DL1,SCN9A,CXCR6,PTPRC,HFE,TARM1,MRC1 |
| GO:0007198 | adenylate cyclase-in | 2 | 4 | 1,51 | 0,0458 | HTR1A,HTR1B |
| GO:0008645 | hexose transmembr | 4 | 37 | 0,85 | 0,0458 | SLC5A9,SLC26A5,EDNRA,SLC2A2 |
| GO:0035585 | calcium-mediated si | 2 | 4 | 1,51 | 0,0458 | CACNA1C,P2RX7 |
| GO:0046684 | response to pyrethr | 2 | 4 | 1,51 | 0,0458 | SCN2B,SCN1B |
| GO:0070837 | dehydroascorbic ac | 2 | 4 | 1,51 | 0,0458 | SLC23A1,SLC2A2 |
| GO:0071918 | urea transmembran | 2 | 4 | 1,51 | 0,0458 | AQP9,SLC14A1 |
| GO:0097091 | synaptic vesicle clus | 2 | 4 | 1,51 | 0,0458 | SYNDIG1,NRXN1 |
| GO:0098815 | modulation of excita | 4 | 37 | 0,85 | 0,0458 | TMEM108,NPY2R,GRIN1,NRXN1 |
| GO:1990410 | adrenomedullin rec | 2 | 4 | 1,51 | 0,0458 | RAMP2,CALCRL |
| GO:0060359 | response to ammon | 7 | 122 | 0,57 | 0,0478 | CHRM3,GABRB2,SLC34A1,CHRNB2,HTR1B,GRIN1,KCNC2 |
| GO:0051930 | regulation of sensor | 4 | 38 | 0,83 | 0,049 | SCN11A,NPY2R,OPRL1,GRIN2A |
| GO:0006833 | water transport | 3 | 18 | 1,03 | 0,0495 | AQP8,AQP9,SLC14A1 |
