## Supplementary material for "Early Reduction of SARS-CoV-2 Replication in Bronchial Epithelium by Kinin B_2_ Receptor Antagonism": Supl. Table 19

**Supplemental Table S19.** Membrane-bound cell surface receptor DEGs, upregulated in SARS-CoV-2 versus medium

| ProbeName | p ([SARS-Cov-: | Regulatic | FC ([SARS-( | GeneSymbo | Description |
|---|---|---|---|---|---|
| A_23_P95536 | 0,032593332 | up | 2,0564296 | ADAM29 | Homo sapiens ADAM metallopeptidase domain 29 (ADAM29), transcript variant 1, mRNA [NM_014269] |
| A_33_P3360296 | 0,04713104 | up | 2,1182783 | ANTXRL | Homo sapiens anthrax toxin receptor-like (ANTXRL), mRNA [NM_001278688] |
| A_33_P3415012 | 0,003580882 | up | 2,2150555 | KCNA6 | Homo sapiens potassium channel, voltage gated shaker related subfamily A, member 6 (KCNA6), mRNA [NM_002235] |
| A_33_P3270104 | 0,036002126 | up | 1,6818469 | FAM26D | Homo sapiens family with sequence similarity 26, member D (FAM26D), transcript variant 1, mRNA [NM_001256887] |
| A_23_P38634 | 0,034794565 | up | 1,6641743 | OR1G1 | Homo sapiens olfactory receptor, family 1, subfamily G, member 1 (OR1G1), mRNA [NM_003555] |
| A_33_P3406836 | 0,011732406 | up | 1,7898222 | HTR1F | Homo sapiens 5-hydroxytryptamine (serotonin) receptor 1F, G protein-coupled (HTR1F), mRNA [NM_000866] |
| A_33_P3416757 | 0,047634322 | up | 1,5079808 | PRLR | Homo sapiens prolactin receptor (PRLR), transcript variant 2, mRNA [NM_001204315] |
| A_24_P945096 | 0,03653614 | up | 1,7702624 | CACNA1I | Homo sapiens calcium channel, voltage-dependent, T type, alpha 1I subunit (CACNA1I), transcript variant 1, mRNA [NM_021096] |
| A_33_P3396244 | 0,035009935 | up | 1,990737 | NPY2R | Homo sapiens neuropeptide Y receptor Y2 (NPY2R), mRNA [NM_000910] |
| A_33_P3340014 | 0,010722341 | up | 2,0883427 | TRO | Homo sapiens trophinin (TRO), transcript variant 3, mRNA [NM_016157] |
| A_21_P0010873 | 0,043899916 | up | 1,5941203 | ANTXRLP1 | Homo sapiens anthrax toxin receptor-like pseudogene 1 (ANTXRLP1), transcript variant 2, non-coding RNA [NR_103828] |
| A_24_P170439 | 0,015414147 | up | 1,8931247 | TREML3P | Homo sapiens triggering receptor expressed on myeloid cells-like 3, pseudogene (TREML3P), non-coding RNA [NR_027256] |
| A_33_P3355856 | 0,026495013 | up | 1,7257988 | | olfactory receptor, family 51, subfamily J, member 1 (gene/pseudogene) [Source:HGNC Symbol;Acc:HGNC:14856] [ENST00000332043] |
| A_33_P3413558 | 0,008619091 | up | 2,6380665 | CD226 | CD226 molecule [Source:HGNC Symbol;Acc:HGNC:16961] [ENST00000280200] |
| A_33_P3241021 | 0,02955563 | up | 2,0316834 | CD69 | CD69 molecule [Source:HGNC Symbol;Acc:HGNC:1694] [ENST00000416624] |
| A_33_P3326271 | 0,011921781 | up | 1,9606916 | OR13C2 | Homo sapiens olfactory receptor, family 13, subfamily C, member 2 (OR13C2), mRNA [NM_001004481] |
| A_33_P3280646 | 0,048862424 | up | 1,7165905 | CASR | Homo sapiens calcium-sensing receptor (CASR), transcript variant 1, mRNA [NM_001178065] |
| A_23_P337867 | 0,041624285 | up | 1,6760936 | TAS2R41 | Homo sapiens taste receptor, type 2, member 41 (TAS2R41), mRNA [NM_176883] |
| A_24_P319113 | 0,031806596 | up | 1,6776376 | P2RX7 | Homo sapiens purinergic receptor P2X, ligand-gated ion channel, 7 (P2RX7), transcript variant 1, mRNA [NM_002562] |
| A_23_P43369 | 0,022677582 | up | 1,7351198 | SIT1 | Homo sapiens signaling threshold regulating transmembrane adaptor 1 (SIT1), mRNA [NM_014450] |
| A_33_P3400239 | 0,025245482 | up | 1,9221019 | SYNDIG1 | PREDICTED: Homo sapiens synapse differentiation inducing 1 (SYNDIG1), transcript variant X1, mRNA [XM_006723626] |
| A_33_P3363725 | 0,03290714 | up | 1,9861467 | OR52N5 | Homo sapiens olfactory receptor, family 52, subfamily N, member 5 (OR52N5), mRNA [NM_001001922] |
| A_33_P3211488 | 0,048390158 | up | 1,5444185 | OR6K3 | Homo sapiens olfactory receptor, family 6, subfamily K, member 3 (OR6K3), mRNA [NM_001005327] |
| A_24_P941896 | 0,04121485 | up | 1,5743759 | GRID1 | Homo sapiens glutamate receptor, ionotropic, delta 1 (GRID1), mRNA [NM_017551] |
| A_33_P3218980 | 0,028221466 | up | 1,6104319 | ENTPD1 | Homo sapiens ectonucleoside triphosphate diphosphohydrolase 1 (ENTPD1), transcript variant 1, mRNA [NM_001776] |
| A_23_P92650 | 1,35E-04 | up | 3,6780155 | SLC25A2 | Homo sapiens solute carrier family 25 (mitochondrial carrier; ornithine transporter) member 2 (SLC25A2), mRNA [NM_031947] |
| A_33_P3230189 | 0,009609546 | up | 1,6858562 | SLITRK6 | Homo sapiens SLIT and NTRK-like family, member 6 (SLITRK6), mRNA [NM_032229] |
| A_32_P183765 | 0,02203102 | up | 2,06349 | ERBB4 | Homo sapiens erb-b2 receptor tyrosine kinase 4 (ERBB4), transcript variant JM-a/CVT-1, mRNA [NM_005235] |

| | | | | | |
|---|---|---|---|---|---|
| A_33_P3400217 | 0,018383147 | up | 1,6902646 | SLC4A1 | Homo sapiens solute carrier family 4 (anion exchanger), member 1 (Diego blood group) (SLC4A1), mRNA [NM_000342] |
| A_33_P3210399 | 0,036284268 | up | 1,846673 | SLC14A1 | Homo sapiens solute carrier family 14 (urea transporter), member 1 (Kidd blood group) (SLC14A1), transcript variant 4, mRNA [NM_001146037] |
| A_23_P318938 | 0,002957786 | up | 2,2908568 | TMEM52B | Homo sapiens transmembrane protein 52B (TMEM52B), transcript variant 1, mRNA [NM_153022] |
| A_33_P3335511 | 0,008867103 | up | 1,8473281 | FCRL5 | Homo sapiens Fc receptor-like 5 (FCRL5), transcript variant 2, mRNA [NM_001195388] |
| A_23_P319423 | 0,040168095 | up | 1,9868454 | KCNK5 | Homo sapiens potassium channel, two pore domain subfamily K, member 5 (KCNK5), mRNA [NM_003740] |
| A_32_P489986 | 0,032472506 | up | 1,8414146 | TMEM232 | Homo sapiens transmembrane protein 232 (TMEM232), mRNA [NM_001039763] |
| A_33_P3261957 | 0,011492129 | up | 2,4471688 | CALCRL | Homo sapiens calcitonin receptor-like (CALCRL), transcript variant 1, mRNA [NM_005795] |
