## Supplementary material for "Early Reduction of SARS-CoV-2 Replication in Bronchial Epithelium by Kinin B_2_ Receptor Antagonism": Supl. Table 20

**Supplemental Table S20.** Cut set: Membrane-bound cell surface receptor DEGs, upregulated in SARS-CoV-2 versus medium and downregulated in SARS-CoV-2 + B2R antagonist versus SARS-CoV-2

| ProbeName | p ([SARS-Co\ | Regulation ([S | FC ([SARS-C | GeneSymbol | Description |
|---|---|---|---|---|---|
| A_23_P9553 | 0,01830276 | down | -2,1320918 | ADAM29 | Homo sapiens ADAM metallopeptidase domain 29 (ADAM29), transcript variant 1, mRNA [NM_014269] |
| A_33_P3360 | 0,03580659 | down | -1,9404445 | ANTXRL | Homo sapiens anthrax toxin receptor-like (ANTXRL), mRNA [NM_001278688] |
| A_33_P3415 | 0,01891622 | down | -2,1078854 | KCNA6 | Homo sapiens potassium channel, voltage gated shaker related subfamily A, member 6 (KCNA6), mRNA [NM_002235] |
| A_33_P3270 | 0,02106279 | down | -1,9156722 | FAM26D | Homo sapiens family with sequence similarity 26, member D (FAM26D), transcript variant 1, mRNA [NM_001256887] |
| A_23_P3863 | 0,04309718 | down | -1,5717353 | OR1G1 | Homo sapiens olfactory receptor, family 1, subfamily G, member 1 (OR1G1), mRNA [NM_003555] |
| A_33_P3406 | 0,00964598 | down | -1,7121874 | HTR1F | Homo sapiens 5-hydroxytryptamine (serotonin) receptor 1F, G protein-coupled (HTR1F), mRNA [NM_000866] |
| A_33_P3416 | 0,03221973 | down | -1,5744605 | PRLR | Homo sapiens prolactin receptor (PRLR), transcript variant 2, mRNA [NM_001204315] |
| A_24_P9450 | 0,04818654 | down | -1,8050053 | CACNA1I | Homo sapiens calcium channel, voltage-dependent, T type, alpha 1I subunit (CACNA1I), transcript variant 1, mRNA [NM_021096] |
| A_33_P3396 | 0,03147338 | down | -1,7422631 | NPY2R | Homo sapiens neuropeptide Y receptor Y2 (NPY2R), mRNA [NM_000910] |
| A_33_P3340 | 0,00952162 | down | -1,9314481 | TRO | Homo sapiens trophinin (TRO), transcript variant 3, mRNA [NM_016157] |
| A_21_P0010 | 0,01194135 | down | -1,8027905 | ANTXRLP1 | Homo sapiens anthrax toxin receptor-like pseudogene 1 (ANTXRLP1), transcript variant 2, non-coding RNA [NR_103828] |
| A_24_P1704 | 0,02440206 | down | -1,7213168 | TREML3P | Homo sapiens triggering receptor expressed on myeloid cells-like 3, pseudogene (TREML3P), non-coding RNA [NR_027256] |
| A_33_P3355 | 0,00441775 | down | -1,965058 | | olfactory receptor, family 51, subfamily J, member 1 (gene/pseudogene) [Source:HGNC Symbol;Acc:HGNC:14856] [ENST00000332043] |
| A_33_P3413 | 0,03426481 | down | -2,0173213 | CD226 | CD226 molecule [Source:HGNC Symbol;Acc:HGNC:16961] [ENST00000280200] |
| A_33_P3241 | 0,01925859 | down | -2,1987698 | CD69 | CD69 molecule [Source:HGNC Symbol;Acc:HGNC:1694] [ENST00000416624] |
| A_33_P3326 | 0,01507 | down | -2,1182187 | OR13C2 | Homo sapiens olfactory receptor, family 13, subfamily C, member 2 (OR13C2), mRNA [NM_001004481] |
| A_33_P3280 | 0,00661566 | down | -2,2763705 | CASR | Homo sapiens calcium-sensing receptor (CASR), transcript variant 1, mRNA [NM_001178065] |
| A_23_P3378 | 0,01912973 | down | -1,8654202 | TAS2R41 | Homo sapiens taste receptor, type 2, member 41 (TAS2R41), mRNA [NM_176883] |
| A_24_P3191 | 0,01615628 | down | -1,7449347 | P2RX7 | Homo sapiens purinergic receptor P2X, ligand-gated ion channel, 7 (P2RX7), transcript variant 1, mRNA [NM_002562] |
| A_23_P4336 | 0,00974178 | down | -1,8951069 | SIT1 | Homo sapiens signaling threshold regulating transmembrane adaptor 1 (SIT1), mRNA [NM_014450] |
| A_33_P3400 | 0,01600916 | down | -2,0678427 | SYNDIG1 | PREDICTED: Homo sapiens synapse differentiation inducing 1 (SYNDIG1), transcript variant X1, mRNA [XM_006723626] |
| A_33_P3363 | 0,00646022 | down | -1,9825865 | OR52N5 | Homo sapiens olfactory receptor, family 52, subfamily N, member 5 (OR52N5), mRNA [NM_001001922] |
| A_33_P3211 | 0,02943458 | down | -1,640576 | OR6K3 | Homo sapiens olfactory receptor, family 6, subfamily K, member 3 (OR6K3), mRNA [NM_001005327] |
| A_24_P9418 | 0,01164032 | down | -1,7608724 | GRID1 | Homo sapiens glutamate receptor, ionotropic, delta 1 (GRID1), mRNA [NM_017551] |
| A_33_P3218 | 0,00842023 | down | -1,7830722 | ENTPD1 | Homo sapiens ectonucleoside triphosphate diphosphohydrolase 1 (ENTPD1), transcript variant 1, mRNA [NM_001776] |
| A_23_P9265 | 0,00807711 | down | -2,16979 | SLC25A2 | Homo sapiens solute carrier family 25 (mitochondrial carrier; ornithine transporter) member 2 (SLC25A2), mRNA [NM_031947] |
| A_33_P3230 | 0,0163531 | down | -1,5824893 | SLITRK6 | Homo sapiens SLIT and NTRK-like family, member 6 (SLITRK6), mRNA [NM_032229] |
| A_32_P1837 | 0,04826975 | down | -2,016431 | ERBB4 | Homo sapiens erb-b2 receptor tyrosine kinase 4 (ERBB4), transcript variant JM-a/CVT-1, mRNA [NM_005235] |
| A_33_P3400 | 0,02282586 | down | -2,310932 | SLC4A1 | Homo sapiens solute carrier family 4 (anion exchanger), member 1 (Diego blood group) (SLC4A1), mRNA [NM_000342] |
| A_33_P3210 | 0,03803759 | down | -1,8543372 | SLC14A1 | Homo sapiens solute carrier family 14 (urea transporter), member 1 (Kidd blood group) (SLC14A1), transcript variant 4, mRNA [NM_001146037] |
| A_23_P3189 | 0,02817906 | down | -2,0037775 | TMEM52B | Homo sapiens transmembrane protein 52B (TMEM52B), transcript variant 1, mRNA [NM_153022] |
| A_33_P3335 | 0,02132568 | down | -1,8054705 | FCRL5 | Homo sapiens Fc receptor-like 5 (FCRL5), transcript variant 2, mRNA [NM_001195388] |
| A_23_P3194 | 0,00103116 | down | -3,493477 | KCNK5 | Homo sapiens potassium channel, two pore domain subfamily K, member 5 (KCNK5), mRNA [NM_003740] |
| A_32_P4899 | 0,02345983 | down | -2,1372492 | TMEM232 | Homo sapiens transmembrane protein 232 (TMEM232), mRNA [NM_001039763] |
| A_33_P3261 | 0,04173563 | down | -1,9484638 | CALCRL | Homo sapiens calcitonin receptor-like (CALCRL), transcript variant 1, mRNA [NM_005795] |
