## Supplementary material for "Early Reduction of SARS-CoV-2 Replication in Bronchial Epithelium by Kinin B_2_ Receptor Antagonism": Supl. Table 21

**Supplemental Table S21.** Reagents

| Method | Reagent / Resource | Company | Identifier |
|---|---|---|---|
| Cell culture | BEGM Bullet kit Medium | Lonza | CC-3170 |
| Cell culture | NHBE-Bronchial Epi Cells | LONZA | CC-2540 |
| Cell culture | Hexa-Peptide von Adam (Tyr-Leu-Tyr....) | Thermo | PEP98UNMOD |
| Cell culture | HOE 140 (B2R antagonist) | SigmaAldrich | H157-250ug |
| Cell culture | 100mg Hydrocortison | sigma aldrich | H0396-100mg |
| Cell culture | antibiotic antimycotic | Thermo | 15240062 |
| ELISA | human ACE2 ELISA Kit | abcam | ab23564g |
| ELISA | Coomassie Plus Bradford Assay Kit | thermo fisher | 23236 |
| RNA extraction | Rneasy Micro Kit (50) | Qiagen | 74004 |
| RNA extraction | QIAshredder (250) | Qiagen | 79656 |
| RNA extraction | Rnase-Free Dnase Set | Qiagen | 79256 |
| Bioanalyzer | agilent rna 6000 nano reagent | agilent | 5067-1512 |
| Microarray | Gene Expression Hybridisierungskit | Agilent | 5188-5242 |
| Microarray | Genexpression Wasch-Paket | Agilent | 5188-5327 |
| Microarray | Sure Print G3 Human GE V2 8x60K Kit | Agilent | G4851 B |
| Microarray | LowInput QuickAmp Labeling Kit One-Color | Agilent | 5190-2305 |
| Microarray | RNA Spike In Kit-One Color | Agilent | 5188-5282 |
| Microarray | Gasketslides | Agilent | G2534-60015 |
| Microarray | Ethanol for molecular biology | vwr/Merck | 1.085.430.250 |
| cDNA Transcription | high capacity cDNA kit | life technologies | 4368814 |
| PCR | FS universal SYBR Green Master Mix | Sigma | 4913850001 |
| PCR | *Human TMPRSS2* fwd AGAGTGCGACTCCTCAGGTA | Metabion | |
| PCR | *Human TMPRSS2* rev AGGCGAACACACCGATTCTC | Metabion | |
| PCR | *Human ACE2* fwd ATGAAGGCCCTCTGCACAAA | Metabion | |
| PCR | *Human ACE2* rev CCAAGCCTCAGCATATTGAACAG | Metabion | |
| PCR | *Human ACTIN* fwd ATTGCCGACAGGATGCAGAA | Metabion | |
| PCR | *Human ACTIN* rev TCTTCATTGTGCTGGGTGCC | Metabion | |
| PCR | *Human HPRT* fwd TGACACTGGCAAAACAATGCA | Metabion | |
| PCR | *Human HPRT* rev GGTCCTTTTCACCAGCAAGCT | Metabion | |
| PCR | *Human 18S* fwd<br>Hs_RRN18S_1_SG QuantiTect Primer Assay | Qiagen | 249900<br>QT00199367 |
| PCR | *Human 18S* rev<br>Hs_RRN18S_1_SG QuantiTect Primer Assay | Qiagen | 249900<br>QT00199367 |
| PCR | *SARS-CoV-2 RdRP* fwd<br>CGTCTGCGGTATGTGGAAAG | Metabion | |
| PCR | *SARS-CoV-2 RdRP* rev<br>TAAGACGGGCTGCACTTACA | Metabion | |
| PCR | *SARS-CoV-2 N1* fwd: GACCCCAAAATCAGCGAAAT | Metabion | |
| PCR | *SARS-CoV-2 N1* rev: TCTGGTTACTGCCAGTTGAATCT[illegible] | Metabion | |
| PCR | *SARS-CoV-2 N1* probe: 5'-FAM-ACC CCG CAT TAC GT[illegible] | Metabion | |
